# Longitudinal Assessment of DNA Repair Signature Trajectory in Prodromal versus Established Parkinson’s Disease

**DOI:** 10.1101/2025.03.19.25324249

**Authors:** Danish Anwer, Nicola Pietro Montaldo, Elva Maria Novoa-del-Toro, Diana Domanska, Hilde Loge Nilsen, Annikka Polster

## Abstract

Parkinson’s disease (PD) is a progressive neurodegenerative disorder characterized by motor and non-motor symptoms. DNA repair dysfunction and integrated stress response (ISR) dysregulation have been suggested to be relevant in PD pathophysiology, but their role during the prodromal phase, before motor symptoms manifest, remains unclear. In this study, we analyzed longitudinal blood transcriptomic data from the Parkinson’s Progression Markers Initiative (PPMI) to investigate how DNA repair and ISR gene expression differ across healthy individuals, prodromal PD patients, and individuals with established PD. Using logistic regression classifiers, a type of supervised machine learning model, we found that DNA repair and ISR gene expression effectively distinguished prodromal PD from healthy individuals, with classification accuracy increasing over time and peaking at later prodromal stages. In contrast, these pathways failed to differentiate established PD from healthy controls, suggesting that DNA repair and ISR dysregulation play a more distinct role early in disease progression. Gene expression variability was high in prodromal PD at baseline but reduced over time, indicating a convergence in gene expression patterns as the disease advances. Notably, 50% of DNA repair genes and 74% of ISR genes exhibited nonlinear expression patterns, with an initial increase followed by a decline. This suggests a transient adaptive response that fades out as PD progresses. Feature importance analysis identified key genes, including ERCC6, PRIMPOL, NEIL2, and NTHL1, as important predictors of prodromal PD. These findings suggest that DNA repair and ISR dysregulation contribute to early-stage PD pathology and may serve as biomarkers for early disease detection and possible treatment avenues during the prodromal phase.

This study highlights the role of DNA repair and ISR pathways in the prodromal phase of PD, demonstrating their potential as early molecular indicators of disease onset. Future research should validate these findings in larger cohorts and explore their relevance for early diagnostics and intervention strategies.

## Introduction

Parkinson’s disease (PD) is a progressive neurodegenerative disorder classically defined by motor symptoms (bradykinesia, tremor, rigidity, postural instability), which emerge only after significant neurodegeneration has occurred^1,2^. Years before this motor symptoms onset, patients often experience a prodromal phase marked by subtle non-motor features such as REM sleep behavior disorder, olfactory loss, constipation, depression, and anxiety.

This temporal progression from prodromal to overt PD suggests a window of opportunity to identify disease-driving molecular processes before extensive dopaminergic (DA) neuron loss. Understanding the molecular changes and pathogenic mechanisms in prodromal PD is therefore critical, as it may allow for diagnosis before significant neuronal loss occurs, when neuroprotective interventions are likely most effective, as well as identify novel treatment targets.

Among the emerging mechanisms implicated in PD pathogenesis is genomic DNA damage and repair dysfunction. Growing evidence suggests that the accumulation of DNA lesions in neurons may play an active role in driving disease onset and progression ^3,4^. Dopaminergic neurons are metabolically active and exposed to high levels of endogenous reactive oxygen species (ROS) as by-products of dopamine metabolism and mitochondrial respiration^5,6^. PD is associated with impaired mitochondrial electron transport and mitophagy, which further exacerbates DNA damage. Over time, this chronic oxidative DNA damage may overwhelm the capacity of the repair pathways^7^.

This results in the accumulation of both single-strand and double-strand breaks (SSBs and DSBs respectively), which are hazardous. In addition to these breaks, a plethora of base lesions can accumulate, each having distinct effects on transcription. In mitochondrial DNA (mtDNA), which has limited DNA repair capacity, the relevance of these might be more important. PD patients, in fact, exhibit a higher burden of somatic mtDNA mutations in striatal nigral neurons compared to age-matched controls^8^.

Cells counteract oxidative DNA damage primarily via the base excision repair (BER) pathway, which recognizes, and excises oxidized bases^9^. The BER pathway is especially important in mitochondria, where it is the only fully functional DNA repair mechanism^10^. However, paradoxically, BER itself can become a source of stress if dysregulated. Our recent study using a *Caenorhabditis elegans* (*C. elegans)* PD model demonstrated that BER activity initiated by the DNA glycosylase NTH-1 leads to accumulation of repair intermediates (single-strand breaks) that trigger neurotoxicity during physiological aging. In this study, reducing NTH-1 levels by knocking out or siRNA mediated knockdown, was actually neuroprotective, preventing neurodegeneration^11^. This highlights that while DNA repair pathways are essential for cell survival, overactivation or lack of coordination in handling toxic intermediates, such as SSBs and DSBs, can cause harm by depleting resources or generating toxic DNA break ends. Consistently, genetic studies in humans have begun to link subtle variants in DNA repair genes to PD risk. For example, rare variants in the BER glycosylase NEIL2 (which, like NTH1, fixes oxidative base lesions) were found enriched in PD patients compared to controls^11^. Furthermore Sanders et al. demonstrated that genetic variants of the BER genes increase risk of PD in combination with pesticides known to affect mitochondrial function ^12^. Taken together, these findings indicate that oxidative DNA damage and an impaired or maladaptive repair response are key contributors to PD pathogenesis from its earliest stages.

A major challenge in studying DNA repair in PD is the inaccessible nature of the affected tissue, as one cannot easily monitor DNA damage or repair activity in relevant cell types in living patients. This has spurred interest in peripheral biomarkers that might reflect central neurodegenerative processes. Therefore, blood-based transcriptomics are of high interest in current PD research. However, previous transcriptomic studies have primarily focused on differentiating PD patients from controls without addressing that in PD patients substantial cell death of up to 80% of relevant cell types had already occurred^2^. However, how these gene expression patterns change during the prodromal phase is largely unknown.

Given the evidence that DNA damage and repair defects are involved early in PD, a logical hypothesis is that prodromal PD patients likely exhibit distinct changes in DNA repair pathways before clinical manifestation. Detecting such changes in peripheral blood could provide a non-invasive marker of ongoing neurodegenerative processes. Additionally, comparing prodromal versus established PD could reveal whether there is an early, perhaps compensatory upregulation of repair mechanisms that later becomes dysfunctional as the disease advances.

By utilizing longitudinal transcriptomics from healthy and prodromal individuals as well as diagnosed PD patients and applying machine learning, this study thus aims to explore the dynamic regulation of DNA repair gene expression in these groups and reveal patterns within curated pathways.

## Materials and methods

### Overview

To investigate how gene expression in DNA repair and stress response pathways changes across Parkinson’s disease stages, we analyzed longitudinal transcriptomic data from blood samples collected in the PPMI cohort. We focused on biologically curated gene sets and used logistic regression, a supervised machine learning method, to assess whether their expression patterns could distinguish between diagnostic groups (healthy, prodromal PD, and established PD) over time. We further evaluated how gene importance and expression variability changed throughout the disease course. This analysis used data openly available from PPMI.

Minor language revisions were supported by ChatGPT (OpenAI, GPT-4), all content was authored, reviewed, and approved by the authors.

### Data

To examine longitudinal gene expression changes across PD progression, transcriptomic data from PPMI^13^ were analyzed across three participant groups: healthy individuals, prodromal PD, and idiopathic PD at four time points: Baseline (BL), after 12 months (M12), after 24 months (M24) and after 36 months (M36). Whole transcriptome sequencing was performed on ribo- and globin-depleted RNA from PaxGene Tubes at HudsonAlpha using Illumina NovaSeq6000. RNA was processed via directional cDNA synthesis and NEB/Kapa library prep, aligned to GRCh38p12 using STAR. Data of the IR3 sequencing version was downloaded as TPM (generated using the Salmon method^14^) from https://ida.loni.usc.edu/.

### Gene sets

We selected five gene sets representing distinct biological processes: 1) mitochondrial DNA damage repair (mtDNArep), 2) general DNA damage repair (DNArep), 3) the integrated stress response (ISR), 4) core Parkinson’s disease-related genes (PD-core), and 5) Parkinson’s disease-associated genes (PD-assoc). The DNArep, mtDNArep, and ISR gene sets were manually curated by HLN and MN, domain experts in DNA repair mechanisms. The PD-core set was compiled through manual curation from publicly available sources, including informational material from the Michael J. Fox Foundation. The PD-assoc list was derived from a recent meta-analysis^15^. The lists are presented in supplementary table S1.

### Data analyses

#### Differential gene expression

Differential gene expression analyses at baseline timepoint were conducted using PyDESeq2^30^, a Python implementation of the DESeq2 methodology. Three comparisons were included, healthy vs established PD, healthy vs prodromal PD and prodromal PD vs established PD. Differential expression was modeled to assess differences between diagnostic groups while accounting for plate, age and sex as potential confounders.

Raw gene-level count data was filtered to include only protein-coding genes by parsing the GTF annotation file (Homo_sapiens.GRCh38.104.gtf) to extract entries where the gene biotype was “protein_coding”. Lowly expressed genes were further removed based on a counts-per-million (CPM) threshold: genes were kept if they had CPM ≥ 1 in at least half of the samples in a given comparison group. Normalization was handled internally via size factor estimation to account for differences in library size and sequencing depth. Significance testing was performed using Wald tests, and p-values were adjusted for multiple testing using the Benjamini-Hochberg method to control the false discovery rate. Genes with adjusted p-values < 0.05 were considered significantly differentially expressed. Both raw and adjusted p-values are reported, and results are visualized through volcano plots.

#### Classifier analysis

To evaluate the differential activity of biological processes represented by gene sets across healthy, prodromal, and established PD states, we performed classification analyses using logistic regression. These logistic regression classifiers are supervised machine learning models that estimate the probability of group membership based on predictor variables such as gene expression. These models are well suited to high-dimensional transcriptomic data and were trained to distinguish between healthy individuals, prodromal PD patients, and individuals with established PD. Performance was evaluated using classification accuracies on bootstrap validation. Classification accuracy is a measure of how well a model correctly identifies or predicts group membership. It is calculated as the proportion of correct predictions out of all predictions made. For example, if a model classifies 80 out of 100 samples correctly, the classification accuracy is 80 percent.

We conducted this analysis for the respective pairwise comparisons (healthy vs. prodromal and prodromal vs. PD) across four time points: BL, M12, M24, M36. To ensure the reliability and robustness of our findings, we performed bootstrap validation by running 1,000 iterations of the logistic regression on randomly split training and test sets for each gene set. In each iteration of the classifier, we randomly selected an equal number of subjects from each group to prevent class imbalance and minimize bias in the results. The data were stratified into training (80%) and testing (20%) sets, with the logistic regression model trained on the training set using Scikit-Learn with default hyperparameters and evaluated on the test set. Classification accuracy was recorded for each iteration and averaged across all iterations to obtain a stable and reliable measure of model performance.

To assess the importance and relative contribution of individual genes, we extracted the genes’ coefficients from the logistic regression model at each iteration. To enhance the robustness of these findings, we calculated the average coefficient value across the 1,000 iterations of each classifier. These average coefficients were then ranked based on their absolute values, with larger absolute values indicating higher importance of the gene in distinguishing between the groups.

To capture shifts in gene importance over time and visualize trends in disease progression, we analyzed the temporal dynamics of gene importance rankings across different time points.

Code is provided at https://github.com/Polster-lab/Longitudinal-DNA-Repair-Parkinson

## Results

### Demographics and Clinical Characteristics

Participants included in this study were obtained from the Parkinson’s Progression Marker Initiative^13^. The cohort consisted of healthy controls (at baseline n=192, 35.4% female), prodromal PD subjects (at baseline n=63, 20.6% female), and established PD patients (at baseline n=412, 34.5% female). Detailed age distributions across the four time points are shown in Figure 1.

**Figure 1:**
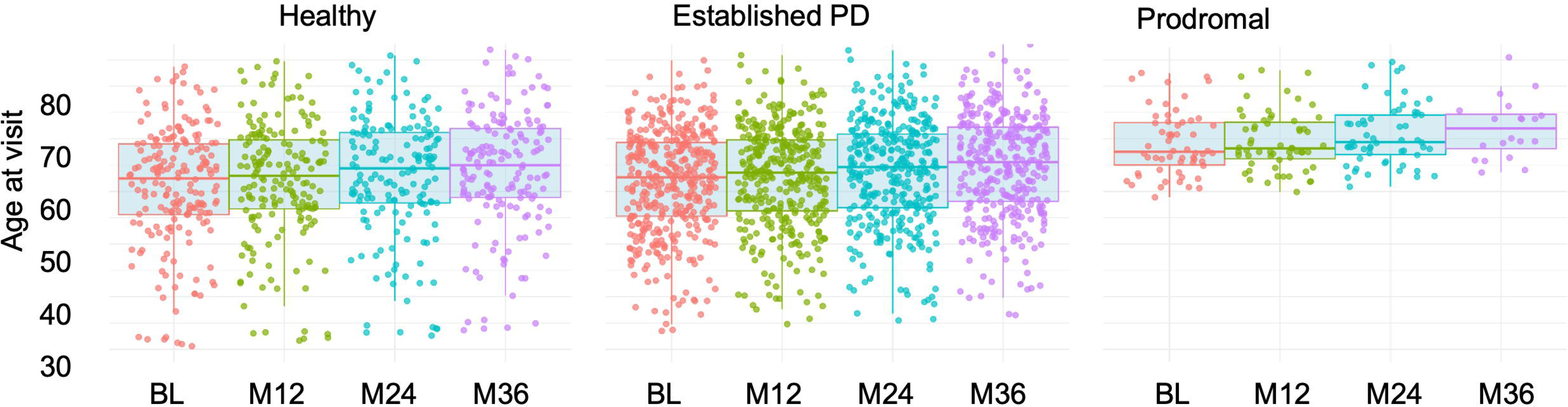
Age distribution across time points by diagnosis. This figure presents the distribution of participant ages across different time points for the three diagnostic groups: Healthy, established Parkinson’s Disease (PD), and prodromal PD. Boxplots depict the median, interquartile range (IQR), and outliers for each group at each visit. Individual data points are overlaid, with colors indicating specific time points: baseline (BL), 12 months (M12), 24 months (M24), and 36 months (M36). The numbers above each boxplot denote the sample size at each visit.

### Differential expression analysis

To orient our gene set–based analyses, we first examined global differential expression patterns across diagnostic groups at baseline (Figure 2). The number of strongly differentially expressed genes increased progressively across comparisons: from no significant changes between healthy and prodromal PD, to extensive changes in prodromal vs. established PD, and healthy vs. established PD. Despite these broad shifts, genes from the mtDNArep, DNArep, and ISR pathways were largely absent from the top 50 differentially expressed genes in each comparison. Only a gene appeared, LITAF(ISR) in healthy vs. established PD.

**Figure 2:**
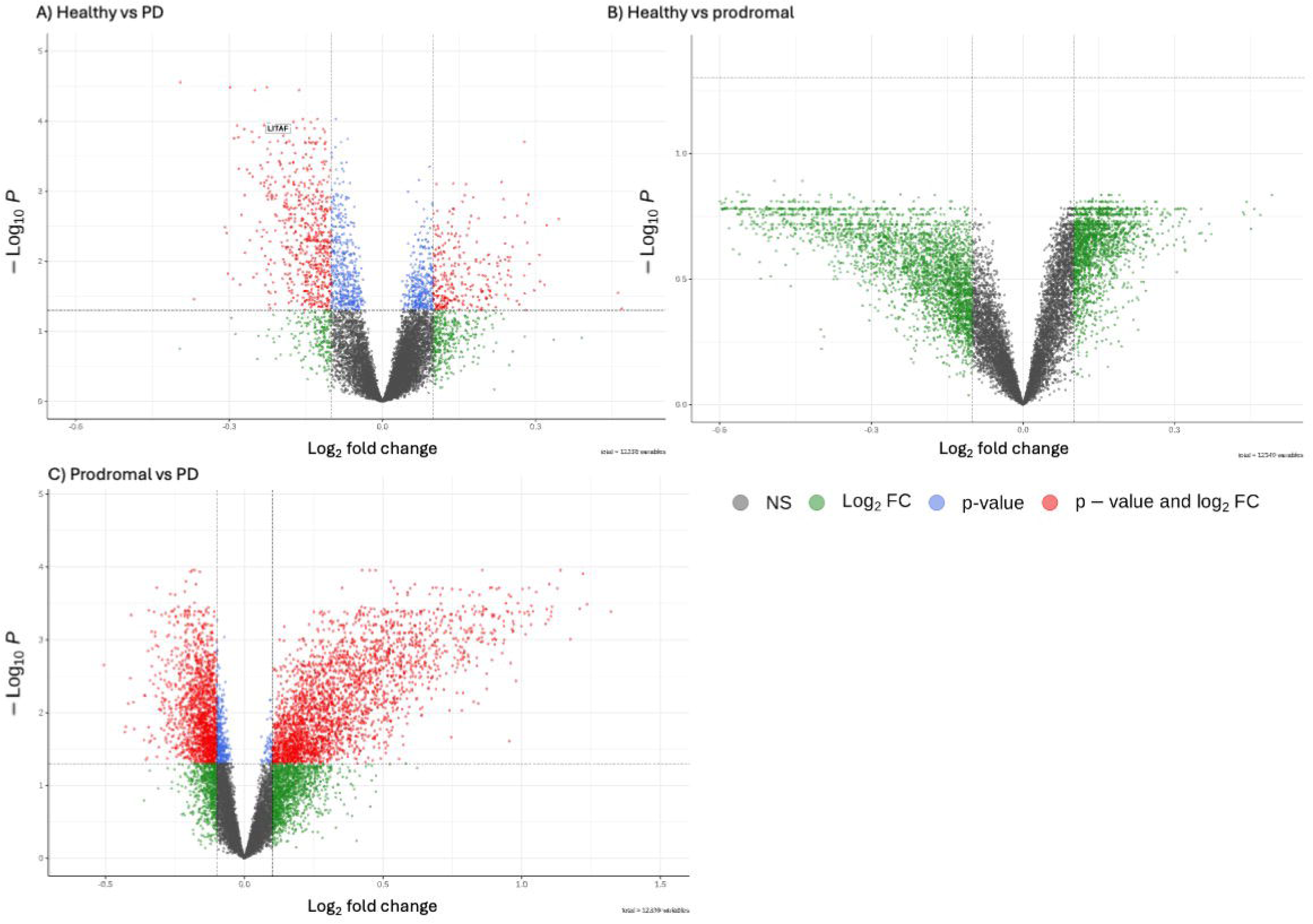
Differential gene expression analyses at baseline Volcano plots illustrate differential gene expression across diagnostic groups at the baseline time point. The x-axis represents log_₂_ fold change, and the y-axis represents –log_₁₀_(p-value), highlighting both the magnitude and statistical significance of expression changes. Each dot corresponds to a gene: red indicates genes meeting both significance and fold change thresholds, blue indicates significance-only, green indicates fold change-only, and gray denotes non-significant genes. A) Comparison between healthy controls and individuals with established Parkinson’s disease (PD) revealed widespread differential expression, suggesting substantial transcriptional alterations associated with disease state. Only one gene (*LITAF*) of the selected gene sets was differentially expressed in this comparison. B) In the comparison between healthy controls and prodromal PD, no genes were differentially expressed, reflecting a subtler transcriptomic shift in early disease stages. C) The contrast between prodromal and established PD showed extensive differential expression, indicating pronounced molecular changes. The relative lack of significant gene expression changes between healthy controls and prodromal PD (Panel B), contrasted with the marked differences observed in established PD (Panels A and C), supports a progressive pattern of transcriptomic dysregulation across the clinical course of Parkinson’s disease.

This limited representation of curated pathway genes among the most strongly differentially expressed genes suggests that their role in PD may be more subtle and relevant later in the disease progression, rather than marked by initial large expression shifts in individual genes. These findings confirmed the need conduct longitudinal analyses to detect coordinated and potentially progressive changes in expression. Full differential expression results are provided in Supplementary Table S2.

### Classification of Disease Stages Using Gene Expression in DNA Repair and ISR Pathways

#### Healthy vs established PD

We evaluated whether gene expression patterns in mitochondrial DNA repair, nuclear DNA repair, or the integrated stress response could distinguish individuals with Parkinson’s disease from healthy controls. Across all time points, classification accuracy ranged from 50 to 64 percent (Figure 3A), indicating performance only slightly above random guessing. These results suggest that peripheral blood expression of these pathways does not provide a consistent or strong enough signal to reliably differentiate individuals with established Parkinson’s disease from healthy individuals. Accuracy did not show any consistent trend over time, further supporting the conclusion that these transcriptional profiles remain relatively stable once the disease is clinically diagnosed and are not sufficient for robust classification.

**Figure 3.**
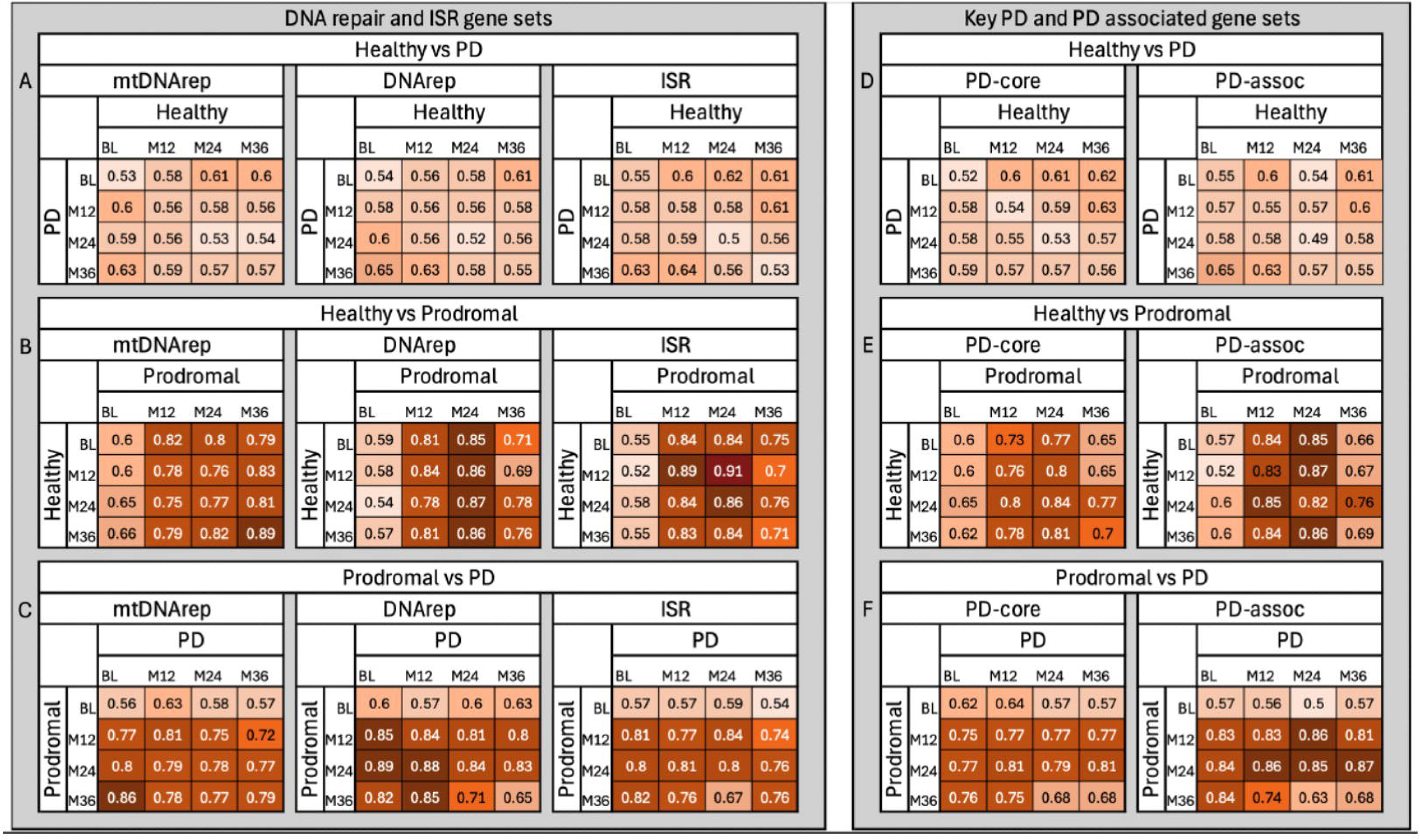
Classification performance of DNA repair, stress response, and PD-related gene sets across Parkinson’s disease stages. This figure illustrates how well predefined gene sets distinguish between healthy individuals, individuals in the prodromal stage of PD, and individuals with established PD, based on gene expression in peripheral blood. We used logistic regression models to test whether expression patterns from specific gene sets could accurately classify individuals into their correct diagnostic group. Panels A–C show results for gene sets involved in mitochondrial DNA repair (mtDNArep), nuclear DNA repair (DNArep), and the integrated stress response (ISR). Panels D–F show results for core Parkinson’s disease-related genes (PD-core) and a broader set of PD-associated genes (PD-assoc) identified from genetic studies. Each heatmap displays classification accuracy, defined as the proportion of individuals correctly identified by the model. For example, an accuracy of 0.80 means that 80% of individuals were assigned to the correct group (e.g., healthy, prodromal, or PD) based on their gene expression profile. Accuracy was assessed at four time points: baseline (BL), 12 months (M12), 24 months (M24), and 36 months (M36). Darker shading indicates higher accuracy. Classification accuracies above 70% can be considered strong, values above 80% indicating high discriminatory power, and accuracies approaching or exceeding 90% reflecting excellent group separation despite biological variability. Gene sets related to DNA repair and ISR showed limited ability to distinguish healthy individuals from PD patients (A), but performed substantially better when distinguishing prodromal cases from either healthy individuals (B) or PD patients (C). A similar trend was observed for PD-core and PD-assoc gene sets (D–F). Notably, classification accuracy increased over time in prodromal individuals, especially for mtDNArep and ISR gene sets, suggesting that molecular signatures become more distinct as disease progresses. These results highlight the potential of blood-based gene expression patterns to detect early, preclinical changes in Parkinson’s disease.

#### Healthy vs. prodromal PD

In contrast, classification accuracy was high when distinguishing healthy individuals from those in the prodromal phase of Parkinson’s disease for all three gene sets, except for the baseline visit (Figure 3B). Accuracy steadily increased over time for the mitochondrial DNA repair gene set, reaching a peak of 0.89 at month 36, and was highest for the integrated stress response gene set at month 24, with an accuracy of 0.91. Gene expression variability was greatest at baseline and decreased over time, with the lowest variance observed at month 24. This pattern suggests that gene expression levels become more uniform among prodromal individuals as the disease advances. The reduction in variability likely contributes to improved classification accuracy and supports the idea that molecular disruptions in these pathways are most dynamic early in the prodromal period, becoming more consistent as individuals approach clinical diagnosis.

#### Prodromal PD vs. established PD

Similarly, classification accuracy between prodromal and established Parkinson’s disease was consistently high at most time points, apart from the baseline visit for the prodromal group (Figure 3C). However, accuracy showed a slight decline at the later time points, suggesting that gene expression differences between these two stages become less pronounced as the disease advances toward clinical diagnosis.

### PD-Associated and Core PD Gene Sets

We also evaluated the performance of Parkinson’s disease-specific gene sets, including a core set of well-established PD-related genes (PD core) and a broader set of PD-associated genes (PD assoc), to compare their classification ability against the DNA repair and integrated stress response pathways. This analysis aimed to determine whether genes directly linked to PD show clearer transcriptional differences than the more general stress and repair pathways.

#### Healthy vs Parkinson’s disease

Neither the PD core nor the PD-associated gene sets could reliably distinguish healthy individuals from those with diagnosed PD in this peripheral blood dataset (Figure 3D). Classification accuracy remained low across all time points, ranging from 49 to 65 percent, indicating limited transcriptional differences in these pathways once PD is clinically established.

#### Healthy vs Prodromal PD

In contrast, both PD core and PD-associated gene sets performed well in distinguishing healthy individuals from those in the prodromal phase (Figure 3E). Accuracy ranged from 65 to 87 percent, with the highest values observed at intermediate time points. The PD assoc gene set showed slightly stronger performance overall. However, as with other gene sets, accuracy was lower at the prodromal baseline and showed a slight decline at later visits.

#### Prodromal vs Established PD

The PD core and PD assoc gene sets also achieved high accuracy when distinguishing between prodromal and established PD, consistent with results from the DNA repair and ISR pathways (Figure 3F). Classification accuracy peaked at month 24 in the prodromal group, while baseline again showed weaker separation between groups.

### Global Gene Expression-Based Classification: Lack of Discrimination Between Healthy and PD

To determine whether any genes or gene combinations could robustly distinguish healthy individuals from established PD at any time point, we additionally conducted a classification analysis on the full dataset. Notably, classification accuracy remained low (53–67%) across timepoints (Supplementary Figure S1), indicating that peripheral blood RNA sequencing does not provide reliable biomarkers for established PD.

### Longitudinal gene expression

We examined how gene expression changed over time within each diagnostic group. In both healthy individuals and those with established Parkinson’s disease, gene expression remained relatively stable across all time points, with only minor changes observed. In contrast, individuals in the prodromal phase showed greater variability. Many genes had the highest expression variability at baseline and became more consistent by month 24 (see Supplementary Figures S2 to S4). This early variability likely contributed to lower classification accuracy at baseline and the improved accuracy observed at later time points.

Notably, many genes in the prodromal group did not follow a simple upward or downward trend. Instead, they displayed dynamic, non-linear patterns over time. About half of the DNA repair genes and nearly three-quarters of the ISR genes showed biphasic trajectories, where expression first increased and then decreased, or vice versa. These results suggest that the prodromal phase is marked by an active but temporary transcriptional response to early disease processes. As the disease progresses, this response appears to decline or collapse, resulting in the more stable but less distinct gene expression patterns seen in established Parkinson’s disease.

### Feature importance

To better understand the classification results, we identified the genes that contributed most to group separation by analyzing feature importance scores from months 12, 24, and 36. Baseline values were excluded due to low classification accuracy. Table 1 details the top 5 genes for each time point across the three gene sets. For each gene set, we further report the most important genes across time points in text. Full rankings are provided in Supplementary Tables S3 to S8.

**Table 1:**
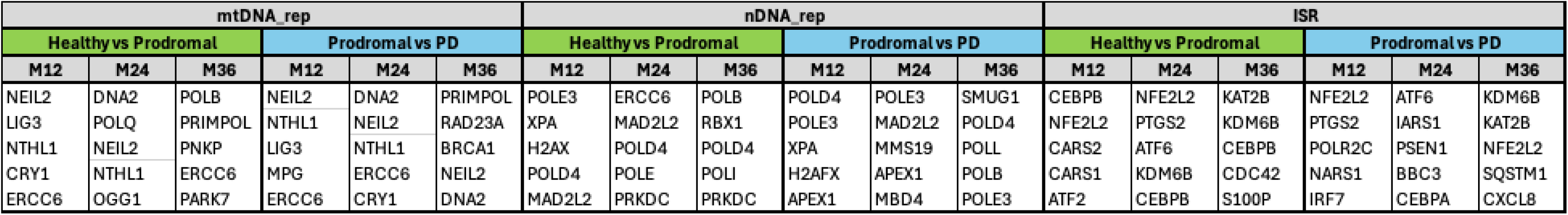
Top five genes ranked by feature importance of the respective timepoint.

#### mtDNA_rep

In the mtDNA_rep gene set, classifying healthy vs prodromal PD, the genes *ERCC6* and *PRIMPOL* consistently ranked in the top 10 in all three timepoints. *NEIL2, NTHL1, RAD23A* and *DNA2* ranked top 10 in M12 and M24, while *MUTYH* ranked top 10 in M24 and M36.

When classifying prodromal vs established PD, *NEIL2* and *ERCC6* consistently ranked in the top 10 in all three timepoints. *ERCC2* and *NTHL1* ranked top 10 in M12 and M24. Notably, *NTHL1* sank to rank 26 in M36. *DNA2* ranked top 10 in M24 and M36.

All rankings are shown in supplementary tables S3 and S4.

#### DNA_rep

In the DNA_rep gene set, classifying healthy vs prodromal PD, the genes *POLD4, XPC, POLE3* and *MAD2L2* consistently ranked in the top 20 in all three timepoints. *H2AX, ERCC6, NEIL2, APEX1, GTF2H1* and *APTX* ranked top 20 in M12 and M24, and *POLB, PRKDC, REV3L* and *POLE4* ranked top 20 in M24 and M36.

When classifying prodromal vs established PD, *POLD4, POLE4* and *ERCC6* consistently ranked in the top 20 in all three timepoints. *RIF1, MAD2L2, REV1, APEX1, POLA1, H2AFX* and *MMS19.* Notably, *APEX1, POLA1, H2AFX* and *MMS19* dropped in rank substantially in M36. *RAD23B, MBD4, PRKDC* and *REV3L* were in the top 20 in both M24 and M36. All rankings are shown in supplementary tables S5 and S6.

#### ISR

In the ISR gene set, classifying healthy vs prodromal PD, the genes *KDM6B, CEBPB, CDC42, S100P, HLA-DRB1, LITAF, CARS2, ATF2, IRF7, PSEN1, ERVW-1, ATF4, NFE2L2, SLC38A2, CREBBP, LARS1, CXCL8, IARS1* and *ATF6* consistently ranked in the top 20 in all three timepoints. *PTGS2, CARS1, CREB1 and RPS6KA3* ranked top 20 in M12 and M24, whereas *IARS1* ranked top 20 in M24 and M36.

When classifying prodromal vs established PD, *NFE2L2, CXCL8, CEBPB, ATF6,* and *IARS1* consistently ranked in the top 20 in all three timepoints. *POLR2C, CARS1, BBC3,* and *CSF1R* ranked top 20 in M12 and M24, whereas *KDM6B, STAT3, RPS6KA3, DISC1* and *GRIN2A* ranked top 20 in M24 and M36. All rankings are shown in supplementary table S7 and S8.

## Discussion

This study investigated longitudinal changes in DNA repair and integrated stress response gene expression during the prodromal stages of PD using peripheral blood transcriptomics. Previous studies on DNA repair dysfunction in PD have predominantly relied on cross-sectional analyses conducted after clinical diagnosis, largely overlooking the critical prodromal period. Consequently, little is known about how gene expression patterns associated with DNA repair dysfunction and ISR evolve before clinical symptoms emerge and how these early molecular changes relate to disease progression. Understanding these dynamic transcriptional changes in the prodromal phase is crucial, as it could reveal early biomarkers of PD and provide insights into potential compensatory mechanisms activated prior to extensive neuronal loss.

Our findings add significant nuance to the evolving understanding that disruptions in the expression of DNA repair genes occur long before PD symptoms manifest. This aligns closely with previous cross-sectional evidence of DNA damage and impaired genome maintenance mechanisms in PD patients^16^. Addressing this important gap, our longitudinal analysis provides novel insights into the evolution of DNA repair dysfunction and integrated stress response (ISR) gene expression throughout the prodromal stages of PD. Utilizing rigorous classification analyses on peripheral blood transcriptomic data, we identified that gene sets related to DNA repair and ISR effectively distinguished prodromal PD individuals from healthy controls and established PD. Conversely, these gene expression signatures did not reliably differentiate patients with clinically established PD from healthy individuals, underscoring that the most pronounced peripheral signals of DNA repair and ISR occur early in disease progression, well before classical motor symptoms manifest.

Our data further highlight substantial transcriptional variability early on in prodromal PD and, importantly, nonlinear expression patterns in many genes. Approximately half of the genes involved in DNA repair pathways and nearly three-quarters of ISR-associated genes demonstrated biphasic expression trajectories. These complex temporal patterns may reflect an initial adaptive response aimed at mitigating accumulating genomic stress, followed by an eventual decline as the compensatory mechanisms become insufficient with disease advancement.

The relative absence of curated DNA repair and ISR genes among the top differentially expressed genes suggests that these pathways may not be driven by large changes in individual gene expression but instead reflect subtle, coordinated shifts across many components. This underscores the limitations of single-gene DE analysis in capturing early disease signatures and reinforces the rationale for pathway-level and integrative modeling approaches.

Feature importance analysis further strengthened our findings by consistently identifying specific DNA repair genes, most importantly ERCC6, PRIMPOL, NEIL2, and NTHL1, as significant and recurring molecular predictors of prodromal PD. ERCC6 and NEIL2 consistently ranked highly across multiple assessments, emphasizing their potential role as robust early biomarkers. Interestingly, NTHL1 emerged as a strong early important feature but declined in importance at later stages, suggesting an important role in the initial prodromal phase, which may reflect an eventually overwhelmed compensatory repair mechanism as disease progresses.

ERCC6, also known as CSB, is crucial for transcription-coupled nucleotide excision repair (TC-NER), targeting actively transcribed genes for repair. It remodels chromatin to facilitate access for repair enzymes and interacts with key factors such as P53 and RNA Polymerase II^17,18^. ERCC6 dysfunction results in defective oxidative DNA repair, contributing to nerve cell death, photosensitivity, and premature aging. Mutations in ERCC6 cause Cockayne syndrome type II (CS-II), a disorder characterized by neurodevelopmental deficits and premature aging^19^. Importantly NER importance is evidenced as Ercc1-mediated DNA repair is necessary for preservation of dopaminergic neurons^20^. Mice heterozygous for Ercc1 defects display signs of dopaminergic pathology and PD patients’ peripheral cells exhibit inefficient nucleotide excision repair^20^.

NEIL2, a DNA glycosylase involved in BER, plays a crucial role in maintaining genomic integrity in both nucleus and mitochondria^10^. NEIL2 associates with RNA polymerase II and other repair proteins, suggesting its involvement in transcription-coupled repair^21^. Iron and copper, which accumulate in neurodegenerative diseases, inhibit NEIL2 activities, potentially exacerbating oxidative genome damage^22^. This inhibition can be reversed by metal chelators, including curcumin, indicating therapeutic potential^22^.

PrimPol, is a recently discovered primase-polymerase, plays crucial roles in both nuclear and mitochondrial DNA maintenance^23^. It functions as a DNA polymerase, capable of extending primers and bypassing various lesions, and as a primase, catalyzing DNA primer formation, and is essential for efficient DNA replication in both the nucleus and mitochondria^23,24^. It is specifically required for replication reinitiation after mtDNA damage^25^. PrimPol’s ability to start DNA chains with deoxynucleotides and bypass common oxidative lesions makes it unique among primases^26^. Given its critical role in bypassing oxidative DNA lesions during mtDNA replication, PrimPol may play a key role in the cellular response to oxidative stress in prodromal PD.

Interestingly, NTHL1 initially emerged as an early prodromal marker, but its relevance sharply declined with disease progression. This is in line with a potentially critical compensatory DNA repair mechanism occurring early in disease pathogenesis and then failing. The *C. elegans* ortholog NTH-1 has been implicated in the age-dependent accumulation of single-stranded DNA breaks, contributing to PD and Alzheimer’s disease (AD) pathology in *C. elegans*. In these disease models, NTH-1 generates strand breaks during physiological aging and its loss is associated with a protective phenotype characterized by reduced proteotoxicity and activation of cellular defenses that improve overall health as well as cognitive function^11,27^. Recent studies have also linked NTHL1 DNA repair to mitochondrial fitness and adaptive stress responses in human cells^28^.

Collectively, our findings clearly demonstrate that dynamic changes in DNA repair dysfunction and ISR pathways occur early in PD pathogenesis and can be reliably captured in peripheral blood, highlighting their promising utility as early-stage molecular biomarkers. The longitudinal transcriptomic changes we report here suggest that the DNA repair perturbations may dynamically evolve as individuals transition from prodromal stages to established PD.

One compelling observation in our analysis is that, in contrast to established PD, prodromal individuals exhibit substantial gene expression volatility and non-linear trajectories, indicating an active biological response aimed at compensating for accruing molecular stress. The biphasic expression patterns observed in half of DNA repair genes during prodromal stages, which were absent in established PD, support this interpretation. We propose that this adaptive transcriptional flexibility reflects a transient attempt to counteract oxidative and genomic stress, aligning with experimental evidence of a compensatory phase before symptom onset. As PD progresses, this response may become overwhelmed, ultimately shifting to diminished homeostatic equilibrium.

Our study has several notable strengths. The longitudinal design enabled us to capture dynamic gene expression changes related to DNA repair dysfunction and ISR during prodromal PD, a perspective often missed by cross-sectional studies. Using the high-quality PPMI dataset, combined with rigorous statistical validation through extensive cross-validation, enhances the reliability and reproducibility of our findings. Additionally, the comprehensive evaluation of multiple biologically relevant gene sets ensured hypothesis-driven analyses, thereby minimizing the potential for statistical biases associated with exploratory analyses or selective reporting of findings.

However, several limitations warrant consideration. Our relatively small sample size may limit the detection of subtle transcriptional changes and reduce the accuracy in capturing disease heterogeneity. Additionally, gene expression in whole blood is only moderately correlated with gene expression in the brain, suggesting peripheral blood samples may be a useful but imperfect proxy for central nervous system processes^29^. Further, variability in gene expression could also be influenced by external factors such as immune status, medication use, or comorbid conditions. Importantly, transcript levels do not directly correlate to protein levels, thus do not provide insights into all potentially relevant factors. Lastly, the demographic characteristics of our cohort potentially limit the generalizability of results.

Despite these limitations, our study contributes significantly to understanding the molecular dynamics of prodromal PD and presents several important areas that merit further investigation. Given the promising biomarkers identified, future research should validate their diagnostic potential in larger, more diverse cohorts, ideally incorporating multi-center studies to enhance generalizability, and further explore the biological mechanisms at play with *in vitro* or *in vivo* models of PD. Additionally, integrating multi-omics approaches, such as proteomics, metabolomics, or epigenomics, could deepen our understanding of DNA repair dysfunction and ISR pathways in prodromal PD and increase mechanistic understanding. Lastly, research exploring interventions targeting the early compensatory mechanisms identified through our longitudinal data could lead to novel therapeutic strategies for delaying PD progression.

In conclusion, this longitudinal study offers novel insights into the dynamics of DNA repair dysfunction and ISR gene expression in prodromal PD. Our findings highlight distinct molecular changes that occur prior to clinical diagnosis, identifying specific genes as promising biomarkers for early detection. While acknowledging its limitations, this study significantly advances our understanding of early-stage PD pathology and lays the groundwork for future research to better explore the variance and heterogeneity in prodromal PD, ultimately advancing clinical applications.

## Data Availability

All data used are available online at https://ida.loni.usc.edu/ or
https://www.ppmi-info.org/access-data-specimens/download-data. No new data was produced.

https://www.ppmi-info.org/access-data-specimens/download-data

## Acknowledgements

Data used in the preparation of this article was obtained on 2025-02-01 from the Parkinson’s Progression Markers Initiative (PPMI) database (www.ppmi-info.org/access-data-specimens/download-data), RRID:SCR_006431. For up-to-date information on the study, visit www.ppmi-info.org.

PPMI – a public-private partnership – is funded by the Michael J. Fox Foundation for Parkinson’s Research, and funding partners; including 4D Pharma, Abbvie, AcureX, Allergan, Amathus Therapeutics, Aligning Science Across Parkinson’s, AskBio, Avid Radiopharmaceuticals, BIAL, BioArctic, Biogen, Biohaven, BioLegend, BlueRock Therapeutics, Bristol-Myers Squibb, Calico Labs, Capsida Biotherapeutics, Celgene, Cerevel Therapeutics, Coave Therapeutics, DaCapo Brainscience, Denali, Edmond J. Safra Foundation, Eli Lilly, Gain Therapeutics, GE HealthCare, Genentech, GSK, Golub Capital, Handl Therapeutics, Insitro, Jazz Pharmaceuticals, Johnson & Johnson Innovative Medicine, Lundbeck, Merck, Meso Scale Discovery, Mission Therapeutics, Neurocrine Biosciences, Neuron23, Neuropore, Pfizer, Piramal, Prevail Therapeutics, Roche, Sanofi, Servier, Sun Pharma Advanced Research Company, Takeda, Teva, UCB, Vanqua Bio, Verily, Voyager Therapeutics, the Weston Family Foundation and Yumanity Therapeutics.

The data handling was enabled by resources provided by the National Academic Infrastructure for Supercomputing in Sweden (NAISS), partially funded by the Swedish Research Council through grant agreement no. 2022-06725. This work was partially supported by the Research Council of Norway through its Centres of Excellence scheme, project number 332713.

We thank OpenAI’s ChatGPT (GPT-4, May 2025) for assistance with minor editorial revisions.

## Author contribution

DA: Study design, differential expression analysis, classifier analysis, interpretation of results, writing of manuscript. NPM: Study design, interpretation of results, writing of manuscript. EN: differential expression analysis. DD: differential expression analysis. HLN: Study design, interpretation of results, writing of manuscript. AP: Study design, data analysis, interpretation of results, writing of manuscript. All authors critically revised the manuscript.

