## Supplementary material for "Longitudinal Assessment of DNA Repair Signature Trajectory in Prodromal versus Established Parkinson’s Disease": Table S1

Table S1: List of the respective gene sets

| mtDNA_rep | DNA_rep | ISR | PD_core | PD_assoc |
|---|---|---|---|---|
| ALKBH1 | APEX1 | AARS2 | ATP13A2 | ASXL3 |
| APEX1 | APEX2 | ACOT11 | CHCHD2 | BAG3 |
| BRCA1 | APLF | APOE | DNAJC6 | BIN3 |
| CRY1 | APTX | ASNS | EIF4G1 | BRIP1 |
| DNA2 | BLM | ATF2 | FBXO7 | BST1 |
| DUT | CETN2 | ATF3 | GBA1 | C5orf24 |
| ERCC2 | DCLRE1A | ATF4 | GIGYF2 | CAB39L |
| ERCC6 | DCLRE1C | ATF5 | HTRA2 | CAMK2D |
| LIG3 | DDB1 | ATF6 | LRP10 | CASC16 |
| MGMT | DDB2 | ATG5 | LRRK2 | CD19 |
| MPG | ERCC1 | BBC3 | PARK7 | CHD9 |
| MUTYH | ERCC2 | BGLAP | PINK1 | CHRNB1 |
| NEIL1 | ERCC3 | CA9 | PLA2G6 | CLCN3 |
| NEIL2 | ERCC4 | CARS1 | PRKN | CRHR1 |
| NTHL1 | ERCC5 | CARS2 | RIC3 | CRLS1 |
| OGG1 | ERCC6 | CASP12 | SNCA | CTSB |
| PARK7 | ERCC8 | CCL2 | SYNJ1 | DLG2 |
| PNKP | FAAP20 | CCNA2 | TMEM230 | DNAH17 |
| POLB | FEN1 | CCND1 | UCHL1 | DYRK1A |
| POLG | GTF2H1 | CDC42 | USP24 | ELOVL7 |
| POLQ | GTF2H2 | CEBPA | VPS13C | FAM171A2 |
| PRIMPOL | GTF2H3 | CEBPB | VPS35 | FAM47E |
| RAD23A | GTF2H4 | CEBPE | | FAM47E-STBD1 |

| | | | | |
|---|---|---|---|---|
| RECQL4 | GTF2H5 | CHAC1 | | FAM49B |
| TP53BP1 | H2AX | CREB1 | | FBRSL1 |
| UNG | HPF1 | CREBBP | | FCGR2A |
| YBX1 | LIG1 | CSF1R | | FGF20 |
| | LIG3 | CTNNB1 | | FYN |
| | LIG4 | CXCL3 | | GAK |
| | MAD2L2 | CXCL8 | | GALC |
| | MBD4 | CYP2E1 | | GBF1 |
| | MCMDC2 | DARS2 | | GCH1 |
| | MMS19 | DDIT3 | | GPNMB |
| | MPG | DDIT4 | | HIP1R |
| | MRE11 | DDR2 | | HLA-DRB5 |
| | MUTYH | DISC1 | | IGSF9B |
| | NAN1 | DKK1 | | INPP5F |
| | NEIL1 | DNAJB9 | | IP6K2 |
| | NEIL2 | DRD2 | | ITGA8 |
| | NEIL3 | EDN1 | | ITPKB |
| | NHEJ1 | EIF2AK2 | | KCNIP3 |
| | NTHL1 | ELANE | | KCNS3 |
| | OGG1 | ERBB2 | | KPNA1 |
| | PARP1 | ERVW-1 | | KRTCAP2 |
| | PARP2 | F7 | | LCORL |
| | PARP3 | FGF19 | | LRRK2 |
| | PARP9 | FGF2 | | MAP4K4 |
| | PNKP | FGF21 | | MBNL2 |
| | POLA1 | GNB1L | | MCCC1 |
| | POLB | GPT2 | | MED12L |

| | | | | |
|---|---|---|---|---|
| | POLD1 | HLA-DRB1 | | MEX3C |
| | POLD2 | HMOX1 | | MIPOL1 |
| | POLD3 | HRK | | NOD2 |
| | POLD4 | HSPA5 | | NUCKS1 |
| | POLE | IARS1 | | PAM |
| | POLE2 | IGFBP1 | | PMVK |
| | POLE3 | IHH | | RAB29 |
| | POLE4 | IL18 | | RETREG3 |
| | POLH | IL1B | | RIMS1 |
| | POLI | IL23A | | RIT2 |
| | POLK | IL6 | | RNF141 |
| | POLL | INHBE | | RPS12 |
| | POLM | INS | | RPS6KL1 |
| | POLN | IRF7 | | SATB1 |
| | POLQ | ITIH3 | | SCAF11 |
| | PRIMPOL | KAT2B | | SCARB2 |
| | PRKDC | KDM6B | | SETD1A |
| | RAD1 | LAMP3 | | SH3GL2 |
| | RAD23A | LARS1 | | SIPA1L2 |
| | RAD23B | LARS2 | | SNCA |
| | RAD50 | LHX2 | | SPPL2B |
| | RAD9A | LITAF | | SPTSSB |
| | RBX1 | MAP1LC3B | | STK39 |
| | REV1 | MCL1 | | SYT17 |
| | REV3L | MTOR | | TMEM163 |
| | RFC4 | NARS1 | | TMEM175 |
| | RIF1 | NARS2 | | TRIM40 |

| | | | | |
|---|---|---|---|---|
| | RNF168 | NDC80 | | UBAP2 |
| | RNF8 | NFE2L2 | | UBTF |
| | SHLD1 | NRP1 | | VAMP4 |
| | SHLD2 | NUPR1 | | VPS13C |
| | SHLD3 | PDGFRA | | WNT3 |
| | SLX4 | PENK | | |
| | SMUG1 | PER2 | | |
| | TDG | PLAT | | |
| | TDP1 | PLAU | | |
| | TP53BP1 | PMAIP1 | | |
| | UNG | POLR2C | | |
| | UVSSA | PPARGC1A | | |
| | XAB2 | PPP1R15A | | |
| | XPA | PRKN | | |
| | XPC | PSEN1 | | |
| | XRCC1 | PTGS2 | | |
| | XRCC4 | PTH | | |
| | XRCC5 | RPS6KA3 | | |
| | XRCC6 | RUNX2 | | |
| | | S100A8 | | |
| | | S100P | | |
| | | SARS2 | | |
| | | SCG2 | | |
| | | SERPINC1 | | |
| | | SIGMAR1 | | |
| | | SIRT1 | | |
| | | SIRT2 | | |

| | | |
|---|---|---|
| | | SIRT4 |
| | | SLC38A2 |
| | | SLC6A4 |
| | | SLC7A11 |
| | | SNCG |
| | | SP7 |
| | | SQSTM1 |
| | | STAT3 |
| | | TARS2 |
| | | TH |
| | | THEG |
| | | TNC |
| | | TNF |
| | | TNFRSF10B |
| | | TNFRSF11A |
| | | TNFSF11 |
| | | TRIB3 |
| | | TRPV6 |
| | | USF1 |
| | | VEGFA |
| | | VIM |
| | | WARS2 |
| | | YARS2 |
