## Supplementary material for "Longitudinal Assessment of DNA Repair Signature Trajectory in Prodromal versus Established Parkinson’s Disease": Figure S1

| | | Healthy | | | |
|---|---|---|---|---|---|
| | | BL | M12 | M24 | M36 |
| PD | BL | 0.54 | 0.62 | 0.65 | 0.65 |
| | M12 | 0.62 | 0.53 | 0.61 | 0.64 |
| | M24 | 0.59 | 0.57 | 0.54 | 0.57 |
| | M36 | 0.67 | 0.6 | 0.59 | 0.54 |

Figure S1: Classification accuracy using the full set of genes for distinguishing Parkinson's Disease (PD) from Healthy individuals across different time points (BL, M12, M24, M36). Each cell represents the percentage of correctly classified individuals, with darker shades indicating higher accuracy.
