## Supplementary material for "Longitudinal Assessment of DNA Repair Signature Trajectory in Prodromal versus Established Parkinson’s Disease": Figure S2

Mean Expression of BRCA1 Over Time Across Groups

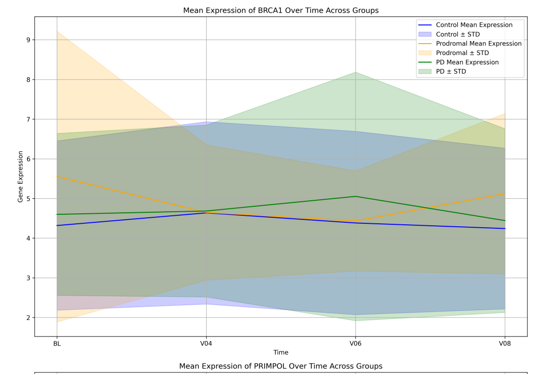


Mean Expression of PRIMPOL Over Time Across Groups

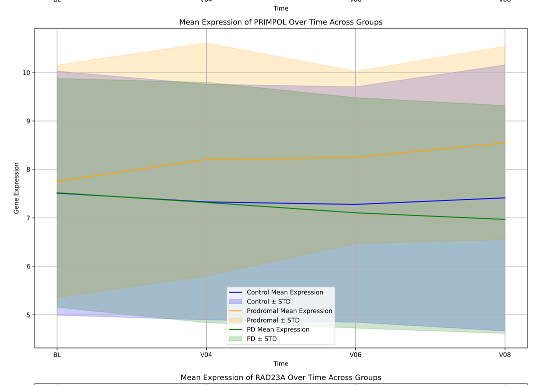

Mean Expression of RAD23A Over Time Across Groups

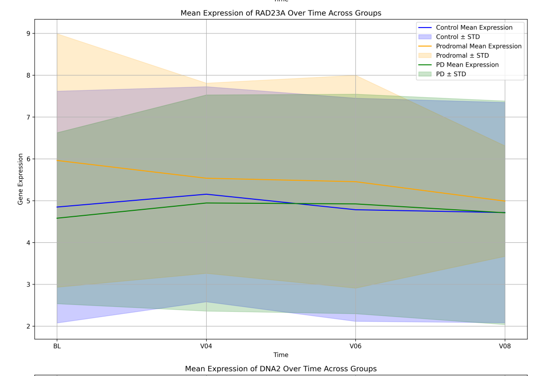

Mean Expression of DNA2 Over Time Across Groups

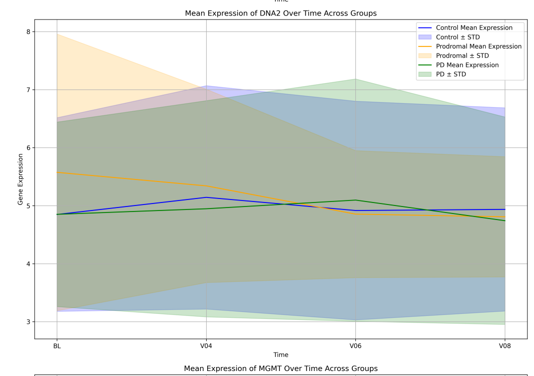

Mean Expression of MGMT Over Time Across Groups

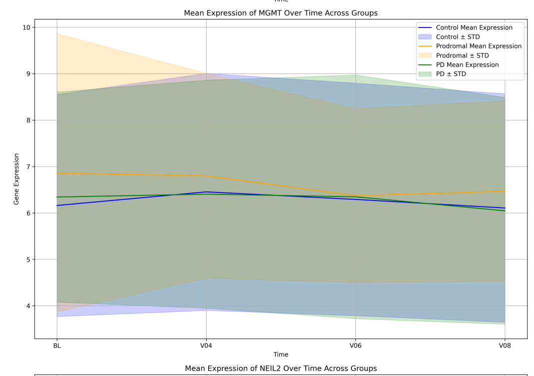

Mean Expression of NEIL2 Over Time Across Groups

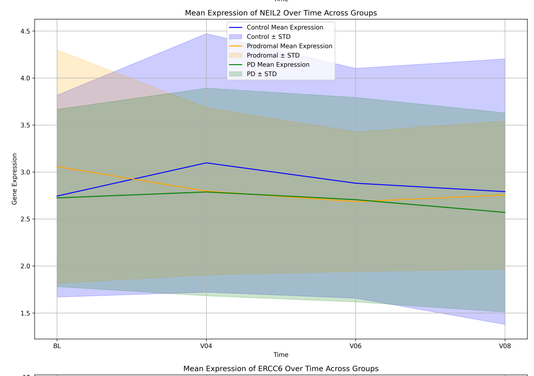

Mean Expression of ERCC6 Over Time Across Groups

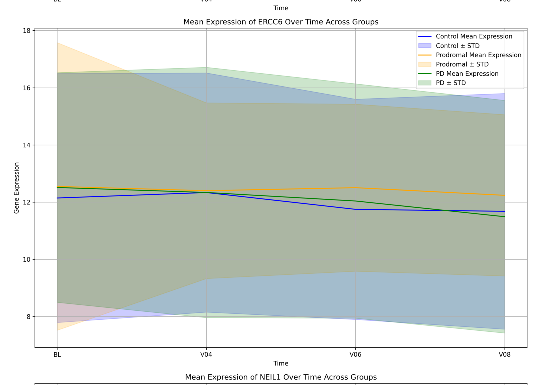

Mean Expression of NEIL1 Over Time Across Groups

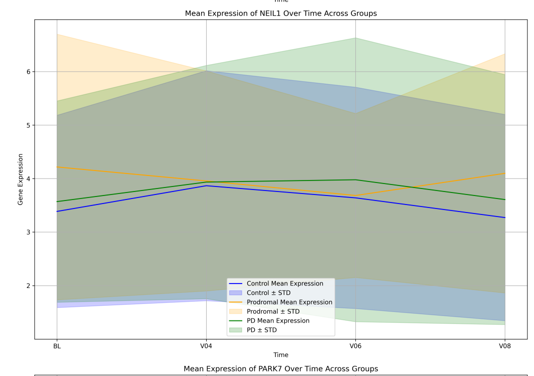

Mean Expression of PARK7 Over Time Across Groups

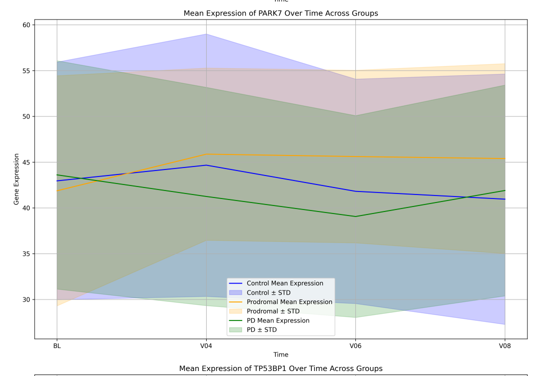

Mean Expression of TP53BP1 Over Time Across Groups

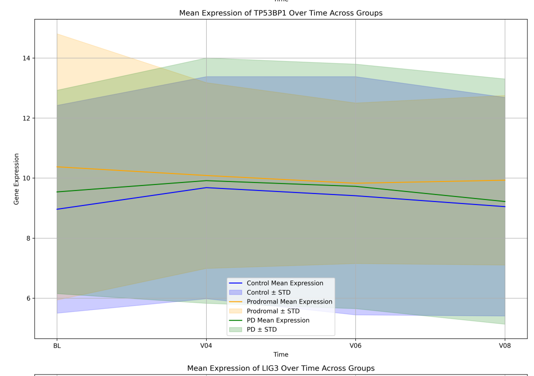

Mean Expression of LIG3 Over Time Across Groups

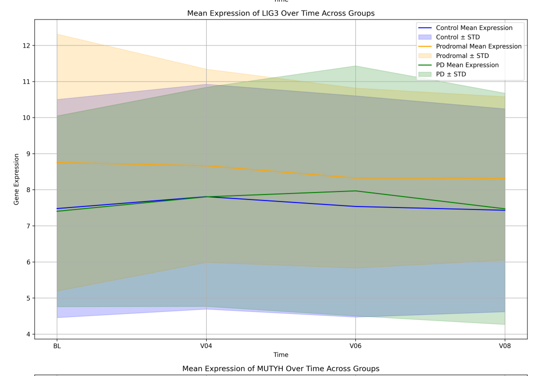

Mean Expression of MUTYH Over Time Across Groups

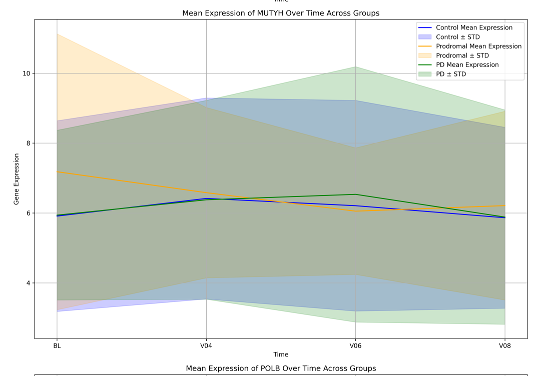

Mean Expression of POLB Over Time Across Groups

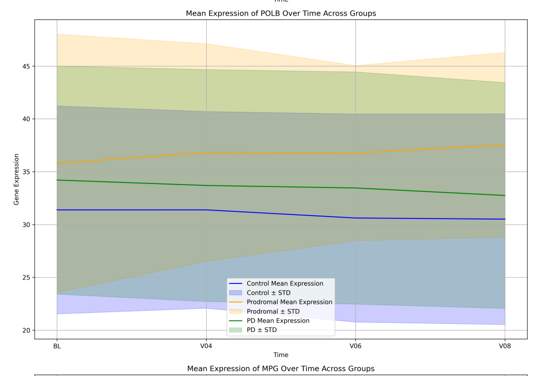

Mean Expression of MPG Over Time Across Groups

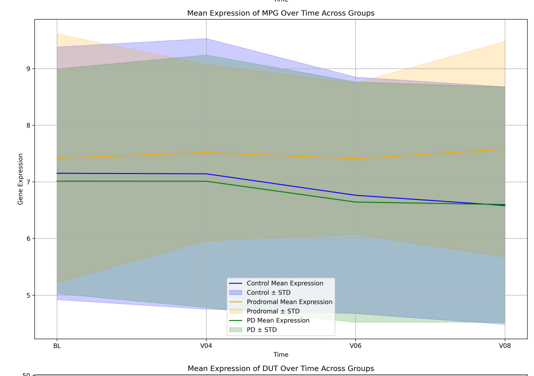

Mean Expression of DUT Over Time Across Groups

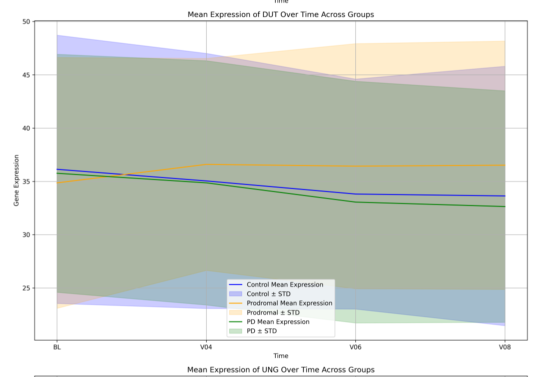

Mean Expression of UNG Over Time Across Groups

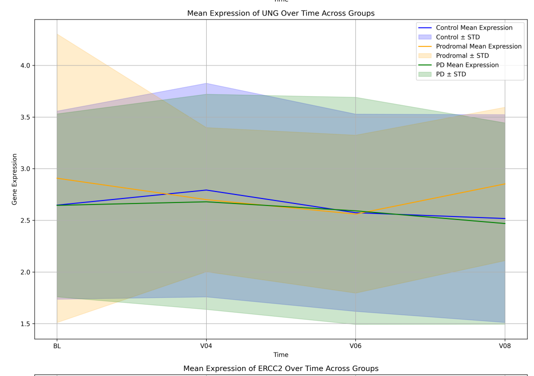

Mean Expression of ERCC2 Over Time Across Groups

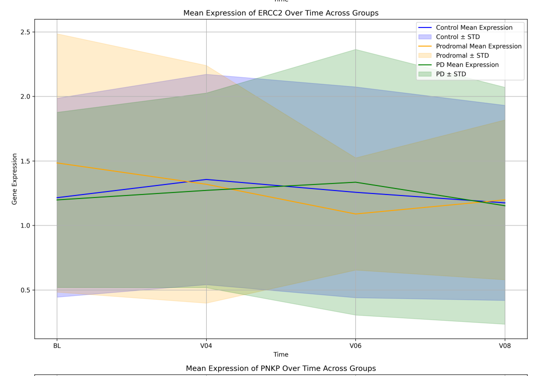

Mean Expression of PNKP Over Time Across Groups

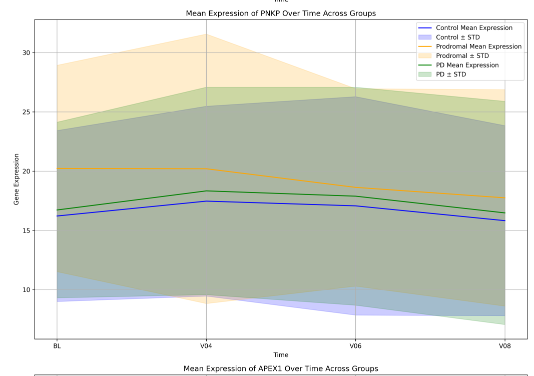

Mean Expression of APEX1 Over Time Across Groups

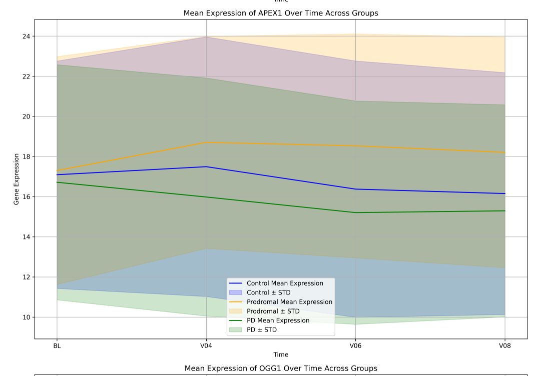

Mean Expression of OGG1 Over Time Across Groups

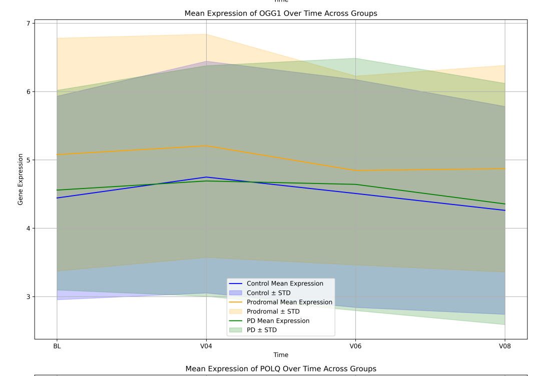

Mean Expression of POLQ Over Time Across Groups

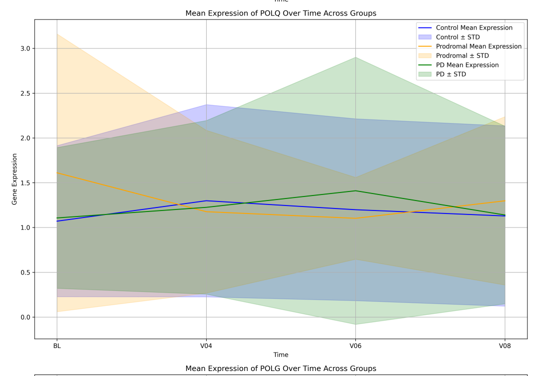

Mean Expression of POLG Over Time Across Groups

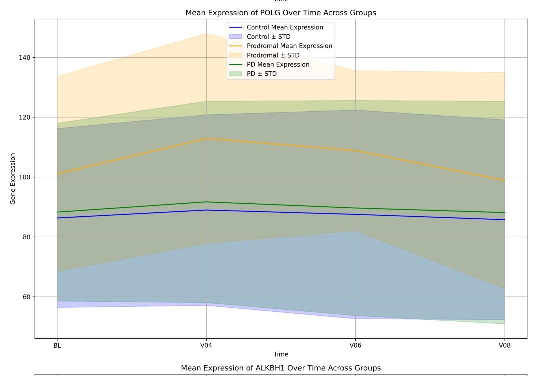

Mean Expression of ALKBH1 Over Time Across Groups

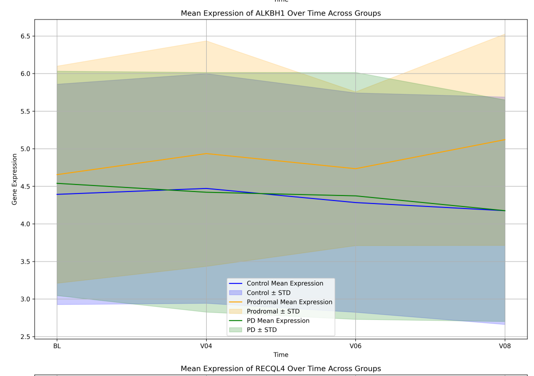

Mean Expression of RECQL4 Over Time Across Groups

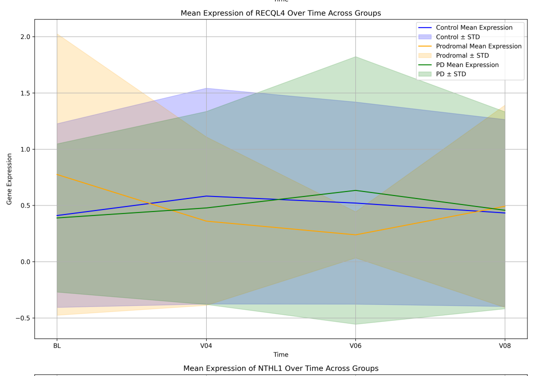

Mean Expression of NTHL1 Over Time Across Groups

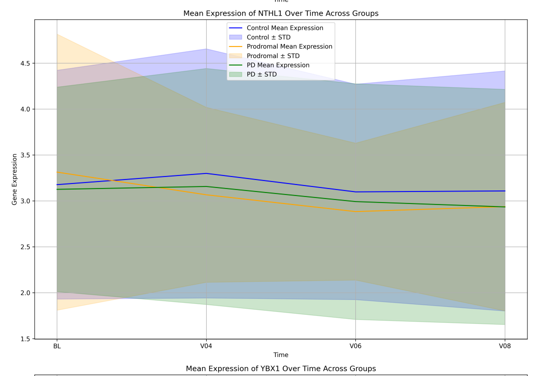

Mean Expression of YBX1 Over Time Across Groups

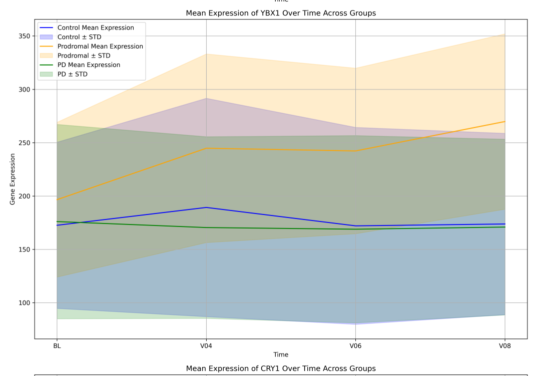

Mean Expression of CRY1 Over Time Across Groups

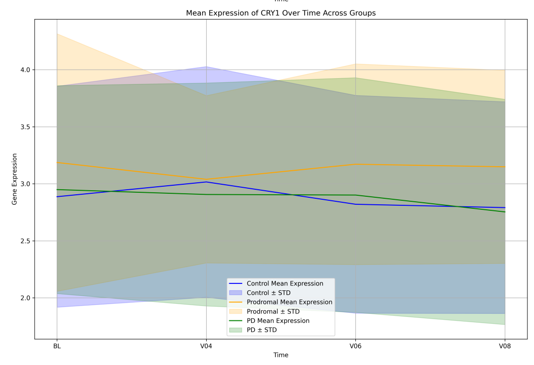
