## Supplementary figures and images for "Longitudinal Assessment of DNA Repair Signature Trajectory in Prodromal versus Established Parkinson’s Disease"

### Figure S3

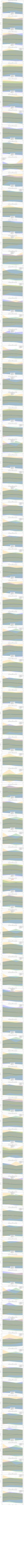
