## Supplementary material for "Longitudinal Assessment of DNA Repair Signature Trajectory in Prodromal versus Established Parkinson’s Disease": Figure S4

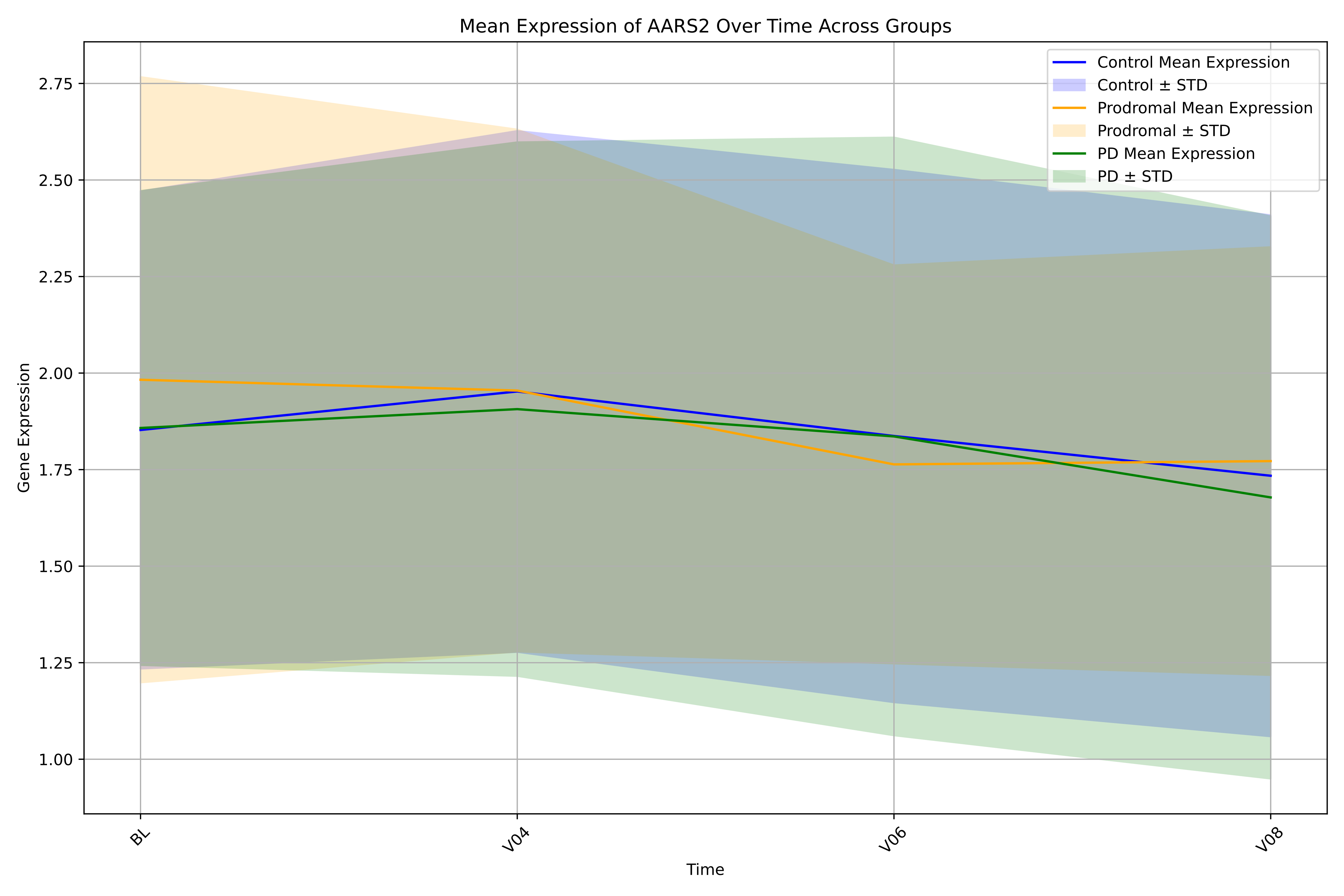

Mean Expression of AARS2 Over Time Across Groups
Control Mean Expression
Control ± STD
Prodromal Mean Expression
Prodromal ± STD
PD Mean Expression
PD ± STD
Gene Expression
2.75
2.50
2.25
2.00
1.75
1.50
1.25
1.00
BL
V04
V06
V08
Time

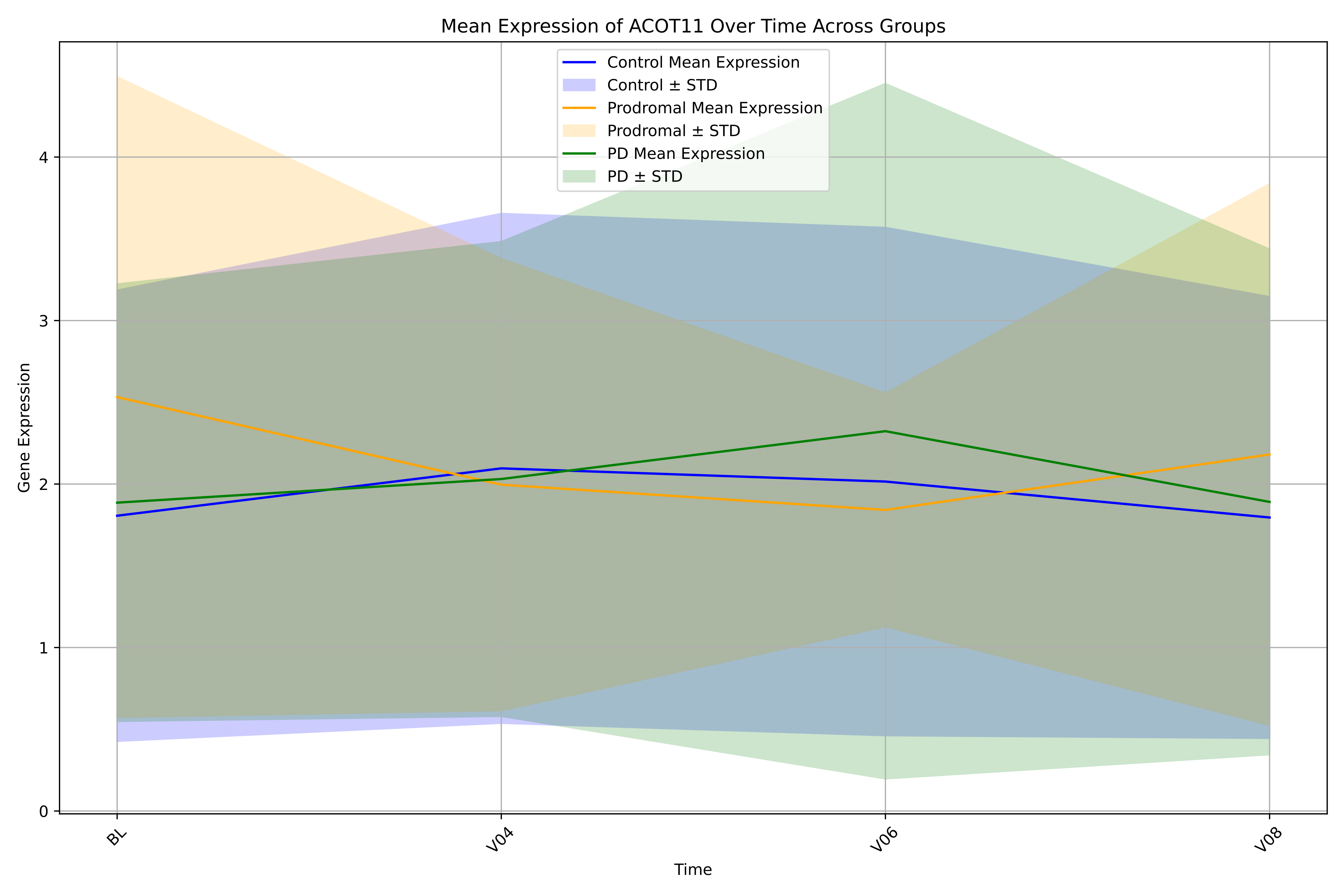
Mean Expression of ACOT11 Over Time Across Groups
Control Mean Expression
Control ± STD
Prodromal Mean Expression
Prodromal ± STD
PD Mean Expression
PD ± STD
Gene Expression
0
1
2
3
4
BL
V04
V06
V08
Time

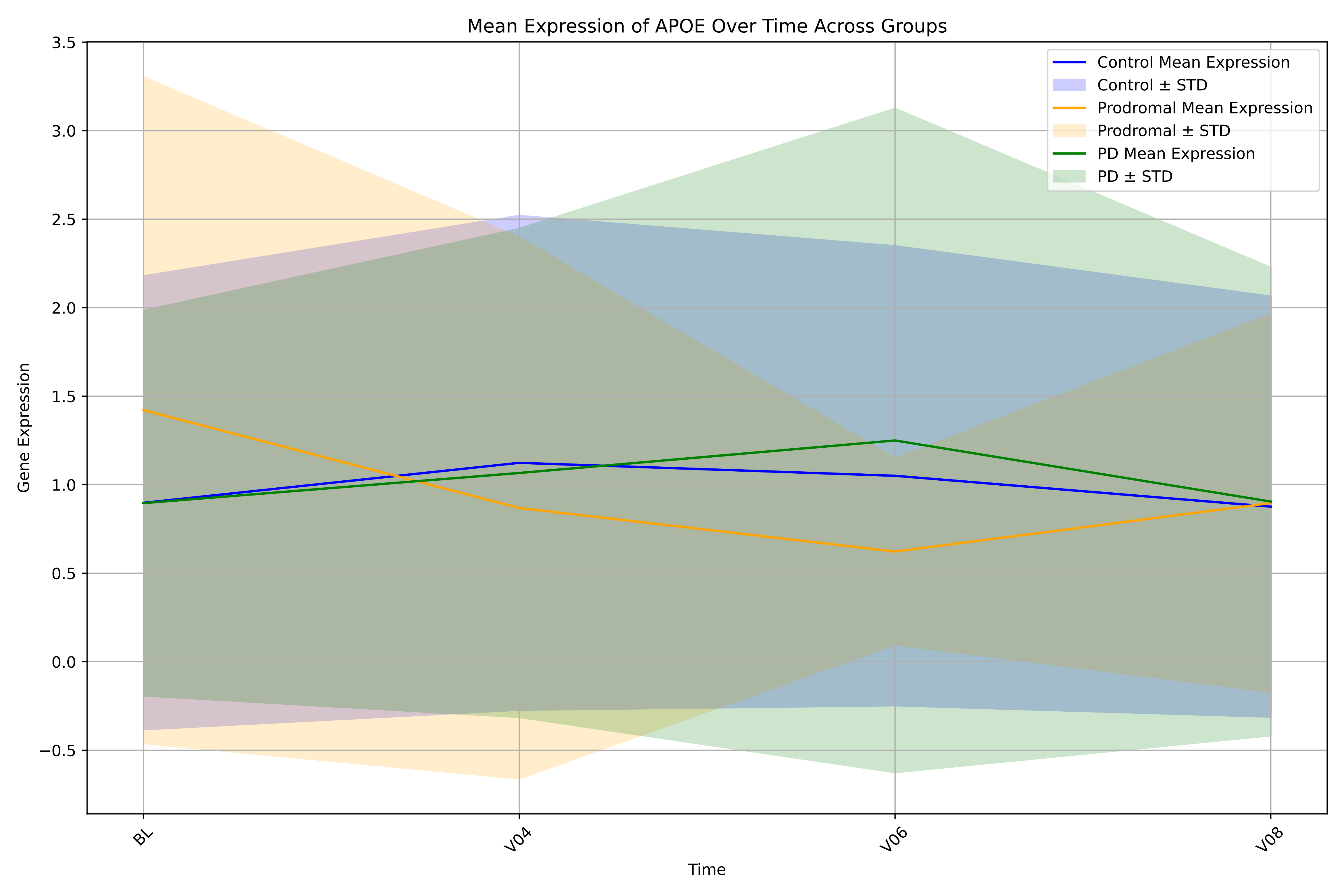

Mean Expression of APOE Over Time Across Groups
Control Mean Expression
Control ± STD
Prodromal Mean Expression
Prodromal ± STD
PD Mean Expression
PD ± STD
3.5
3.0
2.5
2.0
1.5
1.0
0.5
0.0
−0.5
Gene Expression
BL
V04
V06
V08
Time

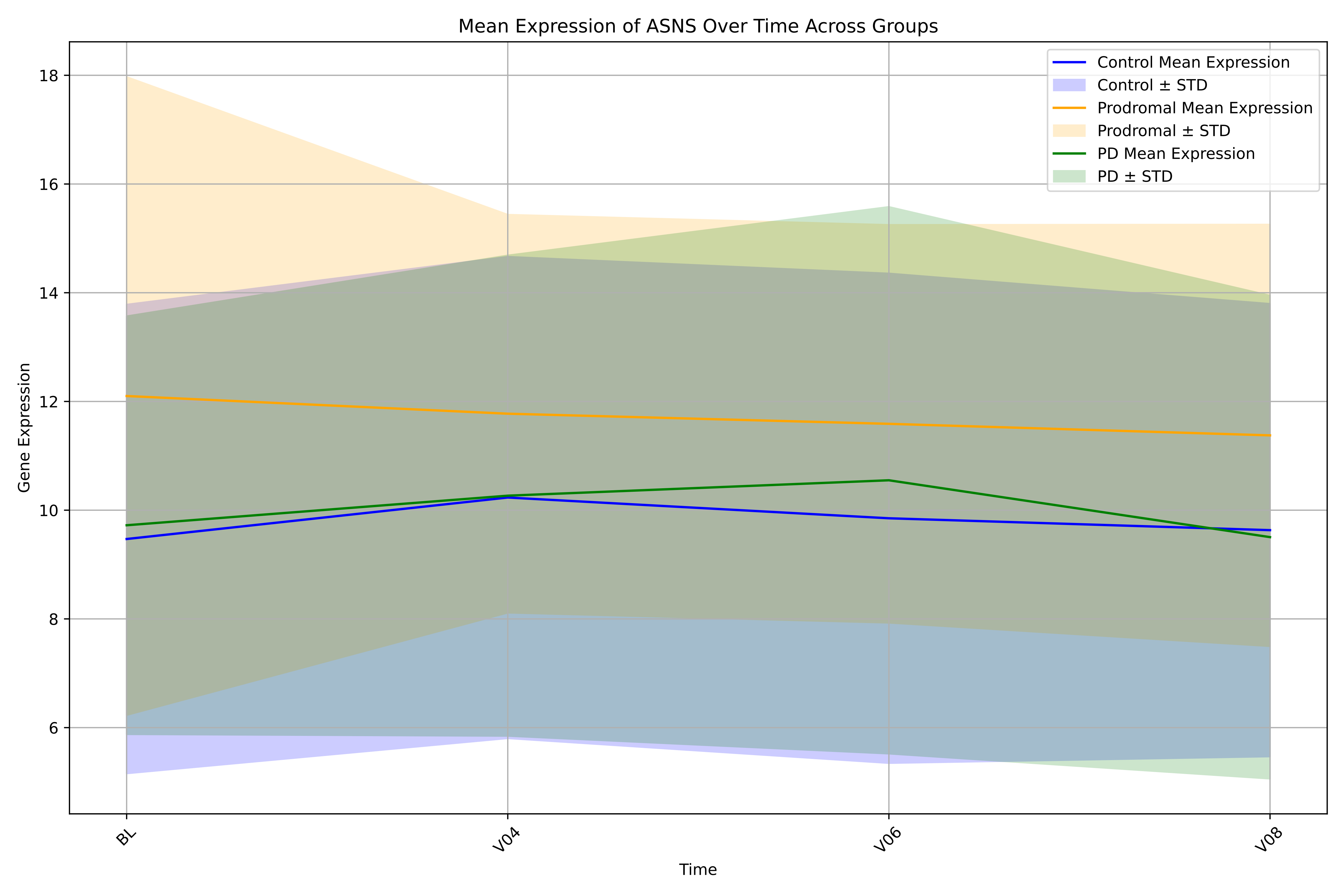

Mean Expression of ASNS Over Time Across Groups
Control Mean Expression
Control ± STD
Prodromal Mean Expression
Prodromal ± STD
PD Mean Expression
PD ± STD
Gene Expression
Time
18
16
14
12
10
8
6
BL
V04
V06
V08

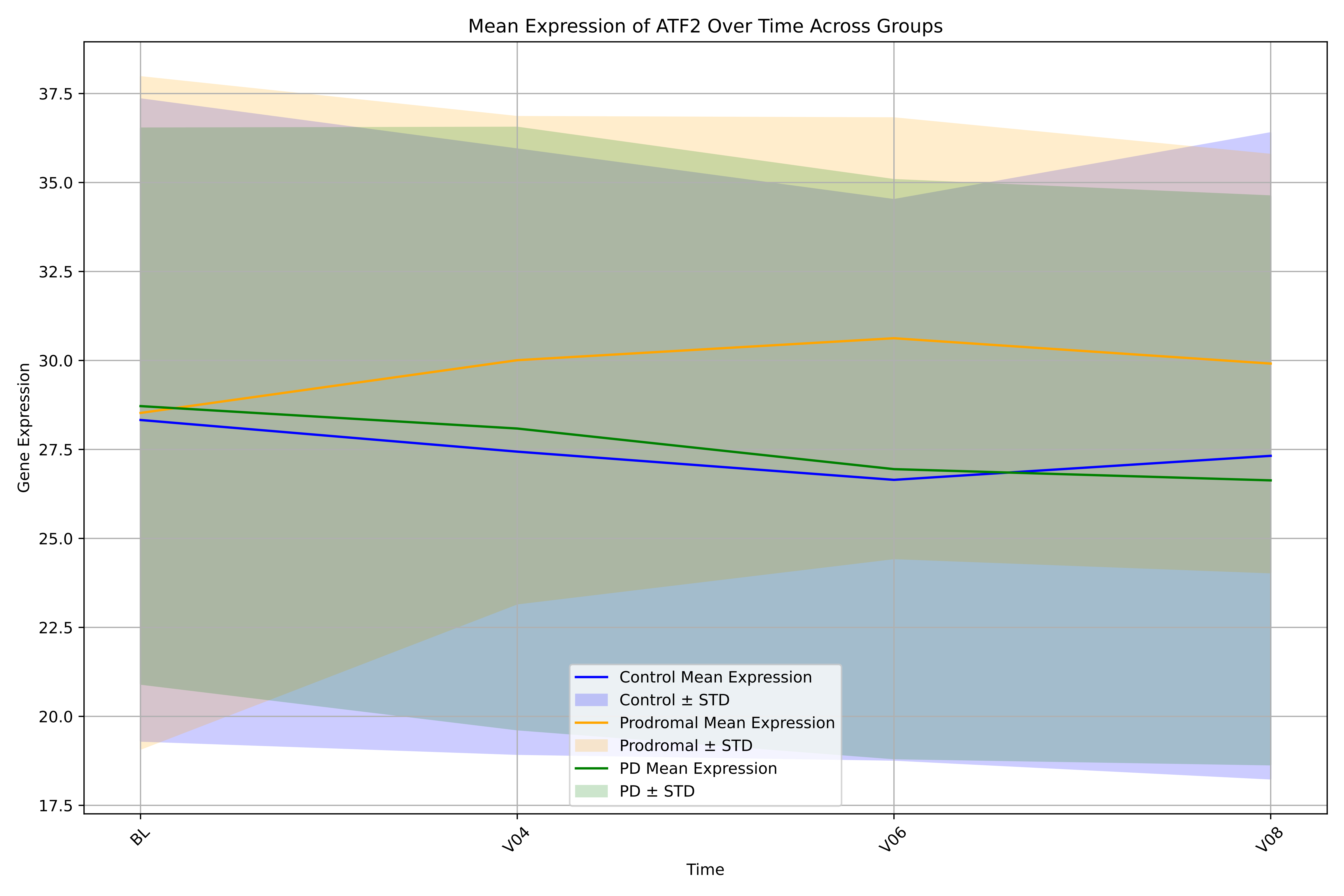

Mean Expression of ATF2 Over Time Across Groups
Gene Expression
Time
BL
V04
V06
V08
17.5
20.0
22.5
25.0
27.5
30.0
32.5
35.0
37.5
Control Mean Expression
Control ± STD
Prodromal Mean Expression
Prodromal ± STD
PD Mean Expression
PD ± STD

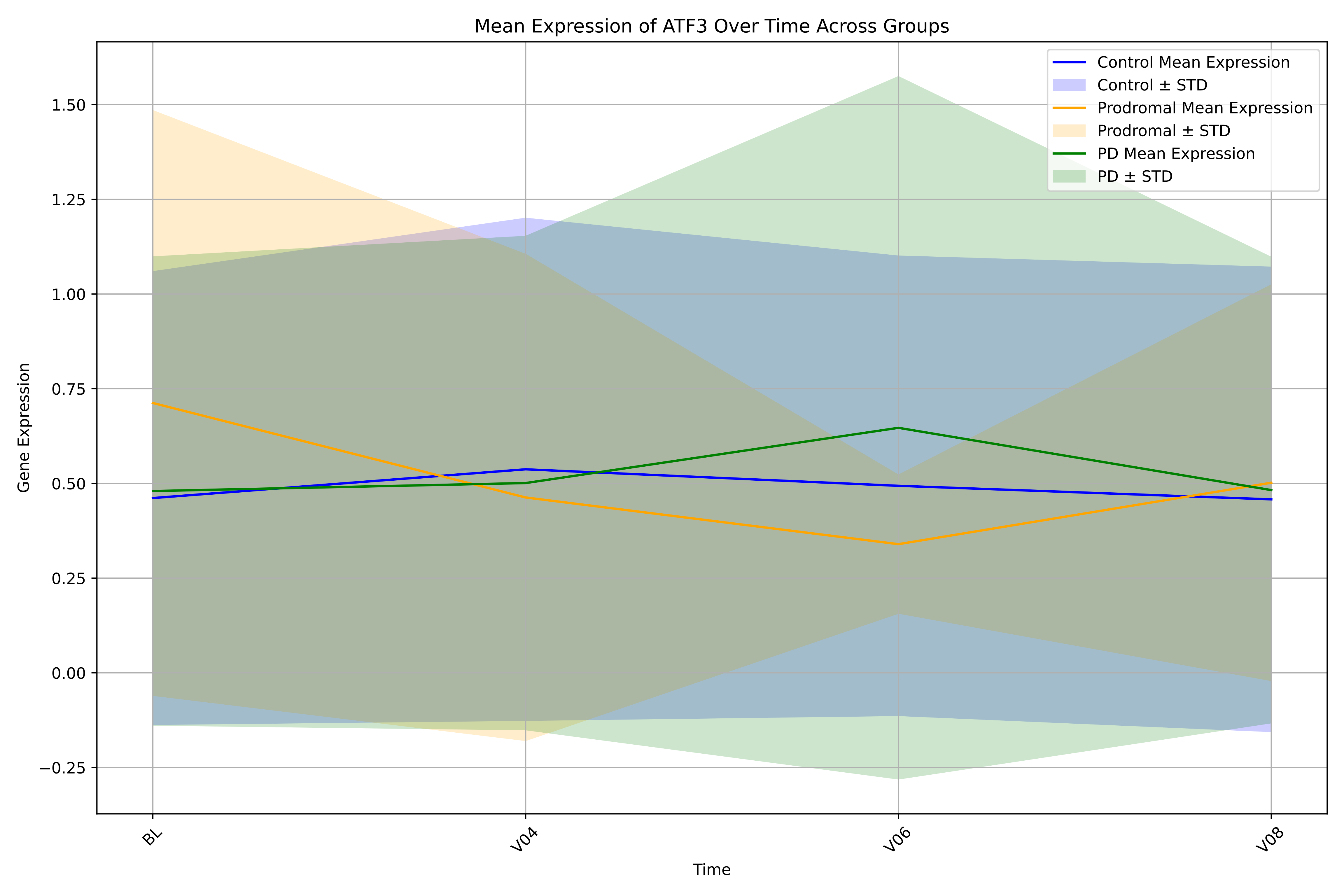

Mean Expression of ATF3 Over Time Across Groups
Control Mean Expression
Control ± STD
Prodromal Mean Expression
Prodromal ± STD
PD Mean Expression
PD ± STD
Gene Expression
1.50
1.25
1.00
0.75
0.50
0.25
0.00
−0.25
BL
V04
V06
V08
Time

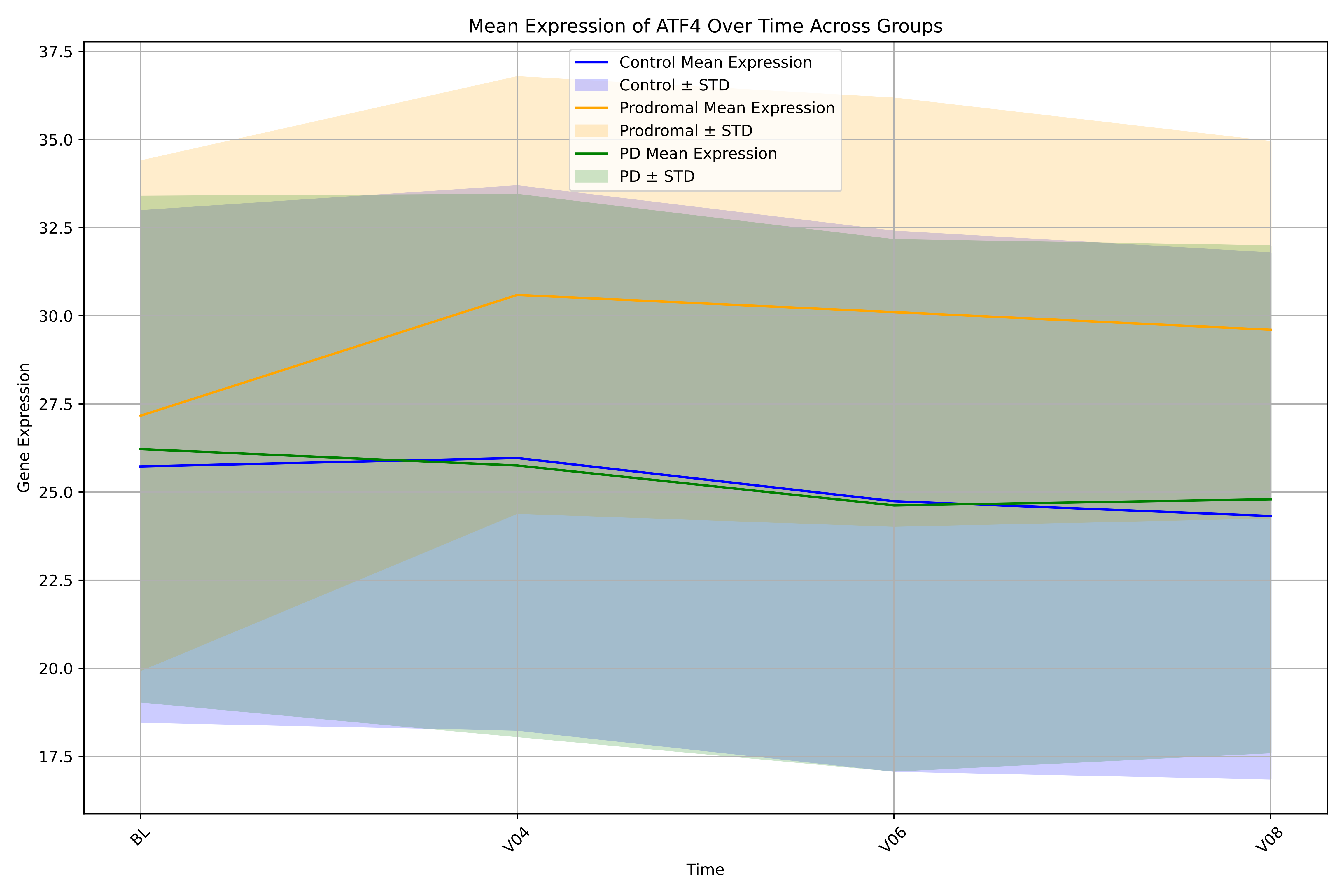

Mean Expression of ATF4 Over Time Across Groups
Control Mean Expression
Control ± STD
Prodromal Mean Expression
Prodromal ± STD
PD Mean Expression
PD ± STD
Gene Expression
37.5
35.0
32.5
30.0
27.5
25.0
22.5
20.0
17.5
BL
V04
V06
V08
Time

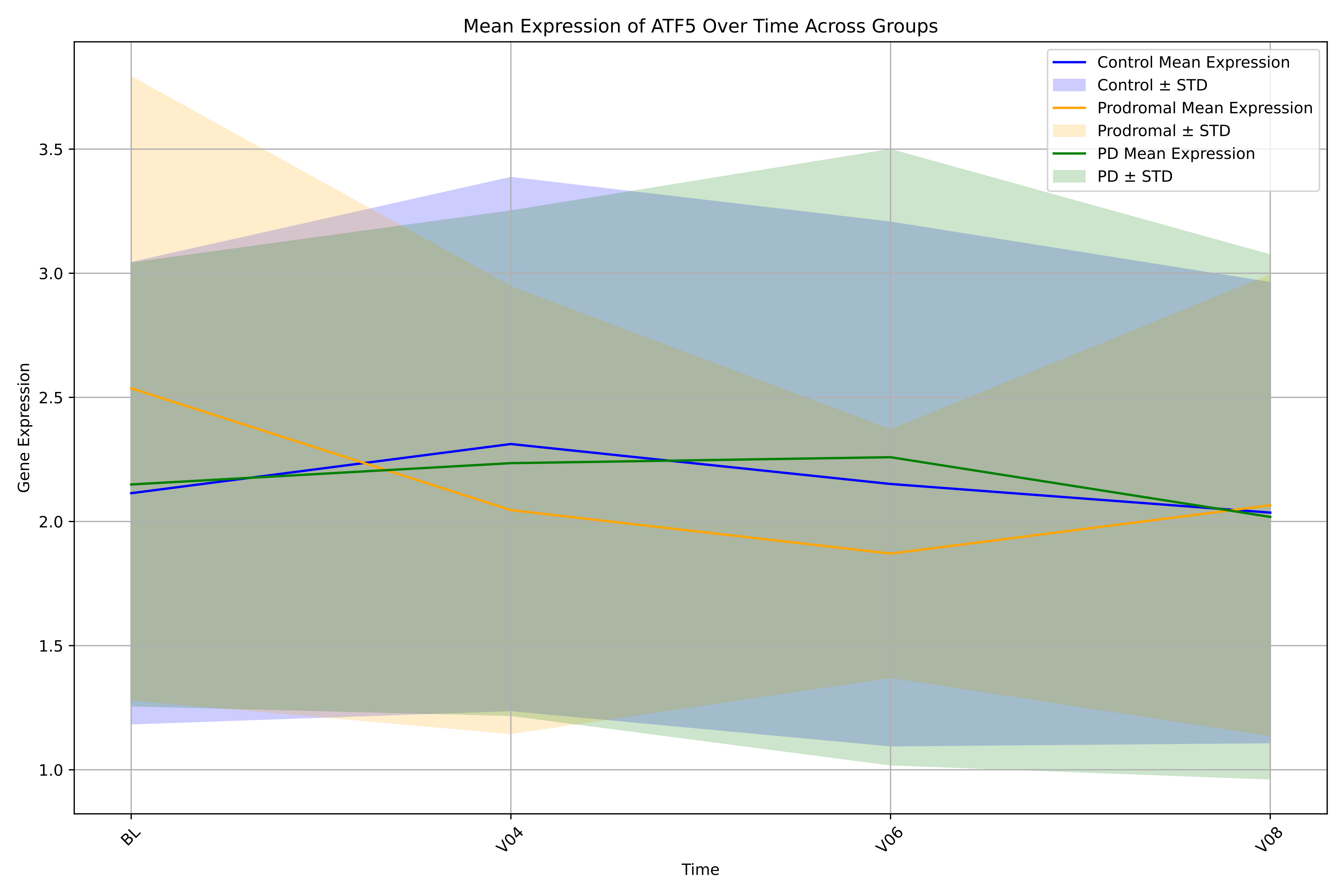
Mean Expression of ATF5 Over Time Across Groups
Control Mean Expression
Control ± STD
Prodromal Mean Expression
Prodromal ± STD
PD Mean Expression
PD ± STD
Gene Expression
3.5
3.0
2.5
2.0
1.5
1.0
BL
V04
V06
V08
Time

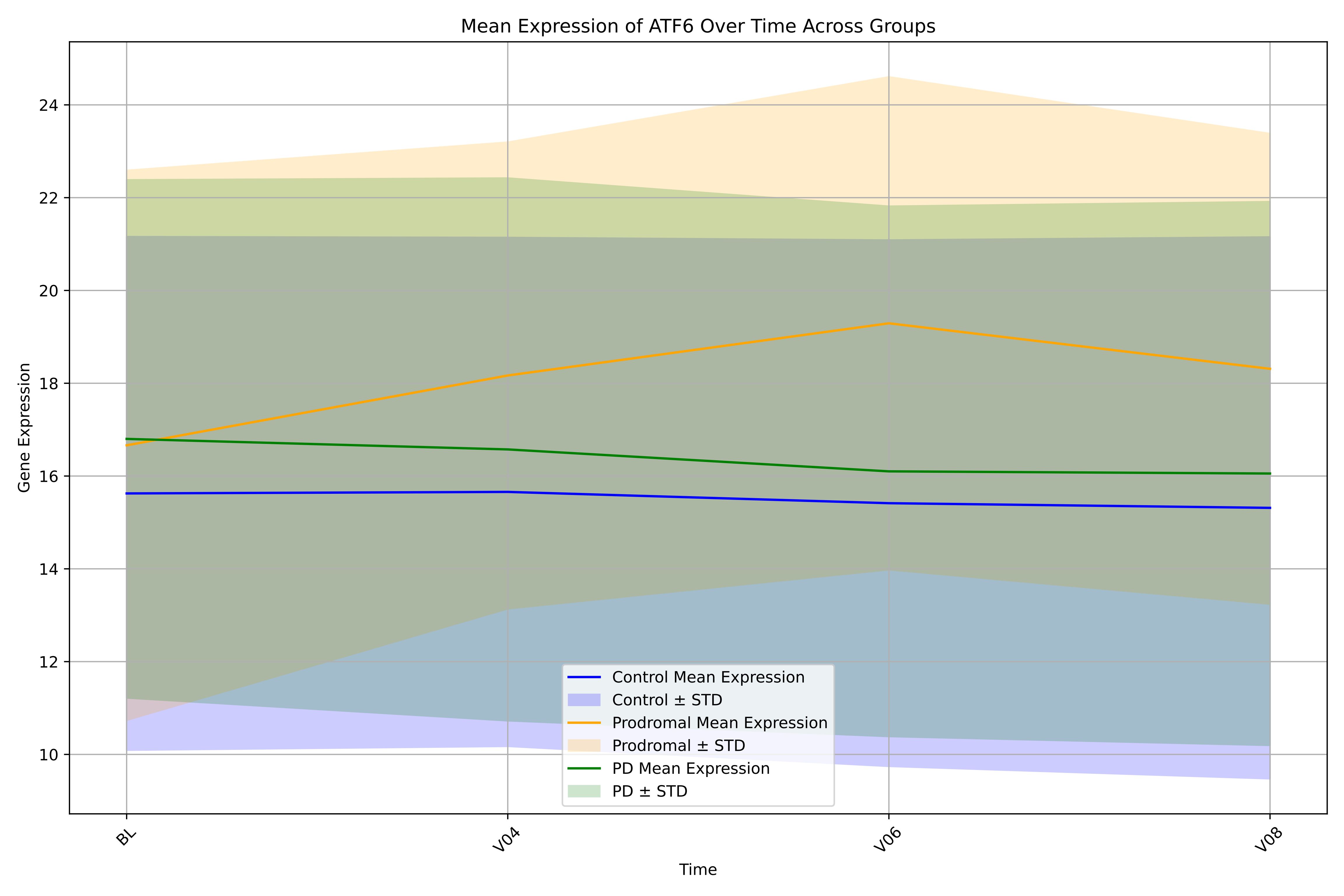

Mean Expression of ATF6 Over Time Across Groups
Gene Expression
Time
24
22
20
18
16
14
12
10
BL
V04
V06
V08
Control Mean Expression
Control ± STD
Prodromal Mean Expression
Prodromal ± STD
PD Mean Expression
PD ± STD

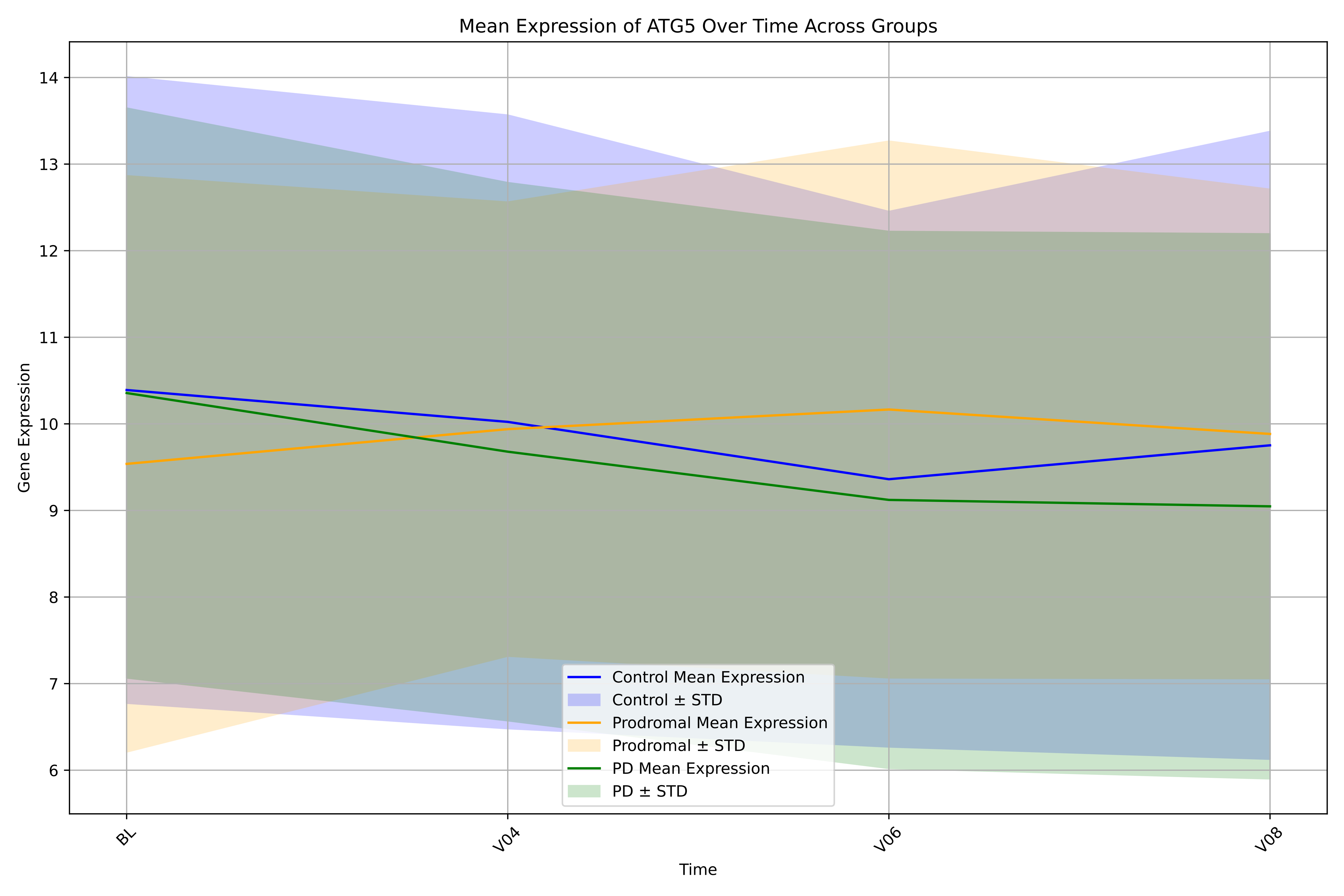
Mean Expression of ATG5 Over Time Across Groups
Gene Expression
Time
BL
V04
V06
V08
14
13
12
11
10
9
8
7
6
Control Mean Expression
Control ± STD
Prodromal Mean Expression
Prodromal ± STD
PD Mean Expression
PD ± STD

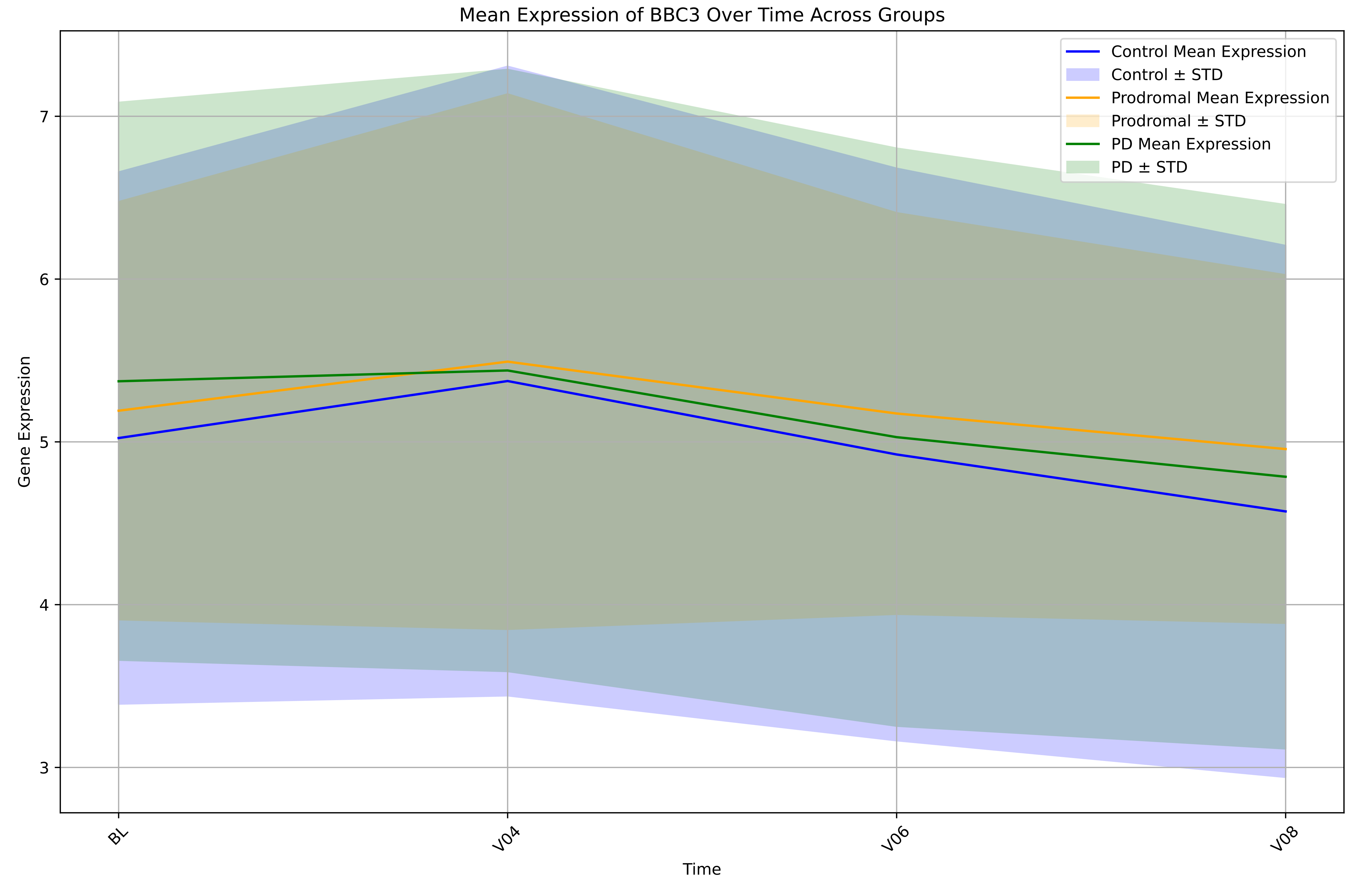

Mean Expression of BBC3 Over Time Across Groups
Control Mean Expression
Control ± STD
Prodromal Mean Expression
Prodromal ± STD
PD Mean Expression
PD ± STD
Gene Expression
7
6
5
4
3
BL
V04
V06
V08
Time

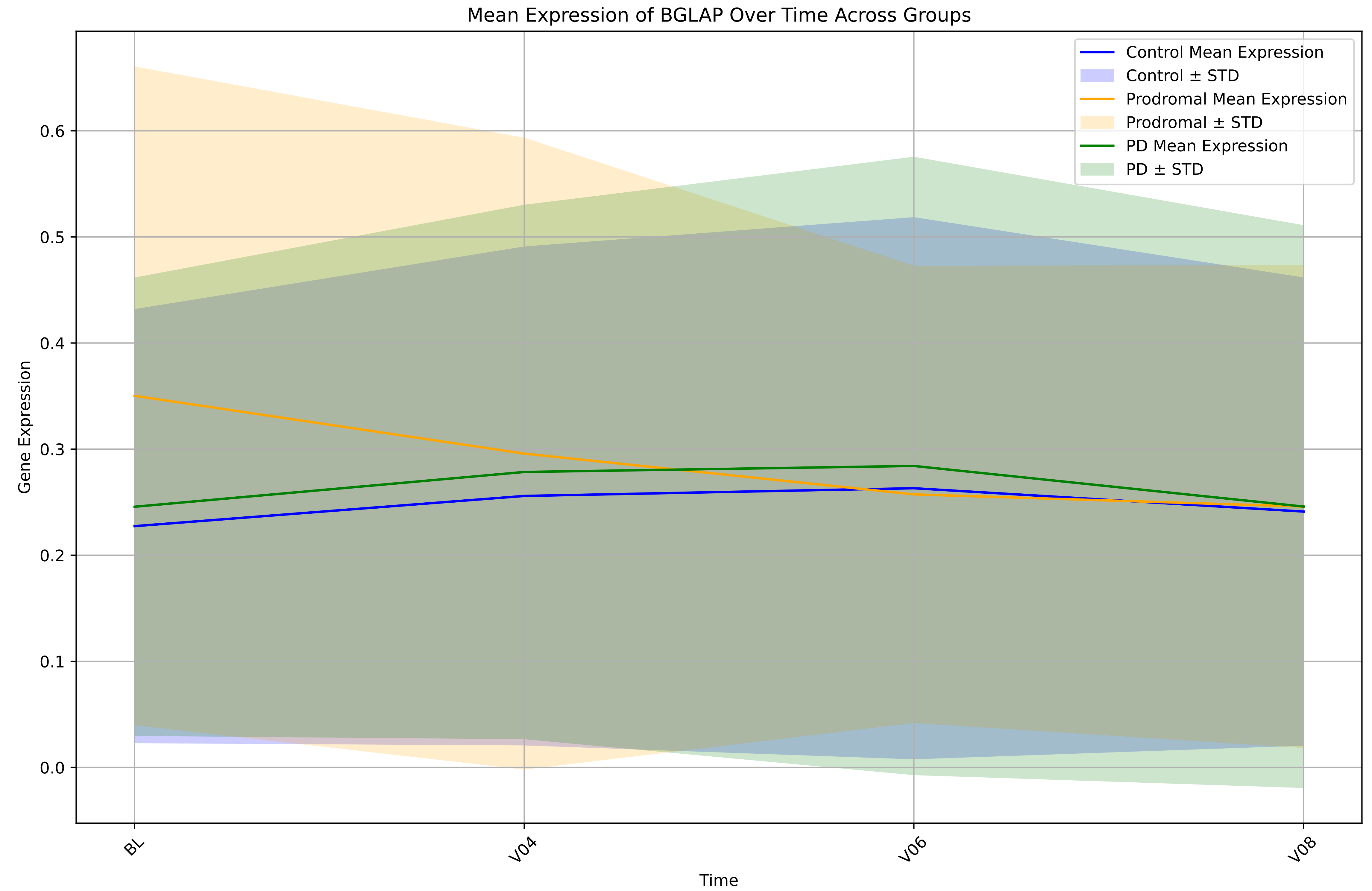
Mean Expression of BGLAP Over Time Across Groups
Control Mean Expression
Control ± STD
Prodromal Mean Expression
Prodromal ± STD
PD Mean Expression
PD ± STD
Gene Expression
0.0
0.1
0.2
0.3
0.4
0.5
0.6
BL
V04
V06
V08
Time

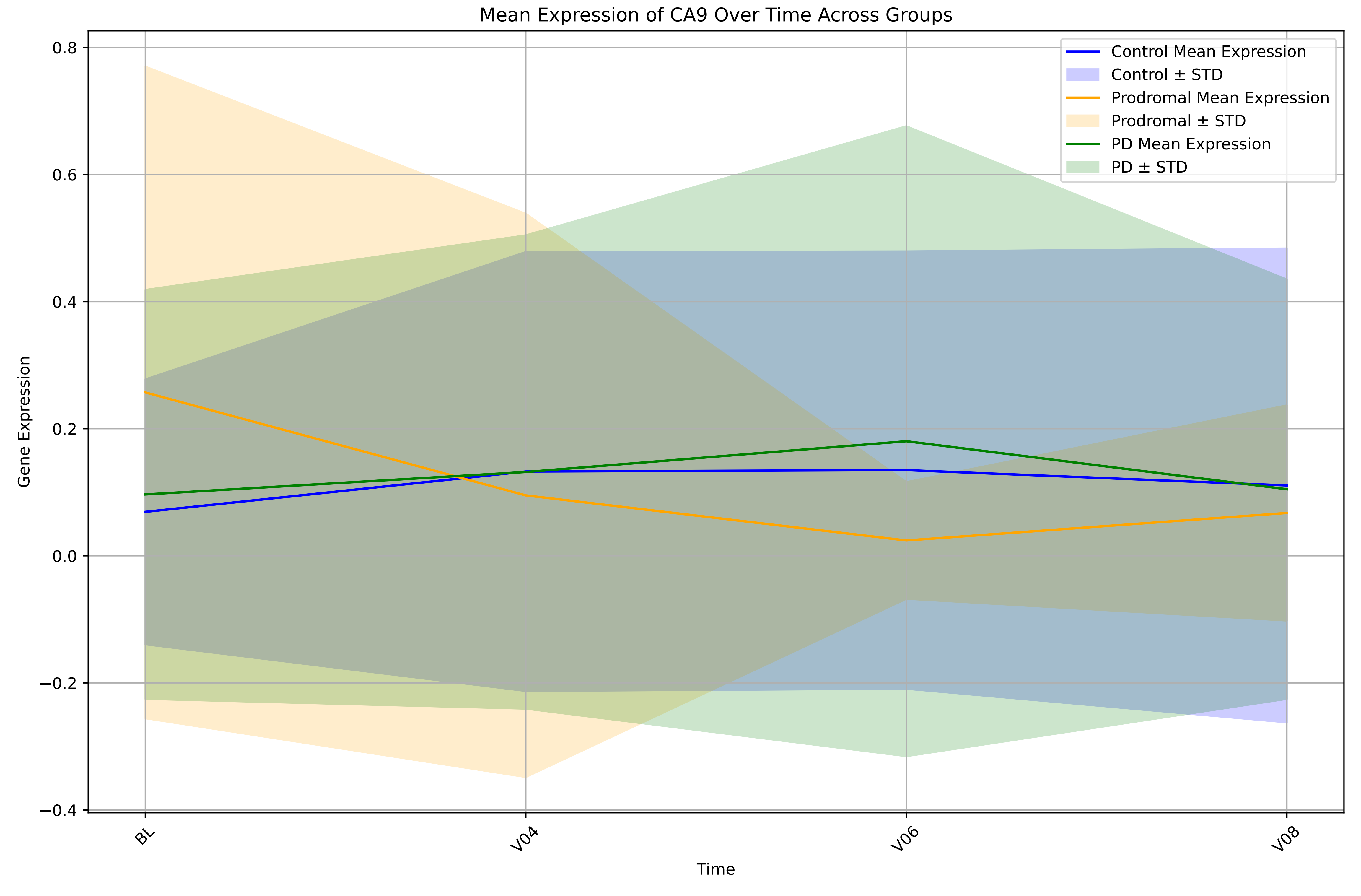

Mean Expression of CA9 Over Time Across Groups
Control Mean Expression
Control ± STD
Prodromal Mean Expression
Prodromal ± STD
PD Mean Expression
PD ± STD
0.8
0.6
0.4
0.2
0.0
−0.2
−0.4
Gene Expression
BL
V04
V06
V08
Time

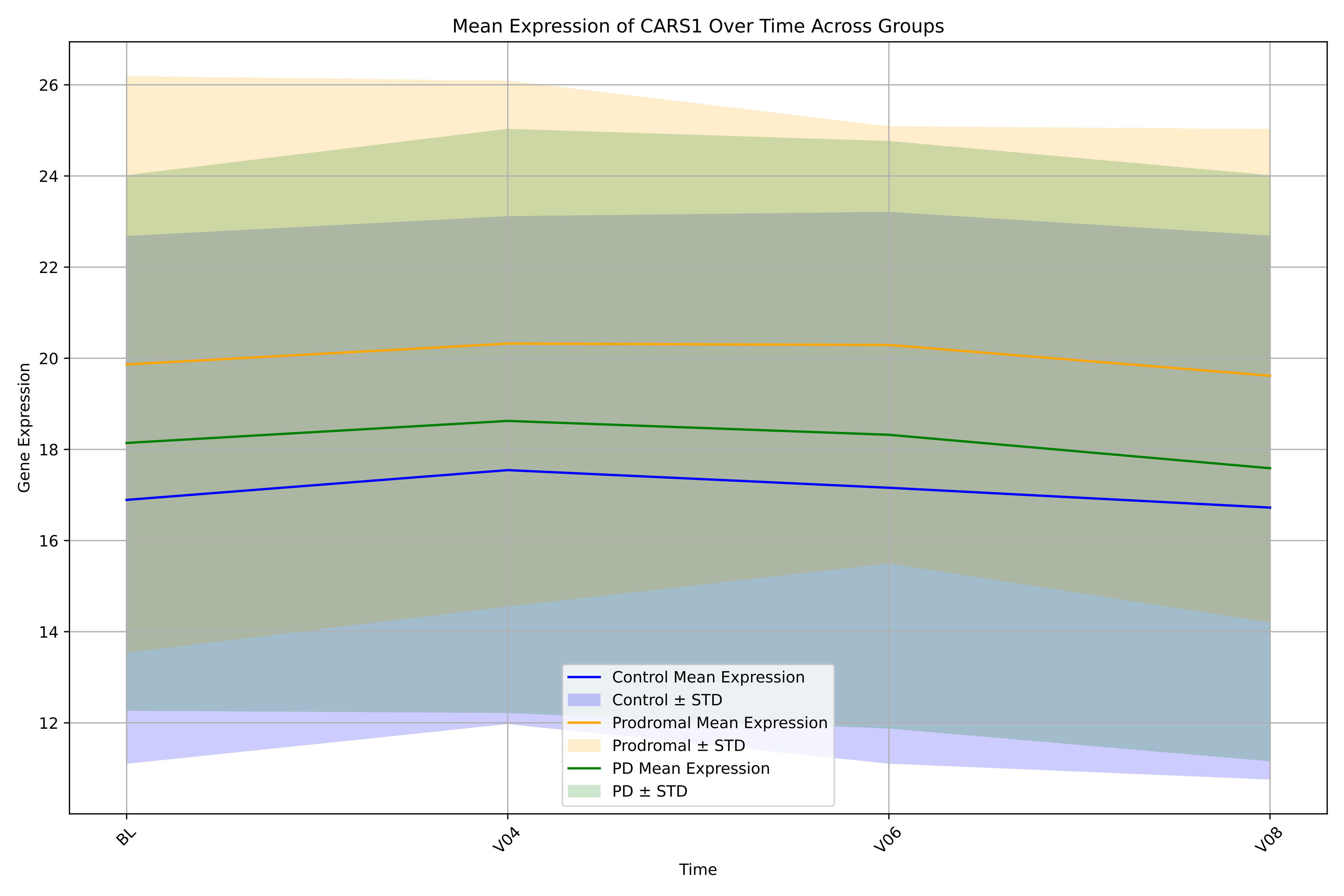

Mean Expression of CARS1 Over Time Across Groups
Gene Expression
Time
BL
V04
V06
V08
26
24
22
20
18
16
14
12
Control Mean Expression
Control ± STD
Prodromal Mean Expression
Prodromal ± STD
PD Mean Expression
PD ± STD

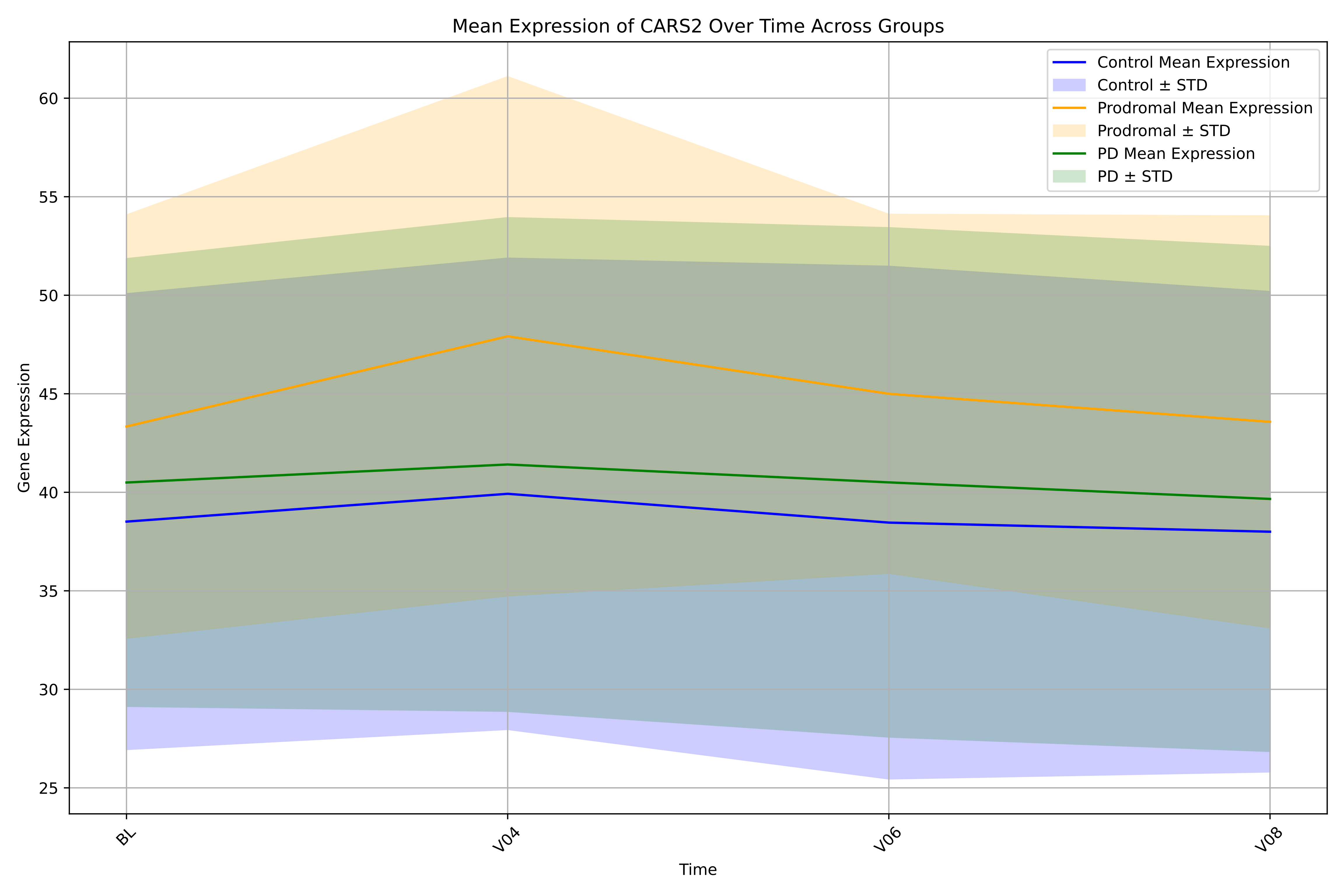

Mean Expression of CARS2 Over Time Across Groups
Control Mean Expression
Control ± STD
Prodromal Mean Expression
Prodromal ± STD
PD Mean Expression
PD ± STD
Gene Expression
Time
BL
V04
V06
V08
25
30
35
40
45
50
55
60

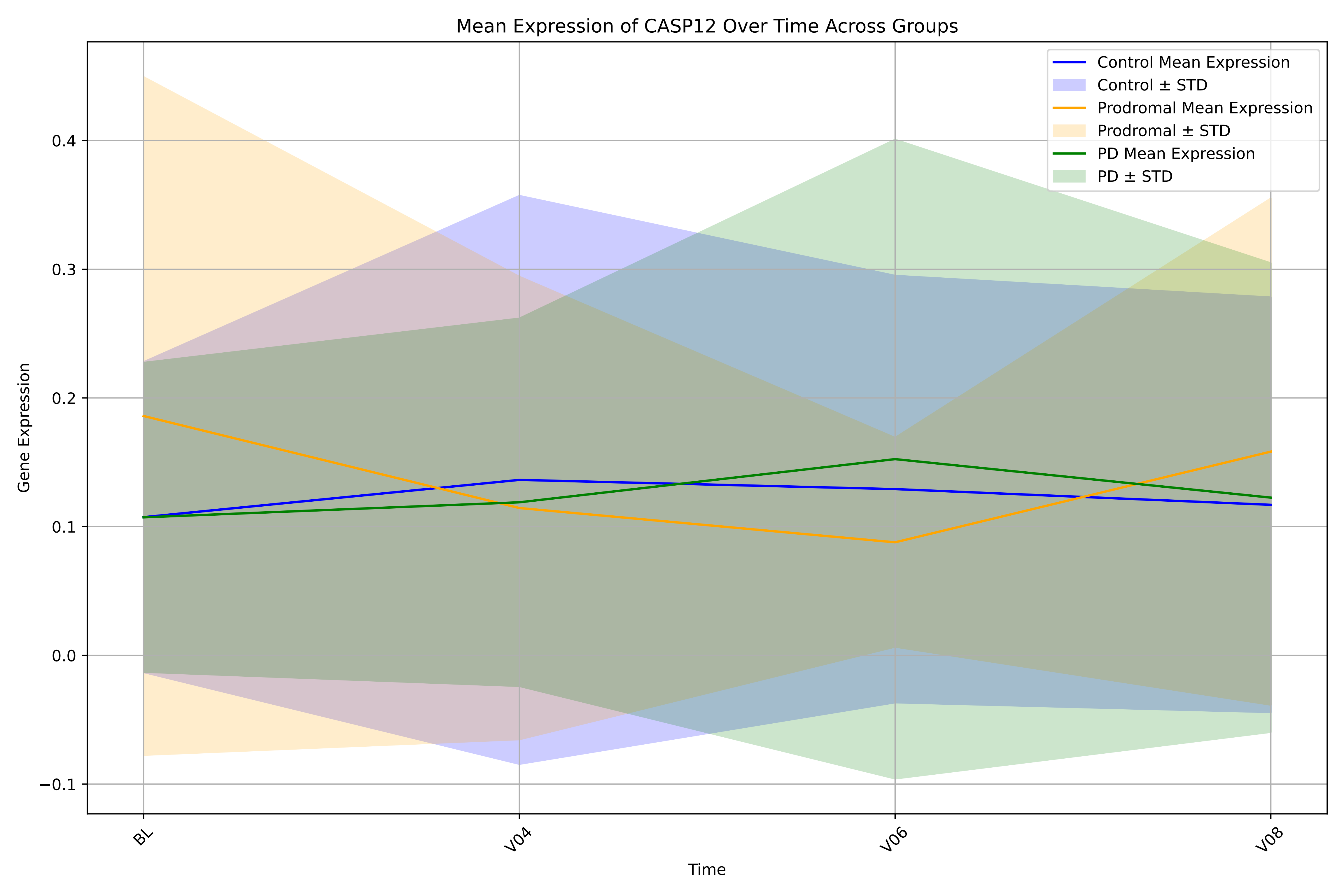

Mean Expression of CASP12 Over Time Across Groups
Control Mean Expression
Control ± STD
Prodromal Mean Expression
Prodromal ± STD
PD Mean Expression
PD ± STD
Gene Expression
0.4
0.3
0.2
0.1
0.0
−0.1
BL
V04
V06
V08
Time

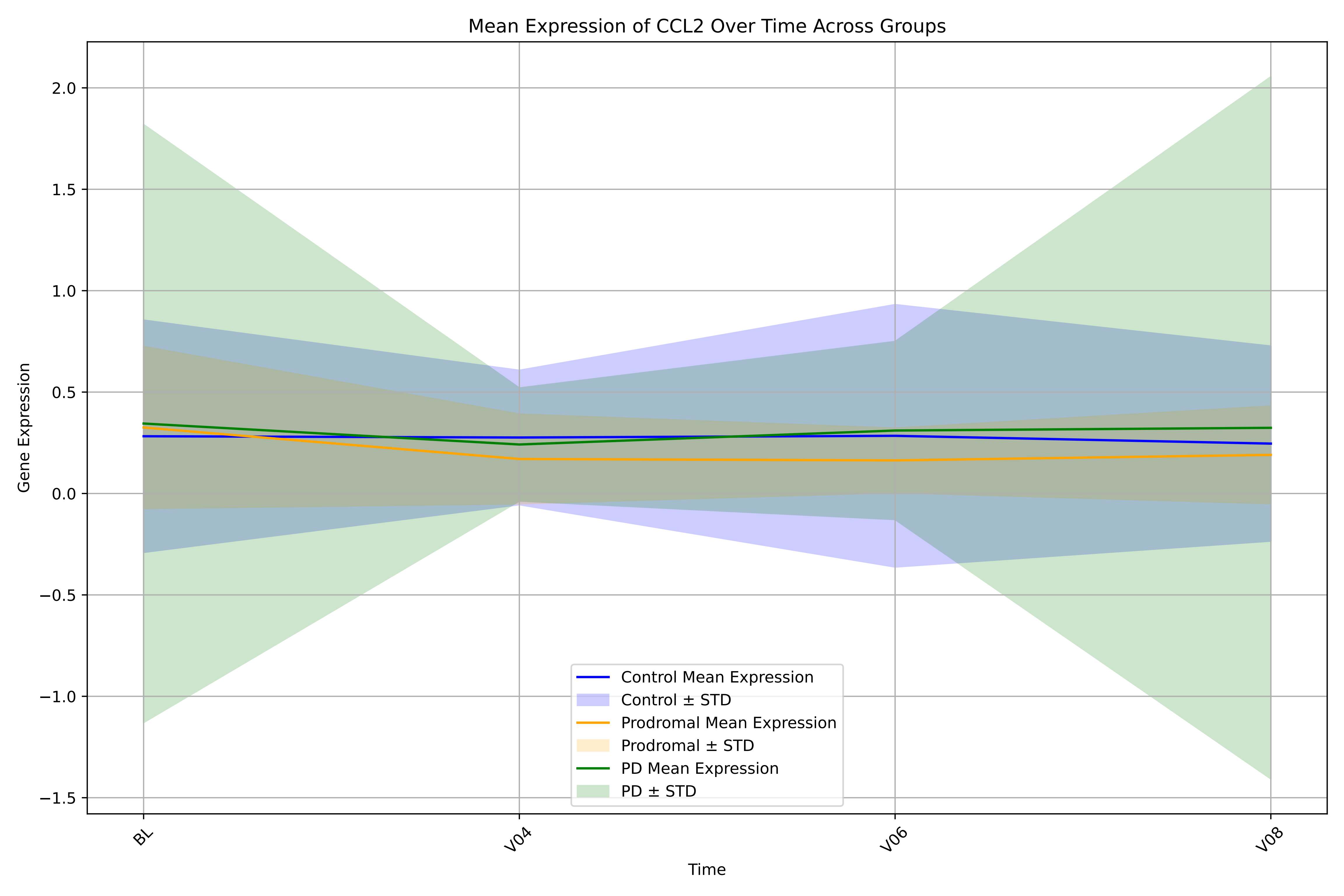
Mean Expression of CCL2 Over Time Across Groups
Gene Expression
Time
BL
V04
V06
V08
2.0
1.5
1.0
0.5
0.0
−0.5
−1.0
−1.5
Control Mean Expression
Control ± STD
Prodromal Mean Expression
Prodromal ± STD
PD Mean Expression
PD ± STD

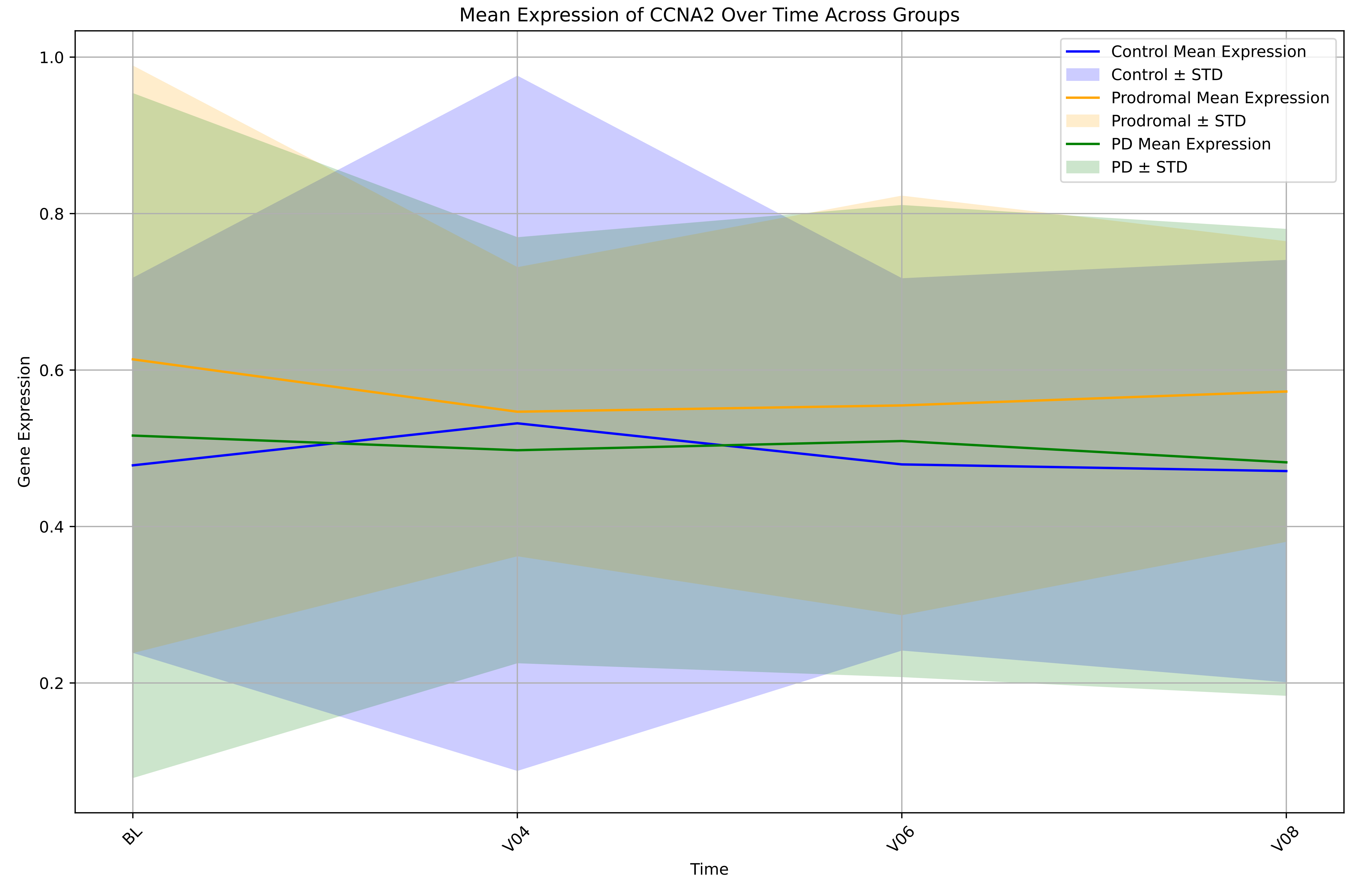

Mean Expression of CCNA2 Over Time Across Groups
Control Mean Expression
Control ± STD
Prodromal Mean Expression
Prodromal ± STD
PD Mean Expression
PD ± STD
Gene Expression
1.0
0.8
0.6
0.4
0.2
BL
V04
V06
V08
Time

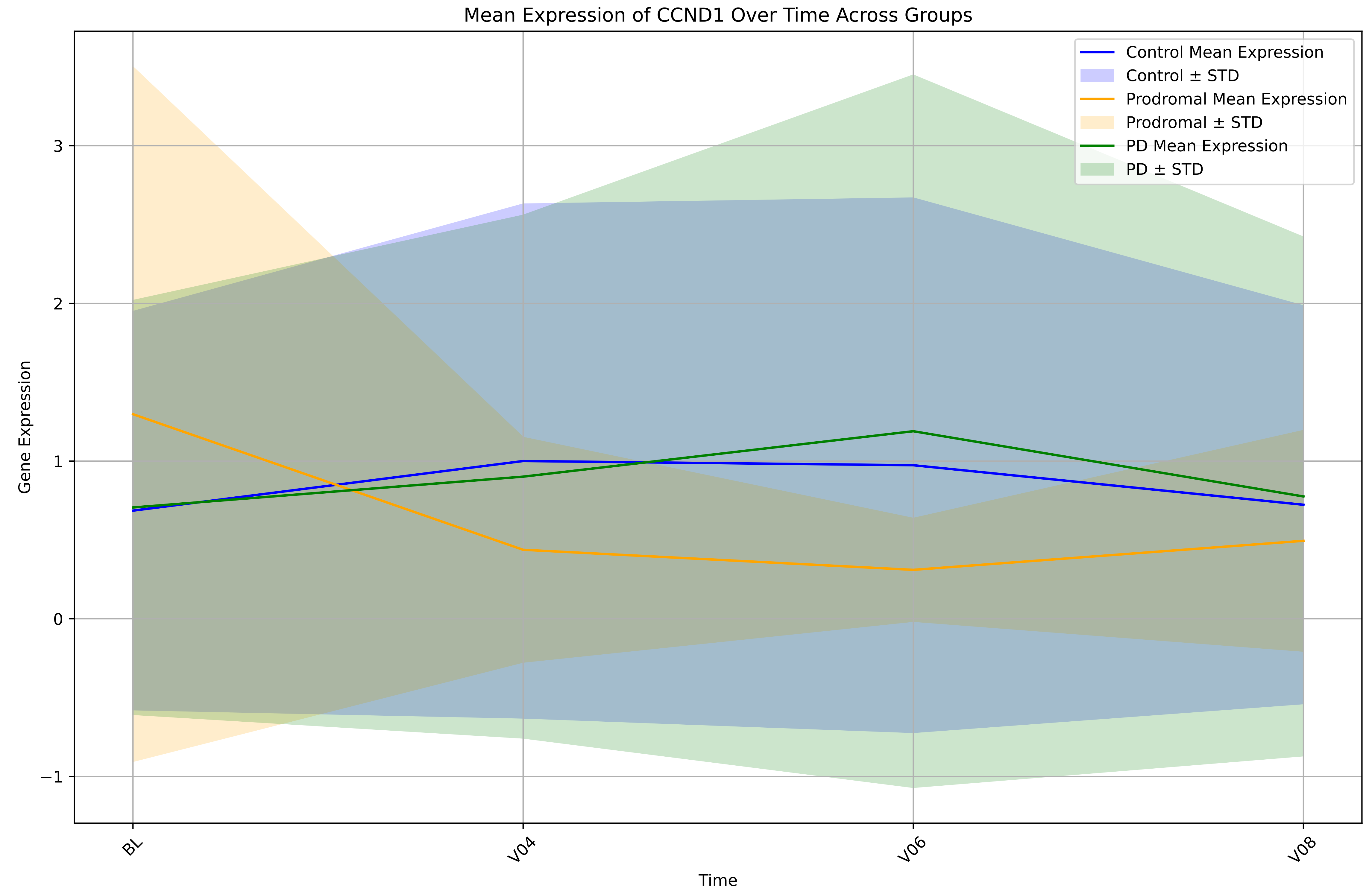

Mean Expression of CCND1 Over Time Across Groups
Control Mean Expression
Control ± STD
Prodromal Mean Expression
Prodromal ± STD
PD Mean Expression
PD ± STD
Gene Expression
3
2
1
0
−1
BL
V04
V06
V08
Time

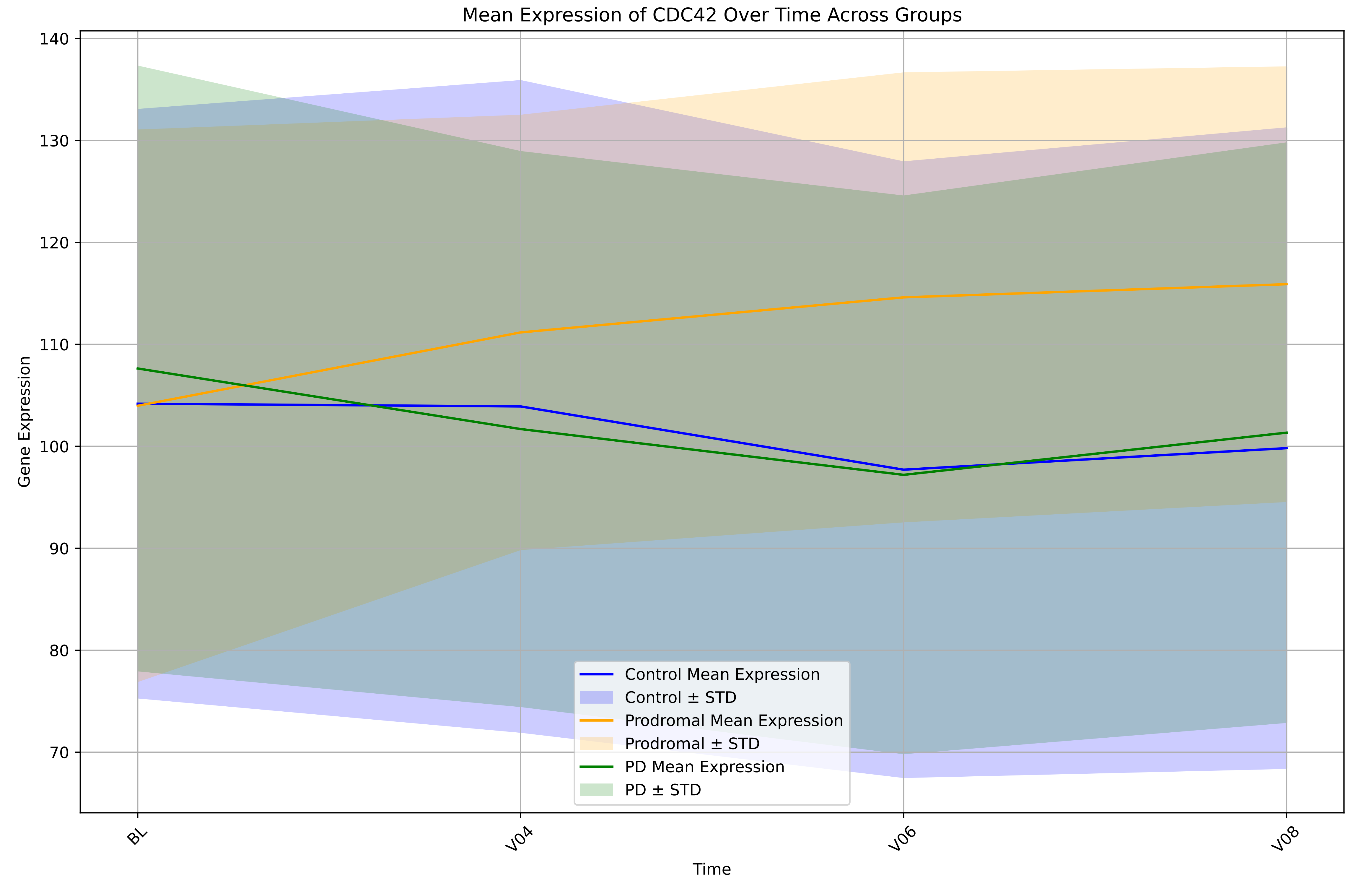
Mean Expression of CDC42 Over Time Across Groups
Gene Expression
Time
BL
V04
V06
V08
140
130
120
110
100
90
80
70
Control Mean Expression
Control ± STD
Prodromal Mean Expression
Prodromal ± STD
PD Mean Expression
PD ± STD

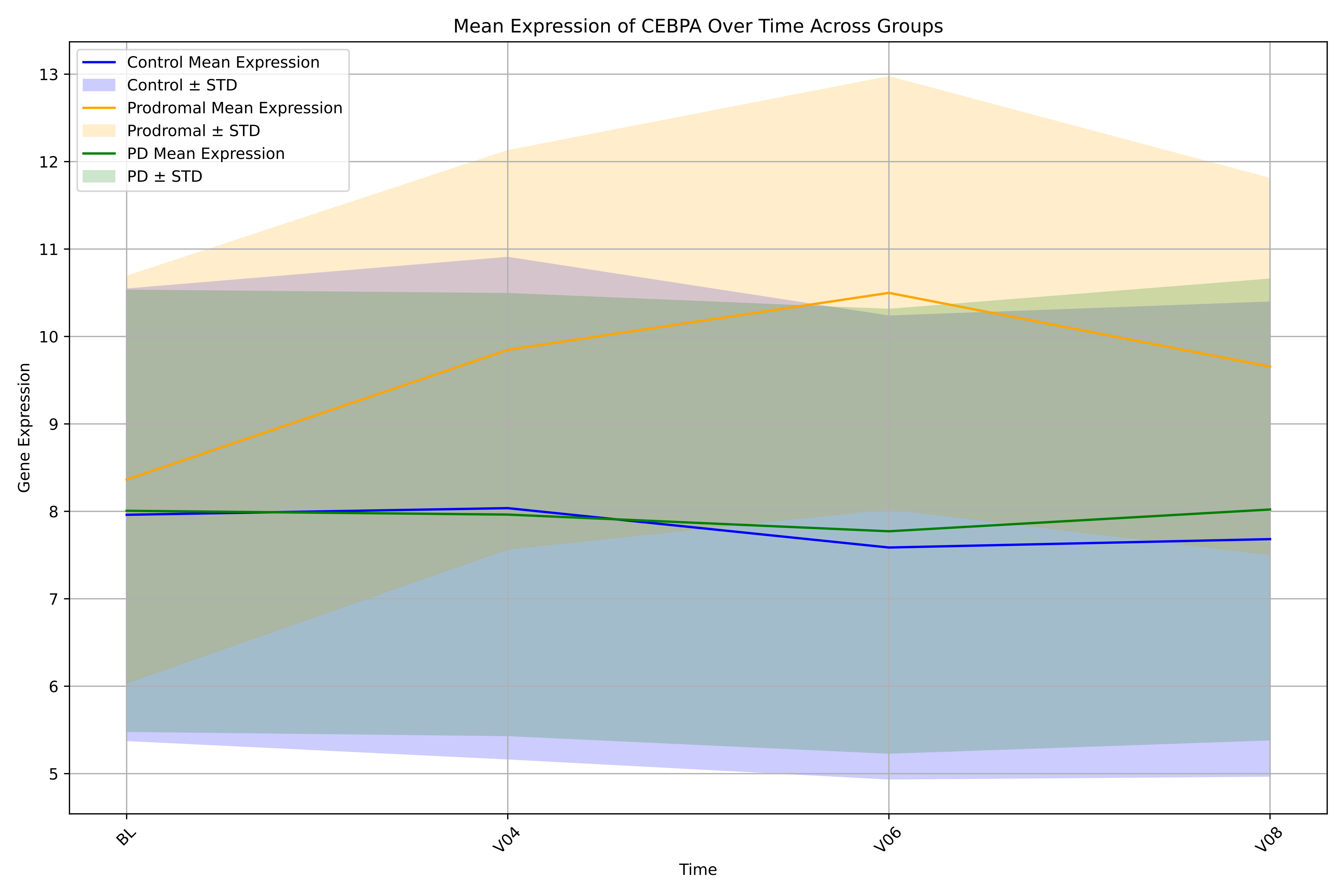

Mean Expression of CEBPA Over Time Across Groups
Control Mean Expression
Control ± STD
Prodromal Mean Expression
Prodromal ± STD
PD Mean Expression
PD ± STD
Gene Expression
Time
BL
V04
V06
V08
5
6
7
8
9
10
11
12
13

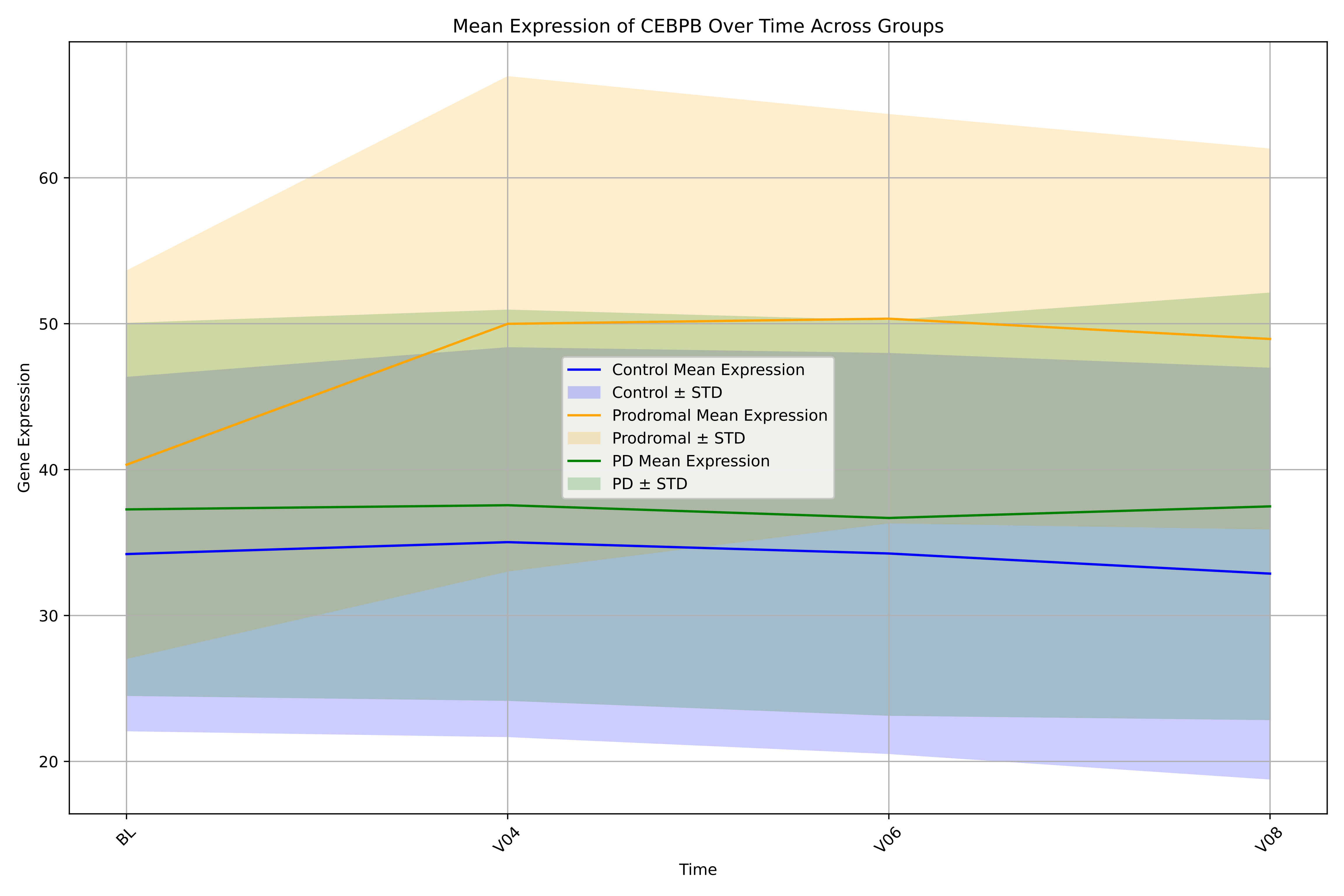

Mean Expression of CEBPB Over Time Across Groups
Gene Expression
Time
20
30
40
50
60
BL
V04
V06
V08
Control Mean Expression
Control ± STD
Prodromal Mean Expression
Prodromal ± STD
PD Mean Expression
PD ± STD

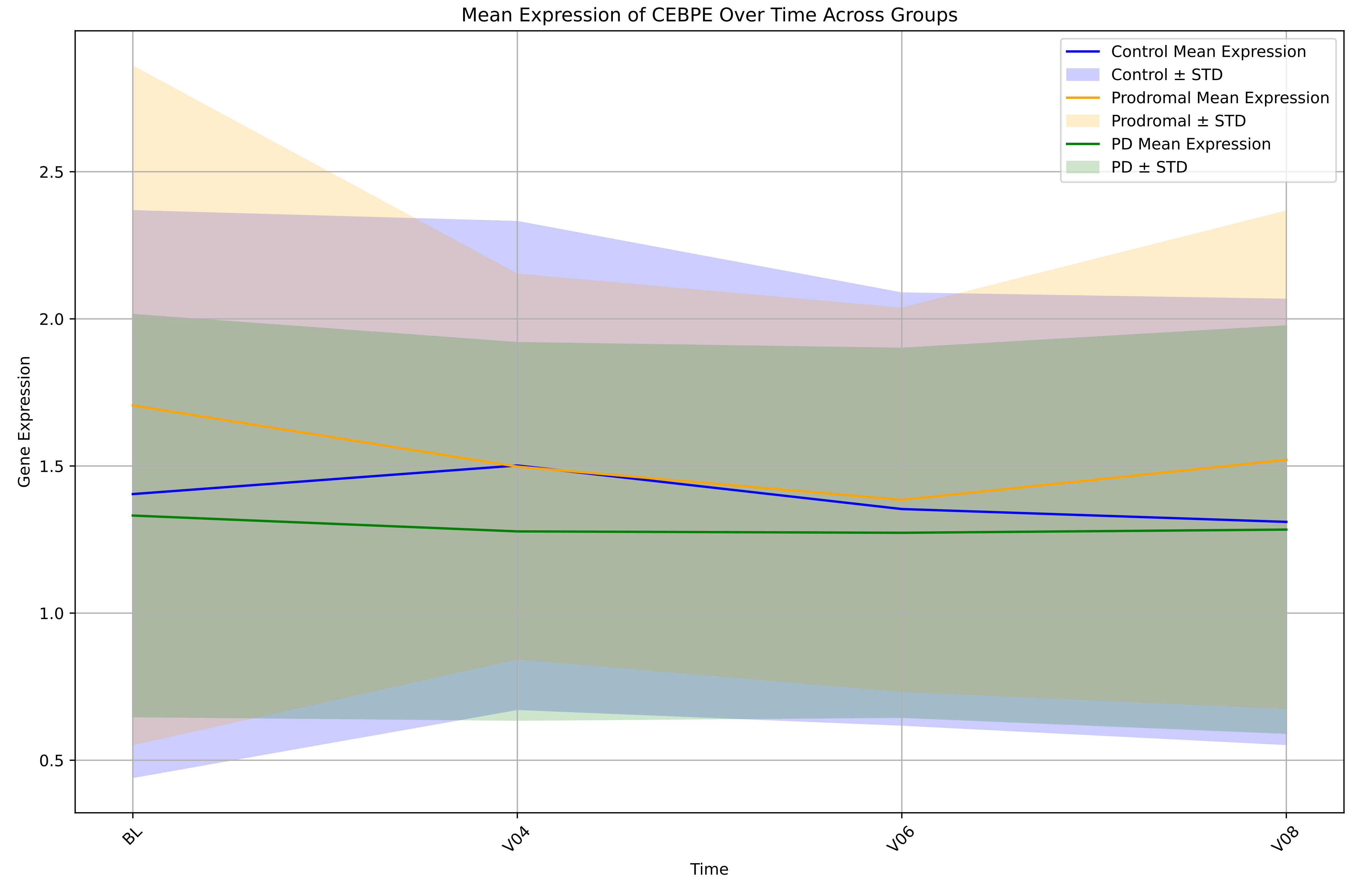

Mean Expression of CEBPE Over Time Across Groups
Control Mean Expression
Control ± STD
Prodromal Mean Expression
Prodromal ± STD
PD Mean Expression
PD ± STD
Gene Expression
2.5
2.0
1.5
1.0
0.5
BL
V04
V06
V08
Time

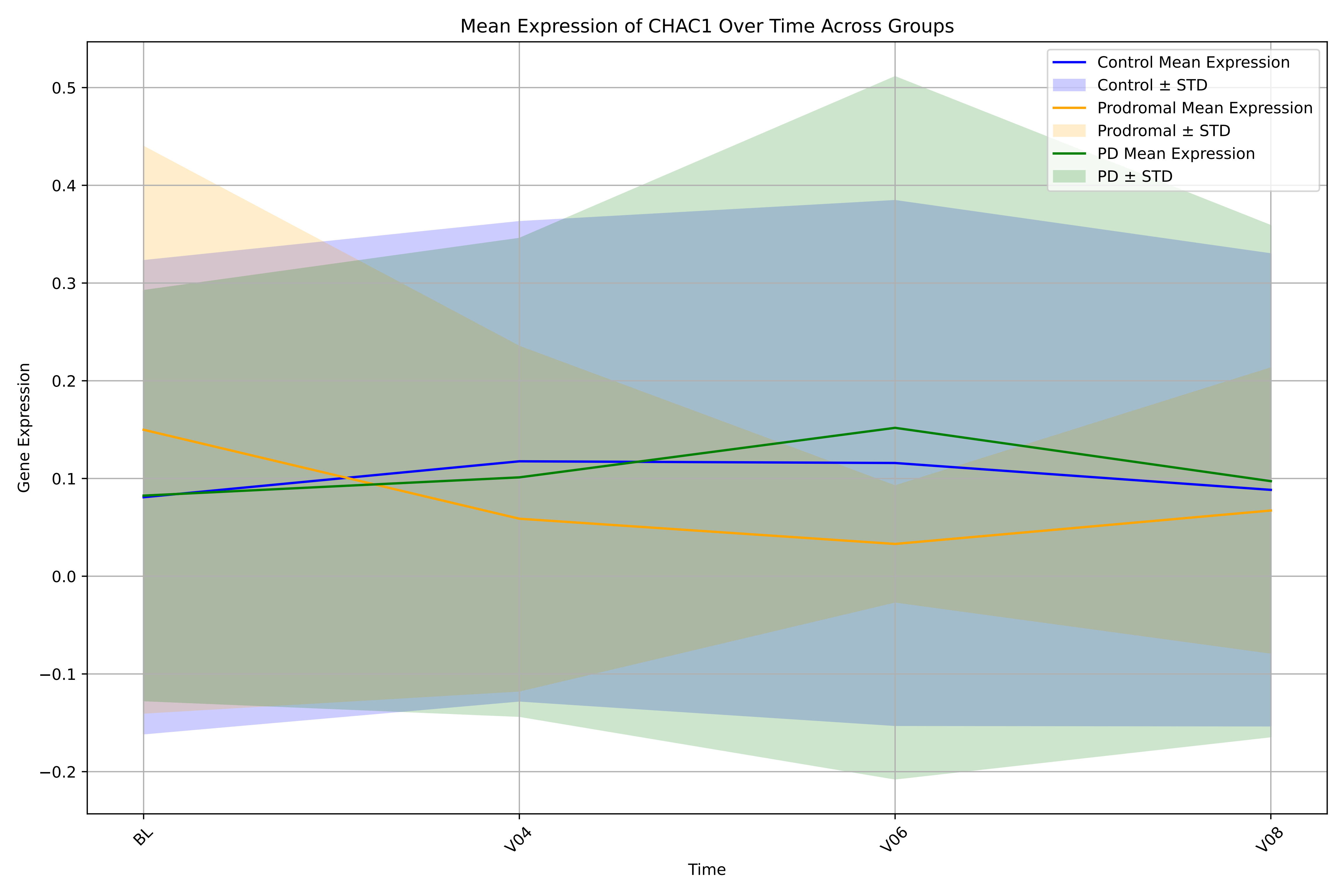

Mean Expression of CHAC1 Over Time Across Groups
Control Mean Expression
Control ± STD
Prodromal Mean Expression
Prodromal ± STD
PD Mean Expression
PD ± STD
Gene Expression
0.5
0.4
0.3
0.2
0.1
0.0
−0.1
−0.2
BL
V04
V06
V08
Time

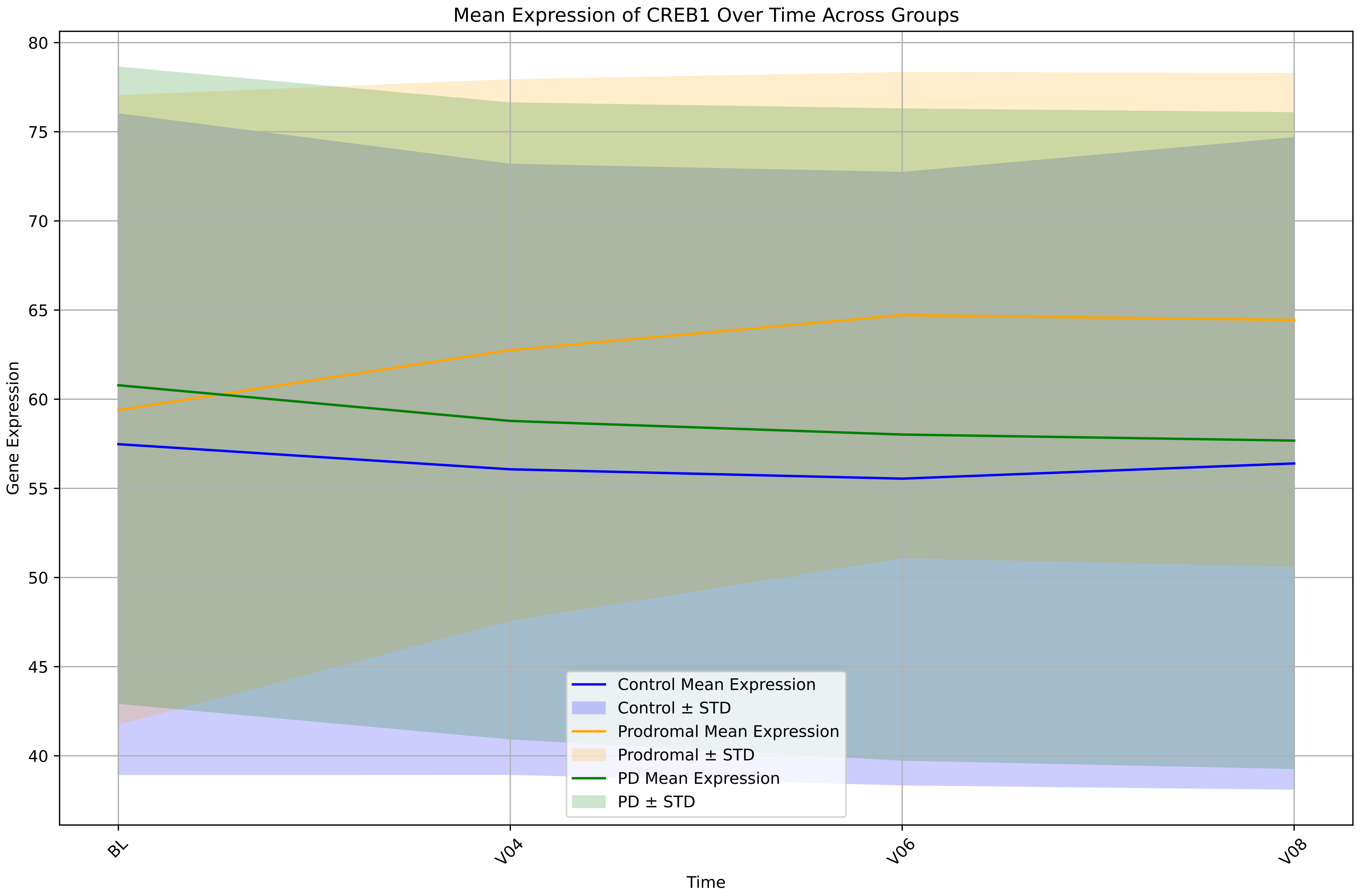

Mean Expression of CREB1 Over Time Across Groups
Gene Expression
Time
80
75
70
65
60
55
50
45
40
BL
V04
V06
V08
Control Mean Expression
Control ± STD
Prodromal Mean Expression
Prodromal ± STD
PD Mean Expression
PD ± STD

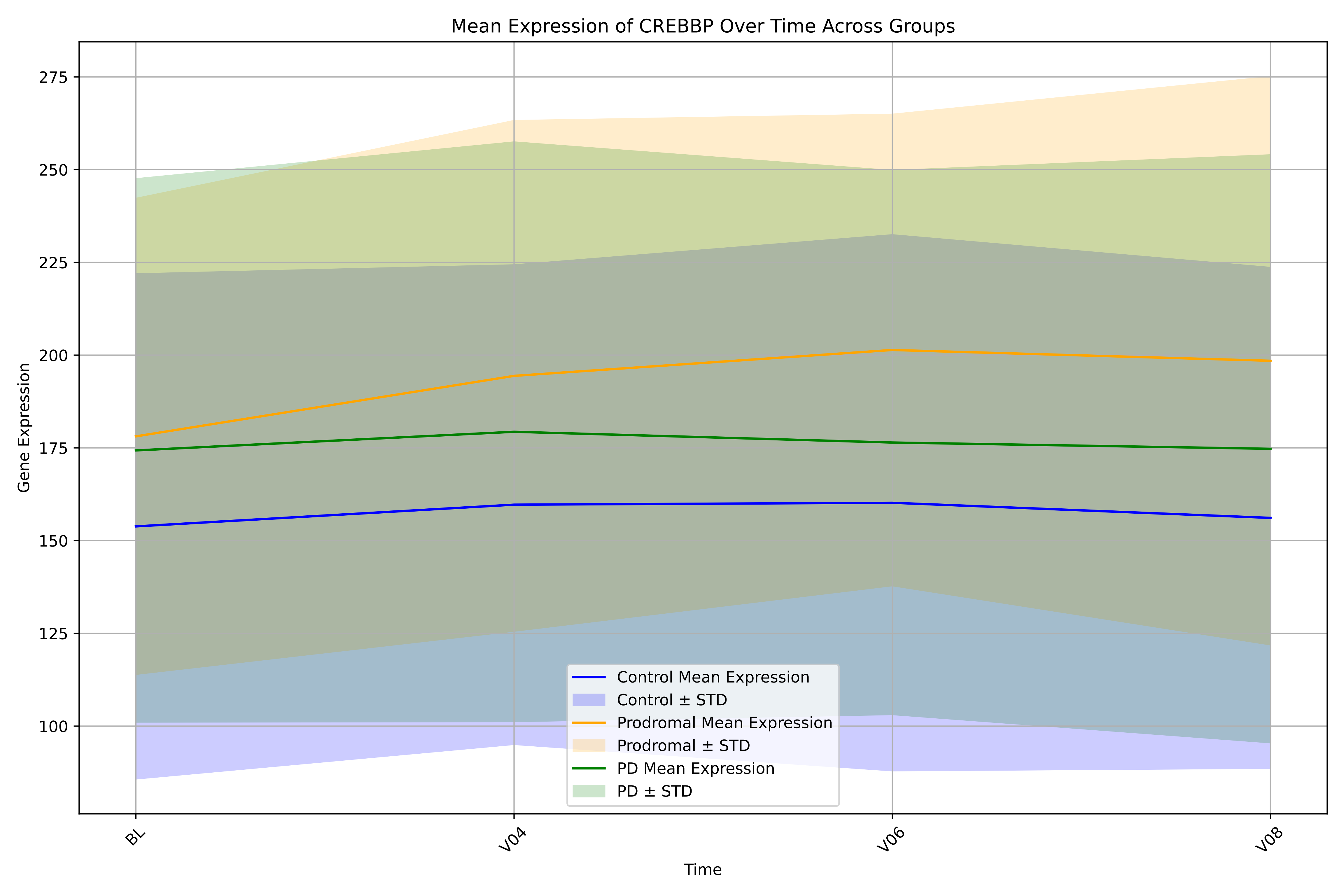
Mean Expression of CREBBP Over Time Across Groups
Gene Expression
Time
275
250
225
200
175
150
125
100
BL
V04
V06
V08
Control Mean Expression
Control ± STD
Prodromal Mean Expression
Prodromal ± STD
PD Mean Expression
PD ± STD

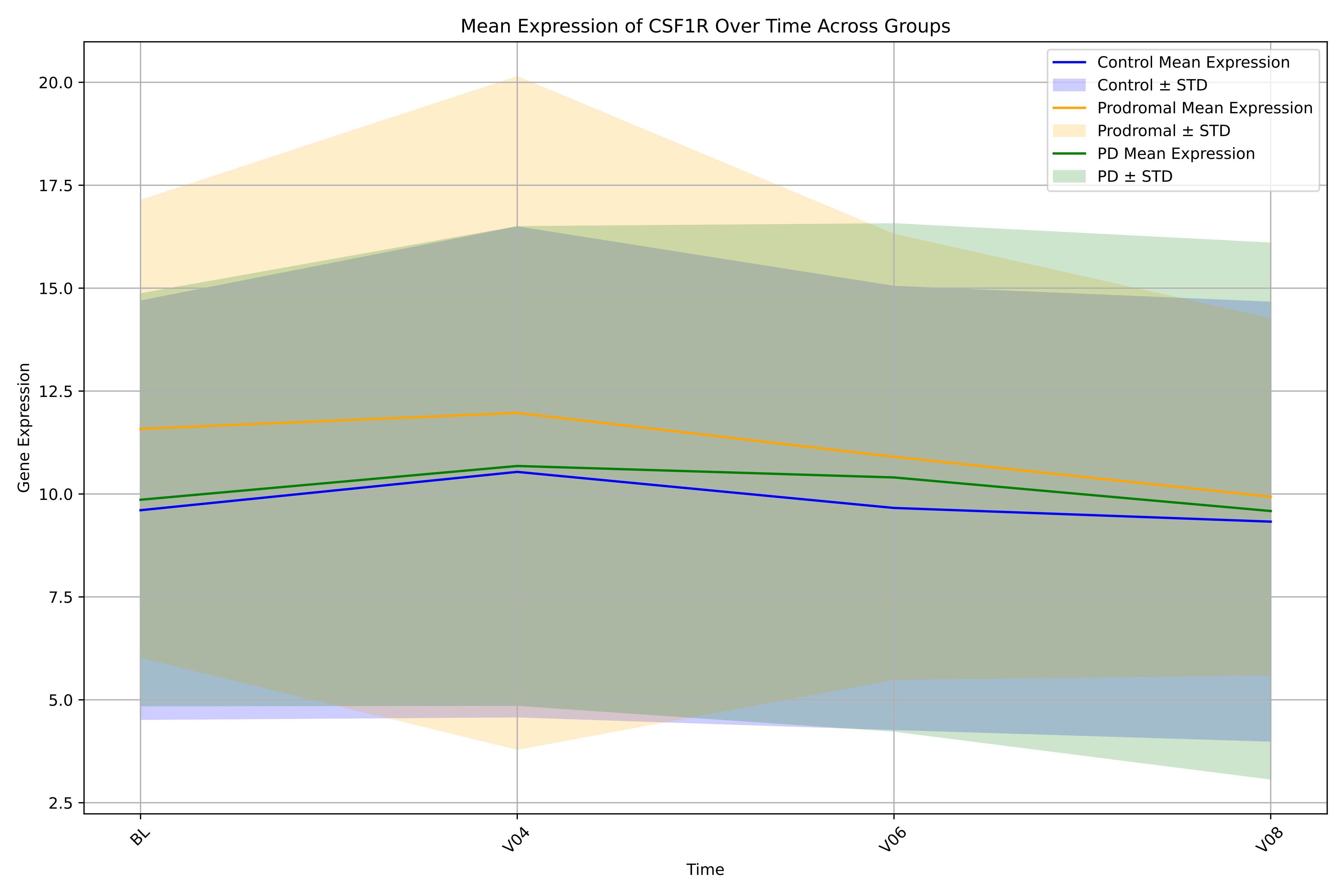

Mean Expression of CSF1R Over Time Across Groups
Control Mean Expression
Control ± STD
Prodromal Mean Expression
Prodromal ± STD
PD Mean Expression
PD ± STD
Gene Expression
20.0
17.5
15.0
12.5
10.0
7.5
5.0
2.5
BL
V04
V06
V08
Time

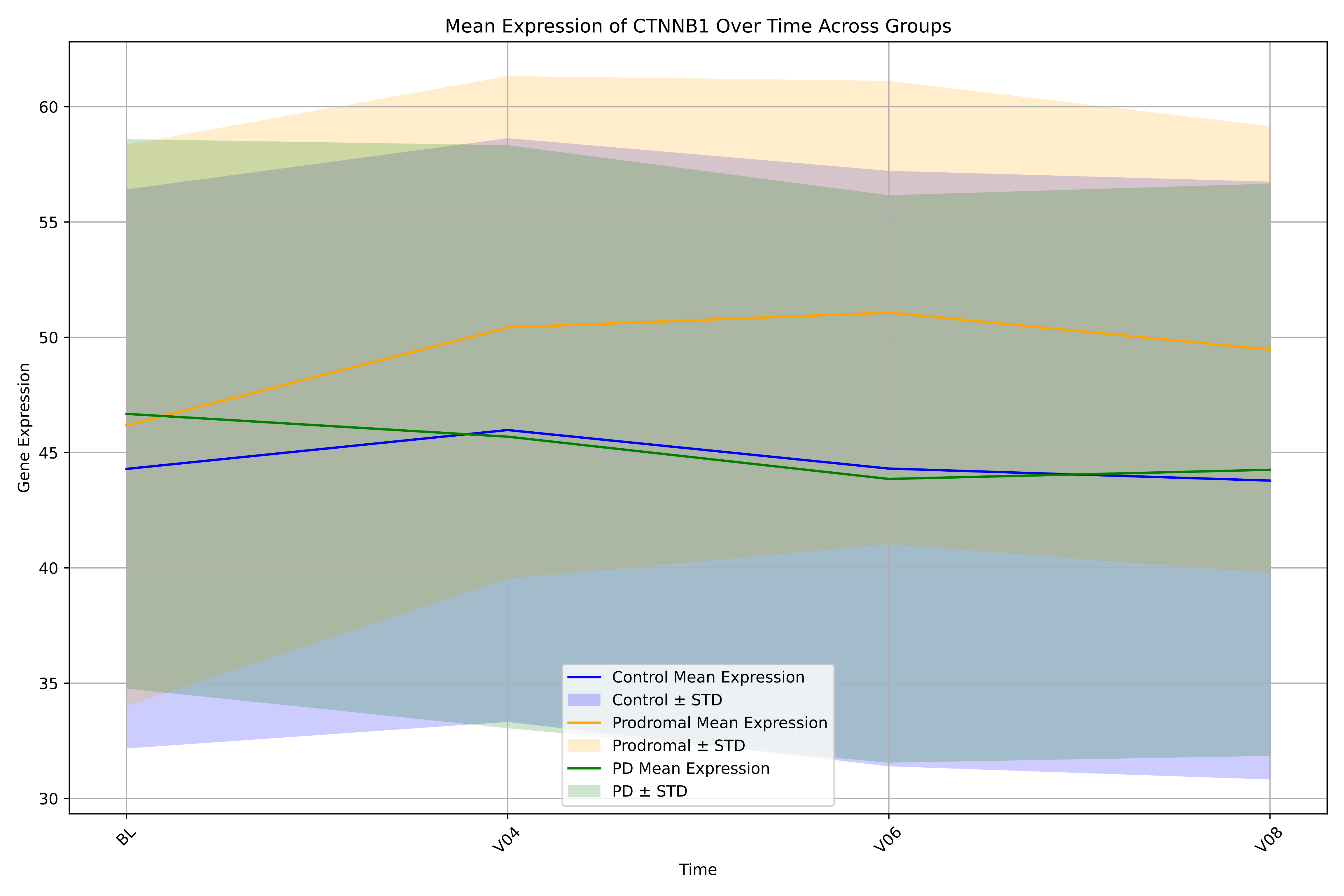

Mean Expression of CTNNB1 Over Time Across Groups
Gene Expression
Time
BL
V04
V06
V08
30
35
40
45
50
55
60
Control Mean Expression
Control ± STD
Prodromal Mean Expression
Prodromal ± STD
PD Mean Expression
PD ± STD

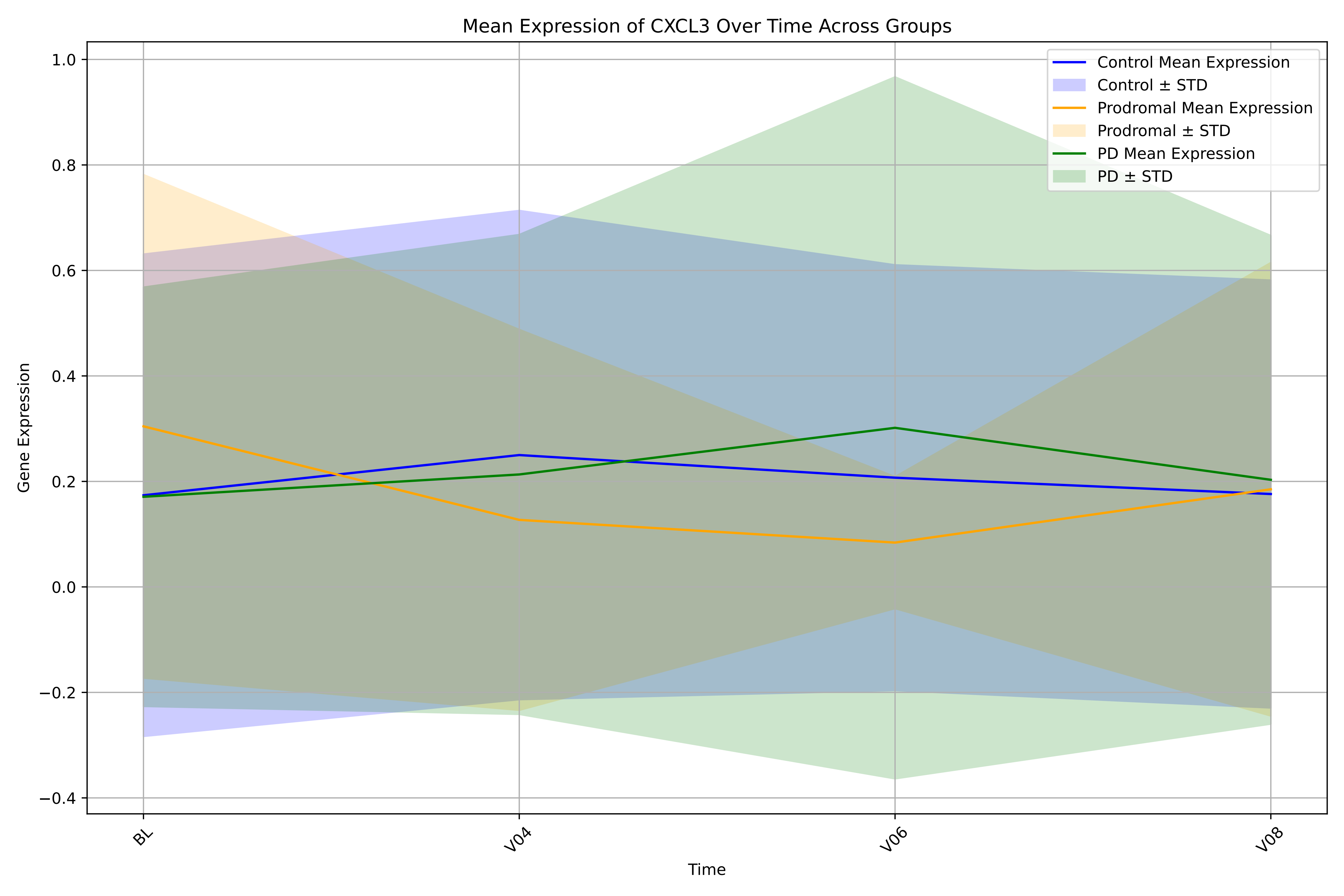

Mean Expression of CXCL3 Over Time Across Groups
Control Mean Expression
Control ± STD
Prodromal Mean Expression
Prodromal ± STD
PD Mean Expression
PD ± STD
Gene Expression
1.0
0.8
0.6
0.4
0.2
0.0
−0.2
−0.4
BL
V04
V06
V08
Time

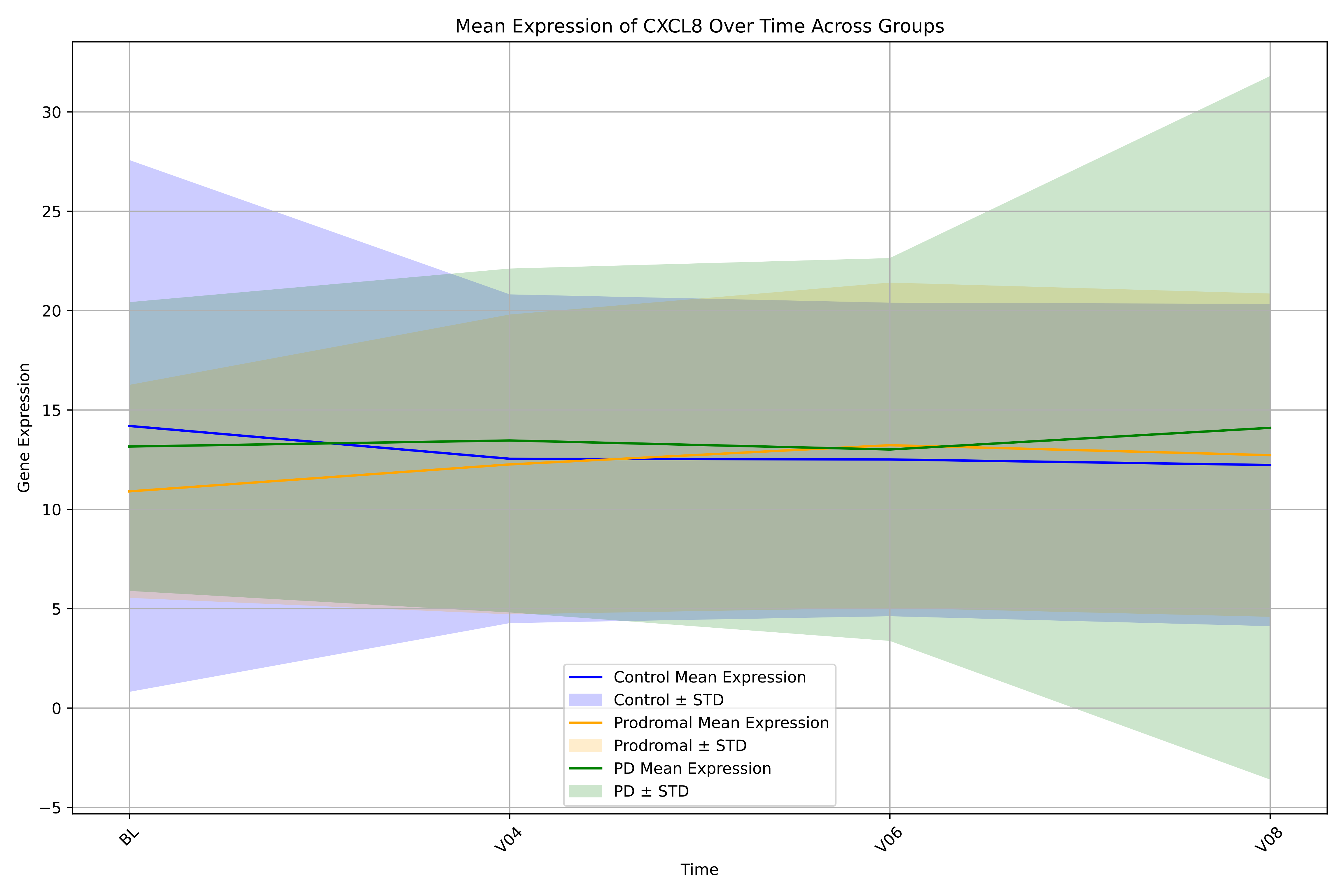

Mean Expression of CXCL8 Over Time Across Groups
Gene Expression
Time
BL
V04
V06
V08
30
25
20
15
10
5
0
−5
Control Mean Expression
Control ± STD
Prodromal Mean Expression
Prodromal ± STD
PD Mean Expression
PD ± STD

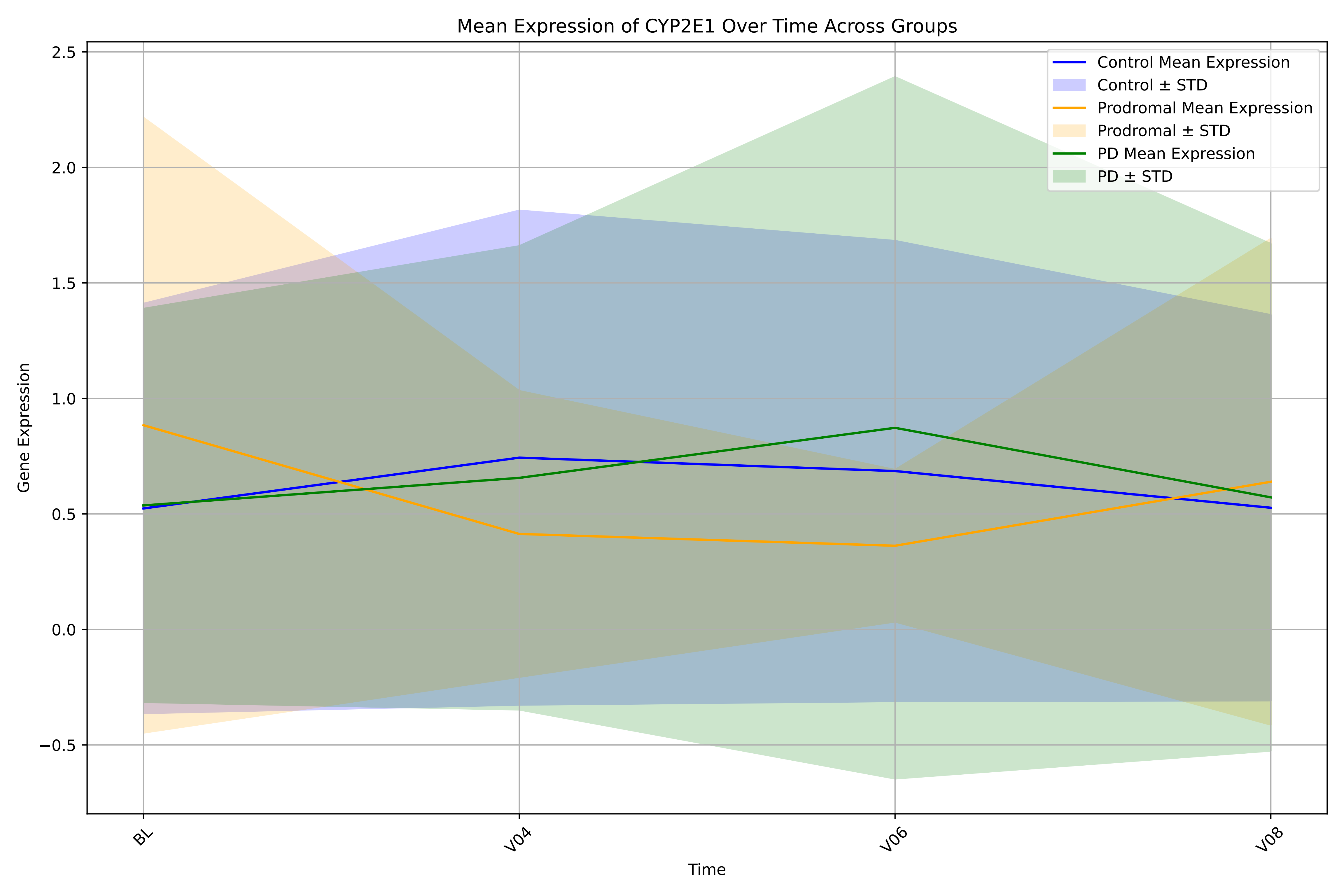

Mean Expression of CYP2E1 Over Time Across Groups
Control Mean Expression
Control ± STD
Prodromal Mean Expression
Prodromal ± STD
PD Mean Expression
PD ± STD
Gene Expression
2.5
2.0
1.5
1.0
0.5
0.0
−0.5
BL
V04
V06
V08
Time

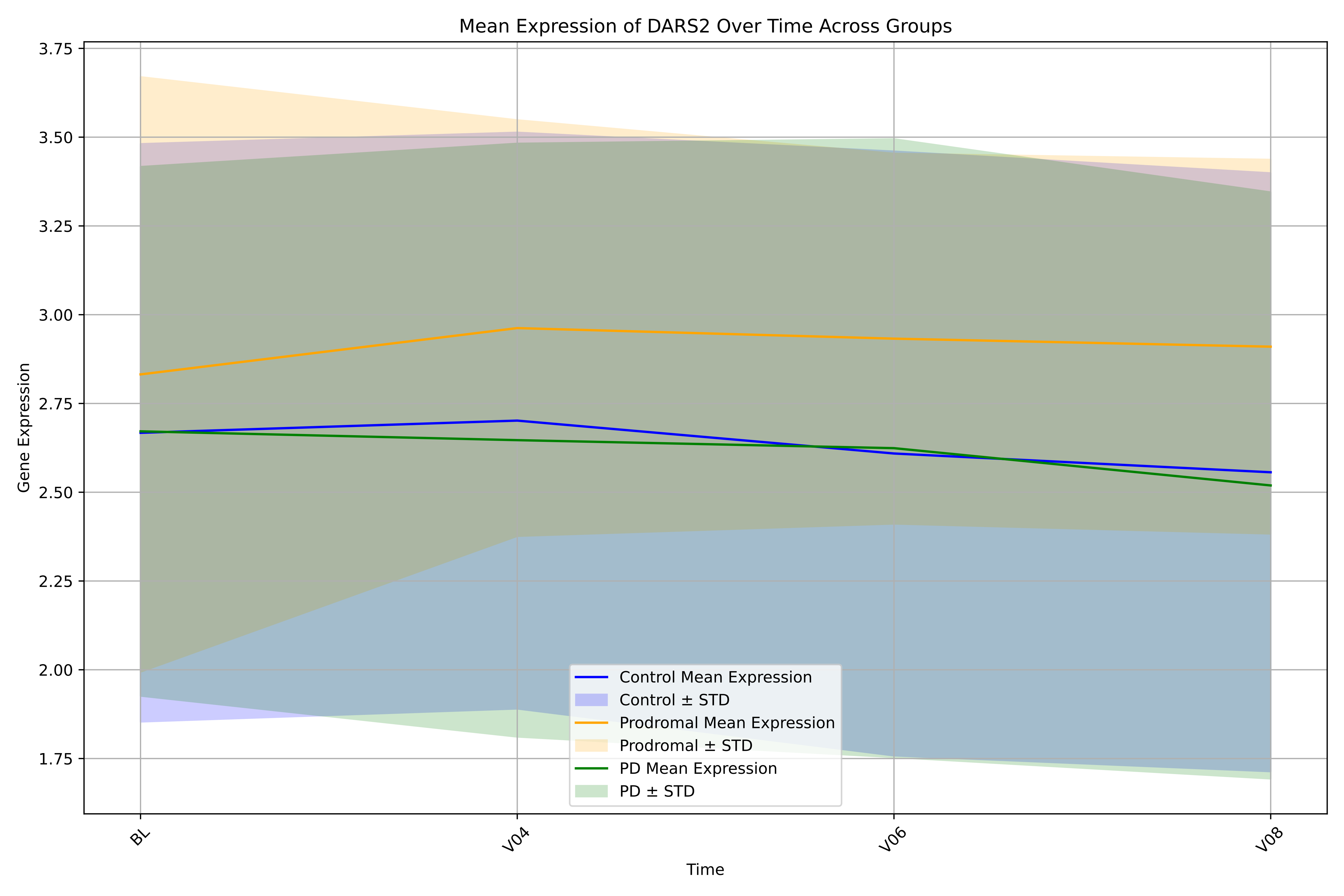

Mean Expression of DARS2 Over Time Across Groups
3.75
3.50
3.25
3.00
2.75
2.50
2.25
2.00
1.75
Gene Expression
BL
V04
V06
V08
Time
Control Mean Expression
Control ± STD
Prodromal Mean Expression
Prodromal ± STD
PD Mean Expression
PD ± STD

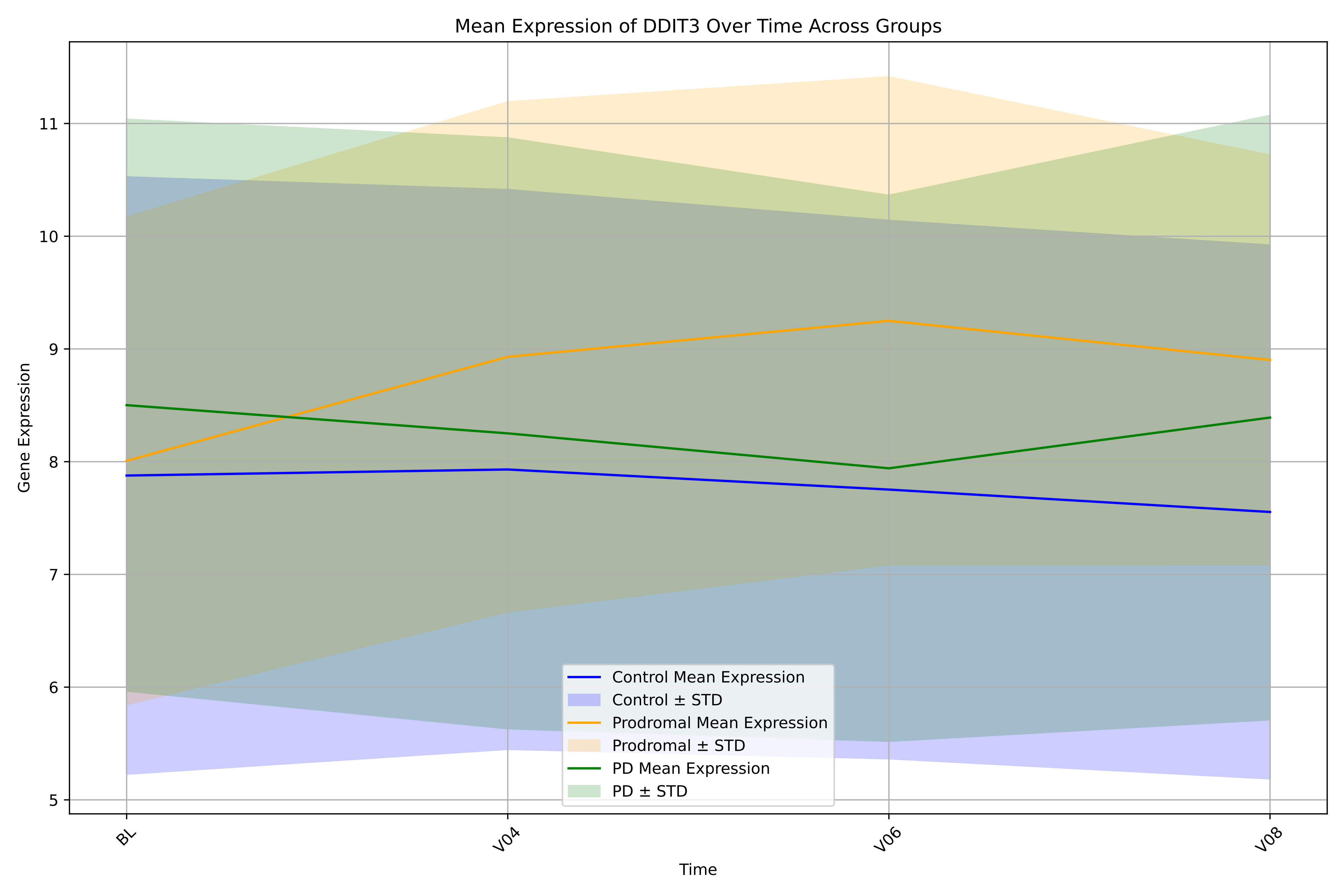

Mean Expression of DDIT3 Over Time Across Groups
Gene Expression
Time
11
10
9
8
7
6
5
BL
V04
V06
V08
Control Mean Expression
Control ± STD
Prodromal Mean Expression
Prodromal ± STD
PD Mean Expression
PD ± STD

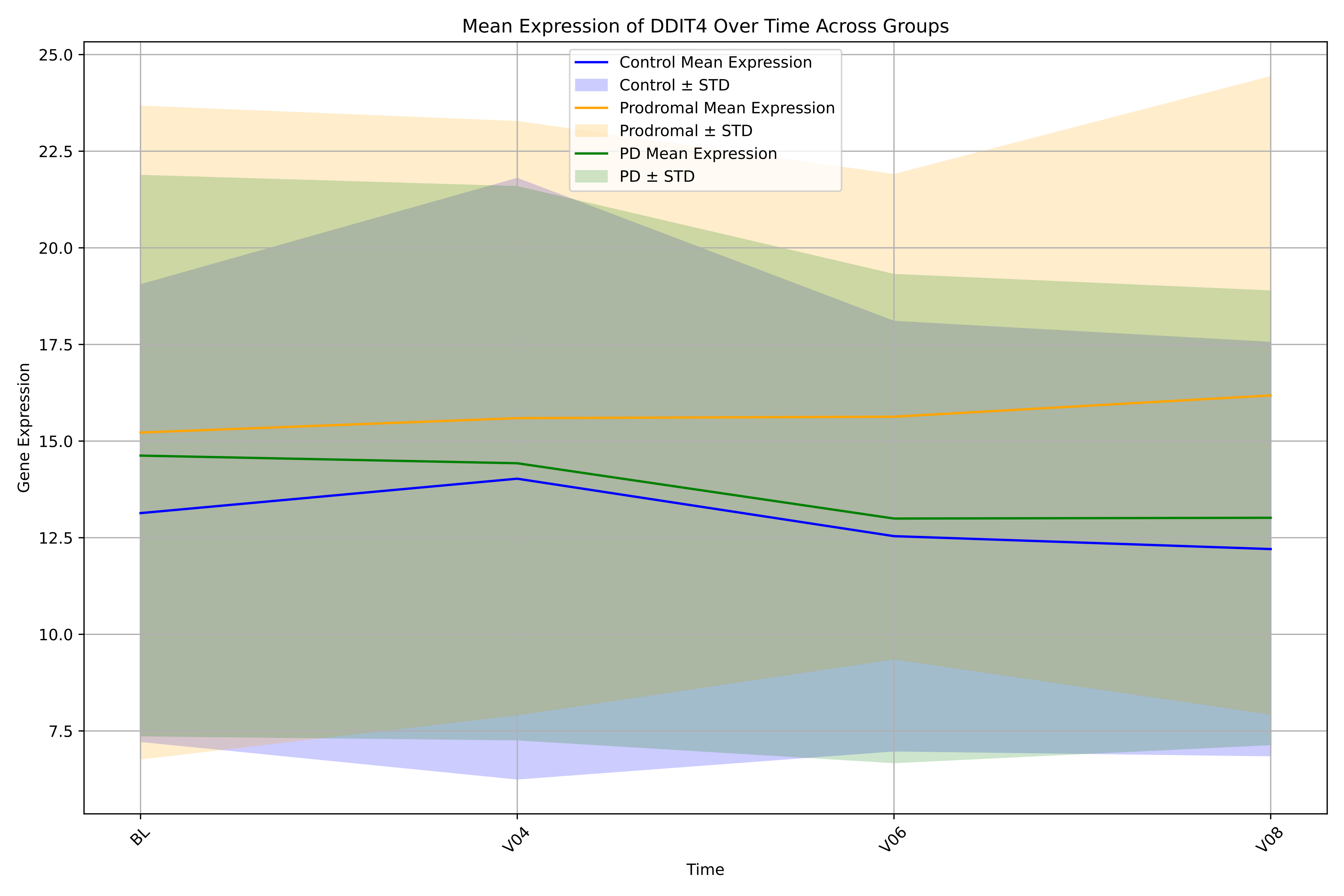

Mean Expression of DDIT4 Over Time Across Groups
Control Mean Expression
Control ± STD
Prodromal Mean Expression
Prodromal ± STD
PD Mean Expression
PD ± STD
Gene Expression
25.0
22.5
20.0
17.5
15.0
12.5
10.0
7.5
BL
V04
V06
V08
Time

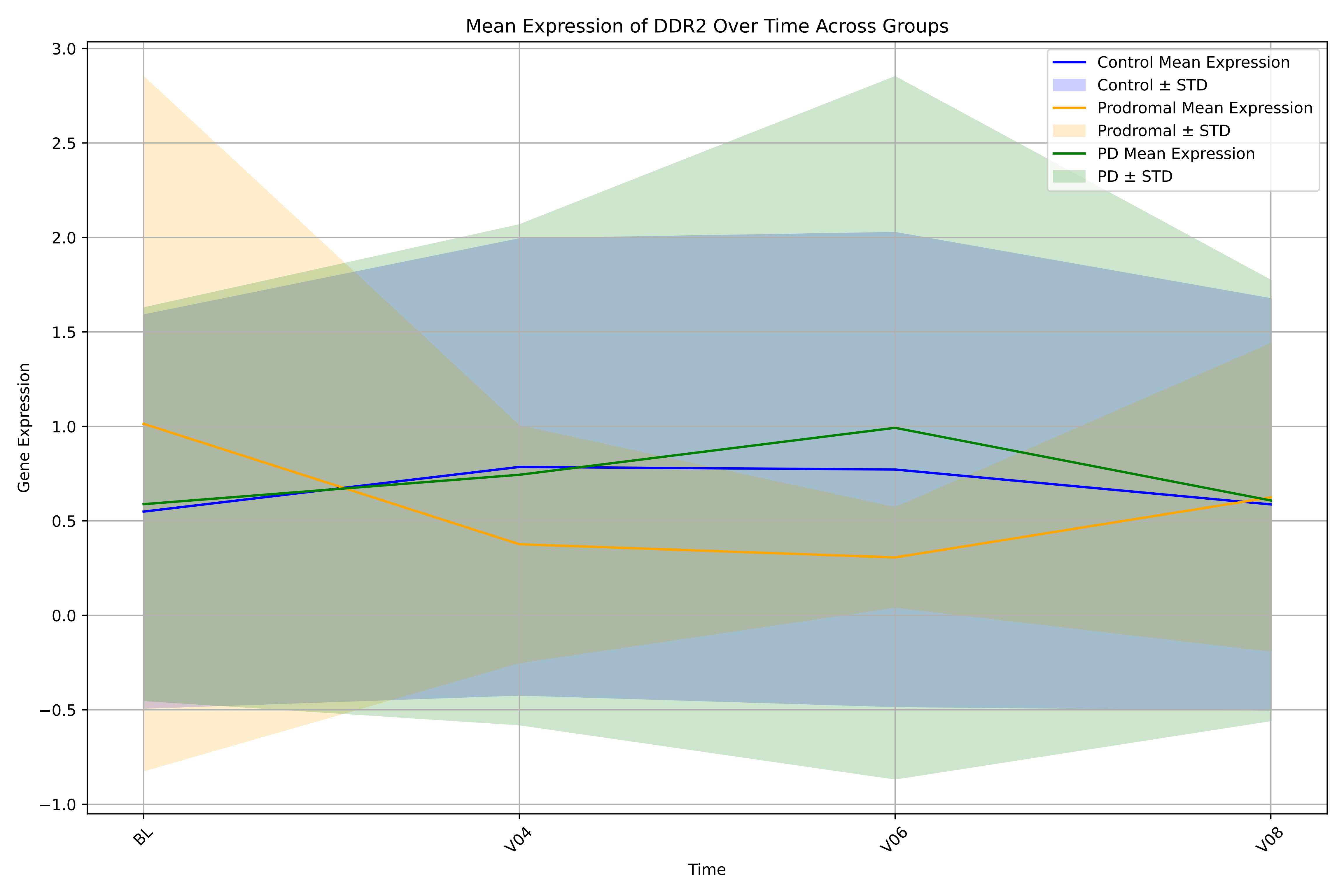

Mean Expression of DDR2 Over Time Across Groups
Control Mean Expression
Control ± STD
Prodromal Mean Expression
Prodromal ± STD
PD Mean Expression
PD ± STD
Gene Expression
Time
3.0
2.5
2.0
1.5
1.0
0.5
0.0
−0.5
−1.0
BL
V04
V06
V08

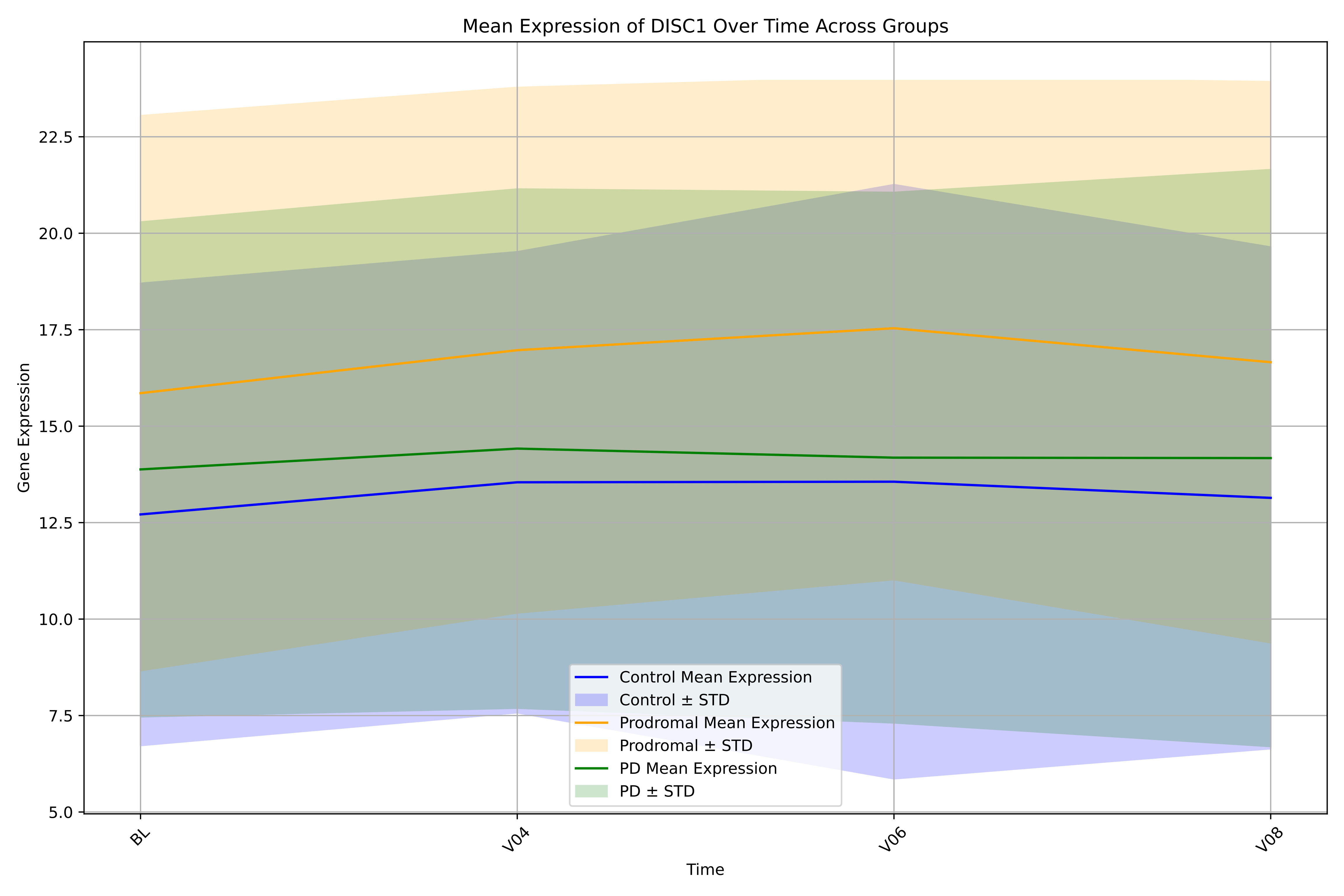
Mean Expression of DISC1 Over Time Across Groups
Gene Expression
22.5
20.0
17.5
15.0
12.5
10.0
7.5
5.0
BL
V04
V06
V08
Time
Control Mean Expression
Control ± STD
Prodromal Mean Expression
Prodromal ± STD
PD Mean Expression
PD ± STD

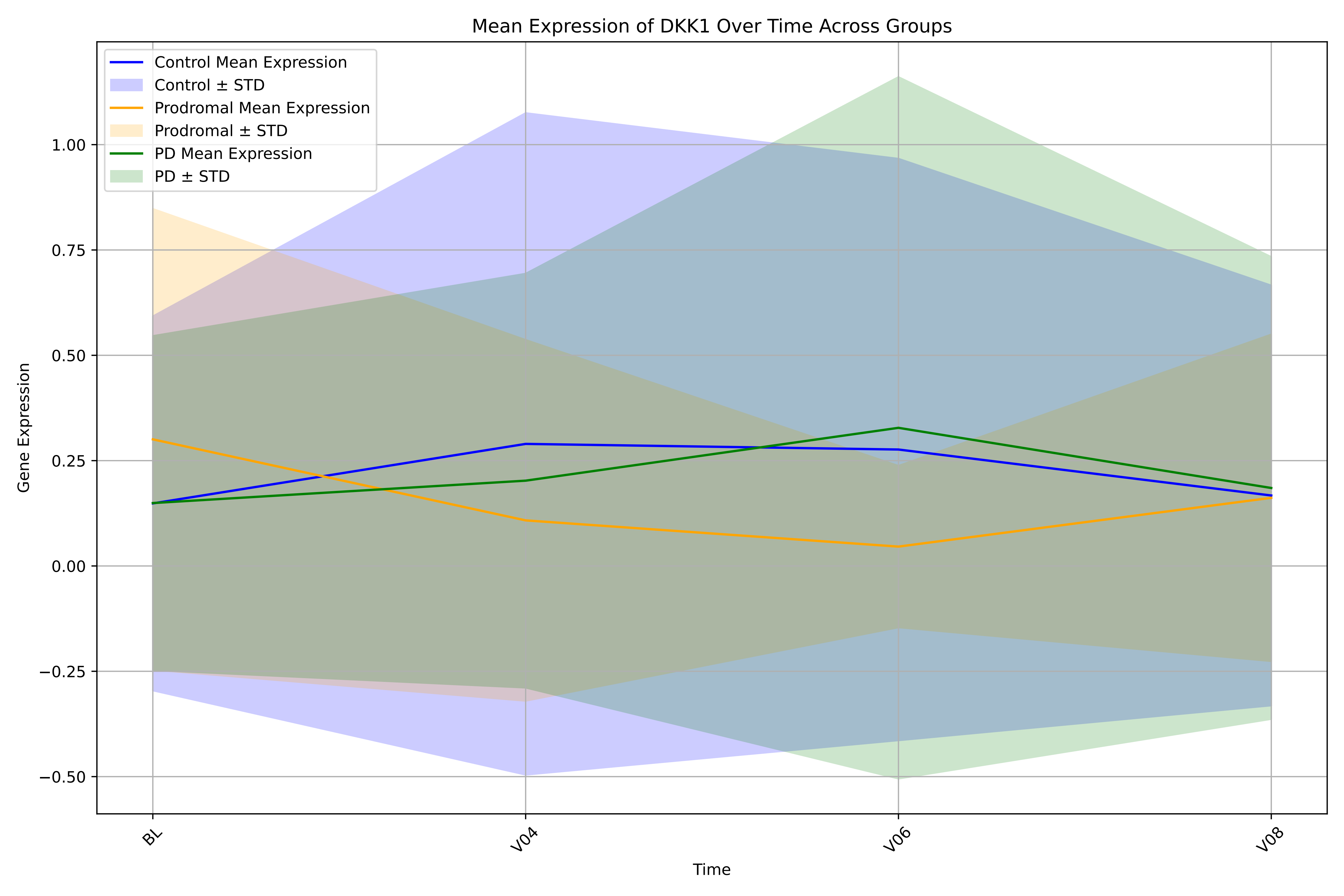

Mean Expression of DKK1 Over Time Across Groups
Control Mean Expression
Control ± STD
Prodromal Mean Expression
Prodromal ± STD
PD Mean Expression
PD ± STD
Gene Expression
1.00
0.75
0.50
0.25
0.00
−0.25
−0.50
BL
V04
V06
V08
Time

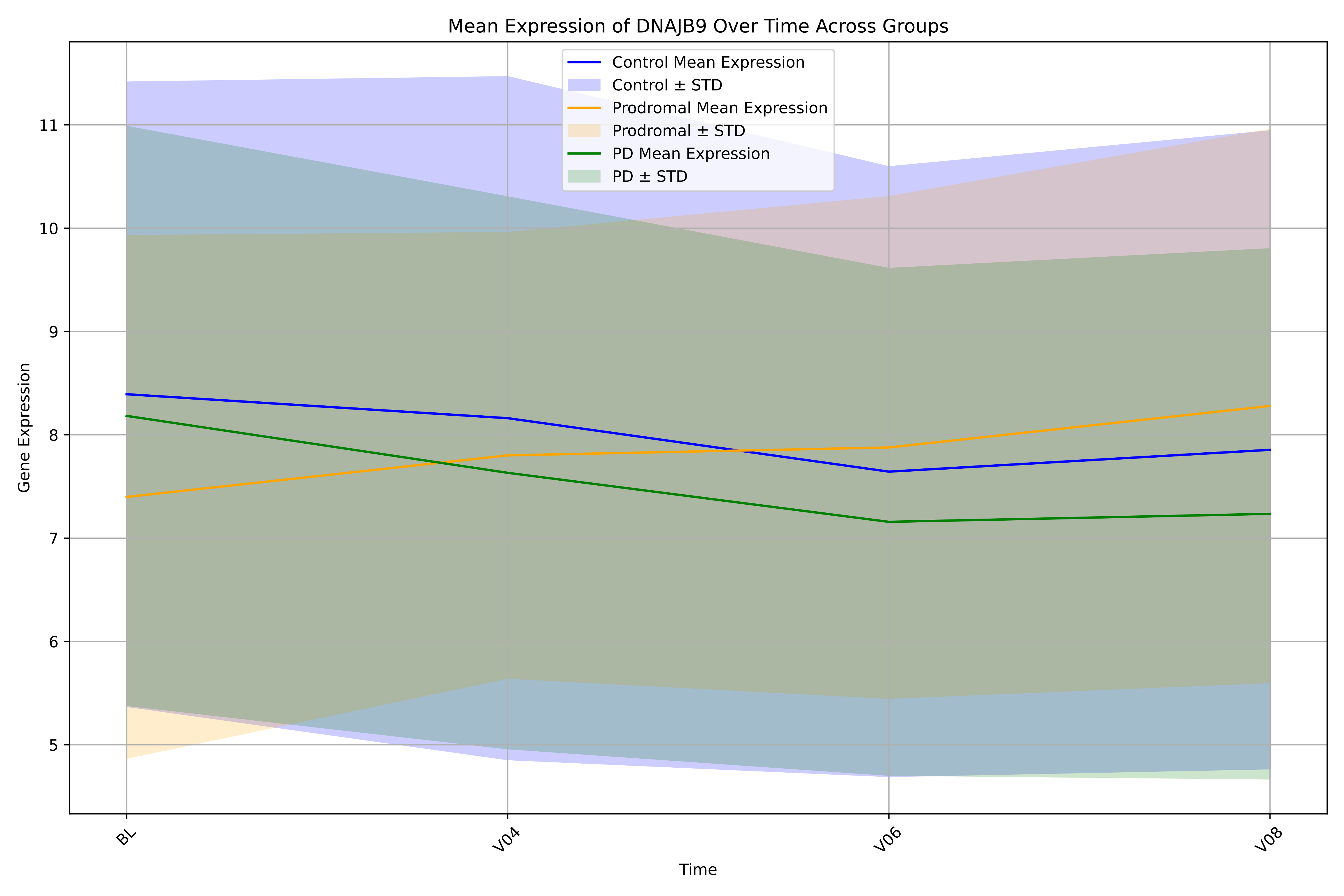

Mean Expression of DNAJB9 Over Time Across Groups
Control Mean Expression
Control ± STD
Prodromal Mean Expression
Prodromal ± STD
PD Mean Expression
PD ± STD
Gene Expression
11
10
9
8
7
6
5
BL
V04
V06
V08
Time

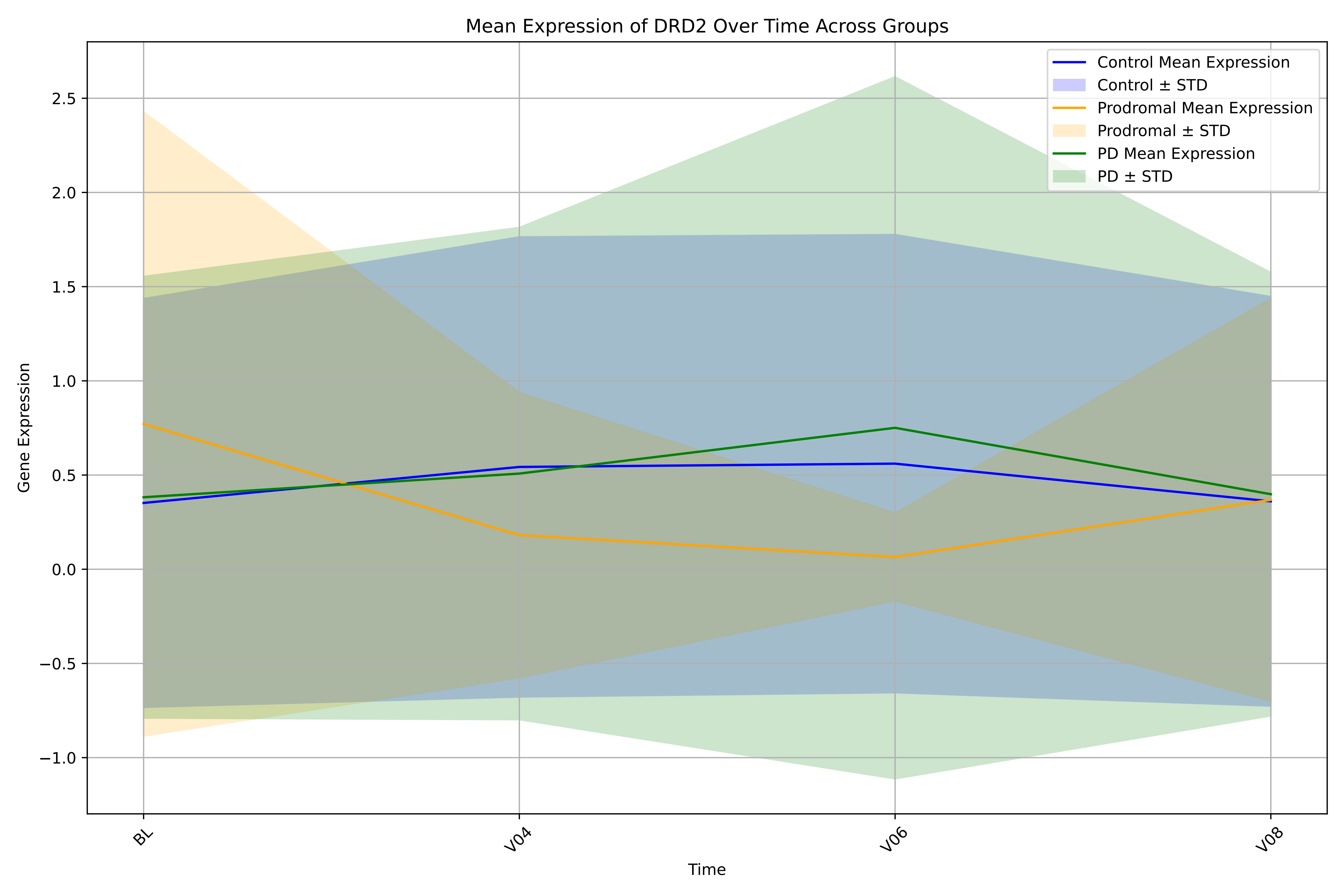

Mean Expression of DRD2 Over Time Across Groups
Control Mean Expression
Control ± STD
Prodromal Mean Expression
Prodromal ± STD
PD Mean Expression
PD ± STD
Gene Expression
2.5
2.0
1.5
1.0
0.5
0.0
−0.5
−1.0
BL
V04
V06
V08
Time

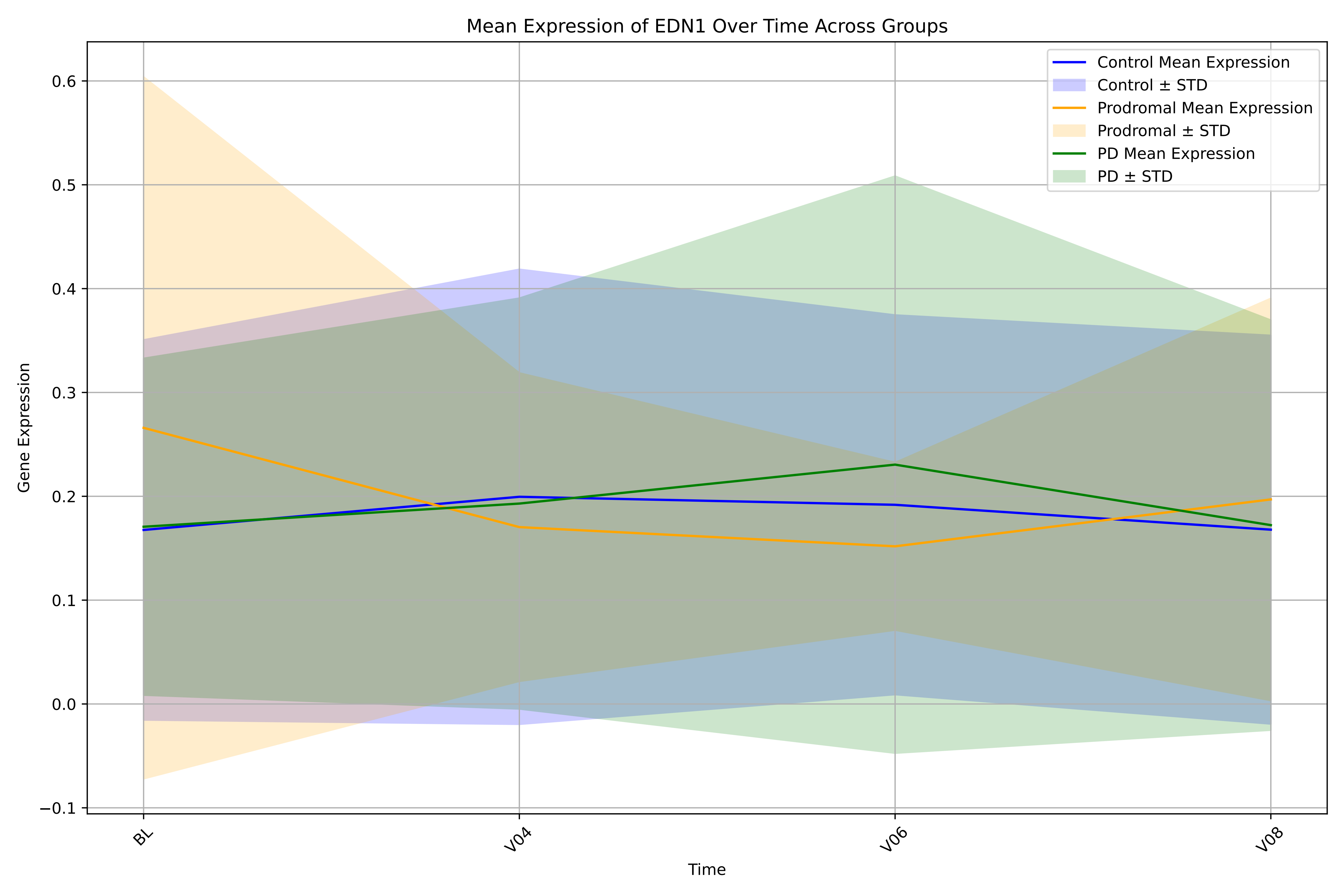

Mean Expression of EDN1 Over Time Across Groups
Control Mean Expression
Control ± STD
Prodromal Mean Expression
Prodromal ± STD
PD Mean Expression
PD ± STD
Gene Expression
0.6
0.5
0.4
0.3
0.2
0.1
0.0
−0.1
BL
V04
V06
V08
Time

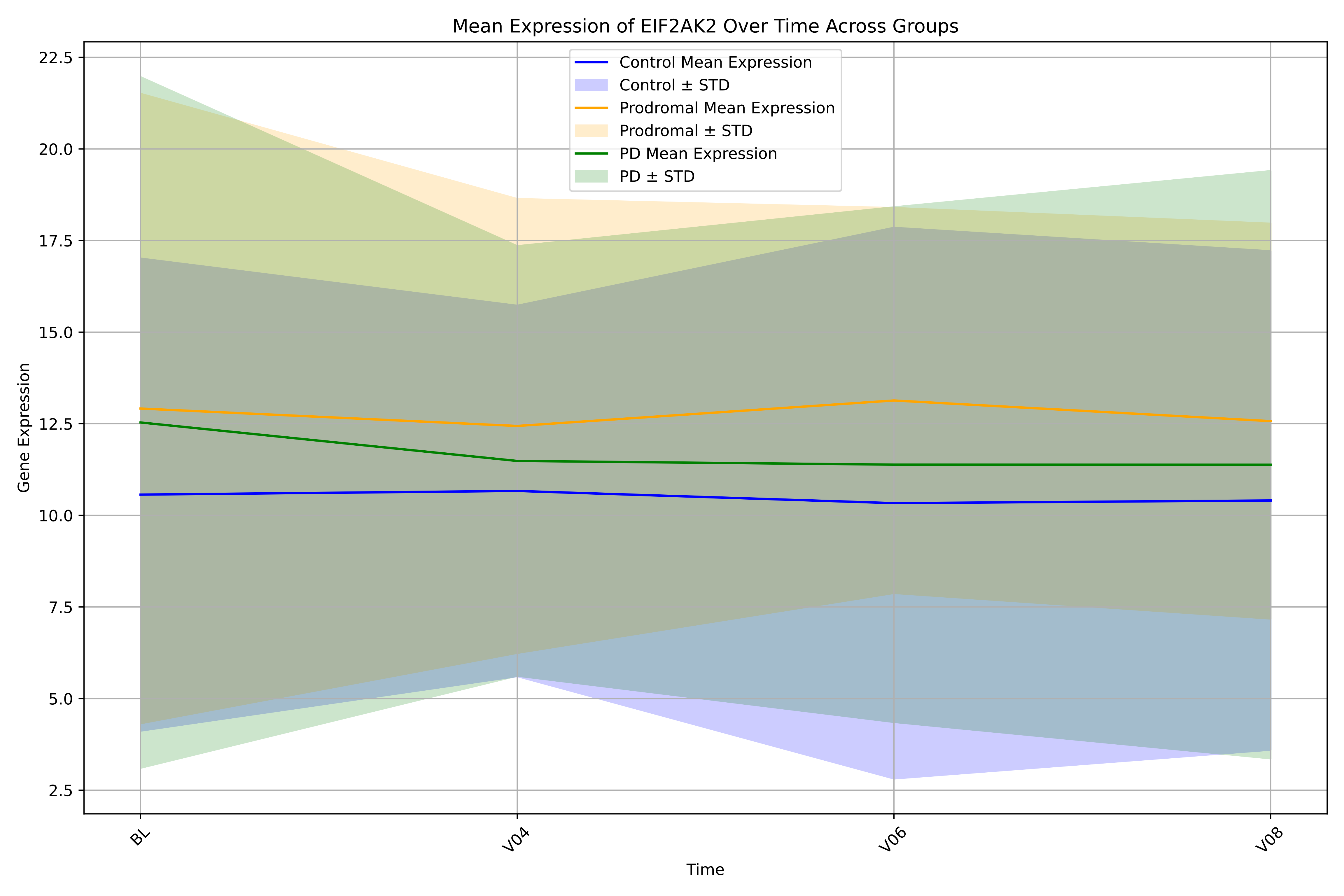

Mean Expression of EIF2AK2 Over Time Across Groups
Control Mean Expression
Control ± STD
Prodromal Mean Expression
Prodromal ± STD
PD Mean Expression
PD ± STD
22.5
20.0
17.5
15.0
12.5
10.0
7.5
5.0
2.5
Gene Expression
BL
V04
V06
V08
Time

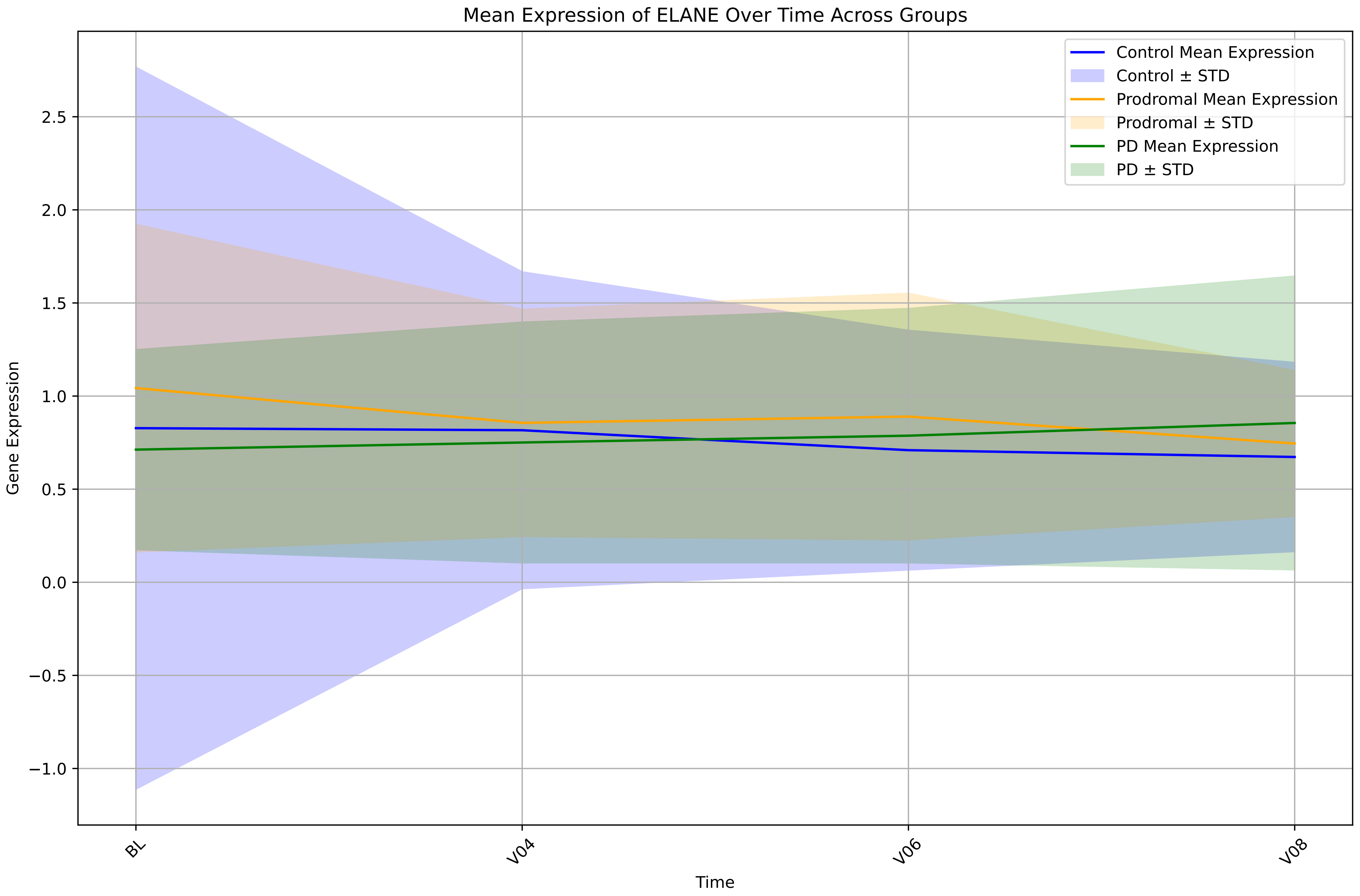

Mean Expression of ELANE Over Time Across Groups
Control Mean Expression
Control ± STD
Prodromal Mean Expression
Prodromal ± STD
PD Mean Expression
PD ± STD
Gene Expression
2.5
2.0
1.5
1.0
0.5
0.0
−0.5
−1.0
BL
V04
V06
V08
Time

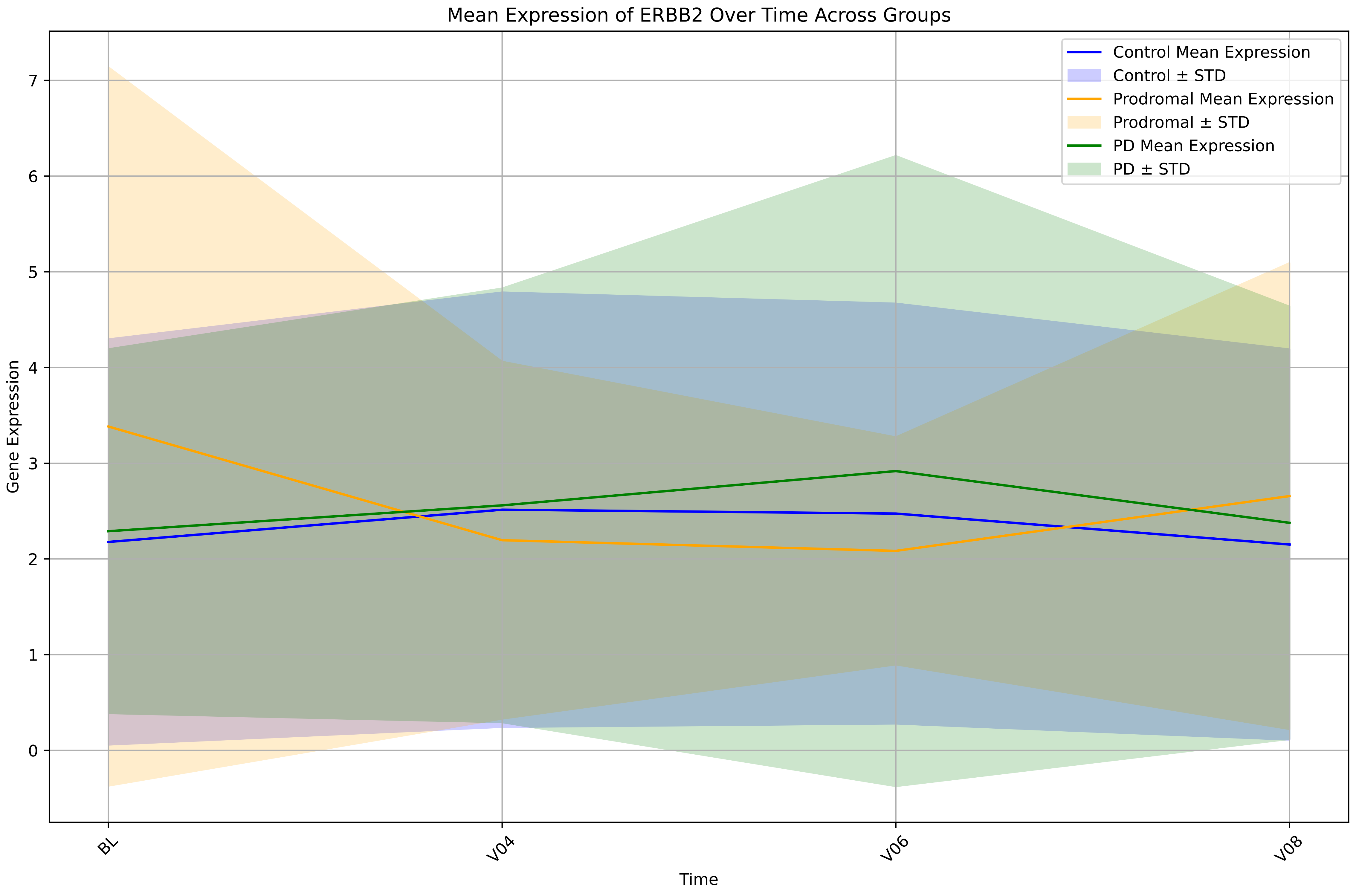

Mean Expression of ERBB2 Over Time Across Groups
Control Mean Expression
Control ± STD
Prodromal Mean Expression
Prodromal ± STD
PD Mean Expression
PD ± STD
Gene Expression
Time
BL
V04
V06
V08
0
1
2
3
4
5
6
7

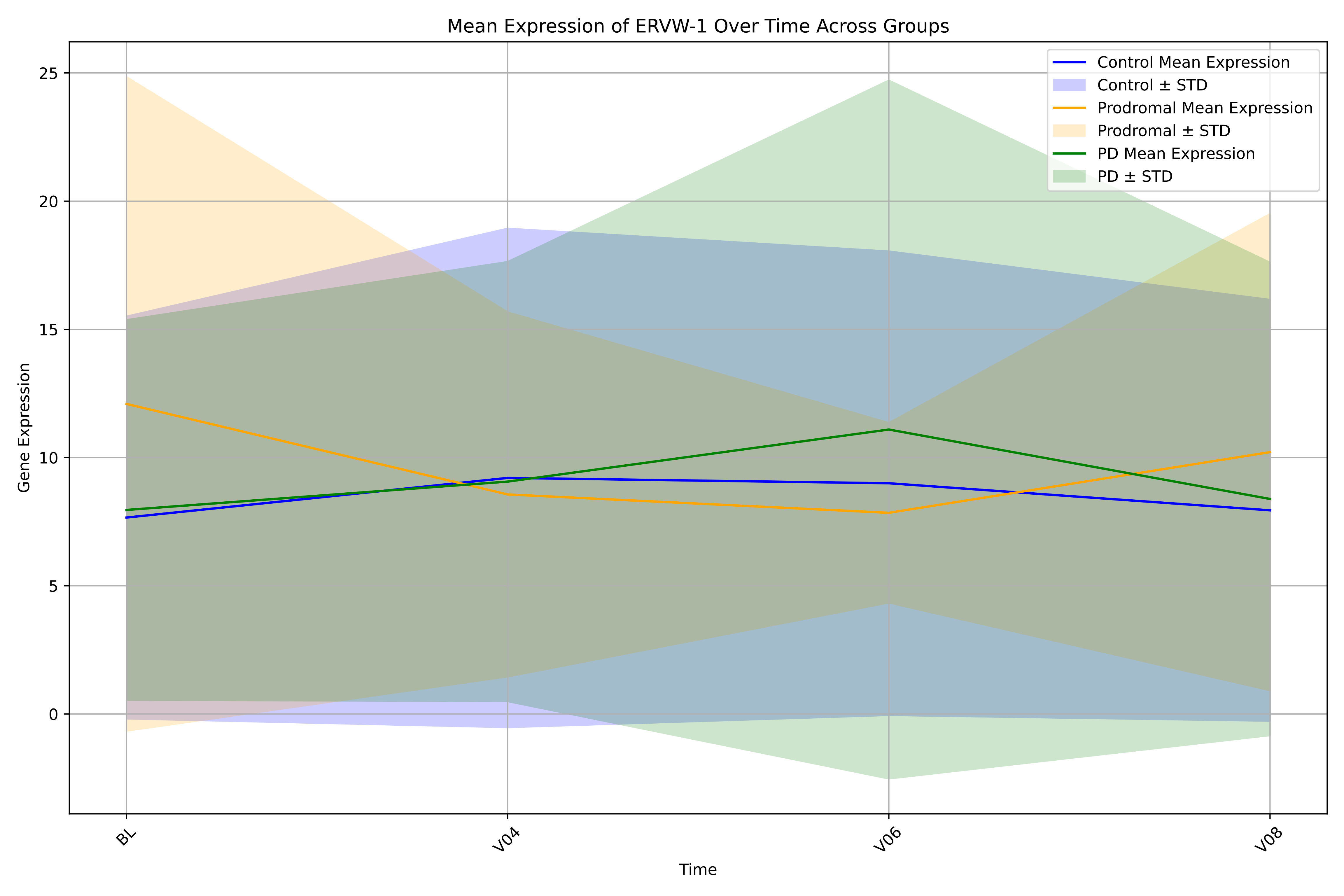

Mean Expression of ERVW-1 Over Time Across Groups
Control Mean Expression
Control ± STD
Prodromal Mean Expression
Prodromal ± STD
PD Mean Expression
PD ± STD
Gene Expression
25
20
15
10
5
0
BL
V04
V06
V08
Time

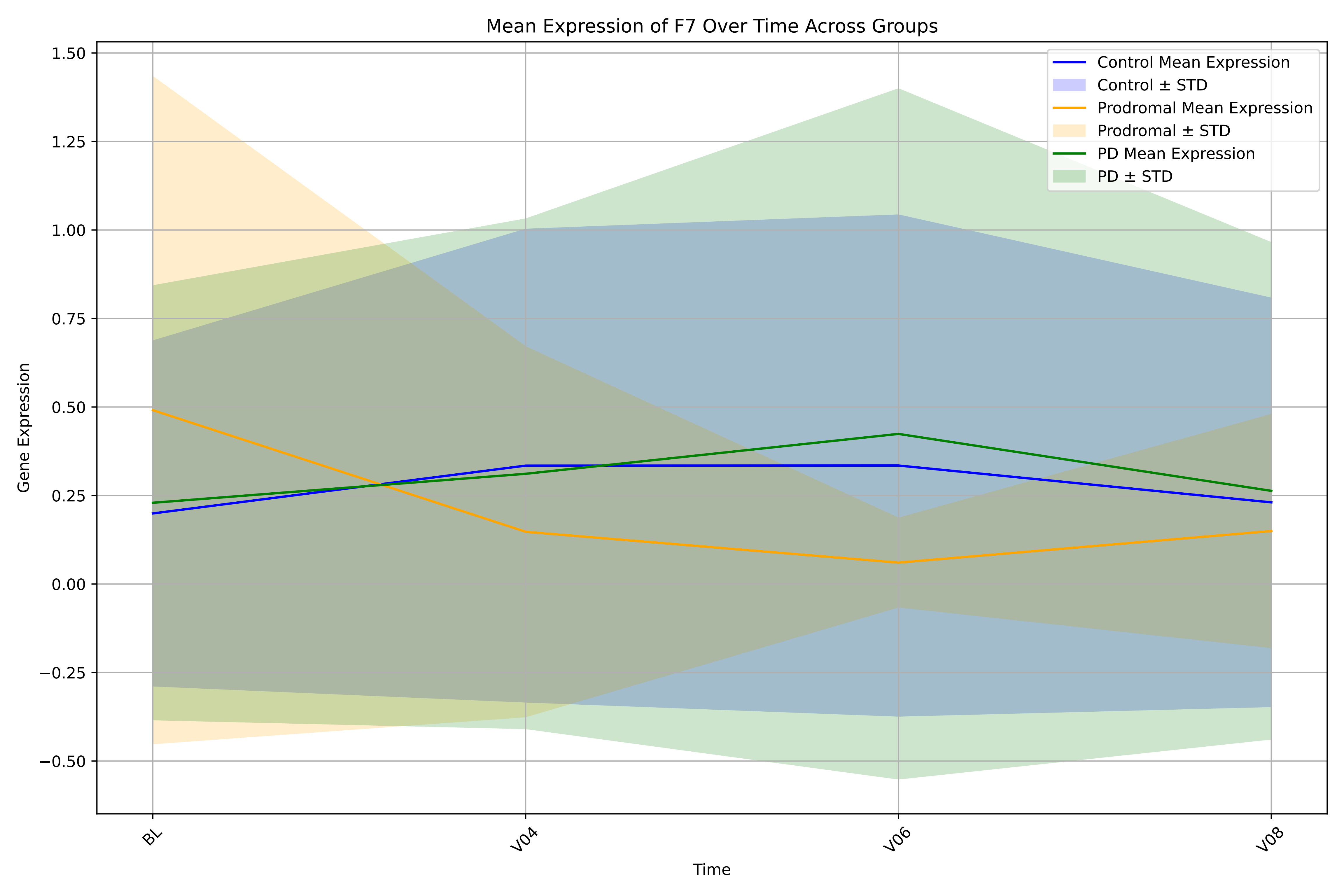

Mean Expression of F7 Over Time Across Groups
Control Mean Expression
Control ± STD
Prodromal Mean Expression
Prodromal ± STD
PD Mean Expression
PD ± STD
Gene Expression
1.50
1.25
1.00
0.75
0.50
0.25
0.00
−0.25
−0.50
BL
V04
V06
V08
Time

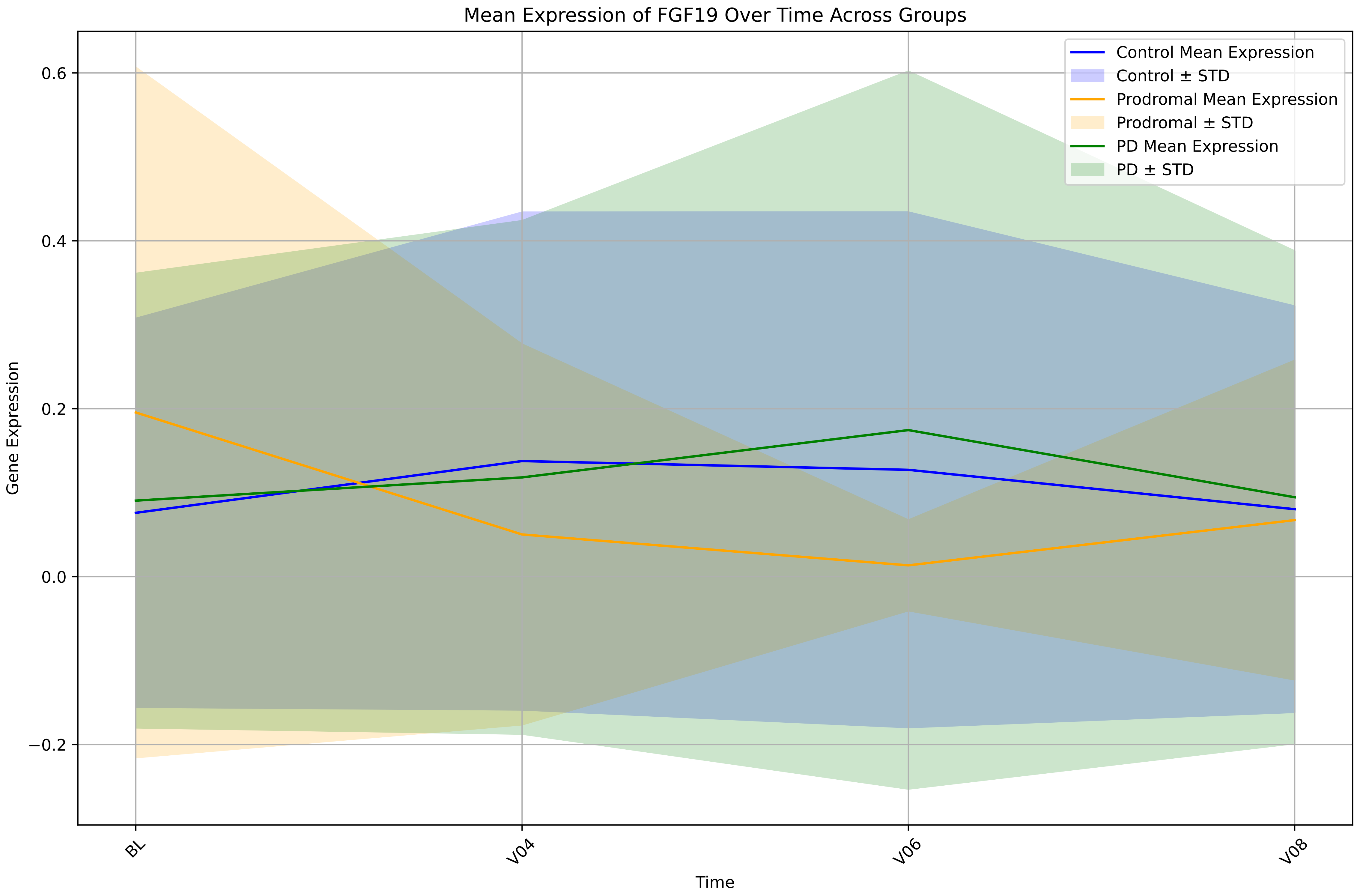

Mean Expression of FGF19 Over Time Across Groups
Control Mean Expression
Control ± STD
Prodromal Mean Expression
Prodromal ± STD
PD Mean Expression
PD ± STD
Gene Expression
0.6
0.4
0.2
0.0
−0.2
BL
V04
V06
V08
Time

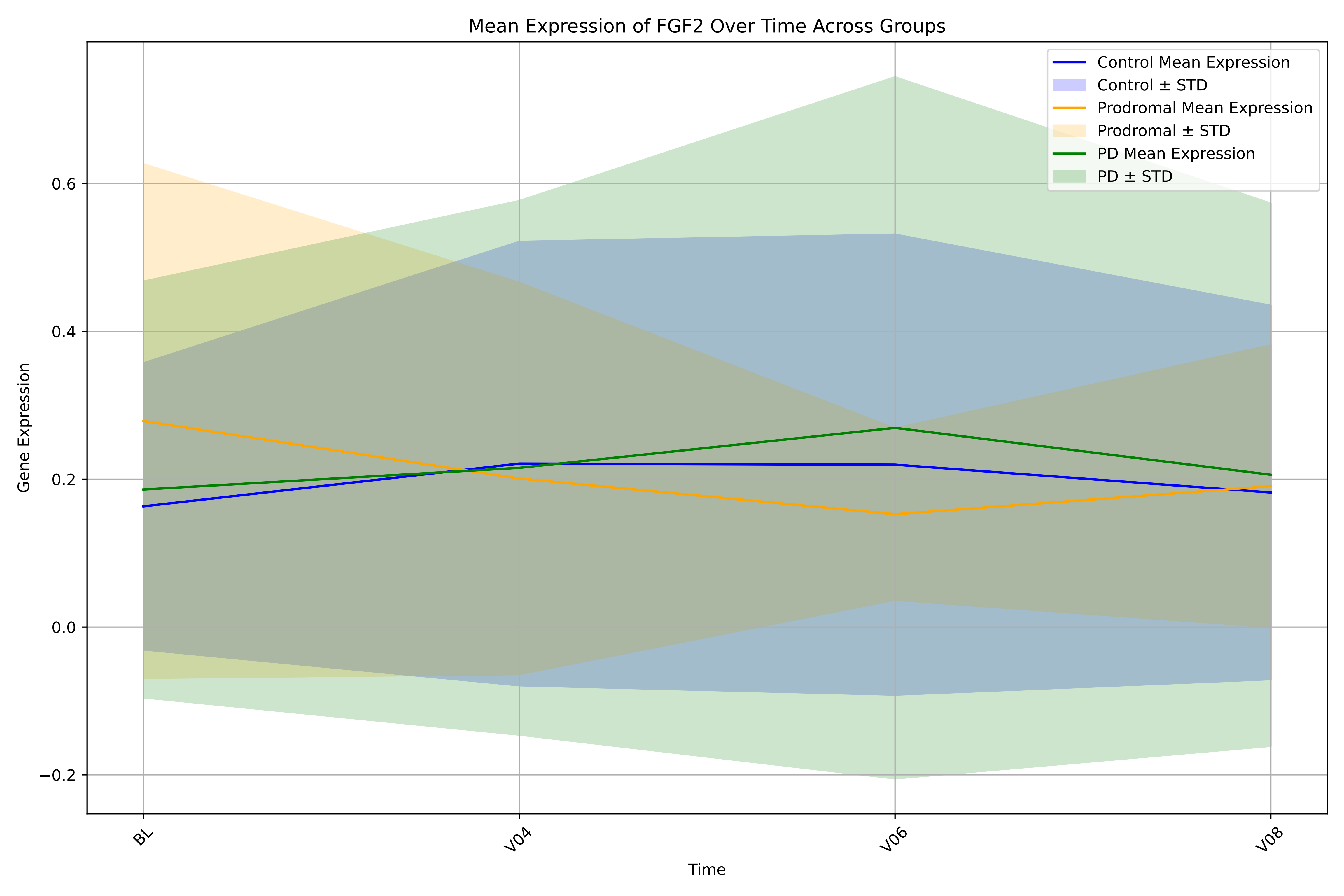
Mean Expression of FGF2 Over Time Across Groups
Control Mean Expression
Control ± STD
Prodromal Mean Expression
Prodromal ± STD
PD Mean Expression
PD ± STD
Gene Expression
0.6
0.4
0.2
0.0
−0.2
BL
V04
V06
V08
Time

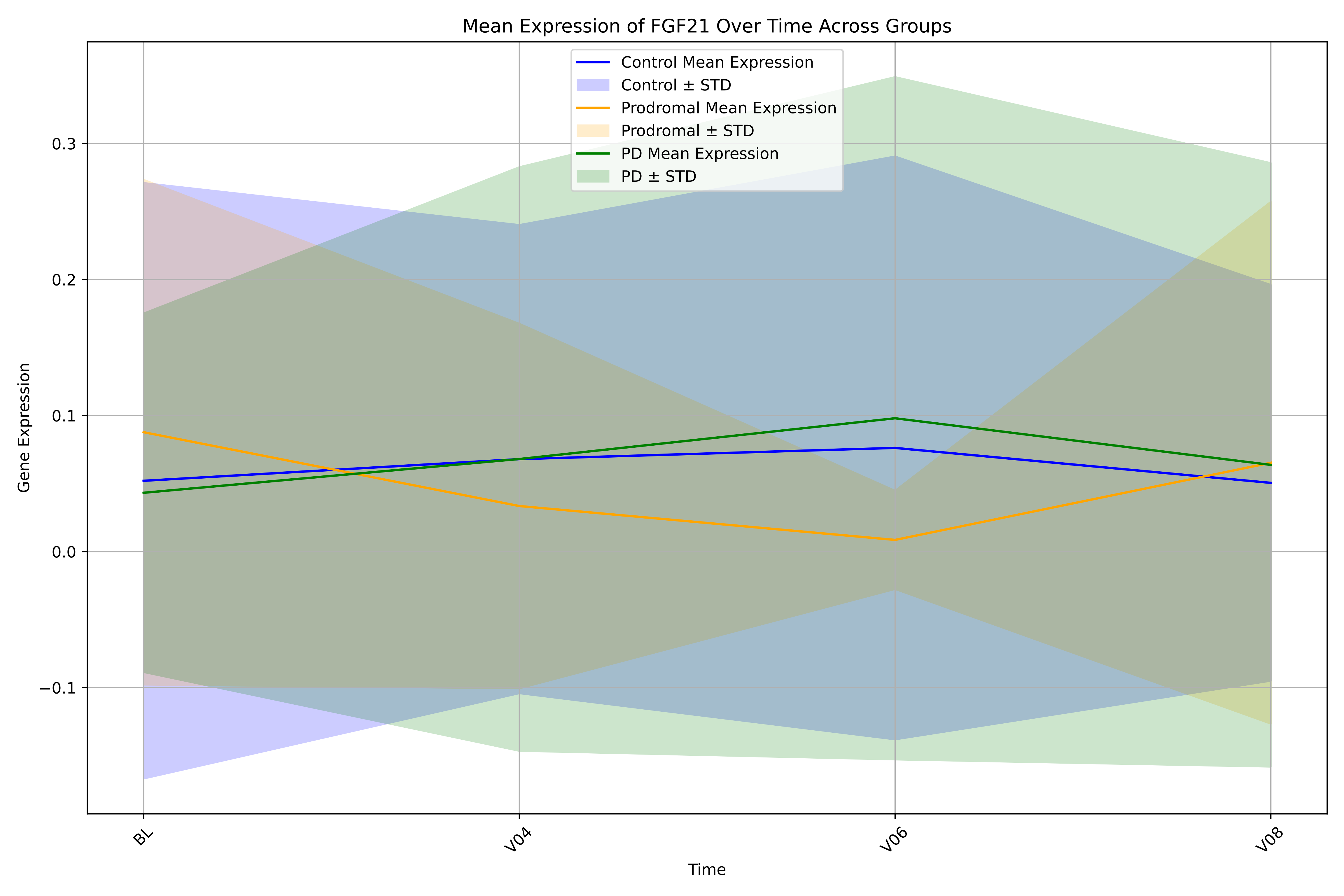
Mean Expression of FGF21 Over Time Across Groups
Control Mean Expression
Control ± STD
Prodromal Mean Expression
Prodromal ± STD
PD Mean Expression
PD ± STD
Gene Expression
0.3
0.2
0.1
0.0
−0.1
BL
V04
V06
V08
Time

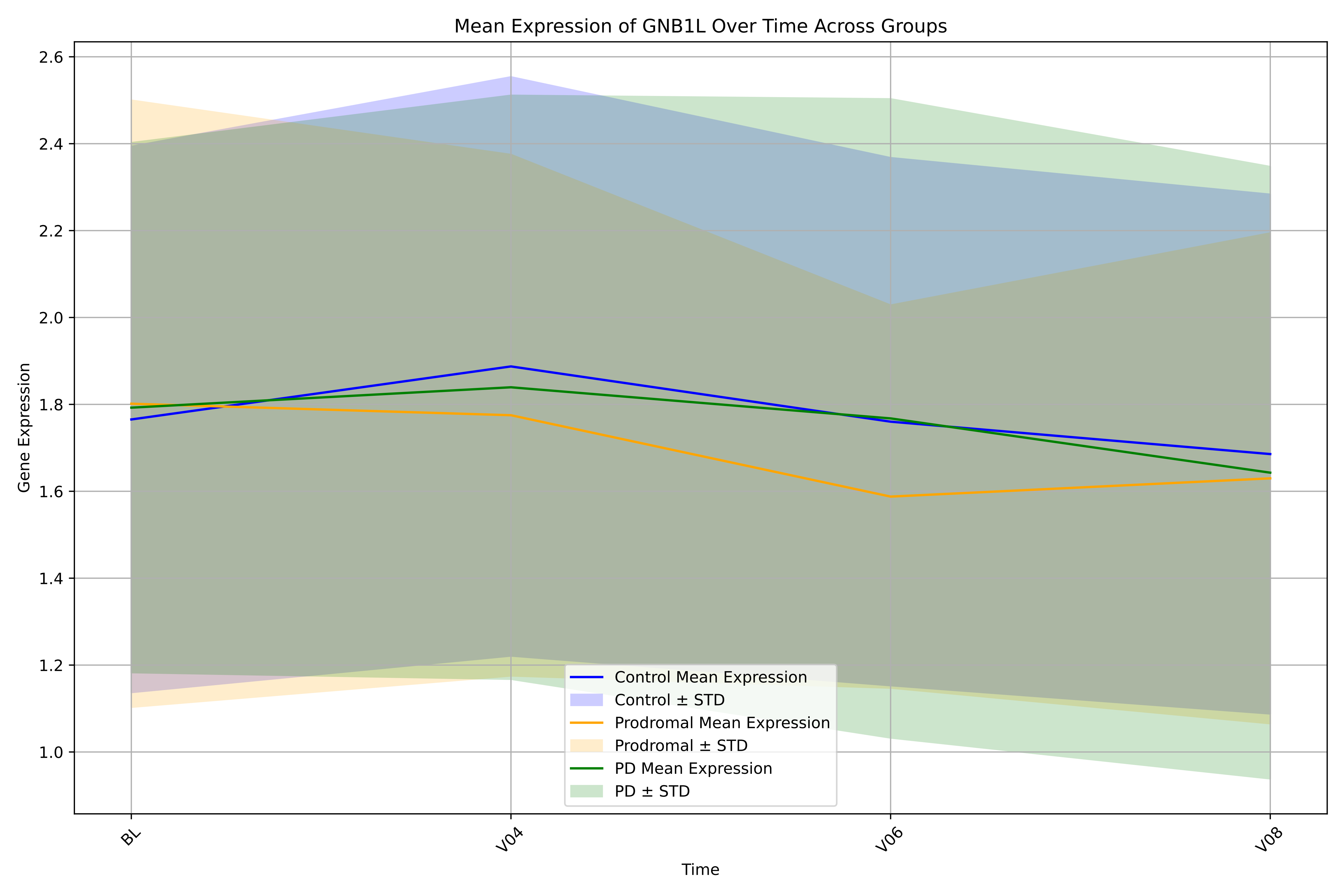

Mean Expression of GNB1L Over Time Across Groups
Gene Expression
Time
2.6
2.4
2.2
2.0
1.8
1.6
1.4
1.2
1.0
BL
V04
V06
V08
Control Mean Expression
Control ± STD
Prodromal Mean Expression
Prodromal ± STD
PD Mean Expression
PD ± STD

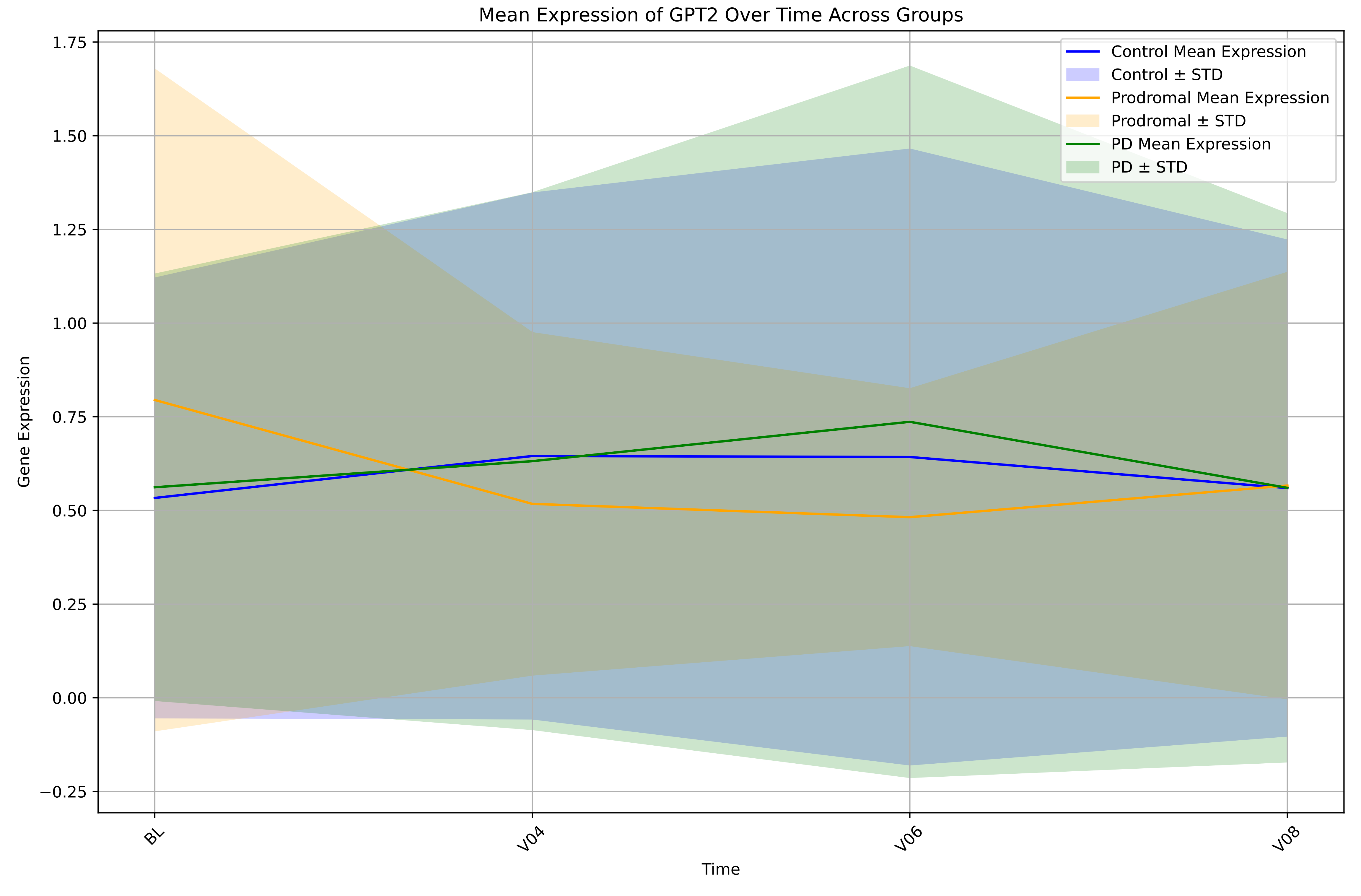

Mean Expression of GPT2 Over Time Across Groups
Control Mean Expression
Control ± STD
Prodromal Mean Expression
Prodromal ± STD
PD Mean Expression
PD ± STD
Gene Expression
1.75
1.50
1.25
1.00
0.75
0.50
0.25
0.00
−0.25
BL
V04
V06
V08
Time

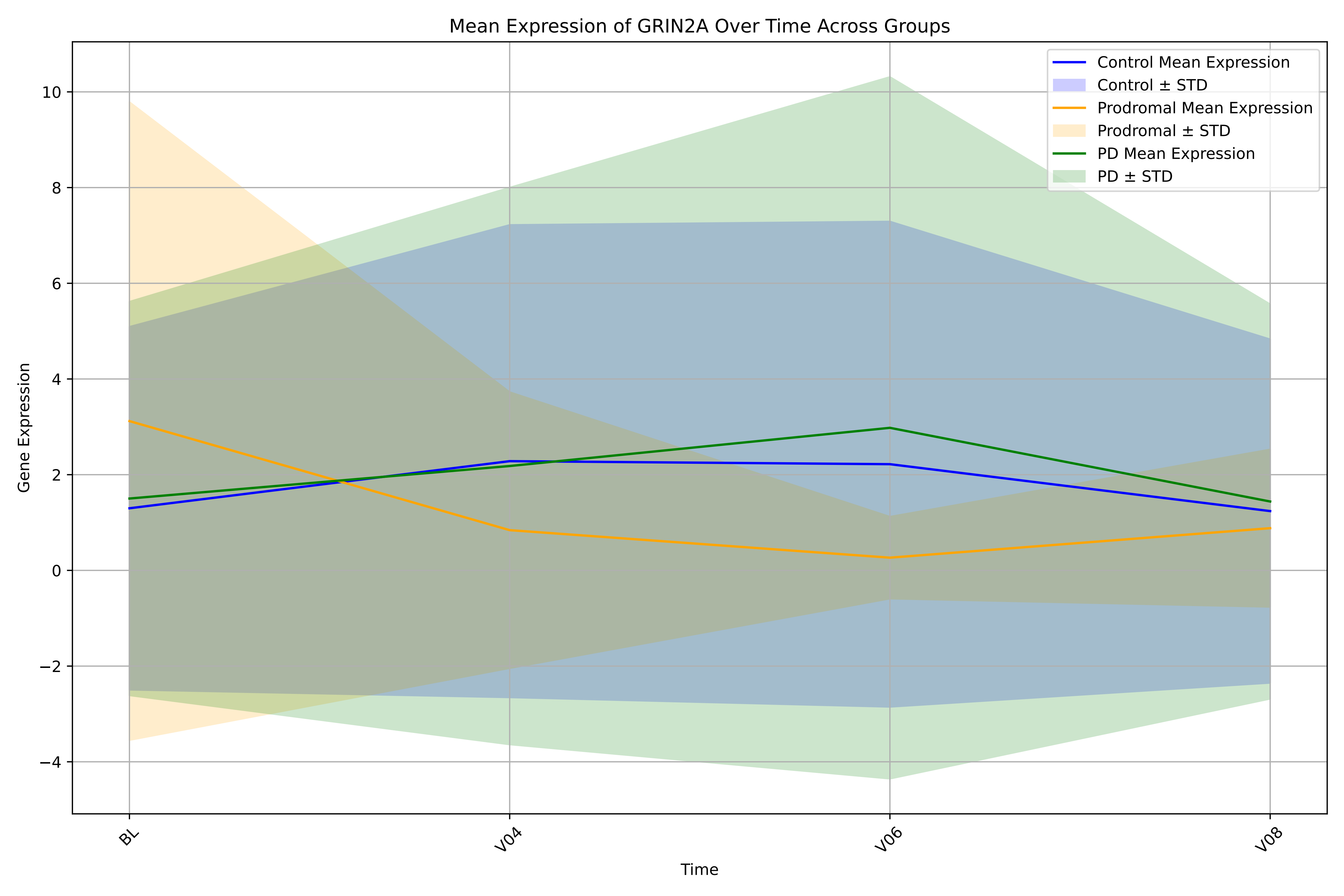

Mean Expression of GRIN2A Over Time Across Groups
Control Mean Expression
Control ± STD
Prodromal Mean Expression
Prodromal ± STD
PD Mean Expression
PD ± STD
10
8
6
4
2
0
−2
−4
Gene Expression
BL
V04
V06
V08
Time

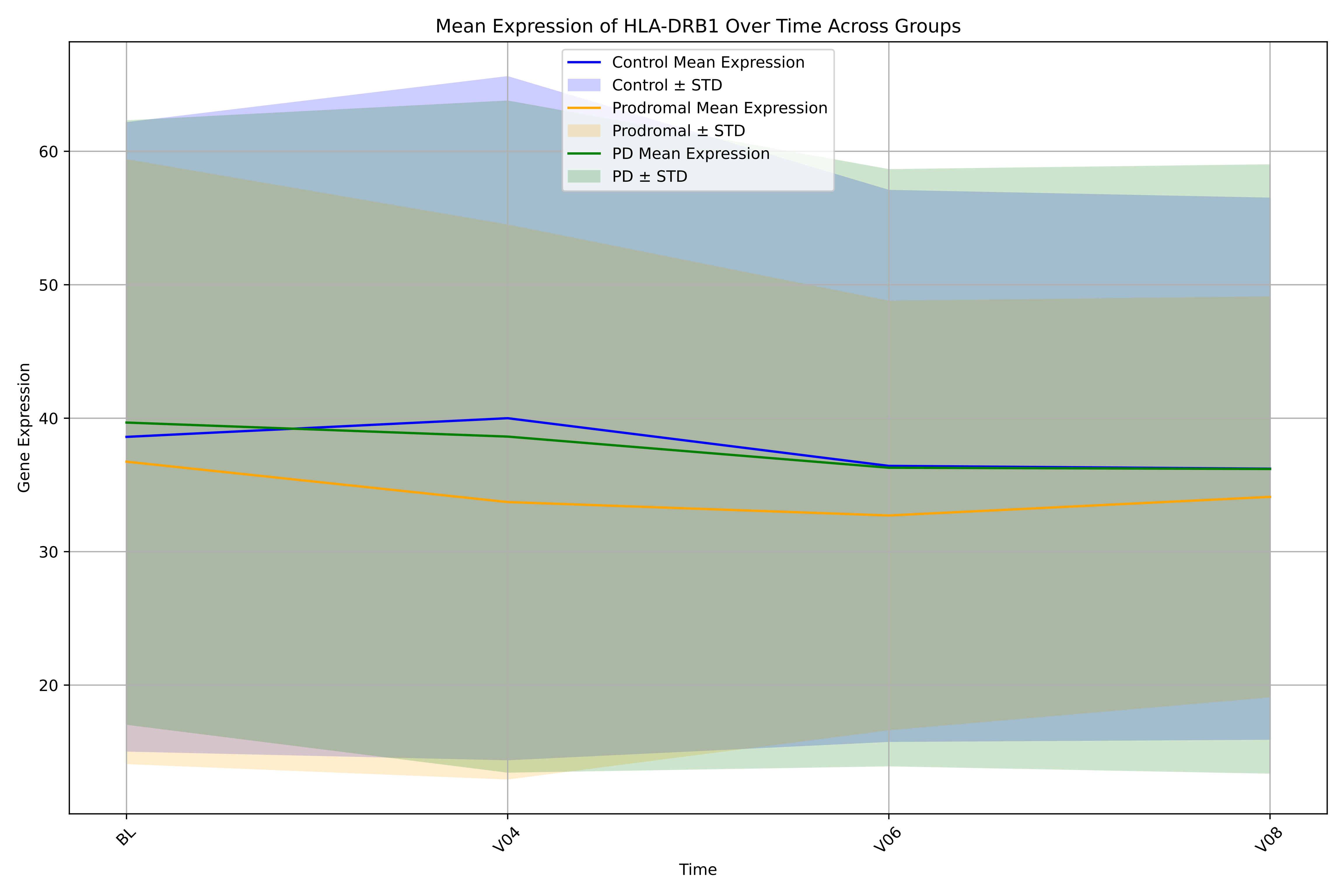
Mean Expression of HLA-DRB1 Over Time Across Groups
Control Mean Expression
Control ± STD
Prodromal Mean Expression
Prodromal ± STD
PD Mean Expression
PD ± STD
Gene Expression
60
50
40
30
20
BL
V04
V06
V08
Time

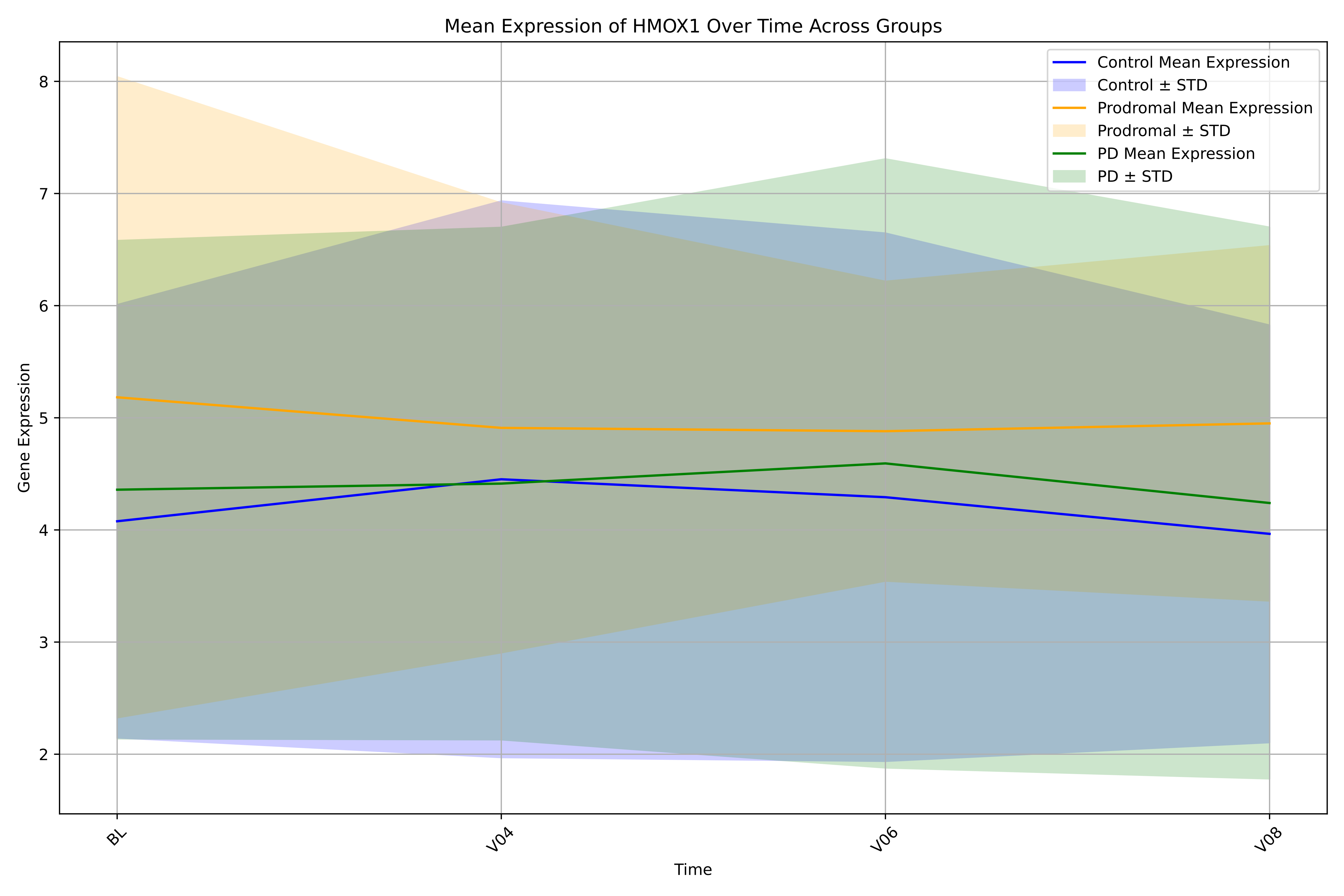
Mean Expression of HMOX1 Over Time Across Groups
Control Mean Expression
Control ± STD
Prodromal Mean Expression
Prodromal ± STD
PD Mean Expression
PD ± STD
Gene Expression
Time
BL
V04
V06
V08
2
3
4
5
6
7
8

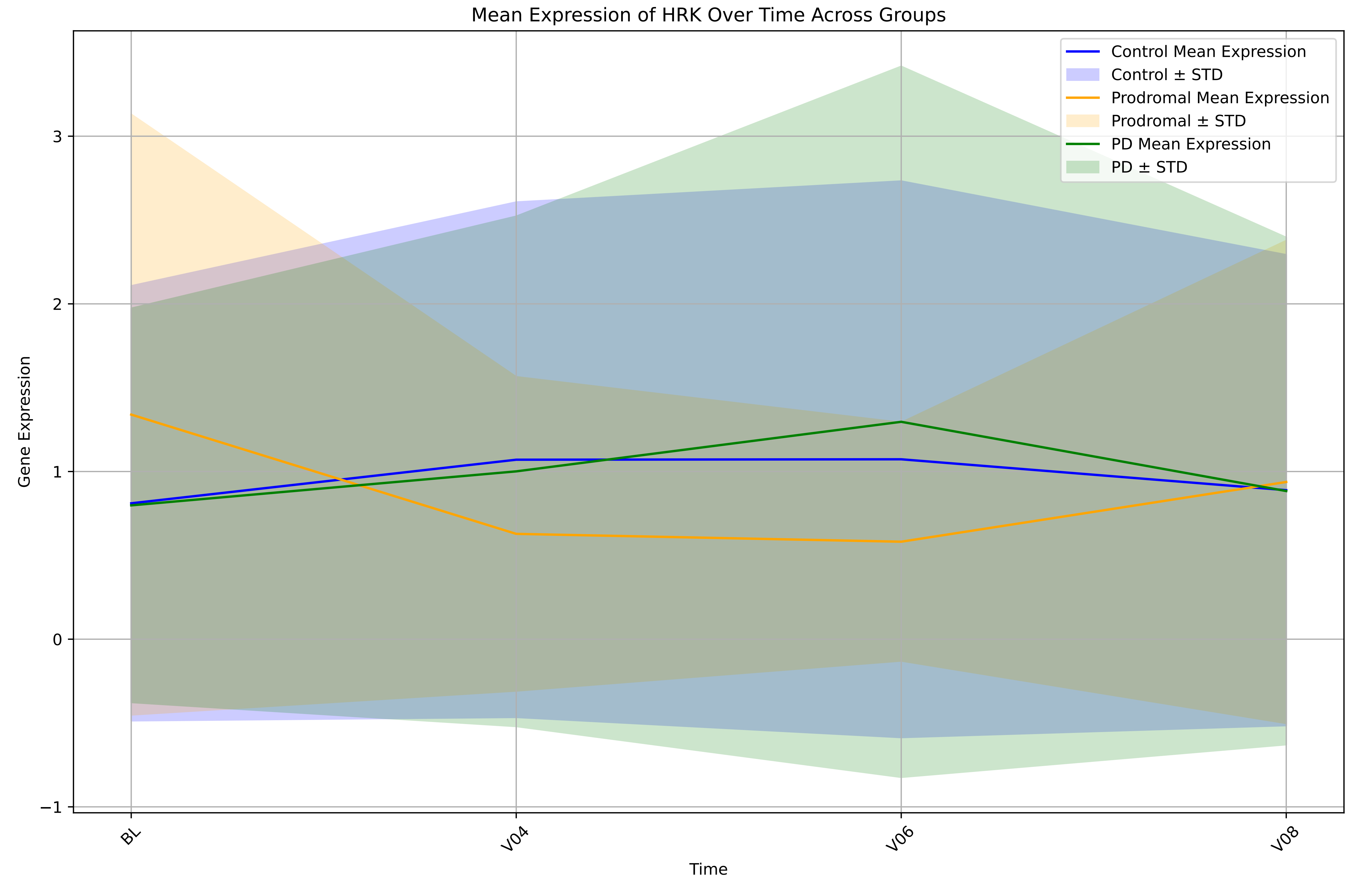
Mean Expression of HRK Over Time Across Groups
Control Mean Expression
Control ± STD
Prodromal Mean Expression
Prodromal ± STD
PD Mean Expression
PD ± STD
Gene Expression
3
2
1
0
−1
BL
V04
V06
V08
Time

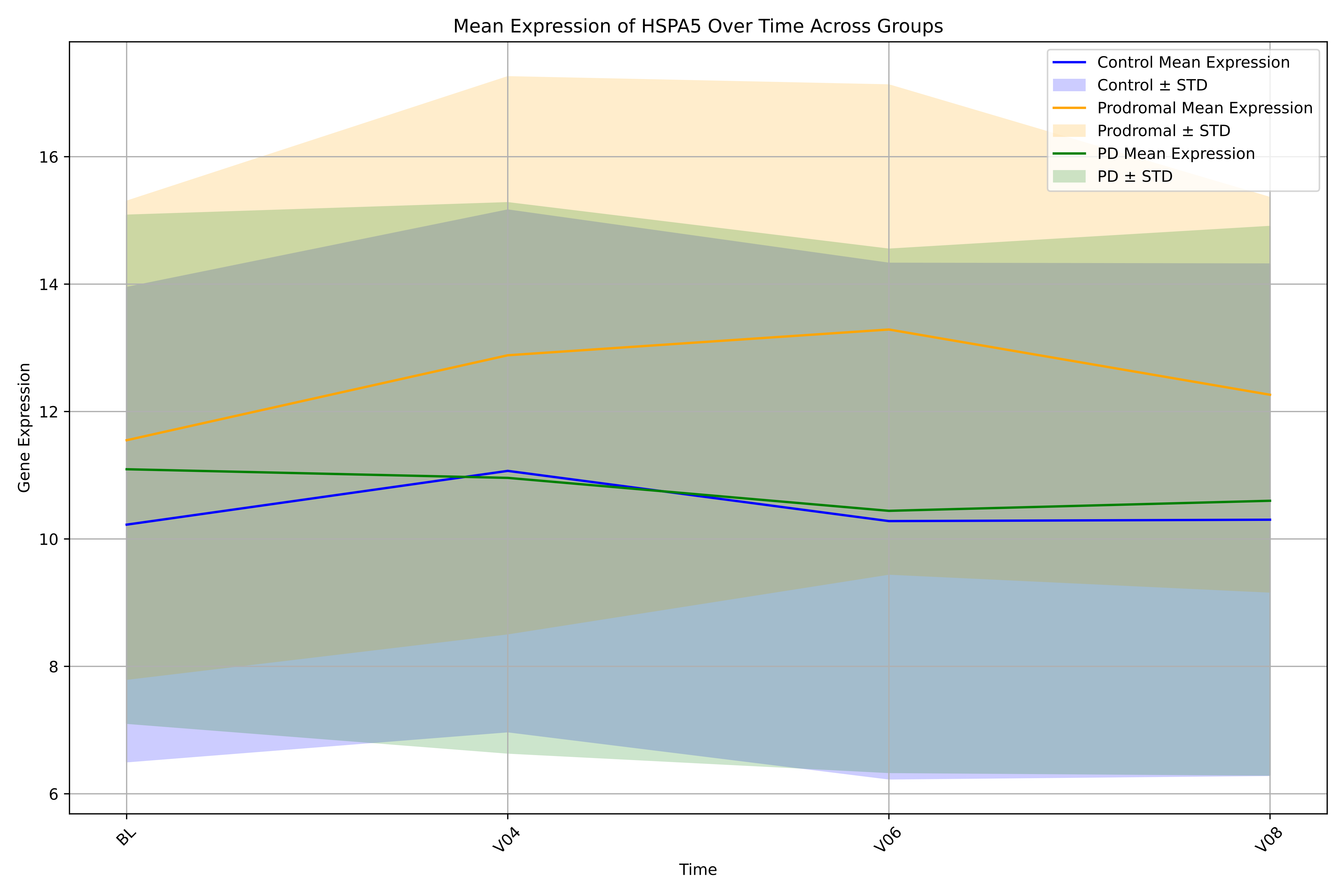

Mean Expression of HSPA5 Over Time Across Groups
Control Mean Expression
Control ± STD
Prodromal Mean Expression
Prodromal ± STD
PD Mean Expression
PD ± STD
Gene Expression
6
8
10
12
14
16
Time
BL
V04
V06
V08

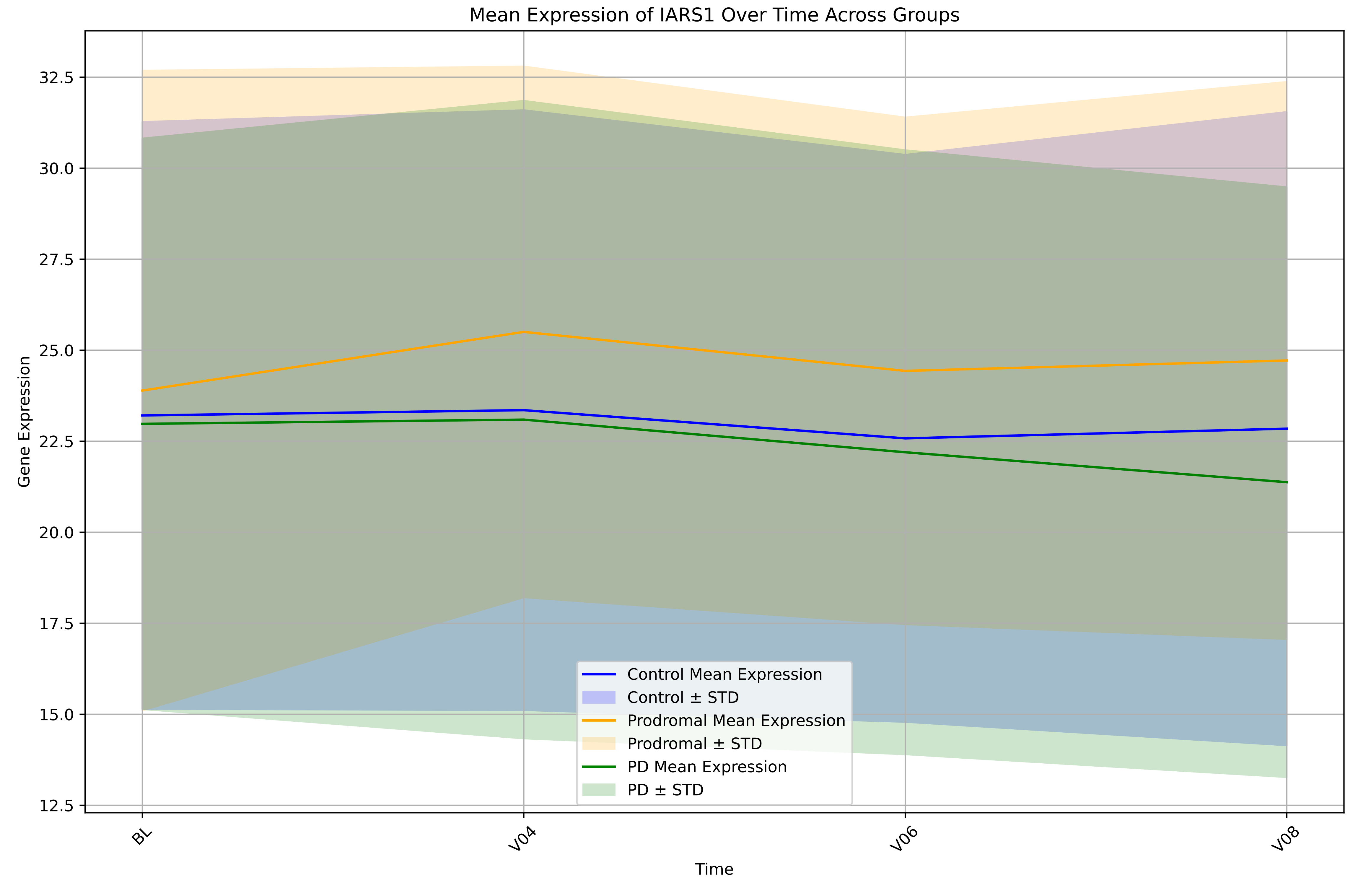

Mean Expression of IARS1 Over Time Across Groups
Gene Expression
Time
BL
V04
V06
V08
12.5
15.0
17.5
20.0
22.5
25.0
27.5
30.0
32.5
Control Mean Expression
Control ± STD
Prodromal Mean Expression
Prodromal ± STD
PD Mean Expression
PD ± STD

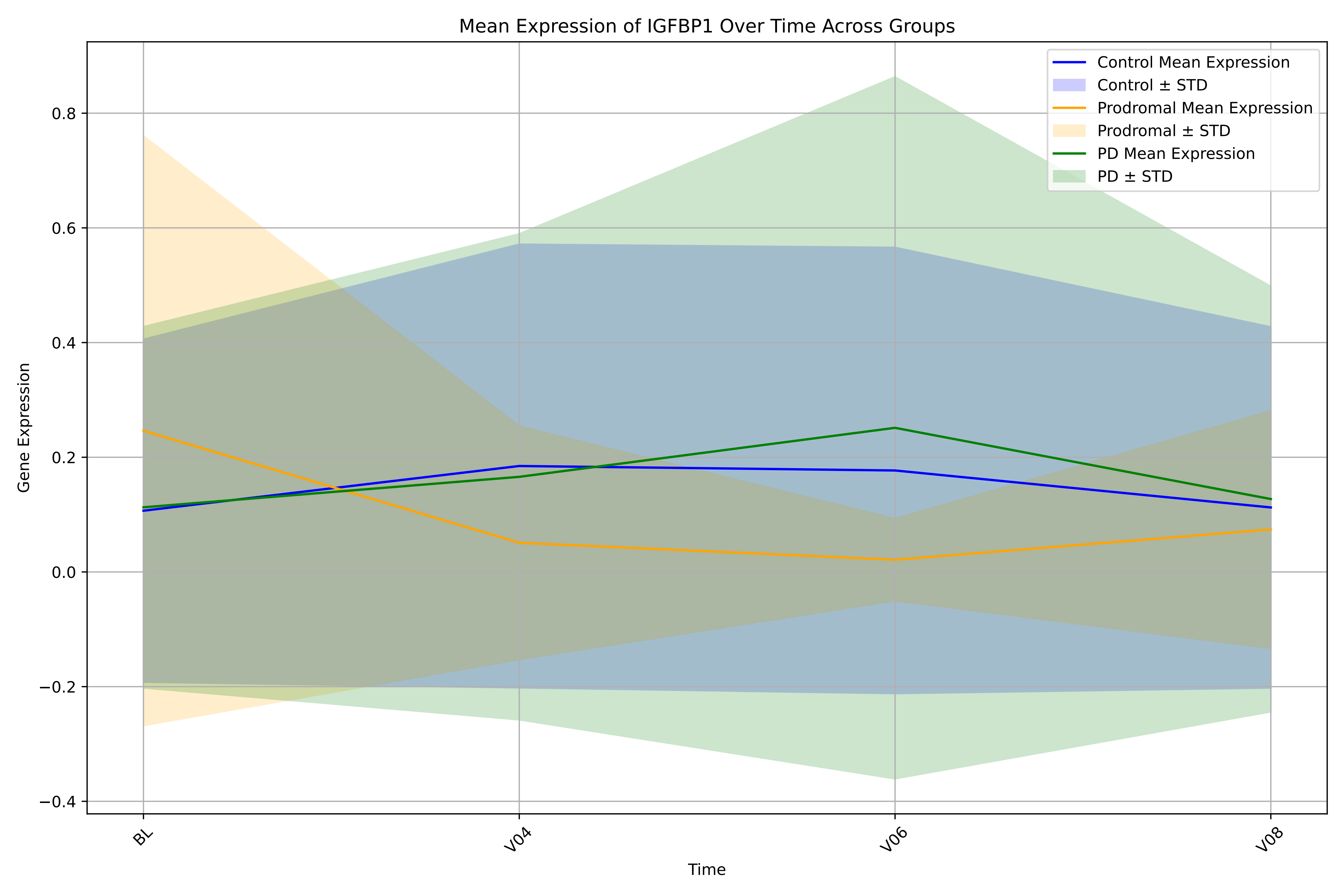

Mean Expression of IGFBP1 Over Time Across Groups
Control Mean Expression
Control ± STD
Prodromal Mean Expression
Prodromal ± STD
PD Mean Expression
PD ± STD
Gene Expression
0.8
0.6
0.4
0.2
0.0
−0.2
−0.4
BL
V04
V06
V08
Time

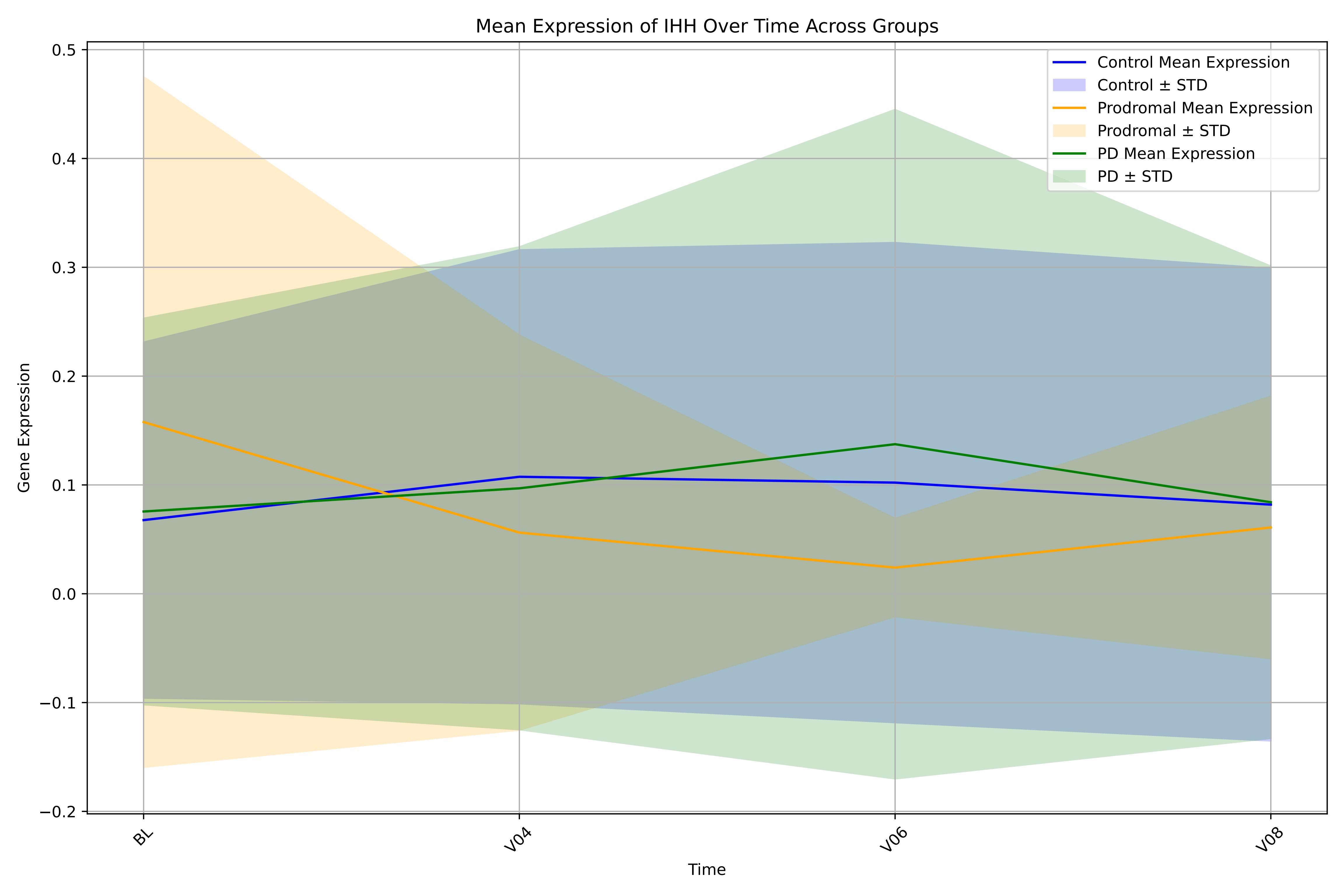

Mean Expression of IHH Over Time Across Groups
Control Mean Expression
Control ± STD
Prodromal Mean Expression
Prodromal ± STD
PD Mean Expression
PD ± STD
Gene Expression
Time
BL
V04
V06
V08
0.5
0.4
0.3
0.2
0.1
0.0
−0.1
−0.2

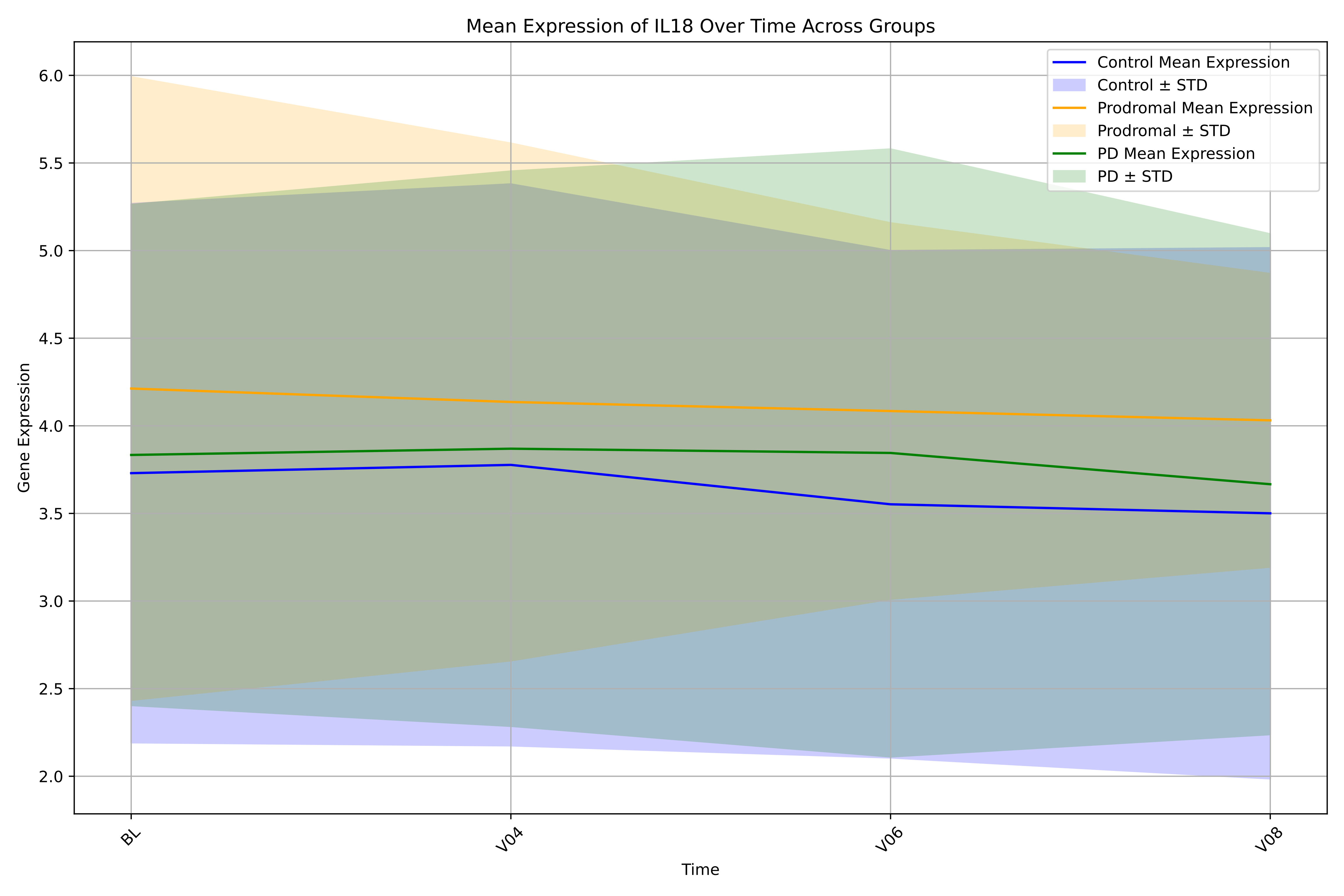

Mean Expression of IL18 Over Time Across Groups
Control Mean Expression
Control ± STD
Prodromal Mean Expression
Prodromal ± STD
PD Mean Expression
PD ± STD
6.0
5.5
5.0
4.5
4.0
3.5
3.0
2.5
2.0
Gene Expression
BL
V04
V06
V08
Time

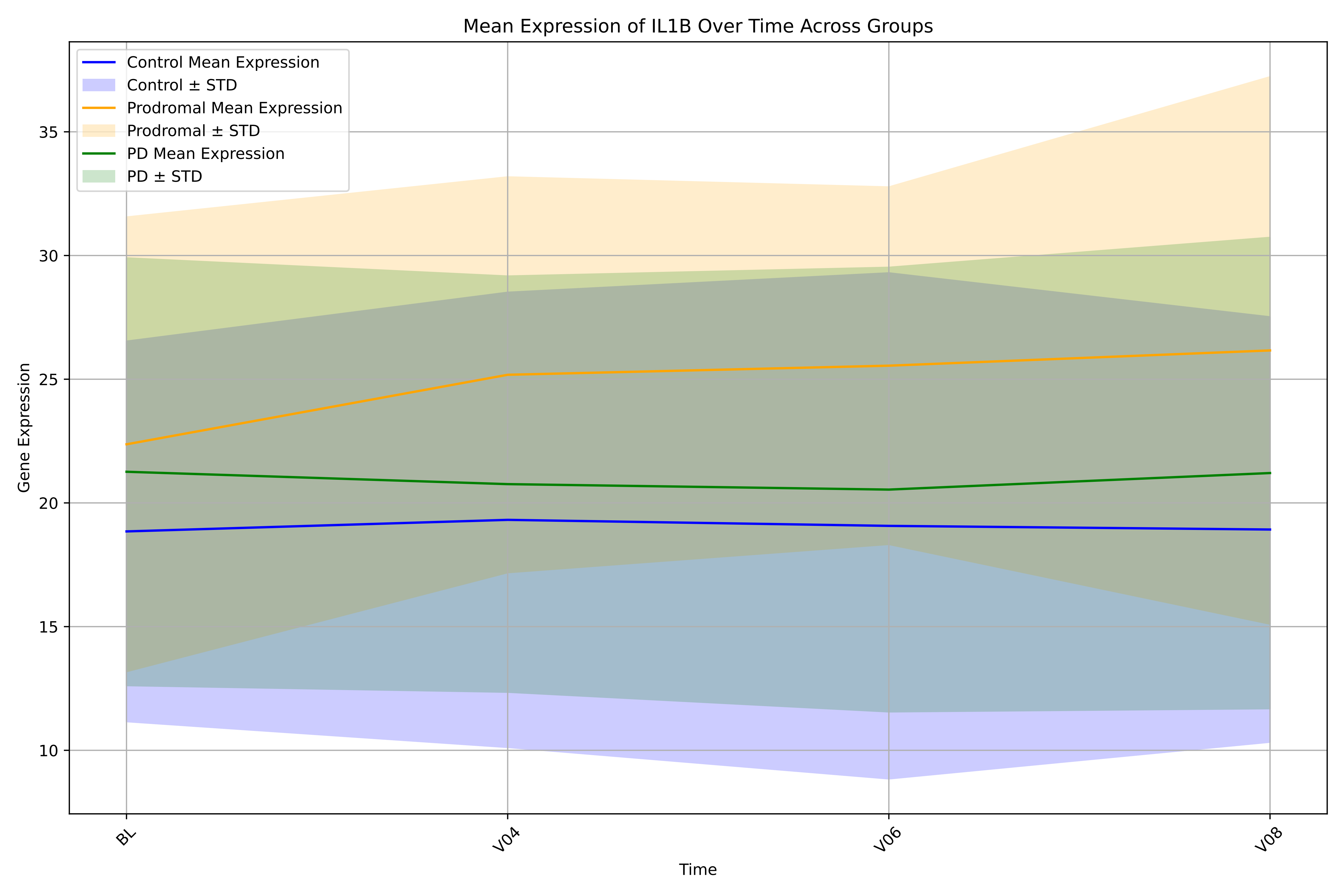

Mean Expression of IL1B Over Time Across Groups
Control Mean Expression
Control ± STD
Prodromal Mean Expression
Prodromal ± STD
PD Mean Expression
PD ± STD
Gene Expression
35
30
25
20
15
10
BL
V04
V06
V08
Time

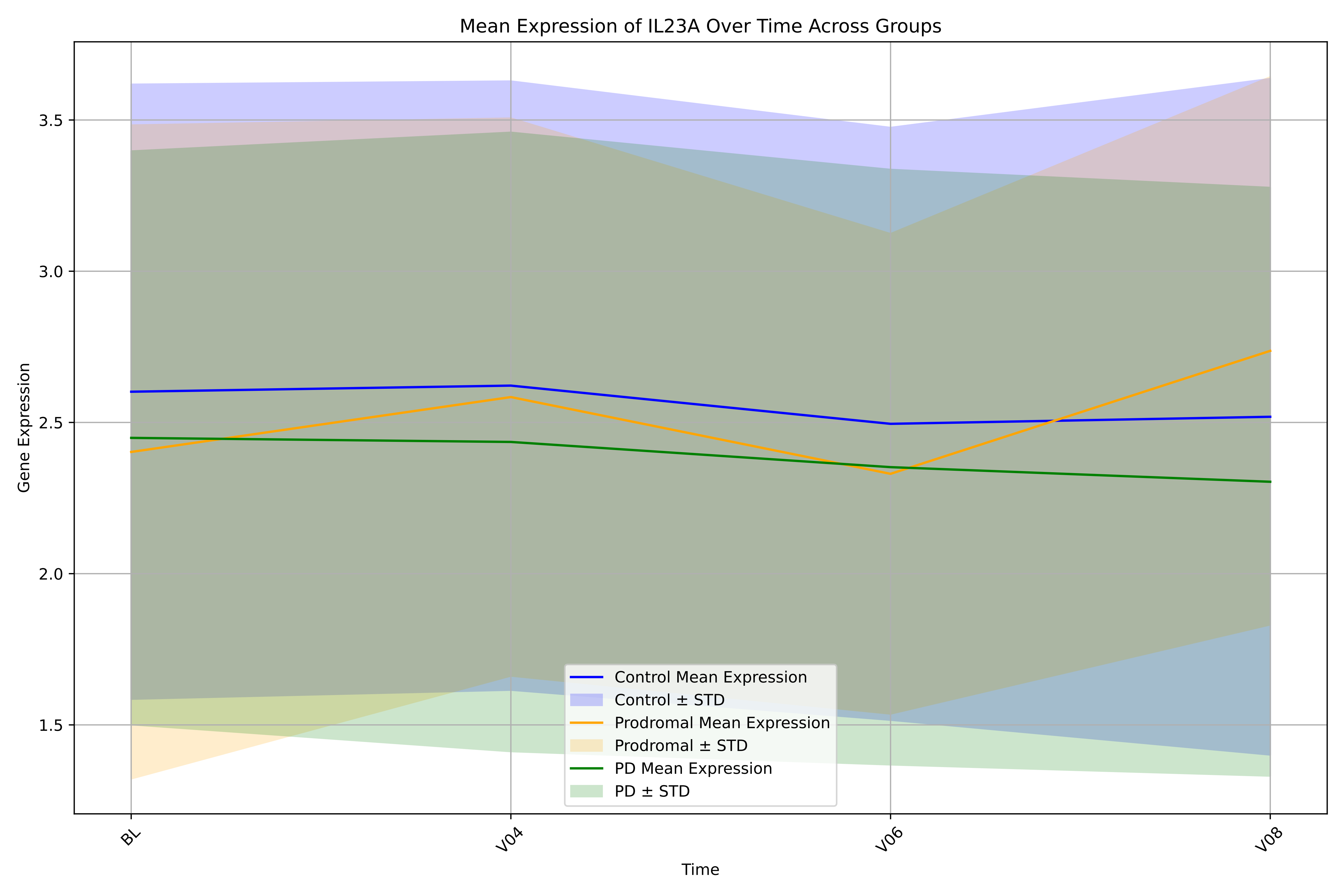

Mean Expression of IL23A Over Time Across Groups
Gene Expression
Time
3.5
3.0
2.5
2.0
1.5
BL
V04
V06
V08
Control Mean Expression
Control ± STD
Prodromal Mean Expression
Prodromal ± STD
PD Mean Expression
PD ± STD

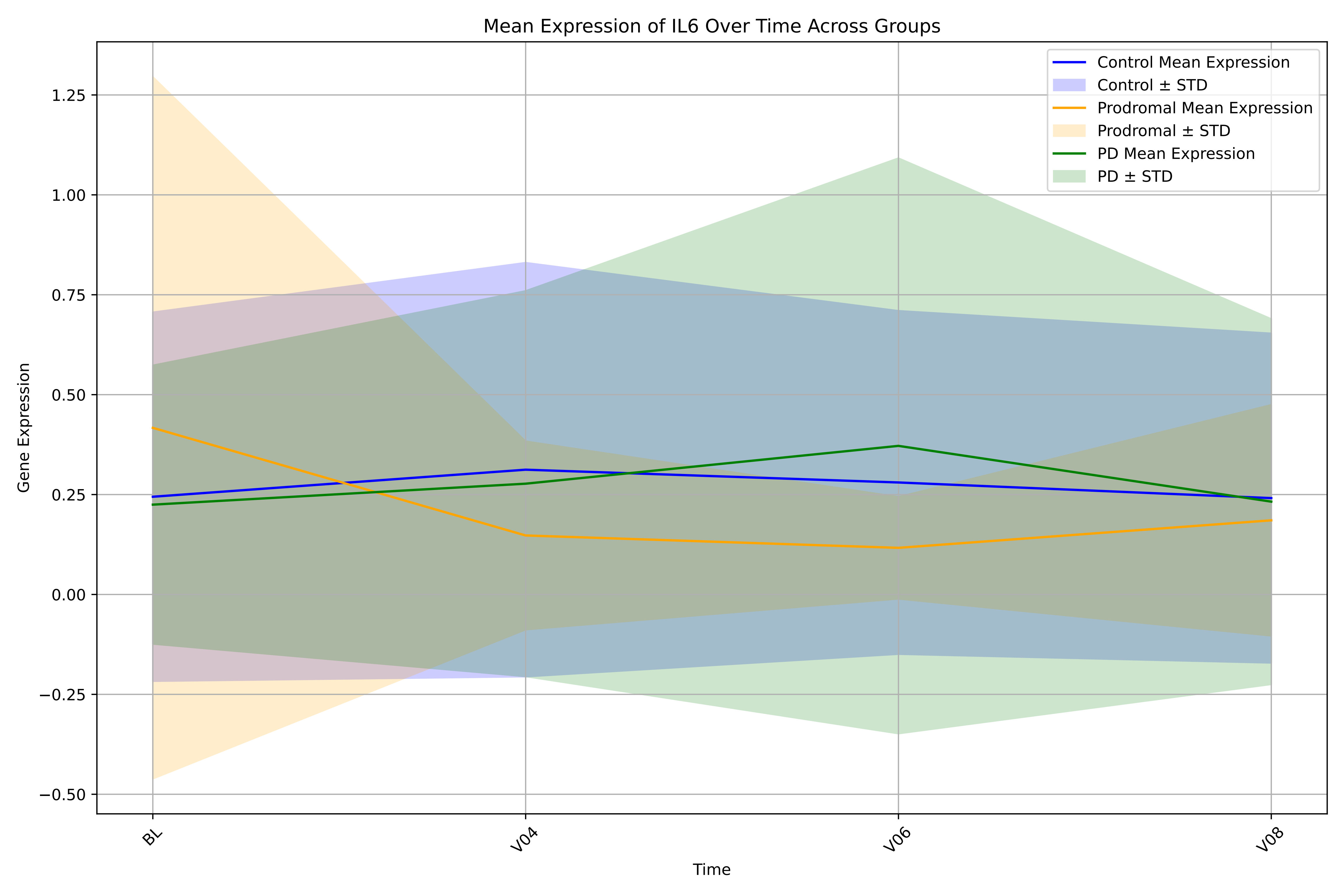

Mean Expression of IL6 Over Time Across Groups
Control Mean Expression
Control ± STD
Prodromal Mean Expression
Prodromal ± STD
PD Mean Expression
PD ± STD
Gene Expression
1.25
1.00
0.75
0.50
0.25
0.00
−0.25
−0.50
BL
V04
V06
V08
Time

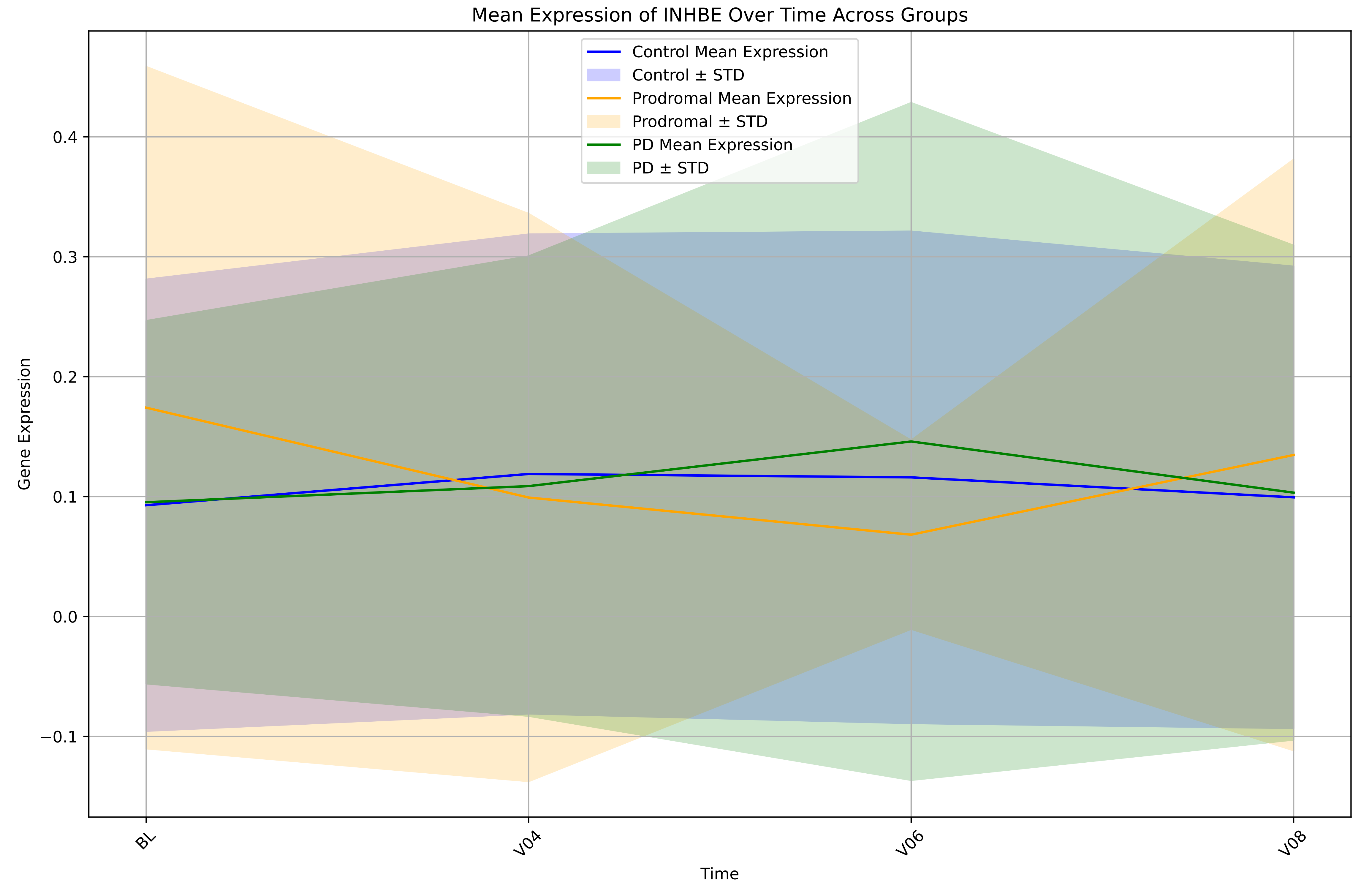

Mean Expression of INHBE Over Time Across Groups
Control Mean Expression
Control ± STD
Prodromal Mean Expression
Prodromal ± STD
PD Mean Expression
PD ± STD
Gene Expression
0.4
0.3
0.2
0.1
0.0
−0.1
BL
V04
V06
V08
Time

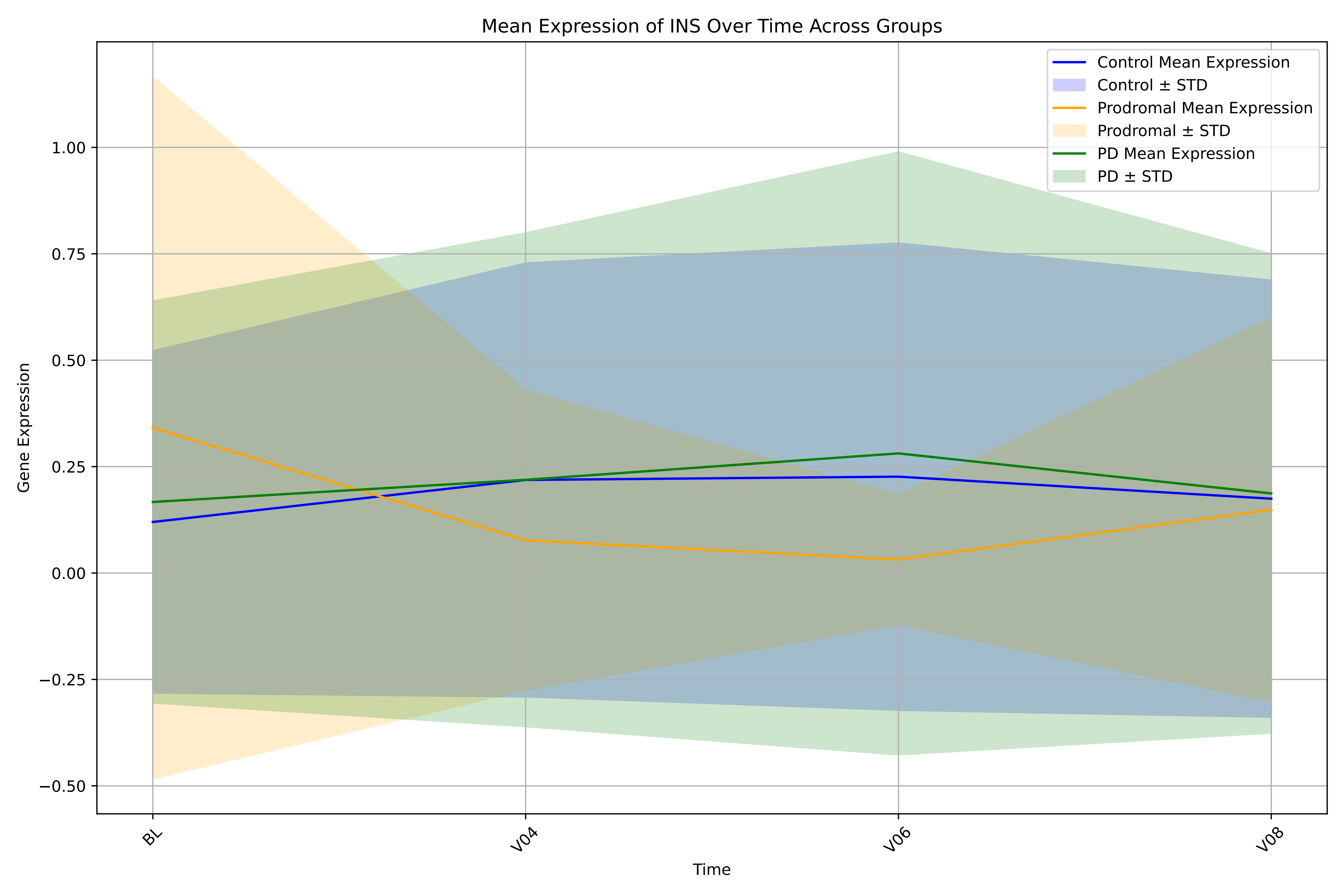

Mean Expression of INS Over Time Across Groups
Control Mean Expression
Control ± STD
Prodromal Mean Expression
Prodromal ± STD
PD Mean Expression
PD ± STD
Gene Expression
1.00
0.75
0.50
0.25
0.00
−0.25
−0.50
BL
V04
V06
V08
Time

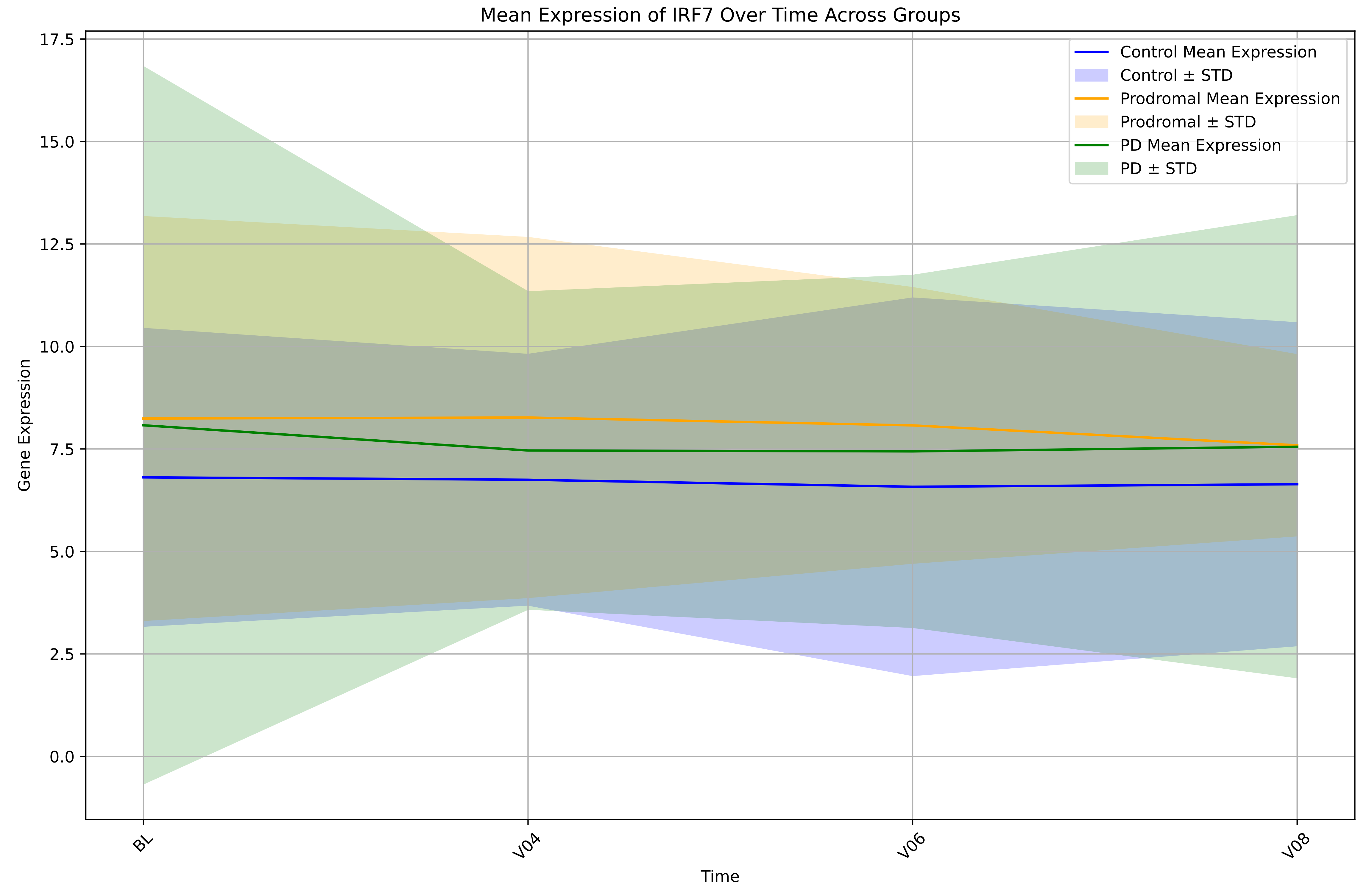
Mean Expression of IRF7 Over Time Across Groups
Control Mean Expression
Control ± STD
Prodromal Mean Expression
Prodromal ± STD
PD Mean Expression
PD ± STD
Gene Expression
17.5
15.0
12.5
10.0
7.5
5.0
2.5
0.0
BL
V04
V06
V08
Time

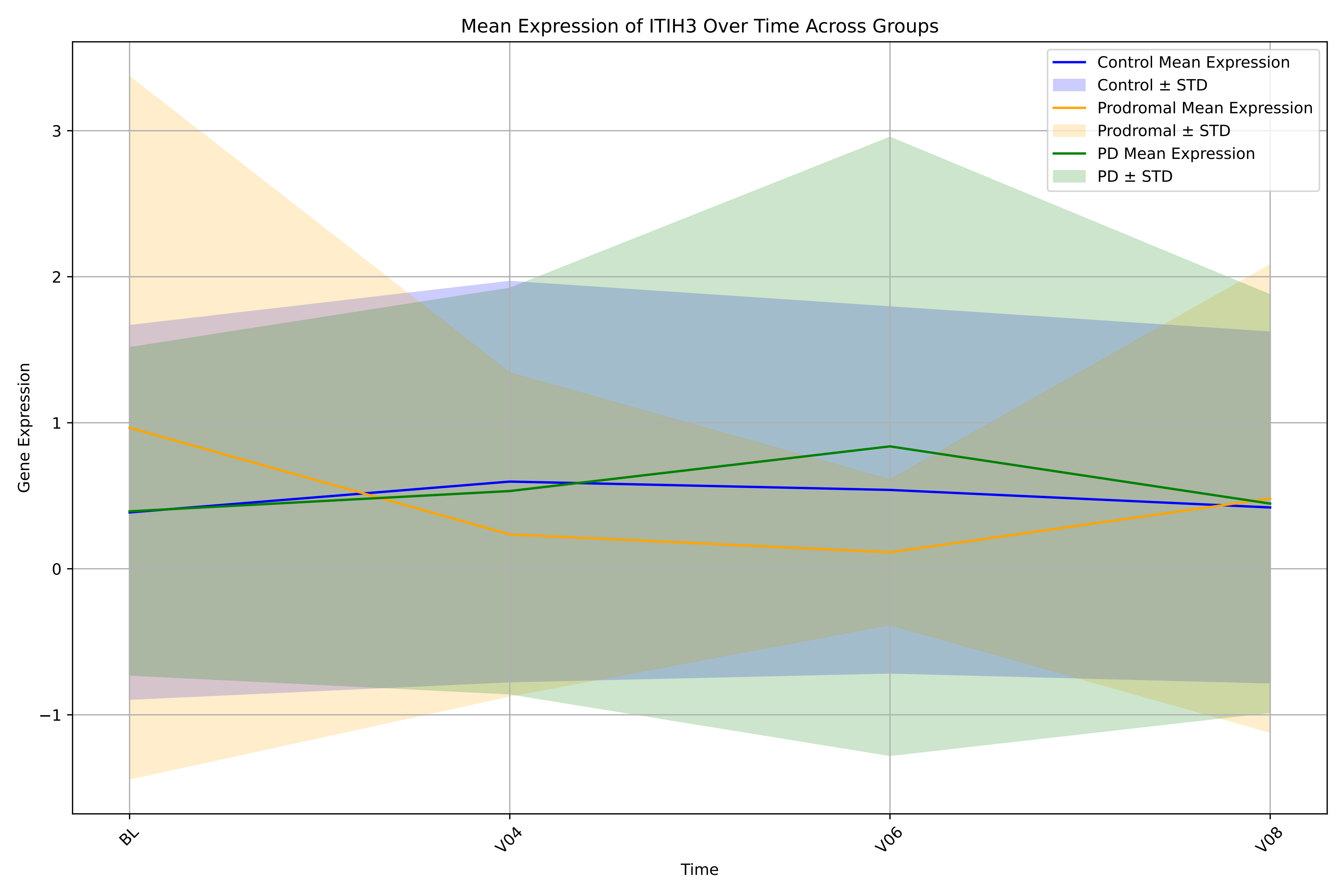
Mean Expression of ITIH3 Over Time Across Groups
Control Mean Expression
Control ± STD
Prodromal Mean Expression
Prodromal ± STD
PD Mean Expression
PD ± STD
Gene Expression
3
2
1
0
−1
BL
V04
V06
V08
Time

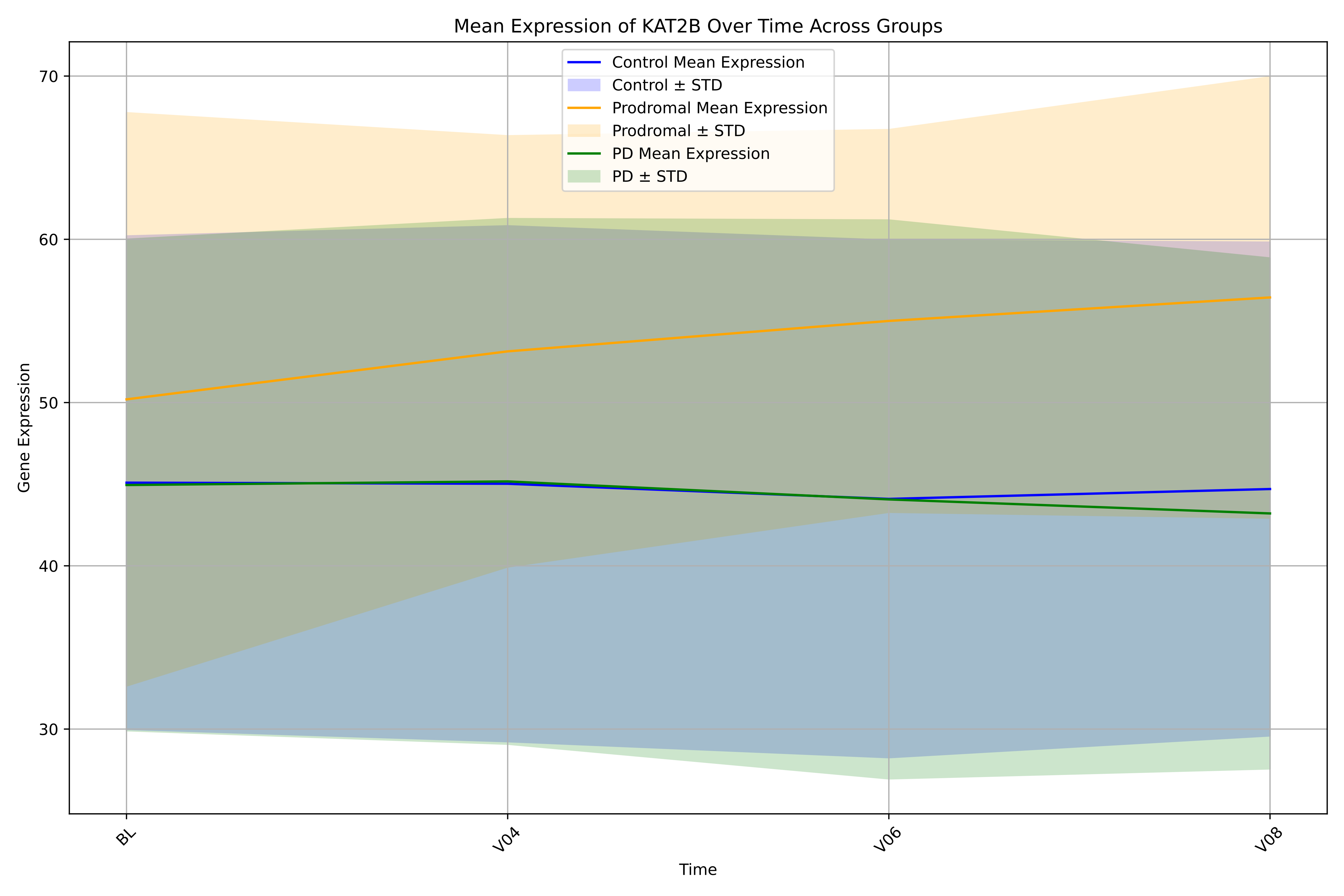

Mean Expression of KAT2B Over Time Across Groups
Control Mean Expression
Control ± STD
Prodromal Mean Expression
Prodromal ± STD
PD Mean Expression
PD ± STD
70
60
50
40
30
Gene Expression
BL
V04
V06
V08
Time

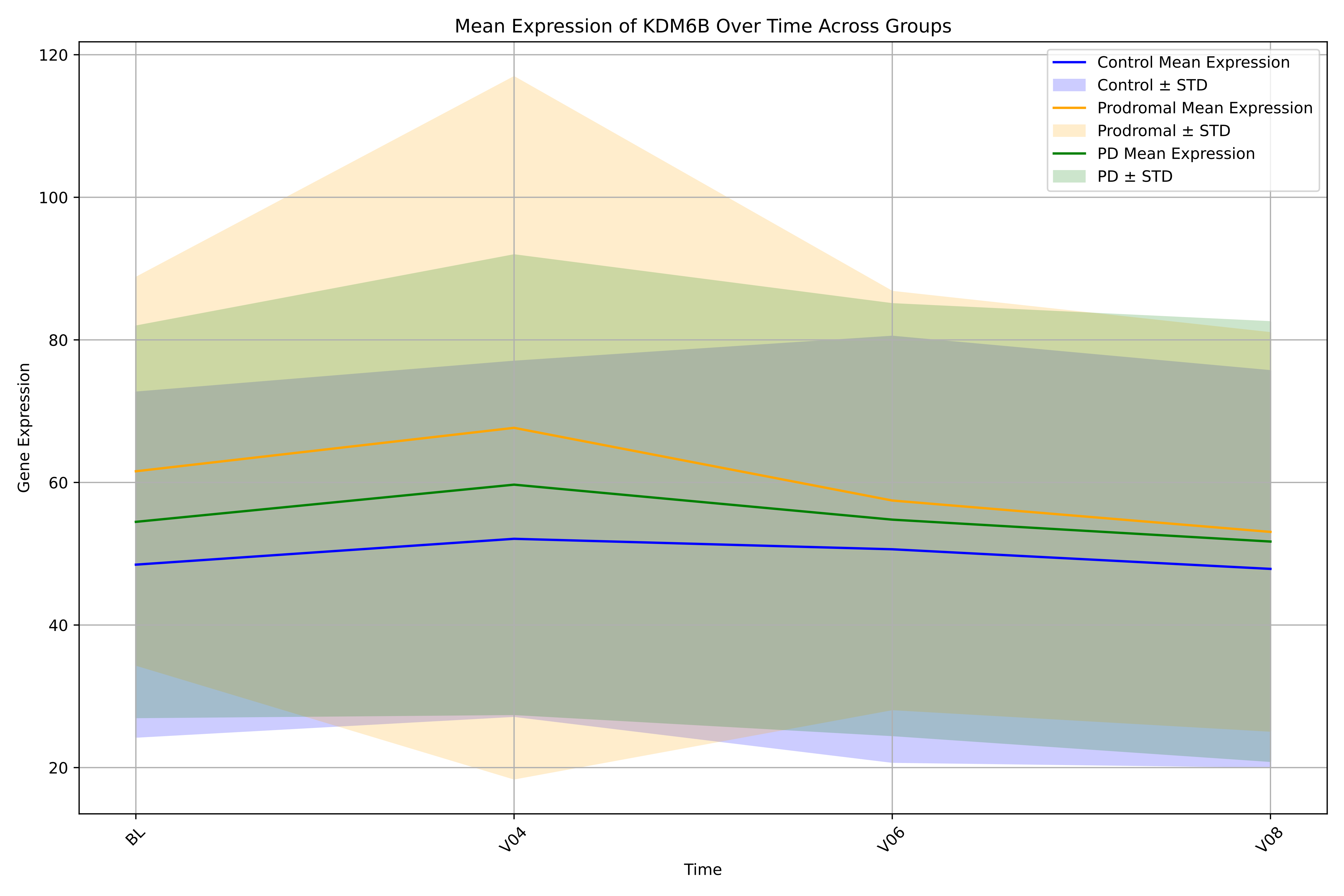

Mean Expression of KDM6B Over Time Across Groups
Control Mean Expression
Control ± STD
Prodromal Mean Expression
Prodromal ± STD
PD Mean Expression
PD ± STD
Gene Expression
Time
120
100
80
60
40
20
BL
V04
V06
V08

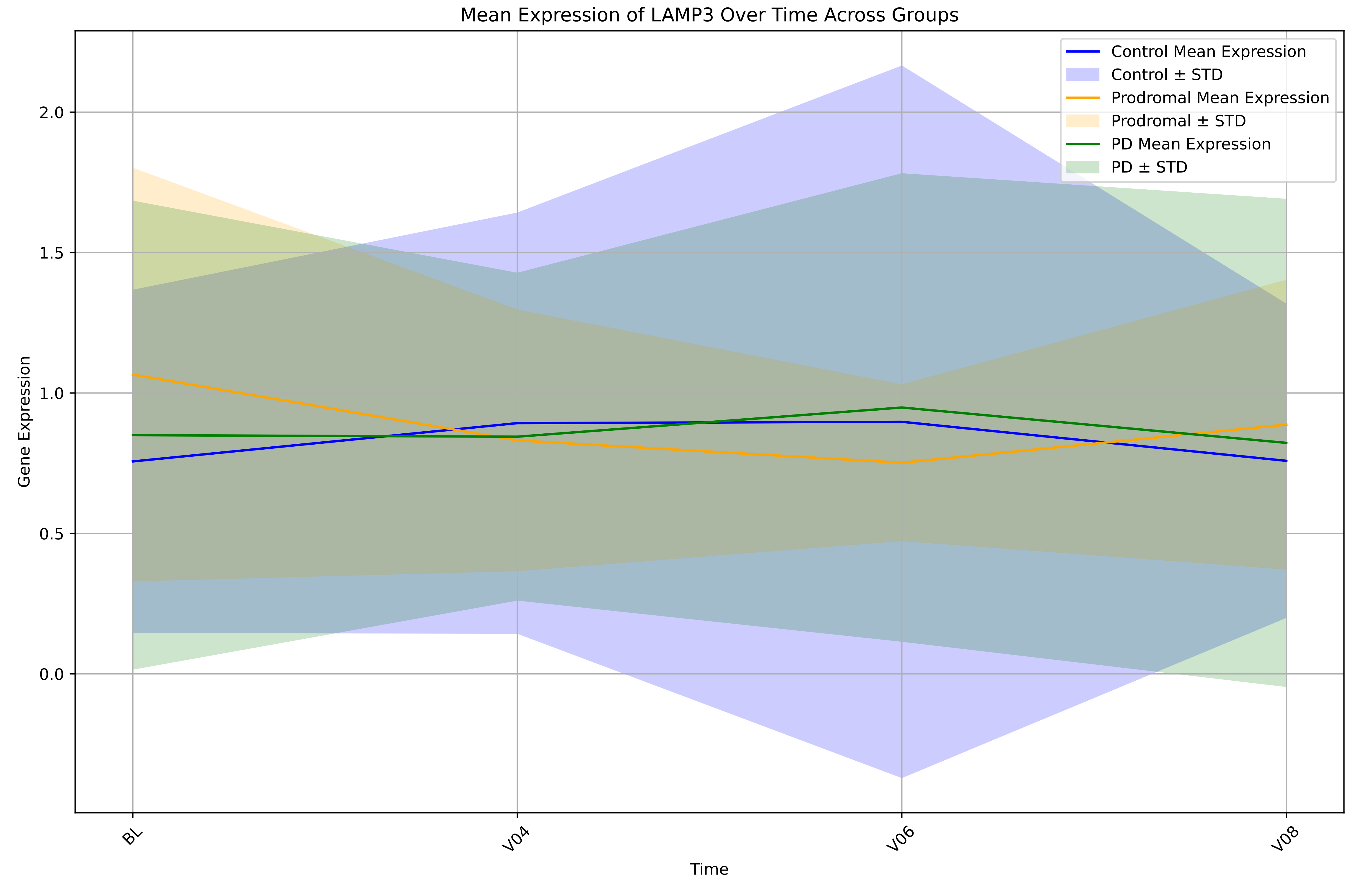
Mean Expression of LAMP3 Over Time Across Groups
Control Mean Expression
Control ± STD
Prodromal Mean Expression
Prodromal ± STD
PD Mean Expression
PD ± STD
Gene Expression
2.0
1.5
1.0
0.5
0.0
BL
V04
V06
V08
Time

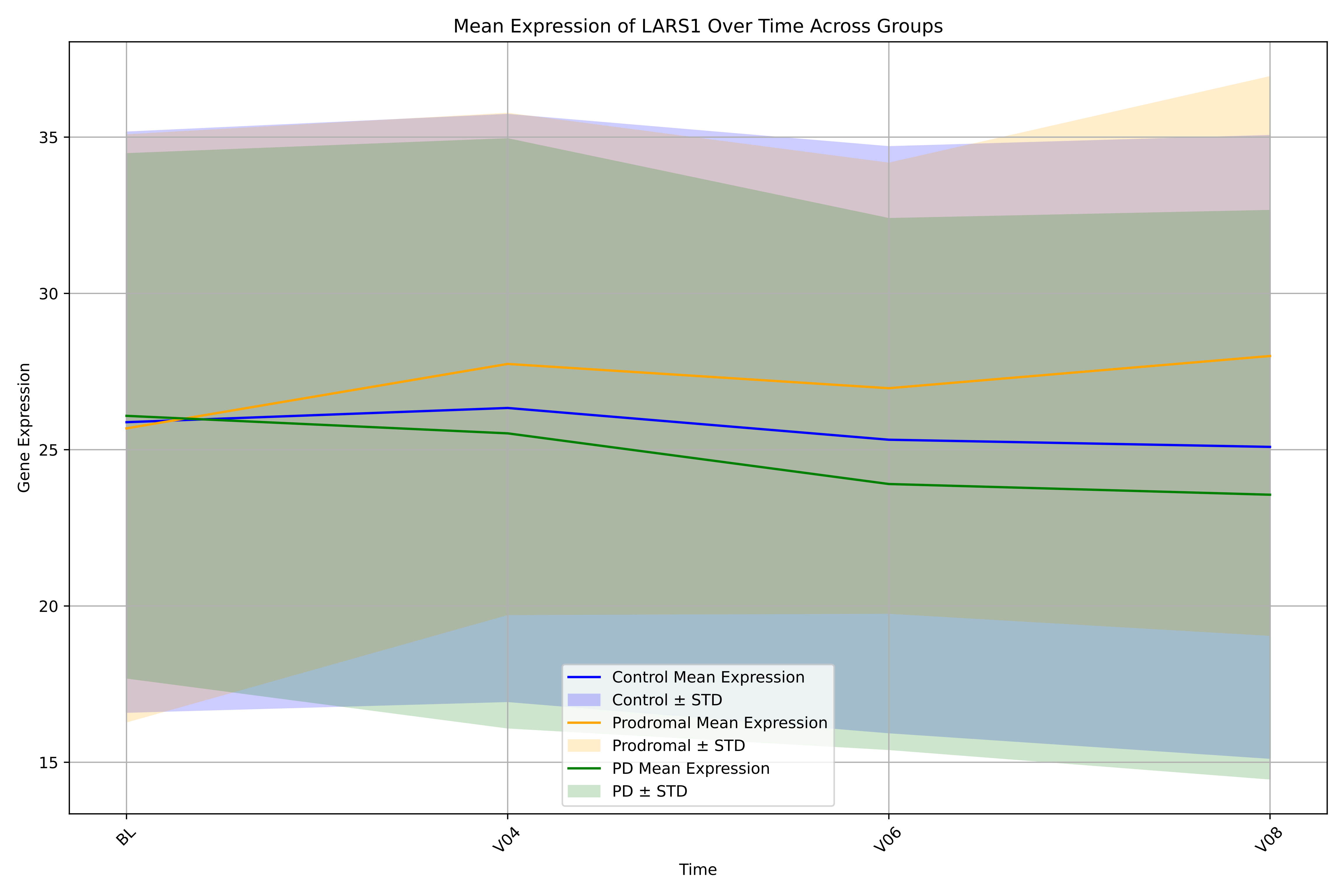
Mean Expression of LARS1 Over Time Across Groups
Gene Expression
Time
35
30
25
20
15
BL
V04
V06
V08
Control Mean Expression
Control ± STD
Prodromal Mean Expression
Prodromal ± STD
PD Mean Expression
PD ± STD

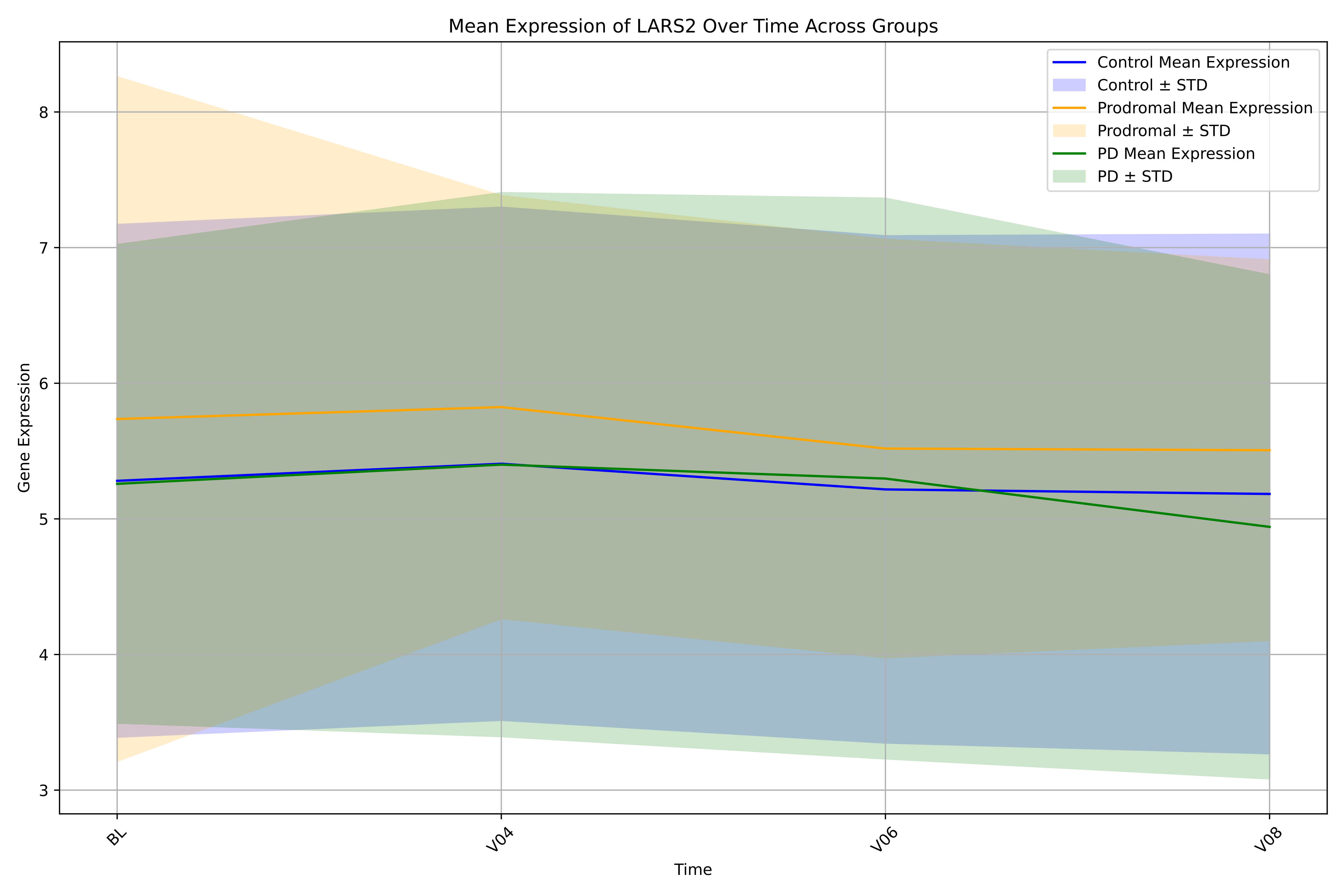
Mean Expression of LARS2 Over Time Across Groups
Control Mean Expression
Control ± STD
Prodromal Mean Expression
Prodromal ± STD
PD Mean Expression
PD ± STD
Gene Expression
3
4
5
6
7
8
Time
BL
V04
V06
V08

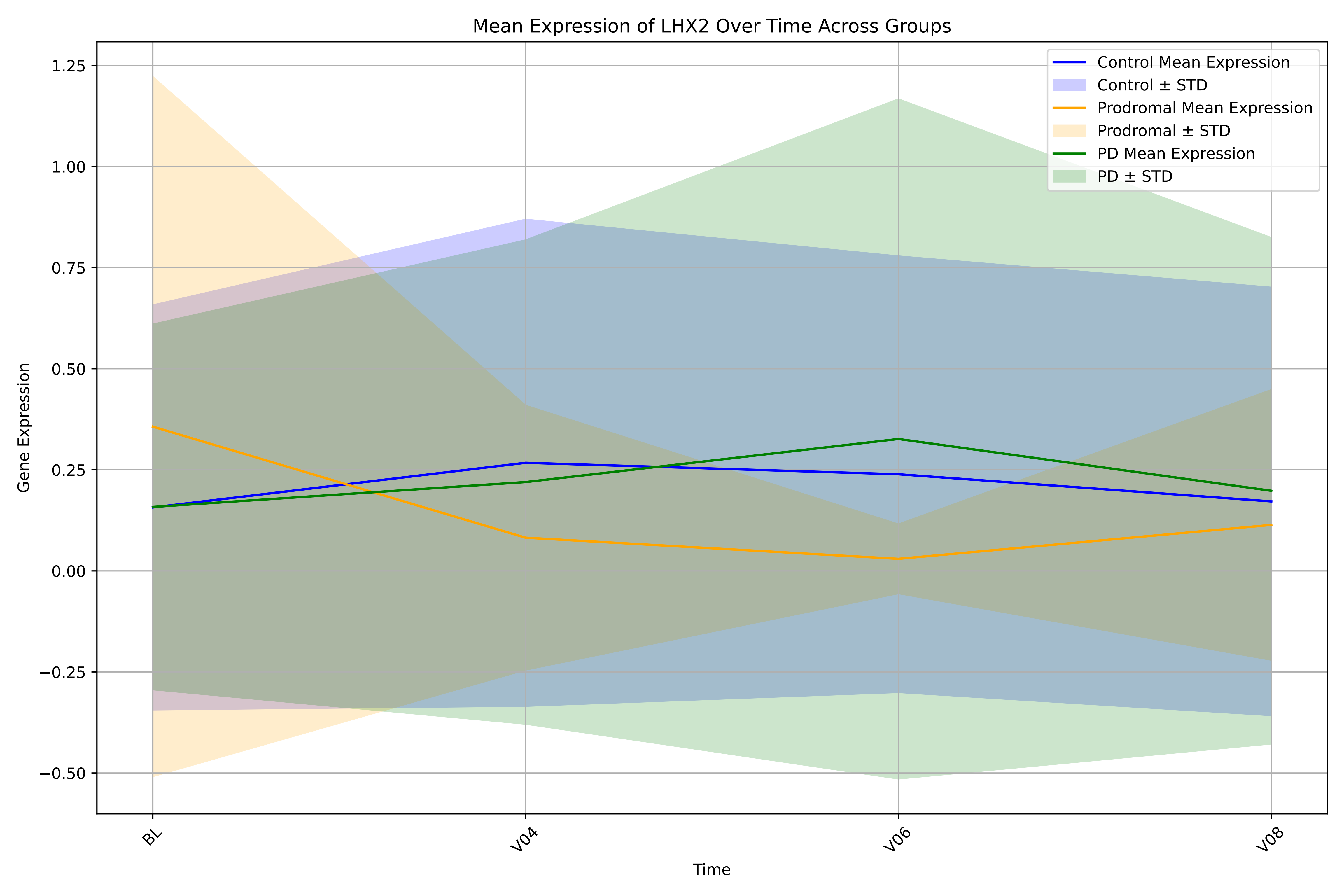

Mean Expression of LHX2 Over Time Across Groups
Control Mean Expression
Control ± STD
Prodromal Mean Expression
Prodromal ± STD
PD Mean Expression
PD ± STD
Gene Expression
Time
1.25
1.00
0.75
0.50
0.25
0.00
−0.25
−0.50
BL
V04
V06
V08

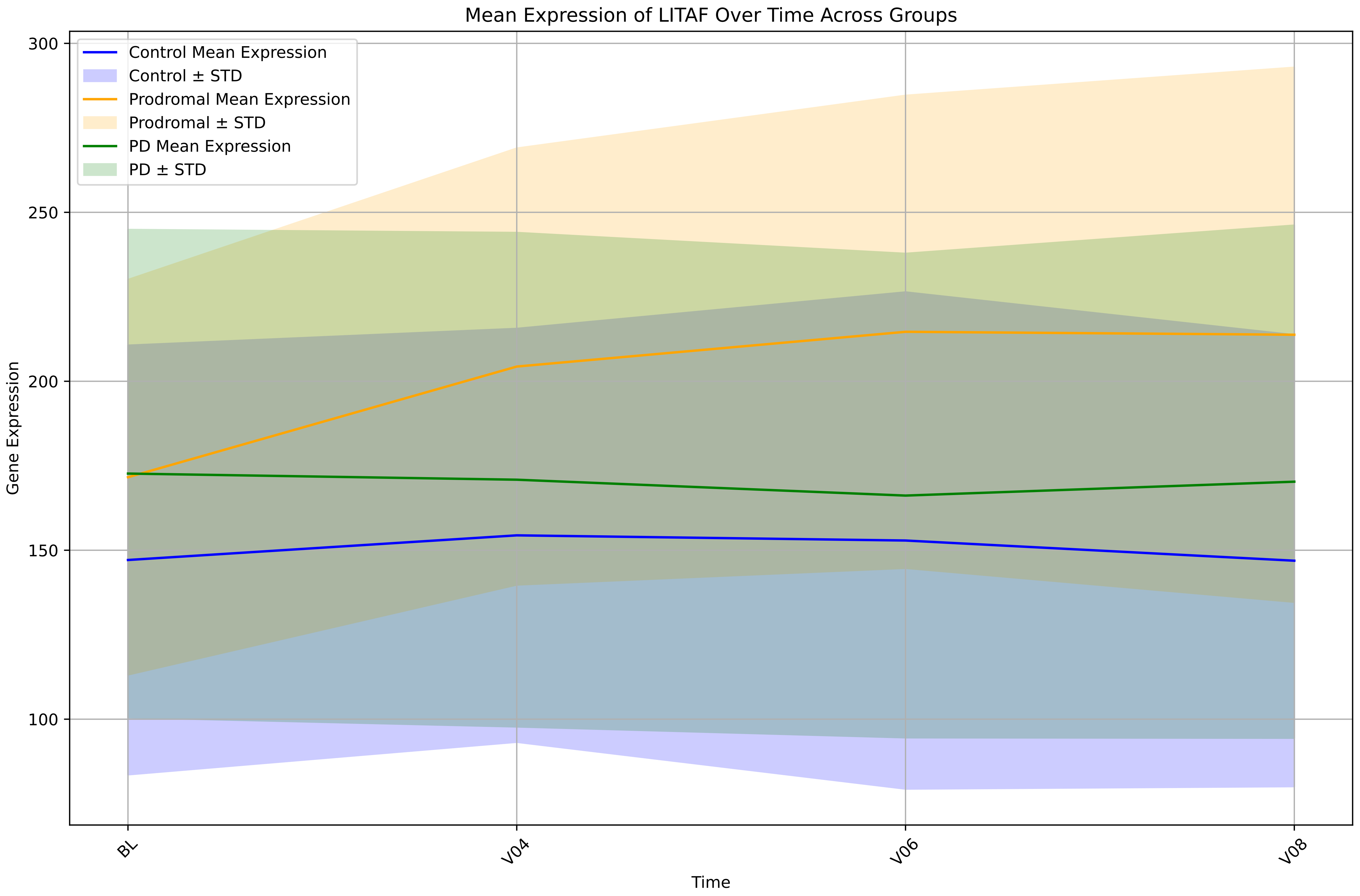

Mean Expression of LITAF Over Time Across Groups
Control Mean Expression
Control ± STD
Prodromal Mean Expression
Prodromal ± STD
PD Mean Expression
PD ± STD
Gene Expression
300
250
200
150
100
BL
V04
V06
V08
Time

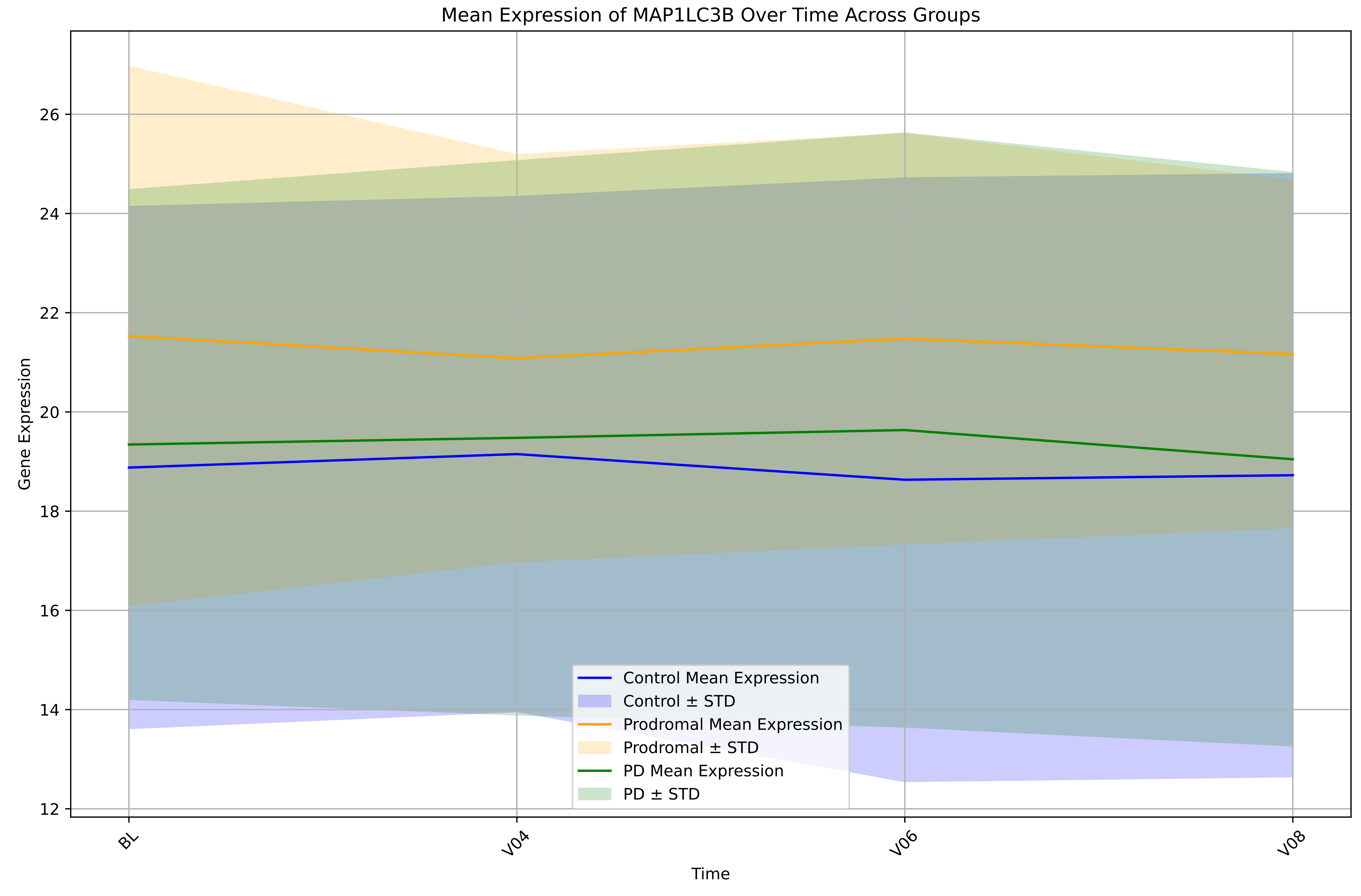

Mean Expression of MAP1LC3B Over Time Across Groups
Gene Expression
Time
12
14
16
18
20
22
24
26
BL
V04
V06
V08
Control Mean Expression
Control ± STD
Prodromal Mean Expression
Prodromal ± STD
PD Mean Expression
PD ± STD

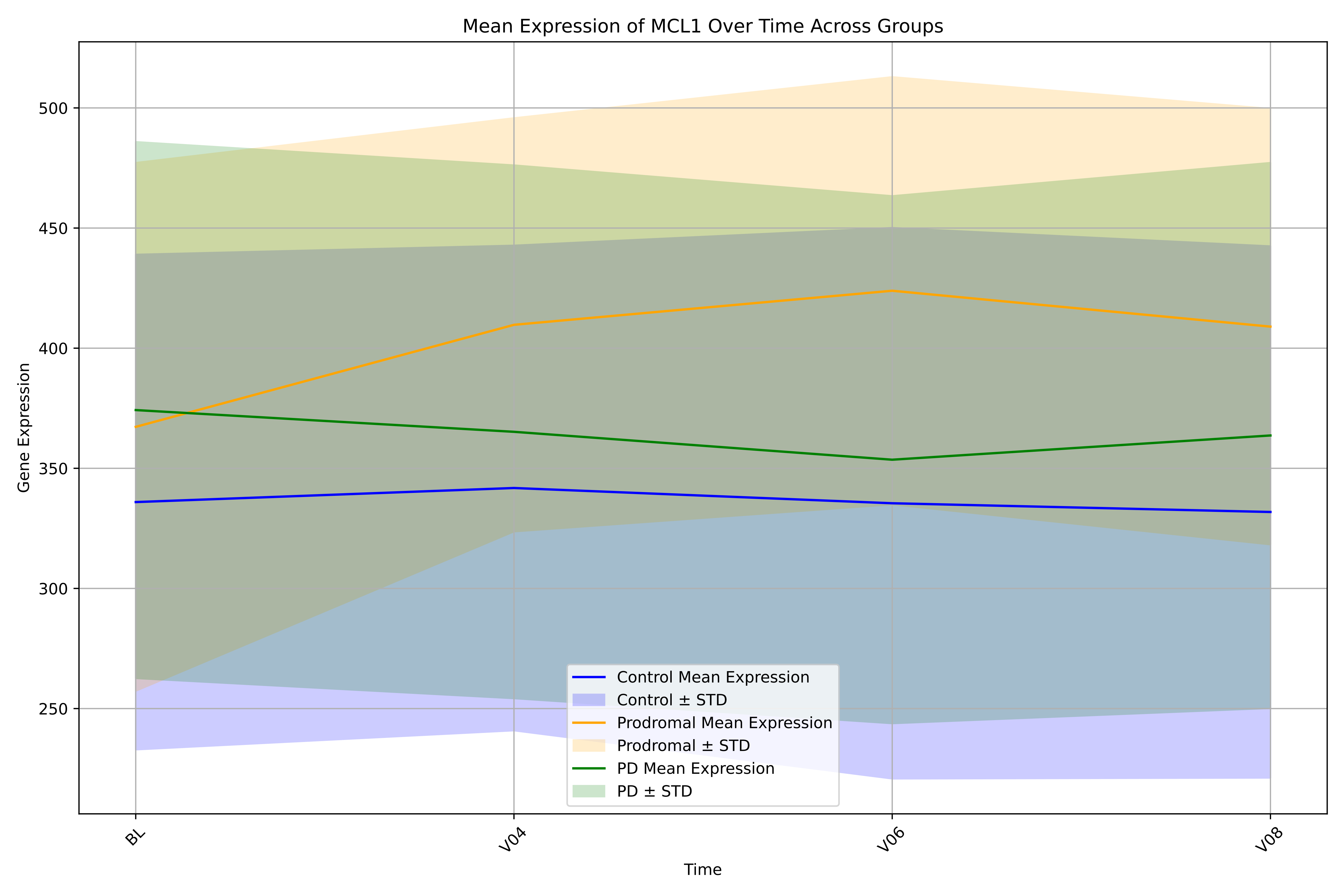
Mean Expression of MCL1 Over Time Across Groups
Gene Expression
Time
500
450
400
350
300
250
BL
V04
V06
V08
Control Mean Expression
Control ± STD
Prodromal Mean Expression
Prodromal ± STD
PD Mean Expression
PD ± STD

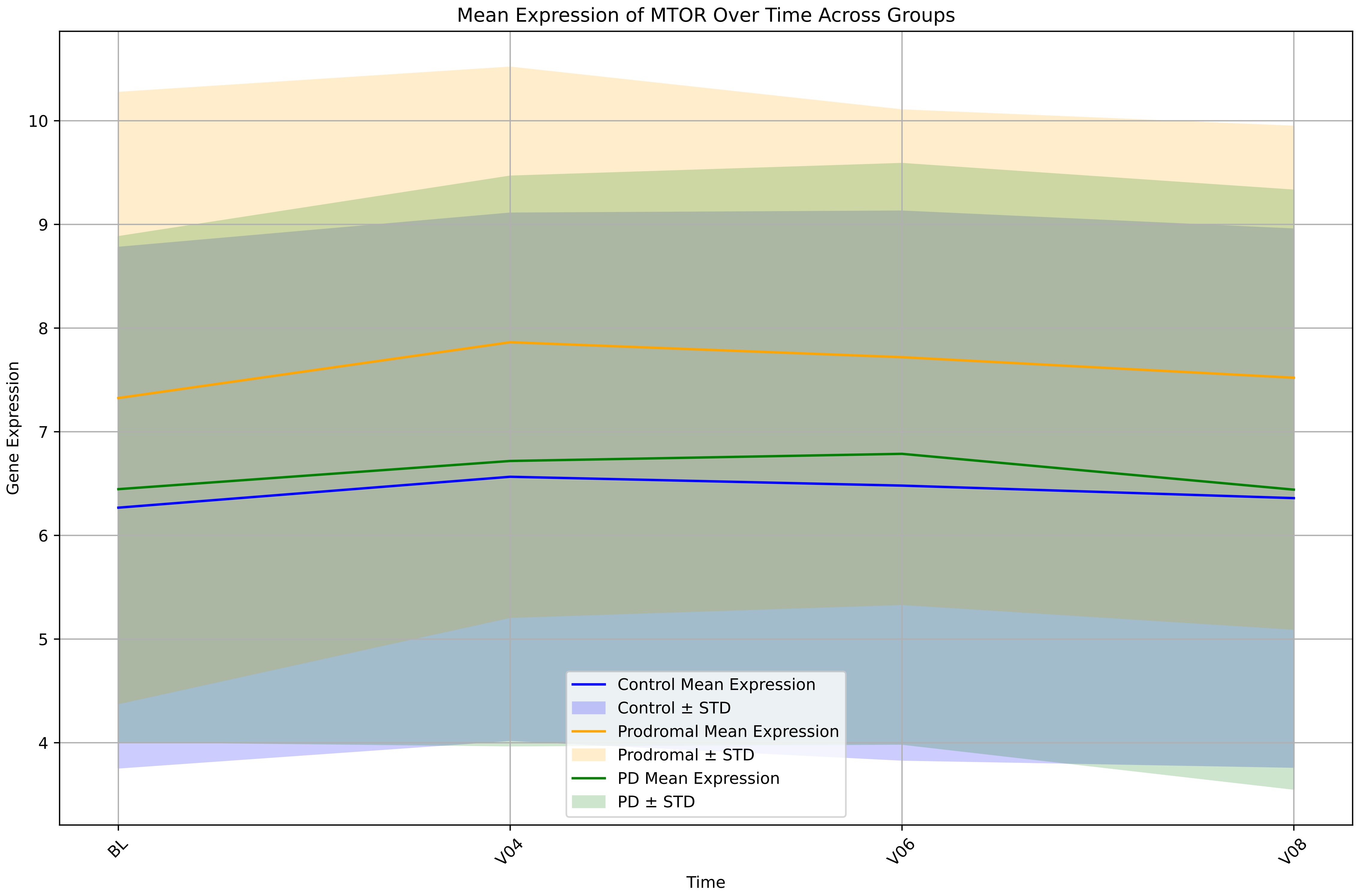

Mean Expression of MTOR Over Time Across Groups
Gene Expression
Time
BL
V04
V06
V08
4
5
6
7
8
9
10
Control Mean Expression
Control ± STD
Prodromal Mean Expression
Prodromal ± STD
PD Mean Expression
PD ± STD

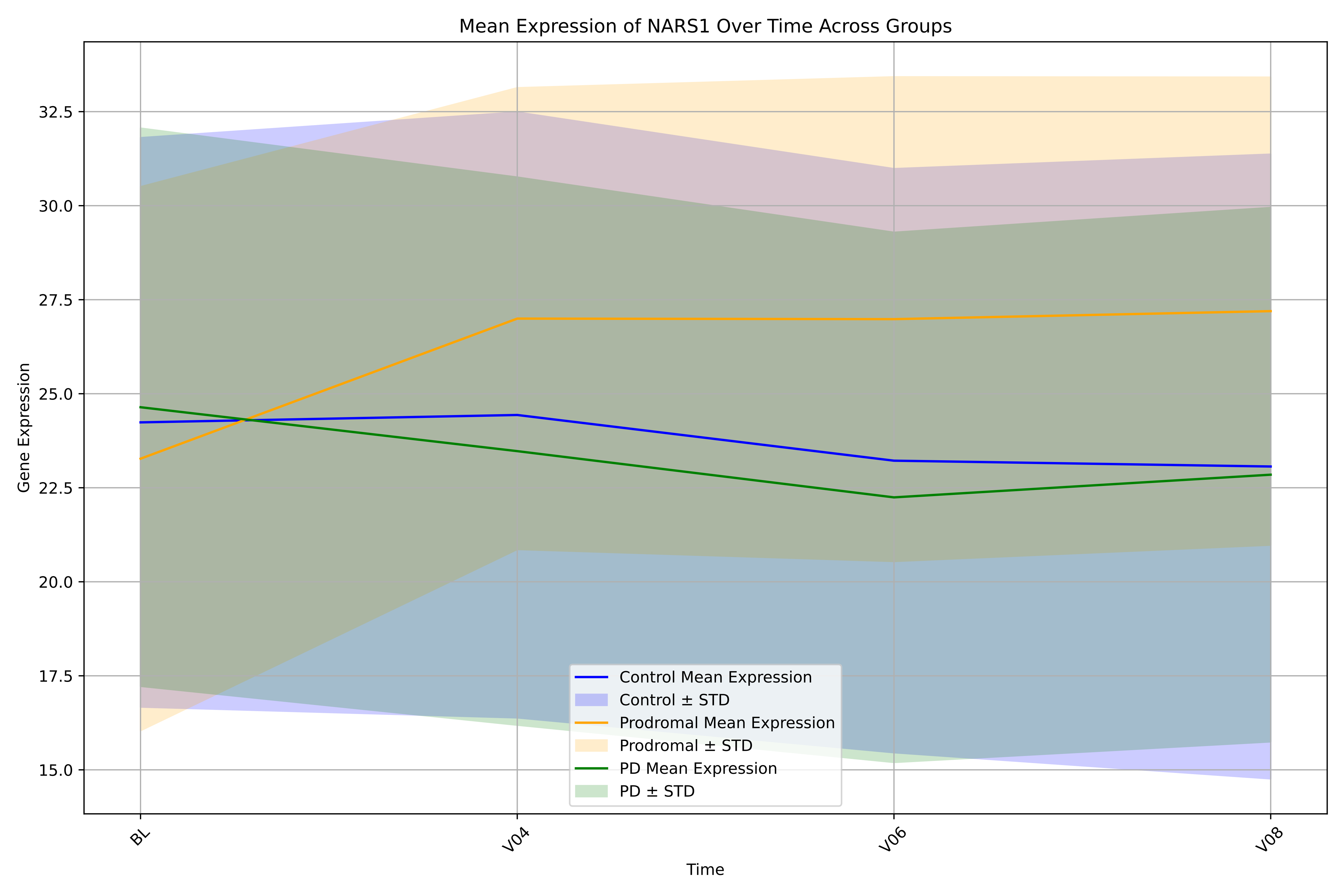
Mean Expression of NARS1 Over Time Across Groups
Gene Expression
32.5
30.0
27.5
25.0
22.5
20.0
17.5
15.0
BL
V04
V06
V08
Time
Control Mean Expression
Control ± STD
Prodromal Mean Expression
Prodromal ± STD
PD Mean Expression
PD ± STD

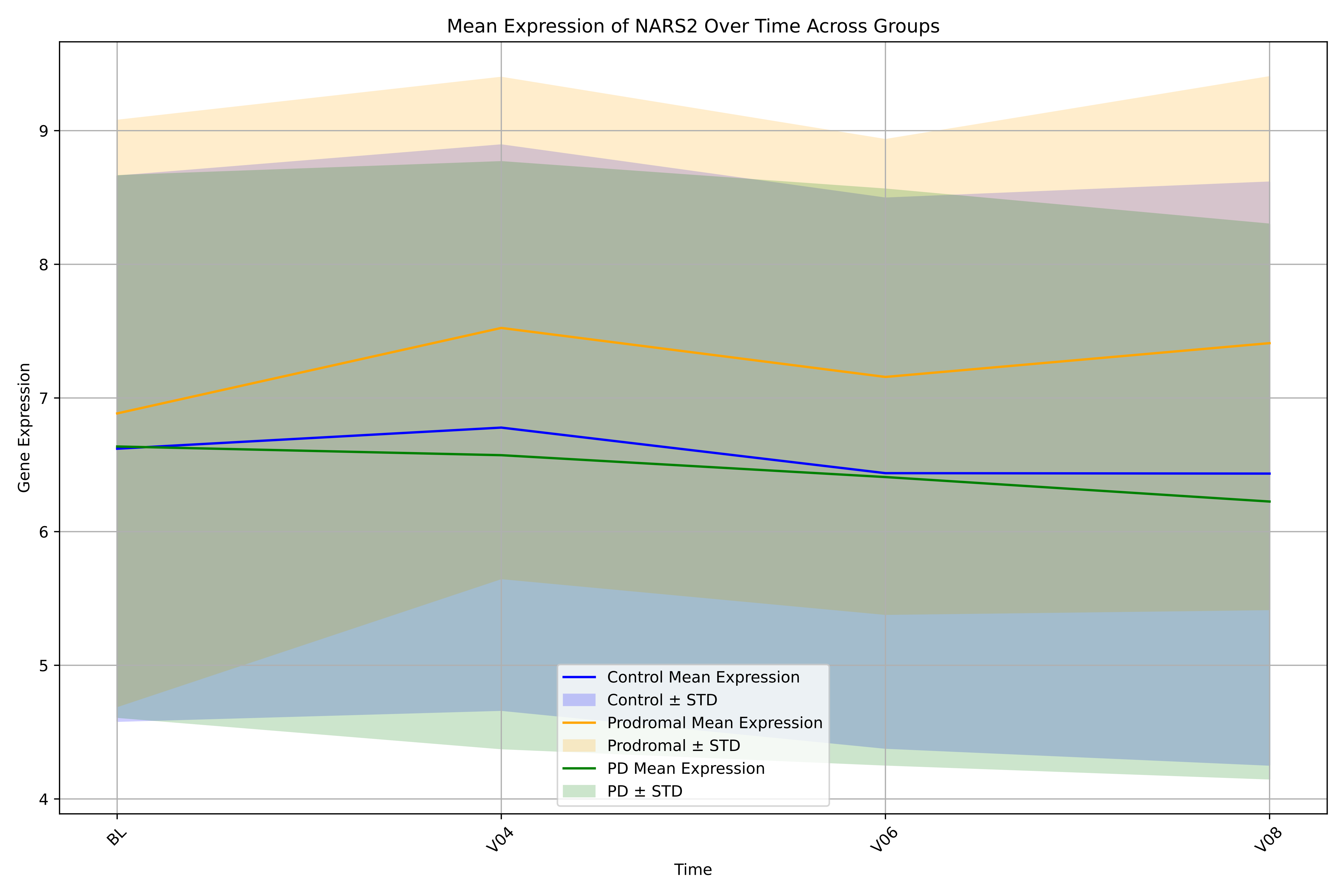
Mean Expression of NARS2 Over Time Across Groups
Gene Expression
Time
BL
V04
V06
V08
4
5
6
7
8
9
Control Mean Expression
Control ± STD
Prodromal Mean Expression
Prodromal ± STD
PD Mean Expression
PD ± STD

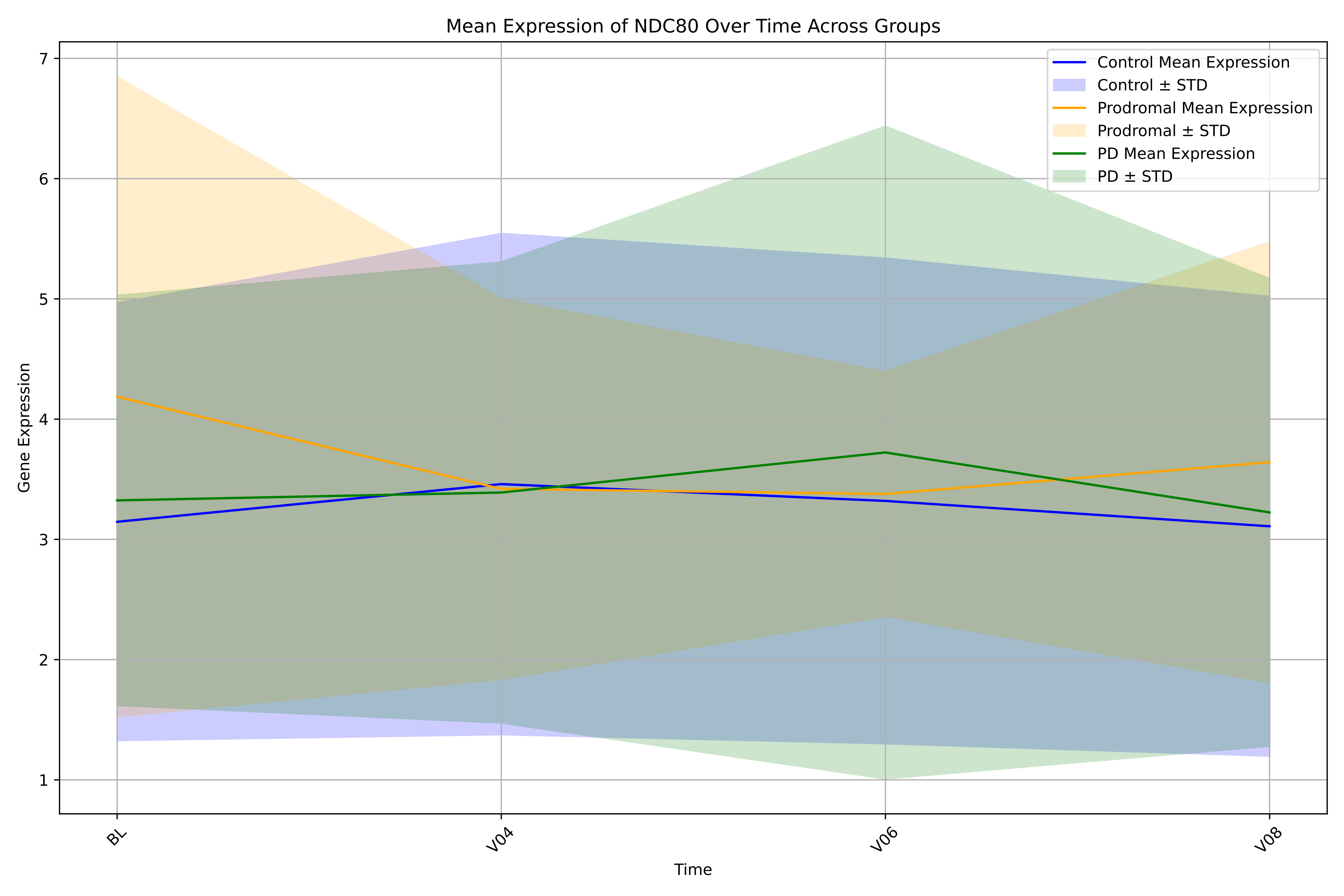

Mean Expression of NDC80 Over Time Across Groups
Control Mean Expression
Control ± STD
Prodromal Mean Expression
Prodromal ± STD
PD Mean Expression
PD ± STD
Gene Expression
1
2
3
4
5
6
7
BL
V04
V06
V08
Time

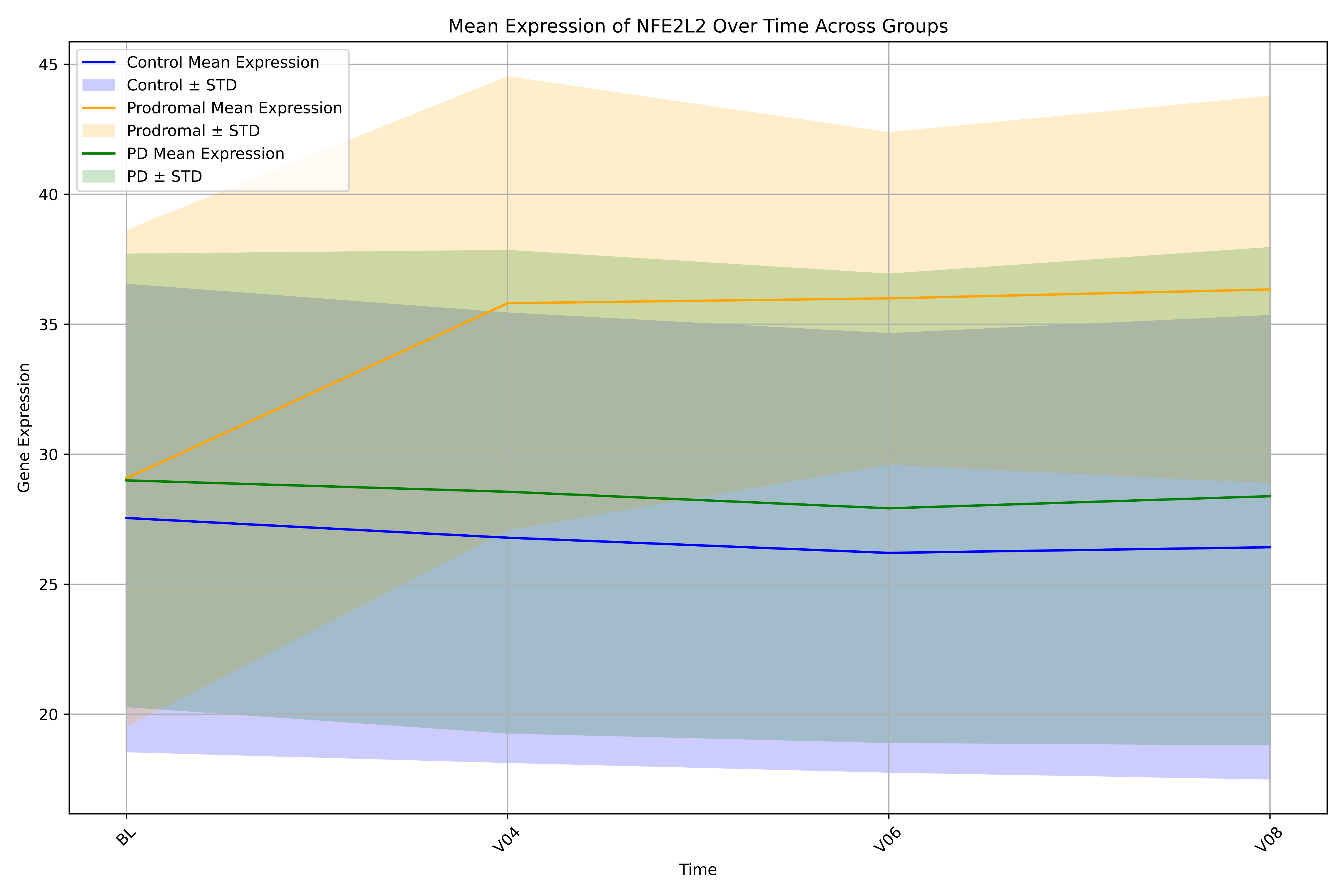
Mean Expression of NFE2L2 Over Time Across Groups
Control Mean Expression
Control ± STD
Prodromal Mean Expression
Prodromal ± STD
PD Mean Expression
PD ± STD
Gene Expression
45
40
35
30
25
20
BL
V04
V06
V08
Time

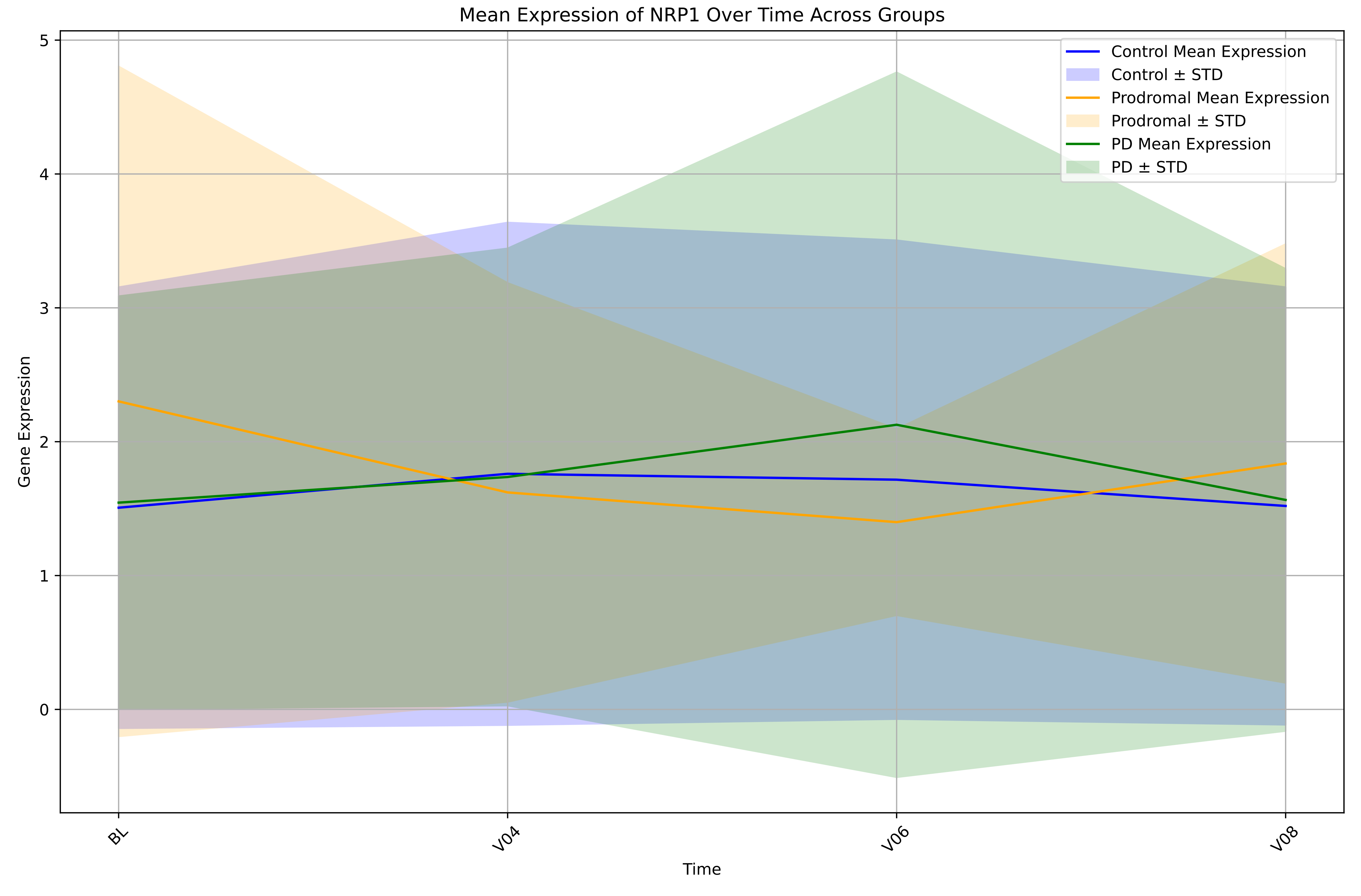

Mean Expression of NRP1 Over Time Across Groups
Control Mean Expression
Control ± STD
Prodromal Mean Expression
Prodromal ± STD
PD Mean Expression
PD ± STD
Gene Expression
0
1
2
3
4
5
BL
V04
V06
V08
Time

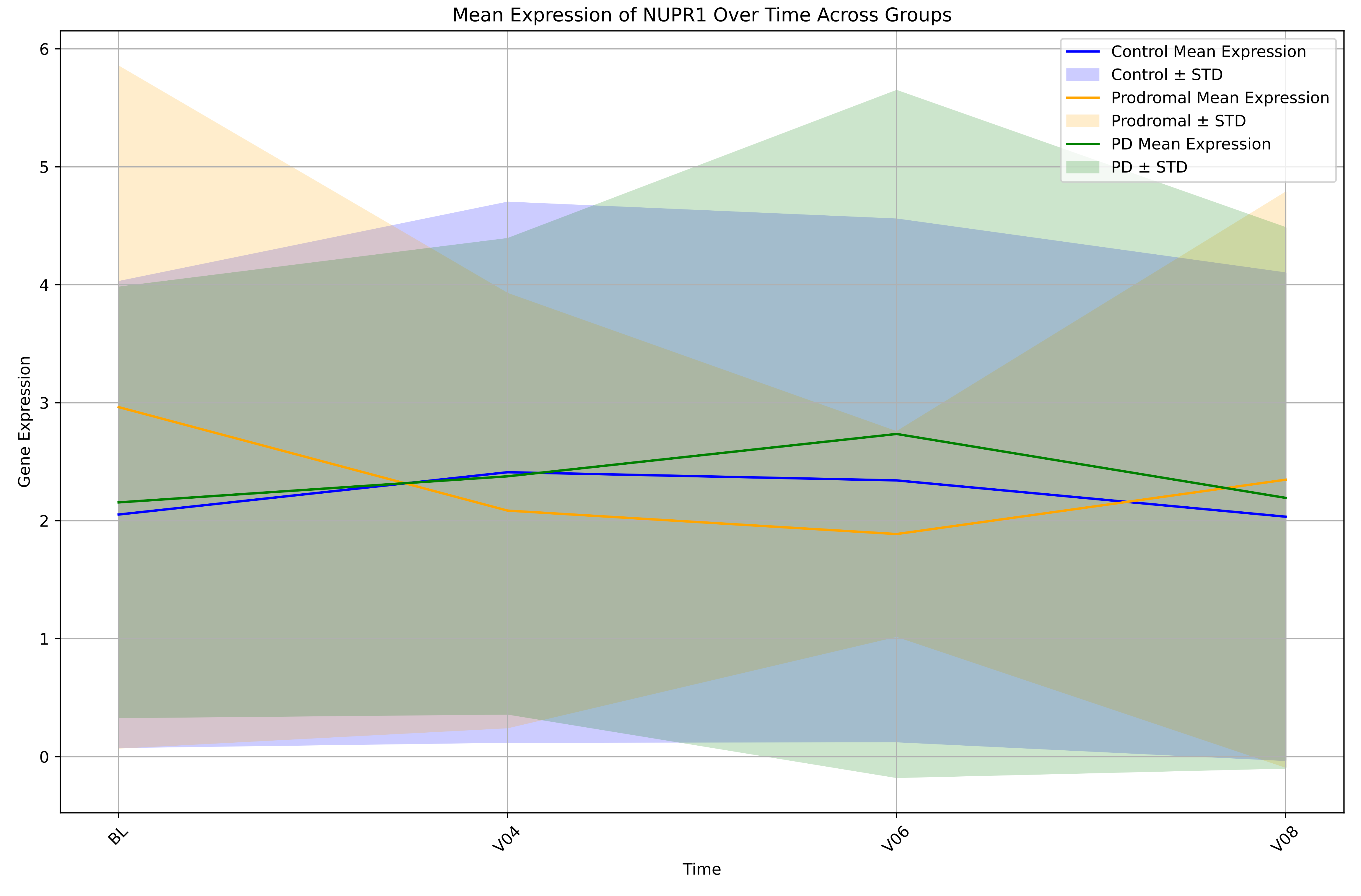

Mean Expression of NUPR1 Over Time Across Groups
Control Mean Expression
Control ± STD
Prodromal Mean Expression
Prodromal ± STD
PD Mean Expression
PD ± STD
Gene Expression
0
1
2
3
4
5
6
BL
V04
V06
V08
Time

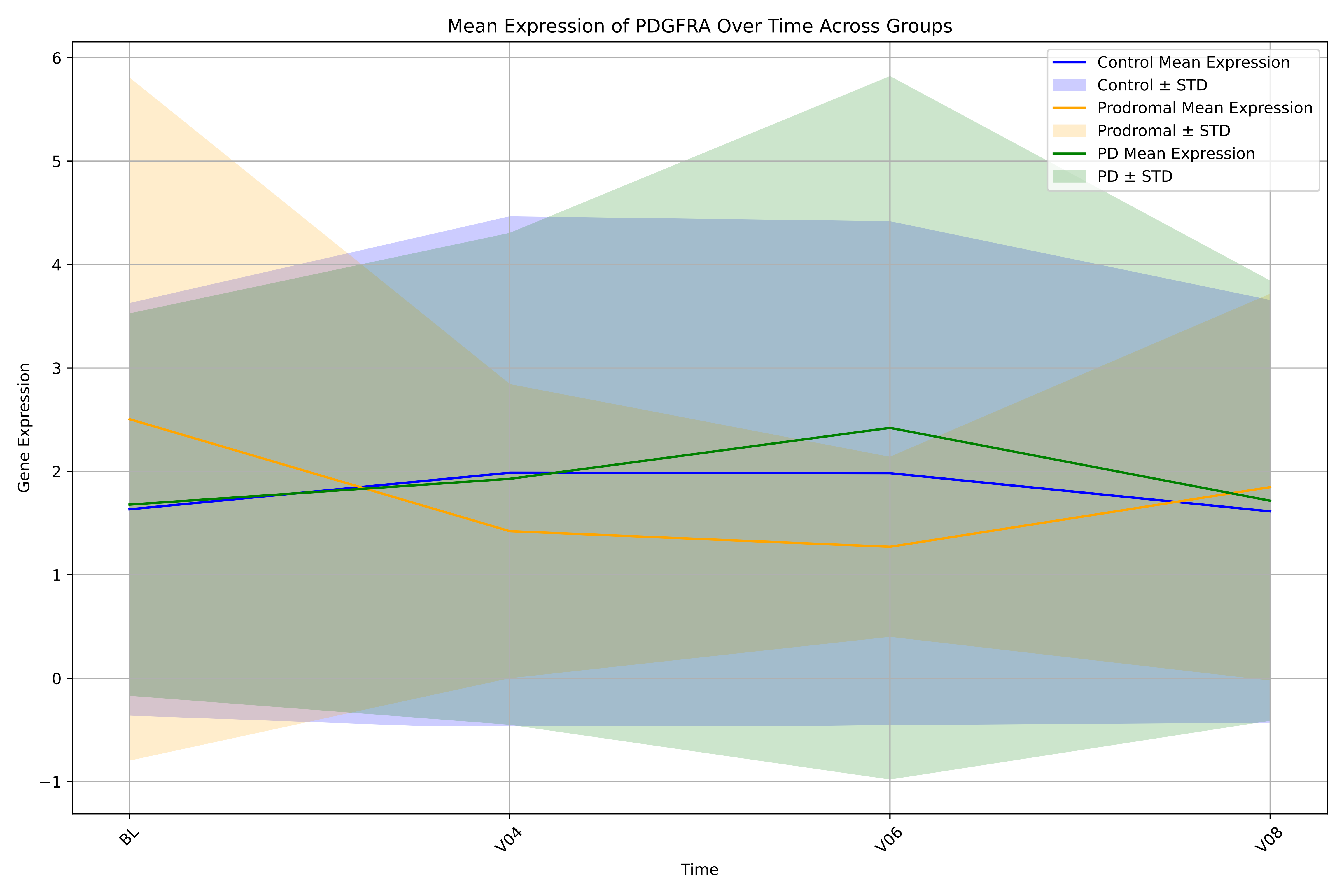
Mean Expression of PDGFRA Over Time Across Groups
Control Mean Expression
Control ± STD
Prodromal Mean Expression
Prodromal ± STD
PD Mean Expression
PD ± STD
Gene Expression
Time
BL
V04
V06
V08
6
5
4
3
2
1
0
−1

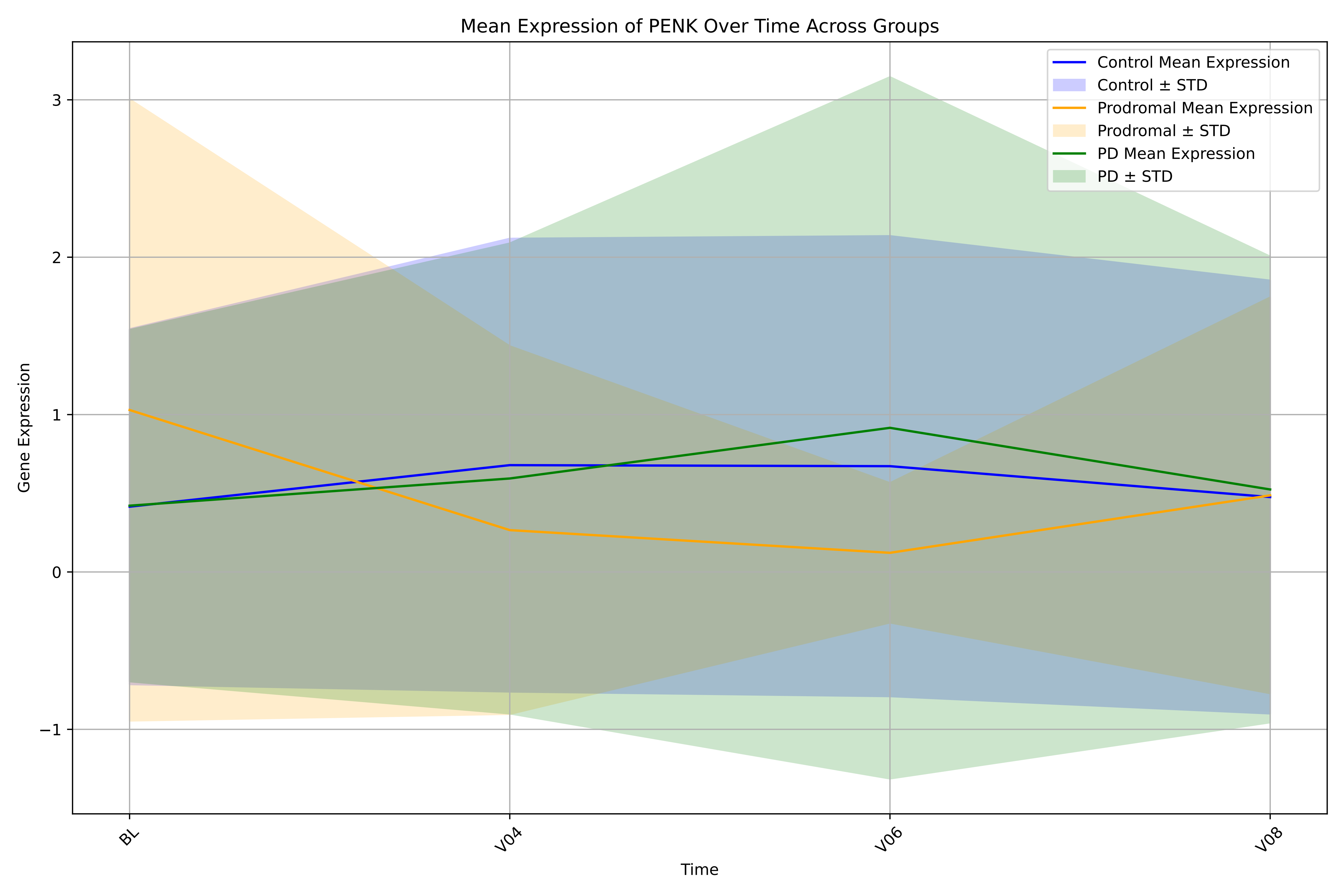

Mean Expression of PENK Over Time Across Groups
Control Mean Expression
Control ± STD
Prodromal Mean Expression
Prodromal ± STD
PD Mean Expression
PD ± STD
Gene Expression
3
2
1
0
−1
BL
V04
V06
V08
Time

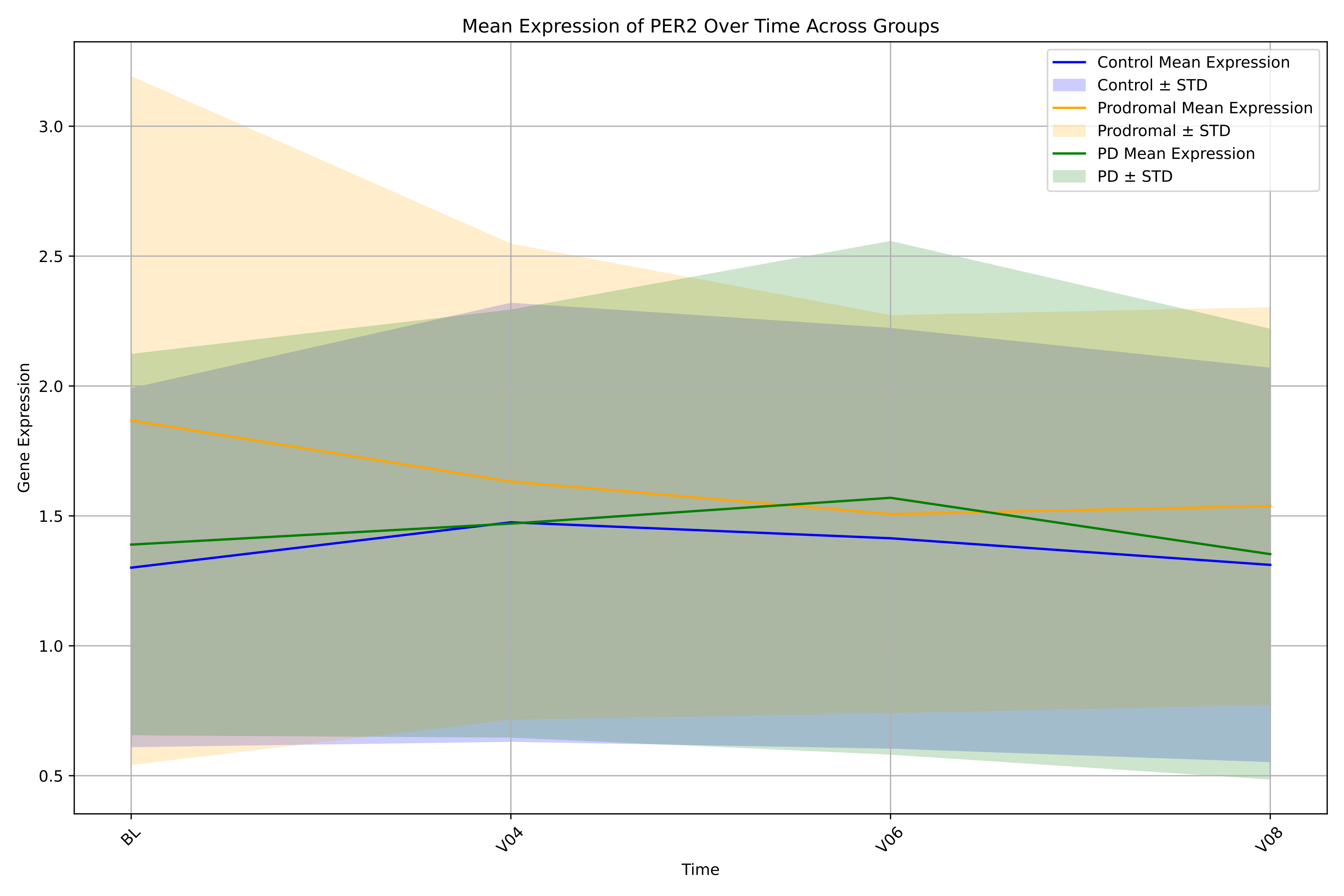
Mean Expression of PER2 Over Time Across Groups
Control Mean Expression
Control ± STD
Prodromal Mean Expression
Prodromal ± STD
PD Mean Expression
PD ± STD
Gene Expression
3.0
2.5
2.0
1.5
1.0
0.5
BL
V04
V06
V08
Time

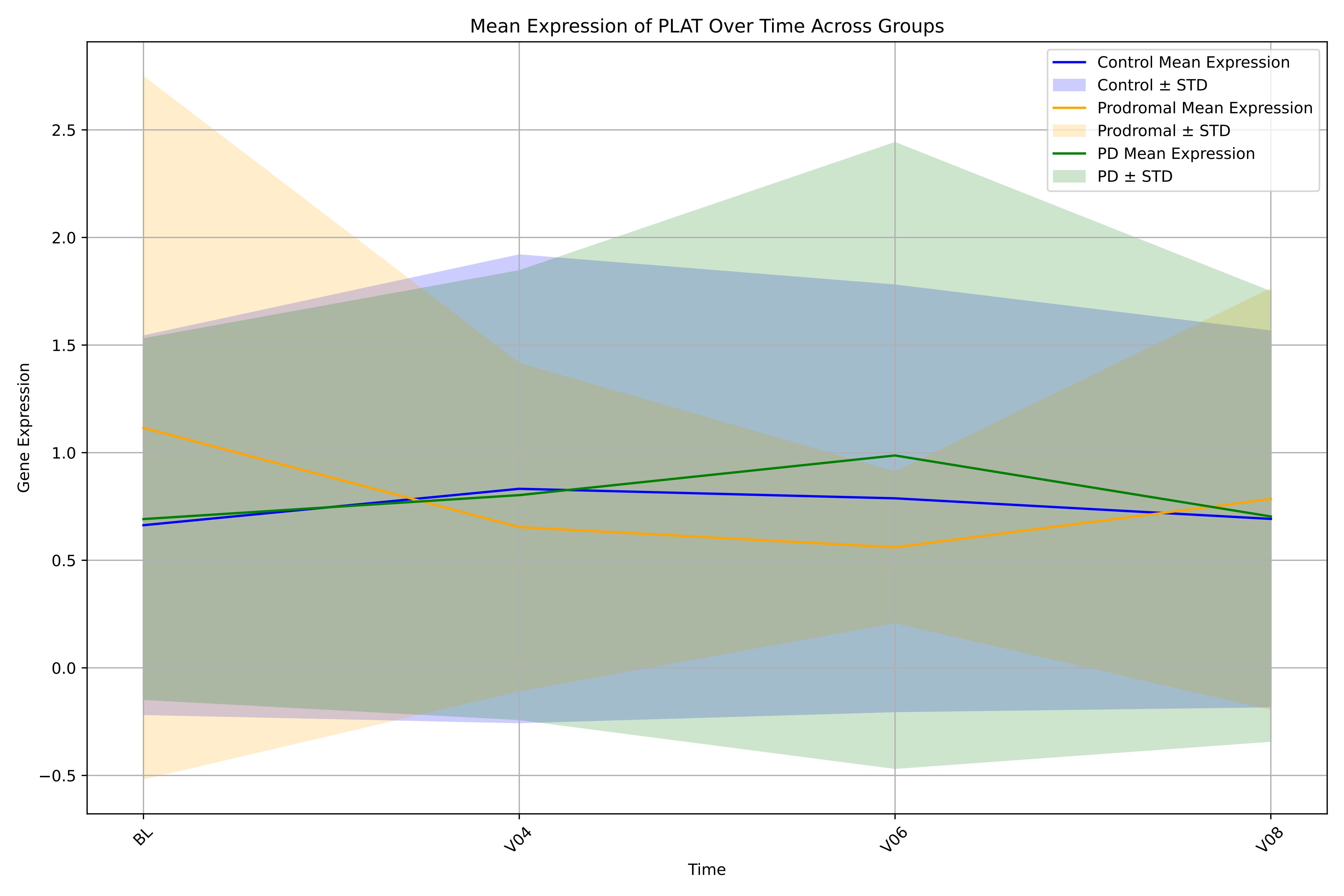
Mean Expression of PLAT Over Time Across Groups
Control Mean Expression
Control ± STD
Prodromal Mean Expression
Prodromal ± STD
PD Mean Expression
PD ± STD
Gene Expression
−0.5
0.0
0.5
1.0
1.5
2.0
2.5
BL
V04
V06
V08
Time

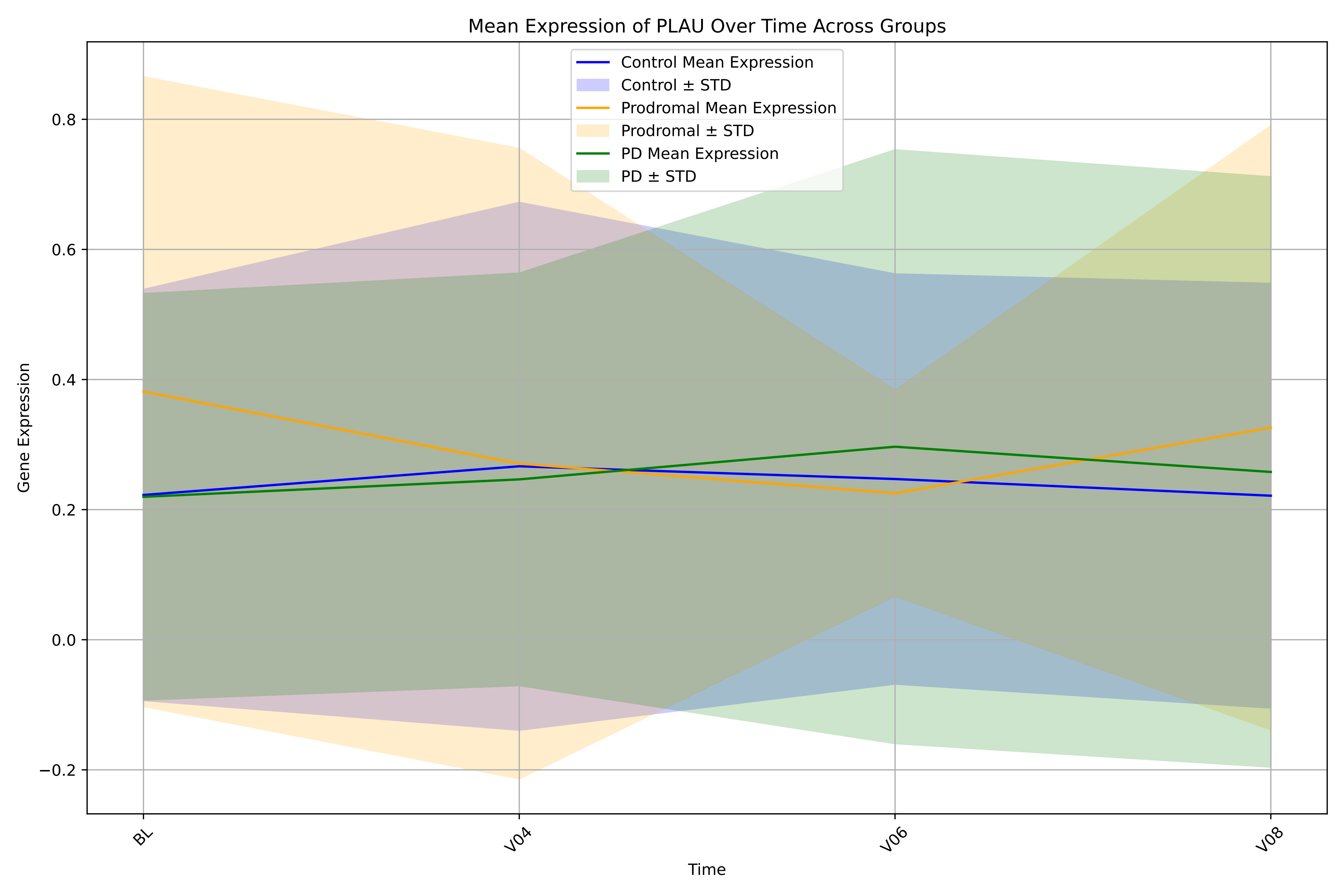

Mean Expression of PLAU Over Time Across Groups
Control Mean Expression
Control ± STD
Prodromal Mean Expression
Prodromal ± STD
PD Mean Expression
PD ± STD
Gene Expression
0.8
0.6
0.4
0.2
0.0
−0.2
BL
V04
V06
V08
Time

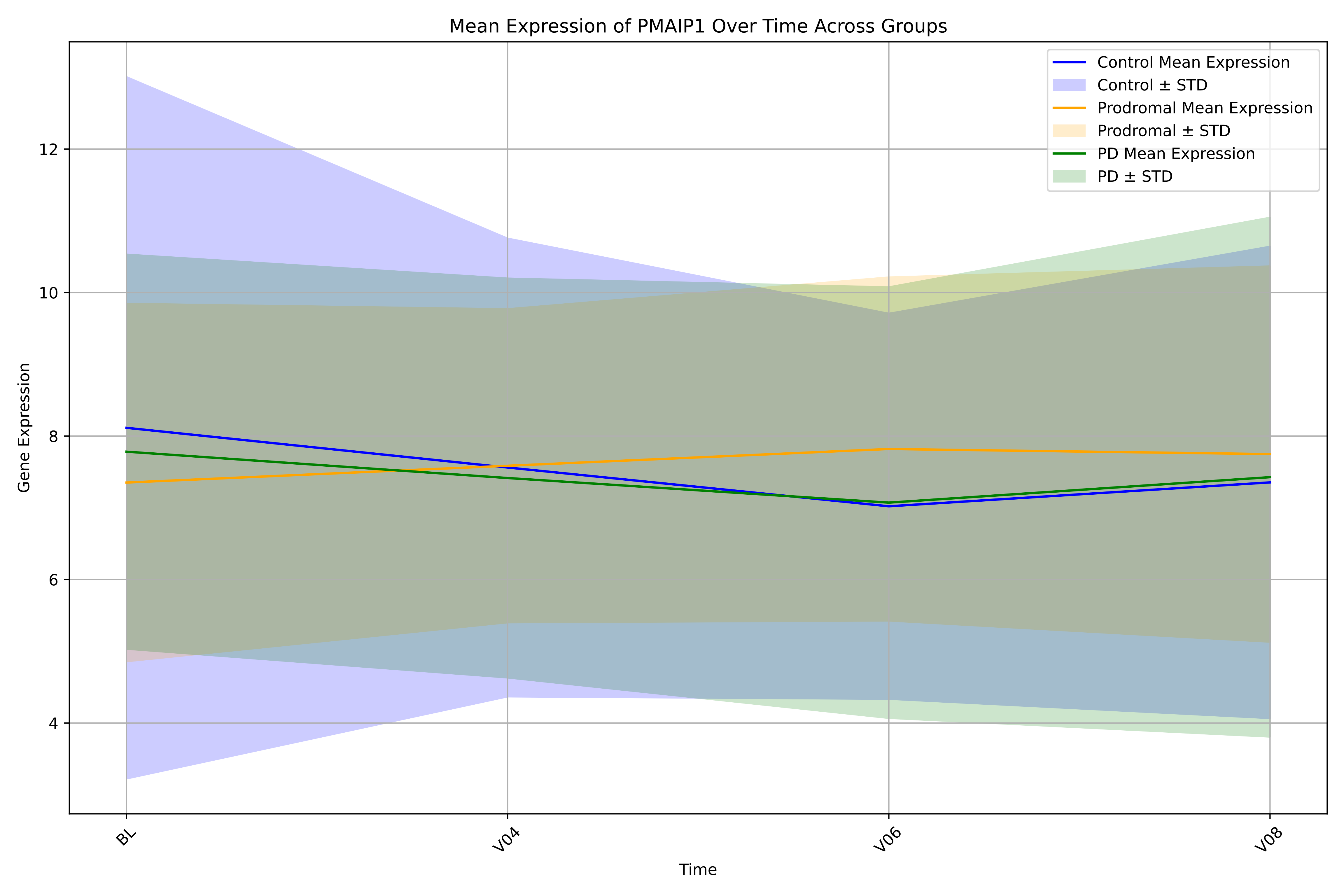

Mean Expression of PMAIP1 Over Time Across Groups
Control Mean Expression
Control ± STD
Prodromal Mean Expression
Prodromal ± STD
PD Mean Expression
PD ± STD
Gene Expression
12
10
8
6
4
BL
V04
V06
V08
Time

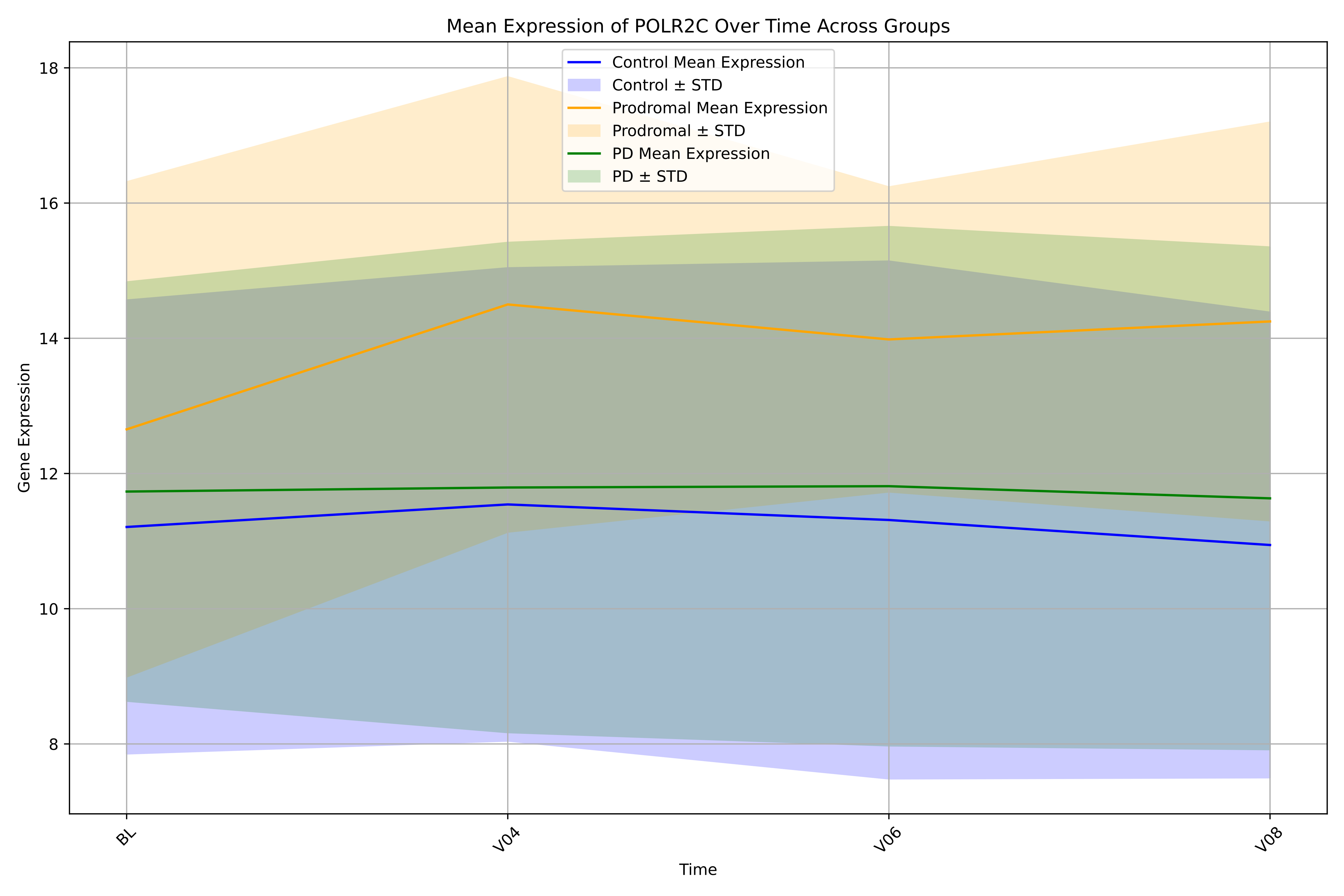

Mean Expression of POLR2C Over Time Across Groups
Control Mean Expression
Control ± STD
Prodromal Mean Expression
Prodromal ± STD
PD Mean Expression
PD ± STD
Gene Expression
18
16
14
12
10
8
BL
V04
V06
V08
Time

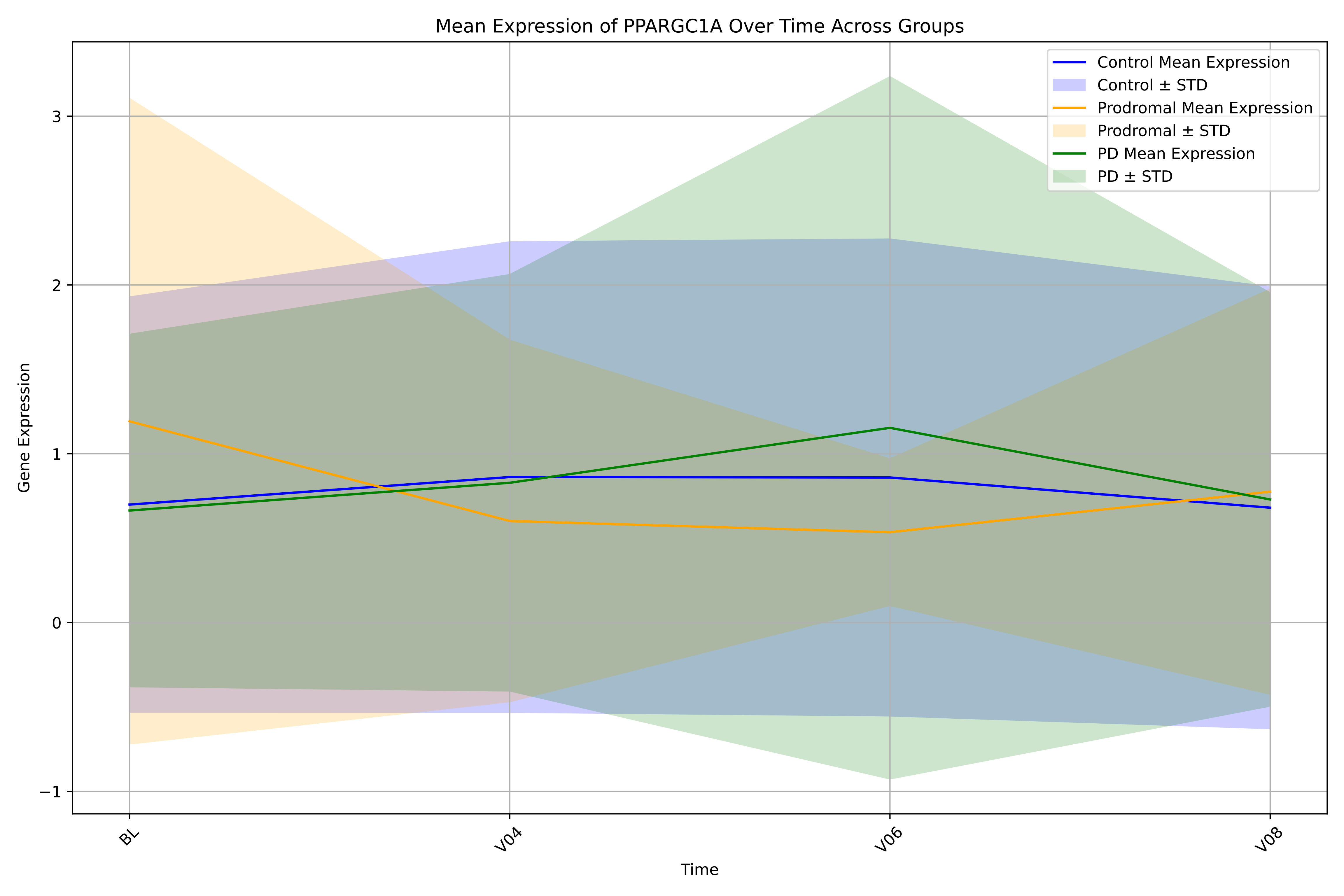
Mean Expression of PPARGC1A Over Time Across Groups
Control Mean Expression
Control ± STD
Prodromal Mean Expression
Prodromal ± STD
PD Mean Expression
PD ± STD
Gene Expression
3
2
1
0
−1
BL
V04
V06
V08
Time

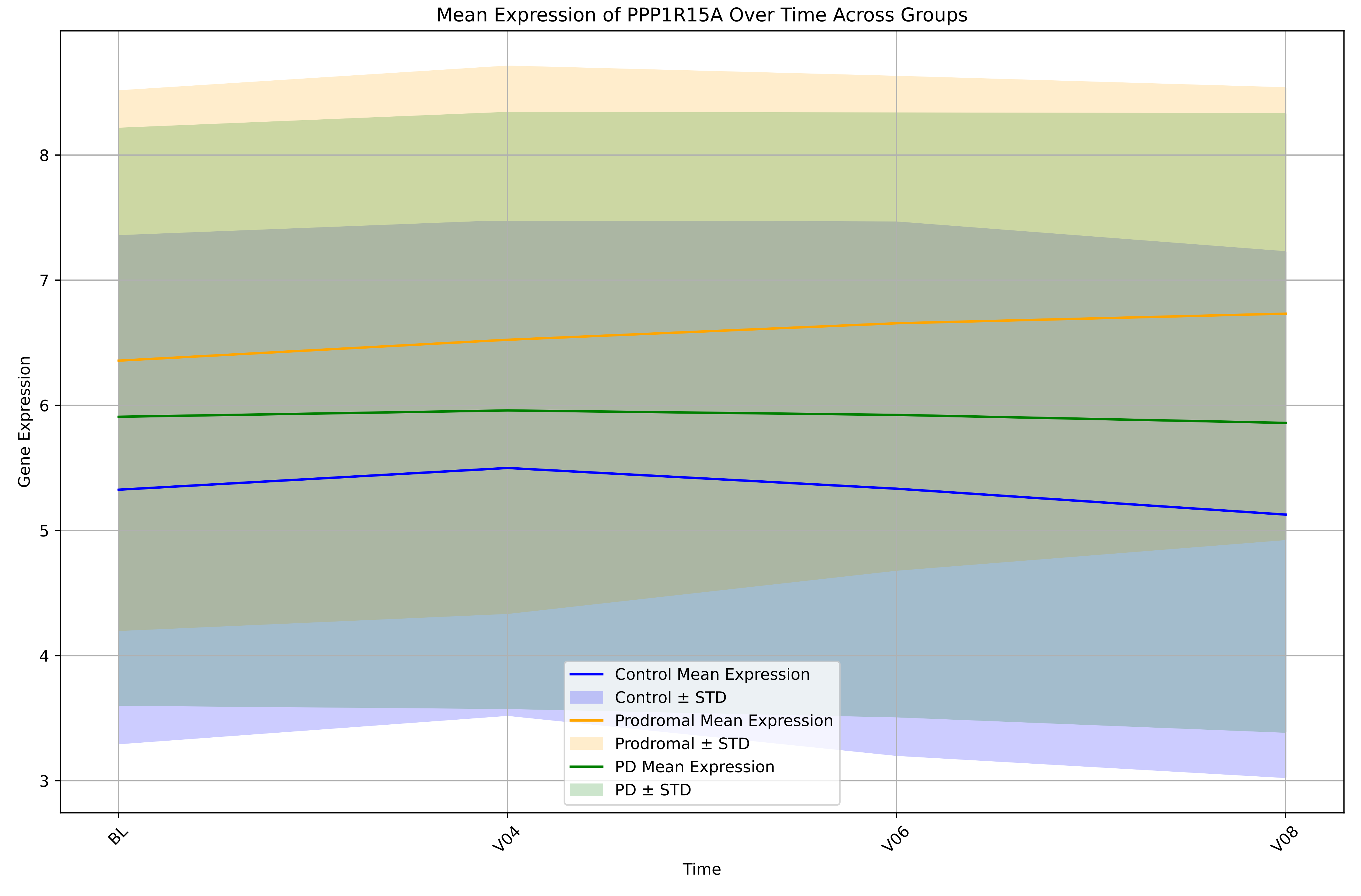

Mean Expression of PPP1R15A Over Time Across Groups
Gene Expression
Time
8
7
6
5
4
3
BL
V04
V06
V08
Control Mean Expression
Control ± STD
Prodromal Mean Expression
Prodromal ± STD
PD Mean Expression
PD ± STD

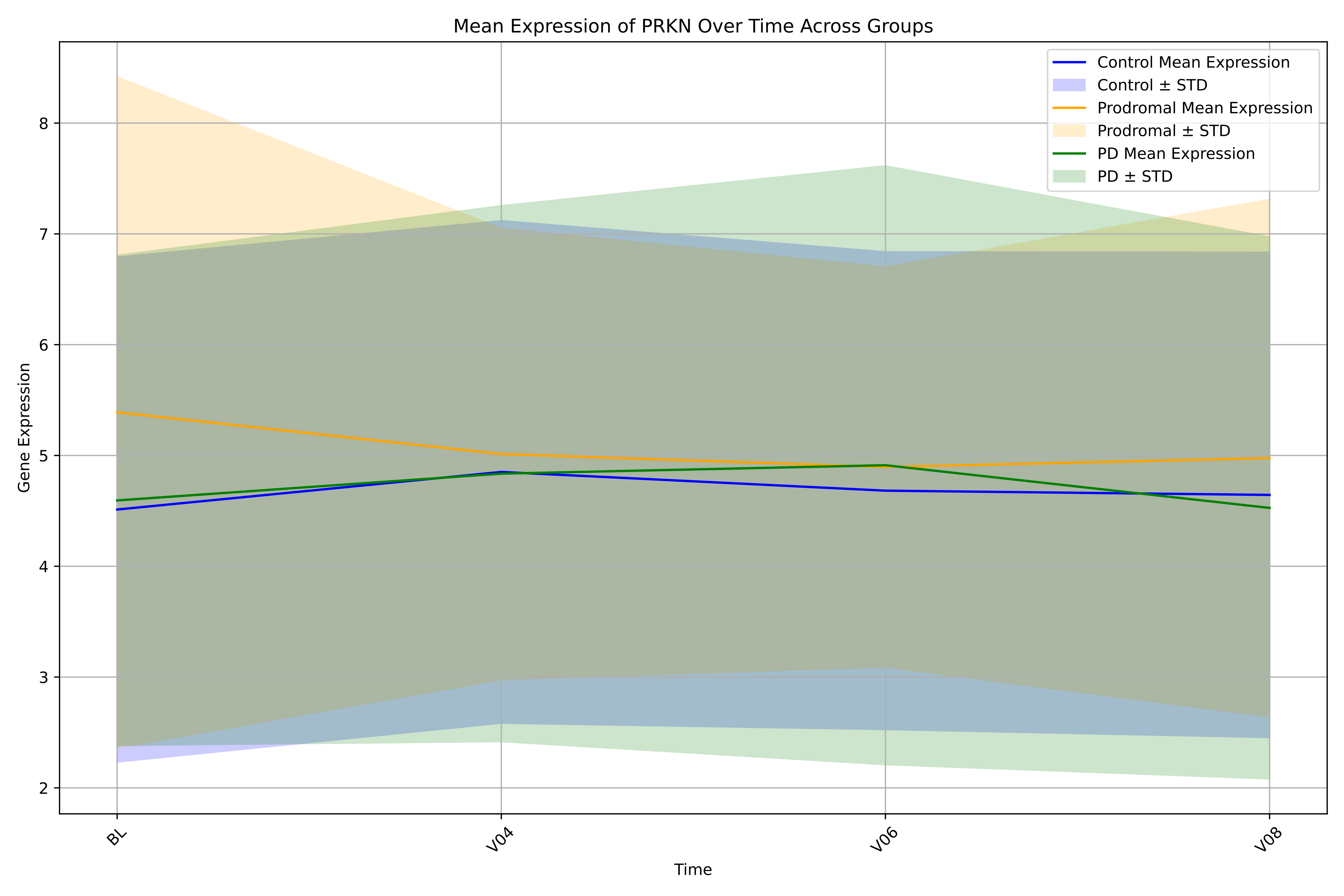

Mean Expression of PRKN Over Time Across Groups
Control Mean Expression
Control ± STD
Prodromal Mean Expression
Prodromal ± STD
PD Mean Expression
PD ± STD
Gene Expression
2
3
4
5
6
7
8
BL
V04
V06
V08
Time

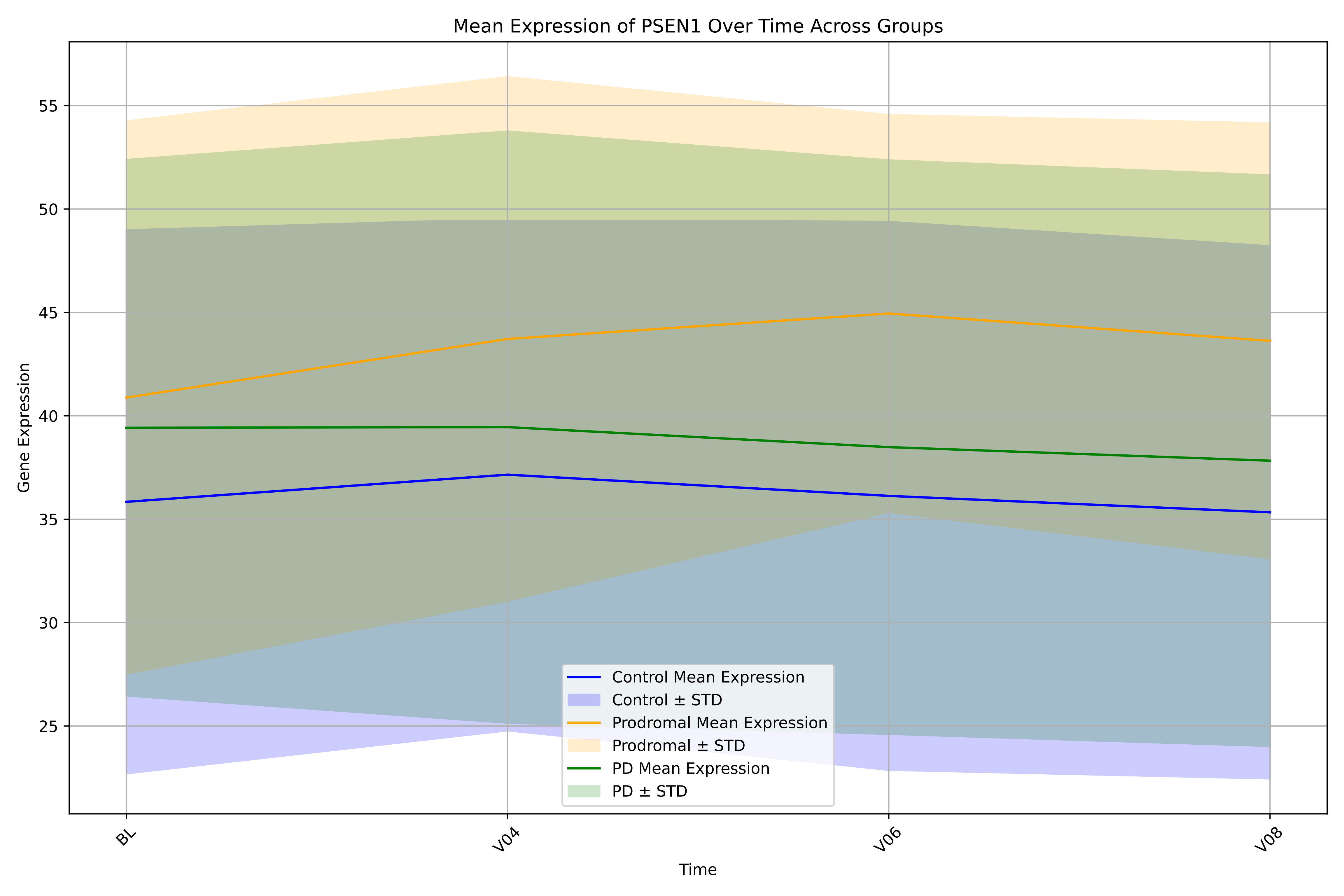
Mean Expression of PSEN1 Over Time Across Groups
Gene Expression
Time
BL
V04
V06
V08
55
50
45
40
35
30
25
Control Mean Expression
Control ± STD
Prodromal Mean Expression
Prodromal ± STD
PD Mean Expression
PD ± STD

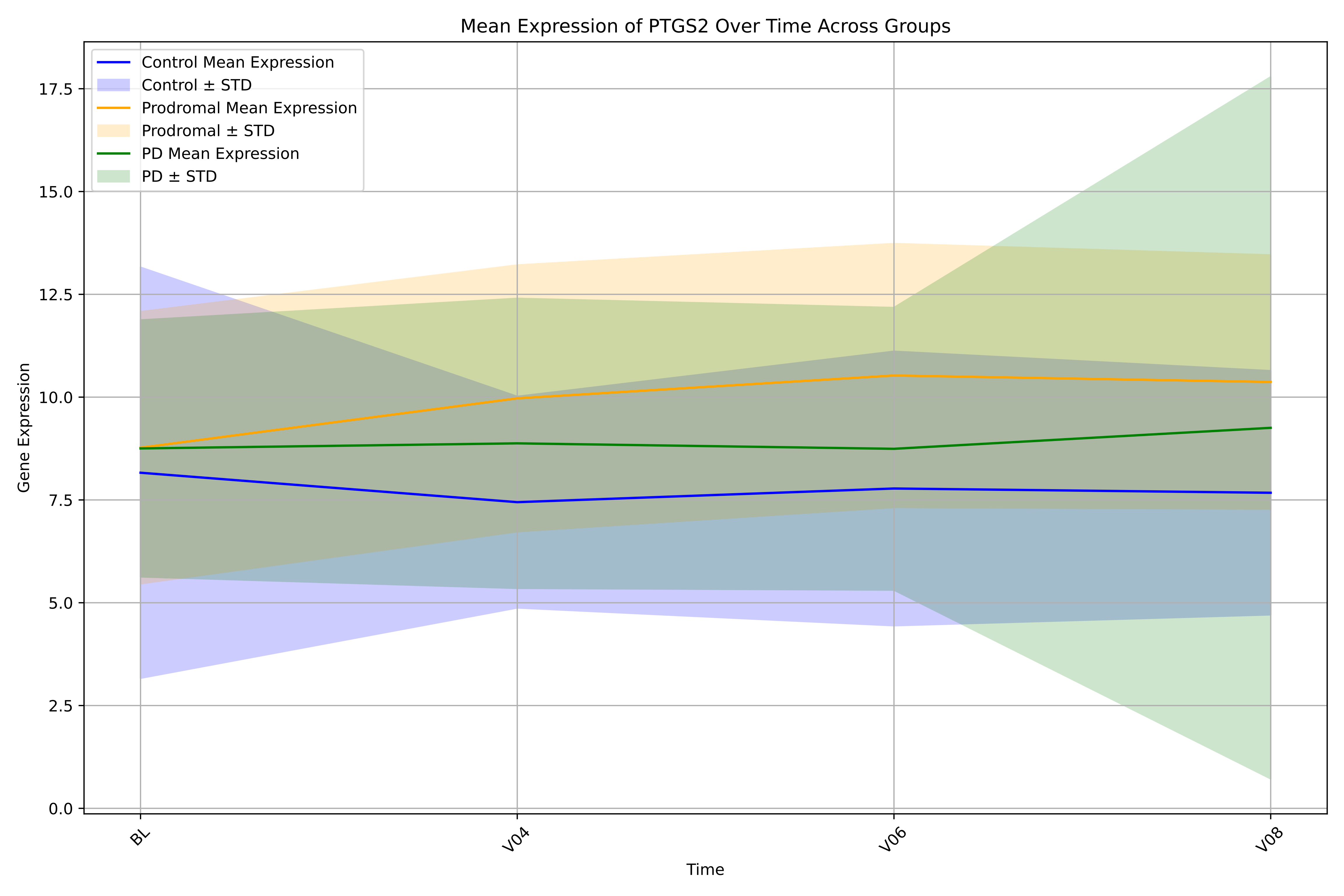
Mean Expression of PTGS2 Over Time Across Groups
Control Mean Expression
Control ± STD
Prodromal Mean Expression
Prodromal ± STD
PD Mean Expression
PD ± STD
Gene Expression
0.0
2.5
5.0
7.5
10.0
12.5
15.0
17.5
BL
V04
V06
V08
Time

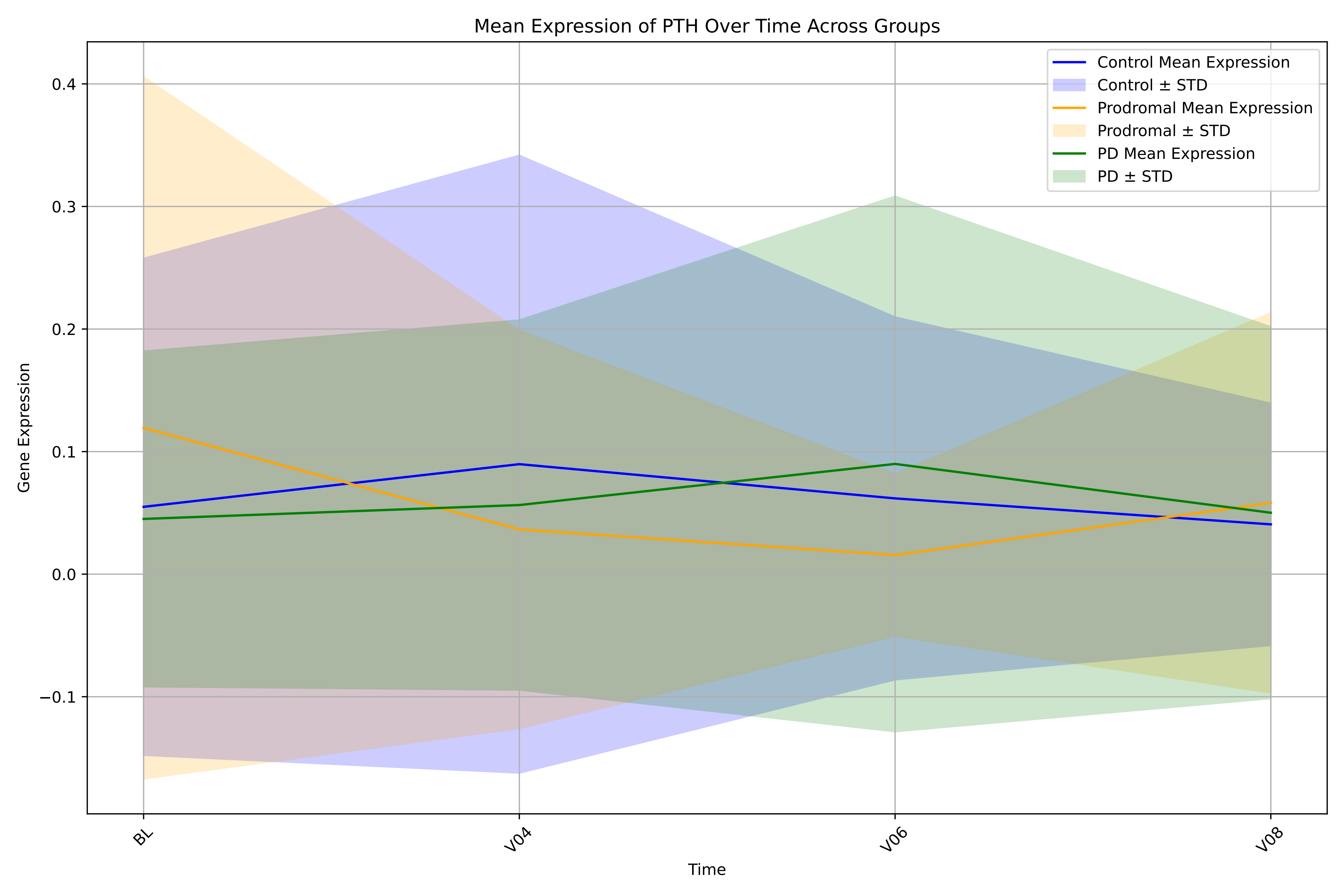

Mean Expression of PTH Over Time Across Groups
Control Mean Expression
Control ± STD
Prodromal Mean Expression
Prodromal ± STD
PD Mean Expression
PD ± STD
Gene Expression
0.4
0.3
0.2
0.1
0.0
−0.1
BL
V04
V06
V08
Time

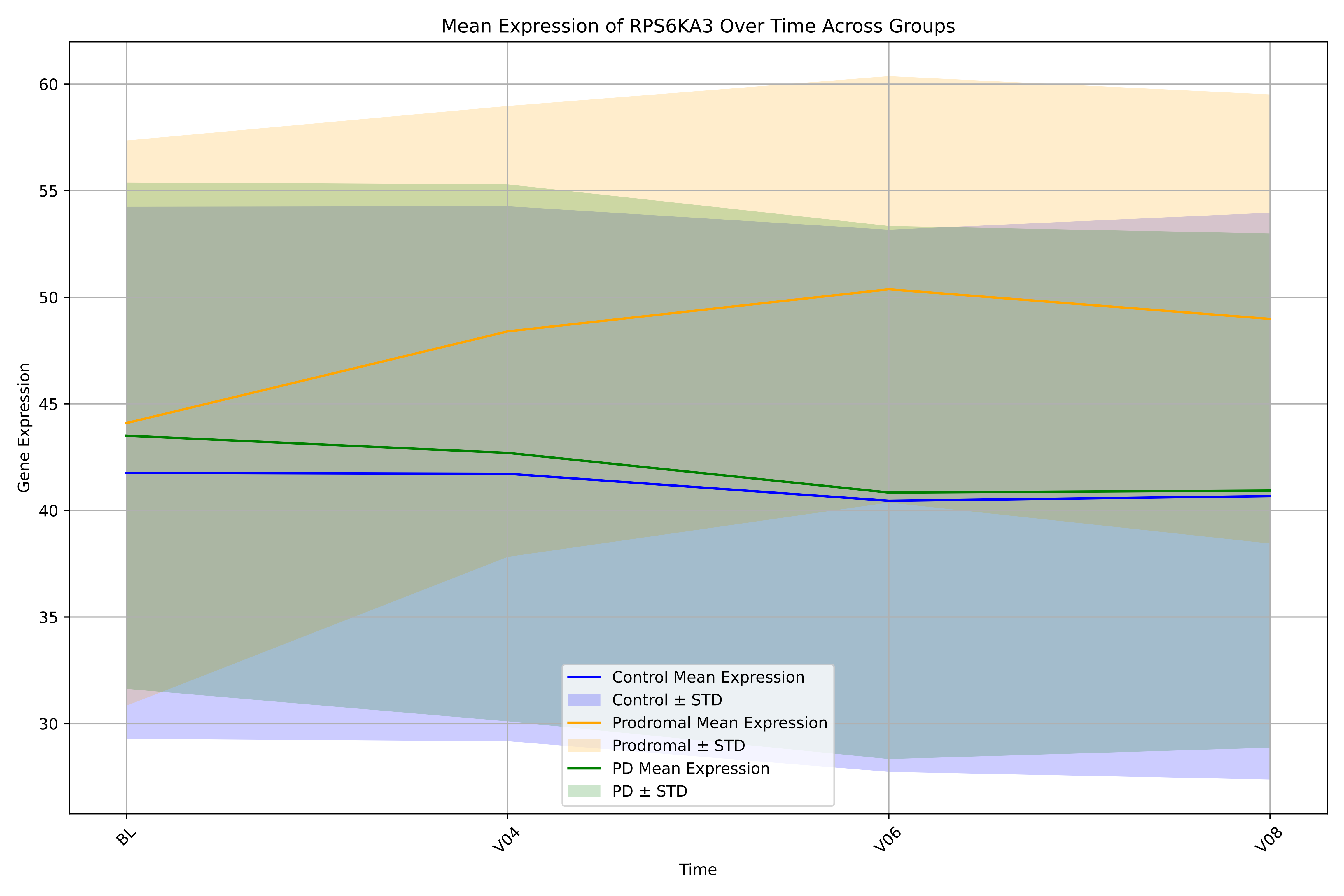
Mean Expression of RPS6KA3 Over Time Across Groups
Gene Expression
Time
60
55
50
45
40
35
30
BL
V04
V06
V08
Control Mean Expression
Control ± STD
Prodromal Mean Expression
Prodromal ± STD
PD Mean Expression
PD ± STD

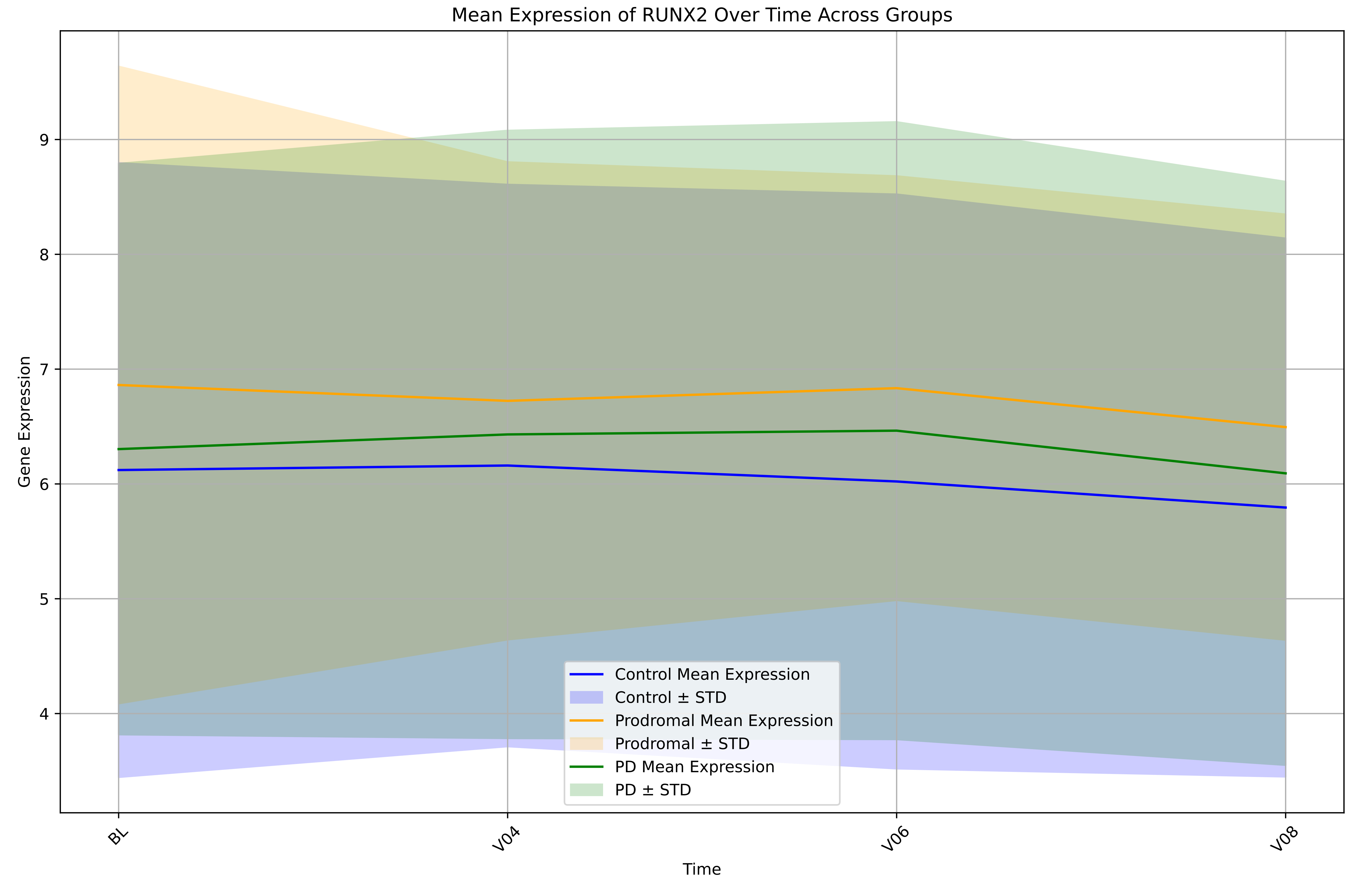

Mean Expression of RUNX2 Over Time Across Groups
Gene Expression
Time
BL
V04
V06
V08
4
5
6
7
8
9
Control Mean Expression
Control ± STD
Prodromal Mean Expression
Prodromal ± STD
PD Mean Expression
PD ± STD

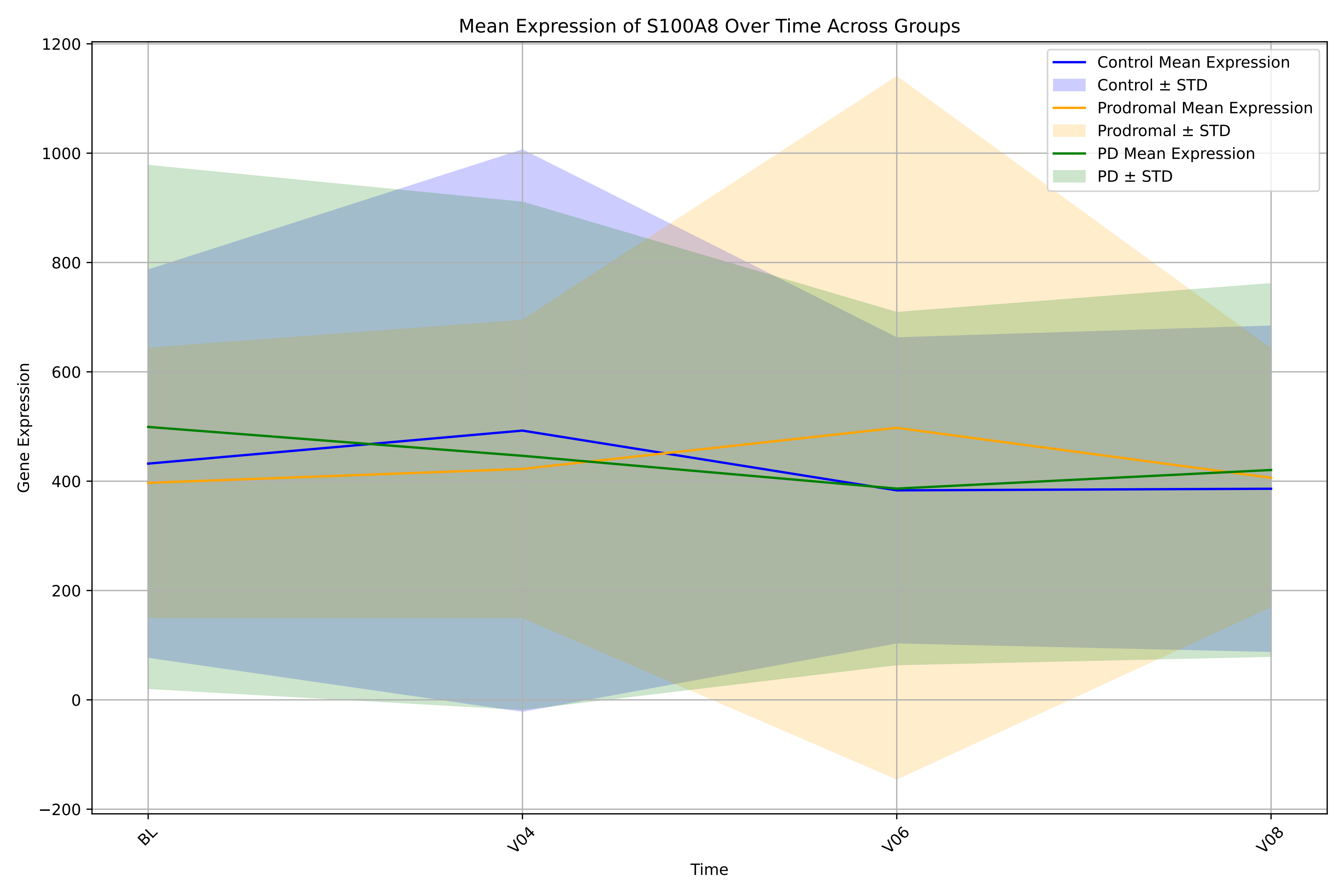
Mean Expression of S100A8 Over Time Across Groups
Control Mean Expression
Control ± STD
Prodromal Mean Expression
Prodromal ± STD
PD Mean Expression
PD ± STD
Gene Expression
1200
1000
800
600
400
200
0
−200
BL
V04
V06
V08
Time

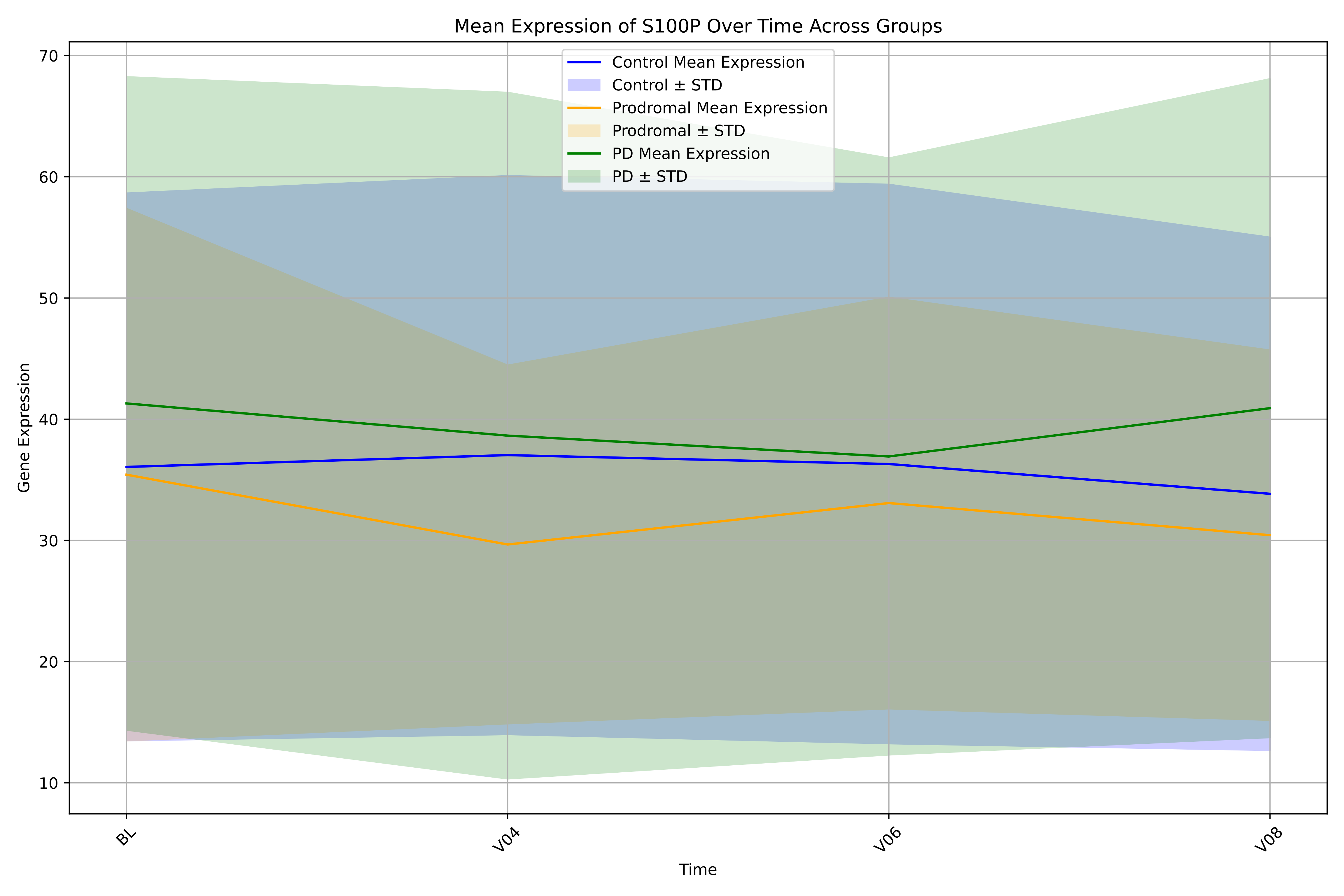
Mean Expression of S100P Over Time Across Groups
Control Mean Expression
Control ± STD
Prodromal Mean Expression
Prodromal ± STD
PD Mean Expression
PD ± STD
Gene Expression
70
60
50
40
30
20
10
BL
V04
V06
V08
Time

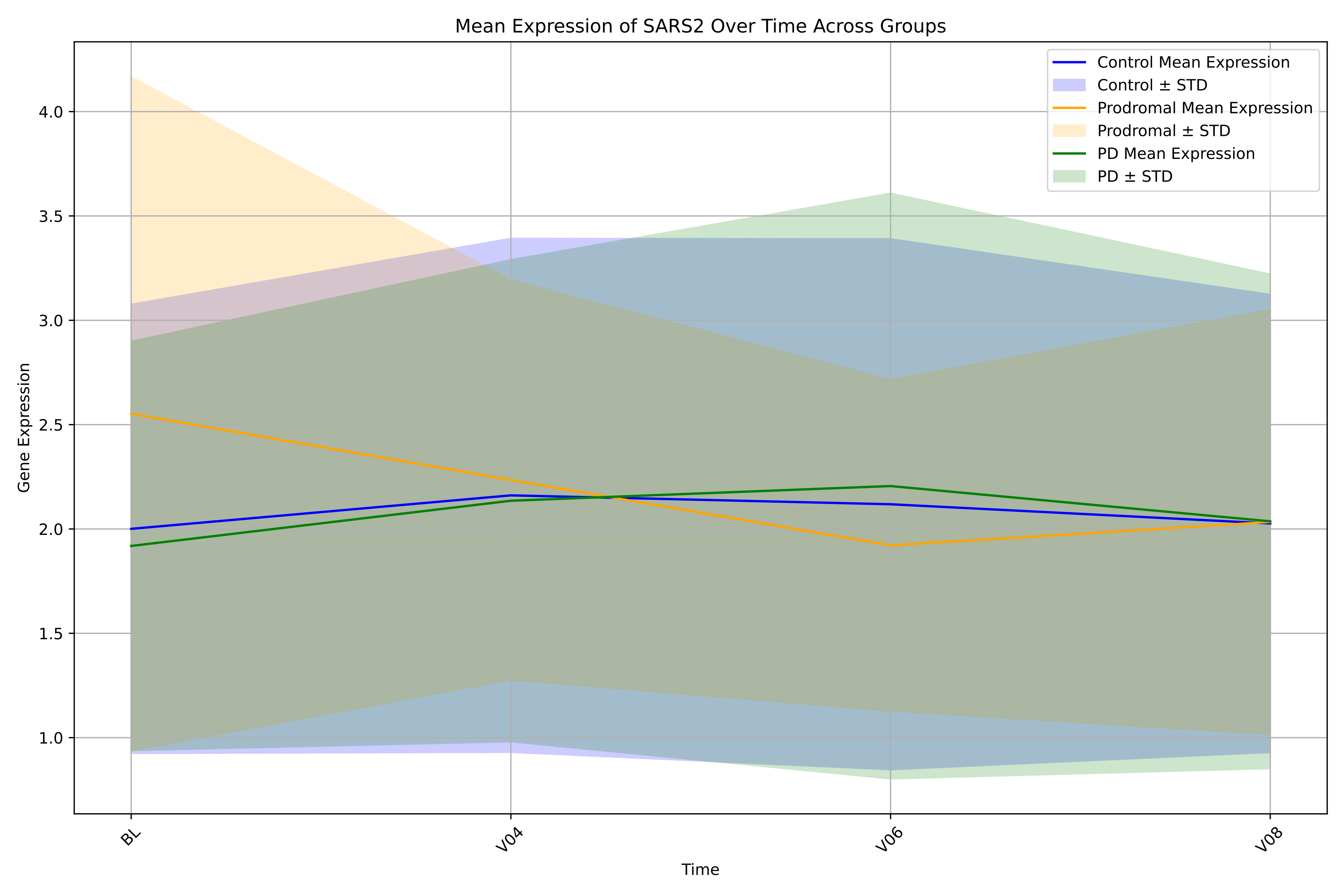

Mean Expression of SARS2 Over Time Across Groups
Control Mean Expression
Control ± STD
Prodromal Mean Expression
Prodromal ± STD
PD Mean Expression
PD ± STD
Gene Expression
4.0
3.5
3.0
2.5
2.0
1.5
1.0
BL
V04
V06
V08
Time

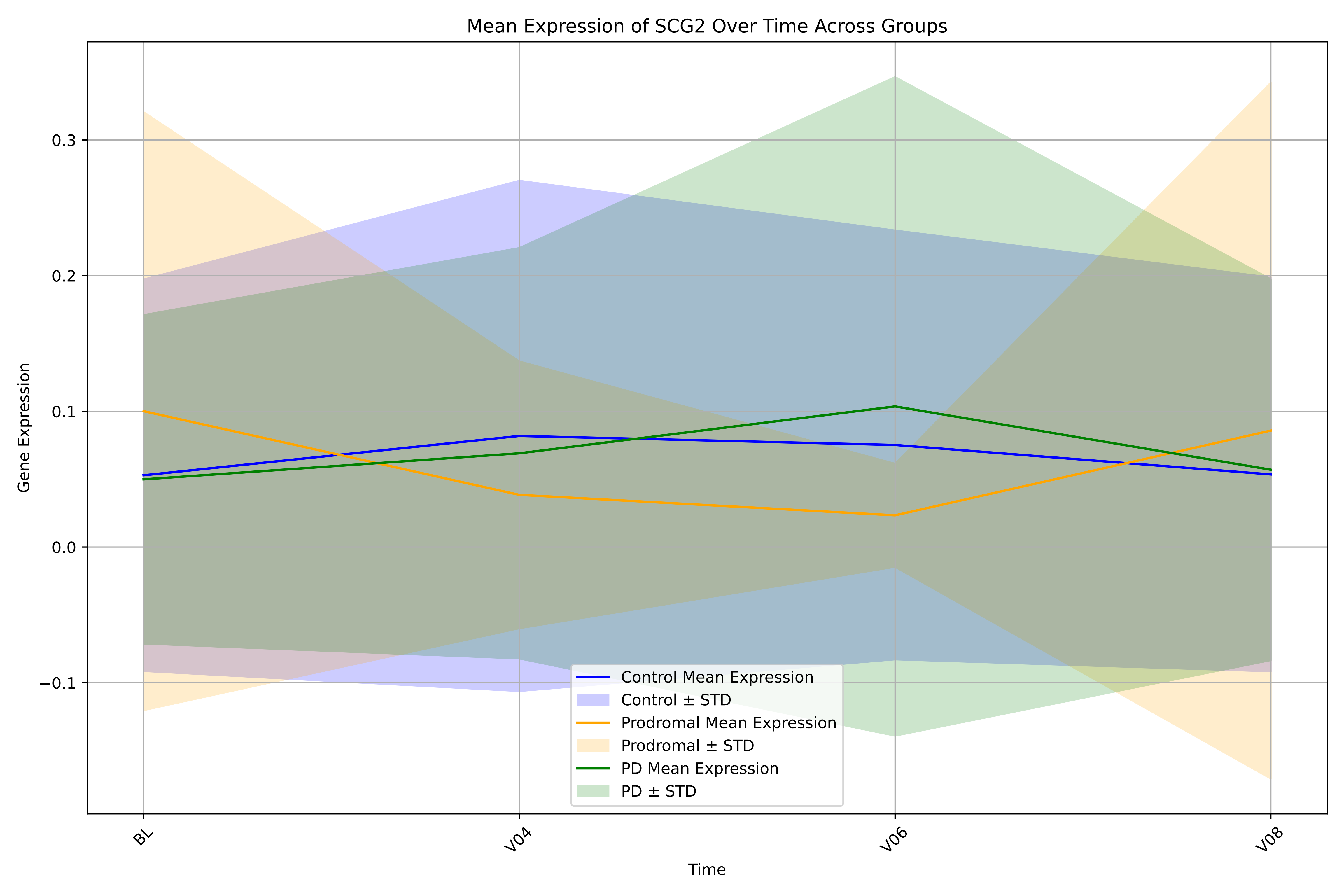

Mean Expression of SCG2 Over Time Across Groups
Gene Expression
Time
BL
V04
V06
V08
0.3
0.2
0.1
0.0
−0.1
Control Mean Expression
Control ± STD
Prodromal Mean Expression
Prodromal ± STD
PD Mean Expression
PD ± STD

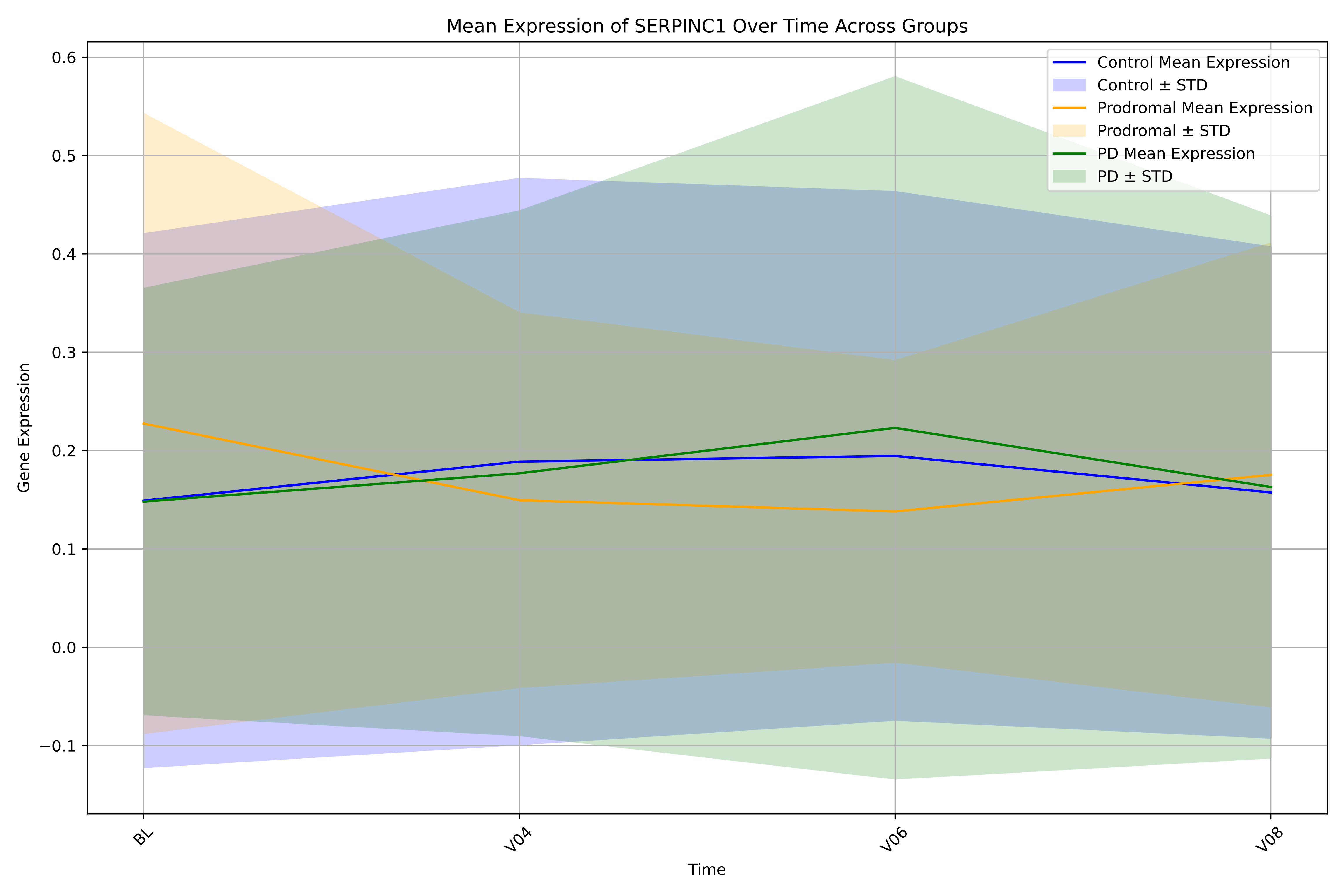

Mean Expression of SERPINC1 Over Time Across Groups
Control Mean Expression
Control ± STD
Prodromal Mean Expression
Prodromal ± STD
PD Mean Expression
PD ± STD
Gene Expression
Time
BL
V04
V06
V08
0.6
0.5
0.4
0.3
0.2
0.1
0.0
−0.1

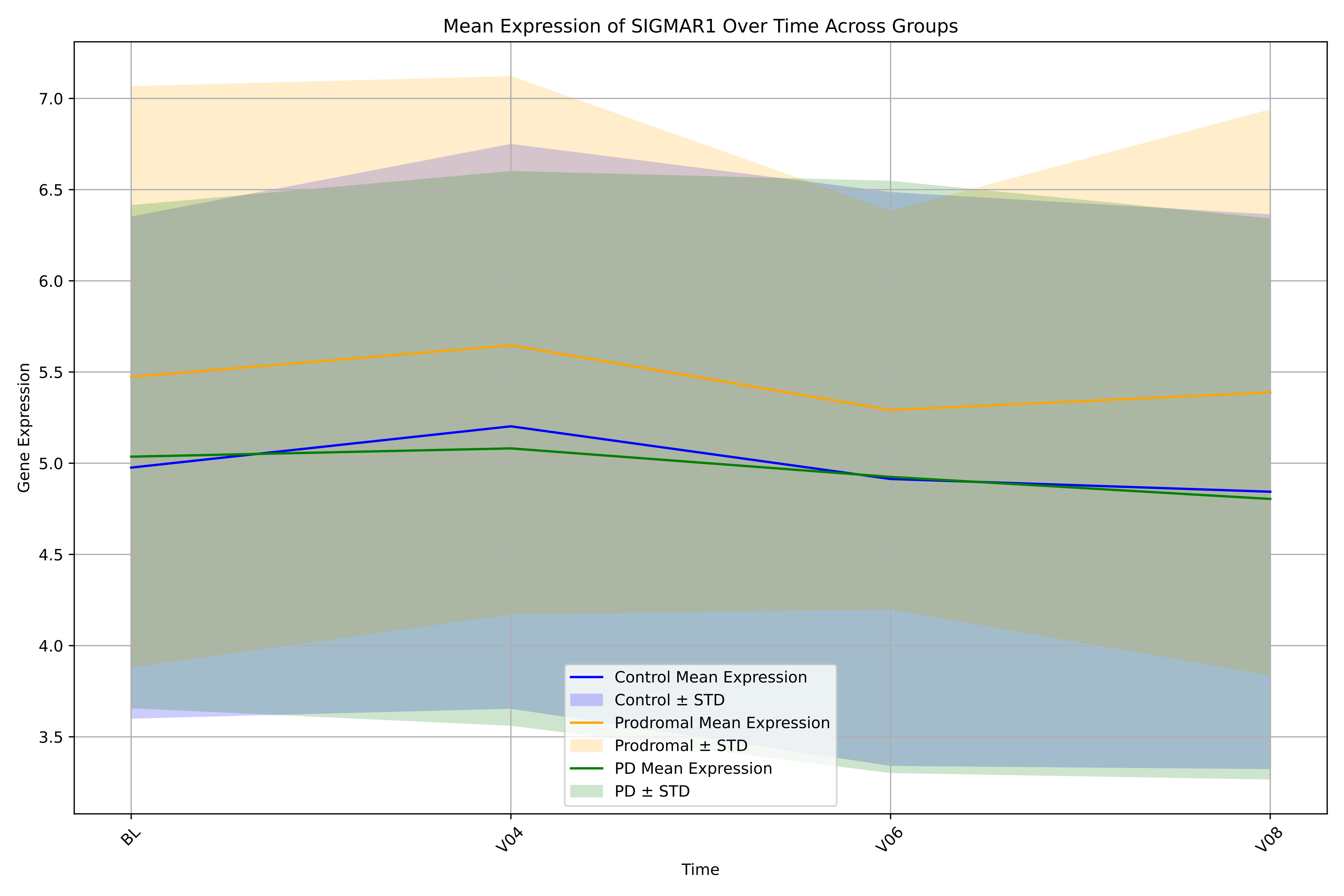
Mean Expression of SIGMAR1 Over Time Across Groups
Gene Expression
Time
7.0
6.5
6.0
5.5
5.0
4.5
4.0
3.5
BL
V04
V06
V08
Control Mean Expression
Control ± STD
Prodromal Mean Expression
Prodromal ± STD
PD Mean Expression
PD ± STD

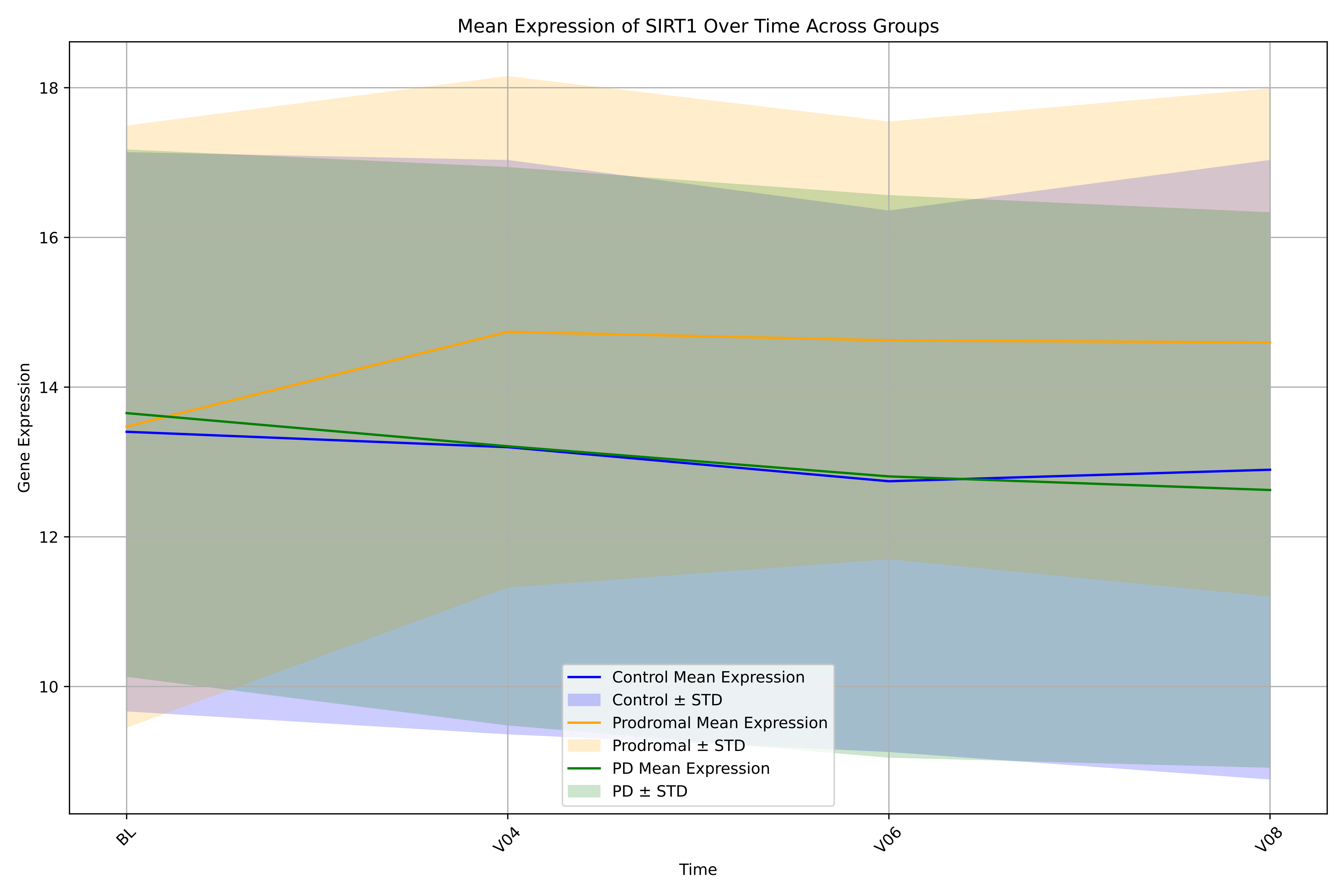

Mean Expression of SIRT1 Over Time Across Groups
Gene Expression
18
16
14
12
10
BL
V04
V06
V08
Time
Control Mean Expression
Control ± STD
Prodromal Mean Expression
Prodromal ± STD
PD Mean Expression
PD ± STD

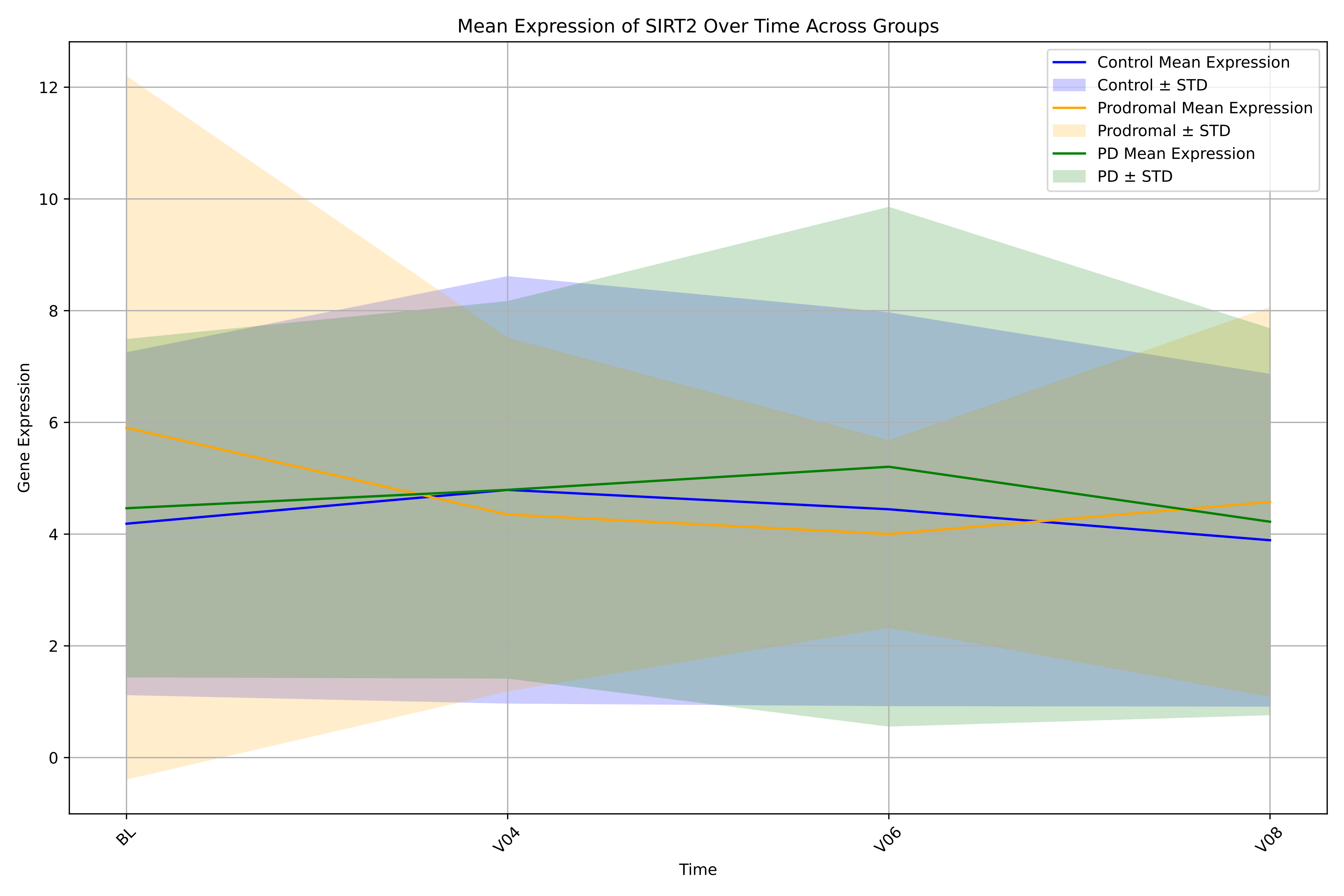

Mean Expression of SIRT2 Over Time Across Groups
Control Mean Expression
Control ± STD
Prodromal Mean Expression
Prodromal ± STD
PD Mean Expression
PD ± STD
Gene Expression
Time
BL
V04
V06
V08
0
2
4
6
8
10
12

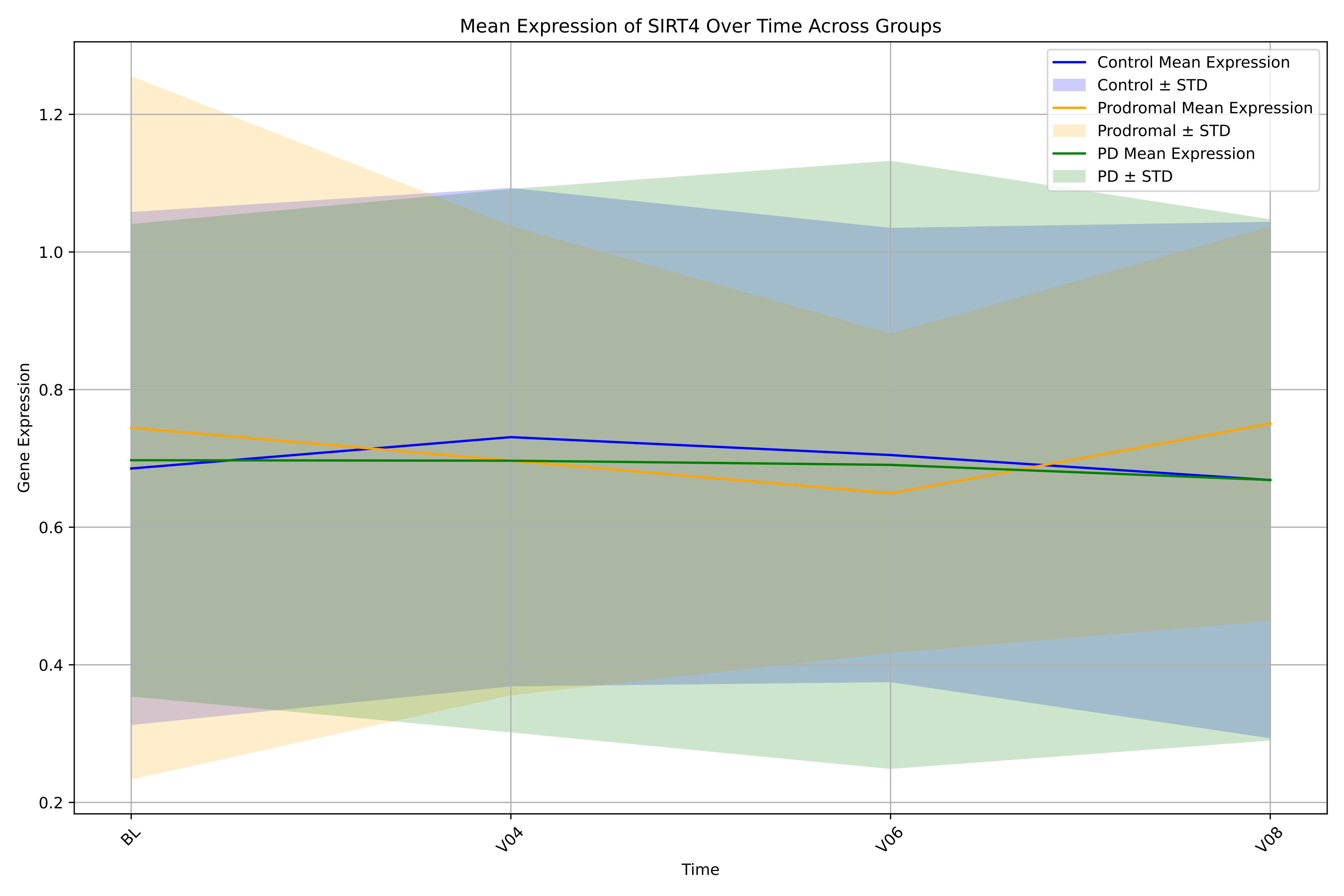
Mean Expression of SIRT4 Over Time Across Groups
Control Mean Expression
Control ± STD
Prodromal Mean Expression
Prodromal ± STD
PD Mean Expression
PD ± STD
Gene Expression
1.2
1.0
0.8
0.6
0.4
0.2
BL
V04
V06
V08
Time

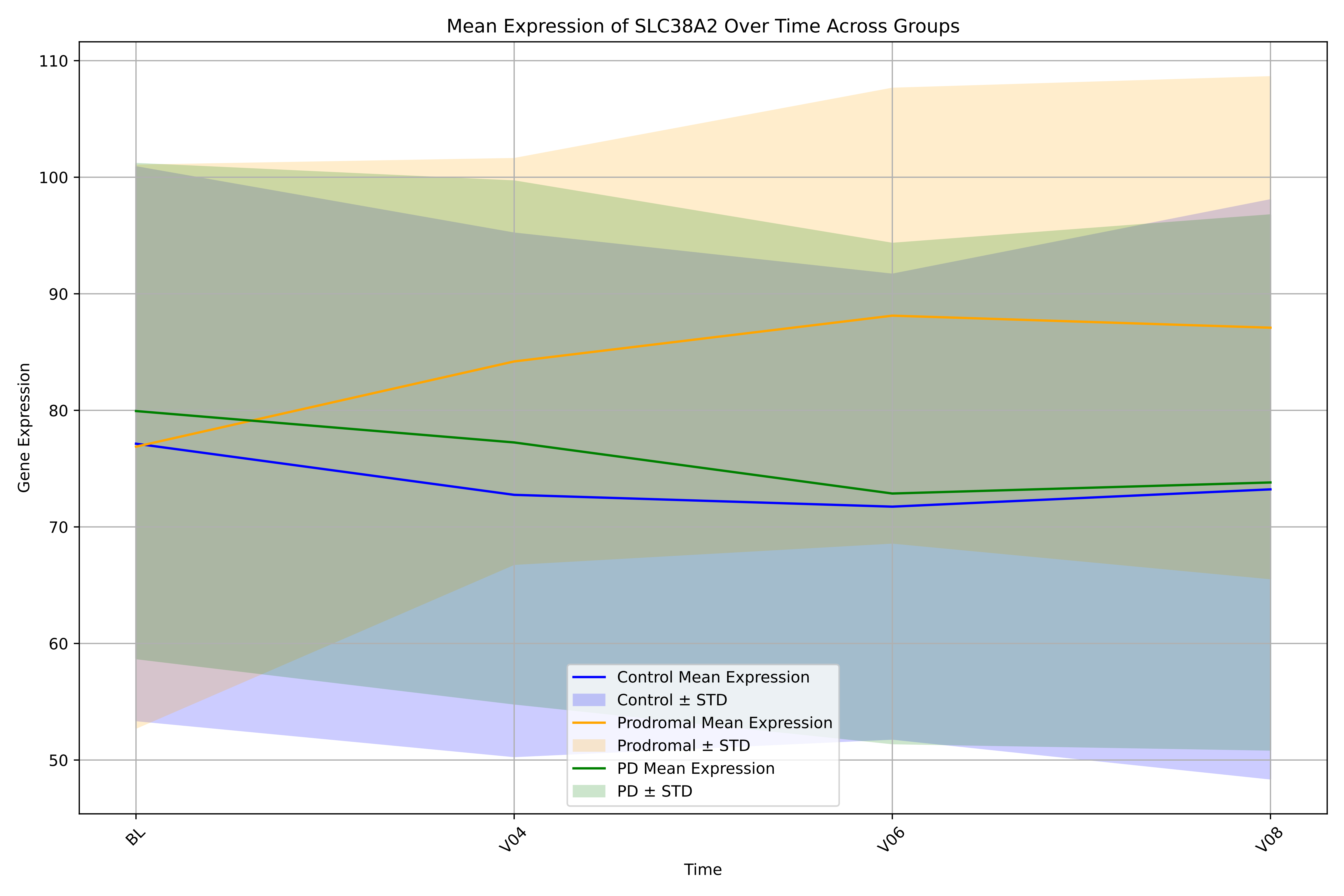

Mean Expression of SLC38A2 Over Time Across Groups
Gene Expression
Time
110
100
90
80
70
60
50
BL
V04
V06
V08
Control Mean Expression
Control ± STD
Prodromal Mean Expression
Prodromal ± STD
PD Mean Expression
PD ± STD

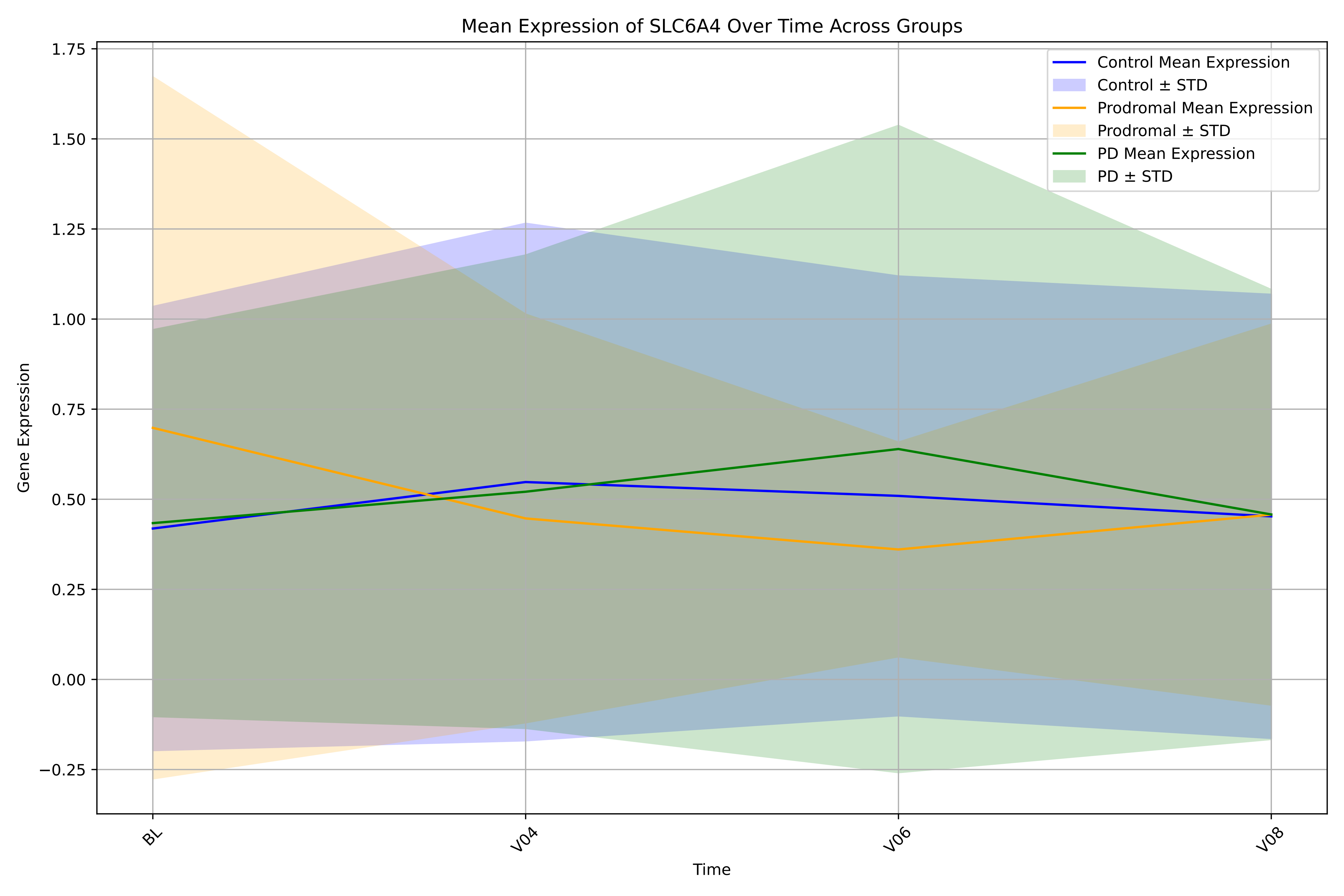

Mean Expression of SLC6A4 Over Time Across Groups
Control Mean Expression
Control ± STD
Prodromal Mean Expression
Prodromal ± STD
PD Mean Expression
PD ± STD
Gene Expression
1.75
1.50
1.25
1.00
0.75
0.50
0.25
0.00
−0.25
BL
V04
V06
V08
Time

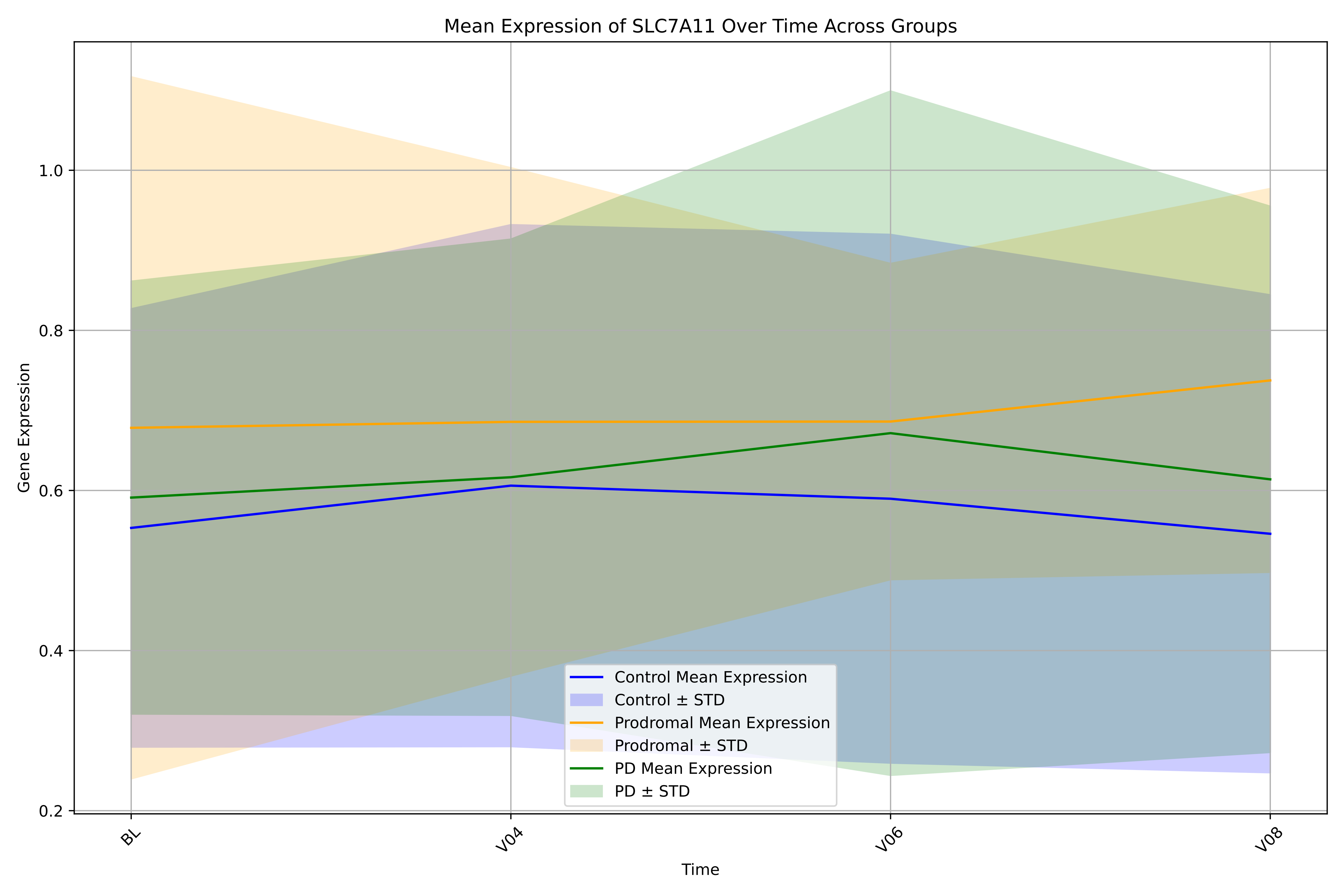

Mean Expression of SLC7A11 Over Time Across Groups
Gene Expression
0.2
0.4
0.6
0.8
1.0
Time
BL
V04
V06
V08
Control Mean Expression
Control ± STD
Prodromal Mean Expression
Prodromal ± STD
PD Mean Expression
PD ± STD

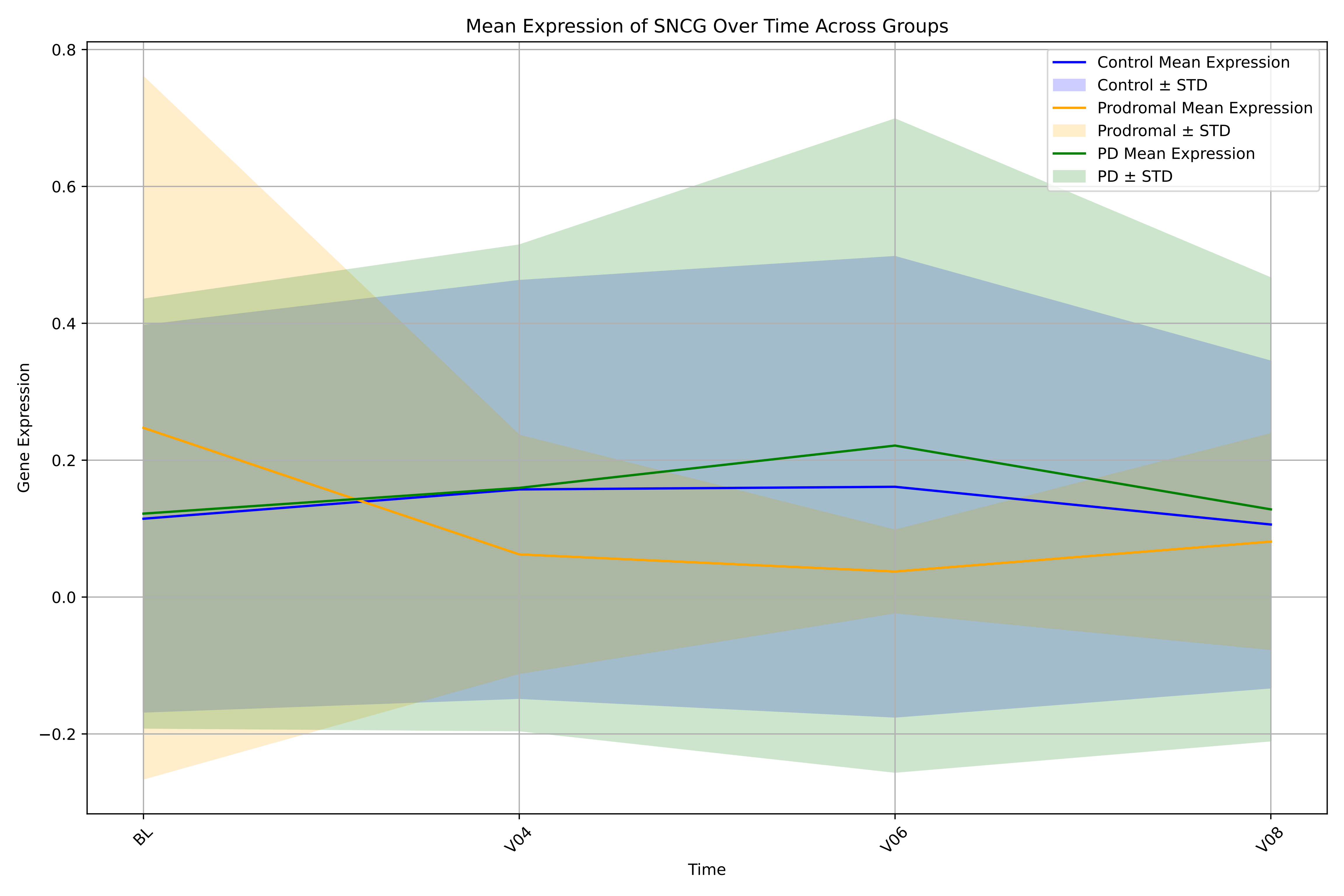

Mean Expression of SNCG Over Time Across Groups
Control Mean Expression
Control ± STD
Prodromal Mean Expression
Prodromal ± STD
PD Mean Expression
PD ± STD
Gene Expression
0.8
0.6
0.4
0.2
0.0
−0.2
BL
V04
V06
V08
Time

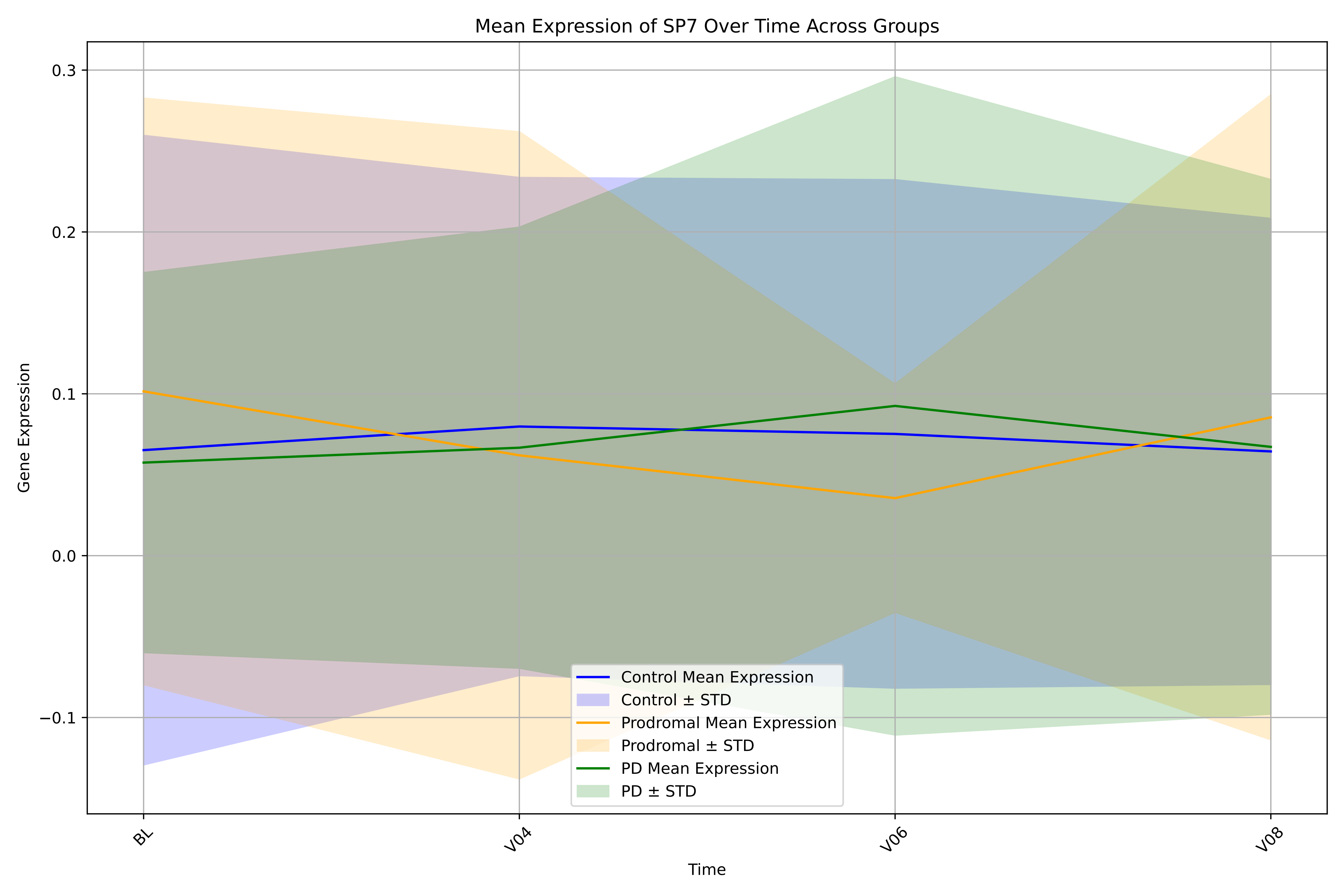

Mean Expression of SP7 Over Time Across Groups
Gene Expression
0.3
0.2
0.1
0.0
−0.1
BL
V04
V06
V08
Time
Control Mean Expression
Control ± STD
Prodromal Mean Expression
Prodromal ± STD
PD Mean Expression
PD ± STD

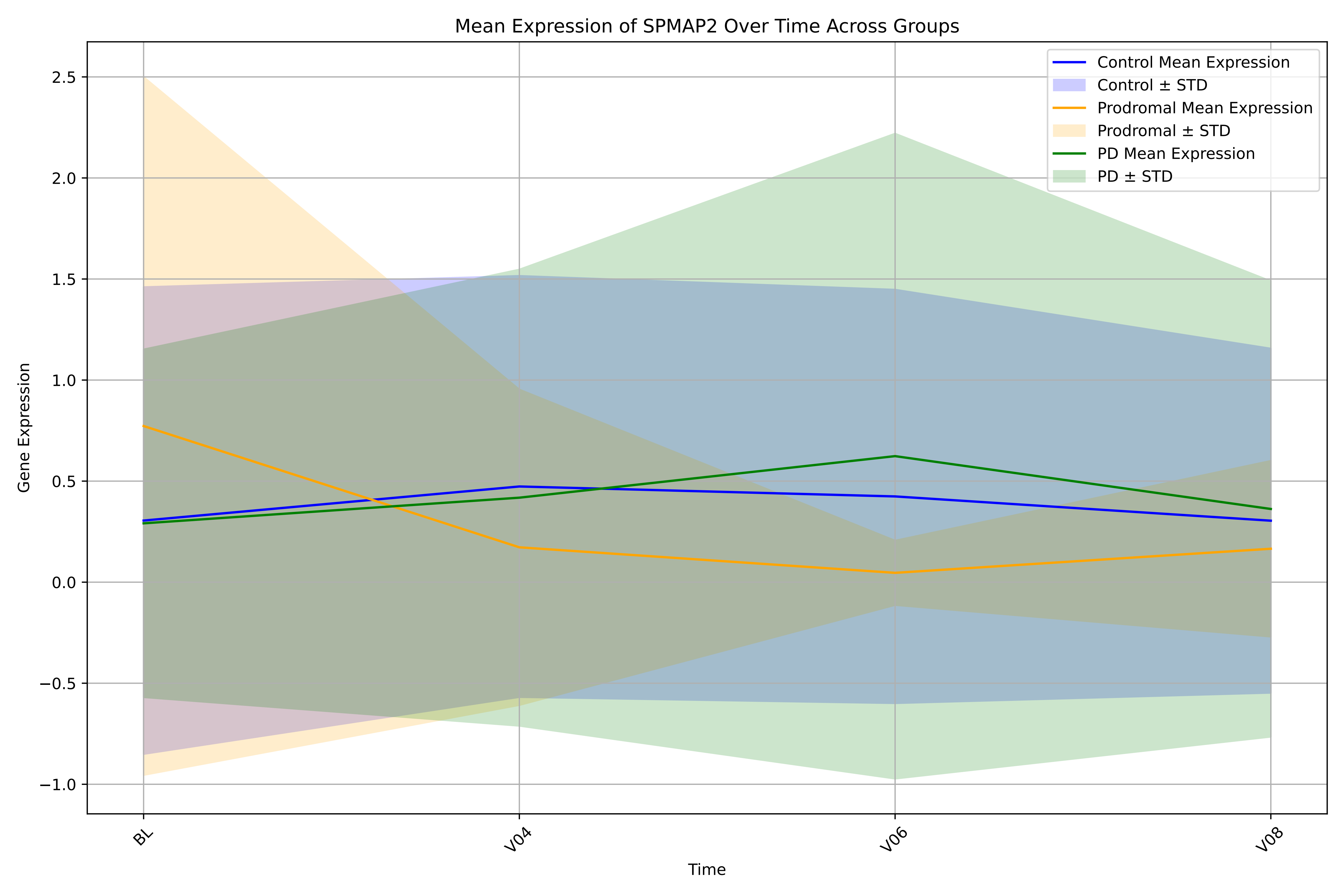

Mean Expression of SPMAP2 Over Time Across Groups
Control Mean Expression
Control ± STD
Prodromal Mean Expression
Prodromal ± STD
PD Mean Expression
PD ± STD
Gene Expression
2.5
2.0
1.5
1.0
0.5
0.0
−0.5
−1.0
BL
V04
V06
V08
Time

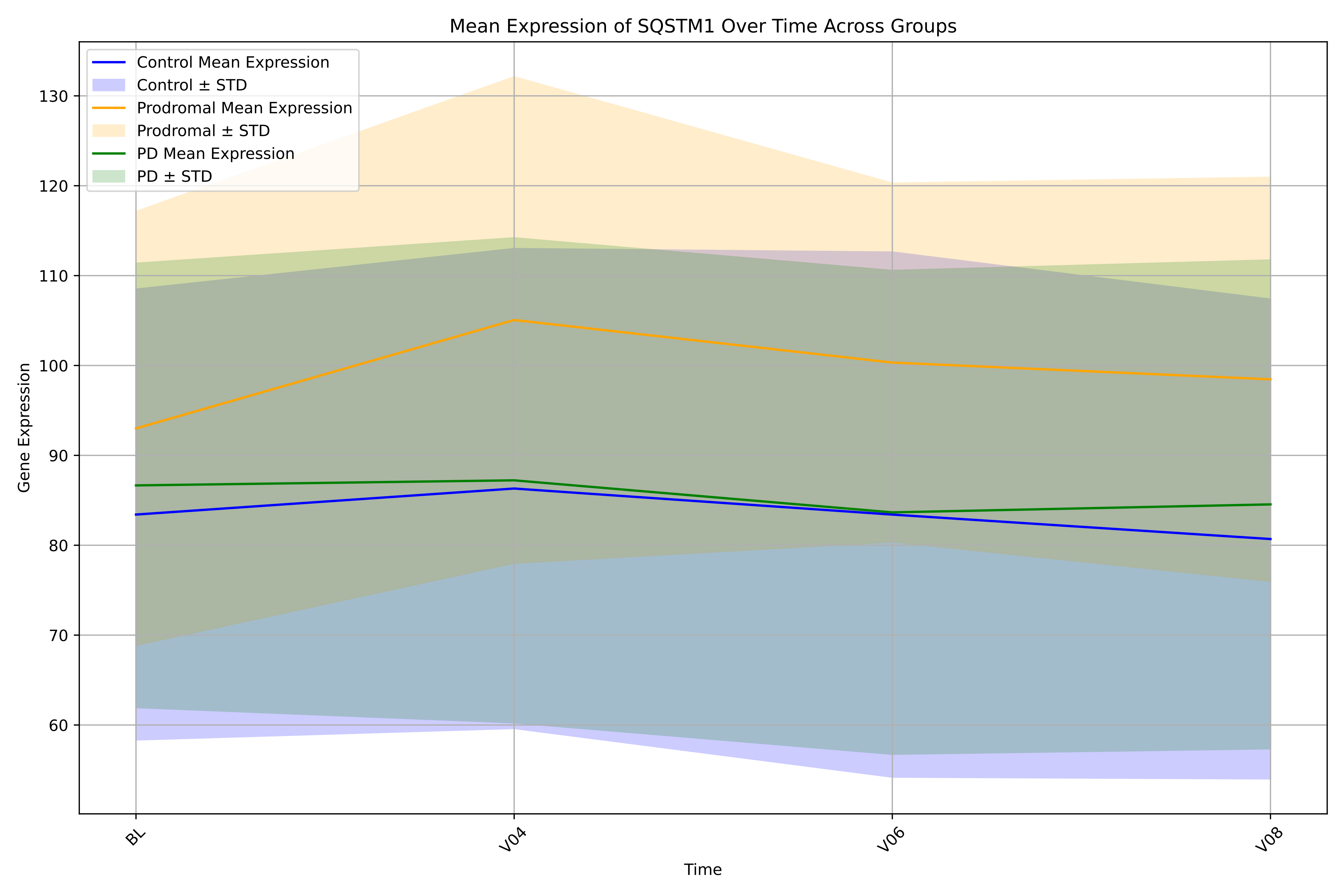

Mean Expression of SQSTM1 Over Time Across Groups
Control Mean Expression
Control ± STD
Prodromal Mean Expression
Prodromal ± STD
PD Mean Expression
PD ± STD
Gene Expression
60
70
80
90
100
110
120
130
BL
V04
V06
V08
Time

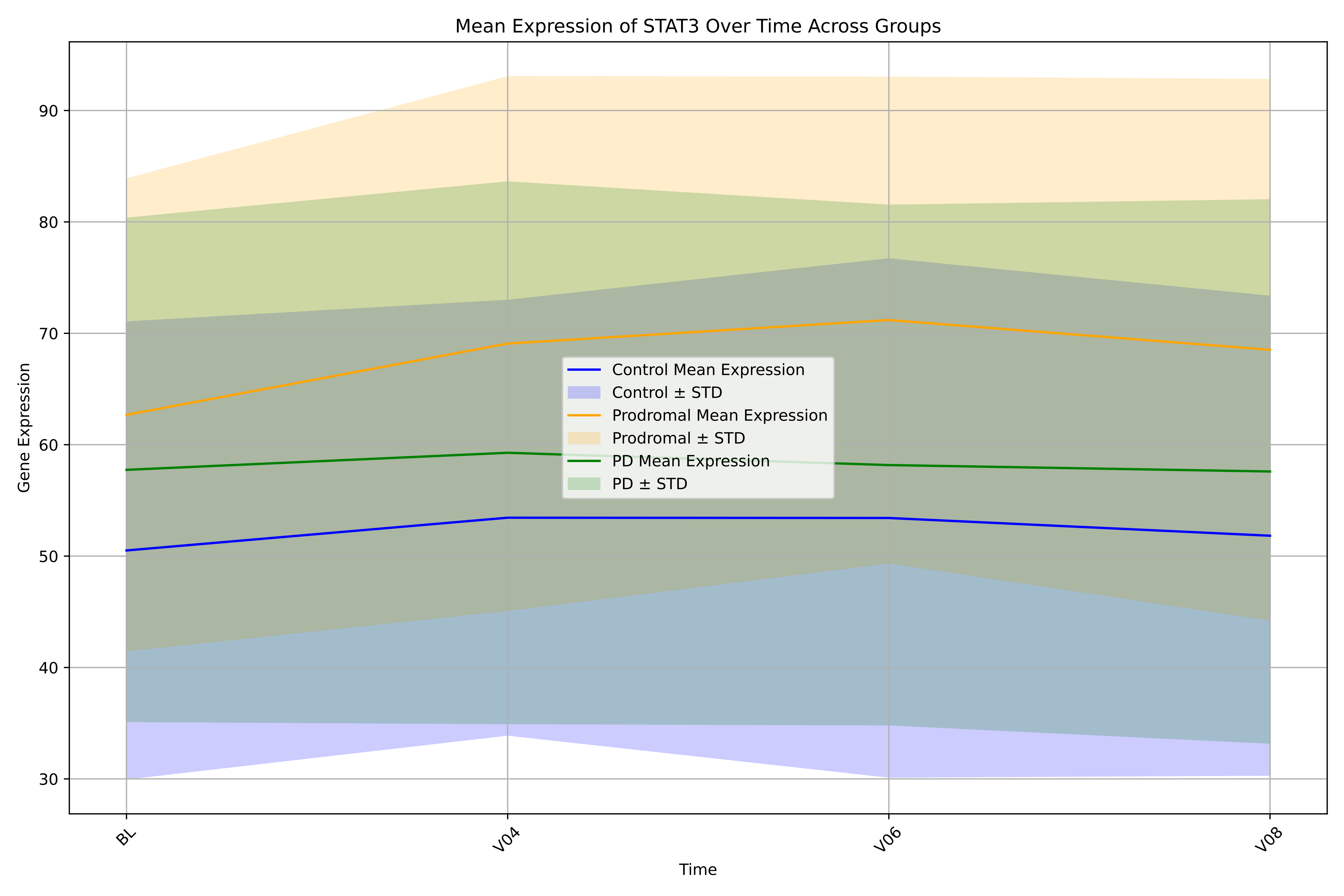

Mean Expression of STAT3 Over Time Across Groups
Control Mean Expression
Control ± STD
Prodromal Mean Expression
Prodromal ± STD
PD Mean Expression
PD ± STD
Gene Expression
90
80
70
60
50
40
30
BL
V04
V06
V08
Time

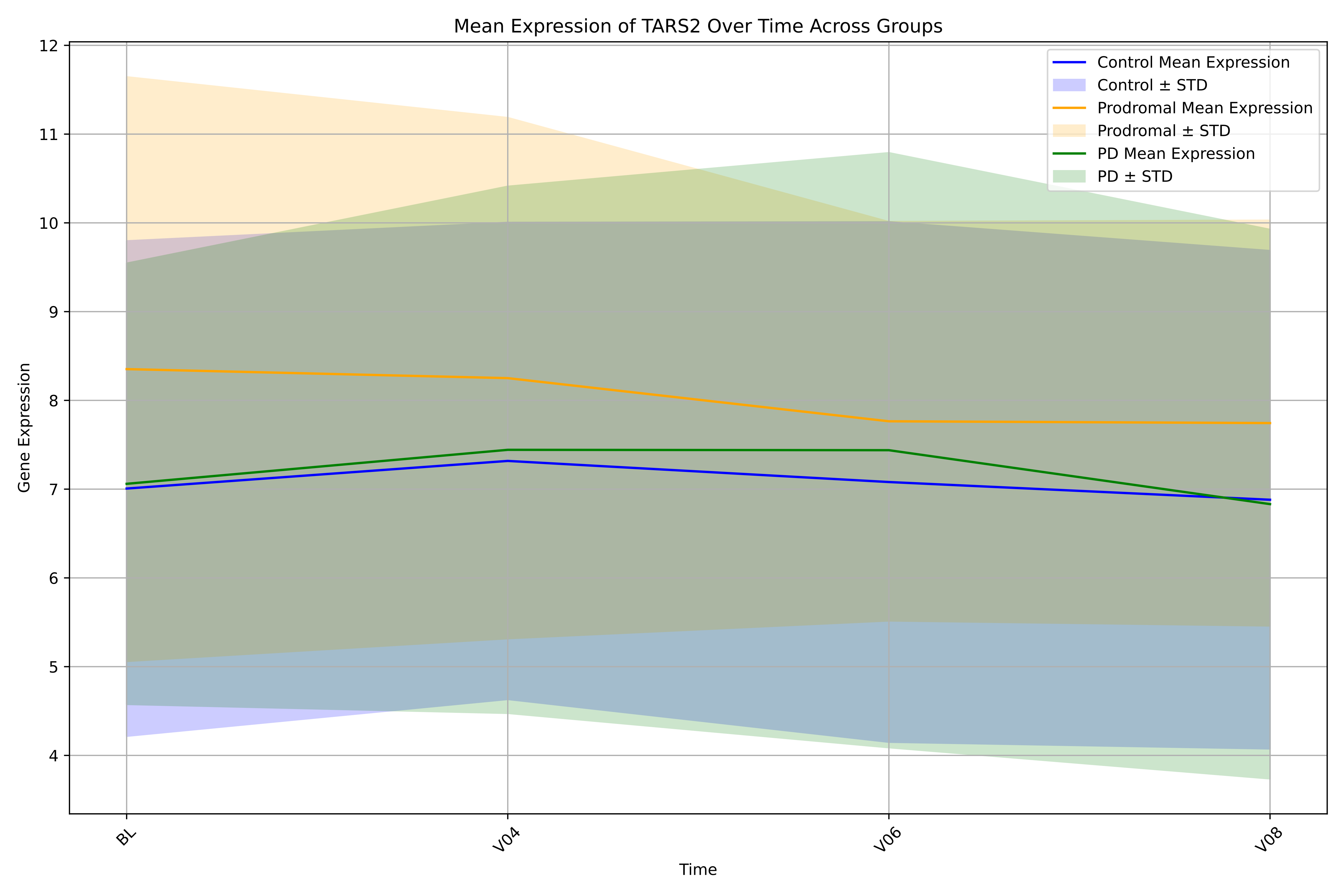

Mean Expression of TARS2 Over Time Across Groups
Control Mean Expression
Control ± STD
Prodromal Mean Expression
Prodromal ± STD
PD Mean Expression
PD ± STD
Gene Expression
Time
BL
V04
V06
V08
4
5
6
7
8
9
10
11
12

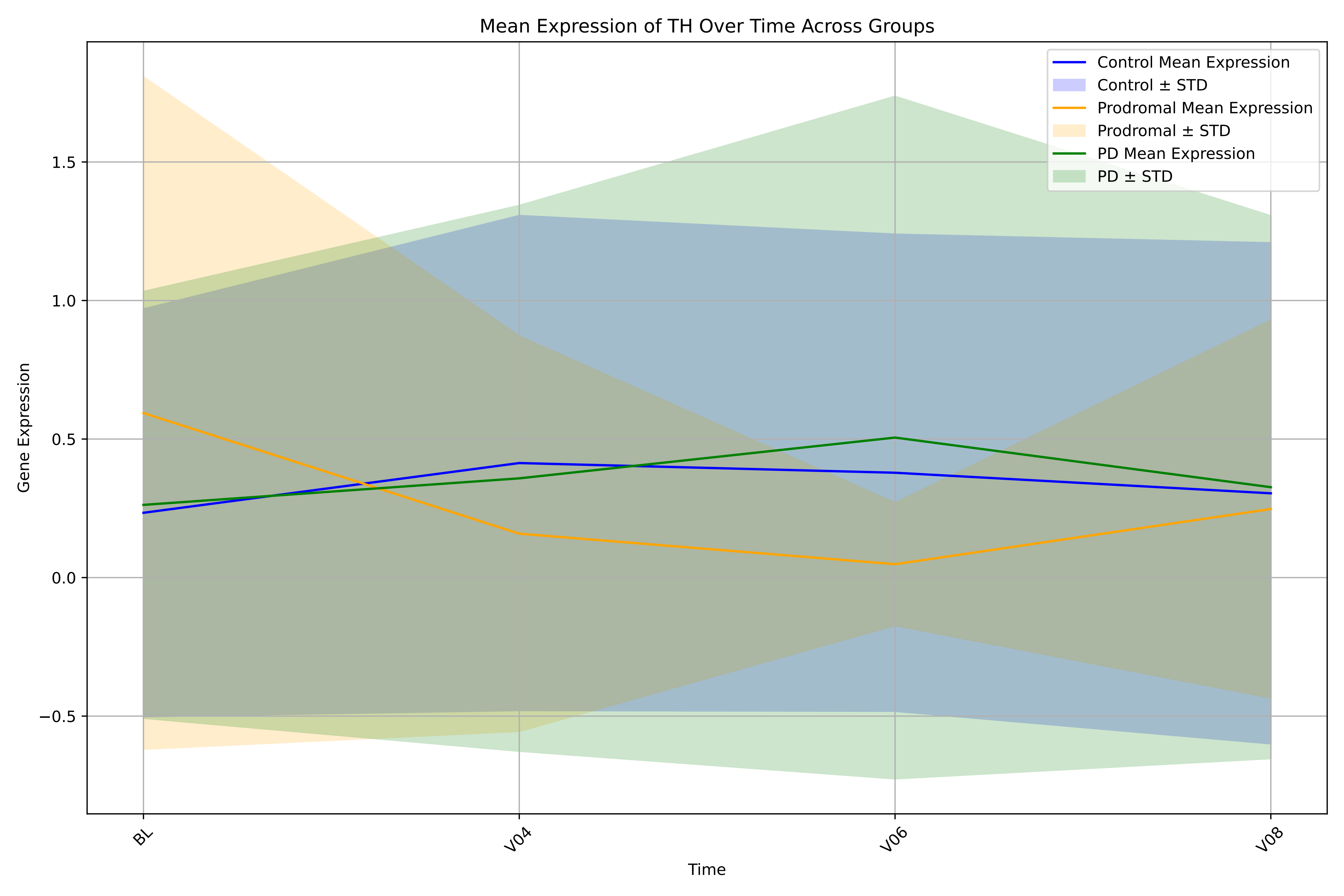

Mean Expression of TH Over Time Across Groups
Control Mean Expression
Control ± STD
Prodromal Mean Expression
Prodromal ± STD
PD Mean Expression
PD ± STD
Gene Expression
1.5
1.0
0.5
0.0
−0.5
BL
V04
V06
V08
Time

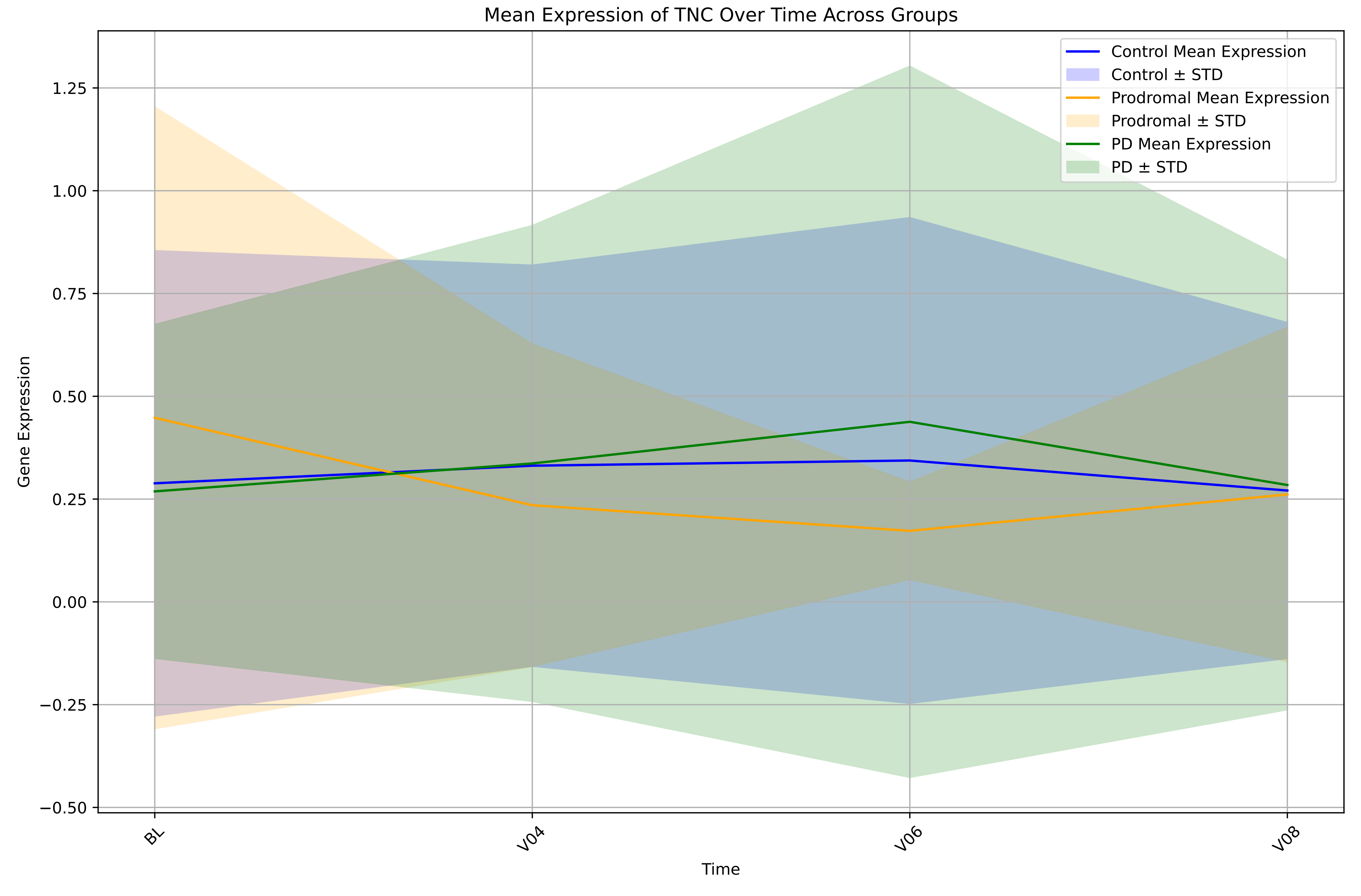

Mean Expression of TNC Over Time Across Groups
Control Mean Expression
Control ± STD
Prodromal Mean Expression
Prodromal ± STD
PD Mean Expression
PD ± STD
Gene Expression
Time
BL
V04
V06
V08
1.25
1.00
0.75
0.50
0.25
0.00
−0.25
−0.50

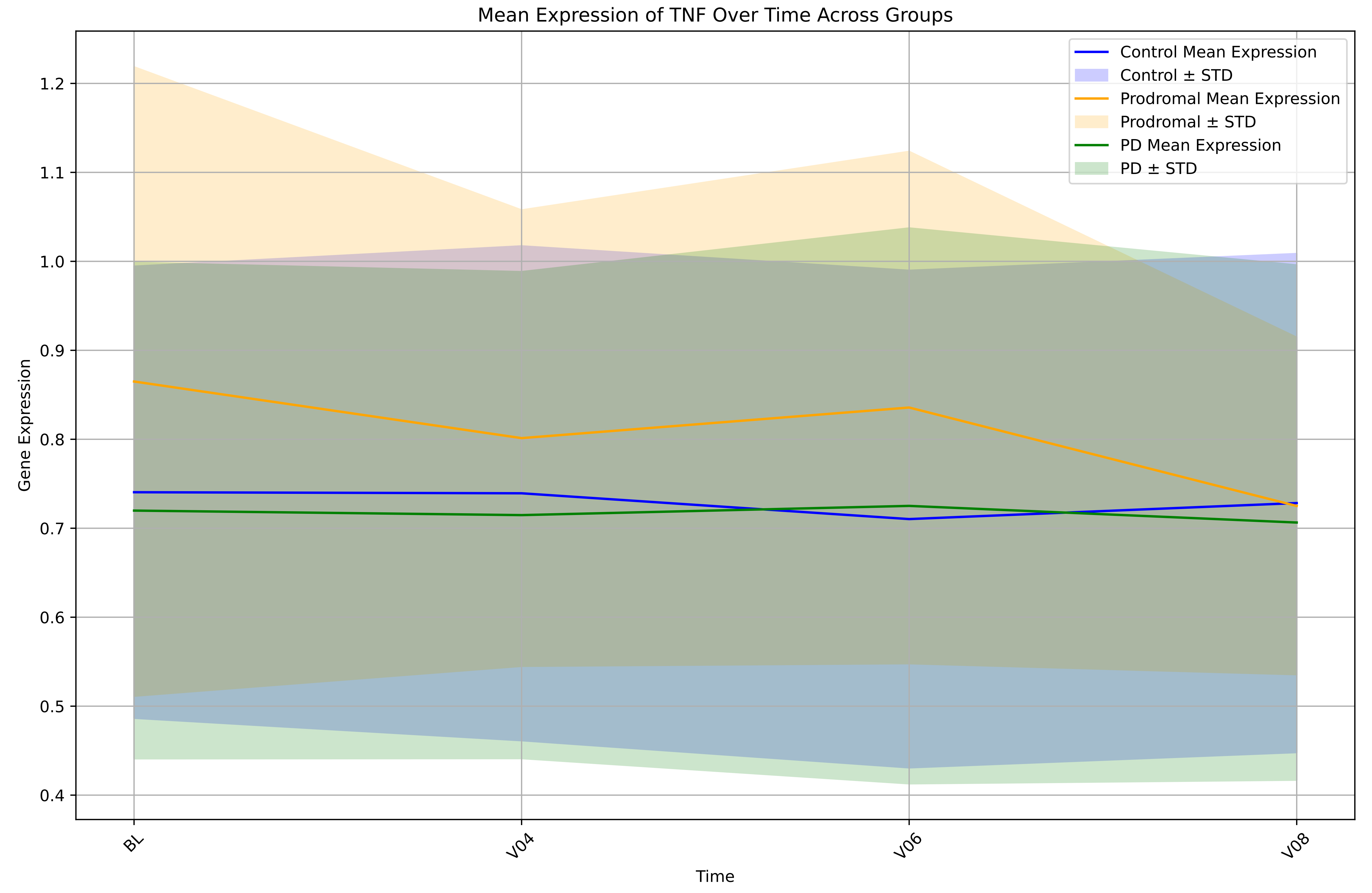
Mean Expression of TNF Over Time Across Groups
Control Mean Expression
Control ± STD
Prodromal Mean Expression
Prodromal ± STD
PD Mean Expression
PD ± STD
Gene Expression
1.2
1.1
1.0
0.9
0.8
0.7
0.6
0.5
0.4
BL
V04
V06
V08
Time

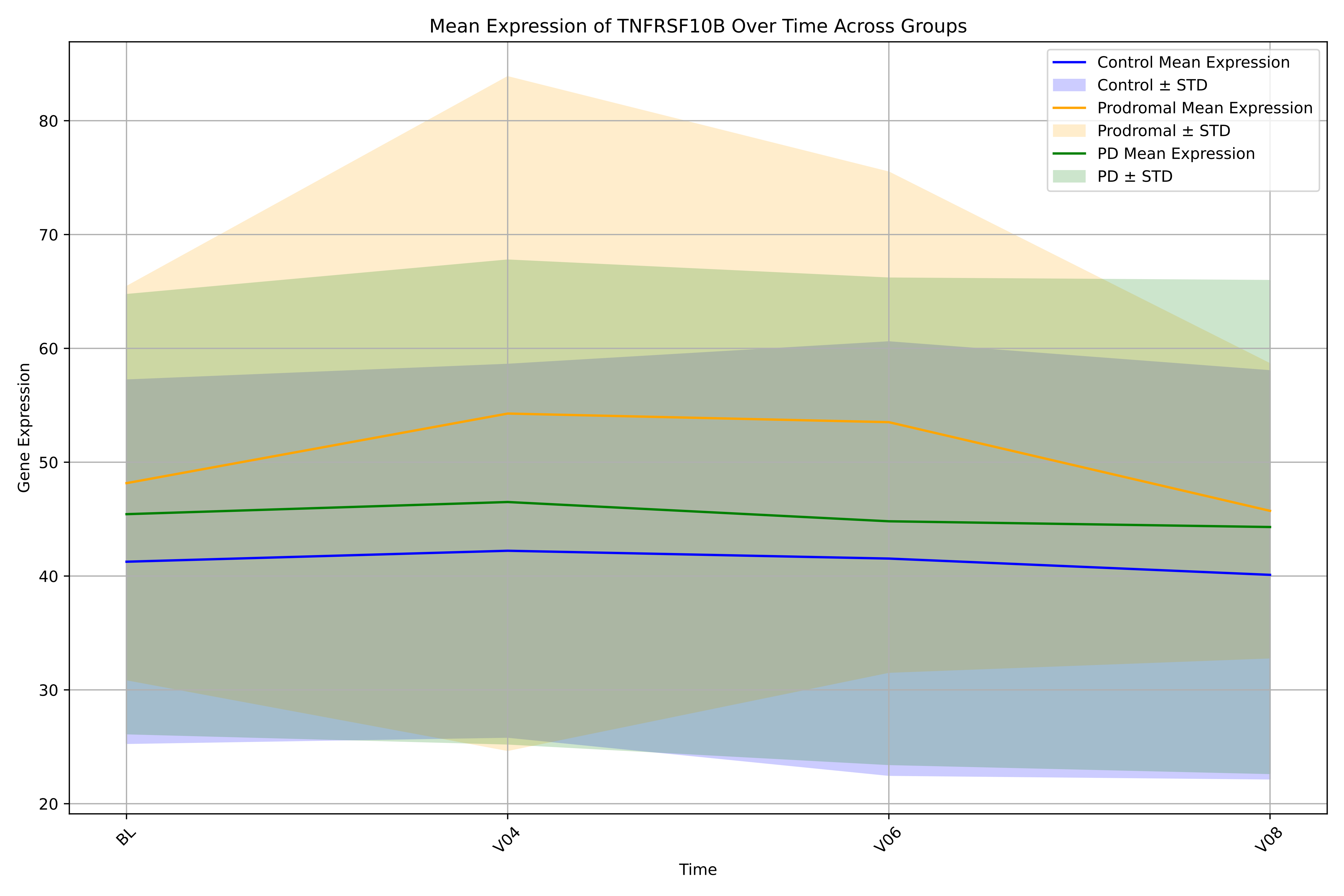

Mean Expression of TNFRSF10B Over Time Across Groups
Control Mean Expression
Control ± STD
Prodromal Mean Expression
Prodromal ± STD
PD Mean Expression
PD ± STD
80
70
60
50
40
30
20
Gene Expression
BL
V04
V06
V08
Time

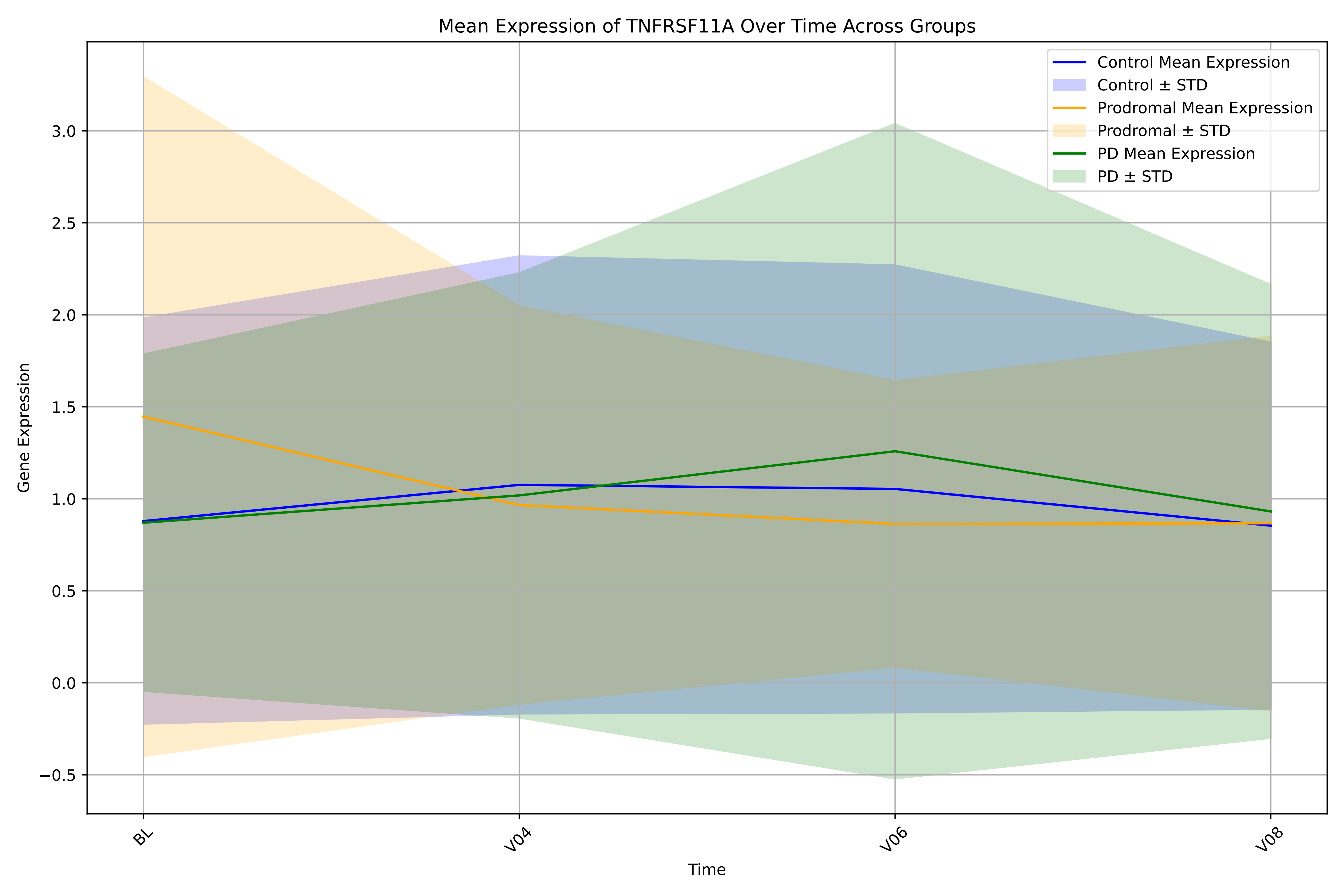

Mean Expression of TNFRSF11A Over Time Across Groups
Control Mean Expression
Control ± STD
Prodromal Mean Expression
Prodromal ± STD
PD Mean Expression
PD ± STD
Gene Expression
3.0
2.5
2.0
1.5
1.0
0.5
0.0
−0.5
BL
V04
V06
V08
Time

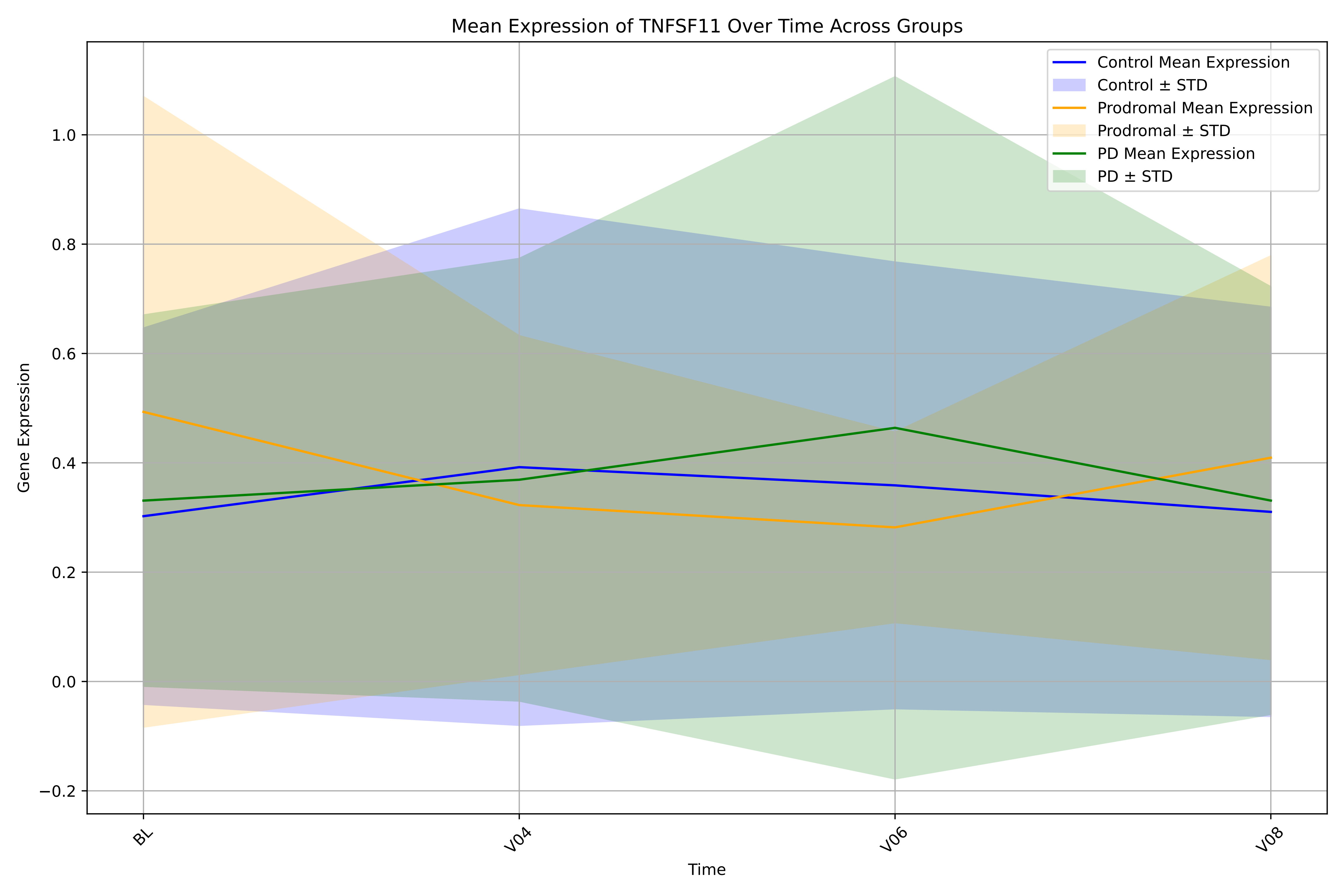

Mean Expression of TNFSF11 Over Time Across Groups
Control Mean Expression
Control ± STD
Prodromal Mean Expression
Prodromal ± STD
PD Mean Expression
PD ± STD
Gene Expression
1.0
0.8
0.6
0.4
0.2
0.0
−0.2
BL
V04
V06
V08
Time

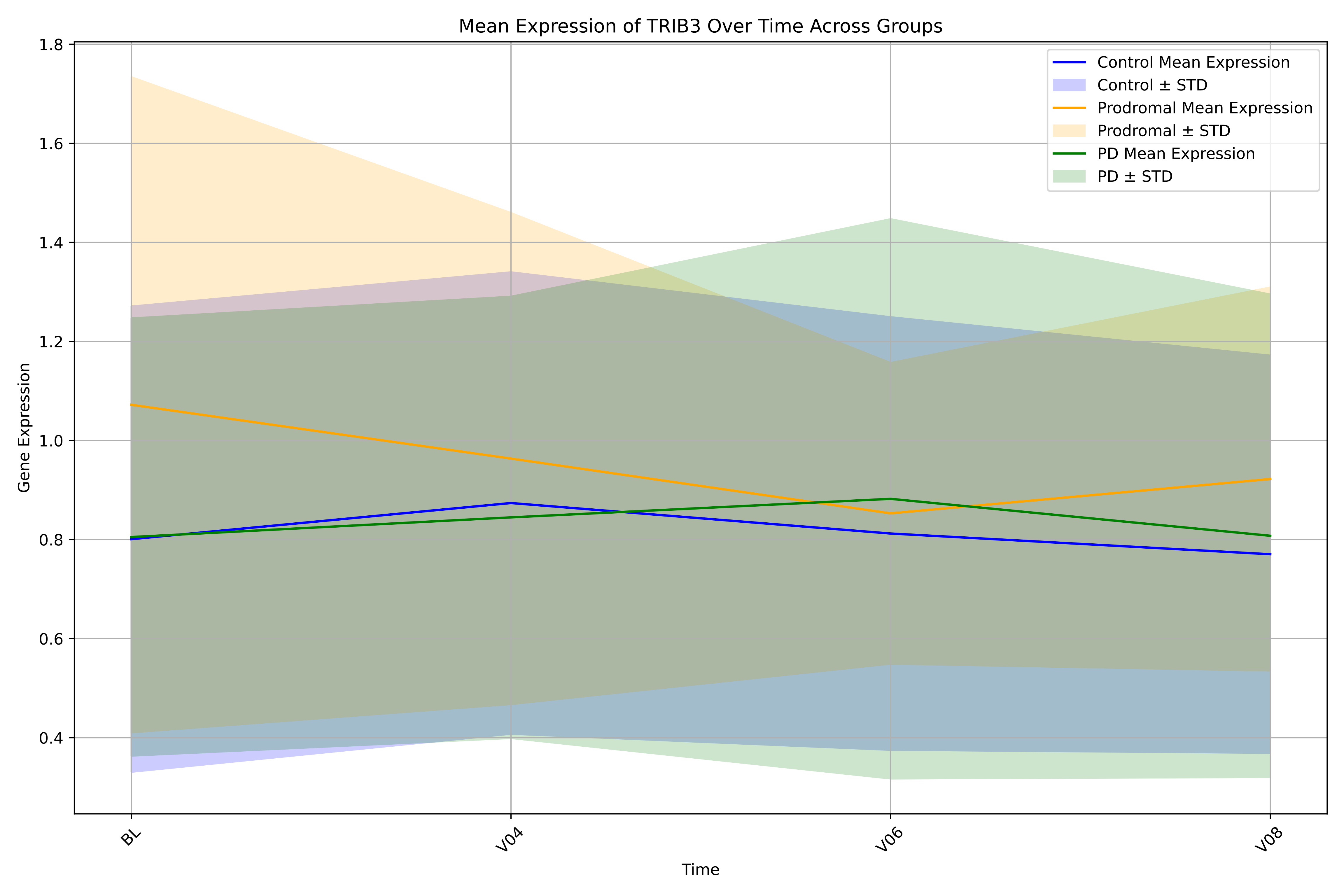

Mean Expression of TRIB3 Over Time Across Groups
Control Mean Expression
Control ± STD
Prodromal Mean Expression
Prodromal ± STD
PD Mean Expression
PD ± STD
Gene Expression
1.8
1.6
1.4
1.2
1.0
0.8
0.6
0.4
BL
V04
V06
V08
Time

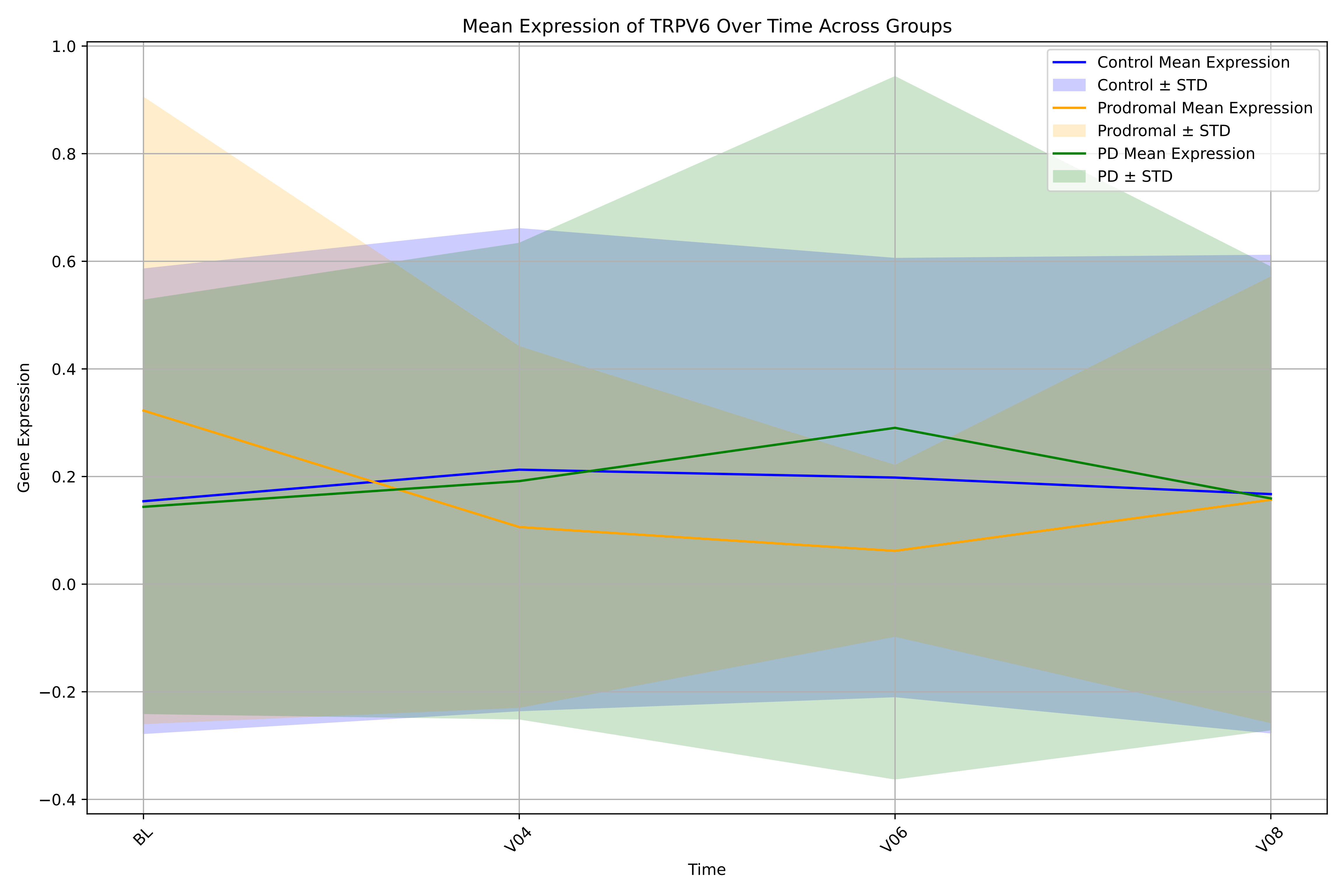

Mean Expression of TRPV6 Over Time Across Groups
Control Mean Expression
Control ± STD
Prodromal Mean Expression
Prodromal ± STD
PD Mean Expression
PD ± STD
Gene Expression
1.0
0.8
0.6
0.4
0.2
0.0
−0.2
−0.4
BL
V04
V06
V08
Time

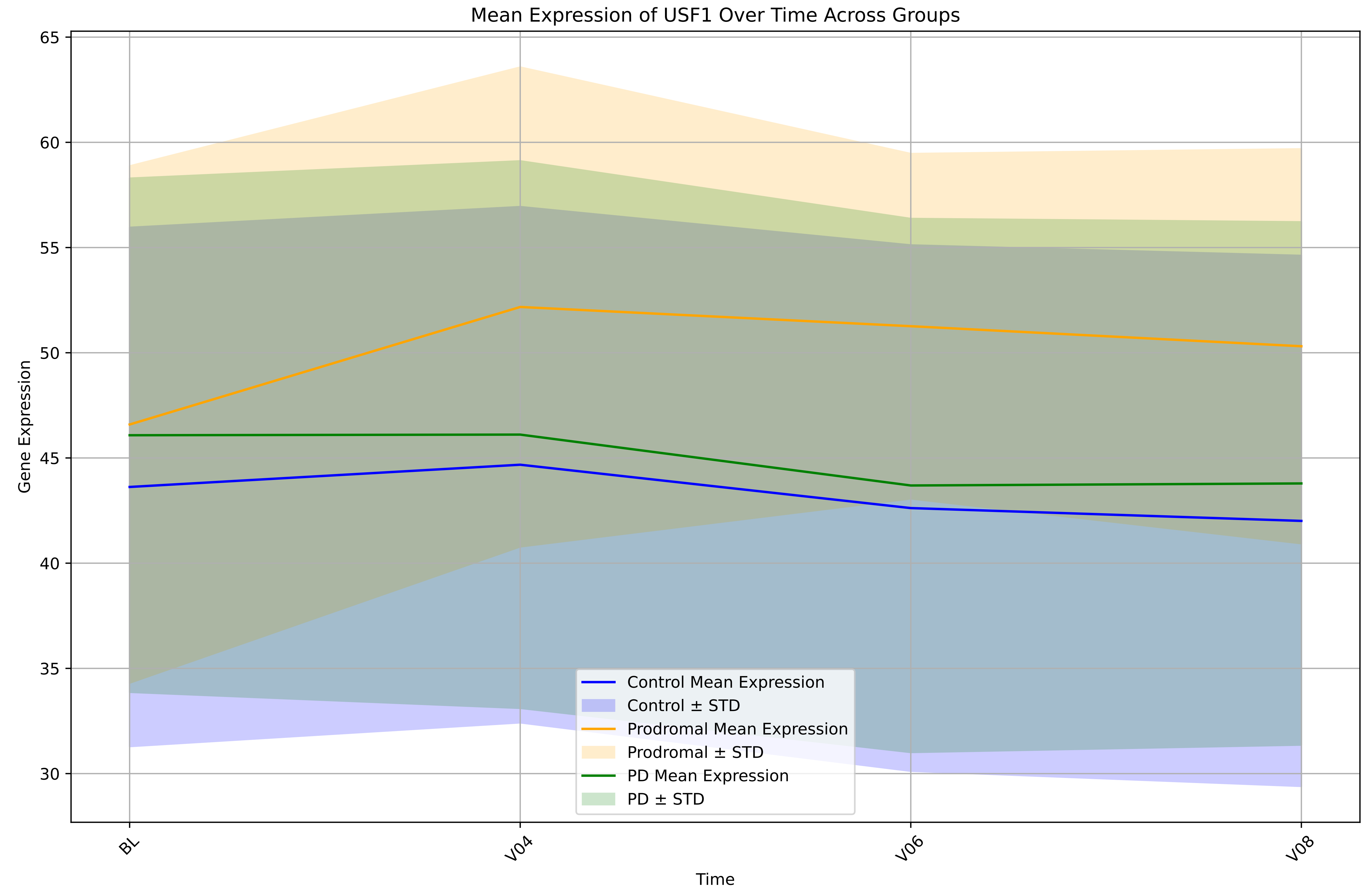

Mean Expression of USF1 Over Time Across Groups
Gene Expression
Time
65
60
55
50
45
40
35
30
BL
V04
V06
V08
Control Mean Expression
Control ± STD
Prodromal Mean Expression
Prodromal ± STD
PD Mean Expression
PD ± STD

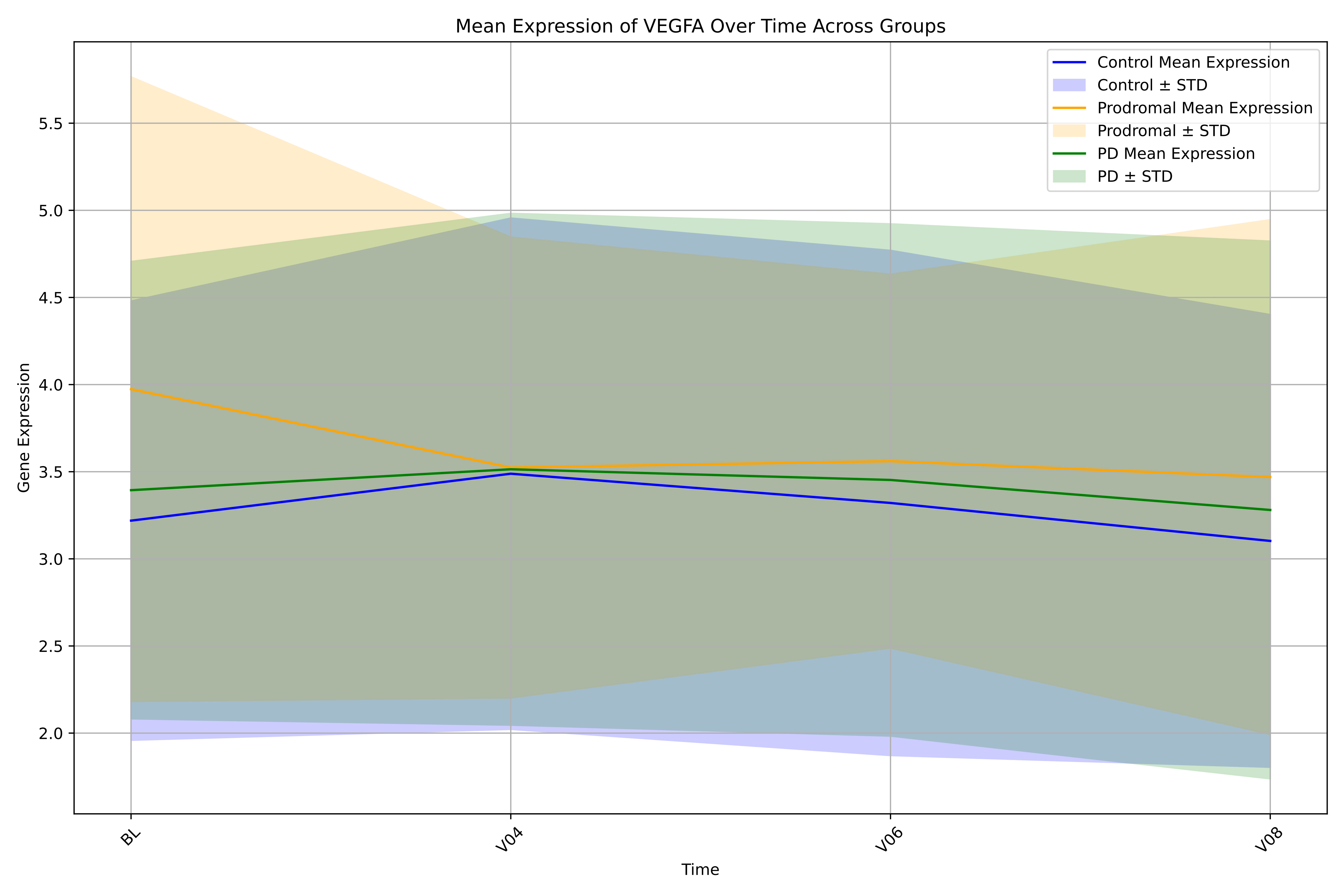

Mean Expression of VEGFA Over Time Across Groups
Control Mean Expression
Control ± STD
Prodromal Mean Expression
Prodromal ± STD
PD Mean Expression
PD ± STD
Gene Expression
2.0
2.5
3.0
3.5
4.0
4.5
5.0
5.5
BL
V04
V06
V08
Time

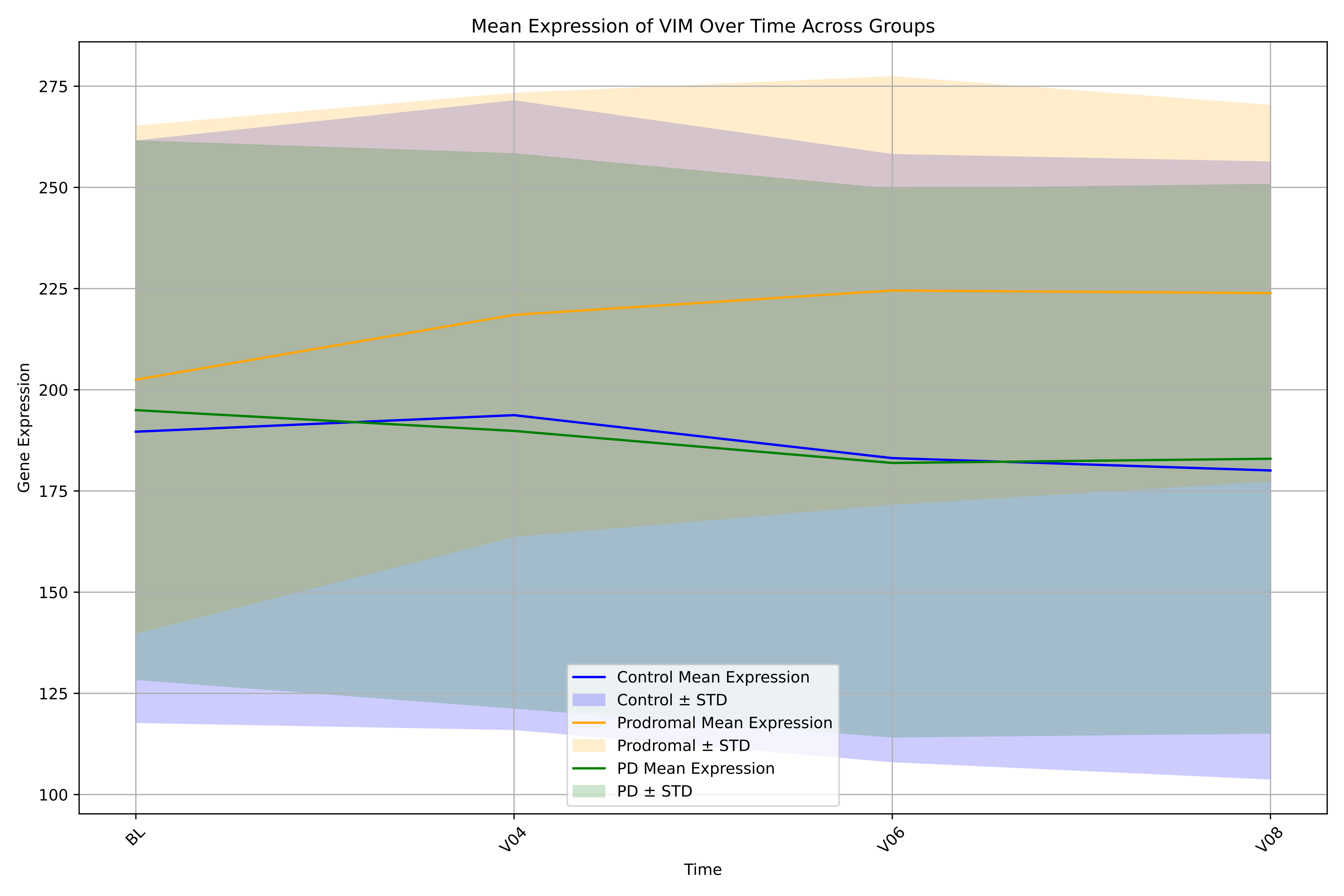

Mean Expression of VIM Over Time Across Groups
Gene Expression
275
250
225
200
175
150
125
100
BL
V04
V06
V08
Time
Control Mean Expression
Control ± STD
Prodromal Mean Expression
Prodromal ± STD
PD Mean Expression
PD ± STD

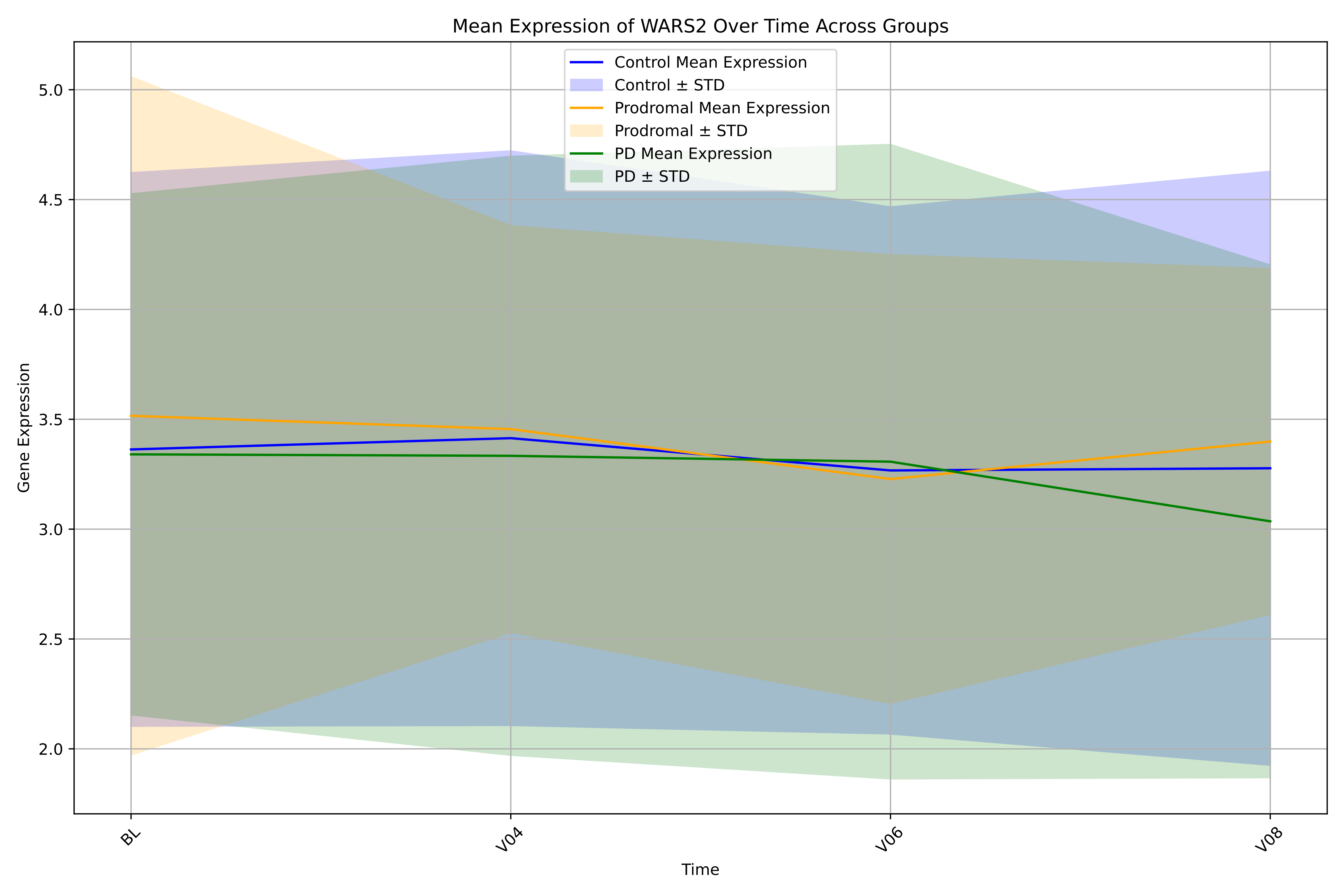

Mean Expression of WARS2 Over Time Across Groups
Control Mean Expression
Control ± STD
Prodromal Mean Expression
Prodromal ± STD
PD Mean Expression
PD ± STD
Gene Expression
Time
5.0
4.5
4.0
3.5
3.0
2.5
2.0
BL
V04
V06
V08

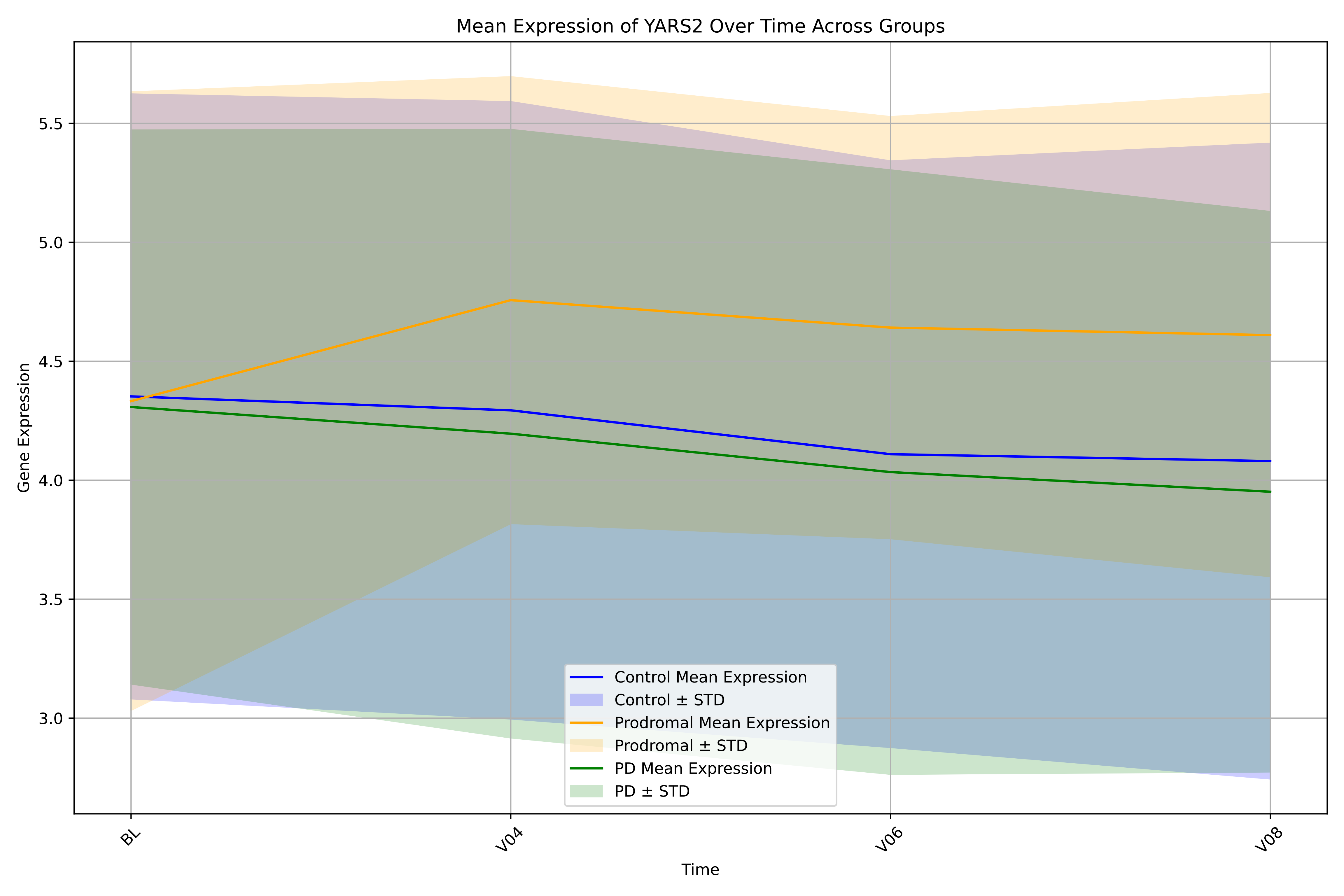

Mean Expression of YARS2 Over Time Across Groups
Gene Expression
Time
BL
V04
V06
V08
3.0
3.5
4.0
4.5
5.0
5.5
Control Mean Expression
Control ± STD
Prodromal Mean Expression
Prodromal ± STD
PD Mean Expression
PD ± STD
