## Supplementary material for "Longitudinal Assessment of DNA Repair Signature Trajectory in Prodromal versus Established Parkinson’s Disease": Table S2

| | baseMean | log2FoldCha | lfcSE | stat | pvalue | padj |
|---|---|---|---|---|---|---|
| BMX | 250,02 | -0,40 | 0,07 | -5,96 | 2,53E-09 | 2,79E-05 |
| PRRG4 | 1326,58 | -0,30 | 0,05 | -5,75 | 8,93E-09 | 3,29E-05 |
| TMCC3 | 3073,26 | -0,23 | 0,04 | -5,81 | 6,20E-09 | 3,29E-05 |
| KPNB1 | 6158,34 | -0,16 | 0,03 | -5,65 | 1,60E-08 | 3,62E-05 |
| LSMEM1 | 349,43 | -0,25 | 0,04 | -5,65 | 1,64E-08 | 3,62E-05 |
| HAPSTR1 | 10103,44 | -0,13 | 0,02 | -5,45 | 5,09E-08 | 9,31E-05 |
| CXorf38 | 1970,79 | -0,09 | 0,02 | -5,41 | 6,37E-08 | 9,31E-05 |
| SLA | 14557,36 | -0,16 | 0,03 | -5,40 | 6,74E-08 | 9,31E-05 |
| RNF149 | 22225,66 | -0,17 | 0,03 | -5,36 | 8,34E-08 | 1,02E-04 |
| RAB27A | 3280,14 | -0,14 | 0,03 | -5,34 | 9,37E-08 | 1,04E-04 |
| MCTP2 | 10101,06 | -0,19 | 0,04 | -5,27 | 1,34E-07 | 1,14E-04 |
| ROPN1L | 418,14 | -0,23 | 0,04 | -5,28 | 1,31E-07 | 1,14E-04 |
| PPP1R3B | 4753,47 | -0,22 | 0,04 | -5,30 | 1,17E-07 | 1,14E-04 |
| FBXL13 | 835,54 | -0,28 | 0,05 | -5,26 | 1,46E-07 | 1,15E-04 |
| MSL1 | 8824,56 | -0,17 | 0,03 | -5,23 | 1,69E-07 | 1,25E-04 |
| SYTL3 | 1490,46 | -0,19 | 0,04 | -5,18 | 2,19E-07 | 1,27E-04 |
| LITAF | 14622,38 | -0,20 | 0,04 | -5,18 | 2,17E-07 | 1,27E-04 |
| CFLAR | 37589,91 | -0,15 | 0,03 | -5,19 | 2,11E-07 | 1,27E-04 |
| RNASEL | 2966,25 | -0,13 | 0,03 | -5,19 | 2,11E-07 | 1,27E-04 |
| TGFA | 867,97 | -0,21 | 0,04 | -5,17 | 2,32E-07 | 1,28E-04 |
| YIPF1 | 646,86 | -0,12 | 0,02 | -5,14 | 2,70E-07 | 1,30E-04 |
| CPD | 8626,77 | -0,19 | 0,04 | -5,15 | 2,61E-07 | 1,30E-04 |
| ACSL1 | 27698,24 | -0,27 | 0,05 | -5,16 | 2,49E-07 | 1,30E-04 |
| CEMIP2 | 6259,91 | -0,11 | 0,02 | -5,11 | 3,21E-07 | 1,42E-04 |
| VNN3P | 3580,01 | -0,26 | 0,05 | -5,11 | 3,17E-07 | 1,42E-04 |
| LMTK2 | 2109,59 | -0,19 | 0,04 | -5,08 | 3,79E-07 | 1,61E-04 |
| FPR2 | 6355,18 | -0,24 | 0,05 | -5,05 | 4,30E-07 | 1,70E-04 |
| TMEM252 | 128,47 | -0,28 | 0,06 | -5,06 | 4,17E-07 | 1,70E-04 |
| CA4 | 352,54 | -0,29 | 0,06 | -5,04 | 4,58E-07 | 1,75E-04 |
| ARHGAP19 | 1422,64 | -0,14 | 0,03 | -5,04 | 4,74E-07 | 1,75E-04 |
| NUP50 | 5703,27 | -0,12 | 0,02 | -5,03 | 4,94E-07 | 1,76E-04 |
| TLK2 | 2454,18 | -0,07 | 0,01 | -5,02 | 5,20E-07 | 1,80E-04 |
| MXD1 | 25392,87 | -0,20 | 0,04 | -4,99 | 5,91E-07 | 1,92E-04 |
| APMAP | 4816,52 | -0,18 | 0,04 | -5,00 | 5,78E-07 | 1,92E-04 |
| CISD2 | 1188,29 | 0,28 | 0,06 | 4,94 | 7,85E-07 | 1,98E-04 |
| RNF169 | 5600,35 | -0,11 | 0,02 | -4,97 | 6,83E-07 | 1,98E-04 |
| DNAJC3 | 3989,74 | -0,15 | 0,03 | -4,96 | 7,20E-07 | 1,98E-04 |
| KIAA0040 | 3916,01 | -0,14 | 0,03 | -4,96 | 6,94E-07 | 1,98E-04 |
| IST1 | 4631,44 | -0,08 | 0,02 | -4,94 | 7,87E-07 | 1,98E-04 |
| AQP9 | 15156,09 | -0,22 | 0,04 | -4,97 | 6,62E-07 | 1,98E-04 |
| CLEC4E | 2770,11 | -0,25 | 0,05 | -4,94 | 7,70E-07 | 1,98E-04 |
| BOD1L1 | 14062,30 | -0,14 | 0,03 | -4,94 | 7,79E-07 | 1,98E-04 |
| TRANK1 | 12888,14 | -0,19 | 0,04 | -4,96 | 7,00E-07 | 1,98E-04 |
| HEBP2 | 3976,65 | -0,13 | 0,03 | -4,94 | 7,90E-07 | 1,98E-04 |
| MAP3K5 | 6523,74 | -0,13 | 0,03 | -4,92 | 8,77E-07 | 2,04E-04 |

| | | | | | | |
|---|---|---|---|---|---|---|
| LCP1 | 45211,17 | -0,14 | 0,03 | -4,92 | 8,85E-07 | 2,04E-04 |
| KIF13A | 3676,16 | -0,19 | 0,04 | -4,92 | 8,50E-07 | 2,04E-04 |
| CKLF | 953,20 | -0,15 | 0,03 | -4,92 | 8,61E-07 | 2,04E-04 |
| KAT6A | 11184,97 | -0,11 | 0,02 | -4,90 | 9,35E-07 | 2,10E-04 |
| ZNF106 | 8970,65 | -0,13 | 0,03 | -4,90 | 9,52E-07 | 2,10E-04 |
| HSDL2 | 2061,97 | -0,17 | 0,04 | -4,89 | 1,00E-06 | 2,17E-04 |
| CWC25 | 1129,15 | -0,09 | 0,02 | -4,87 | 1,10E-06 | 2,34E-04 |
| ZNF200 | 1249,35 | -0,11 | 0,02 | -4,85 | 1,22E-06 | 2,48E-04 |
| DIP2B | 7087,75 | -0,13 | 0,03 | -4,86 | 1,20E-06 | 2,48E-04 |
| CR1 | 6345,37 | -0,23 | 0,05 | -4,85 | 1,24E-06 | 2,48E-04 |
| TMEM81 | 150,04 | -0,10 | 0,02 | -4,82 | 1,42E-06 | 2,81E-04 |
| STX3 | 5516,66 | -0,20 | 0,04 | -4,82 | 1,46E-06 | 2,83E-04 |
| NDEL1 | 6859,35 | -0,17 | 0,04 | -4,81 | 1,49E-06 | 2,84E-04 |
| LMNB1 | 1970,29 | -0,22 | 0,05 | -4,80 | 1,60E-06 | 2,97E-04 |
| RAF1 | 7791,08 | -0,15 | 0,03 | -4,80 | 1,61E-06 | 2,97E-04 |
| STK4 | 21826,34 | -0,10 | 0,02 | -4,77 | 1,83E-06 | 3,27E-04 |
| UBXN2B | 3176,44 | -0,16 | 0,03 | -4,77 | 1,84E-06 | 3,27E-04 |
| CEP63 | 3109,60 | -0,14 | 0,03 | -4,77 | 1,87E-06 | 3,28E-04 |
| ENSG000001 | 51155,41 | -0,20 | 0,04 | -4,74 | 2,14E-06 | 3,69E-04 |
| NRDC | 6161,33 | -0,11 | 0,02 | -4,73 | 2,23E-06 | 3,74E-04 |
| TACC1 | 8101,13 | -0,11 | 0,02 | -4,73 | 2,20E-06 | 3,74E-04 |
| STEAP4 | 12400,31 | -0,22 | 0,05 | -4,72 | 2,36E-06 | 3,84E-04 |
| ABHD5 | 6957,49 | -0,20 | 0,04 | -4,72 | 2,37E-06 | 3,84E-04 |
| IP6K2 | 1226,45 | -0,07 | 0,01 | -4,72 | 2,41E-06 | 3,85E-04 |
| ARHGAP25 | 10476,30 | -0,12 | 0,03 | -4,71 | 2,46E-06 | 3,87E-04 |
| RAB11FIP1 | 10454,92 | -0,17 | 0,04 | -4,71 | 2,49E-06 | 3,87E-04 |
| SRPK1 | 2831,88 | -0,17 | 0,04 | -4,71 | 2,52E-06 | 3,87E-04 |
| SPTY2D1 | 1100,06 | -0,08 | 0,02 | -4,70 | 2,61E-06 | 3,95E-04 |
| TMEM140 | 4541,12 | -0,18 | 0,04 | -4,69 | 2,67E-06 | 3,98E-04 |
| GPAT3 | 1417,43 | -0,20 | 0,04 | -4,69 | 2,78E-06 | 4,10E-04 |
| PSEN1 | 5366,09 | -0,11 | 0,02 | -4,68 | 2,83E-06 | 4,11E-04 |
| B4GALT5 | 3535,31 | -0,20 | 0,04 | -4,68 | 2,94E-06 | 4,22E-04 |
| ENSG000002 | 313,06 | -0,24 | 0,05 | -4,67 | 3,00E-06 | 4,25E-04 |
| TMEM71 | 5223,83 | -0,14 | 0,03 | -4,67 | 3,06E-06 | 4,28E-04 |
| PHF7 | 186,69 | 0,09 | 0,02 | 4,66 | 3,22E-06 | 4,44E-04 |
| GABARAPL1 | 3186,93 | -0,15 | 0,03 | -4,64 | 3,44E-06 | 4,69E-04 |
| MTARC1 | 1346,76 | -0,24 | 0,05 | -4,64 | 3,54E-06 | 4,71E-04 |
| APAF1 | 8747,94 | -0,15 | 0,03 | -4,64 | 3,54E-06 | 4,71E-04 |
| ANKRD13A | 8463,25 | -0,12 | 0,02 | -4,63 | 3,58E-06 | 4,71E-04 |
| NFIL3 | 763,77 | -0,20 | 0,04 | -4,63 | 3,73E-06 | 4,76E-04 |
| RTF1 | 1203,12 | -0,08 | 0,02 | -4,62 | 3,79E-06 | 4,76E-04 |
| STS | 261,62 | -0,14 | 0,03 | -4,62 | 3,79E-06 | 4,76E-04 |
| CRISPLD2 | 3100,55 | -0,28 | 0,06 | -4,62 | 3,79E-06 | 4,76E-04 |
| PLXNC1 | 17639,56 | -0,17 | 0,04 | -4,61 | 3,96E-06 | 4,87E-04 |
| BIN2 | 6399,44 | -0,09 | 0,02 | -4,61 | 3,97E-06 | 4,87E-04 |
| CCN3 | 340,21 | -0,27 | 0,06 | -4,61 | 4,03E-06 | 4,90E-04 |

| | | | | | | |
|---|---|---|---|---|---|---|
| KDM5B | 2367,08 | -0,10 | 0,02 | -4,60 | 4,16E-06 | 5,00E-04 |
| IQGAP1 | 23770,96 | -0,15 | 0,03 | -4,60 | 4,23E-06 | 5,03E-04 |
| RBBP5 | 1778,80 | -0,08 | 0,02 | -4,59 | 4,41E-06 | 5,18E-04 |
| SLC37A3 | 1284,25 | -0,17 | 0,04 | -4,58 | 4,57E-06 | 5,31E-04 |
| KCNJ15 | 7688,27 | -0,26 | 0,06 | -4,57 | 4,82E-06 | 5,55E-04 |
| FCGR2A | 18931,72 | -0,18 | 0,04 | -4,55 | 5,36E-06 | 6,11E-04 |
| CREB5 | 6794,76 | -0,22 | 0,05 | -4,55 | 5,44E-06 | 6,14E-04 |
| HIF1A | 4618,53 | -0,14 | 0,03 | -4,54 | 5,56E-06 | 6,15E-04 |
| TLE4 | 4814,90 | -0,10 | 0,02 | -4,54 | 5,52E-06 | 6,15E-04 |
| MGAM | 10220,98 | -0,25 | 0,06 | -4,54 | 5,65E-06 | 6,18E-04 |
| SLC2A3 | 9806,73 | -0,19 | 0,04 | -4,54 | 5,76E-06 | 6,24E-04 |
| RUBCNL | 2480,42 | -0,22 | 0,05 | -4,53 | 5,87E-06 | 6,29E-04 |
| SERINC3 | 4297,89 | -0,09 | 0,02 | -4,52 | 6,21E-06 | 6,59E-04 |
| CACUL1 | 6407,68 | -0,12 | 0,03 | -4,51 | 6,61E-06 | 6,90E-04 |
| TBL2 | 559,93 | 0,07 | 0,02 | 4,51 | 6,62E-06 | 6,90E-04 |
| FCAR | 1108,60 | -0,23 | 0,05 | -4,50 | 6,71E-06 | 6,93E-04 |
| PHF21A | 4095,53 | -0,14 | 0,03 | -4,50 | 6,81E-06 | 6,97E-04 |
| KIF1B | 2653,55 | -0,18 | 0,04 | -4,50 | 6,92E-06 | 7,01E-04 |
| PLAUR | 1476,27 | -0,18 | 0,04 | -4,48 | 7,53E-06 | 7,35E-04 |
| REPS2 | 3152,85 | -0,21 | 0,05 | -4,48 | 7,55E-06 | 7,35E-04 |
| NRBF2 | 1382,51 | -0,17 | 0,04 | -4,48 | 7,50E-06 | 7,35E-04 |
| SBNO1 | 3570,22 | -0,08 | 0,02 | -4,48 | 7,32E-06 | 7,35E-04 |
| SPAG9 | 9560,97 | -0,12 | 0,03 | -4,48 | 7,58E-06 | 7,35E-04 |
| NPEPPS | 3786,16 | -0,11 | 0,02 | -4,47 | 7,69E-06 | 7,36E-04 |
| USP12 | 3179,90 | 0,23 | 0,05 | 4,47 | 7,72E-06 | 7,36E-04 |
| NEDD9 | 5838,86 | -0,16 | 0,04 | -4,47 | 7,85E-06 | 7,41E-04 |
| HECW2 | 548,59 | -0,23 | 0,05 | -4,47 | 8,00E-06 | 7,49E-04 |
| MME | 15449,87 | -0,24 | 0,05 | -4,46 | 8,34E-06 | 7,63E-04 |
| NBR1 | 5130,61 | -0,09 | 0,02 | -4,46 | 8,24E-06 | 7,63E-04 |
| SENP2 | 1365,65 | -0,06 | 0,01 | -4,46 | 8,35E-06 | 7,63E-04 |
| HSD17B8 | 72,25 | 0,14 | 0,03 | 4,45 | 8,50E-06 | 7,70E-04 |
| MAPK14 | 11919,31 | -0,15 | 0,03 | -4,44 | 8,83E-06 | 7,82E-04 |
| ATP6V1B2 | 15620,14 | -0,14 | 0,03 | -4,44 | 8,88E-06 | 7,82E-04 |
| PPM1D | 815,50 | -0,08 | 0,02 | -4,44 | 8,91E-06 | 7,82E-04 |
| CDKL5 | 921,19 | -0,14 | 0,03 | -4,45 | 8,70E-06 | 7,82E-04 |
| TMEM14C | 422,07 | 0,16 | 0,04 | 4,44 | 9,14E-06 | 7,89E-04 |
| IRF2 | 2788,29 | -0,12 | 0,03 | -4,44 | 9,14E-06 | 7,89E-04 |
| PSMG3 | 275,74 | 0,11 | 0,02 | 4,43 | 9,22E-06 | 7,90E-04 |
| TOR1B | 1177,55 | -0,17 | 0,04 | -4,43 | 9,36E-06 | 7,96E-04 |
| MANSC1 | 563,23 | -0,22 | 0,05 | -4,43 | 9,55E-06 | 8,00E-04 |
| MEGF9 | 10864,11 | -0,17 | 0,04 | -4,43 | 9,55E-06 | 8,00E-04 |
| AHCTF1 | 7768,63 | -0,13 | 0,03 | -4,42 | 9,77E-06 | 8,11E-04 |
| IL6R | 11122,35 | -0,16 | 0,04 | -4,42 | 1,00E-05 | 8,25E-04 |
| SIPA1L2 | 1952,65 | -0,23 | 0,05 | -4,41 | 1,03E-05 | 8,41E-04 |
| C3orf62 | 1202,79 | -0,15 | 0,03 | -4,41 | 1,05E-05 | 8,51E-04 |
| CEP19 | 830,48 | -0,19 | 0,04 | -4,41 | 1,05E-05 | 8,51E-04 |

| | | | | | | |
|---|---|---|---|---|---|---|
| PYGL | 8204,84 | -0,19 | 0,04 | -4,40 | 1,08E-05 | 8,62E-04 |
| BAZ1A | 6621,35 | -0,14 | 0,03 | -4,39 | 1,13E-05 | 8,96E-04 |
| LINC02218 | 327,01 | -0,22 | 0,05 | -4,39 | 1,14E-05 | 8,99E-04 |
| CDK19 | 2437,53 | -0,13 | 0,03 | -4,38 | 1,19E-05 | 9,35E-04 |
| NIN | 15920,86 | -0,10 | 0,02 | -4,38 | 1,21E-05 | 9,44E-04 |
| ZNF438 | 1021,87 | -0,17 | 0,04 | -4,37 | 1,23E-05 | 9,50E-04 |
| MX2 | 6481,38 | -0,22 | 0,05 | -4,36 | 1,31E-05 | 1,00E-03 |
| MCL1 | 59363,61 | -0,13 | 0,03 | -4,36 | 1,31E-05 | 1,00E-03 |
| UBR2 | 14634,34 | -0,12 | 0,03 | -4,35 | 1,34E-05 | 1,01E-03 |
| MAP4K4 | 6815,34 | -0,13 | 0,03 | -4,35 | 1,34E-05 | 1,01E-03 |
| SLC22A15 | 2410,70 | -0,16 | 0,04 | -4,35 | 1,36E-05 | 1,01E-03 |
| CYREN | 873,97 | 0,05 | 0,01 | 4,35 | 1,37E-05 | 1,01E-03 |
| ABCG1 | 1140,64 | -0,22 | 0,05 | -4,34 | 1,45E-05 | 1,07E-03 |
| DOCK5 | 15847,77 | -0,19 | 0,04 | -4,34 | 1,46E-05 | 1,07E-03 |
| STX6 | 2546,70 | -0,09 | 0,02 | -4,33 | 1,50E-05 | 1,09E-03 |
| IL1R2 | 2508,64 | -0,22 | 0,05 | -4,32 | 1,58E-05 | 1,12E-03 |
| LTB4R | 1480,66 | -0,16 | 0,04 | -4,32 | 1,58E-05 | 1,12E-03 |
| RNF11 | 6609,97 | 0,29 | 0,07 | 4,32 | 1,55E-05 | 1,12E-03 |
| STAT3 | 7066,85 | -0,17 | 0,04 | -4,32 | 1,57E-05 | 1,12E-03 |
| DHX8 | 2891,88 | -0,09 | 0,02 | -4,32 | 1,57E-05 | 1,12E-03 |
| NT5C2 | 6912,58 | -0,14 | 0,03 | -4,31 | 1,61E-05 | 1,13E-03 |
| RBM23 | 4128,50 | -0,12 | 0,03 | -4,30 | 1,72E-05 | 1,14E-03 |
| TPST1 | 482,30 | -0,27 | 0,06 | -4,30 | 1,71E-05 | 1,14E-03 |
| SPEN | 6054,33 | -0,08 | 0,02 | -4,30 | 1,67E-05 | 1,14E-03 |
| NHLRC4 | 84,73 | 0,16 | 0,04 | 4,30 | 1,69E-05 | 1,14E-03 |
| PDK3 | 1946,24 | -0,12 | 0,03 | -4,30 | 1,72E-05 | 1,14E-03 |
| HAL | 2709,92 | -0,19 | 0,04 | -4,30 | 1,71E-05 | 1,14E-03 |
| SORL1 | 75720,41 | -0,18 | 0,04 | -4,31 | 1,67E-05 | 1,14E-03 |
| AVIL | 732,80 | -0,21 | 0,05 | -4,31 | 1,67E-05 | 1,14E-03 |
| SP110 | 5370,72 | -0,16 | 0,04 | -4,31 | 1,66E-05 | 1,14E-03 |
| NCF2 | 23904,89 | -0,15 | 0,03 | -4,29 | 1,75E-05 | 1,15E-03 |
| OGA | 17514,31 | -0,09 | 0,02 | -4,29 | 1,77E-05 | 1,16E-03 |
| BCL6 | 6006,36 | -0,23 | 0,05 | -4,29 | 1,79E-05 | 1,17E-03 |
| BTNL8 | 727,54 | -0,22 | 0,05 | -4,28 | 1,84E-05 | 1,19E-03 |
| CLIP1 | 2877,64 | -0,10 | 0,02 | -4,28 | 1,85E-05 | 1,19E-03 |
| AREL1 | 4487,67 | -0,12 | 0,03 | -4,28 | 1,90E-05 | 1,21E-03 |
| CPQ | 3971,69 | -0,15 | 0,03 | -4,28 | 1,90E-05 | 1,21E-03 |
| OSBPL2 | 3838,04 | -0,13 | 0,03 | -4,27 | 1,91E-05 | 1,21E-03 |
| MAPK1 | 12232,42 | -0,10 | 0,02 | -4,27 | 1,94E-05 | 1,21E-03 |
| ACOX1 | 5920,88 | -0,16 | 0,04 | -4,27 | 1,95E-05 | 1,21E-03 |
| BASP1 | 6520,53 | -0,22 | 0,05 | -4,27 | 1,93E-05 | 1,21E-03 |
| CD53 | 13283,58 | -0,08 | 0,02 | -4,26 | 2,01E-05 | 1,22E-03 |
| PGM1 | 814,92 | -0,10 | 0,02 | -4,26 | 2,02E-05 | 1,22E-03 |
| C9orf40 | 261,00 | 0,18 | 0,04 | 4,26 | 2,00E-05 | 1,22E-03 |
| AP1G1 | 4456,55 | -0,07 | 0,02 | -4,26 | 2,02E-05 | 1,22E-03 |
| TOR1AIP2 | 4017,74 | -0,07 | 0,02 | -4,26 | 2,03E-05 | 1,22E-03 |

| | | | | | | |
|---|---|---|---|---|---|---|
| PHTF1 | 1129,76 | -0,11 | 0,03 | -4,27 | 1,98E-05 | 1,22E-03 |
| RALGAPA2 | 5706,93 | -0,15 | 0,04 | -4,26 | 2,06E-05 | 1,23E-03 |
| SAP30L | 1918,45 | -0,07 | 0,02 | -4,25 | 2,10E-05 | 1,25E-03 |
| QPCT | 2504,52 | -0,21 | 0,05 | -4,25 | 2,17E-05 | 1,27E-03 |
| IFNAR1 | 5227,38 | -0,11 | 0,03 | -4,25 | 2,16E-05 | 1,27E-03 |
| GFUS | 751,65 | 0,24 | 0,06 | 4,24 | 2,21E-05 | 1,29E-03 |
| RPGR | 1456,70 | -0,14 | 0,03 | -4,24 | 2,28E-05 | 1,30E-03 |
| IGF2R | 34459,82 | -0,22 | 0,05 | -4,24 | 2,27E-05 | 1,30E-03 |
| RALB | 4092,96 | -0,15 | 0,04 | -4,24 | 2,26E-05 | 1,30E-03 |
| ABCA1 | 4718,66 | -0,22 | 0,05 | -4,24 | 2,24E-05 | 1,30E-03 |
| NFYA | 1906,33 | -0,07 | 0,02 | -4,24 | 2,25E-05 | 1,30E-03 |
| USP10 | 3430,56 | -0,12 | 0,03 | -4,23 | 2,34E-05 | 1,32E-03 |
| GMPR2 | 2011,17 | -0,10 | 0,02 | -4,23 | 2,35E-05 | 1,32E-03 |
| KLHL2 | 3228,54 | -0,14 | 0,03 | -4,23 | 2,38E-05 | 1,33E-03 |
| PPP1R12B | 5896,32 | -0,19 | 0,04 | -4,23 | 2,37E-05 | 1,33E-03 |
| USP9X | 7712,95 | -0,09 | 0,02 | -4,22 | 2,42E-05 | 1,34E-03 |
| EXTL3 | 2615,90 | -0,16 | 0,04 | -4,22 | 2,44E-05 | 1,35E-03 |
| BST1 | 1807,19 | -0,16 | 0,04 | -4,22 | 2,47E-05 | 1,36E-03 |
| MINPP1 | 291,25 | 0,16 | 0,04 | 4,20 | 2,64E-05 | 1,41E-03 |
| UROS | 669,95 | 0,07 | 0,02 | 4,21 | 2,60E-05 | 1,41E-03 |
| NUB1 | 2205,40 | -0,11 | 0,03 | -4,20 | 2,62E-05 | 1,41E-03 |
| JDP2 | 1025,09 | -0,14 | 0,03 | -4,20 | 2,64E-05 | 1,41E-03 |
| ZNF487 | 432,91 | -0,12 | 0,03 | -4,21 | 2,60E-05 | 1,41E-03 |
| JAK1 | 14237,90 | -0,10 | 0,02 | -4,20 | 2,63E-05 | 1,41E-03 |
| NIBAN1 | 22962,37 | -0,19 | 0,05 | -4,20 | 2,72E-05 | 1,44E-03 |
| ARL11 | 1822,34 | -0,13 | 0,03 | -4,19 | 2,82E-05 | 1,48E-03 |
| PTPRE | 9344,59 | -0,13 | 0,03 | -4,19 | 2,82E-05 | 1,48E-03 |
| GFER | 189,84 | 0,10 | 0,02 | 4,19 | 2,83E-05 | 1,48E-03 |
| SLC25A20 | 1005,73 | -0,14 | 0,03 | -4,18 | 2,87E-05 | 1,49E-03 |
| MBNL3 | 18248,81 | 0,28 | 0,07 | 4,18 | 2,87E-05 | 1,49E-03 |
| UIMC1 | 2802,55 | -0,10 | 0,02 | -4,18 | 2,89E-05 | 1,49E-03 |
| HLX | 544,29 | -0,19 | 0,04 | -4,18 | 2,93E-05 | 1,51E-03 |
| YWHAB | 8338,10 | -0,08 | 0,02 | -4,18 | 2,96E-05 | 1,51E-03 |
| LBR | 9389,83 | -0,14 | 0,03 | -4,17 | 3,00E-05 | 1,53E-03 |
| SIPA1L1 | 5586,13 | -0,16 | 0,04 | -4,17 | 3,02E-05 | 1,53E-03 |
| TADA2B | 1925,58 | -0,08 | 0,02 | -4,16 | 3,14E-05 | 1,57E-03 |
| IFRD1 | 6050,68 | -0,16 | 0,04 | -4,16 | 3,14E-05 | 1,57E-03 |
| GRHPR | 826,97 | 0,10 | 0,02 | 4,16 | 3,15E-05 | 1,57E-03 |
| SPATA13 | 3893,62 | -0,09 | 0,02 | -4,16 | 3,22E-05 | 1,59E-03 |
| CRK | 2471,70 | -0,09 | 0,02 | -4,16 | 3,22E-05 | 1,59E-03 |
| RPGRIP1 | 309,22 | -0,19 | 0,05 | -4,16 | 3,21E-05 | 1,59E-03 |
| PACSIN2 | 5697,73 | -0,15 | 0,04 | -4,15 | 3,27E-05 | 1,60E-03 |
| FOSL2 | 4660,97 | -0,19 | 0,05 | -4,15 | 3,36E-05 | 1,64E-03 |
| NCOA1 | 6115,14 | -0,12 | 0,03 | -4,15 | 3,40E-05 | 1,64E-03 |
| KIAA0232 | 6015,44 | -0,12 | 0,03 | -4,15 | 3,39E-05 | 1,64E-03 |
| WDFY3 | 11236,89 | -0,19 | 0,04 | -4,14 | 3,40E-05 | 1,64E-03 |

| | | | | | | |
|---|---|---|---|---|---|---|
| TRIM25 | 7374,08 | -0,19 | 0,05 | -4,14 | 3,45E-05 | 1,66E-03 |
| TP53BP2 | 1365,62 | -0,05 | 0,01 | -4,14 | 3,49E-05 | 1,67E-03 |
| ALOX5AP | 3483,04 | -0,18 | 0,04 | -4,13 | 3,60E-05 | 1,71E-03 |
| CXCR1 | 8442,88 | -0,19 | 0,05 | -4,13 | 3,62E-05 | 1,72E-03 |
| KIF27 | 1242,86 | -0,17 | 0,04 | -4,13 | 3,64E-05 | 1,72E-03 |
| IFNAR2 | 2438,98 | -0,10 | 0,02 | -4,13 | 3,66E-05 | 1,72E-03 |
| MPZL3 | 4992,12 | -0,16 | 0,04 | -4,12 | 3,73E-05 | 1,75E-03 |
| HCLS1 | 18950,10 | -0,12 | 0,03 | -4,12 | 3,83E-05 | 1,76E-03 |
| AIDA | 1419,23 | 0,14 | 0,03 | 4,12 | 3,82E-05 | 1,76E-03 |
| GNAI3 | 8654,79 | -0,10 | 0,02 | -4,12 | 3,80E-05 | 1,76E-03 |
| SEL1L | 5759,88 | -0,09 | 0,02 | -4,12 | 3,80E-05 | 1,76E-03 |
| ST20 | 857,96 | -0,17 | 0,04 | -4,12 | 3,85E-05 | 1,77E-03 |
| MBP | 9811,42 | -0,12 | 0,03 | -4,12 | 3,87E-05 | 1,77E-03 |
| HLA-E | 68127,11 | -0,11 | 0,03 | -4,11 | 3,92E-05 | 1,78E-03 |
| ELF2 | 5373,38 | -0,11 | 0,03 | -4,11 | 3,92E-05 | 1,78E-03 |
| AGO4 | 9579,70 | -0,13 | 0,03 | -4,11 | 3,96E-05 | 1,78E-03 |
| RTN3 | 3355,21 | -0,15 | 0,04 | -4,11 | 3,97E-05 | 1,78E-03 |
| IFRD2 | 397,94 | 0,14 | 0,03 | 4,11 | 3,99E-05 | 1,78E-03 |
| COA4 | 480,69 | 0,08 | 0,02 | 4,11 | 4,02E-05 | 1,79E-03 |
| TRIM38 | 5725,97 | -0,10 | 0,02 | -4,10 | 4,07E-05 | 1,81E-03 |
| PRDM10 | 744,59 | -0,07 | 0,02 | -4,10 | 4,11E-05 | 1,82E-03 |
| FPR1 | 12828,83 | -0,18 | 0,04 | -4,10 | 4,14E-05 | 1,82E-03 |
| ARFGAP3 | 1503,69 | -0,09 | 0,02 | -4,10 | 4,13E-05 | 1,82E-03 |
| PITPNA | 2054,35 | -0,14 | 0,03 | -4,10 | 4,19E-05 | 1,83E-03 |
| N4BP1 | 3604,88 | -0,11 | 0,03 | -4,09 | 4,24E-05 | 1,84E-03 |
| ZBTB34 | 3748,56 | -0,12 | 0,03 | -4,09 | 4,28E-05 | 1,86E-03 |
| TMEM43 | 2870,92 | -0,11 | 0,03 | -4,09 | 4,39E-05 | 1,89E-03 |
| SRSF8 | 1578,21 | 0,11 | 0,03 | 4,08 | 4,43E-05 | 1,91E-03 |
| SNRK | 8528,28 | -0,10 | 0,02 | -4,08 | 4,52E-05 | 1,94E-03 |
| SQLE | 212,96 | 0,13 | 0,03 | 4,08 | 4,54E-05 | 1,94E-03 |
| MTMR10 | 4934,16 | -0,11 | 0,03 | -4,08 | 4,56E-05 | 1,94E-03 |
| CDS2 | 5388,55 | -0,11 | 0,03 | -4,08 | 4,57E-05 | 1,94E-03 |
| NCOA2 | 8105,35 | -0,10 | 0,02 | -4,07 | 4,71E-05 | 1,99E-03 |
| SYAP1 | 1300,84 | -0,08 | 0,02 | -4,06 | 4,88E-05 | 2,05E-03 |
| C3orf18 | 318,02 | 0,10 | 0,02 | 4,06 | 4,93E-05 | 2,06E-03 |
| TSEN34 | 2132,14 | -0,14 | 0,03 | -4,06 | 4,96E-05 | 2,07E-03 |
| CASP4 | 7536,88 | -0,15 | 0,04 | -4,06 | 4,98E-05 | 2,07E-03 |
| NUP214 | 3409,72 | -0,08 | 0,02 | -4,05 | 5,09E-05 | 2,09E-03 |
| MCRIP2 | 190,65 | 0,14 | 0,03 | 4,05 | 5,08E-05 | 2,09E-03 |
| ZNFX1 | 6114,83 | -0,13 | 0,03 | -4,05 | 5,06E-05 | 2,09E-03 |
| TBC1D30 | 230,44 | -0,18 | 0,04 | -4,05 | 5,16E-05 | 2,11E-03 |
| TREM1 | 6352,22 | -0,18 | 0,04 | -4,05 | 5,23E-05 | 2,12E-03 |
| ALPK1 | 6270,39 | -0,17 | 0,04 | -4,05 | 5,21E-05 | 2,12E-03 |
| RASSF2 | 22691,58 | -0,14 | 0,03 | -4,04 | 5,29E-05 | 2,13E-03 |
| FLI1 | 7233,78 | -0,07 | 0,02 | -4,04 | 5,26E-05 | 2,13E-03 |
| MYLIP | 2901,05 | -0,11 | 0,03 | -4,04 | 5,28E-05 | 2,13E-03 |

| | | | | | | |
|---|---|---|---|---|---|---|
| CFAP58 | 147,74 | -0,21 | 0,05 | -4,04 | 5,34E-05 | 2,14E-03 |
| ANXA3 | 1056,85 | -0,28 | 0,07 | -4,04 | 5,44E-05 | 2,17E-03 |
| KIDINS220 | 7582,57 | -0,09 | 0,02 | -4,03 | 5,59E-05 | 2,22E-03 |
| USP4 | 4452,13 | -0,12 | 0,03 | -4,03 | 5,60E-05 | 2,22E-03 |
| TLCD4 | 540,48 | 0,26 | 0,07 | 4,03 | 5,63E-05 | 2,22E-03 |
| PIK3CG | 4171,39 | -0,08 | 0,02 | -4,03 | 5,65E-05 | 2,22E-03 |
| TMBIM6 | 12654,37 | -0,09 | 0,02 | -4,03 | 5,67E-05 | 2,22E-03 |
| VNN2 | 19205,95 | -0,21 | 0,05 | -4,02 | 5,78E-05 | 2,25E-03 |
| SESN3 | 11452,06 | 0,17 | 0,04 | 4,02 | 5,77E-05 | 2,25E-03 |
| TMX4 | 6801,44 | -0,11 | 0,03 | -4,02 | 5,82E-05 | 2,26E-03 |
| SLMAP | 3061,48 | -0,08 | 0,02 | -4,02 | 5,85E-05 | 2,26E-03 |
| TNFSF14 | 2177,94 | -0,13 | 0,03 | -4,01 | 6,13E-05 | 2,35E-03 |
| HECTD2 | 142,13 | 0,24 | 0,06 | 4,01 | 6,14E-05 | 2,35E-03 |
| BRD8 | 993,07 | -0,09 | 0,02 | -4,01 | 6,12E-05 | 2,35E-03 |
| KDM3A | 4536,91 | -0,07 | 0,02 | -4,00 | 6,22E-05 | 2,37E-03 |
| TNFRSF10C | 5514,01 | -0,18 | 0,04 | -4,00 | 6,36E-05 | 2,41E-03 |
| USP32 | 8364,83 | -0,12 | 0,03 | -3,99 | 6,47E-05 | 2,42E-03 |
| LRRC4 | 1292,32 | -0,18 | 0,05 | -3,99 | 6,54E-05 | 2,42E-03 |
| ITPRIP | 3336,07 | -0,16 | 0,04 | -4,00 | 6,40E-05 | 2,42E-03 |
| DNAJB6 | 4149,43 | -0,11 | 0,03 | -3,99 | 6,49E-05 | 2,42E-03 |
| CELF2 | 15442,51 | -0,10 | 0,02 | -3,99 | 6,51E-05 | 2,42E-03 |
| GSTZ1 | 306,71 | 0,09 | 0,02 | 4,00 | 6,45E-05 | 2,42E-03 |
| NHS | 420,15 | -0,17 | 0,04 | -4,00 | 6,43E-05 | 2,42E-03 |
| JAZF1 | 1872,91 | 0,19 | 0,05 | 3,99 | 6,54E-05 | 2,42E-03 |
| DHTKD1 | 1496,36 | -0,09 | 0,02 | -3,99 | 6,57E-05 | 2,42E-03 |
| NPL | 4501,03 | -0,16 | 0,04 | -3,99 | 6,67E-05 | 2,45E-03 |
| TGFBR2 | 10673,18 | -0,08 | 0,02 | -3,99 | 6,70E-05 | 2,45E-03 |
| PLCG2 | 5658,56 | -0,13 | 0,03 | -3,99 | 6,73E-05 | 2,45E-03 |
| OR2W3 | 423,03 | 0,35 | 0,09 | 3,98 | 6,86E-05 | 2,49E-03 |
| ELAC1 | 206,10 | 0,10 | 0,02 | 3,98 | 6,87E-05 | 2,49E-03 |
| PXK | 1414,96 | -0,11 | 0,03 | -3,98 | 6,95E-05 | 2,51E-03 |
| WIPF1 | 12524,56 | -0,10 | 0,02 | -3,98 | 6,98E-05 | 2,51E-03 |
| CSRNP1 | 953,47 | -0,16 | 0,04 | -3,97 | 7,14E-05 | 2,56E-03 |
| F5 | 1695,19 | -0,18 | 0,05 | -3,97 | 7,24E-05 | 2,58E-03 |
| DAPK2 | 2449,89 | -0,19 | 0,05 | -3,97 | 7,22E-05 | 2,58E-03 |
| UBN1 | 4363,54 | -0,20 | 0,05 | -3,97 | 7,33E-05 | 2,61E-03 |
| TNPO3 | 3020,77 | -0,08 | 0,02 | -3,96 | 7,39E-05 | 2,61E-03 |
| ATP6V1A | 5458,67 | -0,13 | 0,03 | -3,96 | 7,37E-05 | 2,61E-03 |
| RGS2 | 14252,20 | -0,20 | 0,05 | -3,96 | 7,45E-05 | 2,62E-03 |
| SULF2 | 4049,89 | -0,16 | 0,04 | -3,96 | 7,49E-05 | 2,63E-03 |
| RTF2 | 4332,95 | -0,11 | 0,03 | -3,96 | 7,55E-05 | 2,64E-03 |
| ADAR | 12763,84 | -0,17 | 0,04 | -3,95 | 7,77E-05 | 2,70E-03 |
| TMEM138 | 616,06 | 0,08 | 0,02 | 3,95 | 7,76E-05 | 2,70E-03 |
| RBM5 | 9728,05 | -0,06 | 0,02 | -3,95 | 7,92E-05 | 2,74E-03 |
| SIGLEC5 | 84,93 | -0,24 | 0,06 | -3,95 | 7,95E-05 | 2,75E-03 |
| AATF | 1611,05 | -0,08 | 0,02 | -3,94 | 8,03E-05 | 2,76E-03 |

| | | | | | | |
|---|---|---|---|---|---|---|
| PTRHD1 | 235,14 | 0,17 | 0,04 | 3,94 | 8,13E-05 | 2,78E-03 |
| ARCN1 | 2317,13 | -0,07 | 0,02 | -3,94 | 8,17E-05 | 2,78E-03 |
| ABCB10 | 1453,81 | 0,13 | 0,03 | 3,94 | 8,11E-05 | 2,78E-03 |
| YOD1 | 11649,01 | 0,28 | 0,07 | 3,94 | 8,15E-05 | 2,78E-03 |
| TBC1D14 | 5384,66 | -0,13 | 0,03 | -3,94 | 8,21E-05 | 2,78E-03 |
| PHF20 | 2931,05 | -0,06 | 0,02 | -3,93 | 8,34E-05 | 2,82E-03 |
| PARP9 | 5179,31 | -0,19 | 0,05 | -3,93 | 8,49E-05 | 2,85E-03 |
| MSL2 | 4411,76 | -0,06 | 0,01 | -3,93 | 8,50E-05 | 2,85E-03 |
| SDCBP | 13555,63 | -0,15 | 0,04 | -3,93 | 8,50E-05 | 2,85E-03 |
| TREML2 | 1641,05 | -0,15 | 0,04 | -3,93 | 8,58E-05 | 2,86E-03 |
| PELI2 | 3890,04 | -0,12 | 0,03 | -3,92 | 8,87E-05 | 2,95E-03 |
| ARHGAP26 | 14600,04 | -0,16 | 0,04 | -3,91 | 9,14E-05 | 3,03E-03 |
| SRPK2 | 5022,20 | -0,10 | 0,03 | -3,91 | 9,26E-05 | 3,04E-03 |
| JAML | 25945,77 | -0,13 | 0,03 | -3,91 | 9,29E-05 | 3,04E-03 |
| TLR6 | 5538,15 | -0,14 | 0,03 | -3,91 | 9,22E-05 | 3,04E-03 |
| RNF114 | 2419,31 | -0,07 | 0,02 | -3,91 | 9,19E-05 | 3,04E-03 |
| LYN | 10914,39 | -0,15 | 0,04 | -3,91 | 9,25E-05 | 3,04E-03 |
| GYPA | 322,66 | 0,32 | 0,08 | 3,90 | 9,42E-05 | 3,06E-03 |
| CDC42 | 795,33 | -0,07 | 0,02 | -3,90 | 9,44E-05 | 3,06E-03 |
| TUT7 | 13001,25 | -0,12 | 0,03 | -3,90 | 9,44E-05 | 3,06E-03 |
| SLC18B1 | 688,89 | 0,11 | 0,03 | 3,90 | 9,50E-05 | 3,07E-03 |
| RAP1GAP | 170,56 | 0,59 | 0,15 | 3,90 | 9,59E-05 | 3,09E-03 |
| AKAP13 | 21707,80 | -0,11 | 0,03 | -3,90 | 9,68E-05 | 3,11E-03 |
| ZNF586 | 1800,03 | -0,08 | 0,02 | -3,90 | 9,77E-05 | 3,13E-03 |
| RBM47 | 4835,86 | -0,16 | 0,04 | -3,89 | 9,84E-05 | 3,14E-03 |
| CDK6 | 2357,15 | 0,10 | 0,03 | 3,89 | 9,90E-05 | 3,15E-03 |
| YKT6 | 1312,40 | -0,06 | 0,01 | -3,89 | 1,02E-04 | 3,18E-03 |
| UPF2 | 3015,52 | -0,09 | 0,02 | -3,89 | 1,02E-04 | 3,18E-03 |
| FBXO38 | 4933,36 | -0,10 | 0,02 | -3,89 | 1,02E-04 | 3,18E-03 |
| DCK | 2131,93 | 0,14 | 0,03 | 3,89 | 1,02E-04 | 3,18E-03 |
| PCYT1A | 2950,86 | -0,09 | 0,02 | -3,89 | 1,02E-04 | 3,18E-03 |
| CD55 | 6427,19 | -0,15 | 0,04 | -3,89 | 1,01E-04 | 3,18E-03 |
| ALDH9A1 | 1394,71 | -0,08 | 0,02 | -3,89 | 1,02E-04 | 3,18E-03 |
| PNO1 | 302,12 | 0,08 | 0,02 | 3,88 | 1,02E-04 | 3,19E-03 |
| YIF1A | 335,46 | 0,08 | 0,02 | 3,88 | 1,03E-04 | 3,19E-03 |
| RNF130 | 6257,44 | -0,11 | 0,03 | -3,88 | 1,03E-04 | 3,19E-03 |
| IRAG1 | 1760,94 | -0,18 | 0,05 | -3,88 | 1,04E-04 | 3,22E-03 |
| TNFAIP6 | 742,66 | -0,31 | 0,08 | -3,88 | 1,06E-04 | 3,25E-03 |
| HSPA6 | 3945,16 | -0,18 | 0,05 | -3,88 | 1,06E-04 | 3,25E-03 |
| UBXN7 | 4077,58 | -0,08 | 0,02 | -3,87 | 1,07E-04 | 3,25E-03 |
| KDM5A | 13266,63 | -0,08 | 0,02 | -3,88 | 1,06E-04 | 3,25E-03 |
| WDR47 | 801,05 | -0,08 | 0,02 | -3,88 | 1,06E-04 | 3,25E-03 |
| CPT1A | 1337,37 | -0,18 | 0,05 | -3,87 | 1,08E-04 | 3,26E-03 |
| DENND5A | 6470,82 | -0,14 | 0,04 | -3,87 | 1,08E-04 | 3,26E-03 |
| DDX60L | 11151,31 | -0,19 | 0,05 | -3,87 | 1,08E-04 | 3,27E-03 |
| HYCC2 | 9002,98 | -0,15 | 0,04 | -3,87 | 1,11E-04 | 3,34E-03 |

| | | | | | | |
|---|---|---|---|---|---|---|
| APRT | 496,17 | 0,09 | 0,02 | 3,86 | 1,11E-04 | 3,34E-03 |
| FUOM | 231,92 | 0,11 | 0,03 | 3,86 | 1,13E-04 | 3,37E-03 |
| FAU | 4840,82 | 0,17 | 0,04 | 3,86 | 1,13E-04 | 3,38E-03 |
| ABHD4 | 619,22 | -0,12 | 0,03 | -3,86 | 1,14E-04 | 3,38E-03 |
| ENTPD1 | 5802,29 | -0,14 | 0,04 | -3,85 | 1,17E-04 | 3,46E-03 |
| PPP1R14B | 193,75 | 0,10 | 0,03 | 3,85 | 1,19E-04 | 3,52E-03 |
| PADI4 | 1694,75 | -0,23 | 0,06 | -3,85 | 1,19E-04 | 3,52E-03 |
| DAZAP2 | 9317,30 | -0,10 | 0,03 | -3,84 | 1,21E-04 | 3,57E-03 |
| PROK2 | 2818,07 | -0,21 | 0,05 | -3,84 | 1,23E-04 | 3,62E-03 |
| APOL2 | 4280,41 | -0,14 | 0,04 | -3,84 | 1,24E-04 | 3,63E-03 |
| MGAM2 | 574,87 | -0,25 | 0,06 | -3,83 | 1,26E-04 | 3,67E-03 |
| HERC3 | 6019,40 | -0,10 | 0,03 | -3,83 | 1,26E-04 | 3,67E-03 |
| TRIM21 | 1897,66 | -0,12 | 0,03 | -3,83 | 1,29E-04 | 3,73E-03 |
| HSPA1A | 1459,65 | -0,17 | 0,04 | -3,83 | 1,29E-04 | 3,73E-03 |
| DGAT2 | 3606,17 | -0,17 | 0,04 | -3,82 | 1,32E-04 | 3,82E-03 |
| APBB1IP | 4140,60 | -0,13 | 0,03 | -3,82 | 1,32E-04 | 3,82E-03 |
| GNS | 5803,42 | -0,12 | 0,03 | -3,82 | 1,33E-04 | 3,82E-03 |
| JPT1 | 1672,11 | -0,12 | 0,03 | -3,82 | 1,33E-04 | 3,83E-03 |
| EIF2AK2 | 3155,07 | -0,23 | 0,06 | -3,82 | 1,35E-04 | 3,86E-03 |
| ANTXR2 | 7689,94 | -0,11 | 0,03 | -3,82 | 1,35E-04 | 3,86E-03 |
| HDHD5 | 503,79 | 0,07 | 0,02 | 3,81 | 1,37E-04 | 3,90E-03 |
| TSPAN17 | 531,04 | 0,09 | 0,02 | 3,81 | 1,39E-04 | 3,94E-03 |
| APPL2 | 1769,19 | -0,09 | 0,02 | -3,81 | 1,40E-04 | 3,95E-03 |
| ALKBH7 | 602,82 | 0,12 | 0,03 | 3,81 | 1,40E-04 | 3,95E-03 |
| LETM2 | 235,45 | -0,14 | 0,04 | -3,81 | 1,39E-04 | 3,95E-03 |
| HSD17B11 | 5783,33 | -0,12 | 0,03 | -3,81 | 1,40E-04 | 3,95E-03 |
| MMP9 | 858,09 | -0,31 | 0,08 | -3,81 | 1,41E-04 | 3,95E-03 |
| ZBP1 | 1807,72 | -0,19 | 0,05 | -3,81 | 1,41E-04 | 3,95E-03 |
| GPR27 | 567,07 | -0,17 | 0,05 | -3,80 | 1,43E-04 | 3,99E-03 |
| TBC1D12 | 301,67 | 0,14 | 0,04 | 3,80 | 1,44E-04 | 4,00E-03 |
| RPL18A | 9398,56 | 0,13 | 0,03 | 3,80 | 1,45E-04 | 4,01E-03 |
| IRF1 | 17710,14 | -0,12 | 0,03 | -3,80 | 1,45E-04 | 4,01E-03 |
| CYRIB | 9044,92 | -0,10 | 0,03 | -3,80 | 1,45E-04 | 4,01E-03 |
| PGM2L1 | 630,47 | 0,14 | 0,04 | 3,80 | 1,46E-04 | 4,03E-03 |
| PPP1R21 | 2255,62 | -0,08 | 0,02 | -3,80 | 1,47E-04 | 4,05E-03 |
| EGLN1 | 6137,29 | -0,13 | 0,03 | -3,79 | 1,48E-04 | 4,05E-03 |
| NENF | 133,51 | 0,10 | 0,03 | 3,79 | 1,49E-04 | 4,09E-03 |
| CDC42SE1 | 15793,65 | -0,08 | 0,02 | -3,79 | 1,51E-04 | 4,12E-03 |
| AVL9 | 4526,16 | -0,06 | 0,02 | -3,79 | 1,52E-04 | 4,12E-03 |
| PYM1 | 411,14 | 0,06 | 0,02 | 3,79 | 1,52E-04 | 4,13E-03 |
| NUDT4 | 2847,79 | 0,17 | 0,04 | 3,78 | 1,56E-04 | 4,22E-03 |
| SOS2 | 4458,45 | -0,12 | 0,03 | -3,78 | 1,57E-04 | 4,25E-03 |
| NUMB | 4509,51 | -0,14 | 0,04 | -3,78 | 1,59E-04 | 4,28E-03 |
| DPP7 | 734,35 | 0,11 | 0,03 | 3,78 | 1,59E-04 | 4,28E-03 |
| IL1B | 1032,61 | -0,16 | 0,04 | -3,77 | 1,60E-04 | 4,30E-03 |
| JARID2 | 3994,09 | -0,13 | 0,03 | -3,77 | 1,61E-04 | 4,31E-03 |

| | | | | | | |
|---|---|---|---|---|---|---|
| TXNIP | 76702,59 | -0,10 | 0,03 | -3,77 | 1,61E-04 | 4,31E-03 |
| TMEM185B | 547,21 | -0,10 | 0,03 | -3,77 | 1,63E-04 | 4,33E-03 |
| RPL15 | 8571,39 | 0,14 | 0,04 | 3,77 | 1,64E-04 | 4,34E-03 |
| ZNF552 | 794,15 | -0,09 | 0,02 | -3,77 | 1,65E-04 | 4,36E-03 |
| RGCC | 968,33 | 0,15 | 0,04 | 3,77 | 1,65E-04 | 4,36E-03 |
| CYRIA | 3777,69 | -0,10 | 0,03 | -3,77 | 1,66E-04 | 4,38E-03 |
| RPS2 | 17319,70 | 0,11 | 0,03 | 3,76 | 1,67E-04 | 4,40E-03 |
| XPO6 | 21640,42 | -0,14 | 0,04 | -3,76 | 1,68E-04 | 4,41E-03 |
| CHSY1 | 2585,41 | -0,13 | 0,04 | -3,76 | 1,69E-04 | 4,42E-03 |
| ASPRV1 | 302,09 | -0,24 | 0,06 | -3,76 | 1,70E-04 | 4,44E-03 |
| BMAL1 | 2666,46 | -0,09 | 0,02 | -3,76 | 1,71E-04 | 4,46E-03 |
| PAK1 | 3375,41 | -0,13 | 0,03 | -3,75 | 1,74E-04 | 4,52E-03 |
| RNF185 | 851,87 | -0,08 | 0,02 | -3,75 | 1,74E-04 | 4,52E-03 |
| ANP32A | 5716,32 | -0,09 | 0,02 | -3,75 | 1,77E-04 | 4,57E-03 |
| CMTM1 | 174,60 | -0,15 | 0,04 | -3,75 | 1,78E-04 | 4,59E-03 |
| IL18R1 | 1537,91 | -0,14 | 0,04 | -3,75 | 1,79E-04 | 4,62E-03 |
| NUP98 | 6711,96 | -0,09 | 0,02 | -3,74 | 1,81E-04 | 4,65E-03 |
| SELL | 20575,26 | -0,14 | 0,04 | -3,74 | 1,84E-04 | 4,71E-03 |
| MYOM1 | 236,01 | -0,15 | 0,04 | -3,73 | 1,88E-04 | 4,82E-03 |
| TMEM42 | 395,98 | 0,09 | 0,02 | 3,73 | 1,91E-04 | 4,85E-03 |
| AIRIM | 307,02 | 0,07 | 0,02 | 3,73 | 1,91E-04 | 4,85E-03 |
| GBP2 | 10539,47 | -0,15 | 0,04 | -3,73 | 1,91E-04 | 4,85E-03 |
| MAP2K4 | 2836,64 | -0,11 | 0,03 | -3,73 | 1,92E-04 | 4,88E-03 |
| NOP16 | 130,35 | 0,08 | 0,02 | 3,73 | 1,93E-04 | 4,89E-03 |
| SLC6A6 | 15040,44 | -0,15 | 0,04 | -3,73 | 1,95E-04 | 4,90E-03 |
| ERGIC1 | 5462,63 | -0,14 | 0,04 | -3,73 | 1,94E-04 | 4,90E-03 |
| NUP153 | 4219,21 | -0,07 | 0,02 | -3,72 | 1,98E-04 | 4,92E-03 |
| MYO5A | 5280,75 | -0,08 | 0,02 | -3,72 | 1,98E-04 | 4,92E-03 |
| PARP4 | 5217,14 | -0,09 | 0,02 | -3,72 | 1,98E-04 | 4,92E-03 |
| TLR4 | 15275,66 | -0,15 | 0,04 | -3,72 | 1,98E-04 | 4,92E-03 |
| FRY | 9177,76 | -0,13 | 0,04 | -3,72 | 1,97E-04 | 4,92E-03 |
| NFYC | 1679,26 | -0,10 | 0,03 | -3,72 | 1,97E-04 | 4,92E-03 |
| TM6SF1 | 1399,04 | -0,13 | 0,03 | -3,72 | 1,99E-04 | 4,92E-03 |
| TXNRD1 | 4560,25 | -0,11 | 0,03 | -3,72 | 2,01E-04 | 4,96E-03 |
| TMOD3 | 4520,79 | -0,06 | 0,02 | -3,72 | 2,01E-04 | 4,96E-03 |
| PPP4R1 | 7283,00 | -0,13 | 0,03 | -3,72 | 2,02E-04 | 4,97E-03 |
| ETHE1 | 194,99 | 0,08 | 0,02 | 3,71 | 2,04E-04 | 4,97E-03 |
| VAV1 | 4823,09 | -0,10 | 0,03 | -3,71 | 2,04E-04 | 4,97E-03 |
| MSMO1 | 462,71 | 0,13 | 0,04 | 3,71 | 2,03E-04 | 4,97E-03 |
| GLT1D1 | 3168,46 | -0,17 | 0,05 | -3,72 | 2,03E-04 | 4,97E-03 |
| CASS4 | 1619,19 | -0,14 | 0,04 | -3,71 | 2,06E-04 | 4,98E-03 |
| SLC26A8 | 224,46 | -0,26 | 0,07 | -3,71 | 2,05E-04 | 4,98E-03 |
| GSK3B | 3021,77 | -0,09 | 0,03 | -3,71 | 2,05E-04 | 4,98E-03 |
| TBC1D1 | 4692,98 | -0,11 | 0,03 | -3,71 | 2,08E-04 | 5,01E-03 |
| NAXD | 736,97 | 0,07 | 0,02 | 3,71 | 2,08E-04 | 5,01E-03 |
| SETX | 18915,29 | -0,09 | 0,02 | -3,71 | 2,08E-04 | 5,01E-03 |

| | | | | | | |
|---|---|---|---|---|---|---|
| IL1RN | 622,27 | -0,19 | 0,05 | -3,71 | 2,11E-04 | 5,04E-03 |
| SLCO4C1 | 1160,89 | -0,14 | 0,04 | -3,71 | 2,10E-04 | 5,04E-03 |
| MKNK1 | 2585,76 | -0,13 | 0,03 | -3,71 | 2,11E-04 | 5,04E-03 |
| RNASET2 | 9251,70 | -0,14 | 0,04 | -3,71 | 2,10E-04 | 5,04E-03 |
| FBXL20 | 2404,13 | -0,12 | 0,03 | -3,70 | 2,12E-04 | 5,05E-03 |
| FBXW2 | 4243,18 | -0,06 | 0,02 | -3,70 | 2,15E-04 | 5,10E-03 |
| TBXAS1 | 7442,34 | -0,13 | 0,04 | -3,70 | 2,15E-04 | 5,10E-03 |
| ALOX5 | 6113,47 | -0,14 | 0,04 | -3,70 | 2,17E-04 | 5,12E-03 |
| RRAGD | 1073,90 | -0,13 | 0,03 | -3,70 | 2,17E-04 | 5,12E-03 |
| SNX27 | 3810,42 | -0,15 | 0,04 | -3,70 | 2,19E-04 | 5,16E-03 |
| ZC3HAV1 | 7161,37 | -0,10 | 0,03 | -3,69 | 2,20E-04 | 5,18E-03 |
| DOCK4 | 1507,44 | -0,22 | 0,06 | -3,69 | 2,22E-04 | 5,22E-03 |
| CHST11 | 5080,61 | -0,11 | 0,03 | -3,69 | 2,25E-04 | 5,26E-03 |
| RNF14 | 2381,90 | 0,15 | 0,04 | 3,69 | 2,29E-04 | 5,27E-03 |
| TMEM209 | 351,87 | 0,09 | 0,02 | 3,69 | 2,28E-04 | 5,27E-03 |
| OSTF1 | 6181,81 | -0,11 | 0,03 | -3,69 | 2,26E-04 | 5,27E-03 |
| NDRG3 | 722,45 | -0,07 | 0,02 | -3,69 | 2,28E-04 | 5,27E-03 |
| TFDP1 | 1685,92 | 0,18 | 0,05 | 3,69 | 2,28E-04 | 5,27E-03 |
| JMJD4 | 566,56 | 0,09 | 0,02 | 3,69 | 2,29E-04 | 5,27E-03 |
| FNDC3B | 6404,42 | -0,12 | 0,03 | -3,68 | 2,29E-04 | 5,27E-03 |
| CIR1 | 1568,13 | -0,11 | 0,03 | -3,69 | 2,26E-04 | 5,27E-03 |
| THOC5 | 1741,68 | -0,13 | 0,03 | -3,68 | 2,30E-04 | 5,29E-03 |
| DICER1 | 11469,88 | -0,10 | 0,03 | -3,68 | 2,31E-04 | 5,30E-03 |
| RC3H1 | 8960,86 | -0,08 | 0,02 | -3,68 | 2,33E-04 | 5,33E-03 |
| KLF6 | 8236,52 | -0,09 | 0,02 | -3,68 | 2,34E-04 | 5,34E-03 |
| CYP4F3 | 6201,48 | -0,20 | 0,05 | -3,68 | 2,36E-04 | 5,37E-03 |
| CAB39 | 7496,16 | -0,09 | 0,02 | -3,68 | 2,37E-04 | 5,38E-03 |
| UBE2Q1 | 1252,73 | -0,06 | 0,02 | -3,68 | 2,38E-04 | 5,39E-03 |
| SEC24D | 1118,93 | -0,08 | 0,02 | -3,67 | 2,38E-04 | 5,40E-03 |
| SP100 | 9583,23 | -0,10 | 0,03 | -3,67 | 2,39E-04 | 5,40E-03 |
| GK | 5728,96 | -0,16 | 0,04 | -3,67 | 2,41E-04 | 5,40E-03 |
| JMJD1C | 13616,20 | -0,11 | 0,03 | -3,67 | 2,41E-04 | 5,40E-03 |
| POLD3 | 774,09 | -0,08 | 0,02 | -3,67 | 2,41E-04 | 5,40E-03 |
| RPS6KA5 | 5846,31 | -0,11 | 0,03 | -3,67 | 2,40E-04 | 5,40E-03 |
| XK | 1834,95 | 0,28 | 0,08 | 3,67 | 2,43E-04 | 5,43E-03 |
| CHST15 | 10089,86 | -0,16 | 0,04 | -3,67 | 2,45E-04 | 5,47E-03 |
| NDUFAF8 | 224,16 | 0,12 | 0,03 | 3,66 | 2,47E-04 | 5,51E-03 |
| ATG14 | 1021,74 | 0,05 | 0,01 | 3,66 | 2,48E-04 | 5,52E-03 |
| KPNA1 | 2828,17 | -0,07 | 0,02 | -3,66 | 2,50E-04 | 5,55E-03 |
| SIAH2 | 3483,80 | 0,24 | 0,07 | 3,66 | 2,51E-04 | 5,56E-03 |
| SAFB | 2722,35 | -0,07 | 0,02 | -3,65 | 2,62E-04 | 5,78E-03 |
| TAOK3 | 4026,79 | -0,06 | 0,02 | -3,65 | 2,63E-04 | 5,80E-03 |
| FLOT1 | 2358,92 | -0,12 | 0,03 | -3,65 | 2,65E-04 | 5,83E-03 |
| ARHGDIB | 9291,99 | -0,08 | 0,02 | -3,65 | 2,66E-04 | 5,85E-03 |
| LNPK | 1536,57 | 0,12 | 0,03 | 3,64 | 2,69E-04 | 5,89E-03 |
| COX18 | 887,62 | 0,08 | 0,02 | 3,64 | 2,71E-04 | 5,93E-03 |

| | | | | | | |
|---|---|---|---|---|---|---|
| TPD52L2 | 3101,08 | -0,11 | 0,03 | -3,64 | 2,73E-04 | 5,96E-03 |
| APLP2 | 6749,30 | -0,11 | 0,03 | -3,64 | 2,76E-04 | 6,01E-03 |
| EPHB1 | 3735,83 | -0,16 | 0,04 | -3,64 | 2,76E-04 | 6,01E-03 |
| ITM2B | 27947,88 | -0,11 | 0,03 | -3,63 | 2,81E-04 | 6,10E-03 |
| CFAP92 | 395,33 | -0,20 | 0,05 | -3,63 | 2,82E-04 | 6,10E-03 |
| ATP5MC1 | 305,16 | 0,08 | 0,02 | 3,63 | 2,86E-04 | 6,13E-03 |
| PRKCB | 11196,36 | -0,09 | 0,03 | -3,63 | 2,86E-04 | 6,13E-03 |
| TDP2 | 2858,93 | -0,11 | 0,03 | -3,63 | 2,84E-04 | 6,13E-03 |
| PHLPP1 | 564,42 | -0,12 | 0,03 | -3,63 | 2,84E-04 | 6,13E-03 |
| PATL1 | 1042,84 | -0,12 | 0,03 | -3,63 | 2,86E-04 | 6,13E-03 |
| IL1RAP | 4688,55 | -0,15 | 0,04 | -3,63 | 2,85E-04 | 6,13E-03 |
| IFI16 | 5922,87 | -0,13 | 0,04 | -3,63 | 2,88E-04 | 6,14E-03 |
| IRF2BP2 | 3377,92 | 0,06 | 0,02 | 3,63 | 2,88E-04 | 6,14E-03 |
| PTPN12 | 2856,65 | -0,10 | 0,03 | -3,62 | 2,90E-04 | 6,17E-03 |
| ANKRD9 | 443,40 | 0,26 | 0,07 | 3,62 | 2,90E-04 | 6,17E-03 |
| PCNX1 | 15679,02 | -0,10 | 0,03 | -3,62 | 2,94E-04 | 6,22E-03 |
| PFKFB3 | 2019,32 | -0,18 | 0,05 | -3,62 | 2,94E-04 | 6,22E-03 |
| LIMK2 | 5028,76 | -0,18 | 0,05 | -3,62 | 2,94E-04 | 6,22E-03 |
| MAP4K5 | 1692,28 | 0,10 | 0,03 | 3,62 | 2,95E-04 | 6,23E-03 |
| NSMCE4A | 917,19 | 0,07 | 0,02 | 3,62 | 2,96E-04 | 6,24E-03 |
| DOCK2 | 15684,57 | -0,10 | 0,03 | -3,62 | 2,97E-04 | 6,24E-03 |
| RPLP0 | 12692,66 | 0,15 | 0,04 | 3,62 | 2,98E-04 | 6,24E-03 |
| LSM4 | 684,10 | 0,08 | 0,02 | 3,62 | 2,98E-04 | 6,24E-03 |
| RIGI | 4938,45 | -0,19 | 0,05 | -3,62 | 3,00E-04 | 6,25E-03 |
| ASH1L | 6537,18 | -0,05 | 0,01 | -3,62 | 2,99E-04 | 6,25E-03 |
| MRPL30 | 505,88 | 0,08 | 0,02 | 3,62 | 3,00E-04 | 6,25E-03 |
| BPHL | 180,35 | 0,10 | 0,03 | 3,61 | 3,02E-04 | 6,28E-03 |
| F2RL1 | 947,57 | -0,19 | 0,05 | -3,61 | 3,04E-04 | 6,29E-03 |
| SH3GLB2 | 1129,75 | 0,11 | 0,03 | 3,61 | 3,04E-04 | 6,29E-03 |
| OSBPL9 | 2448,42 | -0,07 | 0,02 | -3,61 | 3,05E-04 | 6,31E-03 |
| ATP23 | 146,57 | 0,14 | 0,04 | 3,61 | 3,06E-04 | 6,31E-03 |
| TIMM13 | 415,44 | 0,09 | 0,03 | 3,61 | 3,11E-04 | 6,39E-03 |
| MRPL42 | 1383,88 | 0,08 | 0,02 | 3,61 | 3,12E-04 | 6,41E-03 |
| BUD13 | 493,40 | -0,05 | 0,01 | -3,60 | 3,14E-04 | 6,44E-03 |
| GADD45GIP1 | 204,44 | 0,11 | 0,03 | 3,60 | 3,17E-04 | 6,46E-03 |
| BAZ2B | 12872,80 | -0,13 | 0,04 | -3,60 | 3,16E-04 | 6,46E-03 |
| STK11 | 3150,60 | 0,14 | 0,04 | 3,60 | 3,17E-04 | 6,46E-03 |
| DOCK8 | 21166,63 | -0,12 | 0,03 | -3,60 | 3,20E-04 | 6,51E-03 |
| PREX1 | 15717,31 | -0,16 | 0,04 | -3,60 | 3,22E-04 | 6,55E-03 |
| SQOR | 1297,09 | -0,11 | 0,03 | -3,59 | 3,25E-04 | 6,56E-03 |
| FCGR3B | 42293,38 | -0,19 | 0,05 | -3,59 | 3,25E-04 | 6,56E-03 |
| IL1R1 | 927,63 | -0,24 | 0,07 | -3,59 | 3,25E-04 | 6,56E-03 |
| SELENOW | 398,85 | 0,08 | 0,02 | 3,59 | 3,27E-04 | 6,56E-03 |
| LAT2 | 3685,96 | -0,12 | 0,03 | -3,59 | 3,26E-04 | 6,56E-03 |
| CRLF3 | 5281,22 | -0,08 | 0,02 | -3,59 | 3,26E-04 | 6,56E-03 |
| ADO | 575,93 | 0,09 | 0,03 | 3,59 | 3,28E-04 | 6,57E-03 |

| | | | | | | |
|---|---|---|---|---|---|---|
| SLC12A6 | 10653,16 | -0,11 | 0,03 | -3,59 | 3,28E-04 | 6,57E-03 |
| SLCO3A1 | 3011,08 | -0,12 | 0,03 | -3,59 | 3,31E-04 | 6,61E-03 |
| GIT2 | 5079,30 | -0,10 | 0,03 | -3,58 | 3,38E-04 | 6,73E-03 |
| PHACTR1 | 458,80 | -0,16 | 0,05 | -3,58 | 3,38E-04 | 6,73E-03 |
| KRIT1 | 898,43 | 0,10 | 0,03 | 3,58 | 3,39E-04 | 6,74E-03 |
| NADK | 6074,66 | -0,16 | 0,05 | -3,58 | 3,41E-04 | 6,75E-03 |
| SLC25A44 | 1639,37 | -0,13 | 0,04 | -3,58 | 3,41E-04 | 6,75E-03 |
| RAB24 | 767,54 | -0,12 | 0,03 | -3,58 | 3,42E-04 | 6,76E-03 |
| LAMP2 | 9319,45 | -0,12 | 0,03 | -3,58 | 3,45E-04 | 6,81E-03 |
| NAMPT | 64872,60 | -0,18 | 0,05 | -3,58 | 3,49E-04 | 6,88E-03 |
| TNFSF10 | 5590,62 | -0,16 | 0,04 | -3,57 | 3,57E-04 | 7,02E-03 |
| VCPKMT | 1031,83 | -0,09 | 0,03 | -3,57 | 3,59E-04 | 7,04E-03 |
| C6orf89 | 2440,14 | -0,07 | 0,02 | -3,56 | 3,66E-04 | 7,16E-03 |
| COA1 | 2610,12 | -0,10 | 0,03 | -3,56 | 3,66E-04 | 7,16E-03 |
| ELOVL5 | 6829,70 | -0,09 | 0,02 | -3,56 | 3,67E-04 | 7,17E-03 |
| RAPGEF2 | 5013,69 | -0,11 | 0,03 | -3,56 | 3,69E-04 | 7,20E-03 |
| PDE4B | 3864,32 | -0,10 | 0,03 | -3,56 | 3,74E-04 | 7,25E-03 |
| GCLC | 1299,01 | 0,12 | 0,03 | 3,56 | 3,74E-04 | 7,25E-03 |
| IPCEF1 | 3265,09 | -0,08 | 0,02 | -3,56 | 3,74E-04 | 7,25E-03 |
| CARD8 | 6763,52 | -0,09 | 0,03 | -3,56 | 3,75E-04 | 7,25E-03 |
| RFFL | 2693,66 | -0,10 | 0,03 | -3,56 | 3,74E-04 | 7,25E-03 |
| TMCC1 | 3001,20 | -0,12 | 0,03 | -3,55 | 3,81E-04 | 7,35E-03 |
| SMAP2 | 10042,80 | -0,15 | 0,04 | -3,55 | 3,83E-04 | 7,35E-03 |
| C11orf54 | 748,84 | -0,07 | 0,02 | -3,55 | 3,82E-04 | 7,35E-03 |
| HMGCR | 2164,45 | -0,10 | 0,03 | -3,55 | 3,84E-04 | 7,37E-03 |
| TMEM64 | 636,96 | 0,12 | 0,03 | 3,55 | 3,85E-04 | 7,37E-03 |
| RBM3 | 4401,72 | 0,11 | 0,03 | 3,55 | 3,86E-04 | 7,38E-03 |
| LAMP1 | 2667,94 | -0,11 | 0,03 | -3,55 | 3,88E-04 | 7,41E-03 |
| PANK2 | 3188,60 | -0,07 | 0,02 | -3,55 | 3,89E-04 | 7,41E-03 |
| MRPS27 | 818,58 | 0,06 | 0,02 | 3,55 | 3,92E-04 | 7,41E-03 |
| MAK | 703,93 | -0,18 | 0,05 | -3,55 | 3,91E-04 | 7,41E-03 |
| OSBPL11 | 2453,55 | -0,09 | 0,03 | -3,55 | 3,92E-04 | 7,41E-03 |
| RAB31 | 5526,66 | -0,12 | 0,03 | -3,55 | 3,92E-04 | 7,41E-03 |
| PRKDC | 7125,43 | -0,09 | 0,03 | -3,54 | 3,97E-04 | 7,50E-03 |
| PAK2 | 4772,72 | -0,11 | 0,03 | -3,54 | 4,03E-04 | 7,54E-03 |
| TMEM45B | 229,52 | -0,18 | 0,05 | -3,54 | 4,02E-04 | 7,54E-03 |
| STMP1 | 1610,55 | 0,16 | 0,04 | 3,54 | 4,02E-04 | 7,54E-03 |
| LRRFIP1 | 14331,86 | -0,08 | 0,02 | -3,54 | 4,02E-04 | 7,54E-03 |
| TGM3 | 93,10 | -0,27 | 0,08 | -3,54 | 4,02E-04 | 7,54E-03 |
| EIF4G3 | 2133,42 | -0,10 | 0,03 | -3,54 | 4,04E-04 | 7,56E-03 |
| E2F3 | 1191,38 | -0,10 | 0,03 | -3,54 | 4,06E-04 | 7,58E-03 |
| NEK6 | 1784,93 | -0,13 | 0,04 | -3,54 | 4,07E-04 | 7,59E-03 |
| CEP85 | 603,33 | 0,06 | 0,02 | 3,53 | 4,09E-04 | 7,61E-03 |
| SULT1B1 | 4281,57 | -0,16 | 0,05 | -3,53 | 4,11E-04 | 7,63E-03 |
| TRIM39 | 554,44 | -0,06 | 0,02 | -3,53 | 4,14E-04 | 7,67E-03 |
| RNF145 | 3701,80 | -0,07 | 0,02 | -3,53 | 4,14E-04 | 7,67E-03 |

| | | | | | | |
|---|---|---|---|---|---|---|
| ST6GALNAC3 | 263,57 | -0,13 | 0,04 | -3,53 | 4,16E-04 | 7,68E-03 |
| TIMM9 | 84,10 | 0,11 | 0,03 | 3,53 | 4,17E-04 | 7,68E-03 |
| NSFL1C | 2902,43 | -0,12 | 0,03 | -3,53 | 4,16E-04 | 7,68E-03 |
| TAF12 | 701,88 | -0,08 | 0,02 | -3,53 | 4,18E-04 | 7,69E-03 |
| CARF | 712,02 | 0,09 | 0,03 | 3,53 | 4,20E-04 | 7,70E-03 |
| EAF1 | 1324,55 | -0,06 | 0,02 | -3,53 | 4,22E-04 | 7,74E-03 |
| TRPC4AP | 2906,41 | -0,07 | 0,02 | -3,52 | 4,26E-04 | 7,80E-03 |
| IQGAP2 | 7289,11 | -0,09 | 0,03 | -3,52 | 4,28E-04 | 7,82E-03 |
| LRRK2 | 46451,38 | -0,15 | 0,04 | -3,52 | 4,29E-04 | 7,83E-03 |
| PIP4K2C | 624,22 | -0,07 | 0,02 | -3,52 | 4,33E-04 | 7,85E-03 |
| RHBDD1 | 1601,90 | 0,10 | 0,03 | 3,52 | 4,31E-04 | 7,85E-03 |
| RNF170 | 666,45 | 0,08 | 0,02 | 3,52 | 4,32E-04 | 7,85E-03 |
| GTF3A | 1484,23 | 0,09 | 0,03 | 3,52 | 4,33E-04 | 7,85E-03 |
| TBRG4 | 1063,87 | 0,07 | 0,02 | 3,52 | 4,34E-04 | 7,85E-03 |
| IWS1 | 1901,12 | -0,06 | 0,02 | -3,52 | 4,37E-04 | 7,89E-03 |
| DYRK1A | 11539,70 | -0,06 | 0,02 | -3,52 | 4,37E-04 | 7,89E-03 |
| HERPUD2 | 2172,42 | -0,06 | 0,02 | -3,52 | 4,39E-04 | 7,89E-03 |
| SAFB2 | 2604,14 | -0,06 | 0,02 | -3,51 | 4,40E-04 | 7,91E-03 |
| MRPL11 | 289,64 | 0,12 | 0,03 | 3,51 | 4,43E-04 | 7,94E-03 |
| ZNF124 | 409,91 | 0,11 | 0,03 | 3,51 | 4,44E-04 | 7,94E-03 |
| CD52 | 4284,55 | 0,21 | 0,06 | 3,51 | 4,46E-04 | 7,98E-03 |
| CCDC125 | 1553,78 | -0,11 | 0,03 | -3,51 | 4,48E-04 | 8,00E-03 |
| HCAR2 | 713,17 | -0,17 | 0,05 | -3,51 | 4,51E-04 | 8,03E-03 |
| RPL9 | 5210,98 | 0,31 | 0,09 | 3,51 | 4,55E-04 | 8,09E-03 |
| UGGT1 | 4967,34 | -0,06 | 0,02 | -3,51 | 4,54E-04 | 8,09E-03 |
| RPS5 | 6643,47 | 0,15 | 0,04 | 3,50 | 4,61E-04 | 8,19E-03 |
| ZCCHC2 | 2746,76 | -0,17 | 0,05 | -3,50 | 4,63E-04 | 8,20E-03 |
| GAB2 | 2600,47 | -0,18 | 0,05 | -3,50 | 4,65E-04 | 8,22E-03 |
| ZNF33A | 5349,45 | -0,09 | 0,03 | -3,50 | 4,67E-04 | 8,23E-03 |
| ARL4A | 945,50 | 0,21 | 0,06 | 3,50 | 4,67E-04 | 8,23E-03 |
| GPX4 | 1267,39 | 0,10 | 0,03 | 3,50 | 4,70E-04 | 8,26E-03 |
| SSH2 | 17233,48 | -0,14 | 0,04 | -3,50 | 4,70E-04 | 8,26E-03 |
| CFDP1 | 499,58 | -0,06 | 0,02 | -3,50 | 4,72E-04 | 8,28E-03 |
| MRPL43 | 444,17 | 0,06 | 0,02 | 3,49 | 4,74E-04 | 8,31E-03 |
| ACSS3 | 811,34 | -0,17 | 0,05 | -3,49 | 4,77E-04 | 8,34E-03 |
| TMEM154 | 20245,77 | -0,11 | 0,03 | -3,49 | 4,79E-04 | 8,37E-03 |
| CASP5 | 450,65 | -0,23 | 0,07 | -3,49 | 4,81E-04 | 8,39E-03 |
| GPSM2 | 410,60 | -0,12 | 0,04 | -3,49 | 4,85E-04 | 8,44E-03 |
| ATXN7 | 1004,82 | -0,08 | 0,02 | -3,49 | 4,88E-04 | 8,47E-03 |
| ABHD3 | 3153,56 | -0,10 | 0,03 | -3,49 | 4,89E-04 | 8,47E-03 |
| DSTYK | 1691,58 | -0,04 | 0,01 | -3,49 | 4,88E-04 | 8,47E-03 |
| CTBP2 | 2247,60 | -0,10 | 0,03 | -3,49 | 4,91E-04 | 8,50E-03 |
| TRIM27 | 2850,49 | -0,08 | 0,02 | -3,48 | 4,93E-04 | 8,51E-03 |
| UBE2E2 | 219,66 | 0,10 | 0,03 | 3,48 | 4,97E-04 | 8,57E-03 |
| MXI1 | 6686,81 | 0,23 | 0,07 | 3,48 | 5,01E-04 | 8,63E-03 |
| ST6GALNAC4 | 347,72 | 0,25 | 0,07 | 3,48 | 5,06E-04 | 8,69E-03 |

| | | | | | | |
|---|---|---|---|---|---|---|
| FECH | 5241,69 | 0,25 | 0,07 | 3,47 | 5,11E-04 | 8,77E-03 |
| PDK2 | 248,14 | 0,15 | 0,04 | 3,47 | 5,17E-04 | 8,86E-03 |
| MAEA | 1986,95 | -0,11 | 0,03 | -3,47 | 5,27E-04 | 9,01E-03 |
| HK2 | 2108,71 | -0,14 | 0,04 | -3,47 | 5,29E-04 | 9,02E-03 |
| DTX3L | 4147,53 | -0,13 | 0,04 | -3,47 | 5,30E-04 | 9,02E-03 |
| CLTC | 7087,26 | -0,07 | 0,02 | -3,47 | 5,29E-04 | 9,02E-03 |
| RBM33 | 7329,63 | -0,07 | 0,02 | -3,46 | 5,36E-04 | 9,08E-03 |
| BARD1 | 553,59 | 0,12 | 0,03 | 3,46 | 5,35E-04 | 9,08E-03 |
| SSR4 | 842,19 | 0,10 | 0,03 | 3,46 | 5,34E-04 | 9,08E-03 |
| PTGES2 | 391,31 | 0,09 | 0,03 | 3,46 | 5,39E-04 | 9,12E-03 |
| REM2 | 276,57 | -0,17 | 0,05 | -3,46 | 5,41E-04 | 9,14E-03 |
| ARL6IP1 | 2950,00 | 0,13 | 0,04 | 3,46 | 5,42E-04 | 9,15E-03 |
| APOBEC3A | 5098,83 | -0,17 | 0,05 | -3,46 | 5,43E-04 | 9,16E-03 |
| ETS2 | 2264,79 | -0,11 | 0,03 | -3,46 | 5,45E-04 | 9,17E-03 |
| ADAM17 | 2994,53 | -0,08 | 0,02 | -3,45 | 5,51E-04 | 9,24E-03 |
| GRB2 | 8752,46 | -0,09 | 0,03 | -3,45 | 5,51E-04 | 9,24E-03 |
| RABIF | 452,39 | -0,05 | 0,01 | -3,45 | 5,56E-04 | 9,31E-03 |
| APOL1 | 994,02 | -0,17 | 0,05 | -3,45 | 5,58E-04 | 9,32E-03 |
| CMTM2 | 1848,62 | -0,19 | 0,06 | -3,45 | 5,60E-04 | 9,35E-03 |
| GPALPP1 | 897,55 | 0,07 | 0,02 | 3,45 | 5,61E-04 | 9,36E-03 |
| CCDC152 | 152,79 | 0,18 | 0,05 | 3,45 | 5,64E-04 | 9,39E-03 |
| IL16 | 13923,48 | -0,08 | 0,02 | -3,45 | 5,65E-04 | 9,39E-03 |
| POLB | 1028,66 | -0,10 | 0,03 | -3,45 | 5,67E-04 | 9,41E-03 |
| RIOK3 | 10989,15 | 0,20 | 0,06 | 3,45 | 5,69E-04 | 9,43E-03 |
| RPL27 | 3716,09 | 0,24 | 0,07 | 3,45 | 5,70E-04 | 9,44E-03 |
| PRDM5 | 591,14 | -0,22 | 0,06 | -3,44 | 5,73E-04 | 9,47E-03 |
| CCDC30 | 406,43 | -0,12 | 0,03 | -3,44 | 5,74E-04 | 9,47E-03 |
| SLC35A2 | 202,98 | -0,08 | 0,02 | -3,44 | 5,80E-04 | 9,54E-03 |
| MDFIC | 2675,56 | 0,11 | 0,03 | 3,44 | 5,80E-04 | 9,54E-03 |
| CHD8 | 3932,47 | -0,08 | 0,02 | -3,43 | 5,93E-04 | 9,73E-03 |
| RPS18 | 14281,99 | 0,23 | 0,07 | 3,43 | 5,96E-04 | 9,76E-03 |
| ISY1 | 510,44 | -0,06 | 0,02 | -3,43 | 5,96E-04 | 9,76E-03 |
| ATP6V0D1 | 3672,16 | -0,11 | 0,03 | -3,43 | 5,97E-04 | 9,76E-03 |
| INKA2 | 376,71 | -0,14 | 0,04 | -3,43 | 5,99E-04 | 9,78E-03 |
| PLEKHM3 | 1801,17 | -0,09 | 0,03 | -3,43 | 6,02E-04 | 9,81E-03 |
| POLE2 | 135,47 | 0,25 | 0,07 | 3,43 | 6,07E-04 | 9,88E-03 |
| TUFT1 | 159,28 | -0,13 | 0,04 | -3,43 | 6,09E-04 | 9,90E-03 |
| PFKFB4 | 2314,61 | -0,15 | 0,04 | -3,42 | 6,25E-04 | 1,01E-02 |
| DCUN1D3 | 367,01 | -0,09 | 0,03 | -3,42 | 6,25E-04 | 1,01E-02 |
| VPS51 | 2813,77 | 0,07 | 0,02 | 3,42 | 6,24E-04 | 1,01E-02 |
| MTHFS | 312,78 | -0,11 | 0,03 | -3,42 | 6,23E-04 | 1,01E-02 |
| YJU2 | 179,64 | 0,08 | 0,02 | 3,42 | 6,26E-04 | 1,01E-02 |
| CLIC2 | 248,09 | 0,18 | 0,05 | 3,42 | 6,25E-04 | 1,01E-02 |
| RASSF3 | 6013,10 | -0,11 | 0,03 | -3,42 | 6,27E-04 | 1,01E-02 |
| ISG20 | 2839,74 | -0,11 | 0,03 | -3,42 | 6,31E-04 | 1,01E-02 |
| USP8 | 3620,78 | -0,05 | 0,02 | -3,42 | 6,31E-04 | 1,01E-02 |

| | | | | | | |
|---|---|---|---|---|---|---|
| DCAF4L1 | 85,08 | -0,14 | 0,04 | -3,42 | 6,37E-04 | 1,02E-02 |
| ADM | 1059,98 | -0,19 | 0,06 | -3,41 | 6,41E-04 | 1,02E-02 |
| ATP6V1F | 152,40 | 0,08 | 0,02 | 3,41 | 6,41E-04 | 1,02E-02 |
| PPP2R5C | 8862,07 | -0,06 | 0,02 | -3,41 | 6,44E-04 | 1,03E-02 |
| RPIA | 2047,14 | 0,19 | 0,06 | 3,41 | 6,51E-04 | 1,04E-02 |
| MEFV | 6970,96 | -0,14 | 0,04 | -3,41 | 6,56E-04 | 1,04E-02 |
| RAB1A | 2875,52 | -0,09 | 0,03 | -3,41 | 6,57E-04 | 1,04E-02 |
| HARS2 | 1083,79 | -0,06 | 0,02 | -3,40 | 6,64E-04 | 1,05E-02 |
| TOP1 | 2415,64 | -0,08 | 0,02 | -3,40 | 6,64E-04 | 1,05E-02 |
| TECPR2 | 3741,57 | -0,16 | 0,05 | -3,40 | 6,68E-04 | 1,06E-02 |
| NBPF26 | 2024,55 | -0,21 | 0,06 | -3,40 | 6,71E-04 | 1,06E-02 |
| CRAT | 122,98 | 0,23 | 0,07 | 3,40 | 6,73E-04 | 1,06E-02 |
| FSD1L | 329,60 | 0,14 | 0,04 | 3,40 | 6,76E-04 | 1,06E-02 |
| MAF1 | 3435,22 | 0,16 | 0,05 | 3,40 | 6,85E-04 | 1,08E-02 |
| OXLD1 | 776,84 | 0,09 | 0,03 | 3,39 | 6,88E-04 | 1,08E-02 |
| NME8 | 371,13 | -0,16 | 0,05 | -3,39 | 6,92E-04 | 1,08E-02 |
| HSPA5 | 2529,74 | -0,08 | 0,02 | -3,39 | 6,96E-04 | 1,09E-02 |
| KLC3 | 360,87 | 0,29 | 0,08 | 3,39 | 6,96E-04 | 1,09E-02 |
| BNIP3L | 38687,93 | 0,21 | 0,06 | 3,39 | 6,95E-04 | 1,09E-02 |
| C1RL | 2335,13 | -0,12 | 0,04 | -3,39 | 6,98E-04 | 1,09E-02 |
| ADGRG3 | 1402,58 | -0,22 | 0,07 | -3,39 | 7,02E-04 | 1,09E-02 |
| SBF2 | 3995,54 | -0,11 | 0,03 | -3,39 | 7,06E-04 | 1,10E-02 |
| KCNE1 | 769,27 | -0,15 | 0,05 | -3,38 | 7,12E-04 | 1,11E-02 |
| CCPG1 | 2494,02 | -0,11 | 0,03 | -3,38 | 7,15E-04 | 1,11E-02 |
| MTMR3 | 6362,72 | -0,13 | 0,04 | -3,38 | 7,16E-04 | 1,11E-02 |
| FGD4 | 3244,57 | -0,13 | 0,04 | -3,38 | 7,17E-04 | 1,11E-02 |
| ZNF217 | 9209,76 | -0,08 | 0,02 | -3,38 | 7,17E-04 | 1,11E-02 |
| AKR1A1 | 676,98 | 0,06 | 0,02 | 3,38 | 7,21E-04 | 1,11E-02 |
| EIF2D | 808,96 | 0,06 | 0,02 | 3,38 | 7,23E-04 | 1,11E-02 |
| GPR141 | 1012,57 | -0,18 | 0,05 | -3,38 | 7,23E-04 | 1,11E-02 |
| PTPRJ | 10519,10 | -0,13 | 0,04 | -3,38 | 7,24E-04 | 1,11E-02 |
| CCNQ | 322,56 | 0,07 | 0,02 | 3,38 | 7,26E-04 | 1,11E-02 |
| TAP1 | 6516,27 | -0,12 | 0,04 | -3,38 | 7,31E-04 | 1,11E-02 |
| DZIP3 | 729,46 | 0,15 | 0,04 | 3,38 | 7,32E-04 | 1,11E-02 |
| IFT56 | 181,29 | -0,19 | 0,06 | -3,38 | 7,29E-04 | 1,11E-02 |
| APLF | 302,48 | 0,12 | 0,04 | 3,38 | 7,31E-04 | 1,11E-02 |
| CKAP4 | 897,83 | -0,14 | 0,04 | -3,38 | 7,32E-04 | 1,11E-02 |
| CNOT1 | 7657,84 | -0,08 | 0,02 | -3,38 | 7,37E-04 | 1,12E-02 |
| ADAM19 | 3835,10 | -0,13 | 0,04 | -3,37 | 7,46E-04 | 1,13E-02 |
| TM9SF2 | 3416,94 | -0,08 | 0,02 | -3,37 | 7,47E-04 | 1,13E-02 |
| DECR1 | 4197,39 | -0,09 | 0,03 | -3,37 | 7,54E-04 | 1,14E-02 |
| CDKN2AIPNL | 214,02 | 0,09 | 0,03 | 3,37 | 7,53E-04 | 1,14E-02 |
| PRR14L | 4161,38 | -0,05 | 0,02 | -3,37 | 7,57E-04 | 1,14E-02 |
| MZT2B | 1166,00 | 0,14 | 0,04 | 3,37 | 7,65E-04 | 1,14E-02 |
| KIAA0319L | 2582,99 | -0,12 | 0,03 | -3,37 | 7,60E-04 | 1,14E-02 |
| XPC | 2918,08 | -0,07 | 0,02 | -3,36 | 7,66E-04 | 1,14E-02 |

| | | | | | | |
|---|---|---|---|---|---|---|
| SELENOH | 885,82 | 0,10 | 0,03 | 3,37 | 7,65E-04 | 1,14E-02 |
| SLC15A4 | 3752,00 | -0,10 | 0,03 | -3,37 | 7,59E-04 | 1,14E-02 |
| RGL4 | 645,34 | -0,13 | 0,04 | -3,37 | 7,65E-04 | 1,14E-02 |
| FNIP1 | 5354,06 | -0,10 | 0,03 | -3,37 | 7,63E-04 | 1,14E-02 |
| IKBIP | 1376,56 | -0,14 | 0,04 | -3,37 | 7,64E-04 | 1,14E-02 |
| DHRS13 | 885,13 | -0,13 | 0,04 | -3,36 | 7,72E-04 | 1,15E-02 |
| BRCA1 | 522,75 | -0,10 | 0,03 | -3,36 | 7,71E-04 | 1,15E-02 |
| THBD | 868,09 | -0,18 | 0,05 | -3,36 | 7,74E-04 | 1,15E-02 |
| CARD6 | 465,38 | -0,11 | 0,03 | -3,36 | 7,83E-04 | 1,16E-02 |
| GCC1 | 1917,34 | -0,07 | 0,02 | -3,36 | 7,83E-04 | 1,16E-02 |
| PTPRO | 491,76 | 0,14 | 0,04 | 3,36 | 7,86E-04 | 1,16E-02 |
| UBXN6 | 8577,36 | 0,23 | 0,07 | 3,36 | 7,89E-04 | 1,17E-02 |
| C7orf50 | 423,00 | 0,10 | 0,03 | 3,35 | 7,96E-04 | 1,18E-02 |
| TRMT61B | 319,96 | 0,07 | 0,02 | 3,35 | 8,03E-04 | 1,18E-02 |
| LRATD2 | 530,84 | 0,10 | 0,03 | 3,35 | 8,03E-04 | 1,18E-02 |
| SP1 | 6820,32 | -0,09 | 0,03 | -3,35 | 8,02E-04 | 1,18E-02 |
| COPG1 | 2324,42 | -0,07 | 0,02 | -3,35 | 8,06E-04 | 1,18E-02 |
| INPP5A | 995,10 | -0,10 | 0,03 | -3,35 | 8,06E-04 | 1,18E-02 |
| CAP1 | 7735,22 | -0,09 | 0,03 | -3,35 | 8,06E-04 | 1,18E-02 |
| RPL8 | 10472,91 | 0,11 | 0,03 | 3,35 | 8,08E-04 | 1,18E-02 |
| CRLS1 | 945,03 | 0,08 | 0,02 | 3,35 | 8,18E-04 | 1,20E-02 |
| BOLA1 | 161,91 | 0,08 | 0,02 | 3,34 | 8,26E-04 | 1,21E-02 |
| SVIL | 4217,74 | -0,14 | 0,04 | -3,34 | 8,36E-04 | 1,22E-02 |
| HMBS | 206,81 | 0,17 | 0,05 | 3,34 | 8,36E-04 | 1,22E-02 |
| ANAPC10 | 174,73 | 0,10 | 0,03 | 3,34 | 8,42E-04 | 1,22E-02 |
| ZBTB18 | 2645,54 | -0,08 | 0,02 | -3,34 | 8,48E-04 | 1,23E-02 |
| WDR74 | 474,66 | 0,06 | 0,02 | 3,34 | 8,48E-04 | 1,23E-02 |
| CD300A | 3303,79 | -0,10 | 0,03 | -3,34 | 8,47E-04 | 1,23E-02 |
| RIPK1 | 1404,27 | -0,08 | 0,02 | -3,34 | 8,48E-04 | 1,23E-02 |
| FANCF | 321,27 | 0,10 | 0,03 | 3,33 | 8,59E-04 | 1,24E-02 |
| NPTN | 1807,74 | -0,07 | 0,02 | -3,33 | 8,61E-04 | 1,24E-02 |
| IL13RA1 | 3133,22 | -0,13 | 0,04 | -3,33 | 8,63E-04 | 1,24E-02 |
| ZBTB49 | 465,88 | -0,05 | 0,02 | -3,33 | 8,63E-04 | 1,24E-02 |
| TMEM14B | 787,83 | 0,07 | 0,02 | 3,33 | 8,60E-04 | 1,24E-02 |
| SYK | 9598,70 | -0,10 | 0,03 | -3,33 | 8,64E-04 | 1,24E-02 |
| THEMIS2 | 9521,59 | -0,12 | 0,04 | -3,33 | 8,68E-04 | 1,24E-02 |
| FAF2 | 1251,77 | -0,04 | 0,01 | -3,33 | 8,78E-04 | 1,26E-02 |
| MAP2K6 | 2400,09 | -0,09 | 0,03 | -3,33 | 8,79E-04 | 1,26E-02 |
| RNF19B | 1555,73 | -0,13 | 0,04 | -3,33 | 8,84E-04 | 1,26E-02 |
| NLRP12 | 5104,07 | -0,13 | 0,04 | -3,32 | 8,87E-04 | 1,26E-02 |
| PIGW | 143,09 | 0,08 | 0,02 | 3,32 | 8,87E-04 | 1,26E-02 |
| LCMT2 | 392,50 | 0,09 | 0,03 | 3,32 | 8,86E-04 | 1,26E-02 |
| DUS1L | 771,92 | 0,08 | 0,02 | 3,33 | 8,84E-04 | 1,26E-02 |
| SPRYD7 | 146,81 | 0,08 | 0,02 | 3,32 | 8,95E-04 | 1,27E-02 |
| FKBP14 | 205,74 | 0,10 | 0,03 | 3,32 | 8,98E-04 | 1,27E-02 |
| JADE1 | 2647,09 | -0,06 | 0,02 | -3,32 | 9,01E-04 | 1,27E-02 |

| | | | | | | |
|---|---|---|---|---|---|---|
| TBL1X | 2853,84 | -0,14 | 0,04 | -3,32 | 9,02E-04 | 1,27E-02 |
| LRG1 | 1335,65 | -0,16 | 0,05 | -3,32 | 9,14E-04 | 1,29E-02 |
| SNX3 | 3949,35 | 0,17 | 0,05 | 3,31 | 9,17E-04 | 1,29E-02 |
| KCTD3 | 239,52 | 0,10 | 0,03 | 3,31 | 9,24E-04 | 1,30E-02 |
| EIF5A | 1799,95 | 0,08 | 0,02 | 3,31 | 9,26E-04 | 1,30E-02 |
| DYSF | 5450,47 | -0,23 | 0,07 | -3,31 | 9,36E-04 | 1,31E-02 |
| RCL1 | 597,81 | 0,10 | 0,03 | 3,31 | 9,44E-04 | 1,32E-02 |
| VPS8 | 8542,33 | -0,10 | 0,03 | -3,31 | 9,47E-04 | 1,33E-02 |
| ZNF445 | 2772,48 | -0,13 | 0,04 | -3,31 | 9,49E-04 | 1,33E-02 |
| SERPINA1 | 5768,63 | -0,17 | 0,05 | -3,30 | 9,59E-04 | 1,34E-02 |
| CDK11B | 712,46 | -0,08 | 0,02 | -3,30 | 9,59E-04 | 1,34E-02 |
| PELI1 | 9412,87 | -0,11 | 0,03 | -3,30 | 9,58E-04 | 1,34E-02 |
| NABP1 | 21358,62 | -0,12 | 0,04 | -3,30 | 9,61E-04 | 1,34E-02 |
| DNAJC6 | 458,33 | 0,25 | 0,07 | 3,30 | 9,72E-04 | 1,35E-02 |
| KMT2C | 13025,60 | -0,09 | 0,03 | -3,30 | 9,75E-04 | 1,35E-02 |
| LATS2 | 671,66 | -0,10 | 0,03 | -3,30 | 9,77E-04 | 1,36E-02 |
| SH3BP5L | 2262,56 | -0,09 | 0,03 | -3,29 | 9,88E-04 | 1,37E-02 |
| SLC25A36 | 1959,58 | 0,14 | 0,04 | 3,29 | 9,91E-04 | 1,37E-02 |
| MRPL57 | 426,83 | 0,10 | 0,03 | 3,29 | 9,90E-04 | 1,37E-02 |
| MTMR12 | 1164,46 | -0,05 | 0,02 | -3,29 | 9,95E-04 | 1,37E-02 |
| H3-3A | 10928,76 | -0,11 | 0,03 | -3,29 | 1,00E-03 | 1,38E-02 |
| SIVA1 | 700,45 | 0,06 | 0,02 | 3,29 | 1,01E-03 | 1,38E-02 |
| SLC33A1 | 709,06 | 0,07 | 0,02 | 3,29 | 1,02E-03 | 1,39E-02 |
| LPGAT1 | 8045,93 | -0,10 | 0,03 | -3,29 | 1,01E-03 | 1,39E-02 |
| PECAM1 | 10283,83 | -0,11 | 0,03 | -3,28 | 1,02E-03 | 1,40E-02 |
| CARS1 | 1761,97 | -0,07 | 0,02 | -3,28 | 1,02E-03 | 1,40E-02 |
| MFSD3 | 105,65 | 0,10 | 0,03 | 3,28 | 1,03E-03 | 1,41E-02 |
| STK38 | 6241,25 | -0,06 | 0,02 | -3,28 | 1,04E-03 | 1,42E-02 |
| IGSF6 | 8079,57 | -0,12 | 0,04 | -3,28 | 1,04E-03 | 1,42E-02 |
| ZDHHC18 | 5644,17 | -0,15 | 0,04 | -3,28 | 1,05E-03 | 1,43E-02 |
| WDFY4 | 4072,49 | -0,16 | 0,05 | -3,28 | 1,05E-03 | 1,43E-02 |
| RESF1 | 33057,02 | -0,13 | 0,04 | -3,28 | 1,05E-03 | 1,43E-02 |
| KMT2E | 7211,93 | -0,06 | 0,02 | -3,28 | 1,05E-03 | 1,43E-02 |
| LNX2 | 558,94 | 0,07 | 0,02 | 3,28 | 1,05E-03 | 1,43E-02 |
| NLRC4 | 1650,26 | -0,12 | 0,04 | -3,27 | 1,06E-03 | 1,43E-02 |
| NCR1 | 322,39 | -0,14 | 0,04 | -3,27 | 1,07E-03 | 1,44E-02 |
| ICAM3 | 8543,78 | -0,11 | 0,03 | -3,27 | 1,07E-03 | 1,44E-02 |
| KCNE3 | 5387,09 | -0,11 | 0,03 | -3,27 | 1,07E-03 | 1,44E-02 |
| RAD21 | 4508,80 | -0,08 | 0,03 | -3,27 | 1,07E-03 | 1,44E-02 |
| ZDHHC2 | 3053,20 | 0,12 | 0,04 | 3,27 | 1,08E-03 | 1,45E-02 |
| ZNF397 | 2415,92 | -0,05 | 0,02 | -3,27 | 1,08E-03 | 1,45E-02 |
| SLC4A1AP | 909,85 | -0,05 | 0,02 | -3,27 | 1,09E-03 | 1,46E-02 |
| MED13L | 14793,86 | -0,08 | 0,02 | -3,27 | 1,09E-03 | 1,46E-02 |
| TOGARAM1 | 888,83 | 0,06 | 0,02 | 3,27 | 1,09E-03 | 1,46E-02 |
| CCDC174 | 1167,22 | -0,05 | 0,01 | -3,27 | 1,09E-03 | 1,46E-02 |
| BLTP3B | 3281,14 | -0,10 | 0,03 | -3,26 | 1,10E-03 | 1,48E-02 |

| | | | | | | |
|---|---|---|---|---|---|---|
| RLIG1 | 583,54 | 0,14 | 0,04 | 3,26 | 1,11E-03 | 1,48E-02 |
| SH3KBP1 | 3219,43 | -0,09 | 0,03 | -3,26 | 1,11E-03 | 1,48E-02 |
| NME3 | 569,61 | 0,09 | 0,03 | 3,26 | 1,11E-03 | 1,48E-02 |
| ENSG000002 | 117,94 | -0,18 | 0,06 | -3,26 | 1,12E-03 | 1,49E-02 |
| ZNF681 | 253,16 | 0,17 | 0,05 | 3,26 | 1,12E-03 | 1,49E-02 |
| ALG10B | 671,85 | 0,14 | 0,04 | 3,26 | 1,13E-03 | 1,50E-02 |
| IFIT2 | 7776,46 | -0,25 | 0,08 | -3,26 | 1,13E-03 | 1,50E-02 |
| SLC25A25 | 560,90 | 0,06 | 0,02 | 3,25 | 1,13E-03 | 1,50E-02 |
| CREM | 423,37 | -0,07 | 0,02 | -3,25 | 1,14E-03 | 1,50E-02 |
| RBM38 | 6912,82 | 0,23 | 0,07 | 3,25 | 1,14E-03 | 1,50E-02 |
| HERC5 | 1624,28 | -0,30 | 0,09 | -3,25 | 1,14E-03 | 1,50E-02 |
| STK24 | 3755,84 | -0,06 | 0,02 | -3,25 | 1,15E-03 | 1,51E-02 |
| KRT23 | 2277,71 | -0,19 | 0,06 | -3,25 | 1,15E-03 | 1,51E-02 |
| ERV3-1 | 3107,26 | -0,12 | 0,04 | -3,25 | 1,15E-03 | 1,52E-02 |
| CASP9 | 433,91 | -0,09 | 0,03 | -3,25 | 1,16E-03 | 1,53E-02 |
| SPTLC2 | 4877,61 | -0,08 | 0,02 | -3,25 | 1,17E-03 | 1,53E-02 |
| TET2 | 12821,87 | -0,10 | 0,03 | -3,25 | 1,17E-03 | 1,53E-02 |
| MAB21L3 | 73,06 | -0,22 | 0,07 | -3,25 | 1,17E-03 | 1,53E-02 |
| NOD2 | 1695,99 | -0,17 | 0,05 | -3,25 | 1,17E-03 | 1,53E-02 |
| CPPED1 | 7838,65 | -0,12 | 0,04 | -3,25 | 1,17E-03 | 1,53E-02 |
| BIN3 | 1349,74 | -0,09 | 0,03 | -3,24 | 1,18E-03 | 1,53E-02 |
| ZNF517 | 204,69 | 0,09 | 0,03 | 3,24 | 1,18E-03 | 1,54E-02 |
| ARHGAP30 | 14602,62 | -0,08 | 0,02 | -3,24 | 1,18E-03 | 1,54E-02 |
| CREBRF | 8117,73 | -0,09 | 0,03 | -3,24 | 1,18E-03 | 1,54E-02 |
| HIRA | 312,93 | -0,08 | 0,02 | -3,24 | 1,19E-03 | 1,54E-02 |
| HDDC3 | 325,09 | 0,08 | 0,02 | 3,24 | 1,19E-03 | 1,54E-02 |
| H1-4 | 5250,92 | 0,12 | 0,04 | 3,24 | 1,20E-03 | 1,55E-02 |
| RBX1 | 1184,94 | 0,13 | 0,04 | 3,24 | 1,20E-03 | 1,55E-02 |
| PPTC7 | 2950,56 | -0,07 | 0,02 | -3,24 | 1,20E-03 | 1,55E-02 |
| RAB2B | 4165,25 | 0,19 | 0,06 | 3,24 | 1,21E-03 | 1,55E-02 |
| CDA | 485,59 | -0,19 | 0,06 | -3,24 | 1,21E-03 | 1,55E-02 |
| HTATIP2 | 1211,23 | -0,09 | 0,03 | -3,24 | 1,21E-03 | 1,55E-02 |
| IDNK | 281,48 | 0,07 | 0,02 | 3,24 | 1,22E-03 | 1,56E-02 |
| MNDA | 30350,88 | -0,15 | 0,05 | -3,23 | 1,22E-03 | 1,56E-02 |
| RPL13 | 20477,52 | 0,12 | 0,04 | 3,23 | 1,22E-03 | 1,57E-02 |
| C10orf88 | 335,13 | 0,09 | 0,03 | 3,23 | 1,23E-03 | 1,57E-02 |
| TMCC2 | 436,84 | 0,25 | 0,08 | 3,23 | 1,23E-03 | 1,57E-02 |
| MRPL41 | 449,68 | 0,09 | 0,03 | 3,23 | 1,23E-03 | 1,57E-02 |
| CHMP3 | 2227,12 | -0,06 | 0,02 | -3,23 | 1,23E-03 | 1,57E-02 |
| PLXDC2 | 5394,21 | -0,11 | 0,03 | -3,23 | 1,24E-03 | 1,59E-02 |
| ZNF516 | 5159,51 | -0,14 | 0,04 | -3,23 | 1,24E-03 | 1,59E-02 |
| HINT1 | 888,10 | 0,20 | 0,06 | 3,23 | 1,25E-03 | 1,59E-02 |
| NHSL2 | 4587,46 | -0,15 | 0,05 | -3,22 | 1,26E-03 | 1,59E-02 |
| BCL7C | 465,72 | 0,08 | 0,03 | 3,23 | 1,25E-03 | 1,59E-02 |
| ZDHHC4 | 243,18 | 0,05 | 0,02 | 3,23 | 1,26E-03 | 1,59E-02 |
| TOR1A | 920,40 | -0,06 | 0,02 | -3,23 | 1,25E-03 | 1,59E-02 |

| | | | | | | |
|---|---|---|---|---|---|---|
| MBD3 | 703,69 | 0,07 | 0,02 | 3,22 | 1,26E-03 | 1,59E-02 |
| SLC35B4 | 633,69 | 0,05 | 0,02 | 3,23 | 1,26E-03 | 1,59E-02 |
| P4HA2 | 210,26 | -0,15 | 0,05 | -3,22 | 1,26E-03 | 1,59E-02 |
| MID1IP1 | 2856,95 | -0,11 | 0,03 | -3,23 | 1,25E-03 | 1,59E-02 |
| NSD1 | 6548,47 | -0,06 | 0,02 | -3,22 | 1,27E-03 | 1,60E-02 |
| ASB8 | 1374,59 | -0,05 | 0,01 | -3,22 | 1,28E-03 | 1,60E-02 |
| DHRS7 | 2708,55 | -0,10 | 0,03 | -3,22 | 1,27E-03 | 1,60E-02 |
| RPS7 | 2684,17 | 0,28 | 0,09 | 3,22 | 1,28E-03 | 1,60E-02 |
| TFRC | 2861,57 | 0,12 | 0,04 | 3,22 | 1,29E-03 | 1,61E-02 |
| ITCH | 5720,55 | -0,07 | 0,02 | -3,22 | 1,29E-03 | 1,62E-02 |
| SMIM5 | 581,16 | 0,20 | 0,06 | 3,22 | 1,30E-03 | 1,62E-02 |
| TOX4 | 2229,07 | -0,05 | 0,02 | -3,21 | 1,31E-03 | 1,63E-02 |
| NBDY | 114,68 | 0,09 | 0,03 | 3,21 | 1,31E-03 | 1,63E-02 |
| PIK3AP1 | 7050,21 | -0,10 | 0,03 | -3,21 | 1,31E-03 | 1,64E-02 |
| RPL27A | 9674,11 | 0,14 | 0,04 | 3,21 | 1,32E-03 | 1,64E-02 |
| AMIGO2 | 248,16 | 0,09 | 0,03 | 3,21 | 1,32E-03 | 1,64E-02 |
| ARPC2 | 10215,86 | -0,07 | 0,02 | -3,21 | 1,33E-03 | 1,65E-02 |
| GAPVD1 | 4098,14 | -0,06 | 0,02 | -3,21 | 1,34E-03 | 1,65E-02 |
| PLBD1 | 4298,92 | -0,13 | 0,04 | -3,21 | 1,34E-03 | 1,65E-02 |
| DUS4L | 216,35 | 0,08 | 0,02 | 3,21 | 1,34E-03 | 1,65E-02 |
| FAM53C | 2101,16 | -0,14 | 0,04 | -3,21 | 1,34E-03 | 1,65E-02 |
| TAP2 | 1885,59 | -0,11 | 0,04 | -3,21 | 1,35E-03 | 1,67E-02 |
| DUSP1 | 9951,17 | -0,14 | 0,04 | -3,20 | 1,35E-03 | 1,67E-02 |
| PTAFR | 7640,74 | -0,13 | 0,04 | -3,20 | 1,35E-03 | 1,67E-02 |
| OSBP2 | 1254,41 | 0,25 | 0,08 | 3,20 | 1,35E-03 | 1,67E-02 |
| ANO10 | 490,40 | -0,09 | 0,03 | -3,20 | 1,36E-03 | 1,67E-02 |
| CREG1 | 2548,34 | 0,16 | 0,05 | 3,20 | 1,36E-03 | 1,67E-02 |
| POLR2H | 407,28 | 0,06 | 0,02 | 3,20 | 1,36E-03 | 1,67E-02 |
| EIF4E3 | 5367,56 | -0,11 | 0,04 | -3,20 | 1,36E-03 | 1,67E-02 |
| ZNF724 | 75,96 | 0,10 | 0,03 | 3,20 | 1,36E-03 | 1,67E-02 |
| PFDN5 | 2097,49 | 0,22 | 0,07 | 3,20 | 1,37E-03 | 1,67E-02 |
| VPS4B | 4702,62 | -0,08 | 0,02 | -3,20 | 1,37E-03 | 1,68E-02 |
| EZH1 | 2459,70 | -0,07 | 0,02 | -3,20 | 1,37E-03 | 1,68E-02 |
| ECSIT | 241,68 | 0,12 | 0,04 | 3,20 | 1,38E-03 | 1,68E-02 |
| SLC22A4 | 1326,71 | -0,14 | 0,04 | -3,20 | 1,38E-03 | 1,68E-02 |
| MXD4 | 1986,92 | 0,08 | 0,03 | 3,20 | 1,39E-03 | 1,69E-02 |
| OASL | 788,97 | -0,25 | 0,08 | -3,20 | 1,39E-03 | 1,69E-02 |
| IVNS1ABP | 7451,95 | -0,09 | 0,03 | -3,20 | 1,39E-03 | 1,69E-02 |
| METTL17 | 628,42 | 0,06 | 0,02 | 3,19 | 1,41E-03 | 1,71E-02 |
| COA8 | 265,37 | 0,05 | 0,02 | 3,19 | 1,41E-03 | 1,71E-02 |
| EXOSC8 | 569,33 | 0,10 | 0,03 | 3,19 | 1,43E-03 | 1,72E-02 |
| TNRC6B | 11092,84 | -0,09 | 0,03 | -3,19 | 1,42E-03 | 1,72E-02 |
| TRIM5 | 1014,91 | -0,11 | 0,04 | -3,19 | 1,42E-03 | 1,72E-02 |
| ZMYM6 | 916,83 | 0,07 | 0,02 | 3,19 | 1,43E-03 | 1,72E-02 |
| NQO2 | 1520,72 | -0,15 | 0,05 | -3,19 | 1,43E-03 | 1,72E-02 |
| FUNDC2 | 788,50 | 0,10 | 0,03 | 3,19 | 1,43E-03 | 1,72E-02 |

| | | | | | | |
|---|---|---|---|---|---|---|
| TAOK1 | 8431,61 | -0,06 | 0,02 | -3,19 | 1,42E-03 | 1,72E-02 |
| CAMK2D | 2163,48 | 0,10 | 0,03 | 3,19 | 1,43E-03 | 1,72E-02 |
| ECI1 | 344,25 | 0,06 | 0,02 | 3,19 | 1,43E-03 | 1,72E-02 |
| PRADC1 | 127,75 | 0,09 | 0,03 | 3,19 | 1,44E-03 | 1,73E-02 |
| PTPMT1 | 180,22 | 0,06 | 0,02 | 3,19 | 1,45E-03 | 1,73E-02 |
| KCNJ2 | 4996,30 | -0,15 | 0,05 | -3,18 | 1,45E-03 | 1,74E-02 |
| RNF24 | 4311,45 | -0,20 | 0,06 | -3,18 | 1,48E-03 | 1,77E-02 |
| ENSG000002 | 319,65 | -0,18 | 0,06 | -3,18 | 1,48E-03 | 1,77E-02 |
| ZMYM1 | 581,20 | 0,10 | 0,03 | 3,18 | 1,49E-03 | 1,78E-02 |
| TP53BP1 | 1582,18 | -0,07 | 0,02 | -3,18 | 1,50E-03 | 1,78E-02 |
| ANKRD36 | 3201,60 | 0,10 | 0,03 | 3,18 | 1,50E-03 | 1,78E-02 |
| CCNJL | 1383,67 | -0,18 | 0,06 | -3,17 | 1,50E-03 | 1,78E-02 |
| SEC23B | 1684,90 | -0,06 | 0,02 | -3,17 | 1,50E-03 | 1,78E-02 |
| TRAFD1 | 2303,59 | -0,10 | 0,03 | -3,17 | 1,51E-03 | 1,79E-02 |
| IFIT3 | 4894,68 | -0,29 | 0,09 | -3,17 | 1,52E-03 | 1,79E-02 |
| VPS37B | 1092,33 | -0,12 | 0,04 | -3,17 | 1,52E-03 | 1,79E-02 |
| GLRX5 | 4896,17 | 0,23 | 0,07 | 3,17 | 1,51E-03 | 1,79E-02 |
| SEC62 | 4254,98 | 0,13 | 0,04 | 3,17 | 1,52E-03 | 1,79E-02 |
| RFESD | 170,64 | 0,12 | 0,04 | 3,17 | 1,53E-03 | 1,80E-02 |
| TTF1 | 773,64 | -0,06 | 0,02 | -3,17 | 1,54E-03 | 1,81E-02 |
| CCDC28A | 834,47 | -0,07 | 0,02 | -3,17 | 1,54E-03 | 1,81E-02 |
| TARBP1 | 1577,91 | 0,07 | 0,02 | 3,17 | 1,55E-03 | 1,81E-02 |
| LSM7 | 525,47 | 0,12 | 0,04 | 3,17 | 1,55E-03 | 1,81E-02 |
| CDC34 | 5673,04 | 0,21 | 0,07 | 3,17 | 1,55E-03 | 1,81E-02 |
| SGPL1 | 1534,85 | -0,07 | 0,02 | -3,17 | 1,55E-03 | 1,81E-02 |
| ATRAID | 910,66 | 0,07 | 0,02 | 3,16 | 1,56E-03 | 1,82E-02 |
| PRKAR2A | 1481,36 | -0,04 | 0,01 | -3,16 | 1,56E-03 | 1,82E-02 |
| PIH1D1 | 745,44 | 0,06 | 0,02 | 3,16 | 1,56E-03 | 1,82E-02 |
| VPS13B | 10558,27 | -0,07 | 0,02 | -3,16 | 1,57E-03 | 1,82E-02 |
| FUT4 | 657,90 | 0,07 | 0,02 | 3,16 | 1,57E-03 | 1,82E-02 |
| YIPF2 | 338,52 | 0,07 | 0,02 | 3,16 | 1,57E-03 | 1,82E-02 |
| ATG7 | 3019,58 | -0,10 | 0,03 | -3,16 | 1,57E-03 | 1,82E-02 |
| TRMT2A | 536,66 | 0,07 | 0,02 | 3,16 | 1,57E-03 | 1,83E-02 |
| GNG5 | 322,09 | -0,09 | 0,03 | -3,16 | 1,58E-03 | 1,83E-02 |
| DGKE | 1243,75 | 0,09 | 0,03 | 3,16 | 1,60E-03 | 1,85E-02 |
| RBM22 | 897,38 | -0,05 | 0,02 | -3,15 | 1,63E-03 | 1,88E-02 |
| BLTP1 | 15019,16 | -0,09 | 0,03 | -3,15 | 1,63E-03 | 1,88E-02 |
| SETD2 | 4778,91 | -0,05 | 0,02 | -3,15 | 1,63E-03 | 1,88E-02 |
| FARSA | 285,86 | 0,08 | 0,02 | 3,15 | 1,63E-03 | 1,88E-02 |
| MASTL | 459,26 | 0,11 | 0,03 | 3,15 | 1,64E-03 | 1,89E-02 |
| ASB7 | 1110,90 | -0,05 | 0,02 | -3,15 | 1,64E-03 | 1,89E-02 |
| MYC | 509,00 | 0,10 | 0,03 | 3,15 | 1,65E-03 | 1,90E-02 |
| KLHL8 | 1566,19 | -0,09 | 0,03 | -3,15 | 1,65E-03 | 1,90E-02 |
| MXD3 | 1049,33 | -0,13 | 0,04 | -3,15 | 1,66E-03 | 1,91E-02 |
| ALG8 | 409,59 | 0,05 | 0,02 | 3,14 | 1,67E-03 | 1,91E-02 |
| COP1 | 7595,72 | -0,09 | 0,03 | -3,14 | 1,67E-03 | 1,91E-02 |

| | | | | | | |
|---|---|---|---|---|---|---|
| RAB22A | 1906,59 | -0,05 | 0,02 | -3,14 | 1,67E-03 | 1,92E-02 |
| MINDY1 | 1810,81 | -0,15 | 0,05 | -3,14 | 1,68E-03 | 1,92E-02 |
| KLHL42 | 898,68 | 0,06 | 0,02 | 3,14 | 1,68E-03 | 1,92E-02 |
| KAT2B | 6861,75 | 0,13 | 0,04 | 3,14 | 1,68E-03 | 1,92E-02 |
| FGFBP3 | 116,21 | 0,10 | 0,03 | 3,14 | 1,69E-03 | 1,92E-02 |
| ERLIN2 | 914,77 | 0,04 | 0,01 | 3,14 | 1,69E-03 | 1,92E-02 |
| FAM120A | 3838,91 | -0,08 | 0,02 | -3,14 | 1,69E-03 | 1,92E-02 |
| HRH2 | 2438,84 | -0,15 | 0,05 | -3,14 | 1,69E-03 | 1,92E-02 |
| SLC45A4 | 3843,06 | -0,15 | 0,05 | -3,14 | 1,69E-03 | 1,92E-02 |
| CCDC6 | 1161,21 | 0,06 | 0,02 | 3,14 | 1,70E-03 | 1,92E-02 |
| RPL31 | 3197,86 | 0,31 | 0,10 | 3,14 | 1,72E-03 | 1,94E-02 |
| LILRA2 | 2174,82 | -0,14 | 0,04 | -3,14 | 1,72E-03 | 1,94E-02 |
| PPOX | 408,36 | 0,07 | 0,02 | 3,13 | 1,72E-03 | 1,95E-02 |
| CHMP2A | 1288,52 | -0,10 | 0,03 | -3,13 | 1,73E-03 | 1,95E-02 |
| POLR1D | 2113,86 | 0,10 | 0,03 | 3,13 | 1,72E-03 | 1,95E-02 |
| POP5 | 350,94 | 0,08 | 0,03 | 3,13 | 1,73E-03 | 1,95E-02 |
| FKTN | 690,22 | 0,08 | 0,02 | 3,13 | 1,73E-03 | 1,95E-02 |
| NDST1 | 1656,32 | -0,18 | 0,06 | -3,13 | 1,74E-03 | 1,95E-02 |
| SPOP | 2827,30 | -0,05 | 0,02 | -3,13 | 1,74E-03 | 1,95E-02 |
| CCDC91 | 1568,40 | 0,09 | 0,03 | 3,13 | 1,75E-03 | 1,96E-02 |
| SMCHD1 | 59732,96 | -0,12 | 0,04 | -3,13 | 1,76E-03 | 1,97E-02 |
| CD93 | 9849,36 | -0,15 | 0,05 | -3,13 | 1,76E-03 | 1,97E-02 |
| RAB35 | 1110,58 | -0,07 | 0,02 | -3,13 | 1,77E-03 | 1,98E-02 |
| FCGR1BP | 730,77 | -0,21 | 0,07 | -3,12 | 1,78E-03 | 1,99E-02 |
| RASSF5 | 5703,56 | -0,08 | 0,03 | -3,12 | 1,79E-03 | 2,00E-02 |
| PRMT6 | 488,07 | 0,10 | 0,03 | 3,12 | 1,79E-03 | 2,00E-02 |
| ACP1 | 1826,31 | 0,10 | 0,03 | 3,12 | 1,80E-03 | 2,00E-02 |
| MAP9 | 230,91 | 0,16 | 0,05 | 3,12 | 1,80E-03 | 2,01E-02 |
| PPFIA1 | 1867,72 | -0,10 | 0,03 | -3,12 | 1,81E-03 | 2,01E-02 |
| STAT5B | 5926,33 | -0,11 | 0,04 | -3,12 | 1,81E-03 | 2,01E-02 |
| RRP15 | 459,11 | 0,09 | 0,03 | 3,12 | 1,81E-03 | 2,01E-02 |
| MICOS13 | 266,09 | 0,10 | 0,03 | 3,12 | 1,82E-03 | 2,02E-02 |
| NFIX | 315,13 | 0,22 | 0,07 | 3,12 | 1,82E-03 | 2,02E-02 |
| NUP88 | 749,11 | 0,08 | 0,02 | 3,12 | 1,83E-03 | 2,02E-02 |
| PRMT5 | 507,49 | -0,08 | 0,03 | -3,12 | 1,83E-03 | 2,02E-02 |
| NREP | 237,13 | 0,09 | 0,03 | 3,12 | 1,84E-03 | 2,03E-02 |
| MLKL | 2477,06 | -0,10 | 0,03 | -3,11 | 1,86E-03 | 2,05E-02 |
| GAB1 | 1842,99 | -0,10 | 0,03 | -3,11 | 1,86E-03 | 2,05E-02 |
| RIT1 | 2833,18 | -0,10 | 0,03 | -3,11 | 1,86E-03 | 2,05E-02 |
| CCNB1IP1 | 372,63 | 0,09 | 0,03 | 3,11 | 1,88E-03 | 2,06E-02 |
| DNAAF11 | 437,02 | -0,19 | 0,06 | -3,11 | 1,88E-03 | 2,07E-02 |
| MUTYH | 296,20 | 0,06 | 0,02 | 3,11 | 1,89E-03 | 2,07E-02 |
| LANCL1 | 1077,30 | 0,08 | 0,03 | 3,11 | 1,89E-03 | 2,08E-02 |
| ZNF429 | 646,30 | -0,13 | 0,04 | -3,10 | 1,91E-03 | 2,08E-02 |
| PNP | 1617,85 | 0,10 | 0,03 | 3,10 | 1,91E-03 | 2,08E-02 |
| VAV3 | 2741,21 | -0,07 | 0,02 | -3,10 | 1,91E-03 | 2,08E-02 |

| | | | | | | |
|---|---|---|---|---|---|---|
| UQCC4 | 109,50 | 0,09 | 0,03 | 3,10 | 1,90E-03 | 2,08E-02 |
| LRPAP1 | 2950,30 | -0,07 | 0,02 | -3,10 | 1,91E-03 | 2,08E-02 |
| VCP | 4739,80 | -0,08 | 0,02 | -3,10 | 1,91E-03 | 2,08E-02 |
| HSH2D | 1634,97 | -0,16 | 0,05 | -3,10 | 1,92E-03 | 2,09E-02 |
| TBC1D7 | 543,06 | -0,09 | 0,03 | -3,10 | 1,92E-03 | 2,09E-02 |
| SHISA5 | 2118,05 | -0,13 | 0,04 | -3,10 | 1,93E-03 | 2,09E-02 |
| ANKH | 1597,05 | 0,14 | 0,04 | 3,10 | 1,93E-03 | 2,10E-02 |
| THAP7 | 413,13 | 0,07 | 0,02 | 3,10 | 1,95E-03 | 2,11E-02 |
| KICS2 | 399,94 | 0,07 | 0,02 | 3,10 | 1,95E-03 | 2,11E-02 |
| UBE2E3 | 483,91 | 0,08 | 0,03 | 3,10 | 1,96E-03 | 2,12E-02 |
| GOLGB1 | 4720,31 | -0,07 | 0,02 | -3,10 | 1,96E-03 | 2,12E-02 |
| TRIM56 | 5081,93 | -0,08 | 0,03 | -3,09 | 1,97E-03 | 2,13E-02 |
| ARSG | 1740,99 | -0,10 | 0,03 | -3,09 | 1,98E-03 | 2,13E-02 |
| GPX7 | 139,91 | 0,10 | 0,03 | 3,09 | 1,97E-03 | 2,13E-02 |
| GAS2L3 | 88,81 | 0,16 | 0,05 | 3,09 | 1,98E-03 | 2,13E-02 |
| TANC2 | 1425,41 | -0,10 | 0,03 | -3,09 | 1,98E-03 | 2,13E-02 |
| KMT5B | 3071,98 | -0,06 | 0,02 | -3,09 | 1,99E-03 | 2,14E-02 |
| MX1 | 3691,06 | -0,28 | 0,09 | -3,09 | 2,01E-03 | 2,15E-02 |
| MTF1 | 1291,38 | -0,10 | 0,03 | -3,09 | 2,01E-03 | 2,15E-02 |
| TRPM6 | 1423,98 | -0,15 | 0,05 | -3,09 | 2,01E-03 | 2,15E-02 |
| AGBL3 | 215,23 | 0,13 | 0,04 | 3,09 | 2,00E-03 | 2,15E-02 |
| AMN1 | 616,75 | -0,08 | 0,03 | -3,09 | 2,01E-03 | 2,15E-02 |
| RPS8 | 10022,82 | 0,16 | 0,05 | 3,09 | 2,01E-03 | 2,15E-02 |
| PRKAR1A | 11381,76 | -0,10 | 0,03 | -3,09 | 2,01E-03 | 2,15E-02 |
| DHRS9 | 1072,43 | -0,17 | 0,06 | -3,09 | 2,02E-03 | 2,15E-02 |
| SLC1A5 | 1350,96 | 0,20 | 0,07 | 3,09 | 2,02E-03 | 2,15E-02 |
| FASTK | 590,10 | 0,08 | 0,03 | 3,09 | 2,03E-03 | 2,16E-02 |
| DMAC1 | 593,65 | 0,13 | 0,04 | 3,09 | 2,03E-03 | 2,16E-02 |
| UBE2R2 | 1822,85 | -0,10 | 0,03 | -3,08 | 2,04E-03 | 2,17E-02 |
| SLC16A7 | 3090,23 | 0,10 | 0,03 | 3,08 | 2,04E-03 | 2,17E-02 |
| CAMKK2 | 2892,95 | -0,12 | 0,04 | -3,08 | 2,04E-03 | 2,17E-02 |
| EI24 | 598,01 | 0,05 | 0,01 | 3,08 | 2,05E-03 | 2,17E-02 |
| TLE3 | 5499,04 | -0,14 | 0,04 | -3,08 | 2,05E-03 | 2,17E-02 |
| MARCHF5 | 759,17 | -0,06 | 0,02 | -3,08 | 2,05E-03 | 2,17E-02 |
| TENT5C | 25809,46 | 0,25 | 0,08 | 3,08 | 2,05E-03 | 2,17E-02 |
| RPS3A | 6026,16 | 0,32 | 0,10 | 3,08 | 2,06E-03 | 2,17E-02 |
| SMAD5 | 1516,13 | 0,10 | 0,03 | 3,08 | 2,06E-03 | 2,17E-02 |
| SYNJ1 | 2088,48 | -0,07 | 0,02 | -3,08 | 2,07E-03 | 2,18E-02 |
| HCK | 12051,88 | -0,11 | 0,04 | -3,08 | 2,07E-03 | 2,18E-02 |
| PPID | 380,69 | -0,06 | 0,02 | -3,08 | 2,07E-03 | 2,18E-02 |
| GPR15 | 183,83 | 0,24 | 0,08 | 3,08 | 2,08E-03 | 2,18E-02 |
| IRAG2 | 6162,21 | -0,09 | 0,03 | -3,08 | 2,08E-03 | 2,18E-02 |
| GALNT14 | 171,30 | -0,23 | 0,07 | -3,08 | 2,08E-03 | 2,18E-02 |
| AKAP10 | 2735,28 | -0,05 | 0,02 | -3,08 | 2,09E-03 | 2,19E-02 |
| ATF6 | 4948,12 | -0,08 | 0,02 | -3,08 | 2,09E-03 | 2,19E-02 |
| ZNF789 | 489,91 | 0,06 | 0,02 | 3,08 | 2,09E-03 | 2,19E-02 |

| | | | | | | |
|---|---|---|---|---|---|---|
| RPS6 | 12074,33 | 0,18 | 0,06 | 3,08 | 2,09E-03 | 2,19E-02 |
| NIT2 | 664,73 | 0,06 | 0,02 | 3,08 | 2,10E-03 | 2,19E-02 |
| FFAR2 | 2925,98 | -0,15 | 0,05 | -3,08 | 2,10E-03 | 2,19E-02 |
| QARS1 | 1930,46 | 0,05 | 0,02 | 3,07 | 2,11E-03 | 2,20E-02 |
| SHARPIN | 572,33 | 0,14 | 0,04 | 3,07 | 2,13E-03 | 2,22E-02 |
| MTMR4 | 2015,47 | -0,06 | 0,02 | -3,07 | 2,14E-03 | 2,23E-02 |
| SON | 14204,41 | -0,05 | 0,02 | -3,07 | 2,15E-03 | 2,23E-02 |
| SDF2 | 1222,24 | -0,07 | 0,02 | -3,07 | 2,17E-03 | 2,25E-02 |
| DVL1 | 543,19 | 0,06 | 0,02 | 3,07 | 2,17E-03 | 2,25E-02 |
| RPL10 | 15931,62 | 0,10 | 0,03 | 3,06 | 2,18E-03 | 2,26E-02 |
| TPT1 | 11970,56 | 0,21 | 0,07 | 3,06 | 2,20E-03 | 2,27E-02 |
| ARID3A | 1462,30 | -0,13 | 0,04 | -3,06 | 2,20E-03 | 2,27E-02 |
| MPG | 521,96 | 0,06 | 0,02 | 3,06 | 2,20E-03 | 2,27E-02 |
| FBXO4 | 277,88 | 0,12 | 0,04 | 3,06 | 2,20E-03 | 2,27E-02 |
| VASP | 5595,50 | -0,11 | 0,04 | -3,06 | 2,20E-03 | 2,27E-02 |
| USP14 | 862,10 | 0,08 | 0,03 | 3,06 | 2,22E-03 | 2,28E-02 |
| PDCD5 | 428,37 | 0,07 | 0,02 | 3,06 | 2,23E-03 | 2,29E-02 |
| PPM1N | 95,45 | 0,12 | 0,04 | 3,06 | 2,24E-03 | 2,29E-02 |
| WBP1L | 2021,24 | -0,07 | 0,02 | -3,06 | 2,24E-03 | 2,29E-02 |
| CBX1 | 1472,28 | -0,09 | 0,03 | -3,06 | 2,24E-03 | 2,29E-02 |
| LGALS3 | 1414,63 | 0,17 | 0,06 | 3,06 | 2,23E-03 | 2,29E-02 |
| NABP2 | 121,40 | 0,07 | 0,02 | 3,06 | 2,24E-03 | 2,29E-02 |
| STX11 | 2015,36 | -0,13 | 0,04 | -3,06 | 2,23E-03 | 2,29E-02 |
| RPS17 | 302,85 | 0,06 | 0,02 | 3,06 | 2,24E-03 | 2,29E-02 |
| THAP11 | 562,78 | 0,06 | 0,02 | 3,06 | 2,24E-03 | 2,29E-02 |
| BACH1 | 15452,98 | -0,10 | 0,03 | -3,06 | 2,25E-03 | 2,29E-02 |
| TMCO3 | 2011,19 | -0,08 | 0,03 | -3,05 | 2,26E-03 | 2,31E-02 |
| RSBN1L | 3860,30 | -0,07 | 0,02 | -3,05 | 2,27E-03 | 2,31E-02 |
| PGAM5 | 353,04 | 0,06 | 0,02 | 3,05 | 2,27E-03 | 2,31E-02 |
| DHRS12 | 485,11 | -0,13 | 0,04 | -3,05 | 2,28E-03 | 2,31E-02 |
| REX1BD | 690,00 | 0,07 | 0,02 | 3,05 | 2,28E-03 | 2,32E-02 |
| ABCC5 | 2887,29 | -0,11 | 0,04 | -3,05 | 2,30E-03 | 2,33E-02 |
| ZNF18 | 893,71 | -0,05 | 0,02 | -3,05 | 2,30E-03 | 2,33E-02 |
| ZFAND4 | 1049,68 | 0,07 | 0,02 | 3,05 | 2,30E-03 | 2,33E-02 |
| FRAT2 | 8403,84 | -0,12 | 0,04 | -3,05 | 2,31E-03 | 2,33E-02 |
| TMEM91 | 703,34 | -0,11 | 0,03 | -3,05 | 2,31E-03 | 2,34E-02 |
| RAB7A | 1084,99 | -0,13 | 0,04 | -3,05 | 2,32E-03 | 2,34E-02 |
| MPV17 | 732,32 | 0,05 | 0,02 | 3,05 | 2,32E-03 | 2,34E-02 |
| TCF3 | 516,99 | 0,13 | 0,04 | 3,05 | 2,32E-03 | 2,34E-02 |
| CMAS | 537,34 | 0,11 | 0,04 | 3,04 | 2,33E-03 | 2,35E-02 |
| C6orf120 | 937,46 | 0,10 | 0,03 | 3,04 | 2,34E-03 | 2,35E-02 |
| ENTR1 | 514,24 | 0,06 | 0,02 | 3,04 | 2,35E-03 | 2,36E-02 |
| SLC38A6 | 528,24 | 0,08 | 0,03 | 3,04 | 2,36E-03 | 2,36E-02 |
| NMI | 2558,43 | -0,11 | 0,04 | -3,04 | 2,36E-03 | 2,36E-02 |
| PACRGL | 433,22 | 0,07 | 0,02 | 3,04 | 2,36E-03 | 2,36E-02 |
| PLSCR1 | 3501,72 | -0,18 | 0,06 | -3,04 | 2,37E-03 | 2,37E-02 |

| | | | | | | |
|---|---|---|---|---|---|---|
| MED28 | 5253,76 | -0,05 | 0,01 | -3,04 | 2,37E-03 | 2,37E-02 |
| DCUN1D1 | 4581,97 | 0,14 | 0,05 | 3,04 | 2,39E-03 | 2,39E-02 |
| UBR5 | 7634,42 | -0,05 | 0,02 | -3,04 | 2,40E-03 | 2,40E-02 |
| BICD2 | 3126,21 | -0,08 | 0,03 | -3,04 | 2,40E-03 | 2,40E-02 |
| RNF168 | 1562,38 | -0,06 | 0,02 | -3,03 | 2,42E-03 | 2,41E-02 |
| G6PC3 | 126,96 | 0,11 | 0,04 | 3,03 | 2,44E-03 | 2,43E-02 |
| PPP2R2D | 1967,01 | -0,05 | 0,02 | -3,03 | 2,46E-03 | 2,45E-02 |
| MAP10 | 114,64 | 0,14 | 0,05 | 3,03 | 2,47E-03 | 2,45E-02 |
| RACK1 | 14301,02 | 0,08 | 0,03 | 3,03 | 2,48E-03 | 2,47E-02 |
| BRAP | 1404,69 | -0,05 | 0,02 | -3,03 | 2,48E-03 | 2,47E-02 |
| NDUFB7 | 549,64 | 0,10 | 0,03 | 3,02 | 2,49E-03 | 2,47E-02 |
| ISCA1 | 2737,90 | 0,17 | 0,05 | 3,02 | 2,50E-03 | 2,47E-02 |
| PGS1 | 4249,85 | -0,12 | 0,04 | -3,02 | 2,49E-03 | 2,47E-02 |
| BRWD3 | 4616,25 | -0,08 | 0,03 | -3,02 | 2,50E-03 | 2,47E-02 |
| MRPS16 | 1384,75 | 0,06 | 0,02 | 3,02 | 2,51E-03 | 2,48E-02 |
| NECAP1 | 1774,44 | -0,04 | 0,01 | -3,02 | 2,52E-03 | 2,49E-02 |
| H1-10 | 548,35 | 0,13 | 0,04 | 3,02 | 2,52E-03 | 2,49E-02 |
| MRPL34 | 281,31 | 0,08 | 0,03 | 3,02 | 2,52E-03 | 2,49E-02 |
| PPP1R35 | 329,94 | 0,08 | 0,03 | 3,02 | 2,53E-03 | 2,49E-02 |
| VMA21 | 1359,72 | 0,09 | 0,03 | 3,02 | 2,54E-03 | 2,49E-02 |
| CD48 | 5711,62 | 0,11 | 0,04 | 3,02 | 2,53E-03 | 2,49E-02 |
| HERPUD1 | 2015,59 | -0,06 | 0,02 | -3,02 | 2,53E-03 | 2,49E-02 |
| RAD23A | 317,20 | 0,19 | 0,06 | 3,02 | 2,55E-03 | 2,50E-02 |
| UBE2L3 | 1068,07 | -0,05 | 0,02 | -3,02 | 2,54E-03 | 2,50E-02 |
| NCF1 | 4059,95 | -0,14 | 0,05 | -3,01 | 2,57E-03 | 2,52E-02 |
| MRPL27 | 425,06 | 0,07 | 0,02 | 3,01 | 2,57E-03 | 2,52E-02 |
| ZYG11B | 2427,53 | -0,07 | 0,02 | -3,01 | 2,58E-03 | 2,52E-02 |
| FARSB | 593,31 | 0,06 | 0,02 | 3,01 | 2,60E-03 | 2,54E-02 |
| GNA12 | 585,91 | 0,17 | 0,06 | 3,01 | 2,62E-03 | 2,55E-02 |
| GZMK | 710,79 | 0,21 | 0,07 | 3,01 | 2,62E-03 | 2,56E-02 |
| ZNF333 | 1398,02 | -0,07 | 0,02 | -3,01 | 2,62E-03 | 2,56E-02 |
| HBA2 | 79,15 | 0,30 | 0,10 | 3,01 | 2,65E-03 | 2,58E-02 |
| UQCC3 | 181,40 | 0,08 | 0,03 | 3,01 | 2,65E-03 | 2,58E-02 |
| SLC22A1 | 332,29 | -0,18 | 0,06 | -3,00 | 2,67E-03 | 2,60E-02 |
| SIGLEC9 | 1535,98 | -0,13 | 0,04 | -3,00 | 2,68E-03 | 2,60E-02 |
| TPR | 8599,60 | -0,08 | 0,03 | -3,00 | 2,68E-03 | 2,60E-02 |
| TAGAP | 5311,51 | -0,08 | 0,03 | -3,00 | 2,68E-03 | 2,60E-02 |
| TFR2 | 94,21 | 0,21 | 0,07 | 3,00 | 2,69E-03 | 2,61E-02 |
| WDR12 | 451,26 | 0,05 | 0,02 | 3,00 | 2,71E-03 | 2,62E-02 |
| TXN2 | 152,12 | 0,09 | 0,03 | 3,00 | 2,73E-03 | 2,64E-02 |
| KIAA0319 | 286,81 | -0,18 | 0,06 | -3,00 | 2,74E-03 | 2,65E-02 |
| SDHAF2 | 593,56 | -0,06 | 0,02 | -2,99 | 2,76E-03 | 2,65E-02 |
| ATP5MC2 | 1455,73 | 0,06 | 0,02 | 2,99 | 2,75E-03 | 2,65E-02 |
| SOX6 | 834,92 | 0,20 | 0,07 | 2,99 | 2,78E-03 | 2,65E-02 |
| LYRM1 | 643,25 | -0,06 | 0,02 | -2,99 | 2,78E-03 | 2,65E-02 |
| RAB3D | 3036,70 | -0,15 | 0,05 | -2,99 | 2,76E-03 | 2,65E-02 |

| | | | | | | |
|---|---|---|---|---|---|---|
| CSF3R | 33398,10 | -0,15 | 0,05 | -2,99 | 2,77E-03 | 2,65E-02 |
| PNPLA1 | 118,02 | -0,15 | 0,05 | -2,99 | 2,75E-03 | 2,65E-02 |
| F11R | 715,06 | -0,15 | 0,05 | -2,99 | 2,77E-03 | 2,65E-02 |
| ITGA5 | 4174,14 | -0,10 | 0,04 | -2,99 | 2,77E-03 | 2,65E-02 |
| MPHOSPH8 | 2926,29 | -0,04 | 0,01 | -2,99 | 2,76E-03 | 2,65E-02 |
| TOMM40 | 282,90 | 0,08 | 0,03 | 2,99 | 2,77E-03 | 2,65E-02 |
| MTLN | 124,53 | 0,11 | 0,04 | 2,99 | 2,76E-03 | 2,65E-02 |
| AAR2 | 246,63 | 0,06 | 0,02 | 2,99 | 2,77E-03 | 2,65E-02 |
| ZNF846 | 345,10 | 0,07 | 0,02 | 2,99 | 2,78E-03 | 2,65E-02 |
| GEMIN8 | 273,71 | 0,06 | 0,02 | 2,99 | 2,80E-03 | 2,66E-02 |
| RILP | 1349,35 | 0,19 | 0,06 | 2,99 | 2,80E-03 | 2,66E-02 |
| SELPLG | 4535,11 | -0,10 | 0,03 | -2,99 | 2,80E-03 | 2,66E-02 |
| BCL2L1 | 12713,38 | 0,21 | 0,07 | 2,99 | 2,83E-03 | 2,67E-02 |
| DEGS1 | 1569,45 | -0,07 | 0,02 | -2,99 | 2,82E-03 | 2,67E-02 |
| HDAC9 | 1629,27 | 0,12 | 0,04 | 2,99 | 2,82E-03 | 2,67E-02 |
| ZNF555 | 416,86 | 0,09 | 0,03 | 2,99 | 2,83E-03 | 2,67E-02 |
| MICU1 | 1074,37 | -0,07 | 0,02 | -2,99 | 2,83E-03 | 2,67E-02 |
| RPS6KA1 | 4976,12 | -0,09 | 0,03 | -2,99 | 2,82E-03 | 2,67E-02 |
| ATP6V1E1 | 1764,75 | -0,08 | 0,03 | -2,99 | 2,83E-03 | 2,67E-02 |
| DENND3 | 12445,51 | -0,15 | 0,05 | -2,99 | 2,83E-03 | 2,67E-02 |
| CTDP1 | 2379,23 | -0,07 | 0,02 | -2,99 | 2,81E-03 | 2,67E-02 |
| SC5D | 409,28 | 0,11 | 0,04 | 2,99 | 2,83E-03 | 2,67E-02 |
| RALGAPB | 5261,09 | -0,05 | 0,02 | -2,98 | 2,85E-03 | 2,69E-02 |
| TOR1AIP1 | 3389,71 | -0,06 | 0,02 | -2,98 | 2,85E-03 | 2,69E-02 |
| PADI2 | 1938,29 | -0,22 | 0,07 | -2,98 | 2,86E-03 | 2,69E-02 |
| LPCAT1 | 2317,04 | -0,10 | 0,03 | -2,98 | 2,86E-03 | 2,69E-02 |
| C5orf24 | 1220,52 | 0,08 | 0,03 | 2,98 | 2,88E-03 | 2,71E-02 |
| CUL4A | 3125,80 | 0,06 | 0,02 | 2,98 | 2,89E-03 | 2,72E-02 |
| C1QBP | 932,64 | 0,08 | 0,03 | 2,98 | 2,89E-03 | 2,72E-02 |
| SFT2D3 | 575,87 | 0,09 | 0,03 | 2,98 | 2,90E-03 | 2,72E-02 |
| CASP8 | 9520,50 | -0,07 | 0,02 | -2,98 | 2,91E-03 | 2,72E-02 |
| CD36 | 5088,01 | 0,13 | 0,05 | 2,98 | 2,91E-03 | 2,73E-02 |
| OXSM | 180,75 | 0,08 | 0,03 | 2,98 | 2,92E-03 | 2,73E-02 |
| CR1L | 355,84 | 0,19 | 0,07 | 2,98 | 2,93E-03 | 2,74E-02 |
| HSPBAP1 | 1845,33 | -0,08 | 0,03 | -2,97 | 2,93E-03 | 2,74E-02 |
| CACNA1E | 391,12 | -0,21 | 0,07 | -2,97 | 2,94E-03 | 2,74E-02 |
| ZNF639 | 756,63 | 0,08 | 0,03 | 2,97 | 2,94E-03 | 2,74E-02 |
| CAPNS2 | 206,82 | -0,14 | 0,05 | -2,97 | 2,95E-03 | 2,74E-02 |
| DDX49 | 568,71 | 0,05 | 0,02 | 2,97 | 2,95E-03 | 2,74E-02 |
| PWWP2A | 1855,92 | -0,04 | 0,01 | -2,97 | 2,95E-03 | 2,75E-02 |
| PSME3 | 1532,04 | -0,09 | 0,03 | -2,97 | 2,97E-03 | 2,75E-02 |
| CCDC93 | 3546,09 | -0,05 | 0,02 | -2,97 | 3,01E-03 | 2,79E-02 |
| PPP1R15B | 5172,45 | -0,06 | 0,02 | -2,97 | 3,02E-03 | 2,80E-02 |
| GOLGA6L9 | 136,23 | 0,14 | 0,05 | 2,96 | 3,03E-03 | 2,81E-02 |
| RNF182 | 494,66 | 0,46 | 0,16 | 2,96 | 3,04E-03 | 2,81E-02 |
| TMEM9B | 1287,16 | 0,08 | 0,03 | 2,96 | 3,04E-03 | 2,81E-02 |

| | | | | | | |
|---|---|---|---|---|---|---|
| TRIB1 | 778,99 | -0,10 | 0,03 | -2,96 | 3,04E-03 | 2,81E-02 |
| RIPOR2 | 27036,95 | -0,08 | 0,03 | -2,96 | 3,05E-03 | 2,81E-02 |
| RFX7 | 1189,57 | 0,09 | 0,03 | 2,96 | 3,05E-03 | 2,81E-02 |
| GSTCD | 253,80 | 0,08 | 0,03 | 2,96 | 3,06E-03 | 2,82E-02 |
| LRRC8C | 2373,38 | 0,08 | 0,03 | 2,96 | 3,07E-03 | 2,82E-02 |
| KREMEN1 | 638,48 | -0,22 | 0,07 | -2,96 | 3,07E-03 | 2,82E-02 |
| EYS | 91,42 | -0,18 | 0,06 | -2,96 | 3,08E-03 | 2,83E-02 |
| SDHAF4 | 217,84 | 0,07 | 0,02 | 2,96 | 3,08E-03 | 2,83E-02 |
| ARRDC4 | 551,77 | -0,14 | 0,05 | -2,96 | 3,08E-03 | 2,83E-02 |
| GCNT2 | 504,35 | 0,10 | 0,03 | 2,96 | 3,11E-03 | 2,85E-02 |
| GLE1 | 1344,42 | -0,04 | 0,01 | -2,96 | 3,11E-03 | 2,85E-02 |
| SLC6A8 | 934,37 | 0,28 | 0,09 | 2,96 | 3,12E-03 | 2,86E-02 |
| SPIN4 | 172,56 | 0,09 | 0,03 | 2,95 | 3,15E-03 | 2,86E-02 |
| DYNLT1 | 1987,71 | -0,10 | 0,03 | -2,95 | 3,14E-03 | 2,86E-02 |
| ZNF367 | 196,77 | 0,10 | 0,03 | 2,95 | 3,14E-03 | 2,86E-02 |
| TGOLN2 | 8182,44 | -0,09 | 0,03 | -2,95 | 3,15E-03 | 2,86E-02 |
| ABHD2 | 8676,06 | -0,13 | 0,04 | -2,95 | 3,15E-03 | 2,86E-02 |
| HBP1 | 6211,73 | -0,09 | 0,03 | -2,95 | 3,14E-03 | 2,86E-02 |
| MTX2 | 326,94 | 0,10 | 0,03 | 2,95 | 3,13E-03 | 2,86E-02 |
| MIA3 | 2860,88 | -0,05 | 0,02 | -2,95 | 3,15E-03 | 2,86E-02 |
| TP53I13 | 592,84 | 0,08 | 0,03 | 2,95 | 3,16E-03 | 2,87E-02 |
| PRRC1 | 1234,67 | 0,06 | 0,02 | 2,95 | 3,17E-03 | 2,87E-02 |
| DPY19L4 | 637,20 | 0,10 | 0,03 | 2,95 | 3,19E-03 | 2,89E-02 |
| H3-3B | 29288,29 | -0,10 | 0,03 | -2,95 | 3,21E-03 | 2,91E-02 |
| GOLPH3 | 2428,03 | -0,05 | 0,02 | -2,95 | 3,21E-03 | 2,91E-02 |
| NMNAT1 | 465,73 | -0,07 | 0,02 | -2,95 | 3,22E-03 | 2,91E-02 |
| EDEM2 | 1367,18 | -0,06 | 0,02 | -2,95 | 3,22E-03 | 2,91E-02 |
| ZNF558 | 450,78 | 0,07 | 0,03 | 2,95 | 3,22E-03 | 2,91E-02 |
| MCUB | 1499,64 | 0,07 | 0,02 | 2,94 | 3,23E-03 | 2,92E-02 |
| NR2C2 | 3501,67 | -0,08 | 0,03 | -2,94 | 3,24E-03 | 2,93E-02 |
| TLR5 | 1015,71 | -0,15 | 0,05 | -2,94 | 3,27E-03 | 2,94E-02 |
| BYSL | 104,81 | 0,08 | 0,03 | 2,94 | 3,27E-03 | 2,94E-02 |
| SIRPD | 165,65 | -0,14 | 0,05 | -2,94 | 3,27E-03 | 2,95E-02 |
| TFB2M | 207,88 | 0,09 | 0,03 | 2,94 | 3,28E-03 | 2,95E-02 |
| C8orf33 | 561,12 | 0,04 | 0,02 | 2,94 | 3,29E-03 | 2,95E-02 |
| ZNHIT2 | 113,82 | 0,07 | 0,02 | 2,94 | 3,29E-03 | 2,96E-02 |
| DDX3X | 24072,80 | -0,08 | 0,03 | -2,94 | 3,30E-03 | 2,96E-02 |
| ZNF263 | 1025,05 | 0,04 | 0,01 | 2,94 | 3,32E-03 | 2,97E-02 |
| TIGD5 | 252,51 | 0,08 | 0,03 | 2,94 | 3,33E-03 | 2,98E-02 |
| NOXRED1 | 99,40 | -0,08 | 0,03 | -2,94 | 3,33E-03 | 2,98E-02 |
| FASTKD3 | 277,87 | 0,07 | 0,02 | 2,93 | 3,35E-03 | 2,99E-02 |
| IGIP | 669,49 | 0,09 | 0,03 | 2,93 | 3,35E-03 | 2,99E-02 |
| EEF1B2 | 1634,78 | 0,22 | 0,08 | 2,93 | 3,36E-03 | 3,00E-02 |
| MTCH2 | 712,88 | 0,05 | 0,02 | 2,93 | 3,37E-03 | 3,00E-02 |
| TAF6L | 241,40 | 0,06 | 0,02 | 2,93 | 3,38E-03 | 3,01E-02 |
| MANBA | 4229,47 | -0,08 | 0,03 | -2,93 | 3,39E-03 | 3,01E-02 |

| | | | | | | |
|---|---|---|---|---|---|---|
| SLC8A1 | 4144,72 | -0,12 | 0,04 | -2,93 | 3,39E-03 | 3,02E-02 |
| KNL1 | 252,11 | 0,16 | 0,06 | 2,93 | 3,41E-03 | 3,03E-02 |
| DMTN | 5176,43 | 0,24 | 0,08 | 2,93 | 3,41E-03 | 3,03E-02 |
| FUT7 | 1196,86 | -0,12 | 0,04 | -2,93 | 3,42E-03 | 3,03E-02 |
| TESK2 | 430,18 | -0,07 | 0,02 | -2,93 | 3,42E-03 | 3,04E-02 |
| CLCN4 | 448,02 | -0,17 | 0,06 | -2,93 | 3,44E-03 | 3,05E-02 |
| THRAP3 | 5987,24 | -0,07 | 0,02 | -2,92 | 3,45E-03 | 3,05E-02 |
| DCAF12 | 28785,75 | 0,21 | 0,07 | 2,92 | 3,46E-03 | 3,06E-02 |
| ZNF606 | 596,58 | 0,06 | 0,02 | 2,92 | 3,46E-03 | 3,06E-02 |
| RCOR1 | 2271,84 | -0,10 | 0,03 | -2,92 | 3,47E-03 | 3,07E-02 |
| FAHD1 | 470,95 | 0,11 | 0,04 | 2,92 | 3,49E-03 | 3,08E-02 |
| CDK8 | 434,69 | 0,07 | 0,02 | 2,92 | 3,51E-03 | 3,08E-02 |
| LYRM9 | 228,85 | 0,06 | 0,02 | 2,92 | 3,51E-03 | 3,08E-02 |
| FAF1 | 1156,72 | 0,05 | 0,02 | 2,92 | 3,50E-03 | 3,08E-02 |
| DTD2 | 234,29 | 0,07 | 0,03 | 2,92 | 3,50E-03 | 3,08E-02 |
| GCHFR | 182,00 | 0,09 | 0,03 | 2,92 | 3,50E-03 | 3,08E-02 |
| PLIN3 | 1128,18 | -0,10 | 0,03 | -2,92 | 3,51E-03 | 3,08E-02 |
| SOCS6 | 233,74 | 0,10 | 0,04 | 2,92 | 3,50E-03 | 3,08E-02 |
| RAC1 | 2942,29 | -0,07 | 0,03 | -2,92 | 3,51E-03 | 3,08E-02 |
| VTI1A | 2620,27 | -0,06 | 0,02 | -2,92 | 3,52E-03 | 3,08E-02 |
| CPSF6 | 3237,31 | 0,12 | 0,04 | 2,92 | 3,52E-03 | 3,08E-02 |
| RBPJ | 5472,37 | -0,08 | 0,03 | -2,92 | 3,53E-03 | 3,09E-02 |
| LPP | 6171,65 | -0,07 | 0,03 | -2,92 | 3,55E-03 | 3,10E-02 |
| MARCHF2 | 2431,86 | 0,16 | 0,05 | 2,92 | 3,56E-03 | 3,11E-02 |
| SAP30BP | 1950,94 | -0,05 | 0,02 | -2,91 | 3,56E-03 | 3,11E-02 |
| GOLGA7B | 234,99 | 0,11 | 0,04 | 2,91 | 3,56E-03 | 3,11E-02 |
| ENSG000002 | 906,20 | -0,07 | 0,02 | -2,91 | 3,57E-03 | 3,11E-02 |
| PGBD4 | 304,82 | 0,11 | 0,04 | 2,91 | 3,58E-03 | 3,12E-02 |
| SPRED1 | 356,80 | 0,15 | 0,05 | 2,91 | 3,59E-03 | 3,12E-02 |
| RRAGC | 473,08 | -0,06 | 0,02 | -2,91 | 3,60E-03 | 3,12E-02 |
| C11orf71 | 309,12 | 0,10 | 0,03 | 2,91 | 3,59E-03 | 3,12E-02 |
| PPP3CA | 3403,99 | -0,07 | 0,02 | -2,91 | 3,60E-03 | 3,13E-02 |
| ZNF43 | 794,68 | 0,07 | 0,02 | 2,91 | 3,60E-03 | 3,13E-02 |
| ZFP3 | 269,09 | 0,09 | 0,03 | 2,91 | 3,62E-03 | 3,14E-02 |
| PARPBP | 145,43 | 0,12 | 0,04 | 2,91 | 3,62E-03 | 3,14E-02 |
| TMEM245 | 2195,89 | 0,07 | 0,02 | 2,91 | 3,63E-03 | 3,14E-02 |
| DENND4A | 6733,17 | 0,08 | 0,03 | 2,91 | 3,64E-03 | 3,15E-02 |
| PPP1R3D | 1193,88 | -0,08 | 0,03 | -2,91 | 3,64E-03 | 3,15E-02 |
| ALG3 | 175,99 | 0,06 | 0,02 | 2,91 | 3,65E-03 | 3,16E-02 |
| DUSP22 | 2347,70 | -0,06 | 0,02 | -2,91 | 3,67E-03 | 3,16E-02 |
| CANT1 | 1402,89 | -0,12 | 0,04 | -2,91 | 3,67E-03 | 3,16E-02 |
| NSUN7 | 349,77 | -0,14 | 0,05 | -2,90 | 3,69E-03 | 3,17E-02 |
| PLOD1 | 625,93 | -0,12 | 0,04 | -2,90 | 3,69E-03 | 3,17E-02 |
| ZFYVE27 | 1964,22 | -0,05 | 0,02 | -2,90 | 3,71E-03 | 3,18E-02 |
| IFITM1 | 3258,91 | -0,13 | 0,04 | -2,90 | 3,71E-03 | 3,18E-02 |
| NHLRC2 | 2093,01 | 0,07 | 0,03 | 2,90 | 3,70E-03 | 3,18E-02 |

| | | | | | | |
|---|---|---|---|---|---|---|
| TDRD7 | 1400,48 | -0,10 | 0,03 | -2,90 | 3,71E-03 | 3,18E-02 |
| FAHD2A | 300,44 | 0,07 | 0,02 | 2,90 | 3,71E-03 | 3,18E-02 |
| SCOC | 352,86 | 0,18 | 0,06 | 2,90 | 3,72E-03 | 3,18E-02 |
| XRN2 | 4456,25 | -0,06 | 0,02 | -2,90 | 3,71E-03 | 3,18E-02 |
| HADHB | 2370,90 | -0,05 | 0,02 | -2,90 | 3,72E-03 | 3,18E-02 |
| FAM20B | 1424,60 | 0,07 | 0,03 | 2,90 | 3,76E-03 | 3,21E-02 |
| CD46 | 15126,72 | -0,09 | 0,03 | -2,90 | 3,76E-03 | 3,21E-02 |
| TRIM35 | 860,21 | 0,05 | 0,02 | 2,90 | 3,77E-03 | 3,21E-02 |
| RPL35A | 4045,56 | 0,19 | 0,06 | 2,90 | 3,77E-03 | 3,21E-02 |
| ICAM4 | 200,95 | 0,15 | 0,05 | 2,90 | 3,78E-03 | 3,22E-02 |
| ZC3H11A | 120,02 | -0,07 | 0,02 | -2,89 | 3,80E-03 | 3,24E-02 |
| PFDN1 | 167,23 | 0,06 | 0,02 | 2,89 | 3,81E-03 | 3,24E-02 |
| MCRIP1 | 371,74 | 0,06 | 0,02 | 2,89 | 3,81E-03 | 3,24E-02 |
| ANKRD13C | 821,44 | 0,08 | 0,03 | 2,89 | 3,82E-03 | 3,24E-02 |
| NGRN | 767,03 | 0,05 | 0,02 | 2,89 | 3,82E-03 | 3,24E-02 |
| CLASP1 | 3839,22 | -0,07 | 0,02 | -2,89 | 3,83E-03 | 3,25E-02 |
| PUS1 | 444,02 | 0,07 | 0,02 | 2,89 | 3,84E-03 | 3,25E-02 |
| COMMD9 | 657,65 | 0,04 | 0,02 | 2,89 | 3,84E-03 | 3,25E-02 |
| BTN2A1 | 1499,25 | -0,10 | 0,03 | -2,89 | 3,85E-03 | 3,26E-02 |
| ADGRE5 | 9217,59 | -0,13 | 0,04 | -2,89 | 3,86E-03 | 3,26E-02 |
| DRC12 | 109,06 | -0,14 | 0,05 | -2,89 | 3,87E-03 | 3,27E-02 |
| EEF1E1 | 119,37 | 0,12 | 0,04 | 2,89 | 3,88E-03 | 3,27E-02 |
| TNFRSF9 | 394,22 | -0,13 | 0,05 | -2,89 | 3,87E-03 | 3,27E-02 |
| RHAG | 124,80 | 0,20 | 0,07 | 2,89 | 3,89E-03 | 3,28E-02 |
| NIPSNAP2 | 817,62 | 0,05 | 0,02 | 2,89 | 3,89E-03 | 3,28E-02 |
| TLE5 | 1166,03 | 0,11 | 0,04 | 2,89 | 3,90E-03 | 3,28E-02 |
| NCAM1 | 507,50 | -0,16 | 0,05 | -2,89 | 3,91E-03 | 3,29E-02 |
| ZNF714 | 550,22 | 0,08 | 0,03 | 2,89 | 3,91E-03 | 3,29E-02 |
| SLC25A39 | 10213,96 | 0,21 | 0,07 | 2,88 | 3,92E-03 | 3,29E-02 |
| CYB5R4 | 6492,85 | -0,09 | 0,03 | -2,88 | 3,93E-03 | 3,30E-02 |
| RCBTB2 | 3067,85 | -0,08 | 0,03 | -2,88 | 3,94E-03 | 3,31E-02 |
| MTX1 | 1172,80 | -0,11 | 0,04 | -2,88 | 3,97E-03 | 3,31E-02 |
| TSPAN4 | 177,60 | 0,12 | 0,04 | 2,88 | 3,96E-03 | 3,31E-02 |
| AMD1 | 5410,32 | -0,08 | 0,03 | -2,88 | 3,97E-03 | 3,31E-02 |
| SNTB2 | 1567,49 | -0,06 | 0,02 | -2,88 | 3,97E-03 | 3,31E-02 |
| ENSG000002 | 171,86 | -0,17 | 0,06 | -2,88 | 3,97E-03 | 3,31E-02 |
| SHLD2 | 772,59 | 0,12 | 0,04 | 2,88 | 3,95E-03 | 3,31E-02 |
| GIGYF2 | 4740,27 | -0,05 | 0,02 | -2,88 | 3,96E-03 | 3,31E-02 |
| RPS11 | 19466,63 | 0,13 | 0,04 | 2,88 | 4,00E-03 | 3,33E-02 |
| PRCP | 2681,94 | -0,07 | 0,02 | -2,88 | 4,00E-03 | 3,33E-02 |
| USP3 | 6538,77 | -0,08 | 0,03 | -2,88 | 4,01E-03 | 3,33E-02 |
| EIF1AX | 1099,83 | 0,12 | 0,04 | 2,88 | 4,02E-03 | 3,35E-02 |
| ETV3 | 1887,81 | -0,04 | 0,01 | -2,87 | 4,05E-03 | 3,36E-02 |
| DPY19L2 | 84,61 | 0,20 | 0,07 | 2,87 | 4,05E-03 | 3,36E-02 |
| ZNF652 | 4437,82 | -0,08 | 0,03 | -2,87 | 4,05E-03 | 3,36E-02 |
| CCS | 922,49 | 0,10 | 0,03 | 2,87 | 4,09E-03 | 3,38E-02 |

| | | | | | | |
|---|---|---|---|---|---|---|
| SLC25A14 | 264,08 | -0,06 | 0,02 | -2,87 | 4,08E-03 | 3,38E-02 |
| RAPH1 | 230,88 | 0,13 | 0,04 | 2,87 | 4,08E-03 | 3,38E-02 |
| ELMO1 | 5082,23 | -0,08 | 0,03 | -2,87 | 4,10E-03 | 3,39E-02 |
| NIPSNAP3B | 89,20 | 0,14 | 0,05 | 2,87 | 4,13E-03 | 3,41E-02 |
| LRRC58 | 2077,02 | 0,08 | 0,03 | 2,87 | 4,13E-03 | 3,41E-02 |
| LGALS2 | 486,81 | 0,25 | 0,09 | 2,87 | 4,14E-03 | 3,42E-02 |
| CRIP1 | 263,24 | 0,09 | 0,03 | 2,87 | 4,14E-03 | 3,42E-02 |
| MTHFD2L | 500,50 | 0,10 | 0,03 | 2,87 | 4,15E-03 | 3,42E-02 |
| PSME3IP1 | 3170,28 | -0,04 | 0,02 | -2,87 | 4,15E-03 | 3,42E-02 |
| YDJC | 267,59 | 0,07 | 0,03 | 2,87 | 4,16E-03 | 3,42E-02 |
| MYG1 | 417,24 | 0,07 | 0,03 | 2,87 | 4,16E-03 | 3,42E-02 |
| GNG2 | 3237,41 | -0,07 | 0,02 | -2,86 | 4,17E-03 | 3,42E-02 |
| FKBP8 | 22293,10 | 0,20 | 0,07 | 2,87 | 4,17E-03 | 3,42E-02 |
| NAF1 | 452,48 | 0,07 | 0,02 | 2,86 | 4,17E-03 | 3,42E-02 |
| TALDO1 | 11007,99 | -0,11 | 0,04 | -2,86 | 4,20E-03 | 3,44E-02 |
| TP53INP1 | 6252,48 | -0,09 | 0,03 | -2,86 | 4,21E-03 | 3,44E-02 |
| SEC14L1 | 10120,51 | -0,14 | 0,05 | -2,86 | 4,20E-03 | 3,44E-02 |
| PCYT2 | 306,17 | 0,07 | 0,02 | 2,86 | 4,23E-03 | 3,46E-02 |
| HNRNPH2 | 2483,17 | -0,13 | 0,04 | -2,86 | 4,23E-03 | 3,46E-02 |
| ALG10 | 577,29 | 0,08 | 0,03 | 2,86 | 4,23E-03 | 3,46E-02 |
| PICALM | 21215,32 | -0,09 | 0,03 | -2,86 | 4,25E-03 | 3,47E-02 |
| C12orf57 | 872,90 | 0,14 | 0,05 | 2,86 | 4,25E-03 | 3,47E-02 |
| PHTF2 | 1988,94 | 0,10 | 0,03 | 2,86 | 4,27E-03 | 3,48E-02 |
| KBTBD2 | 3908,63 | -0,07 | 0,03 | -2,86 | 4,28E-03 | 3,49E-02 |
| SRGAP2B | 413,45 | -0,09 | 0,03 | -2,86 | 4,29E-03 | 3,49E-02 |
| SPART | 1603,31 | -0,08 | 0,03 | -2,86 | 4,29E-03 | 3,49E-02 |
| RSAD2 | 2403,91 | -0,37 | 0,13 | -2,86 | 4,29E-03 | 3,49E-02 |
| ST13 | 1992,06 | 0,10 | 0,03 | 2,86 | 4,30E-03 | 3,49E-02 |
| PBX2 | 3585,41 | -0,10 | 0,03 | -2,85 | 4,31E-03 | 3,50E-02 |
| NAT9 | 455,67 | 0,05 | 0,02 | 2,85 | 4,32E-03 | 3,51E-02 |
| ZNF658 | 168,26 | 0,14 | 0,05 | 2,85 | 4,37E-03 | 3,54E-02 |
| MYB | 251,06 | 0,10 | 0,04 | 2,85 | 4,37E-03 | 3,54E-02 |
| SECTM1 | 1725,52 | -0,15 | 0,05 | -2,85 | 4,38E-03 | 3,54E-02 |
| SNRNP35 | 744,52 | -0,04 | 0,01 | -2,85 | 4,42E-03 | 3,56E-02 |
| SPG11 | 8755,44 | -0,06 | 0,02 | -2,85 | 4,41E-03 | 3,56E-02 |
| SUSD3 | 433,20 | 0,08 | 0,03 | 2,85 | 4,41E-03 | 3,56E-02 |
| CCM2 | 4286,00 | -0,06 | 0,02 | -2,85 | 4,41E-03 | 3,56E-02 |
| ANTKMT | 328,87 | 0,07 | 0,03 | 2,85 | 4,40E-03 | 3,56E-02 |
| DNAJC19 | 741,73 | 0,10 | 0,04 | 2,85 | 4,41E-03 | 3,56E-02 |
| USP48 | 3575,85 | -0,05 | 0,02 | -2,85 | 4,43E-03 | 3,56E-02 |
| MTG2 | 612,27 | -0,11 | 0,04 | -2,85 | 4,43E-03 | 3,56E-02 |
| ST3GAL4 | 690,43 | -0,14 | 0,05 | -2,85 | 4,44E-03 | 3,57E-02 |
| GZMB | 1252,92 | -0,15 | 0,05 | -2,84 | 4,45E-03 | 3,57E-02 |
| PTMS | 168,93 | 0,19 | 0,07 | 2,84 | 4,46E-03 | 3,57E-02 |
| P2RY1 | 208,12 | 0,12 | 0,04 | 2,84 | 4,46E-03 | 3,57E-02 |
| ZNF736 | 2055,86 | 0,08 | 0,03 | 2,84 | 4,47E-03 | 3,58E-02 |

| | | | | | | |
|---|---|---|---|---|---|---|
| TDRD3 | 856,48 | 0,07 | 0,03 | 2,84 | 4,48E-03 | 3,59E-02 |
| TNFRSF10A | 972,70 | 0,07 | 0,02 | 2,84 | 4,48E-03 | 3,59E-02 |
| IL23A | 140,81 | 0,12 | 0,04 | 2,84 | 4,49E-03 | 3,59E-02 |
| RPSA | 9975,42 | 0,13 | 0,05 | 2,84 | 4,50E-03 | 3,60E-02 |
| C18orf32 | 340,48 | -0,05 | 0,02 | -2,84 | 4,53E-03 | 3,62E-02 |
| SP140 | 2903,65 | -0,10 | 0,03 | -2,84 | 4,55E-03 | 3,63E-02 |
| RPL13A | 30723,38 | 0,13 | 0,05 | 2,84 | 4,55E-03 | 3,63E-02 |
| AMACR | 148,73 | -0,10 | 0,04 | -2,84 | 4,55E-03 | 3,63E-02 |
| STAMBP | 1331,94 | -0,04 | 0,01 | -2,84 | 4,56E-03 | 3,63E-02 |
| ZFAND1 | 627,49 | 0,13 | 0,05 | 2,84 | 4,56E-03 | 3,63E-02 |
| ADGRE2 | 3235,90 | -0,14 | 0,05 | -2,84 | 4,58E-03 | 3,64E-02 |
| PMVK | 235,15 | 0,07 | 0,02 | 2,84 | 4,58E-03 | 3,64E-02 |
| PRMT1 | 450,49 | 0,06 | 0,02 | 2,83 | 4,58E-03 | 3,64E-02 |
| H3C10 | 2495,25 | 0,15 | 0,05 | 2,83 | 4,59E-03 | 3,64E-02 |
| WDR82 | 6102,95 | -0,05 | 0,02 | -2,83 | 4,59E-03 | 3,64E-02 |
| RPS14 | 24820,54 | 0,15 | 0,05 | 2,83 | 4,61E-03 | 3,65E-02 |
| CD58 | 1496,39 | -0,08 | 0,03 | -2,83 | 4,62E-03 | 3,65E-02 |
| MBTPS2 | 352,66 | 0,07 | 0,02 | 2,83 | 4,63E-03 | 3,65E-02 |
| KIAA1586 | 611,96 | 0,16 | 0,06 | 2,83 | 4,62E-03 | 3,65E-02 |
| ARIH1 | 4275,71 | -0,04 | 0,01 | -2,83 | 4,62E-03 | 3,65E-02 |
| TMEM267 | 382,98 | 0,09 | 0,03 | 2,83 | 4,64E-03 | 3,66E-02 |
| FKBP15 | 4017,71 | -0,07 | 0,02 | -2,83 | 4,66E-03 | 3,67E-02 |
| PHC2 | 2279,55 | -0,17 | 0,06 | -2,83 | 4,66E-03 | 3,67E-02 |
| KCMF1 | 2272,84 | -0,04 | 0,02 | -2,83 | 4,66E-03 | 3,67E-02 |
| SPTB | 1755,47 | 0,26 | 0,09 | 2,83 | 4,67E-03 | 3,67E-02 |
| PPP2R5B | 571,04 | 0,14 | 0,05 | 2,83 | 4,67E-03 | 3,67E-02 |
| RNASEH1 | 367,67 | 0,04 | 0,01 | 2,83 | 4,68E-03 | 3,68E-02 |
| DDX17 | 51038,33 | -0,05 | 0,02 | -2,83 | 4,70E-03 | 3,69E-02 |
| LRRC40 | 570,80 | 0,11 | 0,04 | 2,83 | 4,71E-03 | 3,69E-02 |
| SLC16A3 | 8597,65 | -0,11 | 0,04 | -2,83 | 4,70E-03 | 3,69E-02 |
| KCTD20 | 3815,22 | -0,06 | 0,02 | -2,83 | 4,70E-03 | 3,69E-02 |
| EEPD1 | 779,88 | -0,13 | 0,05 | -2,83 | 4,71E-03 | 3,69E-02 |
| RPL19 | 10563,34 | 0,10 | 0,03 | 2,83 | 4,71E-03 | 3,69E-02 |
| THEMIS | 2195,86 | 0,12 | 0,04 | 2,83 | 4,72E-03 | 3,69E-02 |
| NTAN1 | 430,47 | 0,10 | 0,03 | 2,83 | 4,72E-03 | 3,69E-02 |
| WDR11 | 3008,30 | 0,07 | 0,02 | 2,82 | 4,74E-03 | 3,70E-02 |
| KBTBD6 | 517,92 | 0,09 | 0,03 | 2,82 | 4,74E-03 | 3,70E-02 |
| MVP | 6167,95 | -0,08 | 0,03 | -2,82 | 4,75E-03 | 3,71E-02 |
| CALCOCO2 | 2208,50 | -0,08 | 0,03 | -2,82 | 4,76E-03 | 3,71E-02 |
| ADGRE3 | 2645,03 | -0,12 | 0,04 | -2,82 | 4,76E-03 | 3,71E-02 |
| OR52K1 | 359,98 | -0,18 | 0,06 | -2,82 | 4,77E-03 | 3,71E-02 |
| CEACAM1 | 528,30 | -0,15 | 0,05 | -2,82 | 4,79E-03 | 3,72E-02 |
| MRPL20 | 671,70 | 0,06 | 0,02 | 2,82 | 4,79E-03 | 3,72E-02 |
| DUS3L | 394,82 | 0,07 | 0,02 | 2,82 | 4,79E-03 | 3,72E-02 |
| PGM2 | 2197,29 | -0,08 | 0,03 | -2,82 | 4,80E-03 | 3,73E-02 |
| ARMC2 | 376,68 | -0,08 | 0,03 | -2,82 | 4,81E-03 | 3,73E-02 |

| | | | | | | |
|---|---|---|---|---|---|---|
| LPCAT3 | 510,95 | -0,08 | 0,03 | -2,82 | 4,82E-03 | 3,74E-02 |
| CRKL | 3978,02 | -0,06 | 0,02 | -2,82 | 4,86E-03 | 3,76E-02 |
| ELMO2 | 1513,02 | -0,06 | 0,02 | -2,82 | 4,86E-03 | 3,76E-02 |
| RNF213 | 29619,33 | -0,15 | 0,05 | -2,82 | 4,86E-03 | 3,76E-02 |
| ANKRD44 | 23520,16 | -0,06 | 0,02 | -2,81 | 4,94E-03 | 3,79E-02 |
| BNIP1 | 106,17 | 0,08 | 0,03 | 2,81 | 4,95E-03 | 3,79E-02 |
| ANKS1A | 1093,07 | -0,10 | 0,03 | -2,81 | 4,94E-03 | 3,79E-02 |
| EWSR1 | 6724,82 | -0,04 | 0,01 | -2,81 | 4,92E-03 | 3,79E-02 |
| SAAL1 | 310,44 | 0,06 | 0,02 | 2,81 | 4,93E-03 | 3,79E-02 |
| KDM2A | 5322,89 | -0,08 | 0,03 | -2,81 | 4,94E-03 | 3,79E-02 |
| CDKL1 | 506,21 | 0,09 | 0,03 | 2,81 | 4,93E-03 | 3,79E-02 |
| BABAM2 | 607,59 | -0,07 | 0,02 | -2,81 | 4,93E-03 | 3,79E-02 |
| BCAS3 | 1926,75 | -0,06 | 0,02 | -2,81 | 4,94E-03 | 3,79E-02 |
| MRPS26 | 355,83 | 0,06 | 0,02 | 2,81 | 4,94E-03 | 3,79E-02 |
| NDUFAF3 | 597,18 | 0,06 | 0,02 | 2,81 | 4,94E-03 | 3,79E-02 |
| CSF2RB | 7869,03 | -0,12 | 0,04 | -2,81 | 4,94E-03 | 3,79E-02 |
| UBE4B | 1937,59 | -0,09 | 0,03 | -2,81 | 4,95E-03 | 3,79E-02 |
| ADK | 922,65 | 0,09 | 0,03 | 2,81 | 4,95E-03 | 3,79E-02 |
| CCDC47 | 1375,84 | -0,06 | 0,02 | -2,81 | 5,00E-03 | 3,83E-02 |
| TCEANC2 | 1063,55 | -0,03 | 0,01 | -2,81 | 5,01E-03 | 3,83E-02 |
| HECA | 6316,02 | -0,06 | 0,02 | -2,81 | 5,01E-03 | 3,83E-02 |
| LRRFIP2 | 2533,30 | -0,06 | 0,02 | -2,80 | 5,04E-03 | 3,85E-02 |
| PRXL2C | 314,71 | 0,07 | 0,02 | 2,80 | 5,09E-03 | 3,88E-02 |
| ZXDB | 265,67 | 0,07 | 0,03 | 2,80 | 5,09E-03 | 3,88E-02 |
| NCBP3 | 2034,56 | -0,06 | 0,02 | -2,80 | 5,09E-03 | 3,88E-02 |
| ZNF143 | 1379,49 | -0,06 | 0,02 | -2,80 | 5,11E-03 | 3,89E-02 |
| XAF1 | 3145,25 | -0,18 | 0,07 | -2,80 | 5,12E-03 | 3,89E-02 |
| NCF4 | 1668,77 | -0,13 | 0,05 | -2,80 | 5,12E-03 | 3,89E-02 |
| ZNF449 | 347,69 | 0,08 | 0,03 | 2,80 | 5,12E-03 | 3,89E-02 |
| EYA3 | 1073,57 | -0,06 | 0,02 | -2,80 | 5,15E-03 | 3,91E-02 |
| TPM3 | 13119,22 | -0,07 | 0,02 | -2,80 | 5,16E-03 | 3,91E-02 |
| DGLUCY | 5111,60 | -0,11 | 0,04 | -2,80 | 5,16E-03 | 3,92E-02 |
| ARPC1A | 280,17 | -0,05 | 0,02 | -2,80 | 5,19E-03 | 3,93E-02 |
| ACSL3 | 1459,33 | -0,07 | 0,02 | -2,79 | 5,20E-03 | 3,94E-02 |
| MLF2 | 1280,03 | -0,08 | 0,03 | -2,79 | 5,21E-03 | 3,94E-02 |
| CHRNA10 | 115,13 | -0,12 | 0,04 | -2,79 | 5,21E-03 | 3,94E-02 |
| ROGDI | 422,60 | 0,13 | 0,05 | 2,79 | 5,23E-03 | 3,95E-02 |
| TMEM18 | 455,08 | 0,04 | 0,02 | 2,79 | 5,26E-03 | 3,97E-02 |
| ETAA1 | 480,26 | 0,16 | 0,06 | 2,79 | 5,26E-03 | 3,97E-02 |
| ATXN1 | 5474,31 | -0,09 | 0,03 | -2,79 | 5,27E-03 | 3,98E-02 |
| CXCL16 | 546,59 | -0,14 | 0,05 | -2,79 | 5,28E-03 | 3,98E-02 |
| PFKFB2 | 1024,86 | -0,10 | 0,04 | -2,79 | 5,29E-03 | 3,99E-02 |
| OS9 | 2176,34 | -0,09 | 0,03 | -2,79 | 5,30E-03 | 3,99E-02 |
| ERI2 | 188,46 | 0,08 | 0,03 | 2,79 | 5,31E-03 | 3,99E-02 |
| CHD7 | 3044,38 | -0,07 | 0,03 | -2,79 | 5,32E-03 | 3,99E-02 |
| DYNLT2 | 96,53 | 0,13 | 0,05 | 2,79 | 5,32E-03 | 3,99E-02 |

| | | | | | | |
|---|---|---|---|---|---|---|
| RAB21 | 3507,95 | -0,05 | 0,02 | -2,79 | 5,32E-03 | 3,99E-02 |
| HEATR5A | 1028,55 | -0,07 | 0,02 | -2,79 | 5,33E-03 | 4,00E-02 |
| TCERG1 | 2195,52 | 0,07 | 0,02 | 2,79 | 5,34E-03 | 4,00E-02 |
| ACVR2A | 482,02 | 0,09 | 0,03 | 2,79 | 5,34E-03 | 4,00E-02 |
| WIPF2 | 1266,89 | -0,11 | 0,04 | -2,79 | 5,35E-03 | 4,01E-02 |
| FKBP5 | 3622,13 | -0,13 | 0,05 | -2,78 | 5,37E-03 | 4,02E-02 |
| RPL36A | 177,16 | 0,07 | 0,02 | 2,78 | 5,38E-03 | 4,02E-02 |
| POLR2M | 684,86 | 0,11 | 0,04 | 2,78 | 5,39E-03 | 4,02E-02 |
| POC1B | 1249,70 | 0,08 | 0,03 | 2,78 | 5,39E-03 | 4,02E-02 |
| RNASEH2C | 790,29 | 0,06 | 0,02 | 2,78 | 5,40E-03 | 4,02E-02 |
| PCGF1 | 144,54 | 0,05 | 0,02 | 2,78 | 5,40E-03 | 4,02E-02 |
| VAPB | 1672,61 | -0,06 | 0,02 | -2,78 | 5,40E-03 | 4,03E-02 |
| POLD2 | 125,22 | 0,10 | 0,04 | 2,78 | 5,41E-03 | 4,03E-02 |
| NAALADL1 | 155,80 | 0,08 | 0,03 | 2,78 | 5,42E-03 | 4,04E-02 |
| NIPSNAP3A | 412,31 | 0,13 | 0,05 | 2,78 | 5,44E-03 | 4,05E-02 |
| CTSH | 1459,07 | 0,08 | 0,03 | 2,78 | 5,45E-03 | 4,05E-02 |
| MTMR11 | 332,11 | 0,10 | 0,03 | 2,78 | 5,46E-03 | 4,05E-02 |
| PRKAB2 | 1057,84 | 0,05 | 0,02 | 2,78 | 5,48E-03 | 4,07E-02 |
| HIF1AN | 2833,60 | -0,08 | 0,03 | -2,78 | 5,48E-03 | 4,07E-02 |
| NUFIP2 | 9900,85 | -0,06 | 0,02 | -2,78 | 5,49E-03 | 4,07E-02 |
| VDR | 333,94 | -0,13 | 0,05 | -2,78 | 5,49E-03 | 4,07E-02 |
| SIRPB2 | 2430,95 | -0,16 | 0,06 | -2,78 | 5,50E-03 | 4,07E-02 |
| FAM120B | 2240,41 | -0,07 | 0,03 | -2,78 | 5,50E-03 | 4,07E-02 |
| MAPRE1 | 3520,90 | -0,06 | 0,02 | -2,78 | 5,51E-03 | 4,07E-02 |
| ENSG00000２ | 103,24 | -0,14 | 0,05 | -2,78 | 5,50E-03 | 4,07E-02 |
| BCL6-AS1 | 120,73 | -0,21 | 0,08 | -2,78 | 5,51E-03 | 4,07E-02 |
| PLEKHG4 | 191,54 | 0,12 | 0,04 | 2,78 | 5,51E-03 | 4,07E-02 |
| UROD | 696,60 | 0,07 | 0,03 | 2,77 | 5,53E-03 | 4,07E-02 |
| ATP11A | 4080,80 | -0,12 | 0,04 | -2,77 | 5,54E-03 | 4,08E-02 |
| RXYLT1 | 292,19 | 0,07 | 0,02 | 2,77 | 5,54E-03 | 4,08E-02 |
| BPNT2 | 1848,20 | 0,06 | 0,02 | 2,77 | 5,55E-03 | 4,08E-02 |
| RWDD2B | 87,06 | 0,09 | 0,03 | 2,77 | 5,56E-03 | 4,08E-02 |
| H2AJ | 867,50 | 0,12 | 0,04 | 2,77 | 5,57E-03 | 4,09E-02 |
| UBE2G2 | 3039,77 | 0,04 | 0,02 | 2,77 | 5,61E-03 | 4,11E-02 |
| CREB3L4 | 105,49 | 0,07 | 0,03 | 2,77 | 5,61E-03 | 4,11E-02 |
| CRNKL1 | 2118,92 | -0,07 | 0,03 | -2,77 | 5,60E-03 | 4,11E-02 |
| MRPL14 | 310,00 | 0,07 | 0,03 | 2,77 | 5,60E-03 | 4,11E-02 |
| ADIPOR2 | 1537,19 | -0,04 | 0,02 | -2,77 | 5,61E-03 | 4,11E-02 |
| RPL30 | 9567,20 | 0,17 | 0,06 | 2,77 | 5,60E-03 | 4,11E-02 |
| TMEM223 | 238,27 | 0,07 | 0,03 | 2,77 | 5,63E-03 | 4,11E-02 |
| RPP30 | 561,14 | 0,06 | 0,02 | 2,77 | 5,63E-03 | 4,11E-02 |
| GOLGA2 | 1054,59 | -0,08 | 0,03 | -2,77 | 5,62E-03 | 4,11E-02 |
| PAOX | 175,45 | 0,08 | 0,03 | 2,77 | 5,64E-03 | 4,12E-02 |
| RPL34 | 2097,17 | 0,27 | 0,10 | 2,77 | 5,67E-03 | 4,13E-02 |
| IK | 3120,13 | -0,07 | 0,02 | -2,77 | 5,67E-03 | 4,13E-02 |
| MPZL1 | 3018,15 | -0,13 | 0,05 | -2,76 | 5,70E-03 | 4,15E-02 |

| | | | | | | |
|---|---|---|---|---|---|---|
| EXT1 | 819,41 | -0,07 | 0,03 | -2,76 | 5,71E-03 | 4,16E-02 |
| RNF34 | 2237,89 | -0,04 | 0,01 | -2,76 | 5,74E-03 | 4,17E-02 |
| WASL | 649,06 | 0,09 | 0,03 | 2,76 | 5,74E-03 | 4,17E-02 |
| HIVEP1 | 2121,50 | -0,06 | 0,02 | -2,76 | 5,73E-03 | 4,17E-02 |
| TMEM160 | 178,41 | 0,07 | 0,03 | 2,76 | 5,75E-03 | 4,17E-02 |
| MRPS15 | 305,57 | 0,06 | 0,02 | 2,76 | 5,75E-03 | 4,17E-02 |
| GPR180 | 540,13 | 0,11 | 0,04 | 2,76 | 5,76E-03 | 4,17E-02 |
| TMEM128 | 397,07 | 0,08 | 0,03 | 2,76 | 5,77E-03 | 4,17E-02 |
| NAA38 | 518,72 | 0,09 | 0,03 | 2,76 | 5,76E-03 | 4,17E-02 |
| BEND7 | 97,83 | -0,22 | 0,08 | -2,76 | 5,77E-03 | 4,17E-02 |
| BPGM | 3744,31 | 0,21 | 0,08 | 2,76 | 5,78E-03 | 4,18E-02 |
| C2orf69 | 420,27 | 0,10 | 0,03 | 2,76 | 5,78E-03 | 4,18E-02 |
| USF3 | 2797,60 | -0,06 | 0,02 | -2,76 | 5,78E-03 | 4,18E-02 |
| ITPA | 573,04 | 0,05 | 0,02 | 2,76 | 5,80E-03 | 4,19E-02 |
| RHNO1 | 275,44 | 0,07 | 0,02 | 2,76 | 5,81E-03 | 4,19E-02 |
| HNRNPUL2 | 665,32 | -0,05 | 0,02 | -2,76 | 5,82E-03 | 4,19E-02 |
| AASDH | 644,94 | 0,08 | 0,03 | 2,76 | 5,82E-03 | 4,19E-02 |
| RNF187 | 490,09 | 0,16 | 0,06 | 2,76 | 5,83E-03 | 4,20E-02 |
| AP5B1 | 2643,08 | -0,16 | 0,06 | -2,76 | 5,85E-03 | 4,21E-02 |
| USP19 | 2350,84 | -0,07 | 0,03 | -2,75 | 5,91E-03 | 4,24E-02 |
| DDX28 | 339,70 | 0,05 | 0,02 | 2,75 | 5,91E-03 | 4,24E-02 |
| ERN1 | 3261,39 | -0,11 | 0,04 | -2,75 | 5,92E-03 | 4,24E-02 |
| CCDC124 | 135,03 | 0,11 | 0,04 | 2,75 | 5,91E-03 | 4,24E-02 |
| CEACAM4 | 1109,68 | -0,14 | 0,05 | -2,75 | 5,91E-03 | 4,24E-02 |
| PJA2 | 8047,01 | -0,08 | 0,03 | -2,75 | 5,90E-03 | 4,24E-02 |
| ELF1 | 7242,57 | -0,06 | 0,02 | -2,75 | 5,91E-03 | 4,24E-02 |
| SLC49A4 | 1384,50 | -0,08 | 0,03 | -2,75 | 5,92E-03 | 4,24E-02 |
| C4orf46 | 167,24 | 0,07 | 0,03 | 2,75 | 5,94E-03 | 4,25E-02 |
| PHF23 | 1010,11 | -0,06 | 0,02 | -2,75 | 5,97E-03 | 4,27E-02 |
| KLHDC2 | 1575,76 | 0,08 | 0,03 | 2,75 | 5,98E-03 | 4,28E-02 |
| METTL23 | 545,87 | 0,06 | 0,02 | 2,75 | 6,00E-03 | 4,28E-02 |
| USP45 | 586,50 | 0,10 | 0,03 | 2,75 | 6,00E-03 | 4,28E-02 |
| CFL2 | 239,58 | 0,11 | 0,04 | 2,75 | 6,02E-03 | 4,29E-02 |
| UBE4A | 3858,12 | -0,05 | 0,02 | -2,75 | 6,02E-03 | 4,29E-02 |
| PGD | 6565,87 | -0,11 | 0,04 | -2,74 | 6,05E-03 | 4,31E-02 |
| CLNS1A | 1378,72 | 0,07 | 0,02 | 2,74 | 6,07E-03 | 4,32E-02 |
| FAM177A1 | 787,99 | -0,05 | 0,02 | -2,74 | 6,07E-03 | 4,32E-02 |
| ABCC4 | 1381,43 | 0,12 | 0,04 | 2,74 | 6,08E-03 | 4,32E-02 |
| PLEK | 4412,72 | -0,09 | 0,03 | -2,74 | 6,08E-03 | 4,32E-02 |
| CD8B | 790,14 | 0,17 | 0,06 | 2,74 | 6,13E-03 | 4,33E-02 |
| NUDCD2 | 418,38 | 0,09 | 0,03 | 2,74 | 6,13E-03 | 4,33E-02 |
| GBE1 | 1099,86 | -0,08 | 0,03 | -2,74 | 6,13E-03 | 4,33E-02 |
| CASP1 | 4856,07 | -0,10 | 0,04 | -2,74 | 6,13E-03 | 4,33E-02 |
| MRPL13 | 279,30 | 0,12 | 0,05 | 2,74 | 6,11E-03 | 4,33E-02 |
| MFSD9 | 611,96 | -0,12 | 0,04 | -2,74 | 6,12E-03 | 4,33E-02 |
| CSRP1 | 821,14 | 0,04 | 0,02 | 2,74 | 6,10E-03 | 4,33E-02 |

| | | | | | | |
|---|---|---|---|---|---|---|
| GEMIN2 | 159,15 | 0,09 | 0,03 | 2,74 | 6,11E-03 | 4,33E-02 |
| TC2N | 5851,23 | 0,11 | 0,04 | 2,74 | 6,13E-03 | 4,33E-02 |
| CEP57L1 | 261,29 | 0,10 | 0,04 | 2,74 | 6,14E-03 | 4,34E-02 |
| RELL1 | 1573,27 | -0,10 | 0,04 | -2,74 | 6,17E-03 | 4,35E-02 |
| AEN | 854,81 | 0,06 | 0,02 | 2,74 | 6,19E-03 | 4,37E-02 |
| ZNF239 | 104,10 | 0,11 | 0,04 | 2,74 | 6,22E-03 | 4,38E-02 |
| ZNF682 | 198,22 | 0,08 | 0,03 | 2,74 | 6,22E-03 | 4,38E-02 |
| TMEM38B | 315,38 | 0,09 | 0,03 | 2,74 | 6,23E-03 | 4,38E-02 |
| CSKMT | 139,04 | 0,08 | 0,03 | 2,73 | 6,25E-03 | 4,39E-02 |
| MYO1F | 14946,90 | -0,13 | 0,05 | -2,73 | 6,25E-03 | 4,39E-02 |
| IGF1R | 4213,43 | -0,12 | 0,05 | -2,73 | 6,27E-03 | 4,40E-02 |
| CCNC | 1167,68 | 0,11 | 0,04 | 2,73 | 6,29E-03 | 4,41E-02 |
| KLC1 | 3210,91 | -0,08 | 0,03 | -2,73 | 6,30E-03 | 4,41E-02 |
| STOM | 4503,76 | 0,12 | 0,04 | 2,73 | 6,30E-03 | 4,41E-02 |
| LARP4 | 1115,63 | 0,08 | 0,03 | 2,73 | 6,31E-03 | 4,42E-02 |
| TM7SF2 | 203,47 | 0,07 | 0,03 | 2,73 | 6,32E-03 | 4,42E-02 |
| GLYR1 | 2169,60 | -0,05 | 0,02 | -2,73 | 6,33E-03 | 4,43E-02 |
| PHF12 | 2008,19 | -0,12 | 0,04 | -2,73 | 6,34E-03 | 4,43E-02 |
| TSG101 | 1551,10 | -0,05 | 0,02 | -2,73 | 6,36E-03 | 4,44E-02 |
| CD84 | 3314,69 | 0,06 | 0,02 | 2,73 | 6,36E-03 | 4,44E-02 |
| SLC31A2 | 2926,51 | -0,11 | 0,04 | -2,73 | 6,36E-03 | 4,44E-02 |
| PLEKHG3 | 6218,00 | -0,12 | 0,04 | -2,73 | 6,37E-03 | 4,44E-02 |
| DHX16 | 1755,35 | -0,05 | 0,02 | -2,73 | 6,38E-03 | 4,44E-02 |
| NUDT19 | 311,72 | 0,05 | 0,02 | 2,73 | 6,38E-03 | 4,44E-02 |
| GATA1 | 546,73 | 0,19 | 0,07 | 2,73 | 6,39E-03 | 4,45E-02 |
| STK19 | 333,79 | -0,05 | 0,02 | -2,73 | 6,42E-03 | 4,46E-02 |
| KBTBD3 | 230,10 | 0,11 | 0,04 | 2,73 | 6,41E-03 | 4,46E-02 |
| NAGK | 4819,52 | -0,09 | 0,03 | -2,72 | 6,44E-03 | 4,47E-02 |
| PEX10 | 203,13 | 0,06 | 0,02 | 2,72 | 6,45E-03 | 4,47E-02 |
| BRIP1 | 242,18 | 0,12 | 0,04 | 2,72 | 6,45E-03 | 4,47E-02 |
| CHPT1 | 2266,55 | 0,12 | 0,04 | 2,72 | 6,46E-03 | 4,48E-02 |
| TWSG1 | 315,35 | 0,08 | 0,03 | 2,72 | 6,48E-03 | 4,49E-02 |
| SMDT1 | 202,82 | 0,07 | 0,03 | 2,72 | 6,50E-03 | 4,50E-02 |
| TRIM58 | 13635,27 | 0,22 | 0,08 | 2,72 | 6,50E-03 | 4,50E-02 |
| SLC2A6 | 641,47 | 0,09 | 0,03 | 2,72 | 6,55E-03 | 4,52E-02 |
| HMGA1 | 188,01 | 0,14 | 0,05 | 2,72 | 6,55E-03 | 4,52E-02 |
| GDPD1 | 106,44 | 0,09 | 0,03 | 2,72 | 6,54E-03 | 4,52E-02 |
| SEPSECS | 981,08 | 0,07 | 0,03 | 2,72 | 6,57E-03 | 4,53E-02 |
| MTX3 | 938,32 | 0,08 | 0,03 | 2,72 | 6,57E-03 | 4,53E-02 |
| PYURF | 722,43 | 0,12 | 0,04 | 2,72 | 6,59E-03 | 4,54E-02 |
| CCDC85B | 507,63 | 0,08 | 0,03 | 2,72 | 6,59E-03 | 4,54E-02 |
| STYX | 1807,34 | 0,09 | 0,03 | 2,72 | 6,59E-03 | 4,54E-02 |
| PARP14 | 15900,78 | -0,14 | 0,05 | -2,72 | 6,60E-03 | 4,54E-02 |
| P2RX4 | 633,94 | 0,06 | 0,02 | 2,72 | 6,60E-03 | 4,54E-02 |
| SCMH1 | 385,12 | 0,07 | 0,02 | 2,71 | 6,63E-03 | 4,56E-02 |
| ANO9 | 451,56 | 0,12 | 0,04 | 2,71 | 6,67E-03 | 4,58E-02 |

| | | | | | | |
|---|---|---|---|---|---|---|
| H2BC18 | 2812,74 | -0,10 | 0,04 | -2,71 | 6,67E-03 | 4,58E-02 |
| BORCS7 | 375,81 | 0,12 | 0,04 | 2,71 | 6,70E-03 | 4,60E-02 |
| KIAA0753 | 800,53 | -0,06 | 0,02 | -2,71 | 6,71E-03 | 4,60E-02 |
| MRPL58 | 174,69 | 0,07 | 0,03 | 2,71 | 6,74E-03 | 4,62E-02 |
| THUMPD2 | 391,87 | 0,08 | 0,03 | 2,71 | 6,75E-03 | 4,62E-02 |
| SLC7A5 | 197,07 | 0,19 | 0,07 | 2,71 | 6,76E-03 | 4,62E-02 |
| PPIL1 | 185,81 | 0,07 | 0,03 | 2,71 | 6,76E-03 | 4,62E-02 |
| DEDD2 | 2471,92 | -0,07 | 0,03 | -2,71 | 6,76E-03 | 4,62E-02 |
| SNIP1 | 1254,95 | -0,05 | 0,02 | -2,71 | 6,77E-03 | 4,63E-02 |
| MAPK3 | 529,27 | -0,10 | 0,04 | -2,71 | 6,78E-03 | 4,63E-02 |
| MRPL32 | 494,48 | 0,13 | 0,05 | 2,71 | 6,79E-03 | 4,63E-02 |
| COL9A3 | 240,49 | -0,22 | 0,08 | -2,71 | 6,80E-03 | 4,64E-02 |
| NDUFA4 | 823,57 | 0,17 | 0,06 | 2,71 | 6,81E-03 | 4,64E-02 |
| ZFYVE1 | 710,48 | -0,07 | 0,02 | -2,71 | 6,82E-03 | 4,65E-02 |
| HPS1 | 1785,88 | 0,11 | 0,04 | 2,70 | 6,84E-03 | 4,65E-02 |
| RIMKLB | 641,00 | 0,07 | 0,03 | 2,70 | 6,87E-03 | 4,67E-02 |
| ASF1A | 828,02 | 0,11 | 0,04 | 2,70 | 6,87E-03 | 4,67E-02 |
| METRNL | 1456,09 | -0,09 | 0,03 | -2,70 | 6,89E-03 | 4,68E-02 |
| ACSL6 | 480,25 | 0,10 | 0,04 | 2,70 | 6,91E-03 | 4,69E-02 |
| SLC39A7 | 935,03 | -0,06 | 0,02 | -2,70 | 6,93E-03 | 4,70E-02 |
| IMMP1L | 373,17 | 0,09 | 0,03 | 2,70 | 6,94E-03 | 4,71E-02 |
| FBXL8 | 123,73 | 0,09 | 0,03 | 2,70 | 6,95E-03 | 4,71E-02 |
| TLR8 | 5597,44 | -0,10 | 0,04 | -2,70 | 6,97E-03 | 4,72E-02 |
| FLAD1 | 641,55 | 0,04 | 0,02 | 2,70 | 6,98E-03 | 4,72E-02 |
| NEBL | 213,20 | 0,47 | 0,17 | 2,70 | 6,99E-03 | 4,73E-02 |
| PCGF5 | 4941,59 | 0,10 | 0,04 | 2,70 | 7,01E-03 | 4,74E-02 |
| NDC1 | 683,75 | 0,06 | 0,02 | 2,70 | 7,04E-03 | 4,75E-02 |
| PAICS | 389,65 | 0,06 | 0,02 | 2,69 | 7,05E-03 | 4,76E-02 |
| PXMP4 | 394,57 | 0,04 | 0,02 | 2,69 | 7,06E-03 | 4,77E-02 |
| GOLT1B | 1046,01 | 0,12 | 0,04 | 2,69 | 7,07E-03 | 4,77E-02 |
| DOCK11 | 10489,22 | -0,06 | 0,02 | -2,69 | 7,08E-03 | 4,77E-02 |
| RNF217 | 416,24 | 0,10 | 0,04 | 2,69 | 7,09E-03 | 4,77E-02 |
| BOLA3 | 80,79 | 0,10 | 0,04 | 2,69 | 7,12E-03 | 4,79E-02 |
| EEF2 | 24685,97 | 0,07 | 0,03 | 2,69 | 7,13E-03 | 4,80E-02 |
| CLP1 | 413,66 | -0,06 | 0,02 | -2,69 | 7,14E-03 | 4,80E-02 |
| PCED1A | 571,20 | 0,06 | 0,02 | 2,69 | 7,18E-03 | 4,83E-02 |
| DYNLL1 | 693,13 | 0,08 | 0,03 | 2,69 | 7,19E-03 | 4,83E-02 |
| SOAT1 | 2923,75 | -0,05 | 0,02 | -2,69 | 7,20E-03 | 4,83E-02 |
| VAMP4 | 939,48 | 0,10 | 0,04 | 2,69 | 7,24E-03 | 4,86E-02 |
| TCF20 | 3117,62 | -0,07 | 0,03 | -2,68 | 7,26E-03 | 4,86E-02 |
| DNPH1 | 185,20 | 0,10 | 0,04 | 2,68 | 7,26E-03 | 4,86E-02 |
| DARS2 | 426,74 | 0,04 | 0,01 | 2,68 | 7,26E-03 | 4,86E-02 |
| CD59 | 1584,99 | -0,08 | 0,03 | -2,68 | 7,27E-03 | 4,87E-02 |
| RHOU | 607,16 | 0,09 | 0,03 | 2,68 | 7,30E-03 | 4,88E-02 |
| KIF18A | 160,45 | 0,15 | 0,06 | 2,68 | 7,30E-03 | 4,88E-02 |
| CSGALNACT: | 1355,70 | -0,11 | 0,04 | -2,68 | 7,30E-03 | 4,88E-02 |

| | | | | | | |
|---|---|---|---|---|---|---|
| ACP6 | 538,08 | -0,08 | 0,03 | -2,68 | 7,31E-03 | 4,88E-02 |
| TLR1 | 11411,76 | -0,11 | 0,04 | -2,68 | 7,32E-03 | 4,88E-02 |
| ARFIP1 | 2060,98 | -0,09 | 0,03 | -2,68 | 7,33E-03 | 4,88E-02 |
| IGF2BP2 | 840,86 | 0,19 | 0,07 | 2,68 | 7,35E-03 | 4,90E-02 |
| BPTF | 5689,30 | -0,05 | 0,02 | -2,68 | 7,39E-03 | 4,92E-02 |
| TG | 334,22 | -0,15 | 0,06 | -2,68 | 7,40E-03 | 4,92E-02 |
| EPG5 | 4207,21 | -0,07 | 0,03 | -2,68 | 7,43E-03 | 4,94E-02 |
| KDM4A | 1139,62 | -0,05 | 0,02 | -2,68 | 7,44E-03 | 4,94E-02 |
| NFIA | 450,71 | 0,10 | 0,04 | 2,68 | 7,44E-03 | 4,94E-02 |
| GPR174 | 2630,01 | 0,11 | 0,04 | 2,68 | 7,44E-03 | 4,94E-02 |
| ZNF160 | 1484,41 | -0,04 | 0,02 | -2,68 | 7,45E-03 | 4,94E-02 |
| MRPL55 | 262,88 | 0,05 | 0,02 | 2,68 | 7,47E-03 | 4,95E-02 |
| ATP6V1C1 | 1618,21 | -0,07 | 0,03 | -2,68 | 7,47E-03 | 4,95E-02 |
| KRT1 | 1238,63 | 0,28 | 0,10 | 2,68 | 7,46E-03 | 4,95E-02 |
| SNAP23 | 6009,25 | -0,07 | 0,03 | -2,67 | 7,50E-03 | 4,96E-02 |
| SNCA | 36565,95 | 0,20 | 0,07 | 2,67 | 7,50E-03 | 4,96E-02 |
| AHCYL1 | 1601,12 | -0,04 | 0,01 | -2,67 | 7,55E-03 | 4,99E-02 |
| MTERF3 | 274,40 | 0,06 | 0,02 | 2,67 | 7,56E-03 | 4,99E-02 |
| CDH26 | 257,77 | -0,10 | 0,04 | -2,67 | 7,61E-03 | 5,02E-02 |
| TMEM101 | 684,85 | 0,06 | 0,02 | 2,67 | 7,61E-03 | 5,02E-02 |
| DEF6 | 3811,32 | -0,05 | 0,02 | -2,67 | 7,62E-03 | 5,03E-02 |
| NEIL3 | 290,53 | 0,22 | 0,08 | 2,67 | 7,65E-03 | 5,04E-02 |
| RYBP | 3240,96 | -0,07 | 0,03 | -2,67 | 7,65E-03 | 5,04E-02 |
| ZNF551 | 513,18 | 0,06 | 0,02 | 2,67 | 7,67E-03 | 5,05E-02 |
| KNOP1 | 550,09 | 0,05 | 0,02 | 2,67 | 7,69E-03 | 5,06E-02 |
| DYNC1I2 | 1016,60 | 0,06 | 0,02 | 2,66 | 7,71E-03 | 5,07E-02 |
| RPS13 | 6270,93 | 0,12 | 0,05 | 2,66 | 7,72E-03 | 5,07E-02 |
| MOSPD1 | 387,68 | 0,11 | 0,04 | 2,66 | 7,74E-03 | 5,08E-02 |
| PEX7 | 203,11 | 0,06 | 0,02 | 2,66 | 7,74E-03 | 5,08E-02 |
| ELL | 1549,35 | -0,11 | 0,04 | -2,66 | 7,74E-03 | 5,08E-02 |
| PLIN4 | 340,24 | -0,18 | 0,07 | -2,66 | 7,73E-03 | 5,08E-02 |
| DNAJC24 | 765,45 | 0,08 | 0,03 | 2,66 | 7,78E-03 | 5,09E-02 |
| TNFRSF1B | 8579,77 | -0,10 | 0,04 | -2,66 | 7,78E-03 | 5,09E-02 |
| MIGA1 | 1078,27 | 0,06 | 0,02 | 2,66 | 7,78E-03 | 5,09E-02 |
| FRRS1 | 256,42 | 0,13 | 0,05 | 2,66 | 7,81E-03 | 5,11E-02 |
| ZZEF1 | 5712,13 | -0,08 | 0,03 | -2,66 | 7,84E-03 | 5,12E-02 |
| DNM1L | 1604,10 | 0,06 | 0,02 | 2,66 | 7,85E-03 | 5,13E-02 |
| CD83 | 302,49 | 0,10 | 0,04 | 2,66 | 7,85E-03 | 5,13E-02 |
| CHCHD4 | 178,54 | 0,06 | 0,02 | 2,66 | 7,88E-03 | 5,14E-02 |
| SUMO3 | 360,26 | 0,06 | 0,02 | 2,66 | 7,91E-03 | 5,16E-02 |
| SUPT7L | 1220,64 | 0,06 | 0,02 | 2,66 | 7,91E-03 | 5,16E-02 |
| SNX11 | 1206,27 | -0,08 | 0,03 | -2,65 | 7,95E-03 | 5,18E-02 |
| SLC14A1 | 1358,38 | 0,14 | 0,05 | 2,65 | 7,96E-03 | 5,18E-02 |
| NIPBL | 8385,46 | -0,07 | 0,03 | -2,65 | 7,96E-03 | 5,18E-02 |
| IDS | 4067,00 | -0,09 | 0,03 | -2,65 | 7,97E-03 | 5,18E-02 |
| SPIN2B | 88,03 | 0,07 | 0,02 | 2,65 | 7,98E-03 | 5,19E-02 |

| | | | | | | |
|---|---|---|---|---|---|---|
| WDR20 | 1901,92 | -0,05 | 0,02 | -2,65 | 8,00E-03 | 5,19E-02 |
| CYB5D2 | 179,94 | 0,05 | 0,02 | 2,65 | 8,00E-03 | 5,19E-02 |
| NRIP1 | 1712,59 | 0,09 | 0,03 | 2,65 | 8,02E-03 | 5,20E-02 |
| ACTN1 | 3779,30 | -0,14 | 0,05 | -2,65 | 8,02E-03 | 5,20E-02 |
| PPDPF | 209,50 | 0,18 | 0,07 | 2,65 | 8,03E-03 | 5,20E-02 |
| MOB1B | 1479,65 | 0,08 | 0,03 | 2,65 | 8,08E-03 | 5,23E-02 |
| METTL2B | 697,26 | 0,04 | 0,02 | 2,65 | 8,10E-03 | 5,24E-02 |
| SPAG16 | 155,31 | 0,10 | 0,04 | 2,65 | 8,11E-03 | 5,24E-02 |
| ARHGAP15 | 8440,04 | -0,09 | 0,03 | -2,65 | 8,12E-03 | 5,25E-02 |
| ZER1 | 1547,61 | 0,17 | 0,06 | 2,65 | 8,12E-03 | 5,25E-02 |
| NAGLU | 246,32 | 0,07 | 0,03 | 2,65 | 8,13E-03 | 5,25E-02 |
| GTPBP1 | 2223,13 | -0,11 | 0,04 | -2,65 | 8,16E-03 | 5,26E-02 |
| WWP2 | 2530,32 | -0,11 | 0,04 | -2,65 | 8,15E-03 | 5,26E-02 |
| PTBP3 | 10409,02 | -0,06 | 0,02 | -2,64 | 8,18E-03 | 5,27E-02 |
| ANK1 | 1372,57 | 0,20 | 0,08 | 2,64 | 8,18E-03 | 5,27E-02 |
| ALAS1 | 467,19 | -0,08 | 0,03 | -2,64 | 8,20E-03 | 5,28E-02 |
| RAE1 | 757,11 | -0,03 | 0,01 | -2,64 | 8,21E-03 | 5,28E-02 |
| PIN1 | 897,69 | 0,04 | 0,02 | 2,64 | 8,22E-03 | 5,28E-02 |
| GLIPR2 | 5052,17 | -0,08 | 0,03 | -2,64 | 8,22E-03 | 5,28E-02 |
| NFE2L3 | 285,97 | 0,11 | 0,04 | 2,64 | 8,22E-03 | 5,28E-02 |
| ACAP2 | 13469,84 | -0,07 | 0,03 | -2,64 | 8,23E-03 | 5,28E-02 |
| ACTR1B | 1037,93 | 0,04 | 0,02 | 2,64 | 8,24E-03 | 5,28E-02 |
| FAM136A | 395,65 | 0,06 | 0,02 | 2,64 | 8,24E-03 | 5,28E-02 |
| TCEAL1 | 83,59 | 0,12 | 0,05 | 2,64 | 8,26E-03 | 5,29E-02 |
| ZNF251 | 432,75 | 0,05 | 0,02 | 2,64 | 8,27E-03 | 5,29E-02 |
| BANP | 2467,70 | -0,07 | 0,03 | -2,64 | 8,28E-03 | 5,30E-02 |
| ARPC5 | 17360,31 | -0,09 | 0,03 | -2,64 | 8,29E-03 | 5,30E-02 |
| ALKBH3 | 538,46 | 0,05 | 0,02 | 2,64 | 8,29E-03 | 5,30E-02 |
| TCEA1 | 2266,50 | 0,08 | 0,03 | 2,64 | 8,30E-03 | 5,30E-02 |
| FAM135A | 830,81 | 0,08 | 0,03 | 2,64 | 8,31E-03 | 5,31E-02 |
| PTTG2 | 170,17 | -0,13 | 0,05 | -2,64 | 8,35E-03 | 5,33E-02 |
| RNF20 | 2221,20 | -0,06 | 0,02 | -2,64 | 8,40E-03 | 5,35E-02 |
| MOSPD2 | 2504,11 | -0,10 | 0,04 | -2,64 | 8,40E-03 | 5,35E-02 |
| ANKFY1 | 2768,56 | -0,08 | 0,03 | -2,64 | 8,41E-03 | 5,35E-02 |
| TMEM127 | 2651,34 | -0,11 | 0,04 | -2,63 | 8,41E-03 | 5,36E-02 |
| KCTD2 | 631,13 | -0,06 | 0,02 | -2,63 | 8,43E-03 | 5,36E-02 |
| IBA57 | 585,77 | 0,08 | 0,03 | 2,63 | 8,43E-03 | 5,36E-02 |
| STMN3 | 667,21 | 0,08 | 0,03 | 2,63 | 8,48E-03 | 5,39E-02 |
| SRP19 | 628,99 | -0,04 | 0,02 | -2,63 | 8,48E-03 | 5,39E-02 |
| CLEC1A | 98,25 | -0,13 | 0,05 | -2,63 | 8,49E-03 | 5,39E-02 |
| AIM2 | 502,84 | -0,15 | 0,06 | -2,63 | 8,50E-03 | 5,39E-02 |
| NOXA1 | 118,01 | 0,12 | 0,05 | 2,63 | 8,51E-03 | 5,39E-02 |
| DHRS7B | 699,80 | -0,05 | 0,02 | -2,63 | 8,52E-03 | 5,40E-02 |
| ZNF746 | 2334,86 | -0,11 | 0,04 | -2,63 | 8,52E-03 | 5,40E-02 |
| NT5C | 501,64 | 0,06 | 0,02 | 2,63 | 8,53E-03 | 5,40E-02 |
| RPL3 | 12133,07 | 0,11 | 0,04 | 2,63 | 8,55E-03 | 5,40E-02 |

| | | | | | | |
|---|---|---|---|---|---|---|
| AFDN | 814,28 | -0,14 | 0,05 | -2,63 | 8,55E-03 | 5,40E-02 |
| PHB2 | 1549,99 | 0,06 | 0,02 | 2,63 | 8,55E-03 | 5,40E-02 |
| CLK3 | 3709,26 | -0,04 | 0,02 | -2,63 | 8,58E-03 | 5,41E-02 |
| CNPPD1 | 850,74 | 0,13 | 0,05 | 2,63 | 8,57E-03 | 5,41E-02 |
| POLR1F | 366,71 | 0,12 | 0,04 | 2,63 | 8,60E-03 | 5,43E-02 |
| TRAPPC2L | 364,10 | 0,06 | 0,02 | 2,63 | 8,62E-03 | 5,43E-02 |
| NCOA6 | 1270,75 | -0,08 | 0,03 | -2,63 | 8,62E-03 | 5,43E-02 |
| MBLAC2 | 258,71 | 0,11 | 0,04 | 2,62 | 8,67E-03 | 5,46E-02 |
| USP34 | 12125,27 | -0,06 | 0,02 | -2,62 | 8,67E-03 | 5,46E-02 |
| NUDT1 | 286,10 | 0,09 | 0,03 | 2,62 | 8,68E-03 | 5,46E-02 |
| TNS1 | 3938,45 | 0,22 | 0,08 | 2,62 | 8,70E-03 | 5,47E-02 |
| NTNG2 | 902,81 | -0,17 | 0,07 | -2,62 | 8,74E-03 | 5,49E-02 |
| SLAIN1 | 610,96 | 0,07 | 0,03 | 2,62 | 8,76E-03 | 5,50E-02 |
| RILPL2 | 2157,04 | -0,07 | 0,03 | -2,62 | 8,76E-03 | 5,50E-02 |
| ACADSB | 614,73 | 0,05 | 0,02 | 2,62 | 8,78E-03 | 5,51E-02 |
| SLC4A7 | 3278,29 | 0,09 | 0,03 | 2,62 | 8,79E-03 | 5,51E-02 |
| CALCOCO1 | 3139,15 | -0,07 | 0,03 | -2,62 | 8,81E-03 | 5,52E-02 |
| TRIAP1 | 154,67 | 0,14 | 0,05 | 2,62 | 8,81E-03 | 5,52E-02 |
| MAML3 | 664,63 | -0,11 | 0,04 | -2,62 | 8,86E-03 | 5,54E-02 |
| PELI3 | 208,10 | 0,08 | 0,03 | 2,62 | 8,87E-03 | 5,54E-02 |
| TARBP2 | 173,51 | 0,06 | 0,02 | 2,62 | 8,88E-03 | 5,55E-02 |
| PAFAH1B2 | 2559,98 | -0,04 | 0,02 | -2,62 | 8,89E-03 | 5,55E-02 |
| MRPL15 | 203,27 | 0,10 | 0,04 | 2,61 | 8,94E-03 | 5,58E-02 |
| SH3RF3 | 216,70 | -0,16 | 0,06 | -2,61 | 8,95E-03 | 5,58E-02 |
| RAB43 | 398,74 | -0,08 | 0,03 | -2,61 | 8,95E-03 | 5,58E-02 |
| BRAF | 4193,47 | -0,06 | 0,02 | -2,61 | 8,96E-03 | 5,58E-02 |
| COMMD6 | 302,84 | 0,23 | 0,09 | 2,61 | 8,97E-03 | 5,58E-02 |
| NSD3 | 7652,18 | -0,05 | 0,02 | -2,61 | 8,97E-03 | 5,59E-02 |
| SAT1 | 10171,10 | -0,10 | 0,04 | -2,61 | 8,98E-03 | 5,59E-02 |
| TMEM161A | 112,45 | 0,09 | 0,03 | 2,61 | 8,99E-03 | 5,59E-02 |
| ANGEL1 | 572,66 | 0,06 | 0,02 | 2,61 | 9,00E-03 | 5,60E-02 |
| MRPS28 | 246,82 | 0,07 | 0,03 | 2,61 | 9,01E-03 | 5,60E-02 |
| NXT1 | 129,33 | 0,07 | 0,03 | 2,61 | 9,02E-03 | 5,60E-02 |
| EIF1B | 953,86 | 0,08 | 0,03 | 2,61 | 9,03E-03 | 5,60E-02 |
| CCDC88A | 2945,45 | 0,09 | 0,04 | 2,61 | 9,07E-03 | 5,62E-02 |
| EIF1 | 3129,42 | 0,08 | 0,03 | 2,61 | 9,07E-03 | 5,62E-02 |
| MZT2A | 627,55 | 0,08 | 0,03 | 2,61 | 9,08E-03 | 5,62E-02 |
| PNMA1 | 383,58 | 0,06 | 0,02 | 2,61 | 9,09E-03 | 5,63E-02 |
| ABHD16A | 292,40 | -0,06 | 0,02 | -2,61 | 9,09E-03 | 5,63E-02 |
| SGCB | 151,49 | 0,09 | 0,04 | 2,61 | 9,11E-03 | 5,63E-02 |
| PCMTD2 | 3290,74 | 0,05 | 0,02 | 2,61 | 9,13E-03 | 5,65E-02 |
| RAB5A | 1893,29 | -0,06 | 0,02 | -2,61 | 9,16E-03 | 5,66E-02 |
| GPR155 | 3171,34 | -0,07 | 0,03 | -2,60 | 9,19E-03 | 5,68E-02 |
| ACAD11 | 251,85 | 0,06 | 0,02 | 2,60 | 9,20E-03 | 5,68E-02 |
| GUF1 | 1045,62 | 0,08 | 0,03 | 2,60 | 9,23E-03 | 5,69E-02 |
| CREBZF | 4233,29 | 0,08 | 0,03 | 2,60 | 9,25E-03 | 5,70E-02 |

| | | | | | | |
|---|---|---|---|---|---|---|
| SVBP | 383,22 | 0,12 | 0,05 | 2,60 | 9,25E-03 | 5,70E-02 |
| RCSD1 | 7363,46 | -0,06 | 0,02 | -2,60 | 9,28E-03 | 5,71E-02 |
| CENPH | 86,60 | 0,11 | 0,04 | 2,60 | 9,28E-03 | 5,71E-02 |
| IFNGR2 | 4279,66 | -0,08 | 0,03 | -2,60 | 9,34E-03 | 5,74E-02 |
| TESMIN | 197,99 | -0,09 | 0,04 | -2,60 | 9,33E-03 | 5,74E-02 |
| TMED1 | 381,35 | 0,07 | 0,03 | 2,60 | 9,34E-03 | 5,74E-02 |
| MECR | 164,14 | 0,07 | 0,03 | 2,60 | 9,36E-03 | 5,75E-02 |
| UBE2Q2 | 800,41 | 0,07 | 0,03 | 2,60 | 9,37E-03 | 5,75E-02 |
| SLC2A13 | 439,65 | 0,06 | 0,02 | 2,60 | 9,39E-03 | 5,76E-02 |
| ASAH1 | 11042,16 | -0,09 | 0,03 | -2,60 | 9,40E-03 | 5,76E-02 |
| MRPS12 | 246,30 | 0,06 | 0,02 | 2,60 | 9,40E-03 | 5,76E-02 |
| MACROH2A1 | 10970,77 | -0,07 | 0,03 | -2,60 | 9,44E-03 | 5,78E-02 |
| ZFAND3 | 2279,23 | -0,09 | 0,04 | -2,59 | 9,47E-03 | 5,80E-02 |
| OR52K2 | 107,32 | -0,21 | 0,08 | -2,59 | 9,51E-03 | 5,82E-02 |
| CYB5A | 387,29 | 0,07 | 0,03 | 2,59 | 9,58E-03 | 5,86E-02 |
| RPL10A | 8307,65 | 0,12 | 0,04 | 2,59 | 9,60E-03 | 5,86E-02 |
| TRMT61A | 222,00 | 0,06 | 0,02 | 2,59 | 9,64E-03 | 5,88E-02 |
| TSC22D3 | 5173,51 | -0,10 | 0,04 | -2,59 | 9,64E-03 | 5,89E-02 |
| SERTAD2 | 982,78 | -0,06 | 0,02 | -2,59 | 9,66E-03 | 5,89E-02 |
| MCMBP | 3168,74 | -0,06 | 0,02 | -2,59 | 9,66E-03 | 5,89E-02 |
| IP6K1 | 1520,76 | -0,10 | 0,04 | -2,59 | 9,69E-03 | 5,90E-02 |
| RAB34 | 187,24 | 0,09 | 0,03 | 2,59 | 9,70E-03 | 5,90E-02 |
| DYNC1LI1 | 2411,69 | -0,08 | 0,03 | -2,59 | 9,71E-03 | 5,90E-02 |
| KCNMB3 | 100,38 | 0,09 | 0,03 | 2,59 | 9,70E-03 | 5,90E-02 |
| HAGH | 4708,32 | 0,16 | 0,06 | 2,59 | 9,72E-03 | 5,91E-02 |
| PLA2G12A | 601,47 | 0,07 | 0,03 | 2,59 | 9,73E-03 | 5,91E-02 |
| TRMT1 | 528,21 | 0,06 | 0,02 | 2,58 | 9,75E-03 | 5,92E-02 |
| VPS39 | 2099,07 | -0,08 | 0,03 | -2,58 | 9,77E-03 | 5,93E-02 |
| LRRN3 | 299,45 | 0,21 | 0,08 | 2,58 | 9,84E-03 | 5,97E-02 |
| CD24 | 566,39 | 0,16 | 0,06 | 2,58 | 9,90E-03 | 6,00E-02 |
| TRIP4 | 507,21 | -0,04 | 0,02 | -2,58 | 9,92E-03 | 6,01E-02 |
| TEX264 | 1083,17 | 0,04 | 0,01 | 2,58 | 9,92E-03 | 6,01E-02 |
| SIRT5 | 557,05 | 0,04 | 0,02 | 2,58 | 9,94E-03 | 6,01E-02 |
| PLD1 | 736,80 | -0,09 | 0,04 | -2,58 | 9,94E-03 | 6,01E-02 |
| GCLM | 694,81 | 0,07 | 0,03 | 2,58 | 9,96E-03 | 6,02E-02 |
| TMEM50B | 1198,50 | 0,08 | 0,03 | 2,58 | 9,96E-03 | 6,02E-02 |
| FBL | 986,61 | 0,09 | 0,03 | 2,58 | 9,99E-03 | 6,03E-02 |
| NEU1 | 844,94 | -0,06 | 0,02 | -2,57 | 1,00E-02 | 6,05E-02 |
| H1-2 | 6152,16 | 0,10 | 0,04 | 2,57 | 1,00E-02 | 6,05E-02 |
| PRDM8 | 365,04 | -0,08 | 0,03 | -2,57 | 1,00E-02 | 6,05E-02 |
| LETMD1 | 1122,96 | 0,04 | 0,01 | 2,57 | 1,00E-02 | 6,05E-02 |
| RPL18 | 12334,94 | 0,08 | 0,03 | 2,57 | 1,01E-02 | 6,07E-02 |
| NIF3L1 | 434,76 | 0,06 | 0,02 | 2,57 | 1,01E-02 | 6,07E-02 |
| MSRB1 | 3341,46 | -0,10 | 0,04 | -2,57 | 1,01E-02 | 6,07E-02 |
| PSENEN | 337,90 | -0,07 | 0,03 | -2,57 | 1,01E-02 | 6,07E-02 |
| ADAM15 | 426,55 | 0,10 | 0,04 | 2,57 | 1,01E-02 | 6,07E-02 |

| | | | | | | |
|---|---|---|---|---|---|---|
| AKAP7 | 451,55 | 0,11 | 0,04 | 2,57 | 1,02E-02 | 6,13E-02 |
| ZNF326 | 1308,75 | 0,07 | 0,03 | 2,57 | 1,02E-02 | 6,13E-02 |
| POLRMT | 521,88 | 0,08 | 0,03 | 2,57 | 1,02E-02 | 6,13E-02 |
| ACP3 | 1191,60 | -0,10 | 0,04 | -2,57 | 1,03E-02 | 6,16E-02 |
| ADPRS | 697,65 | -0,06 | 0,02 | -2,57 | 1,03E-02 | 6,16E-02 |
| CDK5R1 | 531,79 | -0,11 | 0,04 | -2,57 | 1,03E-02 | 6,16E-02 |
| CBL | 4600,23 | -0,13 | 0,05 | -2,56 | 1,03E-02 | 6,18E-02 |
| AKT1S1 | 372,74 | 0,08 | 0,03 | 2,56 | 1,03E-02 | 6,19E-02 |
| COX11 | 1111,39 | 0,06 | 0,02 | 2,56 | 1,04E-02 | 6,19E-02 |
| TMEM116 | 742,54 | 0,06 | 0,02 | 2,56 | 1,04E-02 | 6,19E-02 |
| TMEM272 | 1019,37 | -0,12 | 0,05 | -2,56 | 1,04E-02 | 6,20E-02 |
| RPL4 | 14616,20 | 0,12 | 0,05 | 2,56 | 1,04E-02 | 6,20E-02 |
| ABHD10 | 844,42 | 0,11 | 0,04 | 2,56 | 1,04E-02 | 6,20E-02 |
| CAMK2G | 2741,71 | -0,10 | 0,04 | -2,56 | 1,05E-02 | 6,23E-02 |
| LIN52 | 300,67 | 0,05 | 0,02 | 2,56 | 1,05E-02 | 6,23E-02 |
| ST8SIA4 | 9694,73 | -0,09 | 0,04 | -2,56 | 1,05E-02 | 6,23E-02 |
| ERCC1 | 583,84 | 0,05 | 0,02 | 2,56 | 1,05E-02 | 6,23E-02 |
| SAR1B | 1476,57 | 0,05 | 0,02 | 2,56 | 1,05E-02 | 6,23E-02 |
| HELZ2 | 2380,46 | -0,17 | 0,06 | -2,56 | 1,05E-02 | 6,24E-02 |
| TBL1XR1 | 8612,28 | 0,08 | 0,03 | 2,56 | 1,05E-02 | 6,24E-02 |
| CCDC50 | 1343,98 | 0,10 | 0,04 | 2,56 | 1,05E-02 | 6,24E-02 |
| AHR | 2087,40 | 0,09 | 0,03 | 2,56 | 1,05E-02 | 6,25E-02 |
| PTPRN2 | 304,36 | -0,13 | 0,05 | -2,56 | 1,05E-02 | 6,26E-02 |
| UBE2O | 1105,01 | 0,17 | 0,07 | 2,56 | 1,06E-02 | 6,27E-02 |
| CERS2 | 2330,65 | -0,06 | 0,02 | -2,56 | 1,06E-02 | 6,27E-02 |
| ZNF786 | 349,49 | 0,05 | 0,02 | 2,56 | 1,06E-02 | 6,27E-02 |
| C5AR1 | 2093,63 | -0,15 | 0,06 | -2,55 | 1,06E-02 | 6,29E-02 |
| KLF7 | 2861,28 | -0,09 | 0,04 | -2,55 | 1,06E-02 | 6,29E-02 |
| SPRYD4 | 339,84 | 0,07 | 0,03 | 2,55 | 1,06E-02 | 6,29E-02 |
| KDM1B | 1633,95 | -0,08 | 0,03 | -2,55 | 1,06E-02 | 6,29E-02 |
| KPNA5 | 1299,72 | 0,12 | 0,05 | 2,55 | 1,07E-02 | 6,30E-02 |
| NDRG2 | 150,97 | 0,11 | 0,04 | 2,55 | 1,07E-02 | 6,30E-02 |
| MLST8 | 145,73 | 0,08 | 0,03 | 2,55 | 1,07E-02 | 6,30E-02 |
| PABIR2 | 842,75 | 0,06 | 0,02 | 2,55 | 1,07E-02 | 6,30E-02 |
| TMTC3 | 829,28 | 0,10 | 0,04 | 2,55 | 1,07E-02 | 6,30E-02 |
| TNFRSF1A | 2770,62 | -0,12 | 0,05 | -2,55 | 1,07E-02 | 6,32E-02 |
| MSL3 | 2426,94 | -0,05 | 0,02 | -2,55 | 1,07E-02 | 6,32E-02 |
| B9D2 | 371,97 | -0,07 | 0,03 | -2,55 | 1,08E-02 | 6,32E-02 |
| IFIT1B | 941,28 | 0,20 | 0,08 | 2,55 | 1,08E-02 | 6,32E-02 |
| ITGA1 | 285,29 | -0,13 | 0,05 | -2,55 | 1,08E-02 | 6,32E-02 |
| FRAT1 | 2510,18 | -0,10 | 0,04 | -2,55 | 1,08E-02 | 6,32E-02 |
| DCP2 | 12037,17 | -0,08 | 0,03 | -2,55 | 1,08E-02 | 6,33E-02 |
| UPF1 | 3308,15 | -0,09 | 0,04 | -2,55 | 1,08E-02 | 6,33E-02 |
| SLC39A8 | 593,00 | 0,08 | 0,03 | 2,55 | 1,08E-02 | 6,33E-02 |
| PYROXD1 | 1130,07 | 0,09 | 0,04 | 2,55 | 1,08E-02 | 6,33E-02 |
| GUK1 | 24482,26 | 0,16 | 0,06 | 2,55 | 1,08E-02 | 6,34E-02 |

| | | | | | | |
|---|---|---|---|---|---|---|
| ZBTB3 | 171,76 | 0,07 | 0,03 | 2,55 | 1,08E-02 | 6,35E-02 |
| ME2 | 3015,93 | -0,05 | 0,02 | -2,55 | 1,09E-02 | 6,36E-02 |
| ZNF675 | 607,03 | 0,08 | 0,03 | 2,55 | 1,09E-02 | 6,36E-02 |
| ENSG000002 | 396,41 | 0,04 | 0,02 | 2,55 | 1,09E-02 | 6,36E-02 |
| CIP2A | 277,06 | 0,10 | 0,04 | 2,55 | 1,09E-02 | 6,36E-02 |
| CFH | 278,05 | 0,19 | 0,07 | 2,55 | 1,09E-02 | 6,36E-02 |
| TUBE1 | 619,84 | 0,09 | 0,04 | 2,55 | 1,09E-02 | 6,37E-02 |
| MAP3K2 | 12709,01 | -0,08 | 0,03 | -2,55 | 1,09E-02 | 6,37E-02 |
| RWDD2A | 93,07 | 0,08 | 0,03 | 2,54 | 1,09E-02 | 6,37E-02 |
| CDC27 | 2905,18 | 0,07 | 0,03 | 2,55 | 1,09E-02 | 6,37E-02 |
| CEP350 | 7357,10 | -0,05 | 0,02 | -2,54 | 1,09E-02 | 6,37E-02 |
| OSER1 | 1682,97 | -0,08 | 0,03 | -2,54 | 1,09E-02 | 6,37E-02 |
| CYTIP | 10338,04 | -0,06 | 0,03 | -2,54 | 1,09E-02 | 6,37E-02 |
| ZNF800 | 2299,24 | 0,07 | 0,03 | 2,54 | 1,10E-02 | 6,38E-02 |
| ZFYVE16 | 5787,35 | -0,09 | 0,04 | -2,54 | 1,10E-02 | 6,40E-02 |
| POLL | 806,45 | 0,11 | 0,04 | 2,54 | 1,10E-02 | 6,40E-02 |
| CMPK1 | 2472,78 | 0,10 | 0,04 | 2,54 | 1,10E-02 | 6,40E-02 |
| KIFC2 | 325,50 | 0,10 | 0,04 | 2,54 | 1,10E-02 | 6,40E-02 |
| DRAM1 | 480,72 | -0,07 | 0,03 | -2,54 | 1,10E-02 | 6,40E-02 |
| TIMM23 | 567,10 | 0,06 | 0,02 | 2,54 | 1,10E-02 | 6,40E-02 |
| WDR36 | 1729,95 | 0,08 | 0,03 | 2,54 | 1,11E-02 | 6,43E-02 |
| ALDH1A2 | 187,86 | -0,19 | 0,07 | -2,54 | 1,11E-02 | 6,43E-02 |
| ZNF121 | 2018,27 | 0,07 | 0,03 | 2,54 | 1,11E-02 | 6,43E-02 |
| KLHDC1 | 304,49 | 0,07 | 0,03 | 2,54 | 1,11E-02 | 6,43E-02 |
| IFIT1 | 2061,20 | -0,30 | 0,12 | -2,54 | 1,11E-02 | 6,44E-02 |
| OAS2 | 2283,67 | -0,16 | 0,06 | -2,54 | 1,11E-02 | 6,44E-02 |
| TMA16 | 568,24 | 0,08 | 0,03 | 2,54 | 1,12E-02 | 6,45E-02 |
| SHF | 87,36 | 0,11 | 0,04 | 2,54 | 1,12E-02 | 6,45E-02 |
| PRRC2C | 9462,77 | -0,06 | 0,02 | -2,54 | 1,12E-02 | 6,46E-02 |
| NDUFS8 | 257,21 | 0,07 | 0,03 | 2,54 | 1,12E-02 | 6,48E-02 |
| MCCC2 | 695,85 | 0,04 | 0,02 | 2,54 | 1,12E-02 | 6,48E-02 |
| NR2C1 | 694,99 | 0,06 | 0,02 | 2,53 | 1,13E-02 | 6,50E-02 |
| COMMD3 | 301,16 | 0,11 | 0,04 | 2,53 | 1,13E-02 | 6,52E-02 |
| MRPS25 | 1668,83 | 0,05 | 0,02 | 2,53 | 1,13E-02 | 6,52E-02 |
| TMEM147 | 280,96 | 0,06 | 0,02 | 2,53 | 1,13E-02 | 6,52E-02 |
| LEPR | 426,17 | 0,14 | 0,06 | 2,53 | 1,14E-02 | 6,53E-02 |
| PPP2R5A | 4252,14 | -0,08 | 0,03 | -2,53 | 1,14E-02 | 6,53E-02 |
| ZNF486 | 800,95 | -0,09 | 0,03 | -2,53 | 1,14E-02 | 6,55E-02 |
| RUNDC3A | 644,78 | 0,20 | 0,08 | 2,53 | 1,15E-02 | 6,59E-02 |
| ZXDA | 292,36 | 0,07 | 0,03 | 2,53 | 1,15E-02 | 6,59E-02 |
| SLC2A1 | 1995,44 | 0,12 | 0,05 | 2,53 | 1,15E-02 | 6,59E-02 |
| MOSMO | 821,92 | 0,07 | 0,03 | 2,53 | 1,15E-02 | 6,59E-02 |
| EID2B | 161,09 | 0,08 | 0,03 | 2,53 | 1,15E-02 | 6,61E-02 |
| KLF1 | 1188,39 | 0,19 | 0,07 | 2,53 | 1,15E-02 | 6,61E-02 |
| HRAS | 90,53 | 0,10 | 0,04 | 2,52 | 1,16E-02 | 6,63E-02 |
| CA1 | 2197,76 | 0,24 | 0,09 | 2,52 | 1,16E-02 | 6,64E-02 |

| | | | | | | |
|---|---|---|---|---|---|---|
| UBR7 | 505,27 | -0,04 | 0,02 | -2,52 | 1,16E-02 | 6,66E-02 |
| PDCD7 | 650,81 | 0,05 | 0,02 | 2,52 | 1,17E-02 | 6,71E-02 |
| HPCAL4 | 135,13 | 0,14 | 0,06 | 2,52 | 1,17E-02 | 6,71E-02 |
| LRR1 | 365,64 | 0,07 | 0,03 | 2,52 | 1,18E-02 | 6,72E-02 |
| EP300 | 5565,07 | -0,08 | 0,03 | -2,52 | 1,18E-02 | 6,72E-02 |
| SIRPB1 | 1659,32 | -0,14 | 0,05 | -2,52 | 1,18E-02 | 6,74E-02 |
| PLAGL1 | 795,02 | -0,09 | 0,04 | -2,52 | 1,18E-02 | 6,75E-02 |
| LYL1 | 2573,20 | 0,12 | 0,05 | 2,52 | 1,18E-02 | 6,75E-02 |
| PIP4P2 | 1842,72 | -0,10 | 0,04 | -2,52 | 1,18E-02 | 6,75E-02 |
| KIAA0825 | 1405,66 | -0,10 | 0,04 | -2,52 | 1,19E-02 | 6,75E-02 |
| ITGAM | 2685,41 | -0,13 | 0,05 | -2,51 | 1,19E-02 | 6,78E-02 |
| GRPEL2 | 783,14 | 0,08 | 0,03 | 2,51 | 1,19E-02 | 6,78E-02 |
| NSMCE3 | 486,05 | 0,06 | 0,02 | 2,51 | 1,19E-02 | 6,78E-02 |
| PTGES3 | 1742,88 | 0,10 | 0,04 | 2,51 | 1,20E-02 | 6,78E-02 |
| DTWD2 | 335,90 | 0,06 | 0,03 | 2,51 | 1,19E-02 | 6,78E-02 |
| LZTS2 | 286,43 | 0,08 | 0,03 | 2,51 | 1,20E-02 | 6,78E-02 |
| ENSG000002 | 132,93 | -0,06 | 0,03 | -2,51 | 1,19E-02 | 6,78E-02 |
| ZNF674 | 515,22 | -0,06 | 0,02 | -2,51 | 1,20E-02 | 6,78E-02 |
| SINHCAF | 1606,56 | 0,07 | 0,03 | 2,51 | 1,20E-02 | 6,78E-02 |
| TRAPPC6A | 360,74 | 0,07 | 0,03 | 2,51 | 1,20E-02 | 6,79E-02 |
| GRAMD1C | 668,67 | -0,13 | 0,05 | -2,51 | 1,20E-02 | 6,79E-02 |
| PRDM1 | 1290,90 | -0,07 | 0,03 | -2,51 | 1,20E-02 | 6,80E-02 |
| CNOT9 | 1329,31 | 0,04 | 0,02 | 2,51 | 1,20E-02 | 6,81E-02 |
| EIF2A | 1558,28 | 0,10 | 0,04 | 2,51 | 1,20E-02 | 6,81E-02 |
| RADX | 235,30 | 0,10 | 0,04 | 2,51 | 1,21E-02 | 6,82E-02 |
| PRPS2 | 269,24 | 0,07 | 0,03 | 2,51 | 1,21E-02 | 6,82E-02 |
| SEMA4A | 1632,61 | -0,12 | 0,05 | -2,51 | 1,21E-02 | 6,82E-02 |
| MAPK6 | 716,42 | 0,07 | 0,03 | 2,51 | 1,21E-02 | 6,83E-02 |
| STAT2 | 3838,28 | -0,12 | 0,05 | -2,51 | 1,21E-02 | 6,83E-02 |
| ACSL4 | 4712,89 | -0,09 | 0,04 | -2,51 | 1,21E-02 | 6,84E-02 |
| PALB2 | 321,74 | 0,05 | 0,02 | 2,51 | 1,22E-02 | 6,85E-02 |
| OSGEPL1 | 314,89 | 0,08 | 0,03 | 2,51 | 1,22E-02 | 6,85E-02 |
| SMG1 | 12384,41 | -0,06 | 0,02 | -2,51 | 1,22E-02 | 6,85E-02 |
| ERMARD | 949,65 | 0,05 | 0,02 | 2,50 | 1,23E-02 | 6,89E-02 |
| TNFAIP8L1 | 292,87 | 0,07 | 0,03 | 2,50 | 1,23E-02 | 6,90E-02 |
| SLIRP | 218,81 | 0,07 | 0,03 | 2,50 | 1,23E-02 | 6,90E-02 |
| PIGB | 2291,96 | -0,09 | 0,04 | -2,50 | 1,23E-02 | 6,90E-02 |
| LYRM2 | 682,33 | 0,10 | 0,04 | 2,50 | 1,23E-02 | 6,91E-02 |
| BCLAF3 | 1222,12 | -0,06 | 0,02 | -2,50 | 1,23E-02 | 6,91E-02 |
| SSH1 | 1433,64 | -0,11 | 0,04 | -2,50 | 1,23E-02 | 6,91E-02 |
| ZNF253 | 432,23 | 0,10 | 0,04 | 2,50 | 1,23E-02 | 6,91E-02 |
| LCP2 | 9886,73 | -0,06 | 0,02 | -2,50 | 1,23E-02 | 6,91E-02 |
| RCN2 | 464,19 | 0,08 | 0,03 | 2,50 | 1,23E-02 | 6,91E-02 |
| TBCEL | 1301,71 | 0,11 | 0,05 | 2,50 | 1,24E-02 | 6,92E-02 |
| SMARCC1 | 2007,65 | -0,05 | 0,02 | -2,50 | 1,24E-02 | 6,94E-02 |
| BMPR2 | 1005,53 | 0,07 | 0,03 | 2,50 | 1,24E-02 | 6,95E-02 |

| | | | | | | |
|---|---|---|---|---|---|---|
| NDRG1 | 1938,89 | -0,06 | 0,03 | -2,50 | 1,24E-02 | 6,95E-02 |
| PFDN4 | 138,75 | 0,14 | 0,05 | 2,50 | 1,25E-02 | 6,96E-02 |
| FGGY | 473,54 | -0,08 | 0,03 | -2,50 | 1,25E-02 | 6,96E-02 |
| EBLN2 | 313,12 | -0,05 | 0,02 | -2,50 | 1,25E-02 | 6,96E-02 |
| POLR3H | 415,52 | 0,08 | 0,03 | 2,50 | 1,25E-02 | 6,96E-02 |
| PPP1R15A | 968,51 | -0,10 | 0,04 | -2,50 | 1,25E-02 | 6,96E-02 |
| RNF144B | 4320,96 | -0,09 | 0,03 | -2,50 | 1,25E-02 | 6,97E-02 |
| RNF126 | 471,95 | 0,08 | 0,03 | 2,50 | 1,25E-02 | 6,98E-02 |
| HDGF | 5528,10 | 0,14 | 0,06 | 2,50 | 1,25E-02 | 6,98E-02 |
| TOP3A | 1536,29 | -0,07 | 0,03 | -2,50 | 1,25E-02 | 6,98E-02 |
| NUDT14 | 86,38 | 0,09 | 0,04 | 2,50 | 1,26E-02 | 6,98E-02 |
| CDCA4 | 260,92 | 0,06 | 0,02 | 2,50 | 1,26E-02 | 6,98E-02 |
| ZNF22 | 527,48 | 0,14 | 0,06 | 2,50 | 1,26E-02 | 6,98E-02 |
| GPR107 | 1857,31 | -0,08 | 0,03 | -2,49 | 1,26E-02 | 6,98E-02 |
| COX7C | 1089,00 | 0,18 | 0,07 | 2,50 | 1,26E-02 | 6,98E-02 |
| DHODH | 246,07 | 0,07 | 0,03 | 2,50 | 1,26E-02 | 6,98E-02 |
| RWDD1 | 363,02 | 0,10 | 0,04 | 2,49 | 1,27E-02 | 7,02E-02 |
| ZBTB10 | 829,34 | 0,10 | 0,04 | 2,49 | 1,27E-02 | 7,03E-02 |
| MRM2 | 456,12 | 0,05 | 0,02 | 2,49 | 1,27E-02 | 7,03E-02 |
| SDHAF1 | 291,14 | 0,07 | 0,03 | 2,49 | 1,27E-02 | 7,03E-02 |
| TAF1 | 3552,59 | -0,05 | 0,02 | -2,49 | 1,27E-02 | 7,03E-02 |
| ZNF850 | 190,12 | 0,07 | 0,03 | 2,49 | 1,27E-02 | 7,03E-02 |
| ZNF564 | 140,50 | -0,07 | 0,03 | -2,49 | 1,27E-02 | 7,03E-02 |
| KIZ | 857,22 | -0,04 | 0,02 | -2,49 | 1,27E-02 | 7,03E-02 |
| COPS2 | 1310,90 | 0,12 | 0,05 | 2,49 | 1,27E-02 | 7,03E-02 |
| HYCC1 | 1998,35 | 0,08 | 0,03 | 2,49 | 1,27E-02 | 7,03E-02 |
| DNAJB4 | 408,90 | 0,08 | 0,03 | 2,49 | 1,28E-02 | 7,04E-02 |
| ZNF678 | 863,20 | 0,11 | 0,05 | 2,49 | 1,28E-02 | 7,04E-02 |
| CNTNAP3 | 2528,14 | -0,18 | 0,07 | -2,49 | 1,28E-02 | 7,05E-02 |
| PSMC1 | 1124,98 | -0,05 | 0,02 | -2,49 | 1,28E-02 | 7,05E-02 |
| L3HYPDH | 560,21 | 0,06 | 0,02 | 2,49 | 1,28E-02 | 7,05E-02 |
| CREBBP | 4403,53 | -0,11 | 0,05 | -2,49 | 1,28E-02 | 7,05E-02 |
| IRAK1BP1 | 154,57 | 0,11 | 0,04 | 2,49 | 1,29E-02 | 7,07E-02 |
| IL10RB | 346,33 | -0,08 | 0,03 | -2,49 | 1,29E-02 | 7,07E-02 |
| SPDYA | 173,21 | -0,09 | 0,04 | -2,49 | 1,29E-02 | 7,07E-02 |
| UQCC6 | 189,16 | 0,07 | 0,03 | 2,49 | 1,29E-02 | 7,07E-02 |
| VSIR | 10944,34 | -0,10 | 0,04 | -2,49 | 1,29E-02 | 7,09E-02 |
| FADD | 1305,38 | -0,06 | 0,02 | -2,49 | 1,29E-02 | 7,09E-02 |
| ACTR2 | 17589,20 | -0,07 | 0,03 | -2,49 | 1,30E-02 | 7,10E-02 |
| KLHL18 | 1272,61 | -0,06 | 0,02 | -2,48 | 1,30E-02 | 7,10E-02 |
| ZNF75A | 1123,10 | 0,06 | 0,03 | 2,48 | 1,30E-02 | 7,11E-02 |
| LYST | 40008,86 | -0,08 | 0,03 | -2,48 | 1,30E-02 | 7,11E-02 |
| ZNF354B | 405,31 | 0,07 | 0,03 | 2,48 | 1,30E-02 | 7,13E-02 |
| KLF4 | 479,95 | 0,09 | 0,04 | 2,48 | 1,30E-02 | 7,13E-02 |
| PSMB9 | 5509,39 | -0,09 | 0,03 | -2,48 | 1,31E-02 | 7,14E-02 |
| TRAM1 | 4283,03 | 0,08 | 0,03 | 2,48 | 1,31E-02 | 7,14E-02 |

| | | | | | | |
|---|---|---|---|---|---|---|
| SFXN5 | 978,88 | -0,08 | 0,03 | -2,48 | 1,31E-02 | 7,14E-02 |
| EXOSC6 | 1253,97 | 0,05 | 0,02 | 2,48 | 1,31E-02 | 7,14E-02 |
| VAPA | 4676,72 | -0,06 | 0,03 | -2,48 | 1,31E-02 | 7,14E-02 |
| POLR2C | 859,42 | -0,04 | 0,02 | -2,48 | 1,31E-02 | 7,15E-02 |
| CPOX | 844,84 | 0,05 | 0,02 | 2,48 | 1,31E-02 | 7,15E-02 |
| C15orf40 | 1204,88 | 0,04 | 0,02 | 2,48 | 1,31E-02 | 7,16E-02 |
| TUBA1A | 4352,71 | -0,08 | 0,03 | -2,48 | 1,32E-02 | 7,17E-02 |
| RSPH3 | 584,62 | -0,08 | 0,03 | -2,48 | 1,32E-02 | 7,17E-02 |
| FBXO9 | 2829,50 | 0,08 | 0,03 | 2,48 | 1,32E-02 | 7,17E-02 |
| SLC11A2 | 1276,25 | 0,04 | 0,02 | 2,48 | 1,32E-02 | 7,18E-02 |
| CD200R1 | 712,48 | 0,10 | 0,04 | 2,48 | 1,33E-02 | 7,20E-02 |
| MRPL17 | 430,02 | 0,07 | 0,03 | 2,48 | 1,33E-02 | 7,22E-02 |
| TTC5 | 1056,73 | 0,04 | 0,02 | 2,48 | 1,33E-02 | 7,22E-02 |
| RPL11 | 5288,60 | 0,17 | 0,07 | 2,48 | 1,33E-02 | 7,22E-02 |
| HAUS4 | 783,36 | -0,10 | 0,04 | -2,48 | 1,33E-02 | 7,22E-02 |
| COX14 | 319,17 | 0,08 | 0,03 | 2,47 | 1,34E-02 | 7,23E-02 |
| TSEN15 | 952,41 | 0,07 | 0,03 | 2,47 | 1,33E-02 | 7,23E-02 |
| DOLK | 130,86 | 0,05 | 0,02 | 2,47 | 1,34E-02 | 7,23E-02 |
| RAB11FIP4 | 4097,25 | -0,07 | 0,03 | -2,47 | 1,34E-02 | 7,24E-02 |
| TPGS1 | 568,63 | 0,05 | 0,02 | 2,47 | 1,34E-02 | 7,24E-02 |
| CDK14 | 1213,33 | -0,11 | 0,04 | -2,47 | 1,34E-02 | 7,25E-02 |
| MCTP1 | 3296,77 | -0,09 | 0,04 | -2,47 | 1,34E-02 | 7,25E-02 |
| MARS1 | 2058,73 | -0,07 | 0,03 | -2,47 | 1,34E-02 | 7,25E-02 |
| NEIL2 | 315,05 | 0,08 | 0,03 | 2,47 | 1,35E-02 | 7,26E-02 |
| IL18RAP | 2456,58 | -0,14 | 0,06 | -2,47 | 1,34E-02 | 7,26E-02 |
| NIT1 | 599,86 | -0,04 | 0,02 | -2,47 | 1,34E-02 | 7,26E-02 |
| NCOA3 | 5350,98 | -0,05 | 0,02 | -2,47 | 1,35E-02 | 7,26E-02 |
| CHIC1 | 731,33 | 0,10 | 0,04 | 2,47 | 1,35E-02 | 7,26E-02 |
| KDM6A | 3053,29 | -0,05 | 0,02 | -2,47 | 1,35E-02 | 7,26E-02 |
| GUCD1 | 7125,85 | 0,14 | 0,06 | 2,47 | 1,35E-02 | 7,27E-02 |
| SRBD1 | 1529,81 | -0,07 | 0,03 | -2,47 | 1,35E-02 | 7,27E-02 |
| PURA | 2964,23 | 0,08 | 0,03 | 2,47 | 1,35E-02 | 7,27E-02 |
| CTNNAL1 | 227,76 | 0,18 | 0,07 | 2,47 | 1,35E-02 | 7,28E-02 |
| BEST1 | 1501,46 | -0,12 | 0,05 | -2,47 | 1,35E-02 | 7,28E-02 |
| MBOAT1 | 1159,55 | -0,06 | 0,02 | -2,47 | 1,36E-02 | 7,28E-02 |
| RPL24 | 4722,41 | 0,15 | 0,06 | 2,47 | 1,36E-02 | 7,28E-02 |
| CCDC14 | 1897,34 | 0,07 | 0,03 | 2,47 | 1,36E-02 | 7,28E-02 |
| L2HGDH | 219,34 | 0,06 | 0,03 | 2,47 | 1,36E-02 | 7,28E-02 |
| GCA | 17359,03 | -0,12 | 0,05 | -2,47 | 1,36E-02 | 7,28E-02 |
| PIWIL4 | 157,70 | 0,09 | 0,04 | 2,47 | 1,36E-02 | 7,29E-02 |
| RFC2 | 446,07 | -0,04 | 0,02 | -2,47 | 1,37E-02 | 7,31E-02 |
| ACOT9 | 1461,10 | -0,07 | 0,03 | -2,47 | 1,37E-02 | 7,31E-02 |
| PIGM | 622,24 | 0,05 | 0,02 | 2,47 | 1,37E-02 | 7,31E-02 |
| RAD17 | 663,57 | 0,06 | 0,02 | 2,47 | 1,37E-02 | 7,31E-02 |
| DNAJC15 | 710,75 | 0,11 | 0,04 | 2,47 | 1,37E-02 | 7,32E-02 |
| GBP1 | 3172,64 | -0,16 | 0,07 | -2,46 | 1,37E-02 | 7,33E-02 |

| | | | | | | |
|---|---|---|---|---|---|---|
| SNX10 | 3277,01 | -0,09 | 0,04 | -2,46 | 1,37E-02 | 7,33E-02 |
| UNC119B | 699,61 | -0,05 | 0,02 | -2,46 | 1,38E-02 | 7,34E-02 |
| ASXL2 | 3141,59 | -0,04 | 0,02 | -2,46 | 1,38E-02 | 7,34E-02 |
| SREK1IP1 | 940,40 | 0,08 | 0,03 | 2,46 | 1,38E-02 | 7,34E-02 |
| CALCRL | 110,57 | 0,13 | 0,05 | 2,46 | 1,38E-02 | 7,34E-02 |
| DLAT | 382,44 | 0,05 | 0,02 | 2,46 | 1,39E-02 | 7,36E-02 |
| MFSD14B | 2553,16 | -0,07 | 0,03 | -2,46 | 1,39E-02 | 7,36E-02 |
| HGH1 | 141,27 | 0,08 | 0,03 | 2,46 | 1,39E-02 | 7,36E-02 |
| NDUFB9 | 995,82 | 0,06 | 0,02 | 2,46 | 1,39E-02 | 7,36E-02 |
| PAPOLG | 1972,38 | -0,04 | 0,02 | -2,46 | 1,38E-02 | 7,36E-02 |
| MAGOHB | 459,94 | 0,08 | 0,03 | 2,46 | 1,38E-02 | 7,36E-02 |
| TCF12 | 2022,42 | 0,04 | 0,02 | 2,46 | 1,38E-02 | 7,36E-02 |
| ZNF688 | 350,32 | 0,05 | 0,02 | 2,46 | 1,39E-02 | 7,36E-02 |
| SVIP | 852,48 | 0,14 | 0,06 | 2,46 | 1,39E-02 | 7,36E-02 |
| COX15 | 741,36 | -0,03 | 0,01 | -2,46 | 1,39E-02 | 7,36E-02 |
| FAM174A | 831,95 | -0,09 | 0,04 | -2,46 | 1,39E-02 | 7,36E-02 |
| ZNF689 | 334,81 | 0,04 | 0,02 | 2,46 | 1,39E-02 | 7,36E-02 |
| MR1 | 2046,61 | -0,05 | 0,02 | -2,46 | 1,39E-02 | 7,36E-02 |
| NDUFB2 | 666,48 | 0,08 | 0,03 | 2,46 | 1,39E-02 | 7,37E-02 |
| SEH1L | 681,14 | 0,04 | 0,02 | 2,46 | 1,39E-02 | 7,37E-02 |
| EIF3J | 1021,85 | 0,07 | 0,03 | 2,46 | 1,40E-02 | 7,38E-02 |
| LPCAT2 | 4228,87 | -0,10 | 0,04 | -2,46 | 1,40E-02 | 7,39E-02 |
| CD180 | 629,99 | 0,10 | 0,04 | 2,46 | 1,40E-02 | 7,39E-02 |
| SFR1 | 159,15 | 0,10 | 0,04 | 2,46 | 1,40E-02 | 7,40E-02 |
| PPP4C | 2108,07 | -0,07 | 0,03 | -2,46 | 1,41E-02 | 7,40E-02 |
| ITGA4 | 12713,56 | 0,08 | 0,03 | 2,46 | 1,41E-02 | 7,40E-02 |
| TSSK6 | 90,46 | 0,07 | 0,03 | 2,46 | 1,41E-02 | 7,40E-02 |
| ZBTB22 | 359,07 | 0,06 | 0,02 | 2,46 | 1,40E-02 | 7,40E-02 |
| YBX3 | 13764,25 | 0,17 | 0,07 | 2,46 | 1,40E-02 | 7,40E-02 |
| CARS2 | 2893,70 | -0,06 | 0,03 | -2,46 | 1,40E-02 | 7,40E-02 |
| TIAL1 | 1956,24 | 0,07 | 0,03 | 2,46 | 1,40E-02 | 7,40E-02 |
| DYNC2LI1 | 101,04 | 0,10 | 0,04 | 2,46 | 1,41E-02 | 7,41E-02 |
| ZC2HC1A | 304,57 | 0,14 | 0,06 | 2,45 | 1,41E-02 | 7,41E-02 |
| TOPORS | 2778,00 | -0,10 | 0,04 | -2,45 | 1,41E-02 | 7,41E-02 |
| C14orf28 | 206,19 | 0,09 | 0,04 | 2,45 | 1,41E-02 | 7,42E-02 |
| C5AR2 | 4658,08 | -0,09 | 0,04 | -2,45 | 1,41E-02 | 7,43E-02 |
| BATF3 | 72,24 | 0,11 | 0,04 | 2,45 | 1,42E-02 | 7,43E-02 |
| STX16 | 3037,11 | -0,06 | 0,02 | -2,45 | 1,42E-02 | 7,43E-02 |
| ISL2 | 116,09 | -0,16 | 0,07 | -2,45 | 1,42E-02 | 7,43E-02 |
| TXNDC12 | 443,82 | -0,06 | 0,02 | -2,45 | 1,42E-02 | 7,43E-02 |
| COPB2 | 3355,95 | -0,05 | 0,02 | -2,45 | 1,42E-02 | 7,43E-02 |
| SNRPA1 | 799,79 | 0,05 | 0,02 | 2,45 | 1,42E-02 | 7,44E-02 |
| NCOR1 | 6138,20 | -0,06 | 0,02 | -2,45 | 1,42E-02 | 7,45E-02 |
| COMT | 626,88 | 0,06 | 0,02 | 2,45 | 1,42E-02 | 7,45E-02 |
| ZPR1 | 874,20 | 0,04 | 0,02 | 2,45 | 1,43E-02 | 7,46E-02 |
| YAE1 | 188,73 | 0,10 | 0,04 | 2,45 | 1,43E-02 | 7,46E-02 |

| | | | | | | |
|---|---|---|---|---|---|---|
| FAM8A1 | 4108,78 | -0,10 | 0,04 | -2,45 | 1,43E-02 | 7,48E-02 |
| CXCL1 | 106,51 | -0,12 | 0,05 | -2,45 | 1,43E-02 | 7,49E-02 |
| CFAP36 | 315,87 | 0,08 | 0,03 | 2,45 | 1,44E-02 | 7,49E-02 |
| PTPA | 781,06 | 0,06 | 0,02 | 2,45 | 1,44E-02 | 7,51E-02 |
| SACS | 2186,73 | 0,12 | 0,05 | 2,45 | 1,44E-02 | 7,51E-02 |
| POLR1C | 377,53 | 0,05 | 0,02 | 2,45 | 1,44E-02 | 7,52E-02 |
| ARK2N | 3436,63 | -0,05 | 0,02 | -2,45 | 1,44E-02 | 7,52E-02 |
| PRDX4 | 231,42 | 0,10 | 0,04 | 2,45 | 1,45E-02 | 7,54E-02 |
| NAA30 | 1160,69 | 0,07 | 0,03 | 2,44 | 1,45E-02 | 7,54E-02 |
| DBP | 406,13 | 0,07 | 0,03 | 2,44 | 1,45E-02 | 7,56E-02 |
| PARN | 1226,36 | -0,03 | 0,01 | -2,44 | 1,45E-02 | 7,56E-02 |
| HIP1 | 1380,65 | -0,17 | 0,07 | -2,44 | 1,46E-02 | 7,56E-02 |
| MPDU1 | 404,86 | 0,05 | 0,02 | 2,44 | 1,46E-02 | 7,56E-02 |
| ATP5MK | 557,80 | 0,10 | 0,04 | 2,44 | 1,46E-02 | 7,57E-02 |
| ZNF485 | 145,98 | 0,07 | 0,03 | 2,44 | 1,46E-02 | 7,57E-02 |
| AOC2 | 211,37 | -0,15 | 0,06 | -2,44 | 1,46E-02 | 7,57E-02 |
| SDR39U1 | 404,89 | 0,05 | 0,02 | 2,44 | 1,46E-02 | 7,57E-02 |
| B3GNTL1 | 851,60 | -0,12 | 0,05 | -2,44 | 1,46E-02 | 7,57E-02 |
| APC | 4123,13 | -0,07 | 0,03 | -2,44 | 1,46E-02 | 7,58E-02 |
| SH2D3A | 136,99 | 0,15 | 0,06 | 2,44 | 1,46E-02 | 7,58E-02 |
| POMGNT1 | 420,00 | 0,05 | 0,02 | 2,44 | 1,46E-02 | 7,58E-02 |
| RPL14 | 8139,03 | 0,09 | 0,04 | 2,44 | 1,47E-02 | 7,58E-02 |
| CCNJ | 468,69 | 0,05 | 0,02 | 2,44 | 1,47E-02 | 7,58E-02 |
| TKTL1 | 115,56 | -0,15 | 0,06 | -2,44 | 1,47E-02 | 7,58E-02 |
| CEP43 | 961,95 | -0,07 | 0,03 | -2,44 | 1,47E-02 | 7,59E-02 |
| ADNP | 3279,39 | -0,04 | 0,02 | -2,44 | 1,47E-02 | 7,59E-02 |
| DPF2 | 963,31 | -0,07 | 0,03 | -2,44 | 1,47E-02 | 7,60E-02 |
| UXT | 815,61 | 0,08 | 0,03 | 2,44 | 1,48E-02 | 7,61E-02 |
| ERLIN1 | 804,12 | -0,06 | 0,02 | -2,44 | 1,48E-02 | 7,61E-02 |
| ZBTB37 | 7631,44 | -0,05 | 0,02 | -2,44 | 1,48E-02 | 7,62E-02 |
| PIGH | 347,41 | 0,04 | 0,02 | 2,44 | 1,48E-02 | 7,62E-02 |
| DGCR6L | 369,81 | 0,06 | 0,02 | 2,44 | 1,48E-02 | 7,62E-02 |
| RPL5 | 7900,32 | 0,12 | 0,05 | 2,44 | 1,48E-02 | 7,63E-02 |
| WDR75 | 1390,91 | 0,07 | 0,03 | 2,44 | 1,49E-02 | 7,63E-02 |
| HIKESHI | 528,79 | 0,06 | 0,02 | 2,44 | 1,49E-02 | 7,63E-02 |
| BDH2 | 167,64 | 0,11 | 0,04 | 2,44 | 1,49E-02 | 7,63E-02 |
| SLC43A1 | 216,35 | 0,09 | 0,04 | 2,44 | 1,48E-02 | 7,63E-02 |
| HPSE | 897,19 | -0,10 | 0,04 | -2,43 | 1,49E-02 | 7,65E-02 |
| CFAP418 | 73,03 | 0,09 | 0,04 | 2,43 | 1,49E-02 | 7,65E-02 |
| ERO1A | 3773,54 | -0,07 | 0,03 | -2,43 | 1,49E-02 | 7,65E-02 |
| NUDT5 | 1730,53 | -0,05 | 0,02 | -2,43 | 1,49E-02 | 7,65E-02 |
| PHLPP2 | 792,94 | 0,08 | 0,03 | 2,43 | 1,50E-02 | 7,66E-02 |
| SRRD | 920,89 | 0,12 | 0,05 | 2,43 | 1,50E-02 | 7,67E-02 |
| VRK3 | 1690,80 | -0,06 | 0,02 | -2,43 | 1,50E-02 | 7,69E-02 |
| PPM1A | 9280,99 | 0,10 | 0,04 | 2,43 | 1,50E-02 | 7,69E-02 |
| PITHD1 | 4213,61 | 0,16 | 0,07 | 2,43 | 1,51E-02 | 7,71E-02 |

| | | | | | | |
|---|---|---|---|---|---|---|
| SLC22A23 | 240,49 | 0,14 | 0,06 | 2,43 | 1,51E-02 | 7,71E-02 |
| NFE2L2 | 2437,34 | -0,06 | 0,02 | -2,43 | 1,51E-02 | 7,71E-02 |
| RSRP1 | 12702,42 | -0,07 | 0,03 | -2,43 | 1,51E-02 | 7,71E-02 |
| LSR | 113,64 | 0,11 | 0,05 | 2,43 | 1,51E-02 | 7,71E-02 |
| FKBP1B | 73,57 | 0,15 | 0,06 | 2,43 | 1,51E-02 | 7,73E-02 |
| CHID1 | 478,45 | 0,07 | 0,03 | 2,43 | 1,52E-02 | 7,73E-02 |
| STOML2 | 985,20 | 0,08 | 0,03 | 2,43 | 1,52E-02 | 7,73E-02 |
| CEACAM3 | 1010,98 | -0,13 | 0,05 | -2,43 | 1,52E-02 | 7,73E-02 |
| NCAPG2 | 497,08 | -0,15 | 0,06 | -2,43 | 1,52E-02 | 7,73E-02 |
| USP7 | 4857,53 | -0,04 | 0,02 | -2,43 | 1,52E-02 | 7,75E-02 |
| ZNF273 | 334,98 | 0,09 | 0,04 | 2,43 | 1,53E-02 | 7,76E-02 |
| CUL5 | 1666,20 | 0,08 | 0,03 | 2,43 | 1,53E-02 | 7,76E-02 |
| CNIH1 | 712,43 | 0,09 | 0,04 | 2,43 | 1,53E-02 | 7,77E-02 |
| GPR146 | 666,07 | 0,19 | 0,08 | 2,43 | 1,53E-02 | 7,78E-02 |
| SMU1 | 2562,14 | -0,04 | 0,02 | -2,42 | 1,54E-02 | 7,80E-02 |
| TMPO | 5880,98 | 0,07 | 0,03 | 2,42 | 1,54E-02 | 7,84E-02 |
| VMP1 | 16966,10 | -0,08 | 0,03 | -2,42 | 1,55E-02 | 7,85E-02 |
| KIF22 | 666,27 | 0,05 | 0,02 | 2,42 | 1,55E-02 | 7,85E-02 |
| KRTCAP2 | 161,76 | 0,06 | 0,02 | 2,42 | 1,55E-02 | 7,86E-02 |
| COPS9 | 342,94 | 0,08 | 0,03 | 2,42 | 1,55E-02 | 7,86E-02 |
| MSR1 | 199,98 | -0,18 | 0,08 | -2,42 | 1,55E-02 | 7,86E-02 |
| TRIM59 | 411,83 | 0,08 | 0,03 | 2,42 | 1,55E-02 | 7,86E-02 |
| RFK | 438,39 | 0,08 | 0,03 | 2,42 | 1,55E-02 | 7,86E-02 |
| SLC35D2 | 212,21 | 0,04 | 0,02 | 2,42 | 1,56E-02 | 7,87E-02 |
| AFF1 | 5534,02 | -0,07 | 0,03 | -2,42 | 1,56E-02 | 7,89E-02 |
| PAIP1 | 770,94 | 0,07 | 0,03 | 2,42 | 1,56E-02 | 7,90E-02 |
| OLA1 | 504,92 | 0,07 | 0,03 | 2,42 | 1,56E-02 | 7,90E-02 |
| DIMT1 | 514,12 | 0,07 | 0,03 | 2,42 | 1,56E-02 | 7,90E-02 |
| MARCHF1 | 3924,48 | 0,10 | 0,04 | 2,42 | 1,57E-02 | 7,90E-02 |
| GPR82 | 88,53 | 0,17 | 0,07 | 2,42 | 1,57E-02 | 7,91E-02 |
| SPON1 | 73,43 | 0,17 | 0,07 | 2,42 | 1,57E-02 | 7,92E-02 |
| SGK1 | 2967,13 | -0,09 | 0,04 | -2,42 | 1,57E-02 | 7,92E-02 |
| WDR1 | 7148,39 | -0,06 | 0,02 | -2,42 | 1,57E-02 | 7,92E-02 |
| NECTIN3 | 112,69 | 0,12 | 0,05 | 2,42 | 1,57E-02 | 7,92E-02 |
| ANO5 | 111,55 | 0,15 | 0,06 | 2,41 | 1,58E-02 | 7,93E-02 |
| NTPCR | 739,64 | 0,07 | 0,03 | 2,41 | 1,58E-02 | 7,93E-02 |
| USP27X | 105,92 | 0,07 | 0,03 | 2,41 | 1,58E-02 | 7,93E-02 |
| MAK16 | 548,62 | 0,07 | 0,03 | 2,41 | 1,58E-02 | 7,94E-02 |
| LIN7A | 2448,61 | -0,11 | 0,05 | -2,41 | 1,58E-02 | 7,94E-02 |
| CPSF4 | 402,37 | 0,04 | 0,02 | 2,41 | 1,59E-02 | 7,95E-02 |
| LRRN1 | 93,17 | -0,14 | 0,06 | -2,41 | 1,59E-02 | 7,95E-02 |
| NOC3L | 753,51 | 0,09 | 0,04 | 2,41 | 1,59E-02 | 7,95E-02 |
| POLE4 | 319,62 | 0,06 | 0,03 | 2,41 | 1,59E-02 | 7,96E-02 |
| MRPL54 | 413,17 | 0,08 | 0,03 | 2,41 | 1,59E-02 | 7,96E-02 |
| CPEB4 | 5751,92 | -0,07 | 0,03 | -2,41 | 1,59E-02 | 7,96E-02 |
| RAP2A | 1686,83 | 0,11 | 0,05 | 2,41 | 1,60E-02 | 7,98E-02 |

| | | | | | | |
|---|---|---|---|---|---|---|
| SLITRK4 | 159,83 | 0,12 | 0,05 | 2,41 | 1,60E-02 | 7,98E-02 |
| PEX3 | 422,06 | 0,09 | 0,04 | 2,41 | 1,60E-02 | 7,98E-02 |
| PIK3CB | 2089,46 | -0,06 | 0,02 | -2,41 | 1,60E-02 | 7,99E-02 |
| SHISA4 | 122,64 | 0,21 | 0,09 | 2,41 | 1,60E-02 | 7,99E-02 |
| GSN | 2103,37 | -0,10 | 0,04 | -2,41 | 1,60E-02 | 7,99E-02 |
| CCDC146 | 515,03 | -0,09 | 0,04 | -2,41 | 1,60E-02 | 7,99E-02 |
| LAPTM5 | 8113,27 | -0,09 | 0,04 | -2,41 | 1,60E-02 | 7,99E-02 |
| FAM117A | 1529,76 | 0,12 | 0,05 | 2,41 | 1,61E-02 | 8,01E-02 |
| ARMC10 | 784,99 | 0,04 | 0,02 | 2,41 | 1,61E-02 | 8,02E-02 |
| TRAF6 | 1412,02 | -0,04 | 0,02 | -2,41 | 1,61E-02 | 8,03E-02 |
| HNRNPK | 15887,69 | -0,06 | 0,02 | -2,41 | 1,61E-02 | 8,03E-02 |
| ACTR5 | 431,93 | 0,04 | 0,02 | 2,41 | 1,61E-02 | 8,04E-02 |
| SUPT3H | 294,47 | 0,08 | 0,03 | 2,41 | 1,62E-02 | 8,04E-02 |
| RHBDD3 | 184,79 | 0,06 | 0,02 | 2,41 | 1,62E-02 | 8,04E-02 |
| SLC39A10 | 2658,62 | 0,08 | 0,03 | 2,40 | 1,62E-02 | 8,05E-02 |
| C5orf34 | 77,12 | 0,09 | 0,04 | 2,40 | 1,62E-02 | 8,05E-02 |
| UBL4A | 651,43 | 0,05 | 0,02 | 2,40 | 1,62E-02 | 8,07E-02 |
| MRPL3 | 836,32 | 0,09 | 0,04 | 2,40 | 1,63E-02 | 8,08E-02 |
| JMJD6 | 984,97 | -0,04 | 0,02 | -2,40 | 1,63E-02 | 8,08E-02 |
| ANXA11 | 5147,50 | -0,10 | 0,04 | -2,40 | 1,63E-02 | 8,09E-02 |
| TMEM183A | 2098,49 | 0,06 | 0,02 | 2,40 | 1,63E-02 | 8,09E-02 |
| ATAD3B | 696,91 | 0,07 | 0,03 | 2,40 | 1,63E-02 | 8,10E-02 |
| MRPS21 | 596,92 | 0,08 | 0,03 | 2,40 | 1,64E-02 | 8,14E-02 |
| ZNF175 | 392,70 | 0,05 | 0,02 | 2,40 | 1,64E-02 | 8,14E-02 |
| CORO1C | 2185,67 | -0,08 | 0,03 | -2,40 | 1,65E-02 | 8,15E-02 |
| HACD2 | 794,39 | 0,05 | 0,02 | 2,40 | 1,65E-02 | 8,16E-02 |
| CMTM6 | 12694,04 | -0,09 | 0,04 | -2,40 | 1,65E-02 | 8,16E-02 |
| ZDBF2 | 401,31 | 0,14 | 0,06 | 2,40 | 1,65E-02 | 8,18E-02 |
| MS4A1 | 2184,86 | 0,14 | 0,06 | 2,40 | 1,66E-02 | 8,19E-02 |
| ZNF783 | 951,72 | 0,05 | 0,02 | 2,40 | 1,66E-02 | 8,19E-02 |
| BTLA | 1045,13 | 0,13 | 0,05 | 2,40 | 1,66E-02 | 8,19E-02 |
| SLC39A3 | 198,92 | 0,07 | 0,03 | 2,39 | 1,66E-02 | 8,21E-02 |
| ANKRD46 | 627,15 | 0,07 | 0,03 | 2,39 | 1,67E-02 | 8,23E-02 |
| TRUB1 | 542,00 | 0,09 | 0,04 | 2,39 | 1,67E-02 | 8,23E-02 |
| LSM5 | 283,24 | 0,10 | 0,04 | 2,39 | 1,67E-02 | 8,23E-02 |
| PRIM1 | 222,81 | 0,08 | 0,03 | 2,39 | 1,67E-02 | 8,23E-02 |
| RBMS1 | 4547,08 | -0,05 | 0,02 | -2,39 | 1,67E-02 | 8,23E-02 |
| ARNT | 1792,53 | -0,04 | 0,02 | -2,39 | 1,67E-02 | 8,23E-02 |
| POU2F1 | 2600,04 | -0,06 | 0,03 | -2,39 | 1,68E-02 | 8,24E-02 |
| SPG21 | 2356,57 | -0,05 | 0,02 | -2,39 | 1,68E-02 | 8,25E-02 |
| SESN2 | 189,33 | -0,10 | 0,04 | -2,39 | 1,68E-02 | 8,25E-02 |
| GYG1 | 800,03 | -0,06 | 0,03 | -2,39 | 1,68E-02 | 8,26E-02 |
| BBIP1 | 1205,21 | -0,05 | 0,02 | -2,39 | 1,69E-02 | 8,29E-02 |
| MMS22L | 993,94 | 0,06 | 0,03 | 2,39 | 1,69E-02 | 8,29E-02 |
| SLC9A8 | 1383,85 | -0,09 | 0,04 | -2,39 | 1,69E-02 | 8,29E-02 |
| FAM216A | 241,11 | 0,07 | 0,03 | 2,39 | 1,69E-02 | 8,29E-02 |

| | | | | | | |
|---|---|---|---|---|---|---|
| RHOA | 8691,24 | -0,05 | 0,02 | -2,39 | 1,69E-02 | 8,30E-02 |
| ENTPD4 | 4196,03 | -0,04 | 0,02 | -2,39 | 1,70E-02 | 8,31E-02 |
| STX12 | 1462,73 | -0,04 | 0,02 | -2,39 | 1,70E-02 | 8,31E-02 |
| KLF12 | 3993,63 | 0,08 | 0,03 | 2,39 | 1,70E-02 | 8,31E-02 |
| KLHL21 | 1890,17 | -0,10 | 0,04 | -2,39 | 1,70E-02 | 8,31E-02 |
| PAF1 | 360,09 | -0,06 | 0,02 | -2,39 | 1,70E-02 | 8,31E-02 |
| LILRB3 | 2755,64 | -0,13 | 0,06 | -2,39 | 1,70E-02 | 8,31E-02 |
| ZDHHC5 | 1945,01 | -0,04 | 0,02 | -2,39 | 1,70E-02 | 8,33E-02 |
| ZNF726 | 97,77 | 0,09 | 0,04 | 2,38 | 1,71E-02 | 8,35E-02 |
| GALNT3 | 1321,99 | -0,08 | 0,03 | -2,38 | 1,71E-02 | 8,35E-02 |
| POP7 | 189,77 | 0,07 | 0,03 | 2,38 | 1,71E-02 | 8,35E-02 |
| COPA | 4812,35 | -0,05 | 0,02 | -2,38 | 1,71E-02 | 8,35E-02 |
| ZNF501 | 140,33 | 0,12 | 0,05 | 2,38 | 1,71E-02 | 8,35E-02 |
| GABARAPL2 | 2261,00 | 0,10 | 0,04 | 2,38 | 1,71E-02 | 8,36E-02 |
| IAH1 | 695,23 | 0,04 | 0,02 | 2,38 | 1,72E-02 | 8,36E-02 |
| THAP2 | 312,33 | 0,09 | 0,04 | 2,38 | 1,72E-02 | 8,38E-02 |
| CRYBG3 | 1223,74 | 0,07 | 0,03 | 2,38 | 1,72E-02 | 8,38E-02 |
| TOE1 | 255,56 | 0,04 | 0,02 | 2,38 | 1,72E-02 | 8,38E-02 |
| SH3GLB1 | 5031,82 | -0,07 | 0,03 | -2,38 | 1,72E-02 | 8,38E-02 |
| CHAMP1 | 260,92 | 0,05 | 0,02 | 2,38 | 1,73E-02 | 8,39E-02 |
| IRF7 | 486,26 | -0,15 | 0,06 | -2,38 | 1,73E-02 | 8,39E-02 |
| SERPINB1 | 4088,33 | -0,07 | 0,03 | -2,38 | 1,73E-02 | 8,39E-02 |
| BTBD10 | 1883,44 | -0,07 | 0,03 | -2,38 | 1,73E-02 | 8,39E-02 |
| OAZ2 | 4283,97 | -0,09 | 0,04 | -2,38 | 1,73E-02 | 8,40E-02 |
| PRR3 | 363,09 | 0,04 | 0,02 | 2,38 | 1,73E-02 | 8,41E-02 |
| CENPX | 272,98 | 0,06 | 0,03 | 2,38 | 1,73E-02 | 8,41E-02 |
| PUS7L | 1912,27 | 0,09 | 0,04 | 2,38 | 1,73E-02 | 8,41E-02 |
| DNAAF10 | 172,24 | 0,05 | 0,02 | 2,38 | 1,74E-02 | 8,42E-02 |
| SLC35A3 | 1438,33 | 0,05 | 0,02 | 2,38 | 1,74E-02 | 8,42E-02 |
| APOOL | 305,57 | 0,05 | 0,02 | 2,38 | 1,74E-02 | 8,43E-02 |
| WLS | 1913,57 | -0,16 | 0,07 | -2,38 | 1,74E-02 | 8,44E-02 |
| TAL1 | 1522,81 | 0,15 | 0,06 | 2,38 | 1,75E-02 | 8,45E-02 |
| OSBPL1A | 403,22 | -0,08 | 0,03 | -2,38 | 1,75E-02 | 8,46E-02 |
| CHD2 | 13827,49 | -0,04 | 0,02 | -2,38 | 1,75E-02 | 8,46E-02 |
| KATNIP | 881,45 | -0,09 | 0,04 | -2,37 | 1,76E-02 | 8,48E-02 |
| ATP5F1D | 90,00 | 0,13 | 0,06 | 2,37 | 1,76E-02 | 8,48E-02 |
| KIN | 1086,37 | -0,05 | 0,02 | -2,37 | 1,76E-02 | 8,49E-02 |
| C15orf61 | 181,15 | 0,09 | 0,04 | 2,37 | 1,76E-02 | 8,49E-02 |
| ZBED5 | 1846,32 | 0,10 | 0,04 | 2,37 | 1,76E-02 | 8,51E-02 |
| RELL2 | 240,74 | 0,07 | 0,03 | 2,37 | 1,77E-02 | 8,51E-02 |
| CCNK | 1244,97 | -0,04 | 0,02 | -2,37 | 1,77E-02 | 8,52E-02 |
| ZNF365 | 77,55 | 0,15 | 0,06 | 2,37 | 1,77E-02 | 8,52E-02 |
| SLC50A1 | 377,48 | 0,05 | 0,02 | 2,37 | 1,77E-02 | 8,52E-02 |
| ZNF572 | 141,13 | 0,10 | 0,04 | 2,37 | 1,77E-02 | 8,52E-02 |
| RPS19 | 10524,57 | 0,09 | 0,04 | 2,37 | 1,78E-02 | 8,54E-02 |
| ZNF451 | 4908,52 | 0,06 | 0,03 | 2,37 | 1,78E-02 | 8,54E-02 |

| | | | | | | |
|---|---|---|---|---|---|---|
| PTER | 607,26 | 0,07 | 0,03 | 2,37 | 1,78E-02 | 8,56E-02 |
| STX10 | 1827,47 | -0,07 | 0,03 | -2,37 | 1,79E-02 | 8,59E-02 |
| TNFSF13B | 3465,97 | -0,12 | 0,05 | -2,37 | 1,79E-02 | 8,60E-02 |
| MFAP1 | 965,35 | -0,07 | 0,03 | -2,37 | 1,79E-02 | 8,61E-02 |
| STAT1 | 13004,00 | -0,11 | 0,05 | -2,37 | 1,80E-02 | 8,62E-02 |
| TIMM44 | 307,09 | 0,05 | 0,02 | 2,37 | 1,80E-02 | 8,62E-02 |
| MUS81 | 635,12 | 0,05 | 0,02 | 2,37 | 1,80E-02 | 8,62E-02 |
| CIAO2A | 692,77 | 0,09 | 0,04 | 2,37 | 1,80E-02 | 8,62E-02 |
| DUSP23 | 232,50 | 0,06 | 0,02 | 2,37 | 1,80E-02 | 8,62E-02 |
| CFAP97 | 1114,08 | 0,08 | 0,04 | 2,36 | 1,80E-02 | 8,63E-02 |
| ACTN4 | 2502,84 | -0,11 | 0,05 | -2,36 | 1,80E-02 | 8,63E-02 |
| ST6GALNAC2 | 506,03 | -0,12 | 0,05 | -2,36 | 1,80E-02 | 8,63E-02 |
| FANCD2 | 763,99 | -0,04 | 0,02 | -2,36 | 1,81E-02 | 8,66E-02 |
| PHF13 | 240,77 | -0,06 | 0,03 | -2,36 | 1,81E-02 | 8,66E-02 |
| NFKBIA | 2616,51 | -0,06 | 0,03 | -2,36 | 1,81E-02 | 8,66E-02 |
| MRTO4 | 347,22 | 0,06 | 0,03 | 2,36 | 1,82E-02 | 8,67E-02 |
| ADISSP | 149,23 | 0,11 | 0,05 | 2,36 | 1,82E-02 | 8,67E-02 |
| ATXN1L | 1499,00 | -0,06 | 0,02 | -2,36 | 1,82E-02 | 8,67E-02 |
| SUPT4H1 | 132,35 | -0,08 | 0,03 | -2,36 | 1,82E-02 | 8,67E-02 |
| VPS50 | 1729,71 | 0,09 | 0,04 | 2,36 | 1,82E-02 | 8,68E-02 |
| CIAO2B | 548,54 | 0,06 | 0,02 | 2,36 | 1,82E-02 | 8,68E-02 |
| LPAR6 | 1221,95 | 0,18 | 0,08 | 2,36 | 1,82E-02 | 8,68E-02 |
| MPI | 407,21 | 0,06 | 0,03 | 2,36 | 1,83E-02 | 8,70E-02 |
| BMI1 | 758,48 | 0,10 | 0,04 | 2,36 | 1,83E-02 | 8,70E-02 |
| GCNA | 252,54 | -0,05 | 0,02 | -2,36 | 1,83E-02 | 8,70E-02 |
| NR6A1 | 262,58 | -0,12 | 0,05 | -2,36 | 1,83E-02 | 8,70E-02 |
| CYTH4 | 11774,18 | -0,07 | 0,03 | -2,36 | 1,83E-02 | 8,71E-02 |
| ECE1 | 2912,69 | -0,11 | 0,05 | -2,36 | 1,84E-02 | 8,72E-02 |
| ZC3HC1 | 274,21 | 0,04 | 0,02 | 2,36 | 1,84E-02 | 8,72E-02 |
| ANKRD16 | 132,56 | 0,07 | 0,03 | 2,36 | 1,84E-02 | 8,73E-02 |
| BBS10 | 649,38 | 0,09 | 0,04 | 2,36 | 1,84E-02 | 8,74E-02 |
| ICE2 | 2018,59 | 0,06 | 0,03 | 2,36 | 1,84E-02 | 8,74E-02 |
| MAPK13 | 1112,03 | -0,13 | 0,06 | -2,36 | 1,85E-02 | 8,75E-02 |
| S100A10 | 754,14 | 0,06 | 0,03 | 2,36 | 1,85E-02 | 8,75E-02 |
| KCTD21 | 263,10 | -0,08 | 0,03 | -2,36 | 1,85E-02 | 8,75E-02 |
| SEC24A | 836,33 | 0,05 | 0,02 | 2,36 | 1,85E-02 | 8,75E-02 |
| CNPY2 | 256,74 | 0,05 | 0,02 | 2,35 | 1,86E-02 | 8,78E-02 |
| TMEM120A | 434,74 | -0,07 | 0,03 | -2,35 | 1,86E-02 | 8,79E-02 |
| MRPS18A | 320,41 | 0,04 | 0,02 | 2,35 | 1,86E-02 | 8,79E-02 |
| ZNF519 | 451,01 | 0,07 | 0,03 | 2,35 | 1,86E-02 | 8,79E-02 |
| TUBA4A | 2282,73 | -0,06 | 0,03 | -2,35 | 1,86E-02 | 8,80E-02 |
| ZNRF2 | 1828,59 | 0,07 | 0,03 | 2,35 | 1,86E-02 | 8,80E-02 |
| POLR2K | 248,02 | 0,18 | 0,08 | 2,35 | 1,87E-02 | 8,81E-02 |
| MTRR | 1240,39 | 0,05 | 0,02 | 2,35 | 1,87E-02 | 8,82E-02 |
| XIAP | 4065,34 | -0,04 | 0,02 | -2,35 | 1,87E-02 | 8,82E-02 |
| INTS6L | 1880,81 | 0,05 | 0,02 | 2,35 | 1,87E-02 | 8,82E-02 |

| | | | | | | |
|---|---|---|---|---|---|---|
| KIF3C | 173,78 | -0,11 | 0,05 | -2,35 | 1,87E-02 | 8,83E-02 |
| RAB20 | 378,13 | -0,11 | 0,05 | -2,35 | 1,88E-02 | 8,83E-02 |
| CYC1 | 741,45 | 0,05 | 0,02 | 2,35 | 1,88E-02 | 8,83E-02 |
| AGTPBP1 | 7389,04 | -0,07 | 0,03 | -2,35 | 1,88E-02 | 8,86E-02 |
| SSBP4 | 387,54 | 0,09 | 0,04 | 2,35 | 1,88E-02 | 8,86E-02 |
| ATP6V0B | 4586,15 | -0,07 | 0,03 | -2,35 | 1,89E-02 | 8,87E-02 |
| ZNF141 | 1111,37 | 0,11 | 0,05 | 2,35 | 1,89E-02 | 8,88E-02 |
| EXOSC5 | 191,02 | 0,05 | 0,02 | 2,35 | 1,89E-02 | 8,89E-02 |
| RBM10 | 1227,99 | -0,05 | 0,02 | -2,35 | 1,89E-02 | 8,89E-02 |
| CERT1 | 4693,72 | -0,05 | 0,02 | -2,35 | 1,90E-02 | 8,89E-02 |
| KDM8 | 198,64 | 0,06 | 0,03 | 2,35 | 1,90E-02 | 8,89E-02 |
| RPL23 | 3272,06 | 0,22 | 0,09 | 2,35 | 1,90E-02 | 8,89E-02 |
| YWHAZ | 16921,41 | -0,04 | 0,02 | -2,35 | 1,90E-02 | 8,91E-02 |
| ADPGK | 5673,84 | -0,04 | 0,02 | -2,35 | 1,90E-02 | 8,91E-02 |
| METTL5 | 247,13 | 0,07 | 0,03 | 2,35 | 1,90E-02 | 8,91E-02 |
| EIF5A2 | 248,89 | 0,10 | 0,04 | 2,34 | 1,90E-02 | 8,91E-02 |
| FANCA | 318,27 | -0,13 | 0,06 | -2,34 | 1,91E-02 | 8,93E-02 |
| ZNF133 | 580,96 | 0,04 | 0,02 | 2,34 | 1,91E-02 | 8,94E-02 |
| SNX1 | 4020,07 | -0,04 | 0,02 | -2,34 | 1,91E-02 | 8,94E-02 |
| YIF1B | 147,64 | 0,09 | 0,04 | 2,34 | 1,92E-02 | 8,97E-02 |
| UST | 296,22 | 0,08 | 0,03 | 2,34 | 1,93E-02 | 9,00E-02 |
| ASAH2B | 104,10 | 0,12 | 0,05 | 2,34 | 1,93E-02 | 9,01E-02 |
| DDAH2 | 711,08 | -0,06 | 0,03 | -2,34 | 1,94E-02 | 9,05E-02 |
| IFT70B | 146,74 | 0,09 | 0,04 | 2,34 | 1,94E-02 | 9,06E-02 |
| USF1 | 3540,07 | -0,06 | 0,02 | -2,34 | 1,95E-02 | 9,08E-02 |
| MTFMT | 514,30 | -0,04 | 0,02 | -2,34 | 1,95E-02 | 9,08E-02 |
| ZNF302 | 978,04 | 0,10 | 0,04 | 2,34 | 1,95E-02 | 9,08E-02 |
| SYPL1 | 748,78 | 0,07 | 0,03 | 2,34 | 1,95E-02 | 9,09E-02 |
| RANBP10 | 1178,85 | 0,14 | 0,06 | 2,33 | 1,95E-02 | 9,10E-02 |
| ZNHIT1 | 745,39 | 0,04 | 0,02 | 2,33 | 1,96E-02 | 9,11E-02 |
| PHOSPHO2 | 97,41 | 0,09 | 0,04 | 2,33 | 1,96E-02 | 9,11E-02 |
| TMEM68 | 442,03 | 0,05 | 0,02 | 2,33 | 1,96E-02 | 9,12E-02 |
| STRN | 3405,88 | -0,05 | 0,02 | -2,33 | 1,97E-02 | 9,13E-02 |
| E2F2 | 1201,40 | 0,18 | 0,08 | 2,33 | 1,97E-02 | 9,13E-02 |
| CRPPA | 122,99 | 0,12 | 0,05 | 2,33 | 1,97E-02 | 9,13E-02 |
| NUDT9 | 281,64 | 0,05 | 0,02 | 2,33 | 1,97E-02 | 9,13E-02 |
| C19orf53 | 311,19 | 0,07 | 0,03 | 2,33 | 1,97E-02 | 9,13E-02 |
| NOTCH2 | 12419,67 | -0,09 | 0,04 | -2,33 | 1,97E-02 | 9,15E-02 |
| MMAB | 233,56 | 0,06 | 0,03 | 2,33 | 1,97E-02 | 9,15E-02 |
| HSF5 | 104,20 | 0,09 | 0,04 | 2,33 | 1,97E-02 | 9,15E-02 |
| FCGR1A | 596,68 | -0,20 | 0,08 | -2,33 | 1,98E-02 | 9,15E-02 |
| PSMD13 | 1850,45 | -0,04 | 0,02 | -2,33 | 1,98E-02 | 9,15E-02 |
| MZT1 | 234,43 | 0,12 | 0,05 | 2,33 | 1,98E-02 | 9,15E-02 |
| ABRAXAS1 | 852,38 | 0,07 | 0,03 | 2,33 | 1,98E-02 | 9,15E-02 |
| PIAS1 | 6030,40 | -0,06 | 0,02 | -2,33 | 1,98E-02 | 9,15E-02 |
| ELP4 | 1745,74 | 0,08 | 0,03 | 2,33 | 1,98E-02 | 9,16E-02 |

| | | | | | | |
|---|---|---|---|---|---|---|
| ETV6 | 1256,25 | -0,11 | 0,05 | -2,33 | 1,98E-02 | 9,16E-02 |
| MSANTD3 | 205,34 | 0,06 | 0,03 | 2,33 | 1,99E-02 | 9,17E-02 |
| SMIM20 | 166,20 | 0,09 | 0,04 | 2,33 | 1,99E-02 | 9,18E-02 |
| DDIT3 | 391,33 | -0,08 | 0,04 | -2,33 | 1,99E-02 | 9,19E-02 |
| MARCHF8 | 11659,00 | 0,15 | 0,06 | 2,33 | 1,99E-02 | 9,19E-02 |
| B4GALT1 | 1864,77 | -0,08 | 0,03 | -2,33 | 1,99E-02 | 9,19E-02 |
| PHF14 | 2227,59 | 0,08 | 0,04 | 2,33 | 2,00E-02 | 9,20E-02 |
| EPM2AIP1 | 2276,76 | 0,07 | 0,03 | 2,33 | 2,00E-02 | 9,20E-02 |
| RFLNB | 1830,03 | -0,11 | 0,05 | -2,33 | 2,00E-02 | 9,20E-02 |
| TSPAN3 | 704,96 | 0,07 | 0,03 | 2,33 | 2,00E-02 | 9,20E-02 |
| RBIS | 483,31 | 0,10 | 0,04 | 2,33 | 2,00E-02 | 9,21E-02 |
| MRM1 | 163,99 | 0,06 | 0,03 | 2,32 | 2,01E-02 | 9,22E-02 |
| RPS6KA4 | 621,33 | 0,07 | 0,03 | 2,32 | 2,01E-02 | 9,22E-02 |
| PDE12 | 1183,90 | 0,03 | 0,01 | 2,32 | 2,01E-02 | 9,22E-02 |
| TMEM67 | 177,85 | 0,08 | 0,03 | 2,32 | 2,01E-02 | 9,22E-02 |
| PRPF18 | 809,39 | -0,07 | 0,03 | -2,33 | 2,01E-02 | 9,22E-02 |
| ZNF587 | 1998,97 | -0,05 | 0,02 | -2,33 | 2,01E-02 | 9,22E-02 |
| ZFP69 | 177,23 | 0,05 | 0,02 | 2,32 | 2,01E-02 | 9,22E-02 |
| FAM168B | 2065,26 | -0,04 | 0,02 | -2,32 | 2,01E-02 | 9,23E-02 |
| ZNF280D | 959,66 | 0,09 | 0,04 | 2,32 | 2,02E-02 | 9,24E-02 |
| VIPAS39 | 906,43 | -0,03 | 0,01 | -2,32 | 2,02E-02 | 9,25E-02 |
| NUS1 | 786,75 | 0,08 | 0,04 | 2,32 | 2,02E-02 | 9,25E-02 |
| ZNF3 | 835,98 | 0,03 | 0,01 | 2,32 | 2,02E-02 | 9,25E-02 |
| MRPL45 | 199,61 | 0,06 | 0,03 | 2,32 | 2,02E-02 | 9,25E-02 |
| UFL1 | 1507,03 | 0,09 | 0,04 | 2,32 | 2,03E-02 | 9,27E-02 |
| EEF1A1 | 129629,62 | 0,13 | 0,05 | 2,32 | 2,03E-02 | 9,28E-02 |
| EXOSC4 | 173,22 | 0,06 | 0,03 | 2,32 | 2,03E-02 | 9,28E-02 |
| RPAIN | 1477,69 | 0,05 | 0,02 | 2,32 | 2,03E-02 | 9,28E-02 |
| ZNF32 | 271,46 | 0,10 | 0,04 | 2,32 | 2,03E-02 | 9,28E-02 |
| XRCC5 | 8658,06 | -0,06 | 0,02 | -2,32 | 2,03E-02 | 9,28E-02 |
| ZNF354C | 510,96 | 0,11 | 0,05 | 2,32 | 2,03E-02 | 9,28E-02 |
| GLCE | 483,48 | 0,06 | 0,03 | 2,32 | 2,04E-02 | 9,29E-02 |
| PIGL | 578,56 | 0,06 | 0,02 | 2,32 | 2,04E-02 | 9,31E-02 |
| TRNAU1AP | 650,36 | 0,03 | 0,01 | 2,32 | 2,04E-02 | 9,31E-02 |
| NUP54 | 837,03 | 0,07 | 0,03 | 2,32 | 2,05E-02 | 9,32E-02 |
| NUAK2 | 1674,21 | -0,11 | 0,05 | -2,32 | 2,05E-02 | 9,32E-02 |
| STK25 | 882,85 | 0,06 | 0,03 | 2,32 | 2,05E-02 | 9,35E-02 |
| MRPS30 | 1490,60 | 0,07 | 0,03 | 2,31 | 2,06E-02 | 9,38E-02 |
| DEF8 | 3396,23 | -0,08 | 0,04 | -2,31 | 2,06E-02 | 9,38E-02 |
| CARD9 | 720,79 | 0,08 | 0,04 | 2,31 | 2,06E-02 | 9,39E-02 |
| COPS8 | 786,94 | 0,07 | 0,03 | 2,31 | 2,07E-02 | 9,39E-02 |
| IFI6 | 2546,50 | -0,22 | 0,10 | -2,31 | 2,07E-02 | 9,40E-02 |
| USB1 | 951,24 | -0,09 | 0,04 | -2,31 | 2,07E-02 | 9,40E-02 |
| FIS1 | 2278,53 | 0,11 | 0,05 | 2,31 | 2,08E-02 | 9,42E-02 |
| TBC1D32 | 633,54 | 0,08 | 0,03 | 2,31 | 2,08E-02 | 9,42E-02 |
| GHDC | 151,38 | 0,10 | 0,04 | 2,31 | 2,08E-02 | 9,42E-02 |

| | | | | | | |
|---|---|---|---|---|---|---|
| ZNF546 | 393,47 | 0,06 | 0,03 | 2,31 | 2,08E-02 | 9,42E-02 |
| INPP5D | 4712,22 | -0,10 | 0,04 | -2,31 | 2,08E-02 | 9,43E-02 |
| FCGR2B | 623,82 | -0,12 | 0,05 | -2,31 | 2,08E-02 | 9,44E-02 |
| UTP15 | 542,01 | 0,07 | 0,03 | 2,31 | 2,08E-02 | 9,44E-02 |
| NDE1 | 2600,77 | -0,10 | 0,04 | -2,31 | 2,09E-02 | 9,45E-02 |
| GCAT | 139,70 | 0,20 | 0,09 | 2,31 | 2,09E-02 | 9,47E-02 |
| PI4K2B | 510,10 | 0,11 | 0,05 | 2,31 | 2,10E-02 | 9,48E-02 |
| GPS1 | 621,91 | 0,05 | 0,02 | 2,31 | 2,10E-02 | 9,48E-02 |
| MTO1 | 1277,04 | 0,04 | 0,02 | 2,31 | 2,10E-02 | 9,48E-02 |
| MRPL9 | 398,87 | 0,05 | 0,02 | 2,31 | 2,10E-02 | 9,48E-02 |
| ZNF680 | 557,01 | 0,11 | 0,05 | 2,31 | 2,10E-02 | 9,48E-02 |
| FPGS | 509,45 | 0,06 | 0,03 | 2,31 | 2,10E-02 | 9,48E-02 |
| ARL5A | 1966,94 | 0,08 | 0,03 | 2,31 | 2,10E-02 | 9,48E-02 |
| FAM98B | 701,74 | 0,07 | 0,03 | 2,31 | 2,10E-02 | 9,48E-02 |
| MFSD14A | 285,34 | -0,06 | 0,03 | -2,31 | 2,11E-02 | 9,49E-02 |
| BAG1 | 6544,86 | 0,14 | 0,06 | 2,31 | 2,11E-02 | 9,50E-02 |
| SIGLEC7 | 328,59 | -0,09 | 0,04 | -2,31 | 2,11E-02 | 9,50E-02 |
| BRIX1 | 367,60 | 0,09 | 0,04 | 2,31 | 2,11E-02 | 9,50E-02 |
| CEP83 | 564,93 | 0,06 | 0,03 | 2,30 | 2,12E-02 | 9,56E-02 |
| ULK2 | 836,41 | 0,05 | 0,02 | 2,30 | 2,13E-02 | 9,56E-02 |
| ZNF653 | 125,40 | 0,08 | 0,04 | 2,30 | 2,13E-02 | 9,56E-02 |
| CEP97 | 1023,25 | -0,06 | 0,03 | -2,30 | 2,13E-02 | 9,56E-02 |
| RNF227 | 113,53 | 0,05 | 0,02 | 2,30 | 2,13E-02 | 9,57E-02 |
| WDR48 | 2207,81 | -0,04 | 0,02 | -2,30 | 2,13E-02 | 9,57E-02 |
| UBQLN1 | 4736,12 | 0,05 | 0,02 | 2,30 | 2,13E-02 | 9,58E-02 |
| NIPAL2 | 539,34 | 0,09 | 0,04 | 2,30 | 2,14E-02 | 9,58E-02 |
| TMEM63B | 880,62 | 0,16 | 0,07 | 2,30 | 2,14E-02 | 9,60E-02 |
| TCEANC | 603,49 | 0,06 | 0,03 | 2,30 | 2,14E-02 | 9,61E-02 |
| ANKRD36C | 2197,33 | 0,07 | 0,03 | 2,30 | 2,15E-02 | 9,61E-02 |
| WASHC2A | 1190,70 | -0,05 | 0,02 | -2,30 | 2,15E-02 | 9,63E-02 |
| MIF4GD | 205,43 | 0,05 | 0,02 | 2,30 | 2,15E-02 | 9,63E-02 |
| C2CD3 | 1440,03 | -0,06 | 0,02 | -2,30 | 2,15E-02 | 9,63E-02 |
| PLXNA2 | 430,16 | -0,12 | 0,05 | -2,30 | 2,15E-02 | 9,64E-02 |
| COPRS | 88,91 | 0,07 | 0,03 | 2,30 | 2,16E-02 | 9,66E-02 |
| PIGX | 1502,12 | -0,07 | 0,03 | -2,30 | 2,16E-02 | 9,66E-02 |
| ZNF281 | 2388,97 | -0,08 | 0,04 | -2,30 | 2,16E-02 | 9,66E-02 |
| CDK4 | 175,83 | 0,06 | 0,02 | 2,30 | 2,16E-02 | 9,66E-02 |
| KCTD17 | 152,81 | 0,06 | 0,03 | 2,30 | 2,16E-02 | 9,66E-02 |
| HCFC2 | 1025,43 | 0,07 | 0,03 | 2,30 | 2,17E-02 | 9,68E-02 |
| SUDS3 | 2875,95 | -0,04 | 0,02 | -2,30 | 2,17E-02 | 9,69E-02 |
| ZNF287 | 240,50 | 0,08 | 0,03 | 2,30 | 2,17E-02 | 9,69E-02 |
| RASSF7 | 294,87 | 0,06 | 0,03 | 2,30 | 2,17E-02 | 9,69E-02 |
| WARS1 | 3848,71 | -0,11 | 0,05 | -2,29 | 2,17E-02 | 9,69E-02 |
| CAPS2 | 240,67 | 0,08 | 0,03 | 2,29 | 2,18E-02 | 9,70E-02 |
| ARHGEF11 | 1157,32 | -0,11 | 0,05 | -2,29 | 2,18E-02 | 9,70E-02 |
| PLIN2 | 814,85 | -0,06 | 0,03 | -2,29 | 2,18E-02 | 9,71E-02 |

| | | | | | | |
|---|---|---|---|---|---|---|
| DUSP28 | 292,61 | 0,04 | 0,02 | 2,29 | 2,18E-02 | 9,71E-02 |
| PSME1 | 4963,14 | -0,07 | 0,03 | -2,29 | 2,18E-02 | 9,71E-02 |
| FKBP9 | 128,72 | -0,13 | 0,06 | -2,29 | 2,19E-02 | 9,71E-02 |
| NUP43 | 1400,01 | 0,05 | 0,02 | 2,29 | 2,19E-02 | 9,73E-02 |
| CDPF1 | 107,63 | 0,06 | 0,03 | 2,29 | 2,19E-02 | 9,74E-02 |
| TMEM106B | 2248,21 | 0,09 | 0,04 | 2,29 | 2,19E-02 | 9,74E-02 |
| HEATR3 | 1011,02 | 0,05 | 0,02 | 2,29 | 2,20E-02 | 9,76E-02 |
| IFIH1 | 1755,18 | -0,12 | 0,05 | -2,29 | 2,20E-02 | 9,77E-02 |
| LXN | 192,66 | -0,09 | 0,04 | -2,29 | 2,20E-02 | 9,77E-02 |
| GTPBP3 | 148,42 | 0,09 | 0,04 | 2,29 | 2,21E-02 | 9,79E-02 |
| MIA2 | 2381,89 | -0,05 | 0,02 | -2,29 | 2,21E-02 | 9,80E-02 |
| RPL12 | 3639,65 | 0,09 | 0,04 | 2,29 | 2,22E-02 | 9,85E-02 |
| EEF2KMT | 121,71 | 0,06 | 0,03 | 2,29 | 2,23E-02 | 9,87E-02 |
| CRTAM | 203,75 | 0,11 | 0,05 | 2,28 | 2,24E-02 | 9,90E-02 |
| NME1 | 105,05 | 0,10 | 0,04 | 2,28 | 2,24E-02 | 9,91E-02 |
| ATP6V0A1 | 1625,91 | -0,07 | 0,03 | -2,28 | 2,24E-02 | 9,91E-02 |
| LSM1 | 447,06 | 0,09 | 0,04 | 2,28 | 2,24E-02 | 9,91E-02 |
| PLEK2 | 458,62 | 0,16 | 0,07 | 2,28 | 2,24E-02 | 9,91E-02 |
| NSMF | 375,32 | 0,10 | 0,04 | 2,28 | 2,25E-02 | 9,92E-02 |
| RAB8A | 1359,72 | -0,06 | 0,03 | -2,28 | 2,25E-02 | 9,93E-02 |
| HSPA1L | 225,70 | -0,06 | 0,03 | -2,28 | 2,25E-02 | 9,95E-02 |
| WBP1 | 332,48 | 0,04 | 0,02 | 2,28 | 2,26E-02 | 9,99E-02 |
| HLA-DQA1 | 1884,45 | 0,11 | 0,05 | 2,28 | 2,27E-02 | 1,00E-01 |
| PABIR1 | 717,41 | 0,03 | 0,01 | 2,28 | 2,27E-02 | 1,00E-01 |
| TMEM218 | 677,56 | 0,04 | 0,02 | 2,28 | 2,27E-02 | 1,00E-01 |
| DBNL | 10903,82 | -0,06 | 0,03 | -2,28 | 2,28E-02 | 1,00E-01 |
| KBTBD8 | 374,79 | 0,15 | 0,06 | 2,28 | 2,28E-02 | 1,00E-01 |
| CUL3 | 3765,98 | -0,06 | 0,02 | -2,28 | 2,28E-02 | 1,00E-01 |
| CENPU | 79,55 | 0,10 | 0,04 | 2,28 | 2,28E-02 | 1,00E-01 |
| ARPC3 | 5211,04 | -0,08 | 0,04 | -2,28 | 2,28E-02 | 1,00E-01 |
| GMPS | 1147,56 | 0,05 | 0,02 | 2,28 | 2,28E-02 | 1,00E-01 |
| PPP2R3C | 1395,28 | -0,08 | 0,03 | -2,28 | 2,29E-02 | 1,01E-01 |
| IFI35 | 865,46 | -0,12 | 0,05 | -2,27 | 2,29E-02 | 1,01E-01 |
| SLC35E3 | 932,05 | -0,08 | 0,03 | -2,27 | 2,30E-02 | 1,01E-01 |
| GNPDA2 | 507,89 | 0,10 | 0,04 | 2,27 | 2,30E-02 | 1,01E-01 |
| ZNF202 | 396,23 | 0,04 | 0,02 | 2,27 | 2,30E-02 | 1,01E-01 |
| MACIR | 212,30 | 0,06 | 0,03 | 2,27 | 2,30E-02 | 1,01E-01 |
| SNX22 | 513,57 | 0,10 | 0,04 | 2,27 | 2,31E-02 | 1,01E-01 |
| HTRA2 | 752,60 | -0,04 | 0,02 | -2,27 | 2,30E-02 | 1,01E-01 |
| RAD51C | 277,11 | 0,07 | 0,03 | 2,27 | 2,31E-02 | 1,01E-01 |
| LIPT2 | 84,09 | 0,08 | 0,04 | 2,27 | 2,31E-02 | 1,01E-01 |
| RBM45 | 676,20 | 0,07 | 0,03 | 2,27 | 2,31E-02 | 1,01E-01 |
| WASHC2C | 1733,77 | -0,06 | 0,03 | -2,27 | 2,32E-02 | 1,01E-01 |
| SLC23A2 | 1807,25 | -0,06 | 0,03 | -2,27 | 2,32E-02 | 1,01E-01 |
| ROCK1 | 7280,40 | -0,07 | 0,03 | -2,27 | 2,32E-02 | 1,01E-01 |
| RP2 | 3410,36 | -0,08 | 0,04 | -2,27 | 2,33E-02 | 1,02E-01 |

| | | | | | | |
|---|---|---|---|---|---|---|
| B4GALT7 | 285,31 | 0,06 | 0,02 | 2,27 | 2,33E-02 | 1,02E-01 |
| UBE3A | 3467,05 | 0,06 | 0,03 | 2,27 | 2,33E-02 | 1,02E-01 |
| YTHDC2 | 2502,12 | 0,06 | 0,03 | 2,27 | 2,33E-02 | 1,02E-01 |
| ZNF557 | 1282,55 | 0,05 | 0,02 | 2,27 | 2,33E-02 | 1,02E-01 |
| REXO4 | 636,65 | 0,04 | 0,02 | 2,27 | 2,33E-02 | 1,02E-01 |
| EIF3K | 2444,03 | 0,06 | 0,02 | 2,27 | 2,33E-02 | 1,02E-01 |
| BTBD6 | 413,67 | 0,05 | 0,02 | 2,27 | 2,34E-02 | 1,02E-01 |
| NAIP | 1535,21 | -0,13 | 0,06 | -2,27 | 2,34E-02 | 1,02E-01 |
| RABGGTB | 1779,53 | 0,07 | 0,03 | 2,27 | 2,34E-02 | 1,02E-01 |
| FAM168A | 1219,45 | -0,06 | 0,03 | -2,27 | 2,34E-02 | 1,02E-01 |
| NASP | 1711,34 | -0,04 | 0,02 | -2,27 | 2,34E-02 | 1,02E-01 |
| OSM | 157,65 | -0,12 | 0,05 | -2,27 | 2,34E-02 | 1,02E-01 |
| MRPL53 | 160,57 | 0,05 | 0,02 | 2,27 | 2,34E-02 | 1,02E-01 |
| MRPS23 | 446,73 | 0,06 | 0,03 | 2,27 | 2,34E-02 | 1,02E-01 |
| MTPAP | 696,77 | 0,03 | 0,01 | 2,27 | 2,35E-02 | 1,02E-01 |
| METTL2A | 351,64 | 0,04 | 0,02 | 2,27 | 2,35E-02 | 1,02E-01 |
| UBAC1 | 881,49 | 0,05 | 0,02 | 2,26 | 2,36E-02 | 1,02E-01 |
| TACO1 | 272,98 | 0,04 | 0,02 | 2,26 | 2,37E-02 | 1,03E-01 |
| TSEN2 | 430,18 | 0,05 | 0,02 | 2,26 | 2,36E-02 | 1,03E-01 |
| BCKDHB | 401,64 | 0,06 | 0,03 | 2,26 | 2,37E-02 | 1,03E-01 |
| MED21 | 903,44 | 0,06 | 0,03 | 2,26 | 2,37E-02 | 1,03E-01 |
| BCAP29 | 624,50 | 0,06 | 0,03 | 2,26 | 2,37E-02 | 1,03E-01 |
| MRPS7 | 454,31 | 0,06 | 0,03 | 2,26 | 2,37E-02 | 1,03E-01 |
| ACP5 | 330,53 | 0,10 | 0,04 | 2,26 | 2,37E-02 | 1,03E-01 |
| GCFC2 | 624,11 | 0,07 | 0,03 | 2,26 | 2,37E-02 | 1,03E-01 |
| RINT1 | 560,33 | 0,04 | 0,02 | 2,26 | 2,38E-02 | 1,03E-01 |
| ADAMTS5 | 121,97 | 0,14 | 0,06 | 2,26 | 2,38E-02 | 1,03E-01 |
| NUTF2 | 656,23 | 0,05 | 0,02 | 2,26 | 2,38E-02 | 1,03E-01 |
| RALY | 2224,50 | -0,06 | 0,02 | -2,26 | 2,38E-02 | 1,03E-01 |
| SMURF1 | 1199,10 | -0,05 | 0,02 | -2,26 | 2,38E-02 | 1,03E-01 |
| P4HTM | 617,72 | 0,05 | 0,02 | 2,26 | 2,38E-02 | 1,03E-01 |
| FOXN3 | 4635,12 | -0,06 | 0,03 | -2,26 | 2,39E-02 | 1,03E-01 |
| CYP26C1 | 83,98 | -0,13 | 0,06 | -2,26 | 2,39E-02 | 1,03E-01 |
| IQCB1 | 1061,53 | 0,07 | 0,03 | 2,26 | 2,39E-02 | 1,03E-01 |
| HCAR3 | 784,97 | -0,12 | 0,05 | -2,26 | 2,39E-02 | 1,03E-01 |
| PLB1 | 864,63 | -0,16 | 0,07 | -2,26 | 2,39E-02 | 1,03E-01 |
| FASTKD2 | 875,76 | 0,07 | 0,03 | 2,26 | 2,40E-02 | 1,03E-01 |
| KBTBD7 | 1397,83 | -0,09 | 0,04 | -2,26 | 2,40E-02 | 1,03E-01 |
| HSPA1B | 326,13 | -0,09 | 0,04 | -2,26 | 2,40E-02 | 1,03E-01 |
| RPS9 | 13284,82 | 0,09 | 0,04 | 2,26 | 2,41E-02 | 1,03E-01 |
| XPO4 | 2021,27 | 0,05 | 0,02 | 2,26 | 2,41E-02 | 1,03E-01 |
| CD2BP2 | 1531,32 | -0,04 | 0,02 | -2,26 | 2,41E-02 | 1,03E-01 |
| SEPHS1 | 634,82 | 0,03 | 0,02 | 2,26 | 2,41E-02 | 1,04E-01 |
| BTBD7 | 1511,03 | -0,03 | 0,02 | -2,25 | 2,41E-02 | 1,04E-01 |
| TMEM199 | 639,54 | -0,03 | 0,01 | -2,25 | 2,42E-02 | 1,04E-01 |
| TMEM14A | 132,95 | 0,09 | 0,04 | 2,25 | 2,42E-02 | 1,04E-01 |

| | | | | | | |
|---|---|---|---|---|---|---|
| ZHX2 | 583,58 | -0,08 | 0,04 | -2,25 | 2,43E-02 | 1,04E-01 |
| PBLD | 101,11 | -0,07 | 0,03 | -2,25 | 2,43E-02 | 1,04E-01 |
| CD2AP | 1510,65 | 0,08 | 0,04 | 2,25 | 2,43E-02 | 1,04E-01 |
| KLF8 | 186,30 | 0,11 | 0,05 | 2,25 | 2,43E-02 | 1,04E-01 |
| ZNF407 | 3408,33 | -0,05 | 0,02 | -2,25 | 2,43E-02 | 1,04E-01 |
| P2RX1 | 770,65 | -0,12 | 0,05 | -2,25 | 2,43E-02 | 1,04E-01 |
| RTN4IP1 | 227,32 | 0,05 | 0,02 | 2,25 | 2,44E-02 | 1,04E-01 |
| TAF9B | 166,60 | 0,10 | 0,05 | 2,25 | 2,43E-02 | 1,04E-01 |
| ZBTB41 | 1098,97 | 0,13 | 0,06 | 2,25 | 2,44E-02 | 1,04E-01 |
| UBE2G1 | 1287,87 | -0,03 | 0,01 | -2,25 | 2,44E-02 | 1,04E-01 |
| ABCD2 | 1332,01 | 0,11 | 0,05 | 2,25 | 2,44E-02 | 1,04E-01 |
| TRPM7 | 3835,62 | 0,06 | 0,03 | 2,25 | 2,45E-02 | 1,04E-01 |
| MBD4 | 1924,11 | -0,06 | 0,03 | -2,25 | 2,45E-02 | 1,05E-01 |
| SAV1 | 403,79 | 0,06 | 0,03 | 2,25 | 2,46E-02 | 1,05E-01 |
| E2F5 | 257,70 | 0,11 | 0,05 | 2,25 | 2,46E-02 | 1,05E-01 |
| MRRF | 552,63 | 0,04 | 0,02 | 2,25 | 2,46E-02 | 1,05E-01 |
| SFXN4 | 199,62 | 0,06 | 0,03 | 2,25 | 2,46E-02 | 1,05E-01 |
| NRAS | 1606,16 | 0,06 | 0,03 | 2,25 | 2,46E-02 | 1,05E-01 |
| KDM3B | 3549,36 | -0,08 | 0,04 | -2,25 | 2,46E-02 | 1,05E-01 |
| NCBP2AS2 | 257,58 | 0,06 | 0,03 | 2,25 | 2,46E-02 | 1,05E-01 |
| CEBPB | 4350,76 | -0,08 | 0,04 | -2,25 | 2,47E-02 | 1,05E-01 |
| ZNF594 | 602,07 | 0,11 | 0,05 | 2,25 | 2,47E-02 | 1,05E-01 |
| IRF9 | 511,33 | -0,10 | 0,04 | -2,24 | 2,48E-02 | 1,05E-01 |
| MSRA | 431,38 | -0,09 | 0,04 | -2,24 | 2,48E-02 | 1,05E-01 |
| SLC30A6 | 1301,44 | 0,05 | 0,02 | 2,24 | 2,48E-02 | 1,05E-01 |
| STRADA | 845,49 | -0,05 | 0,02 | -2,24 | 2,49E-02 | 1,06E-01 |
| METAP1D | 143,17 | 0,08 | 0,04 | 2,24 | 2,49E-02 | 1,06E-01 |
| DGCR2 | 2872,46 | -0,09 | 0,04 | -2,24 | 2,49E-02 | 1,06E-01 |
| TBC1D20 | 1322,98 | -0,04 | 0,02 | -2,24 | 2,50E-02 | 1,06E-01 |
| ZNF510 | 890,26 | 0,09 | 0,04 | 2,24 | 2,50E-02 | 1,06E-01 |
| CLEC7A | 11857,38 | -0,08 | 0,04 | -2,24 | 2,50E-02 | 1,06E-01 |
| UQCC5 | 148,08 | 0,05 | 0,02 | 2,24 | 2,50E-02 | 1,06E-01 |
| FEM1A | 1998,53 | 0,06 | 0,03 | 2,24 | 2,51E-02 | 1,06E-01 |
| ZNF85 | 236,96 | 0,07 | 0,03 | 2,24 | 2,51E-02 | 1,06E-01 |
| ACIN1 | 4746,96 | -0,05 | 0,02 | -2,24 | 2,51E-02 | 1,06E-01 |
| RIPOR3 | 103,72 | 0,17 | 0,07 | 2,24 | 2,51E-02 | 1,06E-01 |
| ARHGAP32 | 350,07 | 0,07 | 0,03 | 2,24 | 2,52E-02 | 1,07E-01 |
| SPACA6 | 248,48 | -0,11 | 0,05 | -2,24 | 2,52E-02 | 1,07E-01 |
| FAT4 | 132,29 | -0,16 | 0,07 | -2,24 | 2,52E-02 | 1,07E-01 |
| ICOS | 606,93 | 0,09 | 0,04 | 2,24 | 2,53E-02 | 1,07E-01 |
| NDUFAB1 | 506,94 | 0,06 | 0,03 | 2,24 | 2,53E-02 | 1,07E-01 |
| SAP130 | 639,84 | -0,06 | 0,03 | -2,24 | 2,53E-02 | 1,07E-01 |
| BICRAL | 1786,52 | -0,06 | 0,03 | -2,24 | 2,53E-02 | 1,07E-01 |
| DCAF17 | 891,84 | 0,05 | 0,02 | 2,24 | 2,53E-02 | 1,07E-01 |
| SPINT1 | 225,77 | -0,10 | 0,05 | -2,24 | 2,53E-02 | 1,07E-01 |
| ZWILCH | 212,18 | 0,08 | 0,03 | 2,24 | 2,54E-02 | 1,07E-01 |

| | | | | | | |
|---|---|---|---|---|---|---|
| RPS3 | 9830,51 | 0,09 | 0,04 | 2,23 | 2,55E-02 | 1,07E-01 |
| MAGEH1 | 213,67 | 0,08 | 0,04 | 2,23 | 2,55E-02 | 1,08E-01 |
| NFRKB | 930,67 | -0,06 | 0,03 | -2,23 | 2,56E-02 | 1,08E-01 |
| DDHD2 | 1549,55 | 0,05 | 0,02 | 2,23 | 2,56E-02 | 1,08E-01 |
| AGPAT5 | 765,45 | 0,08 | 0,04 | 2,23 | 2,56E-02 | 1,08E-01 |
| AP1S2 | 3287,22 | 0,12 | 0,05 | 2,23 | 2,56E-02 | 1,08E-01 |
| CFAP54 | 135,42 | -0,15 | 0,07 | -2,23 | 2,56E-02 | 1,08E-01 |
| CSAD | 799,13 | -0,11 | 0,05 | -2,23 | 2,57E-02 | 1,08E-01 |
| RNF41 | 2185,66 | -0,04 | 0,02 | -2,23 | 2,58E-02 | 1,08E-01 |
| POLR3E | 1127,42 | 0,05 | 0,02 | 2,23 | 2,59E-02 | 1,09E-01 |
| PTGER2 | 796,60 | 0,06 | 0,03 | 2,23 | 2,59E-02 | 1,09E-01 |
| TENM1 | 580,06 | -0,13 | 0,06 | -2,23 | 2,58E-02 | 1,09E-01 |
| TRMT5 | 225,55 | 0,07 | 0,03 | 2,23 | 2,59E-02 | 1,09E-01 |
| ITM2A | 916,74 | 0,13 | 0,06 | 2,23 | 2,59E-02 | 1,09E-01 |
| MTRFR | 559,56 | 0,04 | 0,02 | 2,23 | 2,59E-02 | 1,09E-01 |
| ETFDH | 657,74 | -0,05 | 0,02 | -2,23 | 2,59E-02 | 1,09E-01 |
| TRMT13 | 910,31 | 0,09 | 0,04 | 2,23 | 2,59E-02 | 1,09E-01 |
| SHMT2 | 693,00 | 0,06 | 0,03 | 2,23 | 2,60E-02 | 1,09E-01 |
| RER1 | 2838,45 | -0,04 | 0,02 | -2,23 | 2,60E-02 | 1,09E-01 |
| TMEM161B | 1806,38 | 0,07 | 0,03 | 2,23 | 2,60E-02 | 1,09E-01 |
| RNH1 | 1371,26 | 0,06 | 0,03 | 2,23 | 2,60E-02 | 1,09E-01 |
| TOMM22 | 1098,82 | 0,07 | 0,03 | 2,23 | 2,60E-02 | 1,09E-01 |
| FDX1 | 499,46 | 0,06 | 0,03 | 2,23 | 2,60E-02 | 1,09E-01 |
| DPH2 | 126,55 | 0,08 | 0,03 | 2,23 | 2,60E-02 | 1,09E-01 |
| DUSP12 | 383,41 | 0,05 | 0,02 | 2,23 | 2,60E-02 | 1,09E-01 |
| IRS2 | 1490,51 | -0,10 | 0,04 | -2,22 | 2,61E-02 | 1,09E-01 |
| CGRRF1 | 384,67 | 0,08 | 0,04 | 2,22 | 2,61E-02 | 1,09E-01 |
| MKRN1 | 28856,76 | 0,14 | 0,06 | 2,22 | 2,61E-02 | 1,09E-01 |
| PPP3CC | 1679,96 | 0,06 | 0,03 | 2,22 | 2,61E-02 | 1,09E-01 |
| PDGFRB | 102,50 | -0,18 | 0,08 | -2,22 | 2,61E-02 | 1,09E-01 |
| FXR2 | 588,07 | -0,05 | 0,02 | -2,22 | 2,62E-02 | 1,09E-01 |
| DIPK2A | 1185,76 | 0,06 | 0,03 | 2,22 | 2,62E-02 | 1,09E-01 |
| RBM44 | 188,67 | 0,09 | 0,04 | 2,22 | 2,63E-02 | 1,09E-01 |
| SIGLEC1 | 181,70 | -0,29 | 0,13 | -2,22 | 2,63E-02 | 1,09E-01 |
| RPS10 | 115,68 | 0,05 | 0,02 | 2,22 | 2,63E-02 | 1,09E-01 |
| TAB2 | 4225,81 | -0,04 | 0,02 | -2,22 | 2,63E-02 | 1,09E-01 |
| SLC25A46 | 2102,07 | 0,08 | 0,04 | 2,22 | 2,63E-02 | 1,09E-01 |
| STAT5A | 2865,38 | -0,06 | 0,03 | -2,22 | 2,63E-02 | 1,09E-01 |
| MDM2 | 6776,43 | -0,06 | 0,03 | -2,22 | 2,63E-02 | 1,09E-01 |
| TLR2 | 12749,26 | -0,09 | 0,04 | -2,22 | 2,62E-02 | 1,09E-01 |
| LGR4 | 81,01 | 0,13 | 0,06 | 2,22 | 2,63E-02 | 1,09E-01 |
| RBL2 | 12400,65 | -0,04 | 0,02 | -2,22 | 2,64E-02 | 1,09E-01 |
| PNKD | 269,46 | 0,06 | 0,03 | 2,22 | 2,64E-02 | 1,09E-01 |
| SNX19 | 3335,82 | -0,04 | 0,02 | -2,22 | 2,64E-02 | 1,09E-01 |
| ZNF92 | 397,76 | 0,14 | 0,06 | 2,22 | 2,64E-02 | 1,09E-01 |
| FAM217B | 1270,88 | -0,04 | 0,02 | -2,22 | 2,64E-02 | 1,09E-01 |

| | | | | | | |
|---|---|---|---|---|---|---|
| UBA5 | 876,19 | 0,08 | 0,04 | 2,22 | 2,64E-02 | 1,09E-01 |
| ST7L | 470,84 | 0,04 | 0,02 | 2,22 | 2,65E-02 | 1,10E-01 |
| PRDM2 | 3585,43 | -0,05 | 0,02 | -2,22 | 2,65E-02 | 1,10E-01 |
| RERE | 2649,11 | -0,09 | 0,04 | -2,22 | 2,65E-02 | 1,10E-01 |
| SELENBP1 | 3602,25 | 0,18 | 0,08 | 2,22 | 2,66E-02 | 1,10E-01 |
| TPGS2 | 1702,01 | 0,06 | 0,03 | 2,22 | 2,66E-02 | 1,10E-01 |
| LRRC57 | 696,15 | -0,04 | 0,02 | -2,22 | 2,66E-02 | 1,10E-01 |
| NAP1L1 | 3882,88 | 0,07 | 0,03 | 2,22 | 2,66E-02 | 1,10E-01 |
| NDUFC1 | 344,61 | 0,05 | 0,02 | 2,22 | 2,66E-02 | 1,10E-01 |
| MRPS34 | 439,36 | 0,05 | 0,02 | 2,22 | 2,66E-02 | 1,10E-01 |
| DPM2 | 1280,98 | 0,13 | 0,06 | 2,22 | 2,67E-02 | 1,10E-01 |
| UCKL1 | 266,64 | 0,07 | 0,03 | 2,22 | 2,67E-02 | 1,10E-01 |
| VPS29 | 2116,21 | 0,06 | 0,03 | 2,22 | 2,67E-02 | 1,10E-01 |
| ITGB2 | 20112,32 | -0,08 | 0,04 | -2,22 | 2,67E-02 | 1,10E-01 |
| CSGALNACT2 | 2427,80 | -0,05 | 0,02 | -2,22 | 2,68E-02 | 1,10E-01 |
| SMIM30 | 150,28 | 0,14 | 0,06 | 2,22 | 2,67E-02 | 1,10E-01 |
| SKA3 | 103,74 | 0,18 | 0,08 | 2,22 | 2,67E-02 | 1,10E-01 |
| DISC1 | 1487,81 | -0,08 | 0,04 | -2,22 | 2,67E-02 | 1,10E-01 |
| BAG6 | 2132,86 | 0,12 | 0,06 | 2,22 | 2,67E-02 | 1,10E-01 |
| OGFR | 3215,57 | -0,06 | 0,03 | -2,21 | 2,68E-02 | 1,10E-01 |
| RNF111 | 2452,39 | -0,05 | 0,02 | -2,21 | 2,68E-02 | 1,10E-01 |
| SEC11A | 1130,65 | -0,04 | 0,02 | -2,21 | 2,68E-02 | 1,10E-01 |
| SELENOI | 1096,97 | 0,05 | 0,02 | 2,21 | 2,69E-02 | 1,10E-01 |
| PPP1CB | 8935,48 | 0,10 | 0,04 | 2,21 | 2,69E-02 | 1,10E-01 |
| CYP2R1 | 637,61 | 0,06 | 0,03 | 2,21 | 2,69E-02 | 1,10E-01 |
| GGT7 | 223,13 | 0,08 | 0,04 | 2,21 | 2,70E-02 | 1,10E-01 |
| UGGT2 | 312,47 | 0,10 | 0,04 | 2,21 | 2,69E-02 | 1,10E-01 |
| RGS10 | 1717,10 | 0,11 | 0,05 | 2,21 | 2,70E-02 | 1,10E-01 |
| VTI1B | 2593,47 | 0,07 | 0,03 | 2,21 | 2,70E-02 | 1,10E-01 |
| SPTLC1 | 2037,21 | -0,04 | 0,02 | -2,21 | 2,70E-02 | 1,10E-01 |
| EXTL2 | 151,95 | 0,07 | 0,03 | 2,21 | 2,70E-02 | 1,10E-01 |
| KANK2 | 173,51 | 0,19 | 0,09 | 2,21 | 2,69E-02 | 1,10E-01 |
| RUNX2 | 1397,53 | -0,07 | 0,03 | -2,21 | 2,70E-02 | 1,11E-01 |
| RAP2B | 4225,63 | 0,05 | 0,02 | 2,21 | 2,70E-02 | 1,11E-01 |
| TRAPPC13 | 711,65 | 0,08 | 0,03 | 2,21 | 2,70E-02 | 1,11E-01 |
| NUP58 | 3926,68 | -0,06 | 0,03 | -2,21 | 2,70E-02 | 1,11E-01 |
| ZFAT | 972,33 | -0,05 | 0,02 | -2,21 | 2,71E-02 | 1,11E-01 |
| CTNNB1 | 6992,33 | -0,04 | 0,02 | -2,21 | 2,71E-02 | 1,11E-01 |
| TNF | 85,29 | 0,08 | 0,04 | 2,21 | 2,71E-02 | 1,11E-01 |
| BNIPL | 75,49 | -0,09 | 0,04 | -2,21 | 2,71E-02 | 1,11E-01 |
| REPIN1 | 830,89 | 0,06 | 0,03 | 2,21 | 2,71E-02 | 1,11E-01 |
| TBCK | 2199,30 | 0,07 | 0,03 | 2,21 | 2,72E-02 | 1,11E-01 |
| ANKAR | 649,54 | 0,06 | 0,03 | 2,21 | 2,72E-02 | 1,11E-01 |
| CHML | 1047,31 | 0,12 | 0,05 | 2,21 | 2,73E-02 | 1,11E-01 |
| EIF4H | 5185,44 | -0,04 | 0,02 | -2,21 | 2,73E-02 | 1,11E-01 |
| YJU2B | 418,04 | 0,07 | 0,03 | 2,21 | 2,73E-02 | 1,11E-01 |

| | | | | | | |
|---|---|---|---|---|---|---|
| DNTTIP1 | 782,65 | -0,07 | 0,03 | -2,21 | 2,73E-02 | 1,11E-01 |
| GSPT1 | 10211,72 | 0,13 | 0,06 | 2,21 | 2,74E-02 | 1,11E-01 |
| CD300LB | 837,22 | -0,10 | 0,04 | -2,21 | 2,73E-02 | 1,11E-01 |
| LMAN2 | 2628,59 | -0,04 | 0,02 | -2,21 | 2,74E-02 | 1,11E-01 |
| PAFAH1B1 | 2632,77 | -0,05 | 0,02 | -2,21 | 2,74E-02 | 1,11E-01 |
| WARS2 | 273,28 | 0,05 | 0,02 | 2,21 | 2,74E-02 | 1,11E-01 |
| PAN3 | 5436,86 | -0,05 | 0,02 | -2,20 | 2,75E-02 | 1,12E-01 |
| SLC30A1 | 1292,53 | 0,11 | 0,05 | 2,20 | 2,75E-02 | 1,12E-01 |
| CREBL2 | 1951,57 | 0,06 | 0,03 | 2,20 | 2,75E-02 | 1,12E-01 |
| SSNA1 | 491,28 | 0,05 | 0,02 | 2,20 | 2,75E-02 | 1,12E-01 |
| RPL21 | 8250,24 | 0,18 | 0,08 | 2,20 | 2,76E-02 | 1,12E-01 |
| HAGHL | 104,78 | 0,11 | 0,05 | 2,20 | 2,76E-02 | 1,12E-01 |
| MLLT11 | 98,11 | 0,06 | 0,03 | 2,20 | 2,76E-02 | 1,12E-01 |
| NUDT18 | 265,77 | 0,06 | 0,03 | 2,20 | 2,77E-02 | 1,12E-01 |
| MTBP | 392,48 | 0,07 | 0,03 | 2,20 | 2,77E-02 | 1,12E-01 |
| ZNF813 | 315,37 | 0,10 | 0,05 | 2,20 | 2,77E-02 | 1,12E-01 |
| RCOR3 | 2512,15 | 0,04 | 0,02 | 2,20 | 2,78E-02 | 1,12E-01 |
| GORAB | 379,53 | 0,07 | 0,03 | 2,20 | 2,78E-02 | 1,12E-01 |
| PATZ1 | 588,51 | 0,06 | 0,03 | 2,20 | 2,78E-02 | 1,12E-01 |
| TRIM13 | 1159,12 | 0,06 | 0,03 | 2,20 | 2,78E-02 | 1,12E-01 |
| NAXE | 253,06 | 0,05 | 0,02 | 2,20 | 2,78E-02 | 1,13E-01 |
| GPKOW | 442,50 | -0,04 | 0,02 | -2,20 | 2,79E-02 | 1,13E-01 |
| CRYBG1 | 5421,68 | -0,04 | 0,02 | -2,20 | 2,79E-02 | 1,13E-01 |
| STYXL1 | 434,11 | -0,05 | 0,02 | -2,20 | 2,80E-02 | 1,13E-01 |
| SYNE2 | 23620,89 | -0,06 | 0,03 | -2,20 | 2,80E-02 | 1,13E-01 |
| DMAC2L | 1212,07 | 0,05 | 0,02 | 2,20 | 2,80E-02 | 1,13E-01 |
| FASTKD1 | 707,94 | 0,07 | 0,03 | 2,20 | 2,81E-02 | 1,13E-01 |
| MRPL52 | 372,92 | 0,04 | 0,02 | 2,20 | 2,81E-02 | 1,13E-01 |
| SIN3A | 2078,58 | -0,05 | 0,02 | -2,19 | 2,82E-02 | 1,14E-01 |
| HNMT | 482,69 | 0,10 | 0,04 | 2,19 | 2,82E-02 | 1,14E-01 |
| RPS24 | 3342,81 | 0,13 | 0,06 | 2,19 | 2,83E-02 | 1,14E-01 |
| TADA1 | 322,60 | 0,05 | 0,02 | 2,19 | 2,83E-02 | 1,14E-01 |
| SDE2 | 1965,49 | -0,05 | 0,02 | -2,19 | 2,83E-02 | 1,14E-01 |
| APOL6 | 10445,68 | -0,10 | 0,04 | -2,19 | 2,84E-02 | 1,14E-01 |
| RUFY2 | 721,36 | 0,04 | 0,02 | 2,19 | 2,84E-02 | 1,14E-01 |
| RABGAP1 | 1732,74 | -0,04 | 0,02 | -2,19 | 2,84E-02 | 1,14E-01 |
| DOHH | 188,13 | 0,07 | 0,03 | 2,19 | 2,85E-02 | 1,14E-01 |
| EIF2B2 | 542,85 | 0,04 | 0,02 | 2,19 | 2,86E-02 | 1,15E-01 |
| GID4 | 551,77 | 0,09 | 0,04 | 2,19 | 2,85E-02 | 1,15E-01 |
| MRPS17 | 100,82 | 0,06 | 0,03 | 2,19 | 2,86E-02 | 1,15E-01 |
| FASTKD5 | 736,80 | -0,04 | 0,02 | -2,19 | 2,86E-02 | 1,15E-01 |
| TMOD1 | 954,83 | 0,16 | 0,07 | 2,19 | 2,86E-02 | 1,15E-01 |
| RRP7A | 1298,64 | 0,07 | 0,03 | 2,19 | 2,86E-02 | 1,15E-01 |
| DCUN1D5 | 280,11 | 0,08 | 0,04 | 2,19 | 2,87E-02 | 1,15E-01 |
| ATP7A | 2567,19 | -0,05 | 0,02 | -2,19 | 2,87E-02 | 1,15E-01 |
| TKT | 9611,88 | -0,08 | 0,04 | -2,19 | 2,87E-02 | 1,15E-01 |

| | | | | | | |
|---|---|---|---|---|---|---|
| PEA15 | 530,39 | 0,06 | 0,03 | 2,19 | 2,88E-02 | 1,15E-01 |
| ITSN2 | 9397,92 | -0,05 | 0,02 | -2,19 | 2,88E-02 | 1,15E-01 |
| KIFBP | 376,06 | 0,04 | 0,02 | 2,19 | 2,88E-02 | 1,15E-01 |
| HEXB | 1480,90 | -0,05 | 0,02 | -2,19 | 2,88E-02 | 1,15E-01 |
| INTS6 | 6207,37 | -0,05 | 0,02 | -2,19 | 2,89E-02 | 1,15E-01 |
| MTDH | 2931,72 | 0,05 | 0,02 | 2,19 | 2,89E-02 | 1,15E-01 |
| SIK3 | 2838,78 | -0,08 | 0,04 | -2,19 | 2,89E-02 | 1,15E-01 |
| BCL10 | 1278,75 | -0,04 | 0,02 | -2,18 | 2,89E-02 | 1,16E-01 |
| IFI44 | 1511,63 | -0,23 | 0,10 | -2,18 | 2,89E-02 | 1,16E-01 |
| SASH3 | 3095,12 | -0,06 | 0,03 | -2,18 | 2,91E-02 | 1,16E-01 |
| RPS27A | 6341,86 | 0,14 | 0,06 | 2,18 | 2,91E-02 | 1,16E-01 |
| H1-0 | 781,65 | 0,17 | 0,08 | 2,18 | 2,91E-02 | 1,16E-01 |
| RPUSD4 | 516,88 | 0,03 | 0,02 | 2,18 | 2,91E-02 | 1,16E-01 |
| GBA1 | 268,75 | -0,09 | 0,04 | -2,18 | 2,91E-02 | 1,16E-01 |
| PSTPIP2 | 1513,34 | -0,08 | 0,04 | -2,18 | 2,91E-02 | 1,16E-01 |
| EXOC6 | 1658,42 | -0,07 | 0,03 | -2,18 | 2,91E-02 | 1,16E-01 |
| NFKBIZ | 3915,76 | -0,05 | 0,03 | -2,18 | 2,91E-02 | 1,16E-01 |
| MSRB2 | 358,35 | -0,08 | 0,04 | -2,18 | 2,91E-02 | 1,16E-01 |
| ZNF480 | 445,61 | 0,10 | 0,05 | 2,18 | 2,91E-02 | 1,16E-01 |
| ZNF566 | 347,37 | 0,06 | 0,03 | 2,18 | 2,92E-02 | 1,16E-01 |
| PISD | 2428,92 | -0,10 | 0,05 | -2,18 | 2,92E-02 | 1,16E-01 |
| PANX1 | 538,36 | 0,05 | 0,02 | 2,18 | 2,92E-02 | 1,16E-01 |
| ZNF230 | 813,37 | -0,08 | 0,04 | -2,18 | 2,93E-02 | 1,16E-01 |
| DAAM2 | 241,27 | -0,24 | 0,11 | -2,18 | 2,93E-02 | 1,16E-01 |
| ARL14EP | 1338,39 | 0,10 | 0,04 | 2,18 | 2,93E-02 | 1,16E-01 |
| MYBBP1A | 334,82 | 0,09 | 0,04 | 2,18 | 2,93E-02 | 1,16E-01 |
| IQCG | 321,86 | 0,05 | 0,02 | 2,18 | 2,94E-02 | 1,16E-01 |
| SLC25A43 | 167,31 | 0,05 | 0,02 | 2,18 | 2,94E-02 | 1,16E-01 |
| ENSG000001 | 2356,26 | -0,17 | 0,08 | -2,18 | 2,94E-02 | 1,16E-01 |
| PHF11 | 1711,19 | -0,06 | 0,03 | -2,18 | 2,95E-02 | 1,17E-01 |
| ARHGEF40 | 1841,58 | -0,11 | 0,05 | -2,18 | 2,95E-02 | 1,17E-01 |
| ZNF268 | 679,81 | 0,10 | 0,05 | 2,18 | 2,94E-02 | 1,17E-01 |
| SPOUT1 | 592,16 | 0,04 | 0,02 | 2,18 | 2,95E-02 | 1,17E-01 |
| ZC3H12D | 834,12 | 0,08 | 0,04 | 2,18 | 2,95E-02 | 1,17E-01 |
| MIER3 | 803,51 | 0,09 | 0,04 | 2,18 | 2,95E-02 | 1,17E-01 |
| DGKG | 707,08 | -0,09 | 0,04 | -2,18 | 2,95E-02 | 1,17E-01 |
| PHKA2 | 1335,51 | -0,07 | 0,03 | -2,18 | 2,95E-02 | 1,17E-01 |
| CTBS | 3979,31 | -0,08 | 0,04 | -2,18 | 2,95E-02 | 1,17E-01 |
| NOP53 | 6002,13 | 0,08 | 0,04 | 2,18 | 2,96E-02 | 1,17E-01 |
| RAB5C | 693,32 | -0,07 | 0,03 | -2,18 | 2,96E-02 | 1,17E-01 |
| MTMR1 | 1998,16 | -0,04 | 0,02 | -2,18 | 2,96E-02 | 1,17E-01 |
| RHOBTB3 | 484,15 | 0,10 | 0,04 | 2,17 | 2,96E-02 | 1,17E-01 |
| SLC48A1 | 349,55 | 0,13 | 0,06 | 2,17 | 2,97E-02 | 1,17E-01 |
| ZNF350 | 1051,98 | -0,06 | 0,03 | -2,17 | 2,97E-02 | 1,17E-01 |
| EPB42 | 3094,45 | 0,17 | 0,08 | 2,17 | 2,98E-02 | 1,17E-01 |
| PLPP5 | 507,70 | 0,09 | 0,04 | 2,17 | 2,98E-02 | 1,17E-01 |

| | | | | | | |
|---|---|---|---|---|---|---|
| TRIM24 | 1298,71 | -0,04 | 0,02 | -2,17 | 2,98E-02 | 1,17E-01 |
| CMPK2 | 898,28 | -0,20 | 0,09 | -2,17 | 2,98E-02 | 1,17E-01 |
| IL17RA | 6310,61 | -0,11 | 0,05 | -2,17 | 2,99E-02 | 1,18E-01 |
| PLEKHB2 | 4020,27 | -0,04 | 0,02 | -2,17 | 3,00E-02 | 1,18E-01 |
| C16orf54 | 2995,49 | -0,08 | 0,04 | -2,17 | 3,00E-02 | 1,18E-01 |
| RAD1 | 651,64 | 0,04 | 0,02 | 2,17 | 3,00E-02 | 1,18E-01 |
| SLC25A24 | 1522,75 | 0,06 | 0,03 | 2,17 | 3,00E-02 | 1,18E-01 |
| TTC19 | 1134,49 | 0,04 | 0,02 | 2,17 | 3,00E-02 | 1,18E-01 |
| TRAPPC10 | 3167,43 | -0,05 | 0,02 | -2,17 | 3,01E-02 | 1,18E-01 |
| UHMK1 | 8881,73 | 0,06 | 0,03 | 2,17 | 3,01E-02 | 1,18E-01 |
| IFI27L1 | 74,53 | 0,08 | 0,03 | 2,17 | 3,01E-02 | 1,18E-01 |
| AKIRIN1 | 2739,87 | -0,04 | 0,02 | -2,17 | 3,02E-02 | 1,19E-01 |
| SMIM1 | 226,69 | 0,28 | 0,13 | 2,17 | 3,03E-02 | 1,19E-01 |
| H2AC20 | 6102,20 | 0,09 | 0,04 | 2,17 | 3,03E-02 | 1,19E-01 |
| CLIC4 | 998,23 | 0,08 | 0,04 | 2,17 | 3,03E-02 | 1,19E-01 |
| SLU7 | 1963,44 | -0,08 | 0,04 | -2,17 | 3,03E-02 | 1,19E-01 |
| CARM1 | 458,27 | 0,15 | 0,07 | 2,17 | 3,03E-02 | 1,19E-01 |
| ELP6 | 398,24 | 0,04 | 0,02 | 2,17 | 3,04E-02 | 1,19E-01 |
| RARS1 | 1292,29 | 0,07 | 0,03 | 2,17 | 3,03E-02 | 1,19E-01 |
| PIK3R5 | 3962,78 | -0,09 | 0,04 | -2,16 | 3,05E-02 | 1,19E-01 |
| WDR43 | 1370,38 | 0,06 | 0,03 | 2,16 | 3,05E-02 | 1,19E-01 |
| ZC3H6 | 1695,02 | 0,05 | 0,02 | 2,16 | 3,05E-02 | 1,19E-01 |
| TM2D3 | 1631,86 | -0,03 | 0,02 | -2,16 | 3,06E-02 | 1,19E-01 |
| SLC25A15 | 171,11 | 0,05 | 0,02 | 2,16 | 3,06E-02 | 1,20E-01 |
| BTF3L4 | 1133,24 | 0,06 | 0,03 | 2,16 | 3,07E-02 | 1,20E-01 |
| ERAL1 | 180,34 | 0,04 | 0,02 | 2,16 | 3,07E-02 | 1,20E-01 |
| ETNK1 | 4292,55 | 0,09 | 0,04 | 2,16 | 3,08E-02 | 1,20E-01 |
| C6orf47 | 1075,89 | -0,04 | 0,02 | -2,16 | 3,08E-02 | 1,20E-01 |
| OPTN | 3391,31 | 0,10 | 0,05 | 2,16 | 3,09E-02 | 1,20E-01 |
| TGIF1 | 753,02 | 0,05 | 0,02 | 2,16 | 3,09E-02 | 1,20E-01 |
| EEF1AKMT2 | 360,01 | 0,07 | 0,03 | 2,16 | 3,09E-02 | 1,20E-01 |
| CRTAP | 1786,78 | 0,04 | 0,02 | 2,16 | 3,08E-02 | 1,20E-01 |
| PIK3CD | 5016,44 | -0,10 | 0,05 | -2,16 | 3,08E-02 | 1,20E-01 |
| AP2A1 | 1676,04 | 0,13 | 0,06 | 2,16 | 3,09E-02 | 1,20E-01 |
| CTR9 | 1693,35 | -0,04 | 0,02 | -2,16 | 3,08E-02 | 1,20E-01 |
| DPH5 | 440,76 | 0,07 | 0,03 | 2,16 | 3,09E-02 | 1,20E-01 |
| COX7A2L | 1507,81 | 0,06 | 0,03 | 2,16 | 3,09E-02 | 1,20E-01 |
| PCNP | 2028,17 | 0,08 | 0,04 | 2,16 | 3,09E-02 | 1,20E-01 |
| FKBP1A | 1768,96 | -0,09 | 0,04 | -2,16 | 3,09E-02 | 1,20E-01 |
| LENG9 | 104,63 | 0,07 | 0,03 | 2,16 | 3,09E-02 | 1,20E-01 |
| ABAT | 638,95 | -0,09 | 0,04 | -2,16 | 3,10E-02 | 1,20E-01 |
| DRAM2 | 1788,27 | 0,06 | 0,03 | 2,16 | 3,11E-02 | 1,21E-01 |
| PDSS1 | 153,22 | 0,05 | 0,02 | 2,16 | 3,12E-02 | 1,21E-01 |
| ELOF1 | 565,21 | 0,11 | 0,05 | 2,16 | 3,12E-02 | 1,21E-01 |
| DCLRE1A | 283,53 | 0,08 | 0,04 | 2,15 | 3,12E-02 | 1,21E-01 |
| PPP3CB | 2071,97 | 0,05 | 0,02 | 2,15 | 3,12E-02 | 1,21E-01 |

| | | | | | | |
|---|---|---|---|---|---|---|
| ACVR1B | 678,13 | -0,07 | 0,03 | -2,15 | 3,12E-02 | 1,21E-01 |
| EIF3B | 1840,18 | 0,06 | 0,03 | 2,15 | 3,12E-02 | 1,21E-01 |
| MAPKAPK2 | 1380,31 | -0,08 | 0,04 | -2,15 | 3,13E-02 | 1,21E-01 |
| MEF2D | 1400,10 | -0,09 | 0,04 | -2,15 | 3,13E-02 | 1,21E-01 |
| FAM210B | 8288,31 | 0,16 | 0,07 | 2,15 | 3,14E-02 | 1,21E-01 |
| IFT80 | 456,76 | 0,07 | 0,03 | 2,15 | 3,14E-02 | 1,21E-01 |
| UBXN8 | 97,65 | 0,09 | 0,04 | 2,15 | 3,14E-02 | 1,21E-01 |
| IRAK2 | 224,55 | -0,06 | 0,03 | -2,15 | 3,14E-02 | 1,21E-01 |
| LYRM7 | 918,22 | 0,06 | 0,03 | 2,15 | 3,14E-02 | 1,21E-01 |
| CD27 | 305,02 | 0,09 | 0,04 | 2,15 | 3,14E-02 | 1,21E-01 |
| POR | 647,89 | -0,08 | 0,04 | -2,15 | 3,15E-02 | 1,22E-01 |
| OXSR1 | 2605,89 | -0,04 | 0,02 | -2,15 | 3,15E-02 | 1,22E-01 |
| HARS1 | 1031,23 | -0,03 | 0,01 | -2,15 | 3,16E-02 | 1,22E-01 |
| CASC3 | 2462,60 | -0,09 | 0,04 | -2,15 | 3,16E-02 | 1,22E-01 |
| AGO1 | 2648,83 | -0,07 | 0,03 | -2,15 | 3,16E-02 | 1,22E-01 |
| IFT74 | 278,41 | 0,07 | 0,03 | 2,15 | 3,16E-02 | 1,22E-01 |
| NCBP1 | 1809,89 | 0,04 | 0,02 | 2,15 | 3,17E-02 | 1,22E-01 |
| STK35 | 846,71 | -0,07 | 0,03 | -2,15 | 3,17E-02 | 1,22E-01 |
| CRADD | 444,71 | -0,05 | 0,02 | -2,15 | 3,17E-02 | 1,22E-01 |
| SLBP | 1151,75 | -0,05 | 0,02 | -2,15 | 3,17E-02 | 1,22E-01 |
| ZCCHC4 | 476,72 | 0,05 | 0,02 | 2,15 | 3,17E-02 | 1,22E-01 |
| NAA20 | 484,99 | 0,05 | 0,02 | 2,15 | 3,18E-02 | 1,22E-01 |
| RNF123 | 1679,28 | 0,09 | 0,04 | 2,15 | 3,19E-02 | 1,23E-01 |
| COX7A2 | 1005,06 | 0,11 | 0,05 | 2,15 | 3,19E-02 | 1,23E-01 |
| SEMA4D | 6674,58 | -0,11 | 0,05 | -2,14 | 3,20E-02 | 1,23E-01 |
| TRAK2 | 5109,98 | 0,09 | 0,04 | 2,14 | 3,20E-02 | 1,23E-01 |
| COX7B | 437,87 | 0,10 | 0,05 | 2,14 | 3,20E-02 | 1,23E-01 |
| PHPT1 | 259,97 | 0,06 | 0,03 | 2,14 | 3,20E-02 | 1,23E-01 |
| RPE | 2451,94 | 0,06 | 0,03 | 2,14 | 3,20E-02 | 1,23E-01 |
| IFITM10 | 78,50 | -0,12 | 0,05 | -2,14 | 3,20E-02 | 1,23E-01 |
| ESS2 | 568,56 | -0,05 | 0,02 | -2,14 | 3,20E-02 | 1,23E-01 |
| PIP5K1A | 1260,06 | -0,03 | 0,02 | -2,14 | 3,21E-02 | 1,23E-01 |
| RAB3IL1 | 76,26 | 0,18 | 0,08 | 2,14 | 3,21E-02 | 1,23E-01 |
| SBNO2 | 2720,82 | -0,12 | 0,06 | -2,14 | 3,21E-02 | 1,23E-01 |
| MKLN1 | 8263,32 | -0,05 | 0,03 | -2,14 | 3,21E-02 | 1,23E-01 |
| RAD23B | 3882,60 | -0,05 | 0,02 | -2,14 | 3,21E-02 | 1,23E-01 |
| HELQ | 877,34 | 0,05 | 0,02 | 2,14 | 3,21E-02 | 1,23E-01 |
| ARL13B | 256,01 | 0,05 | 0,02 | 2,14 | 3,22E-02 | 1,23E-01 |
| HNRNPM | 2567,87 | -0,04 | 0,02 | -2,14 | 3,22E-02 | 1,23E-01 |
| HLA-DQA2 | 161,33 | 0,39 | 0,18 | 2,14 | 3,22E-02 | 1,23E-01 |
| BLOC1S4 | 621,21 | 0,05 | 0,02 | 2,14 | 3,22E-02 | 1,23E-01 |
| TSPAN5 | 3691,46 | 0,14 | 0,06 | 2,14 | 3,23E-02 | 1,23E-01 |
| TET3 | 2830,71 | -0,08 | 0,04 | -2,14 | 3,24E-02 | 1,23E-01 |
| KCNH8 | 118,17 | 0,15 | 0,07 | 2,14 | 3,24E-02 | 1,23E-01 |
| SNX20 | 1576,30 | -0,07 | 0,03 | -2,14 | 3,24E-02 | 1,23E-01 |
| HPS6 | 449,33 | 0,05 | 0,02 | 2,14 | 3,24E-02 | 1,23E-01 |

| | | | | | | |
|---|---|---|---|---|---|---|
| KPNA3 | 878,03 | 0,06 | 0,03 | 2,14 | 3,24E-02 | 1,23E-01 |
| FBF1 | 136,38 | 0,05 | 0,02 | 2,14 | 3,24E-02 | 1,23E-01 |
| CLOCK | 989,44 | 0,05 | 0,02 | 2,14 | 3,24E-02 | 1,23E-01 |
| CYTH1 | 10099,84 | -0,05 | 0,02 | -2,14 | 3,24E-02 | 1,23E-01 |
| NDUFAF2 | 108,17 | 0,07 | 0,03 | 2,14 | 3,24E-02 | 1,23E-01 |
| SF3B5 | 806,65 | 0,06 | 0,03 | 2,14 | 3,24E-02 | 1,23E-01 |
| PLPP1 | 125,11 | 0,07 | 0,03 | 2,14 | 3,25E-02 | 1,24E-01 |
| FCER2 | 112,17 | 0,16 | 0,07 | 2,14 | 3,25E-02 | 1,24E-01 |
| CEBPE | 132,56 | 0,11 | 0,05 | 2,14 | 3,26E-02 | 1,24E-01 |
| SLC17A5 | 577,29 | -0,03 | 0,01 | -2,14 | 3,26E-02 | 1,24E-01 |
| PDPK1 | 1717,27 | -0,08 | 0,04 | -2,14 | 3,26E-02 | 1,24E-01 |
| NRDE2 | 1863,50 | -0,04 | 0,02 | -2,14 | 3,26E-02 | 1,24E-01 |
| WDR5B | 597,19 | 0,08 | 0,04 | 2,14 | 3,27E-02 | 1,24E-01 |
| EIF1AD | 705,58 | -0,03 | 0,01 | -2,14 | 3,27E-02 | 1,24E-01 |
| ABCC2 | 211,93 | -0,08 | 0,04 | -2,14 | 3,27E-02 | 1,24E-01 |
| NTHL1 | 155,94 | 0,06 | 0,03 | 2,13 | 3,28E-02 | 1,25E-01 |
| TAF8 | 1575,91 | -0,04 | 0,02 | -2,13 | 3,30E-02 | 1,25E-01 |
| GIMAP5 | 2296,53 | 0,06 | 0,03 | 2,13 | 3,30E-02 | 1,25E-01 |
| SGPP1 | 897,36 | 0,08 | 0,04 | 2,13 | 3,31E-02 | 1,25E-01 |
| SLC10A7 | 525,81 | 0,05 | 0,02 | 2,13 | 3,31E-02 | 1,25E-01 |
| FAN1 | 1073,85 | 0,06 | 0,03 | 2,13 | 3,31E-02 | 1,25E-01 |
| LIN9 | 134,90 | 0,09 | 0,04 | 2,13 | 3,31E-02 | 1,25E-01 |
| SNRNP25 | 154,77 | 0,06 | 0,03 | 2,13 | 3,31E-02 | 1,25E-01 |
| STARD4 | 641,94 | 0,10 | 0,05 | 2,13 | 3,31E-02 | 1,25E-01 |
| NXT2 | 314,75 | 0,12 | 0,05 | 2,13 | 3,32E-02 | 1,25E-01 |
| IER5L | 187,89 | 0,08 | 0,04 | 2,13 | 3,32E-02 | 1,25E-01 |
| GLO1 | 673,60 | 0,08 | 0,04 | 2,13 | 3,32E-02 | 1,25E-01 |
| OSGEP | 1030,16 | 0,04 | 0,02 | 2,13 | 3,32E-02 | 1,26E-01 |
| EPSTI1 | 1841,28 | -0,17 | 0,08 | -2,13 | 3,33E-02 | 1,26E-01 |
| PIDD1 | 149,08 | 0,10 | 0,05 | 2,13 | 3,33E-02 | 1,26E-01 |
| PF4V1 | 101,25 | -0,21 | 0,10 | -2,13 | 3,34E-02 | 1,26E-01 |
| PDIK1L | 421,74 | 0,10 | 0,05 | 2,13 | 3,34E-02 | 1,26E-01 |
| VRK1 | 815,53 | 0,11 | 0,05 | 2,13 | 3,34E-02 | 1,26E-01 |
| CTNNA1 | 2526,59 | -0,07 | 0,03 | -2,13 | 3,34E-02 | 1,26E-01 |
| SEC22A | 526,67 | 0,04 | 0,02 | 2,13 | 3,35E-02 | 1,26E-01 |
| CASD1 | 1605,30 | 0,09 | 0,04 | 2,13 | 3,35E-02 | 1,26E-01 |
| FCGRT | 4218,08 | -0,08 | 0,04 | -2,13 | 3,35E-02 | 1,26E-01 |
| CENPQ | 111,52 | 0,13 | 0,06 | 2,13 | 3,35E-02 | 1,26E-01 |
| MBIP | 318,02 | 0,08 | 0,04 | 2,13 | 3,35E-02 | 1,26E-01 |
| SKP2 | 761,75 | 0,04 | 0,02 | 2,13 | 3,35E-02 | 1,26E-01 |
| ZNF26 | 1476,71 | 0,04 | 0,02 | 2,13 | 3,35E-02 | 1,26E-01 |
| AKAP11 | 3106,26 | 0,06 | 0,03 | 2,13 | 3,35E-02 | 1,26E-01 |
| MPZL2 | 386,89 | -0,09 | 0,04 | -2,13 | 3,36E-02 | 1,26E-01 |
| LASP1 | 3686,21 | -0,09 | 0,04 | -2,12 | 3,36E-02 | 1,26E-01 |
| SPNS3 | 192,35 | 0,11 | 0,05 | 2,12 | 3,36E-02 | 1,26E-01 |
| PARP12 | 2495,48 | -0,09 | 0,04 | -2,12 | 3,37E-02 | 1,27E-01 |

| | | | | | | |
|---|---|---|---|---|---|---|
| RBL1 | 981,76 | 0,06 | 0,03 | 2,12 | 3,38E-02 | 1,27E-01 |
| GNPNAT1 | 302,01 | 0,06 | 0,03 | 2,12 | 3,38E-02 | 1,27E-01 |
| ASPH | 2933,89 | -0,08 | 0,04 | -2,12 | 3,39E-02 | 1,27E-01 |
| TTC1 | 1054,80 | -0,05 | 0,02 | -2,12 | 3,39E-02 | 1,27E-01 |
| TMEM263 | 663,58 | 0,08 | 0,04 | 2,12 | 3,39E-02 | 1,27E-01 |
| ZSCAN30 | 677,71 | 0,05 | 0,02 | 2,12 | 3,39E-02 | 1,27E-01 |
| ZNF182 | 366,57 | -0,04 | 0,02 | -2,12 | 3,40E-02 | 1,27E-01 |
| PDP1 | 1649,97 | 0,06 | 0,03 | 2,12 | 3,40E-02 | 1,27E-01 |
| ZNF428 | 412,34 | 0,05 | 0,03 | 2,12 | 3,43E-02 | 1,28E-01 |
| DTWD1 | 822,71 | 0,06 | 0,03 | 2,12 | 3,42E-02 | 1,28E-01 |
| PLEKHA2 | 3765,55 | -0,04 | 0,02 | -2,12 | 3,42E-02 | 1,28E-01 |
| CEBPZOS | 713,39 | 0,04 | 0,02 | 2,12 | 3,41E-02 | 1,28E-01 |
| TMX3 | 2645,09 | 0,10 | 0,05 | 2,12 | 3,42E-02 | 1,28E-01 |
| ZSWIM7 | 353,83 | 0,04 | 0,02 | 2,12 | 3,42E-02 | 1,28E-01 |
| NSMAF | 1980,82 | -0,04 | 0,02 | -2,12 | 3,43E-02 | 1,28E-01 |
| AP1S1 | 179,50 | 0,07 | 0,03 | 2,12 | 3,43E-02 | 1,28E-01 |
| PHF3 | 6964,51 | -0,06 | 0,03 | -2,12 | 3,43E-02 | 1,28E-01 |
| ADIPOR1 | 29238,30 | 0,14 | 0,07 | 2,12 | 3,43E-02 | 1,28E-01 |
| TIPIN | 88,34 | 0,08 | 0,04 | 2,12 | 3,42E-02 | 1,28E-01 |
| GYPC | 18476,47 | 0,14 | 0,07 | 2,12 | 3,42E-02 | 1,28E-01 |
| PDRG1 | 305,34 | 0,04 | 0,02 | 2,12 | 3,42E-02 | 1,28E-01 |
| CLEC4D | 716,19 | -0,13 | 0,06 | -2,12 | 3,42E-02 | 1,28E-01 |
| NEK3 | 198,46 | 0,06 | 0,03 | 2,12 | 3,43E-02 | 1,28E-01 |
| MRPL4 | 89,61 | 0,10 | 0,05 | 2,12 | 3,43E-02 | 1,28E-01 |
| MBNL1 | 27257,31 | 0,05 | 0,02 | 2,12 | 3,44E-02 | 1,28E-01 |
| GPSM1 | 102,79 | 0,13 | 0,06 | 2,11 | 3,44E-02 | 1,28E-01 |
| ANGEL2 | 2844,72 | 0,06 | 0,03 | 2,11 | 3,45E-02 | 1,28E-01 |
| DCUN1D4 | 888,57 | 0,06 | 0,03 | 2,11 | 3,45E-02 | 1,28E-01 |
| BIRC2 | 4807,74 | 0,09 | 0,04 | 2,11 | 3,45E-02 | 1,28E-01 |
| ARHGAP42 | 129,41 | 0,10 | 0,05 | 2,11 | 3,45E-02 | 1,28E-01 |
| AP1AR | 635,79 | 0,09 | 0,04 | 2,11 | 3,46E-02 | 1,29E-01 |
| ITPRID2 | 3699,04 | -0,06 | 0,03 | -2,11 | 3,46E-02 | 1,29E-01 |
| GMPR | 2131,66 | 0,16 | 0,08 | 2,11 | 3,46E-02 | 1,29E-01 |
| EEF1AKMT1 | 94,23 | 0,07 | 0,03 | 2,11 | 3,46E-02 | 1,29E-01 |
| PNPO | 286,69 | 0,04 | 0,02 | 2,11 | 3,47E-02 | 1,29E-01 |
| TIMP2 | 819,26 | -0,13 | 0,06 | -2,11 | 3,47E-02 | 1,29E-01 |
| CCR7 | 2033,29 | 0,10 | 0,05 | 2,11 | 3,47E-02 | 1,29E-01 |
| FANCM | 565,91 | 0,07 | 0,03 | 2,11 | 3,47E-02 | 1,29E-01 |
| ZZZ3 | 1912,43 | 0,06 | 0,03 | 2,11 | 3,48E-02 | 1,29E-01 |
| HAX1 | 945,79 | 0,05 | 0,02 | 2,11 | 3,48E-02 | 1,29E-01 |
| TXNDC16 | 809,89 | 0,06 | 0,03 | 2,11 | 3,49E-02 | 1,29E-01 |
| DNAJC10 | 2742,94 | 0,07 | 0,03 | 2,11 | 3,49E-02 | 1,29E-01 |
| HPS4 | 931,91 | 0,04 | 0,02 | 2,11 | 3,49E-02 | 1,29E-01 |
| MAP4K3 | 698,90 | 0,06 | 0,03 | 2,11 | 3,50E-02 | 1,29E-01 |
| SPTSSA | 278,20 | 0,08 | 0,04 | 2,11 | 3,51E-02 | 1,29E-01 |
| RPS20 | 9334,21 | 0,13 | 0,06 | 2,11 | 3,50E-02 | 1,29E-01 |

| | | | | | | |
|---|---|---|---|---|---|---|
| BCL2L15 | 101,76 | 0,09 | 0,04 | 2,11 | 3,50E-02 | 1,29E-01 |
| JMY | 1366,31 | 0,05 | 0,02 | 2,11 | 3,51E-02 | 1,29E-01 |
| ZSCAN12 | 415,58 | 0,08 | 0,04 | 2,11 | 3,50E-02 | 1,29E-01 |
| COX10 | 352,94 | 0,04 | 0,02 | 2,11 | 3,50E-02 | 1,29E-01 |
| MOCS3 | 481,76 | 0,05 | 0,02 | 2,11 | 3,51E-02 | 1,29E-01 |
| SKAP2 | 3282,13 | -0,09 | 0,04 | -2,11 | 3,51E-02 | 1,29E-01 |
| RHEB | 841,25 | 0,04 | 0,02 | 2,11 | 3,51E-02 | 1,30E-01 |
| NSUN3 | 1969,27 | 0,07 | 0,03 | 2,11 | 3,52E-02 | 1,30E-01 |
| GPR55 | 120,42 | 0,10 | 0,05 | 2,11 | 3,52E-02 | 1,30E-01 |
| ARL16 | 375,78 | 0,06 | 0,03 | 2,10 | 3,53E-02 | 1,30E-01 |
| WDFY2 | 4620,70 | -0,05 | 0,02 | -2,10 | 3,53E-02 | 1,30E-01 |
| ARL1 | 771,07 | 0,06 | 0,03 | 2,10 | 3,53E-02 | 1,30E-01 |
| DYRK2 | 5789,19 | 0,06 | 0,03 | 2,10 | 3,53E-02 | 1,30E-01 |
| TRIM22 | 10817,38 | -0,10 | 0,05 | -2,10 | 3,53E-02 | 1,30E-01 |
| ZXDC | 2712,67 | -0,06 | 0,03 | -2,10 | 3,54E-02 | 1,30E-01 |
| CPNE8 | 636,27 | 0,07 | 0,03 | 2,10 | 3,54E-02 | 1,30E-01 |
| SMOX | 316,56 | 0,13 | 0,06 | 2,10 | 3,54E-02 | 1,30E-01 |
| ZNF25 | 553,58 | 0,06 | 0,03 | 2,10 | 3,54E-02 | 1,30E-01 |
| ISG20L2 | 991,31 | -0,03 | 0,02 | -2,10 | 3,55E-02 | 1,30E-01 |
| BBLN | 286,83 | 0,05 | 0,02 | 2,10 | 3,55E-02 | 1,30E-01 |
| PPP1R3F | 467,64 | 0,05 | 0,02 | 2,10 | 3,57E-02 | 1,31E-01 |
| ZDHHC21 | 1720,52 | 0,10 | 0,05 | 2,10 | 3,58E-02 | 1,31E-01 |
| CUX1 | 1932,20 | -0,08 | 0,04 | -2,10 | 3,57E-02 | 1,31E-01 |
| ANKLE2 | 2916,76 | -0,03 | 0,01 | -2,10 | 3,58E-02 | 1,31E-01 |
| VPS41 | 3960,34 | -0,05 | 0,02 | -2,10 | 3,59E-02 | 1,31E-01 |
| ORMDL1 | 1953,46 | 0,06 | 0,03 | 2,10 | 3,59E-02 | 1,31E-01 |
| PHF6 | 1085,77 | 0,08 | 0,04 | 2,10 | 3,58E-02 | 1,31E-01 |
| DDX54 | 560,66 | 0,05 | 0,02 | 2,10 | 3,60E-02 | 1,32E-01 |
| MFHAS1 | 837,51 | 0,06 | 0,03 | 2,10 | 3,60E-02 | 1,32E-01 |
| FBXO22 | 1338,55 | 0,06 | 0,03 | 2,10 | 3,60E-02 | 1,32E-01 |
| GLS | 7766,55 | 0,07 | 0,03 | 2,10 | 3,61E-02 | 1,32E-01 |
| NKAP | 490,77 | -0,03 | 0,02 | -2,09 | 3,62E-02 | 1,32E-01 |
| TRIM65 | 393,69 | 0,06 | 0,03 | 2,09 | 3,62E-02 | 1,32E-01 |
| SUN1 | 1551,70 | 0,05 | 0,02 | 2,09 | 3,62E-02 | 1,32E-01 |
| DCTPP1 | 197,94 | 0,06 | 0,03 | 2,10 | 3,62E-02 | 1,32E-01 |
| AQP1 | 79,56 | 0,16 | 0,08 | 2,09 | 3,62E-02 | 1,32E-01 |
| TMEM203 | 580,15 | 0,04 | 0,02 | 2,09 | 3,62E-02 | 1,32E-01 |
| OGFOD3 | 357,86 | 0,05 | 0,02 | 2,09 | 3,62E-02 | 1,32E-01 |
| RARG | 241,30 | 0,08 | 0,04 | 2,10 | 3,61E-02 | 1,32E-01 |
| ZNF599 | 160,59 | 0,06 | 0,03 | 2,10 | 3,62E-02 | 1,32E-01 |
| TUBB2A | 729,23 | 0,34 | 0,16 | 2,09 | 3,62E-02 | 1,32E-01 |
| IPPK | 531,46 | -0,03 | 0,01 | -2,10 | 3,62E-02 | 1,32E-01 |
| ABCB7 | 790,89 | 0,05 | 0,02 | 2,09 | 3,62E-02 | 1,32E-01 |
| HLA-C | 50227,97 | -0,08 | 0,04 | -2,09 | 3,62E-02 | 1,32E-01 |
| DNAJA3 | 847,38 | 0,04 | 0,02 | 2,10 | 3,61E-02 | 1,32E-01 |
| OST4 | 3047,41 | 0,08 | 0,04 | 2,09 | 3,63E-02 | 1,32E-01 |

| | | | | | | |
|---|---|---|---|---|---|---|
| NDUFAF4 | 189,67 | 0,10 | 0,05 | 2,09 | 3,63E-02 | 1,32E-01 |
| NR3C2 | 923,32 | 0,07 | 0,03 | 2,09 | 3,63E-02 | 1,32E-01 |
| PSPH | 73,94 | 0,08 | 0,04 | 2,09 | 3,64E-02 | 1,32E-01 |
| RSL24D1 | 1077,11 | 0,17 | 0,08 | 2,09 | 3,64E-02 | 1,32E-01 |
| IPO5 | 1480,87 | 0,04 | 0,02 | 2,09 | 3,64E-02 | 1,32E-01 |
| CATSPERB | 243,19 | 0,09 | 0,04 | 2,09 | 3,64E-02 | 1,32E-01 |
| ATG16L2 | 10528,90 | -0,11 | 0,05 | -2,09 | 3,64E-02 | 1,32E-01 |
| PDHX | 471,30 | 0,05 | 0,03 | 2,09 | 3,64E-02 | 1,32E-01 |
| MAP2K1 | 1597,43 | -0,04 | 0,02 | -2,09 | 3,65E-02 | 1,32E-01 |
| FAM3C | 745,92 | 0,10 | 0,05 | 2,09 | 3,65E-02 | 1,32E-01 |
| TLR7 | 491,39 | 0,08 | 0,04 | 2,09 | 3,66E-02 | 1,33E-01 |
| RSAD1 | 571,93 | 0,03 | 0,02 | 2,09 | 3,66E-02 | 1,33E-01 |
| P3H1 | 493,89 | 0,04 | 0,02 | 2,09 | 3,66E-02 | 1,33E-01 |
| AKR1E2 | 176,88 | 0,14 | 0,07 | 2,09 | 3,67E-02 | 1,33E-01 |
| MED13 | 7431,58 | -0,04 | 0,02 | -2,09 | 3,66E-02 | 1,33E-01 |
| GTF2H3 | 509,47 | 0,05 | 0,03 | 2,09 | 3,67E-02 | 1,33E-01 |
| PRKRA | 627,94 | 0,04 | 0,02 | 2,09 | 3,68E-02 | 1,33E-01 |
| SRSF2 | 2951,17 | 0,04 | 0,02 | 2,09 | 3,68E-02 | 1,33E-01 |
| HSPBP1 | 98,74 | 0,07 | 0,03 | 2,09 | 3,68E-02 | 1,33E-01 |
| GEMIN6 | 283,72 | 0,06 | 0,03 | 2,09 | 3,69E-02 | 1,33E-01 |
| SPINT2 | 1035,59 | 0,05 | 0,02 | 2,09 | 3,69E-02 | 1,33E-01 |
| IL7 | 103,75 | 0,12 | 0,06 | 2,09 | 3,70E-02 | 1,34E-01 |
| STAG1 | 3514,79 | -0,05 | 0,02 | -2,08 | 3,71E-02 | 1,34E-01 |
| ZNF461 | 314,81 | 0,06 | 0,03 | 2,08 | 3,72E-02 | 1,34E-01 |
| ECHDC3 | 182,26 | -0,11 | 0,05 | -2,08 | 3,73E-02 | 1,35E-01 |
| TBCCD1 | 478,00 | 0,04 | 0,02 | 2,08 | 3,73E-02 | 1,35E-01 |
| ORC5 | 672,32 | 0,07 | 0,03 | 2,08 | 3,73E-02 | 1,35E-01 |
| NT5M | 227,33 | 0,15 | 0,07 | 2,08 | 3,73E-02 | 1,35E-01 |
| ZNF260 | 390,31 | 0,10 | 0,05 | 2,08 | 3,74E-02 | 1,35E-01 |
| GRK5 | 955,15 | -0,04 | 0,02 | -2,08 | 3,75E-02 | 1,35E-01 |
| RUFY3 | 1500,29 | 0,04 | 0,02 | 2,08 | 3,75E-02 | 1,35E-01 |
| WDR89 | 489,40 | 0,12 | 0,06 | 2,08 | 3,75E-02 | 1,35E-01 |
| INO80C | 203,96 | -0,05 | 0,02 | -2,08 | 3,75E-02 | 1,35E-01 |
| ZNF280C | 222,60 | 0,05 | 0,02 | 2,08 | 3,75E-02 | 1,35E-01 |
| B4GALT3 | 374,29 | 0,08 | 0,04 | 2,08 | 3,76E-02 | 1,35E-01 |
| IER3IP1 | 289,54 | 0,10 | 0,05 | 2,08 | 3,77E-02 | 1,36E-01 |
| SETD6 | 324,52 | 0,04 | 0,02 | 2,08 | 3,77E-02 | 1,36E-01 |
| GADD45B | 652,49 | -0,08 | 0,04 | -2,08 | 3,78E-02 | 1,36E-01 |
| SFT2D1 | 1443,13 | -0,05 | 0,02 | -2,08 | 3,79E-02 | 1,36E-01 |
| ZNF324B | 175,59 | 0,05 | 0,02 | 2,08 | 3,79E-02 | 1,36E-01 |
| TRAT1 | 1401,27 | 0,12 | 0,06 | 2,08 | 3,79E-02 | 1,36E-01 |
| COX4I1 | 3243,30 | 0,06 | 0,03 | 2,08 | 3,79E-02 | 1,36E-01 |
| TFIP11 | 1776,16 | -0,05 | 0,02 | -2,08 | 3,79E-02 | 1,36E-01 |
| SNX5 | 2304,33 | 0,04 | 0,02 | 2,08 | 3,79E-02 | 1,36E-01 |
| TRAF1 | 1489,13 | 0,07 | 0,03 | 2,08 | 3,79E-02 | 1,36E-01 |
| NOA1 | 489,05 | 0,03 | 0,01 | 2,07 | 3,80E-02 | 1,36E-01 |

| | | | | | | |
|---|---|---|---|---|---|---|
| IER2 | 1966,98 | -0,06 | 0,03 | -2,07 | 3,81E-02 | 1,37E-01 |
| OSTC | 726,46 | 0,09 | 0,04 | 2,07 | 3,82E-02 | 1,37E-01 |
| PTPRC | 72486,71 | -0,06 | 0,03 | -2,07 | 3,81E-02 | 1,37E-01 |
| RPS27L | 852,57 | 0,07 | 0,04 | 2,07 | 3,82E-02 | 1,37E-01 |
| MAP1LC3A | 81,66 | -0,11 | 0,06 | -2,07 | 3,82E-02 | 1,37E-01 |
| YIPF6 | 1971,43 | 0,06 | 0,03 | 2,07 | 3,82E-02 | 1,37E-01 |
| LILRA5 | 3194,48 | -0,10 | 0,05 | -2,07 | 3,81E-02 | 1,37E-01 |
| SLC35B2 | 432,35 | 0,04 | 0,02 | 2,07 | 3,82E-02 | 1,37E-01 |
| EXOSC1 | 694,24 | 0,04 | 0,02 | 2,07 | 3,83E-02 | 1,37E-01 |
| GLB1 | 1701,00 | -0,05 | 0,02 | -2,07 | 3,83E-02 | 1,37E-01 |
| ATAD2 | 799,86 | 0,07 | 0,03 | 2,07 | 3,83E-02 | 1,37E-01 |
| NKAPD1 | 781,18 | 0,07 | 0,03 | 2,07 | 3,83E-02 | 1,37E-01 |
| NSMCE1 | 533,22 | 0,04 | 0,02 | 2,07 | 3,83E-02 | 1,37E-01 |
| LILRA1 | 1533,55 | -0,08 | 0,04 | -2,07 | 3,83E-02 | 1,37E-01 |
| MARK2 | 1275,66 | -0,11 | 0,05 | -2,07 | 3,84E-02 | 1,37E-01 |
| ZNF37A | 1096,92 | 0,08 | 0,04 | 2,07 | 3,84E-02 | 1,37E-01 |
| TIMM8B | 327,16 | 0,12 | 0,06 | 2,07 | 3,84E-02 | 1,37E-01 |
| STK3 | 645,74 | -0,07 | 0,04 | -2,07 | 3,85E-02 | 1,37E-01 |
| RGL2 | 1836,84 | -0,09 | 0,04 | -2,07 | 3,85E-02 | 1,37E-01 |
| EMG1 | 1022,13 | 0,05 | 0,02 | 2,07 | 3,85E-02 | 1,37E-01 |
| RUBCN | 1117,33 | -0,10 | 0,05 | -2,07 | 3,86E-02 | 1,37E-01 |
| ANPEP | 1231,00 | -0,16 | 0,08 | -2,07 | 3,86E-02 | 1,37E-01 |
| KCNA3 | 1078,46 | 0,07 | 0,03 | 2,07 | 3,86E-02 | 1,38E-01 |
| DCBLD2 | 104,08 | 0,13 | 0,06 | 2,07 | 3,87E-02 | 1,38E-01 |
| HERC1 | 14913,00 | -0,04 | 0,02 | -2,07 | 3,87E-02 | 1,38E-01 |
| QSER1 | 1209,29 | 0,04 | 0,02 | 2,07 | 3,87E-02 | 1,38E-01 |
| SLC25A32 | 327,18 | 0,06 | 0,03 | 2,07 | 3,88E-02 | 1,38E-01 |
| GMFB | 2031,16 | 0,11 | 0,05 | 2,07 | 3,88E-02 | 1,38E-01 |
| RAB5IF | 1256,70 | -0,06 | 0,03 | -2,07 | 3,88E-02 | 1,38E-01 |
| SIRT7 | 1629,09 | -0,05 | 0,02 | -2,07 | 3,88E-02 | 1,38E-01 |
| ZNF101 | 2819,10 | 0,05 | 0,02 | 2,06 | 3,90E-02 | 1,38E-01 |
| E2F4 | 873,91 | 0,07 | 0,03 | 2,06 | 3,90E-02 | 1,38E-01 |
| SYS1 | 540,16 | -0,05 | 0,02 | -2,06 | 3,90E-02 | 1,38E-01 |
| CCZ1 | 568,54 | -0,05 | 0,02 | -2,06 | 3,90E-02 | 1,38E-01 |
| NFKBIB | 304,86 | -0,05 | 0,02 | -2,06 | 3,90E-02 | 1,38E-01 |
| SLC25A12 | 782,40 | 0,04 | 0,02 | 2,06 | 3,92E-02 | 1,39E-01 |
| BCL2 | 5549,26 | 0,07 | 0,03 | 2,06 | 3,93E-02 | 1,39E-01 |
| GPR160 | 756,45 | -0,08 | 0,04 | -2,06 | 3,93E-02 | 1,39E-01 |
| ADORA2A | 169,12 | 0,06 | 0,03 | 2,06 | 3,94E-02 | 1,39E-01 |
| PAQR8 | 625,19 | 0,06 | 0,03 | 2,06 | 3,94E-02 | 1,40E-01 |
| TUBGCP5 | 597,87 | 0,05 | 0,02 | 2,06 | 3,95E-02 | 1,40E-01 |
| EBP | 313,71 | 0,05 | 0,03 | 2,06 | 3,95E-02 | 1,40E-01 |
| ZNF7 | 574,66 | 0,03 | 0,02 | 2,06 | 3,95E-02 | 1,40E-01 |
| PLAG1 | 373,98 | 0,09 | 0,04 | 2,06 | 3,96E-02 | 1,40E-01 |
| PLIN5 | 324,04 | -0,12 | 0,06 | -2,06 | 3,96E-02 | 1,40E-01 |
| THAP9 | 150,76 | 0,09 | 0,04 | 2,06 | 3,96E-02 | 1,40E-01 |

| | | | | | | |
|---|---|---|---|---|---|---|
| FAR2 | 1529,00 | -0,08 | 0,04 | -2,06 | 3,96E-02 | 1,40E-01 |
| AKAP5 | 162,64 | -0,13 | 0,06 | -2,06 | 3,96E-02 | 1,40E-01 |
| DDHD1 | 3706,74 | 0,05 | 0,02 | 2,06 | 3,97E-02 | 1,40E-01 |
| POLD1 | 376,05 | 0,05 | 0,03 | 2,06 | 3,97E-02 | 1,40E-01 |
| OPN3 | 270,45 | 0,05 | 0,02 | 2,06 | 3,97E-02 | 1,40E-01 |
| ERCC8 | 484,74 | 0,07 | 0,03 | 2,06 | 3,98E-02 | 1,40E-01 |
| ILF3 | 3725,23 | 0,04 | 0,02 | 2,06 | 3,98E-02 | 1,40E-01 |
| BABAM1 | 708,96 | 0,08 | 0,04 | 2,06 | 3,98E-02 | 1,40E-01 |
| ACTR1A | 1496,48 | -0,05 | 0,02 | -2,06 | 3,98E-02 | 1,40E-01 |
| FBXO6 | 285,50 | -0,09 | 0,05 | -2,06 | 3,99E-02 | 1,40E-01 |
| ZBTB16 | 423,41 | -0,12 | 0,06 | -2,05 | 3,99E-02 | 1,41E-01 |
| EIF3H | 3476,76 | 0,06 | 0,03 | 2,05 | 3,99E-02 | 1,41E-01 |
| FBXL4 | 2345,50 | 0,05 | 0,02 | 2,05 | 4,00E-02 | 1,41E-01 |
| SLC17A9 | 129,70 | 0,11 | 0,06 | 2,05 | 4,00E-02 | 1,41E-01 |
| AP4S1 | 514,10 | 0,04 | 0,02 | 2,05 | 4,00E-02 | 1,41E-01 |
| SMARCAD1 | 1233,64 | 0,07 | 0,04 | 2,05 | 4,01E-02 | 1,41E-01 |
| UQCR10 | 535,41 | 0,07 | 0,03 | 2,05 | 4,01E-02 | 1,41E-01 |
| SNF8 | 868,10 | 0,03 | 0,01 | 2,05 | 4,01E-02 | 1,41E-01 |
| BTBD2 | 286,15 | 0,10 | 0,05 | 2,05 | 4,01E-02 | 1,41E-01 |
| CHCHD1 | 285,66 | 0,07 | 0,03 | 2,05 | 4,01E-02 | 1,41E-01 |
| NMD3 | 817,66 | 0,10 | 0,05 | 2,05 | 4,01E-02 | 1,41E-01 |
| SLC25A37 | 73347,57 | 0,11 | 0,05 | 2,05 | 4,02E-02 | 1,41E-01 |
| BIRC3 | 6224,80 | 0,11 | 0,05 | 2,05 | 4,02E-02 | 1,41E-01 |
| COX5A | 150,49 | 0,05 | 0,03 | 2,05 | 4,02E-02 | 1,41E-01 |
| RNF7 | 801,09 | 0,05 | 0,02 | 2,05 | 4,02E-02 | 1,41E-01 |
| R3HCC1L | 1486,65 | -0,05 | 0,02 | -2,05 | 4,02E-02 | 1,41E-01 |
| NR1D2 | 1665,12 | 0,07 | 0,03 | 2,05 | 4,03E-02 | 1,41E-01 |
| FCGR3A | 5292,86 | -0,08 | 0,04 | -2,05 | 4,05E-02 | 1,42E-01 |
| TRAPPC12 | 2367,54 | -0,06 | 0,03 | -2,05 | 4,04E-02 | 1,42E-01 |
| TMEM254 | 190,49 | 0,04 | 0,02 | 2,05 | 4,05E-02 | 1,42E-01 |
| ARHGAP9 | 3315,23 | -0,10 | 0,05 | -2,05 | 4,05E-02 | 1,42E-01 |
| FAAP20 | 259,93 | 0,08 | 0,04 | 2,05 | 4,06E-02 | 1,42E-01 |
| ZBTB33 | 1157,45 | 0,09 | 0,04 | 2,05 | 4,07E-02 | 1,42E-01 |
| KLRC2 | 185,28 | -0,21 | 0,10 | -2,05 | 4,07E-02 | 1,42E-01 |
| NDUFS2 | 1288,12 | -0,03 | 0,02 | -2,05 | 4,07E-02 | 1,42E-01 |
| CEP70 | 227,42 | 0,09 | 0,04 | 2,05 | 4,07E-02 | 1,42E-01 |
| SSR2 | 2668,87 | 0,05 | 0,03 | 2,05 | 4,07E-02 | 1,42E-01 |
| FNBP1 | 3706,90 | -0,05 | 0,02 | -2,05 | 4,08E-02 | 1,42E-01 |
| SLC27A5 | 144,97 | 0,07 | 0,03 | 2,05 | 4,08E-02 | 1,42E-01 |
| ZNF300 | 132,02 | 0,12 | 0,06 | 2,05 | 4,08E-02 | 1,43E-01 |
| ZRANB2 | 4228,42 | 0,09 | 0,04 | 2,04 | 4,09E-02 | 1,43E-01 |
| SETDB2 | 1390,53 | 0,04 | 0,02 | 2,04 | 4,09E-02 | 1,43E-01 |
| TTLL1 | 106,45 | 0,06 | 0,03 | 2,04 | 4,09E-02 | 1,43E-01 |
| SESN1 | 1408,26 | 0,07 | 0,03 | 2,04 | 4,09E-02 | 1,43E-01 |
| PRF1 | 1880,30 | -0,11 | 0,05 | -2,04 | 4,10E-02 | 1,43E-01 |
| APOLD1 | 266,76 | -0,07 | 0,03 | -2,04 | 4,10E-02 | 1,43E-01 |

| | | | | | | |
|---|---|---|---|---|---|---|
| SRSF11 | 6251,43 | 0,06 | 0,03 | 2,04 | 4,11E-02 | 1,43E-01 |
| ATP5F1C | 1112,97 | 0,08 | 0,04 | 2,04 | 4,11E-02 | 1,43E-01 |
| MAIP1 | 121,72 | 0,06 | 0,03 | 2,04 | 4,11E-02 | 1,43E-01 |
| H2BC4 | 4001,89 | 0,08 | 0,04 | 2,04 | 4,12E-02 | 1,43E-01 |
| CCNI | 6316,61 | 0,09 | 0,04 | 2,04 | 4,12E-02 | 1,43E-01 |
| TMEM134 | 169,89 | 0,08 | 0,04 | 2,04 | 4,12E-02 | 1,43E-01 |
| PRPF39 | 1004,86 | 0,07 | 0,03 | 2,04 | 4,13E-02 | 1,43E-01 |
| YARS2 | 551,75 | 0,05 | 0,02 | 2,04 | 4,12E-02 | 1,43E-01 |
| ATXN3 | 1917,47 | 0,03 | 0,02 | 2,04 | 4,12E-02 | 1,43E-01 |
| ZBTB24 | 1270,25 | 0,04 | 0,02 | 2,04 | 4,13E-02 | 1,43E-01 |
| RPL7L1 | 2240,29 | 0,04 | 0,02 | 2,04 | 4,12E-02 | 1,43E-01 |
| FOXO3 | 10112,32 | 0,10 | 0,05 | 2,04 | 4,12E-02 | 1,43E-01 |
| C2orf68 | 1871,73 | -0,03 | 0,02 | -2,04 | 4,14E-02 | 1,43E-01 |
| ACER3 | 1089,85 | 0,05 | 0,02 | 2,04 | 4,14E-02 | 1,43E-01 |
| C8orf82 | 214,88 | 0,06 | 0,03 | 2,04 | 4,13E-02 | 1,43E-01 |
| PAQR3 | 464,39 | 0,06 | 0,03 | 2,04 | 4,14E-02 | 1,43E-01 |
| PTGS2 | 1363,64 | -0,09 | 0,05 | -2,04 | 4,14E-02 | 1,43E-01 |
| SRGN | 32575,40 | -0,12 | 0,06 | -2,04 | 4,14E-02 | 1,43E-01 |
| CCDC142 | 1129,60 | 0,04 | 0,02 | 2,04 | 4,14E-02 | 1,43E-01 |
| SRA1 | 594,40 | -0,05 | 0,02 | -2,04 | 4,15E-02 | 1,43E-01 |
| SNX4 | 507,60 | 0,10 | 0,05 | 2,04 | 4,15E-02 | 1,44E-01 |
| GNAQ | 6024,87 | -0,06 | 0,03 | -2,04 | 4,16E-02 | 1,44E-01 |
| BNIP3 | 422,02 | 0,05 | 0,03 | 2,04 | 4,16E-02 | 1,44E-01 |
| EIF3F | 2783,07 | 0,05 | 0,02 | 2,04 | 4,17E-02 | 1,44E-01 |
| RNF175 | 79,13 | 0,13 | 0,06 | 2,04 | 4,17E-02 | 1,44E-01 |
| SLC25A5 | 1566,51 | 0,05 | 0,02 | 2,04 | 4,17E-02 | 1,44E-01 |
| MLH3 | 1272,89 | 0,05 | 0,03 | 2,04 | 4,17E-02 | 1,44E-01 |
| KCTD9 | 996,18 | 0,05 | 0,03 | 2,04 | 4,17E-02 | 1,44E-01 |
| GNAL | 96,12 | -0,14 | 0,07 | -2,04 | 4,17E-02 | 1,44E-01 |
| DNAJA4 | 1088,68 | 0,11 | 0,05 | 2,04 | 4,18E-02 | 1,44E-01 |
| RILPL1 | 138,10 | -0,10 | 0,05 | -2,04 | 4,18E-02 | 1,44E-01 |
| SUSD6 | 2201,95 | -0,10 | 0,05 | -2,04 | 4,18E-02 | 1,44E-01 |
| VMAC | 318,19 | 0,06 | 0,03 | 2,04 | 4,18E-02 | 1,44E-01 |
| WAC | 7116,90 | -0,04 | 0,02 | -2,03 | 4,19E-02 | 1,44E-01 |
| KLHL9 | 2258,94 | 0,08 | 0,04 | 2,03 | 4,19E-02 | 1,44E-01 |
| LRRC69 | 208,75 | 0,05 | 0,03 | 2,03 | 4,19E-02 | 1,44E-01 |
| MRPL35 | 527,25 | 0,06 | 0,03 | 2,03 | 4,20E-02 | 1,44E-01 |
| BORA | 297,58 | 0,07 | 0,03 | 2,03 | 4,20E-02 | 1,45E-01 |
| HBB | 582,48 | 0,22 | 0,11 | 2,03 | 4,21E-02 | 1,45E-01 |
| GNAI2 | 4415,51 | -0,08 | 0,04 | -2,03 | 4,21E-02 | 1,45E-01 |
| OSBP | 1366,65 | -0,03 | 0,01 | -2,03 | 4,21E-02 | 1,45E-01 |
| RBBP4 | 2926,17 | 0,03 | 0,02 | 2,03 | 4,22E-02 | 1,45E-01 |
| BTD | 519,86 | -0,04 | 0,02 | -2,03 | 4,21E-02 | 1,45E-01 |
| IFT70A | 106,76 | 0,09 | 0,04 | 2,03 | 4,22E-02 | 1,45E-01 |
| C18orf54 | 194,47 | 0,08 | 0,04 | 2,03 | 4,22E-02 | 1,45E-01 |
| ZFP90 | 1140,33 | 0,05 | 0,03 | 2,03 | 4,23E-02 | 1,45E-01 |

| | | | | | | |
|---|---|---|---|---|---|---|
| METTL8 | 739,53 | 0,08 | 0,04 | 2,03 | 4,23E-02 | 1,45E-01 |
| IDUA | 208,16 | 0,11 | 0,05 | 2,03 | 4,24E-02 | 1,45E-01 |
| ARL6 | 192,43 | 0,09 | 0,04 | 2,03 | 4,24E-02 | 1,45E-01 |
| SIRPA | 2589,68 | -0,10 | 0,05 | -2,03 | 4,24E-02 | 1,45E-01 |
| PSMD14 | 675,16 | 0,09 | 0,04 | 2,03 | 4,24E-02 | 1,45E-01 |
| MRPL37 | 381,00 | 0,03 | 0,02 | 2,03 | 4,24E-02 | 1,45E-01 |
| UBTD2 | 247,75 | 0,04 | 0,02 | 2,03 | 4,26E-02 | 1,46E-01 |
| ARV1 | 181,06 | 0,06 | 0,03 | 2,03 | 4,25E-02 | 1,46E-01 |
| APPL1 | 2231,58 | 0,05 | 0,03 | 2,03 | 4,26E-02 | 1,46E-01 |
| FAM174C | 187,53 | 0,05 | 0,03 | 2,03 | 4,26E-02 | 1,46E-01 |
| CMTR1 | 2292,53 | -0,06 | 0,03 | -2,03 | 4,26E-02 | 1,46E-01 |
| DTD1 | 384,05 | -0,04 | 0,02 | -2,03 | 4,26E-02 | 1,46E-01 |
| PPP1R7 | 878,11 | -0,04 | 0,02 | -2,03 | 4,26E-02 | 1,46E-01 |
| NDUFS3 | 383,10 | 0,05 | 0,03 | 2,03 | 4,26E-02 | 1,46E-01 |
| UBE2N | 1883,29 | 0,05 | 0,02 | 2,03 | 4,26E-02 | 1,46E-01 |
| RPL22L1 | 242,22 | 0,12 | 0,06 | 2,03 | 4,27E-02 | 1,46E-01 |
| TMEM107 | 102,95 | 0,05 | 0,02 | 2,03 | 4,27E-02 | 1,46E-01 |
| ZNF706 | 1666,43 | 0,05 | 0,02 | 2,03 | 4,28E-02 | 1,46E-01 |
| PLD4 | 225,83 | 0,10 | 0,05 | 2,03 | 4,29E-02 | 1,46E-01 |
| FHIP2A | 2516,56 | -0,06 | 0,03 | -2,02 | 4,30E-02 | 1,47E-01 |
| SPSB3 | 1472,73 | 0,05 | 0,02 | 2,02 | 4,31E-02 | 1,47E-01 |
| FBXL15 | 372,82 | 0,04 | 0,02 | 2,02 | 4,31E-02 | 1,47E-01 |
| NFXL1 | 1099,92 | -0,12 | 0,06 | -2,02 | 4,32E-02 | 1,47E-01 |
| KATNBL1 | 3238,93 | -0,07 | 0,04 | -2,02 | 4,32E-02 | 1,47E-01 |
| ZC3H7A | 1771,03 | -0,03 | 0,02 | -2,02 | 4,31E-02 | 1,47E-01 |
| ZNF91 | 1812,51 | 0,08 | 0,04 | 2,02 | 4,32E-02 | 1,47E-01 |
| UQCRQ | 460,49 | 0,09 | 0,05 | 2,02 | 4,32E-02 | 1,47E-01 |
| ATP6V0E2 | 289,91 | 0,07 | 0,03 | 2,02 | 4,32E-02 | 1,47E-01 |
| ELP5 | 267,73 | 0,04 | 0,02 | 2,02 | 4,32E-02 | 1,47E-01 |
| CFAP45 | 235,13 | -0,11 | 0,05 | -2,02 | 4,32E-02 | 1,47E-01 |
| RBAK | 863,64 | 0,10 | 0,05 | 2,02 | 4,33E-02 | 1,47E-01 |
| PEPD | 675,44 | 0,04 | 0,02 | 2,02 | 4,33E-02 | 1,47E-01 |
| AFF4 | 4045,55 | -0,04 | 0,02 | -2,02 | 4,34E-02 | 1,47E-01 |
| ZNF627 | 362,10 | 0,07 | 0,03 | 2,02 | 4,34E-02 | 1,47E-01 |
| DUSP18 | 364,99 | -0,07 | 0,03 | -2,02 | 4,34E-02 | 1,47E-01 |
| SMIM13 | 327,22 | 0,06 | 0,03 | 2,02 | 4,34E-02 | 1,48E-01 |
| TARS2 | 385,70 | 0,04 | 0,02 | 2,02 | 4,35E-02 | 1,48E-01 |
| DNAJA1 | 2508,16 | -0,10 | 0,05 | -2,02 | 4,35E-02 | 1,48E-01 |
| CEP20 | 498,56 | 0,07 | 0,03 | 2,02 | 4,35E-02 | 1,48E-01 |
| EID1 | 1977,99 | 0,09 | 0,05 | 2,02 | 4,35E-02 | 1,48E-01 |
| NUBP2 | 796,30 | 0,04 | 0,02 | 2,02 | 4,35E-02 | 1,48E-01 |
| SLC19A2 | 226,02 | 0,05 | 0,03 | 2,02 | 4,36E-02 | 1,48E-01 |
| EPS15L1 | 1126,97 | -0,10 | 0,05 | -2,02 | 4,36E-02 | 1,48E-01 |
| ADCK5 | 153,98 | 0,07 | 0,03 | 2,02 | 4,36E-02 | 1,48E-01 |
| WDR13 | 1239,30 | 0,05 | 0,02 | 2,02 | 4,36E-02 | 1,48E-01 |
| ITGB3BP | 379,85 | 0,09 | 0,04 | 2,02 | 4,37E-02 | 1,48E-01 |

| | | | | | | |
|---|---|---|---|---|---|---|
| SLC38A5 | 286,79 | 0,15 | 0,07 | 2,02 | 4,37E-02 | 1,48E-01 |
| DHX34 | 1901,32 | -0,09 | 0,04 | -2,02 | 4,37E-02 | 1,48E-01 |
| ZNF441 | 593,75 | 0,13 | 0,06 | 2,02 | 4,37E-02 | 1,48E-01 |
| ZNF416 | 236,72 | 0,06 | 0,03 | 2,02 | 4,38E-02 | 1,48E-01 |
| SOCS4 | 1438,94 | 0,08 | 0,04 | 2,02 | 4,38E-02 | 1,48E-01 |
| RAD50 | 149,04 | 0,04 | 0,02 | 2,01 | 4,39E-02 | 1,48E-01 |
| CALHM2 | 397,49 | 0,07 | 0,03 | 2,01 | 4,40E-02 | 1,48E-01 |
| ZNF592 | 1873,84 | -0,08 | 0,04 | -2,01 | 4,40E-02 | 1,48E-01 |
| ARMC1 | 702,16 | 0,06 | 0,03 | 2,01 | 4,40E-02 | 1,48E-01 |
| EEA1 | 1255,58 | 0,07 | 0,03 | 2,01 | 4,40E-02 | 1,49E-01 |
| RNF122 | 229,85 | -0,08 | 0,04 | -2,01 | 4,41E-02 | 1,49E-01 |
| PACC1 | 475,05 | 0,03 | 0,02 | 2,01 | 4,41E-02 | 1,49E-01 |
| TSHZ3 | 328,83 | -0,09 | 0,05 | -2,01 | 4,42E-02 | 1,49E-01 |
| CDC5L | 2178,74 | -0,06 | 0,03 | -2,01 | 4,42E-02 | 1,49E-01 |
| ARFGEF1 | 5924,96 | -0,04 | 0,02 | -2,01 | 4,42E-02 | 1,49E-01 |
| LDLR | 266,73 | 0,09 | 0,04 | 2,01 | 4,43E-02 | 1,49E-01 |
| STRADB | 18172,58 | 0,14 | 0,07 | 2,01 | 4,43E-02 | 1,49E-01 |
| PLVAP | 157,35 | 0,17 | 0,08 | 2,01 | 4,43E-02 | 1,49E-01 |
| DCAKD | 306,38 | 0,04 | 0,02 | 2,01 | 4,44E-02 | 1,49E-01 |
| ZNF649 | 605,24 | 0,06 | 0,03 | 2,01 | 4,44E-02 | 1,49E-01 |
| STX4 | 976,11 | -0,05 | 0,02 | -2,01 | 4,44E-02 | 1,49E-01 |
| ZNF16 | 186,61 | 0,05 | 0,02 | 2,01 | 4,45E-02 | 1,50E-01 |
| JMJD8 | 132,84 | 0,08 | 0,04 | 2,01 | 4,46E-02 | 1,50E-01 |
| MRPS18B | 584,95 | 0,05 | 0,02 | 2,01 | 4,46E-02 | 1,50E-01 |
| MCOLN1 | 335,58 | 0,11 | 0,06 | 2,01 | 4,47E-02 | 1,50E-01 |
| ENPP4 | 781,21 | 0,11 | 0,06 | 2,01 | 4,48E-02 | 1,50E-01 |
| EXOSC2 | 921,33 | 0,04 | 0,02 | 2,01 | 4,48E-02 | 1,50E-01 |
| PSMF1 | 2527,91 | 0,11 | 0,05 | 2,01 | 4,48E-02 | 1,50E-01 |
| AKT3 | 2227,26 | 0,05 | 0,03 | 2,01 | 4,49E-02 | 1,50E-01 |
| SREK1 | 4409,57 | 0,06 | 0,03 | 2,01 | 4,49E-02 | 1,50E-01 |
| PUM2 | 7770,73 | -0,04 | 0,02 | -2,01 | 4,49E-02 | 1,50E-01 |
| PHF10 | 1403,83 | 0,06 | 0,03 | 2,00 | 4,50E-02 | 1,51E-01 |
| HLA-B | 74620,33 | -0,06 | 0,03 | -2,00 | 4,50E-02 | 1,51E-01 |
| GBP5 | 12019,20 | -0,13 | 0,07 | -2,00 | 4,50E-02 | 1,51E-01 |
| N4BP2 | 1625,90 | 0,08 | 0,04 | 2,00 | 4,51E-02 | 1,51E-01 |
| INPP4B | 5156,37 | 0,08 | 0,04 | 2,00 | 4,51E-02 | 1,51E-01 |
| TMEM204 | 665,80 | 0,09 | 0,04 | 2,00 | 4,51E-02 | 1,51E-01 |
| VAMP3 | 1894,34 | -0,05 | 0,02 | -2,00 | 4,51E-02 | 1,51E-01 |
| FBXW11 | 2222,96 | -0,03 | 0,02 | -2,00 | 4,52E-02 | 1,51E-01 |
| SKIC8 | 843,04 | 0,03 | 0,02 | 2,00 | 4,52E-02 | 1,51E-01 |
| NDUFAF5 | 506,46 | 0,04 | 0,02 | 2,00 | 4,52E-02 | 1,51E-01 |
| MAX | 3646,14 | -0,04 | 0,02 | -2,00 | 4,52E-02 | 1,51E-01 |
| CNOT10 | 1195,20 | -0,03 | 0,01 | -2,00 | 4,53E-02 | 1,51E-01 |
| GALT | 829,35 | 0,06 | 0,03 | 2,00 | 4,53E-02 | 1,51E-01 |
| H2AZ1 | 3181,63 | -0,08 | 0,04 | -2,00 | 4,53E-02 | 1,51E-01 |
| CISH | 498,86 | 0,11 | 0,05 | 2,00 | 4,53E-02 | 1,51E-01 |

| | | | | | | |
|---|---|---|---|---|---|---|
| MFSD2B | 261,88 | 0,14 | 0,07 | 2,00 | 4,53E-02 | 1,51E-01 |
| LMNA | 308,68 | 0,11 | 0,05 | 2,00 | 4,53E-02 | 1,51E-01 |
| PRMT3 | 828,97 | 0,04 | 0,02 | 2,00 | 4,54E-02 | 1,51E-01 |
| SLC26A2 | 934,92 | 0,04 | 0,02 | 2,00 | 4,54E-02 | 1,51E-01 |
| EMSY | 2255,11 | -0,04 | 0,02 | -2,00 | 4,54E-02 | 1,51E-01 |
| TIMM10B | 578,51 | 0,04 | 0,02 | 2,00 | 4,54E-02 | 1,51E-01 |
| DENND10 | 1722,79 | -0,05 | 0,02 | -2,00 | 4,54E-02 | 1,51E-01 |
| RNF31 | 728,97 | -0,06 | 0,03 | -2,00 | 4,56E-02 | 1,52E-01 |
| SS18 | 2494,10 | 0,05 | 0,02 | 2,00 | 4,57E-02 | 1,52E-01 |
| COL19A1 | 711,49 | 0,12 | 0,06 | 2,00 | 4,57E-02 | 1,52E-01 |
| CSNK1G3 | 1224,11 | 0,09 | 0,04 | 2,00 | 4,57E-02 | 1,52E-01 |
| RFX5 | 1691,76 | 0,04 | 0,02 | 2,00 | 4,57E-02 | 1,52E-01 |
| SRFBP1 | 389,24 | 0,06 | 0,03 | 2,00 | 4,58E-02 | 1,52E-01 |
| ZNF23 | 178,21 | 0,06 | 0,03 | 2,00 | 4,58E-02 | 1,52E-01 |
| YEATS4 | 232,00 | 0,09 | 0,05 | 2,00 | 4,58E-02 | 1,52E-01 |
| FBXL17 | 653,25 | 0,04 | 0,02 | 2,00 | 4,58E-02 | 1,52E-01 |
| PTPN1 | 1958,83 | -0,04 | 0,02 | -2,00 | 4,59E-02 | 1,52E-01 |
| EID2 | 242,39 | 0,06 | 0,03 | 2,00 | 4,59E-02 | 1,52E-01 |
| TAF4B | 346,39 | 0,06 | 0,03 | 2,00 | 4,60E-02 | 1,53E-01 |
| NFKBIE | 458,40 | 0,05 | 0,03 | 2,00 | 4,60E-02 | 1,53E-01 |
| VSTM1 | 226,43 | 0,12 | 0,06 | 1,99 | 4,61E-02 | 1,53E-01 |
| ARRDC2 | 1580,37 | 0,05 | 0,02 | 1,99 | 4,62E-02 | 1,53E-01 |
| RUNDC1 | 549,95 | -0,06 | 0,03 | -1,99 | 4,62E-02 | 1,53E-01 |
| FBXL5 | 16268,07 | -0,08 | 0,04 | -1,99 | 4,62E-02 | 1,53E-01 |
| MAPRE2 | 1732,09 | -0,03 | 0,02 | -1,99 | 4,62E-02 | 1,53E-01 |
| DNAJB12 | 1377,41 | -0,05 | 0,03 | -1,99 | 4,62E-02 | 1,53E-01 |
| SNRPG | 419,44 | 0,10 | 0,05 | 1,99 | 4,63E-02 | 1,53E-01 |
| PTCD3 | 2608,48 | 0,04 | 0,02 | 1,99 | 4,63E-02 | 1,53E-01 |
| ZNF396 | 156,28 | -0,08 | 0,04 | -1,99 | 4,64E-02 | 1,53E-01 |
| FHIP1B | 809,34 | -0,08 | 0,04 | -1,99 | 4,64E-02 | 1,53E-01 |
| CYBRD1 | 1355,66 | -0,08 | 0,04 | -1,99 | 4,64E-02 | 1,54E-01 |
| KAT7 | 3006,94 | -0,04 | 0,02 | -1,99 | 4,65E-02 | 1,54E-01 |
| DPH7 | 542,40 | 0,04 | 0,02 | 1,99 | 4,65E-02 | 1,54E-01 |
| SPIDR | 1966,12 | -0,03 | 0,02 | -1,99 | 4,65E-02 | 1,54E-01 |
| PARP16 | 218,80 | -0,07 | 0,03 | -1,99 | 4,65E-02 | 1,54E-01 |
| AGPS | 1917,64 | 0,06 | 0,03 | 1,99 | 4,65E-02 | 1,54E-01 |
| MRPL22 | 355,34 | 0,05 | 0,03 | 1,99 | 4,66E-02 | 1,54E-01 |
| NF1 | 3196,63 | -0,05 | 0,02 | -1,99 | 4,66E-02 | 1,54E-01 |
| PSMG1 | 274,37 | 0,05 | 0,02 | 1,99 | 4,66E-02 | 1,54E-01 |
| VPS35 | 3425,07 | -0,04 | 0,02 | -1,99 | 4,66E-02 | 1,54E-01 |
| LIPN | 513,89 | -0,12 | 0,06 | -1,99 | 4,67E-02 | 1,54E-01 |
| PCYOX1L | 484,97 | 0,05 | 0,03 | 1,99 | 4,67E-02 | 1,54E-01 |
| TRIM69 | 1732,80 | -0,06 | 0,03 | -1,99 | 4,67E-02 | 1,54E-01 |
| TRAPPC8 | 5204,45 | -0,04 | 0,02 | -1,99 | 4,68E-02 | 1,54E-01 |
| CD6 | 1434,19 | 0,08 | 0,04 | 1,99 | 4,68E-02 | 1,54E-01 |
| ATG13 | 1239,19 | -0,05 | 0,03 | -1,99 | 4,69E-02 | 1,54E-01 |

| | | | | | | |
|---|---|---|---|---|---|---|
| SYF2 | 1675,03 | -0,08 | 0,04 | -1,99 | 4,69E-02 | 1,54E-01 |
| CCSER1 | 256,93 | 0,09 | 0,05 | 1,99 | 4,70E-02 | 1,54E-01 |
| PRKCZ | 560,47 | -0,06 | 0,03 | -1,99 | 4,71E-02 | 1,55E-01 |
| NDUFAF6 | 393,24 | 0,04 | 0,02 | 1,99 | 4,71E-02 | 1,55E-01 |
| C16orf74 | 137,73 | 0,08 | 0,04 | 1,99 | 4,71E-02 | 1,55E-01 |
| SELENOM | 113,94 | 0,08 | 0,04 | 1,99 | 4,71E-02 | 1,55E-01 |
| PAK1IP1 | 167,69 | 0,07 | 0,03 | 1,99 | 4,71E-02 | 1,55E-01 |
| SCYL3 | 1536,50 | -0,04 | 0,02 | -1,98 | 4,72E-02 | 1,55E-01 |
| ZNF548 | 869,47 | 0,05 | 0,02 | 1,98 | 4,72E-02 | 1,55E-01 |
| WWC3 | 2021,61 | -0,08 | 0,04 | -1,98 | 4,72E-02 | 1,55E-01 |
| LHFPL2 | 567,22 | -0,09 | 0,05 | -1,98 | 4,74E-02 | 1,55E-01 |
| WDR18 | 164,52 | 0,06 | 0,03 | 1,98 | 4,74E-02 | 1,55E-01 |
| SLC2A11 | 183,68 | 0,08 | 0,04 | 1,98 | 4,74E-02 | 1,56E-01 |
| DNAJC5 | 3113,93 | -0,07 | 0,04 | -1,98 | 4,75E-02 | 1,56E-01 |
| ZNF468 | 669,75 | 0,07 | 0,04 | 1,98 | 4,76E-02 | 1,56E-01 |
| AOAH | 7471,42 | -0,07 | 0,03 | -1,98 | 4,76E-02 | 1,56E-01 |
| AOC3 | 99,75 | -0,16 | 0,08 | -1,98 | 4,77E-02 | 1,56E-01 |
| ATP5MC3 | 1186,15 | 0,06 | 0,03 | 1,98 | 4,77E-02 | 1,56E-01 |
| TRAPPC5 | 589,24 | -0,05 | 0,03 | -1,98 | 4,78E-02 | 1,56E-01 |
| THEM6 | 236,68 | 0,05 | 0,03 | 1,98 | 4,78E-02 | 1,56E-01 |
| EFCAB2 | 216,32 | -0,08 | 0,04 | -1,98 | 4,79E-02 | 1,57E-01 |
| ODC1 | 5336,43 | 0,11 | 0,05 | 1,98 | 4,79E-02 | 1,57E-01 |
| NUP35 | 452,98 | 0,05 | 0,03 | 1,98 | 4,79E-02 | 1,57E-01 |
| DDB1 | 4047,91 | 0,06 | 0,03 | 1,98 | 4,79E-02 | 1,57E-01 |
| ZNF527 | 322,21 | 0,06 | 0,03 | 1,98 | 4,80E-02 | 1,57E-01 |
| ZFP14 | 845,03 | 0,06 | 0,03 | 1,98 | 4,80E-02 | 1,57E-01 |
| MSANTD4 | 794,25 | 0,07 | 0,04 | 1,98 | 4,80E-02 | 1,57E-01 |
| MRS2 | 622,90 | 0,04 | 0,02 | 1,98 | 4,81E-02 | 1,57E-01 |
| ITPK1 | 1980,83 | -0,10 | 0,05 | -1,98 | 4,81E-02 | 1,57E-01 |
| DCP1B | 382,25 | 0,05 | 0,02 | 1,98 | 4,81E-02 | 1,57E-01 |
| SRP54 | 1562,65 | -0,05 | 0,02 | -1,98 | 4,82E-02 | 1,57E-01 |
| MICU3 | 230,23 | 0,11 | 0,06 | 1,98 | 4,82E-02 | 1,57E-01 |
| ASB13 | 261,30 | 0,05 | 0,02 | 1,98 | 4,82E-02 | 1,57E-01 |
| PRR13 | 644,20 | -0,08 | 0,04 | -1,98 | 4,82E-02 | 1,57E-01 |
| CAMK4 | 4275,49 | 0,08 | 0,04 | 1,98 | 4,83E-02 | 1,57E-01 |
| ZNF614 | 381,88 | 0,07 | 0,04 | 1,97 | 4,83E-02 | 1,57E-01 |
| INPP4A | 7431,99 | -0,05 | 0,02 | -1,97 | 4,84E-02 | 1,57E-01 |
| IQSEC1 | 5784,73 | -0,08 | 0,04 | -1,97 | 4,83E-02 | 1,57E-01 |
| VPS13A | 4001,34 | 0,06 | 0,03 | 1,97 | 4,83E-02 | 1,57E-01 |
| CGAS | 782,02 | -0,04 | 0,02 | -1,97 | 4,84E-02 | 1,57E-01 |
| SLC15A3 | 1355,15 | -0,07 | 0,03 | -1,97 | 4,84E-02 | 1,58E-01 |
| PTTG1IP | 4998,34 | -0,05 | 0,02 | -1,97 | 4,85E-02 | 1,58E-01 |
| ZFP91 | 1463,88 | -0,03 | 0,02 | -1,97 | 4,85E-02 | 1,58E-01 |
| IDH3A | 805,64 | 0,03 | 0,02 | 1,97 | 4,85E-02 | 1,58E-01 |
| LILRA6 | 739,08 | -0,12 | 0,06 | -1,97 | 4,86E-02 | 1,58E-01 |
| KPNA4 | 4365,45 | -0,04 | 0,02 | -1,97 | 4,86E-02 | 1,58E-01 |

| | | | | | | |
|---|---|---|---|---|---|---|
| TGDS | 491,26 | 0,07 | 0,03 | 1,97 | 4,86E-02 | 1,58E-01 |
| GDAP1 | 276,10 | 0,05 | 0,03 | 1,97 | 4,86E-02 | 1,58E-01 |
| UGDH | 329,02 | 0,05 | 0,02 | 1,97 | 4,87E-02 | 1,58E-01 |
| GPR108 | 711,02 | -0,05 | 0,02 | -1,97 | 4,87E-02 | 1,58E-01 |
| KLF5 | 104,65 | -0,09 | 0,05 | -1,97 | 4,88E-02 | 1,58E-01 |
| RNF10 | 10221,41 | 0,13 | 0,07 | 1,97 | 4,88E-02 | 1,58E-01 |
| CD38 | 704,54 | -0,08 | 0,04 | -1,97 | 4,90E-02 | 1,59E-01 |
| HECTD3 | 614,32 | 0,06 | 0,03 | 1,97 | 4,90E-02 | 1,59E-01 |
| CEP68 | 683,78 | 0,05 | 0,03 | 1,97 | 4,90E-02 | 1,59E-01 |
| TNNT1 | 74,64 | 0,19 | 0,10 | 1,97 | 4,90E-02 | 1,59E-01 |
| RALBP1 | 3341,25 | -0,06 | 0,03 | -1,97 | 4,91E-02 | 1,59E-01 |
| UFSP1 | 114,88 | 0,06 | 0,03 | 1,97 | 4,91E-02 | 1,59E-01 |
| GPR18 | 207,24 | 0,09 | 0,05 | 1,97 | 4,91E-02 | 1,59E-01 |
| TMEM243 | 1311,15 | 0,05 | 0,03 | 1,97 | 4,91E-02 | 1,59E-01 |
| SAR1A | 3686,67 | -0,04 | 0,02 | -1,97 | 4,93E-02 | 1,59E-01 |
| TINF2 | 1992,24 | -0,03 | 0,02 | -1,97 | 4,93E-02 | 1,59E-01 |
| COMMD10 | 819,12 | 0,05 | 0,03 | 1,97 | 4,93E-02 | 1,59E-01 |
| UPF3A | 1010,06 | 0,04 | 0,02 | 1,97 | 4,93E-02 | 1,59E-01 |
| CEP295NL | 422,55 | -0,14 | 0,07 | -1,97 | 4,94E-02 | 1,59E-01 |
| ZBTB26 | 440,63 | 0,05 | 0,03 | 1,96 | 4,95E-02 | 1,60E-01 |
| DDIT4 | 1184,58 | -0,10 | 0,05 | -1,96 | 4,95E-02 | 1,60E-01 |
| TBC1D19 | 467,92 | 0,07 | 0,04 | 1,96 | 4,96E-02 | 1,60E-01 |
| H1-3 | 3561,40 | 0,08 | 0,04 | 1,96 | 4,96E-02 | 1,60E-01 |
| NOMO3 | 242,86 | -0,10 | 0,05 | -1,96 | 4,97E-02 | 1,60E-01 |
| GALM | 463,90 | -0,06 | 0,03 | -1,96 | 4,97E-02 | 1,60E-01 |
| MAPRE3 | 75,70 | -0,13 | 0,07 | -1,96 | 4,97E-02 | 1,60E-01 |
| VSIG10 | 168,75 | -0,13 | 0,07 | -1,96 | 4,98E-02 | 1,60E-01 |
| LDAH | 545,34 | 0,05 | 0,02 | 1,96 | 4,98E-02 | 1,60E-01 |
| FLOT2 | 2512,82 | -0,10 | 0,05 | -1,96 | 4,99E-02 | 1,60E-01 |
| RAB30 | 786,80 | 0,08 | 0,04 | 1,96 | 4,98E-02 | 1,60E-01 |
| CDKN2C | 135,22 | 0,08 | 0,04 | 1,96 | 5,00E-02 | 1,61E-01 |
| GALNT7 | 1518,65 | -0,04 | 0,02 | -1,96 | 5,00E-02 | 1,61E-01 |
| DEDD | 287,97 | -0,06 | 0,03 | -1,96 | 5,01E-02 | 1,61E-01 |
| IFT52 | 371,76 | 0,05 | 0,03 | 1,96 | 5,01E-02 | 1,61E-01 |
| DPCD | 179,28 | 0,12 | 0,06 | 1,96 | 5,01E-02 | 1,61E-01 |
| OSGIN2 | 1864,36 | -0,09 | 0,04 | -1,96 | 5,01E-02 | 1,61E-01 |
| IKBKE | 1294,13 | 0,05 | 0,02 | 1,96 | 5,02E-02 | 1,61E-01 |
| GPATCH2 | 965,76 | 0,06 | 0,03 | 1,96 | 5,02E-02 | 1,61E-01 |
| AARS2 | 597,20 | 0,04 | 0,02 | 1,96 | 5,02E-02 | 1,61E-01 |
| BAG4 | 861,51 | 0,06 | 0,03 | 1,96 | 5,02E-02 | 1,61E-01 |
| AAAS | 150,19 | 0,06 | 0,03 | 1,96 | 5,02E-02 | 1,61E-01 |
| ZRANB3 | 214,36 | 0,06 | 0,03 | 1,96 | 5,02E-02 | 1,61E-01 |
| SDHC | 1703,61 | -0,03 | 0,02 | -1,96 | 5,03E-02 | 1,61E-01 |
| TASL | 932,93 | -0,08 | 0,04 | -1,96 | 5,03E-02 | 1,61E-01 |
| UFM1 | 1837,77 | 0,10 | 0,05 | 1,96 | 5,03E-02 | 1,61E-01 |
| ZSWIM6 | 2278,50 | -0,05 | 0,02 | -1,96 | 5,04E-02 | 1,61E-01 |

| | | | | | | |
|---|---|---|---|---|---|---|
| RNF13 | 5972,85 | -0,07 | 0,03 | -1,96 | 5,04E-02 | 1,62E-01 |
| CCR1 | 1618,55 | -0,10 | 0,05 | -1,96 | 5,05E-02 | 1,62E-01 |
| LMBRD2 | 1673,77 | 0,07 | 0,04 | 1,96 | 5,05E-02 | 1,62E-01 |
| GMFG | 5017,22 | -0,09 | 0,04 | -1,96 | 5,05E-02 | 1,62E-01 |
| KCTD12 | 3571,14 | 0,07 | 0,04 | 1,95 | 5,06E-02 | 1,62E-01 |
| FUCA2 | 509,46 | 0,04 | 0,02 | 1,96 | 5,06E-02 | 1,62E-01 |
| ZSWIM8 | 1744,09 | -0,08 | 0,04 | -1,95 | 5,06E-02 | 1,62E-01 |
| CNN2 | 4426,56 | -0,10 | 0,05 | -1,95 | 5,06E-02 | 1,62E-01 |
| ACTR3 | 17781,38 | -0,06 | 0,03 | -1,95 | 5,06E-02 | 1,62E-01 |
| ORC3 | 631,29 | 0,05 | 0,03 | 1,95 | 5,06E-02 | 1,62E-01 |
| FBXO42 | 1453,38 | -0,03 | 0,02 | -1,95 | 5,06E-02 | 1,62E-01 |
| SHLD3 | 135,81 | 0,10 | 0,05 | 1,95 | 5,07E-02 | 1,62E-01 |
| SKP1 | 3196,89 | 0,04 | 0,02 | 1,95 | 5,07E-02 | 1,62E-01 |
| EFHC2 | 210,75 | -0,07 | 0,04 | -1,95 | 5,07E-02 | 1,62E-01 |
| METTL15 | 761,22 | 0,05 | 0,03 | 1,95 | 5,07E-02 | 1,62E-01 |
| CCDC66 | 1235,96 | 0,06 | 0,03 | 1,95 | 5,07E-02 | 1,62E-01 |
| ZNF697 | 325,14 | -0,07 | 0,04 | -1,95 | 5,08E-02 | 1,62E-01 |
| LFNG | 3171,05 | 0,05 | 0,03 | 1,95 | 5,09E-02 | 1,62E-01 |
| TXNDC15 | 973,10 | 0,03 | 0,02 | 1,95 | 5,09E-02 | 1,62E-01 |
| GRPEL1 | 710,32 | 0,04 | 0,02 | 1,95 | 5,10E-02 | 1,62E-01 |
| RAB37 | 1795,64 | -0,05 | 0,03 | -1,95 | 5,10E-02 | 1,62E-01 |
| MRPL21 | 218,64 | 0,05 | 0,03 | 1,95 | 5,11E-02 | 1,63E-01 |
| RNF4 | 2246,26 | -0,03 | 0,02 | -1,95 | 5,11E-02 | 1,63E-01 |
| SRP9 | 1933,22 | 0,10 | 0,05 | 1,95 | 5,12E-02 | 1,63E-01 |
| OLIG1 | 142,87 | 0,11 | 0,06 | 1,95 | 5,12E-02 | 1,63E-01 |
| RBM41 | 789,77 | 0,05 | 0,03 | 1,95 | 5,12E-02 | 1,63E-01 |
| SMIM15 | 727,50 | 0,08 | 0,04 | 1,95 | 5,13E-02 | 1,63E-01 |
| SLC39A1 | 114,63 | 0,05 | 0,03 | 1,95 | 5,14E-02 | 1,63E-01 |
| PDCD2 | 1644,15 | 0,05 | 0,02 | 1,95 | 5,14E-02 | 1,63E-01 |
| JOSD1 | 1205,55 | -0,03 | 0,02 | -1,95 | 5,14E-02 | 1,63E-01 |
| FUT10 | 280,49 | -0,11 | 0,06 | -1,95 | 5,14E-02 | 1,63E-01 |
| LARP4B | 4094,38 | -0,05 | 0,02 | -1,95 | 5,14E-02 | 1,63E-01 |
| INTS14 | 693,00 | 0,03 | 0,02 | 1,95 | 5,15E-02 | 1,63E-01 |
| SLC25A42 | 224,44 | 0,08 | 0,04 | 1,95 | 5,15E-02 | 1,63E-01 |
| FCMR | 1262,94 | 0,08 | 0,04 | 1,95 | 5,15E-02 | 1,63E-01 |
| LSM8 | 1425,93 | 0,07 | 0,03 | 1,95 | 5,16E-02 | 1,63E-01 |
| PITPNB | 874,19 | -0,04 | 0,02 | -1,95 | 5,16E-02 | 1,64E-01 |
| PRH1 | 369,42 | 0,07 | 0,03 | 1,95 | 5,17E-02 | 1,64E-01 |
| ALDH5A1 | 732,34 | 0,10 | 0,05 | 1,95 | 5,17E-02 | 1,64E-01 |
| DENND1B | 2099,25 | 0,07 | 0,04 | 1,95 | 5,17E-02 | 1,64E-01 |
| CACNB4 | 675,79 | -0,12 | 0,06 | -1,95 | 5,18E-02 | 1,64E-01 |
| ZNF567 | 330,79 | 0,07 | 0,04 | 1,94 | 5,18E-02 | 1,64E-01 |
| NDUFS7 | 373,63 | 0,05 | 0,03 | 1,94 | 5,18E-02 | 1,64E-01 |
| TRMT10A | 161,87 | 0,09 | 0,05 | 1,94 | 5,19E-02 | 1,64E-01 |
| MRPL1 | 226,57 | 0,09 | 0,05 | 1,94 | 5,19E-02 | 1,64E-01 |
| HIPK2 | 1634,72 | -0,09 | 0,05 | -1,94 | 5,19E-02 | 1,64E-01 |

| | | | | | | |
|---|---|---|---|---|---|---|
| ENO3 | 130,54 | 0,05 | 0,03 | 1,94 | 5,20E-02 | 1,64E-01 |
| VPS18 | 1062,07 | -0,04 | 0,02 | -1,94 | 5,20E-02 | 1,64E-01 |
| MBOAT7 | 1871,21 | -0,12 | 0,06 | -1,94 | 5,20E-02 | 1,64E-01 |
| C12orf75 | 446,99 | 0,08 | 0,04 | 1,94 | 5,21E-02 | 1,64E-01 |
| ARID3B | 599,54 | -0,09 | 0,05 | -1,94 | 5,21E-02 | 1,64E-01 |
| NCSTN | 1119,80 | -0,07 | 0,03 | -1,94 | 5,21E-02 | 1,65E-01 |
| XYLT2 | 409,91 | 0,05 | 0,03 | 1,94 | 5,22E-02 | 1,65E-01 |
| HNRNPA0 | 1464,94 | 0,05 | 0,02 | 1,94 | 5,22E-02 | 1,65E-01 |
| ALG5 | 384,27 | 0,06 | 0,03 | 1,94 | 5,22E-02 | 1,65E-01 |
| TANGO2 | 2382,23 | 0,09 | 0,05 | 1,94 | 5,22E-02 | 1,65E-01 |
| TMPPE | 685,66 | -0,07 | 0,04 | -1,94 | 5,22E-02 | 1,65E-01 |
| CFP | 4324,54 | -0,07 | 0,03 | -1,94 | 5,22E-02 | 1,65E-01 |
| RMDN1 | 1323,80 | 0,05 | 0,02 | 1,94 | 5,23E-02 | 1,65E-01 |
| OTUD6B | 736,74 | 0,08 | 0,04 | 1,94 | 5,23E-02 | 1,65E-01 |
| SART1 | 1462,15 | -0,06 | 0,03 | -1,94 | 5,23E-02 | 1,65E-01 |
| RWDD3 | 250,72 | 0,06 | 0,03 | 1,94 | 5,23E-02 | 1,65E-01 |
| DYRK3 | 136,30 | 0,13 | 0,07 | 1,94 | 5,23E-02 | 1,65E-01 |
| SLC12A2 | 775,39 | 0,05 | 0,03 | 1,94 | 5,24E-02 | 1,65E-01 |
| TBCB | 949,68 | 0,04 | 0,02 | 1,94 | 5,25E-02 | 1,65E-01 |
| TMEM87B | 1254,96 | 0,04 | 0,02 | 1,94 | 5,25E-02 | 1,65E-01 |
| CCL28 | 176,63 | 0,05 | 0,03 | 1,94 | 5,25E-02 | 1,65E-01 |
| LACTB | 1215,80 | -0,06 | 0,03 | -1,94 | 5,26E-02 | 1,65E-01 |
| ZDHHC20 | 4848,55 | 0,07 | 0,03 | 1,94 | 5,27E-02 | 1,66E-01 |
| PDGFC | 183,63 | 0,08 | 0,04 | 1,94 | 5,28E-02 | 1,66E-01 |
| DDX59 | 840,31 | -0,04 | 0,02 | -1,94 | 5,29E-02 | 1,66E-01 |
| TSC1 | 2023,56 | -0,04 | 0,02 | -1,93 | 5,30E-02 | 1,66E-01 |
| CYP51A1 | 86,14 | 0,06 | 0,03 | 1,93 | 5,30E-02 | 1,66E-01 |
| SCAI | 729,80 | 0,05 | 0,02 | 1,93 | 5,30E-02 | 1,66E-01 |
| DNAJC13 | 3968,81 | -0,04 | 0,02 | -1,94 | 5,30E-02 | 1,66E-01 |
| RFC3 | 300,61 | 0,06 | 0,03 | 1,93 | 5,30E-02 | 1,66E-01 |
| CEP44 | 998,12 | 0,06 | 0,03 | 1,94 | 5,30E-02 | 1,66E-01 |
| OTULIN | 4055,63 | -0,04 | 0,02 | -1,94 | 5,30E-02 | 1,66E-01 |
| LEF1 | 3221,77 | 0,08 | 0,04 | 1,93 | 5,30E-02 | 1,66E-01 |
| CDCA7L | 654,79 | 0,08 | 0,04 | 1,93 | 5,31E-02 | 1,66E-01 |
| ZNF573 | 404,43 | 0,05 | 0,02 | 1,93 | 5,31E-02 | 1,66E-01 |
| ZNF629 | 153,43 | 0,07 | 0,03 | 1,93 | 5,32E-02 | 1,66E-01 |
| RFWD3 | 920,15 | -0,03 | 0,02 | -1,93 | 5,32E-02 | 1,66E-01 |
| TP53I11 | 396,09 | -0,13 | 0,07 | -1,93 | 5,33E-02 | 1,67E-01 |
| CCDC138 | 215,24 | 0,06 | 0,03 | 1,93 | 5,33E-02 | 1,67E-01 |
| RTCA | 1065,30 | 0,03 | 0,01 | 1,93 | 5,33E-02 | 1,67E-01 |
| PHLDA1 | 156,82 | 0,07 | 0,04 | 1,93 | 5,34E-02 | 1,67E-01 |
| GTF2IRD2B | 450,51 | 0,04 | 0,02 | 1,93 | 5,35E-02 | 1,67E-01 |
| THOC6 | 329,85 | 0,04 | 0,02 | 1,93 | 5,36E-02 | 1,67E-01 |
| PMS1 | 801,94 | 0,06 | 0,03 | 1,93 | 5,36E-02 | 1,67E-01 |
| TUBGCP3 | 2267,00 | -0,04 | 0,02 | -1,93 | 5,36E-02 | 1,67E-01 |
| MRPS35 | 553,15 | 0,06 | 0,03 | 1,93 | 5,35E-02 | 1,67E-01 |

| | | | | | | |
|---|---|---|---|---|---|---|
| P2RY13 | 1553,79 | -0,08 | 0,04 | -1,93 | 5,35E-02 | 1,67E-01 |
| CAPN10 | 1042,56 | 0,04 | 0,02 | 1,93 | 5,35E-02 | 1,67E-01 |
| ABCA13 | 417,60 | 0,20 | 0,10 | 1,93 | 5,36E-02 | 1,67E-01 |
| EEF1D | 7255,63 | 0,06 | 0,03 | 1,93 | 5,36E-02 | 1,67E-01 |
| SEC22C | 1303,99 | 0,03 | 0,02 | 1,93 | 5,37E-02 | 1,67E-01 |
| MGAT4B | 841,26 | 0,06 | 0,03 | 1,93 | 5,38E-02 | 1,68E-01 |
| R3HDM4 | 9507,57 | 0,12 | 0,06 | 1,93 | 5,38E-02 | 1,68E-01 |
| GON7 | 178,86 | 0,09 | 0,05 | 1,93 | 5,38E-02 | 1,68E-01 |
| RRM2B | 3063,32 | -0,08 | 0,04 | -1,93 | 5,38E-02 | 1,68E-01 |
| ZNF257 | 229,86 | 0,10 | 0,05 | 1,93 | 5,38E-02 | 1,68E-01 |
| MARCHF3 | 294,30 | 0,07 | 0,04 | 1,93 | 5,39E-02 | 1,68E-01 |
| DHX58 | 637,05 | -0,10 | 0,05 | -1,93 | 5,39E-02 | 1,68E-01 |
| TMEM184B | 510,43 | -0,11 | 0,06 | -1,93 | 5,39E-02 | 1,68E-01 |
| GSTP1 | 1557,64 | 0,05 | 0,03 | 1,93 | 5,39E-02 | 1,68E-01 |
| TIGD6 | 140,86 | 0,06 | 0,03 | 1,93 | 5,40E-02 | 1,68E-01 |
| PLEKHO2 | 1028,93 | -0,10 | 0,05 | -1,93 | 5,39E-02 | 1,68E-01 |
| SPOPL | 6754,41 | -0,08 | 0,04 | -1,93 | 5,40E-02 | 1,68E-01 |
| IMPACT | 536,16 | 0,05 | 0,03 | 1,93 | 5,40E-02 | 1,68E-01 |
| NHP2 | 525,58 | 0,05 | 0,03 | 1,93 | 5,41E-02 | 1,68E-01 |
| WDR81 | 979,81 | 0,06 | 0,03 | 1,93 | 5,42E-02 | 1,68E-01 |
| PIKFYVE | 5566,70 | -0,04 | 0,02 | -1,92 | 5,42E-02 | 1,68E-01 |
| KAZN | 232,39 | -0,17 | 0,09 | -1,92 | 5,43E-02 | 1,68E-01 |
| ZNF787 | 927,35 | -0,05 | 0,03 | -1,92 | 5,43E-02 | 1,68E-01 |
| ZBTB45 | 266,12 | 0,04 | 0,02 | 1,92 | 5,43E-02 | 1,68E-01 |
| ZNF28 | 577,19 | 0,07 | 0,04 | 1,92 | 5,43E-02 | 1,68E-01 |
| UHRF2 | 3575,62 | 0,04 | 0,02 | 1,92 | 5,44E-02 | 1,68E-01 |
| HOOK1 | 297,99 | 0,10 | 0,05 | 1,92 | 5,44E-02 | 1,69E-01 |
| LAIR1 | 1670,71 | 0,05 | 0,02 | 1,92 | 5,44E-02 | 1,69E-01 |
| ARSB | 874,65 | -0,06 | 0,03 | -1,92 | 5,45E-02 | 1,69E-01 |
| HAPLN3 | 583,48 | 0,07 | 0,04 | 1,92 | 5,46E-02 | 1,69E-01 |
| PGRMC2 | 507,71 | 0,03 | 0,02 | 1,92 | 5,47E-02 | 1,69E-01 |
| SLC11A1 | 6756,68 | -0,10 | 0,05 | -1,92 | 5,47E-02 | 1,69E-01 |
| FAM76A | 467,11 | 0,03 | 0,02 | 1,92 | 5,48E-02 | 1,70E-01 |
| LRRC37B | 716,96 | -0,05 | 0,03 | -1,92 | 5,49E-02 | 1,70E-01 |
| TIMM21 | 343,46 | 0,05 | 0,02 | 1,92 | 5,50E-02 | 1,70E-01 |
| TTC38 | 939,29 | -0,07 | 0,04 | -1,92 | 5,50E-02 | 1,70E-01 |
| AIMP2 | 165,31 | 0,04 | 0,02 | 1,92 | 5,51E-02 | 1,70E-01 |
| DUSP7 | 630,10 | 0,05 | 0,02 | 1,92 | 5,51E-02 | 1,70E-01 |
| DALRD3 | 394,96 | 0,04 | 0,02 | 1,92 | 5,52E-02 | 1,70E-01 |
| C11orf58 | 4589,91 | 0,06 | 0,03 | 1,92 | 5,51E-02 | 1,70E-01 |
| POT1 | 695,07 | 0,08 | 0,04 | 1,92 | 5,52E-02 | 1,70E-01 |
| UBA1 | 1932,81 | -0,09 | 0,04 | -1,92 | 5,52E-02 | 1,70E-01 |
| HAUS1 | 258,55 | 0,07 | 0,04 | 1,92 | 5,52E-02 | 1,70E-01 |
| MMP25 | 4065,29 | -0,14 | 0,07 | -1,92 | 5,52E-02 | 1,70E-01 |
| SRSF4 | 4151,13 | -0,03 | 0,02 | -1,92 | 5,53E-02 | 1,70E-01 |
| NCEH1 | 270,02 | 0,04 | 0,02 | 1,92 | 5,53E-02 | 1,70E-01 |

| | | | | | | |
|---|---|---|---|---|---|---|
| KIF15 | 113,23 | 0,14 | 0,07 | 1,92 | 5,53E-02 | 1,70E-01 |
| HACL1 | 838,66 | 0,04 | 0,02 | 1,92 | 5,53E-02 | 1,70E-01 |
| TRAP1 | 596,68 | 0,05 | 0,03 | 1,92 | 5,54E-02 | 1,70E-01 |
| ANKRD11 | 4497,54 | -0,08 | 0,04 | -1,92 | 5,54E-02 | 1,70E-01 |
| MGST3 | 889,70 | 0,04 | 0,02 | 1,92 | 5,53E-02 | 1,70E-01 |
| IFT57 | 477,56 | 0,09 | 0,05 | 1,92 | 5,54E-02 | 1,70E-01 |
| RARA | 1441,48 | -0,10 | 0,05 | -1,92 | 5,54E-02 | 1,70E-01 |
| ERI1 | 747,08 | -0,05 | 0,03 | -1,92 | 5,55E-02 | 1,70E-01 |
| STK32C | 352,32 | 0,05 | 0,03 | 1,92 | 5,55E-02 | 1,70E-01 |
| GSR | 2527,68 | -0,04 | 0,02 | -1,92 | 5,55E-02 | 1,70E-01 |
| BAD | 353,61 | 0,05 | 0,02 | 1,92 | 5,54E-02 | 1,70E-01 |
| RBCK1 | 1313,63 | -0,08 | 0,04 | -1,92 | 5,54E-02 | 1,70E-01 |
| B3GNT8 | 489,07 | -0,08 | 0,04 | -1,91 | 5,55E-02 | 1,70E-01 |
| FYB1 | 17220,77 | -0,06 | 0,03 | -1,91 | 5,56E-02 | 1,71E-01 |
| ATXN10 | 893,25 | 0,04 | 0,02 | 1,91 | 5,56E-02 | 1,71E-01 |
| PA2G4 | 811,01 | 0,06 | 0,03 | 1,91 | 5,56E-02 | 1,71E-01 |
| SH2D1A | 948,20 | 0,10 | 0,05 | 1,91 | 5,56E-02 | 1,71E-01 |
| KBTBD4 | 721,25 | -0,04 | 0,02 | -1,91 | 5,57E-02 | 1,71E-01 |
| TCHP | 1162,14 | -0,03 | 0,02 | -1,91 | 5,57E-02 | 1,71E-01 |
| MAP3K3 | 3149,93 | -0,08 | 0,04 | -1,91 | 5,58E-02 | 1,71E-01 |
| TEFM | 428,25 | 0,05 | 0,02 | 1,91 | 5,59E-02 | 1,71E-01 |
| MBD1 | 1555,61 | -0,03 | 0,02 | -1,91 | 5,59E-02 | 1,71E-01 |
| JAG1 | 255,43 | -0,08 | 0,04 | -1,91 | 5,60E-02 | 1,71E-01 |
| TP53 | 481,01 | 0,06 | 0,03 | 1,91 | 5,60E-02 | 1,71E-01 |
| CNOT8 | 3179,45 | -0,04 | 0,02 | -1,91 | 5,60E-02 | 1,71E-01 |
| ANAPC11 | 464,37 | 0,05 | 0,02 | 1,91 | 5,61E-02 | 1,72E-01 |
| SMARCA2 | 7811,56 | -0,04 | 0,02 | -1,91 | 5,61E-02 | 1,72E-01 |
| SH2D1B | 481,02 | -0,10 | 0,05 | -1,91 | 5,62E-02 | 1,72E-01 |
| CHTOP | 3106,91 | -0,03 | 0,01 | -1,91 | 5,62E-02 | 1,72E-01 |
| NHLRC3 | 1216,85 | 0,04 | 0,02 | 1,91 | 5,63E-02 | 1,72E-01 |
| TNKS2 | 5449,74 | -0,03 | 0,02 | -1,91 | 5,63E-02 | 1,72E-01 |
| MRPL48 | 215,86 | 0,06 | 0,03 | 1,91 | 5,64E-02 | 1,72E-01 |
| ELOVL1 | 877,28 | -0,03 | 0,02 | -1,91 | 5,64E-02 | 1,72E-01 |
| ZMAT3 | 1503,77 | 0,05 | 0,03 | 1,91 | 5,64E-02 | 1,72E-01 |
| PCDH9 | 1290,94 | 0,14 | 0,08 | 1,91 | 5,64E-02 | 1,72E-01 |
| DDX60 | 2823,93 | -0,12 | 0,06 | -1,91 | 5,65E-02 | 1,72E-01 |
| NAV3 | 108,08 | -0,17 | 0,09 | -1,91 | 5,65E-02 | 1,72E-01 |
| PEX11B | 420,12 | 0,03 | 0,02 | 1,91 | 5,65E-02 | 1,72E-01 |
| ZBTB14 | 992,19 | 0,06 | 0,03 | 1,91 | 5,66E-02 | 1,72E-01 |
| ATP6V1H | 1245,36 | -0,03 | 0,02 | -1,91 | 5,66E-02 | 1,72E-01 |
| SNRPF | 293,04 | 0,08 | 0,04 | 1,91 | 5,66E-02 | 1,72E-01 |
| MRPL44 | 789,94 | -0,05 | 0,03 | -1,91 | 5,67E-02 | 1,73E-01 |
| LCORL | 1528,37 | 0,09 | 0,05 | 1,91 | 5,67E-02 | 1,73E-01 |
| CENPB | 1464,87 | 0,05 | 0,03 | 1,91 | 5,67E-02 | 1,73E-01 |
| SRSF7 | 3883,87 | 0,05 | 0,03 | 1,91 | 5,68E-02 | 1,73E-01 |
| GLG1 | 5180,43 | -0,05 | 0,03 | -1,91 | 5,68E-02 | 1,73E-01 |

| | | | | | | |
|---|---|---|---|---|---|---|
| UTRN | 22033,29 | -0,05 | 0,03 | -1,90 | 5,68E-02 | 1,73E-01 |
| CLUAP1 | 356,81 | 0,05 | 0,03 | 1,90 | 5,68E-02 | 1,73E-01 |
| KIAA1958 | 269,80 | -0,08 | 0,04 | -1,90 | 5,70E-02 | 1,73E-01 |
| ZNF644 | 2869,41 | 0,08 | 0,04 | 1,90 | 5,72E-02 | 1,74E-01 |
| PIP4K2A | 13513,07 | 0,09 | 0,05 | 1,90 | 5,72E-02 | 1,74E-01 |
| URI1 | 1754,06 | 0,05 | 0,03 | 1,90 | 5,71E-02 | 1,74E-01 |
| EME2 | 601,40 | 0,07 | 0,04 | 1,90 | 5,72E-02 | 1,74E-01 |
| PRKACB | 3518,33 | 0,09 | 0,05 | 1,90 | 5,72E-02 | 1,74E-01 |
| ZNF880 | 234,26 | 0,06 | 0,03 | 1,90 | 5,73E-02 | 1,74E-01 |
| MORC2 | 1111,36 | 0,04 | 0,02 | 1,90 | 5,74E-02 | 1,74E-01 |
| CARNMT1 | 812,36 | 0,07 | 0,04 | 1,90 | 5,74E-02 | 1,74E-01 |
| RNF220 | 1207,52 | 0,03 | 0,02 | 1,90 | 5,75E-02 | 1,74E-01 |
| TOR3A | 681,25 | 0,04 | 0,02 | 1,90 | 5,76E-02 | 1,74E-01 |
| TTC27 | 660,53 | 0,05 | 0,02 | 1,90 | 5,76E-02 | 1,74E-01 |
| CAAP1 | 677,67 | 0,05 | 0,03 | 1,90 | 5,76E-02 | 1,75E-01 |
| ALAS2 | 41155,77 | 0,18 | 0,10 | 1,90 | 5,77E-02 | 1,75E-01 |
| MPND | 223,49 | 0,05 | 0,02 | 1,90 | 5,78E-02 | 1,75E-01 |
| CTLA4 | 252,96 | 0,09 | 0,05 | 1,90 | 5,79E-02 | 1,75E-01 |
| ARID4B | 6636,06 | -0,05 | 0,03 | -1,90 | 5,80E-02 | 1,75E-01 |
| TXLNB | 78,35 | -0,08 | 0,04 | -1,90 | 5,80E-02 | 1,75E-01 |
| CTSS | 51243,53 | -0,07 | 0,04 | -1,90 | 5,81E-02 | 1,76E-01 |
| SEC61G | 642,34 | 0,07 | 0,04 | 1,89 | 5,82E-02 | 1,76E-01 |
| FAM111A | 3797,59 | -0,04 | 0,02 | -1,89 | 5,82E-02 | 1,76E-01 |
| ZNF181 | 435,90 | 0,10 | 0,05 | 1,89 | 5,82E-02 | 1,76E-01 |
| AHCY | 169,55 | 0,07 | 0,04 | 1,89 | 5,82E-02 | 1,76E-01 |
| HSD17B12 | 656,07 | -0,04 | 0,02 | -1,89 | 5,82E-02 | 1,76E-01 |
| ALKBH2 | 159,69 | 0,06 | 0,03 | 1,89 | 5,81E-02 | 1,76E-01 |
| TSPYL4 | 462,07 | 0,03 | 0,02 | 1,89 | 5,82E-02 | 1,76E-01 |
| WRNIP1 | 1349,53 | 0,02 | 0,01 | 1,89 | 5,81E-02 | 1,76E-01 |
| YIPF4 | 6015,23 | -0,04 | 0,02 | -1,89 | 5,82E-02 | 1,76E-01 |
| POGZ | 2015,47 | -0,05 | 0,02 | -1,89 | 5,83E-02 | 1,76E-01 |
| SF3B2 | 1435,61 | -0,06 | 0,03 | -1,89 | 5,83E-02 | 1,76E-01 |
| MTRES1 | 201,77 | 0,05 | 0,03 | 1,89 | 5,83E-02 | 1,76E-01 |
| DHX38 | 2148,77 | -0,04 | 0,02 | -1,89 | 5,83E-02 | 1,76E-01 |
| FAM3A | 262,93 | 0,05 | 0,03 | 1,89 | 5,84E-02 | 1,76E-01 |
| CYBA | 9671,70 | -0,05 | 0,03 | -1,89 | 5,85E-02 | 1,76E-01 |
| CYSTM1 | 524,16 | -0,09 | 0,05 | -1,89 | 5,85E-02 | 1,76E-01 |
| LYSMD4 | 474,52 | 0,05 | 0,02 | 1,89 | 5,86E-02 | 1,76E-01 |
| CDC14A | 2554,23 | 0,05 | 0,03 | 1,89 | 5,86E-02 | 1,76E-01 |
| SF3A1 | 1297,76 | -0,07 | 0,04 | -1,89 | 5,87E-02 | 1,77E-01 |
| ZNF700 | 887,88 | -0,04 | 0,02 | -1,89 | 5,89E-02 | 1,77E-01 |
| RPUSD2 | 185,77 | 0,05 | 0,03 | 1,89 | 5,90E-02 | 1,77E-01 |
| OAZ1 | 43635,21 | 0,09 | 0,05 | 1,89 | 5,90E-02 | 1,77E-01 |
| SLC38A9 | 1068,83 | 0,04 | 0,02 | 1,89 | 5,90E-02 | 1,77E-01 |
| DYNLT3 | 404,16 | 0,12 | 0,06 | 1,89 | 5,91E-02 | 1,77E-01 |
| UNC119 | 1544,74 | -0,05 | 0,03 | -1,89 | 5,90E-02 | 1,77E-01 |

| | | | | | | |
|---|---|---|---|---|---|---|
| FBXO11 | 3434,39 | -0,05 | 0,02 | -1,89 | 5,91E-02 | 1,77E-01 |
| MCU | 808,66 | -0,04 | 0,02 | -1,89 | 5,91E-02 | 1,77E-01 |
| ANKRD54 | 209,65 | 0,05 | 0,03 | 1,89 | 5,90E-02 | 1,77E-01 |
| CYP20A1 | 907,57 | 0,03 | 0,02 | 1,89 | 5,91E-02 | 1,77E-01 |
| ZNF749 | 164,90 | 0,06 | 0,03 | 1,89 | 5,93E-02 | 1,78E-01 |
| PREPL | 1427,84 | 0,04 | 0,02 | 1,89 | 5,94E-02 | 1,78E-01 |
| PKN2 | 5221,65 | -0,07 | 0,04 | -1,88 | 5,95E-02 | 1,78E-01 |
| G2E3 | 1482,48 | 0,05 | 0,03 | 1,88 | 5,95E-02 | 1,78E-01 |
| ANKRD49 | 771,95 | 0,09 | 0,05 | 1,88 | 5,95E-02 | 1,78E-01 |
| BTNL3 | 79,39 | -0,40 | 0,21 | -1,88 | 5,95E-02 | 1,78E-01 |
| TM9SF1 | 131,07 | -0,04 | 0,02 | -1,88 | 5,96E-02 | 1,78E-01 |
| IPO11 | 610,08 | 0,05 | 0,03 | 1,88 | 5,97E-02 | 1,79E-01 |
| LMLN | 163,19 | 0,06 | 0,03 | 1,88 | 5,97E-02 | 1,79E-01 |
| SLC66A3 | 890,19 | 0,05 | 0,03 | 1,88 | 5,98E-02 | 1,79E-01 |
| CEBPG | 1029,45 | 0,06 | 0,03 | 1,88 | 5,98E-02 | 1,79E-01 |
| RIOX2 | 1091,37 | 0,05 | 0,03 | 1,88 | 5,98E-02 | 1,79E-01 |
| COX20 | 762,65 | 0,05 | 0,02 | 1,88 | 5,98E-02 | 1,79E-01 |
| GRIPAP1 | 1022,61 | -0,07 | 0,04 | -1,88 | 5,99E-02 | 1,79E-01 |
| STK40 | 2245,83 | -0,09 | 0,05 | -1,88 | 6,00E-02 | 1,79E-01 |
| UQCC2 | 319,46 | 0,04 | 0,02 | 1,88 | 6,00E-02 | 1,79E-01 |
| NFE2L1 | 1646,65 | -0,04 | 0,02 | -1,88 | 6,01E-02 | 1,79E-01 |
| SERF2 | 21317,23 | 0,09 | 0,05 | 1,88 | 6,01E-02 | 1,79E-01 |
| TXNL4B | 609,47 | -0,03 | 0,02 | -1,88 | 6,01E-02 | 1,79E-01 |
| ANKZF1 | 2138,30 | -0,04 | 0,02 | -1,88 | 6,01E-02 | 1,79E-01 |
| SPPL2A | 2416,77 | -0,03 | 0,02 | -1,88 | 6,01E-02 | 1,79E-01 |
| ALG6 | 1074,56 | 0,05 | 0,02 | 1,88 | 6,01E-02 | 1,79E-01 |
| CSNK1A1 | 3228,39 | -0,03 | 0,02 | -1,88 | 6,00E-02 | 1,79E-01 |
| MCEE | 317,83 | 0,04 | 0,02 | 1,88 | 6,00E-02 | 1,79E-01 |
| SNAI3 | 180,75 | -0,07 | 0,04 | -1,88 | 6,00E-02 | 1,79E-01 |
| PSMD6 | 1961,81 | -0,03 | 0,02 | -1,88 | 6,01E-02 | 1,79E-01 |
| NUDT6 | 120,58 | 0,07 | 0,04 | 1,88 | 6,01E-02 | 1,79E-01 |
| TNFAIP3 | 1049,24 | -0,06 | 0,03 | -1,88 | 6,02E-02 | 1,79E-01 |
| ZNF140 | 669,00 | 0,04 | 0,02 | 1,88 | 6,02E-02 | 1,79E-01 |
| NAPG | 1491,22 | 0,04 | 0,02 | 1,88 | 6,02E-02 | 1,79E-01 |
| SIT1 | 964,43 | 0,07 | 0,03 | 1,88 | 6,02E-02 | 1,79E-01 |
| HIPK3 | 11401,86 | -0,06 | 0,03 | -1,88 | 6,02E-02 | 1,79E-01 |
| TSPAN2 | 1236,28 | -0,08 | 0,04 | -1,88 | 6,02E-02 | 1,79E-01 |
| REEP3 | 837,55 | 0,06 | 0,03 | 1,88 | 6,03E-02 | 1,79E-01 |
| EMC10 | 1053,02 | 0,04 | 0,02 | 1,88 | 6,03E-02 | 1,79E-01 |
| GYPE | 73,57 | 0,11 | 0,06 | 1,88 | 6,04E-02 | 1,79E-01 |
| EIF2AK1 | 3868,45 | 0,10 | 0,05 | 1,88 | 6,04E-02 | 1,79E-01 |
| DPH6 | 445,23 | 0,07 | 0,04 | 1,88 | 6,04E-02 | 1,79E-01 |
| ZCCHC10 | 223,13 | 0,10 | 0,05 | 1,88 | 6,05E-02 | 1,80E-01 |
| TMEM50A | 3658,40 | -0,05 | 0,03 | -1,88 | 6,06E-02 | 1,80E-01 |
| HDAC4 | 2971,20 | -0,07 | 0,04 | -1,88 | 6,06E-02 | 1,80E-01 |
| SMIM10L1 | 435,18 | 0,07 | 0,04 | 1,88 | 6,06E-02 | 1,80E-01 |

| | | | | | | |
|---|---|---|---|---|---|---|
| TMEM102 | 289,83 | 0,04 | 0,02 | 1,88 | 6,08E-02 | 1,80E-01 |
| SLC2A4RG | 180,67 | 0,09 | 0,05 | 1,88 | 6,08E-02 | 1,80E-01 |
| MRPL36 | 156,12 | 0,07 | 0,04 | 1,87 | 6,08E-02 | 1,80E-01 |
| CXCR4 | 6375,36 | -0,05 | 0,03 | -1,87 | 6,09E-02 | 1,80E-01 |
| TWNK | 259,94 | 0,04 | 0,02 | 1,87 | 6,09E-02 | 1,80E-01 |
| HIBADH | 349,59 | 0,04 | 0,02 | 1,87 | 6,10E-02 | 1,80E-01 |
| CIZ1 | 335,57 | 0,09 | 0,05 | 1,87 | 6,10E-02 | 1,80E-01 |
| RPS6KB2 | 839,00 | 0,04 | 0,02 | 1,87 | 6,09E-02 | 1,80E-01 |
| UBE3D | 415,78 | 0,05 | 0,03 | 1,87 | 6,09E-02 | 1,80E-01 |
| CUTA | 331,64 | 0,04 | 0,02 | 1,87 | 6,09E-02 | 1,80E-01 |
| PPP2CB | 654,68 | -0,04 | 0,02 | -1,87 | 6,10E-02 | 1,80E-01 |
| PANK3 | 3665,78 | 0,05 | 0,03 | 1,87 | 6,10E-02 | 1,80E-01 |
| DPEP3 | 125,00 | -0,12 | 0,06 | -1,87 | 6,11E-02 | 1,80E-01 |
| MS4A3 | 173,66 | 0,17 | 0,09 | 1,87 | 6,11E-02 | 1,81E-01 |
| SUMF2 | 1633,87 | 0,04 | 0,02 | 1,87 | 6,12E-02 | 1,81E-01 |
| PSMB8 | 3458,39 | -0,05 | 0,03 | -1,87 | 6,13E-02 | 1,81E-01 |
| MYCT1 | 74,19 | 0,10 | 0,05 | 1,87 | 6,13E-02 | 1,81E-01 |
| MARS2 | 442,72 | 0,04 | 0,02 | 1,87 | 6,14E-02 | 1,81E-01 |
| HIGD1A | 416,33 | 0,10 | 0,05 | 1,87 | 6,14E-02 | 1,81E-01 |
| PGLYRP1 | 103,34 | -0,12 | 0,07 | -1,87 | 6,14E-02 | 1,81E-01 |
| BLOC1S6 | 2464,23 | 0,06 | 0,03 | 1,87 | 6,14E-02 | 1,81E-01 |
| XKR8 | 615,47 | -0,08 | 0,04 | -1,87 | 6,16E-02 | 1,82E-01 |
| FBXL14 | 415,51 | 0,03 | 0,02 | 1,87 | 6,16E-02 | 1,82E-01 |
| MEF2C | 3442,38 | 0,06 | 0,03 | 1,87 | 6,16E-02 | 1,82E-01 |
| ACSS2 | 587,56 | -0,06 | 0,03 | -1,87 | 6,17E-02 | 1,82E-01 |
| DCAF13 | 833,65 | 0,06 | 0,03 | 1,87 | 6,17E-02 | 1,82E-01 |
| NPC1 | 1659,98 | -0,05 | 0,03 | -1,87 | 6,17E-02 | 1,82E-01 |
| GATD1 | 332,28 | 0,08 | 0,04 | 1,87 | 6,17E-02 | 1,82E-01 |
| LONRF3 | 402,78 | -0,05 | 0,02 | -1,87 | 6,18E-02 | 1,82E-01 |
| DYNC1LI2 | 4591,04 | -0,03 | 0,02 | -1,87 | 6,19E-02 | 1,82E-01 |
| SLC30A9 | 1801,34 | 0,06 | 0,03 | 1,87 | 6,19E-02 | 1,82E-01 |
| CFAP70 | 206,04 | -0,07 | 0,04 | -1,87 | 6,19E-02 | 1,82E-01 |
| CCDC107 | 369,62 | 0,05 | 0,03 | 1,87 | 6,20E-02 | 1,82E-01 |
| ASPM | 192,19 | 0,11 | 0,06 | 1,87 | 6,20E-02 | 1,82E-01 |
| NFKB1 | 3382,56 | -0,04 | 0,02 | -1,87 | 6,21E-02 | 1,82E-01 |
| NUBPL | 310,59 | 0,05 | 0,03 | 1,87 | 6,21E-02 | 1,83E-01 |
| LIPT1 | 139,19 | 0,06 | 0,03 | 1,86 | 6,23E-02 | 1,83E-01 |
| TBC1D4 | 1847,51 | 0,07 | 0,04 | 1,86 | 6,23E-02 | 1,83E-01 |
| ZNF525 | 547,74 | 0,11 | 0,06 | 1,86 | 6,24E-02 | 1,83E-01 |
| ZNF816 | 327,33 | 0,09 | 0,05 | 1,86 | 6,25E-02 | 1,83E-01 |
| MAGEF1 | 210,74 | 0,05 | 0,03 | 1,86 | 6,25E-02 | 1,83E-01 |
| TAB3 | 1926,89 | -0,03 | 0,02 | -1,86 | 6,25E-02 | 1,83E-01 |
| ARPC4 | 5137,77 | -0,04 | 0,02 | -1,86 | 6,25E-02 | 1,83E-01 |
| ERGIC3 | 754,35 | 0,05 | 0,03 | 1,86 | 6,26E-02 | 1,83E-01 |
| CDYL | 1375,60 | 0,08 | 0,04 | 1,86 | 6,26E-02 | 1,83E-01 |
| PALS2 | 710,63 | 0,07 | 0,04 | 1,86 | 6,26E-02 | 1,83E-01 |

| | | | | | | |
|---|---|---|---|---|---|---|
| MANEA | 530,80 | 0,09 | 0,05 | 1,86 | 6,27E-02 | 1,84E-01 |
| GPX1 | 7134,13 | 0,11 | 0,06 | 1,86 | 6,27E-02 | 1,84E-01 |
| VPS26B | 1315,50 | -0,04 | 0,02 | -1,86 | 6,27E-02 | 1,84E-01 |
| CACFD1 | 109,57 | 0,07 | 0,04 | 1,86 | 6,28E-02 | 1,84E-01 |
| TYMP | 5719,81 | -0,09 | 0,05 | -1,86 | 6,28E-02 | 1,84E-01 |
| FUBP1 | 5525,88 | 0,05 | 0,03 | 1,86 | 6,28E-02 | 1,84E-01 |
| GDI1 | 896,95 | -0,06 | 0,03 | -1,86 | 6,28E-02 | 1,84E-01 |
| CRYZL1 | 848,97 | 0,03 | 0,02 | 1,86 | 6,28E-02 | 1,84E-01 |
| BET1 | 686,01 | 0,07 | 0,04 | 1,86 | 6,29E-02 | 1,84E-01 |
| RAVER2 | 155,70 | 0,13 | 0,07 | 1,86 | 6,29E-02 | 1,84E-01 |
| ARHGAP5 | 2001,50 | 0,09 | 0,05 | 1,86 | 6,29E-02 | 1,84E-01 |
| IMP3 | 577,10 | 0,05 | 0,03 | 1,86 | 6,30E-02 | 1,84E-01 |
| P2RY12 | 217,83 | 0,10 | 0,05 | 1,86 | 6,30E-02 | 1,84E-01 |
| HSF1 | 347,37 | 0,08 | 0,05 | 1,86 | 6,30E-02 | 1,84E-01 |
| FIG4 | 1488,99 | -0,04 | 0,02 | -1,86 | 6,31E-02 | 1,84E-01 |
| POLDIP3 | 1827,58 | -0,04 | 0,02 | -1,86 | 6,31E-02 | 1,84E-01 |
| COQ6 | 507,07 | 0,03 | 0,02 | 1,86 | 6,31E-02 | 1,84E-01 |
| CHFR | 2635,31 | -0,05 | 0,03 | -1,86 | 6,31E-02 | 1,84E-01 |
| H2AC8 | 1539,57 | 0,08 | 0,04 | 1,86 | 6,31E-02 | 1,84E-01 |
| ZNF232 | 207,69 | 0,04 | 0,02 | 1,86 | 6,32E-02 | 1,84E-01 |
| CENPI | 92,19 | 0,09 | 0,05 | 1,86 | 6,32E-02 | 1,84E-01 |
| DNAJB2 | 2649,64 | 0,10 | 0,05 | 1,86 | 6,32E-02 | 1,84E-01 |
| AGMAT | 158,65 | 0,07 | 0,04 | 1,86 | 6,33E-02 | 1,84E-01 |
| UFSP2 | 381,55 | 0,05 | 0,03 | 1,86 | 6,33E-02 | 1,84E-01 |
| FLT3LG | 80,61 | 0,12 | 0,07 | 1,86 | 6,33E-02 | 1,84E-01 |
| GTF2B | 834,97 | -0,06 | 0,03 | -1,86 | 6,35E-02 | 1,85E-01 |
| GNB5 | 1210,20 | 0,04 | 0,02 | 1,86 | 6,35E-02 | 1,85E-01 |
| GNL3L | 1397,35 | -0,05 | 0,03 | -1,86 | 6,34E-02 | 1,85E-01 |
| NUDT22 | 443,60 | 0,04 | 0,02 | 1,86 | 6,34E-02 | 1,85E-01 |
| AP3M2 | 899,58 | 0,05 | 0,02 | 1,86 | 6,35E-02 | 1,85E-01 |
| UBTD1 | 82,22 | -0,09 | 0,05 | -1,85 | 6,38E-02 | 1,86E-01 |
| CAMK1 | 558,17 | 0,07 | 0,04 | 1,85 | 6,39E-02 | 1,86E-01 |
| DESI2 | 1078,78 | 0,04 | 0,02 | 1,85 | 6,39E-02 | 1,86E-01 |
| RRBP1 | 1623,73 | -0,05 | 0,03 | -1,85 | 6,39E-02 | 1,86E-01 |
| AEBP2 | 1494,97 | 0,05 | 0,03 | 1,85 | 6,39E-02 | 1,86E-01 |
| PRIM2 | 617,45 | 0,05 | 0,02 | 1,85 | 6,40E-02 | 1,86E-01 |
| RPL23A | 12537,60 | 0,07 | 0,04 | 1,85 | 6,41E-02 | 1,86E-01 |
| YWHAQ | 2095,38 | 0,04 | 0,02 | 1,85 | 6,42E-02 | 1,86E-01 |
| ZNF772 | 431,24 | 0,07 | 0,04 | 1,85 | 6,43E-02 | 1,86E-01 |
| SYNRG | 4674,94 | -0,04 | 0,02 | -1,85 | 6,44E-02 | 1,87E-01 |
| EPM2A | 433,01 | 0,05 | 0,03 | 1,85 | 6,43E-02 | 1,87E-01 |
| PAAF1 | 373,09 | 0,04 | 0,02 | 1,85 | 6,44E-02 | 1,87E-01 |
| ALPK3 | 105,89 | -0,14 | 0,08 | -1,85 | 6,47E-02 | 1,87E-01 |
| EVI5 | 1454,31 | -0,05 | 0,03 | -1,85 | 6,49E-02 | 1,88E-01 |
| ETF1 | 2005,59 | -0,04 | 0,02 | -1,85 | 6,49E-02 | 1,88E-01 |
| ZNF83 | 1057,32 | 0,07 | 0,04 | 1,85 | 6,50E-02 | 1,88E-01 |

| | | | | | | |
|---|---|---|---|---|---|---|
| TCF19 | 115,82 | 0,07 | 0,04 | 1,85 | 6,50E-02 | 1,88E-01 |
| ZNF571 | 216,51 | 0,07 | 0,04 | 1,84 | 6,51E-02 | 1,88E-01 |
| BECN1 | 2934,09 | -0,03 | 0,02 | -1,84 | 6,51E-02 | 1,88E-01 |
| HOMER1 | 103,70 | 0,08 | 0,05 | 1,84 | 6,51E-02 | 1,88E-01 |
| PTMA | 7114,38 | 0,05 | 0,03 | 1,84 | 6,52E-02 | 1,88E-01 |
| ZNF432 | 474,55 | 0,08 | 0,04 | 1,84 | 6,52E-02 | 1,88E-01 |
| BTN3A1 | 7292,86 | -0,06 | 0,03 | -1,84 | 6,53E-02 | 1,89E-01 |
| TAS2R14 | 310,22 | 0,08 | 0,04 | 1,84 | 6,53E-02 | 1,89E-01 |
| IRF2BPL | 1255,32 | -0,07 | 0,04 | -1,84 | 6,54E-02 | 1,89E-01 |
| NBN | 3495,72 | -0,07 | 0,04 | -1,84 | 6,55E-02 | 1,89E-01 |
| GAB3 | 1678,17 | -0,05 | 0,03 | -1,84 | 6,55E-02 | 1,89E-01 |
| CLDND1 | 1985,38 | 0,06 | 0,03 | 1,84 | 6,55E-02 | 1,89E-01 |
| CD3D | 1127,79 | 0,09 | 0,05 | 1,84 | 6,55E-02 | 1,89E-01 |
| SMARCE1 | 1148,20 | 0,05 | 0,03 | 1,84 | 6,56E-02 | 1,89E-01 |
| TEDC1 | 310,52 | 0,06 | 0,03 | 1,84 | 6,58E-02 | 1,90E-01 |
| ZFP69B | 98,67 | -0,04 | 0,02 | -1,84 | 6,58E-02 | 1,90E-01 |
| VPS26C | 2059,77 | -0,02 | 0,01 | -1,84 | 6,58E-02 | 1,90E-01 |
| RCC1L | 540,14 | 0,04 | 0,02 | 1,84 | 6,59E-02 | 1,90E-01 |
| CD226 | 4632,81 | 0,06 | 0,03 | 1,84 | 6,59E-02 | 1,90E-01 |
| UBE2D1 | 2885,79 | -0,09 | 0,05 | -1,84 | 6,59E-02 | 1,90E-01 |
| C16orf86 | 87,33 | 0,05 | 0,03 | 1,84 | 6,60E-02 | 1,90E-01 |
| SNX30 | 2207,46 | -0,04 | 0,02 | -1,84 | 6,61E-02 | 1,90E-01 |
| FUT8 | 1021,60 | 0,05 | 0,03 | 1,84 | 6,60E-02 | 1,90E-01 |
| COMTD1 | 101,03 | 0,06 | 0,03 | 1,84 | 6,61E-02 | 1,90E-01 |
| GPAA1 | 387,60 | 0,05 | 0,03 | 1,84 | 6,60E-02 | 1,90E-01 |
| GBA2 | 1262,70 | -0,05 | 0,03 | -1,84 | 6,62E-02 | 1,90E-01 |
| CELF1 | 6380,03 | -0,04 | 0,02 | -1,84 | 6,63E-02 | 1,90E-01 |
| METTL3 | 1420,31 | 0,03 | 0,02 | 1,84 | 6,63E-02 | 1,90E-01 |
| RDH14 | 246,13 | 0,08 | 0,04 | 1,84 | 6,63E-02 | 1,91E-01 |
| EIF3G | 3233,70 | 0,05 | 0,03 | 1,84 | 6,64E-02 | 1,91E-01 |
| SLC44A2 | 3287,25 | -0,09 | 0,05 | -1,84 | 6,64E-02 | 1,91E-01 |
| GIPC1 | 182,66 | 0,06 | 0,03 | 1,84 | 6,64E-02 | 1,91E-01 |
| NPDC1 | 97,37 | 0,13 | 0,07 | 1,84 | 6,65E-02 | 1,91E-01 |
| GPR183 | 1419,92 | 0,10 | 0,05 | 1,84 | 6,65E-02 | 1,91E-01 |
| CMTR2 | 1420,69 | 0,07 | 0,04 | 1,83 | 6,65E-02 | 1,91E-01 |
| RAB33A | 72,37 | 0,06 | 0,03 | 1,83 | 6,67E-02 | 1,91E-01 |
| CCDC167 | 107,33 | 0,08 | 0,05 | 1,83 | 6,67E-02 | 1,91E-01 |
| GLRX | 2718,13 | -0,05 | 0,03 | -1,83 | 6,67E-02 | 1,91E-01 |
| NBPF10 | 440,46 | -0,06 | 0,03 | -1,83 | 6,68E-02 | 1,91E-01 |
| IDE | 995,58 | 0,03 | 0,01 | 1,83 | 6,68E-02 | 1,91E-01 |
| CCDC170 | 269,90 | -0,07 | 0,04 | -1,83 | 6,68E-02 | 1,91E-01 |
| ITPR2 | 7869,75 | -0,05 | 0,03 | -1,83 | 6,69E-02 | 1,91E-01 |
| HOPX | 608,97 | 0,09 | 0,05 | 1,83 | 6,69E-02 | 1,92E-01 |
| PTK2B | 4295,42 | -0,08 | 0,04 | -1,83 | 6,70E-02 | 1,92E-01 |
| CNPY4 | 403,09 | 0,04 | 0,02 | 1,83 | 6,70E-02 | 1,92E-01 |
| ZNF528 | 814,98 | 0,05 | 0,03 | 1,83 | 6,70E-02 | 1,92E-01 |

| | | | | | | |
|---|---|---|---|---|---|---|
| INTS8 | 2667,14 | -0,03 | 0,02 | -1,83 | 6,71E-02 | 1,92E-01 |
| ETFBKMT | 99,98 | 0,06 | 0,03 | 1,83 | 6,71E-02 | 1,92E-01 |
| ST8SIA1 | 483,62 | 0,08 | 0,05 | 1,83 | 6,72E-02 | 1,92E-01 |
| AMFR | 2662,26 | 0,06 | 0,03 | 1,83 | 6,72E-02 | 1,92E-01 |
| UAP1L1 | 140,59 | 0,11 | 0,06 | 1,83 | 6,73E-02 | 1,92E-01 |
| PSMA6 | 619,53 | 0,06 | 0,03 | 1,83 | 6,74E-02 | 1,92E-01 |
| CKS1B | 97,52 | 0,09 | 0,05 | 1,83 | 6,73E-02 | 1,92E-01 |
| KIAA1191 | 1071,21 | 0,03 | 0,02 | 1,83 | 6,74E-02 | 1,92E-01 |
| NUDT12 | 74,66 | 0,13 | 0,07 | 1,83 | 6,75E-02 | 1,93E-01 |
| REPS1 | 1878,08 | 0,03 | 0,02 | 1,83 | 6,75E-02 | 1,93E-01 |
| RPL6 | 5477,24 | 0,10 | 0,06 | 1,83 | 6,75E-02 | 1,93E-01 |
| DBF4 | 393,82 | 0,06 | 0,03 | 1,83 | 6,76E-02 | 1,93E-01 |
| MIX23 | 135,66 | 0,06 | 0,03 | 1,83 | 6,76E-02 | 1,93E-01 |
| CTDSPL2 | 2461,04 | 0,05 | 0,03 | 1,83 | 6,76E-02 | 1,93E-01 |
| C10orf105 | 154,66 | -0,12 | 0,07 | -1,83 | 6,77E-02 | 1,93E-01 |
| GNA13 | 6234,06 | -0,05 | 0,03 | -1,83 | 6,78E-02 | 1,93E-01 |
| RHD | 92,44 | 0,25 | 0,14 | 1,83 | 6,77E-02 | 1,93E-01 |
| GASK1B | 1336,88 | 0,08 | 0,05 | 1,83 | 6,78E-02 | 1,93E-01 |
| DACT1 | 123,94 | 0,14 | 0,08 | 1,83 | 6,78E-02 | 1,93E-01 |
| SAMD8 | 2819,64 | -0,05 | 0,03 | -1,83 | 6,78E-02 | 1,93E-01 |
| MYD88 | 1871,76 | -0,08 | 0,04 | -1,83 | 6,79E-02 | 1,93E-01 |
| AMIGO1 | 313,71 | 0,07 | 0,04 | 1,83 | 6,79E-02 | 1,93E-01 |
| FAM220A | 320,39 | 0,04 | 0,02 | 1,83 | 6,80E-02 | 1,93E-01 |
| KEL | 164,74 | 0,13 | 0,07 | 1,82 | 6,80E-02 | 1,93E-01 |
| VPS4A | 512,37 | 0,03 | 0,02 | 1,82 | 6,81E-02 | 1,93E-01 |
| CCDC88B | 6099,87 | -0,07 | 0,04 | -1,82 | 6,81E-02 | 1,93E-01 |
| SLC30A7 | 2728,04 | 0,04 | 0,02 | 1,82 | 6,81E-02 | 1,93E-01 |
| NIFK | 516,53 | 0,06 | 0,03 | 1,82 | 6,81E-02 | 1,93E-01 |
| SNRNP200 | 4835,51 | -0,04 | 0,02 | -1,82 | 6,80E-02 | 1,93E-01 |
| TRPC1 | 225,87 | 0,08 | 0,04 | 1,82 | 6,82E-02 | 1,93E-01 |
| RPS4Y1 | 2160,90 | 0,22 | 0,12 | 1,82 | 6,82E-02 | 1,93E-01 |
| RBM19 | 1306,89 | 0,04 | 0,02 | 1,82 | 6,82E-02 | 1,93E-01 |
| KDM4B | 3003,33 | -0,09 | 0,05 | -1,82 | 6,82E-02 | 1,94E-01 |
| NLRP6 | 1081,51 | -0,09 | 0,05 | -1,82 | 6,83E-02 | 1,94E-01 |
| LIN7C | 1856,93 | 0,08 | 0,04 | 1,82 | 6,83E-02 | 1,94E-01 |
| ABCA5 | 2291,23 | 0,06 | 0,03 | 1,82 | 6,84E-02 | 1,94E-01 |
| GRINA | 685,65 | 0,15 | 0,08 | 1,82 | 6,83E-02 | 1,94E-01 |
| LSP1 | 9507,78 | -0,07 | 0,04 | -1,82 | 6,83E-02 | 1,94E-01 |
| FOS | 1278,96 | -0,09 | 0,05 | -1,82 | 6,84E-02 | 1,94E-01 |
| PRKD2 | 1448,71 | -0,09 | 0,05 | -1,82 | 6,84E-02 | 1,94E-01 |
| CHUK | 1708,20 | -0,06 | 0,03 | -1,82 | 6,85E-02 | 1,94E-01 |
| FLII | 3537,63 | -0,06 | 0,03 | -1,82 | 6,85E-02 | 1,94E-01 |
| PTPRK | 216,29 | 0,10 | 0,06 | 1,82 | 6,86E-02 | 1,94E-01 |
| LPIN1 | 1935,38 | 0,04 | 0,02 | 1,82 | 6,86E-02 | 1,94E-01 |
| ABCG2 | 151,20 | 0,13 | 0,07 | 1,82 | 6,86E-02 | 1,94E-01 |
| SUPT6H | 2354,73 | -0,07 | 0,04 | -1,82 | 6,87E-02 | 1,94E-01 |

| | | | | | | |
|---|---|---|---|---|---|---|
| PRDX2 | 1071,40 | 0,11 | 0,06 | 1,82 | 6,88E-02 | 1,94E-01 |
| APBB1 | 496,73 | 0,06 | 0,03 | 1,82 | 6,88E-02 | 1,94E-01 |
| TRIP12 | 12427,81 | -0,04 | 0,02 | -1,82 | 6,88E-02 | 1,94E-01 |
| PDSS2 | 229,50 | 0,03 | 0,02 | 1,82 | 6,89E-02 | 1,95E-01 |
| RENBP | 484,62 | -0,06 | 0,03 | -1,82 | 6,90E-02 | 1,95E-01 |
| NAA25 | 1227,58 | 0,03 | 0,02 | 1,82 | 6,90E-02 | 1,95E-01 |
| DDX42 | 2815,91 | -0,03 | 0,02 | -1,82 | 6,90E-02 | 1,95E-01 |
| GPR171 | 469,61 | 0,09 | 0,05 | 1,82 | 6,91E-02 | 1,95E-01 |
| ENGASE | 1147,81 | 0,06 | 0,03 | 1,82 | 6,92E-02 | 1,95E-01 |
| PRKCD | 1297,02 | -0,11 | 0,06 | -1,82 | 6,92E-02 | 1,95E-01 |
| CSNK1D | 2344,93 | -0,08 | 0,05 | -1,82 | 6,93E-02 | 1,95E-01 |
| IMP4 | 756,23 | 0,03 | 0,02 | 1,82 | 6,94E-02 | 1,95E-01 |
| PSD4 | 5219,20 | -0,07 | 0,04 | -1,82 | 6,94E-02 | 1,96E-01 |
| WDPCP | 689,24 | 0,05 | 0,03 | 1,82 | 6,95E-02 | 1,96E-01 |
| ZNF529 | 700,63 | 0,05 | 0,03 | 1,81 | 6,96E-02 | 1,96E-01 |
| SLC25A33 | 130,75 | 0,04 | 0,02 | 1,81 | 6,96E-02 | 1,96E-01 |
| UBE2K | 1978,73 | 0,05 | 0,03 | 1,81 | 6,97E-02 | 1,96E-01 |
| MAST3 | 2485,25 | -0,10 | 0,06 | -1,81 | 6,98E-02 | 1,96E-01 |
| RAB6A | 3065,77 | 0,05 | 0,03 | 1,81 | 6,98E-02 | 1,96E-01 |
| KIAA1143 | 1383,08 | 0,05 | 0,03 | 1,81 | 6,99E-02 | 1,96E-01 |
| PHF8 | 1579,82 | -0,04 | 0,02 | -1,81 | 7,00E-02 | 1,97E-01 |
| CCNI2 | 83,24 | 0,07 | 0,04 | 1,81 | 7,01E-02 | 1,97E-01 |
| SMYD4 | 739,47 | 0,03 | 0,02 | 1,81 | 7,01E-02 | 1,97E-01 |
| EEIG1 | 678,74 | 0,11 | 0,06 | 1,81 | 7,02E-02 | 1,97E-01 |
| GSTM4 | 191,89 | 0,10 | 0,06 | 1,81 | 7,02E-02 | 1,97E-01 |
| PDCD6 | 2079,50 | -0,03 | 0,01 | -1,81 | 7,03E-02 | 1,97E-01 |
| SLC19A1 | 1941,06 | -0,11 | 0,06 | -1,81 | 7,03E-02 | 1,97E-01 |
| CHST2 | 1256,51 | -0,06 | 0,03 | -1,81 | 7,04E-02 | 1,98E-01 |
| MRPL50 | 489,11 | 0,11 | 0,06 | 1,81 | 7,04E-02 | 1,98E-01 |
| PMAIP1 | 451,89 | 0,10 | 0,05 | 1,81 | 7,04E-02 | 1,98E-01 |
| GTF3C3 | 1198,39 | 0,05 | 0,03 | 1,81 | 7,04E-02 | 1,98E-01 |
| NDUFS6 | 478,74 | 0,04 | 0,02 | 1,81 | 7,05E-02 | 1,98E-01 |
| SELENOK | 1792,66 | 0,10 | 0,05 | 1,81 | 7,06E-02 | 1,98E-01 |
| TIA1 | 4156,46 | 0,06 | 0,03 | 1,81 | 7,06E-02 | 1,98E-01 |
| SERAC1 | 443,46 | -0,05 | 0,03 | -1,81 | 7,07E-02 | 1,98E-01 |
| HERC4 | 6428,04 | -0,04 | 0,02 | -1,81 | 7,08E-02 | 1,98E-01 |
| SYNGR2 | 1232,64 | 0,04 | 0,02 | 1,81 | 7,08E-02 | 1,98E-01 |
| SNX12 | 637,10 | -0,03 | 0,01 | -1,81 | 7,08E-02 | 1,98E-01 |
| FAS | 2445,37 | -0,07 | 0,04 | -1,81 | 7,09E-02 | 1,98E-01 |
| RRP9 | 220,58 | 0,04 | 0,02 | 1,81 | 7,09E-02 | 1,98E-01 |
| CSPP1 | 666,80 | 0,04 | 0,02 | 1,81 | 7,10E-02 | 1,99E-01 |
| SNAP29 | 761,82 | -0,04 | 0,02 | -1,81 | 7,10E-02 | 1,99E-01 |
| PSMB3 | 2803,15 | -0,05 | 0,03 | -1,80 | 7,11E-02 | 1,99E-01 |
| SLA2 | 734,35 | -0,05 | 0,03 | -1,80 | 7,11E-02 | 1,99E-01 |
| SAMHD1 | 9201,66 | -0,05 | 0,03 | -1,80 | 7,12E-02 | 1,99E-01 |
| MTMR14 | 2839,96 | -0,04 | 0,02 | -1,80 | 7,12E-02 | 1,99E-01 |

| | | | | | | |
|---|---|---|---|---|---|---|
| ABCE1 | 1202,78 | 0,07 | 0,04 | 1,80 | 7,13E-02 | 1,99E-01 |
| CD3G | 1649,18 | 0,06 | 0,04 | 1,80 | 7,13E-02 | 1,99E-01 |
| BIRC6 | 15834,34 | -0,04 | 0,02 | -1,80 | 7,13E-02 | 1,99E-01 |
| MAST4 | 587,64 | 0,06 | 0,03 | 1,80 | 7,13E-02 | 1,99E-01 |
| AUP1 | 1298,53 | 0,03 | 0,02 | 1,80 | 7,14E-02 | 1,99E-01 |
| SPHK1 | 148,93 | 0,08 | 0,04 | 1,80 | 7,14E-02 | 1,99E-01 |
| SLC4A1 | 13284,08 | 0,16 | 0,09 | 1,80 | 7,15E-02 | 1,99E-01 |
| DNAJC4 | 137,40 | 0,07 | 0,04 | 1,80 | 7,15E-02 | 1,99E-01 |
| MBNL2 | 1409,65 | 0,06 | 0,03 | 1,80 | 7,15E-02 | 1,99E-01 |
| BICDL1 | 311,13 | 0,08 | 0,04 | 1,80 | 7,16E-02 | 2,00E-01 |
| FEZ2 | 1881,04 | -0,04 | 0,02 | -1,80 | 7,17E-02 | 2,00E-01 |
| MPZ | 243,63 | -0,06 | 0,03 | -1,80 | 7,17E-02 | 2,00E-01 |
| KDM5C | 3320,60 | -0,05 | 0,03 | -1,80 | 7,19E-02 | 2,00E-01 |
| ANXA2R | 573,77 | 0,06 | 0,03 | 1,80 | 7,19E-02 | 2,00E-01 |
| ZNF829 | 214,32 | 0,07 | 0,04 | 1,80 | 7,18E-02 | 2,00E-01 |
| FRMD4B | 721,54 | -0,05 | 0,03 | -1,80 | 7,19E-02 | 2,00E-01 |
| PGAP3 | 180,98 | 0,06 | 0,03 | 1,80 | 7,18E-02 | 2,00E-01 |
| MTIF2 | 671,06 | 0,05 | 0,03 | 1,80 | 7,19E-02 | 2,00E-01 |
| ASAP1 | 3497,39 | -0,07 | 0,04 | -1,80 | 7,19E-02 | 2,00E-01 |
| TTC12 | 864,31 | 0,05 | 0,03 | 1,80 | 7,20E-02 | 2,00E-01 |
| FIRRM | 463,75 | 0,04 | 0,02 | 1,80 | 7,20E-02 | 2,00E-01 |
| TFEC | 1842,21 | 0,11 | 0,06 | 1,80 | 7,20E-02 | 2,00E-01 |
| STX18 | 668,32 | 0,02 | 0,01 | 1,80 | 7,20E-02 | 2,00E-01 |
| PPIL3 | 345,52 | 0,07 | 0,04 | 1,80 | 7,20E-02 | 2,00E-01 |
| BSG | 14750,88 | 0,12 | 0,07 | 1,80 | 7,21E-02 | 2,00E-01 |
| RITA1 | 215,86 | 0,03 | 0,02 | 1,80 | 7,21E-02 | 2,00E-01 |
| NR2C2AP | 225,98 | 0,04 | 0,02 | 1,80 | 7,22E-02 | 2,00E-01 |
| PDK4 | 978,85 | -0,11 | 0,06 | -1,80 | 7,22E-02 | 2,00E-01 |
| RSL1D1 | 1668,22 | 0,05 | 0,03 | 1,80 | 7,23E-02 | 2,01E-01 |
| PTBP2 | 1581,11 | 0,06 | 0,03 | 1,80 | 7,24E-02 | 2,01E-01 |
| ID3 | 125,66 | 0,10 | 0,05 | 1,80 | 7,25E-02 | 2,01E-01 |
| EXD3 | 237,65 | 0,08 | 0,05 | 1,80 | 7,25E-02 | 2,01E-01 |
| TBC1D22B | 619,71 | 0,08 | 0,04 | 1,79 | 7,28E-02 | 2,02E-01 |
| ZNF765 | 560,37 | 0,09 | 0,05 | 1,79 | 7,27E-02 | 2,02E-01 |
| RCAN3 | 2838,66 | 0,06 | 0,04 | 1,79 | 7,29E-02 | 2,02E-01 |
| ZNF569 | 270,86 | 0,07 | 0,04 | 1,79 | 7,28E-02 | 2,02E-01 |
| ZNF845 | 618,86 | 0,10 | 0,06 | 1,79 | 7,28E-02 | 2,02E-01 |
| DGKK | 105,71 | -0,16 | 0,09 | -1,79 | 7,28E-02 | 2,02E-01 |
| SMS | 889,54 | -0,05 | 0,03 | -1,79 | 7,29E-02 | 2,02E-01 |
| COMMD4 | 272,86 | 0,04 | 0,02 | 1,79 | 7,29E-02 | 2,02E-01 |
| R3HDM1 | 784,76 | 0,04 | 0,02 | 1,79 | 7,29E-02 | 2,02E-01 |
| IBTK | 1950,63 | 0,05 | 0,03 | 1,79 | 7,30E-02 | 2,02E-01 |
| SLC12A9 | 3513,65 | -0,07 | 0,04 | -1,79 | 7,31E-02 | 2,02E-01 |
| PROSER1 | 1085,84 | 0,03 | 0,02 | 1,79 | 7,33E-02 | 2,02E-01 |
| NUDT17 | 73,15 | 0,05 | 0,03 | 1,79 | 7,33E-02 | 2,02E-01 |
| CILK1 | 712,05 | 0,03 | 0,01 | 1,79 | 7,33E-02 | 2,02E-01 |

| | | | | | | |
|---|---|---|---|---|---|---|
| RGS14 | 1442,95 | -0,07 | 0,04 | -1,79 | 7,33E-02 | 2,03E-01 |
| NOL11 | 1094,82 | 0,05 | 0,03 | 1,79 | 7,34E-02 | 2,03E-01 |
| ACBD4 | 120,56 | 0,08 | 0,04 | 1,79 | 7,35E-02 | 2,03E-01 |
| CPEB2 | 2137,75 | -0,07 | 0,04 | -1,79 | 7,36E-02 | 2,03E-01 |
| INF2 | 332,42 | 0,12 | 0,07 | 1,79 | 7,36E-02 | 2,03E-01 |
| NDUFV1 | 1016,75 | 0,03 | 0,02 | 1,79 | 7,36E-02 | 2,03E-01 |
| HMOX2 | 911,86 | -0,03 | 0,02 | -1,79 | 7,36E-02 | 2,03E-01 |
| DIAPH1 | 3968,58 | -0,07 | 0,04 | -1,79 | 7,37E-02 | 2,03E-01 |
| ENSG000001 | 444,95 | 0,05 | 0,03 | 1,79 | 7,38E-02 | 2,03E-01 |
| TMEM59 | 3973,70 | -0,05 | 0,03 | -1,79 | 7,38E-02 | 2,03E-01 |
| CHD9 | 3602,25 | 0,04 | 0,02 | 1,79 | 7,39E-02 | 2,03E-01 |
| UNKL | 1316,40 | -0,07 | 0,04 | -1,79 | 7,39E-02 | 2,03E-01 |
| LMAN2L | 319,96 | -0,03 | 0,01 | -1,79 | 7,40E-02 | 2,04E-01 |
| MAFK | 1253,70 | 0,04 | 0,02 | 1,79 | 7,41E-02 | 2,04E-01 |
| ANAPC16 | 3514,06 | 0,04 | 0,02 | 1,79 | 7,41E-02 | 2,04E-01 |
| UBN2 | 2826,83 | -0,03 | 0,02 | -1,79 | 7,42E-02 | 2,04E-01 |
| ITGB7 | 1845,65 | 0,05 | 0,03 | 1,79 | 7,42E-02 | 2,04E-01 |
| MDH1 | 1160,31 | 0,05 | 0,03 | 1,79 | 7,42E-02 | 2,04E-01 |
| ARGLU1 | 8983,34 | 0,06 | 0,03 | 1,79 | 7,42E-02 | 2,04E-01 |
| CPNE1 | 2300,81 | -0,04 | 0,02 | -1,79 | 7,42E-02 | 2,04E-01 |
| RAB4A | 822,88 | 0,03 | 0,02 | 1,78 | 7,43E-02 | 2,04E-01 |
| ZNF195 | 1573,51 | 0,05 | 0,03 | 1,78 | 7,43E-02 | 2,04E-01 |
| ZBTB5 | 522,06 | 0,04 | 0,02 | 1,78 | 7,43E-02 | 2,04E-01 |
| PRPF4 | 798,44 | -0,03 | 0,02 | -1,78 | 7,43E-02 | 2,04E-01 |
| SPATA7 | 141,99 | 0,06 | 0,04 | 1,78 | 7,44E-02 | 2,04E-01 |
| ZNF611 | 1090,42 | -0,04 | 0,02 | -1,78 | 7,44E-02 | 2,04E-01 |
| MAD2L1 | 124,78 | 0,08 | 0,05 | 1,78 | 7,45E-02 | 2,04E-01 |
| IKZF5 | 1009,75 | 0,05 | 0,03 | 1,78 | 7,45E-02 | 2,04E-01 |
| MSN | 5958,25 | -0,08 | 0,05 | -1,78 | 7,45E-02 | 2,04E-01 |
| IMPA2 | 1086,39 | -0,07 | 0,04 | -1,78 | 7,47E-02 | 2,05E-01 |
| SNAPC1 | 139,10 | 0,07 | 0,04 | 1,78 | 7,47E-02 | 2,05E-01 |
| UBQLN2 | 823,92 | 0,05 | 0,03 | 1,78 | 7,48E-02 | 2,05E-01 |
| ADAT2 | 468,56 | 0,04 | 0,02 | 1,78 | 7,48E-02 | 2,05E-01 |
| COX6C | 462,07 | 0,08 | 0,04 | 1,78 | 7,48E-02 | 2,05E-01 |
| PKNOX1 | 603,66 | -0,05 | 0,03 | -1,78 | 7,48E-02 | 2,05E-01 |
| CD300LF | 1343,09 | -0,06 | 0,04 | -1,78 | 7,50E-02 | 2,05E-01 |
| PHETA2 | 126,81 | 0,05 | 0,03 | 1,78 | 7,51E-02 | 2,06E-01 |
| MRPL19 | 660,27 | 0,04 | 0,02 | 1,78 | 7,52E-02 | 2,06E-01 |
| PHF5A | 266,34 | 0,07 | 0,04 | 1,78 | 7,53E-02 | 2,06E-01 |
| FZD5 | 137,85 | 0,09 | 0,05 | 1,78 | 7,54E-02 | 2,06E-01 |
| CD63 | 3123,97 | -0,06 | 0,03 | -1,78 | 7,54E-02 | 2,06E-01 |
| CSTF3 | 826,36 | 0,05 | 0,03 | 1,78 | 7,54E-02 | 2,06E-01 |
| FES | 2541,67 | -0,06 | 0,03 | -1,78 | 7,55E-02 | 2,06E-01 |
| NAE1 | 702,88 | 0,05 | 0,03 | 1,78 | 7,56E-02 | 2,06E-01 |
| PCCB | 515,37 | 0,02 | 0,01 | 1,78 | 7,56E-02 | 2,07E-01 |
| CD81 | 759,59 | 0,07 | 0,04 | 1,78 | 7,57E-02 | 2,07E-01 |

| | | | | | | |
|---|---|---|---|---|---|---|
| RETREG1 | 588,75 | 0,06 | 0,03 | 1,78 | 7,58E-02 | 2,07E-01 |
| WRN | 892,15 | 0,05 | 0,03 | 1,77 | 7,59E-02 | 2,07E-01 |
| HK1 | 2192,68 | 0,08 | 0,04 | 1,77 | 7,61E-02 | 2,08E-01 |
| CC2D2B | 206,10 | -0,09 | 0,05 | -1,77 | 7,63E-02 | 2,08E-01 |
| RABGEF1 | 251,63 | -0,03 | 0,02 | -1,77 | 7,65E-02 | 2,09E-01 |
| CACTIN | 436,83 | 0,07 | 0,04 | 1,77 | 7,65E-02 | 2,09E-01 |
| GTDC1 | 1480,85 | -0,04 | 0,02 | -1,77 | 7,65E-02 | 2,09E-01 |
| PLK4 | 87,21 | 0,08 | 0,05 | 1,77 | 7,66E-02 | 2,09E-01 |
| MYO9B | 5296,41 | -0,09 | 0,05 | -1,77 | 7,66E-02 | 2,09E-01 |
| VEZT | 1282,69 | 0,04 | 0,02 | 1,77 | 7,67E-02 | 2,09E-01 |
| ZNF530 | 271,60 | 0,04 | 0,02 | 1,77 | 7,67E-02 | 2,09E-01 |
| INTS3 | 1479,47 | -0,07 | 0,04 | -1,77 | 7,67E-02 | 2,09E-01 |
| PPIA | 5439,48 | 0,06 | 0,04 | 1,77 | 7,67E-02 | 2,09E-01 |
| UQCRH | 876,44 | 0,08 | 0,05 | 1,77 | 7,68E-02 | 2,09E-01 |
| MGST1 | 166,71 | 0,07 | 0,04 | 1,77 | 7,68E-02 | 2,09E-01 |
| DDX46 | 2698,28 | -0,03 | 0,02 | -1,77 | 7,69E-02 | 2,09E-01 |
| SEC11C | 498,48 | 0,05 | 0,03 | 1,77 | 7,69E-02 | 2,09E-01 |
| DDX23 | 3104,84 | -0,04 | 0,02 | -1,77 | 7,70E-02 | 2,09E-01 |
| ATP6AP2 | 2971,46 | -0,05 | 0,03 | -1,77 | 7,71E-02 | 2,09E-01 |
| TMEM185A | 381,56 | -0,03 | 0,02 | -1,77 | 7,71E-02 | 2,09E-01 |
| DAPK3 | 759,63 | -0,04 | 0,02 | -1,77 | 7,72E-02 | 2,10E-01 |
| KDM7A | 9327,15 | -0,05 | 0,03 | -1,77 | 7,72E-02 | 2,10E-01 |
| UBE2J2 | 1337,69 | -0,03 | 0,02 | -1,77 | 7,72E-02 | 2,10E-01 |
| ALPL | 1248,70 | -0,14 | 0,08 | -1,77 | 7,72E-02 | 2,10E-01 |
| METTL26 | 129,51 | 0,06 | 0,03 | 1,77 | 7,74E-02 | 2,10E-01 |
| OPA3 | 255,95 | 0,06 | 0,03 | 1,77 | 7,74E-02 | 2,10E-01 |
| THUMPD3 | 545,14 | 0,04 | 0,02 | 1,77 | 7,74E-02 | 2,10E-01 |
| CPSF7 | 1072,85 | -0,07 | 0,04 | -1,77 | 7,74E-02 | 2,10E-01 |
| RANBP6 | 1043,19 | 0,09 | 0,05 | 1,77 | 7,74E-02 | 2,10E-01 |
| PILRA | 549,88 | -0,12 | 0,07 | -1,77 | 7,75E-02 | 2,10E-01 |
| PGK1 | 9280,38 | -0,04 | 0,02 | -1,76 | 7,76E-02 | 2,10E-01 |
| TXNRD2 | 704,72 | 0,07 | 0,04 | 1,76 | 7,77E-02 | 2,10E-01 |
| FBXO25 | 768,06 | 0,03 | 0,02 | 1,76 | 7,77E-02 | 2,11E-01 |
| VPS36 | 2224,30 | 0,03 | 0,02 | 1,76 | 7,78E-02 | 2,11E-01 |
| SOCS2 | 305,04 | 0,06 | 0,03 | 1,76 | 7,78E-02 | 2,11E-01 |
| PCBP4 | 121,68 | 0,07 | 0,04 | 1,76 | 7,79E-02 | 2,11E-01 |
| COA7 | 188,25 | 0,05 | 0,03 | 1,76 | 7,80E-02 | 2,11E-01 |
| GNL3 | 826,06 | 0,08 | 0,04 | 1,76 | 7,80E-02 | 2,11E-01 |
| FBXO48 | 253,92 | 0,04 | 0,02 | 1,76 | 7,81E-02 | 2,11E-01 |
| AKR7A2 | 381,10 | 0,04 | 0,02 | 1,76 | 7,81E-02 | 2,11E-01 |
| FBN2 | 556,19 | -0,10 | 0,06 | -1,76 | 7,81E-02 | 2,11E-01 |
| SLC7A1 | 509,49 | 0,06 | 0,04 | 1,76 | 7,82E-02 | 2,11E-01 |
| SYNCRIP | 3338,20 | 0,05 | 0,03 | 1,76 | 7,82E-02 | 2,11E-01 |
| XPR1 | 1580,90 | -0,03 | 0,02 | -1,76 | 7,83E-02 | 2,12E-01 |
| TMEM41A | 707,15 | 0,03 | 0,02 | 1,76 | 7,84E-02 | 2,12E-01 |
| BRD2 | 3719,43 | -0,04 | 0,02 | -1,76 | 7,84E-02 | 2,12E-01 |

| | | | | | | |
|---|---|---|---|---|---|---|
| C10orf143 | 173,05 | 0,06 | 0,03 | 1,76 | 7,85E-02 | 2,12E-01 |
| BZW2 | 559,45 | 0,04 | 0,02 | 1,76 | 7,86E-02 | 2,12E-01 |
| UGP2 | 2456,74 | -0,03 | 0,02 | -1,76 | 7,86E-02 | 2,12E-01 |
| LSG1 | 1025,87 | 0,04 | 0,02 | 1,76 | 7,85E-02 | 2,12E-01 |
| MED25 | 1472,73 | -0,07 | 0,04 | -1,76 | 7,86E-02 | 2,12E-01 |
| LYPLAL1 | 1000,97 | 0,07 | 0,04 | 1,76 | 7,85E-02 | 2,12E-01 |
| SPATA2 | 206,16 | -0,07 | 0,04 | -1,76 | 7,86E-02 | 2,12E-01 |
| DNAAF2 | 196,25 | 0,06 | 0,03 | 1,76 | 7,87E-02 | 2,12E-01 |
| ARRDC5 | 180,97 | 0,08 | 0,05 | 1,76 | 7,87E-02 | 2,12E-01 |
| FBXO45 | 393,12 | 0,05 | 0,03 | 1,76 | 7,88E-02 | 2,12E-01 |
| UBE3B | 1835,53 | -0,04 | 0,02 | -1,76 | 7,88E-02 | 2,12E-01 |
| SNX25 | 507,19 | 0,06 | 0,03 | 1,76 | 7,88E-02 | 2,12E-01 |
| UBA2 | 1646,67 | 0,05 | 0,03 | 1,76 | 7,89E-02 | 2,12E-01 |
| RASGRP1 | 3025,73 | 0,05 | 0,03 | 1,76 | 7,89E-02 | 2,12E-01 |
| AKTIP | 1591,78 | -0,05 | 0,03 | -1,76 | 7,90E-02 | 2,12E-01 |
| TMEM19 | 866,70 | 0,05 | 0,03 | 1,76 | 7,90E-02 | 2,12E-01 |
| PYCR2 | 430,67 | 0,05 | 0,03 | 1,76 | 7,90E-02 | 2,12E-01 |
| SRD5A3 | 156,97 | 0,08 | 0,04 | 1,76 | 7,91E-02 | 2,13E-01 |
| DHRS3 | 360,69 | 0,05 | 0,03 | 1,76 | 7,91E-02 | 2,13E-01 |
| RNF139 | 1382,67 | 0,06 | 0,04 | 1,76 | 7,91E-02 | 2,13E-01 |
| KLF9 | 1004,35 | 0,06 | 0,03 | 1,76 | 7,91E-02 | 2,13E-01 |
| HERC6 | 1012,62 | -0,08 | 0,05 | -1,76 | 7,91E-02 | 2,13E-01 |
| ENSG000002 | 362,14 | -0,05 | 0,03 | -1,76 | 7,92E-02 | 2,13E-01 |
| C19orf12 | 780,82 | 0,03 | 0,02 | 1,76 | 7,92E-02 | 2,13E-01 |
| VILL | 639,66 | 0,05 | 0,03 | 1,76 | 7,92E-02 | 2,13E-01 |
| NLE1 | 322,01 | 0,05 | 0,03 | 1,75 | 7,93E-02 | 2,13E-01 |
| ZNF717 | 330,57 | 0,04 | 0,03 | 1,75 | 7,94E-02 | 2,13E-01 |
| UBE2D2 | 1513,85 | 0,03 | 0,02 | 1,75 | 7,94E-02 | 2,13E-01 |
| PDE3B | 6950,86 | -0,05 | 0,03 | -1,75 | 7,95E-02 | 2,13E-01 |
| DENND1A | 1353,72 | -0,06 | 0,04 | -1,75 | 7,95E-02 | 2,13E-01 |
| NRF1 | 1032,86 | -0,03 | 0,02 | -1,75 | 7,95E-02 | 2,13E-01 |
| ARID4A | 3864,04 | -0,06 | 0,03 | -1,75 | 7,96E-02 | 2,13E-01 |
| SRSF10 | 4099,56 | 0,06 | 0,03 | 1,75 | 7,97E-02 | 2,13E-01 |
| TMSB4Y | 72,42 | 0,07 | 0,04 | 1,75 | 7,98E-02 | 2,14E-01 |
| ELAC2 | 1253,13 | 0,03 | 0,02 | 1,75 | 7,98E-02 | 2,14E-01 |
| UTP20 | 1017,77 | 0,04 | 0,02 | 1,75 | 7,99E-02 | 2,14E-01 |
| ATP6V0A2 | 1618,13 | 0,03 | 0,01 | 1,75 | 7,99E-02 | 2,14E-01 |
| HMGCL | 622,01 | 0,02 | 0,01 | 1,75 | 7,99E-02 | 2,14E-01 |
| N6AMT1 | 515,17 | 0,05 | 0,03 | 1,75 | 7,99E-02 | 2,14E-01 |
| HDAC1 | 3076,80 | -0,03 | 0,02 | -1,75 | 8,00E-02 | 2,14E-01 |
| CLDN12 | 72,94 | 0,07 | 0,04 | 1,75 | 8,00E-02 | 2,14E-01 |
| PAFAH1B3 | 123,03 | 0,04 | 0,03 | 1,75 | 8,00E-02 | 2,14E-01 |
| RBM26 | 4027,30 | 0,04 | 0,02 | 1,75 | 8,02E-02 | 2,14E-01 |
| ITGAV | 946,75 | 0,05 | 0,03 | 1,75 | 8,02E-02 | 2,14E-01 |
| WAS | 846,85 | -0,10 | 0,06 | -1,75 | 8,02E-02 | 2,14E-01 |
| EDEM3 | 4729,24 | -0,05 | 0,03 | -1,75 | 8,03E-02 | 2,14E-01 |

| | | | | | | |
|---|---|---|---|---|---|---|
| OAS1 | 1945,78 | -0,12 | 0,07 | -1,75 | 8,03E-02 | 2,14E-01 |
| ADAM8 | 6555,69 | -0,08 | 0,04 | -1,75 | 8,03E-02 | 2,14E-01 |
| DNAJC27 | 232,72 | 0,04 | 0,02 | 1,75 | 8,03E-02 | 2,14E-01 |
| NOL6 | 327,50 | 0,07 | 0,04 | 1,75 | 8,03E-02 | 2,14E-01 |
| RNF141 | 4038,05 | -0,06 | 0,03 | -1,75 | 8,04E-02 | 2,14E-01 |
| SPIN1 | 764,87 | 0,03 | 0,02 | 1,75 | 8,04E-02 | 2,14E-01 |
| TRA2A | 3753,27 | -0,04 | 0,02 | -1,75 | 8,04E-02 | 2,14E-01 |
| PGM3 | 751,83 | 0,05 | 0,03 | 1,75 | 8,04E-02 | 2,14E-01 |
| VARS2 | 220,75 | 0,08 | 0,04 | 1,75 | 8,05E-02 | 2,14E-01 |
| CEP170 | 3211,21 | -0,04 | 0,02 | -1,75 | 8,05E-02 | 2,14E-01 |
| EOGT | 278,43 | 0,04 | 0,02 | 1,75 | 8,07E-02 | 2,15E-01 |
| GGNBP2 | 2966,67 | -0,03 | 0,02 | -1,75 | 8,07E-02 | 2,15E-01 |
| MAP2K3 | 2841,74 | 0,10 | 0,06 | 1,75 | 8,08E-02 | 2,15E-01 |
| AKT1 | 3508,06 | -0,05 | 0,03 | -1,75 | 8,08E-02 | 2,15E-01 |
| SEC24C | 2086,62 | -0,05 | 0,03 | -1,75 | 8,08E-02 | 2,15E-01 |
| TMEM121B | 184,33 | -0,12 | 0,07 | -1,75 | 8,09E-02 | 2,15E-01 |
| NKRF | 371,31 | 0,04 | 0,02 | 1,75 | 8,09E-02 | 2,15E-01 |
| BTG3 | 135,08 | 0,06 | 0,04 | 1,75 | 8,09E-02 | 2,15E-01 |
| ENSG000002 | 129,50 | -0,20 | 0,11 | -1,74 | 8,10E-02 | 2,15E-01 |
| LY86 | 520,89 | 0,06 | 0,03 | 1,74 | 8,11E-02 | 2,15E-01 |
| ZNF879 | 292,27 | 0,07 | 0,04 | 1,74 | 8,12E-02 | 2,16E-01 |
| BRD10 | 3590,41 | -0,03 | 0,02 | -1,74 | 8,13E-02 | 2,16E-01 |
| MS4A4E | 423,96 | 0,07 | 0,04 | 1,74 | 8,14E-02 | 2,16E-01 |
| PELO | 351,80 | -0,05 | 0,03 | -1,74 | 8,14E-02 | 2,16E-01 |
| CD69 | 399,93 | 0,12 | 0,07 | 1,74 | 8,14E-02 | 2,16E-01 |
| TPRG1 | 313,94 | 0,10 | 0,06 | 1,74 | 8,15E-02 | 2,16E-01 |
| SNAPIN | 407,25 | 0,04 | 0,02 | 1,74 | 8,15E-02 | 2,16E-01 |
| CUL4B | 2277,70 | -0,05 | 0,03 | -1,74 | 8,16E-02 | 2,16E-01 |
| NUFIP1 | 93,18 | 0,06 | 0,03 | 1,74 | 8,17E-02 | 2,17E-01 |
| DHRS4L2 | 173,83 | 0,04 | 0,02 | 1,74 | 8,18E-02 | 2,17E-01 |
| CSNK1E | 299,34 | 0,05 | 0,03 | 1,74 | 8,18E-02 | 2,17E-01 |
| CORIN | 91,51 | -0,18 | 0,10 | -1,74 | 8,18E-02 | 2,17E-01 |
| IRF8 | 2223,54 | 0,05 | 0,03 | 1,74 | 8,19E-02 | 2,17E-01 |
| TOM1 | 1007,31 | -0,06 | 0,04 | -1,74 | 8,19E-02 | 2,17E-01 |
| PAM | 1746,06 | -0,09 | 0,05 | -1,74 | 8,20E-02 | 2,17E-01 |
| ZNF500 | 237,32 | 0,08 | 0,05 | 1,74 | 8,22E-02 | 2,17E-01 |
| SPON2 | 654,38 | -0,10 | 0,06 | -1,74 | 8,23E-02 | 2,18E-01 |
| ZNF524 | 313,94 | 0,04 | 0,02 | 1,74 | 8,24E-02 | 2,18E-01 |
| TOP1MT | 227,38 | 0,07 | 0,04 | 1,74 | 8,24E-02 | 2,18E-01 |
| TAF9 | 404,00 | 0,06 | 0,03 | 1,74 | 8,25E-02 | 2,18E-01 |
| RGS19 | 1734,03 | -0,06 | 0,03 | -1,74 | 8,26E-02 | 2,18E-01 |
| MIIP | 925,01 | 0,05 | 0,03 | 1,74 | 8,27E-02 | 2,19E-01 |
| GRAP | 762,34 | 0,06 | 0,03 | 1,73 | 8,27E-02 | 2,19E-01 |
| PRKCI | 708,87 | 0,05 | 0,03 | 1,73 | 8,28E-02 | 2,19E-01 |
| CD2 | 1216,81 | 0,06 | 0,04 | 1,73 | 8,28E-02 | 2,19E-01 |
| MAN1A1 | 3933,32 | -0,05 | 0,03 | -1,73 | 8,28E-02 | 2,19E-01 |

| | | | | | | |
|---|---|---|---|---|---|---|
| PGAP1 | 457,45 | 0,07 | 0,04 | 1,73 | 8,29E-02 | 2,19E-01 |
| CPSF3 | 713,73 | 0,03 | 0,02 | 1,73 | 8,30E-02 | 2,19E-01 |
| PWWP2B | 280,22 | 0,05 | 0,03 | 1,73 | 8,32E-02 | 2,20E-01 |
| GOPC | 1695,69 | 0,06 | 0,03 | 1,73 | 8,34E-02 | 2,20E-01 |
| MKRN2 | 746,27 | -0,02 | 0,01 | -1,73 | 8,34E-02 | 2,20E-01 |
| DEXI | 548,35 | 0,03 | 0,02 | 1,73 | 8,34E-02 | 2,20E-01 |
| TATDN1 | 292,77 | 0,05 | 0,03 | 1,73 | 8,35E-02 | 2,20E-01 |
| TWF2 | 1698,23 | -0,04 | 0,02 | -1,73 | 8,36E-02 | 2,20E-01 |
| SCARB2 | 1640,64 | -0,04 | 0,02 | -1,73 | 8,36E-02 | 2,20E-01 |
| MRPS5 | 1282,96 | 0,02 | 0,01 | 1,73 | 8,37E-02 | 2,21E-01 |
| RNF138 | 1246,31 | 0,07 | 0,04 | 1,73 | 8,38E-02 | 2,21E-01 |
| AASDHPPT | 1057,30 | 0,06 | 0,03 | 1,73 | 8,40E-02 | 2,21E-01 |
| DSEL | 160,13 | 0,10 | 0,06 | 1,73 | 8,40E-02 | 2,21E-01 |
| ENSG000002 | 98,20 | -0,12 | 0,07 | -1,73 | 8,40E-02 | 2,21E-01 |
| ZNF583 | 232,62 | 0,08 | 0,05 | 1,73 | 8,41E-02 | 2,21E-01 |
| TMEM168 | 1892,95 | 0,05 | 0,03 | 1,73 | 8,41E-02 | 2,21E-01 |
| MPO | 147,95 | 0,14 | 0,08 | 1,73 | 8,42E-02 | 2,21E-01 |
| RAB11B | 447,22 | 0,09 | 0,05 | 1,73 | 8,43E-02 | 2,22E-01 |
| MPHOSPH9 | 1391,60 | 0,04 | 0,02 | 1,73 | 8,43E-02 | 2,22E-01 |
| HIP1R | 914,75 | 0,06 | 0,03 | 1,73 | 8,44E-02 | 2,22E-01 |
| HAUS2 | 497,75 | 0,04 | 0,02 | 1,73 | 8,44E-02 | 2,22E-01 |
| HLTF | 687,04 | 0,06 | 0,04 | 1,73 | 8,45E-02 | 2,22E-01 |
| PXN | 2492,76 | -0,12 | 0,07 | -1,73 | 8,45E-02 | 2,22E-01 |
| EXOC8 | 2543,05 | -0,04 | 0,03 | -1,73 | 8,45E-02 | 2,22E-01 |
| TMEM208 | 415,93 | 0,04 | 0,02 | 1,72 | 8,46E-02 | 2,22E-01 |
| PTGR3 | 2352,11 | 0,03 | 0,02 | 1,72 | 8,46E-02 | 2,22E-01 |
| TACC3 | 1674,26 | -0,08 | 0,05 | -1,72 | 8,46E-02 | 2,22E-01 |
| OTUD4 | 2181,39 | 0,03 | 0,02 | 1,72 | 8,47E-02 | 2,22E-01 |
| NUDCD1 | 468,33 | 0,10 | 0,06 | 1,72 | 8,47E-02 | 2,22E-01 |
| NCKAP1L | 8291,90 | -0,04 | 0,02 | -1,72 | 8,47E-02 | 2,22E-01 |
| ANKRA2 | 958,13 | 0,05 | 0,03 | 1,72 | 8,49E-02 | 2,23E-01 |
| KLHDC8B | 243,48 | -0,08 | 0,05 | -1,72 | 8,50E-02 | 2,23E-01 |
| HMGCS1 | 751,43 | 0,04 | 0,03 | 1,72 | 8,51E-02 | 2,23E-01 |
| TMEM177 | 138,76 | 0,05 | 0,03 | 1,72 | 8,52E-02 | 2,23E-01 |
| ANAPC13 | 707,52 | -0,06 | 0,03 | -1,72 | 8,53E-02 | 2,23E-01 |
| PTPDC1 | 289,31 | 0,05 | 0,03 | 1,72 | 8,53E-02 | 2,23E-01 |
| C19orf38 | 912,05 | -0,06 | 0,03 | -1,72 | 8,55E-02 | 2,24E-01 |
| BOD1 | 500,34 | 0,04 | 0,02 | 1,72 | 8,56E-02 | 2,24E-01 |
| PYGM | 151,14 | -0,09 | 0,05 | -1,72 | 8,56E-02 | 2,24E-01 |
| SRPRA | 1637,65 | -0,05 | 0,03 | -1,72 | 8,56E-02 | 2,24E-01 |
| IPO7 | 2000,53 | 0,05 | 0,03 | 1,72 | 8,57E-02 | 2,24E-01 |
| IGSF8 | 246,16 | 0,08 | 0,05 | 1,72 | 8,57E-02 | 2,24E-01 |
| DTYMK | 180,46 | 0,04 | 0,02 | 1,72 | 8,58E-02 | 2,24E-01 |
| ATAD1 | 1480,75 | 0,07 | 0,04 | 1,72 | 8,59E-02 | 2,24E-01 |
| ZHX1 | 859,23 | 0,07 | 0,04 | 1,72 | 8,60E-02 | 2,25E-01 |
| TM2D2 | 523,40 | 0,04 | 0,02 | 1,72 | 8,61E-02 | 2,25E-01 |

| | | | | | | |
|---|---|---|---|---|---|---|
| SLC25A26 | 652,07 | 0,04 | 0,02 | 1,72 | 8,61E-02 | 2,25E-01 |
| GPR162 | 134,15 | 0,10 | 0,06 | 1,72 | 8,63E-02 | 2,25E-01 |
| RNLS | 91,87 | 0,05 | 0,03 | 1,71 | 8,64E-02 | 2,25E-01 |
| EMC6 | 187,30 | 0,05 | 0,03 | 1,71 | 8,64E-02 | 2,25E-01 |
| TSPAN13 | 181,42 | 0,08 | 0,05 | 1,71 | 8,64E-02 | 2,25E-01 |
| ARMCX5 | 363,44 | 0,04 | 0,02 | 1,71 | 8,65E-02 | 2,26E-01 |
| KLHL24 | 4257,57 | -0,03 | 0,02 | -1,71 | 8,66E-02 | 2,26E-01 |
| DPYD | 11009,99 | -0,05 | 0,03 | -1,71 | 8,66E-02 | 2,26E-01 |
| TCTN3 | 233,25 | 0,04 | 0,02 | 1,71 | 8,67E-02 | 2,26E-01 |
| ITGAE | 390,75 | 0,03 | 0,02 | 1,71 | 8,67E-02 | 2,26E-01 |
| BCL2L11 | 1741,01 | -0,05 | 0,03 | -1,71 | 8,68E-02 | 2,26E-01 |
| EHD1 | 2484,16 | -0,06 | 0,04 | -1,71 | 8,69E-02 | 2,26E-01 |
| CERS5 | 919,65 | -0,03 | 0,02 | -1,71 | 8,69E-02 | 2,26E-01 |
| ZNF77 | 110,54 | 0,04 | 0,02 | 1,71 | 8,69E-02 | 2,26E-01 |
| NSF | 804,18 | -0,03 | 0,02 | -1,71 | 8,71E-02 | 2,26E-01 |
| LAGE3 | 268,90 | 0,05 | 0,03 | 1,71 | 8,70E-02 | 2,26E-01 |
| GPRIN3 | 4571,29 | 0,06 | 0,03 | 1,71 | 8,70E-02 | 2,26E-01 |
| SRSF6 | 4102,45 | 0,04 | 0,02 | 1,71 | 8,72E-02 | 2,27E-01 |
| MTMR6 | 2798,29 | -0,05 | 0,03 | -1,71 | 8,73E-02 | 2,27E-01 |
| NACC2 | 1619,08 | -0,07 | 0,04 | -1,71 | 8,73E-02 | 2,27E-01 |
| BANK1 | 1636,10 | 0,09 | 0,05 | 1,71 | 8,73E-02 | 2,27E-01 |
| SNU13 | 1106,33 | 0,03 | 0,02 | 1,71 | 8,74E-02 | 2,27E-01 |
| ENSG00000２ | 87,87 | -0,07 | 0,04 | -1,71 | 8,75E-02 | 2,27E-01 |
| TMEM120B | 131,42 | 0,08 | 0,05 | 1,71 | 8,75E-02 | 2,27E-01 |
| AP5S1 | 223,30 | 0,04 | 0,03 | 1,71 | 8,77E-02 | 2,28E-01 |
| CCDC134 | 128,26 | 0,07 | 0,04 | 1,71 | 8,77E-02 | 2,28E-01 |
| AKIRIN2 | 1058,44 | -0,04 | 0,02 | -1,71 | 8,77E-02 | 2,28E-01 |
| BAZ2A | 3146,25 | -0,08 | 0,05 | -1,71 | 8,78E-02 | 2,28E-01 |
| TCEAL8 | 261,70 | 0,09 | 0,05 | 1,71 | 8,78E-02 | 2,28E-01 |
| ZDHHC23 | 286,13 | 0,06 | 0,03 | 1,71 | 8,79E-02 | 2,28E-01 |
| VPS13D | 5827,56 | -0,06 | 0,03 | -1,71 | 8,79E-02 | 2,28E-01 |
| NBPF14 | 1689,97 | -0,06 | 0,04 | -1,71 | 8,80E-02 | 2,28E-01 |
| TMEM126B | 537,42 | 0,06 | 0,04 | 1,71 | 8,79E-02 | 2,28E-01 |
| LPIN2 | 4649,53 | -0,06 | 0,03 | -1,71 | 8,80E-02 | 2,28E-01 |
| NANS | 537,25 | -0,04 | 0,02 | -1,71 | 8,81E-02 | 2,28E-01 |
| CDK12 | 3655,71 | -0,03 | 0,02 | -1,71 | 8,82E-02 | 2,28E-01 |
| NLRP3 | 1503,56 | -0,06 | 0,03 | -1,70 | 8,83E-02 | 2,28E-01 |
| LCOR | 6494,59 | -0,03 | 0,02 | -1,70 | 8,83E-02 | 2,28E-01 |
| POLI | 1189,56 | 0,05 | 0,03 | 1,70 | 8,83E-02 | 2,28E-01 |
| GATAD2B | 1557,72 | -0,08 | 0,05 | -1,70 | 8,86E-02 | 2,29E-01 |
| WDR45B | 1311,35 | -0,02 | 0,01 | -1,70 | 8,86E-02 | 2,29E-01 |
| UCHL5 | 1283,44 | 0,07 | 0,04 | 1,70 | 8,86E-02 | 2,29E-01 |
| LIN37 | 178,83 | -0,04 | 0,02 | -1,70 | 8,86E-02 | 2,29E-01 |
| ATP5MJ | 441,61 | 0,08 | 0,05 | 1,70 | 8,86E-02 | 2,29E-01 |
| CBY1 | 229,80 | 0,03 | 0,02 | 1,70 | 8,87E-02 | 2,29E-01 |
| MYO1E | 437,88 | 0,08 | 0,04 | 1,70 | 8,88E-02 | 2,29E-01 |

| | | | | | | |
|---|---|---|---|---|---|---|
| ZNF790 | 247,86 | 0,06 | 0,04 | 1,70 | 8,88E-02 | 2,29E-01 |
| SETD5 | 3707,48 | -0,05 | 0,03 | -1,70 | 8,88E-02 | 2,29E-01 |
| RMND1 | 377,24 | 0,04 | 0,02 | 1,70 | 8,88E-02 | 2,29E-01 |
| ZSWIM3 | 94,72 | -0,04 | 0,03 | -1,70 | 8,89E-02 | 2,30E-01 |
| NIPA2 | 1372,13 | -0,03 | 0,02 | -1,70 | 8,89E-02 | 2,30E-01 |
| H2AX | 154,64 | 0,08 | 0,05 | 1,70 | 8,90E-02 | 2,30E-01 |
| GNB4 | 3735,65 | -0,06 | 0,04 | -1,70 | 8,90E-02 | 2,30E-01 |
| MYO1C | 189,42 | 0,09 | 0,05 | 1,70 | 8,91E-02 | 2,30E-01 |
| DNAJB1 | 1429,32 | -0,03 | 0,02 | -1,70 | 8,91E-02 | 2,30E-01 |
| ZNF493 | 2395,23 | -0,05 | 0,03 | -1,70 | 8,94E-02 | 2,30E-01 |
| GOLGA5 | 1053,73 | -0,03 | 0,02 | -1,70 | 8,94E-02 | 2,30E-01 |
| ANAPC7 | 945,91 | -0,02 | 0,01 | -1,70 | 8,94E-02 | 2,30E-01 |
| POLG2 | 659,64 | 0,03 | 0,02 | 1,70 | 8,94E-02 | 2,30E-01 |
| STRAP | 539,49 | 0,05 | 0,03 | 1,70 | 8,94E-02 | 2,30E-01 |
| SHFL | 542,23 | -0,06 | 0,04 | -1,70 | 8,95E-02 | 2,30E-01 |
| ZNF597 | 416,39 | 0,05 | 0,03 | 1,70 | 8,95E-02 | 2,30E-01 |
| TROAP | 185,94 | 0,12 | 0,07 | 1,70 | 8,95E-02 | 2,30E-01 |
| SSR3 | 2747,18 | 0,05 | 0,03 | 1,70 | 8,96E-02 | 2,31E-01 |
| USP18 | 114,17 | -0,15 | 0,09 | -1,70 | 8,96E-02 | 2,31E-01 |
| TRMU | 909,30 | 0,03 | 0,02 | 1,70 | 8,98E-02 | 2,31E-01 |
| DDX3Y | 2392,11 | 0,18 | 0,11 | 1,70 | 8,98E-02 | 2,31E-01 |
| CASP10 | 2236,70 | -0,04 | 0,02 | -1,70 | 8,98E-02 | 2,31E-01 |
| GDPD3 | 104,25 | -0,11 | 0,06 | -1,70 | 8,98E-02 | 2,31E-01 |
| XRN1 | 8179,00 | -0,05 | 0,03 | -1,70 | 8,99E-02 | 2,31E-01 |
| SNW1 | 1523,95 | -0,04 | 0,03 | -1,70 | 9,00E-02 | 2,31E-01 |
| VWDE | 172,92 | 0,16 | 0,09 | 1,70 | 9,00E-02 | 2,31E-01 |
| TPCN2 | 862,23 | -0,09 | 0,05 | -1,70 | 9,00E-02 | 2,31E-01 |
| MMAA | 350,01 | 0,05 | 0,03 | 1,70 | 9,00E-02 | 2,31E-01 |
| C16orf87 | 325,63 | 0,04 | 0,02 | 1,70 | 9,01E-02 | 2,31E-01 |
| SLC43A2 | 3215,57 | -0,08 | 0,05 | -1,69 | 9,01E-02 | 2,31E-01 |
| CDC123 | 1858,48 | -0,04 | 0,02 | -1,69 | 9,03E-02 | 2,32E-01 |
| ERP29 | 759,56 | 0,04 | 0,02 | 1,69 | 9,03E-02 | 2,32E-01 |
| COA6 | 172,48 | 0,06 | 0,04 | 1,69 | 9,03E-02 | 2,32E-01 |
| ENSG000002 | 119,68 | 0,09 | 0,05 | 1,69 | 9,03E-02 | 2,32E-01 |
| SKIC3 | 3009,83 | 0,05 | 0,03 | 1,69 | 9,04E-02 | 2,32E-01 |
| TMA7 | 788,65 | 0,13 | 0,08 | 1,69 | 9,04E-02 | 2,32E-01 |
| CHIC2 | 2237,03 | -0,05 | 0,03 | -1,69 | 9,06E-02 | 2,32E-01 |
| MIOS | 1147,24 | 0,05 | 0,03 | 1,69 | 9,07E-02 | 2,32E-01 |
| CRYGS | 183,80 | 0,04 | 0,02 | 1,69 | 9,08E-02 | 2,32E-01 |
| AP2S1 | 1081,61 | 0,06 | 0,03 | 1,69 | 9,08E-02 | 2,32E-01 |
| CLEC10A | 187,55 | 0,08 | 0,05 | 1,69 | 9,08E-02 | 2,32E-01 |
| AMPD2 | 2870,30 | -0,08 | 0,05 | -1,69 | 9,09E-02 | 2,33E-01 |
| MYDGF | 605,15 | 0,05 | 0,03 | 1,69 | 9,09E-02 | 2,33E-01 |
| TMPRSS9 | 409,01 | 0,15 | 0,09 | 1,69 | 9,11E-02 | 2,33E-01 |
| FAAP100 | 277,74 | 0,09 | 0,05 | 1,69 | 9,11E-02 | 2,33E-01 |
| PPP2R2A | 1933,54 | -0,04 | 0,02 | -1,69 | 9,11E-02 | 2,33E-01 |

| | | | | | | |
|---|---|---|---|---|---|---|
| THAP12 | 1790,97 | 0,07 | 0,04 | 1,69 | 9,11E-02 | 2,33E-01 |
| LPAR2 | 625,64 | -0,11 | 0,07 | -1,69 | 9,11E-02 | 2,33E-01 |
| RPRD1B | 700,81 | -0,05 | 0,03 | -1,69 | 9,12E-02 | 2,33E-01 |
| SYNJ2BP | 1051,83 | 0,04 | 0,03 | 1,69 | 9,12E-02 | 2,33E-01 |
| PTPRA | 1606,10 | 0,05 | 0,03 | 1,69 | 9,13E-02 | 2,33E-01 |
| GTF2F1 | 932,41 | -0,04 | 0,02 | -1,69 | 9,14E-02 | 2,33E-01 |
| WDFY1 | 2681,74 | -0,05 | 0,03 | -1,69 | 9,13E-02 | 2,33E-01 |
| XPO1 | 5400,15 | 0,04 | 0,02 | 1,69 | 9,14E-02 | 2,33E-01 |
| ZNF384 | 867,83 | -0,04 | 0,02 | -1,69 | 9,14E-02 | 2,33E-01 |
| CD300C | 283,22 | 0,06 | 0,04 | 1,69 | 9,14E-02 | 2,33E-01 |
| EBPL | 223,18 | 0,05 | 0,03 | 1,69 | 9,15E-02 | 2,33E-01 |
| ARHGEF12 | 3716,76 | 0,09 | 0,06 | 1,69 | 9,16E-02 | 2,33E-01 |
| BCAP31 | 1166,30 | -0,03 | 0,02 | -1,69 | 9,16E-02 | 2,33E-01 |
| TAFA2 | 1080,55 | -0,07 | 0,04 | -1,69 | 9,17E-02 | 2,34E-01 |
| ZNF345 | 327,38 | 0,04 | 0,02 | 1,69 | 9,17E-02 | 2,34E-01 |
| EPAS1 | 136,73 | -0,10 | 0,06 | -1,69 | 9,17E-02 | 2,34E-01 |
| ZNF540 | 328,12 | 0,06 | 0,04 | 1,69 | 9,18E-02 | 2,34E-01 |
| KCTD6 | 264,85 | 0,03 | 0,02 | 1,69 | 9,18E-02 | 2,34E-01 |
| STIM1 | 1889,27 | -0,06 | 0,03 | -1,69 | 9,19E-02 | 2,34E-01 |
| POLR3C | 438,62 | 0,02 | 0,01 | 1,68 | 9,22E-02 | 2,35E-01 |
| GLMN | 436,90 | 0,05 | 0,03 | 1,68 | 9,22E-02 | 2,35E-01 |
| TMLHE | 882,19 | -0,04 | 0,02 | -1,68 | 9,23E-02 | 2,35E-01 |
| PDZD8 | 3599,48 | -0,06 | 0,04 | -1,68 | 9,23E-02 | 2,35E-01 |
| TYK2 | 5358,25 | -0,05 | 0,03 | -1,68 | 9,22E-02 | 2,35E-01 |
| LRRC14 | 384,04 | 0,07 | 0,04 | 1,68 | 9,23E-02 | 2,35E-01 |
| FANCL | 226,04 | 0,08 | 0,05 | 1,68 | 9,24E-02 | 2,35E-01 |
| LIAS | 492,95 | 0,03 | 0,02 | 1,68 | 9,25E-02 | 2,35E-01 |
| B3GALNT2 | 367,86 | -0,05 | 0,03 | -1,68 | 9,25E-02 | 2,35E-01 |
| MOV10 | 529,95 | -0,08 | 0,05 | -1,68 | 9,25E-02 | 2,35E-01 |
| ZNG1A | 864,65 | 0,05 | 0,03 | 1,68 | 9,26E-02 | 2,35E-01 |
| RELT | 3605,85 | -0,07 | 0,04 | -1,68 | 9,27E-02 | 2,35E-01 |
| EMP3 | 2738,55 | 0,05 | 0,03 | 1,68 | 9,27E-02 | 2,35E-01 |
| PASK | 1144,70 | 0,06 | 0,03 | 1,68 | 9,29E-02 | 2,36E-01 |
| NUP37 | 317,01 | 0,05 | 0,03 | 1,68 | 9,30E-02 | 2,36E-01 |
| OXCT1 | 887,17 | 0,05 | 0,03 | 1,68 | 9,30E-02 | 2,36E-01 |
| UNC93B1 | 1909,52 | -0,06 | 0,04 | -1,68 | 9,31E-02 | 2,36E-01 |
| VPS13C | 19567,26 | 0,05 | 0,03 | 1,68 | 9,32E-02 | 2,36E-01 |
| CCDC17 | 232,97 | -0,08 | 0,05 | -1,68 | 9,32E-02 | 2,36E-01 |
| C6orf163 | 361,64 | -0,05 | 0,03 | -1,68 | 9,32E-02 | 2,36E-01 |
| TPRA1 | 438,71 | 0,03 | 0,02 | 1,68 | 9,33E-02 | 2,36E-01 |
| MGAT2 | 1724,91 | 0,05 | 0,03 | 1,68 | 9,33E-02 | 2,36E-01 |
| CCNG1 | 1565,18 | 0,06 | 0,04 | 1,68 | 9,33E-02 | 2,36E-01 |
| C9orf85 | 329,49 | 0,03 | 0,02 | 1,68 | 9,34E-02 | 2,36E-01 |
| PEAK1 | 1002,07 | -0,04 | 0,02 | -1,68 | 9,35E-02 | 2,37E-01 |
| DNASE1 | 2109,62 | -0,04 | 0,02 | -1,68 | 9,36E-02 | 2,37E-01 |
| USP9Y | 2002,63 | 0,19 | 0,11 | 1,68 | 9,37E-02 | 2,37E-01 |

| | | | | | | |
|---|---|---|---|---|---|---|
| HLA-DOA | 916,22 | 0,08 | 0,04 | 1,68 | 9,38E-02 | 2,37E-01 |
| TMEM273 | 1273,51 | -0,07 | 0,04 | -1,68 | 9,39E-02 | 2,37E-01 |
| SIGLEC8 | 131,28 | 0,14 | 0,08 | 1,68 | 9,38E-02 | 2,37E-01 |
| DRG2 | 615,11 | 0,05 | 0,03 | 1,68 | 9,38E-02 | 2,37E-01 |
| TMEM33 | 4661,54 | -0,05 | 0,03 | -1,68 | 9,38E-02 | 2,37E-01 |
| NDFIP2 | 166,65 | 0,08 | 0,05 | 1,67 | 9,39E-02 | 2,37E-01 |
| KRBA2 | 132,17 | 0,05 | 0,03 | 1,68 | 9,39E-02 | 2,37E-01 |
| ALG9 | 207,03 | 0,04 | 0,03 | 1,67 | 9,40E-02 | 2,37E-01 |
| SIGIRR | 767,41 | 0,07 | 0,04 | 1,67 | 9,40E-02 | 2,37E-01 |
| SLX9 | 239,24 | 0,03 | 0,02 | 1,67 | 9,40E-02 | 2,37E-01 |
| AKAP1 | 542,66 | 0,04 | 0,02 | 1,67 | 9,41E-02 | 2,37E-01 |
| CGGBP1 | 4912,94 | 0,06 | 0,03 | 1,67 | 9,41E-02 | 2,37E-01 |
| EDF1 | 2263,17 | 0,04 | 0,02 | 1,67 | 9,41E-02 | 2,37E-01 |
| MARCHF7 | 8641,22 | -0,05 | 0,03 | -1,67 | 9,42E-02 | 2,38E-01 |
| JPT2 | 528,17 | 0,03 | 0,02 | 1,67 | 9,42E-02 | 2,38E-01 |
| MRPL46 | 345,39 | 0,04 | 0,02 | 1,67 | 9,43E-02 | 2,38E-01 |
| IL32 | 670,10 | 0,07 | 0,04 | 1,67 | 9,43E-02 | 2,38E-01 |
| CLPB | 765,08 | 0,04 | 0,02 | 1,67 | 9,43E-02 | 2,38E-01 |
| APH1B | 1244,53 | -0,03 | 0,02 | -1,67 | 9,45E-02 | 2,38E-01 |
| UFD1 | 906,76 | -0,03 | 0,02 | -1,67 | 9,44E-02 | 2,38E-01 |
| IRAK4 | 3261,69 | -0,03 | 0,02 | -1,67 | 9,44E-02 | 2,38E-01 |
| RORA | 8511,59 | 0,06 | 0,04 | 1,67 | 9,44E-02 | 2,38E-01 |
| IMMP2L | 528,22 | 0,06 | 0,04 | 1,67 | 9,46E-02 | 2,38E-01 |
| ASB2 | 123,85 | 0,10 | 0,06 | 1,67 | 9,47E-02 | 2,38E-01 |
| NFYB | 462,00 | 0,08 | 0,05 | 1,67 | 9,48E-02 | 2,38E-01 |
| KCTD15 | 311,66 | 0,06 | 0,04 | 1,67 | 9,48E-02 | 2,38E-01 |
| ZFP62 | 1277,12 | 0,06 | 0,03 | 1,67 | 9,49E-02 | 2,38E-01 |
| NOTCH2NLA | 541,53 | -0,07 | 0,04 | -1,67 | 9,49E-02 | 2,38E-01 |
| PDGFB | 77,01 | 0,10 | 0,06 | 1,67 | 9,51E-02 | 2,39E-01 |
| SLX4IP | 509,84 | -0,04 | 0,02 | -1,67 | 9,52E-02 | 2,39E-01 |
| SMPD4 | 402,26 | 0,06 | 0,03 | 1,67 | 9,53E-02 | 2,39E-01 |
| ZNF691 | 289,17 | 0,03 | 0,02 | 1,67 | 9,53E-02 | 2,39E-01 |
| FBXO7 | 19363,83 | 0,10 | 0,06 | 1,67 | 9,53E-02 | 2,39E-01 |
| EAF2 | 426,13 | 0,07 | 0,04 | 1,67 | 9,54E-02 | 2,39E-01 |
| SEMA4B | 1149,55 | -0,08 | 0,05 | -1,67 | 9,54E-02 | 2,39E-01 |
| PAPSS2 | 142,07 | -0,08 | 0,05 | -1,67 | 9,55E-02 | 2,39E-01 |
| TIMM50 | 352,72 | 0,03 | 0,02 | 1,67 | 9,56E-02 | 2,40E-01 |
| KLF10 | 332,25 | 0,06 | 0,04 | 1,67 | 9,57E-02 | 2,40E-01 |
| ARL2BP | 1031,28 | 0,04 | 0,02 | 1,67 | 9,57E-02 | 2,40E-01 |
| CTSD | 1017,40 | -0,06 | 0,04 | -1,67 | 9,57E-02 | 2,40E-01 |
| SHKBP1 | 2060,23 | -0,09 | 0,05 | -1,67 | 9,57E-02 | 2,40E-01 |
| TFEB | 1011,82 | -0,07 | 0,04 | -1,67 | 9,58E-02 | 2,40E-01 |
| ASCC1 | 252,47 | -0,04 | 0,02 | -1,66 | 9,59E-02 | 2,40E-01 |
| HEMK1 | 1207,61 | 0,05 | 0,03 | 1,66 | 9,60E-02 | 2,40E-01 |
| MRPL39 | 292,97 | 0,05 | 0,03 | 1,66 | 9,61E-02 | 2,40E-01 |
| RFC4 | 167,42 | 0,04 | 0,03 | 1,66 | 9,61E-02 | 2,40E-01 |

| | | | | | | |
|---|---|---|---|---|---|---|
| DIAPH2 | 1584,40 | 0,04 | 0,02 | 1,66 | 9,61E-02 | 2,41E-01 |
| MECP2 | 2494,20 | -0,07 | 0,04 | -1,66 | 9,62E-02 | 2,41E-01 |
| SLC2A9 | 316,56 | -0,07 | 0,04 | -1,66 | 9,62E-02 | 2,41E-01 |
| HDAC11 | 138,46 | 0,06 | 0,04 | 1,66 | 9,62E-02 | 2,41E-01 |
| FCGBP | 246,57 | 0,13 | 0,08 | 1,66 | 9,62E-02 | 2,41E-01 |
| MPPE1 | 4151,12 | -0,05 | 0,03 | -1,66 | 9,65E-02 | 2,41E-01 |
| TWF1 | 1113,21 | 0,08 | 0,05 | 1,66 | 9,65E-02 | 2,41E-01 |
| EIF4A3 | 1135,08 | -0,04 | 0,02 | -1,66 | 9,66E-02 | 2,41E-01 |
| MAL | 317,14 | 0,07 | 0,04 | 1,66 | 9,66E-02 | 2,41E-01 |
| NRBP1 | 1165,97 | -0,04 | 0,02 | -1,66 | 9,66E-02 | 2,41E-01 |
| BIN1 | 1603,67 | 0,05 | 0,03 | 1,66 | 9,67E-02 | 2,41E-01 |
| BBOF1 | 517,31 | 0,11 | 0,07 | 1,66 | 9,67E-02 | 2,41E-01 |
| LRRC8D | 1208,53 | 0,03 | 0,02 | 1,66 | 9,68E-02 | 2,42E-01 |
| NSUN6 | 737,03 | 0,04 | 0,02 | 1,66 | 9,69E-02 | 2,42E-01 |
| FLCN | 1357,07 | -0,05 | 0,03 | -1,66 | 9,69E-02 | 2,42E-01 |
| BROX | 3329,94 | -0,04 | 0,03 | -1,66 | 9,70E-02 | 2,42E-01 |
| RUSF1 | 529,81 | 0,05 | 0,03 | 1,66 | 9,70E-02 | 2,42E-01 |
| ZNF248 | 495,07 | 0,04 | 0,03 | 1,66 | 9,71E-02 | 2,42E-01 |
| BBS4 | 391,18 | -0,03 | 0,02 | -1,66 | 9,72E-02 | 2,42E-01 |
| MAN2B2 | 1198,20 | -0,06 | 0,04 | -1,66 | 9,72E-02 | 2,42E-01 |
| SNX8 | 252,59 | 0,06 | 0,04 | 1,66 | 9,72E-02 | 2,42E-01 |
| FAM193A | 1046,97 | -0,07 | 0,04 | -1,66 | 9,72E-02 | 2,42E-01 |
| GTPBP4 | 821,76 | 0,03 | 0,02 | 1,66 | 9,72E-02 | 2,42E-01 |
| TMEM143 | 104,58 | 0,05 | 0,03 | 1,66 | 9,72E-02 | 2,42E-01 |
| DSE | 2139,42 | -0,05 | 0,03 | -1,66 | 9,72E-02 | 2,42E-01 |
| ABHD14B | 194,63 | 0,06 | 0,04 | 1,66 | 9,72E-02 | 2,42E-01 |
| KCTD11 | 121,79 | 0,05 | 0,03 | 1,66 | 9,73E-02 | 2,42E-01 |
| BCAT2 | 100,35 | 0,07 | 0,04 | 1,66 | 9,74E-02 | 2,42E-01 |
| LAMTOR4 | 2118,85 | -0,05 | 0,03 | -1,66 | 9,75E-02 | 2,43E-01 |
| TFE3 | 914,21 | -0,07 | 0,04 | -1,66 | 9,76E-02 | 2,43E-01 |
| FPGT | 627,10 | 0,08 | 0,05 | 1,66 | 9,76E-02 | 2,43E-01 |
| ATR | 3382,63 | 0,04 | 0,03 | 1,66 | 9,78E-02 | 2,43E-01 |
| DHX30 | 931,36 | 0,04 | 0,02 | 1,66 | 9,78E-02 | 2,43E-01 |
| SOX4 | 241,90 | 0,06 | 0,04 | 1,66 | 9,78E-02 | 2,43E-01 |
| TUSC2 | 317,18 | 0,03 | 0,02 | 1,66 | 9,79E-02 | 2,43E-01 |
| SBDS | 462,38 | 0,07 | 0,04 | 1,66 | 9,79E-02 | 2,43E-01 |
| AVEN | 166,70 | 0,04 | 0,02 | 1,65 | 9,79E-02 | 2,43E-01 |
| MRPL18 | 491,51 | 0,04 | 0,03 | 1,65 | 9,81E-02 | 2,43E-01 |
| ZBTB43 | 1084,85 | -0,02 | 0,01 | -1,65 | 9,81E-02 | 2,43E-01 |
| PAXBP1 | 2030,81 | 0,05 | 0,03 | 1,65 | 9,81E-02 | 2,43E-01 |
| CYP27A1 | 738,51 | -0,10 | 0,06 | -1,65 | 9,81E-02 | 2,43E-01 |
| PEF1 | 719,22 | -0,03 | 0,02 | -1,65 | 9,82E-02 | 2,44E-01 |
| AURKA | 129,02 | 0,09 | 0,05 | 1,65 | 9,82E-02 | 2,44E-01 |
| LANCL2 | 306,81 | 0,03 | 0,02 | 1,65 | 9,83E-02 | 2,44E-01 |
| KAT14 | 522,31 | 0,03 | 0,02 | 1,65 | 9,85E-02 | 2,44E-01 |
| HS1BP3 | 609,44 | -0,07 | 0,04 | -1,65 | 9,85E-02 | 2,44E-01 |

| | | | | | | |
|---|---|---|---|---|---|---|
| EIF3E | 4005,75 | 0,09 | 0,06 | 1,65 | 9,85E-02 | 2,44E-01 |
| ALDH3A2 | 1116,19 | 0,02 | 0,01 | 1,65 | 9,87E-02 | 2,44E-01 |
| CA5B | 1296,83 | 0,03 | 0,02 | 1,65 | 9,87E-02 | 2,44E-01 |
| MLLT10 | 1752,70 | 0,03 | 0,02 | 1,65 | 9,87E-02 | 2,44E-01 |
| TNFRSF10B | 2445,89 | -0,04 | 0,03 | -1,65 | 9,89E-02 | 2,45E-01 |
| TMEM129 | 323,09 | 0,05 | 0,03 | 1,65 | 9,89E-02 | 2,45E-01 |
| BCAS4 | 425,00 | 0,07 | 0,04 | 1,65 | 9,89E-02 | 2,45E-01 |
| KCNQ5 | 207,71 | 0,07 | 0,04 | 1,65 | 9,88E-02 | 2,45E-01 |
| MED8 | 772,57 | -0,02 | 0,01 | -1,65 | 9,89E-02 | 2,45E-01 |
| CD177 | 76,38 | -0,21 | 0,13 | -1,65 | 9,90E-02 | 2,45E-01 |
| RPN1 | 4466,64 | -0,04 | 0,02 | -1,65 | 9,91E-02 | 2,45E-01 |
| POGK | 1381,45 | 0,02 | 0,01 | 1,65 | 9,91E-02 | 2,45E-01 |
| NLN | 310,48 | 0,06 | 0,04 | 1,65 | 9,92E-02 | 2,45E-01 |
| STING1 | 494,31 | 0,06 | 0,04 | 1,65 | 9,92E-02 | 2,45E-01 |
| WDR37 | 2634,90 | -0,03 | 0,02 | -1,65 | 9,93E-02 | 2,45E-01 |
| CYB5R3 | 297,05 | 0,09 | 0,05 | 1,65 | 9,93E-02 | 2,45E-01 |
| RHPN1 | 107,69 | 0,09 | 0,06 | 1,65 | 9,94E-02 | 2,45E-01 |
| ICAM1 | 634,99 | -0,06 | 0,04 | -1,65 | 9,95E-02 | 2,45E-01 |
| TAS2R50 | 96,45 | 0,08 | 0,05 | 1,65 | 9,95E-02 | 2,46E-01 |
| ZNF460 | 2781,41 | 0,03 | 0,02 | 1,65 | 9,96E-02 | 2,46E-01 |
| EARS2 | 385,98 | 0,03 | 0,02 | 1,65 | 9,97E-02 | 2,46E-01 |
| ZFY | 553,42 | 0,12 | 0,07 | 1,65 | 9,99E-02 | 2,46E-01 |
| PAWR | 318,39 | 0,09 | 0,05 | 1,65 | 9,99E-02 | 2,46E-01 |
| RBM15B | 1436,86 | 0,02 | 0,01 | 1,64 | 1,00E-01 | 2,47E-01 |
| SWT1 | 871,61 | 0,09 | 0,06 | 1,64 | 1,00E-01 | 2,47E-01 |
| GLRX2 | 625,71 | 0,04 | 0,02 | 1,64 | 1,00E-01 | 2,47E-01 |
| CNR2 | 329,46 | 0,08 | 0,05 | 1,64 | 1,00E-01 | 2,47E-01 |
| ZNF84 | 1003,70 | 0,06 | 0,04 | 1,64 | 1,00E-01 | 2,47E-01 |
| GNPTAB | 2564,07 | -0,04 | 0,02 | -1,64 | 1,00E-01 | 2,47E-01 |
| ZNF383 | 803,29 | 0,03 | 0,02 | 1,64 | 1,01E-01 | 2,47E-01 |
| DDX20 | 703,47 | 0,04 | 0,02 | 1,64 | 1,01E-01 | 2,47E-01 |
| TAF11 | 696,80 | 0,03 | 0,02 | 1,64 | 1,01E-01 | 2,48E-01 |
| MAP3K8 | 1674,68 | -0,03 | 0,02 | -1,64 | 1,01E-01 | 2,48E-01 |
| SLC35A1 | 208,90 | 0,09 | 0,05 | 1,64 | 1,01E-01 | 2,48E-01 |
| STAT6 | 7970,23 | -0,06 | 0,03 | -1,64 | 1,01E-01 | 2,48E-01 |
| QRSL1 | 709,08 | 0,05 | 0,03 | 1,64 | 1,01E-01 | 2,48E-01 |
| TRMT11 | 605,47 | 0,05 | 0,03 | 1,64 | 1,01E-01 | 2,48E-01 |
| KLHL12 | 1368,53 | -0,03 | 0,02 | -1,64 | 1,01E-01 | 2,48E-01 |
| ANKHD1-EIF | 275,36 | -0,05 | 0,03 | -1,64 | 1,01E-01 | 2,48E-01 |
| ZNF254 | 649,11 | 0,07 | 0,04 | 1,64 | 1,01E-01 | 2,48E-01 |
| DTNB | 373,57 | -0,05 | 0,03 | -1,64 | 1,01E-01 | 2,48E-01 |
| TYW5 | 809,50 | 0,04 | 0,02 | 1,64 | 1,01E-01 | 2,48E-01 |
| KLC4 | 289,14 | -0,05 | 0,03 | -1,64 | 1,01E-01 | 2,49E-01 |
| GRIK4 | 86,79 | -0,24 | 0,15 | -1,64 | 1,01E-01 | 2,49E-01 |
| PEMT | 96,94 | 0,06 | 0,03 | 1,64 | 1,01E-01 | 2,49E-01 |
| CNEP1R1 | 1464,85 | -0,05 | 0,03 | -1,64 | 1,02E-01 | 2,49E-01 |

| | | | | | | |
|---|---|---|---|---|---|---|
| PCGF6 | 254,53 | 0,05 | 0,03 | 1,64 | 1,02E-01 | 2,49E-01 |
| FAM200A | 309,55 | 0,05 | 0,03 | 1,64 | 1,02E-01 | 2,49E-01 |
| ESYT2 | 4565,70 | 0,03 | 0,02 | 1,64 | 1,02E-01 | 2,49E-01 |
| TMTC2 | 904,40 | 0,05 | 0,03 | 1,64 | 1,02E-01 | 2,49E-01 |
| FASN | 179,67 | 0,13 | 0,08 | 1,64 | 1,02E-01 | 2,49E-01 |
| EIF3I | 1179,60 | 0,04 | 0,03 | 1,64 | 1,02E-01 | 2,49E-01 |
| TMED7 | 1883,42 | 0,07 | 0,04 | 1,64 | 1,02E-01 | 2,49E-01 |
| PTTG1 | 94,54 | 0,07 | 0,04 | 1,64 | 1,02E-01 | 2,50E-01 |
| PIGP | 246,38 | 0,05 | 0,03 | 1,64 | 1,02E-01 | 2,50E-01 |
| PLCG1 | 2804,44 | 0,05 | 0,03 | 1,63 | 1,02E-01 | 2,50E-01 |
| S100P | 1918,87 | -0,15 | 0,09 | -1,63 | 1,02E-01 | 2,50E-01 |
| SCLT1 | 5151,22 | -0,06 | 0,04 | -1,63 | 1,02E-01 | 2,50E-01 |
| RIC8A | 1686,35 | -0,03 | 0,02 | -1,63 | 1,02E-01 | 2,50E-01 |
| ATXN7L3B | 1840,65 | 0,03 | 0,02 | 1,63 | 1,02E-01 | 2,50E-01 |
| CNKSR2 | 576,63 | 0,07 | 0,04 | 1,63 | 1,02E-01 | 2,50E-01 |
| NDUFB5 | 888,69 | 0,05 | 0,03 | 1,63 | 1,02E-01 | 2,50E-01 |
| GMEB1 | 734,02 | -0,02 | 0,01 | -1,63 | 1,03E-01 | 2,51E-01 |
| CDC26 | 343,33 | -0,05 | 0,03 | -1,63 | 1,03E-01 | 2,51E-01 |
| TFAP4 | 173,40 | 0,07 | 0,05 | 1,63 | 1,03E-01 | 2,51E-01 |
| CDK5RAP3 | 1152,80 | -0,04 | 0,03 | -1,63 | 1,03E-01 | 2,51E-01 |
| PRIMPOL | 451,90 | 0,04 | 0,02 | 1,63 | 1,03E-01 | 2,51E-01 |
| MTM1 | 1270,44 | -0,03 | 0,02 | -1,63 | 1,03E-01 | 2,51E-01 |
| ZSCAN2 | 280,13 | 0,03 | 0,02 | 1,63 | 1,03E-01 | 2,51E-01 |
| GRB10 | 368,63 | -0,09 | 0,06 | -1,63 | 1,03E-01 | 2,51E-01 |
| NUP42 | 935,31 | 0,03 | 0,02 | 1,63 | 1,03E-01 | 2,51E-01 |
| HMGN1 | 2944,77 | 0,05 | 0,03 | 1,63 | 1,03E-01 | 2,51E-01 |
| GNB1 | 3715,01 | -0,05 | 0,03 | -1,63 | 1,03E-01 | 2,51E-01 |
| NEK4 | 910,60 | 0,02 | 0,01 | 1,63 | 1,03E-01 | 2,51E-01 |
| HELLS | 478,73 | 0,06 | 0,03 | 1,63 | 1,03E-01 | 2,51E-01 |
| NUDT16L1 | 475,57 | 0,03 | 0,02 | 1,63 | 1,03E-01 | 2,51E-01 |
| REV3L | 2668,20 | -0,03 | 0,02 | -1,63 | 1,03E-01 | 2,51E-01 |
| CTDSP2 | 4467,31 | -0,07 | 0,04 | -1,63 | 1,03E-01 | 2,52E-01 |
| UBXN2A | 490,67 | 0,03 | 0,02 | 1,63 | 1,03E-01 | 2,52E-01 |
| VKORC1 | 160,25 | 0,04 | 0,02 | 1,63 | 1,03E-01 | 2,52E-01 |
| EMP1 | 82,00 | 0,13 | 0,08 | 1,63 | 1,03E-01 | 2,52E-01 |
| FLYWCH2 | 168,46 | 0,04 | 0,02 | 1,63 | 1,04E-01 | 2,52E-01 |
| AATK | 1060,89 | -0,11 | 0,07 | -1,63 | 1,04E-01 | 2,52E-01 |
| SLC9B2 | 283,92 | 0,04 | 0,03 | 1,63 | 1,04E-01 | 2,52E-01 |
| TBC1D22A | 2503,78 | -0,04 | 0,03 | -1,63 | 1,04E-01 | 2,52E-01 |
| LACTB2 | 279,03 | 0,05 | 0,03 | 1,63 | 1,04E-01 | 2,52E-01 |
| P2RY11 | 152,51 | 0,06 | 0,03 | 1,63 | 1,04E-01 | 2,52E-01 |
| KIF21A | 559,11 | 0,07 | 0,04 | 1,63 | 1,04E-01 | 2,52E-01 |
| THOC1 | 1223,72 | 0,04 | 0,03 | 1,63 | 1,04E-01 | 2,52E-01 |
| EPHB4 | 243,20 | -0,11 | 0,07 | -1,63 | 1,04E-01 | 2,52E-01 |
| RHOH | 3039,44 | 0,04 | 0,03 | 1,63 | 1,04E-01 | 2,52E-01 |
| HSF2 | 405,19 | 0,06 | 0,04 | 1,63 | 1,04E-01 | 2,53E-01 |

| | | | | | | |
|---|---|---|---|---|---|---|
| DR1 | 4317,38 | 0,04 | 0,02 | 1,63 | 1,04E-01 | 2,53E-01 |
| PPP3R1 | 3958,76 | 0,06 | 0,04 | 1,63 | 1,04E-01 | 2,53E-01 |
| ZNF581 | 125,22 | 0,05 | 0,03 | 1,62 | 1,04E-01 | 2,53E-01 |
| RAD18 | 486,53 | 0,03 | 0,02 | 1,62 | 1,04E-01 | 2,53E-01 |
| ZNF511 | 98,05 | 0,04 | 0,03 | 1,62 | 1,04E-01 | 2,53E-01 |
| ERCC2 | 161,34 | 0,07 | 0,04 | 1,62 | 1,04E-01 | 2,53E-01 |
| LIMA1 | 269,21 | 0,05 | 0,03 | 1,62 | 1,04E-01 | 2,53E-01 |
| BRD3 | 878,07 | -0,05 | 0,03 | -1,62 | 1,04E-01 | 2,53E-01 |
| RUFY1 | 2449,89 | -0,04 | 0,03 | -1,62 | 1,05E-01 | 2,53E-01 |
| MGAT1 | 4706,20 | -0,04 | 0,02 | -1,62 | 1,05E-01 | 2,53E-01 |
| POLR1H | 435,57 | 0,03 | 0,02 | 1,62 | 1,05E-01 | 2,53E-01 |
| CAMSAP1 | 841,93 | 0,03 | 0,02 | 1,62 | 1,05E-01 | 2,53E-01 |
| TUBG2 | 107,09 | 0,05 | 0,03 | 1,62 | 1,05E-01 | 2,53E-01 |
| SLC22A16 | 103,59 | 0,10 | 0,06 | 1,62 | 1,05E-01 | 2,54E-01 |
| CCDC15 | 239,42 | 0,07 | 0,04 | 1,62 | 1,05E-01 | 2,54E-01 |
| UBE2E1 | 1074,69 | 0,04 | 0,02 | 1,62 | 1,05E-01 | 2,54E-01 |
| TNFRSF25 | 1097,07 | 0,06 | 0,04 | 1,62 | 1,05E-01 | 2,54E-01 |
| CFAP68 | 84,36 | 0,07 | 0,04 | 1,62 | 1,05E-01 | 2,54E-01 |
| ZC3H12C | 207,55 | 0,07 | 0,04 | 1,62 | 1,05E-01 | 2,54E-01 |
| CD79B | 890,06 | 0,08 | 0,05 | 1,62 | 1,05E-01 | 2,54E-01 |
| IMPDH2 | 853,25 | 0,04 | 0,02 | 1,62 | 1,05E-01 | 2,54E-01 |
| ATP6V0E1 | 3578,04 | -0,05 | 0,03 | -1,62 | 1,05E-01 | 2,54E-01 |
| ERCC4 | 549,48 | 0,03 | 0,02 | 1,62 | 1,05E-01 | 2,54E-01 |
| CCDC115 | 863,55 | 0,03 | 0,02 | 1,62 | 1,05E-01 | 2,54E-01 |
| ZNF562 | 823,76 | 0,03 | 0,02 | 1,62 | 1,05E-01 | 2,54E-01 |
| DNAJC2 | 564,08 | 0,04 | 0,02 | 1,62 | 1,06E-01 | 2,55E-01 |
| ERLEC1 | 686,65 | 0,05 | 0,03 | 1,62 | 1,06E-01 | 2,55E-01 |
| CD28 | 2895,52 | 0,06 | 0,04 | 1,62 | 1,06E-01 | 2,55E-01 |
| ADAM10 | 9336,82 | -0,04 | 0,03 | -1,62 | 1,06E-01 | 2,55E-01 |
| TRIM36 | 75,04 | 0,10 | 0,06 | 1,62 | 1,06E-01 | 2,55E-01 |
| UGT8 | 91,37 | 0,10 | 0,06 | 1,62 | 1,06E-01 | 2,55E-01 |
| PCED1B | 618,63 | 0,06 | 0,04 | 1,62 | 1,06E-01 | 2,55E-01 |
| TRMT6 | 452,60 | 0,03 | 0,02 | 1,62 | 1,06E-01 | 2,55E-01 |
| ABCD3 | 1181,87 | 0,04 | 0,02 | 1,62 | 1,06E-01 | 2,55E-01 |
| TPD52 | 662,95 | 0,06 | 0,04 | 1,62 | 1,06E-01 | 2,55E-01 |
| PIK3C3 | 4134,25 | -0,04 | 0,02 | -1,62 | 1,06E-01 | 2,55E-01 |
| MBD2 | 2217,64 | -0,03 | 0,02 | -1,62 | 1,06E-01 | 2,55E-01 |
| ACSM3 | 359,70 | 0,07 | 0,05 | 1,62 | 1,06E-01 | 2,55E-01 |
| MRPL2 | 120,87 | 0,05 | 0,03 | 1,62 | 1,06E-01 | 2,55E-01 |
| ATG4C | 654,58 | 0,06 | 0,04 | 1,62 | 1,06E-01 | 2,55E-01 |
| SLC9A9 | 986,28 | 0,04 | 0,02 | 1,62 | 1,06E-01 | 2,55E-01 |
| CHD4 | 438,71 | -0,04 | 0,03 | -1,61 | 1,06E-01 | 2,56E-01 |
| HNRNPDL | 3746,99 | 0,03 | 0,02 | 1,61 | 1,06E-01 | 2,56E-01 |
| LMO4 | 938,55 | -0,03 | 0,02 | -1,61 | 1,06E-01 | 2,56E-01 |
| ATP2B1 | 7573,32 | -0,04 | 0,03 | -1,61 | 1,06E-01 | 2,56E-01 |
| POLK | 1831,53 | 0,05 | 0,03 | 1,61 | 1,06E-01 | 2,56E-01 |

| | | | | | | |
|---|---|---|---|---|---|---|
| ATG3 | 2775,83 | -0,05 | 0,03 | -1,61 | 1,07E-01 | 2,56E-01 |
| PER3 | 222,49 | 0,06 | 0,04 | 1,61 | 1,07E-01 | 2,56E-01 |
| HAUS6 | 1225,09 | 0,04 | 0,02 | 1,61 | 1,07E-01 | 2,56E-01 |
| QKI | 10970,96 | -0,05 | 0,03 | -1,61 | 1,07E-01 | 2,56E-01 |
| FUS | 5370,12 | -0,06 | 0,03 | -1,61 | 1,07E-01 | 2,57E-01 |
| ALG2 | 628,86 | 0,03 | 0,02 | 1,61 | 1,07E-01 | 2,57E-01 |
| C1orf162 | 1275,32 | -0,03 | 0,02 | -1,61 | 1,07E-01 | 2,57E-01 |
| RNASE6 | 779,26 | -0,08 | 0,05 | -1,61 | 1,07E-01 | 2,57E-01 |
| KIF20B | 468,12 | 0,09 | 0,06 | 1,61 | 1,07E-01 | 2,57E-01 |
| JRK | 1422,13 | 0,04 | 0,03 | 1,61 | 1,07E-01 | 2,57E-01 |
| TDG | 999,94 | 0,03 | 0,02 | 1,61 | 1,07E-01 | 2,57E-01 |
| SP4 | 1864,52 | 0,04 | 0,02 | 1,61 | 1,07E-01 | 2,58E-01 |
| BID | 2457,41 | -0,04 | 0,03 | -1,61 | 1,07E-01 | 2,58E-01 |
| CSTF2 | 308,16 | 0,03 | 0,02 | 1,61 | 1,08E-01 | 2,58E-01 |
| KRBOX5 | 911,61 | 0,05 | 0,03 | 1,61 | 1,08E-01 | 2,58E-01 |
| TCF7 | 2681,59 | 0,08 | 0,05 | 1,61 | 1,08E-01 | 2,58E-01 |
| METTL21A | 503,13 | 0,03 | 0,02 | 1,61 | 1,08E-01 | 2,58E-01 |
| RHOG | 693,70 | -0,08 | 0,05 | -1,61 | 1,08E-01 | 2,58E-01 |
| S1PR1 | 3329,35 | 0,05 | 0,03 | 1,61 | 1,08E-01 | 2,58E-01 |
| CMIP | 1345,82 | -0,10 | 0,06 | -1,61 | 1,08E-01 | 2,58E-01 |
| QSOX1 | 702,24 | -0,07 | 0,04 | -1,61 | 1,08E-01 | 2,58E-01 |
| YEATS2 | 1688,90 | -0,03 | 0,02 | -1,61 | 1,08E-01 | 2,59E-01 |
| LRRCC1 | 132,30 | 0,08 | 0,05 | 1,61 | 1,08E-01 | 2,59E-01 |
| TMEM260 | 2282,83 | -0,04 | 0,02 | -1,61 | 1,08E-01 | 2,59E-01 |
| CD40LG | 326,29 | 0,06 | 0,03 | 1,60 | 1,09E-01 | 2,59E-01 |
| REL | 2898,40 | -0,03 | 0,02 | -1,60 | 1,09E-01 | 2,60E-01 |
| ACTR6 | 520,35 | 0,08 | 0,05 | 1,60 | 1,09E-01 | 2,60E-01 |
| IRF3 | 1134,46 | 0,04 | 0,02 | 1,60 | 1,09E-01 | 2,60E-01 |
| NUP107 | 1195,41 | 0,04 | 0,03 | 1,60 | 1,09E-01 | 2,60E-01 |
| SLIT1 | 83,11 | -0,16 | 0,10 | -1,60 | 1,09E-01 | 2,60E-01 |
| RHOC | 453,04 | 0,05 | 0,03 | 1,60 | 1,09E-01 | 2,60E-01 |
| CNOT11 | 625,13 | 0,02 | 0,01 | 1,60 | 1,09E-01 | 2,60E-01 |
| BRCA2 | 282,85 | 0,08 | 0,05 | 1,60 | 1,09E-01 | 2,60E-01 |
| GBP4 | 2668,02 | -0,08 | 0,05 | -1,60 | 1,09E-01 | 2,60E-01 |
| TIMMDC1 | 1123,17 | 0,03 | 0,02 | 1,60 | 1,09E-01 | 2,60E-01 |
| CRACR2A | 1356,65 | -0,04 | 0,03 | -1,60 | 1,09E-01 | 2,60E-01 |
| DMXL2 | 11054,30 | -0,06 | 0,04 | -1,60 | 1,09E-01 | 2,60E-01 |
| ARAP1 | 7581,21 | -0,09 | 0,06 | -1,60 | 1,09E-01 | 2,60E-01 |
| ANAPC4 | 1549,75 | 0,04 | 0,02 | 1,60 | 1,09E-01 | 2,60E-01 |
| URM1 | 137,45 | 0,06 | 0,04 | 1,60 | 1,09E-01 | 2,60E-01 |
| RXRA | 5140,56 | -0,07 | 0,05 | -1,60 | 1,09E-01 | 2,60E-01 |
| ZNF274 | 1691,82 | -0,03 | 0,02 | -1,60 | 1,09E-01 | 2,60E-01 |
| PLRG1 | 1103,70 | 0,06 | 0,04 | 1,60 | 1,09E-01 | 2,61E-01 |
| PTEN | 24788,90 | -0,06 | 0,04 | -1,60 | 1,10E-01 | 2,61E-01 |
| INTS13 | 601,97 | 0,04 | 0,03 | 1,60 | 1,10E-01 | 2,61E-01 |
| LCN2 | 104,09 | 0,12 | 0,08 | 1,60 | 1,10E-01 | 2,61E-01 |

| | | | | | | |
|---|---|---|---|---|---|---|
| PPP1R12A | 8670,37 | -0,05 | 0,03 | -1,60 | 1,10E-01 | 2,61E-01 |
| NUDT3 | 2246,97 | -0,03 | 0,02 | -1,60 | 1,10E-01 | 2,61E-01 |
| PDZK1IP1 | 647,81 | 0,11 | 0,07 | 1,60 | 1,10E-01 | 2,61E-01 |
| GRWD1 | 311,72 | 0,04 | 0,03 | 1,60 | 1,10E-01 | 2,62E-01 |
| ANKHD1 | 977,60 | -0,04 | 0,02 | -1,60 | 1,10E-01 | 2,62E-01 |
| ZNF805 | 704,75 | 0,03 | 0,02 | 1,60 | 1,10E-01 | 2,62E-01 |
| NEK1 | 789,39 | 0,04 | 0,03 | 1,60 | 1,10E-01 | 2,62E-01 |
| OTUD5 | 1118,93 | -0,06 | 0,04 | -1,60 | 1,10E-01 | 2,62E-01 |
| FIBP | 360,01 | 0,03 | 0,02 | 1,60 | 1,10E-01 | 2,62E-01 |
| NDUFB10 | 560,00 | 0,04 | 0,03 | 1,60 | 1,10E-01 | 2,62E-01 |
| LRPPRC | 2184,44 | 0,04 | 0,03 | 1,60 | 1,11E-01 | 2,62E-01 |
| NDUFB1 | 795,67 | 0,05 | 0,03 | 1,60 | 1,11E-01 | 2,62E-01 |
| RABL3 | 577,94 | 0,05 | 0,03 | 1,60 | 1,10E-01 | 2,62E-01 |
| EEIG2 | 1095,75 | 0,04 | 0,02 | 1,60 | 1,11E-01 | 2,62E-01 |
| KDM6B | 2262,13 | -0,11 | 0,07 | -1,59 | 1,11E-01 | 2,63E-01 |
| WDHD1 | 186,63 | 0,07 | 0,04 | 1,59 | 1,11E-01 | 2,63E-01 |
| NDUFA6 | 415,32 | 0,08 | 0,05 | 1,59 | 1,11E-01 | 2,63E-01 |
| SLAMF1 | 633,63 | 0,05 | 0,03 | 1,59 | 1,11E-01 | 2,63E-01 |
| GRAMD1B | 457,07 | -0,09 | 0,06 | -1,59 | 1,11E-01 | 2,63E-01 |
| PLPPR2 | 380,31 | -0,10 | 0,06 | -1,59 | 1,11E-01 | 2,63E-01 |
| NUBP1 | 674,20 | 0,03 | 0,02 | 1,59 | 1,11E-01 | 2,63E-01 |
| SURF2 | 163,17 | 0,03 | 0,02 | 1,59 | 1,11E-01 | 2,63E-01 |
| ZBTB6 | 1038,25 | 0,07 | 0,05 | 1,59 | 1,11E-01 | 2,63E-01 |
| ZNF184 | 414,68 | 0,07 | 0,04 | 1,59 | 1,11E-01 | 2,63E-01 |
| LRRC45 | 313,50 | 0,04 | 0,02 | 1,59 | 1,11E-01 | 2,63E-01 |
| SMCR8 | 2214,77 | -0,04 | 0,03 | -1,59 | 1,11E-01 | 2,63E-01 |
| ZNF502 | 144,58 | 0,09 | 0,06 | 1,59 | 1,11E-01 | 2,63E-01 |
| ABL2 | 1192,96 | -0,04 | 0,02 | -1,59 | 1,11E-01 | 2,64E-01 |
| FEM1B | 2941,75 | 0,04 | 0,03 | 1,59 | 1,11E-01 | 2,64E-01 |
| SFXN2 | 252,92 | 0,06 | 0,04 | 1,59 | 1,12E-01 | 2,64E-01 |
| WASHC3 | 897,78 | 0,04 | 0,02 | 1,59 | 1,12E-01 | 2,64E-01 |
| MPLKIP | 646,38 | 0,03 | 0,02 | 1,59 | 1,12E-01 | 2,64E-01 |
| OCIAD2 | 238,29 | 0,06 | 0,04 | 1,59 | 1,12E-01 | 2,64E-01 |
| COQ10B | 1509,01 | -0,05 | 0,03 | -1,59 | 1,12E-01 | 2,64E-01 |
| TRIOBP | 1697,65 | -0,06 | 0,04 | -1,59 | 1,12E-01 | 2,64E-01 |
| GCNT4 | 756,55 | 0,07 | 0,05 | 1,59 | 1,12E-01 | 2,64E-01 |
| ZNF446 | 125,71 | 0,06 | 0,04 | 1,59 | 1,12E-01 | 2,64E-01 |
| MTERF1 | 737,65 | 0,06 | 0,04 | 1,59 | 1,12E-01 | 2,64E-01 |
| ENSG00000２ | 403,22 | -0,06 | 0,04 | -1,59 | 1,12E-01 | 2,64E-01 |
| OFD1 | 2077,75 | 0,04 | 0,02 | 1,59 | 1,12E-01 | 2,65E-01 |
| NUDT21 | 1922,12 | 0,06 | 0,04 | 1,59 | 1,12E-01 | 2,65E-01 |
| BAZ1B | 2457,01 | -0,03 | 0,02 | -1,59 | 1,12E-01 | 2,65E-01 |
| SYT17 | 97,56 | 0,11 | 0,07 | 1,59 | 1,12E-01 | 2,65E-01 |
| GNLY | 2390,93 | -0,10 | 0,07 | -1,59 | 1,12E-01 | 2,65E-01 |
| DOK3 | 1927,53 | -0,08 | 0,05 | -1,59 | 1,13E-01 | 2,65E-01 |
| HACD4 | 3954,81 | -0,04 | 0,03 | -1,59 | 1,13E-01 | 2,65E-01 |

| | | | | | | |
|---|---|---|---|---|---|---|
| NARF | 1410,22 | -0,06 | 0,04 | -1,59 | 1,13E-01 | 2,66E-01 |
| PARP10 | 2051,77 | -0,06 | 0,04 | -1,59 | 1,13E-01 | 2,66E-01 |
| NDUFS1 | 2696,55 | -0,02 | 0,01 | -1,59 | 1,13E-01 | 2,66E-01 |
| UBXN10 | 123,99 | 0,09 | 0,05 | 1,59 | 1,13E-01 | 2,66E-01 |
| AIP | 379,18 | 0,04 | 0,03 | 1,58 | 1,13E-01 | 2,66E-01 |
| AVPR1A | 88,88 | 0,10 | 0,06 | 1,58 | 1,13E-01 | 2,66E-01 |
| AGL | 1155,15 | 0,04 | 0,03 | 1,58 | 1,13E-01 | 2,66E-01 |
| PABPC1L | 338,14 | -0,09 | 0,06 | -1,58 | 1,13E-01 | 2,66E-01 |
| RRAGB | 554,87 | 0,03 | 0,02 | 1,58 | 1,13E-01 | 2,66E-01 |
| MON1B | 1160,77 | -0,04 | 0,03 | -1,58 | 1,13E-01 | 2,67E-01 |
| ZNF641 | 3780,49 | -0,05 | 0,03 | -1,58 | 1,13E-01 | 2,67E-01 |
| PUSL1 | 97,22 | 0,05 | 0,03 | 1,58 | 1,13E-01 | 2,67E-01 |
| FURIN | 1378,52 | 0,10 | 0,06 | 1,58 | 1,14E-01 | 2,67E-01 |
| GOLGA8N | 265,14 | -0,07 | 0,04 | -1,58 | 1,14E-01 | 2,67E-01 |
| ZMYM4 | 1250,43 | 0,03 | 0,02 | 1,58 | 1,14E-01 | 2,67E-01 |
| VARS1 | 292,29 | 0,07 | 0,04 | 1,58 | 1,14E-01 | 2,67E-01 |
| TMEM164 | 2360,63 | -0,10 | 0,06 | -1,58 | 1,14E-01 | 2,67E-01 |
| TMEM230 | 1227,16 | 0,03 | 0,02 | 1,58 | 1,14E-01 | 2,67E-01 |
| TAGLN2 | 12383,57 | -0,06 | 0,04 | -1,58 | 1,14E-01 | 2,67E-01 |
| ATP2B4 | 3954,34 | -0,05 | 0,03 | -1,58 | 1,14E-01 | 2,67E-01 |
| NAIF1 | 388,99 | -0,03 | 0,02 | -1,58 | 1,14E-01 | 2,67E-01 |
| STT3B | 5500,58 | 0,05 | 0,03 | 1,58 | 1,14E-01 | 2,67E-01 |
| COG6 | 856,41 | 0,06 | 0,04 | 1,58 | 1,14E-01 | 2,67E-01 |
| CCNG2 | 2552,56 | -0,04 | 0,03 | -1,58 | 1,14E-01 | 2,67E-01 |
| MRFAP1L2 | 674,07 | 0,04 | 0,03 | 1,58 | 1,14E-01 | 2,67E-01 |
| ZMYND19 | 210,86 | 0,03 | 0,02 | 1,58 | 1,14E-01 | 2,67E-01 |
| ENTPD5 | 479,20 | 0,06 | 0,04 | 1,58 | 1,14E-01 | 2,67E-01 |
| FAM210A | 326,13 | 0,04 | 0,02 | 1,58 | 1,14E-01 | 2,67E-01 |
| NFAT5 | 5354,84 | -0,05 | 0,03 | -1,58 | 1,14E-01 | 2,67E-01 |
| FER | 812,25 | 0,04 | 0,03 | 1,58 | 1,14E-01 | 2,67E-01 |
| CALM1 | 8886,62 | -0,03 | 0,02 | -1,58 | 1,14E-01 | 2,68E-01 |
| EFCAB7 | 243,39 | 0,06 | 0,04 | 1,58 | 1,14E-01 | 2,68E-01 |
| FAM131A | 550,50 | -0,05 | 0,03 | -1,58 | 1,14E-01 | 2,68E-01 |
| EEFSEC | 184,07 | 0,05 | 0,03 | 1,58 | 1,14E-01 | 2,68E-01 |
| HLA-DPB1 | 4758,09 | 0,05 | 0,03 | 1,58 | 1,15E-01 | 2,68E-01 |
| C3orf38 | 1114,67 | 0,06 | 0,04 | 1,58 | 1,15E-01 | 2,68E-01 |
| OPA1 | 3432,34 | 0,05 | 0,03 | 1,58 | 1,15E-01 | 2,68E-01 |
| CINP | 573,96 | -0,02 | 0,02 | -1,58 | 1,15E-01 | 2,68E-01 |
| BRAT1 | 802,05 | 0,04 | 0,03 | 1,58 | 1,15E-01 | 2,68E-01 |
| TCEA2 | 154,34 | 0,06 | 0,04 | 1,58 | 1,15E-01 | 2,68E-01 |
| ACAD8 | 725,34 | -0,03 | 0,02 | -1,58 | 1,15E-01 | 2,68E-01 |
| ACTR3B | 259,52 | -0,04 | 0,03 | -1,58 | 1,15E-01 | 2,68E-01 |
| TAPBPL | 1052,83 | -0,04 | 0,03 | -1,58 | 1,15E-01 | 2,68E-01 |
| MMUT | 547,73 | 0,05 | 0,03 | 1,58 | 1,15E-01 | 2,68E-01 |
| ATAD5 | 325,39 | 0,05 | 0,03 | 1,58 | 1,15E-01 | 2,68E-01 |
| CLDN5 | 200,09 | 0,10 | 0,07 | 1,58 | 1,15E-01 | 2,68E-01 |

| | | | | | | |
|---|---|---|---|---|---|---|
| ABI3 | 435,41 | 0,06 | 0,04 | 1,58 | 1,15E-01 | 2,69E-01 |
| CTSZ | 4784,19 | -0,04 | 0,03 | -1,58 | 1,15E-01 | 2,69E-01 |
| SNX16 | 480,01 | 0,08 | 0,05 | 1,58 | 1,15E-01 | 2,69E-01 |
| RANBP1 | 552,66 | 0,04 | 0,02 | 1,58 | 1,15E-01 | 2,69E-01 |
| TECR | 238,21 | 0,06 | 0,04 | 1,58 | 1,15E-01 | 2,69E-01 |
| MIS12 | 630,03 | 0,05 | 0,03 | 1,57 | 1,15E-01 | 2,69E-01 |
| SMYD2 | 1076,48 | 0,03 | 0,02 | 1,57 | 1,15E-01 | 2,69E-01 |
| MAN2C1 | 1675,66 | 0,06 | 0,04 | 1,57 | 1,15E-01 | 2,69E-01 |
| JOSD2 | 132,73 | 0,05 | 0,03 | 1,57 | 1,15E-01 | 2,69E-01 |
| PML | 651,21 | -0,11 | 0,07 | -1,57 | 1,16E-01 | 2,69E-01 |
| SDAD1 | 957,93 | 0,04 | 0,03 | 1,57 | 1,16E-01 | 2,69E-01 |
| ARFRP1 | 1413,19 | 0,03 | 0,02 | 1,57 | 1,16E-01 | 2,69E-01 |
| NR3C1 | 8552,43 | -0,04 | 0,02 | -1,57 | 1,16E-01 | 2,70E-01 |
| FCHSD2 | 1464,33 | -0,05 | 0,03 | -1,57 | 1,16E-01 | 2,70E-01 |
| TRUB2 | 406,31 | 0,03 | 0,02 | 1,57 | 1,16E-01 | 2,70E-01 |
| TMUB2 | 652,85 | -0,05 | 0,03 | -1,57 | 1,16E-01 | 2,70E-01 |
| TIMM8A | 123,69 | 0,04 | 0,02 | 1,57 | 1,16E-01 | 2,70E-01 |
| CD163 | 1086,61 | -0,07 | 0,04 | -1,57 | 1,16E-01 | 2,70E-01 |
| TST | 443,29 | -0,05 | 0,03 | -1,57 | 1,16E-01 | 2,70E-01 |
| UBE2V2 | 610,06 | 0,05 | 0,03 | 1,57 | 1,16E-01 | 2,70E-01 |
| ANAPC15 | 135,99 | 0,04 | 0,03 | 1,57 | 1,16E-01 | 2,70E-01 |
| KRR1 | 1376,00 | 0,06 | 0,04 | 1,57 | 1,16E-01 | 2,70E-01 |
| BRI3 | 1209,69 | -0,05 | 0,03 | -1,57 | 1,17E-01 | 2,71E-01 |
| ATP5PO | 439,07 | 0,06 | 0,04 | 1,57 | 1,17E-01 | 2,71E-01 |
| R3HCC1 | 446,13 | 0,03 | 0,02 | 1,57 | 1,17E-01 | 2,71E-01 |
| CD82 | 1832,69 | -0,05 | 0,03 | -1,57 | 1,17E-01 | 2,71E-01 |
| MRC2 | 785,94 | 0,11 | 0,07 | 1,57 | 1,17E-01 | 2,71E-01 |
| RBMX | 3507,94 | 0,03 | 0,02 | 1,57 | 1,17E-01 | 2,71E-01 |
| ZNF624 | 313,31 | 0,07 | 0,04 | 1,57 | 1,17E-01 | 2,71E-01 |
| CMSS1 | 273,82 | 0,04 | 0,03 | 1,57 | 1,17E-01 | 2,71E-01 |
| MAP2K2 | 1036,64 | 0,06 | 0,04 | 1,57 | 1,17E-01 | 2,71E-01 |
| PEBP1 | 1015,38 | 0,05 | 0,03 | 1,57 | 1,17E-01 | 2,71E-01 |
| COMMD8 | 330,74 | 0,12 | 0,07 | 1,57 | 1,17E-01 | 2,71E-01 |
| CCDC69 | 2245,49 | -0,06 | 0,04 | -1,57 | 1,17E-01 | 2,72E-01 |
| CEP76 | 252,03 | 0,04 | 0,02 | 1,57 | 1,17E-01 | 2,72E-01 |
| SLC9B1 | 95,82 | 0,06 | 0,04 | 1,57 | 1,17E-01 | 2,72E-01 |
| NUDT15 | 112,40 | 0,05 | 0,03 | 1,57 | 1,17E-01 | 2,72E-01 |
| ZNF490 | 247,68 | -0,02 | 0,02 | -1,57 | 1,18E-01 | 2,72E-01 |
| SNAPC3 | 2250,44 | 0,03 | 0,02 | 1,57 | 1,18E-01 | 2,72E-01 |
| NOSIP | 2010,15 | 0,04 | 0,02 | 1,56 | 1,18E-01 | 2,72E-01 |
| DNAJC17 | 508,71 | 0,03 | 0,02 | 1,56 | 1,18E-01 | 2,72E-01 |
| ATG4D | 321,20 | 0,04 | 0,03 | 1,56 | 1,18E-01 | 2,72E-01 |
| KIFAP3 | 943,29 | 0,05 | 0,03 | 1,56 | 1,18E-01 | 2,72E-01 |
| TBKBP1 | 271,25 | -0,09 | 0,06 | -1,56 | 1,18E-01 | 2,72E-01 |
| PLEKHG2 | 219,53 | 0,08 | 0,05 | 1,56 | 1,18E-01 | 2,73E-01 |
| KLC2 | 167,03 | 0,07 | 0,05 | 1,56 | 1,18E-01 | 2,73E-01 |

| | | | | | | |
|---|---|---|---|---|---|---|
| DNA2 | 310,33 | 0,06 | 0,04 | 1,56 | 1,18E-01 | 2,73E-01 |
| TIAM2 | 584,30 | -0,06 | 0,04 | -1,56 | 1,18E-01 | 2,73E-01 |
| SMYD5 | 165,48 | 0,07 | 0,04 | 1,56 | 1,18E-01 | 2,73E-01 |
| PLEKHJ1 | 766,78 | 0,03 | 0,02 | 1,56 | 1,19E-01 | 2,74E-01 |
| RPL7 | 7540,75 | 0,18 | 0,11 | 1,56 | 1,19E-01 | 2,74E-01 |
| RIOX1 | 611,67 | 0,03 | 0,02 | 1,56 | 1,19E-01 | 2,74E-01 |
| CEP290 | 957,77 | 0,07 | 0,04 | 1,56 | 1,19E-01 | 2,74E-01 |
| MOB3A | 2530,03 | -0,09 | 0,05 | -1,56 | 1,19E-01 | 2,74E-01 |
| SNN | 3273,35 | -0,05 | 0,03 | -1,56 | 1,19E-01 | 2,74E-01 |
| SCAF8 | 3185,32 | -0,02 | 0,02 | -1,56 | 1,19E-01 | 2,74E-01 |
| RPL17 | 469,60 | 0,06 | 0,04 | 1,56 | 1,19E-01 | 2,74E-01 |
| ERMP1 | 1218,97 | -0,04 | 0,03 | -1,56 | 1,19E-01 | 2,74E-01 |
| ZNF791 | 1455,76 | 0,05 | 0,03 | 1,56 | 1,19E-01 | 2,75E-01 |
| DBT | 845,90 | 0,02 | 0,01 | 1,56 | 1,19E-01 | 2,75E-01 |
| PRR5 | 506,24 | 0,07 | 0,04 | 1,56 | 1,19E-01 | 2,75E-01 |
| MLX | 1379,20 | -0,03 | 0,02 | -1,56 | 1,19E-01 | 2,75E-01 |
| ARMCX4 | 304,44 | -0,06 | 0,04 | -1,56 | 1,19E-01 | 2,75E-01 |
| TRAPPC14 | 1047,45 | -0,06 | 0,04 | -1,56 | 1,19E-01 | 2,75E-01 |
| RAB3GAP1 | 2386,89 | -0,03 | 0,02 | -1,56 | 1,20E-01 | 2,75E-01 |
| HDHD2 | 610,40 | 0,04 | 0,02 | 1,56 | 1,20E-01 | 2,76E-01 |
| FKBP3 | 475,58 | 0,06 | 0,04 | 1,56 | 1,20E-01 | 2,76E-01 |
| MAPDA | 186,82 | 0,04 | 0,03 | 1,56 | 1,20E-01 | 2,76E-01 |
| PFKL | 1977,31 | 0,04 | 0,03 | 1,56 | 1,20E-01 | 2,76E-01 |
| QTRT1 | 215,37 | 0,04 | 0,03 | 1,56 | 1,20E-01 | 2,76E-01 |
| FHIT | 537,37 | 0,08 | 0,05 | 1,55 | 1,20E-01 | 2,76E-01 |
| CCDC43 | 459,83 | 0,05 | 0,03 | 1,55 | 1,20E-01 | 2,76E-01 |
| LZTFL1 | 425,94 | 0,04 | 0,03 | 1,55 | 1,20E-01 | 2,76E-01 |
| DCAF5 | 1818,52 | -0,03 | 0,02 | -1,55 | 1,20E-01 | 2,76E-01 |
| DGKQ | 616,36 | 0,07 | 0,05 | 1,55 | 1,20E-01 | 2,76E-01 |
| TXNL1 | 989,06 | 0,03 | 0,02 | 1,55 | 1,20E-01 | 2,77E-01 |
| LRRC61 | 123,07 | -0,05 | 0,03 | -1,55 | 1,20E-01 | 2,77E-01 |
| GFM1 | 1392,38 | 0,03 | 0,02 | 1,55 | 1,21E-01 | 2,77E-01 |
| MID2 | 151,37 | 0,05 | 0,03 | 1,55 | 1,21E-01 | 2,77E-01 |
| NCAPG | 83,72 | 0,12 | 0,08 | 1,55 | 1,21E-01 | 2,77E-01 |
| TMEM156 | 386,61 | 0,05 | 0,03 | 1,55 | 1,21E-01 | 2,78E-01 |
| CEP85L | 3508,70 | 0,05 | 0,03 | 1,55 | 1,21E-01 | 2,78E-01 |
| TASOR | 4876,26 | 0,04 | 0,03 | 1,55 | 1,21E-01 | 2,78E-01 |
| SS18L2 | 272,36 | 0,07 | 0,05 | 1,55 | 1,21E-01 | 2,78E-01 |
| ANKRD10 | 5863,69 | 0,04 | 0,03 | 1,55 | 1,21E-01 | 2,78E-01 |
| ILVBL | 272,53 | 0,04 | 0,03 | 1,55 | 1,21E-01 | 2,78E-01 |
| TSPAN32 | 1117,49 | 0,06 | 0,04 | 1,55 | 1,21E-01 | 2,78E-01 |
| AFG1L | 213,36 | 0,04 | 0,02 | 1,55 | 1,21E-01 | 2,78E-01 |
| STX16-NPEPL | 130,74 | -0,07 | 0,04 | -1,55 | 1,22E-01 | 2,78E-01 |
| SASS6 | 237,70 | 0,05 | 0,03 | 1,55 | 1,22E-01 | 2,78E-01 |
| SH2D3C | 1514,40 | -0,07 | 0,04 | -1,55 | 1,22E-01 | 2,78E-01 |
| EXOC5 | 2446,28 | 0,05 | 0,03 | 1,55 | 1,22E-01 | 2,78E-01 |

| | | | | | | |
|---|---|---|---|---|---|---|
| ATMIN | 1287,45 | 0,03 | 0,02 | 1,55 | 1,22E-01 | 2,79E-01 |
| BCORL1 | 240,62 | -0,09 | 0,06 | -1,55 | 1,22E-01 | 2,79E-01 |
| P2RX7 | 662,52 | 0,05 | 0,03 | 1,55 | 1,22E-01 | 2,79E-01 |
| KAT8 | 1721,92 | -0,03 | 0,02 | -1,55 | 1,22E-01 | 2,79E-01 |
| URB2 | 483,50 | 0,03 | 0,02 | 1,55 | 1,22E-01 | 2,79E-01 |
| GET3 | 909,52 | 0,05 | 0,03 | 1,55 | 1,22E-01 | 2,79E-01 |
| STIP1 | 1340,54 | -0,04 | 0,02 | -1,55 | 1,22E-01 | 2,79E-01 |
| CD14 | 4489,60 | -0,07 | 0,04 | -1,55 | 1,22E-01 | 2,79E-01 |
| VPS45 | 903,56 | 0,03 | 0,02 | 1,55 | 1,22E-01 | 2,79E-01 |
| C21orf91 | 2150,33 | 0,08 | 0,05 | 1,55 | 1,22E-01 | 2,79E-01 |
| CTSO | 849,04 | 0,04 | 0,03 | 1,55 | 1,22E-01 | 2,79E-01 |
| DPEP2 | 3191,75 | -0,07 | 0,04 | -1,54 | 1,23E-01 | 2,80E-01 |
| COPS4 | 454,90 | 0,07 | 0,04 | 1,54 | 1,23E-01 | 2,80E-01 |
| MAZ | 543,27 | 0,08 | 0,05 | 1,54 | 1,23E-01 | 2,80E-01 |
| ARMC7 | 218,83 | -0,05 | 0,03 | -1,54 | 1,23E-01 | 2,80E-01 |
| FOXO3B | 92,29 | 0,05 | 0,03 | 1,54 | 1,23E-01 | 2,80E-01 |
| ZNF559 | 639,95 | -0,04 | 0,03 | -1,54 | 1,23E-01 | 2,80E-01 |
| STK10 | 7290,81 | -0,07 | 0,04 | -1,54 | 1,23E-01 | 2,80E-01 |
| POLR3F | 268,29 | 0,04 | 0,02 | 1,54 | 1,23E-01 | 2,80E-01 |
| BBC3 | 503,18 | -0,05 | 0,03 | -1,54 | 1,23E-01 | 2,80E-01 |
| MFSD8 | 1472,21 | 0,03 | 0,02 | 1,54 | 1,23E-01 | 2,80E-01 |
| MRPS33 | 295,62 | 0,07 | 0,04 | 1,54 | 1,23E-01 | 2,80E-01 |
| TNFAIP8L2 | 1610,04 | -0,03 | 0,02 | -1,54 | 1,23E-01 | 2,80E-01 |
| TIPARP | 1902,95 | -0,04 | 0,03 | -1,54 | 1,23E-01 | 2,81E-01 |
| NDUFA5 | 597,72 | 0,08 | 0,05 | 1,54 | 1,23E-01 | 2,81E-01 |
| PPP6R3 | 6774,95 | -0,02 | 0,02 | -1,54 | 1,23E-01 | 2,81E-01 |
| NPM3 | 139,72 | 0,04 | 0,03 | 1,54 | 1,24E-01 | 2,81E-01 |
| CCAR1 | 1941,18 | 0,02 | 0,02 | 1,54 | 1,24E-01 | 2,81E-01 |
| C5orf22 | 726,94 | 0,03 | 0,02 | 1,54 | 1,24E-01 | 2,81E-01 |
| CEP295 | 1651,62 | -0,03 | 0,02 | -1,54 | 1,24E-01 | 2,82E-01 |
| PRX | 91,59 | 0,08 | 0,05 | 1,54 | 1,24E-01 | 2,82E-01 |
| PSMD12 | 1427,62 | 0,03 | 0,02 | 1,54 | 1,24E-01 | 2,82E-01 |
| VCF1 | 4905,47 | 0,08 | 0,05 | 1,54 | 1,24E-01 | 2,82E-01 |
| SLC30A4 | 275,16 | 0,05 | 0,03 | 1,54 | 1,24E-01 | 2,83E-01 |
| ASPSCR1 | 297,93 | 0,09 | 0,06 | 1,54 | 1,24E-01 | 2,83E-01 |
| ARF3 | 1145,39 | -0,07 | 0,05 | -1,54 | 1,24E-01 | 2,83E-01 |
| SMG7 | 2008,65 | -0,04 | 0,03 | -1,54 | 1,24E-01 | 2,83E-01 |
| AP3B1 | 4244,16 | -0,04 | 0,03 | -1,54 | 1,24E-01 | 2,83E-01 |
| HLA-DMB | 3006,05 | 0,05 | 0,03 | 1,54 | 1,24E-01 | 2,83E-01 |
| TTC9C | 578,38 | 0,02 | 0,01 | 1,54 | 1,25E-01 | 2,83E-01 |
| WDR3 | 910,02 | 0,04 | 0,02 | 1,54 | 1,25E-01 | 2,83E-01 |
| CYTH2 | 877,14 | -0,04 | 0,03 | -1,54 | 1,25E-01 | 2,83E-01 |
| ZNF131 | 1338,24 | 0,03 | 0,02 | 1,54 | 1,25E-01 | 2,83E-01 |
| MAN1B1 | 1341,80 | 0,03 | 0,02 | 1,53 | 1,25E-01 | 2,83E-01 |
| MRM3 | 323,35 | 0,03 | 0,02 | 1,53 | 1,25E-01 | 2,83E-01 |
| CHCHD2 | 2281,21 | 0,05 | 0,03 | 1,53 | 1,25E-01 | 2,83E-01 |

| | | | | | | |
|---|---|---|---|---|---|---|
| IER5 | 2140,84 | 0,04 | 0,03 | 1,53 | 1,25E-01 | 2,83E-01 |
| RFNG | 119,84 | 0,08 | 0,05 | 1,53 | 1,25E-01 | 2,83E-01 |
| HLA-DPA1 | 5034,55 | 0,05 | 0,03 | 1,53 | 1,25E-01 | 2,83E-01 |
| OLIG2 | 112,94 | 0,13 | 0,08 | 1,53 | 1,25E-01 | 2,83E-01 |
| KLHL14 | 177,89 | 0,11 | 0,07 | 1,53 | 1,25E-01 | 2,84E-01 |
| DDI2 | 2270,00 | 0,03 | 0,02 | 1,53 | 1,25E-01 | 2,84E-01 |
| PPIF | 1048,96 | -0,05 | 0,03 | -1,53 | 1,25E-01 | 2,84E-01 |
| GATA3 | 175,26 | 0,06 | 0,04 | 1,53 | 1,25E-01 | 2,84E-01 |
| B3GLCT | 254,68 | 0,04 | 0,02 | 1,53 | 1,25E-01 | 2,84E-01 |
| C1orf21 | 804,83 | -0,08 | 0,05 | -1,53 | 1,25E-01 | 2,84E-01 |
| ALKBH8 | 564,65 | 0,04 | 0,03 | 1,53 | 1,26E-01 | 2,84E-01 |
| NPHP3 | 543,83 | 0,04 | 0,02 | 1,53 | 1,26E-01 | 2,84E-01 |
| SUPV3L1 | 570,60 | 0,03 | 0,02 | 1,53 | 1,26E-01 | 2,84E-01 |
| PARP8 | 15142,37 | -0,05 | 0,03 | -1,53 | 1,26E-01 | 2,84E-01 |
| UEVLD | 751,70 | -0,03 | 0,02 | -1,53 | 1,26E-01 | 2,84E-01 |
| TSR3 | 641,15 | 0,03 | 0,02 | 1,53 | 1,26E-01 | 2,84E-01 |
| IGFBP3 | 165,14 | 0,09 | 0,06 | 1,53 | 1,26E-01 | 2,84E-01 |
| ZNF33B | 1507,26 | 0,04 | 0,03 | 1,53 | 1,26E-01 | 2,84E-01 |
| SOWAHC | 114,65 | 0,07 | 0,04 | 1,53 | 1,26E-01 | 2,84E-01 |
| POLR2D | 775,54 | 0,02 | 0,01 | 1,53 | 1,26E-01 | 2,84E-01 |
| CCDC86 | 94,33 | 0,06 | 0,04 | 1,53 | 1,26E-01 | 2,84E-01 |
| INTS4 | 1414,56 | -0,02 | 0,01 | -1,53 | 1,26E-01 | 2,84E-01 |
| SIAH1 | 956,69 | -0,03 | 0,02 | -1,53 | 1,26E-01 | 2,84E-01 |
| MAML1 | 1600,23 | -0,06 | 0,04 | -1,53 | 1,26E-01 | 2,85E-01 |
| CROCC | 663,13 | 0,06 | 0,04 | 1,53 | 1,26E-01 | 2,85E-01 |
| H3C11 | 488,16 | 0,06 | 0,04 | 1,53 | 1,26E-01 | 2,85E-01 |
| WDR4 | 271,24 | 0,05 | 0,03 | 1,53 | 1,26E-01 | 2,85E-01 |
| IRAK3 | 6706,31 | -0,06 | 0,04 | -1,53 | 1,26E-01 | 2,85E-01 |
| MED17 | 1466,16 | -0,02 | 0,01 | -1,53 | 1,27E-01 | 2,86E-01 |
| ZC3H8 | 544,91 | 0,03 | 0,02 | 1,53 | 1,27E-01 | 2,86E-01 |
| AKR1B1 | 401,28 | 0,03 | 0,02 | 1,53 | 1,27E-01 | 2,86E-01 |
| DAPP1 | 4741,11 | -0,05 | 0,04 | -1,53 | 1,27E-01 | 2,86E-01 |
| EXOSC7 | 649,80 | 0,03 | 0,02 | 1,53 | 1,27E-01 | 2,86E-01 |
| KNTC1 | 1063,19 | 0,04 | 0,02 | 1,52 | 1,27E-01 | 2,87E-01 |
| SCML1 | 332,56 | 0,07 | 0,05 | 1,52 | 1,27E-01 | 2,87E-01 |
| SLC2A5 | 86,10 | 0,11 | 0,07 | 1,52 | 1,27E-01 | 2,87E-01 |
| SIRT3 | 262,48 | 0,04 | 0,03 | 1,52 | 1,27E-01 | 2,87E-01 |
| FGL2 | 22047,41 | -0,07 | 0,05 | -1,52 | 1,27E-01 | 2,87E-01 |
| TOPBP1 | 2272,55 | -0,03 | 0,02 | -1,52 | 1,28E-01 | 2,87E-01 |
| LIMD1 | 1607,19 | 0,04 | 0,03 | 1,52 | 1,28E-01 | 2,87E-01 |
| IMPDH1 | 1706,64 | -0,07 | 0,05 | -1,52 | 1,28E-01 | 2,87E-01 |
| RHOT1 | 2384,42 | -0,04 | 0,03 | -1,52 | 1,28E-01 | 2,87E-01 |
| ARSD | 446,54 | -0,04 | 0,03 | -1,52 | 1,28E-01 | 2,87E-01 |
| NDUFS4 | 242,55 | 0,06 | 0,04 | 1,52 | 1,28E-01 | 2,87E-01 |
| ODF2L | 2478,61 | 0,05 | 0,04 | 1,52 | 1,28E-01 | 2,87E-01 |
| SNX24 | 170,59 | 0,04 | 0,03 | 1,52 | 1,28E-01 | 2,87E-01 |

| | | | | | | |
|---|---|---|---|---|---|---|
| TELO2 | 305,05 | 0,07 | 0,05 | 1,52 | 1,28E-01 | 2,87E-01 |
| NCKIPSD | 128,93 | 0,07 | 0,04 | 1,52 | 1,28E-01 | 2,87E-01 |
| AFTPH | 3562,50 | -0,04 | 0,03 | -1,52 | 1,28E-01 | 2,87E-01 |
| TCL1A | 582,00 | 0,11 | 0,07 | 1,52 | 1,28E-01 | 2,87E-01 |
| NSUN4 | 443,15 | 0,04 | 0,02 | 1,52 | 1,28E-01 | 2,87E-01 |
| NFAM1 | 2937,61 | -0,11 | 0,07 | -1,52 | 1,28E-01 | 2,87E-01 |
| TADA2A | 601,59 | 0,02 | 0,02 | 1,52 | 1,28E-01 | 2,87E-01 |
| EMC9 | 90,08 | 0,04 | 0,03 | 1,52 | 1,28E-01 | 2,88E-01 |
| MIPEP | 279,20 | 0,04 | 0,03 | 1,52 | 1,28E-01 | 2,88E-01 |
| ASCC2 | 4671,75 | 0,10 | 0,06 | 1,52 | 1,28E-01 | 2,88E-01 |
| SIKE1 | 1457,71 | 0,05 | 0,03 | 1,52 | 1,28E-01 | 2,88E-01 |
| INTS2 | 968,37 | 0,03 | 0,02 | 1,52 | 1,28E-01 | 2,88E-01 |
| RAN | 2166,75 | 0,05 | 0,03 | 1,52 | 1,29E-01 | 2,88E-01 |
| SUCO | 2480,07 | 0,05 | 0,03 | 1,52 | 1,29E-01 | 2,88E-01 |
| CARMIL2 | 293,90 | 0,09 | 0,06 | 1,52 | 1,29E-01 | 2,89E-01 |
| DHX29 | 1066,30 | 0,04 | 0,03 | 1,52 | 1,29E-01 | 2,89E-01 |
| PTPN4 | 5821,88 | 0,05 | 0,04 | 1,52 | 1,29E-01 | 2,89E-01 |
| SLC46A3 | 760,83 | -0,03 | 0,02 | -1,52 | 1,29E-01 | 2,89E-01 |
| SNTB1 | 1086,58 | 0,03 | 0,02 | 1,52 | 1,29E-01 | 2,89E-01 |
| DDX56 | 889,00 | 0,03 | 0,02 | 1,52 | 1,29E-01 | 2,89E-01 |
| DOCK10 | 7922,75 | 0,04 | 0,02 | 1,52 | 1,29E-01 | 2,89E-01 |
| DNAJC18 | 128,45 | 0,04 | 0,03 | 1,52 | 1,29E-01 | 2,89E-01 |
| WAPL | 5045,78 | -0,04 | 0,02 | -1,52 | 1,29E-01 | 2,89E-01 |
| ANKIB1 | 2173,13 | 0,03 | 0,02 | 1,52 | 1,30E-01 | 2,90E-01 |
| RPP40 | 88,85 | 0,05 | 0,03 | 1,52 | 1,30E-01 | 2,90E-01 |
| STK38L | 2894,78 | -0,05 | 0,03 | -1,52 | 1,30E-01 | 2,90E-01 |
| ALKBH6 | 100,91 | 0,04 | 0,03 | 1,51 | 1,30E-01 | 2,90E-01 |
| ZNF317 | 1310,54 | -0,02 | 0,01 | -1,51 | 1,30E-01 | 2,90E-01 |
| ATP6AP1 | 764,97 | -0,05 | 0,03 | -1,51 | 1,30E-01 | 2,90E-01 |
| SRGAP2 | 1082,72 | -0,06 | 0,04 | -1,51 | 1,30E-01 | 2,91E-01 |
| AMMECR1 | 646,96 | 0,03 | 0,02 | 1,51 | 1,30E-01 | 2,91E-01 |
| CSNK1G1 | 699,09 | -0,04 | 0,03 | -1,51 | 1,30E-01 | 2,91E-01 |
| RAB11FIP2 | 1707,31 | 0,05 | 0,03 | 1,51 | 1,30E-01 | 2,91E-01 |
| R3HDM2 | 786,71 | -0,05 | 0,04 | -1,51 | 1,30E-01 | 2,91E-01 |
| GMDS | 351,37 | -0,06 | 0,04 | -1,51 | 1,30E-01 | 2,91E-01 |
| QSOX2 | 477,94 | 0,05 | 0,04 | 1,51 | 1,30E-01 | 2,91E-01 |
| TBC1D5 | 3531,51 | -0,03 | 0,02 | -1,51 | 1,31E-01 | 2,91E-01 |
| PIGU | 251,26 | 0,03 | 0,02 | 1,51 | 1,31E-01 | 2,91E-01 |
| PIGK | 788,66 | 0,07 | 0,05 | 1,51 | 1,31E-01 | 2,91E-01 |
| KDM5D | 2458,48 | 0,19 | 0,12 | 1,51 | 1,31E-01 | 2,91E-01 |
| MFSD6 | 1306,25 | 0,03 | 0,02 | 1,51 | 1,31E-01 | 2,91E-01 |
| SH3RF1 | 133,99 | 0,07 | 0,05 | 1,51 | 1,31E-01 | 2,91E-01 |
| SURF1 | 497,66 | 0,03 | 0,02 | 1,51 | 1,31E-01 | 2,92E-01 |
| SUPT16H | 2046,01 | -0,03 | 0,02 | -1,51 | 1,31E-01 | 2,92E-01 |
| ATIC | 427,20 | 0,04 | 0,03 | 1,51 | 1,31E-01 | 2,92E-01 |
| HBM | 201,13 | 0,12 | 0,08 | 1,51 | 1,31E-01 | 2,92E-01 |

| | | | | | | |
|---|---|---|---|---|---|---|
| ZNF646 | 1595,44 | -0,05 | 0,04 | -1,51 | 1,31E-01 | 2,92E-01 |
| ZCCHC7 | 1872,31 | 0,05 | 0,03 | 1,51 | 1,31E-01 | 2,92E-01 |
| ARRB2 | 4161,58 | -0,07 | 0,04 | -1,51 | 1,31E-01 | 2,92E-01 |
| RAB2A | 2920,74 | -0,04 | 0,03 | -1,51 | 1,32E-01 | 2,92E-01 |
| MOCS2 | 321,94 | 0,05 | 0,03 | 1,51 | 1,32E-01 | 2,92E-01 |
| PSAP | 13983,17 | -0,06 | 0,04 | -1,51 | 1,32E-01 | 2,92E-01 |
| ZNF804A | 91,74 | 0,06 | 0,04 | 1,51 | 1,32E-01 | 2,93E-01 |
| LZTR1 | 440,86 | 0,05 | 0,03 | 1,51 | 1,32E-01 | 2,93E-01 |
| SLC41A3 | 411,82 | 0,04 | 0,02 | 1,51 | 1,32E-01 | 2,93E-01 |
| ENOPH1 | 491,31 | 0,04 | 0,03 | 1,51 | 1,32E-01 | 2,93E-01 |
| ZNF283 | 543,38 | 0,04 | 0,03 | 1,51 | 1,32E-01 | 2,93E-01 |
| RYK | 1408,93 | 0,03 | 0,02 | 1,51 | 1,32E-01 | 2,93E-01 |
| ATPAF1 | 699,40 | 0,02 | 0,02 | 1,51 | 1,32E-01 | 2,93E-01 |
| TOMM20 | 1941,88 | 0,05 | 0,03 | 1,51 | 1,32E-01 | 2,93E-01 |
| NHERF1 | 2153,14 | -0,05 | 0,03 | -1,51 | 1,32E-01 | 2,93E-01 |
| HSPA13 | 980,64 | 0,08 | 0,05 | 1,51 | 1,32E-01 | 2,93E-01 |
| CES4A | 144,58 | 0,05 | 0,04 | 1,51 | 1,32E-01 | 2,93E-01 |
| H2BC5 | 2026,83 | 0,06 | 0,04 | 1,51 | 1,32E-01 | 2,93E-01 |
| AFF2 | 256,36 | -0,07 | 0,05 | -1,50 | 1,32E-01 | 2,94E-01 |
| ABCA10 | 161,17 | 0,07 | 0,05 | 1,50 | 1,32E-01 | 2,94E-01 |
| FBXW4 | 1516,96 | 0,03 | 0,02 | 1,50 | 1,33E-01 | 2,94E-01 |
| GALNT12 | 179,97 | 0,05 | 0,03 | 1,50 | 1,33E-01 | 2,94E-01 |
| ERP44 | 1673,03 | -0,04 | 0,02 | -1,50 | 1,33E-01 | 2,94E-01 |
| HAUS5 | 152,39 | 0,06 | 0,04 | 1,50 | 1,33E-01 | 2,95E-01 |
| COA3 | 374,68 | 0,04 | 0,03 | 1,50 | 1,33E-01 | 2,95E-01 |
| NLRP1 | 5698,73 | -0,08 | 0,06 | -1,50 | 1,33E-01 | 2,95E-01 |
| PCBD1 | 127,51 | 0,04 | 0,03 | 1,50 | 1,33E-01 | 2,95E-01 |
| ZNF711 | 78,78 | 0,08 | 0,06 | 1,50 | 1,33E-01 | 2,95E-01 |
| RAP1GDS1 | 1264,50 | 0,04 | 0,02 | 1,50 | 1,33E-01 | 2,95E-01 |
| JRKL | 328,25 | 0,07 | 0,05 | 1,50 | 1,33E-01 | 2,95E-01 |
| MTAP | 1210,03 | 0,03 | 0,02 | 1,50 | 1,33E-01 | 2,95E-01 |
| GRHL1 | 108,84 | 0,09 | 0,06 | 1,50 | 1,33E-01 | 2,95E-01 |
| LRRC8A | 608,97 | 0,05 | 0,04 | 1,50 | 1,33E-01 | 2,95E-01 |
| TANGO6 | 1142,94 | 0,02 | 0,02 | 1,50 | 1,34E-01 | 2,95E-01 |
| MRPL24 | 345,46 | 0,04 | 0,03 | 1,50 | 1,34E-01 | 2,96E-01 |
| PPA2 | 1208,37 | 0,04 | 0,02 | 1,50 | 1,34E-01 | 2,96E-01 |
| TARDBP | 1549,52 | -0,03 | 0,02 | -1,50 | 1,34E-01 | 2,96E-01 |
| BCL2L13 | 1388,65 | 0,04 | 0,03 | 1,50 | 1,34E-01 | 2,96E-01 |
| PRNP | 1312,01 | 0,04 | 0,03 | 1,50 | 1,34E-01 | 2,96E-01 |
| NME4 | 276,97 | 0,06 | 0,04 | 1,50 | 1,34E-01 | 2,97E-01 |
| PAPOLA | 9903,70 | -0,04 | 0,02 | -1,50 | 1,34E-01 | 2,97E-01 |
| ARPC1B | 1149,71 | -0,04 | 0,02 | -1,50 | 1,34E-01 | 2,97E-01 |
| ANKRD6 | 299,84 | -0,04 | 0,03 | -1,50 | 1,35E-01 | 2,97E-01 |
| RAP1GAP2 | 2636,16 | -0,07 | 0,05 | -1,50 | 1,34E-01 | 2,97E-01 |
| PUF60 | 1066,67 | 0,02 | 0,02 | 1,50 | 1,35E-01 | 2,97E-01 |
| ADAP2 | 228,80 | 0,06 | 0,04 | 1,50 | 1,34E-01 | 2,97E-01 |

| | | | | | | |
|---|---|---|---|---|---|---|
| C5orf63 | 417,67 | 0,05 | 0,03 | 1,50 | 1,35E-01 | 2,97E-01 |
| GGCT | 126,09 | 0,05 | 0,03 | 1,50 | 1,35E-01 | 2,97E-01 |
| GALC | 2003,66 | -0,03 | 0,02 | -1,50 | 1,35E-01 | 2,97E-01 |
| NADK2 | 517,75 | 0,04 | 0,03 | 1,50 | 1,35E-01 | 2,97E-01 |
| FAM24B | 131,78 | 0,05 | 0,03 | 1,50 | 1,35E-01 | 2,97E-01 |
| BLTP2 | 4730,84 | -0,03 | 0,02 | -1,49 | 1,35E-01 | 2,98E-01 |
| BPNT1 | 450,45 | 0,04 | 0,03 | 1,49 | 1,35E-01 | 2,98E-01 |
| USP22 | 1942,55 | -0,06 | 0,04 | -1,49 | 1,35E-01 | 2,98E-01 |
| TMEM192 | 508,67 | 0,02 | 0,01 | 1,49 | 1,35E-01 | 2,98E-01 |
| SRM | 77,33 | 0,08 | 0,05 | 1,49 | 1,35E-01 | 2,98E-01 |
| LSM6 | 1082,59 | 0,04 | 0,03 | 1,49 | 1,35E-01 | 2,98E-01 |
| LRP10 | 3232,28 | -0,08 | 0,06 | -1,49 | 1,35E-01 | 2,98E-01 |
| ARHGEF6 | 4958,03 | -0,03 | 0,02 | -1,49 | 1,36E-01 | 2,98E-01 |
| JAK3 | 3097,81 | -0,08 | 0,06 | -1,49 | 1,36E-01 | 2,98E-01 |
| TMEM167B | 1965,14 | -0,05 | 0,03 | -1,49 | 1,36E-01 | 2,98E-01 |
| CCDC149 | 314,96 | -0,07 | 0,05 | -1,49 | 1,36E-01 | 2,98E-01 |
| NSA2 | 1043,74 | 0,08 | 0,05 | 1,49 | 1,36E-01 | 2,98E-01 |
| SPAG1 | 262,68 | -0,06 | 0,04 | -1,49 | 1,36E-01 | 2,99E-01 |
| COPS3 | 1172,46 | 0,04 | 0,03 | 1,49 | 1,36E-01 | 2,99E-01 |
| ZNF256 | 173,09 | 0,05 | 0,03 | 1,49 | 1,36E-01 | 2,99E-01 |
| GON4L | 2157,75 | -0,04 | 0,02 | -1,49 | 1,36E-01 | 2,99E-01 |
| METAP1 | 934,64 | 0,03 | 0,02 | 1,49 | 1,36E-01 | 2,99E-01 |
| RHOT2 | 1097,93 | 0,04 | 0,03 | 1,49 | 1,36E-01 | 2,99E-01 |
| GABRR2 | 74,71 | -0,08 | 0,06 | -1,49 | 1,36E-01 | 2,99E-01 |
| EXOSC9 | 710,18 | 0,04 | 0,03 | 1,49 | 1,36E-01 | 2,99E-01 |
| NOP58 | 1431,47 | 0,05 | 0,04 | 1,49 | 1,36E-01 | 2,99E-01 |
| SH3GL1 | 508,12 | 0,06 | 0,04 | 1,49 | 1,37E-01 | 2,99E-01 |
| TPP2 | 4615,97 | 0,04 | 0,02 | 1,49 | 1,37E-01 | 2,99E-01 |
| FBXW5 | 1737,12 | 0,03 | 0,02 | 1,49 | 1,37E-01 | 3,00E-01 |
| THNSL1 | 160,31 | 0,07 | 0,05 | 1,49 | 1,37E-01 | 3,00E-01 |
| TLN1 | 14749,76 | -0,08 | 0,05 | -1,49 | 1,37E-01 | 3,00E-01 |
| PCID2 | 1503,03 | 0,03 | 0,02 | 1,49 | 1,37E-01 | 3,00E-01 |
| MAF | 891,06 | 0,05 | 0,03 | 1,49 | 1,37E-01 | 3,00E-01 |
| SNRPD1 | 420,59 | 0,05 | 0,03 | 1,49 | 1,37E-01 | 3,00E-01 |
| GLI4 | 157,99 | 0,08 | 0,05 | 1,49 | 1,37E-01 | 3,00E-01 |
| CELSR3 | 108,68 | 0,11 | 0,07 | 1,49 | 1,37E-01 | 3,00E-01 |
| ZDHHC7 | 3784,17 | -0,04 | 0,03 | -1,49 | 1,37E-01 | 3,00E-01 |
| PPT1 | 4421,32 | -0,06 | 0,04 | -1,49 | 1,37E-01 | 3,01E-01 |
| CLEC6A | 94,70 | -0,09 | 0,06 | -1,49 | 1,38E-01 | 3,01E-01 |
| EIF4G2 | 8417,47 | -0,03 | 0,02 | -1,48 | 1,38E-01 | 3,01E-01 |
| LAPTM4B | 107,56 | 0,07 | 0,04 | 1,48 | 1,38E-01 | 3,01E-01 |
| MAML2 | 1516,77 | -0,03 | 0,02 | -1,48 | 1,38E-01 | 3,01E-01 |
| FRA10AC1 | 993,55 | 0,05 | 0,03 | 1,48 | 1,38E-01 | 3,01E-01 |
| TEX10 | 914,41 | 0,03 | 0,02 | 1,48 | 1,38E-01 | 3,01E-01 |
| MSTO1 | 147,41 | 0,04 | 0,03 | 1,48 | 1,38E-01 | 3,01E-01 |
| PHF20L1 | 10018,76 | -0,05 | 0,04 | -1,48 | 1,38E-01 | 3,01E-01 |

| | | | | | | |
|---|---|---|---|---|---|---|
| CD101 | 601,48 | 0,07 | 0,05 | 1,48 | 1,38E-01 | 3,02E-01 |
| LILRB2 | 2006,24 | -0,08 | 0,05 | -1,48 | 1,38E-01 | 3,02E-01 |
| SAMD9L | 6366,83 | -0,09 | 0,06 | -1,48 | 1,38E-01 | 3,02E-01 |
| ACYP1 | 221,17 | 0,04 | 0,03 | 1,48 | 1,39E-01 | 3,02E-01 |
| C4BPA | 110,98 | 0,25 | 0,17 | 1,48 | 1,39E-01 | 3,02E-01 |
| GPR34 | 150,30 | 0,10 | 0,07 | 1,48 | 1,39E-01 | 3,02E-01 |
| USP39 | 1051,02 | -0,02 | 0,01 | -1,48 | 1,39E-01 | 3,02E-01 |
| MRPL16 | 419,48 | 0,03 | 0,02 | 1,48 | 1,39E-01 | 3,02E-01 |
| TNFRSF8 | 291,77 | -0,05 | 0,03 | -1,48 | 1,39E-01 | 3,02E-01 |
| METTL25B | 318,59 | 0,03 | 0,02 | 1,48 | 1,39E-01 | 3,02E-01 |
| LYSMD3 | 1513,00 | 0,08 | 0,05 | 1,48 | 1,39E-01 | 3,02E-01 |
| GMCL1 | 1908,73 | -0,04 | 0,02 | -1,48 | 1,39E-01 | 3,02E-01 |
| ATG2A | 954,87 | -0,10 | 0,07 | -1,48 | 1,39E-01 | 3,02E-01 |
| VPS25 | 481,03 | 0,03 | 0,02 | 1,48 | 1,39E-01 | 3,02E-01 |
| CHM | 895,81 | 0,05 | 0,03 | 1,48 | 1,39E-01 | 3,02E-01 |
| BRI3BP | 1101,73 | 0,03 | 0,02 | 1,48 | 1,39E-01 | 3,02E-01 |
| NIPSNAP1 | 197,64 | 0,04 | 0,03 | 1,48 | 1,39E-01 | 3,03E-01 |
| SNRPA | 191,58 | 0,05 | 0,04 | 1,48 | 1,40E-01 | 3,04E-01 |
| AHNAK | 61854,45 | 0,03 | 0,02 | 1,48 | 1,40E-01 | 3,04E-01 |
| OXR1 | 2307,74 | 0,06 | 0,04 | 1,48 | 1,40E-01 | 3,04E-01 |
| COQ8B | 649,34 | -0,03 | 0,02 | -1,48 | 1,40E-01 | 3,04E-01 |
| ZNF780B | 2393,75 | 0,06 | 0,04 | 1,48 | 1,40E-01 | 3,04E-01 |
| MCRS1 | 401,66 | 0,04 | 0,03 | 1,48 | 1,40E-01 | 3,04E-01 |
| DUSP10 | 305,30 | -0,03 | 0,02 | -1,48 | 1,40E-01 | 3,05E-01 |
| MTFR1 | 187,13 | 0,04 | 0,03 | 1,47 | 1,40E-01 | 3,05E-01 |
| N4BP2L2 | 12902,54 | -0,03 | 0,02 | -1,47 | 1,40E-01 | 3,05E-01 |
| TMEM9 | 255,77 | 0,04 | 0,03 | 1,47 | 1,41E-01 | 3,05E-01 |
| BOP1 | 97,49 | 0,07 | 0,05 | 1,47 | 1,41E-01 | 3,06E-01 |
| CDKN1A | 240,30 | 0,08 | 0,06 | 1,47 | 1,41E-01 | 3,06E-01 |
| ZC3H13 | 2773,45 | -0,03 | 0,02 | -1,47 | 1,41E-01 | 3,06E-01 |
| ASGR1 | 230,43 | 0,05 | 0,04 | 1,47 | 1,41E-01 | 3,06E-01 |
| DNPEP | 615,43 | 0,03 | 0,02 | 1,47 | 1,41E-01 | 3,07E-01 |
| GIMAP4 | 11258,68 | -0,05 | 0,04 | -1,47 | 1,41E-01 | 3,07E-01 |
| LMF1 | 309,22 | 0,07 | 0,04 | 1,47 | 1,41E-01 | 3,07E-01 |
| KCNMB1 | 123,22 | 0,07 | 0,05 | 1,47 | 1,42E-01 | 3,07E-01 |
| FGD3 | 3825,85 | -0,08 | 0,05 | -1,47 | 1,42E-01 | 3,07E-01 |
| PABPN1 | 1310,02 | -0,02 | 0,02 | -1,47 | 1,42E-01 | 3,07E-01 |
| NMUR1 | 211,10 | -0,08 | 0,06 | -1,47 | 1,42E-01 | 3,07E-01 |
| CUEDC2 | 284,85 | 0,03 | 0,02 | 1,47 | 1,42E-01 | 3,08E-01 |
| FERRY3 | 570,01 | 0,04 | 0,03 | 1,47 | 1,42E-01 | 3,08E-01 |
| GATB | 349,62 | 0,04 | 0,03 | 1,47 | 1,42E-01 | 3,08E-01 |
| PM20D2 | 627,22 | 0,04 | 0,02 | 1,47 | 1,42E-01 | 3,08E-01 |
| ACOT2 | 137,36 | 0,04 | 0,03 | 1,47 | 1,42E-01 | 3,08E-01 |
| FAM98C | 365,27 | 0,04 | 0,03 | 1,47 | 1,42E-01 | 3,08E-01 |
| ZFX | 3597,36 | -0,03 | 0,02 | -1,47 | 1,42E-01 | 3,08E-01 |
| SLC25A51 | 661,84 | 0,03 | 0,02 | 1,47 | 1,43E-01 | 3,09E-01 |

| | | | | | | |
|---|---|---|---|---|---|---|
| UMAD1 | 685,75 | -0,03 | 0,02 | -1,47 | 1,43E-01 | 3,09E-01 |
| PIK3R4 | 1720,99 | -0,02 | 0,02 | -1,47 | 1,43E-01 | 3,09E-01 |
| SYDE2 | 140,16 | 0,07 | 0,05 | 1,47 | 1,43E-01 | 3,09E-01 |
| NBEAL1 | 1229,69 | 0,03 | 0,02 | 1,46 | 1,43E-01 | 3,09E-01 |
| ZFP30 | 544,28 | 0,07 | 0,05 | 1,46 | 1,43E-01 | 3,09E-01 |
| PAX5 | 539,31 | 0,10 | 0,07 | 1,46 | 1,43E-01 | 3,09E-01 |
| UXS1 | 1383,95 | 0,03 | 0,02 | 1,46 | 1,43E-01 | 3,09E-01 |
| ENTPD6 | 640,62 | 0,04 | 0,03 | 1,46 | 1,43E-01 | 3,09E-01 |
| HELZ | 6868,71 | -0,03 | 0,02 | -1,46 | 1,43E-01 | 3,09E-01 |
| FANCB | 227,95 | 0,05 | 0,04 | 1,46 | 1,43E-01 | 3,09E-01 |
| PHF2 | 894,19 | -0,06 | 0,04 | -1,46 | 1,43E-01 | 3,09E-01 |
| SOCS3 | 1027,19 | -0,08 | 0,05 | -1,46 | 1,43E-01 | 3,10E-01 |
| NLRX1 | 346,28 | -0,07 | 0,05 | -1,46 | 1,43E-01 | 3,10E-01 |
| ZNF235 | 588,19 | 0,03 | 0,02 | 1,46 | 1,43E-01 | 3,10E-01 |
| DCAF7 | 1695,22 | -0,05 | 0,03 | -1,46 | 1,43E-01 | 3,10E-01 |
| ELK4 | 4563,78 | 0,03 | 0,02 | 1,46 | 1,43E-01 | 3,10E-01 |
| IGFLR1 | 189,13 | 0,04 | 0,03 | 1,46 | 1,43E-01 | 3,10E-01 |
| CHCHD3 | 993,84 | 0,03 | 0,02 | 1,46 | 1,44E-01 | 3,10E-01 |
| COL18A1 | 1613,97 | -0,09 | 0,06 | -1,46 | 1,44E-01 | 3,10E-01 |
| BLVRB | 8807,50 | 0,10 | 0,07 | 1,46 | 1,44E-01 | 3,10E-01 |
| CREB1 | 6733,24 | -0,04 | 0,03 | -1,46 | 1,44E-01 | 3,10E-01 |
| JADE3 | 85,51 | -0,08 | 0,05 | -1,46 | 1,44E-01 | 3,10E-01 |
| RNF144A | 1243,12 | -0,06 | 0,04 | -1,46 | 1,44E-01 | 3,10E-01 |
| FH | 611,09 | 0,03 | 0,02 | 1,46 | 1,44E-01 | 3,11E-01 |
| ZC3H7B | 791,80 | 0,05 | 0,04 | 1,46 | 1,44E-01 | 3,11E-01 |
| RCN1 | 151,81 | 0,05 | 0,03 | 1,46 | 1,44E-01 | 3,11E-01 |
| ZNF563 | 129,87 | 0,06 | 0,04 | 1,46 | 1,45E-01 | 3,11E-01 |
| ARL5B | 1273,30 | 0,06 | 0,04 | 1,46 | 1,45E-01 | 3,11E-01 |
| ZNF19 | 125,06 | 0,07 | 0,05 | 1,46 | 1,45E-01 | 3,11E-01 |
| ZNF398 | 1157,59 | -0,02 | 0,02 | -1,46 | 1,45E-01 | 3,12E-01 |
| MKKS | 477,14 | 0,04 | 0,03 | 1,46 | 1,45E-01 | 3,12E-01 |
| SARAF | 10195,36 | -0,04 | 0,03 | -1,46 | 1,45E-01 | 3,12E-01 |
| MAN2A2 | 3944,50 | -0,05 | 0,03 | -1,46 | 1,45E-01 | 3,12E-01 |
| FAM76B | 1674,25 | 0,06 | 0,04 | 1,46 | 1,45E-01 | 3,12E-01 |
| CHRNB1 | 215,51 | 0,04 | 0,03 | 1,46 | 1,45E-01 | 3,12E-01 |
| CCT2 | 1261,04 | 0,06 | 0,04 | 1,46 | 1,45E-01 | 3,12E-01 |
| BMPR1A | 469,95 | 0,05 | 0,03 | 1,46 | 1,45E-01 | 3,12E-01 |
| ZNF785 | 225,22 | 0,06 | 0,04 | 1,46 | 1,45E-01 | 3,12E-01 |
| CCL3 | 81,78 | -0,08 | 0,06 | -1,46 | 1,45E-01 | 3,12E-01 |
| MEX3C | 2148,57 | 0,04 | 0,03 | 1,46 | 1,45E-01 | 3,12E-01 |
| ZMPSTE24 | 1682,07 | -0,04 | 0,03 | -1,46 | 1,45E-01 | 3,12E-01 |
| MAN2A1 | 4785,18 | 0,03 | 0,02 | 1,46 | 1,45E-01 | 3,12E-01 |
| RIC8B | 839,41 | 0,03 | 0,02 | 1,46 | 1,45E-01 | 3,12E-01 |
| KCTD5 | 875,59 | -0,02 | 0,02 | -1,46 | 1,45E-01 | 3,12E-01 |
| GPAM | 473,31 | 0,04 | 0,03 | 1,46 | 1,46E-01 | 3,12E-01 |
| ANKRD13D | 2809,17 | -0,06 | 0,04 | -1,46 | 1,46E-01 | 3,12E-01 |

| | | | | | | |
|---|---|---|---|---|---|---|
| CSTB | 431,82 | 0,04 | 0,03 | 1,45 | 1,46E-01 | 3,13E-01 |
| PDK1 | 3842,79 | -0,04 | 0,03 | -1,45 | 1,46E-01 | 3,13E-01 |
| MAPKAP1 | 2057,18 | -0,02 | 0,01 | -1,45 | 1,46E-01 | 3,13E-01 |
| PHF19 | 728,28 | 0,04 | 0,03 | 1,45 | 1,46E-01 | 3,13E-01 |
| CEP152 | 653,34 | -0,04 | 0,03 | -1,45 | 1,46E-01 | 3,13E-01 |
| SETDB1 | 864,44 | -0,05 | 0,03 | -1,45 | 1,46E-01 | 3,13E-01 |
| CRIPT | 729,75 | -0,03 | 0,02 | -1,45 | 1,46E-01 | 3,14E-01 |
| TOGARAM2 | 324,58 | -0,08 | 0,06 | -1,45 | 1,47E-01 | 3,14E-01 |
| B3GAT1 | 250,39 | -0,12 | 0,09 | -1,45 | 1,47E-01 | 3,14E-01 |
| SCN9A | 203,40 | -0,08 | 0,06 | -1,45 | 1,47E-01 | 3,14E-01 |
| COIL | 505,47 | 0,04 | 0,03 | 1,45 | 1,47E-01 | 3,14E-01 |
| AURKAIP1 | 1225,58 | -0,03 | 0,02 | -1,45 | 1,47E-01 | 3,14E-01 |
| ARHGEF2 | 4524,77 | -0,05 | 0,04 | -1,45 | 1,47E-01 | 3,14E-01 |
| ADH5 | 1272,14 | 0,05 | 0,03 | 1,45 | 1,47E-01 | 3,14E-01 |
| PNRC1 | 5206,05 | 0,04 | 0,03 | 1,45 | 1,47E-01 | 3,14E-01 |
| EIF1AY | 872,68 | 0,16 | 0,11 | 1,45 | 1,47E-01 | 3,14E-01 |
| DCUN1D2 | 754,32 | -0,02 | 0,02 | -1,45 | 1,47E-01 | 3,14E-01 |
| PPM1F | 4495,44 | -0,06 | 0,04 | -1,45 | 1,47E-01 | 3,14E-01 |
| TPP1 | 1018,50 | -0,06 | 0,04 | -1,45 | 1,47E-01 | 3,14E-01 |
| UBE2F | 605,53 | -0,03 | 0,02 | -1,45 | 1,47E-01 | 3,14E-01 |
| SCN3A | 190,54 | 0,11 | 0,07 | 1,45 | 1,47E-01 | 3,15E-01 |
| PPFIA4 | 88,42 | 0,13 | 0,09 | 1,45 | 1,47E-01 | 3,15E-01 |
| OBI1 | 562,42 | 0,07 | 0,05 | 1,45 | 1,47E-01 | 3,15E-01 |
| ADAMTS6 | 140,58 | 0,09 | 0,06 | 1,45 | 1,47E-01 | 3,15E-01 |
| FAM107B | 4665,94 | -0,02 | 0,02 | -1,45 | 1,48E-01 | 3,15E-01 |
| ACTL6A | 720,62 | 0,04 | 0,03 | 1,45 | 1,48E-01 | 3,15E-01 |
| LGALS9 | 463,32 | -0,08 | 0,06 | -1,45 | 1,48E-01 | 3,15E-01 |
| ZCRB1 | 211,06 | 0,05 | 0,03 | 1,45 | 1,48E-01 | 3,15E-01 |
| ORMDL3 | 3685,47 | 0,05 | 0,04 | 1,45 | 1,48E-01 | 3,15E-01 |
| PTGIR | 174,90 | 0,06 | 0,04 | 1,45 | 1,48E-01 | 3,15E-01 |
| APEH | 1329,78 | 0,02 | 0,02 | 1,45 | 1,48E-01 | 3,15E-01 |
| ARMC6 | 307,75 | 0,03 | 0,02 | 1,45 | 1,48E-01 | 3,16E-01 |
| NMT2 | 702,23 | 0,04 | 0,03 | 1,45 | 1,48E-01 | 3,16E-01 |
| C17orf75 | 363,96 | 0,03 | 0,02 | 1,45 | 1,48E-01 | 3,16E-01 |
| RAPGEFL1 | 125,57 | -0,11 | 0,07 | -1,45 | 1,48E-01 | 3,16E-01 |
| SHPRH | 3312,98 | 0,04 | 0,03 | 1,44 | 1,48E-01 | 3,16E-01 |
| RPUSD3 | 490,66 | 0,03 | 0,02 | 1,44 | 1,49E-01 | 3,17E-01 |
| EIF4A1 | 570,69 | 0,04 | 0,02 | 1,44 | 1,49E-01 | 3,17E-01 |
| TRAF5 | 2357,12 | 0,04 | 0,03 | 1,44 | 1,49E-01 | 3,17E-01 |
| SAT2 | 156,06 | 0,05 | 0,03 | 1,44 | 1,49E-01 | 3,17E-01 |
| TXNL4A | 1256,78 | 0,02 | 0,01 | 1,44 | 1,49E-01 | 3,17E-01 |
| DPP4 | 1075,57 | 0,05 | 0,04 | 1,44 | 1,49E-01 | 3,17E-01 |
| TMEM205 | 418,51 | 0,03 | 0,02 | 1,44 | 1,49E-01 | 3,17E-01 |
| MLLT3 | 1227,82 | 0,05 | 0,03 | 1,44 | 1,49E-01 | 3,17E-01 |
| GMPPB | 554,52 | 0,03 | 0,02 | 1,44 | 1,49E-01 | 3,17E-01 |
| EPS15 | 4790,46 | -0,03 | 0,02 | -1,44 | 1,49E-01 | 3,17E-01 |

| | | | | | | |
|---|---|---|---|---|---|---|
| NOP10 | 1130,91 | -0,05 | 0,04 | -1,44 | 1,50E-01 | 3,18E-01 |
| EIF3M | 2308,65 | 0,04 | 0,03 | 1,44 | 1,49E-01 | 3,18E-01 |
| ARK2C | 159,23 | -0,11 | 0,08 | -1,44 | 1,50E-01 | 3,18E-01 |
| RRS1 | 253,52 | 0,04 | 0,02 | 1,44 | 1,49E-01 | 3,18E-01 |
| FHDC1 | 90,50 | -0,09 | 0,06 | -1,44 | 1,50E-01 | 3,18E-01 |
| KIF21B | 2617,84 | -0,07 | 0,05 | -1,44 | 1,50E-01 | 3,18E-01 |
| DNAJC16 | 737,30 | 0,02 | 0,02 | 1,44 | 1,50E-01 | 3,18E-01 |
| SCD | 86,63 | 0,08 | 0,05 | 1,44 | 1,50E-01 | 3,18E-01 |
| FCHO2 | 1951,47 | -0,05 | 0,04 | -1,44 | 1,50E-01 | 3,18E-01 |
| ZBTB21 | 709,17 | 0,03 | 0,02 | 1,44 | 1,50E-01 | 3,18E-01 |
| KAT5 | 893,17 | -0,02 | 0,02 | -1,44 | 1,50E-01 | 3,18E-01 |
| PTDSS2 | 189,37 | 0,05 | 0,04 | 1,44 | 1,50E-01 | 3,18E-01 |
| MANBAL | 356,12 | -0,03 | 0,02 | -1,44 | 1,50E-01 | 3,19E-01 |
| GGACT | 198,94 | 0,04 | 0,03 | 1,44 | 1,50E-01 | 3,19E-01 |
| FBXO5 | 198,16 | 0,05 | 0,04 | 1,44 | 1,51E-01 | 3,19E-01 |
| GOLGA7 | 1485,10 | -0,03 | 0,02 | -1,44 | 1,51E-01 | 3,19E-01 |
| SLC25A40 | 1318,76 | 0,05 | 0,03 | 1,44 | 1,51E-01 | 3,19E-01 |
| RAP1A | 3824,31 | -0,04 | 0,02 | -1,44 | 1,51E-01 | 3,19E-01 |
| PSMD11 | 987,88 | -0,02 | 0,01 | -1,44 | 1,51E-01 | 3,19E-01 |
| CAPS | 376,83 | 0,05 | 0,03 | 1,44 | 1,51E-01 | 3,19E-01 |
| LYSET | 162,40 | 0,04 | 0,03 | 1,44 | 1,51E-01 | 3,19E-01 |
| MTMR9 | 1644,93 | 0,03 | 0,02 | 1,44 | 1,51E-01 | 3,19E-01 |
| SEMA4C | 518,72 | 0,04 | 0,03 | 1,44 | 1,51E-01 | 3,20E-01 |
| TPRKB | 314,43 | 0,04 | 0,03 | 1,44 | 1,51E-01 | 3,20E-01 |
| EFHD2 | 3743,04 | -0,06 | 0,04 | -1,43 | 1,51E-01 | 3,20E-01 |
| ANKS3 | 254,82 | 0,05 | 0,03 | 1,43 | 1,52E-01 | 3,21E-01 |
| ALG14 | 107,08 | 0,05 | 0,03 | 1,43 | 1,52E-01 | 3,21E-01 |
| PACS1 | 2769,21 | -0,07 | 0,05 | -1,43 | 1,52E-01 | 3,21E-01 |
| MYO19 | 770,06 | 0,04 | 0,03 | 1,43 | 1,52E-01 | 3,21E-01 |
| PARVG | 3851,33 | -0,04 | 0,03 | -1,43 | 1,52E-01 | 3,21E-01 |
| RPS6KA3 | 8256,80 | -0,03 | 0,02 | -1,43 | 1,52E-01 | 3,21E-01 |
| ETFRF1 | 326,04 | 0,06 | 0,04 | 1,43 | 1,52E-01 | 3,21E-01 |
| PRKAB1 | 919,84 | -0,02 | 0,02 | -1,43 | 1,52E-01 | 3,22E-01 |
| NANP | 130,57 | 0,04 | 0,03 | 1,43 | 1,52E-01 | 3,22E-01 |
| PANK1 | 109,27 | 0,07 | 0,05 | 1,43 | 1,53E-01 | 3,22E-01 |
| C1QTNF6 | 417,89 | 0,04 | 0,03 | 1,43 | 1,53E-01 | 3,23E-01 |
| RRAS2 | 785,69 | 0,06 | 0,04 | 1,43 | 1,53E-01 | 3,23E-01 |
| PHB1 | 1077,28 | 0,04 | 0,03 | 1,43 | 1,53E-01 | 3,23E-01 |
| PIK3IP1 | 4443,06 | 0,04 | 0,03 | 1,43 | 1,53E-01 | 3,23E-01 |
| SLC6A12 | 119,21 | -0,10 | 0,07 | -1,43 | 1,53E-01 | 3,23E-01 |
| COX17 | 418,88 | 0,03 | 0,02 | 1,43 | 1,53E-01 | 3,24E-01 |
| WTAP | 4737,05 | -0,04 | 0,03 | -1,43 | 1,53E-01 | 3,24E-01 |
| PLBD2 | 190,74 | 0,08 | 0,06 | 1,43 | 1,54E-01 | 3,24E-01 |
| ZNF596 | 100,01 | 0,07 | 0,05 | 1,43 | 1,54E-01 | 3,24E-01 |
| RPA1 | 1073,96 | 0,04 | 0,03 | 1,43 | 1,54E-01 | 3,24E-01 |
| RALGDS | 491,69 | 0,06 | 0,04 | 1,43 | 1,54E-01 | 3,25E-01 |

| | | | | | | |
|---|---|---|---|---|---|---|
| ZNF426 | 811,90 | 0,04 | 0,03 | 1,43 | 1,54E-01 | 3,25E-01 |
| POLR2J3.1 | 108,10 | 0,10 | 0,07 | 1,43 | 1,54E-01 | 3,25E-01 |
| BTN3A3 | 2973,86 | -0,04 | 0,03 | -1,42 | 1,54E-01 | 3,25E-01 |
| CCDC90B | 903,82 | 0,04 | 0,03 | 1,42 | 1,54E-01 | 3,25E-01 |
| CLN5 | 767,37 | 0,03 | 0,02 | 1,42 | 1,55E-01 | 3,25E-01 |
| BACH2 | 1500,83 | 0,06 | 0,04 | 1,42 | 1,55E-01 | 3,26E-01 |
| RICTOR | 14299,22 | -0,05 | 0,04 | -1,42 | 1,55E-01 | 3,26E-01 |
| C5 | 235,12 | -0,05 | 0,03 | -1,42 | 1,55E-01 | 3,26E-01 |
| TIMM17A | 601,00 | 0,04 | 0,03 | 1,42 | 1,55E-01 | 3,26E-01 |
| RTRAF | 1083,44 | 0,05 | 0,03 | 1,42 | 1,55E-01 | 3,26E-01 |
| USP30 | 287,61 | -0,03 | 0,02 | -1,42 | 1,55E-01 | 3,26E-01 |
| THTPA | 156,99 | 0,04 | 0,03 | 1,42 | 1,55E-01 | 3,26E-01 |
| ZBTB42 | 127,84 | 0,07 | 0,05 | 1,42 | 1,55E-01 | 3,26E-01 |
| GALNT5 | 87,75 | 0,10 | 0,07 | 1,42 | 1,55E-01 | 3,26E-01 |
| GFM2 | 844,48 | 0,04 | 0,03 | 1,42 | 1,55E-01 | 3,26E-01 |
| FGR | 9907,09 | -0,04 | 0,03 | -1,42 | 1,55E-01 | 3,26E-01 |
| ARSK | 216,73 | 0,04 | 0,03 | 1,42 | 1,55E-01 | 3,26E-01 |
| NINJ1 | 919,63 | -0,07 | 0,05 | -1,42 | 1,55E-01 | 3,26E-01 |
| ENSG000002 | 229,97 | 0,05 | 0,03 | 1,42 | 1,55E-01 | 3,26E-01 |
| BCL7B | 297,29 | 0,03 | 0,02 | 1,42 | 1,55E-01 | 3,26E-01 |
| SMIM3 | 327,47 | -0,05 | 0,03 | -1,42 | 1,55E-01 | 3,26E-01 |
| B3GAT3 | 340,69 | 0,04 | 0,02 | 1,42 | 1,55E-01 | 3,26E-01 |
| RPS4X | 10935,57 | 0,05 | 0,04 | 1,42 | 1,55E-01 | 3,26E-01 |
| SGTB | 1750,03 | 0,04 | 0,03 | 1,42 | 1,55E-01 | 3,26E-01 |
| PRUNE2 | 96,52 | -0,16 | 0,11 | -1,42 | 1,55E-01 | 3,26E-01 |
| TTC33 | 564,96 | 0,07 | 0,05 | 1,42 | 1,56E-01 | 3,26E-01 |
| AKNA | 7906,83 | -0,08 | 0,05 | -1,42 | 1,56E-01 | 3,26E-01 |
| PDXDC1 | 2162,85 | -0,03 | 0,02 | -1,42 | 1,56E-01 | 3,26E-01 |
| DIDO1 | 4383,65 | -0,02 | 0,02 | -1,42 | 1,56E-01 | 3,27E-01 |
| MOB2 | 1409,09 | 0,03 | 0,02 | 1,42 | 1,56E-01 | 3,27E-01 |
| BTBD9 | 1302,80 | -0,03 | 0,02 | -1,42 | 1,56E-01 | 3,27E-01 |
| DNAJB9 | 545,84 | 0,07 | 0,05 | 1,42 | 1,56E-01 | 3,27E-01 |
| IFNGR1 | 6228,07 | -0,06 | 0,04 | -1,42 | 1,57E-01 | 3,28E-01 |
| ATP6V1D | 1003,31 | -0,04 | 0,03 | -1,42 | 1,57E-01 | 3,28E-01 |
| DCAF16 | 1943,77 | 0,02 | 0,02 | 1,42 | 1,57E-01 | 3,28E-01 |
| PEX1 | 1111,86 | 0,03 | 0,02 | 1,42 | 1,57E-01 | 3,28E-01 |
| COQ8A | 1357,11 | -0,04 | 0,03 | -1,42 | 1,57E-01 | 3,28E-01 |
| DCTN1 | 1564,75 | -0,04 | 0,03 | -1,42 | 1,57E-01 | 3,28E-01 |
| WDR24 | 88,10 | 0,07 | 0,05 | 1,42 | 1,57E-01 | 3,28E-01 |
| CCDC51 | 139,81 | 0,03 | 0,02 | 1,42 | 1,57E-01 | 3,28E-01 |
| ZNF550 | 678,00 | 0,04 | 0,03 | 1,42 | 1,57E-01 | 3,28E-01 |
| CCT5 | 3007,60 | -0,03 | 0,02 | -1,41 | 1,57E-01 | 3,28E-01 |
| DHX35 | 615,34 | 0,02 | 0,02 | 1,41 | 1,57E-01 | 3,28E-01 |
| ATP8B2 | 3991,77 | 0,04 | 0,03 | 1,41 | 1,57E-01 | 3,28E-01 |
| MED1 | 1646,37 | -0,01 | 0,01 | -1,41 | 1,57E-01 | 3,29E-01 |
| ULK3 | 290,45 | 0,06 | 0,04 | 1,41 | 1,57E-01 | 3,29E-01 |

| | | | | | | |
|---|---|---|---|---|---|---|
| CHMP5 | 801,40 | -0,08 | 0,06 | -1,41 | 1,57E-01 | 3,29E-01 |
| CACNB3 | 72,30 | 0,09 | 0,06 | 1,41 | 1,57E-01 | 3,29E-01 |
| B3GALT6 | 513,53 | 0,03 | 0,02 | 1,41 | 1,57E-01 | 3,29E-01 |
| WRAP53 | 129,78 | 0,03 | 0,02 | 1,41 | 1,58E-01 | 3,29E-01 |
| ELK3 | 756,54 | 0,03 | 0,02 | 1,41 | 1,58E-01 | 3,29E-01 |
| IL12RB2 | 166,16 | -0,08 | 0,06 | -1,41 | 1,58E-01 | 3,29E-01 |
| TIGD1 | 476,03 | 0,06 | 0,05 | 1,41 | 1,58E-01 | 3,29E-01 |
| ZNF513 | 323,50 | 0,02 | 0,02 | 1,41 | 1,58E-01 | 3,29E-01 |
| CDK2AP1 | 297,58 | 0,04 | 0,03 | 1,41 | 1,58E-01 | 3,29E-01 |
| GCSAM | 402,82 | 0,05 | 0,04 | 1,41 | 1,58E-01 | 3,29E-01 |
| BACE2 | 353,60 | 0,07 | 0,05 | 1,41 | 1,58E-01 | 3,30E-01 |
| MDH2 | 490,73 | 0,03 | 0,02 | 1,41 | 1,58E-01 | 3,30E-01 |
| NADSYN1 | 2692,60 | -0,04 | 0,03 | -1,41 | 1,58E-01 | 3,30E-01 |
| POU5F2 | 1128,63 | -0,06 | 0,04 | -1,41 | 1,58E-01 | 3,30E-01 |
| TP53RK | 1014,07 | 0,04 | 0,03 | 1,41 | 1,58E-01 | 3,30E-01 |
| DDX1 | 1076,92 | 0,04 | 0,03 | 1,41 | 1,58E-01 | 3,30E-01 |
| SWSAP1 | 99,91 | 0,04 | 0,03 | 1,41 | 1,58E-01 | 3,30E-01 |
| ZNF668 | 112,49 | 0,05 | 0,03 | 1,41 | 1,58E-01 | 3,30E-01 |
| SGCE | 100,96 | 0,13 | 0,09 | 1,41 | 1,59E-01 | 3,30E-01 |
| YME1L1 | 4527,51 | -0,03 | 0,02 | -1,41 | 1,59E-01 | 3,30E-01 |
| MUL1 | 514,19 | -0,02 | 0,02 | -1,41 | 1,59E-01 | 3,30E-01 |
| RETREG2 | 2514,89 | 0,04 | 0,03 | 1,41 | 1,59E-01 | 3,30E-01 |
| LYPLA1 | 1847,52 | 0,04 | 0,03 | 1,41 | 1,59E-01 | 3,31E-01 |
| NOP14 | 779,81 | 0,03 | 0,02 | 1,41 | 1,59E-01 | 3,31E-01 |
| SEC61A2 | 490,82 | -0,04 | 0,03 | -1,41 | 1,59E-01 | 3,31E-01 |
| ERH | 826,71 | 0,08 | 0,06 | 1,41 | 1,59E-01 | 3,31E-01 |
| ARL2 | 163,52 | 0,03 | 0,02 | 1,41 | 1,59E-01 | 3,31E-01 |
| GPATCH4 | 347,50 | 0,04 | 0,03 | 1,41 | 1,59E-01 | 3,31E-01 |
| TXNDC9 | 507,31 | 0,07 | 0,05 | 1,41 | 1,60E-01 | 3,31E-01 |
| PRPSAP2 | 1054,98 | 0,02 | 0,02 | 1,41 | 1,60E-01 | 3,32E-01 |
| CSRNP2 | 531,82 | -0,03 | 0,02 | -1,41 | 1,60E-01 | 3,32E-01 |
| RBM12B | 3832,71 | 0,06 | 0,04 | 1,41 | 1,60E-01 | 3,32E-01 |
| ARHGEF7 | 2143,48 | -0,03 | 0,02 | -1,41 | 1,60E-01 | 3,32E-01 |
| CEPT1 | 2617,03 | 0,04 | 0,03 | 1,41 | 1,60E-01 | 3,32E-01 |
| AGFG2 | 352,02 | 0,05 | 0,04 | 1,41 | 1,60E-01 | 3,32E-01 |
| BAG2 | 275,54 | 0,05 | 0,04 | 1,41 | 1,60E-01 | 3,32E-01 |
| USP44 | 156,36 | 0,08 | 0,06 | 1,41 | 1,60E-01 | 3,32E-01 |
| COPS7A | 595,98 | -0,03 | 0,02 | -1,40 | 1,60E-01 | 3,32E-01 |
| GOSR1 | 1270,05 | -0,02 | 0,02 | -1,40 | 1,60E-01 | 3,32E-01 |
| PRPF3 | 1852,40 | -0,02 | 0,01 | -1,40 | 1,60E-01 | 3,32E-01 |
| STXBP3 | 4391,12 | -0,05 | 0,03 | -1,40 | 1,60E-01 | 3,33E-01 |
| CDKN2AIP | 1055,77 | 0,05 | 0,04 | 1,40 | 1,61E-01 | 3,33E-01 |
| ZNHIT6 | 746,23 | 0,04 | 0,03 | 1,40 | 1,61E-01 | 3,33E-01 |
| PBX1 | 358,51 | 0,09 | 0,07 | 1,40 | 1,61E-01 | 3,33E-01 |
| DYNC2H1 | 481,97 | 0,05 | 0,04 | 1,40 | 1,61E-01 | 3,33E-01 |
| UBR4 | 7120,86 | -0,05 | 0,04 | -1,40 | 1,61E-01 | 3,33E-01 |

| | | | | | | |
|---|---|---|---|---|---|---|
| ANKRD39 | 148,84 | 0,03 | 0,02 | 1,40 | 1,61E-01 | 3,33E-01 |
| CACNA1D | 225,09 | -0,11 | 0,08 | -1,40 | 1,61E-01 | 3,33E-01 |
| OPLAH | 123,95 | -0,09 | 0,07 | -1,40 | 1,61E-01 | 3,33E-01 |
| AGO3 | 3893,24 | -0,03 | 0,02 | -1,40 | 1,61E-01 | 3,33E-01 |
| KHDRBS2 | 126,51 | 0,09 | 0,06 | 1,40 | 1,61E-01 | 3,33E-01 |
| MIDN | 1332,65 | -0,07 | 0,05 | -1,40 | 1,61E-01 | 3,33E-01 |
| NDUFV3 | 1112,61 | 0,02 | 0,02 | 1,40 | 1,61E-01 | 3,34E-01 |
| MINDY3 | 1165,05 | 0,04 | 0,03 | 1,40 | 1,61E-01 | 3,34E-01 |
| H2AC25 | 324,81 | 0,06 | 0,05 | 1,40 | 1,61E-01 | 3,34E-01 |
| FMR1 | 3342,65 | -0,04 | 0,03 | -1,40 | 1,61E-01 | 3,34E-01 |
| FADS2 | 334,02 | -0,14 | 0,10 | -1,40 | 1,62E-01 | 3,34E-01 |
| ERAP2 | 3368,64 | 0,07 | 0,05 | 1,40 | 1,61E-01 | 3,34E-01 |
| OMA1 | 1315,81 | 0,04 | 0,03 | 1,40 | 1,62E-01 | 3,34E-01 |
| B3GALT9 | 121,82 | 0,04 | 0,03 | 1,40 | 1,62E-01 | 3,34E-01 |
| CCR5 | 711,86 | 0,07 | 0,05 | 1,40 | 1,62E-01 | 3,34E-01 |
| OPRL1 | 247,18 | -0,06 | 0,04 | -1,40 | 1,62E-01 | 3,34E-01 |
| G6PD | 674,84 | -0,07 | 0,05 | -1,40 | 1,62E-01 | 3,35E-01 |
| ARID5A | 636,28 | -0,06 | 0,04 | -1,40 | 1,62E-01 | 3,35E-01 |
| FNIP2 | 926,75 | 0,03 | 0,02 | 1,40 | 1,62E-01 | 3,35E-01 |
| ZNF222 | 115,23 | -0,05 | 0,04 | -1,40 | 1,62E-01 | 3,35E-01 |
| CYB5R1 | 762,32 | -0,03 | 0,02 | -1,40 | 1,63E-01 | 3,35E-01 |
| RPA3 | 253,64 | 0,05 | 0,04 | 1,40 | 1,63E-01 | 3,35E-01 |
| CASP8AP2 | 1467,96 | 0,06 | 0,05 | 1,40 | 1,63E-01 | 3,35E-01 |
| FBH1 | 1833,19 | -0,03 | 0,02 | -1,40 | 1,63E-01 | 3,36E-01 |
| KYAT3 | 575,78 | -0,03 | 0,02 | -1,40 | 1,63E-01 | 3,36E-01 |
| INAFM1 | 133,18 | -0,05 | 0,03 | -1,39 | 1,63E-01 | 3,36E-01 |
| TOMM5 | 205,45 | 0,04 | 0,03 | 1,39 | 1,63E-01 | 3,36E-01 |
| DPY19L1 | 815,15 | 0,04 | 0,03 | 1,39 | 1,63E-01 | 3,36E-01 |
| CD5 | 633,89 | 0,06 | 0,04 | 1,39 | 1,63E-01 | 3,36E-01 |
| EIF2B3 | 355,58 | 0,02 | 0,02 | 1,39 | 1,63E-01 | 3,36E-01 |
| CRYL1 | 343,79 | 0,03 | 0,02 | 1,39 | 1,63E-01 | 3,36E-01 |
| MACC1 | 94,51 | 0,08 | 0,05 | 1,39 | 1,63E-01 | 3,37E-01 |
| SUZ12 | 1308,48 | 0,05 | 0,04 | 1,39 | 1,64E-01 | 3,37E-01 |
| SYCP2 | 410,08 | 0,06 | 0,05 | 1,39 | 1,64E-01 | 3,37E-01 |
| MSH3 | 850,07 | 0,03 | 0,02 | 1,39 | 1,64E-01 | 3,37E-01 |
| EDA | 90,89 | 0,10 | 0,07 | 1,39 | 1,64E-01 | 3,37E-01 |
| CYFIP2 | 7598,86 | -0,04 | 0,03 | -1,39 | 1,64E-01 | 3,37E-01 |
| SLC26A11 | 283,65 | 0,05 | 0,04 | 1,39 | 1,64E-01 | 3,37E-01 |
| FOXJ3 | 2147,81 | -0,02 | 0,02 | -1,39 | 1,64E-01 | 3,37E-01 |
| GPR19 | 74,23 | 0,05 | 0,04 | 1,39 | 1,64E-01 | 3,37E-01 |
| SELENOS | 495,72 | 0,03 | 0,02 | 1,39 | 1,64E-01 | 3,38E-01 |
| ZNF226 | 1340,57 | 0,05 | 0,04 | 1,39 | 1,64E-01 | 3,38E-01 |
| ZNF484 | 958,15 | 0,06 | 0,05 | 1,39 | 1,64E-01 | 3,38E-01 |
| TSC22D1 | 1202,50 | 0,04 | 0,03 | 1,39 | 1,65E-01 | 3,38E-01 |
| HDX | 113,15 | 0,05 | 0,04 | 1,39 | 1,65E-01 | 3,38E-01 |
| ZNF138 | 414,26 | 0,06 | 0,04 | 1,39 | 1,65E-01 | 3,39E-01 |

| | | | | | | |
|---|---|---|---|---|---|---|
| BST2 | 1674,95 | -0,06 | 0,04 | -1,39 | 1,65E-01 | 3,39E-01 |
| ABCB1 | 668,04 | -0,05 | 0,04 | -1,39 | 1,65E-01 | 3,39E-01 |
| ARHGAP11A | 193,42 | 0,06 | 0,05 | 1,39 | 1,65E-01 | 3,39E-01 |
| NCAPD2 | 1070,80 | -0,04 | 0,03 | -1,39 | 1,65E-01 | 3,40E-01 |
| PRSS33 | 99,84 | 0,13 | 0,09 | 1,39 | 1,66E-01 | 3,40E-01 |
| CLEC4A | 310,30 | -0,06 | 0,04 | -1,39 | 1,66E-01 | 3,40E-01 |
| CCDC65 | 144,23 | 0,06 | 0,05 | 1,39 | 1,66E-01 | 3,40E-01 |
| MYCBP2 | 14467,98 | -0,03 | 0,02 | -1,39 | 1,66E-01 | 3,40E-01 |
| EXOC6B | 695,22 | 0,03 | 0,02 | 1,38 | 1,66E-01 | 3,41E-01 |
| NAP1L4 | 5476,94 | 0,03 | 0,02 | 1,38 | 1,66E-01 | 3,41E-01 |
| MIGA2 | 615,13 | 0,04 | 0,03 | 1,38 | 1,66E-01 | 3,41E-01 |
| MPC2 | 934,46 | 0,04 | 0,03 | 1,38 | 1,66E-01 | 3,41E-01 |
| RLIM | 5583,75 | -0,04 | 0,03 | -1,38 | 1,66E-01 | 3,41E-01 |
| TMEM259 | 1222,05 | 0,06 | 0,04 | 1,38 | 1,67E-01 | 3,42E-01 |
| POU6F1 | 180,75 | 0,06 | 0,05 | 1,38 | 1,67E-01 | 3,42E-01 |
| GOLGA1 | 762,27 | -0,03 | 0,02 | -1,38 | 1,67E-01 | 3,42E-01 |
| AKIP1 | 193,32 | 0,03 | 0,02 | 1,38 | 1,67E-01 | 3,42E-01 |
| N4BP3 | 94,07 | 0,09 | 0,06 | 1,38 | 1,67E-01 | 3,42E-01 |
| DNMBP | 405,88 | -0,06 | 0,04 | -1,38 | 1,67E-01 | 3,42E-01 |
| ZNF512B | 147,40 | 0,09 | 0,07 | 1,38 | 1,67E-01 | 3,42E-01 |
| TMEM62 | 787,91 | 0,03 | 0,02 | 1,38 | 1,67E-01 | 3,42E-01 |
| FBXO34 | 1328,28 | -0,02 | 0,02 | -1,38 | 1,67E-01 | 3,42E-01 |
| KLF13 | 6477,25 | -0,04 | 0,03 | -1,38 | 1,67E-01 | 3,42E-01 |
| CACNB1 | 146,21 | 0,06 | 0,05 | 1,38 | 1,67E-01 | 3,42E-01 |
| JAKMIP2 | 466,31 | 0,07 | 0,05 | 1,38 | 1,67E-01 | 3,42E-01 |
| NUDT16 | 1687,74 | -0,03 | 0,02 | -1,38 | 1,67E-01 | 3,42E-01 |
| NOB1 | 148,18 | 0,04 | 0,03 | 1,38 | 1,68E-01 | 3,42E-01 |
| TNFAIP1 | 393,73 | -0,03 | 0,02 | -1,38 | 1,68E-01 | 3,43E-01 |
| C4orf3 | 3202,07 | -0,05 | 0,04 | -1,38 | 1,68E-01 | 3,43E-01 |
| ZMYND11 | 1429,18 | 0,03 | 0,03 | 1,38 | 1,68E-01 | 3,43E-01 |
| ZNF585B | 328,17 | 0,03 | 0,02 | 1,38 | 1,68E-01 | 3,43E-01 |
| MFSD11 | 785,33 | -0,02 | 0,01 | -1,38 | 1,68E-01 | 3,43E-01 |
| MYH9 | 16354,97 | -0,07 | 0,05 | -1,38 | 1,68E-01 | 3,43E-01 |
| ZSCAN21 | 153,61 | 0,02 | 0,02 | 1,38 | 1,68E-01 | 3,43E-01 |
| FADS1 | 344,90 | 0,05 | 0,04 | 1,38 | 1,68E-01 | 3,43E-01 |
| LRCH4 | 1065,82 | -0,05 | 0,03 | -1,38 | 1,68E-01 | 3,44E-01 |
| RBBP8 | 304,57 | 0,06 | 0,04 | 1,38 | 1,68E-01 | 3,44E-01 |
| MED19 | 114,01 | 0,03 | 0,02 | 1,38 | 1,69E-01 | 3,44E-01 |
| HMGB2 | 2020,07 | -0,08 | 0,06 | -1,38 | 1,69E-01 | 3,44E-01 |
| LTBP3 | 422,99 | 0,09 | 0,06 | 1,38 | 1,68E-01 | 3,44E-01 |
| CLC | 1981,32 | 0,11 | 0,08 | 1,38 | 1,69E-01 | 3,44E-01 |
| MEN1 | 171,11 | 0,06 | 0,04 | 1,38 | 1,69E-01 | 3,44E-01 |
| SLC25A19 | 180,88 | 0,03 | 0,03 | 1,38 | 1,69E-01 | 3,44E-01 |
| MRPL40 | 143,80 | 0,06 | 0,04 | 1,38 | 1,69E-01 | 3,44E-01 |
| ZNF532 | 419,53 | 0,05 | 0,04 | 1,38 | 1,69E-01 | 3,44E-01 |
| DLST | 1631,84 | -0,02 | 0,02 | -1,37 | 1,69E-01 | 3,44E-01 |

| | | | | | | |
|---|---|---|---|---|---|---|
| CAND1 | 3438,34 | 0,04 | 0,03 | 1,37 | 1,69E-01 | 3,45E-01 |
| CDC16 | 1413,17 | 0,03 | 0,02 | 1,37 | 1,70E-01 | 3,45E-01 |
| YWHAE | 824,91 | -0,03 | 0,02 | -1,37 | 1,70E-01 | 3,45E-01 |
| ORC2 | 1264,68 | -0,02 | 0,02 | -1,37 | 1,70E-01 | 3,46E-01 |
| MYO9A | 2344,84 | 0,03 | 0,02 | 1,37 | 1,70E-01 | 3,46E-01 |
| ZUP1 | 466,48 | 0,05 | 0,03 | 1,37 | 1,70E-01 | 3,46E-01 |
| CDC42BPA | 381,01 | 0,08 | 0,06 | 1,37 | 1,70E-01 | 3,46E-01 |
| SPATS2L | 207,34 | -0,10 | 0,08 | -1,37 | 1,71E-01 | 3,47E-01 |
| EBAG9 | 469,92 | 0,05 | 0,03 | 1,37 | 1,71E-01 | 3,47E-01 |
| GPATCH11 | 601,42 | 0,06 | 0,04 | 1,37 | 1,71E-01 | 3,47E-01 |
| ESD | 944,90 | 0,04 | 0,03 | 1,37 | 1,71E-01 | 3,47E-01 |
| POLE3 | 856,10 | 0,03 | 0,02 | 1,37 | 1,71E-01 | 3,47E-01 |
| DHX37 | 296,96 | 0,06 | 0,04 | 1,37 | 1,71E-01 | 3,47E-01 |
| NCBP2 | 1362,58 | 0,03 | 0,02 | 1,37 | 1,71E-01 | 3,47E-01 |
| RAC2 | 2440,56 | -0,06 | 0,04 | -1,37 | 1,71E-01 | 3,47E-01 |
| CXCL8 | 630,18 | 0,11 | 0,08 | 1,37 | 1,71E-01 | 3,47E-01 |
| GRAMD1A | 1394,65 | -0,07 | 0,05 | -1,37 | 1,71E-01 | 3,47E-01 |
| CCSAP | 1709,65 | -0,04 | 0,03 | -1,37 | 1,71E-01 | 3,47E-01 |
| WASHC5 | 1678,13 | -0,02 | 0,01 | -1,37 | 1,71E-01 | 3,47E-01 |
| SEPTIN1 | 1448,86 | 0,03 | 0,03 | 1,37 | 1,71E-01 | 3,48E-01 |
| SPN | 2625,00 | 0,05 | 0,04 | 1,37 | 1,71E-01 | 3,48E-01 |
| FAAH | 187,52 | 0,07 | 0,05 | 1,37 | 1,71E-01 | 3,48E-01 |
| DUT | 1532,65 | 0,05 | 0,03 | 1,37 | 1,71E-01 | 3,48E-01 |
| CLEC2D | 8005,74 | 0,04 | 0,03 | 1,37 | 1,72E-01 | 3,48E-01 |
| PSMD8 | 594,44 | 0,02 | 0,02 | 1,37 | 1,72E-01 | 3,49E-01 |
| NOTCH1 | 3769,65 | -0,09 | 0,07 | -1,37 | 1,72E-01 | 3,49E-01 |
| RASGRP4 | 842,67 | -0,10 | 0,07 | -1,37 | 1,72E-01 | 3,49E-01 |
| TGM2 | 168,30 | 0,11 | 0,08 | 1,37 | 1,72E-01 | 3,49E-01 |
| LIPA | 1590,77 | 0,05 | 0,03 | 1,37 | 1,72E-01 | 3,49E-01 |
| ZNF860 | 198,90 | 0,10 | 0,07 | 1,37 | 1,72E-01 | 3,49E-01 |
| UGCG | 1590,37 | 0,05 | 0,04 | 1,37 | 1,72E-01 | 3,49E-01 |
| C12orf43 | 263,64 | 0,02 | 0,02 | 1,37 | 1,72E-01 | 3,49E-01 |
| ATP9B | 3034,73 | 0,03 | 0,02 | 1,36 | 1,72E-01 | 3,49E-01 |
| TMEM123 | 7398,10 | 0,07 | 0,05 | 1,36 | 1,72E-01 | 3,49E-01 |
| DPY19L3 | 1367,57 | -0,04 | 0,03 | -1,36 | 1,72E-01 | 3,49E-01 |
| STXBP4 | 522,44 | 0,04 | 0,03 | 1,36 | 1,72E-01 | 3,49E-01 |
| COBLL1 | 642,22 | 0,07 | 0,05 | 1,36 | 1,73E-01 | 3,49E-01 |
| FNTB | 204,67 | -0,04 | 0,03 | -1,36 | 1,73E-01 | 3,49E-01 |
| FZD1 | 225,42 | 0,05 | 0,04 | 1,36 | 1,73E-01 | 3,49E-01 |
| AKAP12 | 240,73 | -0,14 | 0,10 | -1,36 | 1,73E-01 | 3,49E-01 |
| ZNF419 | 364,62 | 0,03 | 0,02 | 1,36 | 1,73E-01 | 3,49E-01 |
| APIP | 404,21 | 0,05 | 0,03 | 1,36 | 1,73E-01 | 3,49E-01 |
| ZNF506 | 2586,73 | 0,03 | 0,02 | 1,36 | 1,73E-01 | 3,50E-01 |
| GNG11 | 559,24 | 0,10 | 0,07 | 1,36 | 1,73E-01 | 3,50E-01 |
| CDK13 | 7069,82 | -0,02 | 0,01 | -1,36 | 1,73E-01 | 3,50E-01 |
| GBF1 | 1243,35 | -0,05 | 0,03 | -1,36 | 1,73E-01 | 3,50E-01 |

| | | | | | | |
|---|---|---|---|---|---|---|
| UQCRFS1 | 933,06 | 0,03 | 0,02 | 1,36 | 1,73E-01 | 3,50E-01 |
| ECD | 1087,72 | -0,02 | 0,01 | -1,36 | 1,74E-01 | 3,50E-01 |
| GSE1 | 1570,66 | 0,05 | 0,04 | 1,36 | 1,74E-01 | 3,50E-01 |
| FAM221A | 186,39 | 0,04 | 0,03 | 1,36 | 1,74E-01 | 3,50E-01 |
| RNF157 | 404,58 | 0,06 | 0,05 | 1,36 | 1,74E-01 | 3,50E-01 |
| LACC1 | 162,56 | 0,06 | 0,04 | 1,36 | 1,74E-01 | 3,50E-01 |
| RTEL1 | 76,40 | 0,06 | 0,04 | 1,36 | 1,74E-01 | 3,50E-01 |
| LDLRAP1 | 1297,40 | 0,06 | 0,04 | 1,36 | 1,74E-01 | 3,50E-01 |
| RHOBTB2 | 223,18 | 0,06 | 0,05 | 1,36 | 1,74E-01 | 3,51E-01 |
| METRN | 76,67 | 0,06 | 0,05 | 1,36 | 1,74E-01 | 3,51E-01 |
| RPS19BP1 | 679,60 | 0,03 | 0,02 | 1,36 | 1,74E-01 | 3,51E-01 |
| ARHGAP12 | 1480,92 | 0,04 | 0,03 | 1,36 | 1,74E-01 | 3,51E-01 |
| RPS25 | 6318,04 | 0,08 | 0,06 | 1,36 | 1,74E-01 | 3,51E-01 |
| SUCLG2 | 1547,32 | 0,03 | 0,02 | 1,36 | 1,74E-01 | 3,51E-01 |
| SPX | 87,47 | 0,10 | 0,07 | 1,36 | 1,74E-01 | 3,51E-01 |
| SF3A2 | 334,88 | 0,05 | 0,03 | 1,36 | 1,74E-01 | 3,51E-01 |
| SIGLEC10 | 2004,08 | -0,06 | 0,04 | -1,36 | 1,74E-01 | 3,51E-01 |
| SLC7A11 | 137,40 | -0,07 | 0,05 | -1,36 | 1,75E-01 | 3,51E-01 |
| GLUD1 | 3225,82 | 0,03 | 0,02 | 1,36 | 1,75E-01 | 3,52E-01 |
| MTFR1L | 729,22 | 0,02 | 0,02 | 1,36 | 1,75E-01 | 3,52E-01 |
| AGTRAP | 870,48 | -0,05 | 0,04 | -1,36 | 1,75E-01 | 3,52E-01 |
| VOPP1 | 2220,46 | -0,04 | 0,03 | -1,36 | 1,75E-01 | 3,52E-01 |
| FCRL3 | 1531,64 | 0,07 | 0,05 | 1,36 | 1,75E-01 | 3,52E-01 |
| CDK10 | 507,96 | 0,05 | 0,04 | 1,36 | 1,75E-01 | 3,52E-01 |
| RMND5B | 510,49 | 0,03 | 0,02 | 1,36 | 1,75E-01 | 3,52E-01 |
| POLR2J3 | 416,85 | 0,07 | 0,05 | 1,36 | 1,75E-01 | 3,52E-01 |
| STK17B | 20606,77 | -0,05 | 0,03 | -1,36 | 1,75E-01 | 3,52E-01 |
| ENSA | 1450,69 | -0,02 | 0,02 | -1,36 | 1,75E-01 | 3,52E-01 |
| SLC38A1 | 10511,13 | 0,04 | 0,03 | 1,36 | 1,75E-01 | 3,52E-01 |
| STAB1 | 1406,87 | 0,08 | 0,06 | 1,36 | 1,75E-01 | 3,52E-01 |
| COQ2 | 553,01 | -0,03 | 0,02 | -1,35 | 1,76E-01 | 3,52E-01 |
| BCDIN3D | 329,73 | 0,04 | 0,03 | 1,35 | 1,76E-01 | 3,53E-01 |
| SLC20A1 | 3207,87 | -0,03 | 0,03 | -1,35 | 1,76E-01 | 3,53E-01 |
| GAN | 979,63 | 0,02 | 0,02 | 1,35 | 1,76E-01 | 3,53E-01 |
| TSPAN31 | 515,40 | 0,02 | 0,02 | 1,35 | 1,76E-01 | 3,53E-01 |
| ZNF792 | 757,33 | 0,03 | 0,02 | 1,35 | 1,76E-01 | 3,53E-01 |
| CMC1 | 992,26 | 0,06 | 0,04 | 1,35 | 1,76E-01 | 3,54E-01 |
| IL6ST | 6202,20 | 0,05 | 0,04 | 1,35 | 1,76E-01 | 3,54E-01 |
| ZCCHC18 | 113,39 | 0,06 | 0,05 | 1,35 | 1,76E-01 | 3,54E-01 |
| GIMAP2 | 2315,56 | 0,07 | 0,05 | 1,35 | 1,76E-01 | 3,54E-01 |
| C9orf78 | 7130,61 | 0,09 | 0,07 | 1,35 | 1,77E-01 | 3,54E-01 |
| MFSD12 | 462,55 | 0,06 | 0,05 | 1,35 | 1,77E-01 | 3,54E-01 |
| EBNA1BP2 | 372,46 | 0,04 | 0,03 | 1,35 | 1,77E-01 | 3,54E-01 |
| CNST | 2415,03 | 0,03 | 0,02 | 1,35 | 1,77E-01 | 3,54E-01 |
| B3GALT2 | 73,33 | 0,10 | 0,08 | 1,35 | 1,77E-01 | 3,54E-01 |
| SNX14 | 2446,02 | 0,04 | 0,03 | 1,35 | 1,77E-01 | 3,54E-01 |

| | | | | | | |
|---|---|---|---|---|---|---|
| ZNF891 | 1004,11 | 0,03 | 0,02 | 1,35 | 1,77E-01 | 3,54E-01 |
| KIT | 120,47 | -0,07 | 0,05 | -1,35 | 1,77E-01 | 3,55E-01 |
| NT5C3B | 179,76 | 0,06 | 0,04 | 1,35 | 1,77E-01 | 3,55E-01 |
| CCDC71 | 441,50 | 0,03 | 0,02 | 1,35 | 1,77E-01 | 3,55E-01 |
| BUD31 | 821,09 | -0,03 | 0,02 | -1,35 | 1,77E-01 | 3,55E-01 |
| FDXR | 111,93 | 0,05 | 0,04 | 1,35 | 1,78E-01 | 3,55E-01 |
| NOC2L | 390,04 | 0,05 | 0,04 | 1,35 | 1,78E-01 | 3,55E-01 |
| CAMLG | 605,43 | 0,04 | 0,03 | 1,35 | 1,78E-01 | 3,55E-01 |
| RXRB | 960,87 | -0,04 | 0,03 | -1,35 | 1,78E-01 | 3,56E-01 |
| SERTAD1 | 387,35 | -0,03 | 0,03 | -1,35 | 1,78E-01 | 3,56E-01 |
| SMN1 | 110,54 | 0,04 | 0,03 | 1,35 | 1,78E-01 | 3,56E-01 |
| CAPRIN2 | 1258,21 | 0,03 | 0,02 | 1,35 | 1,78E-01 | 3,56E-01 |
| EIF3L | 2584,01 | 0,03 | 0,02 | 1,35 | 1,78E-01 | 3,56E-01 |
| EZR | 3618,23 | 0,04 | 0,03 | 1,35 | 1,78E-01 | 3,56E-01 |
| TNFAIP8 | 5309,81 | 0,03 | 0,02 | 1,35 | 1,78E-01 | 3,56E-01 |
| SMPD2 | 222,74 | 0,04 | 0,03 | 1,35 | 1,78E-01 | 3,56E-01 |
| DYRK4 | 302,77 | 0,04 | 0,03 | 1,35 | 1,78E-01 | 3,56E-01 |
| ZNF561 | 778,27 | 0,03 | 0,02 | 1,35 | 1,78E-01 | 3,56E-01 |
| MSH6 | 1358,22 | -0,03 | 0,02 | -1,35 | 1,79E-01 | 3,56E-01 |
| GTPBP8 | 473,79 | 0,04 | 0,03 | 1,35 | 1,79E-01 | 3,56E-01 |
| PDE6B | 153,18 | 0,05 | 0,04 | 1,34 | 1,79E-01 | 3,56E-01 |
| TMT1A | 2558,17 | 0,04 | 0,03 | 1,34 | 1,79E-01 | 3,56E-01 |
| TARS1 | 1098,94 | 0,03 | 0,03 | 1,34 | 1,79E-01 | 3,57E-01 |
| SERPINI1 | 173,20 | 0,06 | 0,04 | 1,34 | 1,79E-01 | 3,57E-01 |
| FRYL | 8073,44 | -0,03 | 0,02 | -1,34 | 1,79E-01 | 3,57E-01 |
| RIDA | 84,82 | 0,06 | 0,04 | 1,34 | 1,79E-01 | 3,57E-01 |
| ZNF69 | 164,03 | 0,04 | 0,03 | 1,34 | 1,79E-01 | 3,57E-01 |
| ZNF518A | 2912,55 | 0,05 | 0,04 | 1,34 | 1,79E-01 | 3,57E-01 |
| ADHFE1 | 146,13 | 0,04 | 0,03 | 1,34 | 1,79E-01 | 3,57E-01 |
| GIT1 | 343,96 | 0,06 | 0,04 | 1,34 | 1,79E-01 | 3,57E-01 |
| ITGAX | 6245,92 | -0,10 | 0,07 | -1,34 | 1,79E-01 | 3,57E-01 |
| RTN1 | 547,90 | 0,05 | 0,04 | 1,34 | 1,79E-01 | 3,57E-01 |
| HEATR6 | 547,46 | 0,03 | 0,02 | 1,34 | 1,79E-01 | 3,57E-01 |
| SMURF2 | 1916,89 | -0,02 | 0,02 | -1,34 | 1,80E-01 | 3,57E-01 |
| BTF3 | 3366,79 | 0,05 | 0,04 | 1,34 | 1,80E-01 | 3,57E-01 |
| HACE1 | 750,75 | 0,03 | 0,02 | 1,34 | 1,80E-01 | 3,57E-01 |
| TEX30 | 154,25 | 0,05 | 0,04 | 1,34 | 1,80E-01 | 3,57E-01 |
| PKD1 | 626,75 | 0,09 | 0,07 | 1,34 | 1,80E-01 | 3,57E-01 |
| RAB33B | 1127,38 | -0,05 | 0,04 | -1,34 | 1,80E-01 | 3,57E-01 |
| PIGF | 690,44 | 0,04 | 0,03 | 1,34 | 1,80E-01 | 3,58E-01 |
| AMDHD2 | 752,71 | 0,04 | 0,03 | 1,34 | 1,80E-01 | 3,58E-01 |
| TRIB2 | 1980,20 | 0,04 | 0,03 | 1,34 | 1,80E-01 | 3,58E-01 |
| ZNF776 | 1293,41 | -0,04 | 0,03 | -1,34 | 1,80E-01 | 3,58E-01 |
| ZNF197 | 736,10 | 0,04 | 0,03 | 1,34 | 1,81E-01 | 3,59E-01 |
| OGFRL1 | 18904,17 | -0,06 | 0,04 | -1,34 | 1,81E-01 | 3,59E-01 |
| CNOT6 | 1796,05 | 0,03 | 0,02 | 1,34 | 1,81E-01 | 3,59E-01 |

| | | | | | | |
|---|---|---|---|---|---|---|
| LARS2 | 834,92 | 0,02 | 0,02 | 1,34 | 1,81E-01 | 3,59E-01 |
| KLF11 | 482,10 | 0,03 | 0,03 | 1,34 | 1,81E-01 | 3,59E-01 |
| E2F6 | 246,20 | 0,03 | 0,02 | 1,34 | 1,81E-01 | 3,59E-01 |
| CEP57 | 1503,28 | 0,05 | 0,04 | 1,34 | 1,81E-01 | 3,59E-01 |
| TMEM248 | 2497,21 | -0,02 | 0,01 | -1,34 | 1,81E-01 | 3,59E-01 |
| CACNA2D4 | 230,09 | 0,05 | 0,04 | 1,34 | 1,81E-01 | 3,60E-01 |
| PANX2 | 123,26 | -0,11 | 0,08 | -1,34 | 1,81E-01 | 3,60E-01 |
| EFNB1 | 124,10 | 0,04 | 0,03 | 1,34 | 1,81E-01 | 3,60E-01 |
| LONP1 | 874,49 | 0,03 | 0,02 | 1,34 | 1,82E-01 | 3,60E-01 |
| PITPNM2 | 277,40 | 0,08 | 0,06 | 1,34 | 1,81E-01 | 3,60E-01 |
| ARHGAP27 | 3248,86 | -0,06 | 0,05 | -1,34 | 1,81E-01 | 3,60E-01 |
| COTL1 | 5010,75 | -0,06 | 0,04 | -1,34 | 1,82E-01 | 3,60E-01 |
| EFCAB13 | 362,27 | 0,07 | 0,05 | 1,33 | 1,82E-01 | 3,60E-01 |
| ZNF823 | 104,64 | 0,05 | 0,04 | 1,34 | 1,82E-01 | 3,60E-01 |
| TTC8 | 106,79 | 0,06 | 0,04 | 1,33 | 1,82E-01 | 3,61E-01 |
| USP6NL | 1135,37 | 0,04 | 0,03 | 1,33 | 1,82E-01 | 3,61E-01 |
| JAGN1 | 347,02 | 0,02 | 0,02 | 1,33 | 1,83E-01 | 3,61E-01 |
| CASK | 1438,46 | 0,03 | 0,02 | 1,33 | 1,83E-01 | 3,61E-01 |
| MCCC1 | 815,00 | 0,02 | 0,02 | 1,33 | 1,83E-01 | 3,61E-01 |
| PRKG2 | 71,75 | 0,11 | 0,08 | 1,33 | 1,83E-01 | 3,61E-01 |
| KLRC4 | 222,44 | -0,09 | 0,07 | -1,33 | 1,83E-01 | 3,61E-01 |
| KLHL26 | 103,86 | 0,07 | 0,05 | 1,33 | 1,83E-01 | 3,62E-01 |
| PLEC | 9441,25 | 0,06 | 0,04 | 1,33 | 1,83E-01 | 3,62E-01 |
| ITPR3 | 939,01 | 0,07 | 0,05 | 1,33 | 1,83E-01 | 3,63E-01 |
| CSNK1G2 | 2570,07 | -0,04 | 0,03 | -1,33 | 1,84E-01 | 3,63E-01 |
| NFATC2IP | 1517,72 | 0,03 | 0,02 | 1,33 | 1,84E-01 | 3,63E-01 |
| ING3 | 1001,82 | -0,04 | 0,03 | -1,33 | 1,84E-01 | 3,63E-01 |
| RABL6 | 503,51 | 0,05 | 0,04 | 1,33 | 1,84E-01 | 3,63E-01 |
| SOCS5 | 754,08 | 0,04 | 0,03 | 1,33 | 1,84E-01 | 3,63E-01 |
| GCNT1 | 833,92 | 0,04 | 0,03 | 1,33 | 1,84E-01 | 3,63E-01 |
| TMBIM1 | 3734,02 | -0,03 | 0,02 | -1,33 | 1,84E-01 | 3,63E-01 |
| NT5C3A | 1908,49 | -0,05 | 0,04 | -1,33 | 1,84E-01 | 3,63E-01 |
| MAT2A | 2359,38 | 0,02 | 0,02 | 1,33 | 1,84E-01 | 3,63E-01 |
| UBE2S | 344,67 | 0,04 | 0,03 | 1,33 | 1,84E-01 | 3,63E-01 |
| SAYSD1 | 422,36 | 0,03 | 0,02 | 1,33 | 1,84E-01 | 3,63E-01 |
| CENPK | 464,01 | 0,10 | 0,08 | 1,33 | 1,84E-01 | 3,63E-01 |
| AMZ2 | 853,80 | 0,02 | 0,02 | 1,33 | 1,84E-01 | 3,63E-01 |
| TRMO | 1556,72 | -0,03 | 0,02 | -1,33 | 1,84E-01 | 3,64E-01 |
| SLC16A10 | 311,31 | 0,06 | 0,05 | 1,33 | 1,84E-01 | 3,64E-01 |
| XCL2 | 116,83 | 0,10 | 0,07 | 1,33 | 1,85E-01 | 3,64E-01 |
| TRNT1 | 1170,26 | 0,03 | 0,03 | 1,33 | 1,85E-01 | 3,64E-01 |
| GRAP2 | 2699,09 | 0,03 | 0,03 | 1,33 | 1,85E-01 | 3,64E-01 |
| ZC3HAV1L | 80,17 | 0,06 | 0,05 | 1,33 | 1,85E-01 | 3,64E-01 |
| PBX4 | 101,66 | 0,06 | 0,04 | 1,33 | 1,85E-01 | 3,64E-01 |
| RGS3 | 1457,05 | -0,04 | 0,03 | -1,33 | 1,85E-01 | 3,64E-01 |
| PDLIM7 | 574,42 | -0,09 | 0,07 | -1,33 | 1,85E-01 | 3,64E-01 |

| | | | | | | |
|---|---|---|---|---|---|---|
| ARAP3 | 598,39 | -0,10 | 0,07 | -1,33 | 1,85E-01 | 3,65E-01 |
| CSTF1 | 714,83 | -0,02 | 0,01 | -1,33 | 1,85E-01 | 3,65E-01 |
| LMAN1 | 1781,38 | 0,04 | 0,03 | 1,32 | 1,85E-01 | 3,65E-01 |
| MED12 | 833,38 | -0,08 | 0,06 | -1,32 | 1,86E-01 | 3,65E-01 |
| SAMD4B | 737,93 | -0,07 | 0,05 | -1,32 | 1,86E-01 | 3,65E-01 |
| MREG | 206,71 | 0,03 | 0,02 | 1,32 | 1,86E-01 | 3,65E-01 |
| EZH2 | 322,76 | 0,04 | 0,03 | 1,32 | 1,86E-01 | 3,65E-01 |
| ADAM9 | 979,99 | -0,04 | 0,03 | -1,32 | 1,86E-01 | 3,65E-01 |
| ZNF608 | 165,90 | -0,08 | 0,06 | -1,32 | 1,86E-01 | 3,65E-01 |
| EPHX2 | 434,50 | 0,06 | 0,05 | 1,32 | 1,86E-01 | 3,66E-01 |
| BRMS1L | 108,45 | 0,05 | 0,04 | 1,32 | 1,86E-01 | 3,66E-01 |
| NMRAL1 | 601,84 | 0,03 | 0,02 | 1,32 | 1,86E-01 | 3,66E-01 |
| ZSCAN32 | 777,23 | -0,02 | 0,01 | -1,32 | 1,86E-01 | 3,66E-01 |
| SLC41A2 | 91,46 | 0,06 | 0,04 | 1,32 | 1,87E-01 | 3,67E-01 |
| MMP8 | 147,86 | 0,15 | 0,12 | 1,32 | 1,87E-01 | 3,67E-01 |
| RNPEP | 859,48 | -0,06 | 0,04 | -1,32 | 1,87E-01 | 3,67E-01 |
| TBPL1 | 951,16 | 0,04 | 0,03 | 1,32 | 1,87E-01 | 3,67E-01 |
| PHETA1 | 92,79 | 0,07 | 0,05 | 1,32 | 1,87E-01 | 3,67E-01 |
| RPAP2 | 1313,73 | 0,04 | 0,03 | 1,32 | 1,87E-01 | 3,67E-01 |
| DENND2D | 3514,40 | 0,03 | 0,02 | 1,32 | 1,87E-01 | 3,67E-01 |
| GINM1 | 694,68 | -0,03 | 0,02 | -1,32 | 1,87E-01 | 3,67E-01 |
| MINDY2 | 1787,86 | 0,04 | 0,03 | 1,32 | 1,87E-01 | 3,67E-01 |
| ZNF600 | 728,12 | 0,06 | 0,04 | 1,32 | 1,87E-01 | 3,67E-01 |
| TMEM258 | 671,64 | 0,04 | 0,03 | 1,32 | 1,87E-01 | 3,67E-01 |
| SLC25A22 | 157,33 | 0,05 | 0,04 | 1,32 | 1,87E-01 | 3,67E-01 |
| PDCD10 | 1890,63 | 0,06 | 0,04 | 1,32 | 1,87E-01 | 3,67E-01 |
| CYP4V2 | 1239,83 | 0,04 | 0,03 | 1,32 | 1,87E-01 | 3,67E-01 |
| BCL11B | 4268,83 | 0,04 | 0,03 | 1,32 | 1,88E-01 | 3,68E-01 |
| AK3 | 700,04 | 0,03 | 0,03 | 1,32 | 1,88E-01 | 3,68E-01 |
| UBASH3A | 1003,46 | 0,05 | 0,03 | 1,32 | 1,88E-01 | 3,68E-01 |
| FBXO33 | 1989,48 | -0,03 | 0,03 | -1,32 | 1,88E-01 | 3,68E-01 |
| PARP11 | 1182,88 | -0,03 | 0,02 | -1,32 | 1,88E-01 | 3,68E-01 |
| ATF5 | 80,17 | 0,07 | 0,05 | 1,32 | 1,88E-01 | 3,68E-01 |
| MDM4 | 11286,46 | -0,03 | 0,02 | -1,32 | 1,88E-01 | 3,68E-01 |
| MYO5C | 87,07 | -0,11 | 0,09 | -1,32 | 1,88E-01 | 3,68E-01 |
| RBMXL1 | 1066,82 | -0,04 | 0,03 | -1,32 | 1,88E-01 | 3,68E-01 |
| SMIM12 | 1314,11 | 0,01 | 0,01 | 1,32 | 1,88E-01 | 3,68E-01 |
| FOXN2 | 3913,35 | 0,05 | 0,03 | 1,32 | 1,88E-01 | 3,68E-01 |
| RIPK3 | 228,97 | -0,05 | 0,04 | -1,32 | 1,88E-01 | 3,69E-01 |
| MPC1 | 649,56 | 0,03 | 0,02 | 1,32 | 1,89E-01 | 3,69E-01 |
| ZBTB25 | 2479,60 | 0,03 | 0,02 | 1,31 | 1,89E-01 | 3,69E-01 |
| MCAT | 141,74 | 0,03 | 0,02 | 1,31 | 1,89E-01 | 3,69E-01 |
| LY9 | 909,25 | 0,05 | 0,03 | 1,31 | 1,89E-01 | 3,69E-01 |
| TTC21B | 1510,27 | 0,03 | 0,03 | 1,31 | 1,89E-01 | 3,69E-01 |
| SH3BP5 | 3064,53 | -0,03 | 0,02 | -1,31 | 1,89E-01 | 3,69E-01 |
| SIGMAR1 | 326,84 | 0,03 | 0,02 | 1,31 | 1,89E-01 | 3,69E-01 |

| | | | | | | |
|---|---|---|---|---|---|---|
| LMBR1L | 1436,44 | -0,03 | 0,02 | -1,31 | 1,89E-01 | 3,69E-01 |
| GPANK1 | 676,30 | -0,02 | 0,02 | -1,31 | 1,89E-01 | 3,69E-01 |
| SLC35C1 | 163,90 | 0,06 | 0,05 | 1,31 | 1,89E-01 | 3,70E-01 |
| KIAA0513 | 3108,48 | -0,06 | 0,05 | -1,31 | 1,89E-01 | 3,70E-01 |
| NAPEPLD | 555,66 | 0,03 | 0,02 | 1,31 | 1,90E-01 | 3,70E-01 |
| SLC25A45 | 394,34 | 0,07 | 0,05 | 1,31 | 1,90E-01 | 3,70E-01 |
| WDR90 | 110,26 | 0,10 | 0,08 | 1,31 | 1,90E-01 | 3,70E-01 |
| RTTN | 1810,43 | 0,03 | 0,02 | 1,31 | 1,90E-01 | 3,70E-01 |
| DIABLO | 325,13 | 0,02 | 0,02 | 1,31 | 1,90E-01 | 3,70E-01 |
| ZGRF1 | 471,61 | 0,04 | 0,03 | 1,31 | 1,90E-01 | 3,71E-01 |
| MMADHC | 1771,00 | -0,04 | 0,03 | -1,31 | 1,90E-01 | 3,71E-01 |
| ACRBP | 241,65 | 0,06 | 0,05 | 1,31 | 1,90E-01 | 3,71E-01 |
| IREB2 | 2920,45 | -0,02 | 0,02 | -1,31 | 1,90E-01 | 3,71E-01 |
| TMED3 | 893,74 | 0,03 | 0,02 | 1,31 | 1,90E-01 | 3,71E-01 |
| TAF4 | 420,30 | -0,06 | 0,05 | -1,31 | 1,90E-01 | 3,71E-01 |
| SERPINH1 | 190,22 | 0,04 | 0,03 | 1,31 | 1,91E-01 | 3,71E-01 |
| FLNB | 1319,23 | 0,05 | 0,04 | 1,31 | 1,91E-01 | 3,72E-01 |
| TRIM4 | 1399,46 | -0,02 | 0,01 | -1,31 | 1,91E-01 | 3,72E-01 |
| HSPA14 | 357,29 | 0,05 | 0,04 | 1,31 | 1,91E-01 | 3,72E-01 |
| TCF25 | 2501,54 | -0,03 | 0,02 | -1,31 | 1,91E-01 | 3,72E-01 |
| IL27RA | 1053,22 | 0,03 | 0,02 | 1,31 | 1,91E-01 | 3,72E-01 |
| MRTFB | 626,48 | 0,04 | 0,03 | 1,31 | 1,91E-01 | 3,72E-01 |
| MALT1 | 3797,50 | 0,03 | 0,03 | 1,31 | 1,91E-01 | 3,72E-01 |
| GATC | 626,80 | 0,02 | 0,02 | 1,31 | 1,92E-01 | 3,73E-01 |
| TRIM10 | 540,59 | 0,09 | 0,07 | 1,31 | 1,92E-01 | 3,73E-01 |
| STK17A | 4367,37 | -0,03 | 0,03 | -1,31 | 1,92E-01 | 3,73E-01 |
| USP28 | 1359,15 | -0,04 | 0,03 | -1,31 | 1,92E-01 | 3,73E-01 |
| RHOF | 560,49 | 0,05 | 0,04 | 1,30 | 1,92E-01 | 3,73E-01 |
| RAB18 | 3551,75 | -0,04 | 0,03 | -1,30 | 1,92E-01 | 3,73E-01 |
| PPP1R2 | 3416,69 | 0,04 | 0,03 | 1,30 | 1,92E-01 | 3,73E-01 |
| PEX16 | 894,17 | 0,02 | 0,02 | 1,30 | 1,92E-01 | 3,74E-01 |
| CCNYL1 | 475,70 | 0,03 | 0,02 | 1,30 | 1,92E-01 | 3,74E-01 |
| ENDOG | 118,08 | 0,04 | 0,03 | 1,30 | 1,92E-01 | 3,74E-01 |
| SLC35B3 | 1049,75 | 0,04 | 0,03 | 1,30 | 1,92E-01 | 3,74E-01 |
| DDX47 | 180,64 | 0,03 | 0,02 | 1,30 | 1,93E-01 | 3,74E-01 |
| SLC39A9 | 1836,89 | -0,02 | 0,01 | -1,30 | 1,93E-01 | 3,74E-01 |
| NAA15 | 1882,89 | 0,04 | 0,03 | 1,30 | 1,93E-01 | 3,74E-01 |
| EXOC3 | 1248,16 | -0,05 | 0,04 | -1,30 | 1,93E-01 | 3,74E-01 |
| ASCC3 | 3255,26 | 0,05 | 0,04 | 1,30 | 1,93E-01 | 3,74E-01 |
| SMIM19 | 213,37 | 0,04 | 0,03 | 1,30 | 1,93E-01 | 3,74E-01 |
| MEIS1 | 215,39 | 0,07 | 0,06 | 1,30 | 1,93E-01 | 3,74E-01 |
| PPAN | 85,77 | 0,04 | 0,03 | 1,30 | 1,93E-01 | 3,74E-01 |
| MMD | 1218,98 | 0,06 | 0,04 | 1,30 | 1,93E-01 | 3,74E-01 |
| NAA35 | 874,90 | 0,03 | 0,02 | 1,30 | 1,93E-01 | 3,74E-01 |
| FAM91A1 | 5078,97 | -0,04 | 0,03 | -1,30 | 1,93E-01 | 3,75E-01 |
| RASSF1 | 3088,72 | 0,03 | 0,02 | 1,30 | 1,93E-01 | 3,75E-01 |

| | | | | | | |
|---|---|---|---|---|---|---|
| CD79A | 180,83 | 0,09 | 0,07 | 1,30 | 1,93E-01 | 3,75E-01 |
| SH3YL1 | 1618,88 | 0,04 | 0,03 | 1,30 | 1,93E-01 | 3,75E-01 |
| ZNF410 | 406,86 | -0,02 | 0,01 | -1,30 | 1,94E-01 | 3,75E-01 |
| INTS10 | 1663,20 | 0,02 | 0,01 | 1,30 | 1,94E-01 | 3,75E-01 |
| TAF6 | 177,59 | 0,06 | 0,05 | 1,30 | 1,94E-01 | 3,75E-01 |
| ADCY4 | 116,94 | -0,09 | 0,07 | -1,30 | 1,94E-01 | 3,75E-01 |
| ATP10A | 1074,71 | 0,04 | 0,03 | 1,30 | 1,94E-01 | 3,75E-01 |
| ZNF570 | 400,92 | 0,04 | 0,03 | 1,30 | 1,94E-01 | 3,75E-01 |
| TTC39B | 2048,24 | 0,04 | 0,03 | 1,30 | 1,94E-01 | 3,75E-01 |
| VPS53 | 1821,85 | -0,04 | 0,03 | -1,30 | 1,94E-01 | 3,75E-01 |
| PRDX6 | 5616,44 | 0,08 | 0,06 | 1,30 | 1,94E-01 | 3,75E-01 |
| TAS2R20 | 188,26 | 0,06 | 0,05 | 1,30 | 1,94E-01 | 3,75E-01 |
| PPCDC | 973,99 | -0,06 | 0,05 | -1,30 | 1,94E-01 | 3,75E-01 |
| BRK1 | 1829,47 | 0,04 | 0,03 | 1,30 | 1,94E-01 | 3,75E-01 |
| SEMA3C | 475,64 | -0,05 | 0,04 | -1,30 | 1,94E-01 | 3,75E-01 |
| PCF11 | 7045,02 | -0,03 | 0,02 | -1,30 | 1,94E-01 | 3,75E-01 |
| RCCD1 | 183,34 | 0,06 | 0,04 | 1,30 | 1,94E-01 | 3,75E-01 |
| RECK | 342,52 | 0,02 | 0,02 | 1,30 | 1,94E-01 | 3,76E-01 |
| CMTM7 | 962,43 | -0,03 | 0,03 | -1,30 | 1,95E-01 | 3,76E-01 |
| LENG1 | 517,25 | -0,02 | 0,02 | -1,30 | 1,95E-01 | 3,76E-01 |
| ANXA4 | 821,35 | 0,03 | 0,02 | 1,30 | 1,95E-01 | 3,76E-01 |
| TUT1 | 94,08 | 0,05 | 0,04 | 1,30 | 1,95E-01 | 3,76E-01 |
| BNC2 | 215,95 | -0,09 | 0,07 | -1,30 | 1,95E-01 | 3,76E-01 |
| RPF2 | 226,06 | 0,04 | 0,03 | 1,30 | 1,95E-01 | 3,77E-01 |
| LAP3 | 2086,08 | -0,08 | 0,06 | -1,29 | 1,95E-01 | 3,77E-01 |
| PCBP1 | 2176,74 | 0,07 | 0,05 | 1,29 | 1,95E-01 | 3,77E-01 |
| IKBKG | 317,36 | -0,05 | 0,03 | -1,29 | 1,95E-01 | 3,77E-01 |
| RNF5 | 706,42 | 0,04 | 0,03 | 1,29 | 1,95E-01 | 3,77E-01 |
| DOK2 | 1370,67 | 0,03 | 0,03 | 1,29 | 1,96E-01 | 3,77E-01 |
| ANO8 | 195,00 | 0,06 | 0,05 | 1,29 | 1,96E-01 | 3,77E-01 |
| PIGA | 595,73 | 0,04 | 0,03 | 1,29 | 1,96E-01 | 3,77E-01 |
| MTMR2 | 463,35 | 0,03 | 0,02 | 1,29 | 1,96E-01 | 3,77E-01 |
| MTA1 | 375,65 | 0,07 | 0,05 | 1,29 | 1,96E-01 | 3,77E-01 |
| SURF6 | 934,06 | 0,02 | 0,02 | 1,29 | 1,96E-01 | 3,78E-01 |
| C6orf136 | 120,31 | 0,04 | 0,03 | 1,29 | 1,96E-01 | 3,78E-01 |
| HSP90B1 | 2277,22 | -0,03 | 0,02 | -1,29 | 1,96E-01 | 3,78E-01 |
| SMC2 | 498,09 | 0,05 | 0,04 | 1,29 | 1,97E-01 | 3,79E-01 |
| TESC | 2021,44 | 0,08 | 0,06 | 1,29 | 1,97E-01 | 3,79E-01 |
| AGAP2 | 983,16 | -0,06 | 0,04 | -1,29 | 1,97E-01 | 3,79E-01 |
| INSIG1 | 1278,29 | 0,03 | 0,02 | 1,29 | 1,97E-01 | 3,79E-01 |
| BTBD3 | 197,98 | 0,04 | 0,03 | 1,29 | 1,97E-01 | 3,79E-01 |
| GVQW3 | 222,53 | 0,04 | 0,03 | 1,29 | 1,97E-01 | 3,79E-01 |
| METTL6 | 471,27 | 0,02 | 0,02 | 1,29 | 1,97E-01 | 3,80E-01 |
| ZIK1 | 267,96 | 0,04 | 0,03 | 1,29 | 1,97E-01 | 3,80E-01 |
| GPC2 | 107,75 | 0,05 | 0,04 | 1,29 | 1,98E-01 | 3,80E-01 |
| TMEM242 | 442,84 | 0,02 | 0,02 | 1,29 | 1,98E-01 | 3,80E-01 |

| | | | | | | |
|---|---|---|---|---|---|---|
| FTCDNL1 | 107,83 | 0,04 | 0,03 | 1,29 | 1,98E-01 | 3,81E-01 |
| FAM124B | 84,35 | 0,06 | 0,04 | 1,29 | 1,98E-01 | 3,81E-01 |
| GIMAP8 | 2051,29 | -0,03 | 0,03 | -1,29 | 1,98E-01 | 3,81E-01 |
| HSD17B10 | 496,42 | 0,03 | 0,03 | 1,29 | 1,98E-01 | 3,81E-01 |
| ZNF343 | 269,23 | 0,03 | 0,02 | 1,29 | 1,98E-01 | 3,81E-01 |
| PRELID2 | 88,77 | 0,06 | 0,05 | 1,29 | 1,98E-01 | 3,81E-01 |
| EDRF1 | 1912,08 | -0,02 | 0,01 | -1,29 | 1,98E-01 | 3,81E-01 |
| ORAI3 | 667,04 | -0,03 | 0,02 | -1,29 | 1,99E-01 | 3,81E-01 |
| EHBP1L1 | 8064,28 | -0,05 | 0,04 | -1,29 | 1,99E-01 | 3,81E-01 |
| VIRMA | 2098,95 | -0,02 | 0,02 | -1,29 | 1,99E-01 | 3,81E-01 |
| SORBS3 | 164,12 | 0,07 | 0,05 | 1,28 | 1,99E-01 | 3,81E-01 |
| CIMAP1B | 545,55 | -0,07 | 0,06 | -1,28 | 1,99E-01 | 3,82E-01 |
| ORAI2 | 3469,11 | -0,05 | 0,04 | -1,28 | 1,99E-01 | 3,82E-01 |
| GIN1 | 355,69 | 0,05 | 0,04 | 1,28 | 1,99E-01 | 3,82E-01 |
| PIM1 | 12614,50 | 0,07 | 0,05 | 1,28 | 1,99E-01 | 3,82E-01 |
| SLC36A4 | 1252,93 | 0,05 | 0,04 | 1,28 | 1,99E-01 | 3,82E-01 |
| TTLL5 | 671,56 | -0,04 | 0,03 | -1,28 | 2,00E-01 | 3,82E-01 |
| CCDC7 | 895,11 | 0,05 | 0,04 | 1,28 | 2,00E-01 | 3,83E-01 |
| GGT1 | 243,69 | -0,05 | 0,04 | -1,28 | 2,00E-01 | 3,83E-01 |
| TNPO2 | 464,93 | 0,05 | 0,04 | 1,28 | 2,00E-01 | 3,83E-01 |
| PPP1CC | 3882,27 | 0,03 | 0,03 | 1,28 | 2,00E-01 | 3,83E-01 |
| EIF4ENIF1 | 696,22 | -0,02 | 0,02 | -1,28 | 2,00E-01 | 3,83E-01 |
| FAM219B | 1093,44 | 0,02 | 0,01 | 1,28 | 2,00E-01 | 3,83E-01 |
| ALS2CL | 326,62 | 0,07 | 0,05 | 1,28 | 2,00E-01 | 3,83E-01 |
| CFAP410 | 318,31 | 0,04 | 0,03 | 1,28 | 2,00E-01 | 3,83E-01 |
| ENSG000002 | 134,66 | 0,09 | 0,07 | 1,28 | 2,00E-01 | 3,83E-01 |
| YBEY | 226,80 | 0,04 | 0,03 | 1,28 | 2,00E-01 | 3,84E-01 |
| PRMT7 | 823,26 | 0,03 | 0,02 | 1,28 | 2,01E-01 | 3,84E-01 |
| METTL22 | 994,70 | -0,04 | 0,03 | -1,28 | 2,01E-01 | 3,84E-01 |
| PPME1 | 394,68 | -0,02 | 0,02 | -1,28 | 2,01E-01 | 3,84E-01 |
| KDELR2 | 794,44 | 0,02 | 0,01 | 1,28 | 2,01E-01 | 3,84E-01 |
| COQ4 | 245,61 | 0,05 | 0,04 | 1,28 | 2,01E-01 | 3,84E-01 |
| SLK | 4373,78 | -0,05 | 0,04 | -1,28 | 2,01E-01 | 3,85E-01 |
| ACLY | 2237,69 | -0,03 | 0,02 | -1,28 | 2,01E-01 | 3,85E-01 |
| FLT3 | 224,00 | -0,07 | 0,05 | -1,28 | 2,01E-01 | 3,85E-01 |
| TAF2 | 1888,58 | 0,03 | 0,02 | 1,28 | 2,01E-01 | 3,85E-01 |
| TTC21A | 193,48 | -0,08 | 0,06 | -1,28 | 2,01E-01 | 3,85E-01 |
| CMKLR1 | 548,97 | -0,06 | 0,05 | -1,28 | 2,01E-01 | 3,85E-01 |
| AP5M1 | 2662,13 | 0,05 | 0,04 | 1,28 | 2,01E-01 | 3,85E-01 |
| L3MBTL2 | 473,40 | 0,03 | 0,02 | 1,28 | 2,01E-01 | 3,85E-01 |
| WDCP | 320,42 | 0,03 | 0,02 | 1,28 | 2,01E-01 | 3,85E-01 |
| TAS2R46 | 133,76 | 0,06 | 0,05 | 1,28 | 2,01E-01 | 3,85E-01 |
| ZNF587B | 1017,79 | -0,02 | 0,02 | -1,28 | 2,02E-01 | 3,85E-01 |
| MTHFSD | 595,86 | -0,02 | 0,02 | -1,28 | 2,02E-01 | 3,85E-01 |
| POLR2E | 504,66 | 0,04 | 0,03 | 1,28 | 2,02E-01 | 3,85E-01 |
| ARL8A | 410,99 | -0,05 | 0,04 | -1,28 | 2,02E-01 | 3,85E-01 |

| | | | | | | |
|---|---|---|---|---|---|---|
| LRRK1 | 1005,33 | -0,06 | 0,04 | -1,28 | 2,02E-01 | 3,85E-01 |
| DNAJB5 | 85,63 | -0,08 | 0,07 | -1,28 | 2,02E-01 | 3,85E-01 |
| UBAP2 | 940,28 | 0,04 | 0,03 | 1,28 | 2,02E-01 | 3,85E-01 |
| ZNF696 | 276,30 | 0,03 | 0,03 | 1,28 | 2,02E-01 | 3,85E-01 |
| CPEB3 | 412,55 | -0,03 | 0,02 | -1,28 | 2,02E-01 | 3,85E-01 |
| PKIA | 295,53 | 0,06 | 0,05 | 1,28 | 2,02E-01 | 3,85E-01 |
| BIVM | 187,17 | 0,04 | 0,03 | 1,28 | 2,02E-01 | 3,85E-01 |
| IL10RA | 8090,45 | 0,03 | 0,02 | 1,28 | 2,02E-01 | 3,85E-01 |
| ZBTB1 | 2146,36 | 0,05 | 0,04 | 1,27 | 2,02E-01 | 3,85E-01 |
| TPRN | 92,82 | 0,06 | 0,05 | 1,27 | 2,02E-01 | 3,85E-01 |
| DCLRE1B | 146,57 | 0,03 | 0,02 | 1,27 | 2,02E-01 | 3,85E-01 |
| RAB12 | 693,39 | 0,03 | 0,02 | 1,27 | 2,02E-01 | 3,85E-01 |
| POGLUT1 | 1054,62 | 0,02 | 0,02 | 1,27 | 2,03E-01 | 3,85E-01 |
| ZNF568 | 467,76 | 0,04 | 0,03 | 1,27 | 2,03E-01 | 3,86E-01 |
| CRACDL | 81,16 | -0,09 | 0,07 | -1,27 | 2,03E-01 | 3,86E-01 |
| TP53INP2 | 164,39 | -0,08 | 0,07 | -1,27 | 2,03E-01 | 3,86E-01 |
| LPAR1 | 309,18 | 0,07 | 0,05 | 1,27 | 2,03E-01 | 3,86E-01 |
| ACAA1 | 1750,24 | -0,03 | 0,03 | -1,27 | 2,03E-01 | 3,86E-01 |
| TOMM70 | 1284,74 | 0,04 | 0,03 | 1,27 | 2,03E-01 | 3,86E-01 |
| CENPT | 165,33 | 0,06 | 0,05 | 1,27 | 2,03E-01 | 3,86E-01 |
| ZNF440 | 679,79 | -0,03 | 0,02 | -1,27 | 2,03E-01 | 3,86E-01 |
| TRMT12 | 556,78 | 0,04 | 0,03 | 1,27 | 2,03E-01 | 3,86E-01 |
| RSRC1 | 1009,15 | 0,04 | 0,03 | 1,27 | 2,03E-01 | 3,86E-01 |
| FIGNL1 | 408,47 | 0,05 | 0,04 | 1,27 | 2,03E-01 | 3,86E-01 |
| GIPR | 92,26 | 0,07 | 0,06 | 1,27 | 2,03E-01 | 3,86E-01 |
| MRPS10 | 719,26 | 0,03 | 0,03 | 1,27 | 2,03E-01 | 3,86E-01 |
| ARFGEF2 | 2293,77 | 0,02 | 0,02 | 1,27 | 2,03E-01 | 3,86E-01 |
| KIF3B | 985,41 | -0,02 | 0,02 | -1,27 | 2,03E-01 | 3,86E-01 |
| FBXL12 | 292,29 | 0,03 | 0,02 | 1,27 | 2,04E-01 | 3,86E-01 |
| SLC25A30 | 645,28 | 0,02 | 0,01 | 1,27 | 2,04E-01 | 3,87E-01 |
| THOC7 | 323,87 | 0,04 | 0,03 | 1,27 | 2,04E-01 | 3,87E-01 |
| NARS2 | 618,45 | 0,03 | 0,02 | 1,27 | 2,04E-01 | 3,88E-01 |
| UCP3 | 103,53 | -0,05 | 0,04 | -1,27 | 2,04E-01 | 3,88E-01 |
| RDH11 | 1110,81 | 0,03 | 0,02 | 1,27 | 2,04E-01 | 3,88E-01 |
| LTF | 526,49 | 0,13 | 0,11 | 1,27 | 2,05E-01 | 3,88E-01 |
| STOML1 | 208,10 | 0,05 | 0,04 | 1,27 | 2,05E-01 | 3,88E-01 |
| SPHK2 | 183,12 | 0,05 | 0,04 | 1,27 | 2,05E-01 | 3,88E-01 |
| CENPP | 144,87 | 0,06 | 0,05 | 1,27 | 2,05E-01 | 3,89E-01 |
| PLEKHF2 | 1674,80 | 0,05 | 0,04 | 1,27 | 2,05E-01 | 3,89E-01 |
| ABHD17B | 674,80 | -0,02 | 0,02 | -1,27 | 2,05E-01 | 3,89E-01 |
| ALAD | 712,96 | 0,03 | 0,02 | 1,27 | 2,05E-01 | 3,89E-01 |
| TTC14 | 3856,25 | 0,05 | 0,04 | 1,27 | 2,06E-01 | 3,90E-01 |
| MMS19 | 1120,49 | 0,03 | 0,02 | 1,27 | 2,06E-01 | 3,90E-01 |
| MRPS14 | 575,22 | 0,03 | 0,03 | 1,27 | 2,06E-01 | 3,90E-01 |
| DIPK1A | 286,74 | 0,04 | 0,03 | 1,26 | 2,06E-01 | 3,90E-01 |
| TSPYL5 | 103,00 | 0,06 | 0,05 | 1,26 | 2,06E-01 | 3,90E-01 |

| | | | | | | |
|---|---|---|---|---|---|---|
| PDAP1 | 772,46 | -0,02 | 0,02 | -1,26 | 2,06E-01 | 3,90E-01 |
| LLPH | 887,32 | -0,03 | 0,02 | -1,26 | 2,06E-01 | 3,90E-01 |
| PDIA3 | 1279,83 | -0,03 | 0,02 | -1,26 | 2,06E-01 | 3,90E-01 |
| XPNPEP1 | 1683,63 | 0,02 | 0,02 | 1,26 | 2,07E-01 | 3,91E-01 |
| HNRNPC | 14275,61 | -0,03 | 0,02 | -1,26 | 2,07E-01 | 3,91E-01 |
| RASGRP3 | 425,36 | 0,06 | 0,05 | 1,26 | 2,07E-01 | 3,91E-01 |
| CLYBL | 225,27 | 0,04 | 0,03 | 1,26 | 2,07E-01 | 3,91E-01 |
| CTSB | 10007,95 | 0,06 | 0,04 | 1,26 | 2,07E-01 | 3,91E-01 |
| HNRNPH1 | 17122,38 | 0,03 | 0,02 | 1,26 | 2,07E-01 | 3,92E-01 |
| MKRN3 | 86,24 | 0,10 | 0,08 | 1,26 | 2,07E-01 | 3,92E-01 |
| FOXO4 | 3634,84 | 0,08 | 0,06 | 1,26 | 2,08E-01 | 3,92E-01 |
| CHD1L | 1032,70 | 0,02 | 0,02 | 1,26 | 2,08E-01 | 3,92E-01 |
| SSBP1 | 1354,64 | 0,03 | 0,03 | 1,26 | 2,08E-01 | 3,93E-01 |
| GCDH | 462,95 | 0,03 | 0,02 | 1,26 | 2,08E-01 | 3,93E-01 |
| MAP4K1 | 436,15 | 0,06 | 0,05 | 1,26 | 2,08E-01 | 3,93E-01 |
| LIN54 | 1264,80 | -0,02 | 0,02 | -1,26 | 2,08E-01 | 3,93E-01 |
| GLA | 487,15 | -0,03 | 0,03 | -1,26 | 2,09E-01 | 3,93E-01 |
| KDSR | 515,76 | 0,03 | 0,02 | 1,26 | 2,09E-01 | 3,94E-01 |
| SCFD2 | 770,59 | 0,03 | 0,02 | 1,26 | 2,09E-01 | 3,94E-01 |
| CEACAM8 | 110,66 | 0,12 | 0,10 | 1,26 | 2,09E-01 | 3,94E-01 |
| TRMT10C | 350,29 | 0,07 | 0,05 | 1,26 | 2,09E-01 | 3,94E-01 |
| SRP72 | 1518,82 | 0,04 | 0,03 | 1,26 | 2,09E-01 | 3,94E-01 |
| ST3GAL6 | 1450,92 | -0,05 | 0,04 | -1,26 | 2,09E-01 | 3,94E-01 |
| MFGE8 | 173,52 | 0,07 | 0,05 | 1,26 | 2,09E-01 | 3,94E-01 |
| PDXK | 1839,27 | -0,04 | 0,04 | -1,26 | 2,09E-01 | 3,94E-01 |
| KHNYN | 5182,53 | -0,02 | 0,02 | -1,26 | 2,09E-01 | 3,94E-01 |
| DRAXIN | 135,19 | -0,07 | 0,05 | -1,26 | 2,09E-01 | 3,94E-01 |
| WDR76 | 319,88 | 0,04 | 0,03 | 1,26 | 2,09E-01 | 3,94E-01 |
| RAD51D | 632,24 | 0,03 | 0,02 | 1,26 | 2,09E-01 | 3,94E-01 |
| HORMAD1 | 257,36 | -0,07 | 0,05 | -1,26 | 2,09E-01 | 3,94E-01 |
| TIGD2 | 162,48 | 0,06 | 0,05 | 1,25 | 2,10E-01 | 3,95E-01 |
| NOPCHAP1 | 905,82 | 0,04 | 0,03 | 1,25 | 2,10E-01 | 3,95E-01 |
| EXPH5 | 146,24 | 0,06 | 0,05 | 1,25 | 2,10E-01 | 3,95E-01 |
| SGK3 | 1156,50 | 0,02 | 0,02 | 1,25 | 2,10E-01 | 3,95E-01 |
| MFSD2A | 111,55 | -0,05 | 0,04 | -1,25 | 2,10E-01 | 3,96E-01 |
| POMK | 526,60 | -0,02 | 0,02 | -1,25 | 2,10E-01 | 3,96E-01 |
| SERPINF1 | 74,65 | 0,07 | 0,05 | 1,25 | 2,11E-01 | 3,96E-01 |
| CRISP3 | 98,61 | 0,13 | 0,10 | 1,25 | 2,11E-01 | 3,96E-01 |
| CZIB | 656,60 | 0,02 | 0,01 | 1,25 | 2,11E-01 | 3,96E-01 |
| GAK | 2252,96 | -0,06 | 0,05 | -1,25 | 2,11E-01 | 3,97E-01 |
| VRK2 | 974,84 | 0,05 | 0,04 | 1,25 | 2,11E-01 | 3,97E-01 |
| ZBTB47 | 338,60 | -0,04 | 0,03 | -1,25 | 2,11E-01 | 3,97E-01 |
| PID1 | 351,76 | 0,06 | 0,05 | 1,25 | 2,11E-01 | 3,97E-01 |
| ZNG1B | 1425,66 | 0,04 | 0,03 | 1,25 | 2,12E-01 | 3,97E-01 |
| ATP5IF1 | 368,69 | 0,04 | 0,03 | 1,25 | 2,12E-01 | 3,97E-01 |
| RNF38 | 1920,90 | -0,03 | 0,02 | -1,25 | 2,12E-01 | 3,98E-01 |

| | | | | | | |
|---|---|---|---|---|---|---|
| RHOB | 2913,35 | -0,05 | 0,04 | -1,25 | 2,12E-01 | 3,98E-01 |
| PRXL2B | 156,05 | 0,04 | 0,03 | 1,25 | 2,12E-01 | 3,98E-01 |
| PLEKHA1 | 2958,58 | 0,04 | 0,03 | 1,25 | 2,12E-01 | 3,98E-01 |
| TRPS1 | 2548,52 | 0,03 | 0,02 | 1,25 | 2,12E-01 | 3,98E-01 |
| CPA3 | 140,89 | -0,13 | 0,10 | -1,25 | 2,12E-01 | 3,98E-01 |
| NAGPA | 383,36 | 0,03 | 0,03 | 1,25 | 2,12E-01 | 3,98E-01 |
| BCKDK | 525,31 | -0,04 | 0,03 | -1,25 | 2,13E-01 | 3,99E-01 |
| ZAP70 | 3858,49 | 0,04 | 0,03 | 1,25 | 2,13E-01 | 3,99E-01 |
| ANAPC2 | 657,08 | 0,05 | 0,04 | 1,25 | 2,13E-01 | 3,99E-01 |
| ARL17B | 107,62 | 0,09 | 0,07 | 1,25 | 2,13E-01 | 3,99E-01 |
| FANCE | 149,76 | 0,03 | 0,02 | 1,24 | 2,13E-01 | 3,99E-01 |
| ACTG1 | 21186,86 | -0,03 | 0,02 | -1,24 | 2,13E-01 | 3,99E-01 |
| CYB561A3 | 1058,12 | 0,04 | 0,03 | 1,24 | 2,13E-01 | 3,99E-01 |
| SGMS1 | 1904,69 | -0,02 | 0,02 | -1,24 | 2,13E-01 | 3,99E-01 |
| IPMK | 3259,65 | -0,06 | 0,05 | -1,24 | 2,13E-01 | 3,99E-01 |
| MAN1A2 | 3187,25 | 0,04 | 0,03 | 1,24 | 2,13E-01 | 3,99E-01 |
| USP20 | 1284,75 | 0,03 | 0,02 | 1,24 | 2,13E-01 | 3,99E-01 |
| C21orf58 | 82,56 | 0,06 | 0,05 | 1,24 | 2,13E-01 | 4,00E-01 |
| CCDC25 | 484,97 | 0,03 | 0,03 | 1,24 | 2,14E-01 | 4,00E-01 |
| DGAT1 | 853,83 | -0,04 | 0,03 | -1,24 | 2,14E-01 | 4,00E-01 |
| POLR2B | 4394,42 | -0,04 | 0,03 | -1,24 | 2,14E-01 | 4,00E-01 |
| IARS1 | 2067,04 | 0,02 | 0,02 | 1,24 | 2,14E-01 | 4,00E-01 |
| MAP3K11 | 2818,81 | -0,05 | 0,04 | -1,24 | 2,15E-01 | 4,01E-01 |
| TUBB4B | 1450,51 | 0,03 | 0,03 | 1,24 | 2,15E-01 | 4,01E-01 |
| SNX21 | 142,95 | 0,05 | 0,04 | 1,24 | 2,15E-01 | 4,01E-01 |
| DESI1 | 671,22 | 0,03 | 0,03 | 1,24 | 2,15E-01 | 4,01E-01 |
| ZNF582 | 129,75 | 0,04 | 0,04 | 1,24 | 2,15E-01 | 4,01E-01 |
| PBXIP1 | 4904,94 | -0,04 | 0,03 | -1,24 | 2,15E-01 | 4,01E-01 |
| PRPF6 | 1801,91 | -0,03 | 0,02 | -1,24 | 2,15E-01 | 4,01E-01 |
| FNBP4 | 3890,09 | 0,02 | 0,02 | 1,24 | 2,15E-01 | 4,01E-01 |
| NTMT1 | 219,06 | 0,03 | 0,02 | 1,24 | 2,15E-01 | 4,01E-01 |
| ACKR3 | 131,77 | 0,04 | 0,03 | 1,24 | 2,15E-01 | 4,02E-01 |
| NUP155 | 1259,23 | 0,02 | 0,01 | 1,24 | 2,15E-01 | 4,02E-01 |
| NDUFA2 | 746,77 | 0,04 | 0,03 | 1,24 | 2,15E-01 | 4,02E-01 |
| BTAF1 | 5811,68 | 0,03 | 0,02 | 1,24 | 2,15E-01 | 4,02E-01 |
| ZC3H14 | 1348,92 | 0,02 | 0,02 | 1,24 | 2,16E-01 | 4,02E-01 |
| AGK | 1199,99 | 0,03 | 0,02 | 1,24 | 2,16E-01 | 4,02E-01 |
| FAM13B | 2817,19 | -0,02 | 0,02 | -1,24 | 2,16E-01 | 4,02E-01 |
| TNFRSF12A | 231,02 | -0,05 | 0,04 | -1,24 | 2,16E-01 | 4,02E-01 |
| ZNF740 | 2366,55 | -0,02 | 0,01 | -1,24 | 2,16E-01 | 4,02E-01 |
| ARHGEF19 | 97,55 | 0,06 | 0,05 | 1,24 | 2,16E-01 | 4,02E-01 |
| ABRACL | 672,78 | 0,05 | 0,04 | 1,24 | 2,16E-01 | 4,02E-01 |
| PRKCE | 822,26 | -0,05 | 0,04 | -1,24 | 2,16E-01 | 4,02E-01 |
| SART3 | 2293,04 | -0,02 | 0,02 | -1,24 | 2,16E-01 | 4,02E-01 |
| KATNAL1 | 499,51 | 0,04 | 0,03 | 1,24 | 2,16E-01 | 4,03E-01 |
| S1PR5 | 1118,99 | -0,06 | 0,05 | -1,24 | 2,17E-01 | 4,03E-01 |

| | | | | | | |
|---|---|---|---|---|---|---|
| TAOK2 | 1062,57 | -0,04 | 0,03 | -1,24 | 2,17E-01 | 4,03E-01 |
| RSF1 | 3511,19 | -0,03 | 0,02 | -1,24 | 2,17E-01 | 4,03E-01 |
| ACAA2 | 623,44 | -0,03 | 0,03 | -1,23 | 2,17E-01 | 4,04E-01 |
| CD22 | 642,95 | 0,07 | 0,06 | 1,23 | 2,17E-01 | 4,04E-01 |
| ANKUB1 | 125,29 | -0,07 | 0,05 | -1,23 | 2,17E-01 | 4,04E-01 |
| ACOT13 | 295,31 | -0,03 | 0,02 | -1,23 | 2,17E-01 | 4,04E-01 |
| GPBP1 | 4711,41 | -0,05 | 0,04 | -1,23 | 2,17E-01 | 4,04E-01 |
| CTU2 | 92,22 | 0,05 | 0,04 | 1,23 | 2,17E-01 | 4,04E-01 |
| PLAC8 | 3910,99 | 0,03 | 0,02 | 1,23 | 2,17E-01 | 4,04E-01 |
| ADD2 | 281,15 | 0,07 | 0,05 | 1,23 | 2,17E-01 | 4,04E-01 |
| EIF4EBP2 | 4067,97 | -0,04 | 0,03 | -1,23 | 2,17E-01 | 4,04E-01 |
| EVL | 7460,95 | 0,04 | 0,03 | 1,23 | 2,17E-01 | 4,04E-01 |
| KCTD7 | 1275,41 | 0,03 | 0,02 | 1,23 | 2,18E-01 | 4,05E-01 |
| PYCARD | 1647,25 | -0,04 | 0,03 | -1,23 | 2,18E-01 | 4,05E-01 |
| ZNF526 | 162,50 | 0,05 | 0,04 | 1,23 | 2,18E-01 | 4,05E-01 |
| NPM1 | 4861,76 | 0,06 | 0,05 | 1,23 | 2,18E-01 | 4,05E-01 |
| EIF4E2 | 2525,97 | -0,02 | 0,02 | -1,23 | 2,18E-01 | 4,05E-01 |
| WBP2NL | 124,68 | 0,04 | 0,03 | 1,23 | 2,18E-01 | 4,05E-01 |
| SH3TC1 | 1490,63 | 0,04 | 0,03 | 1,23 | 2,18E-01 | 4,05E-01 |
| DHPS | 140,23 | 0,04 | 0,03 | 1,23 | 2,18E-01 | 4,05E-01 |
| WDR19 | 488,03 | 0,03 | 0,02 | 1,23 | 2,18E-01 | 4,05E-01 |
| SLC31A1 | 928,93 | -0,04 | 0,03 | -1,23 | 2,18E-01 | 4,05E-01 |
| PPRC1 | 215,68 | 0,06 | 0,05 | 1,23 | 2,18E-01 | 4,05E-01 |
| TASP1 | 539,56 | 0,03 | 0,02 | 1,23 | 2,18E-01 | 4,05E-01 |
| HACD3 | 630,83 | 0,03 | 0,02 | 1,23 | 2,19E-01 | 4,05E-01 |
| SUCLA2 | 707,27 | 0,03 | 0,02 | 1,23 | 2,19E-01 | 4,05E-01 |
| RNF40 | 2382,29 | -0,04 | 0,03 | -1,23 | 2,19E-01 | 4,06E-01 |
| DNAJC21 | 1248,70 | 0,03 | 0,02 | 1,23 | 2,19E-01 | 4,06E-01 |
| KRI1 | 613,43 | 0,03 | 0,03 | 1,23 | 2,19E-01 | 4,06E-01 |
| NIBAN2 | 583,15 | 0,04 | 0,04 | 1,23 | 2,19E-01 | 4,06E-01 |
| DERA | 582,74 | 0,02 | 0,02 | 1,23 | 2,19E-01 | 4,06E-01 |
| TMEM135 | 443,79 | 0,04 | 0,03 | 1,23 | 2,19E-01 | 4,06E-01 |
| DTNBP1 | 455,64 | -0,03 | 0,02 | -1,23 | 2,19E-01 | 4,06E-01 |
| SAMD12 | 431,67 | 0,07 | 0,05 | 1,23 | 2,19E-01 | 4,07E-01 |
| MT-ND6 | 194,67 | 0,14 | 0,11 | 1,23 | 2,20E-01 | 4,07E-01 |
| NIP7 | 630,15 | 0,03 | 0,02 | 1,23 | 2,20E-01 | 4,07E-01 |
| ARMH1 | 110,47 | 0,06 | 0,05 | 1,23 | 2,20E-01 | 4,07E-01 |
| KCNN4 | 283,29 | 0,06 | 0,05 | 1,23 | 2,20E-01 | 4,07E-01 |
| ZSWIM1 | 135,98 | 0,04 | 0,03 | 1,23 | 2,20E-01 | 4,07E-01 |
| PLEKHM1 | 1299,29 | -0,06 | 0,05 | -1,23 | 2,20E-01 | 4,07E-01 |
| EMB | 7692,59 | 0,04 | 0,03 | 1,23 | 2,20E-01 | 4,07E-01 |
| BCAT1 | 465,16 | -0,08 | 0,06 | -1,23 | 2,20E-01 | 4,07E-01 |
| HK3 | 5423,45 | -0,05 | 0,04 | -1,23 | 2,20E-01 | 4,07E-01 |
| RFX3 | 1845,41 | 0,03 | 0,03 | 1,23 | 2,20E-01 | 4,07E-01 |
| SEMA4F | 322,93 | 0,03 | 0,03 | 1,23 | 2,20E-01 | 4,07E-01 |
| ARMC5 | 97,77 | 0,06 | 0,05 | 1,23 | 2,20E-01 | 4,07E-01 |

| | | | | | | |
|---|---|---|---|---|---|---|
| H1-5 | 779,12 | 0,07 | 0,06 | 1,23 | 2,20E-01 | 4,07E-01 |
| NPAT | 2734,63 | 0,03 | 0,02 | 1,23 | 2,20E-01 | 4,07E-01 |
| CCDC18 | 605,02 | 0,05 | 0,04 | 1,22 | 2,21E-01 | 4,08E-01 |
| TMC6 | 3401,59 | 0,05 | 0,04 | 1,22 | 2,21E-01 | 4,08E-01 |
| RCC1 | 266,40 | 0,03 | 0,02 | 1,22 | 2,21E-01 | 4,08E-01 |
| MED26 | 201,20 | -0,04 | 0,03 | -1,22 | 2,21E-01 | 4,08E-01 |
| SOD1 | 1287,16 | 0,05 | 0,04 | 1,22 | 2,21E-01 | 4,08E-01 |
| COMMD7 | 97,95 | 0,03 | 0,03 | 1,22 | 2,21E-01 | 4,08E-01 |
| CHST7 | 586,58 | -0,05 | 0,04 | -1,22 | 2,21E-01 | 4,08E-01 |
| PIGBOS1 | 245,10 | -0,03 | 0,02 | -1,22 | 2,21E-01 | 4,09E-01 |
| LRRC27 | 213,79 | 0,04 | 0,03 | 1,22 | 2,22E-01 | 4,09E-01 |
| CADM1 | 101,53 | -0,10 | 0,08 | -1,22 | 2,21E-01 | 4,09E-01 |
| CNOT7 | 2625,76 | 0,04 | 0,03 | 1,22 | 2,21E-01 | 4,09E-01 |
| SDC4 | 74,44 | 0,07 | 0,06 | 1,22 | 2,22E-01 | 4,09E-01 |
| PTPN11 | 2269,96 | -0,02 | 0,02 | -1,22 | 2,22E-01 | 4,09E-01 |
| RBSN | 1390,84 | -0,02 | 0,01 | -1,22 | 2,22E-01 | 4,09E-01 |
| PSTPIP1 | 4026,17 | -0,04 | 0,03 | -1,22 | 2,22E-01 | 4,09E-01 |
| PIGT | 979,36 | -0,03 | 0,02 | -1,22 | 2,22E-01 | 4,09E-01 |
| SDR42E1 | 351,91 | 0,04 | 0,03 | 1,22 | 2,22E-01 | 4,09E-01 |
| MYO1G | 6152,43 | 0,03 | 0,02 | 1,22 | 2,22E-01 | 4,09E-01 |
| TBL3 | 322,04 | 0,03 | 0,03 | 1,22 | 2,22E-01 | 4,09E-01 |
| BRPF3 | 693,57 | -0,04 | 0,03 | -1,22 | 2,22E-01 | 4,09E-01 |
| PPP1R11 | 343,93 | -0,03 | 0,02 | -1,22 | 2,22E-01 | 4,09E-01 |
| PIGQ | 211,29 | 0,06 | 0,05 | 1,22 | 2,23E-01 | 4,10E-01 |
| TMOD2 | 2675,71 | -0,04 | 0,03 | -1,22 | 2,22E-01 | 4,10E-01 |
| ITFG2 | 1335,58 | 0,02 | 0,02 | 1,22 | 2,22E-01 | 4,10E-01 |
| TPK1 | 1152,19 | -0,03 | 0,02 | -1,22 | 2,23E-01 | 4,10E-01 |
| DCAF10 | 1867,12 | 0,02 | 0,02 | 1,22 | 2,23E-01 | 4,10E-01 |
| ST6GALNAC6 | 276,93 | 0,04 | 0,03 | 1,22 | 2,23E-01 | 4,10E-01 |
| IL4R | 6061,80 | -0,04 | 0,04 | -1,22 | 2,23E-01 | 4,10E-01 |
| CPNE2 | 337,66 | -0,05 | 0,04 | -1,22 | 2,23E-01 | 4,10E-01 |
| TRAF3IP2 | 143,39 | -0,04 | 0,04 | -1,22 | 2,23E-01 | 4,10E-01 |
| NIBAN3 | 1268,71 | 0,07 | 0,06 | 1,22 | 2,23E-01 | 4,10E-01 |
| CORO1A | 12525,46 | -0,04 | 0,03 | -1,22 | 2,24E-01 | 4,11E-01 |
| ROCK2 | 2046,85 | -0,03 | 0,03 | -1,22 | 2,24E-01 | 4,11E-01 |
| ZNF10 | 277,16 | 0,04 | 0,03 | 1,22 | 2,24E-01 | 4,11E-01 |
| CAMKMT | 611,53 | 0,03 | 0,03 | 1,22 | 2,24E-01 | 4,11E-01 |
| KAT2A | 190,07 | 0,06 | 0,05 | 1,22 | 2,24E-01 | 4,11E-01 |
| STXBP5 | 6407,45 | -0,04 | 0,03 | -1,22 | 2,24E-01 | 4,11E-01 |
| SELENOT | 2783,77 | 0,05 | 0,04 | 1,22 | 2,24E-01 | 4,11E-01 |
| RFXAP | 282,43 | 0,03 | 0,02 | 1,22 | 2,24E-01 | 4,11E-01 |
| ITGA6 | 2687,28 | 0,04 | 0,03 | 1,22 | 2,24E-01 | 4,11E-01 |
| HSPA4 | 2099,42 | -0,03 | 0,03 | -1,22 | 2,24E-01 | 4,12E-01 |
| EXT2 | 1097,33 | -0,02 | 0,02 | -1,21 | 2,25E-01 | 4,12E-01 |
| ZNF90 | 135,32 | 0,05 | 0,04 | 1,21 | 2,25E-01 | 4,12E-01 |
| YY1 | 3527,17 | -0,01 | 0,01 | -1,21 | 2,25E-01 | 4,13E-01 |

| | | | | | | |
|---|---|---|---|---|---|---|
| GBP3 | 1170,29 | -0,05 | 0,04 | -1,21 | 2,25E-01 | 4,13E-01 |
| BLCAP | 1686,98 | -0,04 | 0,03 | -1,21 | 2,25E-01 | 4,13E-01 |
| ING4 | 880,36 | -0,02 | 0,02 | -1,21 | 2,25E-01 | 4,13E-01 |
| FRS2 | 1193,71 | 0,03 | 0,02 | 1,21 | 2,25E-01 | 4,13E-01 |
| CCNY | 3727,05 | -0,02 | 0,02 | -1,21 | 2,26E-01 | 4,13E-01 |
| SPIB | 169,14 | 0,06 | 0,05 | 1,21 | 2,26E-01 | 4,13E-01 |
| ZC3H12A | 528,86 | 0,03 | 0,02 | 1,21 | 2,26E-01 | 4,14E-01 |
| ZKSCAN8 | 1759,82 | 0,02 | 0,02 | 1,21 | 2,26E-01 | 4,14E-01 |
| SNX13 | 5608,49 | -0,04 | 0,03 | -1,21 | 2,26E-01 | 4,14E-01 |
| RSPRY1 | 1279,95 | -0,02 | 0,01 | -1,21 | 2,26E-01 | 4,14E-01 |
| CD109 | 94,67 | 0,08 | 0,07 | 1,21 | 2,26E-01 | 4,14E-01 |
| RAB29 | 1865,65 | 0,03 | 0,02 | 1,21 | 2,26E-01 | 4,14E-01 |
| TTC4 | 80,30 | 0,03 | 0,03 | 1,21 | 2,26E-01 | 4,14E-01 |
| ATP6V1G1 | 1861,74 | 0,05 | 0,05 | 1,21 | 2,26E-01 | 4,14E-01 |
| ZBTB4 | 946,09 | 0,04 | 0,04 | 1,21 | 2,26E-01 | 4,14E-01 |
| YIPF3 | 455,23 | -0,05 | 0,04 | -1,21 | 2,26E-01 | 4,14E-01 |
| ALDH1A1 | 771,44 | -0,05 | 0,05 | -1,21 | 2,26E-01 | 4,14E-01 |
| WEE1 | 308,41 | 0,05 | 0,04 | 1,21 | 2,27E-01 | 4,14E-01 |
| ELF4 | 847,90 | -0,07 | 0,06 | -1,21 | 2,27E-01 | 4,14E-01 |
| FAM43A | 445,02 | 0,04 | 0,03 | 1,21 | 2,27E-01 | 4,14E-01 |
| ZNF577 | 468,77 | 0,03 | 0,02 | 1,21 | 2,27E-01 | 4,14E-01 |
| AFG2A | 1058,28 | 0,03 | 0,03 | 1,21 | 2,27E-01 | 4,15E-01 |
| CNBP | 8748,08 | -0,03 | 0,03 | -1,21 | 2,27E-01 | 4,15E-01 |
| AIMP1 | 517,99 | 0,05 | 0,04 | 1,21 | 2,27E-01 | 4,15E-01 |
| SDHB | 1212,04 | -0,02 | 0,02 | -1,21 | 2,27E-01 | 4,15E-01 |
| BLOC1S5 | 489,29 | 0,03 | 0,02 | 1,21 | 2,28E-01 | 4,16E-01 |
| NT5E | 242,94 | 0,07 | 0,06 | 1,21 | 2,28E-01 | 4,16E-01 |
| CFAP119 | 115,49 | -0,04 | 0,03 | -1,21 | 2,28E-01 | 4,16E-01 |
| CNIH4 | 1230,56 | -0,03 | 0,03 | -1,21 | 2,28E-01 | 4,16E-01 |
| ZNF747 | 191,67 | 0,02 | 0,02 | 1,21 | 2,28E-01 | 4,16E-01 |
| GSTO1 | 1287,75 | 0,04 | 0,03 | 1,21 | 2,28E-01 | 4,16E-01 |
| AGPAT4 | 890,75 | -0,04 | 0,03 | -1,20 | 2,28E-01 | 4,16E-01 |
| ACOT11 | 105,69 | 0,10 | 0,08 | 1,20 | 2,28E-01 | 4,17E-01 |
| OTUD7A | 191,12 | 0,06 | 0,05 | 1,20 | 2,29E-01 | 4,17E-01 |
| H2BC6 | 638,29 | 0,05 | 0,04 | 1,20 | 2,29E-01 | 4,17E-01 |
| CFD | 3834,31 | -0,10 | 0,08 | -1,20 | 2,29E-01 | 4,18E-01 |
| NECTIN1 | 135,74 | -0,08 | 0,07 | -1,20 | 2,29E-01 | 4,18E-01 |
| AOPEP | 668,75 | -0,04 | 0,03 | -1,20 | 2,29E-01 | 4,18E-01 |
| GTF2H2 | 340,74 | -0,09 | 0,08 | -1,20 | 2,29E-01 | 4,18E-01 |
| MARF1 | 5748,57 | -0,05 | 0,04 | -1,20 | 2,30E-01 | 4,18E-01 |
| IFT140 | 156,38 | 0,06 | 0,05 | 1,20 | 2,30E-01 | 4,19E-01 |
| ALDH6A1 | 384,86 | 0,03 | 0,02 | 1,20 | 2,30E-01 | 4,19E-01 |
| UVSSA | 2009,80 | -0,05 | 0,04 | -1,20 | 2,30E-01 | 4,19E-01 |
| LGALS8 | 3127,95 | -0,02 | 0,02 | -1,20 | 2,30E-01 | 4,19E-01 |
| POFUT1 | 495,71 | 0,03 | 0,03 | 1,20 | 2,30E-01 | 4,19E-01 |
| DNAI4 | 88,22 | 0,06 | 0,05 | 1,20 | 2,30E-01 | 4,19E-01 |

| | | | | | | |
|---|---|---|---|---|---|---|
| ADI1 | 997,30 | 0,02 | 0,02 | 1,20 | 2,30E-01 | 4,19E-01 |
| ELK1 | 254,26 | 0,03 | 0,02 | 1,20 | 2,31E-01 | 4,20E-01 |
| C1D | 518,82 | -0,05 | 0,04 | -1,20 | 2,31E-01 | 4,20E-01 |
| RASGRP2 | 5554,56 | 0,03 | 0,02 | 1,20 | 2,31E-01 | 4,20E-01 |
| ZNF470 | 208,26 | 0,05 | 0,04 | 1,20 | 2,31E-01 | 4,21E-01 |
| CLEC17A | 195,90 | 0,07 | 0,06 | 1,20 | 2,31E-01 | 4,21E-01 |
| TSC22D2 | 1083,05 | 0,02 | 0,02 | 1,20 | 2,31E-01 | 4,21E-01 |
| WDR45 | 2515,18 | 0,05 | 0,04 | 1,20 | 2,32E-01 | 4,22E-01 |
| TTI1 | 1020,18 | -0,02 | 0,01 | -1,19 | 2,32E-01 | 4,22E-01 |
| YTHDF3 | 4015,33 | -0,03 | 0,02 | -1,19 | 2,32E-01 | 4,22E-01 |
| UTP25 | 857,19 | 0,03 | 0,02 | 1,19 | 2,33E-01 | 4,23E-01 |
| SLC6A9 | 120,64 | 0,10 | 0,09 | 1,19 | 2,33E-01 | 4,23E-01 |
| EIF2S1 | 1427,29 | 0,04 | 0,03 | 1,19 | 2,33E-01 | 4,23E-01 |
| IFT27 | 357,59 | 0,03 | 0,02 | 1,19 | 2,33E-01 | 4,23E-01 |
| RTP4 | 403,59 | -0,07 | 0,06 | -1,19 | 2,33E-01 | 4,23E-01 |
| ANK3 | 2171,59 | 0,05 | 0,04 | 1,19 | 2,33E-01 | 4,23E-01 |
| GATM | 107,82 | 0,07 | 0,06 | 1,19 | 2,33E-01 | 4,23E-01 |
| BTBD1 | 1637,36 | 0,03 | 0,03 | 1,19 | 2,33E-01 | 4,23E-01 |
| PELP1 | 220,15 | 0,05 | 0,04 | 1,19 | 2,33E-01 | 4,23E-01 |
| PNMA3 | 213,31 | 0,08 | 0,07 | 1,19 | 2,33E-01 | 4,23E-01 |
| ARSA | 223,72 | -0,09 | 0,08 | -1,19 | 2,33E-01 | 4,23E-01 |
| ZNF395 | 301,55 | 0,05 | 0,04 | 1,19 | 2,33E-01 | 4,23E-01 |
| ATXN2 | 1205,28 | -0,04 | 0,03 | -1,19 | 2,33E-01 | 4,23E-01 |
| PLCB2 | 3762,60 | -0,06 | 0,05 | -1,19 | 2,33E-01 | 4,23E-01 |
| ZNF234 | 659,45 | 0,02 | 0,02 | 1,19 | 2,34E-01 | 4,24E-01 |
| GTF2H2C | 419,33 | -0,03 | 0,03 | -1,19 | 2,34E-01 | 4,24E-01 |
| UBIAD1 | 510,86 | 0,02 | 0,02 | 1,19 | 2,34E-01 | 4,24E-01 |
| PKIG | 179,41 | 0,06 | 0,05 | 1,19 | 2,34E-01 | 4,24E-01 |
| TRA2B | 6727,50 | 0,03 | 0,02 | 1,19 | 2,34E-01 | 4,25E-01 |
| PDCD4 | 7101,75 | -0,03 | 0,03 | -1,19 | 2,34E-01 | 4,25E-01 |
| GGCX | 660,53 | -0,03 | 0,02 | -1,19 | 2,35E-01 | 4,25E-01 |
| CNPY3 | 1431,88 | -0,04 | 0,03 | -1,19 | 2,35E-01 | 4,25E-01 |
| RFX2 | 372,51 | -0,07 | 0,06 | -1,19 | 2,35E-01 | 4,25E-01 |
| HEATR1 | 2168,23 | 0,03 | 0,02 | 1,19 | 2,35E-01 | 4,25E-01 |
| DLG1 | 2188,51 | 0,02 | 0,02 | 1,19 | 2,35E-01 | 4,25E-01 |
| LSM14A | 4096,97 | -0,02 | 0,02 | -1,19 | 2,35E-01 | 4,25E-01 |
| PNPLA2 | 840,87 | 0,07 | 0,06 | 1,19 | 2,35E-01 | 4,25E-01 |
| VPS28 | 1421,81 | 0,04 | 0,03 | 1,19 | 2,35E-01 | 4,25E-01 |
| GLTP | 1020,31 | -0,02 | 0,02 | -1,19 | 2,35E-01 | 4,25E-01 |
| IQCE | 735,30 | -0,04 | 0,04 | -1,19 | 2,35E-01 | 4,25E-01 |
| GARRE1 | 1059,10 | -0,03 | 0,03 | -1,19 | 2,35E-01 | 4,25E-01 |
| ADTRP | 414,32 | 0,08 | 0,07 | 1,19 | 2,35E-01 | 4,25E-01 |
| ZNF331 | 520,03 | 0,03 | 0,02 | 1,19 | 2,35E-01 | 4,26E-01 |
| MAP2K7 | 939,98 | 0,05 | 0,04 | 1,19 | 2,35E-01 | 4,26E-01 |
| TMEM41B | 1158,47 | 0,03 | 0,02 | 1,19 | 2,36E-01 | 4,26E-01 |
| CLIP4 | 2188,92 | 0,03 | 0,02 | 1,19 | 2,36E-01 | 4,26E-01 |

| | | | | | | |
|---|---|---|---|---|---|---|
| SLFN12L | 4188,50 | 0,04 | 0,03 | 1,19 | 2,36E-01 | 4,26E-01 |
| RB1CC1 | 5106,29 | -0,04 | 0,04 | -1,19 | 2,36E-01 | 4,26E-01 |
| PSMD5 | 917,69 | 0,02 | 0,02 | 1,18 | 2,36E-01 | 4,26E-01 |
| CSNK2B | 201,83 | -0,04 | 0,03 | -1,18 | 2,36E-01 | 4,26E-01 |
| ERCC6 | 2297,42 | -0,02 | 0,02 | -1,18 | 2,36E-01 | 4,26E-01 |
| RAB27B | 1464,68 | 0,06 | 0,05 | 1,18 | 2,36E-01 | 4,26E-01 |
| SMAD4 | 4590,82 | -0,02 | 0,02 | -1,18 | 2,36E-01 | 4,26E-01 |
| GCN1 | 2068,32 | 0,03 | 0,03 | 1,18 | 2,36E-01 | 4,27E-01 |
| TAMALIN | 101,78 | 0,06 | 0,05 | 1,18 | 2,37E-01 | 4,27E-01 |
| ECI2 | 560,69 | 0,02 | 0,02 | 1,18 | 2,37E-01 | 4,27E-01 |
| GRN | 1724,87 | -0,06 | 0,05 | -1,18 | 2,37E-01 | 4,27E-01 |
| RNF207 | 76,17 | 0,10 | 0,08 | 1,18 | 2,37E-01 | 4,27E-01 |
| NUCKS1 | 4173,87 | 0,03 | 0,03 | 1,18 | 2,37E-01 | 4,27E-01 |
| NFU1 | 247,97 | 0,03 | 0,02 | 1,18 | 2,37E-01 | 4,27E-01 |
| CHMP1B | 3860,06 | -0,04 | 0,03 | -1,18 | 2,37E-01 | 4,27E-01 |
| KDM2B | 801,12 | 0,04 | 0,03 | 1,18 | 2,37E-01 | 4,27E-01 |
| TFB1M | 614,58 | 0,03 | 0,02 | 1,18 | 2,37E-01 | 4,27E-01 |
| OSBPL5 | 736,71 | -0,06 | 0,05 | -1,18 | 2,37E-01 | 4,28E-01 |
| ORC4 | 1712,04 | 0,04 | 0,03 | 1,18 | 2,37E-01 | 4,28E-01 |
| RALGAPA1 | 2962,38 | 0,03 | 0,02 | 1,18 | 2,37E-01 | 4,28E-01 |
| HYI | 142,09 | 0,04 | 0,04 | 1,18 | 2,38E-01 | 4,28E-01 |
| MRPS18C | 726,07 | 0,05 | 0,04 | 1,18 | 2,38E-01 | 4,28E-01 |
| SLC37A4 | 186,15 | 0,03 | 0,03 | 1,18 | 2,38E-01 | 4,28E-01 |
| STX5 | 768,72 | -0,02 | 0,02 | -1,18 | 2,38E-01 | 4,28E-01 |
| PPP2R5E | 1388,56 | 0,03 | 0,02 | 1,18 | 2,38E-01 | 4,28E-01 |
| SH3BGRL | 4160,59 | 0,03 | 0,03 | 1,18 | 2,38E-01 | 4,28E-01 |
| CREB3 | 273,07 | 0,02 | 0,02 | 1,18 | 2,38E-01 | 4,28E-01 |
| GLT8D1 | 644,60 | 0,02 | 0,01 | 1,18 | 2,38E-01 | 4,28E-01 |
| PSMG4 | 1254,09 | 0,03 | 0,02 | 1,18 | 2,38E-01 | 4,28E-01 |
| SLC35A5 | 1445,56 | -0,03 | 0,02 | -1,18 | 2,39E-01 | 4,29E-01 |
| SP2 | 189,01 | -0,05 | 0,04 | -1,18 | 2,39E-01 | 4,29E-01 |
| KIF13B | 1596,24 | -0,03 | 0,03 | -1,18 | 2,39E-01 | 4,29E-01 |
| ZMIZ1 | 1504,50 | -0,06 | 0,05 | -1,18 | 2,39E-01 | 4,29E-01 |
| REXO2 | 730,15 | 0,04 | 0,03 | 1,18 | 2,39E-01 | 4,29E-01 |
| LIPE | 80,17 | 0,06 | 0,05 | 1,18 | 2,39E-01 | 4,29E-01 |
| ATP7B | 126,56 | -0,07 | 0,06 | -1,18 | 2,39E-01 | 4,29E-01 |
| RNF2 | 769,04 | 0,03 | 0,03 | 1,18 | 2,39E-01 | 4,29E-01 |
| CCNT1 | 2435,05 | -0,02 | 0,02 | -1,18 | 2,39E-01 | 4,29E-01 |
| WIPI2 | 2094,33 | -0,03 | 0,03 | -1,18 | 2,39E-01 | 4,29E-01 |
| MPP7 | 1915,45 | -0,03 | 0,03 | -1,18 | 2,39E-01 | 4,29E-01 |
| ICE1 | 2014,13 | 0,02 | 0,02 | 1,18 | 2,39E-01 | 4,29E-01 |
| IGLL5 | 88,07 | 0,10 | 0,08 | 1,18 | 2,39E-01 | 4,29E-01 |
| UPB1 | 94,93 | 0,07 | 0,06 | 1,18 | 2,39E-01 | 4,29E-01 |
| ZNF598 | 252,64 | 0,05 | 0,04 | 1,18 | 2,39E-01 | 4,29E-01 |
| C2orf88 | 833,19 | 0,06 | 0,05 | 1,18 | 2,39E-01 | 4,29E-01 |
| UTP18 | 949,13 | 0,03 | 0,02 | 1,18 | 2,39E-01 | 4,29E-01 |

| | | | | | | |
|---|---|---|---|---|---|---|
| ZNF420 | 373,65 | 0,04 | 0,04 | 1,18 | 2,40E-01 | 4,30E-01 |
| NEK8 | 79,95 | 0,06 | 0,05 | 1,18 | 2,40E-01 | 4,30E-01 |
| HDGFL2 | 608,99 | -0,03 | 0,03 | -1,17 | 2,40E-01 | 4,30E-01 |
| CCL4L2 | 98,09 | 0,09 | 0,07 | 1,17 | 2,40E-01 | 4,30E-01 |
| GPS2 | 133,00 | 0,04 | 0,03 | 1,17 | 2,40E-01 | 4,30E-01 |
| FUNDC1 | 142,18 | 0,04 | 0,03 | 1,17 | 2,40E-01 | 4,30E-01 |
| DARS1 | 1349,24 | 0,04 | 0,03 | 1,17 | 2,40E-01 | 4,30E-01 |
| ALG13 | 2382,85 | 0,03 | 0,03 | 1,17 | 2,40E-01 | 4,30E-01 |
| ZFC3H1 | 5901,83 | -0,03 | 0,02 | -1,17 | 2,40E-01 | 4,30E-01 |
| CHKA | 190,71 | 0,04 | 0,04 | 1,17 | 2,41E-01 | 4,31E-01 |
| PGP | 444,24 | 0,02 | 0,02 | 1,17 | 2,41E-01 | 4,31E-01 |
| GPR137 | 188,42 | 0,04 | 0,03 | 1,17 | 2,41E-01 | 4,31E-01 |
| MAP3K21 | 107,95 | 0,06 | 0,05 | 1,17 | 2,41E-01 | 4,31E-01 |
| CD40 | 331,71 | 0,04 | 0,04 | 1,17 | 2,41E-01 | 4,31E-01 |
| MRPS11 | 627,32 | 0,01 | 0,01 | 1,17 | 2,41E-01 | 4,31E-01 |
| SNED1 | 340,66 | 0,07 | 0,06 | 1,17 | 2,41E-01 | 4,31E-01 |
| HEBP1 | 193,37 | 0,07 | 0,06 | 1,17 | 2,41E-01 | 4,31E-01 |
| CDH1 | 146,43 | 0,09 | 0,07 | 1,17 | 2,41E-01 | 4,31E-01 |
| LUC7L2 | 1590,10 | 0,03 | 0,02 | 1,17 | 2,41E-01 | 4,31E-01 |
| C2 | 92,70 | 0,04 | 0,04 | 1,17 | 2,41E-01 | 4,31E-01 |
| PTCD2 | 439,10 | 0,03 | 0,02 | 1,17 | 2,41E-01 | 4,31E-01 |
| SPI1 | 1078,86 | -0,07 | 0,06 | -1,17 | 2,41E-01 | 4,31E-01 |
| MAP3K10 | 134,29 | 0,06 | 0,05 | 1,17 | 2,42E-01 | 4,32E-01 |
| WDR91 | 798,16 | -0,03 | 0,03 | -1,17 | 2,42E-01 | 4,32E-01 |
| MRI1 | 634,39 | 0,04 | 0,03 | 1,17 | 2,42E-01 | 4,32E-01 |
| FMNL1 | 3408,90 | -0,09 | 0,08 | -1,17 | 2,42E-01 | 4,32E-01 |
| RNF113A | 448,85 | 0,03 | 0,02 | 1,17 | 2,42E-01 | 4,32E-01 |
| CCL4 | 463,47 | -0,06 | 0,05 | -1,17 | 2,42E-01 | 4,32E-01 |
| HMGN4 | 1596,58 | 0,03 | 0,03 | 1,17 | 2,42E-01 | 4,32E-01 |
| EID3 | 245,82 | -0,03 | 0,02 | -1,17 | 2,42E-01 | 4,32E-01 |
| SPAG7 | 250,17 | 0,03 | 0,03 | 1,17 | 2,42E-01 | 4,32E-01 |
| THG1L | 246,58 | 0,03 | 0,03 | 1,17 | 2,42E-01 | 4,32E-01 |
| ABCA7 | 2268,17 | -0,07 | 0,06 | -1,17 | 2,42E-01 | 4,32E-01 |
| DMXL1 | 4383,45 | 0,03 | 0,03 | 1,17 | 2,42E-01 | 4,32E-01 |
| SMARCD2 | 2300,14 | -0,02 | 0,02 | -1,17 | 2,42E-01 | 4,32E-01 |
| CYB561D1 | 1008,35 | -0,03 | 0,03 | -1,17 | 2,43E-01 | 4,32E-01 |
| MACROD2 | 118,78 | 0,10 | 0,09 | 1,17 | 2,43E-01 | 4,32E-01 |
| TBK1 | 1546,46 | -0,04 | 0,04 | -1,17 | 2,43E-01 | 4,32E-01 |
| LYZ | 79673,25 | 0,06 | 0,05 | 1,17 | 2,43E-01 | 4,32E-01 |
| MMP24OS | 525,60 | -0,04 | 0,04 | -1,17 | 2,43E-01 | 4,32E-01 |
| STIL | 447,02 | 0,07 | 0,06 | 1,17 | 2,43E-01 | 4,32E-01 |
| PSMC4 | 979,01 | -0,02 | 0,02 | -1,17 | 2,43E-01 | 4,33E-01 |
| MED24 | 383,71 | 0,04 | 0,03 | 1,17 | 2,43E-01 | 4,33E-01 |
| AGO2 | 4717,51 | -0,05 | 0,04 | -1,17 | 2,43E-01 | 4,33E-01 |
| DCTD | 582,38 | 0,02 | 0,01 | 1,17 | 2,43E-01 | 4,33E-01 |
| TMIGD2 | 191,62 | 0,06 | 0,05 | 1,17 | 2,43E-01 | 4,33E-01 |

| | | | | | | |
|---|---|---|---|---|---|---|
| TAPBP | 2717,13 | -0,06 | 0,05 | -1,17 | 2,43E-01 | 4,33E-01 |
| ATXN7L3 | 1852,09 | -0,03 | 0,02 | -1,17 | 2,44E-01 | 4,33E-01 |
| GPA33 | 193,66 | 0,05 | 0,04 | 1,17 | 2,44E-01 | 4,33E-01 |
| KIF14 | 123,08 | 0,09 | 0,07 | 1,17 | 2,44E-01 | 4,33E-01 |
| AKAP8L | 1413,47 | -0,03 | 0,02 | -1,17 | 2,44E-01 | 4,34E-01 |
| PHLDB3 | 175,75 | 0,04 | 0,04 | 1,16 | 2,44E-01 | 4,34E-01 |
| JUNB | 4158,97 | -0,04 | 0,04 | -1,16 | 2,44E-01 | 4,34E-01 |
| ING5 | 1131,19 | 0,02 | 0,02 | 1,16 | 2,44E-01 | 4,34E-01 |
| PCK2 | 195,58 | 0,05 | 0,05 | 1,16 | 2,44E-01 | 4,34E-01 |
| RING1 | 446,99 | 0,02 | 0,02 | 1,16 | 2,44E-01 | 4,34E-01 |
| HMGN3 | 477,99 | 0,04 | 0,03 | 1,16 | 2,45E-01 | 4,35E-01 |
| FAM133B | 730,42 | 0,04 | 0,03 | 1,16 | 2,45E-01 | 4,35E-01 |
| CHORDC1 | 2702,69 | 0,04 | 0,03 | 1,16 | 2,45E-01 | 4,35E-01 |
| ACVR1C | 211,24 | 0,06 | 0,05 | 1,16 | 2,45E-01 | 4,35E-01 |
| LCK | 4139,69 | 0,03 | 0,03 | 1,16 | 2,45E-01 | 4,35E-01 |
| MGLL | 189,37 | 0,07 | 0,06 | 1,16 | 2,45E-01 | 4,36E-01 |
| ALMS1 | 2420,58 | 0,03 | 0,02 | 1,16 | 2,45E-01 | 4,36E-01 |
| CENPE | 143,23 | 0,08 | 0,07 | 1,16 | 2,45E-01 | 4,36E-01 |
| TIMM23B | 571,84 | 0,02 | 0,02 | 1,16 | 2,46E-01 | 4,36E-01 |
| PEX13 | 1179,76 | 0,03 | 0,02 | 1,16 | 2,46E-01 | 4,36E-01 |
| THAP5 | 1296,62 | 0,05 | 0,04 | 1,16 | 2,46E-01 | 4,36E-01 |
| VNN1 | 748,58 | -0,08 | 0,07 | -1,16 | 2,46E-01 | 4,36E-01 |
| AP4M1 | 121,56 | 0,04 | 0,03 | 1,16 | 2,46E-01 | 4,36E-01 |
| CROT | 447,72 | 0,03 | 0,03 | 1,16 | 2,46E-01 | 4,37E-01 |
| ZFTA | 143,59 | 0,05 | 0,04 | 1,16 | 2,46E-01 | 4,37E-01 |
| ZNF382 | 267,59 | 0,04 | 0,03 | 1,16 | 2,46E-01 | 4,37E-01 |
| ARL4C | 4256,74 | 0,03 | 0,03 | 1,16 | 2,46E-01 | 4,37E-01 |
| METTL9 | 3883,76 | 0,04 | 0,03 | 1,16 | 2,47E-01 | 4,37E-01 |
| CIAO3 | 288,32 | 0,04 | 0,04 | 1,16 | 2,47E-01 | 4,37E-01 |
| CD320 | 86,45 | 0,06 | 0,05 | 1,16 | 2,47E-01 | 4,37E-01 |
| CD7 | 143,20 | 0,06 | 0,05 | 1,16 | 2,47E-01 | 4,37E-01 |
| NBEAL2 | 5357,93 | -0,08 | 0,07 | -1,16 | 2,47E-01 | 4,37E-01 |
| ISCA2 | 558,93 | 0,03 | 0,03 | 1,16 | 2,47E-01 | 4,37E-01 |
| C1QA | 118,52 | -0,09 | 0,07 | -1,16 | 2,47E-01 | 4,37E-01 |
| WSB1 | 7978,74 | -0,03 | 0,03 | -1,16 | 2,47E-01 | 4,37E-01 |
| KIAA1328 | 538,06 | 0,03 | 0,02 | 1,16 | 2,47E-01 | 4,37E-01 |
| TRIM41 | 1026,05 | -0,02 | 0,02 | -1,16 | 2,47E-01 | 4,37E-01 |
| KLF3 | 6644,71 | -0,02 | 0,02 | -1,16 | 2,47E-01 | 4,37E-01 |
| POLR3K | 195,66 | 0,02 | 0,02 | 1,16 | 2,47E-01 | 4,37E-01 |
| ANKRD17 | 3776,22 | -0,03 | 0,02 | -1,16 | 2,47E-01 | 4,37E-01 |
| BTG2 | 1658,98 | -0,05 | 0,04 | -1,16 | 2,47E-01 | 4,37E-01 |
| UBXN11 | 680,06 | 0,06 | 0,05 | 1,16 | 2,47E-01 | 4,37E-01 |
| LARP1B | 464,15 | 0,04 | 0,03 | 1,16 | 2,48E-01 | 4,38E-01 |
| LGALSL | 602,79 | -0,06 | 0,05 | -1,16 | 2,48E-01 | 4,38E-01 |
| FNBP1L | 284,55 | 0,04 | 0,04 | 1,16 | 2,48E-01 | 4,38E-01 |
| ZNF66 | 636,75 | -0,05 | 0,04 | -1,16 | 2,48E-01 | 4,38E-01 |

| | | | | | | |
|---|---|---|---|---|---|---|
| STRBP | 1104,32 | 0,05 | 0,04 | 1,16 | 2,48E-01 | 4,38E-01 |
| NINL | 120,82 | -0,10 | 0,08 | -1,15 | 2,48E-01 | 4,38E-01 |
| SLC36A1 | 1414,66 | -0,05 | 0,04 | -1,15 | 2,48E-01 | 4,39E-01 |
| LY6E | 452,90 | -0,09 | 0,08 | -1,15 | 2,48E-01 | 4,39E-01 |
| HLA-DQB1 | 2902,31 | 0,10 | 0,09 | 1,15 | 2,49E-01 | 4,39E-01 |
| HNRNPU | 24578,30 | -0,02 | 0,02 | -1,15 | 2,49E-01 | 4,39E-01 |
| SLC20A2 | 763,41 | 0,02 | 0,02 | 1,15 | 2,49E-01 | 4,39E-01 |
| ZNF554 | 176,53 | 0,03 | 0,03 | 1,15 | 2,49E-01 | 4,39E-01 |
| U2SURP | 4431,69 | 0,04 | 0,03 | 1,15 | 2,49E-01 | 4,39E-01 |
| FXYD5 | 641,02 | 0,03 | 0,03 | 1,15 | 2,49E-01 | 4,39E-01 |
| CDK7 | 502,31 | -0,02 | 0,02 | -1,15 | 2,49E-01 | 4,39E-01 |
| SWAP70 | 1634,92 | 0,04 | 0,04 | 1,15 | 2,49E-01 | 4,39E-01 |
| LRWD1 | 386,20 | -0,05 | 0,04 | -1,15 | 2,49E-01 | 4,40E-01 |
| AP2M1 | 2026,56 | 0,05 | 0,04 | 1,15 | 2,49E-01 | 4,40E-01 |
| RETSAT | 459,74 | -0,02 | 0,02 | -1,15 | 2,49E-01 | 4,40E-01 |
| SNUPN | 249,67 | 0,02 | 0,02 | 1,15 | 2,49E-01 | 4,40E-01 |
| BCL2L2 | 269,12 | 0,03 | 0,03 | 1,15 | 2,49E-01 | 4,40E-01 |
| EIF5B | 1050,55 | 0,02 | 0,02 | 1,15 | 2,50E-01 | 4,40E-01 |
| SEC14L4 | 84,81 | 0,09 | 0,08 | 1,15 | 2,50E-01 | 4,40E-01 |
| EPB41 | 46558,12 | 0,07 | 0,06 | 1,15 | 2,50E-01 | 4,40E-01 |
| TEX9 | 103,90 | 0,08 | 0,07 | 1,15 | 2,50E-01 | 4,40E-01 |
| CACNA2D3 | 190,53 | 0,05 | 0,04 | 1,15 | 2,50E-01 | 4,40E-01 |
| FBXO30 | 1677,12 | 0,04 | 0,04 | 1,15 | 2,50E-01 | 4,41E-01 |
| ABHD8 | 131,61 | 0,03 | 0,03 | 1,15 | 2,50E-01 | 4,41E-01 |
| H2AC11 | 1482,14 | 0,06 | 0,05 | 1,15 | 2,51E-01 | 4,41E-01 |
| ICAM2 | 1102,09 | 0,02 | 0,02 | 1,15 | 2,51E-01 | 4,41E-01 |
| TBC1D24 | 109,97 | 0,07 | 0,06 | 1,15 | 2,51E-01 | 4,41E-01 |
| SLC4A2 | 390,62 | 0,05 | 0,04 | 1,15 | 2,51E-01 | 4,42E-01 |
| PSMB5 | 443,68 | 0,02 | 0,02 | 1,15 | 2,51E-01 | 4,42E-01 |
| ATP13A1 | 564,16 | 0,05 | 0,04 | 1,15 | 2,51E-01 | 4,42E-01 |
| EXOC2 | 1976,96 | 0,02 | 0,02 | 1,15 | 2,51E-01 | 4,42E-01 |
| MARCHF9 | 218,25 | 0,04 | 0,04 | 1,15 | 2,52E-01 | 4,43E-01 |
| ATOX1 | 561,39 | -0,03 | 0,02 | -1,15 | 2,52E-01 | 4,43E-01 |
| PACS2 | 434,59 | 0,07 | 0,06 | 1,15 | 2,52E-01 | 4,43E-01 |
| GPLD1 | 87,79 | 0,08 | 0,07 | 1,15 | 2,52E-01 | 4,43E-01 |
| GART | 1210,60 | 0,02 | 0,01 | 1,15 | 2,52E-01 | 4,43E-01 |
| EBF1 | 297,82 | 0,07 | 0,06 | 1,15 | 2,52E-01 | 4,43E-01 |
| DAAM1 | 520,04 | 0,03 | 0,02 | 1,15 | 2,52E-01 | 4,43E-01 |
| THUMPD1 | 2294,55 | 0,04 | 0,03 | 1,14 | 2,52E-01 | 4,43E-01 |
| CASP2 | 1608,83 | -0,03 | 0,03 | -1,14 | 2,52E-01 | 4,43E-01 |
| ZKSCAN1 | 2798,31 | -0,01 | 0,01 | -1,14 | 2,53E-01 | 4,43E-01 |
| EPHA1 | 195,64 | 0,06 | 0,05 | 1,14 | 2,53E-01 | 4,43E-01 |
| PALS1 | 463,56 | 0,03 | 0,03 | 1,14 | 2,53E-01 | 4,43E-01 |
| TNFRSF13C | 172,40 | 0,08 | 0,07 | 1,14 | 2,53E-01 | 4,43E-01 |
| TMEM268 | 435,28 | -0,05 | 0,04 | -1,14 | 2,53E-01 | 4,43E-01 |
| INIP | 967,73 | -0,02 | 0,02 | -1,14 | 2,53E-01 | 4,43E-01 |

| | | | | | | |
|---|---|---|---|---|---|---|
| NDUFB6 | 615,04 | -0,04 | 0,03 | -1,14 | 2,52E-01 | 4,43E-01 |
| POMT1 | 510,55 | 0,03 | 0,02 | 1,14 | 2,53E-01 | 4,43E-01 |
| GLMP | 113,06 | 0,05 | 0,05 | 1,14 | 2,53E-01 | 4,43E-01 |
| PPIH | 361,25 | 0,02 | 0,02 | 1,14 | 2,53E-01 | 4,43E-01 |
| ZFAND5 | 5442,08 | 0,03 | 0,03 | 1,14 | 2,53E-01 | 4,43E-01 |
| SLC25A38 | 818,21 | 0,03 | 0,02 | 1,14 | 2,53E-01 | 4,43E-01 |
| ZKSCAN4 | 408,89 | -0,02 | 0,02 | -1,14 | 2,53E-01 | 4,44E-01 |
| ZDHHC8 | 217,65 | 0,07 | 0,06 | 1,14 | 2,53E-01 | 4,44E-01 |
| TBCD | 1394,45 | 0,04 | 0,03 | 1,14 | 2,53E-01 | 4,44E-01 |
| ARMH3 | 1019,49 | 0,02 | 0,02 | 1,14 | 2,54E-01 | 4,44E-01 |
| NAV1 | 222,99 | 0,07 | 0,06 | 1,14 | 2,54E-01 | 4,44E-01 |
| ABTB3 | 591,43 | -0,04 | 0,03 | -1,14 | 2,54E-01 | 4,45E-01 |
| GPD2 | 1519,52 | -0,03 | 0,03 | -1,14 | 2,54E-01 | 4,45E-01 |
| TTYH3 | 435,36 | 0,08 | 0,07 | 1,14 | 2,55E-01 | 4,45E-01 |
| ATG101 | 558,37 | 0,03 | 0,02 | 1,14 | 2,55E-01 | 4,45E-01 |
| SPECC1 | 4852,89 | 0,06 | 0,05 | 1,14 | 2,54E-01 | 4,45E-01 |
| ZNF888 | 287,15 | 0,04 | 0,04 | 1,14 | 2,55E-01 | 4,45E-01 |
| TMUB1 | 240,12 | 0,03 | 0,02 | 1,14 | 2,55E-01 | 4,45E-01 |
| ZNF12 | 1811,89 | 0,04 | 0,03 | 1,14 | 2,54E-01 | 4,45E-01 |
| GAS7 | 2458,39 | -0,05 | 0,05 | -1,14 | 2,55E-01 | 4,45E-01 |
| PCBP2 | 4658,73 | 0,03 | 0,03 | 1,14 | 2,55E-01 | 4,45E-01 |
| STARD3NL | 816,62 | -0,03 | 0,03 | -1,14 | 2,55E-01 | 4,46E-01 |
| ALKBH4 | 183,55 | 0,02 | 0,02 | 1,14 | 2,55E-01 | 4,46E-01 |
| HMGB1 | 5535,78 | 0,06 | 0,05 | 1,14 | 2,55E-01 | 4,46E-01 |
| C1orf52 | 584,59 | 0,03 | 0,02 | 1,14 | 2,55E-01 | 4,46E-01 |
| SLC40A1 | 3190,03 | -0,04 | 0,04 | -1,14 | 2,55E-01 | 4,46E-01 |
| DSC2 | 1371,46 | -0,07 | 0,06 | -1,14 | 2,55E-01 | 4,46E-01 |
| MYNN | 1088,96 | 0,05 | 0,04 | 1,14 | 2,56E-01 | 4,46E-01 |
| ARFGAP2 | 2469,86 | -0,02 | 0,02 | -1,14 | 2,56E-01 | 4,46E-01 |
| TMED9 | 1325,48 | 0,02 | 0,02 | 1,14 | 2,56E-01 | 4,46E-01 |
| NDUFAF1 | 410,29 | -0,03 | 0,02 | -1,14 | 2,56E-01 | 4,47E-01 |
| UCP2 | 1419,40 | 0,07 | 0,06 | 1,14 | 2,56E-01 | 4,47E-01 |
| CCDC137 | 258,02 | 0,02 | 0,02 | 1,14 | 2,56E-01 | 4,47E-01 |
| GPN3 | 274,75 | 0,04 | 0,04 | 1,13 | 2,56E-01 | 4,47E-01 |
| PHAX | 698,87 | 0,04 | 0,03 | 1,13 | 2,57E-01 | 4,48E-01 |
| P2RY10 | 1116,98 | 0,04 | 0,04 | 1,13 | 2,57E-01 | 4,48E-01 |
| RPF1 | 667,02 | 0,04 | 0,04 | 1,13 | 2,57E-01 | 4,48E-01 |
| MED14 | 1525,73 | -0,02 | 0,02 | -1,13 | 2,57E-01 | 4,48E-01 |
| MRPS22 | 580,10 | 0,03 | 0,03 | 1,13 | 2,57E-01 | 4,48E-01 |
| DNAJB14 | 4835,89 | 0,05 | 0,04 | 1,13 | 2,57E-01 | 4,48E-01 |
| STX7 | 10897,65 | -0,03 | 0,03 | -1,13 | 2,57E-01 | 4,48E-01 |
| ADCY3 | 208,84 | 0,05 | 0,05 | 1,13 | 2,58E-01 | 4,49E-01 |
| UBR1 | 3041,18 | -0,02 | 0,02 | -1,13 | 2,58E-01 | 4,49E-01 |
| ZNF277 | 638,60 | 0,04 | 0,03 | 1,13 | 2,58E-01 | 4,49E-01 |
| C3AR1 | 326,98 | -0,05 | 0,05 | -1,13 | 2,58E-01 | 4,49E-01 |
| GNPTG | 824,99 | 0,02 | 0,02 | 1,13 | 2,58E-01 | 4,49E-01 |

| | | | | | | |
|---|---|---|---|---|---|---|
| NUCB1 | 1075,67 | 0,06 | 0,05 | 1,13 | 2,58E-01 | 4,49E-01 |
| LYRM4 | 337,55 | 0,03 | 0,03 | 1,13 | 2,58E-01 | 4,49E-01 |
| NT5DC1 | 723,21 | 0,03 | 0,02 | 1,13 | 2,58E-01 | 4,49E-01 |
| KLRC4-KLRK1 | 256,48 | -0,08 | 0,07 | -1,13 | 2,58E-01 | 4,49E-01 |
| SMPDL3A | 196,48 | -0,04 | 0,04 | -1,13 | 2,58E-01 | 4,49E-01 |
| HSCB | 153,05 | 0,03 | 0,03 | 1,13 | 2,58E-01 | 4,49E-01 |
| PDE1B | 225,48 | -0,06 | 0,05 | -1,13 | 2,58E-01 | 4,49E-01 |
| XPO7 | 3209,83 | 0,04 | 0,04 | 1,13 | 2,58E-01 | 4,49E-01 |
| COPZ1 | 937,02 | 0,02 | 0,02 | 1,13 | 2,58E-01 | 4,49E-01 |
| ANXA2 | 5420,48 | 0,03 | 0,03 | 1,13 | 2,58E-01 | 4,49E-01 |
| DNMT3A | 1658,47 | -0,04 | 0,03 | -1,13 | 2,59E-01 | 4,50E-01 |
| TES | 3397,78 | -0,03 | 0,02 | -1,13 | 2,59E-01 | 4,50E-01 |
| MBTPS1 | 3021,93 | 0,02 | 0,02 | 1,13 | 2,59E-01 | 4,50E-01 |
| ACADM | 822,46 | 0,04 | 0,04 | 1,13 | 2,59E-01 | 4,50E-01 |
| KLHL15 | 650,88 | 0,03 | 0,03 | 1,13 | 2,59E-01 | 4,50E-01 |
| AGGF1 | 681,93 | 0,03 | 0,03 | 1,13 | 2,59E-01 | 4,50E-01 |
| TMEM237 | 105,49 | 0,05 | 0,05 | 1,13 | 2,59E-01 | 4,50E-01 |
| SGO2 | 116,14 | 0,07 | 0,06 | 1,13 | 2,60E-01 | 4,51E-01 |
| LRRC25 | 1440,91 | -0,06 | 0,05 | -1,13 | 2,60E-01 | 4,51E-01 |
| TENT4B | 1187,33 | -0,02 | 0,02 | -1,13 | 2,60E-01 | 4,51E-01 |
| FKBP11 | 284,53 | -0,05 | 0,05 | -1,13 | 2,60E-01 | 4,51E-01 |
| NFATC3 | 3484,30 | -0,02 | 0,02 | -1,13 | 2,60E-01 | 4,51E-01 |
| NRCAM | 89,96 | 0,12 | 0,10 | 1,13 | 2,60E-01 | 4,51E-01 |
| RNF8 | 442,16 | 0,02 | 0,02 | 1,13 | 2,60E-01 | 4,51E-01 |
| BDP1 | 3675,57 | 0,03 | 0,03 | 1,13 | 2,60E-01 | 4,51E-01 |
| PRKAG2 | 1033,30 | -0,02 | 0,02 | -1,13 | 2,60E-01 | 4,51E-01 |
| STARD3 | 729,00 | -0,04 | 0,03 | -1,13 | 2,60E-01 | 4,52E-01 |
| NDUFB4 | 833,28 | 0,03 | 0,03 | 1,13 | 2,61E-01 | 4,52E-01 |
| FAM193B | 804,16 | -0,07 | 0,06 | -1,12 | 2,61E-01 | 4,52E-01 |
| CATSPER1 | 99,31 | 0,04 | 0,03 | 1,12 | 2,61E-01 | 4,52E-01 |
| SAMD1 | 74,07 | 0,06 | 0,06 | 1,12 | 2,61E-01 | 4,52E-01 |
| ZNF669 | 284,18 | 0,04 | 0,03 | 1,12 | 2,61E-01 | 4,52E-01 |
| MTSS1 | 1801,57 | 0,04 | 0,03 | 1,12 | 2,61E-01 | 4,53E-01 |
| GTF3C6 | 330,76 | 0,03 | 0,03 | 1,12 | 2,61E-01 | 4,53E-01 |
| TYSND1 | 655,92 | 0,03 | 0,02 | 1,12 | 2,62E-01 | 4,53E-01 |
| SH2D2A | 151,04 | -0,07 | 0,07 | -1,12 | 2,62E-01 | 4,53E-01 |
| TRAF4 | 173,45 | 0,04 | 0,03 | 1,12 | 2,62E-01 | 4,53E-01 |
| UBE2D4 | 140,73 | 0,03 | 0,03 | 1,12 | 2,62E-01 | 4,53E-01 |
| SPECC1L | 332,57 | -0,03 | 0,03 | -1,12 | 2,62E-01 | 4,54E-01 |
| EDEM1 | 3883,17 | -0,03 | 0,02 | -1,12 | 2,63E-01 | 4,54E-01 |
| PPP1CA | 1988,17 | -0,03 | 0,02 | -1,12 | 2,63E-01 | 4,54E-01 |
| RTN2 | 92,68 | -0,06 | 0,05 | -1,12 | 2,63E-01 | 4,54E-01 |
| CSE1L | 1320,93 | 0,03 | 0,02 | 1,12 | 2,63E-01 | 4,54E-01 |
| CLTCL1 | 343,08 | 0,05 | 0,05 | 1,12 | 2,63E-01 | 4,55E-01 |
| ATP8B4 | 1734,50 | 0,04 | 0,04 | 1,12 | 2,63E-01 | 4,55E-01 |
| GPATCH1 | 372,57 | -0,02 | 0,02 | -1,12 | 2,63E-01 | 4,55E-01 |

| | | | | | | |
|---|---|---|---|---|---|---|
| SPIN3 | 591,54 | 0,03 | 0,03 | 1,12 | 2,63E-01 | 4,55E-01 |
| SETD1A | 150,42 | 0,07 | 0,07 | 1,12 | 2,63E-01 | 4,55E-01 |
| FCHO1 | 1137,04 | -0,04 | 0,04 | -1,12 | 2,63E-01 | 4,55E-01 |
| ZNF622 | 1128,83 | -0,03 | 0,03 | -1,12 | 2,63E-01 | 4,55E-01 |
| ZDHHC3 | 1653,38 | -0,02 | 0,02 | -1,12 | 2,63E-01 | 4,55E-01 |
| MAD2L2 | 289,79 | 0,02 | 0,02 | 1,12 | 2,63E-01 | 4,55E-01 |
| POLH | 552,37 | 0,02 | 0,02 | 1,12 | 2,64E-01 | 4,55E-01 |
| PPP1R3E | 673,14 | 0,03 | 0,02 | 1,12 | 2,64E-01 | 4,55E-01 |
| BCL9L | 812,76 | 0,07 | 0,06 | 1,12 | 2,64E-01 | 4,55E-01 |
| GIMAP1 | 2498,11 | 0,02 | 0,02 | 1,12 | 2,64E-01 | 4,55E-01 |
| GSPT2 | 133,13 | 0,04 | 0,03 | 1,12 | 2,64E-01 | 4,56E-01 |
| LSM14B | 906,36 | -0,02 | 0,02 | -1,12 | 2,64E-01 | 4,56E-01 |
| MCTS1 | 425,90 | 0,03 | 0,03 | 1,12 | 2,64E-01 | 4,56E-01 |
| PLGRKT | 287,83 | 0,03 | 0,02 | 1,12 | 2,64E-01 | 4,56E-01 |
| PSMB10 | 1619,79 | -0,03 | 0,03 | -1,12 | 2,65E-01 | 4,56E-01 |
| NBEA | 376,04 | 0,05 | 0,04 | 1,12 | 2,65E-01 | 4,56E-01 |
| NOL8 | 1511,02 | 0,02 | 0,02 | 1,12 | 2,65E-01 | 4,56E-01 |
| SNX18 | 6064,47 | -0,03 | 0,03 | -1,12 | 2,65E-01 | 4,56E-01 |
| RUSC1 | 80,33 | 0,05 | 0,05 | 1,12 | 2,65E-01 | 4,56E-01 |
| PSMB7 | 1153,90 | 0,02 | 0,02 | 1,12 | 2,65E-01 | 4,56E-01 |
| GDAP2 | 2654,09 | -0,02 | 0,02 | -1,12 | 2,65E-01 | 4,56E-01 |
| ZKSCAN5 | 627,82 | -0,02 | 0,02 | -1,11 | 2,65E-01 | 4,57E-01 |
| MAGOH | 440,15 | -0,04 | 0,04 | -1,11 | 2,65E-01 | 4,57E-01 |
| ZFP82 | 259,84 | 0,05 | 0,04 | 1,11 | 2,65E-01 | 4,57E-01 |
| CHMP4A | 163,41 | 0,03 | 0,02 | 1,11 | 2,65E-01 | 4,57E-01 |
| ANKRD23 | 93,61 | 0,03 | 0,03 | 1,11 | 2,66E-01 | 4,57E-01 |
| CTNNBL1 | 1227,40 | -0,02 | 0,02 | -1,11 | 2,66E-01 | 4,57E-01 |
| RAB4B | 152,83 | -0,04 | 0,04 | -1,11 | 2,66E-01 | 4,57E-01 |
| DTX2 | 274,04 | -0,06 | 0,06 | -1,11 | 2,66E-01 | 4,57E-01 |
| TAF1A | 313,74 | 0,03 | 0,03 | 1,11 | 2,66E-01 | 4,57E-01 |
| FGFRL1 | 167,45 | 0,05 | 0,04 | 1,11 | 2,66E-01 | 4,57E-01 |
| TMX1 | 1753,42 | 0,06 | 0,05 | 1,11 | 2,66E-01 | 4,57E-01 |
| ANKMY1 | 1146,46 | 0,04 | 0,03 | 1,11 | 2,66E-01 | 4,57E-01 |
| CCDC82 | 1981,74 | 0,04 | 0,04 | 1,11 | 2,66E-01 | 4,57E-01 |
| LRRC59 | 1218,67 | -0,02 | 0,02 | -1,11 | 2,66E-01 | 4,58E-01 |
| CDHR2 | 114,89 | 0,06 | 0,05 | 1,11 | 2,66E-01 | 4,58E-01 |
| MFSD4B | 1231,50 | 0,02 | 0,02 | 1,11 | 2,66E-01 | 4,58E-01 |
| FHL3 | 391,94 | -0,06 | 0,05 | -1,11 | 2,67E-01 | 4,58E-01 |
| SF1 | 3897,43 | -0,04 | 0,03 | -1,11 | 2,67E-01 | 4,58E-01 |
| MAP3K6 | 124,62 | 0,05 | 0,05 | 1,11 | 2,67E-01 | 4,58E-01 |
| SLC25A1 | 240,10 | 0,03 | 0,03 | 1,11 | 2,67E-01 | 4,58E-01 |
| DIS3L2 | 924,39 | 0,03 | 0,02 | 1,11 | 2,67E-01 | 4,58E-01 |
| YTHDC1 | 5575,91 | -0,03 | 0,03 | -1,11 | 2,67E-01 | 4,58E-01 |
| KIAA2013 | 1131,85 | -0,03 | 0,03 | -1,11 | 2,67E-01 | 4,58E-01 |
| EFCAB14 | 3223,52 | -0,01 | 0,01 | -1,11 | 2,67E-01 | 4,59E-01 |
| PDP2 | 290,54 | 0,03 | 0,02 | 1,11 | 2,67E-01 | 4,59E-01 |

| | | | | | | |
|---|---|---|---|---|---|---|
| ZNF761 | 334,79 | 0,04 | 0,04 | 1,11 | 2,67E-01 | 4,59E-01 |
| ATOSA | 3939,72 | -0,03 | 0,03 | -1,11 | 2,67E-01 | 4,59E-01 |
| TFPT | 247,65 | -0,03 | 0,02 | -1,11 | 2,67E-01 | 4,59E-01 |
| TSR1 | 781,93 | 0,02 | 0,02 | 1,11 | 2,68E-01 | 4,59E-01 |
| TTYH2 | 258,31 | 0,05 | 0,05 | 1,11 | 2,68E-01 | 4,59E-01 |
| SECISBP2 | 5035,53 | -0,02 | 0,02 | -1,11 | 2,68E-01 | 4,59E-01 |
| PPA1 | 600,95 | 0,05 | 0,05 | 1,11 | 2,68E-01 | 4,60E-01 |
| TIRAP | 362,54 | -0,04 | 0,04 | -1,11 | 2,68E-01 | 4,60E-01 |
| PLXNB2 | 869,34 | 0,08 | 0,07 | 1,11 | 2,68E-01 | 4,60E-01 |
| GCSAML | 91,81 | -0,09 | 0,08 | -1,11 | 2,69E-01 | 4,60E-01 |
| RAD52 | 331,86 | 0,03 | 0,02 | 1,11 | 2,69E-01 | 4,60E-01 |
| ABCC3 | 435,72 | 0,07 | 0,07 | 1,11 | 2,69E-01 | 4,60E-01 |
| NOM1 | 948,06 | 0,02 | 0,02 | 1,10 | 2,69E-01 | 4,61E-01 |
| SENP6 | 5745,86 | -0,03 | 0,03 | -1,10 | 2,69E-01 | 4,61E-01 |
| SBK1 | 133,31 | 0,08 | 0,07 | 1,10 | 2,70E-01 | 4,61E-01 |
| HSBP1L1 | 136,41 | 0,04 | 0,04 | 1,10 | 2,70E-01 | 4,62E-01 |
| RIPK2 | 631,44 | -0,03 | 0,03 | -1,10 | 2,70E-01 | 4,62E-01 |
| BLK | 606,14 | 0,06 | 0,06 | 1,10 | 2,70E-01 | 4,62E-01 |
| FTL | 44760,18 | 0,05 | 0,04 | 1,10 | 2,70E-01 | 4,62E-01 |
| C12orf76 | 494,99 | 0,02 | 0,02 | 1,10 | 2,70E-01 | 4,62E-01 |
| CAPN12 | 151,15 | 0,06 | 0,05 | 1,10 | 2,70E-01 | 4,62E-01 |
| CAPN5 | 158,84 | 0,07 | 0,07 | 1,10 | 2,70E-01 | 4,62E-01 |
| TBC1D2B | 3083,14 | -0,03 | 0,02 | -1,10 | 2,70E-01 | 4,62E-01 |
| CLEC9A | 80,65 | -0,07 | 0,07 | -1,10 | 2,70E-01 | 4,62E-01 |
| ZBTB7A | 628,70 | 0,05 | 0,05 | 1,10 | 2,71E-01 | 4,63E-01 |
| IKZF1 | 11698,88 | -0,03 | 0,02 | -1,10 | 2,71E-01 | 4,63E-01 |
| USP37 | 1267,54 | 0,02 | 0,02 | 1,10 | 2,71E-01 | 4,63E-01 |
| TCEA3 | 140,39 | 0,06 | 0,05 | 1,10 | 2,71E-01 | 4,63E-01 |
| BLM | 331,62 | 0,03 | 0,02 | 1,10 | 2,71E-01 | 4,63E-01 |
| NCKAP1 | 349,99 | 0,04 | 0,04 | 1,10 | 2,71E-01 | 4,63E-01 |
| MYL12A | 8340,69 | -0,03 | 0,03 | -1,10 | 2,71E-01 | 4,63E-01 |
| NVL | 748,86 | 0,02 | 0,02 | 1,10 | 2,71E-01 | 4,63E-01 |
| GALNS | 743,52 | -0,06 | 0,06 | -1,10 | 2,72E-01 | 4,64E-01 |
| PAQR4 | 72,92 | 0,05 | 0,05 | 1,10 | 2,72E-01 | 4,64E-01 |
| ADCY7 | 7112,97 | -0,03 | 0,03 | -1,10 | 2,72E-01 | 4,64E-01 |
| SHMT1 | 229,84 | 0,04 | 0,04 | 1,10 | 2,72E-01 | 4,64E-01 |
| EMILIN2 | 2125,20 | -0,04 | 0,03 | -1,10 | 2,72E-01 | 4,64E-01 |
| EPOR | 124,40 | 0,05 | 0,05 | 1,10 | 2,72E-01 | 4,64E-01 |
| DFFA | 1132,93 | 0,02 | 0,02 | 1,10 | 2,72E-01 | 4,64E-01 |
| SLC10A3 | 180,15 | 0,04 | 0,04 | 1,10 | 2,72E-01 | 4,64E-01 |
| TUT4 | 3171,46 | -0,02 | 0,02 | -1,10 | 2,73E-01 | 4,65E-01 |
| ERBB2 | 214,29 | -0,06 | 0,06 | -1,10 | 2,73E-01 | 4,65E-01 |
| ATG16L1 | 1841,85 | -0,02 | 0,02 | -1,10 | 2,73E-01 | 4,65E-01 |
| PHC3 | 8626,29 | 0,02 | 0,02 | 1,10 | 2,73E-01 | 4,66E-01 |
| CCDC77 | 266,04 | -0,02 | 0,02 | -1,10 | 2,73E-01 | 4,66E-01 |
| ITGB1BP1 | 1079,03 | 0,02 | 0,02 | 1,10 | 2,73E-01 | 4,66E-01 |

| | | | | | | |
|---|---|---|---|---|---|---|
| MRPL47 | 203,38 | 0,06 | 0,05 | 1,09 | 2,74E-01 | 4,66E-01 |
| ACADVL | 2316,32 | -0,03 | 0,03 | -1,09 | 2,74E-01 | 4,66E-01 |
| REC8 | 174,51 | -0,08 | 0,07 | -1,09 | 2,74E-01 | 4,67E-01 |
| C1orf216 | 247,76 | 0,03 | 0,03 | 1,09 | 2,74E-01 | 4,67E-01 |
| PREB | 435,60 | 0,02 | 0,02 | 1,09 | 2,74E-01 | 4,67E-01 |
| ASTE1 | 699,69 | 0,03 | 0,03 | 1,09 | 2,74E-01 | 4,67E-01 |
| FLYWCH1 | 584,25 | 0,05 | 0,04 | 1,09 | 2,74E-01 | 4,67E-01 |
| RUVBL2 | 481,87 | 0,02 | 0,02 | 1,09 | 2,75E-01 | 4,67E-01 |
| EIF2AK3 | 1168,67 | 0,02 | 0,02 | 1,09 | 2,75E-01 | 4,67E-01 |
| MEAF6 | 1163,41 | 0,02 | 0,02 | 1,09 | 2,75E-01 | 4,68E-01 |
| MTRF1 | 525,62 | 0,02 | 0,02 | 1,09 | 2,75E-01 | 4,68E-01 |
| SREBF2 | 603,60 | 0,06 | 0,05 | 1,09 | 2,75E-01 | 4,68E-01 |
| BCL7A | 121,90 | 0,06 | 0,05 | 1,09 | 2,75E-01 | 4,68E-01 |
| ARHGEF37 | 127,68 | 0,08 | 0,08 | 1,09 | 2,75E-01 | 4,68E-01 |
| SEPTIN10 | 110,40 | 0,05 | 0,05 | 1,09 | 2,75E-01 | 4,68E-01 |
| TMEM106C | 298,92 | 0,03 | 0,02 | 1,09 | 2,75E-01 | 4,68E-01 |
| DBI | 424,37 | 0,05 | 0,05 | 1,09 | 2,75E-01 | 4,68E-01 |
| ZNF442 | 90,46 | 0,06 | 0,05 | 1,09 | 2,75E-01 | 4,68E-01 |
| GTPBP10 | 1293,67 | 0,03 | 0,03 | 1,09 | 2,76E-01 | 4,68E-01 |
| DERL1 | 2295,28 | -0,02 | 0,01 | -1,09 | 2,76E-01 | 4,68E-01 |
| CAMKK1 | 390,00 | -0,06 | 0,05 | -1,09 | 2,76E-01 | 4,68E-01 |
| CXCR5 | 291,83 | 0,06 | 0,05 | 1,09 | 2,76E-01 | 4,68E-01 |
| MKS1 | 171,77 | 0,03 | 0,03 | 1,09 | 2,76E-01 | 4,68E-01 |
| PI16 | 121,21 | 0,06 | 0,05 | 1,09 | 2,76E-01 | 4,68E-01 |
| ACSF3 | 1590,95 | 0,03 | 0,03 | 1,09 | 2,76E-01 | 4,68E-01 |
| PLGLB1 | 130,89 | -0,05 | 0,05 | -1,09 | 2,76E-01 | 4,68E-01 |
| CD4 | 1223,85 | 0,05 | 0,04 | 1,09 | 2,76E-01 | 4,69E-01 |
| ATF2 | 2571,42 | 0,03 | 0,03 | 1,09 | 2,76E-01 | 4,69E-01 |
| NET1 | 287,84 | 0,04 | 0,04 | 1,09 | 2,77E-01 | 4,70E-01 |
| GNB2 | 3196,72 | -0,04 | 0,04 | -1,09 | 2,77E-01 | 4,70E-01 |
| UNG | 270,73 | 0,03 | 0,02 | 1,09 | 2,77E-01 | 4,70E-01 |
| CD3E | 662,94 | 0,04 | 0,04 | 1,09 | 2,77E-01 | 4,70E-01 |
| LY75 | 1028,66 | -0,03 | 0,03 | -1,09 | 2,78E-01 | 4,71E-01 |
| LBHD1 | 225,86 | 0,03 | 0,03 | 1,09 | 2,78E-01 | 4,71E-01 |
| RIOK2 | 566,35 | 0,05 | 0,04 | 1,09 | 2,78E-01 | 4,71E-01 |
| RP9 | 149,59 | 0,03 | 0,02 | 1,09 | 2,78E-01 | 4,71E-01 |
| BEGAIN | 77,74 | 0,13 | 0,12 | 1,09 | 2,78E-01 | 4,71E-01 |
| IL11RA | 404,48 | 0,05 | 0,05 | 1,08 | 2,78E-01 | 4,71E-01 |
| ATG5 | 999,92 | 0,04 | 0,04 | 1,08 | 2,78E-01 | 4,71E-01 |
| GRSF1 | 1770,40 | 0,02 | 0,02 | 1,08 | 2,79E-01 | 4,72E-01 |
| NXPE3 | 1559,02 | -0,02 | 0,02 | -1,08 | 2,79E-01 | 4,72E-01 |
| PPIE | 782,02 | 0,03 | 0,03 | 1,08 | 2,79E-01 | 4,72E-01 |
| VPS37A | 1236,63 | 0,02 | 0,02 | 1,08 | 2,79E-01 | 4,72E-01 |
| MRTFA | 1504,84 | -0,04 | 0,04 | -1,08 | 2,79E-01 | 4,72E-01 |
| TCIRG1 | 3757,66 | -0,05 | 0,04 | -1,08 | 2,79E-01 | 4,72E-01 |
| NID1 | 137,50 | 0,07 | 0,07 | 1,08 | 2,79E-01 | 4,72E-01 |

| | | | | | | |
|---|---|---|---|---|---|---|
| CST7 | 387,56 | -0,05 | 0,05 | -1,08 | 2,79E-01 | 4,72E-01 |
| NDUFS5 | 325,15 | 0,06 | 0,06 | 1,08 | 2,79E-01 | 4,72E-01 |
| GABPA | 1878,61 | 0,04 | 0,04 | 1,08 | 2,79E-01 | 4,73E-01 |
| PRUNE1 | 674,21 | -0,02 | 0,02 | -1,08 | 2,80E-01 | 4,73E-01 |
| INCENP | 282,18 | -0,03 | 0,03 | -1,08 | 2,80E-01 | 4,73E-01 |
| GPN2 | 709,56 | 0,02 | 0,02 | 1,08 | 2,80E-01 | 4,74E-01 |
| STAMBPL1 | 1177,38 | 0,03 | 0,03 | 1,08 | 2,80E-01 | 4,74E-01 |
| GORASP1 | 714,86 | -0,03 | 0,02 | -1,08 | 2,81E-01 | 4,74E-01 |
| N4BP2L1 | 1343,56 | 0,02 | 0,02 | 1,08 | 2,81E-01 | 4,74E-01 |
| GMIP | 2316,89 | -0,07 | 0,06 | -1,08 | 2,81E-01 | 4,74E-01 |
| CAPN2 | 3797,90 | 0,02 | 0,02 | 1,08 | 2,81E-01 | 4,74E-01 |
| TUBGCP6 | 1699,38 | 0,04 | 0,04 | 1,08 | 2,81E-01 | 4,75E-01 |
| EPB41L2 | 481,23 | 0,04 | 0,03 | 1,08 | 2,82E-01 | 4,75E-01 |
| ARG1 | 281,19 | -0,08 | 0,08 | -1,08 | 2,82E-01 | 4,76E-01 |
| CENPL | 206,78 | 0,03 | 0,02 | 1,08 | 2,82E-01 | 4,76E-01 |
| NLK | 962,37 | 0,02 | 0,02 | 1,08 | 2,82E-01 | 4,76E-01 |
| TUBD1 | 354,78 | 0,02 | 0,02 | 1,08 | 2,82E-01 | 4,76E-01 |
| ZNF35 | 234,04 | 0,03 | 0,03 | 1,08 | 2,82E-01 | 4,76E-01 |
| RABGGTA | 92,33 | 0,06 | 0,05 | 1,08 | 2,82E-01 | 4,76E-01 |
| DHX32 | 350,01 | 0,03 | 0,02 | 1,08 | 2,82E-01 | 4,76E-01 |
| SCAF11 | 10236,21 | -0,03 | 0,03 | -1,08 | 2,82E-01 | 4,76E-01 |
| ERG28 | 166,93 | 0,02 | 0,02 | 1,08 | 2,82E-01 | 4,76E-01 |
| LPAR5 | 201,72 | 0,04 | 0,04 | 1,08 | 2,82E-01 | 4,76E-01 |
| KMT2D | 4446,30 | -0,07 | 0,06 | -1,07 | 2,82E-01 | 4,76E-01 |
| TCAF2 | 184,41 | 0,04 | 0,03 | 1,07 | 2,82E-01 | 4,76E-01 |
| RRP36 | 717,65 | 0,02 | 0,02 | 1,07 | 2,83E-01 | 4,76E-01 |
| ITLN1 | 100,02 | 0,11 | 0,10 | 1,07 | 2,83E-01 | 4,76E-01 |
| TREML1 | 168,43 | 0,08 | 0,07 | 1,07 | 2,83E-01 | 4,76E-01 |
| PF4 | 2290,81 | 0,07 | 0,07 | 1,07 | 2,83E-01 | 4,76E-01 |
| CISD1 | 239,07 | 0,05 | 0,04 | 1,07 | 2,83E-01 | 4,76E-01 |
| CYCS | 1542,03 | 0,04 | 0,03 | 1,07 | 2,83E-01 | 4,76E-01 |
| DOT1L | 317,50 | 0,07 | 0,07 | 1,07 | 2,83E-01 | 4,77E-01 |
| CCDC157 | 76,86 | 0,06 | 0,05 | 1,07 | 2,83E-01 | 4,77E-01 |
| IL21R | 669,38 | 0,03 | 0,03 | 1,07 | 2,83E-01 | 4,77E-01 |
| TANK | 6101,97 | -0,03 | 0,03 | -1,07 | 2,83E-01 | 4,77E-01 |
| BUB3 | 2828,51 | 0,02 | 0,02 | 1,07 | 2,83E-01 | 4,77E-01 |
| CHPF2 | 1221,89 | -0,04 | 0,04 | -1,07 | 2,83E-01 | 4,77E-01 |
| CCDC171 | 152,14 | 0,05 | 0,05 | 1,07 | 2,83E-01 | 4,77E-01 |
| ACAD9 | 998,70 | 0,02 | 0,02 | 1,07 | 2,83E-01 | 4,77E-01 |
| CETN3 | 180,53 | 0,06 | 0,06 | 1,07 | 2,84E-01 | 4,77E-01 |
| MEGF8 | 228,70 | 0,06 | 0,06 | 1,07 | 2,84E-01 | 4,77E-01 |
| KCNMA1 | 163,77 | -0,15 | 0,14 | -1,07 | 2,84E-01 | 4,77E-01 |
| AZI2 | 948,52 | 0,05 | 0,05 | 1,07 | 2,84E-01 | 4,77E-01 |
| TMEM144 | 395,77 | 0,05 | 0,05 | 1,07 | 2,84E-01 | 4,77E-01 |
| PSMG2 | 778,54 | 0,02 | 0,02 | 1,07 | 2,84E-01 | 4,77E-01 |
| VCF2 | 109,75 | 0,05 | 0,04 | 1,07 | 2,84E-01 | 4,77E-01 |

| | | | | | | |
|---|---|---|---|---|---|---|
| IL12RB1 | 605,07 | -0,03 | 0,03 | -1,07 | 2,84E-01 | 4,77E-01 |
| TPM4 | 5457,90 | -0,03 | 0,02 | -1,07 | 2,84E-01 | 4,77E-01 |
| DSN1 | 269,81 | -0,02 | 0,02 | -1,07 | 2,84E-01 | 4,77E-01 |
| ZNF839 | 480,72 | -0,02 | 0,02 | -1,07 | 2,84E-01 | 4,78E-01 |
| NRXN1 | 162,37 | -0,08 | 0,08 | -1,07 | 2,85E-01 | 4,78E-01 |
| DOCK7 | 580,93 | 0,04 | 0,03 | 1,07 | 2,85E-01 | 4,78E-01 |
| NAB2 | 226,12 | 0,03 | 0,03 | 1,07 | 2,85E-01 | 4,78E-01 |
| MTA3 | 351,24 | 0,03 | 0,03 | 1,07 | 2,85E-01 | 4,78E-01 |
| POMGNT2 | 82,88 | 0,04 | 0,04 | 1,07 | 2,85E-01 | 4,79E-01 |
| CARD19 | 728,82 | 0,03 | 0,03 | 1,07 | 2,85E-01 | 4,79E-01 |
| RNPEPL1 | 764,56 | 0,07 | 0,06 | 1,07 | 2,85E-01 | 4,79E-01 |
| NUCB2 | 693,97 | 0,03 | 0,03 | 1,07 | 2,86E-01 | 4,79E-01 |
| FGD2 | 1379,35 | 0,05 | 0,05 | 1,07 | 2,86E-01 | 4,79E-01 |
| PFAS | 158,06 | 0,06 | 0,05 | 1,07 | 2,85E-01 | 4,79E-01 |
| PCNX2 | 1663,11 | 0,03 | 0,03 | 1,07 | 2,86E-01 | 4,79E-01 |
| TMEM86A | 90,74 | 0,04 | 0,04 | 1,07 | 2,86E-01 | 4,79E-01 |
| RHOQ | 2663,37 | -0,02 | 0,02 | -1,07 | 2,86E-01 | 4,79E-01 |
| COG2 | 1227,29 | 0,02 | 0,02 | 1,07 | 2,86E-01 | 4,79E-01 |
| RAB9A | 624,12 | -0,02 | 0,02 | -1,07 | 2,86E-01 | 4,79E-01 |
| TOP2A | 133,01 | 0,06 | 0,06 | 1,07 | 2,86E-01 | 4,80E-01 |
| CUTC | 685,10 | -0,03 | 0,03 | -1,07 | 2,86E-01 | 4,80E-01 |
| NGDN | 549,23 | 0,03 | 0,03 | 1,07 | 2,87E-01 | 4,80E-01 |
| SETMAR | 126,78 | 0,03 | 0,03 | 1,07 | 2,87E-01 | 4,80E-01 |
| FOLR3 | 699,93 | -0,10 | 0,09 | -1,07 | 2,87E-01 | 4,80E-01 |
| DSTN | 549,28 | 0,02 | 0,02 | 1,06 | 2,87E-01 | 4,80E-01 |
| ERF | 347,52 | -0,05 | 0,04 | -1,06 | 2,87E-01 | 4,81E-01 |
| TYW3 | 537,65 | 0,04 | 0,04 | 1,06 | 2,87E-01 | 4,81E-01 |
| NRG1 | 274,36 | 0,09 | 0,08 | 1,06 | 2,87E-01 | 4,81E-01 |
| TMEM70 | 549,68 | 0,03 | 0,03 | 1,06 | 2,87E-01 | 4,81E-01 |
| U2AF2 | 1401,72 | -0,03 | 0,03 | -1,06 | 2,88E-01 | 4,81E-01 |
| ZNF721 | 754,87 | 0,06 | 0,05 | 1,06 | 2,88E-01 | 4,81E-01 |
| ASL | 300,04 | -0,02 | 0,02 | -1,06 | 2,88E-01 | 4,81E-01 |
| STAP1 | 272,87 | 0,06 | 0,06 | 1,06 | 2,88E-01 | 4,82E-01 |
| SYNM | 317,14 | -0,09 | 0,08 | -1,06 | 2,88E-01 | 4,82E-01 |
| TRPV2 | 1147,68 | 0,03 | 0,03 | 1,06 | 2,88E-01 | 4,82E-01 |
| CRCP | 827,33 | -0,02 | 0,02 | -1,06 | 2,89E-01 | 4,82E-01 |
| KLHL17 | 121,91 | 0,05 | 0,05 | 1,06 | 2,88E-01 | 4,82E-01 |
| EIF2B1 | 974,77 | 0,01 | 0,01 | 1,06 | 2,89E-01 | 4,82E-01 |
| ZNF589 | 1016,34 | 0,02 | 0,02 | 1,06 | 2,89E-01 | 4,82E-01 |
| RPUSD1 | 149,13 | 0,04 | 0,04 | 1,06 | 2,89E-01 | 4,82E-01 |
| SULT1A1 | 1476,61 | 0,06 | 0,05 | 1,06 | 2,89E-01 | 4,82E-01 |
| ZYX | 1330,16 | -0,07 | 0,07 | -1,06 | 2,89E-01 | 4,82E-01 |
| CCDC59 | 679,39 | 0,05 | 0,05 | 1,06 | 2,89E-01 | 4,82E-01 |
| CCDC186 | 2705,57 | -0,04 | 0,04 | -1,06 | 2,89E-01 | 4,82E-01 |
| CAPN7 | 2555,92 | 0,03 | 0,02 | 1,06 | 2,89E-01 | 4,82E-01 |
| NOP56 | 1342,39 | 0,02 | 0,02 | 1,06 | 2,89E-01 | 4,82E-01 |

| | | | | | | |
|---|---|---|---|---|---|---|
| VPS16 | 1215,93 | -0,02 | 0,02 | -1,06 | 2,89E-01 | 4,82E-01 |
| ALDH16A1 | 732,90 | 0,03 | 0,03 | 1,06 | 2,89E-01 | 4,83E-01 |
| LTB | 4415,90 | 0,03 | 0,03 | 1,06 | 2,89E-01 | 4,83E-01 |
| ZFP36 | 1441,69 | -0,04 | 0,04 | -1,06 | 2,89E-01 | 4,83E-01 |
| CBX8 | 139,78 | -0,04 | 0,04 | -1,06 | 2,90E-01 | 4,83E-01 |
| ARFIP2 | 444,56 | 0,01 | 0,01 | 1,06 | 2,90E-01 | 4,83E-01 |
| TRIM66 | 924,41 | -0,05 | 0,04 | -1,06 | 2,90E-01 | 4,83E-01 |
| GATAD2A | 1415,24 | -0,05 | 0,05 | -1,06 | 2,90E-01 | 4,83E-01 |
| POLR2J2 | 98,77 | 0,07 | 0,06 | 1,06 | 2,90E-01 | 4,83E-01 |
| JAM3 | 427,55 | -0,06 | 0,05 | -1,06 | 2,90E-01 | 4,83E-01 |
| CYSLTR2 | 714,13 | 0,05 | 0,04 | 1,06 | 2,90E-01 | 4,84E-01 |
| SPOCK2 | 2991,16 | 0,04 | 0,04 | 1,06 | 2,90E-01 | 4,84E-01 |
| NEDD4L | 365,36 | 0,06 | 0,06 | 1,06 | 2,90E-01 | 4,84E-01 |
| HPRT1 | 123,21 | 0,04 | 0,04 | 1,06 | 2,91E-01 | 4,84E-01 |
| KMO | 353,79 | 0,04 | 0,04 | 1,06 | 2,91E-01 | 4,84E-01 |
| STARD7 | 3141,61 | 0,02 | 0,02 | 1,06 | 2,91E-01 | 4,84E-01 |
| SCAP | 1216,36 | -0,05 | 0,05 | -1,06 | 2,91E-01 | 4,84E-01 |
| CLK2 | 927,53 | -0,02 | 0,02 | -1,06 | 2,91E-01 | 4,84E-01 |
| ZNF267 | 3003,66 | -0,05 | 0,04 | -1,06 | 2,91E-01 | 4,84E-01 |
| SLAIN2 | 1782,70 | -0,02 | 0,02 | -1,06 | 2,91E-01 | 4,84E-01 |
| KANTR | 902,58 | 0,03 | 0,03 | 1,05 | 2,92E-01 | 4,85E-01 |
| MRAS | 91,32 | 0,07 | 0,07 | 1,05 | 2,92E-01 | 4,85E-01 |
| ANAPC1 | 1582,98 | 0,02 | 0,02 | 1,05 | 2,92E-01 | 4,85E-01 |
| RNF146 | 2363,47 | -0,03 | 0,03 | -1,05 | 2,92E-01 | 4,86E-01 |
| RAB11A | 1986,95 | -0,03 | 0,03 | -1,05 | 2,92E-01 | 4,86E-01 |
| CIDEB | 630,58 | -0,03 | 0,03 | -1,05 | 2,92E-01 | 4,86E-01 |
| THYN1 | 399,32 | 0,02 | 0,02 | 1,05 | 2,92E-01 | 4,86E-01 |
| BHLHE40 | 854,70 | -0,03 | 0,03 | -1,05 | 2,92E-01 | 4,86E-01 |
| ACTR3C | 492,90 | -0,03 | 0,03 | -1,05 | 2,93E-01 | 4,86E-01 |
| SLC29A3 | 116,83 | 0,04 | 0,04 | 1,05 | 2,93E-01 | 4,86E-01 |
| CHD1 | 6425,60 | -0,03 | 0,03 | -1,05 | 2,93E-01 | 4,86E-01 |
| MOAP1 | 1011,83 | -0,02 | 0,02 | -1,05 | 2,93E-01 | 4,87E-01 |
| CCT6B | 80,13 | 0,05 | 0,04 | 1,05 | 2,93E-01 | 4,87E-01 |
| IKBKB | 2935,79 | -0,03 | 0,03 | -1,05 | 2,93E-01 | 4,87E-01 |
| ABT1 | 1447,85 | 0,02 | 0,02 | 1,05 | 2,93E-01 | 4,87E-01 |
| SMC1A | 2168,63 | -0,02 | 0,02 | -1,05 | 2,93E-01 | 4,87E-01 |
| C11orf21 | 3145,93 | 0,03 | 0,03 | 1,05 | 2,93E-01 | 4,87E-01 |
| BCL2A1 | 626,48 | -0,09 | 0,08 | -1,05 | 2,94E-01 | 4,87E-01 |
| PPP2CA | 2609,07 | -0,03 | 0,02 | -1,05 | 2,94E-01 | 4,87E-01 |
| ZNF81 | 1136,68 | -0,02 | 0,02 | -1,05 | 2,94E-01 | 4,88E-01 |
| PES1 | 794,14 | 0,02 | 0,02 | 1,05 | 2,94E-01 | 4,88E-01 |
| CIITA | 2596,28 | 0,06 | 0,06 | 1,05 | 2,94E-01 | 4,88E-01 |
| COQ10A | 180,88 | 0,02 | 0,02 | 1,05 | 2,94E-01 | 4,88E-01 |
| MILR1 | 421,26 | -0,05 | 0,04 | -1,05 | 2,95E-01 | 4,88E-01 |
| KGD4 | 355,29 | 0,03 | 0,03 | 1,05 | 2,95E-01 | 4,88E-01 |
| METAP2 | 812,81 | 0,04 | 0,04 | 1,05 | 2,95E-01 | 4,89E-01 |

| | | | | | | |
|---|---|---|---|---|---|---|
| RABEPK | 366,18 | -0,02 | 0,02 | -1,05 | 2,95E-01 | 4,89E-01 |
| CD8A | 1714,54 | 0,07 | 0,06 | 1,05 | 2,95E-01 | 4,89E-01 |
| BAK1 | 652,25 | -0,03 | 0,03 | -1,05 | 2,95E-01 | 4,89E-01 |
| LRRC41 | 664,36 | 0,02 | 0,02 | 1,05 | 2,95E-01 | 4,89E-01 |
| CCDC12 | 841,56 | 0,02 | 0,02 | 1,05 | 2,96E-01 | 4,89E-01 |
| TMTC1 | 480,69 | -0,11 | 0,10 | -1,05 | 2,96E-01 | 4,89E-01 |
| WDR77 | 423,61 | 0,02 | 0,02 | 1,05 | 2,96E-01 | 4,89E-01 |
| MFN1 | 2548,12 | -0,03 | 0,03 | -1,05 | 2,96E-01 | 4,89E-01 |
| WBP11 | 434,69 | 0,03 | 0,03 | 1,05 | 2,96E-01 | 4,89E-01 |
| PLD6 | 161,32 | 0,04 | 0,04 | 1,05 | 2,96E-01 | 4,89E-01 |
| CHURC1 | 2110,68 | 0,03 | 0,03 | 1,05 | 2,96E-01 | 4,89E-01 |
| ACBD6 | 1294,87 | 0,02 | 0,02 | 1,05 | 2,96E-01 | 4,89E-01 |
| CAPNS1 | 469,72 | 0,04 | 0,04 | 1,05 | 2,96E-01 | 4,89E-01 |
| IFT25 | 551,88 | 0,02 | 0,02 | 1,05 | 2,96E-01 | 4,89E-01 |
| HMGXB3 | 1128,81 | 0,02 | 0,02 | 1,04 | 2,96E-01 | 4,90E-01 |
| LTC4S | 129,75 | -0,04 | 0,04 | -1,04 | 2,96E-01 | 4,90E-01 |
| MRPS31 | 635,37 | 0,03 | 0,03 | 1,04 | 2,96E-01 | 4,90E-01 |
| HLA-DOB | 218,33 | 0,06 | 0,06 | 1,04 | 2,96E-01 | 4,90E-01 |
| KLHL32 | 79,12 | 0,04 | 0,04 | 1,04 | 2,96E-01 | 4,90E-01 |
| MBD6 | 412,33 | -0,08 | 0,08 | -1,04 | 2,97E-01 | 4,90E-01 |
| ARID1A | 3156,12 | -0,04 | 0,04 | -1,04 | 2,97E-01 | 4,90E-01 |
| RAI1 | 336,42 | 0,05 | 0,05 | 1,04 | 2,97E-01 | 4,90E-01 |
| LSM11 | 341,42 | 0,02 | 0,02 | 1,04 | 2,97E-01 | 4,90E-01 |
| ABI1 | 5347,98 | -0,03 | 0,03 | -1,04 | 2,97E-01 | 4,90E-01 |
| PRPF8 | 4312,58 | -0,04 | 0,03 | -1,04 | 2,97E-01 | 4,90E-01 |
| AFG2B | 730,23 | -0,03 | 0,03 | -1,04 | 2,97E-01 | 4,90E-01 |
| UNC13D | 2095,41 | -0,06 | 0,06 | -1,04 | 2,97E-01 | 4,91E-01 |
| ZSCAN16 | 302,74 | -0,03 | 0,03 | -1,04 | 2,97E-01 | 4,91E-01 |
| ZNF180 | 614,82 | 0,03 | 0,03 | 1,04 | 2,97E-01 | 4,91E-01 |
| STIM2 | 1908,63 | 0,02 | 0,02 | 1,04 | 2,98E-01 | 4,91E-01 |
| TMED10 | 3728,74 | 0,02 | 0,02 | 1,04 | 2,98E-01 | 4,91E-01 |
| SLC4A4 | 228,04 | 0,06 | 0,05 | 1,04 | 2,98E-01 | 4,91E-01 |
| UBR3 | 3809,97 | -0,02 | 0,02 | -1,04 | 2,98E-01 | 4,91E-01 |
| TIMM22 | 379,52 | 0,02 | 0,02 | 1,04 | 2,98E-01 | 4,92E-01 |
| SH2B2 | 258,32 | -0,06 | 0,05 | -1,04 | 2,98E-01 | 4,92E-01 |
| RASA1 | 2478,99 | 0,02 | 0,02 | 1,04 | 2,99E-01 | 4,93E-01 |
| WDR46 | 667,52 | 0,02 | 0,02 | 1,04 | 2,99E-01 | 4,93E-01 |
| CCNB1 | 97,38 | 0,05 | 0,05 | 1,04 | 2,99E-01 | 4,93E-01 |
| HDAC6 | 951,01 | 0,03 | 0,03 | 1,04 | 2,99E-01 | 4,93E-01 |
| IL7R | 19738,27 | 0,04 | 0,03 | 1,04 | 3,00E-01 | 4,94E-01 |
| DHX36 | 2742,49 | 0,02 | 0,02 | 1,04 | 3,00E-01 | 4,94E-01 |
| ISCU | 3157,11 | 0,03 | 0,03 | 1,04 | 3,00E-01 | 4,94E-01 |
| RAB5B | 1440,69 | -0,05 | 0,05 | -1,04 | 3,00E-01 | 4,94E-01 |
| PHYH | 115,44 | 0,04 | 0,03 | 1,04 | 3,00E-01 | 4,94E-01 |
| NOL7 | 809,97 | 0,02 | 0,02 | 1,04 | 3,00E-01 | 4,94E-01 |
| RABEP2 | 547,31 | 0,03 | 0,03 | 1,04 | 3,00E-01 | 4,94E-01 |

| | | | | | | |
|---|---|---|---|---|---|---|
| SPEF2 | 500,34 | 0,04 | 0,04 | 1,04 | 3,00E-01 | 4,94E-01 |
| ZNF467 | 533,93 | -0,06 | 0,06 | -1,04 | 3,01E-01 | 4,94E-01 |
| TNKS | 2544,32 | -0,02 | 0,02 | -1,04 | 3,01E-01 | 4,94E-01 |
| PEX2 | 1429,64 | 0,03 | 0,03 | 1,03 | 3,01E-01 | 4,95E-01 |
| STX2 | 557,12 | 0,04 | 0,04 | 1,03 | 3,01E-01 | 4,95E-01 |
| STAG2 | 10247,06 | -0,04 | 0,04 | -1,03 | 3,01E-01 | 4,95E-01 |
| C5orf15 | 891,15 | 0,03 | 0,03 | 1,03 | 3,01E-01 | 4,95E-01 |
| CPSF1 | 636,72 | 0,05 | 0,05 | 1,03 | 3,02E-01 | 4,96E-01 |
| NRXN3 | 92,04 | 0,12 | 0,11 | 1,03 | 3,02E-01 | 4,96E-01 |
| C2orf49 | 978,87 | 0,03 | 0,02 | 1,03 | 3,02E-01 | 4,96E-01 |
| PIGC | 1479,27 | 0,06 | 0,06 | 1,03 | 3,02E-01 | 4,96E-01 |
| BRD7 | 1892,44 | -0,02 | 0,02 | -1,03 | 3,02E-01 | 4,96E-01 |
| FBXO8 | 729,23 | 0,04 | 0,04 | 1,03 | 3,02E-01 | 4,96E-01 |
| PIGG | 2033,12 | -0,02 | 0,02 | -1,03 | 3,02E-01 | 4,96E-01 |
| ZNF347 | 533,25 | 0,03 | 0,03 | 1,03 | 3,02E-01 | 4,96E-01 |
| FCRL1 | 2043,54 | 0,07 | 0,06 | 1,03 | 3,02E-01 | 4,96E-01 |
| ATF7IP | 7919,91 | -0,02 | 0,02 | -1,03 | 3,02E-01 | 4,96E-01 |
| DDX11 | 516,19 | 0,08 | 0,07 | 1,03 | 3,02E-01 | 4,96E-01 |
| GDPGP1 | 73,70 | 0,04 | 0,04 | 1,03 | 3,02E-01 | 4,96E-01 |
| PLEKHO1 | 3405,81 | -0,03 | 0,02 | -1,03 | 3,03E-01 | 4,96E-01 |
| SLC25A28 | 633,86 | -0,02 | 0,02 | -1,03 | 3,03E-01 | 4,97E-01 |
| CDV3 | 6175,91 | -0,03 | 0,03 | -1,03 | 3,03E-01 | 4,97E-01 |
| RBM6 | 3040,63 | -0,03 | 0,02 | -1,03 | 3,03E-01 | 4,97E-01 |
| TKFC | 185,99 | 0,04 | 0,04 | 1,03 | 3,03E-01 | 4,98E-01 |
| GZMM | 1043,58 | 0,04 | 0,04 | 1,03 | 3,04E-01 | 4,98E-01 |
| RANBP3 | 738,03 | -0,04 | 0,04 | -1,03 | 3,04E-01 | 4,98E-01 |
| LMBRD1 | 3134,43 | -0,03 | 0,03 | -1,03 | 3,04E-01 | 4,99E-01 |
| THAP1 | 319,48 | 0,03 | 0,03 | 1,03 | 3,04E-01 | 4,99E-01 |
| IFI30 | 247,57 | 0,04 | 0,04 | 1,03 | 3,04E-01 | 4,99E-01 |
| GPATCH8 | 2652,69 | -0,02 | 0,02 | -1,03 | 3,04E-01 | 4,99E-01 |
| PRKAR1B | 143,88 | 0,06 | 0,06 | 1,03 | 3,04E-01 | 4,99E-01 |
| ALG1 | 406,61 | 0,02 | 0,02 | 1,03 | 3,04E-01 | 4,99E-01 |
| NEDD4 | 318,28 | 0,04 | 0,04 | 1,03 | 3,04E-01 | 4,99E-01 |
| ANGPT1 | 352,84 | -0,06 | 0,06 | -1,03 | 3,04E-01 | 4,99E-01 |
| STK16 | 636,34 | 0,02 | 0,02 | 1,03 | 3,05E-01 | 4,99E-01 |
| ATL2 | 1219,27 | 0,03 | 0,03 | 1,03 | 3,05E-01 | 4,99E-01 |
| MYCBP | 413,66 | -0,03 | 0,03 | -1,03 | 3,05E-01 | 4,99E-01 |
| ZBTB44 | 4240,91 | 0,02 | 0,02 | 1,03 | 3,05E-01 | 4,99E-01 |
| ARIH2 | 3042,22 | -0,01 | 0,01 | -1,03 | 3,05E-01 | 5,00E-01 |
| FUCA1 | 604,19 | 0,03 | 0,03 | 1,02 | 3,06E-01 | 5,00E-01 |
| GGA1 | 662,57 | 0,04 | 0,04 | 1,02 | 3,06E-01 | 5,00E-01 |
| LUC7L3 | 5791,17 | 0,03 | 0,03 | 1,02 | 3,06E-01 | 5,00E-01 |
| DMAP1 | 902,07 | 0,02 | 0,02 | 1,02 | 3,06E-01 | 5,00E-01 |
| CIB1 | 1028,95 | 0,03 | 0,02 | 1,02 | 3,06E-01 | 5,00E-01 |
| GLOD4 | 1170,41 | 0,02 | 0,02 | 1,02 | 3,06E-01 | 5,00E-01 |
| COQ3 | 71,70 | 0,03 | 0,03 | 1,02 | 3,06E-01 | 5,00E-01 |

| | | | | | | |
|---|---|---|---|---|---|---|
| DUSP11 | 1206,04 | -0,02 | 0,02 | -1,02 | 3,06E-01 | 5,00E-01 |
| SPTA1 | 461,25 | 0,07 | 0,07 | 1,02 | 3,06E-01 | 5,00E-01 |
| TMCO4 | 458,10 | 0,03 | 0,03 | 1,02 | 3,06E-01 | 5,00E-01 |
| CIAO1 | 1838,62 | -0,01 | 0,01 | -1,02 | 3,06E-01 | 5,00E-01 |
| DGKH | 1283,06 | 0,03 | 0,03 | 1,02 | 3,06E-01 | 5,00E-01 |
| CTCF | 1172,98 | -0,02 | 0,02 | -1,02 | 3,06E-01 | 5,00E-01 |
| RPA2 | 1360,30 | -0,03 | 0,03 | -1,02 | 3,06E-01 | 5,00E-01 |
| DDOST | 2252,59 | 0,02 | 0,02 | 1,02 | 3,06E-01 | 5,00E-01 |
| ZFYVE9 | 260,36 | 0,04 | 0,04 | 1,02 | 3,06E-01 | 5,00E-01 |
| ABCF3 | 834,02 | -0,02 | 0,02 | -1,02 | 3,06E-01 | 5,00E-01 |
| CLUH | 249,22 | 0,05 | 0,05 | 1,02 | 3,06E-01 | 5,00E-01 |
| ST3GAL2 | 2458,40 | -0,04 | 0,04 | -1,02 | 3,06E-01 | 5,00E-01 |
| AGAP1 | 365,14 | -0,07 | 0,07 | -1,02 | 3,07E-01 | 5,00E-01 |
| PARVB | 226,71 | 0,06 | 0,05 | 1,02 | 3,07E-01 | 5,00E-01 |
| DNM3 | 471,81 | 0,04 | 0,04 | 1,02 | 3,07E-01 | 5,01E-01 |
| ITM2C | 196,32 | 0,04 | 0,04 | 1,02 | 3,07E-01 | 5,01E-01 |
| CPSF2 | 2771,80 | -0,02 | 0,02 | -1,02 | 3,07E-01 | 5,01E-01 |
| MAGEE1 | 81,12 | 0,04 | 0,04 | 1,02 | 3,07E-01 | 5,01E-01 |
| LIX1L | 1169,26 | -0,02 | 0,02 | -1,02 | 3,07E-01 | 5,01E-01 |
| SORT1 | 1284,32 | -0,04 | 0,04 | -1,02 | 3,07E-01 | 5,01E-01 |
| LETM1 | 788,96 | 0,02 | 0,02 | 1,02 | 3,08E-01 | 5,01E-01 |
| H2BC15 | 346,35 | 0,03 | 0,03 | 1,02 | 3,08E-01 | 5,01E-01 |
| PIGN | 1777,14 | 0,02 | 0,02 | 1,02 | 3,08E-01 | 5,01E-01 |
| ZNF48 | 147,25 | 0,03 | 0,03 | 1,02 | 3,08E-01 | 5,01E-01 |
| ADPRM | 471,15 | 0,04 | 0,04 | 1,02 | 3,08E-01 | 5,02E-01 |
| HSP90AB1 | 2225,88 | 0,03 | 0,02 | 1,02 | 3,08E-01 | 5,02E-01 |
| CCT3 | 2056,29 | 0,02 | 0,02 | 1,02 | 3,09E-01 | 5,02E-01 |
| SEC14L5 | 82,44 | 0,10 | 0,10 | 1,02 | 3,09E-01 | 5,02E-01 |
| RBM15 | 2499,97 | 0,02 | 0,02 | 1,02 | 3,09E-01 | 5,03E-01 |
| BBS12 | 110,49 | 0,04 | 0,04 | 1,02 | 3,09E-01 | 5,03E-01 |
| PRPF38B | 3170,31 | 0,03 | 0,03 | 1,02 | 3,09E-01 | 5,03E-01 |
| NDC80 | 148,92 | -0,04 | 0,04 | -1,02 | 3,09E-01 | 5,03E-01 |
| CD96 | 2908,77 | 0,03 | 0,03 | 1,02 | 3,09E-01 | 5,03E-01 |
| CEP192 | 4128,29 | 0,02 | 0,02 | 1,02 | 3,09E-01 | 5,03E-01 |
| PPARGC1B | 451,14 | 0,04 | 0,04 | 1,02 | 3,10E-01 | 5,04E-01 |
| SLC7A6OS | 1311,84 | -0,01 | 0,01 | -1,02 | 3,10E-01 | 5,04E-01 |
| SLC25A4 | 120,23 | 0,04 | 0,04 | 1,02 | 3,10E-01 | 5,04E-01 |
| PRR12 | 230,91 | 0,09 | 0,09 | 1,02 | 3,10E-01 | 5,04E-01 |
| G3BP1 | 5299,92 | 0,02 | 0,02 | 1,02 | 3,10E-01 | 5,04E-01 |
| TSPYL1 | 2596,95 | 0,02 | 0,02 | 1,02 | 3,10E-01 | 5,04E-01 |
| HLA-F | 6074,83 | -0,03 | 0,03 | -1,02 | 3,10E-01 | 5,04E-01 |
| CLASP2 | 3132,83 | 0,02 | 0,02 | 1,02 | 3,10E-01 | 5,04E-01 |
| SPAST | 2069,27 | -0,03 | 0,03 | -1,01 | 3,10E-01 | 5,04E-01 |
| PMM1 | 161,92 | 0,03 | 0,03 | 1,01 | 3,10E-01 | 5,04E-01 |
| FAM53B | 2425,39 | -0,03 | 0,03 | -1,01 | 3,10E-01 | 5,04E-01 |
| PDE7B | 91,90 | 0,05 | 0,05 | 1,01 | 3,11E-01 | 5,05E-01 |

| | | | | | | |
|---|---|---|---|---|---|---|
| USP46 | 495,04 | 0,03 | 0,03 | 1,01 | 3,11E-01 | 5,05E-01 |
| RPTOR | 843,61 | 0,04 | 0,04 | 1,01 | 3,11E-01 | 5,05E-01 |
| GIMAP6 | 7036,16 | 0,03 | 0,03 | 1,01 | 3,11E-01 | 5,05E-01 |
| ALG11 | 996,30 | 0,02 | 0,02 | 1,01 | 3,11E-01 | 5,05E-01 |
| CHEK1 | 84,09 | 0,06 | 0,06 | 1,01 | 3,11E-01 | 5,05E-01 |
| ZNF662 | 74,94 | 0,06 | 0,06 | 1,01 | 3,12E-01 | 5,06E-01 |
| TSN | 1374,32 | 0,03 | 0,03 | 1,01 | 3,12E-01 | 5,06E-01 |
| SWI5 | 98,49 | 0,02 | 0,02 | 1,01 | 3,12E-01 | 5,06E-01 |
| TAB1 | 347,17 | 0,04 | 0,04 | 1,01 | 3,12E-01 | 5,06E-01 |
| GZF1 | 636,03 | 0,02 | 0,02 | 1,01 | 3,12E-01 | 5,06E-01 |
| STYK1 | 94,33 | 0,07 | 0,07 | 1,01 | 3,12E-01 | 5,06E-01 |
| KYNU | 1847,71 | -0,04 | 0,04 | -1,01 | 3,13E-01 | 5,07E-01 |
| SEPTIN9 | 3337,99 | 0,04 | 0,04 | 1,01 | 3,13E-01 | 5,07E-01 |
| DAP3 | 1459,49 | 0,02 | 0,01 | 1,01 | 3,13E-01 | 5,07E-01 |
| RBBP9 | 318,52 | 0,02 | 0,02 | 1,01 | 3,13E-01 | 5,07E-01 |
| TRMT2B | 1199,91 | 0,02 | 0,02 | 1,01 | 3,13E-01 | 5,07E-01 |
| F2R | 618,21 | -0,04 | 0,04 | -1,01 | 3,13E-01 | 5,07E-01 |
| ADPRHL1 | 76,23 | -0,09 | 0,09 | -1,01 | 3,13E-01 | 5,07E-01 |
| MPHOSPH6 | 446,67 | 0,03 | 0,03 | 1,01 | 3,13E-01 | 5,07E-01 |
| ARL3 | 166,51 | -0,02 | 0,02 | -1,01 | 3,13E-01 | 5,07E-01 |
| UBAP2L | 1673,31 | -0,02 | 0,02 | -1,01 | 3,13E-01 | 5,07E-01 |
| CPAMD8 | 145,02 | 0,07 | 0,07 | 1,01 | 3,14E-01 | 5,07E-01 |
| TPMT | 392,75 | 0,04 | 0,04 | 1,01 | 3,14E-01 | 5,07E-01 |
| TSPAN33 | 378,73 | 0,05 | 0,05 | 1,01 | 3,14E-01 | 5,07E-01 |
| SEPTIN7 | 6955,11 | -0,03 | 0,03 | -1,01 | 3,14E-01 | 5,07E-01 |
| NFS1 | 390,25 | 0,02 | 0,02 | 1,01 | 3,14E-01 | 5,08E-01 |
| NPC2 | 1769,82 | -0,03 | 0,03 | -1,01 | 3,15E-01 | 5,08E-01 |
| CCDC121 | 74,48 | 0,05 | 0,05 | 1,01 | 3,15E-01 | 5,08E-01 |
| AQP3 | 310,59 | 0,04 | 0,04 | 1,01 | 3,15E-01 | 5,08E-01 |
| RAMAC | 507,20 | -0,03 | 0,03 | -1,01 | 3,15E-01 | 5,08E-01 |
| BEND5 | 90,04 | 0,05 | 0,05 | 1,01 | 3,15E-01 | 5,08E-01 |
| YWHAH | 412,37 | 0,03 | 0,03 | 1,01 | 3,15E-01 | 5,08E-01 |
| CPNE5 | 201,49 | 0,05 | 0,05 | 1,01 | 3,15E-01 | 5,08E-01 |
| ZFYVE21 | 420,37 | 0,02 | 0,02 | 1,01 | 3,15E-01 | 5,08E-01 |
| SARM1 | 532,97 | 0,04 | 0,04 | 1,01 | 3,15E-01 | 5,08E-01 |
| VPS54 | 1179,88 | 0,03 | 0,03 | 1,01 | 3,15E-01 | 5,09E-01 |
| PDE6D | 530,22 | -0,01 | 0,01 | -1,00 | 3,15E-01 | 5,09E-01 |
| CCDC71L | 871,25 | -0,03 | 0,03 | -1,00 | 3,15E-01 | 5,09E-01 |
| SNX9 | 615,78 | 0,03 | 0,03 | 1,00 | 3,15E-01 | 5,09E-01 |
| EXOSC10 | 1092,48 | 0,02 | 0,02 | 1,00 | 3,16E-01 | 5,09E-01 |
| AFMID | 103,91 | 0,04 | 0,04 | 1,00 | 3,16E-01 | 5,09E-01 |
| DPH1 | 184,10 | 0,04 | 0,04 | 1,00 | 3,16E-01 | 5,09E-01 |
| USP15 | 23552,98 | -0,04 | 0,04 | -1,00 | 3,16E-01 | 5,09E-01 |
| RETREG3 | 2577,57 | -0,01 | 0,01 | -1,00 | 3,16E-01 | 5,09E-01 |
| CASZ1 | 235,31 | -0,06 | 0,06 | -1,00 | 3,16E-01 | 5,09E-01 |
| PLCH2 | 223,87 | 0,07 | 0,07 | 1,00 | 3,16E-01 | 5,09E-01 |

| | | | | | | |
|---|---|---|---|---|---|---|
| SND1 | 2726,86 | 0,02 | 0,02 | 1,00 | 3,16E-01 | 5,09E-01 |
| GTPBP2 | 913,74 | -0,04 | 0,04 | -1,00 | 3,16E-01 | 5,09E-01 |
| FAM117B | 1192,37 | 0,03 | 0,03 | 1,00 | 3,16E-01 | 5,10E-01 |
| CDC42BPG | 86,79 | 0,07 | 0,07 | 1,00 | 3,16E-01 | 5,10E-01 |
| VWA8 | 1529,78 | 0,02 | 0,02 | 1,00 | 3,16E-01 | 5,10E-01 |
| SCRN3 | 516,27 | 0,02 | 0,02 | 1,00 | 3,16E-01 | 5,10E-01 |
| PI4KA | 2111,51 | -0,04 | 0,04 | -1,00 | 3,17E-01 | 5,10E-01 |
| VPS33A | 727,52 | -0,01 | 0,01 | -1,00 | 3,17E-01 | 5,10E-01 |
| CRY2 | 850,81 | 0,02 | 0,02 | 1,00 | 3,17E-01 | 5,10E-01 |
| KLRD1 | 5332,63 | -0,05 | 0,05 | -1,00 | 3,17E-01 | 5,10E-01 |
| TTC39C | 2011,86 | 0,02 | 0,02 | 1,00 | 3,17E-01 | 5,10E-01 |
| ST3GAL5 | 1609,65 | 0,03 | 0,03 | 1,00 | 3,17E-01 | 5,10E-01 |
| MTRF1L | 550,95 | 0,03 | 0,03 | 1,00 | 3,17E-01 | 5,10E-01 |
| RALGPS2 | 3273,90 | 0,05 | 0,05 | 1,00 | 3,17E-01 | 5,11E-01 |
| PLA2G4A | 317,83 | 0,04 | 0,04 | 1,00 | 3,18E-01 | 5,11E-01 |
| CENPC | 2649,29 | 0,03 | 0,03 | 1,00 | 3,18E-01 | 5,11E-01 |
| COL9A2 | 182,50 | 0,06 | 0,06 | 1,00 | 3,18E-01 | 5,11E-01 |
| POLR3D | 413,75 | 0,03 | 0,03 | 1,00 | 3,18E-01 | 5,12E-01 |
| TMEM126A | 142,40 | 0,05 | 0,05 | 1,00 | 3,18E-01 | 5,12E-01 |
| SATB1 | 4046,01 | 0,03 | 0,03 | 1,00 | 3,18E-01 | 5,12E-01 |
| GHRL | 152,26 | -0,05 | 0,05 | -1,00 | 3,18E-01 | 5,12E-01 |
| ZNF76 | 546,63 | 0,03 | 0,03 | 1,00 | 3,18E-01 | 5,12E-01 |
| GORASP2 | 544,58 | 0,02 | 0,02 | 1,00 | 3,19E-01 | 5,12E-01 |
| DNAJC7 | 2198,96 | -0,01 | 0,01 | -1,00 | 3,19E-01 | 5,12E-01 |
| UBA6 | 2841,88 | 0,03 | 0,03 | 1,00 | 3,19E-01 | 5,12E-01 |
| DENND6A | 2162,84 | 0,02 | 0,02 | 1,00 | 3,19E-01 | 5,12E-01 |
| IVD | 860,02 | 0,03 | 0,03 | 1,00 | 3,19E-01 | 5,12E-01 |
| IFIT5 | 1914,66 | -0,07 | 0,07 | -1,00 | 3,19E-01 | 5,12E-01 |
| TMEM80 | 363,27 | 0,03 | 0,03 | 1,00 | 3,19E-01 | 5,12E-01 |
| NXF1 | 1730,38 | -0,03 | 0,03 | -1,00 | 3,19E-01 | 5,12E-01 |
| GFPT1 | 1143,41 | 0,02 | 0,02 | 1,00 | 3,19E-01 | 5,12E-01 |
| ENSG000002 | 72,31 | -0,06 | 0,07 | -1,00 | 3,19E-01 | 5,12E-01 |
| UBAP1 | 1486,13 | -0,03 | 0,04 | -1,00 | 3,20E-01 | 5,13E-01 |
| THBS1 | 1154,69 | -0,07 | 0,07 | -0,99 | 3,20E-01 | 5,13E-01 |
| XRCC3 | 161,13 | 0,05 | 0,05 | 0,99 | 3,20E-01 | 5,13E-01 |
| MEI1 | 401,83 | 0,03 | 0,03 | 0,99 | 3,20E-01 | 5,13E-01 |
| LAPTM4A | 1448,77 | -0,03 | 0,03 | -0,99 | 3,20E-01 | 5,13E-01 |
| ZNF417 | 1324,38 | -0,02 | 0,02 | -0,99 | 3,21E-01 | 5,14E-01 |
| LTBR | 557,14 | -0,04 | 0,04 | -0,99 | 3,21E-01 | 5,14E-01 |
| HBS1L | 2067,31 | 0,03 | 0,03 | 0,99 | 3,21E-01 | 5,14E-01 |
| TAF1D | 4097,19 | 0,05 | 0,05 | 0,99 | 3,21E-01 | 5,14E-01 |
| DPP8 | 3917,41 | -0,02 | 0,02 | -0,99 | 3,21E-01 | 5,14E-01 |
| GRAMD2B | 151,35 | 0,03 | 0,03 | 0,99 | 3,21E-01 | 5,14E-01 |
| FDFT1 | 2038,32 | -0,02 | 0,02 | -0,99 | 3,21E-01 | 5,14E-01 |
| EIF2B5 | 1210,90 | 0,01 | 0,02 | 0,99 | 3,22E-01 | 5,15E-01 |
| REXO1 | 307,07 | 0,06 | 0,07 | 0,99 | 3,22E-01 | 5,15E-01 |

| | | | | | | |
|---|---|---|---|---|---|---|
| VIPR1 | 428,28 | 0,05 | 0,05 | 0,99 | 3,22E-01 | 5,15E-01 |
| SERINC1 | 8592,10 | -0,04 | 0,04 | -0,99 | 3,22E-01 | 5,15E-01 |
| ZNF654 | 1444,75 | 0,05 | 0,05 | 0,99 | 3,22E-01 | 5,15E-01 |
| MATR3 | 1822,47 | 0,02 | 0,02 | 0,99 | 3,22E-01 | 5,15E-01 |
| GANC | 1720,40 | 0,02 | 0,02 | 0,99 | 3,22E-01 | 5,15E-01 |
| RIC3 | 706,32 | 0,04 | 0,04 | 0,99 | 3,22E-01 | 5,16E-01 |
| ZNF841 | 899,41 | -0,02 | 0,02 | -0,99 | 3,22E-01 | 5,16E-01 |
| TRAPPC9 | 1772,36 | -0,03 | 0,03 | -0,99 | 3,22E-01 | 5,16E-01 |
| MAD1L1 | 1416,54 | 0,02 | 0,02 | 0,99 | 3,23E-01 | 5,16E-01 |
| ZDHHC16 | 500,45 | 0,02 | 0,02 | 0,99 | 3,23E-01 | 5,16E-01 |
| SSX2IP | 573,28 | 0,04 | 0,04 | 0,99 | 3,23E-01 | 5,16E-01 |
| SURF4 | 1978,73 | -0,02 | 0,02 | -0,99 | 3,23E-01 | 5,16E-01 |
| ARL6IP6 | 1043,71 | 0,03 | 0,03 | 0,99 | 3,23E-01 | 5,16E-01 |
| FCRL2 | 830,54 | 0,07 | 0,07 | 0,99 | 3,23E-01 | 5,17E-01 |
| NDUFA9 | 913,40 | 0,02 | 0,02 | 0,99 | 3,24E-01 | 5,18E-01 |
| TSPAN18 | 193,49 | 0,05 | 0,05 | 0,99 | 3,24E-01 | 5,18E-01 |
| TTC31 | 365,83 | 0,03 | 0,03 | 0,99 | 3,24E-01 | 5,18E-01 |
| BCCIP | 622,38 | 0,02 | 0,02 | 0,99 | 3,24E-01 | 5,18E-01 |
| MMRN1 | 366,87 | 0,07 | 0,07 | 0,99 | 3,24E-01 | 5,18E-01 |
| ACAT2 | 397,19 | 0,03 | 0,03 | 0,98 | 3,25E-01 | 5,19E-01 |
| TRAPPC2 | 1066,00 | 0,02 | 0,02 | 0,98 | 3,25E-01 | 5,19E-01 |
| ZNG1C | 593,26 | -0,03 | 0,03 | -0,98 | 3,25E-01 | 5,19E-01 |
| ATP5PD | 927,54 | 0,03 | 0,03 | 0,98 | 3,25E-01 | 5,19E-01 |
| DOLPP1 | 162,59 | 0,02 | 0,02 | 0,98 | 3,25E-01 | 5,19E-01 |
| SAMSN1 | 1334,33 | -0,03 | 0,04 | -0,98 | 3,25E-01 | 5,19E-01 |
| CRBN | 3352,34 | 0,03 | 0,03 | 0,98 | 3,26E-01 | 5,20E-01 |
| IPO9 | 1685,76 | 0,02 | 0,02 | 0,98 | 3,26E-01 | 5,20E-01 |
| C14orf93 | 305,30 | -0,02 | 0,02 | -0,98 | 3,26E-01 | 5,20E-01 |
| ANKRD28 | 1943,35 | 0,03 | 0,03 | 0,98 | 3,26E-01 | 5,20E-01 |
| RRAGA | 542,88 | 0,02 | 0,02 | 0,98 | 3,26E-01 | 5,20E-01 |
| ELAPOR2 | 388,22 | -0,06 | 0,06 | -0,98 | 3,26E-01 | 5,20E-01 |
| RPAP1 | 274,27 | 0,04 | 0,04 | 0,98 | 3,26E-01 | 5,20E-01 |
| TXN | 747,05 | -0,06 | 0,06 | -0,98 | 3,26E-01 | 5,20E-01 |
| ATP1B1 | 96,68 | 0,05 | 0,05 | 0,98 | 3,27E-01 | 5,21E-01 |
| PIGS | 558,87 | -0,02 | 0,02 | -0,98 | 3,27E-01 | 5,21E-01 |
| TRABD2A | 2498,39 | 0,04 | 0,04 | 0,98 | 3,27E-01 | 5,21E-01 |
| ALOX15 | 293,45 | 0,10 | 0,10 | 0,98 | 3,27E-01 | 5,21E-01 |
| SNRPE | 349,20 | 0,05 | 0,05 | 0,98 | 3,27E-01 | 5,21E-01 |
| DDX19B | 221,98 | -0,02 | 0,02 | -0,98 | 3,27E-01 | 5,21E-01 |
| NECAP2 | 1925,40 | 0,02 | 0,02 | 0,98 | 3,27E-01 | 5,21E-01 |
| EIF4E | 1415,09 | 0,03 | 0,03 | 0,98 | 3,27E-01 | 5,21E-01 |
| SUGT1 | 2375,54 | 0,02 | 0,02 | 0,98 | 3,28E-01 | 5,21E-01 |
| SKA2 | 269,38 | 0,04 | 0,04 | 0,98 | 3,28E-01 | 5,21E-01 |
| H2AC4 | 188,31 | 0,07 | 0,07 | 0,98 | 3,28E-01 | 5,21E-01 |
| PGPEP1 | 780,99 | 0,03 | 0,03 | 0,98 | 3,28E-01 | 5,21E-01 |
| BCS1L | 232,00 | 0,02 | 0,02 | 0,98 | 3,28E-01 | 5,21E-01 |

| | | | | | | |
|---|---|---|---|---|---|---|
| GEMIN7 | 242,74 | 0,02 | 0,02 | 0,98 | 3,28E-01 | 5,22E-01 |
| TTLL4 | 468,70 | -0,04 | 0,04 | -0,98 | 3,28E-01 | 5,22E-01 |
| H2BC21 | 2977,67 | -0,03 | 0,03 | -0,98 | 3,28E-01 | 5,22E-01 |
| TMEM234 | 353,65 | -0,03 | 0,03 | -0,98 | 3,28E-01 | 5,22E-01 |
| SPATA2L | 140,67 | -0,04 | 0,04 | -0,98 | 3,28E-01 | 5,22E-01 |
| NUDT13 | 87,07 | 0,03 | 0,03 | 0,98 | 3,28E-01 | 5,22E-01 |
| C11orf24 | 168,92 | 0,03 | 0,03 | 0,98 | 3,29E-01 | 5,23E-01 |
| MTHFR | 885,12 | 0,04 | 0,04 | 0,98 | 3,29E-01 | 5,23E-01 |
| ANKRD18A | 128,43 | 0,05 | 0,05 | 0,98 | 3,29E-01 | 5,23E-01 |
| ZNF664 | 1205,89 | 0,02 | 0,02 | 0,98 | 3,29E-01 | 5,23E-01 |
| GATAD1 | 1165,47 | 0,01 | 0,02 | 0,98 | 3,29E-01 | 5,23E-01 |
| SLC46A1 | 155,94 | 0,04 | 0,04 | 0,98 | 3,29E-01 | 5,23E-01 |
| COLQ | 236,61 | -0,05 | 0,05 | -0,98 | 3,30E-01 | 5,23E-01 |
| ARMCX6 | 310,48 | 0,03 | 0,03 | 0,97 | 3,30E-01 | 5,23E-01 |
| SRSF5 | 11272,69 | -0,02 | 0,02 | -0,97 | 3,30E-01 | 5,23E-01 |
| RAB40C | 330,57 | -0,03 | 0,03 | -0,97 | 3,30E-01 | 5,24E-01 |
| GSTM2 | 163,52 | 0,06 | 0,06 | 0,97 | 3,30E-01 | 5,24E-01 |
| EPS8 | 134,46 | 0,06 | 0,06 | 0,97 | 3,30E-01 | 5,24E-01 |
| RWDD4 | 323,25 | 0,03 | 0,03 | 0,97 | 3,30E-01 | 5,24E-01 |
| CD19 | 170,87 | 0,06 | 0,06 | 0,97 | 3,30E-01 | 5,24E-01 |
| ADGRF3 | 81,34 | 0,05 | 0,05 | 0,97 | 3,30E-01 | 5,24E-01 |
| NBPF11 | 717,32 | 0,03 | 0,03 | 0,97 | 3,30E-01 | 5,24E-01 |
| SNX6 | 2452,38 | -0,03 | 0,03 | -0,97 | 3,30E-01 | 5,24E-01 |
| FLACC1 | 342,75 | 0,04 | 0,05 | 0,97 | 3,31E-01 | 5,24E-01 |
| ALYREF | 648,60 | 0,02 | 0,02 | 0,97 | 3,31E-01 | 5,24E-01 |
| GPN1 | 432,40 | 0,02 | 0,02 | 0,97 | 3,31E-01 | 5,25E-01 |
| GZMA | 882,12 | 0,07 | 0,07 | 0,97 | 3,31E-01 | 5,25E-01 |
| EMC7 | 486,02 | -0,02 | 0,02 | -0,97 | 3,31E-01 | 5,25E-01 |
| ANKEF1 | 209,93 | 0,03 | 0,03 | 0,97 | 3,31E-01 | 5,25E-01 |
| ANKDD1A | 223,85 | -0,06 | 0,07 | -0,97 | 3,32E-01 | 5,25E-01 |
| PIPOX | 218,19 | -0,02 | 0,02 | -0,97 | 3,32E-01 | 5,26E-01 |
| WDR86 | 74,33 | 0,07 | 0,07 | 0,97 | 3,32E-01 | 5,26E-01 |
| STXBP1 | 99,25 | 0,06 | 0,07 | 0,97 | 3,32E-01 | 5,26E-01 |
| EHMT1 | 2682,19 | -0,03 | 0,03 | -0,97 | 3,32E-01 | 5,26E-01 |
| TMEM169 | 128,12 | -0,02 | 0,02 | -0,97 | 3,32E-01 | 5,26E-01 |
| TRIM33 | 5482,72 | -0,02 | 0,02 | -0,97 | 3,32E-01 | 5,26E-01 |
| ZNHIT3 | 606,89 | 0,02 | 0,02 | 0,97 | 3,33E-01 | 5,26E-01 |
| ZNF616 | 297,50 | 0,03 | 0,03 | 0,97 | 3,33E-01 | 5,26E-01 |
| ARL8B | 1629,61 | 0,02 | 0,02 | 0,97 | 3,33E-01 | 5,26E-01 |
| DHRS4 | 321,77 | 0,02 | 0,02 | 0,97 | 3,33E-01 | 5,26E-01 |
| NEURL1 | 181,90 | 0,05 | 0,05 | 0,97 | 3,33E-01 | 5,26E-01 |
| CDYL2 | 355,62 | 0,03 | 0,03 | 0,97 | 3,33E-01 | 5,26E-01 |
| AMER1 | 261,26 | 0,03 | 0,03 | 0,97 | 3,33E-01 | 5,26E-01 |
| AUTS2 | 613,73 | -0,05 | 0,05 | -0,97 | 3,33E-01 | 5,26E-01 |
| SH3BP1 | 245,55 | 0,04 | 0,04 | 0,97 | 3,33E-01 | 5,26E-01 |
| PLEKHA8 | 801,43 | 0,02 | 0,02 | 0,97 | 3,33E-01 | 5,26E-01 |

| | | | | | | |
|---|---|---|---|---|---|---|
| TLK1 | 2368,97 | 0,03 | 0,03 | 0,97 | 3,33E-01 | 5,27E-01 |
| BPI | 119,50 | 0,09 | 0,09 | 0,97 | 3,33E-01 | 5,27E-01 |
| UTP23 | 827,47 | 0,03 | 0,03 | 0,97 | 3,34E-01 | 5,27E-01 |
| MAN1C1 | 372,71 | 0,05 | 0,05 | 0,97 | 3,34E-01 | 5,27E-01 |
| CLU | 374,22 | 0,07 | 0,08 | 0,97 | 3,34E-01 | 5,27E-01 |
| SHISAL2A | 276,67 | 0,05 | 0,05 | 0,97 | 3,34E-01 | 5,27E-01 |
| ZNF852 | 603,56 | -0,02 | 0,02 | -0,97 | 3,34E-01 | 5,27E-01 |
| GTF3C1 | 1357,99 | -0,03 | 0,03 | -0,97 | 3,34E-01 | 5,27E-01 |
| MED11 | 454,58 | -0,02 | 0,02 | -0,97 | 3,34E-01 | 5,27E-01 |
| THRA | 378,59 | 0,03 | 0,04 | 0,97 | 3,34E-01 | 5,28E-01 |
| NELFE | 451,22 | 0,02 | 0,02 | 0,97 | 3,34E-01 | 5,28E-01 |
| NAA50 | 3991,37 | -0,03 | 0,03 | -0,97 | 3,35E-01 | 5,28E-01 |
| PLPP6 | 362,57 | 0,03 | 0,03 | 0,96 | 3,35E-01 | 5,28E-01 |
| NEDD1 | 1032,00 | 0,03 | 0,03 | 0,96 | 3,35E-01 | 5,28E-01 |
| GOSR2 | 2092,67 | 0,02 | 0,02 | 0,96 | 3,35E-01 | 5,28E-01 |
| FAM98A | 523,62 | 0,02 | 0,02 | 0,96 | 3,35E-01 | 5,28E-01 |
| CARNS1 | 83,91 | 0,07 | 0,07 | 0,96 | 3,35E-01 | 5,28E-01 |
| OTUD1 | 725,97 | 0,03 | 0,03 | 0,96 | 3,35E-01 | 5,29E-01 |
| TRIM26 | 829,91 | -0,03 | 0,03 | -0,96 | 3,35E-01 | 5,29E-01 |
| ZBTB40 | 2048,39 | 0,02 | 0,02 | 0,96 | 3,35E-01 | 5,29E-01 |
| SMPD3 | 255,51 | 0,08 | 0,08 | 0,96 | 3,36E-01 | 5,29E-01 |
| OVGP1 | 77,41 | 0,04 | 0,05 | 0,96 | 3,36E-01 | 5,29E-01 |
| WASHC1 | 150,56 | 0,06 | 0,06 | 0,96 | 3,36E-01 | 5,29E-01 |
| ZSCAN9 | 400,44 | -0,02 | 0,02 | -0,96 | 3,36E-01 | 5,29E-01 |
| TRPT1 | 99,04 | 0,03 | 0,03 | 0,96 | 3,36E-01 | 5,29E-01 |
| RPAP3 | 1414,85 | -0,03 | 0,03 | -0,96 | 3,36E-01 | 5,29E-01 |
| NOL10 | 722,12 | 0,02 | 0,02 | 0,96 | 3,36E-01 | 5,29E-01 |
| ZFP1 | 213,55 | 0,03 | 0,03 | 0,96 | 3,36E-01 | 5,29E-01 |
| HUS1 | 898,46 | 0,02 | 0,02 | 0,96 | 3,36E-01 | 5,29E-01 |
| WDR83 | 190,33 | 0,02 | 0,02 | 0,96 | 3,36E-01 | 5,29E-01 |
| PPP1R16A | 101,36 | 0,05 | 0,05 | 0,96 | 3,37E-01 | 5,30E-01 |
| DCAF6 | 1951,43 | 0,02 | 0,02 | 0,96 | 3,37E-01 | 5,30E-01 |
| PRAM1 | 543,95 | -0,07 | 0,07 | -0,96 | 3,37E-01 | 5,30E-01 |
| PUM1 | 3072,13 | -0,02 | 0,02 | -0,96 | 3,37E-01 | 5,30E-01 |
| SDHA | 2202,89 | -0,01 | 0,02 | -0,96 | 3,37E-01 | 5,30E-01 |
| ZMYND8 | 2148,34 | -0,02 | 0,02 | -0,96 | 3,38E-01 | 5,31E-01 |
| TCAF1 | 248,02 | 0,02 | 0,02 | 0,96 | 3,38E-01 | 5,31E-01 |
| TMF1 | 3842,95 | -0,03 | 0,03 | -0,96 | 3,38E-01 | 5,31E-01 |
| SMARCB1 | 502,88 | 0,04 | 0,05 | 0,96 | 3,38E-01 | 5,31E-01 |
| CCND3 | 3130,64 | -0,03 | 0,03 | -0,96 | 3,38E-01 | 5,31E-01 |
| ENDOD1 | 1377,80 | 0,04 | 0,04 | 0,96 | 3,38E-01 | 5,31E-01 |
| PIEZO1 | 1513,02 | 0,06 | 0,06 | 0,96 | 3,38E-01 | 5,31E-01 |
| SSR1 | 4089,37 | -0,02 | 0,02 | -0,96 | 3,38E-01 | 5,32E-01 |
| NDUFA12 | 649,29 | 0,03 | 0,03 | 0,96 | 3,38E-01 | 5,32E-01 |
| TRAPPC6B | 1380,36 | 0,03 | 0,03 | 0,96 | 3,39E-01 | 5,32E-01 |
| GLIPR1 | 8397,84 | -0,03 | 0,04 | -0,96 | 3,39E-01 | 5,32E-01 |

| | | | | | | |
|---|---|---|---|---|---|---|
| GGA3 | 848,37 | -0,04 | 0,04 | -0,96 | 3,39E-01 | 5,32E-01 |
| NPIPB12 | 105,97 | -0,06 | 0,06 | -0,96 | 3,39E-01 | 5,32E-01 |
| PDS5A | 4003,73 | 0,02 | 0,02 | 0,96 | 3,39E-01 | 5,32E-01 |
| PWP1 | 845,04 | 0,02 | 0,03 | 0,96 | 3,39E-01 | 5,32E-01 |
| RTN4 | 2414,63 | -0,02 | 0,03 | -0,96 | 3,39E-01 | 5,32E-01 |
| NEURL4 | 503,40 | 0,02 | 0,02 | 0,96 | 3,39E-01 | 5,33E-01 |
| TMCO1 | 1243,23 | 0,05 | 0,05 | 0,96 | 3,39E-01 | 5,33E-01 |
| ENO2 | 305,60 | 0,03 | 0,04 | 0,95 | 3,40E-01 | 5,33E-01 |
| RBP7 | 158,71 | -0,05 | 0,05 | -0,95 | 3,40E-01 | 5,33E-01 |
| TENT2 | 4500,53 | 0,02 | 0,02 | 0,95 | 3,40E-01 | 5,33E-01 |
| TNIP3 | 84,00 | 0,07 | 0,07 | 0,95 | 3,40E-01 | 5,33E-01 |
| ATP5PF | 202,92 | 0,04 | 0,04 | 0,95 | 3,40E-01 | 5,33E-01 |
| ATL3 | 2136,07 | 0,02 | 0,02 | 0,95 | 3,40E-01 | 5,33E-01 |
| TRAM2 | 512,79 | -0,03 | 0,03 | -0,95 | 3,40E-01 | 5,33E-01 |
| FMNL2 | 185,29 | -0,05 | 0,05 | -0,95 | 3,40E-01 | 5,33E-01 |
| VPS11 | 928,80 | -0,02 | 0,02 | -0,95 | 3,41E-01 | 5,34E-01 |
| PRKCH | 6656,78 | -0,02 | 0,03 | -0,95 | 3,41E-01 | 5,34E-01 |
| SCAMP4 | 105,21 | 0,07 | 0,07 | 0,95 | 3,41E-01 | 5,34E-01 |
| SNAPC5 | 443,73 | 0,02 | 0,02 | 0,95 | 3,41E-01 | 5,35E-01 |
| TBC1D9B | 831,89 | 0,05 | 0,05 | 0,95 | 3,41E-01 | 5,35E-01 |
| GRK2 | 4878,25 | -0,04 | 0,04 | -0,95 | 3,42E-01 | 5,35E-01 |
| SERBP1 | 5838,37 | 0,02 | 0,02 | 0,95 | 3,42E-01 | 5,36E-01 |
| EFNA4 | 113,15 | -0,02 | 0,02 | -0,95 | 3,42E-01 | 5,36E-01 |
| PIBF1 | 1107,23 | 0,04 | 0,04 | 0,95 | 3,42E-01 | 5,36E-01 |
| PRDX5 | 3891,17 | 0,05 | 0,05 | 0,95 | 3,43E-01 | 5,36E-01 |
| POLM | 614,49 | 0,03 | 0,03 | 0,95 | 3,43E-01 | 5,36E-01 |
| COL13A1 | 85,12 | -0,10 | 0,11 | -0,95 | 3,43E-01 | 5,36E-01 |
| ASRGL1 | 259,07 | 0,03 | 0,03 | 0,95 | 3,43E-01 | 5,36E-01 |
| KCNC4 | 246,44 | 0,07 | 0,07 | 0,95 | 3,43E-01 | 5,36E-01 |
| CCNT2 | 4397,07 | 0,03 | 0,03 | 0,95 | 3,43E-01 | 5,36E-01 |
| ABR | 1771,66 | -0,05 | 0,05 | -0,95 | 3,43E-01 | 5,36E-01 |
| ZNF701 | 1067,03 | -0,03 | 0,03 | -0,95 | 3,43E-01 | 5,36E-01 |
| YBX1 | 12652,76 | 0,05 | 0,05 | 0,95 | 3,44E-01 | 5,37E-01 |
| MDM1 | 955,35 | 0,03 | 0,03 | 0,95 | 3,44E-01 | 5,37E-01 |
| ATP11B | 8613,24 | -0,03 | 0,03 | -0,95 | 3,44E-01 | 5,37E-01 |
| CLTA | 1112,32 | -0,02 | 0,02 | -0,95 | 3,44E-01 | 5,37E-01 |
| VCPIP1 | 7514,14 | -0,03 | 0,03 | -0,95 | 3,44E-01 | 5,37E-01 |
| COMMD2 | 934,65 | 0,03 | 0,03 | 0,95 | 3,44E-01 | 5,37E-01 |
| TNFRSF4 | 114,32 | 0,05 | 0,05 | 0,95 | 3,44E-01 | 5,38E-01 |
| ESRRA | 1533,72 | -0,02 | 0,02 | -0,95 | 3,44E-01 | 5,38E-01 |
| EXOSC3 | 855,58 | 0,02 | 0,02 | 0,95 | 3,45E-01 | 5,38E-01 |
| ATG2B | 3516,17 | -0,02 | 0,02 | -0,95 | 3,45E-01 | 5,38E-01 |
| ZNF329 | 371,54 | 0,03 | 0,03 | 0,94 | 3,45E-01 | 5,38E-01 |
| STRIP1 | 1085,03 | -0,03 | 0,03 | -0,94 | 3,45E-01 | 5,38E-01 |
| SLC66A2 | 1515,95 | 0,04 | 0,04 | 0,94 | 3,45E-01 | 5,38E-01 |
| PLEKHG1 | 476,39 | 0,05 | 0,06 | 0,94 | 3,45E-01 | 5,38E-01 |

| | | | | | | |
|---|---|---|---|---|---|---|
| SNRPD3 | 968,75 | 0,02 | 0,02 | 0,94 | 3,45E-01 | 5,39E-01 |
| ECHDC2 | 894,04 | 0,04 | 0,04 | 0,94 | 3,45E-01 | 5,39E-01 |
| MCPH1 | 1110,81 | 0,02 | 0,02 | 0,94 | 3,46E-01 | 5,39E-01 |
| ATAD3A | 152,23 | 0,03 | 0,03 | 0,94 | 3,46E-01 | 5,39E-01 |
| VBP1 | 675,69 | 0,05 | 0,06 | 0,94 | 3,46E-01 | 5,39E-01 |
| GTF2H5 | 235,73 | 0,04 | 0,04 | 0,94 | 3,46E-01 | 5,39E-01 |
| GNAI1 | 88,81 | 0,08 | 0,08 | 0,94 | 3,46E-01 | 5,39E-01 |
| IQCN | 258,36 | -0,06 | 0,07 | -0,94 | 3,46E-01 | 5,39E-01 |
| PSMA7 | 2078,69 | 0,02 | 0,02 | 0,94 | 3,46E-01 | 5,39E-01 |
| CCSER2 | 2918,50 | 0,03 | 0,03 | 0,94 | 3,46E-01 | 5,39E-01 |
| AXIN2 | 233,65 | 0,04 | 0,04 | 0,94 | 3,46E-01 | 5,39E-01 |
| SEC24B | 1782,54 | -0,01 | 0,01 | -0,94 | 3,46E-01 | 5,39E-01 |
| ITGA2B | 683,85 | 0,08 | 0,09 | 0,94 | 3,47E-01 | 5,40E-01 |
| RIF1 | 5011,44 | 0,03 | 0,03 | 0,94 | 3,47E-01 | 5,40E-01 |
| MYBPC3 | 205,97 | -0,06 | 0,06 | -0,94 | 3,47E-01 | 5,40E-01 |
| MDC1 | 412,11 | 0,03 | 0,03 | 0,94 | 3,47E-01 | 5,40E-01 |
| ADD1 | 4428,49 | -0,03 | 0,04 | -0,94 | 3,47E-01 | 5,40E-01 |
| MNAT1 | 408,16 | 0,03 | 0,03 | 0,94 | 3,48E-01 | 5,41E-01 |
| PLD2 | 192,97 | 0,05 | 0,05 | 0,94 | 3,48E-01 | 5,41E-01 |
| CD44 | 9824,38 | 0,02 | 0,02 | 0,94 | 3,48E-01 | 5,41E-01 |
| BSCL2 | 105,05 | 0,02 | 0,02 | 0,94 | 3,48E-01 | 5,41E-01 |
| VCL | 4795,17 | -0,03 | 0,04 | -0,94 | 3,48E-01 | 5,41E-01 |
| MAP3K7 | 2399,98 | 0,02 | 0,03 | 0,94 | 3,48E-01 | 5,42E-01 |
| HAUS3 | 1018,24 | 0,03 | 0,03 | 0,94 | 3,48E-01 | 5,42E-01 |
| PTPN2 | 2096,43 | 0,02 | 0,02 | 0,94 | 3,48E-01 | 5,42E-01 |
| FBXL3 | 4044,76 | -0,03 | 0,03 | -0,94 | 3,48E-01 | 5,42E-01 |
| ABCB8 | 292,96 | 0,04 | 0,04 | 0,94 | 3,48E-01 | 5,42E-01 |
| HECTD4 | 3693,71 | 0,03 | 0,03 | 0,94 | 3,48E-01 | 5,42E-01 |
| S1PR4 | 2526,64 | -0,03 | 0,03 | -0,94 | 3,49E-01 | 5,42E-01 |
| GEN1 | 616,00 | 0,04 | 0,04 | 0,94 | 3,49E-01 | 5,42E-01 |
| PARD6B | 215,41 | 0,02 | 0,03 | 0,94 | 3,49E-01 | 5,42E-01 |
| ZBTB7B | 2109,46 | -0,04 | 0,04 | -0,94 | 3,49E-01 | 5,42E-01 |
| DNAJC14 | 798,84 | -0,02 | 0,02 | -0,94 | 3,49E-01 | 5,43E-01 |
| PIAS4 | 470,31 | -0,03 | 0,03 | -0,93 | 3,50E-01 | 5,43E-01 |
| VAC14 | 638,31 | 0,03 | 0,04 | 0,93 | 3,50E-01 | 5,43E-01 |
| HINFP | 438,39 | 0,03 | 0,03 | 0,93 | 3,50E-01 | 5,43E-01 |
| USP25 | 5410,24 | -0,03 | 0,03 | -0,93 | 3,50E-01 | 5,43E-01 |
| TAS2R13 | 137,70 | 0,05 | 0,05 | 0,93 | 3,50E-01 | 5,43E-01 |
| PTRH1 | 138,18 | 0,03 | 0,03 | 0,93 | 3,50E-01 | 5,43E-01 |
| SIK2 | 677,32 | -0,02 | 0,02 | -0,93 | 3,50E-01 | 5,43E-01 |
| PIP5K1B | 525,31 | 0,03 | 0,03 | 0,93 | 3,50E-01 | 5,43E-01 |
| POLR2A | 1492,92 | -0,06 | 0,06 | -0,93 | 3,50E-01 | 5,43E-01 |
| MTHFD1L | 130,94 | 0,05 | 0,06 | 0,93 | 3,50E-01 | 5,43E-01 |
| PRR5L | 2496,69 | -0,04 | 0,04 | -0,93 | 3,50E-01 | 5,43E-01 |
| DYNC1H1 | 7381,34 | -0,03 | 0,03 | -0,93 | 3,50E-01 | 5,43E-01 |
| ATRX | 6665,47 | -0,03 | 0,03 | -0,93 | 3,50E-01 | 5,43E-01 |

| | | | | | | |
|---|---|---|---|---|---|---|
| UQCC1 | 698,84 | 0,02 | 0,02 | 0,93 | 3,51E-01 | 5,44E-01 |
| S100A8 | 10433,77 | -0,08 | 0,09 | -0,93 | 3,51E-01 | 5,44E-01 |
| UVRAG | 2250,03 | -0,02 | 0,02 | -0,93 | 3,51E-01 | 5,44E-01 |
| STXBP2 | 758,93 | -0,05 | 0,05 | -0,93 | 3,51E-01 | 5,44E-01 |
| CBX4 | 489,71 | -0,03 | 0,03 | -0,93 | 3,51E-01 | 5,44E-01 |
| CDH23 | 2078,97 | -0,03 | 0,04 | -0,93 | 3,51E-01 | 5,44E-01 |
| ZNF684 | 148,76 | -0,03 | 0,04 | -0,93 | 3,51E-01 | 5,44E-01 |
| ZNF620 | 115,07 | 0,02 | 0,03 | 0,93 | 3,52E-01 | 5,44E-01 |
| CCND2 | 3324,51 | -0,02 | 0,03 | -0,93 | 3,52E-01 | 5,45E-01 |
| GOT2 | 391,93 | -0,02 | 0,02 | -0,93 | 3,52E-01 | 5,45E-01 |
| KLHL11 | 429,91 | 0,02 | 0,02 | 0,93 | 3,52E-01 | 5,45E-01 |
| HDLBP | 4727,87 | -0,03 | 0,03 | -0,93 | 3,52E-01 | 5,45E-01 |
| KIF23 | 95,05 | 0,05 | 0,06 | 0,93 | 3,52E-01 | 5,45E-01 |
| B4GALT6 | 114,67 | 0,04 | 0,05 | 0,93 | 3,52E-01 | 5,45E-01 |
| MAGED2 | 1109,69 | 0,02 | 0,02 | 0,93 | 3,52E-01 | 5,45E-01 |
| CENATAC | 1106,47 | 0,02 | 0,02 | 0,93 | 3,52E-01 | 5,45E-01 |
| IKZF2 | 2889,17 | 0,04 | 0,05 | 0,93 | 3,53E-01 | 5,45E-01 |
| TET1 | 173,17 | 0,03 | 0,03 | 0,93 | 3,53E-01 | 5,45E-01 |
| AHCYL2 | 717,97 | -0,02 | 0,02 | -0,93 | 3,53E-01 | 5,46E-01 |
| KLHDC10 | 1552,53 | 0,01 | 0,01 | 0,93 | 3,53E-01 | 5,46E-01 |
| AAGAB | 1080,83 | -0,02 | 0,02 | -0,93 | 3,53E-01 | 5,46E-01 |
| TMED2 | 2504,04 | 0,03 | 0,03 | 0,93 | 3,54E-01 | 5,47E-01 |
| MAP7 | 138,05 | -0,04 | 0,05 | -0,93 | 3,54E-01 | 5,47E-01 |
| KMT5A | 335,84 | -0,03 | 0,03 | -0,93 | 3,54E-01 | 5,47E-01 |
| LOXHD1 | 387,94 | -0,07 | 0,08 | -0,93 | 3,54E-01 | 5,47E-01 |
| CYBC1 | 2880,77 | -0,04 | 0,05 | -0,93 | 3,55E-01 | 5,48E-01 |
| LRRC8B | 1216,13 | 0,03 | 0,03 | 0,93 | 3,55E-01 | 5,48E-01 |
| CDK5RAP2 | 2391,98 | -0,02 | 0,02 | -0,93 | 3,55E-01 | 5,48E-01 |
| ACADS | 313,46 | 0,02 | 0,02 | 0,93 | 3,55E-01 | 5,48E-01 |
| CDKAL1 | 1241,25 | 0,02 | 0,02 | 0,93 | 3,55E-01 | 5,48E-01 |
| DOK1 | 1037,67 | -0,02 | 0,02 | -0,93 | 3,55E-01 | 5,48E-01 |
| MICB | 795,31 | -0,03 | 0,03 | -0,93 | 3,55E-01 | 5,48E-01 |
| DDX6 | 18493,31 | -0,02 | 0,02 | -0,92 | 3,55E-01 | 5,48E-01 |
| DNM2 | 4280,22 | -0,03 | 0,04 | -0,92 | 3,55E-01 | 5,48E-01 |
| D2HGDH | 431,36 | 0,04 | 0,04 | 0,92 | 3,55E-01 | 5,48E-01 |
| FAM185A | 320,06 | 0,02 | 0,02 | 0,92 | 3,55E-01 | 5,48E-01 |
| ATG4B | 709,31 | -0,03 | 0,03 | -0,92 | 3,56E-01 | 5,48E-01 |
| TRIM32 | 429,91 | 0,02 | 0,02 | 0,92 | 3,56E-01 | 5,48E-01 |
| KPNA6 | 1862,49 | -0,02 | 0,02 | -0,92 | 3,56E-01 | 5,49E-01 |
| RAP1B | 7176,41 | 0,02 | 0,02 | 0,92 | 3,56E-01 | 5,49E-01 |
| WDR97 | 74,87 | 0,07 | 0,07 | 0,92 | 3,56E-01 | 5,49E-01 |
| LRBA | 7811,87 | 0,02 | 0,02 | 0,92 | 3,56E-01 | 5,49E-01 |
| SYNGAP1 | 221,17 | 0,06 | 0,07 | 0,92 | 3,56E-01 | 5,49E-01 |
| AGPAT3 | 1362,43 | 0,03 | 0,04 | 0,92 | 3,56E-01 | 5,49E-01 |
| CHMP7 | 3145,69 | 0,02 | 0,03 | 0,92 | 3,56E-01 | 5,49E-01 |
| SEL1L3 | 2515,94 | 0,03 | 0,03 | 0,92 | 3,56E-01 | 5,49E-01 |

| | | | | | | |
|---|---|---|---|---|---|---|
| EPHB6 | 193,51 | 0,05 | 0,05 | 0,92 | 3,57E-01 | 5,50E-01 |
| NONO | 2132,13 | 0,02 | 0,02 | 0,92 | 3,57E-01 | 5,50E-01 |
| ABCC10 | 909,22 | 0,03 | 0,03 | 0,92 | 3,57E-01 | 5,50E-01 |
| C17orf67 | 231,59 | 0,02 | 0,02 | 0,92 | 3,57E-01 | 5,50E-01 |
| RNF135 | 693,53 | -0,02 | 0,03 | -0,92 | 3,57E-01 | 5,50E-01 |
| GFOD2 | 885,43 | 0,03 | 0,03 | 0,92 | 3,57E-01 | 5,50E-01 |
| ASB3 | 358,79 | -0,02 | 0,02 | -0,92 | 3,57E-01 | 5,50E-01 |
| PLCL1 | 626,06 | 0,03 | 0,04 | 0,92 | 3,58E-01 | 5,50E-01 |
| TEX2 | 590,99 | 0,02 | 0,02 | 0,92 | 3,58E-01 | 5,50E-01 |
| CENPV | 90,54 | 0,03 | 0,04 | 0,92 | 3,58E-01 | 5,51E-01 |
| TJP3 | 84,48 | 0,05 | 0,06 | 0,92 | 3,58E-01 | 5,51E-01 |
| TMEM117 | 154,68 | 0,03 | 0,03 | 0,92 | 3,58E-01 | 5,51E-01 |
| TSSC4 | 1041,81 | 0,02 | 0,02 | 0,92 | 3,58E-01 | 5,51E-01 |
| CATSPERG | 261,12 | 0,04 | 0,04 | 0,92 | 3,59E-01 | 5,51E-01 |
| GMPPA | 189,38 | 0,03 | 0,04 | 0,92 | 3,59E-01 | 5,52E-01 |
| TMEM39A | 1139,90 | 0,02 | 0,02 | 0,92 | 3,59E-01 | 5,52E-01 |
| IRF2BP1 | 431,49 | 0,02 | 0,02 | 0,92 | 3,59E-01 | 5,52E-01 |
| RAP2C | 1495,58 | 0,04 | 0,04 | 0,92 | 3,60E-01 | 5,52E-01 |
| ABTB1 | 4189,89 | -0,05 | 0,06 | -0,92 | 3,60E-01 | 5,53E-01 |
| CEP41 | 229,17 | 0,03 | 0,03 | 0,92 | 3,60E-01 | 5,53E-01 |
| CCZ1B | 605,66 | 0,03 | 0,03 | 0,92 | 3,60E-01 | 5,53E-01 |
| ARHGAP21 | 939,28 | 0,02 | 0,02 | 0,92 | 3,60E-01 | 5,53E-01 |
| DNAJB11 | 731,44 | -0,02 | 0,02 | -0,91 | 3,60E-01 | 5,53E-01 |
| SCAND1 | 468,08 | 0,02 | 0,02 | 0,91 | 3,60E-01 | 5,53E-01 |
| CTPS2 | 297,16 | 0,02 | 0,02 | 0,91 | 3,60E-01 | 5,53E-01 |
| FBRS | 1543,23 | -0,04 | 0,04 | -0,91 | 3,60E-01 | 5,53E-01 |
| RPS23 | 8405,56 | 0,06 | 0,06 | 0,91 | 3,61E-01 | 5,54E-01 |
| THEM4 | 475,29 | 0,03 | 0,04 | 0,91 | 3,61E-01 | 5,54E-01 |
| SSH3 | 478,17 | -0,03 | 0,04 | -0,91 | 3,62E-01 | 5,55E-01 |
| METTL16 | 572,31 | 0,03 | 0,03 | 0,91 | 3,62E-01 | 5,55E-01 |
| KIF16B | 681,06 | -0,03 | 0,03 | -0,91 | 3,62E-01 | 5,55E-01 |
| PNN | 2606,79 | -0,02 | 0,02 | -0,91 | 3,62E-01 | 5,56E-01 |
| SQSTM1 | 5371,61 | -0,02 | 0,02 | -0,91 | 3,63E-01 | 5,56E-01 |
| EXOC4 | 2822,69 | -0,02 | 0,02 | -0,91 | 3,63E-01 | 5,56E-01 |
| LMO7 | 497,04 | 0,03 | 0,03 | 0,91 | 3,63E-01 | 5,56E-01 |
| RPP38 | 342,15 | 0,02 | 0,02 | 0,91 | 3,63E-01 | 5,56E-01 |
| LRRC37A3 | 302,62 | -0,05 | 0,06 | -0,91 | 3,63E-01 | 5,56E-01 |
| ZNF275 | 549,43 | 0,03 | 0,03 | 0,91 | 3,64E-01 | 5,57E-01 |
| FTSJ1 | 217,81 | 0,02 | 0,03 | 0,91 | 3,64E-01 | 5,57E-01 |
| HP1BP3 | 6527,83 | -0,01 | 0,01 | -0,91 | 3,64E-01 | 5,57E-01 |
| NME6 | 1066,02 | -0,01 | 0,01 | -0,91 | 3,64E-01 | 5,58E-01 |
| C2orf76 | 159,41 | 0,03 | 0,03 | 0,91 | 3,64E-01 | 5,58E-01 |
| AGAP9 | 130,14 | 0,06 | 0,07 | 0,91 | 3,64E-01 | 5,58E-01 |
| MRNIP | 485,94 | 0,04 | 0,04 | 0,91 | 3,64E-01 | 5,58E-01 |
| TBC1D8 | 868,59 | -0,03 | 0,04 | -0,91 | 3,65E-01 | 5,58E-01 |
| JHY | 91,83 | 0,04 | 0,04 | 0,91 | 3,65E-01 | 5,58E-01 |

| | | | | | | |
|---|---|---|---|---|---|---|
| CD1C | 298,62 | 0,04 | 0,04 | 0,91 | 3,65E-01 | 5,58E-01 |
| ZNF605 | 555,00 | 0,03 | 0,04 | 0,91 | 3,65E-01 | 5,58E-01 |
| TSHZ2 | 462,58 | 0,05 | 0,05 | 0,91 | 3,65E-01 | 5,58E-01 |
| SLC29A1 | 127,97 | 0,07 | 0,08 | 0,91 | 3,65E-01 | 5,58E-01 |
| TUBB1 | 3029,67 | 0,05 | 0,06 | 0,91 | 3,65E-01 | 5,59E-01 |
| CUZD1 | 137,25 | 0,03 | 0,03 | 0,90 | 3,66E-01 | 5,59E-01 |
| TRIM37 | 945,96 | 0,02 | 0,02 | 0,90 | 3,66E-01 | 5,59E-01 |
| KTN1 | 3048,74 | 0,03 | 0,03 | 0,90 | 3,66E-01 | 5,59E-01 |
| CR2 | 169,04 | 0,05 | 0,06 | 0,90 | 3,66E-01 | 5,59E-01 |
| RAB39A | 125,19 | 0,04 | 0,04 | 0,90 | 3,66E-01 | 5,59E-01 |
| FCSK | 199,87 | 0,05 | 0,06 | 0,90 | 3,66E-01 | 5,59E-01 |
| PCGF3 | 1607,47 | -0,02 | 0,03 | -0,90 | 3,66E-01 | 5,59E-01 |
| CERK | 4137,02 | -0,01 | 0,02 | -0,90 | 3,66E-01 | 5,59E-01 |
| ZNF638 | 8787,21 | -0,02 | 0,03 | -0,90 | 3,66E-01 | 5,59E-01 |
| DDX51 | 169,74 | 0,05 | 0,05 | 0,90 | 3,66E-01 | 5,59E-01 |
| EMC4 | 923,22 | 0,02 | 0,02 | 0,90 | 3,66E-01 | 5,59E-01 |
| NRROS | 186,62 | 0,05 | 0,06 | 0,90 | 3,66E-01 | 5,59E-01 |
| CLK4 | 4064,08 | -0,03 | 0,03 | -0,90 | 3,67E-01 | 5,60E-01 |
| TCP11L1 | 566,21 | -0,02 | 0,03 | -0,90 | 3,67E-01 | 5,60E-01 |
| MRPL49 | 688,87 | 0,01 | 0,01 | 0,90 | 3,67E-01 | 5,60E-01 |
| PATJ | 2580,55 | 0,03 | 0,03 | 0,90 | 3,67E-01 | 5,60E-01 |
| KIF5B | 4634,35 | -0,03 | 0,03 | -0,90 | 3,67E-01 | 5,60E-01 |
| ANXA6 | 7434,88 | 0,02 | 0,03 | 0,90 | 3,67E-01 | 5,60E-01 |
| EGLN2 | 1580,05 | -0,03 | 0,03 | -0,90 | 3,67E-01 | 5,60E-01 |
| GK5 | 1913,99 | -0,03 | 0,03 | -0,90 | 3,67E-01 | 5,60E-01 |
| DGKD | 2571,19 | -0,04 | 0,04 | -0,90 | 3,67E-01 | 5,60E-01 |
| SCO1 | 995,53 | 0,01 | 0,02 | 0,90 | 3,67E-01 | 5,60E-01 |
| DLG5 | 243,24 | -0,06 | 0,07 | -0,90 | 3,68E-01 | 5,60E-01 |
| HLA-DRB5 | 1534,98 | 0,10 | 0,12 | 0,90 | 3,68E-01 | 5,60E-01 |
| DST | 1038,74 | 0,04 | 0,04 | 0,90 | 3,68E-01 | 5,61E-01 |
| ZNF136 | 965,04 | -0,02 | 0,02 | -0,90 | 3,68E-01 | 5,61E-01 |
| PDE8A | 1146,36 | 0,02 | 0,02 | 0,90 | 3,68E-01 | 5,61E-01 |
| PRKACA | 556,00 | -0,05 | 0,06 | -0,90 | 3,68E-01 | 5,61E-01 |
| SPPL2B | 321,11 | 0,05 | 0,05 | 0,90 | 3,68E-01 | 5,61E-01 |
| ZNF146 | 1239,71 | 0,04 | 0,04 | 0,90 | 3,68E-01 | 5,61E-01 |
| CDK2AP2 | 87,89 | 0,05 | 0,05 | 0,90 | 3,68E-01 | 5,61E-01 |
| RBM8A | 2644,57 | -0,02 | 0,02 | -0,90 | 3,69E-01 | 5,61E-01 |
| FBXO3 | 3038,98 | 0,02 | 0,02 | 0,90 | 3,69E-01 | 5,62E-01 |
| MBD5 | 3686,21 | -0,02 | 0,03 | -0,90 | 3,69E-01 | 5,62E-01 |
| HSBP1 | 1216,96 | 0,03 | 0,04 | 0,90 | 3,69E-01 | 5,62E-01 |
| FBXL6 | 152,44 | 0,03 | 0,04 | 0,90 | 3,69E-01 | 5,62E-01 |
| EFL1 | 615,77 | 0,02 | 0,02 | 0,90 | 3,69E-01 | 5,62E-01 |
| UTY | 4529,75 | 0,10 | 0,11 | 0,90 | 3,69E-01 | 5,62E-01 |
| CORO1B | 965,27 | 0,02 | 0,02 | 0,90 | 3,69E-01 | 5,62E-01 |
| TEF | 208,18 | 0,03 | 0,03 | 0,90 | 3,69E-01 | 5,62E-01 |
| AFG3L2 | 1130,50 | 0,01 | 0,02 | 0,90 | 3,69E-01 | 5,62E-01 |

| | | | | | | |
|---|---|---|---|---|---|---|
| TUBG1 | 78,39 | 0,03 | 0,03 | 0,90 | 3,70E-01 | 5,62E-01 |
| ZNF408 | 257,91 | -0,02 | 0,02 | -0,90 | 3,70E-01 | 5,62E-01 |
| TCTA | 575,84 | 0,02 | 0,02 | 0,90 | 3,70E-01 | 5,62E-01 |
| SEC22B | 3638,77 | -0,02 | 0,03 | -0,90 | 3,70E-01 | 5,63E-01 |
| UTP14C | 187,46 | 0,03 | 0,03 | 0,90 | 3,71E-01 | 5,63E-01 |
| KANSL1 | 7097,52 | -0,03 | 0,04 | -0,90 | 3,71E-01 | 5,63E-01 |
| SCAMP1 | 1295,37 | 0,03 | 0,03 | 0,89 | 3,71E-01 | 5,64E-01 |
| TMEM182 | 152,55 | 0,03 | 0,04 | 0,89 | 3,71E-01 | 5,64E-01 |
| PSMB4 | 2468,57 | 0,02 | 0,02 | 0,89 | 3,71E-01 | 5,64E-01 |
| FAM89B | 101,25 | 0,03 | 0,03 | 0,89 | 3,71E-01 | 5,64E-01 |
| RASGRF2 | 1245,02 | 0,03 | 0,03 | 0,89 | 3,72E-01 | 5,64E-01 |
| ABCB4 | 289,15 | 0,05 | 0,06 | 0,89 | 3,72E-01 | 5,64E-01 |
| GPATCH2L | 6125,52 | -0,01 | 0,02 | -0,89 | 3,72E-01 | 5,64E-01 |
| POFUT2 | 414,57 | -0,04 | 0,04 | -0,89 | 3,72E-01 | 5,64E-01 |
| TRMT44 | 574,99 | 0,02 | 0,02 | 0,89 | 3,72E-01 | 5,64E-01 |
| CCDC191 | 468,72 | 0,02 | 0,03 | 0,89 | 3,72E-01 | 5,65E-01 |
| MARK4 | 251,21 | 0,04 | 0,05 | 0,89 | 3,72E-01 | 5,65E-01 |
| SAMM50 | 673,65 | 0,01 | 0,01 | 0,89 | 3,72E-01 | 5,65E-01 |
| CD72 | 296,86 | 0,05 | 0,05 | 0,89 | 3,72E-01 | 5,65E-01 |
| SCAMP3 | 303,98 | 0,02 | 0,02 | 0,89 | 3,73E-01 | 5,65E-01 |
| CA13 | 226,73 | 0,02 | 0,03 | 0,89 | 3,73E-01 | 5,65E-01 |
| METTL13 | 755,41 | 0,01 | 0,02 | 0,89 | 3,73E-01 | 5,66E-01 |
| WRAP73 | 578,98 | -0,02 | 0,03 | -0,89 | 3,73E-01 | 5,66E-01 |
| ZNF655 | 4380,23 | 0,01 | 0,02 | 0,89 | 3,73E-01 | 5,66E-01 |
| TTL | 771,09 | 0,02 | 0,02 | 0,89 | 3,74E-01 | 5,66E-01 |
| LRSAM1 | 468,77 | -0,03 | 0,04 | -0,89 | 3,74E-01 | 5,66E-01 |
| PPP1R37 | 188,22 | 0,04 | 0,05 | 0,89 | 3,74E-01 | 5,66E-01 |
| STARD8 | 124,30 | -0,05 | 0,06 | -0,89 | 3,74E-01 | 5,66E-01 |
| RAPGEF1 | 2321,84 | -0,04 | 0,05 | -0,89 | 3,74E-01 | 5,66E-01 |
| GNL1 | 896,81 | 0,01 | 0,02 | 0,89 | 3,74E-01 | 5,67E-01 |
| MYBL1 | 2435,44 | 0,04 | 0,05 | 0,89 | 3,74E-01 | 5,67E-01 |
| HDGFL3 | 260,62 | 0,05 | 0,06 | 0,89 | 3,74E-01 | 5,67E-01 |
| POLE | 868,18 | 0,03 | 0,03 | 0,89 | 3,74E-01 | 5,67E-01 |
| TDRKH | 212,46 | 0,02 | 0,03 | 0,89 | 3,75E-01 | 5,67E-01 |
| TOP6BL | 259,19 | 0,04 | 0,04 | 0,89 | 3,75E-01 | 5,67E-01 |
| NEXN | 121,83 | -0,05 | 0,05 | -0,89 | 3,75E-01 | 5,68E-01 |
| ZNF17 | 398,82 | 0,03 | 0,04 | 0,89 | 3,75E-01 | 5,68E-01 |
| VAT1 | 156,89 | 0,04 | 0,04 | 0,89 | 3,75E-01 | 5,68E-01 |
| TRIM44 | 2631,66 | 0,02 | 0,02 | 0,89 | 3,76E-01 | 5,68E-01 |
| ITK | 6358,65 | 0,02 | 0,03 | 0,89 | 3,76E-01 | 5,68E-01 |
| NEK10 | 94,63 | 0,06 | 0,06 | 0,89 | 3,76E-01 | 5,68E-01 |
| AHSA1 | 1046,83 | -0,02 | 0,02 | -0,89 | 3,76E-01 | 5,68E-01 |
| TRIR | 1150,34 | 0,02 | 0,02 | 0,89 | 3,76E-01 | 5,68E-01 |
| POMP | 732,30 | -0,03 | 0,03 | -0,89 | 3,76E-01 | 5,68E-01 |
| SLC16A1 | 346,66 | 0,02 | 0,02 | 0,89 | 3,76E-01 | 5,68E-01 |
| PRDM4 | 963,82 | -0,01 | 0,01 | -0,89 | 3,76E-01 | 5,68E-01 |

| | | | | | | |
|---|---|---|---|---|---|---|
| WDR6 | 1345,74 | 0,03 | 0,03 | 0,88 | 3,76E-01 | 5,69E-01 |
| ASPHD2 | 128,60 | -0,03 | 0,04 | -0,88 | 3,76E-01 | 5,69E-01 |
| SETD7 | 1172,35 | 0,02 | 0,02 | 0,88 | 3,76E-01 | 5,69E-01 |
| WDR49 | 244,98 | 0,04 | 0,05 | 0,88 | 3,76E-01 | 5,69E-01 |
| GSDMB | 1224,08 | 0,03 | 0,04 | 0,88 | 3,77E-01 | 5,69E-01 |
| GPAT4 | 2002,96 | -0,01 | 0,01 | -0,88 | 3,77E-01 | 5,69E-01 |
| TRAPPC3 | 1090,58 | -0,01 | 0,01 | -0,88 | 3,78E-01 | 5,70E-01 |
| SERTAD3 | 276,27 | -0,02 | 0,03 | -0,88 | 3,77E-01 | 5,70E-01 |
| ICA1L | 133,91 | 0,04 | 0,05 | 0,88 | 3,78E-01 | 5,70E-01 |
| CAPN1 | 1357,75 | -0,03 | 0,04 | -0,88 | 3,78E-01 | 5,70E-01 |
| PPFIBP1 | 165,10 | 0,04 | 0,05 | 0,88 | 3,78E-01 | 5,71E-01 |
| EVA1B | 165,07 | 0,03 | 0,04 | 0,88 | 3,78E-01 | 5,71E-01 |
| SERPINB8 | 583,31 | -0,02 | 0,02 | -0,88 | 3,78E-01 | 5,71E-01 |
| TATDN3 | 566,54 | -0,02 | 0,02 | -0,88 | 3,79E-01 | 5,71E-01 |
| ATF1 | 702,14 | 0,02 | 0,02 | 0,88 | 3,79E-01 | 5,71E-01 |
| FOXP1 | 8027,48 | -0,02 | 0,02 | -0,88 | 3,79E-01 | 5,71E-01 |
| ZNF41 | 545,84 | 0,01 | 0,01 | 0,88 | 3,79E-01 | 5,71E-01 |
| AGAP3 | 278,13 | 0,05 | 0,06 | 0,88 | 3,79E-01 | 5,71E-01 |
| TFAM | 1542,22 | 0,03 | 0,04 | 0,88 | 3,79E-01 | 5,71E-01 |
| PAN2 | 846,39 | 0,03 | 0,03 | 0,88 | 3,79E-01 | 5,71E-01 |
| SLC27A4 | 216,96 | 0,02 | 0,03 | 0,88 | 3,79E-01 | 5,72E-01 |
| POLA1 | 639,41 | 0,02 | 0,02 | 0,88 | 3,79E-01 | 5,72E-01 |
| HPGD | 169,97 | -0,05 | 0,05 | -0,88 | 3,79E-01 | 5,72E-01 |
| RANGAP1 | 680,13 | 0,03 | 0,03 | 0,88 | 3,80E-01 | 5,72E-01 |
| RRP1 | 320,97 | 0,02 | 0,02 | 0,88 | 3,80E-01 | 5,72E-01 |
| ZNF34 | 162,80 | 0,02 | 0,02 | 0,88 | 3,80E-01 | 5,72E-01 |
| CES2 | 495,73 | 0,02 | 0,02 | 0,88 | 3,80E-01 | 5,73E-01 |
| CYP2U1 | 346,32 | 0,02 | 0,03 | 0,88 | 3,80E-01 | 5,73E-01 |
| CPTP | 76,56 | 0,05 | 0,06 | 0,88 | 3,81E-01 | 5,73E-01 |
| ARMCX2 | 87,55 | 0,04 | 0,04 | 0,88 | 3,81E-01 | 5,73E-01 |
| NR4A2 | 95,29 | 0,03 | 0,04 | 0,88 | 3,81E-01 | 5,73E-01 |
| NDUFB3 | 257,21 | -0,06 | 0,07 | -0,88 | 3,81E-01 | 5,74E-01 |
| TCF4 | 1426,73 | 0,04 | 0,05 | 0,88 | 3,81E-01 | 5,74E-01 |
| TK2 | 961,30 | -0,03 | 0,03 | -0,88 | 3,81E-01 | 5,74E-01 |
| TGFBR1 | 5710,14 | -0,03 | 0,03 | -0,88 | 3,81E-01 | 5,74E-01 |
| ZMAT1 | 1156,33 | 0,02 | 0,03 | 0,88 | 3,81E-01 | 5,74E-01 |
| ZNF773 | 325,85 | 0,02 | 0,02 | 0,87 | 3,82E-01 | 5,74E-01 |
| SPSB2 | 400,17 | 0,03 | 0,03 | 0,87 | 3,82E-01 | 5,74E-01 |
| GSTK1 | 2584,17 | 0,02 | 0,02 | 0,87 | 3,82E-01 | 5,75E-01 |
| FAM204A | 2074,08 | 0,02 | 0,02 | 0,87 | 3,82E-01 | 5,75E-01 |
| TPCN1 | 414,33 | 0,05 | 0,06 | 0,87 | 3,82E-01 | 5,75E-01 |
| PRKAR2B | 2178,91 | 0,05 | 0,06 | 0,87 | 3,83E-01 | 5,75E-01 |
| ENSG000002 | 146,72 | -0,04 | 0,05 | -0,87 | 3,83E-01 | 5,75E-01 |
| LIMS1 | 4393,65 | 0,03 | 0,03 | 0,87 | 3,83E-01 | 5,75E-01 |
| ITPRIPL2 | 782,96 | 0,02 | 0,03 | 0,87 | 3,83E-01 | 5,75E-01 |
| ARHGAP24 | 1086,09 | -0,04 | 0,04 | -0,87 | 3,83E-01 | 5,75E-01 |

| | | | | | | |
|---|---|---|---|---|---|---|
| NCOA7 | 1506,66 | -0,02 | 0,02 | -0,87 | 3,83E-01 | 5,75E-01 |
| GID8 | 1600,22 | 0,02 | 0,02 | 0,87 | 3,83E-01 | 5,75E-01 |
| FKBP2 | 474,59 | 0,02 | 0,02 | 0,87 | 3,83E-01 | 5,75E-01 |
| HDAC3 | 647,63 | -0,02 | 0,02 | -0,87 | 3,84E-01 | 5,76E-01 |
| AGER | 240,29 | -0,05 | 0,06 | -0,87 | 3,84E-01 | 5,76E-01 |
| KCTD10 | 783,80 | -0,02 | 0,03 | -0,87 | 3,84E-01 | 5,76E-01 |
| ADCK2 | 298,70 | 0,02 | 0,02 | 0,87 | 3,84E-01 | 5,76E-01 |
| RANBP17 | 91,01 | 0,08 | 0,09 | 0,87 | 3,84E-01 | 5,76E-01 |
| SASH1 | 240,22 | 0,05 | 0,06 | 0,87 | 3,84E-01 | 5,76E-01 |
| POLA2 | 111,36 | -0,03 | 0,04 | -0,87 | 3,85E-01 | 5,77E-01 |
| HABP4 | 447,12 | 0,02 | 0,03 | 0,87 | 3,85E-01 | 5,77E-01 |
| USPL1 | 1256,63 | 0,02 | 0,02 | 0,87 | 3,85E-01 | 5,77E-01 |
| WBP2 | 1157,57 | 0,04 | 0,05 | 0,87 | 3,84E-01 | 5,77E-01 |
| PSMB6 | 660,61 | 0,02 | 0,03 | 0,87 | 3,85E-01 | 5,77E-01 |
| UBE2D3 | 16643,65 | -0,02 | 0,03 | -0,87 | 3,85E-01 | 5,77E-01 |
| UTP4 | 308,88 | 0,02 | 0,02 | 0,87 | 3,85E-01 | 5,77E-01 |
| MAP3K20 | 950,15 | -0,03 | 0,03 | -0,87 | 3,85E-01 | 5,77E-01 |
| HIVEP3 | 353,83 | 0,05 | 0,06 | 0,87 | 3,85E-01 | 5,77E-01 |
| CREB3L2 | 870,27 | -0,03 | 0,03 | -0,87 | 3,85E-01 | 5,77E-01 |
| MFN2 | 3306,33 | -0,03 | 0,04 | -0,87 | 3,86E-01 | 5,77E-01 |
| ZFAND2B | 514,17 | 0,02 | 0,02 | 0,87 | 3,86E-01 | 5,78E-01 |
| IRF5 | 589,49 | 0,03 | 0,04 | 0,87 | 3,86E-01 | 5,78E-01 |
| NRBP2 | 118,07 | 0,05 | 0,05 | 0,87 | 3,86E-01 | 5,78E-01 |
| RPS6KC1 | 1191,03 | 0,02 | 0,02 | 0,87 | 3,86E-01 | 5,78E-01 |
| GSKIP | 640,56 | 0,04 | 0,05 | 0,87 | 3,86E-01 | 5,78E-01 |
| TRMT112 | 1425,86 | 0,03 | 0,03 | 0,87 | 3,86E-01 | 5,78E-01 |
| GABPB1 | 766,95 | 0,02 | 0,03 | 0,87 | 3,86E-01 | 5,78E-01 |
| CCNH | 4227,32 | -0,02 | 0,03 | -0,87 | 3,87E-01 | 5,78E-01 |
| CFL1 | 2557,21 | -0,03 | 0,04 | -0,87 | 3,87E-01 | 5,78E-01 |
| UBQLN4 | 170,79 | 0,03 | 0,04 | 0,87 | 3,87E-01 | 5,78E-01 |
| LANCL3 | 144,14 | 0,04 | 0,04 | 0,87 | 3,87E-01 | 5,79E-01 |
| CNOT2 | 4514,69 | 0,02 | 0,02 | 0,86 | 3,87E-01 | 5,79E-01 |
| ZFP36L1 | 2738,09 | -0,03 | 0,03 | -0,86 | 3,87E-01 | 5,79E-01 |
| COPB1 | 2828,78 | -0,02 | 0,03 | -0,86 | 3,88E-01 | 5,80E-01 |
| CHD6 | 3541,72 | 0,02 | 0,02 | 0,86 | 3,89E-01 | 5,81E-01 |
| KPNA2 | 102,30 | 0,03 | 0,04 | 0,86 | 3,89E-01 | 5,81E-01 |
| HIC1 | 139,39 | 0,05 | 0,06 | 0,86 | 3,89E-01 | 5,81E-01 |
| RAB44 | 344,80 | 0,05 | 0,06 | 0,86 | 3,89E-01 | 5,81E-01 |
| VAMP1 | 1000,35 | -0,02 | 0,03 | -0,86 | 3,89E-01 | 5,81E-01 |
| IL15 | 628,36 | 0,03 | 0,03 | 0,86 | 3,89E-01 | 5,82E-01 |
| ST7 | 194,46 | 0,02 | 0,03 | 0,86 | 3,89E-01 | 5,82E-01 |
| RCN3 | 188,24 | -0,07 | 0,09 | -0,86 | 3,90E-01 | 5,82E-01 |
| SSBP2 | 2026,59 | 0,02 | 0,03 | 0,86 | 3,90E-01 | 5,82E-01 |
| EVI2A | 2062,81 | 0,08 | 0,09 | 0,86 | 3,90E-01 | 5,82E-01 |
| CLIP2 | 225,63 | 0,05 | 0,06 | 0,86 | 3,90E-01 | 5,82E-01 |
| ADSL | 387,26 | 0,02 | 0,02 | 0,86 | 3,90E-01 | 5,82E-01 |

| | | | | | | |
|---|---|---|---|---|---|---|
| SLC22A5 | 296,46 | 0,03 | 0,04 | 0,86 | 3,90E-01 | 5,82E-01 |
| MAU2 | 1620,36 | -0,04 | 0,05 | -0,86 | 3,90E-01 | 5,82E-01 |
| MTR | 3465,10 | 0,02 | 0,02 | 0,86 | 3,91E-01 | 5,83E-01 |
| NARS1 | 2364,41 | -0,02 | 0,02 | -0,86 | 3,91E-01 | 5,83E-01 |
| CLIC5 | 182,98 | 0,04 | 0,05 | 0,86 | 3,91E-01 | 5,83E-01 |
| SRI | 1148,74 | 0,02 | 0,03 | 0,86 | 3,91E-01 | 5,83E-01 |
| RPP25L | 363,72 | 0,02 | 0,02 | 0,86 | 3,91E-01 | 5,83E-01 |
| TSPOAP1 | 453,18 | -0,06 | 0,07 | -0,86 | 3,91E-01 | 5,83E-01 |
| FAM169A | 658,34 | 0,03 | 0,04 | 0,86 | 3,91E-01 | 5,83E-01 |
| ZMAT5 | 289,22 | -0,02 | 0,02 | -0,86 | 3,91E-01 | 5,83E-01 |
| SIAE | 181,03 | 0,03 | 0,04 | 0,86 | 3,91E-01 | 5,83E-01 |
| DHRS11 | 72,55 | 0,04 | 0,05 | 0,86 | 3,92E-01 | 5,83E-01 |
| EIF4A2 | 9224,66 | 0,03 | 0,04 | 0,86 | 3,92E-01 | 5,83E-01 |
| RALA | 455,86 | 0,02 | 0,02 | 0,86 | 3,92E-01 | 5,83E-01 |
| AFF3 | 1080,56 | 0,05 | 0,06 | 0,86 | 3,92E-01 | 5,83E-01 |
| CATSPER2 | 289,41 | 0,04 | 0,04 | 0,86 | 3,92E-01 | 5,83E-01 |
| EOMES | 768,21 | 0,04 | 0,05 | 0,86 | 3,92E-01 | 5,84E-01 |
| SYNE1 | 19551,15 | -0,03 | 0,03 | -0,86 | 3,92E-01 | 5,84E-01 |
| NCR3 | 125,04 | 0,04 | 0,04 | 0,86 | 3,92E-01 | 5,84E-01 |
| BAHD1 | 540,59 | 0,02 | 0,03 | 0,86 | 3,92E-01 | 5,84E-01 |
| PSKH1 | 339,59 | 0,03 | 0,03 | 0,85 | 3,93E-01 | 5,85E-01 |
| COG1 | 663,16 | 0,02 | 0,02 | 0,85 | 3,93E-01 | 5,85E-01 |
| ZNF574 | 202,26 | -0,03 | 0,03 | -0,85 | 3,93E-01 | 5,85E-01 |
| LAMTOR2 | 580,42 | 0,02 | 0,03 | 0,85 | 3,93E-01 | 5,85E-01 |
| PAM16 | 204,39 | -0,05 | 0,05 | -0,85 | 3,93E-01 | 5,85E-01 |
| SLFN13 | 1458,24 | -0,03 | 0,03 | -0,85 | 3,93E-01 | 5,85E-01 |
| MCM9 | 1909,38 | -0,01 | 0,02 | -0,85 | 3,94E-01 | 5,86E-01 |
| ZDHHC12 | 91,48 | 0,04 | 0,05 | 0,85 | 3,94E-01 | 5,86E-01 |
| ACYP2 | 358,25 | 0,02 | 0,02 | 0,85 | 3,94E-01 | 5,86E-01 |
| ANXA7 | 2529,77 | -0,02 | 0,02 | -0,85 | 3,94E-01 | 5,86E-01 |
| NOG | 215,95 | 0,06 | 0,07 | 0,85 | 3,94E-01 | 5,86E-01 |
| MGRN1 | 1248,31 | -0,05 | 0,06 | -0,85 | 3,94E-01 | 5,86E-01 |
| FCN1 | 21391,07 | -0,03 | 0,04 | -0,85 | 3,94E-01 | 5,86E-01 |
| EOLA2 | 326,58 | 0,02 | 0,02 | 0,85 | 3,94E-01 | 5,86E-01 |
| CHTF18 | 185,27 | 0,04 | 0,05 | 0,85 | 3,95E-01 | 5,86E-01 |
| TDP1 | 919,88 | 0,02 | 0,02 | 0,85 | 3,95E-01 | 5,87E-01 |
| SFXN1 | 1125,27 | 0,02 | 0,02 | 0,85 | 3,95E-01 | 5,87E-01 |
| SUSD1 | 1276,60 | -0,02 | 0,03 | -0,85 | 3,95E-01 | 5,87E-01 |
| ANKRD26 | 642,37 | 0,03 | 0,04 | 0,85 | 3,96E-01 | 5,87E-01 |
| RHBDD2 | 252,19 | -0,03 | 0,04 | -0,85 | 3,96E-01 | 5,87E-01 |
| HNRNPR | 4759,42 | 0,02 | 0,03 | 0,85 | 3,96E-01 | 5,87E-01 |
| ENSG000002 | 442,41 | 0,07 | 0,08 | 0,85 | 3,96E-01 | 5,88E-01 |
| TMEM167A | 2970,42 | 0,04 | 0,04 | 0,85 | 3,96E-01 | 5,88E-01 |
| DNAJC11 | 619,18 | 0,02 | 0,02 | 0,85 | 3,97E-01 | 5,88E-01 |
| TMEM109 | 326,31 | 0,02 | 0,03 | 0,85 | 3,97E-01 | 5,89E-01 |
| NUDC | 636,07 | -0,02 | 0,02 | -0,85 | 3,97E-01 | 5,89E-01 |

| | | | | | | |
|---|---|---|---|---|---|---|
| FAM219A | 227,54 | -0,04 | 0,05 | -0,85 | 3,97E-01 | 5,89E-01 |
| ACACA | 833,86 | 0,03 | 0,03 | 0,85 | 3,97E-01 | 5,89E-01 |
| WSB2 | 980,83 | -0,02 | 0,03 | -0,85 | 3,97E-01 | 5,89E-01 |
| KRAS | 2815,09 | -0,02 | 0,03 | -0,85 | 3,97E-01 | 5,89E-01 |
| H3C1 | 271,86 | 0,04 | 0,05 | 0,85 | 3,97E-01 | 5,89E-01 |
| PHACTR2 | 1717,25 | 0,02 | 0,02 | 0,85 | 3,98E-01 | 5,89E-01 |
| RALGPS1 | 593,70 | -0,03 | 0,03 | -0,84 | 3,99E-01 | 5,90E-01 |
| ACSF2 | 304,45 | 0,03 | 0,03 | 0,84 | 3,99E-01 | 5,90E-01 |
| SUV39H2 | 103,83 | 0,03 | 0,04 | 0,84 | 3,99E-01 | 5,90E-01 |
| TP53I3 | 128,49 | 0,03 | 0,04 | 0,84 | 3,99E-01 | 5,90E-01 |
| FMO4 | 141,51 | 0,02 | 0,03 | 0,84 | 3,99E-01 | 5,90E-01 |
| RBM25 | 7558,69 | -0,02 | 0,02 | -0,84 | 3,99E-01 | 5,90E-01 |
| KDELR1 | 155,26 | 0,04 | 0,05 | 0,84 | 3,98E-01 | 5,90E-01 |
| ZNF132 | 310,21 | 0,03 | 0,04 | 0,84 | 3,99E-01 | 5,90E-01 |
| FAM174B | 353,55 | -0,04 | 0,05 | -0,84 | 3,99E-01 | 5,90E-01 |
| KBTBD11 | 1066,56 | -0,02 | 0,03 | -0,84 | 3,98E-01 | 5,90E-01 |
| SKI | 1039,40 | -0,05 | 0,06 | -0,84 | 3,99E-01 | 5,90E-01 |
| MGAT5 | 2678,10 | -0,02 | 0,02 | -0,84 | 3,99E-01 | 5,90E-01 |
| ACAP3 | 787,97 | 0,04 | 0,04 | 0,84 | 3,99E-01 | 5,91E-01 |
| SELENON | 108,67 | 0,06 | 0,07 | 0,84 | 3,99E-01 | 5,91E-01 |
| CDIN1 | 317,93 | 0,03 | 0,03 | 0,84 | 4,00E-01 | 5,91E-01 |
| RECQL | 1873,08 | 0,04 | 0,05 | 0,84 | 4,00E-01 | 5,91E-01 |
| RNASEH2B | 4351,98 | 0,02 | 0,02 | 0,84 | 4,00E-01 | 5,91E-01 |
| CEP164 | 951,83 | -0,03 | 0,03 | -0,84 | 4,00E-01 | 5,91E-01 |
| NAGA | 983,88 | 0,02 | 0,03 | 0,84 | 4,00E-01 | 5,91E-01 |
| CDKN2B | 153,35 | -0,04 | 0,04 | -0,84 | 4,00E-01 | 5,92E-01 |
| PAIP2B | 410,98 | 0,03 | 0,03 | 0,84 | 4,00E-01 | 5,92E-01 |
| TNS3 | 750,50 | -0,04 | 0,05 | -0,84 | 4,00E-01 | 5,92E-01 |
| ZNF799 | 330,62 | 0,02 | 0,03 | 0,84 | 4,00E-01 | 5,92E-01 |
| NBAS | 3158,70 | 0,02 | 0,02 | 0,84 | 4,00E-01 | 5,92E-01 |
| ACTB | 26653,33 | -0,04 | 0,05 | -0,84 | 4,01E-01 | 5,92E-01 |
| MAP3K12 | 226,75 | 0,03 | 0,04 | 0,84 | 4,01E-01 | 5,92E-01 |
| C2CD5 | 2645,76 | 0,02 | 0,02 | 0,84 | 4,01E-01 | 5,92E-01 |
| IGSF9B | 137,35 | 0,06 | 0,08 | 0,84 | 4,01E-01 | 5,92E-01 |
| LY6G5B | 72,93 | -0,05 | 0,06 | -0,84 | 4,01E-01 | 5,92E-01 |
| URB1 | 973,55 | 0,02 | 0,03 | 0,84 | 4,01E-01 | 5,92E-01 |
| ZCCHC3 | 1079,30 | 0,01 | 0,02 | 0,84 | 4,01E-01 | 5,92E-01 |
| SPCS1 | 372,99 | 0,02 | 0,02 | 0,84 | 4,01E-01 | 5,92E-01 |
| TESK1 | 139,68 | 0,03 | 0,04 | 0,84 | 4,02E-01 | 5,92E-01 |
| FBXO41 | 231,82 | 0,05 | 0,06 | 0,84 | 4,02E-01 | 5,92E-01 |
| FNTA | 2641,17 | 0,02 | 0,02 | 0,84 | 4,02E-01 | 5,93E-01 |
| CLHC1 | 243,36 | 0,04 | 0,05 | 0,84 | 4,02E-01 | 5,93E-01 |
| HSD17B4 | 2553,74 | -0,02 | 0,03 | -0,84 | 4,02E-01 | 5,93E-01 |
| IDO1 | 212,15 | 0,07 | 0,09 | 0,84 | 4,02E-01 | 5,93E-01 |
| KIF11 | 123,65 | 0,04 | 0,05 | 0,84 | 4,02E-01 | 5,93E-01 |
| TUBGCP4 | 561,12 | 0,02 | 0,03 | 0,84 | 4,02E-01 | 5,93E-01 |

| | | | | | | |
|---|---|---|---|---|---|---|
| SPATA20 | 212,59 | 0,05 | 0,06 | 0,84 | 4,03E-01 | 5,93E-01 |
| FZR1 | 1352,24 | 0,03 | 0,03 | 0,84 | 4,03E-01 | 5,94E-01 |
| RRP1B | 658,45 | 0,02 | 0,03 | 0,84 | 4,03E-01 | 5,94E-01 |
| PTPRS | 138,70 | -0,08 | 0,09 | -0,84 | 4,03E-01 | 5,94E-01 |
| AGRN | 113,14 | -0,10 | 0,12 | -0,84 | 4,03E-01 | 5,94E-01 |
| UBALD2 | 935,16 | 0,03 | 0,04 | 0,84 | 4,03E-01 | 5,94E-01 |
| ILK | 924,12 | -0,03 | 0,03 | -0,84 | 4,03E-01 | 5,94E-01 |
| SRPRB | 870,70 | 0,02 | 0,03 | 0,84 | 4,03E-01 | 5,94E-01 |
| MOB3B | 272,35 | 0,03 | 0,03 | 0,84 | 4,03E-01 | 5,94E-01 |
| ATN1 | 72,53 | 0,07 | 0,08 | 0,84 | 4,04E-01 | 5,94E-01 |
| RBM14 | 1731,52 | -0,03 | 0,03 | -0,84 | 4,04E-01 | 5,94E-01 |
| TRIM52 | 1723,25 | 0,02 | 0,02 | 0,83 | 4,04E-01 | 5,94E-01 |
| PFKP | 1049,05 | 0,02 | 0,03 | 0,83 | 4,04E-01 | 5,94E-01 |
| RSBN1 | 2186,95 | 0,02 | 0,02 | 0,83 | 4,04E-01 | 5,94E-01 |
| DNAH7 | 94,98 | -0,08 | 0,09 | -0,83 | 4,04E-01 | 5,95E-01 |
| ZNF576 | 177,54 | 0,01 | 0,02 | 0,83 | 4,04E-01 | 5,95E-01 |
| MPHOSPH10 | 662,13 | 0,02 | 0,03 | 0,83 | 4,04E-01 | 5,95E-01 |
| HPF1 | 190,35 | 0,03 | 0,03 | 0,83 | 4,04E-01 | 5,95E-01 |
| TBC1D9 | 1340,38 | 0,03 | 0,03 | 0,83 | 4,05E-01 | 5,95E-01 |
| PSME2 | 486,42 | -0,03 | 0,03 | -0,83 | 4,05E-01 | 5,95E-01 |
| MT1F | 78,97 | -0,03 | 0,04 | -0,83 | 4,05E-01 | 5,95E-01 |
| EED | 1144,48 | 0,03 | 0,03 | 0,83 | 4,05E-01 | 5,95E-01 |
| PHKG2 | 355,01 | -0,02 | 0,03 | -0,83 | 4,05E-01 | 5,95E-01 |
| DHCR24 | 118,42 | 0,05 | 0,06 | 0,83 | 4,05E-01 | 5,95E-01 |
| VPS9D1 | 455,35 | -0,04 | 0,05 | -0,83 | 4,05E-01 | 5,95E-01 |
| SLC25A11 | 726,04 | -0,01 | 0,02 | -0,83 | 4,05E-01 | 5,96E-01 |
| DVL2 | 177,83 | 0,04 | 0,04 | 0,83 | 4,06E-01 | 5,96E-01 |
| TMEM106A | 350,57 | -0,04 | 0,05 | -0,83 | 4,06E-01 | 5,96E-01 |
| COL24A1 | 86,69 | 0,06 | 0,08 | 0,83 | 4,06E-01 | 5,96E-01 |
| RANBP9 | 1219,10 | 0,02 | 0,03 | 0,83 | 4,06E-01 | 5,96E-01 |
| IFT88 | 554,80 | -0,02 | 0,03 | -0,83 | 4,06E-01 | 5,96E-01 |
| SLC27A1 | 142,99 | 0,05 | 0,06 | 0,83 | 4,06E-01 | 5,96E-01 |
| DNAAF5 | 197,23 | 0,04 | 0,05 | 0,83 | 4,06E-01 | 5,96E-01 |
| FAM162A | 278,07 | 0,02 | 0,03 | 0,83 | 4,06E-01 | 5,96E-01 |
| BATF | 188,65 | 0,03 | 0,03 | 0,83 | 4,06E-01 | 5,96E-01 |
| METTL18 | 141,86 | 0,06 | 0,07 | 0,83 | 4,06E-01 | 5,96E-01 |
| TMEM179B | 264,82 | -0,02 | 0,03 | -0,83 | 4,07E-01 | 5,97E-01 |
| ERC1 | 1581,24 | -0,02 | 0,03 | -0,83 | 4,07E-01 | 5,97E-01 |
| GAPT | 1332,21 | 0,04 | 0,05 | 0,83 | 4,07E-01 | 5,97E-01 |
| TVP23C | 515,89 | -0,02 | 0,03 | -0,83 | 4,07E-01 | 5,97E-01 |
| PPIP5K1 | 151,69 | 0,02 | 0,03 | 0,83 | 4,07E-01 | 5,97E-01 |
| BACE1 | 72,78 | 0,06 | 0,07 | 0,83 | 4,07E-01 | 5,97E-01 |
| HNRNPAB | 1250,93 | -0,02 | 0,02 | -0,83 | 4,07E-01 | 5,97E-01 |
| RSRC2 | 3400,84 | -0,02 | 0,02 | -0,83 | 4,08E-01 | 5,97E-01 |
| RAB28 | 677,80 | 0,02 | 0,02 | 0,83 | 4,08E-01 | 5,97E-01 |
| CA8 | 105,60 | -0,07 | 0,08 | -0,83 | 4,08E-01 | 5,98E-01 |

| | | | | | | |
|---|---|---|---|---|---|---|
| NBPF9 | 359,22 | 0,03 | 0,04 | 0,83 | 4,08E-01 | 5,98E-01 |
| CLK1 | 7565,20 | -0,03 | 0,03 | -0,83 | 4,09E-01 | 5,98E-01 |
| ABCD1 | 255,80 | -0,04 | 0,05 | -0,83 | 4,09E-01 | 5,99E-01 |
| SLC25A16 | 928,89 | 0,01 | 0,01 | 0,83 | 4,09E-01 | 5,99E-01 |
| DUSP14 | 74,62 | -0,03 | 0,04 | -0,83 | 4,09E-01 | 5,99E-01 |
| PILRB | 495,16 | -0,05 | 0,06 | -0,83 | 4,09E-01 | 5,99E-01 |
| GSS | 306,66 | 0,02 | 0,02 | 0,83 | 4,09E-01 | 5,99E-01 |
| C1orf159 | 177,81 | 0,04 | 0,05 | 0,83 | 4,09E-01 | 5,99E-01 |
| PRELID1 | 2165,77 | -0,02 | 0,03 | -0,82 | 4,09E-01 | 5,99E-01 |
| MED12L | 139,56 | 0,05 | 0,06 | 0,83 | 4,09E-01 | 5,99E-01 |
| GLYCTK | 401,03 | 0,03 | 0,04 | 0,82 | 4,10E-01 | 5,99E-01 |
| ZCCHC9 | 760,26 | 0,02 | 0,03 | 0,82 | 4,10E-01 | 5,99E-01 |
| MTF2 | 1751,36 | 0,02 | 0,03 | 0,82 | 4,10E-01 | 5,99E-01 |
| REEP4 | 110,91 | 0,05 | 0,06 | 0,82 | 4,10E-01 | 6,00E-01 |
| CWC22 | 1127,43 | -0,03 | 0,03 | -0,82 | 4,10E-01 | 6,00E-01 |
| CYBB | 20090,73 | 0,03 | 0,04 | 0,82 | 4,11E-01 | 6,00E-01 |
| FAM83G | 119,06 | 0,05 | 0,06 | 0,82 | 4,11E-01 | 6,00E-01 |
| RUNX1 | 2987,16 | -0,02 | 0,02 | -0,82 | 4,11E-01 | 6,00E-01 |
| TTC32 | 739,37 | 0,03 | 0,04 | 0,82 | 4,11E-01 | 6,00E-01 |
| VSIG1 | 292,21 | 0,03 | 0,04 | 0,82 | 4,10E-01 | 6,00E-01 |
| UBC | 11885,51 | -0,02 | 0,03 | -0,82 | 4,11E-01 | 6,00E-01 |
| UBXN4 | 3645,98 | -0,02 | 0,03 | -0,82 | 4,11E-01 | 6,00E-01 |
| TRERF1 | 1678,73 | -0,03 | 0,03 | -0,82 | 4,11E-01 | 6,00E-01 |
| CBFB | 2263,50 | 0,02 | 0,02 | 0,82 | 4,11E-01 | 6,00E-01 |
| LRRC1 | 134,81 | 0,03 | 0,04 | 0,82 | 4,11E-01 | 6,00E-01 |
| DCP1A | 933,50 | -0,01 | 0,02 | -0,82 | 4,11E-01 | 6,00E-01 |
| TCAIM | 699,50 | 0,02 | 0,02 | 0,82 | 4,11E-01 | 6,00E-01 |
| SREBF1 | 401,27 | 0,05 | 0,06 | 0,82 | 4,11E-01 | 6,00E-01 |
| TRIM14 | 1051,24 | -0,04 | 0,04 | -0,82 | 4,11E-01 | 6,01E-01 |
| FAM120AOS | 1868,31 | -0,01 | 0,02 | -0,82 | 4,12E-01 | 6,01E-01 |
| METTL25 | 611,78 | 0,02 | 0,03 | 0,82 | 4,12E-01 | 6,01E-01 |
| ZDHHC13 | 988,58 | -0,02 | 0,02 | -0,82 | 4,12E-01 | 6,01E-01 |
| EML5 | 396,40 | 0,03 | 0,04 | 0,82 | 4,12E-01 | 6,01E-01 |
| ILKAP | 706,16 | 0,02 | 0,02 | 0,82 | 4,12E-01 | 6,01E-01 |
| HOXB3 | 116,05 | -0,06 | 0,07 | -0,82 | 4,12E-01 | 6,01E-01 |
| ENSG000002 | 149,71 | 0,04 | 0,04 | 0,82 | 4,12E-01 | 6,01E-01 |
| HIPK1 | 8064,09 | -0,02 | 0,03 | -0,82 | 4,12E-01 | 6,01E-01 |
| ADRB2 | 1340,23 | 0,03 | 0,03 | 0,82 | 4,12E-01 | 6,01E-01 |
| ZBTB17 | 607,38 | -0,02 | 0,02 | -0,82 | 4,13E-01 | 6,01E-01 |
| PPM1M | 1797,31 | -0,02 | 0,02 | -0,82 | 4,13E-01 | 6,01E-01 |
| IMPG2 | 204,13 | 0,03 | 0,03 | 0,82 | 4,13E-01 | 6,01E-01 |
| CHCHD7 | 866,47 | 0,02 | 0,02 | 0,82 | 4,13E-01 | 6,01E-01 |
| SCIMP | 862,16 | 0,03 | 0,03 | 0,82 | 4,13E-01 | 6,01E-01 |
| SH2B3 | 2517,04 | -0,03 | 0,04 | -0,82 | 4,13E-01 | 6,01E-01 |
| MYO10 | 96,75 | -0,08 | 0,09 | -0,82 | 4,13E-01 | 6,02E-01 |
| SMIM29 | 209,63 | 0,02 | 0,02 | 0,82 | 4,13E-01 | 6,02E-01 |

| | | | | | | |
|---|---|---|---|---|---|---|
| TGFBI | 1834,01 | 0,03 | 0,04 | 0,82 | 4,14E-01 | 6,02E-01 |
| FTSJ3 | 1018,81 | -0,02 | 0,02 | -0,82 | 4,14E-01 | 6,02E-01 |
| EMC1 | 600,87 | 0,02 | 0,03 | 0,82 | 4,14E-01 | 6,03E-01 |
| FHL1 | 361,67 | -0,03 | 0,04 | -0,82 | 4,14E-01 | 6,03E-01 |
| ARHGEF10L | 619,91 | 0,03 | 0,04 | 0,82 | 4,15E-01 | 6,03E-01 |
| ZNF793 | 288,94 | 0,03 | 0,04 | 0,82 | 4,15E-01 | 6,03E-01 |
| S100Z | 306,37 | -0,03 | 0,04 | -0,82 | 4,15E-01 | 6,04E-01 |
| ACO2 | 602,99 | 0,03 | 0,03 | 0,82 | 4,15E-01 | 6,04E-01 |
| DHX57 | 844,83 | 0,01 | 0,02 | 0,81 | 4,15E-01 | 6,04E-01 |
| TJAP1 | 268,73 | 0,03 | 0,04 | 0,81 | 4,16E-01 | 6,04E-01 |
| HMBOX1 | 1915,09 | -0,02 | 0,02 | -0,81 | 4,16E-01 | 6,05E-01 |
| EHBP1 | 748,30 | 0,02 | 0,02 | 0,81 | 4,16E-01 | 6,05E-01 |
| KCNAB2 | 2975,14 | -0,03 | 0,04 | -0,81 | 4,16E-01 | 6,05E-01 |
| PDPR | 1129,04 | 0,04 | 0,05 | 0,81 | 4,16E-01 | 6,05E-01 |
| JCHAIN | 1648,00 | 0,08 | 0,10 | 0,81 | 4,16E-01 | 6,05E-01 |
| ZNF692 | 458,80 | -0,02 | 0,03 | -0,81 | 4,16E-01 | 6,05E-01 |
| SLFN5 | 6987,98 | 0,02 | 0,03 | 0,81 | 4,17E-01 | 6,05E-01 |
| MLH1 | 1008,67 | 0,01 | 0,02 | 0,81 | 4,17E-01 | 6,05E-01 |
| S100PBP | 2040,89 | -0,01 | 0,02 | -0,81 | 4,17E-01 | 6,06E-01 |
| RAB3GAP2 | 3021,09 | -0,02 | 0,02 | -0,81 | 4,17E-01 | 6,06E-01 |
| GOT1 | 193,31 | -0,02 | 0,02 | -0,81 | 4,17E-01 | 6,06E-01 |
| RIC1 | 3441,85 | -0,02 | 0,02 | -0,81 | 4,17E-01 | 6,06E-01 |
| RPP14 | 688,56 | 0,01 | 0,01 | 0,81 | 4,17E-01 | 6,06E-01 |
| ZNF585A | 337,05 | -0,02 | 0,02 | -0,81 | 4,17E-01 | 6,06E-01 |
| RASA2 | 4714,86 | -0,02 | 0,03 | -0,81 | 4,17E-01 | 6,06E-01 |
| NAT10 | 1068,22 | 0,02 | 0,02 | 0,81 | 4,17E-01 | 6,06E-01 |
| ZMAT2 | 2884,21 | -0,04 | 0,05 | -0,81 | 4,18E-01 | 6,06E-01 |
| CNOT3 | 269,00 | -0,05 | 0,06 | -0,81 | 4,18E-01 | 6,06E-01 |
| PBRM1 | 3489,45 | -0,01 | 0,02 | -0,81 | 4,18E-01 | 6,06E-01 |
| DPY30 | 335,75 | 0,03 | 0,04 | 0,81 | 4,18E-01 | 6,06E-01 |
| NFX1 | 1376,78 | 0,01 | 0,02 | 0,81 | 4,18E-01 | 6,06E-01 |
| RPGRIP1L | 199,30 | 0,03 | 0,04 | 0,81 | 4,18E-01 | 6,06E-01 |
| SLC46A2 | 257,64 | 0,03 | 0,04 | 0,81 | 4,18E-01 | 6,06E-01 |
| NAP1L2 | 176,22 | 0,04 | 0,05 | 0,81 | 4,18E-01 | 6,07E-01 |
| SYTL4 | 108,81 | -0,05 | 0,07 | -0,81 | 4,19E-01 | 6,07E-01 |
| TNFSF8 | 1099,81 | -0,02 | 0,03 | -0,81 | 4,19E-01 | 6,07E-01 |
| PIK3C2B | 499,39 | 0,03 | 0,04 | 0,81 | 4,19E-01 | 6,07E-01 |
| SMAD3 | 1795,69 | 0,03 | 0,04 | 0,81 | 4,19E-01 | 6,07E-01 |
| SHLD1 | 290,77 | 0,03 | 0,03 | 0,81 | 4,19E-01 | 6,07E-01 |
| USP42 | 388,81 | -0,03 | 0,03 | -0,81 | 4,19E-01 | 6,07E-01 |
| TOR4A | 366,97 | 0,04 | 0,05 | 0,81 | 4,19E-01 | 6,07E-01 |
| EML4 | 5185,01 | 0,02 | 0,02 | 0,81 | 4,20E-01 | 6,07E-01 |
| TMX2 | 641,21 | 0,01 | 0,01 | 0,81 | 4,20E-01 | 6,07E-01 |
| NFKBIL1 | 89,73 | 0,02 | 0,03 | 0,81 | 4,20E-01 | 6,07E-01 |
| DNAL1 | 257,29 | 0,02 | 0,02 | 0,81 | 4,20E-01 | 6,08E-01 |
| UFC1 | 643,35 | 0,03 | 0,03 | 0,81 | 4,20E-01 | 6,08E-01 |

| | | | | | | |
|---|---|---|---|---|---|---|
| CD300E | 1257,80 | -0,04 | 0,04 | -0,81 | 4,20E-01 | 6,08E-01 |
| WDR70 | 1157,96 | 0,02 | 0,02 | 0,81 | 4,20E-01 | 6,08E-01 |
| FAM20C | 96,68 | 0,06 | 0,07 | 0,81 | 4,20E-01 | 6,08E-01 |
| ABHD13 | 2232,02 | 0,04 | 0,05 | 0,81 | 4,21E-01 | 6,08E-01 |
| SMG8 | 628,44 | 0,02 | 0,02 | 0,81 | 4,21E-01 | 6,09E-01 |
| PTPN7 | 561,89 | 0,03 | 0,04 | 0,80 | 4,21E-01 | 6,09E-01 |
| MITF | 151,43 | 0,04 | 0,04 | 0,80 | 4,21E-01 | 6,09E-01 |
| PDLIM2 | 1116,54 | -0,03 | 0,04 | -0,80 | 4,21E-01 | 6,09E-01 |
| PLPBP | 881,91 | 0,01 | 0,02 | 0,80 | 4,21E-01 | 6,09E-01 |
| ATRIP | 225,40 | 0,02 | 0,02 | 0,80 | 4,21E-01 | 6,09E-01 |
| PLXNA4 | 226,94 | -0,05 | 0,07 | -0,80 | 4,21E-01 | 6,09E-01 |
| RNF25 | 252,39 | 0,01 | 0,01 | 0,80 | 4,21E-01 | 6,09E-01 |
| GFI1B | 287,81 | 0,04 | 0,06 | 0,80 | 4,21E-01 | 6,09E-01 |
| PUM3 | 749,64 | 0,02 | 0,03 | 0,80 | 4,22E-01 | 6,10E-01 |
| SNRPN | 307,34 | 0,02 | 0,02 | 0,80 | 4,23E-01 | 6,10E-01 |
| SEC16A | 1483,56 | -0,04 | 0,05 | -0,80 | 4,23E-01 | 6,10E-01 |
| RNF166 | 1453,58 | -0,04 | 0,05 | -0,80 | 4,23E-01 | 6,10E-01 |
| VTA1 | 962,70 | 0,03 | 0,03 | 0,80 | 4,23E-01 | 6,10E-01 |
| ID2 | 1119,44 | 0,02 | 0,03 | 0,80 | 4,23E-01 | 6,10E-01 |
| MOB4 | 710,23 | 0,02 | 0,03 | 0,80 | 4,23E-01 | 6,10E-01 |
| DCPS | 323,62 | -0,02 | 0,02 | -0,80 | 4,23E-01 | 6,10E-01 |
| COQ5 | 325,49 | 0,03 | 0,03 | 0,80 | 4,23E-01 | 6,11E-01 |
| CAPZB | 1181,77 | -0,03 | 0,04 | -0,80 | 4,23E-01 | 6,11E-01 |
| FXN | 321,72 | 0,02 | 0,03 | 0,80 | 4,23E-01 | 6,11E-01 |
| ATP5F1A | 5510,45 | 0,02 | 0,02 | 0,80 | 4,23E-01 | 6,11E-01 |
| POP4 | 620,52 | 0,01 | 0,02 | 0,80 | 4,24E-01 | 6,11E-01 |
| NMT1 | 1496,81 | -0,01 | 0,02 | -0,80 | 4,24E-01 | 6,11E-01 |
| CPM | 180,18 | -0,03 | 0,04 | -0,80 | 4,24E-01 | 6,11E-01 |
| C7orf25 | 137,52 | -0,03 | 0,04 | -0,80 | 4,24E-01 | 6,11E-01 |
| BRF1 | 970,93 | 0,02 | 0,03 | 0,80 | 4,24E-01 | 6,11E-01 |
| DHFR2 | 431,84 | 0,03 | 0,03 | 0,80 | 4,24E-01 | 6,11E-01 |
| ZNF304 | 497,03 | 0,02 | 0,03 | 0,80 | 4,24E-01 | 6,12E-01 |
| FRMD8 | 841,13 | -0,03 | 0,04 | -0,80 | 4,25E-01 | 6,12E-01 |
| CBR1 | 403,59 | 0,02 | 0,03 | 0,80 | 4,25E-01 | 6,12E-01 |
| IFT122 | 396,69 | -0,02 | 0,03 | -0,80 | 4,25E-01 | 6,12E-01 |
| IDH3G | 611,31 | 0,02 | 0,02 | 0,80 | 4,25E-01 | 6,13E-01 |
| PDLIM1 | 374,65 | 0,03 | 0,04 | 0,80 | 4,25E-01 | 6,13E-01 |
| NFE2 | 5616,77 | -0,03 | 0,04 | -0,80 | 4,25E-01 | 6,13E-01 |
| THOP1 | 218,97 | 0,03 | 0,04 | 0,80 | 4,25E-01 | 6,13E-01 |
| TPRG1L | 1217,79 | -0,02 | 0,03 | -0,80 | 4,26E-01 | 6,13E-01 |
| TAS2R4 | 211,58 | -0,03 | 0,04 | -0,80 | 4,26E-01 | 6,13E-01 |
| TRAF3 | 1350,95 | 0,02 | 0,02 | 0,80 | 4,26E-01 | 6,13E-01 |
| GYS1 | 348,50 | 0,03 | 0,04 | 0,80 | 4,26E-01 | 6,13E-01 |
| MFNG | 957,64 | 0,03 | 0,03 | 0,80 | 4,26E-01 | 6,13E-01 |
| ECPAS | 3202,93 | -0,01 | 0,02 | -0,80 | 4,26E-01 | 6,13E-01 |
| OLFM4 | 127,56 | -0,11 | 0,13 | -0,80 | 4,26E-01 | 6,13E-01 |

| | | | | | | |
|---|---|---|---|---|---|---|
| MICAL3 | 908,34 | 0,03 | 0,04 | 0,80 | 4,26E-01 | 6,13E-01 |
| CDRT4 | 77,20 | -0,05 | 0,06 | -0,80 | 4,26E-01 | 6,13E-01 |
| METTL4 | 831,85 | -0,02 | 0,03 | -0,80 | 4,26E-01 | 6,13E-01 |
| NBPF12 | 1240,44 | -0,02 | 0,03 | -0,80 | 4,26E-01 | 6,13E-01 |
| SMAD1 | 138,34 | 0,04 | 0,06 | 0,80 | 4,27E-01 | 6,13E-01 |
| LRP6 | 202,07 | 0,05 | 0,06 | 0,79 | 4,27E-01 | 6,14E-01 |
| ZNF71 | 290,57 | 0,02 | 0,02 | 0,79 | 4,27E-01 | 6,14E-01 |
| BCO2 | 76,59 | -0,04 | 0,05 | -0,79 | 4,27E-01 | 6,14E-01 |
| DCTN4 | 2103,78 | -0,01 | 0,02 | -0,79 | 4,28E-01 | 6,15E-01 |
| PPAT | 361,32 | 0,02 | 0,03 | 0,79 | 4,28E-01 | 6,15E-01 |
| STAC3 | 123,59 | -0,03 | 0,04 | -0,79 | 4,28E-01 | 6,15E-01 |
| SEPTIN11 | 1102,42 | 0,02 | 0,02 | 0,79 | 4,28E-01 | 6,15E-01 |
| CDK9 | 1356,33 | -0,01 | 0,02 | -0,79 | 4,29E-01 | 6,15E-01 |
| TYW1B | 295,09 | -0,02 | 0,03 | -0,79 | 4,29E-01 | 6,16E-01 |
| PSPC1 | 3735,84 | -0,01 | 0,02 | -0,79 | 4,29E-01 | 6,16E-01 |
| KIF3A | 562,03 | 0,02 | 0,03 | 0,79 | 4,29E-01 | 6,16E-01 |
| RBM43 | 458,29 | 0,02 | 0,03 | 0,79 | 4,29E-01 | 6,16E-01 |
| HNRNPF | 4806,93 | -0,02 | 0,02 | -0,79 | 4,29E-01 | 6,16E-01 |
| COA5 | 1157,71 | 0,02 | 0,03 | 0,79 | 4,30E-01 | 6,17E-01 |
| TSGA10 | 422,35 | 0,03 | 0,04 | 0,79 | 4,30E-01 | 6,17E-01 |
| RACGAP1 | 297,75 | -0,02 | 0,02 | -0,79 | 4,30E-01 | 6,17E-01 |
| POLR1E | 404,82 | 0,02 | 0,02 | 0,79 | 4,30E-01 | 6,17E-01 |
| CYB561 | 361,01 | 0,03 | 0,04 | 0,79 | 4,30E-01 | 6,17E-01 |
| NDFIP1 | 2975,40 | 0,02 | 0,02 | 0,79 | 4,30E-01 | 6,17E-01 |
| PRR14 | 401,92 | -0,03 | 0,04 | -0,79 | 4,30E-01 | 6,17E-01 |
| CD247 | 4229,56 | -0,02 | 0,03 | -0,79 | 4,30E-01 | 6,17E-01 |
| CCDC122 | 169,16 | 0,05 | 0,06 | 0,79 | 4,31E-01 | 6,17E-01 |
| SIPA1 | 2188,44 | -0,04 | 0,05 | -0,79 | 4,31E-01 | 6,17E-01 |
| ABITRAM | 202,74 | 0,02 | 0,02 | 0,79 | 4,31E-01 | 6,17E-01 |
| TSTD2 | 624,34 | 0,01 | 0,02 | 0,79 | 4,31E-01 | 6,17E-01 |
| CARHSP1 | 997,19 | -0,02 | 0,02 | -0,79 | 4,31E-01 | 6,17E-01 |
| TNNI2 | 84,35 | -0,03 | 0,04 | -0,79 | 4,30E-01 | 6,17E-01 |
| HINT3 | 978,63 | -0,02 | 0,03 | -0,79 | 4,31E-01 | 6,17E-01 |
| AP3S2 | 564,53 | -0,02 | 0,03 | -0,79 | 4,32E-01 | 6,18E-01 |
| CIAPIN1 | 493,29 | 0,01 | 0,01 | 0,79 | 4,32E-01 | 6,18E-01 |
| DYRK1B | 269,24 | -0,03 | 0,04 | -0,79 | 4,32E-01 | 6,18E-01 |
| NEPRO | 1437,91 | 0,02 | 0,02 | 0,79 | 4,32E-01 | 6,18E-01 |
| LTK | 158,26 | 0,05 | 0,06 | 0,79 | 4,32E-01 | 6,19E-01 |
| PCMTD1 | 6375,33 | 0,03 | 0,04 | 0,79 | 4,32E-01 | 6,19E-01 |
| CDC37L1 | 636,88 | 0,03 | 0,04 | 0,78 | 4,32E-01 | 6,19E-01 |
| PKM | 2692,44 | 0,03 | 0,04 | 0,78 | 4,33E-01 | 6,19E-01 |
| MIEN1 | 93,33 | 0,03 | 0,04 | 0,78 | 4,33E-01 | 6,19E-01 |
| ZNF2 | 228,39 | -0,01 | 0,01 | -0,78 | 4,33E-01 | 6,19E-01 |
| MFSD10 | 437,73 | 0,03 | 0,03 | 0,78 | 4,33E-01 | 6,19E-01 |
| GIMAP7 | 2975,76 | 0,04 | 0,05 | 0,78 | 4,33E-01 | 6,20E-01 |
| CUL1 | 1515,97 | -0,01 | 0,02 | -0,78 | 4,34E-01 | 6,20E-01 |

| | | | | | | |
|---|---|---|---|---|---|---|
| GLUL | 28995,58 | -0,03 | 0,04 | -0,78 | 4,34E-01 | 6,20E-01 |
| ARRDC1 | 621,94 | -0,03 | 0,04 | -0,78 | 4,34E-01 | 6,21E-01 |
| ZNF777 | 82,56 | 0,05 | 0,06 | 0,78 | 4,34E-01 | 6,21E-01 |
| INTS5 | 215,63 | 0,03 | 0,04 | 0,78 | 4,34E-01 | 6,21E-01 |
| RBM18 | 763,57 | -0,02 | 0,02 | -0,78 | 4,35E-01 | 6,21E-01 |
| SERPINB6 | 1551,40 | -0,02 | 0,03 | -0,78 | 4,35E-01 | 6,21E-01 |
| FADS3 | 86,84 | 0,04 | 0,06 | 0,78 | 4,35E-01 | 6,22E-01 |
| CLCN7 | 635,38 | 0,05 | 0,06 | 0,78 | 4,36E-01 | 6,22E-01 |
| FGFR1OP2 | 3454,70 | 0,03 | 0,04 | 0,78 | 4,36E-01 | 6,22E-01 |
| SPICE1 | 297,89 | 0,02 | 0,03 | 0,78 | 4,36E-01 | 6,22E-01 |
| TTLL12 | 225,63 | 0,03 | 0,04 | 0,78 | 4,36E-01 | 6,22E-01 |
| MRPL23 | 149,35 | 0,05 | 0,06 | 0,78 | 4,36E-01 | 6,22E-01 |
| ACAT1 | 333,88 | 0,03 | 0,04 | 0,78 | 4,36E-01 | 6,22E-01 |
| M6PR | 4036,93 | 0,02 | 0,02 | 0,78 | 4,36E-01 | 6,22E-01 |
| CCNL1 | 9816,14 | -0,02 | 0,03 | -0,78 | 4,36E-01 | 6,22E-01 |
| HS2ST1 | 994,47 | 0,02 | 0,03 | 0,78 | 4,36E-01 | 6,22E-01 |
| TRADD | 1596,39 | 0,02 | 0,02 | 0,78 | 4,36E-01 | 6,22E-01 |
| TNFSF4 | 331,62 | 0,03 | 0,04 | 0,78 | 4,37E-01 | 6,23E-01 |
| DBF4B | 132,58 | -0,04 | 0,05 | -0,78 | 4,37E-01 | 6,23E-01 |
| TMEM79 | 80,60 | -0,03 | 0,04 | -0,78 | 4,37E-01 | 6,23E-01 |
| BDH1 | 303,81 | 0,03 | 0,04 | 0,78 | 4,37E-01 | 6,23E-01 |
| SCRN2 | 235,96 | 0,02 | 0,03 | 0,78 | 4,37E-01 | 6,24E-01 |
| DHX33 | 371,01 | 0,01 | 0,02 | 0,78 | 4,37E-01 | 6,24E-01 |
| STARD10 | 125,18 | -0,04 | 0,05 | -0,78 | 4,37E-01 | 6,24E-01 |
| SERP1 | 4007,31 | 0,03 | 0,03 | 0,78 | 4,38E-01 | 6,24E-01 |
| PSMD10 | 681,49 | 0,03 | 0,04 | 0,78 | 4,38E-01 | 6,24E-01 |
| SLC4A8 | 525,01 | 0,03 | 0,04 | 0,78 | 4,38E-01 | 6,24E-01 |
| BTBD19 | 83,15 | -0,05 | 0,07 | -0,78 | 4,38E-01 | 6,24E-01 |
| ATP2C1 | 2271,26 | 0,02 | 0,02 | 0,77 | 4,38E-01 | 6,25E-01 |
| IL18BP | 237,65 | 0,04 | 0,05 | 0,77 | 4,38E-01 | 6,25E-01 |
| CDKN1B | 6931,85 | -0,02 | 0,03 | -0,77 | 4,39E-01 | 6,25E-01 |
| CCDC112 | 266,70 | 0,03 | 0,04 | 0,77 | 4,39E-01 | 6,25E-01 |
| CTBP1 | 3993,07 | -0,01 | 0,02 | -0,77 | 4,39E-01 | 6,25E-01 |
| FAM32A | 1498,76 | -0,02 | 0,03 | -0,77 | 4,39E-01 | 6,25E-01 |
| TGFB1 | 836,73 | 0,05 | 0,07 | 0,77 | 4,39E-01 | 6,25E-01 |
| POU2F2 | 732,72 | 0,04 | 0,05 | 0,77 | 4,39E-01 | 6,25E-01 |
| NSL1 | 1085,44 | 0,02 | 0,03 | 0,77 | 4,40E-01 | 6,26E-01 |
| PRDX1 | 1834,91 | 0,03 | 0,04 | 0,77 | 4,40E-01 | 6,26E-01 |
| ARL17A | 260,42 | 0,06 | 0,08 | 0,77 | 4,40E-01 | 6,26E-01 |
| HMOX1 | 310,37 | -0,03 | 0,04 | -0,77 | 4,40E-01 | 6,26E-01 |
| VENTX | 91,79 | 0,05 | 0,06 | 0,77 | 4,40E-01 | 6,26E-01 |
| FZD3 | 256,05 | 0,03 | 0,04 | 0,77 | 4,40E-01 | 6,26E-01 |
| PTDSS1 | 2062,26 | 0,01 | 0,02 | 0,77 | 4,40E-01 | 6,26E-01 |
| SPINDOC | 83,09 | 0,03 | 0,04 | 0,77 | 4,40E-01 | 6,26E-01 |
| CRIP2 | 108,20 | 0,05 | 0,07 | 0,77 | 4,40E-01 | 6,26E-01 |
| H2BC8 | 719,10 | 0,03 | 0,04 | 0,77 | 4,40E-01 | 6,26E-01 |

| | | | | | | |
|---|---|---|---|---|---|---|
| PTPN13 | 358,34 | 0,03 | 0,04 | 0,77 | 4,40E-01 | 6,26E-01 |
| NCOR2 | 1100,41 | 0,04 | 0,05 | 0,77 | 4,41E-01 | 6,27E-01 |
| FBXO44 | 227,05 | 0,03 | 0,04 | 0,77 | 4,41E-01 | 6,27E-01 |
| B3GNT5 | 608,02 | -0,03 | 0,04 | -0,77 | 4,41E-01 | 6,27E-01 |
| UPRT | 610,50 | 0,02 | 0,02 | 0,77 | 4,41E-01 | 6,27E-01 |
| NCAPH2 | 599,32 | 0,02 | 0,02 | 0,77 | 4,42E-01 | 6,27E-01 |
| LHPP | 179,28 | 0,03 | 0,04 | 0,77 | 4,42E-01 | 6,27E-01 |
| ETFA | 1714,42 | 0,02 | 0,02 | 0,77 | 4,42E-01 | 6,27E-01 |
| NCAPD3 | 909,83 | -0,02 | 0,02 | -0,77 | 4,42E-01 | 6,27E-01 |
| MRPS9 | 451,89 | 0,02 | 0,03 | 0,77 | 4,42E-01 | 6,27E-01 |
| TRMT1L | 781,95 | 0,02 | 0,03 | 0,77 | 4,42E-01 | 6,27E-01 |
| FN3KRP | 340,97 | 0,02 | 0,02 | 0,77 | 4,42E-01 | 6,27E-01 |
| SRCAP | 429,46 | -0,03 | 0,03 | -0,77 | 4,42E-01 | 6,28E-01 |
| NEMP2 | 763,74 | 0,02 | 0,03 | 0,77 | 4,43E-01 | 6,28E-01 |
| FAT1 | 81,50 | 0,08 | 0,11 | 0,77 | 4,43E-01 | 6,29E-01 |
| ACP2 | 149,69 | 0,03 | 0,04 | 0,77 | 4,43E-01 | 6,29E-01 |
| DACH1 | 160,88 | 0,04 | 0,05 | 0,77 | 4,43E-01 | 6,29E-01 |
| LRIF1 | 799,59 | -0,02 | 0,03 | -0,77 | 4,43E-01 | 6,29E-01 |
| LEMD2 | 2464,84 | -0,01 | 0,01 | -0,77 | 4,44E-01 | 6,30E-01 |
| FLT4 | 110,37 | 0,07 | 0,09 | 0,77 | 4,44E-01 | 6,30E-01 |
| LYAR | 693,70 | 0,03 | 0,03 | 0,77 | 4,44E-01 | 6,30E-01 |
| RANBP2 | 5744,75 | -0,02 | 0,03 | -0,77 | 4,44E-01 | 6,30E-01 |
| TTC7A | 1784,59 | 0,02 | 0,03 | 0,76 | 4,45E-01 | 6,30E-01 |
| CHEK2 | 198,18 | 0,02 | 0,03 | 0,76 | 4,45E-01 | 6,31E-01 |
| NLRC3 | 3021,92 | 0,02 | 0,03 | 0,76 | 4,45E-01 | 6,31E-01 |
| SOCS7 | 675,89 | -0,01 | 0,02 | -0,76 | 4,45E-01 | 6,31E-01 |
| PWP2 | 95,36 | -0,10 | 0,12 | -0,76 | 4,45E-01 | 6,31E-01 |
| LILRB4 | 118,96 | 0,04 | 0,06 | 0,76 | 4,45E-01 | 6,31E-01 |
| SPCS3 | 7649,35 | 0,03 | 0,03 | 0,76 | 4,45E-01 | 6,31E-01 |
| LMO2 | 2058,53 | -0,03 | 0,03 | -0,76 | 4,46E-01 | 6,32E-01 |
| CITED2 | 941,07 | 0,01 | 0,02 | 0,76 | 4,46E-01 | 6,32E-01 |
| SH3PXD2A | 307,92 | 0,04 | 0,05 | 0,76 | 4,46E-01 | 6,32E-01 |
| CA2 | 469,89 | 0,04 | 0,06 | 0,76 | 4,46E-01 | 6,32E-01 |
| WDR5 | 358,06 | 0,02 | 0,02 | 0,76 | 4,46E-01 | 6,32E-01 |
| TOMM40L | 286,56 | 0,01 | 0,02 | 0,76 | 4,46E-01 | 6,32E-01 |
| SCNM1 | 240,25 | 0,02 | 0,03 | 0,76 | 4,46E-01 | 6,32E-01 |
| AIF1 | 1539,26 | -0,04 | 0,05 | -0,76 | 4,47E-01 | 6,32E-01 |
| MFSD13A | 108,95 | -0,04 | 0,05 | -0,76 | 4,47E-01 | 6,32E-01 |
| PEX19 | 494,53 | -0,01 | 0,01 | -0,76 | 4,47E-01 | 6,32E-01 |
| CCDC144A | 729,81 | 0,12 | 0,15 | 0,76 | 4,47E-01 | 6,32E-01 |
| USP49 | 460,32 | -0,02 | 0,02 | -0,76 | 4,47E-01 | 6,32E-01 |
| ILF2 | 1399,39 | 0,02 | 0,03 | 0,76 | 4,47E-01 | 6,32E-01 |
| KLF2 | 3392,22 | 0,02 | 0,03 | 0,76 | 4,47E-01 | 6,32E-01 |
| POLD4 | 881,57 | 0,02 | 0,02 | 0,76 | 4,47E-01 | 6,32E-01 |
| PGAM1 | 1045,10 | -0,02 | 0,02 | -0,76 | 4,47E-01 | 6,32E-01 |
| TEP1 | 3616,23 | -0,02 | 0,03 | -0,76 | 4,48E-01 | 6,33E-01 |

| | | | | | | |
|---|---|---|---|---|---|---|
| RNMT | 1161,19 | 0,02 | 0,02 | 0,76 | 4,48E-01 | 6,33E-01 |
| CLCN6 | 554,21 | 0,03 | 0,04 | 0,76 | 4,48E-01 | 6,33E-01 |
| LEO1 | 518,48 | -0,02 | 0,02 | -0,76 | 4,48E-01 | 6,33E-01 |
| MRPS2 | 177,79 | 0,02 | 0,03 | 0,76 | 4,48E-01 | 6,33E-01 |
| ENDOV | 215,82 | 0,03 | 0,03 | 0,76 | 4,48E-01 | 6,33E-01 |
| TRIM2 | 191,64 | 0,03 | 0,05 | 0,76 | 4,49E-01 | 6,34E-01 |
| DDX50 | 902,58 | 0,03 | 0,03 | 0,76 | 4,49E-01 | 6,34E-01 |
| SIRPG | 160,76 | 0,03 | 0,04 | 0,76 | 4,49E-01 | 6,34E-01 |
| CTSK | 152,57 | -0,03 | 0,04 | -0,76 | 4,49E-01 | 6,34E-01 |
| ACOX3 | 510,34 | -0,01 | 0,02 | -0,76 | 4,49E-01 | 6,34E-01 |
| DDX10 | 820,81 | 0,02 | 0,02 | 0,76 | 4,49E-01 | 6,34E-01 |
| PPP6R1 | 1107,00 | 0,04 | 0,06 | 0,76 | 4,49E-01 | 6,34E-01 |
| MOB3C | 1493,22 | -0,02 | 0,02 | -0,76 | 4,49E-01 | 6,34E-01 |
| NRP1 | 113,10 | -0,06 | 0,08 | -0,76 | 4,50E-01 | 6,34E-01 |
| FITM2 | 234,25 | 0,02 | 0,03 | 0,76 | 4,50E-01 | 6,35E-01 |
| ARMCX3 | 871,00 | -0,02 | 0,02 | -0,76 | 4,50E-01 | 6,35E-01 |
| NELL2 | 1982,32 | 0,03 | 0,05 | 0,76 | 4,50E-01 | 6,35E-01 |
| LYPLA2 | 392,59 | -0,02 | 0,02 | -0,75 | 4,51E-01 | 6,36E-01 |
| ZNF236 | 1437,24 | 0,01 | 0,02 | 0,75 | 4,51E-01 | 6,36E-01 |
| MCM8 | 495,29 | 0,02 | 0,02 | 0,75 | 4,51E-01 | 6,36E-01 |
| ZNF770 | 2576,56 | 0,02 | 0,03 | 0,75 | 4,51E-01 | 6,36E-01 |
| CTSF | 119,85 | 0,03 | 0,04 | 0,75 | 4,52E-01 | 6,36E-01 |
| CEBPZ | 1040,92 | 0,04 | 0,05 | 0,75 | 4,51E-01 | 6,36E-01 |
| NKD1 | 108,65 | 0,06 | 0,08 | 0,75 | 4,51E-01 | 6,36E-01 |
| SHQ1 | 604,19 | 0,01 | 0,02 | 0,75 | 4,52E-01 | 6,36E-01 |
| MRPL33 | 514,43 | 0,02 | 0,03 | 0,75 | 4,52E-01 | 6,36E-01 |
| ZNF439 | 743,62 | 0,02 | 0,03 | 0,75 | 4,51E-01 | 6,36E-01 |
| OSBPL10 | 595,53 | 0,05 | 0,07 | 0,75 | 4,52E-01 | 6,36E-01 |
| CAD | 474,25 | 0,03 | 0,04 | 0,75 | 4,52E-01 | 6,36E-01 |
| SMIM14 | 1875,82 | 0,02 | 0,02 | 0,75 | 4,52E-01 | 6,36E-01 |
| IFI27 | 467,52 | 0,11 | 0,15 | 0,75 | 4,52E-01 | 6,37E-01 |
| CCDC102B | 129,60 | 0,03 | 0,04 | 0,75 | 4,52E-01 | 6,37E-01 |
| TRIM28 | 919,01 | 0,03 | 0,04 | 0,75 | 4,53E-01 | 6,37E-01 |
| PRELID3B | 1323,29 | 0,03 | 0,03 | 0,75 | 4,53E-01 | 6,37E-01 |
| MAPK8 | 1642,07 | 0,02 | 0,02 | 0,75 | 4,53E-01 | 6,38E-01 |
| TRIM8 | 1819,35 | -0,03 | 0,04 | -0,75 | 4,54E-01 | 6,38E-01 |
| ADAM28 | 1663,57 | 0,03 | 0,05 | 0,75 | 4,54E-01 | 6,38E-01 |
| CCR4 | 1268,73 | 0,03 | 0,04 | 0,75 | 4,54E-01 | 6,38E-01 |
| OTUD7B | 595,08 | 0,02 | 0,02 | 0,75 | 4,54E-01 | 6,39E-01 |
| TAS2R3 | 136,20 | -0,03 | 0,04 | -0,75 | 4,54E-01 | 6,39E-01 |
| SECISBP2L | 2414,27 | -0,01 | 0,02 | -0,75 | 4,54E-01 | 6,39E-01 |
| SPTBN1 | 4784,65 | 0,02 | 0,03 | 0,75 | 4,55E-01 | 6,39E-01 |
| CDK17 | 3699,92 | -0,02 | 0,02 | -0,75 | 4,55E-01 | 6,39E-01 |
| CRY1 | 373,71 | 0,02 | 0,02 | 0,75 | 4,55E-01 | 6,39E-01 |
| MED7 | 516,37 | 0,03 | 0,04 | 0,75 | 4,55E-01 | 6,39E-01 |
| LINS1 | 2131,30 | 0,02 | 0,03 | 0,75 | 4,55E-01 | 6,39E-01 |

| | | | | | | |
|---|---|---|---|---|---|---|
| EOLA1 | 360,57 | 0,01 | 0,02 | 0,75 | 4,55E-01 | 6,39E-01 |
| FMNL3 | 923,67 | 0,03 | 0,04 | 0,75 | 4,55E-01 | 6,39E-01 |
| SCAF4 | 797,59 | 0,02 | 0,02 | 0,75 | 4,55E-01 | 6,40E-01 |
| ZEB1 | 2455,56 | -0,02 | 0,02 | -0,75 | 4,56E-01 | 6,40E-01 |
| MYO6 | 360,65 | 0,04 | 0,06 | 0,75 | 4,56E-01 | 6,40E-01 |
| ACSS1 | 1478,54 | 0,02 | 0,02 | 0,75 | 4,56E-01 | 6,40E-01 |
| LRRC37A | 121,85 | -0,04 | 0,05 | -0,75 | 4,56E-01 | 6,40E-01 |
| DDIAS | 164,22 | -0,03 | 0,04 | -0,75 | 4,56E-01 | 6,40E-01 |
| TAS2R30 | 411,76 | 0,03 | 0,04 | 0,75 | 4,56E-01 | 6,40E-01 |
| SLC5A6 | 212,69 | 0,03 | 0,04 | 0,74 | 4,56E-01 | 6,40E-01 |
| EGLN3 | 129,42 | 0,04 | 0,06 | 0,74 | 4,57E-01 | 6,41E-01 |
| PUDP | 249,83 | 0,01 | 0,02 | 0,74 | 4,57E-01 | 6,41E-01 |
| P2RX5 | 159,45 | 0,04 | 0,05 | 0,74 | 4,57E-01 | 6,41E-01 |
| TSTD1 | 586,19 | 0,02 | 0,02 | 0,74 | 4,57E-01 | 6,42E-01 |
| TIMM17B | 517,78 | 0,02 | 0,02 | 0,74 | 4,57E-01 | 6,42E-01 |
| LRRC28 | 552,08 | 0,02 | 0,02 | 0,74 | 4,58E-01 | 6,42E-01 |
| LTBP4 | 260,48 | 0,04 | 0,06 | 0,74 | 4,58E-01 | 6,42E-01 |
| PSD3 | 142,16 | 0,05 | 0,07 | 0,74 | 4,58E-01 | 6,42E-01 |
| VEZF1 | 2368,15 | -0,02 | 0,02 | -0,74 | 4,58E-01 | 6,42E-01 |
| HCFC1 | 634,51 | 0,04 | 0,05 | 0,74 | 4,58E-01 | 6,42E-01 |
| TTC16 | 296,35 | 0,04 | 0,06 | 0,74 | 4,58E-01 | 6,42E-01 |
| C2orf42 | 361,43 | 0,01 | 0,01 | 0,74 | 4,58E-01 | 6,42E-01 |
| CASP6 | 307,22 | 0,02 | 0,03 | 0,74 | 4,58E-01 | 6,42E-01 |
| LSM12 | 201,15 | -0,01 | 0,02 | -0,74 | 4,58E-01 | 6,42E-01 |
| AP1S3 | 126,17 | 0,03 | 0,05 | 0,74 | 4,58E-01 | 6,42E-01 |
| TXNDC17 | 355,19 | 0,03 | 0,04 | 0,74 | 4,59E-01 | 6,42E-01 |
| MFF | 1551,16 | 0,02 | 0,03 | 0,74 | 4,59E-01 | 6,42E-01 |
| SETD3 | 1608,68 | 0,01 | 0,02 | 0,74 | 4,59E-01 | 6,42E-01 |
| ADGRD1 | 182,71 | 0,06 | 0,08 | 0,74 | 4,59E-01 | 6,42E-01 |
| TMEM175 | 263,08 | 0,03 | 0,05 | 0,74 | 4,59E-01 | 6,43E-01 |
| CXCR6 | 304,24 | 0,05 | 0,06 | 0,74 | 4,60E-01 | 6,43E-01 |
| LIMD2 | 6463,60 | 0,02 | 0,02 | 0,74 | 4,60E-01 | 6,43E-01 |
| EIF3D | 3077,73 | 0,02 | 0,02 | 0,74 | 4,60E-01 | 6,43E-01 |
| NIPA1 | 617,22 | 0,02 | 0,03 | 0,74 | 4,60E-01 | 6,44E-01 |
| SDC2 | 87,50 | -0,05 | 0,07 | -0,74 | 4,60E-01 | 6,44E-01 |
| ATP13A2 | 223,70 | 0,04 | 0,06 | 0,74 | 4,60E-01 | 6,44E-01 |
| DDA1 | 251,03 | 0,02 | 0,03 | 0,74 | 4,60E-01 | 6,44E-01 |
| TMED8 | 1870,13 | -0,02 | 0,02 | -0,74 | 4,60E-01 | 6,44E-01 |
| UMPS | 668,74 | 0,02 | 0,02 | 0,74 | 4,61E-01 | 6,44E-01 |
| NUDT2 | 214,53 | 0,02 | 0,03 | 0,74 | 4,61E-01 | 6,44E-01 |
| H2AC13 | 871,75 | 0,04 | 0,05 | 0,74 | 4,61E-01 | 6,44E-01 |
| FNDC3A | 2852,41 | -0,02 | 0,02 | -0,74 | 4,61E-01 | 6,44E-01 |
| SETD9 | 112,68 | 0,03 | 0,05 | 0,74 | 4,61E-01 | 6,44E-01 |
| MAGT1 | 2478,76 | 0,02 | 0,03 | 0,74 | 4,61E-01 | 6,45E-01 |
| CDC25B | 903,00 | 0,04 | 0,05 | 0,74 | 4,62E-01 | 6,45E-01 |
| RGL1 | 135,84 | -0,03 | 0,04 | -0,74 | 4,62E-01 | 6,45E-01 |

| | | | | | | |
|---|---|---|---|---|---|---|
| KIF1C | 483,18 | -0,03 | 0,04 | -0,74 | 4,62E-01 | 6,45E-01 |
| BFAR | 665,94 | 0,01 | 0,02 | 0,74 | 4,62E-01 | 6,45E-01 |
| TMEM219 | 441,06 | 0,01 | 0,02 | 0,74 | 4,62E-01 | 6,45E-01 |
| BEX2 | 91,04 | 0,04 | 0,06 | 0,74 | 4,62E-01 | 6,45E-01 |
| API5 | 2720,59 | 0,02 | 0,03 | 0,74 | 4,62E-01 | 6,45E-01 |
| LRRC23 | 95,34 | 0,03 | 0,04 | 0,74 | 4,62E-01 | 6,45E-01 |
| RUVBL1 | 693,51 | 0,02 | 0,02 | 0,74 | 4,62E-01 | 6,45E-01 |
| SIRT1 | 1484,33 | -0,02 | 0,02 | -0,73 | 4,62E-01 | 6,45E-01 |
| CYB5RL | 142,80 | 0,03 | 0,05 | 0,73 | 4,62E-01 | 6,45E-01 |
| CD200 | 160,09 | 0,05 | 0,07 | 0,73 | 4,63E-01 | 6,45E-01 |
| CHMP6 | 252,85 | 0,02 | 0,03 | 0,73 | 4,63E-01 | 6,45E-01 |
| HGS | 839,62 | -0,04 | 0,05 | -0,73 | 4,63E-01 | 6,45E-01 |
| TMEM94 | 319,37 | 0,04 | 0,05 | 0,73 | 4,63E-01 | 6,45E-01 |
| SIPA1L3 | 574,05 | 0,04 | 0,05 | 0,73 | 4,63E-01 | 6,46E-01 |
| SCRIB | 314,88 | 0,04 | 0,06 | 0,73 | 4,63E-01 | 6,46E-01 |
| TVP23B | 984,23 | -0,02 | 0,03 | -0,73 | 4,63E-01 | 6,46E-01 |
| TMEM131L | 4747,83 | -0,02 | 0,02 | -0,73 | 4,63E-01 | 6,46E-01 |
| FAM234A | 72,85 | 0,04 | 0,05 | 0,73 | 4,64E-01 | 6,46E-01 |
| CELSR1 | 159,83 | 0,07 | 0,10 | 0,73 | 4,64E-01 | 6,47E-01 |
| SEC23A | 2235,46 | -0,01 | 0,02 | -0,73 | 4,64E-01 | 6,47E-01 |
| RGS6 | 186,20 | 0,05 | 0,07 | 0,73 | 4,65E-01 | 6,47E-01 |
| LPXN | 1384,38 | 0,01 | 0,02 | 0,73 | 4,65E-01 | 6,47E-01 |
| DUSP3 | 906,75 | -0,02 | 0,03 | -0,73 | 4,65E-01 | 6,47E-01 |
| CNTROB | 277,66 | 0,02 | 0,03 | 0,73 | 4,65E-01 | 6,47E-01 |
| GTF2A2 | 768,36 | 0,02 | 0,03 | 0,73 | 4,65E-01 | 6,47E-01 |
| ATF7 | 448,51 | -0,04 | 0,05 | -0,73 | 4,65E-01 | 6,47E-01 |
| VPS37C | 312,96 | -0,03 | 0,04 | -0,73 | 4,65E-01 | 6,47E-01 |
| NOL12 | 179,13 | -0,03 | 0,04 | -0,73 | 4,65E-01 | 6,47E-01 |
| HGSNAT | 1173,23 | -0,02 | 0,03 | -0,73 | 4,65E-01 | 6,48E-01 |
| NOC4L | 85,61 | 0,04 | 0,05 | 0,73 | 4,66E-01 | 6,48E-01 |
| SARNP | 195,15 | -0,01 | 0,02 | -0,73 | 4,67E-01 | 6,49E-01 |
| SLC25A3 | 3173,28 | 0,01 | 0,01 | 0,73 | 4,67E-01 | 6,49E-01 |
| LUZP1 | 853,23 | -0,01 | 0,02 | -0,73 | 4,67E-01 | 6,50E-01 |
| PLXDC1 | 161,79 | 0,04 | 0,06 | 0,73 | 4,67E-01 | 6,50E-01 |
| SMC3 | 1600,71 | -0,03 | 0,04 | -0,73 | 4,67E-01 | 6,50E-01 |
| SLC7A6 | 2084,07 | 0,02 | 0,03 | 0,73 | 4,68E-01 | 6,50E-01 |
| NACA | 9274,94 | 0,03 | 0,04 | 0,73 | 4,68E-01 | 6,50E-01 |
| USP16 | 1792,79 | 0,02 | 0,03 | 0,73 | 4,68E-01 | 6,50E-01 |
| ZNF385A | 207,19 | 0,05 | 0,06 | 0,73 | 4,68E-01 | 6,50E-01 |
| SFSWAP | 2561,14 | -0,01 | 0,02 | -0,73 | 4,68E-01 | 6,51E-01 |
| IPO8 | 2458,21 | -0,01 | 0,02 | -0,73 | 4,68E-01 | 6,51E-01 |
| SNRNP27 | 305,66 | 0,02 | 0,03 | 0,72 | 4,68E-01 | 6,51E-01 |
| VAV2 | 286,44 | 0,04 | 0,05 | 0,73 | 4,68E-01 | 6,51E-01 |
| MTREX | 2346,40 | 0,03 | 0,04 | 0,72 | 4,69E-01 | 6,51E-01 |
| PIP5K1C | 327,57 | 0,05 | 0,06 | 0,72 | 4,69E-01 | 6,51E-01 |
| POLDIP2 | 851,99 | 0,01 | 0,02 | 0,72 | 4,69E-01 | 6,51E-01 |

| | | | | | | |
|---|---|---|---|---|---|---|
| ALDOA | 249,20 | -0,02 | 0,03 | -0,72 | 4,69E-01 | 6,51E-01 |
| ZNF8 | 372,58 | -0,02 | 0,03 | -0,72 | 4,69E-01 | 6,51E-01 |
| SMC5 | 2408,84 | 0,02 | 0,03 | 0,72 | 4,69E-01 | 6,52E-01 |
| SMIM8 | 778,02 | -0,02 | 0,03 | -0,72 | 4,70E-01 | 6,52E-01 |
| GNL2 | 725,46 | 0,01 | 0,02 | 0,72 | 4,69E-01 | 6,52E-01 |
| POLQ | 139,13 | 0,04 | 0,05 | 0,72 | 4,70E-01 | 6,52E-01 |
| STPG1 | 100,55 | 0,04 | 0,05 | 0,72 | 4,70E-01 | 6,52E-01 |
| SP140L | 1988,66 | -0,02 | 0,02 | -0,72 | 4,70E-01 | 6,52E-01 |
| XPOT | 1632,13 | 0,02 | 0,02 | 0,72 | 4,70E-01 | 6,52E-01 |
| LUC7L | 1392,60 | 0,02 | 0,03 | 0,72 | 4,70E-01 | 6,52E-01 |
| SLC38A7 | 139,70 | 0,03 | 0,05 | 0,72 | 4,70E-01 | 6,52E-01 |
| SNPH | 159,14 | 0,04 | 0,05 | 0,72 | 4,71E-01 | 6,52E-01 |
| ZNF677 | 173,40 | 0,03 | 0,05 | 0,72 | 4,71E-01 | 6,53E-01 |
| PCSK7 | 4618,83 | 0,02 | 0,03 | 0,72 | 4,71E-01 | 6,53E-01 |
| KLHL3 | 614,91 | 0,03 | 0,04 | 0,72 | 4,71E-01 | 6,53E-01 |
| STT3A | 1406,19 | -0,01 | 0,01 | -0,72 | 4,71E-01 | 6,53E-01 |
| DERL2 | 1210,53 | -0,01 | 0,02 | -0,72 | 4,71E-01 | 6,53E-01 |
| BMP6 | 204,27 | -0,04 | 0,05 | -0,72 | 4,71E-01 | 6,53E-01 |
| MIEF2 | 362,00 | 0,01 | 0,02 | 0,72 | 4,71E-01 | 6,53E-01 |
| ASNSD1 | 955,85 | 0,04 | 0,06 | 0,72 | 4,71E-01 | 6,53E-01 |
| PNPLA7 | 112,32 | 0,07 | 0,09 | 0,72 | 4,71E-01 | 6,53E-01 |
| CAMK1D | 2317,19 | -0,03 | 0,04 | -0,72 | 4,71E-01 | 6,53E-01 |
| CDC23 | 635,58 | 0,01 | 0,02 | 0,72 | 4,72E-01 | 6,53E-01 |
| CEP120 | 2084,37 | 0,02 | 0,03 | 0,72 | 4,72E-01 | 6,53E-01 |
| ZNF507 | 1685,15 | 0,02 | 0,02 | 0,72 | 4,72E-01 | 6,53E-01 |
| GPR137B | 190,81 | 0,02 | 0,03 | 0,72 | 4,72E-01 | 6,53E-01 |
| MSRB3 | 196,46 | -0,03 | 0,05 | -0,72 | 4,72E-01 | 6,53E-01 |
| PUS7 | 165,51 | -0,02 | 0,03 | -0,72 | 4,72E-01 | 6,53E-01 |
| TMEM69 | 367,57 | 0,01 | 0,02 | 0,72 | 4,72E-01 | 6,53E-01 |
| ZNF185 | 1190,73 | -0,03 | 0,05 | -0,72 | 4,72E-01 | 6,53E-01 |
| MED16 | 376,98 | 0,04 | 0,06 | 0,72 | 4,72E-01 | 6,53E-01 |
| BLOC1S3 | 204,76 | 0,03 | 0,04 | 0,72 | 4,72E-01 | 6,54E-01 |
| CAVIN2 | 2225,30 | 0,04 | 0,06 | 0,72 | 4,73E-01 | 6,54E-01 |
| PIGO | 398,71 | 0,02 | 0,03 | 0,72 | 4,73E-01 | 6,54E-01 |
| SLC25A17 | 398,94 | 0,01 | 0,02 | 0,72 | 4,73E-01 | 6,54E-01 |
| LIG3 | 909,17 | 0,02 | 0,02 | 0,72 | 4,73E-01 | 6,54E-01 |
| TCN1 | 133,43 | 0,05 | 0,07 | 0,72 | 4,73E-01 | 6,54E-01 |
| GPATCH3 | 259,37 | -0,01 | 0,02 | -0,72 | 4,73E-01 | 6,54E-01 |
| TMED4 | 1993,68 | 0,01 | 0,01 | 0,72 | 4,73E-01 | 6,54E-01 |
| EML2 | 695,62 | 0,01 | 0,02 | 0,72 | 4,73E-01 | 6,54E-01 |
| BBS7 | 310,68 | 0,02 | 0,03 | 0,72 | 4,74E-01 | 6,54E-01 |
| ARHGEF28 | 99,81 | -0,07 | 0,10 | -0,72 | 4,74E-01 | 6,54E-01 |
| FAM151B | 210,43 | -0,03 | 0,04 | -0,72 | 4,74E-01 | 6,54E-01 |
| PPM1G | 1289,15 | -0,02 | 0,03 | -0,72 | 4,74E-01 | 6,54E-01 |
| KLLN | 77,92 | -0,03 | 0,04 | -0,72 | 4,74E-01 | 6,55E-01 |
| PSMD1 | 1894,77 | -0,01 | 0,02 | -0,72 | 4,75E-01 | 6,55E-01 |

| | | | | | | |
|---|---|---|---|---|---|---|
| WWP1 | 2304,37 | 0,02 | 0,03 | 0,71 | 4,75E-01 | 6,55E-01 |
| RLF | 2272,03 | -0,02 | 0,03 | -0,71 | 4,75E-01 | 6,55E-01 |
| ARHGAP31 | 634,41 | 0,02 | 0,03 | 0,71 | 4,75E-01 | 6,56E-01 |
| CAT | 6682,86 | -0,03 | 0,04 | -0,71 | 4,75E-01 | 6,56E-01 |
| WDR44 | 570,39 | 0,01 | 0,02 | 0,71 | 4,76E-01 | 6,56E-01 |
| DDX27 | 1037,19 | -0,01 | 0,02 | -0,71 | 4,76E-01 | 6,57E-01 |
| KSR1 | 1300,42 | -0,04 | 0,05 | -0,71 | 4,76E-01 | 6,57E-01 |
| GALNT2 | 839,42 | -0,02 | 0,03 | -0,71 | 4,76E-01 | 6,57E-01 |
| CHN2 | 455,04 | -0,03 | 0,04 | -0,71 | 4,77E-01 | 6,58E-01 |
| ZNF225 | 442,31 | 0,02 | 0,02 | 0,71 | 4,77E-01 | 6,58E-01 |
| EML6 | 193,71 | -0,05 | 0,07 | -0,71 | 4,77E-01 | 6,58E-01 |
| ELP2 | 2569,28 | 0,01 | 0,02 | 0,71 | 4,77E-01 | 6,58E-01 |
| EPS8L2 | 88,02 | 0,03 | 0,05 | 0,71 | 4,77E-01 | 6,58E-01 |
| SLFN14 | 228,69 | 0,05 | 0,06 | 0,71 | 4,77E-01 | 6,58E-01 |
| COX19 | 1247,36 | -0,01 | 0,02 | -0,71 | 4,77E-01 | 6,58E-01 |
| C1orf43 | 1354,63 | 0,01 | 0,02 | 0,71 | 4,78E-01 | 6,58E-01 |
| GFOD1 | 1291,09 | -0,02 | 0,03 | -0,71 | 4,78E-01 | 6,58E-01 |
| DDX39A | 782,74 | 0,01 | 0,02 | 0,71 | 4,78E-01 | 6,58E-01 |
| LST1 | 5307,22 | -0,02 | 0,03 | -0,71 | 4,78E-01 | 6,58E-01 |
| ZCCHC8 | 991,75 | 0,01 | 0,02 | 0,71 | 4,78E-01 | 6,58E-01 |
| PDE7A | 7122,50 | 0,02 | 0,03 | 0,71 | 4,78E-01 | 6,58E-01 |
| CCDC61 | 113,82 | 0,03 | 0,05 | 0,71 | 4,78E-01 | 6,58E-01 |
| LOXL2 | 145,31 | -0,03 | 0,04 | -0,71 | 4,78E-01 | 6,58E-01 |
| MACO1 | 743,17 | 0,01 | 0,01 | 0,71 | 4,78E-01 | 6,58E-01 |
| NUP85 | 591,31 | 0,02 | 0,03 | 0,71 | 4,78E-01 | 6,58E-01 |
| IPP | 406,91 | -0,01 | 0,02 | -0,71 | 4,79E-01 | 6,59E-01 |
| MPRIP | 4567,46 | 0,02 | 0,03 | 0,71 | 4,79E-01 | 6,59E-01 |
| PAXIP1 | 717,17 | -0,01 | 0,02 | -0,71 | 4,79E-01 | 6,59E-01 |
| SUPT20H | 3851,17 | -0,02 | 0,03 | -0,71 | 4,79E-01 | 6,59E-01 |
| GSK3A | 448,14 | 0,02 | 0,04 | 0,71 | 4,79E-01 | 6,59E-01 |
| CASP3 | 1173,73 | 0,03 | 0,05 | 0,71 | 4,79E-01 | 6,59E-01 |
| CNTRL | 3742,61 | -0,02 | 0,02 | -0,71 | 4,79E-01 | 6,59E-01 |
| ZNF738 | 294,85 | 0,02 | 0,02 | 0,71 | 4,80E-01 | 6,60E-01 |
| ZNF322 | 533,03 | 0,03 | 0,04 | 0,71 | 4,80E-01 | 6,60E-01 |
| DCTN6 | 193,27 | 0,02 | 0,03 | 0,71 | 4,80E-01 | 6,60E-01 |
| TESPA1 | 1578,84 | 0,02 | 0,03 | 0,71 | 4,80E-01 | 6,60E-01 |
| AP5Z1 | 749,47 | -0,03 | 0,04 | -0,71 | 4,80E-01 | 6,60E-01 |
| MRPL28 | 608,31 | -0,02 | 0,02 | -0,71 | 4,81E-01 | 6,61E-01 |
| MTURN | 3152,31 | -0,02 | 0,03 | -0,71 | 4,81E-01 | 6,61E-01 |
| PSRC1 | 118,79 | -0,02 | 0,03 | -0,71 | 4,81E-01 | 6,61E-01 |
| KDM4C | 4388,53 | 0,02 | 0,02 | 0,71 | 4,81E-01 | 6,61E-01 |
| BUD23 | 487,62 | 0,01 | 0,02 | 0,70 | 4,81E-01 | 6,61E-01 |
| GALK2 | 1197,39 | -0,01 | 0,02 | -0,70 | 4,81E-01 | 6,61E-01 |
| TNFAIP2 | 7831,16 | -0,04 | 0,05 | -0,70 | 4,81E-01 | 6,61E-01 |
| KANSL2 | 1174,88 | 0,01 | 0,02 | 0,70 | 4,81E-01 | 6,61E-01 |
| SRSF3 | 7800,76 | 0,02 | 0,03 | 0,70 | 4,81E-01 | 6,61E-01 |

| | | | | | | |
|---|---|---|---|---|---|---|
| MYCL | 430,21 | 0,02 | 0,03 | 0,70 | 4,81E-01 | 6,61E-01 |
| STON2 | 555,71 | 0,03 | 0,05 | 0,70 | 4,81E-01 | 6,61E-01 |
| KLHL5 | 1360,52 | 0,02 | 0,03 | 0,70 | 4,82E-01 | 6,61E-01 |
| TUBB6 | 167,83 | 0,04 | 0,05 | 0,70 | 4,82E-01 | 6,61E-01 |
| THAP6 | 786,33 | 0,02 | 0,03 | 0,70 | 4,82E-01 | 6,61E-01 |
| YPEL5 | 3593,51 | -0,03 | 0,04 | -0,70 | 4,82E-01 | 6,61E-01 |
| TTC17 | 5114,93 | 0,01 | 0,02 | 0,70 | 4,82E-01 | 6,61E-01 |
| PPP1R10 | 2113,83 | -0,02 | 0,02 | -0,70 | 4,82E-01 | 6,61E-01 |
| MYL6 | 9812,09 | 0,04 | 0,06 | 0,70 | 4,82E-01 | 6,61E-01 |
| MARCKSL1 | 98,53 | 0,04 | 0,05 | 0,70 | 4,82E-01 | 6,61E-01 |
| SLC24A3 | 129,67 | -0,05 | 0,07 | -0,70 | 4,82E-01 | 6,61E-01 |
| XPA | 604,33 | 0,02 | 0,03 | 0,70 | 4,83E-01 | 6,62E-01 |
| LY96 | 574,83 | -0,04 | 0,05 | -0,70 | 4,83E-01 | 6,62E-01 |
| GABBR1 | 634,09 | 0,04 | 0,06 | 0,70 | 4,83E-01 | 6,62E-01 |
| PLA2G4C | 88,42 | 0,07 | 0,10 | 0,70 | 4,83E-01 | 6,62E-01 |
| AZIN1 | 4234,70 | -0,02 | 0,03 | -0,70 | 4,83E-01 | 6,62E-01 |
| MORF4L1 | 4299,51 | -0,01 | 0,02 | -0,70 | 4,83E-01 | 6,62E-01 |
| PAG1 | 8315,70 | 0,02 | 0,02 | 0,70 | 4,83E-01 | 6,62E-01 |
| CLIC1 | 8675,85 | -0,02 | 0,04 | -0,70 | 4,84E-01 | 6,63E-01 |
| ST14 | 254,72 | -0,04 | 0,05 | -0,70 | 4,84E-01 | 6,63E-01 |
| HKDC1 | 177,89 | 0,04 | 0,06 | 0,70 | 4,84E-01 | 6,63E-01 |
| NME7 | 239,54 | 0,02 | 0,03 | 0,70 | 4,84E-01 | 6,63E-01 |
| TMED5 | 3068,76 | 0,02 | 0,03 | 0,70 | 4,84E-01 | 6,63E-01 |
| C20orf204 | 83,44 | 0,04 | 0,06 | 0,70 | 4,84E-01 | 6,63E-01 |
| ZFYVE19 | 379,02 | 0,02 | 0,02 | 0,70 | 4,85E-01 | 6,63E-01 |
| FOXO1 | 2161,67 | 0,01 | 0,02 | 0,70 | 4,85E-01 | 6,63E-01 |
| RIMOC1 | 888,45 | 0,01 | 0,02 | 0,70 | 4,85E-01 | 6,63E-01 |
| MYEF2 | 515,82 | 0,03 | 0,05 | 0,70 | 4,85E-01 | 6,63E-01 |
| RIPOR1 | 777,61 | 0,03 | 0,04 | 0,70 | 4,85E-01 | 6,63E-01 |
| FHIP2B | 277,44 | 0,04 | 0,05 | 0,70 | 4,85E-01 | 6,63E-01 |
| MTG1 | 206,07 | 0,02 | 0,03 | 0,70 | 4,85E-01 | 6,63E-01 |
| PDIA6 | 2220,38 | -0,02 | 0,03 | -0,70 | 4,85E-01 | 6,63E-01 |
| ISOC1 | 369,84 | -0,02 | 0,03 | -0,70 | 4,85E-01 | 6,63E-01 |
| XRCC6 | 4759,69 | -0,02 | 0,03 | -0,70 | 4,85E-01 | 6,64E-01 |
| MFSD1 | 4353,73 | -0,02 | 0,03 | -0,70 | 4,86E-01 | 6,65E-01 |
| UBE3C | 3194,49 | -0,01 | 0,01 | -0,70 | 4,86E-01 | 6,65E-01 |
| PPM1H | 174,08 | -0,04 | 0,05 | -0,70 | 4,87E-01 | 6,65E-01 |
| FBRSL1 | 314,60 | 0,05 | 0,08 | 0,70 | 4,87E-01 | 6,65E-01 |
| SP3 | 9913,85 | -0,02 | 0,03 | -0,69 | 4,87E-01 | 6,66E-01 |
| PAIP2 | 2472,46 | -0,02 | 0,03 | -0,69 | 4,87E-01 | 6,66E-01 |
| ELMOD2 | 1478,08 | 0,03 | 0,04 | 0,69 | 4,87E-01 | 6,66E-01 |
| TUBA1C | 623,19 | -0,02 | 0,02 | -0,69 | 4,87E-01 | 6,66E-01 |
| TATDN2 | 917,64 | -0,02 | 0,02 | -0,69 | 4,88E-01 | 6,66E-01 |
| PABPC4 | 1343,86 | 0,01 | 0,02 | 0,69 | 4,88E-01 | 6,66E-01 |
| FAM227B | 147,01 | 0,03 | 0,05 | 0,69 | 4,88E-01 | 6,66E-01 |
| ZNF491 | 103,26 | 0,02 | 0,03 | 0,69 | 4,88E-01 | 6,66E-01 |

| | | | | | | |
|---|---|---|---|---|---|---|
| LRP12 | 146,23 | 0,03 | 0,04 | 0,69 | 4,88E-01 | 6,66E-01 |
| HLA-A | 40507,42 | -0,03 | 0,05 | -0,69 | 4,88E-01 | 6,66E-01 |
| LGR6 | 112,52 | -0,05 | 0,07 | -0,69 | 4,88E-01 | 6,66E-01 |
| TAS2R19 | 251,41 | 0,03 | 0,04 | 0,69 | 4,88E-01 | 6,66E-01 |
| TMEM220 | 73,73 | 0,04 | 0,05 | 0,69 | 4,88E-01 | 6,66E-01 |
| DENND11 | 3185,99 | 0,02 | 0,02 | 0,69 | 4,88E-01 | 6,66E-01 |
| ARF6 | 4594,54 | 0,02 | 0,02 | 0,69 | 4,89E-01 | 6,66E-01 |
| TMEM30A | 5039,97 | -0,02 | 0,03 | -0,69 | 4,89E-01 | 6,66E-01 |
| CHN1 | 80,33 | 0,04 | 0,06 | 0,69 | 4,89E-01 | 6,66E-01 |
| GOLGA3 | 2644,98 | -0,02 | 0,03 | -0,69 | 4,89E-01 | 6,66E-01 |
| ZNF473 | 308,40 | -0,01 | 0,02 | -0,69 | 4,89E-01 | 6,66E-01 |
| ESAM | 103,58 | 0,04 | 0,06 | 0,69 | 4,89E-01 | 6,66E-01 |
| KCNH3 | 76,87 | -0,06 | 0,08 | -0,69 | 4,89E-01 | 6,66E-01 |
| MICA | 506,15 | -0,02 | 0,02 | -0,69 | 4,89E-01 | 6,67E-01 |
| SDHAF3 | 144,59 | 0,02 | 0,03 | 0,69 | 4,90E-01 | 6,67E-01 |
| ZNF319 | 326,08 | -0,03 | 0,04 | -0,69 | 4,90E-01 | 6,68E-01 |
| KLHL20 | 1066,25 | 0,02 | 0,02 | 0,69 | 4,90E-01 | 6,68E-01 |
| ZKSCAN2 | 188,32 | 0,02 | 0,03 | 0,69 | 4,90E-01 | 6,68E-01 |
| CEBPA | 1260,83 | 0,02 | 0,03 | 0,69 | 4,90E-01 | 6,68E-01 |
| APP | 1699,71 | -0,02 | 0,03 | -0,69 | 4,91E-01 | 6,68E-01 |
| SHOC2 | 4881,71 | -0,02 | 0,04 | -0,69 | 4,91E-01 | 6,68E-01 |
| SOWAHD | 281,61 | -0,02 | 0,03 | -0,69 | 4,91E-01 | 6,68E-01 |
| FASLG | 210,82 | -0,03 | 0,04 | -0,69 | 4,91E-01 | 6,68E-01 |
| ARHGAP45 | 9723,93 | -0,02 | 0,04 | -0,69 | 4,91E-01 | 6,68E-01 |
| TPH1 | 73,48 | 0,04 | 0,06 | 0,69 | 4,91E-01 | 6,68E-01 |
| UBLCP1 | 1021,08 | -0,02 | 0,03 | -0,69 | 4,91E-01 | 6,68E-01 |
| FBLN7 | 149,28 | 0,03 | 0,05 | 0,69 | 4,91E-01 | 6,68E-01 |
| PSMD2 | 2118,77 | -0,01 | 0,02 | -0,69 | 4,91E-01 | 6,68E-01 |
| HAVCR2 | 777,55 | -0,02 | 0,03 | -0,69 | 4,92E-01 | 6,69E-01 |
| GUSB | 502,40 | 0,02 | 0,02 | 0,69 | 4,92E-01 | 6,69E-01 |
| EHHADH | 83,93 | 0,03 | 0,05 | 0,69 | 4,92E-01 | 6,69E-01 |
| RFXANK | 662,32 | 0,02 | 0,02 | 0,69 | 4,92E-01 | 6,69E-01 |
| BAG3 | 96,98 | 0,03 | 0,04 | 0,69 | 4,92E-01 | 6,69E-01 |
| MEPCE | 767,22 | 0,02 | 0,02 | 0,69 | 4,93E-01 | 6,70E-01 |
| ARRDC3 | 10358,65 | -0,03 | 0,05 | -0,69 | 4,93E-01 | 6,70E-01 |
| SFXN3 | 612,38 | 0,02 | 0,02 | 0,69 | 4,93E-01 | 6,70E-01 |
| SYNE3 | 4558,63 | 0,02 | 0,03 | 0,69 | 4,93E-01 | 6,70E-01 |
| ETFB | 327,05 | 0,01 | 0,02 | 0,69 | 4,93E-01 | 6,70E-01 |
| MTHFD2 | 872,00 | 0,03 | 0,04 | 0,69 | 4,93E-01 | 6,70E-01 |
| THOC2 | 2863,43 | 0,02 | 0,03 | 0,69 | 4,93E-01 | 6,70E-01 |
| TSNARE1 | 285,87 | 0,03 | 0,04 | 0,69 | 4,93E-01 | 6,70E-01 |
| QRICH1 | 1596,44 | -0,01 | 0,02 | -0,69 | 4,93E-01 | 6,70E-01 |
| EPHB2 | 75,33 | -0,08 | 0,12 | -0,69 | 4,93E-01 | 6,70E-01 |
| TPX2 | 73,12 | 0,04 | 0,06 | 0,68 | 4,93E-01 | 6,70E-01 |
| FUZ | 307,49 | -0,02 | 0,03 | -0,68 | 4,93E-01 | 6,70E-01 |
| MAPKAPK3 | 2921,40 | -0,02 | 0,03 | -0,68 | 4,94E-01 | 6,70E-01 |

| | | | | | | |
|---|---|---|---|---|---|---|
| PMPCA | 423,40 | 0,02 | 0,02 | 0,68 | 4,94E-01 | 6,70E-01 |
| C1GALT1 | 3201,98 | -0,02 | 0,03 | -0,68 | 4,94E-01 | 6,70E-01 |
| PPP2R5D | 200,85 | -0,03 | 0,04 | -0,68 | 4,94E-01 | 6,70E-01 |
| VEGFA | 400,10 | -0,03 | 0,04 | -0,68 | 4,94E-01 | 6,70E-01 |
| IFT46 | 143,74 | 0,02 | 0,02 | 0,68 | 4,94E-01 | 6,70E-01 |
| SUSD4 | 130,12 | 0,04 | 0,06 | 0,68 | 4,94E-01 | 6,70E-01 |
| CCR3 | 1054,22 | -0,04 | 0,06 | -0,68 | 4,94E-01 | 6,70E-01 |
| ATE1 | 995,32 | 0,01 | 0,02 | 0,68 | 4,94E-01 | 6,71E-01 |
| ZNF250 | 411,81 | 0,01 | 0,02 | 0,68 | 4,95E-01 | 6,71E-01 |
| TAX1BP1 | 4772,53 | -0,03 | 0,04 | -0,68 | 4,95E-01 | 6,71E-01 |
| SMC6 | 1491,96 | 0,02 | 0,04 | 0,68 | 4,95E-01 | 6,71E-01 |
| SPATS2 | 151,27 | -0,03 | 0,04 | -0,68 | 4,95E-01 | 6,71E-01 |
| SGF29 | 258,14 | 0,01 | 0,02 | 0,68 | 4,96E-01 | 6,72E-01 |
| USP38 | 1836,44 | -0,01 | 0,02 | -0,68 | 4,96E-01 | 6,72E-01 |
| SUV39H1 | 141,52 | -0,01 | 0,02 | -0,68 | 4,96E-01 | 6,72E-01 |
| FGFR1 | 143,56 | 0,05 | 0,08 | 0,68 | 4,96E-01 | 6,72E-01 |
| YPEL3 | 2348,77 | 0,04 | 0,05 | 0,68 | 4,96E-01 | 6,72E-01 |
| ABLIM3 | 86,35 | 0,06 | 0,09 | 0,68 | 4,96E-01 | 6,72E-01 |
| CLNK | 80,31 | -0,07 | 0,11 | -0,68 | 4,97E-01 | 6,73E-01 |
| APBB3 | 410,70 | -0,02 | 0,03 | -0,68 | 4,97E-01 | 6,73E-01 |
| DFFB | 436,65 | 0,01 | 0,02 | 0,68 | 4,97E-01 | 6,73E-01 |
| DPYSL2 | 2292,67 | 0,02 | 0,03 | 0,68 | 4,97E-01 | 6,73E-01 |
| FEZ1 | 106,43 | -0,05 | 0,07 | -0,68 | 4,97E-01 | 6,73E-01 |
| ENSG000002 | 224,86 | 0,02 | 0,02 | 0,68 | 4,98E-01 | 6,73E-01 |
| USP47 | 3441,17 | 0,02 | 0,03 | 0,68 | 4,98E-01 | 6,74E-01 |
| SLC25A13 | 661,30 | -0,01 | 0,02 | -0,68 | 4,98E-01 | 6,74E-01 |
| HYAL2 | 163,59 | 0,02 | 0,03 | 0,68 | 4,98E-01 | 6,74E-01 |
| CTDNEP1 | 593,78 | 0,03 | 0,04 | 0,68 | 4,98E-01 | 6,74E-01 |
| ENKD1 | 156,01 | -0,02 | 0,03 | -0,68 | 4,98E-01 | 6,74E-01 |
| TMEM229B | 510,67 | 0,02 | 0,03 | 0,68 | 4,98E-01 | 6,74E-01 |
| IMPA1 | 952,34 | 0,02 | 0,03 | 0,68 | 4,98E-01 | 6,74E-01 |
| PHOSPHO1 | 3175,35 | 0,04 | 0,06 | 0,68 | 4,98E-01 | 6,74E-01 |
| ELOA | 615,47 | -0,01 | 0,02 | -0,68 | 4,99E-01 | 6,74E-01 |
| COG4 | 1422,78 | -0,01 | 0,01 | -0,68 | 4,99E-01 | 6,74E-01 |
| MPEG1 | 7324,16 | -0,02 | 0,03 | -0,68 | 4,99E-01 | 6,74E-01 |
| TNKS1BP1 | 114,07 | 0,04 | 0,07 | 0,68 | 4,99E-01 | 6,74E-01 |
| ZNF337 | 1111,27 | 0,01 | 0,02 | 0,68 | 4,98E-01 | 6,74E-01 |
| SENP1 | 985,19 | 0,01 | 0,01 | 0,68 | 4,99E-01 | 6,74E-01 |
| BRWD1 | 8060,92 | 0,02 | 0,03 | 0,68 | 4,99E-01 | 6,74E-01 |
| PDCD11 | 1176,21 | 0,01 | 0,02 | 0,68 | 4,99E-01 | 6,74E-01 |
| PSMC3 | 965,04 | 0,02 | 0,03 | 0,68 | 4,99E-01 | 6,74E-01 |
| HOXB4 | 77,89 | -0,03 | 0,04 | -0,68 | 4,99E-01 | 6,74E-01 |
| IKZF3 | 4911,91 | 0,02 | 0,03 | 0,68 | 4,99E-01 | 6,74E-01 |
| NOL9 | 909,25 | 0,02 | 0,03 | 0,68 | 4,99E-01 | 6,74E-01 |
| ZNF44 | 1257,51 | 0,01 | 0,02 | 0,67 | 5,00E-01 | 6,75E-01 |
| CCNF | 95,19 | -0,03 | 0,04 | -0,67 | 5,00E-01 | 6,75E-01 |

| | | | | | | |
|---|---|---|---|---|---|---|
| ARL10 | 541,57 | 0,02 | 0,03 | 0,67 | 5,00E-01 | 6,75E-01 |
| ZNF549 | 664,80 | 0,01 | 0,02 | 0,67 | 5,00E-01 | 6,75E-01 |
| TRIB3 | 99,58 | 0,03 | 0,04 | 0,67 | 5,01E-01 | 6,75E-01 |
| DHX9 | 5014,86 | -0,01 | 0,02 | -0,67 | 5,01E-01 | 6,75E-01 |
| RAB6B | 99,99 | 0,05 | 0,08 | 0,67 | 5,01E-01 | 6,75E-01 |
| RPN2 | 1931,13 | -0,01 | 0,02 | -0,67 | 5,01E-01 | 6,75E-01 |
| RNF216 | 2241,54 | -0,01 | 0,02 | -0,67 | 5,01E-01 | 6,75E-01 |
| HSP90AA1 | 3296,14 | -0,04 | 0,06 | -0,67 | 5,01E-01 | 6,75E-01 |
| MYL12B | 4083,41 | -0,03 | 0,04 | -0,67 | 5,01E-01 | 6,75E-01 |
| ZNF213 | 224,15 | 0,03 | 0,04 | 0,67 | 5,01E-01 | 6,75E-01 |
| SLC35F2 | 153,98 | 0,02 | 0,04 | 0,67 | 5,01E-01 | 6,75E-01 |
| SPRTN | 877,46 | -0,02 | 0,02 | -0,67 | 5,01E-01 | 6,75E-01 |
| KCNQ1 | 691,43 | -0,03 | 0,04 | -0,67 | 5,02E-01 | 6,76E-01 |
| GPR35 | 176,31 | 0,03 | 0,05 | 0,67 | 5,02E-01 | 6,76E-01 |
| PNPLA6 | 912,85 | 0,03 | 0,05 | 0,67 | 5,02E-01 | 6,77E-01 |
| PDZD4 | 492,72 | -0,03 | 0,05 | -0,67 | 5,03E-01 | 6,78E-01 |
| ASCL2 | 299,74 | -0,03 | 0,04 | -0,67 | 5,03E-01 | 6,78E-01 |
| TRIM16 | 202,77 | 0,02 | 0,03 | 0,67 | 5,03E-01 | 6,78E-01 |
| PSMD3 | 791,17 | 0,01 | 0,02 | 0,67 | 5,04E-01 | 6,78E-01 |
| KLRK1 | 448,44 | -0,03 | 0,05 | -0,67 | 5,04E-01 | 6,78E-01 |
| CTPS1 | 691,14 | 0,02 | 0,03 | 0,67 | 5,04E-01 | 6,78E-01 |
| SYTL1 | 1813,65 | 0,02 | 0,03 | 0,67 | 5,04E-01 | 6,78E-01 |
| TMBIM4 | 1579,07 | -0,02 | 0,03 | -0,67 | 5,04E-01 | 6,78E-01 |
| ZFTRAF1 | 979,63 | -0,03 | 0,04 | -0,67 | 5,04E-01 | 6,78E-01 |
| TAF10 | 897,99 | -0,01 | 0,02 | -0,67 | 5,04E-01 | 6,79E-01 |
| GSTM1 | 72,70 | 0,22 | 0,33 | 0,67 | 5,05E-01 | 6,79E-01 |
| CRTC1 | 319,51 | 0,04 | 0,05 | 0,67 | 5,05E-01 | 6,79E-01 |
| RELB | 335,65 | 0,02 | 0,04 | 0,67 | 5,05E-01 | 6,79E-01 |
| RAD54B | 73,23 | 0,03 | 0,05 | 0,67 | 5,05E-01 | 6,79E-01 |
| PFN1 | 3571,71 | -0,02 | 0,03 | -0,67 | 5,06E-01 | 6,80E-01 |
| GNE | 532,55 | 0,01 | 0,01 | 0,67 | 5,05E-01 | 6,80E-01 |
| B3GALT4 | 807,53 | 0,01 | 0,02 | 0,67 | 5,06E-01 | 6,80E-01 |
| ELOC | 479,26 | 0,02 | 0,02 | 0,67 | 5,06E-01 | 6,80E-01 |
| DET1 | 213,22 | 0,02 | 0,03 | 0,67 | 5,06E-01 | 6,80E-01 |
| ZNF615 | 465,99 | 0,02 | 0,04 | 0,67 | 5,06E-01 | 6,80E-01 |
| SLC4A10 | 354,37 | 0,06 | 0,09 | 0,66 | 5,06E-01 | 6,80E-01 |
| ZNF135 | 83,30 | 0,04 | 0,07 | 0,66 | 5,06E-01 | 6,80E-01 |
| CHI3L2 | 238,15 | 0,03 | 0,05 | 0,66 | 5,06E-01 | 6,80E-01 |
| SAP30 | 221,46 | 0,02 | 0,03 | 0,66 | 5,07E-01 | 6,80E-01 |
| CDC42SE2 | 10102,16 | 0,02 | 0,03 | 0,66 | 5,06E-01 | 6,80E-01 |
| PTGDR2 | 433,29 | 0,04 | 0,06 | 0,66 | 5,07E-01 | 6,81E-01 |
| CTNND1 | 745,12 | 0,03 | 0,04 | 0,66 | 5,07E-01 | 6,81E-01 |
| NDUFV2 | 468,24 | 0,01 | 0,02 | 0,66 | 5,07E-01 | 6,81E-01 |
| CARD11 | 1112,71 | 0,03 | 0,04 | 0,66 | 5,07E-01 | 6,81E-01 |
| RFC1 | 1793,57 | -0,01 | 0,02 | -0,66 | 5,07E-01 | 6,81E-01 |
| HEMGN | 3100,48 | 0,05 | 0,07 | 0,66 | 5,07E-01 | 6,81E-01 |

| | | | | | | |
|---|---|---|---|---|---|---|
| ZNF618 | 134,15 | -0,04 | 0,07 | -0,66 | 5,08E-01 | 6,81E-01 |
| TERF1 | 965,95 | -0,01 | 0,02 | -0,66 | 5,08E-01 | 6,81E-01 |
| WIPI1 | 930,12 | -0,02 | 0,03 | -0,66 | 5,08E-01 | 6,81E-01 |
| SEC23IP | 978,59 | -0,01 | 0,01 | -0,66 | 5,08E-01 | 6,81E-01 |
| ARL6IP5 | 4394,33 | 0,02 | 0,03 | 0,66 | 5,08E-01 | 6,81E-01 |
| TAF1C | 204,09 | 0,04 | 0,06 | 0,66 | 5,08E-01 | 6,81E-01 |
| PPP2R1A | 1618,33 | -0,01 | 0,02 | -0,66 | 5,08E-01 | 6,81E-01 |
| COMMD1 | 337,40 | 0,01 | 0,02 | 0,66 | 5,08E-01 | 6,82E-01 |
| UBE2H | 6447,92 | 0,03 | 0,04 | 0,66 | 5,09E-01 | 6,83E-01 |
| SEPTIN2 | 5832,48 | -0,01 | 0,02 | -0,66 | 5,09E-01 | 6,83E-01 |
| AGPAT1 | 184,58 | 0,03 | 0,05 | 0,66 | 5,10E-01 | 6,83E-01 |
| PCMT1 | 1481,34 | -0,02 | 0,03 | -0,66 | 5,10E-01 | 6,84E-01 |
| MED29 | 688,12 | 0,01 | 0,02 | 0,66 | 5,10E-01 | 6,84E-01 |
| KDM1A | 1239,16 | 0,01 | 0,02 | 0,66 | 5,10E-01 | 6,84E-01 |
| DVL3 | 355,79 | -0,03 | 0,05 | -0,66 | 5,11E-01 | 6,84E-01 |
| SLC43A3 | 272,01 | 0,02 | 0,03 | 0,66 | 5,11E-01 | 6,84E-01 |
| GTF2E2 | 869,83 | -0,01 | 0,02 | -0,66 | 5,11E-01 | 6,84E-01 |
| COPG2 | 414,13 | 0,01 | 0,02 | 0,66 | 5,11E-01 | 6,84E-01 |
| ULK1 | 700,12 | -0,05 | 0,07 | -0,66 | 5,11E-01 | 6,84E-01 |
| RBFA | 195,84 | 0,02 | 0,02 | 0,66 | 5,11E-01 | 6,84E-01 |
| EPRS1 | 2230,18 | 0,02 | 0,03 | 0,66 | 5,10E-01 | 6,84E-01 |
| B4GALNT3 | 169,32 | 0,06 | 0,10 | 0,66 | 5,10E-01 | 6,84E-01 |
| CBLB | 2910,28 | 0,02 | 0,03 | 0,66 | 5,11E-01 | 6,84E-01 |
| POM121C | 525,69 | -0,03 | 0,05 | -0,66 | 5,11E-01 | 6,84E-01 |
| EIPR1 | 686,26 | -0,01 | 0,02 | -0,66 | 5,11E-01 | 6,84E-01 |
| PDZD11 | 100,58 | 0,01 | 0,02 | 0,66 | 5,11E-01 | 6,84E-01 |
| NAA60 | 544,20 | -0,02 | 0,03 | -0,66 | 5,11E-01 | 6,84E-01 |
| RBFOX2 | 81,60 | -0,07 | 0,10 | -0,66 | 5,11E-01 | 6,84E-01 |
| TNRC6A | 3054,82 | -0,02 | 0,02 | -0,66 | 5,11E-01 | 6,84E-01 |
| PC | 81,12 | 0,05 | 0,07 | 0,66 | 5,11E-01 | 6,84E-01 |
| ZDHHC17 | 4286,72 | -0,02 | 0,03 | -0,66 | 5,11E-01 | 6,84E-01 |
| PURB | 3965,48 | 0,02 | 0,03 | 0,66 | 5,12E-01 | 6,84E-01 |
| UCHL3 | 103,40 | 0,02 | 0,03 | 0,66 | 5,12E-01 | 6,84E-01 |
| LRIG1 | 1115,23 | 0,02 | 0,02 | 0,66 | 5,12E-01 | 6,84E-01 |
| GOLGA8B | 1453,35 | 0,03 | 0,05 | 0,65 | 5,12E-01 | 6,85E-01 |
| VDAC1 | 796,48 | 0,01 | 0,02 | 0,65 | 5,13E-01 | 6,85E-01 |
| ENOSF1 | 940,71 | 0,03 | 0,04 | 0,65 | 5,13E-01 | 6,85E-01 |
| MRGBP | 378,49 | 0,01 | 0,02 | 0,65 | 5,13E-01 | 6,86E-01 |
| SH3BP2 | 4251,01 | -0,03 | 0,05 | -0,65 | 5,13E-01 | 6,86E-01 |
| RAB3IP | 883,47 | 0,02 | 0,03 | 0,65 | 5,13E-01 | 6,86E-01 |
| CCDC102A | 198,37 | -0,02 | 0,03 | -0,65 | 5,14E-01 | 6,86E-01 |
| CEP126 | 177,55 | 0,03 | 0,04 | 0,65 | 5,14E-01 | 6,86E-01 |
| NKTR | 13845,56 | 0,02 | 0,02 | 0,65 | 5,14E-01 | 6,86E-01 |
| EHMT2 | 527,37 | 0,02 | 0,03 | 0,65 | 5,14E-01 | 6,86E-01 |
| PPIAL4H | 84,51 | 0,05 | 0,07 | 0,65 | 5,14E-01 | 6,86E-01 |
| EIF2B4 | 401,04 | 0,01 | 0,02 | 0,65 | 5,14E-01 | 6,86E-01 |

| | | | | | | |
|---|---|---|---|---|---|---|
| CTC1 | 1970,64 | -0,02 | 0,04 | -0,65 | 5,14E-01 | 6,86E-01 |
| RBM7 | 717,16 | 0,02 | 0,03 | 0,65 | 5,14E-01 | 6,86E-01 |
| PER1 | 161,86 | 0,05 | 0,07 | 0,65 | 5,14E-01 | 6,86E-01 |
| FBXO28 | 1617,54 | 0,02 | 0,02 | 0,65 | 5,14E-01 | 6,86E-01 |
| MAP7D3 | 313,26 | 0,01 | 0,02 | 0,65 | 5,14E-01 | 6,86E-01 |
| ZC3H4 | 935,24 | -0,02 | 0,03 | -0,65 | 5,15E-01 | 6,87E-01 |
| CTSC | 6753,07 | -0,02 | 0,03 | -0,65 | 5,15E-01 | 6,87E-01 |
| C15orf39 | 1772,36 | -0,04 | 0,06 | -0,65 | 5,15E-01 | 6,87E-01 |
| RAB8B | 7128,81 | -0,03 | 0,04 | -0,65 | 5,16E-01 | 6,87E-01 |
| COPS7B | 595,19 | -0,02 | 0,03 | -0,65 | 5,16E-01 | 6,87E-01 |
| NOTCH4 | 75,79 | 0,05 | 0,07 | 0,65 | 5,16E-01 | 6,87E-01 |
| C13orf46 | 266,61 | 0,04 | 0,06 | 0,65 | 5,16E-01 | 6,87E-01 |
| SH2B1 | 477,42 | 0,03 | 0,05 | 0,65 | 5,16E-01 | 6,87E-01 |
| HADH | 479,28 | 0,02 | 0,03 | 0,65 | 5,16E-01 | 6,87E-01 |
| TRIP6 | 123,91 | -0,02 | 0,04 | -0,65 | 5,16E-01 | 6,87E-01 |
| SSPN | 89,12 | -0,04 | 0,06 | -0,65 | 5,16E-01 | 6,87E-01 |
| ATG12 | 3121,57 | 0,02 | 0,03 | 0,65 | 5,16E-01 | 6,87E-01 |
| NEMP1 | 910,72 | 0,02 | 0,02 | 0,65 | 5,16E-01 | 6,87E-01 |
| IL2RA | 335,89 | 0,02 | 0,04 | 0,65 | 5,16E-01 | 6,88E-01 |
| STARD5 | 84,35 | 0,03 | 0,04 | 0,65 | 5,17E-01 | 6,88E-01 |
| ABCB9 | 90,55 | 0,05 | 0,07 | 0,65 | 5,16E-01 | 6,88E-01 |
| UBE2Z | 1310,08 | -0,02 | 0,03 | -0,65 | 5,17E-01 | 6,88E-01 |
| ADGRG1 | 642,84 | -0,04 | 0,07 | -0,65 | 5,17E-01 | 6,88E-01 |
| MORF4L2 | 1255,44 | 0,02 | 0,03 | 0,65 | 5,17E-01 | 6,88E-01 |
| MLEC | 1982,12 | -0,01 | 0,02 | -0,65 | 5,17E-01 | 6,88E-01 |
| FYTTD1 | 1747,83 | 0,02 | 0,03 | 0,65 | 5,17E-01 | 6,88E-01 |
| RINL | 931,17 | 0,01 | 0,02 | 0,65 | 5,17E-01 | 6,88E-01 |
| BBS9 | 876,19 | 0,02 | 0,02 | 0,65 | 5,17E-01 | 6,88E-01 |
| GGA2 | 2423,88 | 0,02 | 0,02 | 0,65 | 5,18E-01 | 6,88E-01 |
| WWC2 | 324,05 | 0,02 | 0,03 | 0,65 | 5,18E-01 | 6,89E-01 |
| ITGA3 | 96,04 | 0,05 | 0,07 | 0,65 | 5,18E-01 | 6,89E-01 |
| PCBP3 | 100,38 | -0,05 | 0,08 | -0,65 | 5,18E-01 | 6,89E-01 |
| ZSCAN5A | 283,33 | 0,02 | 0,03 | 0,65 | 5,18E-01 | 6,89E-01 |
| RPL22 | 2759,58 | 0,03 | 0,04 | 0,65 | 5,18E-01 | 6,89E-01 |
| HOMER2 | 198,46 | 0,03 | 0,04 | 0,65 | 5,18E-01 | 6,89E-01 |
| TAFAZZIN | 619,57 | 0,02 | 0,03 | 0,65 | 5,18E-01 | 6,89E-01 |
| ZC3H18 | 671,44 | -0,02 | 0,04 | -0,65 | 5,18E-01 | 6,89E-01 |
| ABHD17A | 3682,82 | 0,02 | 0,02 | 0,65 | 5,19E-01 | 6,89E-01 |
| MTCL2 | 296,43 | 0,04 | 0,06 | 0,65 | 5,19E-01 | 6,89E-01 |
| CCDC28B | 124,31 | 0,02 | 0,03 | 0,65 | 5,19E-01 | 6,89E-01 |
| PEX6 | 773,00 | -0,02 | 0,03 | -0,65 | 5,19E-01 | 6,89E-01 |
| SCP2 | 1710,80 | 0,02 | 0,03 | 0,64 | 5,19E-01 | 6,89E-01 |
| ZNF518B | 1822,24 | -0,02 | 0,03 | -0,64 | 5,19E-01 | 6,89E-01 |
| HMGN2 | 3941,02 | 0,01 | 0,02 | 0,64 | 5,19E-01 | 6,89E-01 |
| MIEF1 | 565,55 | 0,01 | 0,02 | 0,64 | 5,20E-01 | 6,91E-01 |
| CLECL1P | 135,84 | 0,04 | 0,07 | 0,64 | 5,21E-01 | 6,91E-01 |

| | | | | | | |
|---|---|---|---|---|---|---|
| GM2A | 1404,97 | -0,03 | 0,04 | -0,64 | 5,21E-01 | 6,91E-01 |
| CNNM3 | 647,05 | -0,02 | 0,03 | -0,64 | 5,21E-01 | 6,91E-01 |
| CLDND2 | 177,09 | 0,03 | 0,05 | 0,64 | 5,21E-01 | 6,91E-01 |
| KHDC4 | 1567,97 | 0,01 | 0,02 | 0,64 | 5,21E-01 | 6,92E-01 |
| OBSCN | 1510,92 | 0,04 | 0,06 | 0,64 | 5,22E-01 | 6,92E-01 |
| PLEKHB1 | 166,18 | 0,03 | 0,04 | 0,64 | 5,22E-01 | 6,92E-01 |
| TMTC4 | 550,84 | -0,02 | 0,03 | -0,64 | 5,22E-01 | 6,92E-01 |
| PTOV1 | 850,13 | -0,02 | 0,03 | -0,64 | 5,22E-01 | 6,92E-01 |
| FMO5 | 341,95 | 0,02 | 0,03 | 0,64 | 5,22E-01 | 6,92E-01 |
| RBM28 | 1066,34 | -0,02 | 0,03 | -0,64 | 5,22E-01 | 6,92E-01 |
| ADAM22 | 365,33 | 0,02 | 0,04 | 0,64 | 5,23E-01 | 6,92E-01 |
| WDR7 | 2942,05 | 0,01 | 0,02 | 0,64 | 5,23E-01 | 6,92E-01 |
| CNTLN | 425,43 | 0,02 | 0,04 | 0,64 | 5,22E-01 | 6,92E-01 |
| GDE1 | 1737,60 | -0,02 | 0,03 | -0,64 | 5,23E-01 | 6,92E-01 |
| APPBP2 | 2155,39 | -0,01 | 0,02 | -0,64 | 5,23E-01 | 6,92E-01 |
| IFT20 | 320,76 | 0,02 | 0,02 | 0,64 | 5,22E-01 | 6,92E-01 |
| MPP1 | 9899,48 | 0,04 | 0,06 | 0,64 | 5,22E-01 | 6,92E-01 |
| RNFT1 | 686,39 | 0,02 | 0,03 | 0,64 | 5,22E-01 | 6,92E-01 |
| ERCC6L2 | 2206,61 | 0,02 | 0,03 | 0,64 | 5,22E-01 | 6,92E-01 |
| FBLN5 | 105,58 | 0,04 | 0,06 | 0,64 | 5,23E-01 | 6,92E-01 |
| FAAH2 | 263,52 | -0,03 | 0,05 | -0,64 | 5,23E-01 | 6,92E-01 |
| BRPF1 | 830,22 | -0,01 | 0,02 | -0,64 | 5,23E-01 | 6,92E-01 |
| ZNF14 | 539,90 | 0,03 | 0,05 | 0,64 | 5,23E-01 | 6,92E-01 |
| L3MBTL4 | 283,18 | -0,03 | 0,04 | -0,64 | 5,23E-01 | 6,93E-01 |
| SIDT1 | 1656,84 | -0,02 | 0,03 | -0,64 | 5,23E-01 | 6,93E-01 |
| TAPT1 | 1943,78 | -0,01 | 0,02 | -0,64 | 5,24E-01 | 6,93E-01 |
| OXNAD1 | 5486,54 | 0,02 | 0,04 | 0,64 | 5,24E-01 | 6,94E-01 |
| SNRNP48 | 1104,41 | 0,02 | 0,03 | 0,64 | 5,24E-01 | 6,94E-01 |
| PLOD3 | 267,02 | 0,02 | 0,04 | 0,64 | 5,25E-01 | 6,94E-01 |
| PGRMC1 | 1059,04 | 0,03 | 0,04 | 0,64 | 5,25E-01 | 6,94E-01 |
| DLG4 | 165,26 | 0,04 | 0,06 | 0,64 | 5,25E-01 | 6,94E-01 |
| GMEB2 | 421,49 | -0,02 | 0,04 | -0,64 | 5,25E-01 | 6,94E-01 |
| GOLGA8A | 1236,09 | 0,03 | 0,04 | 0,64 | 5,25E-01 | 6,94E-01 |
| SF3B4 | 183,66 | -0,02 | 0,03 | -0,64 | 5,25E-01 | 6,94E-01 |
| HDC | 292,27 | -0,07 | 0,11 | -0,64 | 5,25E-01 | 6,94E-01 |
| FAM114A1 | 106,31 | -0,02 | 0,04 | -0,64 | 5,25E-01 | 6,94E-01 |
| CHI3L1 | 2301,64 | -0,05 | 0,08 | -0,63 | 5,26E-01 | 6,94E-01 |
| AP1M1 | 1240,74 | -0,02 | 0,03 | -0,63 | 5,26E-01 | 6,94E-01 |
| LLGL1 | 293,23 | 0,02 | 0,04 | 0,63 | 5,26E-01 | 6,95E-01 |
| ZNF496 | 215,69 | -0,04 | 0,06 | -0,63 | 5,26E-01 | 6,95E-01 |
| SLC26A6 | 255,79 | 0,02 | 0,04 | 0,63 | 5,26E-01 | 6,95E-01 |
| AGBL5 | 115,19 | 0,03 | 0,04 | 0,63 | 5,26E-01 | 6,95E-01 |
| PTCRA | 163,93 | 0,04 | 0,06 | 0,63 | 5,26E-01 | 6,95E-01 |
| FAM89A | 102,56 | 0,02 | 0,04 | 0,63 | 5,26E-01 | 6,95E-01 |
| GMNN | 97,10 | 0,02 | 0,03 | 0,63 | 5,27E-01 | 6,95E-01 |
| BORCS8 | 303,89 | -0,03 | 0,05 | -0,63 | 5,27E-01 | 6,96E-01 |

| | | | | | | |
|---|---|---|---|---|---|---|
| NRP2 | 94,36 | 0,06 | 0,09 | 0,63 | 5,27E-01 | 6,96E-01 |
| TCTN2 | 90,30 | 0,03 | 0,05 | 0,63 | 5,28E-01 | 6,96E-01 |
| SMIM7 | 1108,66 | 0,01 | 0,02 | 0,63 | 5,28E-01 | 6,96E-01 |
| SEPHS2 | 567,89 | -0,02 | 0,03 | -0,63 | 5,27E-01 | 6,96E-01 |
| ZBTB8OS | 462,31 | 0,01 | 0,02 | 0,63 | 5,28E-01 | 6,96E-01 |
| DGKA | 3565,16 | 0,02 | 0,03 | 0,63 | 5,28E-01 | 6,96E-01 |
| MGST2 | 200,51 | -0,02 | 0,03 | -0,63 | 5,28E-01 | 6,97E-01 |
| OSCAR | 340,47 | -0,03 | 0,05 | -0,63 | 5,28E-01 | 6,97E-01 |
| RABGAP1L | 9638,84 | -0,01 | 0,02 | -0,63 | 5,28E-01 | 6,97E-01 |
| CXCL5 | 327,59 | 0,05 | 0,07 | 0,63 | 5,29E-01 | 6,97E-01 |
| ZNF431 | 2320,95 | 0,01 | 0,02 | 0,63 | 5,29E-01 | 6,97E-01 |
| ATOSB | 1359,11 | 0,03 | 0,04 | 0,63 | 5,29E-01 | 6,97E-01 |
| LRFN1 | 119,92 | -0,04 | 0,07 | -0,63 | 5,29E-01 | 6,98E-01 |
| CDK5RAP1 | 797,45 | 0,01 | 0,01 | 0,63 | 5,29E-01 | 6,98E-01 |
| USP1 | 1592,48 | 0,03 | 0,05 | 0,63 | 5,30E-01 | 6,98E-01 |
| MTERF2 | 382,83 | 0,02 | 0,02 | 0,63 | 5,30E-01 | 6,98E-01 |
| BLOC1S2 | 651,41 | 0,03 | 0,05 | 0,63 | 5,30E-01 | 6,98E-01 |
| EMC3 | 3010,14 | 0,03 | 0,05 | 0,63 | 5,30E-01 | 6,98E-01 |
| CLPX | 1867,78 | -0,01 | 0,02 | -0,63 | 5,30E-01 | 6,98E-01 |
| SIMC1 | 280,23 | 0,02 | 0,03 | 0,63 | 5,30E-01 | 6,98E-01 |
| ACAP1 | 2641,19 | -0,02 | 0,04 | -0,63 | 5,30E-01 | 6,98E-01 |
| KIF2A | 2745,27 | 0,02 | 0,03 | 0,63 | 5,31E-01 | 6,99E-01 |
| PON2 | 247,26 | 0,02 | 0,03 | 0,63 | 5,31E-01 | 6,99E-01 |
| CKAP2 | 439,99 | 0,02 | 0,03 | 0,63 | 5,31E-01 | 6,99E-01 |
| PLXNA3 | 323,68 | 0,04 | 0,06 | 0,63 | 5,31E-01 | 6,99E-01 |
| INPP5E | 167,00 | 0,03 | 0,06 | 0,63 | 5,31E-01 | 6,99E-01 |
| ADGRB1 | 87,90 | -0,07 | 0,12 | -0,63 | 5,31E-01 | 6,99E-01 |
| BORCS5 | 578,80 | -0,02 | 0,03 | -0,63 | 5,32E-01 | 7,00E-01 |
| CUEDC1 | 327,75 | -0,03 | 0,05 | -0,63 | 5,32E-01 | 7,00E-01 |
| PTCD1 | 102,97 | 0,02 | 0,03 | 0,63 | 5,32E-01 | 7,00E-01 |
| ZNF276 | 3931,25 | -0,01 | 0,02 | -0,63 | 5,32E-01 | 7,00E-01 |
| CDCA7 | 73,44 | 0,03 | 0,06 | 0,62 | 5,32E-01 | 7,00E-01 |
| LRIG2 | 1304,85 | 0,01 | 0,02 | 0,62 | 5,32E-01 | 7,00E-01 |
| DNAJC1 | 698,88 | -0,02 | 0,02 | -0,62 | 5,32E-01 | 7,00E-01 |
| ENPP5 | 128,34 | 0,03 | 0,05 | 0,62 | 5,33E-01 | 7,00E-01 |
| RNF167 | 378,63 | -0,02 | 0,04 | -0,62 | 5,33E-01 | 7,01E-01 |
| PTP4A3 | 522,56 | -0,02 | 0,04 | -0,62 | 5,33E-01 | 7,01E-01 |
| MATK | 726,33 | 0,03 | 0,04 | 0,62 | 5,33E-01 | 7,01E-01 |
| ZNF212 | 391,60 | -0,01 | 0,02 | -0,62 | 5,33E-01 | 7,01E-01 |
| PPWD1 | 2047,96 | 0,02 | 0,03 | 0,62 | 5,33E-01 | 7,01E-01 |
| PRKAA1 | 3810,25 | -0,01 | 0,02 | -0,62 | 5,33E-01 | 7,01E-01 |
| GPRASP3 | 97,87 | -0,02 | 0,04 | -0,62 | 5,34E-01 | 7,01E-01 |
| BLNK | 309,68 | 0,03 | 0,05 | 0,62 | 5,34E-01 | 7,01E-01 |
| CYP4F12 | 189,18 | -0,04 | 0,06 | -0,62 | 5,34E-01 | 7,01E-01 |
| PARP2 | 122,10 | 0,02 | 0,03 | 0,62 | 5,34E-01 | 7,01E-01 |
| CCR2 | 3830,72 | 0,02 | 0,04 | 0,62 | 5,34E-01 | 7,01E-01 |

| | | | | | | |
|---|---|---|---|---|---|---|
| KXD1 | 732,43 | 0,02 | 0,03 | 0,62 | 5,34E-01 | 7,01E-01 |
| CCP110 | 959,16 | -0,02 | 0,03 | -0,62 | 5,34E-01 | 7,02E-01 |
| FUBP3 | 1346,35 | 0,01 | 0,01 | 0,62 | 5,35E-01 | 7,02E-01 |
| SRRM1 | 2015,77 | 0,01 | 0,02 | 0,62 | 5,35E-01 | 7,02E-01 |
| ZBTB2 | 417,47 | -0,01 | 0,02 | -0,62 | 5,35E-01 | 7,02E-01 |
| JUP | 104,76 | 0,06 | 0,10 | 0,62 | 5,35E-01 | 7,02E-01 |
| MROH6 | 77,72 | 0,04 | 0,07 | 0,62 | 5,35E-01 | 7,02E-01 |
| MVK | 189,39 | 0,02 | 0,04 | 0,62 | 5,36E-01 | 7,03E-01 |
| EMD | 204,80 | 0,02 | 0,03 | 0,62 | 5,36E-01 | 7,03E-01 |
| EAPP | 1021,61 | -0,02 | 0,03 | -0,62 | 5,36E-01 | 7,03E-01 |
| ZFYVE26 | 1918,96 | -0,02 | 0,03 | -0,62 | 5,36E-01 | 7,03E-01 |
| TBX21 | 678,07 | -0,03 | 0,05 | -0,62 | 5,37E-01 | 7,03E-01 |
| PDE5A | 943,55 | 0,03 | 0,05 | 0,62 | 5,37E-01 | 7,03E-01 |
| LRP8 | 334,78 | 0,02 | 0,03 | 0,62 | 5,37E-01 | 7,04E-01 |
| PALLD | 197,65 | -0,03 | 0,05 | -0,62 | 5,37E-01 | 7,04E-01 |
| BICD1 | 437,46 | 0,02 | 0,03 | 0,62 | 5,37E-01 | 7,04E-01 |
| SUMO2 | 2573,79 | 0,02 | 0,03 | 0,62 | 5,38E-01 | 7,04E-01 |
| C1GALT1C1 | 468,87 | -0,03 | 0,04 | -0,62 | 5,38E-01 | 7,04E-01 |
| FBXW7 | 3966,83 | 0,01 | 0,02 | 0,62 | 5,38E-01 | 7,05E-01 |
| ZNF45 | 732,18 | 0,01 | 0,02 | 0,62 | 5,38E-01 | 7,05E-01 |
| WHAMM | 960,07 | 0,01 | 0,02 | 0,62 | 5,38E-01 | 7,05E-01 |
| CLN3 | 366,00 | -0,02 | 0,04 | -0,62 | 5,38E-01 | 7,05E-01 |
| ARHGEF3 | 2719,22 | -0,01 | 0,02 | -0,61 | 5,39E-01 | 7,05E-01 |
| SAP18 | 2376,12 | -0,02 | 0,03 | -0,61 | 5,39E-01 | 7,05E-01 |
| PCIF1 | 948,02 | -0,02 | 0,03 | -0,61 | 5,39E-01 | 7,05E-01 |
| DHX40 | 1491,67 | -0,02 | 0,03 | -0,61 | 5,39E-01 | 7,05E-01 |
| PMS2 | 452,40 | 0,01 | 0,01 | 0,61 | 5,39E-01 | 7,05E-01 |
| PPP6C | 1806,11 | -0,01 | 0,02 | -0,61 | 5,39E-01 | 7,05E-01 |
| VPS26A | 852,42 | 0,02 | 0,03 | 0,61 | 5,39E-01 | 7,06E-01 |
| ANXA1 | 5012,77 | 0,03 | 0,05 | 0,61 | 5,40E-01 | 7,06E-01 |
| SHC1 | 674,55 | -0,02 | 0,03 | -0,61 | 5,40E-01 | 7,06E-01 |
| PPP1R18 | 1467,70 | -0,04 | 0,06 | -0,61 | 5,40E-01 | 7,07E-01 |
| PRPF38A | 1451,33 | -0,01 | 0,02 | -0,61 | 5,41E-01 | 7,07E-01 |
| DNAH10 | 112,43 | -0,07 | 0,12 | -0,61 | 5,41E-01 | 7,07E-01 |
| GPR89A | 374,58 | 0,02 | 0,03 | 0,61 | 5,41E-01 | 7,07E-01 |
| GAPDH | 10822,05 | 0,02 | 0,02 | 0,61 | 5,41E-01 | 7,07E-01 |
| TMEM165 | 3100,84 | -0,01 | 0,02 | -0,61 | 5,40E-01 | 7,07E-01 |
| ZNF621 | 814,12 | 0,01 | 0,02 | 0,61 | 5,40E-01 | 7,07E-01 |
| SLAMF7 | 1103,71 | -0,02 | 0,04 | -0,61 | 5,41E-01 | 7,07E-01 |
| BSDC1 | 2694,08 | -0,01 | 0,02 | -0,61 | 5,41E-01 | 7,07E-01 |
| B3GNT9 | 108,76 | -0,02 | 0,04 | -0,61 | 5,41E-01 | 7,07E-01 |
| DBR1 | 634,59 | 0,01 | 0,02 | 0,61 | 5,41E-01 | 7,07E-01 |
| ADA | 322,40 | 0,02 | 0,03 | 0,61 | 5,41E-01 | 7,07E-01 |
| NAPB | 992,05 | -0,01 | 0,02 | -0,61 | 5,41E-01 | 7,07E-01 |
| MVD | 377,35 | 0,02 | 0,03 | 0,61 | 5,41E-01 | 7,07E-01 |
| HM13 | 1218,13 | -0,03 | 0,04 | -0,61 | 5,41E-01 | 7,07E-01 |

| | | | | | | |
|---|---|---|---|---|---|---|
| PRMT9 | 512,08 | 0,01 | 0,02 | 0,61 | 5,41E-01 | 7,07E-01 |
| PRICKLE3 | 96,80 | -0,03 | 0,05 | -0,61 | 5,42E-01 | 7,07E-01 |
| LDB1 | 1247,64 | -0,02 | 0,03 | -0,61 | 5,42E-01 | 7,07E-01 |
| DZANK1 | 159,03 | -0,03 | 0,04 | -0,61 | 5,42E-01 | 7,07E-01 |
| NOLC1 | 586,20 | 0,01 | 0,02 | 0,61 | 5,42E-01 | 7,07E-01 |
| NOMO1 | 352,23 | -0,02 | 0,04 | -0,61 | 5,42E-01 | 7,07E-01 |
| ZNF70 | 200,76 | 0,02 | 0,04 | 0,61 | 5,43E-01 | 7,08E-01 |
| PICK1 | 283,71 | 0,02 | 0,03 | 0,61 | 5,43E-01 | 7,08E-01 |
| YPEL2 | 2144,06 | -0,01 | 0,02 | -0,61 | 5,43E-01 | 7,08E-01 |
| SCAMP2 | 901,37 | -0,02 | 0,03 | -0,61 | 5,43E-01 | 7,08E-01 |
| TRIM23 | 1772,01 | 0,02 | 0,04 | 0,61 | 5,43E-01 | 7,08E-01 |
| FAM111B | 77,07 | 0,05 | 0,08 | 0,61 | 5,43E-01 | 7,08E-01 |
| EPN2 | 287,57 | -0,03 | 0,05 | -0,61 | 5,43E-01 | 7,08E-01 |
| NCOA4 | 39238,33 | 0,03 | 0,04 | 0,61 | 5,44E-01 | 7,09E-01 |
| SORCS3 | 140,45 | -0,05 | 0,08 | -0,61 | 5,44E-01 | 7,09E-01 |
| PUS10 | 509,49 | -0,02 | 0,03 | -0,61 | 5,44E-01 | 7,09E-01 |
| RABL2B | 229,77 | 0,03 | 0,04 | 0,61 | 5,44E-01 | 7,09E-01 |
| SLC9A7 | 905,81 | 0,02 | 0,04 | 0,61 | 5,44E-01 | 7,09E-01 |
| DHX15 | 5246,35 | 0,02 | 0,03 | 0,61 | 5,44E-01 | 7,09E-01 |
| STAM2 | 2468,83 | -0,02 | 0,03 | -0,61 | 5,44E-01 | 7,09E-01 |
| LRRC47 | 945,33 | 0,01 | 0,01 | 0,61 | 5,45E-01 | 7,09E-01 |
| PROS1 | 191,34 | -0,05 | 0,07 | -0,61 | 5,45E-01 | 7,09E-01 |
| ZNF766 | 742,81 | 0,01 | 0,02 | 0,61 | 5,45E-01 | 7,09E-01 |
| CD9 | 695,63 | 0,04 | 0,06 | 0,61 | 5,45E-01 | 7,09E-01 |
| HSPE1 | 212,71 | 0,02 | 0,04 | 0,61 | 5,45E-01 | 7,09E-01 |
| TRAK1 | 1273,85 | -0,02 | 0,03 | -0,61 | 5,45E-01 | 7,09E-01 |
| RMDN3 | 502,20 | 0,01 | 0,02 | 0,61 | 5,45E-01 | 7,09E-01 |
| ADARB2 | 120,25 | -0,10 | 0,17 | -0,61 | 5,45E-01 | 7,10E-01 |
| ZFHX3 | 411,51 | 0,03 | 0,06 | 0,60 | 5,45E-01 | 7,10E-01 |
| SOS1 | 3107,75 | 0,02 | 0,03 | 0,60 | 5,46E-01 | 7,10E-01 |
| GPR157 | 181,55 | 0,02 | 0,04 | 0,60 | 5,46E-01 | 7,10E-01 |
| TIAM1 | 2169,60 | 0,02 | 0,03 | 0,60 | 5,46E-01 | 7,10E-01 |
| ATL1 | 125,59 | 0,03 | 0,04 | 0,60 | 5,46E-01 | 7,11E-01 |
| FYN | 7349,43 | -0,02 | 0,03 | -0,60 | 5,47E-01 | 7,11E-01 |
| AGPAT2 | 509,81 | -0,02 | 0,03 | -0,60 | 5,47E-01 | 7,11E-01 |
| SCYL1 | 854,56 | -0,02 | 0,03 | -0,60 | 5,47E-01 | 7,11E-01 |
| TRIT1 | 592,61 | 0,01 | 0,02 | 0,60 | 5,47E-01 | 7,12E-01 |
| IL15RA | 265,29 | -0,02 | 0,03 | -0,60 | 5,48E-01 | 7,12E-01 |
| EFCAB8 | 101,10 | -0,04 | 0,07 | -0,60 | 5,48E-01 | 7,12E-01 |
| TAF5 | 285,44 | -0,01 | 0,02 | -0,60 | 5,48E-01 | 7,12E-01 |
| KLHL22 | 1013,00 | -0,01 | 0,02 | -0,60 | 5,48E-01 | 7,12E-01 |
| ZNF565 | 121,27 | 0,02 | 0,03 | 0,60 | 5,48E-01 | 7,12E-01 |
| PTPN22 | 1450,27 | 0,02 | 0,04 | 0,60 | 5,48E-01 | 7,12E-01 |
| ATP2A2 | 2604,66 | -0,01 | 0,02 | -0,60 | 5,48E-01 | 7,12E-01 |
| B4GALT4 | 819,82 | -0,01 | 0,02 | -0,60 | 5,48E-01 | 7,12E-01 |
| SNAP47 | 490,53 | 0,01 | 0,02 | 0,60 | 5,49E-01 | 7,13E-01 |

| | | | | | | |
|---|---|---|---|---|---|---|
| ARVCF | 126,94 | -0,04 | 0,06 | -0,60 | 5,49E-01 | 7,13E-01 |
| SNRNP40 | 1056,62 | 0,01 | 0,02 | 0,60 | 5,49E-01 | 7,13E-01 |
| AK2 | 2892,71 | 0,01 | 0,02 | 0,60 | 5,50E-01 | 7,13E-01 |
| IL5RA | 753,77 | 0,04 | 0,07 | 0,60 | 5,50E-01 | 7,13E-01 |
| NSD2 | 1496,11 | 0,01 | 0,02 | 0,60 | 5,50E-01 | 7,13E-01 |
| SMARCA4 | 950,56 | 0,03 | 0,05 | 0,60 | 5,50E-01 | 7,14E-01 |
| MS4A4A | 164,30 | -0,03 | 0,06 | -0,60 | 5,50E-01 | 7,14E-01 |
| PPP1R9B | 2529,09 | 0,02 | 0,04 | 0,60 | 5,50E-01 | 7,14E-01 |
| TTC13 | 2137,49 | 0,01 | 0,02 | 0,60 | 5,51E-01 | 7,14E-01 |
| IQSEC2 | 138,04 | 0,04 | 0,06 | 0,60 | 5,51E-01 | 7,14E-01 |
| THOC3 | 710,94 | 0,02 | 0,03 | 0,60 | 5,51E-01 | 7,14E-01 |
| DBN1 | 122,52 | -0,04 | 0,06 | -0,60 | 5,51E-01 | 7,14E-01 |
| SEC13 | 1199,99 | -0,01 | 0,01 | -0,60 | 5,51E-01 | 7,14E-01 |
| MOB1A | 9321,61 | -0,01 | 0,02 | -0,60 | 5,51E-01 | 7,15E-01 |
| BAIAP3 | 244,42 | 0,04 | 0,07 | 0,60 | 5,51E-01 | 7,15E-01 |
| FAM114A2 | 773,98 | 0,01 | 0,02 | 0,60 | 5,51E-01 | 7,15E-01 |
| IGF2BP3 | 109,52 | 0,04 | 0,07 | 0,60 | 5,52E-01 | 7,15E-01 |
| CMTM3 | 907,03 | 0,01 | 0,02 | 0,60 | 5,52E-01 | 7,15E-01 |
| DDX52 | 1074,31 | -0,01 | 0,02 | -0,59 | 5,52E-01 | 7,15E-01 |
| ZNF93 | 208,89 | -0,02 | 0,03 | -0,59 | 5,52E-01 | 7,15E-01 |
| SEC31A | 2229,20 | 0,01 | 0,02 | 0,59 | 5,53E-01 | 7,16E-01 |
| LLGL2 | 490,10 | 0,03 | 0,04 | 0,59 | 5,53E-01 | 7,16E-01 |
| SPTBN5 | 151,05 | -0,05 | 0,09 | -0,59 | 5,53E-01 | 7,17E-01 |
| BAHCC1 | 79,49 | 0,07 | 0,11 | 0,59 | 5,54E-01 | 7,17E-01 |
| AKR1C3 | 189,84 | -0,04 | 0,06 | -0,59 | 5,54E-01 | 7,17E-01 |
| UNC45A | 807,12 | 0,01 | 0,02 | 0,59 | 5,54E-01 | 7,17E-01 |
| XNDC1N | 550,50 | 0,01 | 0,03 | 0,59 | 5,54E-01 | 7,17E-01 |
| NLRC5 | 5404,99 | -0,03 | 0,05 | -0,59 | 5,54E-01 | 7,17E-01 |
| PSTK | 131,37 | 0,02 | 0,03 | 0,59 | 5,54E-01 | 7,17E-01 |
| MITD1 | 901,17 | 0,01 | 0,02 | 0,59 | 5,54E-01 | 7,17E-01 |
| ENSG000002 | 246,64 | 0,04 | 0,06 | 0,59 | 5,54E-01 | 7,17E-01 |
| TGFBRAP1 | 646,74 | 0,01 | 0,02 | 0,59 | 5,54E-01 | 7,17E-01 |
| DYNLL2 | 3218,33 | -0,01 | 0,02 | -0,59 | 5,54E-01 | 7,17E-01 |
| STRN4 | 1124,12 | -0,02 | 0,04 | -0,59 | 5,54E-01 | 7,17E-01 |
| EPB41L4A | 194,60 | -0,04 | 0,06 | -0,59 | 5,54E-01 | 7,17E-01 |
| EVI5L | 147,49 | 0,03 | 0,06 | 0,59 | 5,55E-01 | 7,17E-01 |
| CIC | 559,58 | 0,04 | 0,07 | 0,59 | 5,55E-01 | 7,17E-01 |
| CLPTM1L | 1561,25 | -0,01 | 0,02 | -0,59 | 5,55E-01 | 7,18E-01 |
| CAMTA1 | 465,07 | 0,02 | 0,03 | 0,59 | 5,55E-01 | 7,18E-01 |
| SLC18A2 | 105,30 | -0,03 | 0,06 | -0,59 | 5,55E-01 | 7,18E-01 |
| SRXN1 | 609,30 | -0,03 | 0,05 | -0,59 | 5,55E-01 | 7,18E-01 |
| LARGE1 | 226,05 | -0,04 | 0,07 | -0,59 | 5,55E-01 | 7,18E-01 |
| OSBPL3 | 1314,14 | 0,01 | 0,02 | 0,59 | 5,56E-01 | 7,18E-01 |
| SDF2L1 | 239,99 | 0,02 | 0,03 | 0,59 | 5,56E-01 | 7,18E-01 |
| NAB1 | 2051,13 | -0,02 | 0,03 | -0,59 | 5,56E-01 | 7,18E-01 |
| TSHZ1 | 529,62 | 0,02 | 0,03 | 0,59 | 5,56E-01 | 7,18E-01 |

| | | | | | | |
|---|---|---|---|---|---|---|
| MPST | 446,17 | 0,02 | 0,03 | 0,59 | 5,56E-01 | 7,18E-01 |
| USP24 | 7055,95 | -0,01 | 0,02 | -0,59 | 5,56E-01 | 7,18E-01 |
| HFE | 214,24 | 0,01 | 0,02 | 0,59 | 5,56E-01 | 7,18E-01 |
| ZNF282 | 493,91 | 0,01 | 0,02 | 0,59 | 5,57E-01 | 7,19E-01 |
| ZNF318 | 1366,65 | 0,01 | 0,02 | 0,59 | 5,57E-01 | 7,19E-01 |
| NOP9 | 1465,73 | 0,01 | 0,02 | 0,59 | 5,57E-01 | 7,19E-01 |
| GPR75 | 98,45 | -0,02 | 0,03 | -0,59 | 5,57E-01 | 7,20E-01 |
| ANXA5 | 3242,71 | -0,02 | 0,03 | -0,59 | 5,58E-01 | 7,20E-01 |
| TFCP2 | 712,96 | -0,01 | 0,02 | -0,59 | 5,58E-01 | 7,20E-01 |
| HYLS1 | 160,13 | 0,02 | 0,03 | 0,59 | 5,58E-01 | 7,20E-01 |
| ARF4 | 1170,42 | -0,01 | 0,02 | -0,59 | 5,58E-01 | 7,21E-01 |
| AP2B1 | 3968,16 | -0,01 | 0,02 | -0,58 | 5,59E-01 | 7,21E-01 |
| ANKRD12 | 6467,34 | 0,02 | 0,04 | 0,58 | 5,59E-01 | 7,21E-01 |
| SF3B3 | 3431,59 | 0,01 | 0,02 | 0,58 | 5,59E-01 | 7,21E-01 |
| TRAF3IP3 | 5475,73 | 0,01 | 0,02 | 0,58 | 5,59E-01 | 7,21E-01 |
| COL6A3 | 153,35 | 0,05 | 0,09 | 0,58 | 5,59E-01 | 7,21E-01 |
| DHDDS | 446,41 | 0,01 | 0,02 | 0,58 | 5,59E-01 | 7,21E-01 |
| TRRAP | 3435,44 | -0,02 | 0,04 | -0,58 | 5,60E-01 | 7,21E-01 |
| SERINC5 | 3175,13 | -0,02 | 0,04 | -0,58 | 5,60E-01 | 7,21E-01 |
| YPEL1 | 418,68 | -0,02 | 0,03 | -0,58 | 5,60E-01 | 7,22E-01 |
| CAPN15 | 480,84 | 0,04 | 0,07 | 0,58 | 5,60E-01 | 7,22E-01 |
| WDSUB1 | 261,42 | 0,01 | 0,02 | 0,58 | 5,60E-01 | 7,22E-01 |
| TMEM176A | 161,65 | -0,08 | 0,13 | -0,58 | 5,60E-01 | 7,22E-01 |
| CCT4 | 1795,65 | 0,02 | 0,03 | 0,58 | 5,61E-01 | 7,22E-01 |
| RTKN2 | 478,43 | 0,03 | 0,06 | 0,58 | 5,61E-01 | 7,22E-01 |
| TTPAL | 864,82 | -0,01 | 0,02 | -0,58 | 5,61E-01 | 7,23E-01 |
| MLC1 | 183,92 | -0,03 | 0,05 | -0,58 | 5,61E-01 | 7,23E-01 |
| TSNAX | 1379,82 | 0,02 | 0,04 | 0,58 | 5,61E-01 | 7,23E-01 |
| PPP4R3A | 5122,24 | -0,01 | 0,02 | -0,58 | 5,61E-01 | 7,23E-01 |
| REST | 2985,16 | 0,01 | 0,02 | 0,58 | 5,62E-01 | 7,23E-01 |
| MCM7 | 929,58 | 0,01 | 0,02 | 0,58 | 5,62E-01 | 7,23E-01 |
| CLEC12B | 296,07 | 0,05 | 0,08 | 0,58 | 5,62E-01 | 7,23E-01 |
| FAM13A | 1041,30 | -0,02 | 0,04 | -0,58 | 5,62E-01 | 7,23E-01 |
| PMPCB | 1462,86 | 0,01 | 0,02 | 0,58 | 5,62E-01 | 7,23E-01 |
| CDK5 | 150,88 | 0,01 | 0,02 | 0,58 | 5,62E-01 | 7,23E-01 |
| CLEC12A | 4679,45 | 0,05 | 0,09 | 0,58 | 5,62E-01 | 7,23E-01 |
| ENC1 | 908,66 | -0,04 | 0,06 | -0,58 | 5,62E-01 | 7,23E-01 |
| LZIC | 660,37 | 0,01 | 0,02 | 0,58 | 5,63E-01 | 7,23E-01 |
| ZNF189 | 867,15 | 0,02 | 0,03 | 0,58 | 5,63E-01 | 7,24E-01 |
| PIM2 | 1521,28 | -0,02 | 0,03 | -0,58 | 5,63E-01 | 7,24E-01 |
| OTULINL | 1741,06 | -0,02 | 0,03 | -0,58 | 5,63E-01 | 7,24E-01 |
| KCNIP2 | 95,48 | 0,03 | 0,06 | 0,58 | 5,63E-01 | 7,24E-01 |
| MICOS10 | 499,65 | 0,01 | 0,02 | 0,58 | 5,63E-01 | 7,24E-01 |
| PPIG | 2317,60 | 0,03 | 0,05 | 0,58 | 5,64E-01 | 7,24E-01 |
| FBXW8 | 350,89 | 0,02 | 0,03 | 0,58 | 5,64E-01 | 7,24E-01 |
| THBS3 | 489,34 | -0,02 | 0,03 | -0,58 | 5,64E-01 | 7,24E-01 |

| | | | | | | |
|---|---|---|---|---|---|---|
| NKIRAS2 | 443,48 | 0,01 | 0,02 | 0,58 | 5,64E-01 | 7,24E-01 |
| MED6 | 983,54 | -0,01 | 0,02 | -0,58 | 5,65E-01 | 7,25E-01 |
| SGSM2 | 806,17 | 0,02 | 0,04 | 0,58 | 5,65E-01 | 7,25E-01 |
| MLYCD | 992,03 | 0,02 | 0,03 | 0,58 | 5,65E-01 | 7,25E-01 |
| MYSM1 | 3500,62 | 0,01 | 0,03 | 0,58 | 5,65E-01 | 7,25E-01 |
| MCM6 | 479,22 | 0,01 | 0,02 | 0,58 | 5,65E-01 | 7,25E-01 |
| TMEM214 | 270,16 | 0,02 | 0,04 | 0,58 | 5,65E-01 | 7,25E-01 |
| CXXC5 | 475,14 | 0,02 | 0,04 | 0,58 | 5,65E-01 | 7,25E-01 |
| TCOF1 | 651,38 | 0,02 | 0,03 | 0,58 | 5,65E-01 | 7,25E-01 |
| DCHS1 | 120,78 | 0,05 | 0,08 | 0,58 | 5,65E-01 | 7,25E-01 |
| DUSP16 | 707,09 | -0,02 | 0,03 | -0,58 | 5,65E-01 | 7,25E-01 |
| TSPAN14 | 3872,14 | -0,02 | 0,03 | -0,57 | 5,65E-01 | 7,25E-01 |
| UPF3B | 413,56 | -0,01 | 0,02 | -0,57 | 5,66E-01 | 7,25E-01 |
| E4F1 | 850,77 | 0,01 | 0,02 | 0,57 | 5,66E-01 | 7,25E-01 |
| ELAPOR1 | 1217,53 | -0,05 | 0,08 | -0,57 | 5,66E-01 | 7,25E-01 |
| LSM3 | 276,80 | 0,02 | 0,04 | 0,57 | 5,66E-01 | 7,25E-01 |
| CHAF1B | 74,11 | -0,03 | 0,05 | -0,57 | 5,66E-01 | 7,26E-01 |
| TOP3B | 769,80 | 0,01 | 0,02 | 0,57 | 5,66E-01 | 7,26E-01 |
| CNTF | 103,65 | -0,01 | 0,02 | -0,57 | 5,66E-01 | 7,26E-01 |
| ZFP28 | 211,63 | 0,02 | 0,04 | 0,57 | 5,67E-01 | 7,26E-01 |
| APEX2 | 177,53 | 0,02 | 0,03 | 0,57 | 5,67E-01 | 7,26E-01 |
| SRD5A1 | 556,11 | 0,02 | 0,03 | 0,57 | 5,67E-01 | 7,27E-01 |
| CD47 | 9737,27 | 0,01 | 0,02 | 0,57 | 5,67E-01 | 7,27E-01 |
| SIRT2 | 339,34 | -0,02 | 0,04 | -0,57 | 5,67E-01 | 7,27E-01 |
| INTS12 | 352,49 | 0,01 | 0,02 | 0,57 | 5,67E-01 | 7,27E-01 |
| EXO5 | 281,29 | 0,01 | 0,02 | 0,57 | 5,68E-01 | 7,27E-01 |
| ITGAL | 5136,43 | -0,02 | 0,04 | -0,57 | 5,68E-01 | 7,27E-01 |
| DISP1 | 134,49 | 0,02 | 0,03 | 0,57 | 5,68E-01 | 7,27E-01 |
| CTSA | 763,34 | -0,03 | 0,05 | -0,57 | 5,68E-01 | 7,27E-01 |
| ARHGAP4 | 3327,63 | 0,02 | 0,03 | 0,57 | 5,68E-01 | 7,27E-01 |
| KANSL3 | 1436,98 | -0,02 | 0,03 | -0,57 | 5,69E-01 | 7,28E-01 |
| ACER2 | 98,97 | 0,02 | 0,04 | 0,57 | 5,69E-01 | 7,28E-01 |
| ELP1 | 1849,69 | -0,01 | 0,02 | -0,57 | 5,69E-01 | 7,28E-01 |
| TNRC6C | 1838,15 | -0,02 | 0,03 | -0,57 | 5,69E-01 | 7,28E-01 |
| TRAF2 | 422,96 | 0,01 | 0,02 | 0,57 | 5,69E-01 | 7,28E-01 |
| SRRM2 | 5819,78 | -0,03 | 0,06 | -0,57 | 5,69E-01 | 7,28E-01 |
| ZNF107 | 1393,99 | 0,02 | 0,04 | 0,57 | 5,69E-01 | 7,28E-01 |
| GPI | 644,94 | 0,02 | 0,04 | 0,57 | 5,69E-01 | 7,28E-01 |
| ZNF227 | 1005,95 | 0,02 | 0,03 | 0,57 | 5,69E-01 | 7,28E-01 |
| TMC8 | 2576,90 | 0,03 | 0,05 | 0,57 | 5,69E-01 | 7,28E-01 |
| EHD3 | 296,37 | 0,03 | 0,05 | 0,57 | 5,70E-01 | 7,28E-01 |
| TTLL3 | 775,39 | -0,03 | 0,06 | -0,57 | 5,70E-01 | 7,28E-01 |
| HARBI1 | 146,24 | -0,01 | 0,02 | -0,57 | 5,70E-01 | 7,29E-01 |
| MCM2 | 232,37 | 0,02 | 0,04 | 0,57 | 5,70E-01 | 7,29E-01 |
| BLMH | 504,93 | 0,01 | 0,02 | 0,57 | 5,70E-01 | 7,29E-01 |
| CERS6 | 1277,84 | 0,01 | 0,02 | 0,57 | 5,70E-01 | 7,29E-01 |

| | | | | | | |
|---|---|---|---|---|---|---|
| PKN1 | 1031,33 | 0,03 | 0,05 | 0,57 | 5,71E-01 | 7,30E-01 |
| WDR41 | 1160,89 | -0,01 | 0,02 | -0,57 | 5,71E-01 | 7,30E-01 |
| MSH2 | 783,30 | 0,02 | 0,03 | 0,57 | 5,71E-01 | 7,30E-01 |
| LIG1 | 326,71 | 0,02 | 0,04 | 0,57 | 5,71E-01 | 7,30E-01 |
| ABCC6 | 95,06 | -0,05 | 0,08 | -0,57 | 5,71E-01 | 7,30E-01 |
| ABHD6 | 253,95 | -0,01 | 0,02 | -0,57 | 5,72E-01 | 7,30E-01 |
| MYL5 | 88,72 | 0,03 | 0,05 | 0,57 | 5,72E-01 | 7,30E-01 |
| DNAH1 | 2171,50 | -0,03 | 0,05 | -0,57 | 5,72E-01 | 7,30E-01 |
| SAMD4A | 300,98 | -0,03 | 0,06 | -0,57 | 5,72E-01 | 7,30E-01 |
| RBM17 | 1660,62 | 0,01 | 0,02 | 0,56 | 5,72E-01 | 7,30E-01 |
| RBM4 | 1538,58 | -0,01 | 0,02 | -0,56 | 5,72E-01 | 7,30E-01 |
| MON1A | 167,26 | 0,01 | 0,02 | 0,56 | 5,73E-01 | 7,30E-01 |
| ABCA3 | 100,76 | 0,04 | 0,07 | 0,56 | 5,73E-01 | 7,30E-01 |
| INO80 | 1522,87 | -0,01 | 0,02 | -0,56 | 5,73E-01 | 7,30E-01 |
| EXOC1 | 2607,84 | -0,02 | 0,03 | -0,56 | 5,72E-01 | 7,30E-01 |
| CIMAP2 | 85,68 | -0,04 | 0,07 | -0,56 | 5,72E-01 | 7,30E-01 |
| FGFBP2 | 816,58 | -0,03 | 0,06 | -0,56 | 5,73E-01 | 7,30E-01 |
| TAF5L | 655,69 | 0,01 | 0,01 | 0,56 | 5,73E-01 | 7,30E-01 |
| GATA2 | 252,33 | -0,06 | 0,10 | -0,56 | 5,73E-01 | 7,31E-01 |
| NEO1 | 265,78 | -0,03 | 0,05 | -0,56 | 5,74E-01 | 7,31E-01 |
| MCOLN2 | 537,80 | 0,03 | 0,05 | 0,56 | 5,74E-01 | 7,31E-01 |
| SLC38A10 | 1969,75 | -0,02 | 0,04 | -0,56 | 5,74E-01 | 7,32E-01 |
| ADAMTSL4 | 182,96 | 0,05 | 0,08 | 0,56 | 5,74E-01 | 7,32E-01 |
| FKBP4 | 377,01 | -0,01 | 0,02 | -0,56 | 5,74E-01 | 7,32E-01 |
| HHEX | 720,58 | 0,02 | 0,03 | 0,56 | 5,75E-01 | 7,32E-01 |
| COG8 | 939,18 | 0,01 | 0,01 | 0,56 | 5,75E-01 | 7,33E-01 |
| CBX3 | 2129,62 | 0,02 | 0,04 | 0,56 | 5,75E-01 | 7,33E-01 |
| FCHSD1 | 728,22 | -0,02 | 0,04 | -0,56 | 5,75E-01 | 7,33E-01 |
| KLHDC3 | 1194,28 | 0,01 | 0,01 | 0,56 | 5,75E-01 | 7,33E-01 |
| CLINT1 | 3103,96 | 0,01 | 0,02 | 0,56 | 5,76E-01 | 7,33E-01 |
| PDE3A | 91,40 | 0,04 | 0,08 | 0,56 | 5,76E-01 | 7,33E-01 |
| RIOK1 | 527,61 | 0,01 | 0,02 | 0,56 | 5,76E-01 | 7,33E-01 |
| GALNT6 | 674,28 | 0,02 | 0,03 | 0,56 | 5,76E-01 | 7,33E-01 |
| AGAP6 | 452,13 | 0,02 | 0,03 | 0,56 | 5,76E-01 | 7,34E-01 |
| ELOVL7 | 440,82 | 0,03 | 0,06 | 0,56 | 5,77E-01 | 7,34E-01 |
| TMEM131 | 2766,88 | -0,01 | 0,02 | -0,56 | 5,77E-01 | 7,34E-01 |
| ANKMY2 | 468,56 | 0,01 | 0,03 | 0,56 | 5,77E-01 | 7,34E-01 |
| XYLT1 | 1634,46 | -0,02 | 0,03 | -0,56 | 5,77E-01 | 7,34E-01 |
| CD74 | 4527,35 | 0,03 | 0,06 | 0,56 | 5,77E-01 | 7,34E-01 |
| ACD | 119,78 | 0,02 | 0,04 | 0,56 | 5,77E-01 | 7,34E-01 |
| ZNF148 | 4491,15 | -0,02 | 0,03 | -0,56 | 5,78E-01 | 7,35E-01 |
| FGFR2 | 136,52 | -0,05 | 0,09 | -0,56 | 5,78E-01 | 7,35E-01 |
| ZFPM1 | 160,07 | 0,04 | 0,07 | 0,56 | 5,78E-01 | 7,35E-01 |
| CWC27 | 505,05 | 0,02 | 0,03 | 0,56 | 5,78E-01 | 7,35E-01 |
| USP13 | 335,89 | 0,01 | 0,03 | 0,56 | 5,78E-01 | 7,35E-01 |
| ISYNA1 | 78,34 | 0,03 | 0,05 | 0,56 | 5,78E-01 | 7,35E-01 |

| | | | | | | |
|---|---|---|---|---|---|---|
| L3MBTL3 | 1175,30 | 0,01 | 0,02 | 0,56 | 5,78E-01 | 7,35E-01 |
| KAT6B | 2331,82 | -0,01 | 0,02 | -0,56 | 5,78E-01 | 7,35E-01 |
| ZNF626 | 392,06 | 0,02 | 0,04 | 0,56 | 5,78E-01 | 7,35E-01 |
| ERAP1 | 3518,50 | -0,01 | 0,02 | -0,56 | 5,79E-01 | 7,35E-01 |
| TBX19 | 139,99 | -0,03 | 0,05 | -0,56 | 5,79E-01 | 7,35E-01 |
| DDX55 | 623,61 | 0,01 | 0,02 | 0,55 | 5,79E-01 | 7,35E-01 |
| CNP | 429,51 | -0,02 | 0,03 | -0,56 | 5,79E-01 | 7,35E-01 |
| HYPK | 333,67 | -0,01 | 0,03 | -0,55 | 5,79E-01 | 7,35E-01 |
| CBLL1 | 1145,44 | 0,02 | 0,03 | 0,55 | 5,79E-01 | 7,35E-01 |
| AARS1 | 752,10 | 0,01 | 0,02 | 0,55 | 5,79E-01 | 7,35E-01 |
| ATPAF2 | 389,24 | 0,01 | 0,02 | 0,56 | 5,79E-01 | 7,35E-01 |
| ALCAM | 1000,15 | 0,02 | 0,04 | 0,55 | 5,79E-01 | 7,35E-01 |
| TLNRD1 | 540,77 | 0,01 | 0,03 | 0,55 | 5,79E-01 | 7,35E-01 |
| RBM39 | 18261,04 | -0,02 | 0,03 | -0,55 | 5,79E-01 | 7,35E-01 |
| TBC1D2 | 394,91 | -0,02 | 0,04 | -0,55 | 5,79E-01 | 7,35E-01 |
| FAXDC2 | 1360,23 | 0,04 | 0,07 | 0,55 | 5,80E-01 | 7,36E-01 |
| UHRF1 | 72,03 | -0,04 | 0,07 | -0,55 | 5,80E-01 | 7,36E-01 |
| CLCN5 | 470,62 | -0,02 | 0,03 | -0,55 | 5,80E-01 | 7,36E-01 |
| DCTN3 | 675,65 | -0,01 | 0,02 | -0,55 | 5,81E-01 | 7,36E-01 |
| TRIM11 | 930,96 | -0,02 | 0,03 | -0,55 | 5,80E-01 | 7,36E-01 |
| WBP4 | 449,89 | 0,02 | 0,03 | 0,55 | 5,81E-01 | 7,36E-01 |
| ALDH18A1 | 456,48 | 0,01 | 0,02 | 0,55 | 5,81E-01 | 7,37E-01 |
| ZNF512 | 714,50 | 0,01 | 0,02 | 0,55 | 5,81E-01 | 7,37E-01 |
| MOGS | 536,92 | 0,02 | 0,03 | 0,55 | 5,81E-01 | 7,37E-01 |
| NFIC | 315,13 | 0,04 | 0,07 | 0,55 | 5,81E-01 | 7,37E-01 |
| TSPO | 3840,10 | -0,02 | 0,03 | -0,55 | 5,82E-01 | 7,37E-01 |
| ADAM12 | 104,30 | -0,05 | 0,09 | -0,55 | 5,82E-01 | 7,37E-01 |
| SBF1 | 1537,12 | 0,03 | 0,06 | 0,55 | 5,82E-01 | 7,37E-01 |
| FAM184A | 125,34 | 0,03 | 0,05 | 0,55 | 5,82E-01 | 7,38E-01 |
| TMEM150B | 77,85 | 0,03 | 0,06 | 0,55 | 5,82E-01 | 7,38E-01 |
| CCAR2 | 646,65 | 0,02 | 0,04 | 0,55 | 5,82E-01 | 7,38E-01 |
| AGA | 534,97 | 0,02 | 0,03 | 0,55 | 5,83E-01 | 7,38E-01 |
| PLEKHA5 | 278,41 | 0,03 | 0,05 | 0,55 | 5,83E-01 | 7,38E-01 |
| MORC4 | 212,14 | 0,02 | 0,03 | 0,55 | 5,83E-01 | 7,38E-01 |
| ZNF286A | 283,73 | 0,02 | 0,03 | 0,55 | 5,83E-01 | 7,38E-01 |
| CXXC1 | 867,12 | -0,01 | 0,02 | -0,55 | 5,83E-01 | 7,38E-01 |
| MYO7A | 109,98 | -0,06 | 0,11 | -0,55 | 5,84E-01 | 7,39E-01 |
| SCAPER | 1624,97 | -0,01 | 0,03 | -0,55 | 5,84E-01 | 7,39E-01 |
| GALE | 150,31 | 0,01 | 0,02 | 0,55 | 5,84E-01 | 7,39E-01 |
| SLC66A1 | 123,39 | -0,02 | 0,04 | -0,55 | 5,84E-01 | 7,39E-01 |
| LDHB | 780,92 | 0,02 | 0,05 | 0,55 | 5,84E-01 | 7,39E-01 |
| ZRSR2 | 715,20 | -0,01 | 0,02 | -0,55 | 5,84E-01 | 7,39E-01 |
| SKIL | 1794,99 | 0,02 | 0,04 | 0,55 | 5,85E-01 | 7,39E-01 |
| MACF1 | 28144,60 | -0,01 | 0,02 | -0,55 | 5,85E-01 | 7,39E-01 |
| IDH2 | 901,02 | 0,01 | 0,02 | 0,55 | 5,85E-01 | 7,40E-01 |
| RNF103 | 1382,71 | -0,01 | 0,02 | -0,55 | 5,85E-01 | 7,40E-01 |

| | | | | | | |
|---|---|---|---|---|---|---|
| TGFBR3 | 1432,77 | -0,03 | 0,05 | -0,55 | 5,85E-01 | 7,40E-01 |
| DAG1 | 273,81 | 0,02 | 0,03 | 0,55 | 5,85E-01 | 7,40E-01 |
| MMGT1 | 791,71 | 0,01 | 0,02 | 0,55 | 5,85E-01 | 7,40E-01 |
| ICMT | 812,61 | 0,01 | 0,02 | 0,55 | 5,86E-01 | 7,40E-01 |
| NLRP2 | 143,74 | -0,04 | 0,08 | -0,54 | 5,86E-01 | 7,41E-01 |
| NAAA | 2158,62 | -0,02 | 0,03 | -0,54 | 5,86E-01 | 7,41E-01 |
| SLC39A13 | 169,31 | 0,02 | 0,04 | 0,54 | 5,86E-01 | 7,41E-01 |
| CCNDBP1 | 5636,25 | -0,02 | 0,03 | -0,54 | 5,87E-01 | 7,41E-01 |
| WASF2 | 3429,98 | 0,03 | 0,05 | 0,54 | 5,87E-01 | 7,41E-01 |
| SLC9A1 | 194,43 | -0,04 | 0,07 | -0,54 | 5,87E-01 | 7,41E-01 |
| RPL36AL | 2216,94 | -0,03 | 0,05 | -0,54 | 5,87E-01 | 7,41E-01 |
| L3MBTL1 | 429,17 | 0,02 | 0,04 | 0,54 | 5,87E-01 | 7,41E-01 |
| CNOT4 | 1228,66 | 0,01 | 0,02 | 0,54 | 5,87E-01 | 7,41E-01 |
| OXA1L | 1627,52 | 0,01 | 0,02 | 0,54 | 5,87E-01 | 7,41E-01 |
| PTPN6 | 2810,29 | -0,03 | 0,05 | -0,54 | 5,87E-01 | 7,41E-01 |
| UBXN1 | 1926,52 | 0,01 | 0,02 | 0,54 | 5,87E-01 | 7,41E-01 |
| GTF2IRD2 | 181,80 | -0,02 | 0,04 | -0,54 | 5,87E-01 | 7,41E-01 |
| ZFYVE28 | 365,99 | 0,03 | 0,05 | 0,54 | 5,87E-01 | 7,41E-01 |
| WDR35 | 536,00 | 0,02 | 0,03 | 0,54 | 5,88E-01 | 7,42E-01 |
| CBR4 | 1577,05 | 0,01 | 0,03 | 0,54 | 5,88E-01 | 7,42E-01 |
| CALM3 | 1244,88 | -0,02 | 0,03 | -0,54 | 5,88E-01 | 7,42E-01 |
| HSPD1 | 1503,18 | -0,01 | 0,03 | -0,54 | 5,88E-01 | 7,42E-01 |
| SLC12A7 | 688,76 | 0,04 | 0,07 | 0,54 | 5,88E-01 | 7,42E-01 |
| TOR2A | 114,06 | 0,03 | 0,05 | 0,54 | 5,88E-01 | 7,42E-01 |
| SRGAP1 | 72,81 | -0,07 | 0,14 | -0,54 | 5,88E-01 | 7,42E-01 |
| ATF4 | 2583,20 | 0,01 | 0,02 | 0,54 | 5,88E-01 | 7,42E-01 |
| TNRC18 | 1889,08 | -0,04 | 0,07 | -0,54 | 5,88E-01 | 7,42E-01 |
| CDK20 | 87,42 | 0,03 | 0,05 | 0,54 | 5,89E-01 | 7,42E-01 |
| CBX6 | 1121,74 | 0,02 | 0,03 | 0,54 | 5,89E-01 | 7,43E-01 |
| IPO13 | 212,59 | -0,02 | 0,04 | -0,54 | 5,89E-01 | 7,43E-01 |
| COG5 | 2493,33 | 0,01 | 0,03 | 0,54 | 5,89E-01 | 7,43E-01 |
| DDX31 | 477,97 | 0,01 | 0,03 | 0,54 | 5,90E-01 | 7,43E-01 |
| MKI67 | 290,17 | 0,03 | 0,06 | 0,54 | 5,89E-01 | 7,43E-01 |
| DENND4C | 3209,27 | 0,01 | 0,02 | 0,54 | 5,90E-01 | 7,44E-01 |
| PLD5P1 | 169,77 | 0,02 | 0,04 | 0,54 | 5,90E-01 | 7,44E-01 |
| ECHDC1 | 1514,10 | 0,02 | 0,04 | 0,54 | 5,90E-01 | 7,44E-01 |
| BAP1 | 1306,00 | -0,01 | 0,02 | -0,54 | 5,91E-01 | 7,44E-01 |
| TOM1L2 | 689,47 | -0,02 | 0,04 | -0,54 | 5,91E-01 | 7,44E-01 |
| KRT73 | 177,81 | -0,05 | 0,10 | -0,54 | 5,91E-01 | 7,45E-01 |
| TNIP2 | 1058,00 | -0,01 | 0,02 | -0,54 | 5,92E-01 | 7,45E-01 |
| SMAD7 | 558,88 | 0,02 | 0,03 | 0,54 | 5,92E-01 | 7,45E-01 |
| TCEAL3 | 104,88 | 0,02 | 0,04 | 0,54 | 5,92E-01 | 7,45E-01 |
| MYO1B | 89,49 | 0,05 | 0,08 | 0,54 | 5,92E-01 | 7,45E-01 |
| GOLM1 | 208,63 | -0,03 | 0,05 | -0,54 | 5,92E-01 | 7,45E-01 |
| DCXR | 340,63 | -0,01 | 0,03 | -0,54 | 5,92E-01 | 7,45E-01 |
| RAB39B | 368,49 | 0,01 | 0,03 | 0,54 | 5,92E-01 | 7,45E-01 |

| | | | | | | |
|---|---|---|---|---|---|---|
| ZC3H15 | 1055,29 | 0,03 | 0,05 | 0,54 | 5,92E-01 | 7,45E-01 |
| NCALD | 760,86 | -0,02 | 0,04 | -0,54 | 5,93E-01 | 7,45E-01 |
| ZSCAN22 | 144,26 | 0,01 | 0,02 | 0,53 | 5,93E-01 | 7,45E-01 |
| TFG | 1486,98 | -0,01 | 0,02 | -0,53 | 5,93E-01 | 7,46E-01 |
| SANBR | 351,35 | 0,01 | 0,02 | 0,53 | 5,93E-01 | 7,46E-01 |
| CHCHD6 | 152,11 | 0,02 | 0,03 | 0,53 | 5,93E-01 | 7,46E-01 |
| ZSCAN20 | 161,69 | 0,02 | 0,03 | 0,53 | 5,94E-01 | 7,46E-01 |
| WDR17 | 89,28 | 0,03 | 0,05 | 0,53 | 5,94E-01 | 7,46E-01 |
| ZNF862 | 1067,66 | 0,01 | 0,02 | 0,53 | 5,94E-01 | 7,46E-01 |
| ERCC5 | 654,57 | 0,01 | 0,02 | 0,53 | 5,94E-01 | 7,46E-01 |
| MEA1 | 427,08 | 0,01 | 0,02 | 0,53 | 5,94E-01 | 7,46E-01 |
| MAP3K7CL | 1013,72 | 0,03 | 0,05 | 0,53 | 5,94E-01 | 7,46E-01 |
| PLD3 | 472,26 | 0,02 | 0,04 | 0,53 | 5,94E-01 | 7,46E-01 |
| GTF2H1 | 1443,03 | 0,01 | 0,02 | 0,53 | 5,94E-01 | 7,46E-01 |
| TBC1D16 | 103,31 | 0,04 | 0,07 | 0,53 | 5,94E-01 | 7,46E-01 |
| MIER1 | 5077,32 | -0,02 | 0,04 | -0,53 | 5,95E-01 | 7,47E-01 |
| RTL6 | 370,15 | 0,02 | 0,04 | 0,53 | 5,95E-01 | 7,47E-01 |
| GLCCI1 | 1195,07 | -0,01 | 0,02 | -0,53 | 5,95E-01 | 7,47E-01 |
| PRP4K | 3566,03 | 0,01 | 0,02 | 0,53 | 5,95E-01 | 7,47E-01 |
| RBM42 | 156,30 | 0,02 | 0,04 | 0,53 | 5,95E-01 | 7,47E-01 |
| TBC1D10B | 420,27 | -0,03 | 0,06 | -0,53 | 5,95E-01 | 7,47E-01 |
| FAM241A | 442,12 | 0,01 | 0,03 | 0,53 | 5,95E-01 | 7,47E-01 |
| COASY | 202,81 | -0,03 | 0,05 | -0,53 | 5,95E-01 | 7,47E-01 |
| UQCRC2 | 2664,81 | 0,01 | 0,02 | 0,53 | 5,95E-01 | 7,47E-01 |
| HS6ST1 | 232,76 | -0,02 | 0,04 | -0,53 | 5,96E-01 | 7,47E-01 |
| PNPT1 | 702,87 | -0,02 | 0,04 | -0,53 | 5,96E-01 | 7,47E-01 |
| PSMD9 | 313,16 | -0,01 | 0,02 | -0,53 | 5,96E-01 | 7,48E-01 |
| RIN1 | 81,47 | 0,03 | 0,06 | 0,53 | 5,96E-01 | 7,48E-01 |
| LAS1L | 746,72 | 0,01 | 0,02 | 0,53 | 5,96E-01 | 7,48E-01 |
| TMEM63A | 2958,92 | 0,03 | 0,05 | 0,53 | 5,96E-01 | 7,48E-01 |
| TAF15 | 2034,75 | -0,01 | 0,02 | -0,53 | 5,96E-01 | 7,48E-01 |
| GDF11 | 416,36 | 0,01 | 0,03 | 0,53 | 5,97E-01 | 7,48E-01 |
| SLC25A53 | 583,48 | -0,01 | 0,02 | -0,53 | 5,97E-01 | 7,48E-01 |
| USP53 | 1483,27 | 0,03 | 0,05 | 0,53 | 5,97E-01 | 7,48E-01 |
| ABTB2 | 139,72 | -0,04 | 0,07 | -0,53 | 5,97E-01 | 7,48E-01 |
| ZNF764 | 125,70 | 0,02 | 0,04 | 0,53 | 5,97E-01 | 7,48E-01 |
| SMARCC2 | 1175,18 | -0,02 | 0,03 | -0,53 | 5,98E-01 | 7,49E-01 |
| TBC1D31 | 1129,18 | 0,01 | 0,03 | 0,53 | 5,98E-01 | 7,49E-01 |
| MIS18BP1 | 2577,03 | 0,02 | 0,04 | 0,53 | 5,98E-01 | 7,49E-01 |
| SMARCD1 | 599,12 | 0,01 | 0,02 | 0,53 | 5,98E-01 | 7,49E-01 |
| TSFM | 260,11 | 0,01 | 0,02 | 0,53 | 5,98E-01 | 7,49E-01 |
| NELFCD | 881,57 | 0,01 | 0,02 | 0,53 | 5,98E-01 | 7,49E-01 |
| PRAG1 | 147,63 | 0,03 | 0,05 | 0,53 | 5,98E-01 | 7,49E-01 |
| CWC15 | 597,77 | 0,02 | 0,03 | 0,53 | 5,99E-01 | 7,49E-01 |
| SMAD2 | 8611,34 | -0,01 | 0,02 | -0,53 | 5,99E-01 | 7,49E-01 |
| OTUB1 | 604,91 | -0,01 | 0,02 | -0,53 | 5,99E-01 | 7,49E-01 |

| | | | | | | |
|---|---|---|---|---|---|---|
| OCIAD1 | 1606,67 | 0,01 | 0,02 | 0,53 | 5,99E-01 | 7,50E-01 |
| ARHGDIA | 612,14 | 0,03 | 0,05 | 0,53 | 5,99E-01 | 7,50E-01 |
| OGDH | 1497,22 | -0,02 | 0,03 | -0,53 | 5,99E-01 | 7,50E-01 |
| TRIO | 1828,73 | 0,02 | 0,03 | 0,53 | 6,00E-01 | 7,50E-01 |
| ABLIM1 | 4146,53 | 0,02 | 0,03 | 0,52 | 6,00E-01 | 7,50E-01 |
| ORAI1 | 457,35 | 0,02 | 0,03 | 0,52 | 6,00E-01 | 7,50E-01 |
| FYCO1 | 1343,60 | 0,02 | 0,03 | 0,52 | 6,00E-01 | 7,50E-01 |
| DNHD1 | 924,75 | 0,03 | 0,05 | 0,52 | 6,00E-01 | 7,50E-01 |
| APOBEC3G | 1169,99 | 0,02 | 0,03 | 0,52 | 6,00E-01 | 7,51E-01 |
| WDR55 | 1174,65 | -0,01 | 0,02 | -0,52 | 6,00E-01 | 7,51E-01 |
| RGS9 | 130,06 | -0,04 | 0,07 | -0,52 | 6,01E-01 | 7,51E-01 |
| MC1R | 218,68 | -0,02 | 0,04 | -0,52 | 6,01E-01 | 7,51E-01 |
| NPEPL1 | 332,12 | -0,03 | 0,05 | -0,52 | 6,01E-01 | 7,51E-01 |
| FAM118B | 581,32 | 0,01 | 0,02 | 0,52 | 6,02E-01 | 7,52E-01 |
| ACVR1 | 438,62 | 0,01 | 0,02 | 0,52 | 6,02E-01 | 7,52E-01 |
| CABIN1 | 1763,83 | 0,02 | 0,04 | 0,52 | 6,02E-01 | 7,52E-01 |
| PLA2G15 | 90,01 | 0,03 | 0,05 | 0,52 | 6,03E-01 | 7,52E-01 |
| IFT172 | 379,51 | -0,02 | 0,03 | -0,52 | 6,02E-01 | 7,52E-01 |
| DHRS1 | 195,74 | -0,02 | 0,03 | -0,52 | 6,03E-01 | 7,52E-01 |
| AK6 | 84,57 | 0,03 | 0,05 | 0,52 | 6,03E-01 | 7,53E-01 |
| SMG5 | 1182,39 | -0,02 | 0,04 | -0,52 | 6,03E-01 | 7,53E-01 |
| BEX4 | 311,80 | 0,02 | 0,03 | 0,52 | 6,03E-01 | 7,53E-01 |
| AGAP4 | 319,40 | -0,02 | 0,03 | -0,52 | 6,04E-01 | 7,53E-01 |
| ERBIN | 11867,49 | -0,02 | 0,03 | -0,52 | 6,04E-01 | 7,53E-01 |
| TNK2 | 1946,18 | -0,03 | 0,06 | -0,52 | 6,04E-01 | 7,53E-01 |
| EXD2 | 295,42 | 0,01 | 0,02 | 0,52 | 6,04E-01 | 7,53E-01 |
| CX3CR1 | 2469,05 | -0,02 | 0,04 | -0,52 | 6,04E-01 | 7,53E-01 |
| ECH1 | 304,15 | -0,02 | 0,03 | -0,52 | 6,04E-01 | 7,53E-01 |
| ENSG000002 | 123,36 | 0,09 | 0,17 | 0,52 | 6,04E-01 | 7,53E-01 |
| NEIL1 | 223,16 | -0,02 | 0,05 | -0,52 | 6,04E-01 | 7,54E-01 |
| NBPF19 | 3436,84 | -0,02 | 0,04 | -0,52 | 6,05E-01 | 7,54E-01 |
| ABRAXAS2 | 284,36 | 0,01 | 0,02 | 0,52 | 6,05E-01 | 7,54E-01 |
| KATNB1 | 267,88 | -0,02 | 0,04 | -0,52 | 6,05E-01 | 7,54E-01 |
| CDC7 | 288,59 | 0,02 | 0,03 | 0,52 | 6,05E-01 | 7,54E-01 |
| RAB11FIP5 | 143,28 | 0,03 | 0,06 | 0,52 | 6,05E-01 | 7,54E-01 |
| ARHGAP6 | 131,28 | -0,03 | 0,06 | -0,52 | 6,06E-01 | 7,55E-01 |
| PARP15 | 2173,56 | 0,02 | 0,04 | 0,52 | 6,06E-01 | 7,55E-01 |
| PVR | 322,40 | -0,02 | 0,04 | -0,52 | 6,06E-01 | 7,55E-01 |
| DHCR7 | 88,55 | -0,03 | 0,05 | -0,52 | 6,06E-01 | 7,55E-01 |
| PEX12 | 142,95 | 0,01 | 0,03 | 0,52 | 6,06E-01 | 7,55E-01 |
| PGGT1B | 3137,71 | 0,02 | 0,03 | 0,52 | 6,06E-01 | 7,55E-01 |
| ZSCAN18 | 616,43 | 0,02 | 0,04 | 0,52 | 6,06E-01 | 7,55E-01 |
| HERC2 | 2155,62 | -0,02 | 0,03 | -0,52 | 6,06E-01 | 7,55E-01 |
| TLE1 | 320,91 | 0,02 | 0,04 | 0,52 | 6,06E-01 | 7,55E-01 |
| KLHL28 | 2120,83 | 0,02 | 0,03 | 0,52 | 6,06E-01 | 7,55E-01 |
| PPFIBP2 | 793,06 | 0,02 | 0,03 | 0,51 | 6,07E-01 | 7,55E-01 |

| | | | | | | |
|---|---|---|---|---|---|---|
| ENY2 | 808,43 | 0,02 | 0,03 | 0,51 | 6,07E-01 | 7,56E-01 |
| PPP2R1B | 1366,44 | -0,01 | 0,02 | -0,51 | 6,08E-01 | 7,56E-01 |
| MROH1 | 472,04 | 0,03 | 0,06 | 0,51 | 6,08E-01 | 7,56E-01 |
| INTS15 | 696,05 | 0,01 | 0,02 | 0,51 | 6,08E-01 | 7,56E-01 |
| CD160 | 273,34 | -0,03 | 0,06 | -0,51 | 6,08E-01 | 7,56E-01 |
| MSS51 | 271,35 | -0,01 | 0,03 | -0,51 | 6,08E-01 | 7,56E-01 |
| ZNF264 | 2872,42 | -0,01 | 0,02 | -0,51 | 6,08E-01 | 7,56E-01 |
| ZNF284 | 87,33 | 0,01 | 0,02 | 0,51 | 6,08E-01 | 7,56E-01 |
| CLMN | 868,28 | -0,02 | 0,04 | -0,51 | 6,08E-01 | 7,56E-01 |
| DYNC2I1 | 502,41 | 0,01 | 0,03 | 0,51 | 6,08E-01 | 7,56E-01 |
| YIPF5 | 970,47 | 0,02 | 0,03 | 0,51 | 6,08E-01 | 7,56E-01 |
| SZRD1 | 1468,81 | -0,01 | 0,02 | -0,51 | 6,09E-01 | 7,57E-01 |
| TIMM29 | 698,21 | 0,01 | 0,02 | 0,51 | 6,09E-01 | 7,57E-01 |
| PCDH1 | 85,05 | 0,04 | 0,07 | 0,51 | 6,09E-01 | 7,57E-01 |
| H2AC15 | 332,51 | 0,02 | 0,04 | 0,51 | 6,09E-01 | 7,57E-01 |
| IRS1 | 92,88 | 0,03 | 0,06 | 0,51 | 6,09E-01 | 7,57E-01 |
| MICAL1 | 1092,99 | -0,03 | 0,06 | -0,51 | 6,09E-01 | 7,57E-01 |
| SRSF9 | 2534,32 | -0,01 | 0,02 | -0,51 | 6,09E-01 | 7,57E-01 |
| MORC3 | 4244,33 | 0,02 | 0,04 | 0,51 | 6,10E-01 | 7,57E-01 |
| PLAA | 758,31 | -0,01 | 0,02 | -0,51 | 6,10E-01 | 7,58E-01 |
| GNG7 | 169,73 | 0,03 | 0,05 | 0,51 | 6,10E-01 | 7,58E-01 |
| SETD4 | 694,41 | -0,01 | 0,02 | -0,51 | 6,10E-01 | 7,58E-01 |
| BNIP2 | 6115,15 | -0,02 | 0,03 | -0,51 | 6,11E-01 | 7,58E-01 |
| CCL5 | 4532,00 | 0,02 | 0,05 | 0,51 | 6,11E-01 | 7,58E-01 |
| ALG12 | 397,77 | -0,01 | 0,02 | -0,51 | 6,11E-01 | 7,59E-01 |
| NR1H3 | 102,77 | 0,02 | 0,03 | 0,51 | 6,12E-01 | 7,59E-01 |
| ZNF221 | 86,05 | 0,02 | 0,04 | 0,51 | 6,12E-01 | 7,59E-01 |
| MS4A2 | 165,51 | -0,05 | 0,10 | -0,51 | 6,12E-01 | 7,60E-01 |
| TMEM38A | 85,66 | -0,02 | 0,04 | -0,51 | 6,12E-01 | 7,60E-01 |
| AIFM1 | 378,35 | -0,01 | 0,02 | -0,51 | 6,12E-01 | 7,60E-01 |
| HEXIM1 | 1365,28 | 0,01 | 0,02 | 0,51 | 6,13E-01 | 7,60E-01 |
| SLC6A16 | 164,22 | -0,02 | 0,04 | -0,51 | 6,13E-01 | 7,61E-01 |
| STIMATE | 87,54 | -0,02 | 0,04 | -0,51 | 6,14E-01 | 7,61E-01 |
| CYB561D2 | 409,27 | 0,01 | 0,02 | 0,50 | 6,14E-01 | 7,61E-01 |
| HAT1 | 662,96 | -0,02 | 0,05 | -0,50 | 6,14E-01 | 7,61E-01 |
| CDIP1 | 242,87 | 0,02 | 0,04 | 0,50 | 6,14E-01 | 7,62E-01 |
| AAK1 | 5522,76 | -0,02 | 0,03 | -0,50 | 6,15E-01 | 7,62E-01 |
| KIAA0930 | 941,22 | 0,03 | 0,05 | 0,50 | 6,15E-01 | 7,62E-01 |
| CCR6 | 143,84 | 0,02 | 0,04 | 0,50 | 6,15E-01 | 7,62E-01 |
| HSD17B7 | 538,92 | 0,01 | 0,02 | 0,50 | 6,15E-01 | 7,63E-01 |
| SESTD1 | 1175,73 | -0,02 | 0,03 | -0,50 | 6,15E-01 | 7,63E-01 |
| IFT81 | 82,33 | -0,02 | 0,05 | -0,50 | 6,16E-01 | 7,63E-01 |
| NUGGC | 279,15 | 0,03 | 0,06 | 0,50 | 6,16E-01 | 7,63E-01 |
| GUCY1A1 | 276,79 | 0,02 | 0,05 | 0,50 | 6,17E-01 | 7,64E-01 |
| KCTD13 | 763,98 | -0,02 | 0,03 | -0,50 | 6,17E-01 | 7,64E-01 |
| ZNF699 | 482,91 | 0,01 | 0,03 | 0,50 | 6,17E-01 | 7,64E-01 |

| | | | | | | |
|---|---|---|---|---|---|---|
| BRCC3 | 586,27 | -0,01 | 0,02 | -0,50 | 6,17E-01 | 7,64E-01 |
| CERS4 | 368,74 | -0,02 | 0,03 | -0,50 | 6,17E-01 | 7,65E-01 |
| CFAP298 | 264,54 | 0,01 | 0,02 | 0,50 | 6,18E-01 | 7,65E-01 |
| HUWE1 | 4759,35 | -0,02 | 0,04 | -0,50 | 6,18E-01 | 7,65E-01 |
| SMG9 | 302,76 | 0,02 | 0,04 | 0,50 | 6,18E-01 | 7,65E-01 |
| STAM | 973,82 | 0,01 | 0,02 | 0,50 | 6,18E-01 | 7,65E-01 |
| HAVCR1 | 81,09 | -0,02 | 0,05 | -0,50 | 6,18E-01 | 7,65E-01 |
| ABHD12 | 334,78 | 0,01 | 0,03 | 0,50 | 6,18E-01 | 7,65E-01 |
| GNPAT | 1280,61 | -0,01 | 0,02 | -0,50 | 6,19E-01 | 7,66E-01 |
| ODR4 | 854,57 | 0,02 | 0,03 | 0,50 | 6,19E-01 | 7,66E-01 |
| COQ9 | 567,09 | 0,01 | 0,01 | 0,50 | 6,19E-01 | 7,66E-01 |
| TBCA | 428,33 | 0,01 | 0,03 | 0,50 | 6,19E-01 | 7,66E-01 |
| FAM120C | 308,18 | 0,01 | 0,02 | 0,50 | 6,20E-01 | 7,66E-01 |
| ZNF285 | 78,55 | 0,03 | 0,06 | 0,50 | 6,20E-01 | 7,67E-01 |
| ATP11C | 1986,97 | 0,01 | 0,02 | 0,50 | 6,20E-01 | 7,67E-01 |
| NHSL1 | 82,27 | -0,04 | 0,08 | -0,50 | 6,20E-01 | 7,67E-01 |
| RAB11FIP3 | 392,85 | 0,02 | 0,04 | 0,50 | 6,20E-01 | 7,67E-01 |
| RIN3 | 1286,30 | -0,03 | 0,06 | -0,50 | 6,20E-01 | 7,67E-01 |
| ABI2 | 896,00 | 0,01 | 0,03 | 0,49 | 6,21E-01 | 7,67E-01 |
| GPHN | 191,54 | 0,02 | 0,04 | 0,49 | 6,21E-01 | 7,67E-01 |
| SSTR3 | 171,42 | 0,03 | 0,05 | 0,49 | 6,21E-01 | 7,68E-01 |
| USO1 | 2482,81 | -0,01 | 0,03 | -0,49 | 6,21E-01 | 7,68E-01 |
| SLC35F6 | 371,56 | -0,02 | 0,04 | -0,49 | 6,21E-01 | 7,68E-01 |
| CMTM5 | 180,65 | 0,03 | 0,07 | 0,49 | 6,21E-01 | 7,68E-01 |
| PDCL | 755,24 | 0,01 | 0,02 | 0,49 | 6,22E-01 | 7,68E-01 |
| ATPSCKMT | 439,37 | -0,01 | 0,02 | -0,49 | 6,22E-01 | 7,68E-01 |
| SLC37A1 | 671,98 | -0,01 | 0,02 | -0,49 | 6,22E-01 | 7,68E-01 |
| CEP95 | 1303,90 | 0,01 | 0,02 | 0,49 | 6,22E-01 | 7,68E-01 |
| WDR25 | 214,54 | -0,01 | 0,03 | -0,49 | 6,22E-01 | 7,68E-01 |
| ADAM20 | 117,20 | -0,02 | 0,05 | -0,49 | 6,22E-01 | 7,68E-01 |
| PDE4D | 1505,68 | 0,02 | 0,03 | 0,49 | 6,23E-01 | 7,68E-01 |
| ENSG000002 | 270,46 | 0,01 | 0,03 | 0,49 | 6,23E-01 | 7,68E-01 |
| PECR | 522,40 | -0,01 | 0,02 | -0,49 | 6,23E-01 | 7,68E-01 |
| UBE2M | 698,84 | 0,01 | 0,03 | 0,49 | 6,23E-01 | 7,69E-01 |
| INSR | 533,31 | 0,02 | 0,04 | 0,49 | 6,23E-01 | 7,69E-01 |
| RO60 | 4666,00 | 0,02 | 0,03 | 0,49 | 6,23E-01 | 7,69E-01 |
| ADNP2 | 583,51 | -0,01 | 0,02 | -0,49 | 6,24E-01 | 7,69E-01 |
| ZNF142 | 342,62 | 0,02 | 0,05 | 0,49 | 6,24E-01 | 7,69E-01 |
| NUP205 | 2286,89 | 0,01 | 0,02 | 0,49 | 6,24E-01 | 7,70E-01 |
| ADGRL1 | 113,39 | 0,04 | 0,08 | 0,49 | 6,25E-01 | 7,70E-01 |
| DDX19A | 314,61 | 0,01 | 0,02 | 0,49 | 6,25E-01 | 7,70E-01 |
| DOCK3 | 125,25 | 0,03 | 0,06 | 0,49 | 6,26E-01 | 7,71E-01 |
| NBPF20 | 99,85 | 0,02 | 0,04 | 0,49 | 6,25E-01 | 7,71E-01 |
| DMWD | 199,03 | 0,02 | 0,05 | 0,49 | 6,25E-01 | 7,71E-01 |
| FDPS | 1168,32 | 0,01 | 0,01 | 0,49 | 6,26E-01 | 7,71E-01 |
| HEATR5B | 3801,57 | -0,01 | 0,02 | -0,49 | 6,26E-01 | 7,71E-01 |

| | | | | | | |
|---|---|---|---|---|---|---|
| RCC2 | 1929,20 | 0,01 | 0,02 | 0,49 | 6,26E-01 | 7,71E-01 |
| KLRB1 | 689,77 | 0,03 | 0,07 | 0,49 | 6,26E-01 | 7,71E-01 |
| ZSWIM4 | 78,30 | 0,03 | 0,06 | 0,49 | 6,26E-01 | 7,71E-01 |
| NPHP4 | 258,33 | -0,03 | 0,07 | -0,49 | 6,26E-01 | 7,71E-01 |
| ESYT1 | 2453,54 | 0,02 | 0,03 | 0,49 | 6,27E-01 | 7,72E-01 |
| C1orf198 | 247,49 | 0,03 | 0,06 | 0,49 | 6,27E-01 | 7,72E-01 |
| DAB2 | 463,01 | -0,02 | 0,03 | -0,49 | 6,27E-01 | 7,72E-01 |
| DNAAF9 | 683,20 | 0,01 | 0,03 | 0,49 | 6,27E-01 | 7,72E-01 |
| PDHA1 | 817,81 | 0,01 | 0,01 | 0,49 | 6,27E-01 | 7,72E-01 |
| STEAP1B | 117,20 | 0,03 | 0,07 | 0,49 | 6,27E-01 | 7,72E-01 |
| POLR1A | 930,52 | 0,01 | 0,03 | 0,49 | 6,27E-01 | 7,72E-01 |
| HIRIP3 | 217,27 | 0,01 | 0,03 | 0,49 | 6,27E-01 | 7,72E-01 |
| ZNF211 | 769,93 | 0,01 | 0,02 | 0,49 | 6,28E-01 | 7,72E-01 |
| PAPSS1 | 1614,25 | -0,02 | 0,04 | -0,49 | 6,28E-01 | 7,72E-01 |
| NISCH | 1639,28 | 0,01 | 0,03 | 0,48 | 6,28E-01 | 7,72E-01 |
| ERP27 | 483,27 | -0,01 | 0,03 | -0,48 | 6,28E-01 | 7,72E-01 |
| PIK3R1 | 5387,71 | -0,01 | 0,03 | -0,48 | 6,28E-01 | 7,72E-01 |
| ASB1 | 1155,90 | 0,01 | 0,02 | 0,48 | 6,28E-01 | 7,72E-01 |
| CFAP251 | 73,55 | -0,04 | 0,08 | -0,48 | 6,28E-01 | 7,72E-01 |
| WNK1 | 8669,06 | 0,02 | 0,04 | 0,48 | 6,28E-01 | 7,72E-01 |
| SCARF1 | 1021,06 | -0,03 | 0,06 | -0,48 | 6,28E-01 | 7,72E-01 |
| LTB4R2 | 323,39 | 0,01 | 0,03 | 0,48 | 6,29E-01 | 7,72E-01 |
| RCHY1 | 1322,48 | 0,02 | 0,03 | 0,48 | 6,29E-01 | 7,72E-01 |
| PIAS2 | 2086,74 | 0,01 | 0,03 | 0,48 | 6,29E-01 | 7,72E-01 |
| RPS6KB1 | 1545,98 | -0,01 | 0,01 | -0,48 | 6,29E-01 | 7,72E-01 |
| EIF4B | 14894,33 | 0,01 | 0,03 | 0,48 | 6,29E-01 | 7,72E-01 |
| SPATA6 | 352,87 | -0,02 | 0,04 | -0,48 | 6,29E-01 | 7,72E-01 |
| MCM3 | 997,70 | -0,01 | 0,02 | -0,48 | 6,29E-01 | 7,72E-01 |
| VKORC1L1 | 645,58 | 0,01 | 0,02 | 0,48 | 6,29E-01 | 7,73E-01 |
| ARHGAP22 | 158,79 | 0,03 | 0,07 | 0,48 | 6,29E-01 | 7,73E-01 |
| POPDC2 | 85,02 | -0,02 | 0,03 | -0,48 | 6,29E-01 | 7,73E-01 |
| ATP9A | 309,06 | -0,03 | 0,05 | -0,48 | 6,30E-01 | 7,73E-01 |
| ZNF623 | 1022,12 | 0,01 | 0,02 | 0,48 | 6,30E-01 | 7,73E-01 |
| RRP8 | 921,92 | 0,01 | 0,02 | 0,48 | 6,30E-01 | 7,74E-01 |
| SLC35E1 | 767,27 | -0,02 | 0,03 | -0,48 | 6,31E-01 | 7,74E-01 |
| KLHL36 | 2847,14 | 0,01 | 0,02 | 0,48 | 6,31E-01 | 7,74E-01 |
| TBC1D10C | 3617,79 | 0,01 | 0,02 | 0,48 | 6,31E-01 | 7,74E-01 |
| ZNF547 | 101,92 | -0,02 | 0,04 | -0,48 | 6,31E-01 | 7,74E-01 |
| PDIA5 | 105,76 | 0,02 | 0,04 | 0,48 | 6,31E-01 | 7,74E-01 |
| C2CD2L | 373,99 | 0,02 | 0,05 | 0,48 | 6,31E-01 | 7,74E-01 |
| CCDC180 | 100,36 | -0,03 | 0,06 | -0,48 | 6,31E-01 | 7,74E-01 |
| MAPK7 | 176,96 | 0,02 | 0,04 | 0,48 | 6,31E-01 | 7,74E-01 |
| YWHAG | 1438,32 | -0,01 | 0,02 | -0,48 | 6,32E-01 | 7,75E-01 |
| DCBLD1 | 226,59 | -0,02 | 0,04 | -0,48 | 6,32E-01 | 7,75E-01 |
| SCCPDH | 331,73 | 0,02 | 0,04 | 0,48 | 6,32E-01 | 7,75E-01 |
| RASA3 | 3538,27 | 0,02 | 0,03 | 0,48 | 6,32E-01 | 7,75E-01 |

| | | | | | | |
|---|---|---|---|---|---|---|
| MTMR8 | 231,60 | 0,01 | 0,03 | 0,48 | 6,33E-01 | 7,75E-01 |
| RREB1 | 2341,69 | -0,02 | 0,03 | -0,48 | 6,33E-01 | 7,75E-01 |
| NKX3-1 | 237,53 | -0,06 | 0,14 | -0,48 | 6,33E-01 | 7,75E-01 |
| NNT | 2124,05 | 0,01 | 0,02 | 0,48 | 6,33E-01 | 7,76E-01 |
| TECPR1 | 1890,22 | 0,01 | 0,03 | 0,48 | 6,33E-01 | 7,76E-01 |
| MTHFD1 | 603,54 | 0,01 | 0,02 | 0,48 | 6,33E-01 | 7,76E-01 |
| FAM222B | 110,89 | -0,02 | 0,04 | -0,48 | 6,33E-01 | 7,76E-01 |
| COCH | 87,66 | -0,04 | 0,09 | -0,48 | 6,34E-01 | 7,76E-01 |
| ACACB | 556,33 | -0,02 | 0,04 | -0,48 | 6,33E-01 | 7,76E-01 |
| TAS2R31 | 296,08 | 0,02 | 0,04 | 0,48 | 6,33E-01 | 7,76E-01 |
| EMC2 | 1075,18 | 0,02 | 0,04 | 0,48 | 6,34E-01 | 7,76E-01 |
| KIFC3 | 224,42 | -0,03 | 0,06 | -0,48 | 6,34E-01 | 7,76E-01 |
| UBE2W | 3430,59 | -0,02 | 0,04 | -0,48 | 6,34E-01 | 7,76E-01 |
| GGPS1 | 320,84 | 0,01 | 0,03 | 0,48 | 6,34E-01 | 7,76E-01 |
| NOL4L | 1324,31 | -0,02 | 0,04 | -0,48 | 6,34E-01 | 7,76E-01 |
| CWF19L1 | 1331,94 | -0,01 | 0,01 | -0,48 | 6,35E-01 | 7,76E-01 |
| CAB39L | 183,61 | -0,01 | 0,03 | -0,48 | 6,34E-01 | 7,76E-01 |
| ANKS6 | 242,58 | 0,02 | 0,04 | 0,48 | 6,34E-01 | 7,76E-01 |
| ST6GAL1 | 3734,18 | 0,01 | 0,03 | 0,48 | 6,35E-01 | 7,76E-01 |
| FTO | 1513,48 | -0,01 | 0,03 | -0,48 | 6,34E-01 | 7,76E-01 |
| ZNF808 | 1082,42 | -0,01 | 0,02 | -0,48 | 6,35E-01 | 7,76E-01 |
| EVA1C | 201,48 | -0,02 | 0,04 | -0,48 | 6,34E-01 | 7,76E-01 |
| MBOAT2 | 2749,18 | -0,02 | 0,04 | -0,48 | 6,35E-01 | 7,76E-01 |
| PNPLA4 | 97,27 | 0,01 | 0,03 | 0,47 | 6,35E-01 | 7,76E-01 |
| PTAR1 | 6692,71 | 0,02 | 0,04 | 0,47 | 6,35E-01 | 7,76E-01 |
| MYO7B | 126,04 | 0,03 | 0,07 | 0,47 | 6,36E-01 | 7,77E-01 |
| DLD | 1704,69 | -0,01 | 0,03 | -0,47 | 6,36E-01 | 7,77E-01 |
| TMEM250 | 1186,88 | 0,01 | 0,02 | 0,47 | 6,36E-01 | 7,77E-01 |
| YES1 | 355,43 | -0,02 | 0,03 | -0,47 | 6,36E-01 | 7,77E-01 |
| ABCD4 | 592,41 | 0,01 | 0,03 | 0,47 | 6,36E-01 | 7,77E-01 |
| EXOG | 560,41 | -0,01 | 0,03 | -0,47 | 6,37E-01 | 7,78E-01 |
| C1orf115 | 74,50 | -0,02 | 0,05 | -0,47 | 6,37E-01 | 7,78E-01 |
| SMN2 | 106,51 | -0,03 | 0,06 | -0,47 | 6,37E-01 | 7,78E-01 |
| RPH3AL | 84,11 | -0,04 | 0,08 | -0,47 | 6,37E-01 | 7,78E-01 |
| CYTH3 | 317,26 | -0,01 | 0,03 | -0,47 | 6,37E-01 | 7,78E-01 |
| CHRAC1 | 238,43 | -0,01 | 0,02 | -0,47 | 6,38E-01 | 7,79E-01 |
| NCR3LG1 | 149,95 | 0,02 | 0,04 | 0,47 | 6,38E-01 | 7,79E-01 |
| CS | 965,55 | 0,01 | 0,03 | 0,47 | 6,38E-01 | 7,79E-01 |
| CYSLTR1 | 1022,09 | -0,01 | 0,03 | -0,47 | 6,38E-01 | 7,79E-01 |
| OSTM1 | 2629,70 | -0,02 | 0,03 | -0,47 | 6,38E-01 | 7,79E-01 |
| PUS3 | 180,98 | 0,01 | 0,03 | 0,47 | 6,39E-01 | 7,79E-01 |
| CMTM4 | 283,23 | -0,01 | 0,03 | -0,47 | 6,39E-01 | 7,79E-01 |
| ZNF544 | 501,93 | 0,01 | 0,02 | 0,47 | 6,39E-01 | 7,79E-01 |
| FAM177B | 119,63 | -0,02 | 0,05 | -0,47 | 6,39E-01 | 7,79E-01 |
| NUP210 | 3243,81 | 0,02 | 0,04 | 0,47 | 6,38E-01 | 7,79E-01 |
| GNRH1 | 116,69 | 0,01 | 0,02 | 0,47 | 6,39E-01 | 7,79E-01 |

| | | | | | | |
|---|---|---|---|---|---|---|
| TMEM241 | 121,53 | -0,02 | 0,05 | -0,47 | 6,39E-01 | 7,79E-01 |
| FHIP1A | 77,68 | -0,03 | 0,07 | -0,47 | 6,39E-01 | 7,79E-01 |
| FMN1 | 670,32 | -0,05 | 0,10 | -0,47 | 6,39E-01 | 7,79E-01 |
| SIDT2 | 566,63 | 0,03 | 0,06 | 0,47 | 6,40E-01 | 7,80E-01 |
| GALNT11 | 1000,06 | -0,01 | 0,02 | -0,47 | 6,40E-01 | 7,80E-01 |
| GRK6 | 3742,63 | -0,01 | 0,03 | -0,47 | 6,40E-01 | 7,80E-01 |
| PHIP | 9313,83 | -0,02 | 0,03 | -0,47 | 6,41E-01 | 7,81E-01 |
| SPATA33 | 129,96 | -0,01 | 0,03 | -0,47 | 6,41E-01 | 7,81E-01 |
| HNRNPL | 3021,90 | -0,01 | 0,01 | -0,47 | 6,41E-01 | 7,81E-01 |
| KATNA1 | 388,61 | 0,01 | 0,02 | 0,47 | 6,41E-01 | 7,81E-01 |
| CKAP5 | 1231,47 | -0,01 | 0,02 | -0,47 | 6,41E-01 | 7,81E-01 |
| FBXO21 | 1163,38 | 0,01 | 0,02 | 0,47 | 6,41E-01 | 7,81E-01 |
| TMEM222 | 321,44 | 0,01 | 0,03 | 0,47 | 6,41E-01 | 7,81E-01 |
| PTBP1 | 2004,69 | -0,01 | 0,03 | -0,47 | 6,42E-01 | 7,82E-01 |
| UBA3 | 3267,43 | 0,02 | 0,03 | 0,46 | 6,42E-01 | 7,82E-01 |
| BEX3 | 269,26 | 0,02 | 0,05 | 0,46 | 6,43E-01 | 7,82E-01 |
| MARK3 | 5361,11 | -0,01 | 0,02 | -0,46 | 6,43E-01 | 7,82E-01 |
| AIG1 | 396,70 | -0,01 | 0,02 | -0,46 | 6,43E-01 | 7,82E-01 |
| CTDSPL | 220,35 | 0,03 | 0,07 | 0,46 | 6,43E-01 | 7,82E-01 |
| KALRN | 194,08 | 0,04 | 0,09 | 0,46 | 6,43E-01 | 7,83E-01 |
| ZNF835 | 93,17 | 0,02 | 0,05 | 0,46 | 6,43E-01 | 7,83E-01 |
| TLR3 | 84,29 | 0,03 | 0,06 | 0,46 | 6,43E-01 | 7,83E-01 |
| AKT2 | 2741,25 | 0,01 | 0,02 | 0,46 | 6,43E-01 | 7,83E-01 |
| POLR2G | 921,97 | 0,01 | 0,02 | 0,46 | 6,44E-01 | 7,83E-01 |
| LEAP2 | 87,35 | -0,01 | 0,03 | -0,46 | 6,44E-01 | 7,83E-01 |
| P4HB | 2261,97 | -0,01 | 0,02 | -0,46 | 6,44E-01 | 7,83E-01 |
| AMMECR1L | 710,38 | -0,01 | 0,02 | -0,46 | 6,44E-01 | 7,83E-01 |
| CDK11A | 199,77 | -0,02 | 0,04 | -0,46 | 6,44E-01 | 7,83E-01 |
| RNF215 | 149,52 | 0,02 | 0,04 | 0,46 | 6,44E-01 | 7,83E-01 |
| LONRF1 | 960,29 | 0,01 | 0,03 | 0,46 | 6,44E-01 | 7,83E-01 |
| ANKDD1B | 75,36 | -0,02 | 0,05 | -0,46 | 6,44E-01 | 7,83E-01 |
| PSMA3 | 792,48 | -0,02 | 0,03 | -0,46 | 6,45E-01 | 7,83E-01 |
| NUDT7 | 71,99 | 0,02 | 0,04 | 0,46 | 6,45E-01 | 7,84E-01 |
| DNAH6 | 148,50 | 0,04 | 0,08 | 0,46 | 6,45E-01 | 7,84E-01 |
| PCNT | 1869,91 | 0,01 | 0,03 | 0,46 | 6,46E-01 | 7,84E-01 |
| ZNG1F | 558,04 | -0,02 | 0,04 | -0,46 | 6,46E-01 | 7,84E-01 |
| SUMF1 | 1407,84 | 0,01 | 0,02 | 0,46 | 6,46E-01 | 7,85E-01 |
| STUB1 | 535,44 | 0,01 | 0,02 | 0,46 | 6,47E-01 | 7,85E-01 |
| TSPYL2 | 365,31 | -0,01 | 0,03 | -0,46 | 6,47E-01 | 7,86E-01 |
| ZC3H10 | 698,51 | 0,01 | 0,01 | 0,46 | 6,47E-01 | 7,86E-01 |
| NCL | 3858,91 | -0,01 | 0,03 | -0,46 | 6,48E-01 | 7,86E-01 |
| CPLANE1 | 1019,94 | 0,01 | 0,03 | 0,46 | 6,48E-01 | 7,87E-01 |
| CTSW | 378,98 | 0,03 | 0,06 | 0,46 | 6,48E-01 | 7,87E-01 |
| BTBD8 | 321,20 | 0,02 | 0,03 | 0,46 | 6,48E-01 | 7,87E-01 |
| DNAL4 | 132,10 | 0,01 | 0,02 | 0,46 | 6,48E-01 | 7,87E-01 |
| TAS2R43 | 89,30 | 0,05 | 0,11 | 0,46 | 6,49E-01 | 7,87E-01 |

| | | | | | | |
|---|---|---|---|---|---|---|
| TBC1D23 | 2345,37 | -0,01 | 0,03 | -0,46 | 6,49E-01 | 7,87E-01 |
| SPDL1 | 203,17 | 0,01 | 0,03 | 0,46 | 6,49E-01 | 7,87E-01 |
| SUOX | 142,81 | -0,02 | 0,04 | -0,46 | 6,49E-01 | 7,87E-01 |
| HGF | 523,70 | 0,02 | 0,04 | 0,46 | 6,49E-01 | 7,87E-01 |
| SRC | 140,81 | 0,04 | 0,08 | 0,46 | 6,49E-01 | 7,87E-01 |
| ZC3H3 | 410,23 | -0,02 | 0,05 | -0,46 | 6,49E-01 | 7,87E-01 |
| MS4A6A | 4963,49 | 0,02 | 0,04 | 0,46 | 6,49E-01 | 7,87E-01 |
| TMEM170A | 1673,67 | -0,01 | 0,02 | -0,45 | 6,49E-01 | 7,87E-01 |
| ZBTB38 | 2433,97 | 0,01 | 0,03 | 0,45 | 6,49E-01 | 7,87E-01 |
| FARS2 | 677,39 | 0,01 | 0,02 | 0,45 | 6,49E-01 | 7,87E-01 |
| SIN3B | 618,67 | -0,03 | 0,06 | -0,45 | 6,50E-01 | 7,87E-01 |
| HIVEP2 | 3356,16 | 0,01 | 0,02 | 0,45 | 6,50E-01 | 7,88E-01 |
| GADD45A | 285,71 | 0,02 | 0,04 | 0,45 | 6,50E-01 | 7,88E-01 |
| MYO18A | 1744,62 | 0,02 | 0,04 | 0,45 | 6,50E-01 | 7,88E-01 |
| GPCPD1 | 11604,71 | -0,01 | 0,03 | -0,45 | 6,50E-01 | 7,88E-01 |
| GHITM | 4942,15 | -0,01 | 0,03 | -0,45 | 6,50E-01 | 7,88E-01 |
| ME3 | 89,97 | 0,03 | 0,06 | 0,45 | 6,51E-01 | 7,88E-01 |
| MVB12A | 78,64 | 0,03 | 0,06 | 0,45 | 6,51E-01 | 7,88E-01 |
| AP1B1 | 548,39 | 0,02 | 0,05 | 0,45 | 6,52E-01 | 7,89E-01 |
| ZBED10P | 189,13 | -0,02 | 0,04 | -0,45 | 6,52E-01 | 7,89E-01 |
| XRRA1 | 746,58 | -0,03 | 0,07 | -0,45 | 6,52E-01 | 7,89E-01 |
| ALKBH1 | 379,56 | 0,01 | 0,01 | 0,45 | 6,52E-01 | 7,89E-01 |
| UBOX5 | 195,52 | -0,02 | 0,04 | -0,45 | 6,52E-01 | 7,89E-01 |
| SUN2 | 8711,03 | 0,01 | 0,02 | 0,45 | 6,52E-01 | 7,89E-01 |
| POLG | 3321,11 | -0,01 | 0,02 | -0,45 | 6,52E-01 | 7,89E-01 |
| HADHA | 3763,96 | -0,01 | 0,02 | -0,45 | 6,52E-01 | 7,89E-01 |
| ZNF30 | 113,74 | 0,02 | 0,04 | 0,45 | 6,53E-01 | 7,89E-01 |
| HP | 100,64 | -0,04 | 0,09 | -0,45 | 6,53E-01 | 7,90E-01 |
| NSDHL | 135,34 | 0,01 | 0,02 | 0,45 | 6,53E-01 | 7,90E-01 |
| MED22 | 1293,44 | 0,01 | 0,02 | 0,45 | 6,53E-01 | 7,90E-01 |
| DNAH8 | 97,11 | -0,05 | 0,12 | -0,45 | 6,53E-01 | 7,90E-01 |
| CLEC16A | 1345,29 | 0,02 | 0,03 | 0,45 | 6,53E-01 | 7,90E-01 |
| HAUS8 | 186,14 | -0,01 | 0,02 | -0,45 | 6,53E-01 | 7,90E-01 |
| KCNH2 | 93,67 | 0,03 | 0,08 | 0,45 | 6,53E-01 | 7,90E-01 |
| FEN1 | 218,53 | -0,01 | 0,03 | -0,45 | 6,53E-01 | 7,90E-01 |
| RMI1 | 371,39 | -0,01 | 0,03 | -0,45 | 6,54E-01 | 7,90E-01 |
| NCKAP5L | 256,36 | 0,02 | 0,05 | 0,45 | 6,54E-01 | 7,90E-01 |
| HELB | 4489,74 | -0,01 | 0,02 | -0,45 | 6,54E-01 | 7,90E-01 |
| RBM4B | 403,12 | -0,01 | 0,01 | -0,45 | 6,54E-01 | 7,90E-01 |
| LIMK1 | 246,88 | 0,02 | 0,05 | 0,45 | 6,54E-01 | 7,90E-01 |
| SUGP2 | 2100,33 | 0,01 | 0,03 | 0,45 | 6,54E-01 | 7,90E-01 |
| RBM27 | 961,65 | 0,01 | 0,03 | 0,45 | 6,54E-01 | 7,90E-01 |
| PRKN | 175,65 | 0,02 | 0,04 | 0,45 | 6,55E-01 | 7,91E-01 |
| SEMA7A | 294,46 | 0,02 | 0,04 | 0,45 | 6,55E-01 | 7,91E-01 |
| SEPTIN8 | 173,50 | 0,02 | 0,04 | 0,45 | 6,55E-01 | 7,91E-01 |
| UBALD1 | 234,39 | 0,02 | 0,04 | 0,45 | 6,55E-01 | 7,91E-01 |

| | | | | | | |
|---|---|---|---|---|---|---|
| CNDP2 | 2185,10 | -0,01 | 0,02 | -0,45 | 6,55E-01 | 7,91E-01 |
| AHDC1 | 119,23 | 0,03 | 0,07 | 0,45 | 6,55E-01 | 7,91E-01 |
| DELE1 | 1140,87 | -0,01 | 0,03 | -0,45 | 6,56E-01 | 7,92E-01 |
| ZNF543 | 394,44 | 0,01 | 0,02 | 0,44 | 6,56E-01 | 7,92E-01 |
| HMG20B | 372,31 | 0,01 | 0,03 | 0,44 | 6,57E-01 | 7,92E-01 |
| STRN3 | 1433,05 | 0,01 | 0,02 | 0,44 | 6,57E-01 | 7,92E-01 |
| RASGEF1B | 746,03 | 0,01 | 0,03 | 0,44 | 6,57E-01 | 7,92E-01 |
| SLC38A2 | 7099,05 | -0,02 | 0,04 | -0,44 | 6,57E-01 | 7,92E-01 |
| ARPP19 | 4825,10 | 0,01 | 0,03 | 0,44 | 6,57E-01 | 7,92E-01 |
| INPP1 | 303,72 | -0,01 | 0,03 | -0,44 | 6,57E-01 | 7,92E-01 |
| SMNDC1 | 1095,31 | 0,01 | 0,03 | 0,44 | 6,57E-01 | 7,92E-01 |
| TJP2 | 178,74 | -0,02 | 0,05 | -0,44 | 6,57E-01 | 7,92E-01 |
| PRCC | 422,14 | -0,01 | 0,02 | -0,44 | 6,57E-01 | 7,92E-01 |
| FANCI | 394,39 | -0,01 | 0,03 | -0,44 | 6,58E-01 | 7,93E-01 |
| PPP2R2B | 400,99 | -0,02 | 0,05 | -0,44 | 6,58E-01 | 7,93E-01 |
| MGAT3 | 106,06 | 0,03 | 0,06 | 0,44 | 6,58E-01 | 7,93E-01 |
| CCHCR1 | 266,20 | -0,01 | 0,03 | -0,44 | 6,58E-01 | 7,93E-01 |
| UBE2J1 | 2622,38 | 0,01 | 0,03 | 0,44 | 6,58E-01 | 7,93E-01 |
| RASSF4 | 1236,73 | 0,02 | 0,04 | 0,44 | 6,59E-01 | 7,94E-01 |
| SKAP1 | 818,64 | 0,02 | 0,04 | 0,44 | 6,59E-01 | 7,94E-01 |
| DTX4 | 275,97 | -0,02 | 0,04 | -0,44 | 6,59E-01 | 7,94E-01 |
| PSMD4 | 3008,07 | -0,01 | 0,03 | -0,44 | 6,59E-01 | 7,94E-01 |
| EDC4 | 468,96 | 0,02 | 0,04 | 0,44 | 6,59E-01 | 7,94E-01 |
| FAM50B | 186,09 | 0,01 | 0,03 | 0,44 | 6,59E-01 | 7,94E-01 |
| ADARB1 | 865,09 | 0,02 | 0,04 | 0,44 | 6,59E-01 | 7,94E-01 |
| MORN3 | 137,28 | 0,02 | 0,04 | 0,44 | 6,60E-01 | 7,94E-01 |
| MTERF4 | 1988,59 | 0,01 | 0,02 | 0,44 | 6,60E-01 | 7,94E-01 |
| XKR6 | 327,93 | -0,02 | 0,03 | -0,44 | 6,60E-01 | 7,94E-01 |
| CLASRP | 378,07 | -0,02 | 0,05 | -0,44 | 6,60E-01 | 7,94E-01 |
| SMG6 | 1052,45 | -0,02 | 0,03 | -0,44 | 6,60E-01 | 7,94E-01 |
| EPPK1 | 267,32 | -0,03 | 0,06 | -0,44 | 6,61E-01 | 7,94E-01 |
| PATL2 | 1263,15 | -0,02 | 0,04 | -0,44 | 6,60E-01 | 7,94E-01 |
| CENPO | 165,16 | -0,01 | 0,03 | -0,44 | 6,60E-01 | 7,94E-01 |
| CCT6A | 1819,94 | 0,01 | 0,03 | 0,44 | 6,60E-01 | 7,94E-01 |
| ABHD15 | 596,39 | 0,01 | 0,02 | 0,44 | 6,60E-01 | 7,94E-01 |
| HECTD1 | 6127,64 | -0,01 | 0,02 | -0,44 | 6,60E-01 | 7,94E-01 |
| RUSC2 | 119,24 | 0,03 | 0,06 | 0,44 | 6,61E-01 | 7,94E-01 |
| ZNF320 | 110,95 | 0,02 | 0,03 | 0,44 | 6,61E-01 | 7,94E-01 |
| PIK3CA | 3007,62 | -0,01 | 0,03 | -0,44 | 6,60E-01 | 7,94E-01 |
| NUP62 | 634,26 | 0,02 | 0,03 | 0,44 | 6,60E-01 | 7,94E-01 |
| CUL7 | 250,25 | 0,02 | 0,04 | 0,44 | 6,61E-01 | 7,94E-01 |
| AUH | 848,36 | -0,01 | 0,02 | -0,44 | 6,61E-01 | 7,94E-01 |
| PNRC2 | 3872,89 | 0,02 | 0,04 | 0,44 | 6,61E-01 | 7,94E-01 |
| CD164 | 9018,68 | 0,01 | 0,03 | 0,44 | 6,61E-01 | 7,94E-01 |
| NAPA | 1013,44 | -0,02 | 0,05 | -0,44 | 6,61E-01 | 7,95E-01 |
| GTF2F2 | 624,30 | -0,01 | 0,03 | -0,44 | 6,62E-01 | 7,95E-01 |

| | | | | | | |
|---|---|---|---|---|---|---|
| DPF3 | 105,97 | 0,03 | 0,07 | 0,44 | 6,62E-01 | 7,95E-01 |
| TXNRD3 | 78,80 | 0,03 | 0,06 | 0,44 | 6,62E-01 | 7,95E-01 |
| PLXNA1 | 207,67 | 0,03 | 0,07 | 0,44 | 6,62E-01 | 7,95E-01 |
| PRKX | 1968,78 | -0,01 | 0,02 | -0,44 | 6,62E-01 | 7,95E-01 |
| ZNF341 | 110,26 | -0,03 | 0,06 | -0,44 | 6,62E-01 | 7,95E-01 |
| SF3A3 | 1654,38 | -0,01 | 0,02 | -0,44 | 6,62E-01 | 7,95E-01 |
| HTATSF1 | 836,65 | -0,01 | 0,03 | -0,44 | 6,62E-01 | 7,95E-01 |
| TRIM62 | 163,54 | -0,03 | 0,06 | -0,44 | 6,62E-01 | 7,95E-01 |
| FZD6 | 123,23 | 0,02 | 0,05 | 0,44 | 6,62E-01 | 7,95E-01 |
| GSAP | 3386,56 | 0,01 | 0,03 | 0,44 | 6,62E-01 | 7,95E-01 |
| ENKUR | 129,17 | 0,03 | 0,06 | 0,44 | 6,62E-01 | 7,95E-01 |
| SF3B1 | 21147,96 | -0,01 | 0,03 | -0,44 | 6,63E-01 | 7,95E-01 |
| VWA5A | 241,04 | 0,01 | 0,03 | 0,44 | 6,63E-01 | 7,95E-01 |
| SSBP3 | 565,38 | 0,02 | 0,05 | 0,44 | 6,63E-01 | 7,96E-01 |
| SENP5 | 3168,69 | -0,01 | 0,01 | -0,43 | 6,64E-01 | 7,96E-01 |
| QTRT2 | 1146,67 | -0,01 | 0,01 | -0,43 | 6,64E-01 | 7,96E-01 |
| HIC2 | 390,92 | 0,01 | 0,02 | 0,43 | 6,64E-01 | 7,96E-01 |
| TRAPPC11 | 2163,07 | -0,01 | 0,02 | -0,43 | 6,64E-01 | 7,96E-01 |
| ENSG000002 | 84,66 | 0,02 | 0,04 | 0,43 | 6,65E-01 | 7,97E-01 |
| ARID5B | 2093,80 | 0,01 | 0,03 | 0,43 | 6,65E-01 | 7,97E-01 |
| AP3M1 | 1435,75 | 0,01 | 0,02 | 0,43 | 6,65E-01 | 7,97E-01 |
| ANKRD50 | 856,27 | 0,01 | 0,03 | 0,43 | 6,65E-01 | 7,97E-01 |
| SPPL3 | 836,31 | -0,01 | 0,03 | -0,43 | 6,65E-01 | 7,97E-01 |
| CEP128 | 689,99 | 0,01 | 0,03 | 0,43 | 6,66E-01 | 7,97E-01 |
| SCRN1 | 911,99 | 0,01 | 0,03 | 0,43 | 6,66E-01 | 7,97E-01 |
| BMF | 538,64 | 0,02 | 0,04 | 0,43 | 6,66E-01 | 7,97E-01 |
| PCCA | 572,53 | -0,01 | 0,03 | -0,43 | 6,65E-01 | 7,97E-01 |
| LMBR1 | 2543,13 | 0,01 | 0,02 | 0,43 | 6,66E-01 | 7,98E-01 |
| SARS1 | 2291,22 | -0,01 | 0,02 | -0,43 | 6,66E-01 | 7,98E-01 |
| UACA | 154,95 | -0,03 | 0,07 | -0,43 | 6,67E-01 | 7,98E-01 |
| MSANTD2 | 622,94 | 0,01 | 0,02 | 0,43 | 6,67E-01 | 7,98E-01 |
| PINK1 | 3837,07 | 0,02 | 0,05 | 0,43 | 6,66E-01 | 7,98E-01 |
| SLC5A3 | 4574,62 | -0,01 | 0,03 | -0,43 | 6,67E-01 | 7,98E-01 |
| EFTUD2 | 1292,33 | 0,01 | 0,02 | 0,43 | 6,67E-01 | 7,98E-01 |
| TOX | 219,62 | 0,02 | 0,04 | 0,43 | 6,66E-01 | 7,98E-01 |
| NF2 | 832,09 | 0,01 | 0,03 | 0,43 | 6,67E-01 | 7,98E-01 |
| EHD4 | 695,17 | -0,01 | 0,02 | -0,43 | 6,67E-01 | 7,98E-01 |
| CENPJ | 286,05 | 0,01 | 0,03 | 0,43 | 6,67E-01 | 7,98E-01 |
| DENR | 1162,97 | -0,01 | 0,02 | -0,43 | 6,68E-01 | 7,99E-01 |
| PRSS23 | 498,85 | -0,03 | 0,06 | -0,43 | 6,68E-01 | 7,99E-01 |
| ITPR1 | 6513,43 | -0,01 | 0,02 | -0,43 | 6,68E-01 | 7,99E-01 |
| UBASH3B | 1203,71 | -0,01 | 0,02 | -0,43 | 6,68E-01 | 7,99E-01 |
| TTLL11 | 145,65 | 0,01 | 0,03 | 0,43 | 6,69E-01 | 8,00E-01 |
| SERPINB9 | 2463,75 | 0,01 | 0,02 | 0,43 | 6,69E-01 | 8,00E-01 |
| NCK1 | 1133,81 | 0,01 | 0,03 | 0,43 | 6,69E-01 | 8,00E-01 |
| OAF | 131,65 | 0,02 | 0,05 | 0,43 | 6,70E-01 | 8,00E-01 |

| | | | | | | |
|---|---|---|---|---|---|---|
| PYHIN1 | 1603,70 | -0,02 | 0,04 | -0,43 | 6,70E-01 | 8,01E-01 |
| ZNF316 | 1606,32 | -0,01 | 0,03 | -0,43 | 6,70E-01 | 8,01E-01 |
| TIGD7 | 478,68 | 0,02 | 0,05 | 0,43 | 6,70E-01 | 8,01E-01 |
| GANAB | 1697,41 | -0,01 | 0,03 | -0,43 | 6,71E-01 | 8,01E-01 |
| RDH13 | 259,27 | 0,01 | 0,04 | 0,42 | 6,71E-01 | 8,01E-01 |
| CCDC88C | 4699,47 | 0,01 | 0,03 | 0,42 | 6,71E-01 | 8,02E-01 |
| SKIC2 | 878,94 | -0,01 | 0,03 | -0,42 | 6,71E-01 | 8,02E-01 |
| SYT11 | 677,75 | 0,01 | 0,03 | 0,42 | 6,71E-01 | 8,02E-01 |
| DENND5B | 283,05 | 0,02 | 0,05 | 0,42 | 6,72E-01 | 8,02E-01 |
| YLPM1 | 3199,70 | -0,01 | 0,02 | -0,42 | 6,72E-01 | 8,03E-01 |
| RNASEK | 129,35 | 0,01 | 0,03 | 0,42 | 6,72E-01 | 8,03E-01 |
| TEC | 424,54 | 0,01 | 0,03 | 0,42 | 6,72E-01 | 8,03E-01 |
| DIS3 | 2330,54 | 0,01 | 0,03 | 0,42 | 6,72E-01 | 8,03E-01 |
| ACTR8 | 1247,26 | -0,01 | 0,02 | -0,42 | 6,73E-01 | 8,03E-01 |
| CD151 | 491,67 | -0,02 | 0,05 | -0,42 | 6,73E-01 | 8,03E-01 |
| ABL1 | 354,03 | -0,02 | 0,04 | -0,42 | 6,73E-01 | 8,03E-01 |
| GTF2I | 932,55 | -0,01 | 0,02 | -0,42 | 6,73E-01 | 8,03E-01 |
| ZNF613 | 282,79 | 0,02 | 0,04 | 0,42 | 6,73E-01 | 8,03E-01 |
| ZNF718 | 646,62 | 0,01 | 0,03 | 0,42 | 6,74E-01 | 8,04E-01 |
| DKC1 | 846,86 | -0,01 | 0,02 | -0,42 | 6,74E-01 | 8,04E-01 |
| BTN3A2 | 4207,00 | -0,02 | 0,04 | -0,42 | 6,74E-01 | 8,04E-01 |
| ARL15 | 1072,97 | 0,01 | 0,03 | 0,42 | 6,74E-01 | 8,04E-01 |
| FAM234B | 214,32 | -0,01 | 0,03 | -0,42 | 6,74E-01 | 8,04E-01 |
| COG3 | 2235,04 | 0,01 | 0,02 | 0,42 | 6,74E-01 | 8,04E-01 |
| TNFRSF18 | 103,65 | -0,02 | 0,05 | -0,42 | 6,75E-01 | 8,04E-01 |
| VPS33B | 102,26 | -0,01 | 0,02 | -0,42 | 6,75E-01 | 8,04E-01 |
| NDUFA8 | 246,66 | 0,01 | 0,03 | 0,42 | 6,75E-01 | 8,04E-01 |
| QDPR | 117,78 | 0,02 | 0,04 | 0,42 | 6,75E-01 | 8,04E-01 |
| RCAN1 | 326,95 | -0,01 | 0,02 | -0,42 | 6,75E-01 | 8,04E-01 |
| TNXB | 122,76 | 0,04 | 0,09 | 0,42 | 6,75E-01 | 8,04E-01 |
| KEAP1 | 513,90 | 0,01 | 0,02 | 0,42 | 6,75E-01 | 8,04E-01 |
| ZFP41 | 128,12 | 0,02 | 0,05 | 0,42 | 6,75E-01 | 8,04E-01 |
| TOB2 | 329,35 | -0,02 | 0,04 | -0,42 | 6,76E-01 | 8,05E-01 |
| NELFB | 941,35 | 0,01 | 0,02 | 0,42 | 6,76E-01 | 8,05E-01 |
| GTF3C2 | 796,63 | -0,01 | 0,03 | -0,42 | 6,76E-01 | 8,05E-01 |
| OSBPL8 | 14839,17 | -0,02 | 0,04 | -0,42 | 6,76E-01 | 8,05E-01 |
| LIG4 | 597,76 | 0,02 | 0,05 | 0,42 | 6,76E-01 | 8,05E-01 |
| HUS1B | 79,43 | -0,02 | 0,04 | -0,42 | 6,77E-01 | 8,05E-01 |
| RNF214 | 309,09 | 0,01 | 0,03 | 0,42 | 6,77E-01 | 8,05E-01 |
| SIL1 | 569,83 | 0,01 | 0,02 | 0,42 | 6,77E-01 | 8,05E-01 |
| PEDS1 | 239,54 | 0,01 | 0,04 | 0,42 | 6,77E-01 | 8,05E-01 |
| UBP1 | 1935,69 | 0,01 | 0,02 | 0,42 | 6,77E-01 | 8,06E-01 |
| GALNT10 | 1978,87 | -0,01 | 0,03 | -0,42 | 6,78E-01 | 8,06E-01 |
| RHOBTB1 | 206,73 | -0,02 | 0,06 | -0,42 | 6,78E-01 | 8,06E-01 |
| ARPC5L | 633,27 | 0,01 | 0,02 | 0,42 | 6,78E-01 | 8,06E-01 |
| ARHGAP1 | 594,14 | -0,02 | 0,05 | -0,42 | 6,78E-01 | 8,06E-01 |

| | | | | | | |
|---|---|---|---|---|---|---|
| ESF1 | 361,61 | 0,02 | 0,06 | 0,42 | 6,78E-01 | 8,07E-01 |
| TUBA1B | 3351,66 | -0,01 | 0,02 | -0,41 | 6,78E-01 | 8,07E-01 |
| LSM10 | 425,07 | -0,01 | 0,03 | -0,41 | 6,79E-01 | 8,07E-01 |
| PSMA2 | 397,64 | 0,01 | 0,02 | 0,41 | 6,79E-01 | 8,07E-01 |
| RASGEF1A | 230,82 | -0,02 | 0,04 | -0,41 | 6,79E-01 | 8,07E-01 |
| ZNF821 | 158,42 | 0,01 | 0,03 | 0,41 | 6,79E-01 | 8,07E-01 |
| OGG1 | 580,63 | 0,01 | 0,03 | 0,41 | 6,79E-01 | 8,07E-01 |
| MB21D2 | 124,19 | 0,02 | 0,04 | 0,41 | 6,80E-01 | 8,08E-01 |
| LARP1 | 2138,37 | 0,01 | 0,04 | 0,41 | 6,80E-01 | 8,08E-01 |
| ARMC8 | 2699,54 | -0,01 | 0,02 | -0,41 | 6,80E-01 | 8,08E-01 |
| TXLNA | 1050,36 | 0,01 | 0,02 | 0,41 | 6,80E-01 | 8,08E-01 |
| PGAP2 | 169,96 | -0,01 | 0,03 | -0,41 | 6,80E-01 | 8,08E-01 |
| PAK4 | 79,17 | 0,03 | 0,08 | 0,41 | 6,80E-01 | 8,08E-01 |
| COL7A1 | 92,25 | -0,04 | 0,10 | -0,41 | 6,80E-01 | 8,08E-01 |
| DNAJC25 | 154,20 | 0,01 | 0,03 | 0,41 | 6,81E-01 | 8,08E-01 |
| MAPK8IP3 | 2219,88 | 0,02 | 0,04 | 0,41 | 6,81E-01 | 8,08E-01 |
| HNRNPLL | 2104,29 | -0,01 | 0,03 | -0,41 | 6,81E-01 | 8,08E-01 |
| PBX3 | 718,11 | 0,01 | 0,02 | 0,41 | 6,81E-01 | 8,08E-01 |
| LATS1 | 1748,28 | 0,01 | 0,02 | 0,41 | 6,81E-01 | 8,08E-01 |
| PSMB2 | 618,36 | 0,01 | 0,02 | 0,41 | 6,81E-01 | 8,09E-01 |
| DIP2A | 2911,15 | -0,02 | 0,04 | -0,41 | 6,81E-01 | 8,09E-01 |
| MED20 | 412,35 | -0,01 | 0,01 | -0,41 | 6,82E-01 | 8,09E-01 |
| IGFBP7 | 329,49 | 0,02 | 0,04 | 0,41 | 6,82E-01 | 8,09E-01 |
| TAMM41 | 433,22 | -0,02 | 0,04 | -0,41 | 6,82E-01 | 8,09E-01 |
| FIP1L1 | 1315,34 | 0,01 | 0,02 | 0,41 | 6,82E-01 | 8,09E-01 |
| ZNF362 | 310,57 | 0,02 | 0,04 | 0,41 | 6,82E-01 | 8,09E-01 |
| INVS | 923,00 | 0,01 | 0,02 | 0,41 | 6,83E-01 | 8,11E-01 |
| DNAJC30 | 91,79 | 0,02 | 0,04 | 0,41 | 6,84E-01 | 8,11E-01 |
| ELL2 | 599,57 | 0,01 | 0,03 | 0,41 | 6,84E-01 | 8,11E-01 |
| ADRM1 | 408,73 | 0,01 | 0,03 | 0,41 | 6,84E-01 | 8,11E-01 |
| ENO1 | 7717,17 | -0,01 | 0,03 | -0,41 | 6,84E-01 | 8,11E-01 |
| PCNA | 591,12 | 0,01 | 0,03 | 0,41 | 6,84E-01 | 8,11E-01 |
| ITGB5 | 376,22 | 0,03 | 0,07 | 0,41 | 6,84E-01 | 8,11E-01 |
| ZDHHC6 | 1403,20 | 0,01 | 0,02 | 0,41 | 6,84E-01 | 8,11E-01 |
| ATP5PB | 2189,01 | 0,02 | 0,04 | 0,41 | 6,84E-01 | 8,11E-01 |
| MTA2 | 2202,95 | -0,01 | 0,02 | -0,41 | 6,85E-01 | 8,11E-01 |
| IFFO1 | 687,49 | -0,02 | 0,04 | -0,41 | 6,85E-01 | 8,11E-01 |
| SLC39A6 | 704,77 | -0,01 | 0,02 | -0,41 | 6,85E-01 | 8,11E-01 |
| SSRP1 | 730,92 | 0,01 | 0,02 | 0,41 | 6,85E-01 | 8,11E-01 |
| KRBA1 | 128,55 | 0,02 | 0,05 | 0,41 | 6,85E-01 | 8,11E-01 |
| C2orf92 | 139,41 | 0,02 | 0,04 | 0,41 | 6,85E-01 | 8,11E-01 |
| VGLL4 | 613,74 | -0,01 | 0,02 | -0,40 | 6,86E-01 | 8,12E-01 |
| RGPD8 | 300,87 | -0,02 | 0,05 | -0,40 | 6,86E-01 | 8,12E-01 |
| DLG3 | 252,12 | -0,02 | 0,04 | -0,40 | 6,86E-01 | 8,12E-01 |
| EPB41L5 | 330,60 | -0,01 | 0,03 | -0,40 | 6,87E-01 | 8,13E-01 |
| MARCKS | 2702,10 | -0,02 | 0,04 | -0,40 | 6,87E-01 | 8,13E-01 |

| | | | | | | |
|---|---|---|---|---|---|---|
| ZNF667 | 84,91 | 0,03 | 0,08 | 0,40 | 6,87E-01 | 8,13E-01 |
| C8orf88 | 75,10 | 0,02 | 0,05 | 0,40 | 6,88E-01 | 8,14E-01 |
| AZIN2 | 143,98 | 0,02 | 0,04 | 0,40 | 6,88E-01 | 8,14E-01 |
| PROSER3 | 97,03 | 0,03 | 0,06 | 0,40 | 6,88E-01 | 8,14E-01 |
| SHTN1 | 570,24 | 0,02 | 0,04 | 0,40 | 6,89E-01 | 8,14E-01 |
| ZSWIM5 | 134,30 | 0,02 | 0,05 | 0,40 | 6,88E-01 | 8,14E-01 |
| ABHD18 | 1530,91 | 0,01 | 0,02 | 0,40 | 6,89E-01 | 8,14E-01 |
| PEX26 | 1318,64 | 0,01 | 0,03 | 0,40 | 6,89E-01 | 8,14E-01 |
| BBX | 4386,74 | -0,01 | 0,02 | -0,40 | 6,89E-01 | 8,14E-01 |
| ENSG000002 | 278,53 | -0,02 | 0,05 | -0,40 | 6,89E-01 | 8,14E-01 |
| PRPF19 | 1344,14 | 0,01 | 0,02 | 0,40 | 6,88E-01 | 8,14E-01 |
| ANKRD40 | 521,66 | 0,01 | 0,02 | 0,40 | 6,89E-01 | 8,14E-01 |
| ATG9A | 453,58 | 0,01 | 0,04 | 0,40 | 6,89E-01 | 8,14E-01 |
| ZNF782 | 840,73 | 0,01 | 0,02 | 0,40 | 6,89E-01 | 8,14E-01 |
| ACBD3 | 1684,57 | 0,01 | 0,02 | 0,40 | 6,89E-01 | 8,14E-01 |
| ZNF831 | 1846,44 | 0,01 | 0,03 | 0,40 | 6,89E-01 | 8,15E-01 |
| PHF1 | 631,66 | -0,01 | 0,03 | -0,40 | 6,90E-01 | 8,15E-01 |
| KCNK6 | 602,98 | 0,01 | 0,03 | 0,40 | 6,90E-01 | 8,15E-01 |
| PSMC2 | 810,28 | 0,01 | 0,04 | 0,40 | 6,90E-01 | 8,15E-01 |
| TRIM68 | 319,77 | -0,01 | 0,02 | -0,40 | 6,90E-01 | 8,15E-01 |
| BRD3OS | 117,34 | -0,01 | 0,04 | -0,40 | 6,90E-01 | 8,15E-01 |
| PLXND1 | 588,01 | -0,03 | 0,06 | -0,40 | 6,91E-01 | 8,15E-01 |
| HHAT | 224,03 | 0,02 | 0,04 | 0,40 | 6,91E-01 | 8,16E-01 |
| DLEC1 | 161,69 | 0,02 | 0,05 | 0,40 | 6,91E-01 | 8,16E-01 |
| PARK7 | 1797,41 | 0,01 | 0,03 | 0,40 | 6,91E-01 | 8,16E-01 |
| TAF1B | 607,02 | 0,01 | 0,03 | 0,40 | 6,91E-01 | 8,16E-01 |
| LTV1 | 226,96 | 0,01 | 0,03 | 0,40 | 6,91E-01 | 8,16E-01 |
| INPP5B | 1664,70 | -0,01 | 0,02 | -0,40 | 6,91E-01 | 8,16E-01 |
| LOXL3 | 206,88 | 0,02 | 0,04 | 0,40 | 6,92E-01 | 8,16E-01 |
| NUDCD3 | 2496,20 | 0,01 | 0,02 | 0,40 | 6,92E-01 | 8,16E-01 |
| PIK3R6 | 324,60 | 0,02 | 0,06 | 0,40 | 6,92E-01 | 8,16E-01 |
| TM9SF3 | 4531,19 | 0,01 | 0,03 | 0,40 | 6,92E-01 | 8,16E-01 |
| NKG7 | 594,74 | 0,02 | 0,06 | 0,40 | 6,92E-01 | 8,17E-01 |
| EIF3A | 6235,22 | -0,01 | 0,02 | -0,40 | 6,93E-01 | 8,17E-01 |
| SSU72 | 2169,68 | -0,01 | 0,02 | -0,40 | 6,92E-01 | 8,17E-01 |
| SEC61A1 | 2909,14 | -0,01 | 0,02 | -0,40 | 6,92E-01 | 8,17E-01 |
| MON2 | 6549,05 | -0,01 | 0,02 | -0,39 | 6,93E-01 | 8,17E-01 |
| CNIH3 | 190,64 | 0,02 | 0,04 | 0,39 | 6,93E-01 | 8,17E-01 |
| PDS5B | 3271,82 | -0,01 | 0,03 | -0,39 | 6,94E-01 | 8,17E-01 |
| GDPD5 | 419,07 | 0,02 | 0,05 | 0,39 | 6,94E-01 | 8,18E-01 |
| THADA | 3078,48 | -0,01 | 0,02 | -0,39 | 6,94E-01 | 8,18E-01 |
| OARD1 | 1119,29 | 0,01 | 0,02 | 0,39 | 6,94E-01 | 8,18E-01 |
| KNSTRN | 310,41 | 0,01 | 0,03 | 0,39 | 6,95E-01 | 8,18E-01 |
| KANK1 | 372,21 | -0,02 | 0,05 | -0,39 | 6,95E-01 | 8,18E-01 |
| HNRNPD | 3969,24 | -0,01 | 0,02 | -0,39 | 6,95E-01 | 8,19E-01 |
| MIDEAS | 2185,79 | -0,01 | 0,04 | -0,39 | 6,96E-01 | 8,19E-01 |

| | | | | | | |
|---|---|---|---|---|---|---|
| SFPQ | 10392,00 | -0,01 | 0,02 | -0,39 | 6,96E-01 | 8,19E-01 |
| PTPN9 | 698,61 | -0,01 | 0,02 | -0,39 | 6,96E-01 | 8,19E-01 |
| PDE4A | 97,50 | -0,03 | 0,07 | -0,39 | 6,95E-01 | 8,19E-01 |
| SLFN12 | 785,10 | 0,01 | 0,04 | 0,39 | 6,96E-01 | 8,19E-01 |
| PTPRF | 93,92 | 0,04 | 0,11 | 0,39 | 6,96E-01 | 8,20E-01 |
| RORC | 103,18 | 0,03 | 0,06 | 0,39 | 6,96E-01 | 8,20E-01 |
| DCTN2 | 597,46 | -0,01 | 0,03 | -0,39 | 6,97E-01 | 8,20E-01 |
| SUB1 | 2580,38 | 0,02 | 0,04 | 0,39 | 6,97E-01 | 8,20E-01 |
| TBC1D15 | 2139,02 | -0,01 | 0,03 | -0,39 | 6,97E-01 | 8,20E-01 |
| INPP5K | 1316,51 | 0,01 | 0,03 | 0,39 | 6,97E-01 | 8,20E-01 |
| SYMPK | 788,60 | 0,02 | 0,05 | 0,39 | 6,97E-01 | 8,20E-01 |
| WDTC1 | 2186,48 | -0,02 | 0,04 | -0,39 | 6,97E-01 | 8,20E-01 |
| BMS1 | 1342,96 | 0,01 | 0,02 | 0,39 | 6,98E-01 | 8,21E-01 |
| NUP210L | 97,49 | 0,04 | 0,11 | 0,39 | 6,98E-01 | 8,21E-01 |
| TAFA1 | 87,06 | 0,02 | 0,05 | 0,39 | 6,98E-01 | 8,21E-01 |
| CALM2 | 10922,07 | -0,01 | 0,04 | -0,39 | 6,98E-01 | 8,21E-01 |
| ADAP1 | 691,34 | 0,01 | 0,03 | 0,39 | 6,98E-01 | 8,21E-01 |
| FAM228B | 410,92 | 0,01 | 0,02 | 0,39 | 6,99E-01 | 8,21E-01 |
| GPR65 | 2459,65 | 0,02 | 0,04 | 0,39 | 6,99E-01 | 8,21E-01 |
| DOP1B | 910,69 | -0,02 | 0,04 | -0,39 | 6,99E-01 | 8,22E-01 |
| F8 | 117,87 | -0,02 | 0,04 | -0,39 | 6,99E-01 | 8,22E-01 |
| MEST | 137,21 | 0,02 | 0,05 | 0,39 | 6,99E-01 | 8,22E-01 |
| SMPD1 | 159,12 | 0,02 | 0,04 | 0,39 | 6,99E-01 | 8,22E-01 |
| UBA7 | 1360,13 | -0,01 | 0,04 | -0,39 | 6,99E-01 | 8,22E-01 |
| PABPC1 | 8780,77 | 0,01 | 0,03 | 0,39 | 7,00E-01 | 8,22E-01 |
| SEPTIN6 | 2476,69 | 0,01 | 0,03 | 0,39 | 7,00E-01 | 8,22E-01 |
| RFC5 | 297,27 | 0,01 | 0,01 | 0,39 | 7,00E-01 | 8,22E-01 |
| SYNPO2 | 77,53 | 0,03 | 0,08 | 0,39 | 7,00E-01 | 8,22E-01 |
| IFT43 | 166,73 | 0,01 | 0,02 | 0,39 | 7,00E-01 | 8,22E-01 |
| DAP | 635,85 | -0,02 | 0,04 | -0,38 | 7,00E-01 | 8,22E-01 |
| ATP8A1 | 5380,41 | 0,01 | 0,03 | 0,38 | 7,01E-01 | 8,22E-01 |
| TRAPPC4 | 956,12 | -0,01 | 0,02 | -0,38 | 7,01E-01 | 8,22E-01 |
| TMEM104 | 321,07 | 0,02 | 0,04 | 0,38 | 7,01E-01 | 8,22E-01 |
| SFMBT1 | 642,08 | 0,01 | 0,03 | 0,38 | 7,01E-01 | 8,22E-01 |
| MAFF | 185,42 | -0,01 | 0,04 | -0,38 | 7,01E-01 | 8,22E-01 |
| APH1A | 2182,95 | 0,01 | 0,02 | 0,38 | 7,01E-01 | 8,22E-01 |
| NDUFAF7 | 770,84 | 0,01 | 0,02 | 0,38 | 7,01E-01 | 8,22E-01 |
| CCT7 | 2137,07 | -0,01 | 0,02 | -0,38 | 7,01E-01 | 8,23E-01 |
| TARS3 | 875,70 | 0,01 | 0,02 | 0,38 | 7,02E-01 | 8,23E-01 |
| PTP4A2 | 6737,14 | 0,01 | 0,02 | 0,38 | 7,02E-01 | 8,23E-01 |
| CHERP | 221,55 | 0,02 | 0,05 | 0,38 | 7,03E-01 | 8,24E-01 |
| ZNF324 | 582,38 | -0,01 | 0,02 | -0,38 | 7,03E-01 | 8,24E-01 |
| BEND2 | 256,95 | -0,02 | 0,06 | -0,38 | 7,03E-01 | 8,24E-01 |
| HIBCH | 983,18 | -0,01 | 0,02 | -0,38 | 7,03E-01 | 8,25E-01 |
| ZMYM3 | 239,94 | 0,02 | 0,04 | 0,38 | 7,04E-01 | 8,25E-01 |
| COPE | 1053,73 | -0,01 | 0,02 | -0,38 | 7,04E-01 | 8,25E-01 |

| | | | | | | |
|---|---|---|---|---|---|---|
| DDB2 | 345,62 | 0,01 | 0,04 | 0,38 | 7,04E-01 | 8,25E-01 |
| RCE1 | 187,85 | 0,01 | 0,02 | 0,38 | 7,04E-01 | 8,25E-01 |
| MTUS1 | 138,71 | 0,03 | 0,08 | 0,38 | 7,04E-01 | 8,25E-01 |
| PEX14 | 206,25 | 0,01 | 0,03 | 0,38 | 7,04E-01 | 8,25E-01 |
| PRLR | 133,52 | 0,02 | 0,06 | 0,38 | 7,05E-01 | 8,26E-01 |
| ZNF483 | 335,17 | 0,01 | 0,03 | 0,38 | 7,05E-01 | 8,26E-01 |
| DDX5 | 27214,12 | -0,01 | 0,02 | -0,38 | 7,05E-01 | 8,26E-01 |
| PNPLA8 | 1852,85 | 0,02 | 0,05 | 0,38 | 7,06E-01 | 8,26E-01 |
| GRK3 | 2964,79 | 0,01 | 0,03 | 0,38 | 7,06E-01 | 8,27E-01 |
| ERVK3-1 | 851,19 | 0,01 | 0,02 | 0,38 | 7,06E-01 | 8,27E-01 |
| MAPK1IP1L | 1644,79 | -0,01 | 0,02 | -0,38 | 7,06E-01 | 8,27E-01 |
| ZNF707 | 370,23 | 0,01 | 0,03 | 0,38 | 7,06E-01 | 8,27E-01 |
| CCDC22 | 395,52 | 0,01 | 0,02 | 0,38 | 7,06E-01 | 8,27E-01 |
| PFKM | 296,71 | 0,01 | 0,03 | 0,38 | 7,06E-01 | 8,27E-01 |
| MAP4 | 1167,29 | 0,01 | 0,03 | 0,38 | 7,07E-01 | 8,27E-01 |
| TASOR2 | 3625,78 | 0,01 | 0,02 | 0,38 | 7,07E-01 | 8,27E-01 |
| ARHGAP10 | 494,13 | 0,01 | 0,03 | 0,38 | 7,07E-01 | 8,27E-01 |
| SORD | 322,80 | 0,01 | 0,04 | 0,38 | 7,07E-01 | 8,27E-01 |
| CDIPT | 1551,88 | 0,01 | 0,03 | 0,38 | 7,07E-01 | 8,27E-01 |
| PLCB3 | 159,25 | 0,02 | 0,05 | 0,38 | 7,07E-01 | 8,27E-01 |
| NEK11 | 160,38 | -0,01 | 0,04 | -0,38 | 7,07E-01 | 8,27E-01 |
| NSMCE2 | 643,43 | 0,01 | 0,02 | 0,38 | 7,07E-01 | 8,27E-01 |
| CPED1 | 357,51 | 0,02 | 0,04 | 0,38 | 7,07E-01 | 8,27E-01 |
| BCL11A | 2390,19 | 0,02 | 0,04 | 0,38 | 7,07E-01 | 8,27E-01 |
| GOLGA4 | 3861,32 | -0,01 | 0,03 | -0,38 | 7,08E-01 | 8,27E-01 |
| ITGA9 | 101,61 | 0,03 | 0,09 | 0,37 | 7,08E-01 | 8,27E-01 |
| SGMS2 | 821,71 | 0,01 | 0,04 | 0,37 | 7,08E-01 | 8,27E-01 |
| CACNA1I | 313,22 | 0,03 | 0,07 | 0,37 | 7,08E-01 | 8,28E-01 |
| TRIP11 | 2430,33 | 0,01 | 0,03 | 0,37 | 7,09E-01 | 8,28E-01 |
| MFSD5 | 447,61 | -0,01 | 0,02 | -0,37 | 7,09E-01 | 8,28E-01 |
| CDKN2D | 2429,54 | 0,01 | 0,04 | 0,37 | 7,09E-01 | 8,28E-01 |
| VAMP2 | 378,14 | 0,02 | 0,05 | 0,37 | 7,10E-01 | 8,29E-01 |
| PHYKPL | 3186,37 | -0,01 | 0,03 | -0,37 | 7,10E-01 | 8,29E-01 |
| NUMBL | 75,63 | 0,02 | 0,07 | 0,37 | 7,10E-01 | 8,29E-01 |
| CAPN3 | 152,52 | -0,02 | 0,05 | -0,37 | 7,10E-01 | 8,29E-01 |
| LGALS12 | 178,34 | 0,02 | 0,06 | 0,37 | 7,10E-01 | 8,29E-01 |
| GKAP1 | 214,73 | -0,01 | 0,03 | -0,37 | 7,10E-01 | 8,29E-01 |
| GLRX3 | 911,20 | 0,01 | 0,02 | 0,37 | 7,10E-01 | 8,29E-01 |
| SCAF1 | 315,70 | 0,02 | 0,06 | 0,37 | 7,11E-01 | 8,29E-01 |
| IL18 | 178,86 | -0,01 | 0,04 | -0,37 | 7,10E-01 | 8,29E-01 |
| BTRC | 505,81 | -0,01 | 0,02 | -0,37 | 7,11E-01 | 8,29E-01 |
| NDST2 | 91,95 | -0,02 | 0,04 | -0,37 | 7,11E-01 | 8,29E-01 |
| TSR2 | 460,92 | 0,01 | 0,02 | 0,37 | 7,11E-01 | 8,30E-01 |
| FANCG | 166,15 | 0,01 | 0,03 | 0,37 | 7,11E-01 | 8,30E-01 |
| ARID1B | 6765,89 | -0,01 | 0,02 | -0,37 | 7,12E-01 | 8,30E-01 |
| CNTNAP1 | 83,97 | 0,03 | 0,07 | 0,37 | 7,12E-01 | 8,30E-01 |

| | | | | | | |
|---|---|---|---|---|---|---|
| ADAMTS10 | 296,72 | 0,02 | 0,05 | 0,37 | 7,12E-01 | 8,30E-01 |
| PNKP | 977,61 | -0,01 | 0,04 | -0,37 | 7,12E-01 | 8,30E-01 |
| NUP133 | 1475,84 | -0,01 | 0,01 | -0,37 | 7,12E-01 | 8,30E-01 |
| SUGP1 | 321,52 | -0,01 | 0,04 | -0,37 | 7,12E-01 | 8,31E-01 |
| RNF121 | 467,64 | -0,01 | 0,02 | -0,37 | 7,13E-01 | 8,31E-01 |
| DIS3L | 840,36 | 0,01 | 0,02 | 0,37 | 7,13E-01 | 8,31E-01 |
| ABCC1 | 1206,59 | 0,02 | 0,04 | 0,37 | 7,13E-01 | 8,31E-01 |
| GPBAR1 | 189,77 | 0,01 | 0,04 | 0,37 | 7,13E-01 | 8,31E-01 |
| ABCA2 | 790,31 | -0,02 | 0,06 | -0,37 | 7,13E-01 | 8,31E-01 |
| CWF19L2 | 1657,57 | -0,01 | 0,04 | -0,37 | 7,14E-01 | 8,31E-01 |
| PKD1L1 | 87,59 | -0,03 | 0,09 | -0,37 | 7,14E-01 | 8,32E-01 |
| AP4E1 | 1303,21 | 0,01 | 0,02 | 0,37 | 7,14E-01 | 8,32E-01 |
| FAM229A | 190,12 | -0,02 | 0,05 | -0,37 | 7,15E-01 | 8,32E-01 |
| PPP5C | 602,64 | 0,01 | 0,03 | 0,37 | 7,15E-01 | 8,32E-01 |
| FAM149B1 | 525,01 | 0,00 | 0,01 | 0,37 | 7,15E-01 | 8,32E-01 |
| DMAC2 | 105,06 | 0,01 | 0,03 | 0,37 | 7,15E-01 | 8,32E-01 |
| MADD | 2012,25 | -0,01 | 0,03 | -0,36 | 7,15E-01 | 8,32E-01 |
| INTS1 | 941,91 | 0,02 | 0,06 | 0,37 | 7,15E-01 | 8,32E-01 |
| SMCO4 | 271,15 | -0,01 | 0,04 | -0,37 | 7,15E-01 | 8,32E-01 |
| RFTN1 | 1245,76 | 0,01 | 0,02 | 0,37 | 7,15E-01 | 8,32E-01 |
| SS18L1 | 486,50 | 0,01 | 0,02 | 0,37 | 7,15E-01 | 8,32E-01 |
| IFNLR1 | 154,06 | 0,02 | 0,05 | 0,36 | 7,15E-01 | 8,33E-01 |
| ZNF266 | 2030,32 | -0,01 | 0,02 | -0,36 | 7,16E-01 | 8,33E-01 |
| FBXO31 | 350,61 | 0,01 | 0,04 | 0,36 | 7,16E-01 | 8,33E-01 |
| TYW1 | 614,01 | 0,01 | 0,02 | 0,36 | 7,16E-01 | 8,33E-01 |
| ALDH1B1 | 125,09 | 0,01 | 0,03 | 0,36 | 7,16E-01 | 8,33E-01 |
| ANKRD33B | 333,75 | -0,02 | 0,04 | -0,36 | 7,16E-01 | 8,33E-01 |
| QNG1 | 356,34 | 0,01 | 0,02 | 0,36 | 7,16E-01 | 8,33E-01 |
| C1orf131 | 517,77 | 0,01 | 0,02 | 0,36 | 7,16E-01 | 8,33E-01 |
| CSTF2T | 1397,07 | 0,01 | 0,03 | 0,36 | 7,16E-01 | 8,33E-01 |
| NUP93 | 990,62 | 0,01 | 0,02 | 0,36 | 7,16E-01 | 8,33E-01 |
| CSTPP1 | 137,79 | 0,02 | 0,04 | 0,36 | 7,17E-01 | 8,33E-01 |
| CNR1 | 79,66 | 0,03 | 0,07 | 0,36 | 7,17E-01 | 8,34E-01 |
| AP3S1 | 791,98 | 0,01 | 0,03 | 0,36 | 7,17E-01 | 8,34E-01 |
| ZBTB20 | 3817,93 | 0,01 | 0,02 | 0,36 | 7,18E-01 | 8,34E-01 |
| TOB1 | 618,01 | 0,01 | 0,02 | 0,36 | 7,18E-01 | 8,34E-01 |
| ELAVL1 | 686,43 | 0,01 | 0,02 | 0,36 | 7,18E-01 | 8,34E-01 |
| H6PD | 1037,79 | 0,02 | 0,05 | 0,36 | 7,18E-01 | 8,34E-01 |
| TMEM11 | 239,40 | -0,01 | 0,03 | -0,36 | 7,18E-01 | 8,34E-01 |
| C4orf50 | 172,07 | 0,03 | 0,09 | 0,36 | 7,19E-01 | 8,35E-01 |
| KRT72 | 116,97 | -0,04 | 0,10 | -0,36 | 7,19E-01 | 8,35E-01 |
| MED9 | 253,78 | 0,01 | 0,02 | 0,36 | 7,19E-01 | 8,35E-01 |
| NOMO2 | 436,03 | -0,01 | 0,03 | -0,36 | 7,19E-01 | 8,35E-01 |
| CMC2 | 614,14 | 0,01 | 0,02 | 0,36 | 7,19E-01 | 8,35E-01 |
| MZB1 | 135,14 | -0,02 | 0,07 | -0,36 | 7,20E-01 | 8,35E-01 |
| XXYLT1 | 241,99 | 0,01 | 0,03 | 0,36 | 7,20E-01 | 8,36E-01 |

| | | | | | | |
|---|---|---|---|---|---|---|
| CC2D1A | 135,11 | 0,02 | 0,07 | 0,36 | 7,20E-01 | 8,36E-01 |
| FOXK2 | 779,73 | -0,01 | 0,03 | -0,36 | 7,20E-01 | 8,36E-01 |
| BMP1 | 88,95 | 0,02 | 0,07 | 0,36 | 7,21E-01 | 8,36E-01 |
| MEAK7 | 424,25 | -0,01 | 0,03 | -0,36 | 7,21E-01 | 8,36E-01 |
| LBH | 5727,01 | 0,01 | 0,03 | 0,36 | 7,21E-01 | 8,36E-01 |
| CYB5B | 1114,32 | 0,01 | 0,02 | 0,36 | 7,21E-01 | 8,36E-01 |
| PGGHG | 2160,17 | -0,02 | 0,06 | -0,36 | 7,21E-01 | 8,36E-01 |
| UTP6 | 1845,04 | -0,01 | 0,02 | -0,36 | 7,21E-01 | 8,36E-01 |
| CASP7 | 620,46 | 0,01 | 0,03 | 0,36 | 7,21E-01 | 8,36E-01 |
| INAFM2 | 1248,59 | 0,01 | 0,03 | 0,36 | 7,21E-01 | 8,36E-01 |
| UCK1 | 403,65 | -0,01 | 0,02 | -0,36 | 7,21E-01 | 8,36E-01 |
| ENSG000002 | 118,10 | 0,01 | 0,03 | 0,36 | 7,21E-01 | 8,36E-01 |
| SLC4A5 | 88,96 | -0,01 | 0,04 | -0,36 | 7,22E-01 | 8,36E-01 |
| ZCCHC14 | 556,62 | 0,01 | 0,02 | 0,36 | 7,22E-01 | 8,36E-01 |
| PSIP1 | 3358,43 | -0,01 | 0,03 | -0,36 | 7,22E-01 | 8,36E-01 |
| SORBS1 | 80,76 | 0,03 | 0,08 | 0,36 | 7,22E-01 | 8,37E-01 |
| TTC3 | 5269,46 | 0,01 | 0,02 | 0,36 | 7,22E-01 | 8,37E-01 |
| SUPT5H | 1400,92 | -0,01 | 0,03 | -0,36 | 7,23E-01 | 8,37E-01 |
| PITPNC1 | 1759,22 | -0,01 | 0,02 | -0,36 | 7,22E-01 | 8,37E-01 |
| PRORP | 477,17 | -0,01 | 0,03 | -0,36 | 7,22E-01 | 8,37E-01 |
| ATG10 | 284,80 | 0,01 | 0,03 | 0,36 | 7,22E-01 | 8,37E-01 |
| ZSCAN26 | 436,81 | 0,01 | 0,02 | 0,36 | 7,22E-01 | 8,37E-01 |
| MSANTD1 | 80,54 | -0,03 | 0,08 | -0,36 | 7,23E-01 | 8,37E-01 |
| ENOX2 | 454,40 | 0,01 | 0,02 | 0,35 | 7,23E-01 | 8,37E-01 |
| ACCS | 565,61 | -0,02 | 0,07 | -0,35 | 7,23E-01 | 8,37E-01 |
| USP31 | 419,24 | -0,01 | 0,03 | -0,35 | 7,24E-01 | 8,38E-01 |
| TOMM34 | 224,16 | 0,01 | 0,03 | 0,35 | 7,24E-01 | 8,38E-01 |
| TRAF7 | 428,93 | 0,01 | 0,04 | 0,35 | 7,24E-01 | 8,38E-01 |
| NDUFC2 | 397,42 | 0,01 | 0,03 | 0,35 | 7,24E-01 | 8,38E-01 |
| TTC9 | 495,01 | -0,01 | 0,04 | -0,35 | 7,24E-01 | 8,38E-01 |
| THAP3 | 278,34 | 0,01 | 0,02 | 0,35 | 7,25E-01 | 8,38E-01 |
| TFPI | 115,24 | 0,02 | 0,06 | 0,35 | 7,25E-01 | 8,38E-01 |
| SRGAP2C | 572,29 | 0,01 | 0,03 | 0,35 | 7,25E-01 | 8,38E-01 |
| MGA | 4462,13 | 0,01 | 0,02 | 0,35 | 7,25E-01 | 8,38E-01 |
| POLN | 75,61 | 0,02 | 0,06 | 0,35 | 7,25E-01 | 8,38E-01 |
| COL6A2 | 359,74 | 0,02 | 0,06 | 0,35 | 7,25E-01 | 8,38E-01 |
| SETBP1 | 598,25 | -0,02 | 0,04 | -0,35 | 7,25E-01 | 8,38E-01 |
| ZNF830 | 741,46 | 0,01 | 0,04 | 0,35 | 7,25E-01 | 8,38E-01 |
| ATG4A | 332,54 | -0,01 | 0,02 | -0,35 | 7,25E-01 | 8,38E-01 |
| DDX21 | 2821,04 | 0,01 | 0,03 | 0,35 | 7,25E-01 | 8,38E-01 |
| WDR26 | 10264,82 | 0,01 | 0,03 | 0,35 | 7,24E-01 | 8,38E-01 |
| OLFM2 | 94,80 | 0,02 | 0,04 | 0,35 | 7,25E-01 | 8,38E-01 |
| IRF4 | 662,20 | 0,01 | 0,03 | 0,35 | 7,25E-01 | 8,38E-01 |
| U2AF1 | 390,06 | -0,04 | 0,11 | -0,35 | 7,25E-01 | 8,38E-01 |
| ATP1B3 | 497,48 | -0,01 | 0,02 | -0,35 | 7,25E-01 | 8,38E-01 |
| QRICH2 | 141,28 | 0,01 | 0,04 | 0,35 | 7,25E-01 | 8,38E-01 |

| | | | | | | |
|---|---|---|---|---|---|---|
| ELMO3 | 95,87 | 0,02 | 0,05 | 0,35 | 7,26E-01 | 8,38E-01 |
| STMN1 | 349,48 | 0,01 | 0,04 | 0,35 | 7,26E-01 | 8,38E-01 |
| BRF2 | 411,63 | -0,01 | 0,02 | -0,35 | 7,26E-01 | 8,39E-01 |
| MFAP3 | 1293,16 | -0,01 | 0,02 | -0,35 | 7,27E-01 | 8,39E-01 |
| GDI2 | 4817,28 | 0,01 | 0,03 | 0,35 | 7,27E-01 | 8,39E-01 |
| TTF2 | 657,47 | 0,01 | 0,03 | 0,35 | 7,27E-01 | 8,39E-01 |
| TMEM216 | 220,52 | 0,01 | 0,02 | 0,35 | 7,27E-01 | 8,39E-01 |
| DAD1 | 1330,10 | 0,01 | 0,03 | 0,35 | 7,27E-01 | 8,39E-01 |
| IARS2 | 1403,60 | -0,01 | 0,02 | -0,35 | 7,27E-01 | 8,39E-01 |
| CCDC126 | 564,37 | -0,01 | 0,04 | -0,35 | 7,28E-01 | 8,40E-01 |
| PCTP | 1310,59 | 0,01 | 0,04 | 0,35 | 7,28E-01 | 8,40E-01 |
| PMM2 | 748,44 | 0,01 | 0,02 | 0,35 | 7,28E-01 | 8,40E-01 |
| BTN2A2 | 508,79 | 0,01 | 0,03 | 0,35 | 7,28E-01 | 8,40E-01 |
| ZNF780A | 2054,36 | 0,01 | 0,04 | 0,35 | 7,28E-01 | 8,40E-01 |
| BTK | 2095,39 | -0,01 | 0,02 | -0,35 | 7,28E-01 | 8,40E-01 |
| PSMC6 | 1434,91 | 0,01 | 0,04 | 0,35 | 7,29E-01 | 8,40E-01 |
| STAU1 | 1347,24 | 0,01 | 0,04 | 0,35 | 7,29E-01 | 8,40E-01 |
| PLK3 | 392,57 | -0,02 | 0,04 | -0,35 | 7,29E-01 | 8,40E-01 |
| EDAR | 168,03 | 0,02 | 0,06 | 0,35 | 7,29E-01 | 8,40E-01 |
| DRAP1 | 359,29 | 0,01 | 0,03 | 0,35 | 7,29E-01 | 8,40E-01 |
| BZW1 | 3752,94 | -0,01 | 0,03 | -0,35 | 7,29E-01 | 8,40E-01 |
| COL5A3 | 115,88 | -0,04 | 0,12 | -0,35 | 7,29E-01 | 8,41E-01 |
| CDR2 | 757,94 | 0,01 | 0,03 | 0,35 | 7,29E-01 | 8,41E-01 |
| PJA1 | 367,55 | 0,01 | 0,03 | 0,35 | 7,30E-01 | 8,41E-01 |
| NECAB3 | 122,88 | 0,01 | 0,04 | 0,35 | 7,30E-01 | 8,41E-01 |
| SLC16A5 | 332,73 | -0,02 | 0,05 | -0,35 | 7,30E-01 | 8,41E-01 |
| STN1 | 1195,93 | -0,01 | 0,02 | -0,35 | 7,30E-01 | 8,41E-01 |
| SLC45A3 | 158,53 | -0,03 | 0,08 | -0,35 | 7,30E-01 | 8,41E-01 |
| RPRD1A | 1931,21 | -0,01 | 0,02 | -0,34 | 7,30E-01 | 8,41E-01 |
| RARS2 | 1234,04 | 0,01 | 0,02 | 0,34 | 7,31E-01 | 8,41E-01 |
| ZNF443 | 149,07 | 0,02 | 0,05 | 0,34 | 7,31E-01 | 8,42E-01 |
| FCRLA | 330,27 | 0,02 | 0,07 | 0,34 | 7,31E-01 | 8,42E-01 |
| CHST10 | 92,01 | -0,02 | 0,06 | -0,34 | 7,32E-01 | 8,42E-01 |
| BLZF1 | 886,30 | -0,01 | 0,04 | -0,34 | 7,32E-01 | 8,42E-01 |
| MOK | 76,81 | -0,03 | 0,09 | -0,34 | 7,32E-01 | 8,42E-01 |
| PPM1B | 1908,57 | 0,01 | 0,04 | 0,34 | 7,32E-01 | 8,42E-01 |
| PTCH1 | 550,03 | 0,02 | 0,05 | 0,34 | 7,32E-01 | 8,42E-01 |
| PSMA5 | 1954,62 | 0,01 | 0,02 | 0,34 | 7,32E-01 | 8,42E-01 |
| BRICD5 | 141,25 | 0,01 | 0,04 | 0,34 | 7,32E-01 | 8,43E-01 |
| ASH2L | 976,71 | -0,01 | 0,02 | -0,34 | 7,32E-01 | 8,43E-01 |
| ZFR | 3257,56 | 0,01 | 0,02 | 0,34 | 7,33E-01 | 8,43E-01 |
| WASHC4 | 10579,36 | -0,01 | 0,04 | -0,34 | 7,33E-01 | 8,43E-01 |
| AAMDC | 91,81 | -0,01 | 0,03 | -0,34 | 7,33E-01 | 8,43E-01 |
| SPTAN1 | 3790,22 | -0,01 | 0,04 | -0,34 | 7,33E-01 | 8,43E-01 |
| CNTNAP3C | 469,31 | -0,03 | 0,08 | -0,34 | 7,33E-01 | 8,43E-01 |
| BTG1 | 5430,25 | -0,01 | 0,03 | -0,34 | 7,33E-01 | 8,43E-01 |

| | | | | | | |
|---|---|---|---|---|---|---|
| ZNF354A | 301,39 | -0,02 | 0,05 | -0,34 | 7,34E-01 | 8,44E-01 |
| EPC2 | 2365,85 | -0,01 | 0,02 | -0,34 | 7,34E-01 | 8,44E-01 |
| XPO5 | 647,57 | 0,01 | 0,03 | 0,34 | 7,34E-01 | 8,44E-01 |
| ATG9B | 73,84 | 0,02 | 0,07 | 0,34 | 7,34E-01 | 8,44E-01 |
| FIZ1 | 108,54 | 0,02 | 0,05 | 0,34 | 7,34E-01 | 8,44E-01 |
| TCP1 | 2568,94 | -0,01 | 0,02 | -0,34 | 7,34E-01 | 8,44E-01 |
| DCAF1 | 1230,04 | 0,01 | 0,02 | 0,34 | 7,34E-01 | 8,44E-01 |
| CAPG | 1002,80 | 0,01 | 0,03 | 0,34 | 7,35E-01 | 8,44E-01 |
| CAPZA2 | 4773,17 | -0,02 | 0,05 | -0,34 | 7,35E-01 | 8,44E-01 |
| UQCRC1 | 1398,00 | 0,01 | 0,02 | 0,34 | 7,35E-01 | 8,44E-01 |
| NAA40 | 196,29 | 0,02 | 0,06 | 0,34 | 7,35E-01 | 8,44E-01 |
| FAM118A | 1365,39 | -0,02 | 0,06 | -0,34 | 7,35E-01 | 8,44E-01 |
| DDX24 | 3004,34 | 0,01 | 0,02 | 0,34 | 7,36E-01 | 8,45E-01 |
| ZNF436 | 461,41 | -0,01 | 0,02 | -0,34 | 7,36E-01 | 8,45E-01 |
| CSF1 | 179,88 | -0,02 | 0,06 | -0,34 | 7,36E-01 | 8,45E-01 |
| GPR132 | 1231,57 | -0,01 | 0,02 | -0,34 | 7,36E-01 | 8,45E-01 |
| TMEM181 | 2033,91 | 0,01 | 0,02 | 0,34 | 7,37E-01 | 8,46E-01 |
| CCDC141 | 822,39 | 0,02 | 0,05 | 0,34 | 7,38E-01 | 8,46E-01 |
| RBMS2 | 231,89 | -0,02 | 0,06 | -0,33 | 7,38E-01 | 8,46E-01 |
| EIF2S2 | 1551,10 | -0,01 | 0,03 | -0,33 | 7,38E-01 | 8,46E-01 |
| PABIR3 | 321,14 | -0,01 | 0,03 | -0,33 | 7,38E-01 | 8,46E-01 |
| RBBP6 | 2771,33 | 0,01 | 0,02 | 0,33 | 7,38E-01 | 8,46E-01 |
| HDHD3 | 155,27 | 0,01 | 0,03 | 0,33 | 7,38E-01 | 8,46E-01 |
| NKIRAS1 | 107,18 | -0,01 | 0,03 | -0,33 | 7,38E-01 | 8,46E-01 |
| CCDC32 | 702,33 | 0,01 | 0,02 | 0,33 | 7,38E-01 | 8,47E-01 |
| UNK | 539,28 | 0,01 | 0,03 | 0,33 | 7,38E-01 | 8,47E-01 |
| HSPA9 | 2748,43 | 0,01 | 0,02 | 0,33 | 7,39E-01 | 8,47E-01 |
| RSKR | 221,33 | -0,02 | 0,05 | -0,33 | 7,39E-01 | 8,47E-01 |
| OSBPL7 | 270,78 | 0,01 | 0,04 | 0,33 | 7,39E-01 | 8,47E-01 |
| NAA10 | 259,55 | -0,01 | 0,03 | -0,33 | 7,39E-01 | 8,47E-01 |
| UNC50 | 1056,35 | -0,01 | 0,02 | -0,33 | 7,39E-01 | 8,47E-01 |
| PPP1R13B | 225,70 | 0,01 | 0,04 | 0,33 | 7,40E-01 | 8,47E-01 |
| ZSCAN29 | 983,41 | 0,01 | 0,02 | 0,33 | 7,40E-01 | 8,48E-01 |
| LRRC37A2 | 510,96 | -0,01 | 0,04 | -0,33 | 7,40E-01 | 8,48E-01 |
| MXRA7 | 256,23 | 0,03 | 0,08 | 0,33 | 7,40E-01 | 8,48E-01 |
| CFAP20 | 384,77 | -0,01 | 0,02 | -0,33 | 7,40E-01 | 8,48E-01 |
| SEM1 | 829,88 | 0,01 | 0,03 | 0,33 | 7,41E-01 | 8,48E-01 |
| AFAP1L2 | 75,90 | 0,02 | 0,06 | 0,33 | 7,41E-01 | 8,48E-01 |
| TCF7L2 | 547,24 | -0,01 | 0,04 | -0,33 | 7,41E-01 | 8,48E-01 |
| AK9 | 511,99 | 0,01 | 0,03 | 0,33 | 7,41E-01 | 8,48E-01 |
| CIRBP | 8832,90 | 0,01 | 0,02 | 0,33 | 7,41E-01 | 8,48E-01 |
| RABL2A | 227,19 | 0,01 | 0,04 | 0,33 | 7,41E-01 | 8,48E-01 |
| EGF | 188,00 | -0,02 | 0,07 | -0,33 | 7,41E-01 | 8,48E-01 |
| ZNF471 | 74,84 | 0,02 | 0,07 | 0,33 | 7,42E-01 | 8,49E-01 |
| ZNF117 | 3318,69 | -0,02 | 0,06 | -0,33 | 7,42E-01 | 8,49E-01 |
| TTBK2 | 1794,70 | 0,00 | 0,01 | 0,33 | 7,42E-01 | 8,49E-01 |

| | | | | | | |
|---|---|---|---|---|---|---|
| CD1D | 1321,32 | 0,01 | 0,03 | 0,33 | 7,42E-01 | 8,49E-01 |
| DMPK | 108,67 | 0,02 | 0,07 | 0,33 | 7,42E-01 | 8,49E-01 |
| NBPF1 | 561,68 | 0,01 | 0,04 | 0,33 | 7,42E-01 | 8,49E-01 |
| ZNF444 | 224,09 | 0,02 | 0,05 | 0,33 | 7,42E-01 | 8,49E-01 |
| TMEM8B | 150,45 | 0,02 | 0,06 | 0,33 | 7,43E-01 | 8,49E-01 |
| CHIT1 | 166,71 | -0,01 | 0,04 | -0,33 | 7,43E-01 | 8,49E-01 |
| CEP162 | 627,39 | -0,01 | 0,04 | -0,33 | 7,43E-01 | 8,49E-01 |
| CLDN15 | 156,22 | 0,02 | 0,05 | 0,33 | 7,43E-01 | 8,49E-01 |
| ERGIC2 | 1756,73 | -0,01 | 0,04 | -0,33 | 7,43E-01 | 8,49E-01 |
| APOBEC3F | 151,24 | 0,01 | 0,05 | 0,33 | 7,43E-01 | 8,49E-01 |
| CHMP2B | 884,80 | -0,01 | 0,03 | -0,33 | 7,44E-01 | 8,50E-01 |
| AP2A2 | 740,58 | -0,01 | 0,04 | -0,33 | 7,44E-01 | 8,50E-01 |
| MED4 | 1043,27 | -0,01 | 0,02 | -0,33 | 7,44E-01 | 8,50E-01 |
| LASP1NB | 176,43 | -0,01 | 0,03 | -0,33 | 7,44E-01 | 8,50E-01 |
| NPRL3 | 2288,18 | 0,04 | 0,11 | 0,33 | 7,45E-01 | 8,51E-01 |
| GTF2A1 | 2535,33 | 0,01 | 0,03 | 0,32 | 7,45E-01 | 8,51E-01 |
| SELENOF | 3885,08 | 0,01 | 0,04 | 0,32 | 7,46E-01 | 8,52E-01 |
| ZMIZ2 | 693,46 | 0,02 | 0,05 | 0,32 | 7,46E-01 | 8,52E-01 |
| TMEM186 | 185,85 | 0,01 | 0,02 | 0,32 | 7,46E-01 | 8,52E-01 |
| PPP4R2 | 3214,37 | 0,01 | 0,04 | 0,32 | 7,46E-01 | 8,52E-01 |
| NDUFA10 | 2250,21 | 0,01 | 0,02 | 0,32 | 7,46E-01 | 8,52E-01 |
| BEND4 | 148,71 | 0,02 | 0,07 | 0,32 | 7,47E-01 | 8,52E-01 |
| SH3D21 | 110,03 | -0,02 | 0,05 | -0,32 | 7,47E-01 | 8,53E-01 |
| CLN6 | 326,12 | 0,01 | 0,04 | 0,32 | 7,47E-01 | 8,53E-01 |
| PRDX3 | 1596,15 | -0,01 | 0,03 | -0,32 | 7,47E-01 | 8,53E-01 |
| UBE2I | 1668,16 | 0,01 | 0,02 | 0,32 | 7,47E-01 | 8,53E-01 |
| NT5DC3 | 289,43 | -0,01 | 0,05 | -0,32 | 7,47E-01 | 8,53E-01 |
| ELOVL6 | 209,31 | -0,01 | 0,04 | -0,32 | 7,48E-01 | 8,53E-01 |
| ARF5 | 510,00 | 0,01 | 0,02 | 0,32 | 7,48E-01 | 8,53E-01 |
| NUP160 | 1616,83 | 0,01 | 0,02 | 0,32 | 7,49E-01 | 8,54E-01 |
| TRIM9 | 75,20 | -0,02 | 0,08 | -0,32 | 7,49E-01 | 8,54E-01 |
| RNF115 | 1751,81 | 0,00 | 0,01 | -0,32 | 7,50E-01 | 8,55E-01 |
| SLF2 | 2025,47 | 0,01 | 0,02 | 0,32 | 7,50E-01 | 8,55E-01 |
| C1QTNF3 | 120,14 | 0,01 | 0,03 | 0,32 | 7,50E-01 | 8,55E-01 |
| GNAS | 5688,57 | 0,01 | 0,04 | 0,32 | 7,50E-01 | 8,55E-01 |
| PARL | 184,55 | 0,01 | 0,02 | 0,32 | 7,51E-01 | 8,56E-01 |
| MS4A7 | 2655,73 | 0,01 | 0,04 | 0,32 | 7,51E-01 | 8,56E-01 |
| ADAMTS1 | 229,14 | -0,02 | 0,06 | -0,32 | 7,51E-01 | 8,56E-01 |
| PWWP3A | 377,66 | 0,01 | 0,03 | 0,32 | 7,51E-01 | 8,56E-01 |
| CEP72 | 220,46 | 0,01 | 0,04 | 0,32 | 7,52E-01 | 8,57E-01 |
| ZNF737 | 844,01 | 0,01 | 0,04 | 0,32 | 7,52E-01 | 8,57E-01 |
| PRICKLE1 | 189,94 | 0,01 | 0,05 | 0,32 | 7,52E-01 | 8,57E-01 |
| PPM1K | 3509,34 | 0,01 | 0,03 | 0,32 | 7,52E-01 | 8,57E-01 |
| CCDC117 | 1134,62 | -0,01 | 0,02 | -0,32 | 7,52E-01 | 8,57E-01 |
| ZFAND2A | 343,57 | -0,01 | 0,02 | -0,32 | 7,52E-01 | 8,57E-01 |
| ARHGAP35 | 1533,85 | -0,01 | 0,02 | -0,31 | 7,53E-01 | 8,57E-01 |

| | | | | | | |
|---|---|---|---|---|---|---|
| GPR89B | 558,08 | 0,01 | 0,02 | 0,31 | 7,53E-01 | 8,57E-01 |
| FRS3 | 73,78 | 0,01 | 0,04 | 0,31 | 7,53E-01 | 8,58E-01 |
| TEX261 | 324,10 | 0,01 | 0,04 | 0,31 | 7,53E-01 | 8,58E-01 |
| UNC13B | 109,06 | -0,03 | 0,09 | -0,31 | 7,54E-01 | 8,58E-01 |
| C6orf62 | 11864,82 | 0,01 | 0,02 | 0,31 | 7,54E-01 | 8,58E-01 |
| KIF5C | 379,35 | 0,01 | 0,04 | 0,31 | 7,54E-01 | 8,58E-01 |
| ZFAND6 | 3339,09 | 0,01 | 0,03 | 0,31 | 7,54E-01 | 8,58E-01 |
| VDAC2 | 930,58 | 0,00 | 0,01 | -0,31 | 7,54E-01 | 8,58E-01 |
| COLGALT2 | 151,22 | -0,02 | 0,08 | -0,31 | 7,54E-01 | 8,58E-01 |
| HRH4 | 451,15 | 0,02 | 0,07 | 0,31 | 7,54E-01 | 8,58E-01 |
| VPS52 | 914,12 | 0,01 | 0,02 | 0,31 | 7,54E-01 | 8,58E-01 |
| RNPS1 | 1030,79 | 0,01 | 0,03 | 0,31 | 7,54E-01 | 8,58E-01 |
| TAGLN | 436,58 | 0,02 | 0,06 | 0,31 | 7,54E-01 | 8,58E-01 |
| SCYL2 | 2791,33 | -0,01 | 0,02 | -0,31 | 7,55E-01 | 8,58E-01 |
| ADGRE1 | 1303,01 | -0,02 | 0,05 | -0,31 | 7,56E-01 | 8,60E-01 |
| KHSRP | 639,93 | 0,01 | 0,04 | 0,31 | 7,56E-01 | 8,60E-01 |
| RGS18 | 5003,47 | 0,02 | 0,06 | 0,31 | 7,56E-01 | 8,60E-01 |
| SGTA | 180,28 | 0,02 | 0,05 | 0,31 | 7,56E-01 | 8,60E-01 |
| CLTB | 1145,62 | 0,01 | 0,03 | 0,31 | 7,57E-01 | 8,60E-01 |
| VIM | 19328,73 | -0,01 | 0,03 | -0,31 | 7,56E-01 | 8,60E-01 |
| MCM3AP | 2284,04 | -0,01 | 0,03 | -0,31 | 7,58E-01 | 8,61E-01 |
| CDKN1C | 448,82 | 0,02 | 0,06 | 0,31 | 7,58E-01 | 8,61E-01 |
| ASAP2 | 168,80 | 0,02 | 0,06 | 0,31 | 7,58E-01 | 8,61E-01 |
| COLGALT1 | 1533,26 | -0,01 | 0,03 | -0,31 | 7,58E-01 | 8,61E-01 |
| TUFM | 1477,90 | 0,01 | 0,02 | 0,31 | 7,58E-01 | 8,61E-01 |
| NMNAT3 | 175,01 | 0,01 | 0,05 | 0,31 | 7,58E-01 | 8,61E-01 |
| LPCAT4 | 350,03 | 0,01 | 0,03 | 0,31 | 7,59E-01 | 8,62E-01 |
| HDAC2 | 938,92 | 0,01 | 0,03 | 0,31 | 7,59E-01 | 8,62E-01 |
| CETN2 | 241,62 | 0,01 | 0,04 | 0,31 | 7,59E-01 | 8,62E-01 |
| TUBB | 3170,44 | 0,01 | 0,02 | 0,31 | 7,59E-01 | 8,62E-01 |
| ATP5F1B | 6179,75 | 0,01 | 0,02 | 0,31 | 7,59E-01 | 8,62E-01 |
| APBA3 | 401,64 | 0,01 | 0,03 | 0,31 | 7,59E-01 | 8,62E-01 |
| NSRP1 | 842,62 | 0,00 | 0,01 | 0,31 | 7,60E-01 | 8,62E-01 |
| HOXB2 | 126,01 | 0,02 | 0,05 | 0,31 | 7,60E-01 | 8,62E-01 |
| PSMC5 | 520,61 | -0,01 | 0,02 | -0,31 | 7,60E-01 | 8,63E-01 |
| ATP13A4 | 111,29 | -0,02 | 0,06 | -0,30 | 7,60E-01 | 8,63E-01 |
| NCOA5 | 675,52 | -0,01 | 0,02 | -0,30 | 7,61E-01 | 8,63E-01 |
| NEK9 | 2770,85 | 0,00 | 0,01 | -0,30 | 7,61E-01 | 8,63E-01 |
| ITGA10 | 89,75 | -0,02 | 0,05 | -0,30 | 7,61E-01 | 8,63E-01 |
| CLSPN | 82,58 | -0,02 | 0,06 | -0,30 | 7,61E-01 | 8,63E-01 |
| CAPZA1 | 8372,26 | -0,01 | 0,04 | -0,30 | 7,61E-01 | 8,63E-01 |
| GDF7 | 152,15 | 0,03 | 0,10 | 0,30 | 7,61E-01 | 8,63E-01 |
| MRPS6 | 355,14 | -0,01 | 0,03 | -0,30 | 7,62E-01 | 8,63E-01 |
| SLC52A2 | 194,10 | 0,01 | 0,03 | 0,30 | 7,62E-01 | 8,63E-01 |
| ZNF20 | 77,15 | -0,01 | 0,03 | -0,30 | 7,62E-01 | 8,64E-01 |
| RASAL2 | 79,07 | 0,03 | 0,09 | 0,30 | 7,62E-01 | 8,64E-01 |

| | | | | | | |
|---|---|---|---|---|---|---|
| ATAD2B | 4644,28 | 0,01 | 0,02 | 0,30 | 7,62E-01 | 8,64E-01 |
| CHAF1A | 177,39 | 0,01 | 0,03 | 0,30 | 7,62E-01 | 8,64E-01 |
| PTGR2 | 90,14 | 0,01 | 0,05 | 0,30 | 7,63E-01 | 8,64E-01 |
| PPP1R8 | 1093,18 | 0,01 | 0,02 | 0,30 | 7,63E-01 | 8,64E-01 |
| ZNF763 | 175,63 | -0,01 | 0,03 | -0,30 | 7,62E-01 | 8,64E-01 |
| NFKB2 | 1259,66 | -0,01 | 0,04 | -0,30 | 7,63E-01 | 8,64E-01 |
| ENSG000002 | 242,16 | 0,04 | 0,14 | 0,30 | 7,63E-01 | 8,64E-01 |
| ZMYM2 | 5584,73 | -0,01 | 0,02 | -0,30 | 7,63E-01 | 8,64E-01 |
| ZCWPW1 | 196,09 | 0,01 | 0,02 | 0,30 | 7,63E-01 | 8,64E-01 |
| HDDC2 | 581,38 | 0,01 | 0,03 | 0,30 | 7,64E-01 | 8,64E-01 |
| PRPF40A | 4550,77 | 0,01 | 0,03 | 0,30 | 7,64E-01 | 8,65E-01 |
| PPIP5K2 | 4799,54 | 0,01 | 0,04 | 0,30 | 7,64E-01 | 8,65E-01 |
| NCDN | 102,12 | 0,02 | 0,06 | 0,30 | 7,64E-01 | 8,65E-01 |
| MALSU1 | 435,52 | 0,01 | 0,02 | 0,30 | 7,65E-01 | 8,65E-01 |
| NEU3 | 670,95 | -0,01 | 0,03 | -0,30 | 7,65E-01 | 8,65E-01 |
| YARS1 | 949,52 | 0,01 | 0,02 | 0,30 | 7,65E-01 | 8,65E-01 |
| ATF6B | 562,21 | -0,01 | 0,05 | -0,30 | 7,65E-01 | 8,65E-01 |
| MED10 | 546,82 | 0,01 | 0,02 | 0,30 | 7,65E-01 | 8,65E-01 |
| SLC38A11 | 162,62 | 0,02 | 0,08 | 0,30 | 7,65E-01 | 8,65E-01 |
| HLA-DMA | 933,40 | 0,01 | 0,03 | 0,30 | 7,65E-01 | 8,65E-01 |
| ZNF514 | 925,32 | 0,01 | 0,03 | 0,30 | 7,66E-01 | 8,66E-01 |
| H2AZ2 | 2731,43 | 0,00 | 0,01 | -0,30 | 7,66E-01 | 8,66E-01 |
| MARCO | 226,89 | -0,02 | 0,07 | -0,30 | 7,66E-01 | 8,66E-01 |
| TRIM3 | 136,01 | 0,01 | 0,05 | 0,30 | 7,66E-01 | 8,66E-01 |
| SMARCA5 | 3097,49 | 0,01 | 0,04 | 0,30 | 7,66E-01 | 8,66E-01 |
| PPP1R26 | 216,57 | 0,01 | 0,04 | 0,30 | 7,66E-01 | 8,66E-01 |
| CSK | 2469,19 | -0,01 | 0,05 | -0,30 | 7,66E-01 | 8,66E-01 |
| ST3GAL3 | 477,16 | -0,01 | 0,04 | -0,30 | 7,66E-01 | 8,66E-01 |
| CALR | 1578,43 | 0,01 | 0,03 | 0,30 | 7,66E-01 | 8,66E-01 |
| ABCA6 | 166,42 | -0,03 | 0,11 | -0,30 | 7,66E-01 | 8,66E-01 |
| LNPEP | 12317,87 | 0,01 | 0,02 | 0,30 | 7,67E-01 | 8,66E-01 |
| ZNF687 | 690,99 | -0,01 | 0,04 | -0,30 | 7,67E-01 | 8,67E-01 |
| CUL9 | 1041,28 | 0,01 | 0,04 | 0,30 | 7,68E-01 | 8,67E-01 |
| FRMD4A | 179,50 | 0,03 | 0,09 | 0,30 | 7,68E-01 | 8,67E-01 |
| CRELD2 | 416,47 | -0,01 | 0,02 | -0,30 | 7,68E-01 | 8,67E-01 |
| ZNF292 | 7234,30 | 0,01 | 0,04 | 0,30 | 7,68E-01 | 8,67E-01 |
| SRP14 | 1605,91 | 0,02 | 0,05 | 0,29 | 7,69E-01 | 8,68E-01 |
| NEXMIF | 85,72 | 0,02 | 0,06 | 0,29 | 7,69E-01 | 8,68E-01 |
| SLC15A2 | 662,19 | 0,01 | 0,04 | 0,29 | 7,69E-01 | 8,68E-01 |
| HLA-DRB1 | 4865,57 | -0,01 | 0,04 | -0,29 | 7,70E-01 | 8,69E-01 |
| ERI3 | 497,60 | 0,01 | 0,03 | 0,29 | 7,70E-01 | 8,69E-01 |
| PIK3C2A | 2469,40 | 0,01 | 0,02 | 0,29 | 7,70E-01 | 8,69E-01 |
| TENM4 | 102,12 | 0,04 | 0,15 | 0,29 | 7,71E-01 | 8,69E-01 |
| NPLOC4 | 2400,33 | -0,01 | 0,03 | -0,29 | 7,70E-01 | 8,69E-01 |
| AQR | 3419,36 | -0,01 | 0,02 | -0,29 | 7,71E-01 | 8,70E-01 |
| AFAP1 | 357,17 | -0,02 | 0,06 | -0,29 | 7,71E-01 | 8,70E-01 |

| | | | | | | |
|---|---|---|---|---|---|---|
| USP5 | 814,74 | 0,01 | 0,02 | 0,29 | 7,71E-01 | 8,70E-01 |
| KLRF1 | 1035,79 | -0,02 | 0,06 | -0,29 | 7,71E-01 | 8,70E-01 |
| G3BP2 | 4271,37 | -0,01 | 0,02 | -0,29 | 7,72E-01 | 8,70E-01 |
| SCARB1 | 167,78 | 0,02 | 0,06 | 0,29 | 7,72E-01 | 8,71E-01 |
| BCL9 | 139,70 | 0,01 | 0,05 | 0,29 | 7,73E-01 | 8,71E-01 |
| KLRC1 | 373,49 | 0,02 | 0,07 | 0,29 | 7,73E-01 | 8,71E-01 |
| PGAP6 | 615,39 | 0,02 | 0,05 | 0,29 | 7,73E-01 | 8,71E-01 |
| PTPN23 | 315,68 | 0,01 | 0,05 | 0,29 | 7,73E-01 | 8,71E-01 |
| TBC1D25 | 297,11 | -0,01 | 0,04 | -0,29 | 7,73E-01 | 8,71E-01 |
| ACVR2B | 673,51 | 0,01 | 0,04 | 0,29 | 7,73E-01 | 8,71E-01 |
| YY1AP1 | 1958,59 | 0,01 | 0,03 | 0,29 | 7,74E-01 | 8,72E-01 |
| TUG1 | 8145,69 | -0,01 | 0,02 | -0,29 | 7,74E-01 | 8,72E-01 |
| AKAP8 | 1271,25 | 0,00 | 0,01 | 0,29 | 7,74E-01 | 8,72E-01 |
| SZT2 | 2106,31 | -0,01 | 0,04 | -0,29 | 7,74E-01 | 8,72E-01 |
| ZNF584 | 140,92 | -0,01 | 0,02 | -0,29 | 7,75E-01 | 8,73E-01 |
| KLF16 | 392,51 | -0,01 | 0,04 | -0,29 | 7,75E-01 | 8,73E-01 |
| MCF2L2 | 297,15 | 0,02 | 0,05 | 0,29 | 7,75E-01 | 8,73E-01 |
| ADPRH | 255,79 | 0,01 | 0,03 | 0,29 | 7,75E-01 | 8,73E-01 |
| COX5B | 1851,47 | 0,01 | 0,03 | 0,29 | 7,75E-01 | 8,73E-01 |
| ARHGEF18 | 257,17 | 0,01 | 0,04 | 0,28 | 7,76E-01 | 8,73E-01 |
| BRD1 | 1418,38 | -0,01 | 0,03 | -0,28 | 7,76E-01 | 8,73E-01 |
| ARHGAP18 | 886,27 | -0,01 | 0,03 | -0,28 | 7,76E-01 | 8,74E-01 |
| LRP1 | 2467,16 | 0,02 | 0,07 | 0,28 | 7,77E-01 | 8,74E-01 |
| DCAF4 | 194,14 | 0,01 | 0,04 | 0,28 | 7,77E-01 | 8,74E-01 |
| SLF1 | 1062,51 | 0,01 | 0,04 | 0,28 | 7,77E-01 | 8,74E-01 |
| CCDC159 | 271,20 | -0,01 | 0,03 | -0,28 | 7,77E-01 | 8,74E-01 |
| TLN2 | 163,65 | 0,03 | 0,10 | 0,28 | 7,78E-01 | 8,74E-01 |
| C22orf39 | 797,07 | 0,00 | 0,01 | -0,28 | 7,77E-01 | 8,74E-01 |
| GNGT2 | 124,60 | -0,01 | 0,03 | -0,28 | 7,77E-01 | 8,74E-01 |
| TOP2B | 5794,49 | -0,01 | 0,03 | -0,28 | 7,77E-01 | 8,74E-01 |
| PIGV | 491,24 | 0,00 | 0,02 | -0,28 | 7,77E-01 | 8,74E-01 |
| WDR27 | 879,85 | 0,01 | 0,04 | 0,28 | 7,78E-01 | 8,74E-01 |
| ZNF207 | 12432,69 | -0,01 | 0,02 | -0,28 | 7,77E-01 | 8,74E-01 |
| BAIAP2 | 109,27 | -0,02 | 0,09 | -0,28 | 7,77E-01 | 8,74E-01 |
| ADSS2 | 2481,71 | 0,01 | 0,02 | 0,28 | 7,78E-01 | 8,74E-01 |
| TMEM255B | 139,88 | -0,01 | 0,05 | -0,28 | 7,78E-01 | 8,74E-01 |
| FOXJ2 | 562,60 | -0,01 | 0,04 | -0,28 | 7,78E-01 | 8,74E-01 |
| WDR73 | 985,73 | 0,00 | 0,01 | -0,28 | 7,77E-01 | 8,74E-01 |
| MTCH1 | 2190,38 | -0,01 | 0,02 | -0,28 | 7,77E-01 | 8,74E-01 |
| TTC34 | 94,44 | 0,02 | 0,08 | 0,28 | 7,76E-01 | 8,74E-01 |
| POLR2J | 548,06 | 0,01 | 0,02 | 0,28 | 7,78E-01 | 8,74E-01 |
| RIMS3 | 89,10 | 0,02 | 0,05 | 0,28 | 7,78E-01 | 8,74E-01 |
| HLCS | 503,22 | 0,01 | 0,03 | 0,28 | 7,78E-01 | 8,74E-01 |
| ZNF208 | 93,53 | 0,02 | 0,07 | 0,28 | 7,78E-01 | 8,74E-01 |
| PHGDH | 80,60 | 0,03 | 0,09 | 0,28 | 7,79E-01 | 8,74E-01 |
| APBA2 | 876,43 | 0,01 | 0,04 | 0,28 | 7,79E-01 | 8,74E-01 |

| | | | | | | |
|---|---|---|---|---|---|---|
| ATM | 28388,40 | -0,01 | 0,03 | -0,28 | 7,79E-01 | 8,74E-01 |
| GUCY1B1 | 233,21 | 0,01 | 0,05 | 0,28 | 7,79E-01 | 8,75E-01 |
| PXYLP1 | 502,54 | -0,01 | 0,03 | -0,28 | 7,79E-01 | 8,75E-01 |
| NPR2 | 86,05 | -0,01 | 0,05 | -0,28 | 7,80E-01 | 8,75E-01 |
| FAM78A | 1772,41 | 0,01 | 0,03 | 0,28 | 7,80E-01 | 8,75E-01 |
| FARP2 | 750,41 | 0,01 | 0,03 | 0,28 | 7,80E-01 | 8,75E-01 |
| ZNF154 | 564,72 | 0,01 | 0,05 | 0,28 | 7,80E-01 | 8,75E-01 |
| SPRING1 | 1208,05 | 0,00 | 0,01 | 0,28 | 7,80E-01 | 8,75E-01 |
| PLEKHA7 | 100,48 | 0,02 | 0,08 | 0,28 | 7,80E-01 | 8,75E-01 |
| DROSHA | 954,71 | -0,01 | 0,02 | -0,28 | 7,80E-01 | 8,75E-01 |
| EXOC7 | 2245,31 | -0,01 | 0,03 | -0,28 | 7,80E-01 | 8,75E-01 |
| RDH10 | 188,82 | -0,01 | 0,03 | -0,28 | 7,81E-01 | 8,75E-01 |
| ASXL1 | 2357,11 | -0,01 | 0,02 | -0,28 | 7,81E-01 | 8,75E-01 |
| MAP3K9 | 172,66 | -0,02 | 0,06 | -0,28 | 7,81E-01 | 8,75E-01 |
| ARPIN | 271,36 | 0,02 | 0,07 | 0,28 | 7,81E-01 | 8,75E-01 |
| MRPL10 | 227,32 | 0,01 | 0,02 | 0,28 | 7,81E-01 | 8,75E-01 |
| DDX41 | 935,75 | -0,01 | 0,02 | -0,28 | 7,81E-01 | 8,75E-01 |
| XYLB | 114,90 | 0,01 | 0,05 | 0,28 | 7,81E-01 | 8,75E-01 |
| TNIK | 2271,56 | 0,01 | 0,02 | 0,28 | 7,81E-01 | 8,75E-01 |
| RAB14 | 2766,44 | -0,01 | 0,02 | -0,28 | 7,82E-01 | 8,76E-01 |
| ELMOD3 | 813,04 | 0,00 | 0,02 | 0,28 | 7,82E-01 | 8,76E-01 |
| PEX5 | 530,47 | 0,00 | 0,02 | 0,28 | 7,82E-01 | 8,76E-01 |
| GPR135 | 139,91 | 0,01 | 0,04 | 0,28 | 7,82E-01 | 8,76E-01 |
| PLA2G7 | 251,80 | 0,01 | 0,05 | 0,28 | 7,82E-01 | 8,76E-01 |
| NTAQ1 | 194,06 | 0,01 | 0,03 | 0,28 | 7,81E-01 | 8,76E-01 |
| UBB | 57536,76 | 0,02 | 0,07 | 0,28 | 7,82E-01 | 8,76E-01 |
| SNX33 | 176,54 | -0,01 | 0,04 | -0,28 | 7,82E-01 | 8,76E-01 |
| C9orf72 | 7571,66 | 0,01 | 0,04 | 0,28 | 7,82E-01 | 8,76E-01 |
| TIPRL | 1028,21 | 0,01 | 0,03 | 0,28 | 7,82E-01 | 8,76E-01 |
| SMYD3 | 630,69 | -0,01 | 0,04 | -0,28 | 7,83E-01 | 8,76E-01 |
| JKAMP | 1389,60 | -0,01 | 0,03 | -0,28 | 7,83E-01 | 8,76E-01 |
| NDOR1 | 239,04 | 0,01 | 0,04 | 0,28 | 7,83E-01 | 8,76E-01 |
| DNAJC8 | 1218,23 | -0,01 | 0,03 | -0,28 | 7,83E-01 | 8,76E-01 |
| LMF2 | 609,87 | 0,01 | 0,04 | 0,28 | 7,83E-01 | 8,76E-01 |
| NPIPB4 | 234,67 | -0,02 | 0,06 | -0,27 | 7,84E-01 | 8,77E-01 |
| DXO | 103,44 | 0,01 | 0,03 | 0,27 | 7,85E-01 | 8,78E-01 |
| DENND4B | 2040,84 | -0,01 | 0,05 | -0,27 | 7,85E-01 | 8,78E-01 |
| UBTF | 1980,90 | -0,01 | 0,02 | -0,27 | 7,85E-01 | 8,78E-01 |
| GPBP1L1 | 3363,76 | 0,00 | 0,02 | -0,27 | 7,85E-01 | 8,78E-01 |
| PLEKHF1 | 514,74 | -0,01 | 0,04 | -0,27 | 7,85E-01 | 8,78E-01 |
| TULP3 | 86,86 | 0,01 | 0,03 | 0,27 | 7,85E-01 | 8,78E-01 |
| COMMD5 | 566,11 | 0,00 | 0,01 | 0,27 | 7,85E-01 | 8,78E-01 |
| FERMT3 | 1614,86 | 0,01 | 0,04 | 0,27 | 7,86E-01 | 8,78E-01 |
| PHACTR4 | 458,00 | -0,01 | 0,03 | -0,27 | 7,86E-01 | 8,78E-01 |
| LAIR2 | 75,16 | -0,02 | 0,07 | -0,27 | 7,86E-01 | 8,78E-01 |
| NSUN2 | 2297,35 | 0,01 | 0,02 | 0,27 | 7,86E-01 | 8,78E-01 |

| | | | | | | |
|---|---|---|---|---|---|---|
| TBC1D13 | 250,60 | 0,01 | 0,04 | 0,27 | 7,86E-01 | 8,79E-01 |
| ZW10 | 462,05 | 0,01 | 0,02 | 0,27 | 7,87E-01 | 8,79E-01 |
| SDF4 | 863,43 | 0,01 | 0,03 | 0,27 | 7,87E-01 | 8,79E-01 |
| MTOR | 1886,54 | -0,01 | 0,03 | -0,27 | 7,87E-01 | 8,79E-01 |
| CCDC9 | 489,80 | -0,01 | 0,03 | -0,27 | 7,88E-01 | 8,80E-01 |
| PKHD1L1 | 240,14 | 0,02 | 0,07 | 0,27 | 7,88E-01 | 8,80E-01 |
| ADA2 | 3311,13 | -0,01 | 0,04 | -0,27 | 7,88E-01 | 8,80E-01 |
| PAFAH2 | 762,98 | -0,01 | 0,02 | -0,27 | 7,88E-01 | 8,80E-01 |
| INPP5F | 733,95 | -0,01 | 0,04 | -0,27 | 7,88E-01 | 8,80E-01 |
| PLCB1 | 1449,18 | -0,01 | 0,03 | -0,27 | 7,88E-01 | 8,80E-01 |
| KCTD18 | 847,12 | 0,00 | 0,01 | 0,27 | 7,89E-01 | 8,80E-01 |
| NOD1 | 735,21 | -0,01 | 0,03 | -0,27 | 7,90E-01 | 8,82E-01 |
| INTS9 | 454,45 | 0,00 | 0,02 | -0,27 | 7,90E-01 | 8,82E-01 |
| DIXDC1 | 112,71 | -0,02 | 0,06 | -0,27 | 7,90E-01 | 8,82E-01 |
| SACM1L | 3953,38 | 0,01 | 0,03 | 0,27 | 7,90E-01 | 8,82E-01 |
| TRPM2 | 287,55 | -0,02 | 0,06 | -0,27 | 7,90E-01 | 8,82E-01 |
| FAM110A | 369,78 | 0,01 | 0,04 | 0,27 | 7,91E-01 | 8,82E-01 |
| TPM1 | 887,00 | 0,01 | 0,04 | 0,27 | 7,91E-01 | 8,82E-01 |
| ADAT1 | 1472,43 | 0,01 | 0,02 | 0,27 | 7,91E-01 | 8,82E-01 |
| SMARCAL1 | 489,47 | -0,01 | 0,02 | -0,26 | 7,92E-01 | 8,83E-01 |
| H2BC13 | 197,63 | 0,01 | 0,06 | 0,26 | 7,92E-01 | 8,83E-01 |
| ZHX3 | 288,62 | -0,01 | 0,04 | -0,26 | 7,92E-01 | 8,83E-01 |
| CEP112 | 75,57 | 0,02 | 0,09 | 0,26 | 7,92E-01 | 8,83E-01 |
| MTIF3 | 630,41 | -0,01 | 0,02 | -0,26 | 7,92E-01 | 8,83E-01 |
| EMC8 | 240,77 | 0,01 | 0,03 | 0,26 | 7,92E-01 | 8,83E-01 |
| CTIF | 145,41 | -0,02 | 0,07 | -0,26 | 7,92E-01 | 8,83E-01 |
| SLC49A3 | 291,56 | -0,01 | 0,04 | -0,26 | 7,93E-01 | 8,83E-01 |
| MGMT | 423,34 | -0,01 | 0,02 | -0,26 | 7,93E-01 | 8,84E-01 |
| UTP14A | 382,41 | 0,01 | 0,02 | 0,26 | 7,93E-01 | 8,84E-01 |
| YAF2 | 2024,46 | 0,01 | 0,02 | 0,26 | 7,93E-01 | 8,84E-01 |
| MAP2K5 | 677,93 | -0,01 | 0,03 | -0,26 | 7,94E-01 | 8,85E-01 |
| MAP1LC3B | 1849,73 | -0,01 | 0,03 | -0,26 | 7,95E-01 | 8,85E-01 |
| NACC1 | 262,97 | 0,01 | 0,05 | 0,26 | 7,95E-01 | 8,85E-01 |
| CD33 | 503,10 | 0,01 | 0,04 | 0,26 | 7,95E-01 | 8,85E-01 |
| FAAP24 | 89,38 | -0,01 | 0,04 | -0,26 | 7,95E-01 | 8,85E-01 |
| PLA2G6 | 449,34 | 0,01 | 0,04 | 0,26 | 7,95E-01 | 8,85E-01 |
| MATCAP1 | 84,83 | -0,01 | 0,05 | -0,26 | 7,95E-01 | 8,85E-01 |
| PPP1R12C | 992,70 | 0,01 | 0,05 | 0,26 | 7,95E-01 | 8,85E-01 |
| KLHL6 | 2575,43 | -0,01 | 0,02 | -0,26 | 7,95E-01 | 8,85E-01 |
| PRDM11 | 232,02 | 0,01 | 0,05 | 0,26 | 7,96E-01 | 8,86E-01 |
| MAGED1 | 148,48 | 0,01 | 0,04 | 0,26 | 7,96E-01 | 8,86E-01 |
| STAU2 | 1508,55 | -0,01 | 0,02 | -0,26 | 7,96E-01 | 8,86E-01 |
| POC5 | 606,01 | -0,01 | 0,02 | -0,26 | 7,96E-01 | 8,86E-01 |
| LTBP2 | 220,78 | 0,02 | 0,06 | 0,26 | 7,96E-01 | 8,86E-01 |
| TENT5A | 1973,12 | -0,01 | 0,03 | -0,26 | 7,97E-01 | 8,86E-01 |
| ATF7IP2 | 1000,71 | 0,01 | 0,03 | 0,26 | 7,97E-01 | 8,86E-01 |

| | | | | | | |
|---|---|---|---|---|---|---|
| IL2RB | 635,76 | -0,01 | 0,06 | -0,26 | 7,97E-01 | 8,86E-01 |
| ZNG1E | 1096,45 | -0,01 | 0,03 | -0,26 | 7,97E-01 | 8,87E-01 |
| CCDC92 | 543,71 | 0,01 | 0,02 | 0,26 | 7,97E-01 | 8,87E-01 |
| ITSN1 | 497,84 | 0,01 | 0,05 | 0,26 | 7,97E-01 | 8,87E-01 |
| PGBD2 | 507,66 | 0,00 | 0,02 | -0,26 | 7,97E-01 | 8,87E-01 |
| FLVCR1 | 1020,65 | -0,01 | 0,03 | -0,26 | 7,98E-01 | 8,87E-01 |
| SFI1 | 2110,68 | 0,01 | 0,03 | 0,26 | 7,98E-01 | 8,87E-01 |
| CDON | 73,17 | 0,02 | 0,08 | 0,26 | 7,98E-01 | 8,87E-01 |
| PPP6R2 | 1383,14 | -0,01 | 0,04 | -0,26 | 7,98E-01 | 8,87E-01 |
| ULK4 | 1054,70 | -0,01 | 0,02 | -0,26 | 7,99E-01 | 8,87E-01 |
| ZNF778 | 690,55 | 0,00 | 0,01 | 0,25 | 7,99E-01 | 8,88E-01 |
| ASGR2 | 109,45 | 0,02 | 0,06 | 0,25 | 7,99E-01 | 8,88E-01 |
| ZMYM5 | 870,32 | -0,01 | 0,03 | -0,25 | 8,00E-01 | 8,88E-01 |
| C18orf21 | 209,80 | 0,01 | 0,03 | 0,25 | 8,00E-01 | 8,88E-01 |
| UPP1 | 1157,56 | -0,01 | 0,03 | -0,25 | 8,00E-01 | 8,89E-01 |
| RBKS | 97,76 | -0,01 | 0,03 | -0,25 | 8,01E-01 | 8,89E-01 |
| CTSL | 297,71 | 0,01 | 0,05 | 0,25 | 8,01E-01 | 8,89E-01 |
| PDIA4 | 1087,58 | -0,01 | 0,02 | -0,25 | 8,01E-01 | 8,89E-01 |
| KANSL1L | 1160,76 | -0,01 | 0,03 | -0,25 | 8,01E-01 | 8,89E-01 |
| ZSWIM9 | 140,99 | 0,01 | 0,04 | 0,25 | 8,01E-01 | 8,89E-01 |
| SCPEP1 | 2342,60 | -0,01 | 0,03 | -0,25 | 8,02E-01 | 8,90E-01 |
| GPM6A | 100,50 | 0,03 | 0,12 | 0,25 | 8,02E-01 | 8,90E-01 |
| ODF2 | 706,08 | -0,01 | 0,02 | -0,25 | 8,02E-01 | 8,90E-01 |
| IL1RL1 | 480,13 | -0,02 | 0,08 | -0,25 | 8,02E-01 | 8,90E-01 |
| SDK1 | 78,22 | -0,03 | 0,13 | -0,25 | 8,02E-01 | 8,90E-01 |
| SF3B6 | 481,66 | -0,01 | 0,06 | -0,25 | 8,02E-01 | 8,90E-01 |
| GPR68 | 265,85 | 0,01 | 0,04 | 0,25 | 8,02E-01 | 8,90E-01 |
| H2AC6 | 11657,11 | 0,01 | 0,04 | 0,25 | 8,03E-01 | 8,90E-01 |
| SELP | 371,09 | -0,01 | 0,05 | -0,25 | 8,03E-01 | 8,91E-01 |
| CTTNBP2NL | 121,84 | 0,01 | 0,04 | 0,25 | 8,04E-01 | 8,91E-01 |
| MCFD2 | 957,52 | 0,00 | 0,01 | 0,25 | 8,04E-01 | 8,91E-01 |
| NFATC2 | 1433,80 | -0,01 | 0,03 | -0,25 | 8,04E-01 | 8,91E-01 |
| CUL2 | 734,98 | 0,01 | 0,02 | 0,25 | 8,04E-01 | 8,91E-01 |
| TMCO6 | 415,42 | -0,01 | 0,03 | -0,25 | 8,04E-01 | 8,91E-01 |
| PLEKHA3 | 2266,18 | 0,00 | 0,02 | 0,25 | 8,04E-01 | 8,91E-01 |
| C4orf33 | 596,15 | 0,01 | 0,03 | 0,25 | 8,04E-01 | 8,91E-01 |
| F13A1 | 5540,43 | -0,01 | 0,06 | -0,25 | 8,04E-01 | 8,91E-01 |
| CEP78 | 2223,20 | -0,01 | 0,04 | -0,25 | 8,05E-01 | 8,92E-01 |
| ARMT1 | 753,12 | 0,01 | 0,04 | 0,25 | 8,06E-01 | 8,93E-01 |
| ZNF892 | 140,38 | -0,01 | 0,03 | -0,25 | 8,06E-01 | 8,93E-01 |
| ZNF74 | 241,18 | -0,01 | 0,03 | -0,25 | 8,06E-01 | 8,93E-01 |
| EP400 | 1468,38 | -0,01 | 0,05 | -0,25 | 8,06E-01 | 8,93E-01 |
| GNA15 | 373,14 | 0,01 | 0,03 | 0,24 | 8,08E-01 | 8,94E-01 |
| RGS12 | 285,45 | -0,01 | 0,05 | -0,24 | 8,08E-01 | 8,94E-01 |
| UTP11 | 502,71 | 0,01 | 0,02 | 0,24 | 8,07E-01 | 8,94E-01 |
| USP54 | 140,72 | 0,01 | 0,05 | 0,24 | 8,07E-01 | 8,94E-01 |

| | | | | | | |
|---|---|---|---|---|---|---|
| MEMO1 | 642,00 | 0,01 | 0,02 | 0,24 | 8,08E-01 | 8,94E-01 |
| C1orf174 | 579,10 | 0,00 | 0,01 | 0,24 | 8,08E-01 | 8,94E-01 |
| RYR1 | 106,34 | 0,03 | 0,12 | 0,24 | 8,08E-01 | 8,94E-01 |
| RBM34 | 93,64 | -0,01 | 0,02 | -0,24 | 8,07E-01 | 8,94E-01 |
| GPR52 | 131,70 | -0,01 | 0,05 | -0,24 | 8,08E-01 | 8,94E-01 |
| SMC4 | 1385,40 | -0,01 | 0,03 | -0,24 | 8,07E-01 | 8,94E-01 |
| MLXIP | 1235,34 | 0,01 | 0,06 | 0,24 | 8,07E-01 | 8,94E-01 |
| H2AC12 | 346,28 | 0,02 | 0,07 | 0,24 | 8,08E-01 | 8,94E-01 |
| ENPP3 | 93,99 | -0,02 | 0,08 | -0,24 | 8,07E-01 | 8,94E-01 |
| TNIP1 | 3885,54 | -0,01 | 0,03 | -0,24 | 8,07E-01 | 8,94E-01 |
| MYO16 | 81,94 | -0,03 | 0,12 | -0,24 | 8,08E-01 | 8,94E-01 |
| ZNRF1 | 150,59 | -0,01 | 0,05 | -0,24 | 8,08E-01 | 8,94E-01 |
| FAM50A | 436,01 | 0,00 | 0,02 | -0,24 | 8,08E-01 | 8,94E-01 |
| KMT2B | 579,60 | -0,02 | 0,08 | -0,24 | 8,08E-01 | 8,94E-01 |
| MYO15B | 2083,19 | 0,01 | 0,06 | 0,24 | 8,08E-01 | 8,94E-01 |
| MINK1 | 622,18 | 0,02 | 0,06 | 0,24 | 8,09E-01 | 8,94E-01 |
| ANKRD55 | 161,04 | 0,01 | 0,05 | 0,24 | 8,09E-01 | 8,94E-01 |
| INTS7 | 727,92 | -0,01 | 0,02 | -0,24 | 8,09E-01 | 8,94E-01 |
| MYH3 | 231,24 | 0,01 | 0,05 | 0,24 | 8,09E-01 | 8,94E-01 |
| VANGL1 | 204,92 | 0,01 | 0,04 | 0,24 | 8,10E-01 | 8,94E-01 |
| COL4A4 | 165,36 | 0,02 | 0,08 | 0,24 | 8,10E-01 | 8,95E-01 |
| TTC22 | 318,50 | 0,01 | 0,04 | 0,24 | 8,11E-01 | 8,96E-01 |
| IFFO2 | 997,22 | 0,01 | 0,03 | 0,24 | 8,11E-01 | 8,96E-01 |
| C8orf76 | 191,76 | 0,00 | 0,01 | -0,24 | 8,11E-01 | 8,96E-01 |
| SLTM | 4345,37 | -0,01 | 0,03 | -0,24 | 8,11E-01 | 8,96E-01 |
| MAVS | 2189,28 | -0,01 | 0,04 | -0,24 | 8,11E-01 | 8,96E-01 |
| CES1 | 284,33 | -0,02 | 0,08 | -0,24 | 8,12E-01 | 8,96E-01 |
| PRXL2A | 115,47 | 0,01 | 0,04 | 0,24 | 8,12E-01 | 8,96E-01 |
| CNTNAP2 | 108,32 | 0,03 | 0,12 | 0,24 | 8,12E-01 | 8,96E-01 |
| AK5 | 422,36 | -0,01 | 0,05 | -0,24 | 8,12E-01 | 8,96E-01 |
| UBFD1 | 704,83 | 0,01 | 0,02 | 0,24 | 8,13E-01 | 8,97E-01 |
| CHMP4B | 791,94 | 0,01 | 0,03 | 0,24 | 8,13E-01 | 8,97E-01 |
| RGMB | 196,71 | -0,01 | 0,05 | -0,24 | 8,13E-01 | 8,97E-01 |
| GET1 | 441,14 | -0,01 | 0,03 | -0,24 | 8,14E-01 | 8,98E-01 |
| CLEC2B | 1677,61 | -0,01 | 0,06 | -0,24 | 8,14E-01 | 8,98E-01 |
| HCCS | 294,01 | 0,01 | 0,02 | 0,24 | 8,14E-01 | 8,98E-01 |
| MYLK | 569,64 | -0,01 | 0,06 | -0,24 | 8,14E-01 | 8,98E-01 |
| KMT5C | 129,26 | 0,01 | 0,05 | 0,24 | 8,14E-01 | 8,98E-01 |
| TBCC | 1616,02 | -0,01 | 0,02 | -0,24 | 8,14E-01 | 8,98E-01 |
| B4GAT1 | 74,31 | 0,01 | 0,04 | 0,23 | 8,15E-01 | 8,99E-01 |
| FRG1 | 544,21 | 0,01 | 0,04 | 0,23 | 8,15E-01 | 8,99E-01 |
| DEK | 2991,54 | 0,01 | 0,05 | 0,23 | 8,16E-01 | 8,99E-01 |
| SETD1B | 760,04 | -0,02 | 0,07 | -0,23 | 8,16E-01 | 8,99E-01 |
| DPP9 | 682,67 | -0,01 | 0,04 | -0,23 | 8,16E-01 | 8,99E-01 |
| KMT2A | 4034,14 | -0,01 | 0,03 | -0,23 | 8,16E-01 | 8,99E-01 |
| PIP4K2B | 1628,99 | 0,00 | 0,02 | -0,23 | 8,16E-01 | 8,99E-01 |

| | | | | | | |
|---|---|---|---|---|---|---|
| MTCL1 | 97,48 | -0,03 | 0,12 | -0,23 | 8,16E-01 | 8,99E-01 |
| ARHGEF9 | 965,53 | -0,01 | 0,02 | -0,23 | 8,16E-01 | 8,99E-01 |
| MRE11 | 2059,80 | 0,00 | 0,02 | 0,23 | 8,16E-01 | 8,99E-01 |
| PLAAT3 | 157,78 | 0,01 | 0,04 | 0,23 | 8,16E-01 | 8,99E-01 |
| USP11 | 311,53 | -0,01 | 0,04 | -0,23 | 8,17E-01 | 9,00E-01 |
| TMEM176B | 204,02 | -0,03 | 0,14 | -0,23 | 8,17E-01 | 9,00E-01 |
| CUBN | 849,61 | -0,01 | 0,04 | -0,23 | 8,17E-01 | 9,00E-01 |
| PFN2 | 97,16 | -0,01 | 0,05 | -0,23 | 8,17E-01 | 9,00E-01 |
| GOLM2 | 3385,51 | -0,01 | 0,03 | -0,23 | 8,17E-01 | 9,00E-01 |
| TBC1D8B | 92,72 | -0,01 | 0,05 | -0,23 | 8,17E-01 | 9,00E-01 |
| PCNX4 | 3231,88 | 0,00 | 0,02 | -0,23 | 8,17E-01 | 9,00E-01 |
| C14orf132 | 133,67 | 0,02 | 0,07 | 0,23 | 8,18E-01 | 9,00E-01 |
| WDR53 | 223,41 | 0,00 | 0,02 | 0,23 | 8,18E-01 | 9,00E-01 |
| NEFL | 84,34 | 0,02 | 0,08 | 0,23 | 8,18E-01 | 9,00E-01 |
| SAMD9 | 5112,82 | 0,02 | 0,08 | 0,23 | 8,18E-01 | 9,00E-01 |
| NSG1 | 133,09 | -0,02 | 0,07 | -0,23 | 8,19E-01 | 9,00E-01 |
| PPM1L | 935,80 | -0,01 | 0,03 | -0,23 | 8,18E-01 | 9,00E-01 |
| SCML4 | 1753,43 | -0,01 | 0,03 | -0,23 | 8,19E-01 | 9,00E-01 |
| CFAP44 | 362,60 | -0,01 | 0,03 | -0,23 | 8,19E-01 | 9,00E-01 |
| SDHD | 1341,80 | 0,01 | 0,02 | 0,23 | 8,19E-01 | 9,00E-01 |
| FZD2 | 78,36 | -0,01 | 0,05 | -0,23 | 8,19E-01 | 9,00E-01 |
| HEG1 | 742,02 | -0,01 | 0,03 | -0,23 | 8,18E-01 | 9,00E-01 |
| PARG | 1245,25 | 0,00 | 0,02 | -0,23 | 8,19E-01 | 9,00E-01 |
| CYLD | 12772,55 | -0,01 | 0,03 | -0,23 | 8,19E-01 | 9,00E-01 |
| ZNF330 | 887,13 | 0,01 | 0,02 | 0,23 | 8,19E-01 | 9,00E-01 |
| CDHR3 | 425,00 | 0,01 | 0,04 | 0,23 | 8,19E-01 | 9,01E-01 |
| GNAZ | 413,17 | 0,01 | 0,06 | 0,23 | 8,19E-01 | 9,01E-01 |
| C1orf50 | 297,84 | 0,00 | 0,02 | 0,23 | 8,20E-01 | 9,01E-01 |
| FAH | 233,73 | 0,01 | 0,04 | 0,23 | 8,20E-01 | 9,01E-01 |
| SLC41A1 | 183,75 | 0,01 | 0,05 | 0,23 | 8,20E-01 | 9,01E-01 |
| CDAN1 | 320,64 | -0,01 | 0,04 | -0,23 | 8,20E-01 | 9,01E-01 |
| ZNF665 | 148,59 | 0,01 | 0,04 | 0,23 | 8,20E-01 | 9,01E-01 |
| CEP135 | 1143,36 | -0,01 | 0,03 | -0,23 | 8,21E-01 | 9,02E-01 |
| CD302 | 2440,97 | -0,01 | 0,04 | -0,23 | 8,21E-01 | 9,02E-01 |
| HDAC5 | 937,51 | -0,01 | 0,04 | -0,23 | 8,21E-01 | 9,02E-01 |
| DCAF8 | 2624,76 | 0,00 | 0,02 | -0,23 | 8,22E-01 | 9,02E-01 |
| RFX1 | 199,83 | -0,02 | 0,07 | -0,22 | 8,22E-01 | 9,03E-01 |
| CACNA2D2 | 109,57 | -0,02 | 0,08 | -0,22 | 8,22E-01 | 9,03E-01 |
| GSDMD | 2603,19 | -0,01 | 0,02 | -0,22 | 8,22E-01 | 9,03E-01 |
| WDR33 | 3386,81 | 0,00 | 0,02 | 0,22 | 8,22E-01 | 9,03E-01 |
| SPMIP4 | 77,65 | -0,01 | 0,04 | -0,22 | 8,22E-01 | 9,03E-01 |
| ARFGAP1 | 662,35 | 0,01 | 0,05 | 0,22 | 8,23E-01 | 9,03E-01 |
| IDI1 | 1223,94 | 0,01 | 0,05 | 0,22 | 8,23E-01 | 9,03E-01 |
| SPIRE1 | 161,31 | -0,01 | 0,04 | -0,22 | 8,24E-01 | 9,04E-01 |
| ADGRV1 | 99,54 | 0,03 | 0,14 | 0,22 | 8,24E-01 | 9,04E-01 |
| KARS1 | 2160,22 | 0,01 | 0,02 | 0,22 | 8,24E-01 | 9,04E-01 |

| | | | | | | |
|---|---|---|---|---|---|---|
| SGSM3 | 86,95 | -0,01 | 0,04 | -0,22 | 8,24E-01 | 9,04E-01 |
| ERO1B | 1258,65 | -0,01 | 0,02 | -0,22 | 8,24E-01 | 9,04E-01 |
| PYROXD2 | 451,04 | 0,01 | 0,03 | 0,22 | 8,24E-01 | 9,04E-01 |
| PSMA1 | 880,68 | 0,00 | 0,02 | 0,22 | 8,24E-01 | 9,04E-01 |
| MAFB | 552,36 | -0,01 | 0,04 | -0,22 | 8,24E-01 | 9,04E-01 |
| PYGO2 | 378,10 | 0,01 | 0,04 | 0,22 | 8,25E-01 | 9,04E-01 |
| AXIN1 | 1061,35 | -0,01 | 0,03 | -0,22 | 8,25E-01 | 9,04E-01 |
| BLTP3A | 790,51 | 0,01 | 0,03 | 0,22 | 8,25E-01 | 9,04E-01 |
| SLC44A1 | 1473,78 | 0,00 | 0,02 | 0,22 | 8,25E-01 | 9,04E-01 |
| MS4A14 | 384,97 | -0,01 | 0,05 | -0,22 | 8,25E-01 | 9,04E-01 |
| DEAF1 | 304,71 | 0,01 | 0,04 | 0,22 | 8,25E-01 | 9,04E-01 |
| RDX | 634,37 | 0,01 | 0,03 | 0,22 | 8,25E-01 | 9,04E-01 |
| PDGFD | 324,89 | -0,01 | 0,05 | -0,22 | 8,26E-01 | 9,04E-01 |
| FHOD1 | 315,09 | -0,01 | 0,05 | -0,22 | 8,26E-01 | 9,04E-01 |
| EIF6 | 828,83 | -0,01 | 0,02 | -0,22 | 8,25E-01 | 9,04E-01 |
| APTX | 539,25 | 0,00 | 0,01 | 0,22 | 8,26E-01 | 9,05E-01 |
| ADCY9 | 393,49 | -0,01 | 0,04 | -0,22 | 8,26E-01 | 9,05E-01 |
| LEPROT | 1313,04 | 0,01 | 0,03 | 0,22 | 8,26E-01 | 9,05E-01 |
| TFDP2 | 1437,11 | 0,00 | 0,02 | 0,22 | 8,27E-01 | 9,05E-01 |
| PRKD3 | 1834,11 | 0,00 | 0,02 | 0,22 | 8,27E-01 | 9,05E-01 |
| CRAMP1 | 333,09 | 0,01 | 0,04 | 0,22 | 8,27E-01 | 9,06E-01 |
| ACO1 | 383,63 | -0,01 | 0,02 | -0,22 | 8,28E-01 | 9,06E-01 |
| PCSK5 | 754,89 | 0,01 | 0,03 | 0,22 | 8,28E-01 | 9,06E-01 |
| POGLUT3 | 227,03 | -0,01 | 0,03 | -0,22 | 8,28E-01 | 9,06E-01 |
| BRD4 | 1658,23 | 0,01 | 0,05 | 0,22 | 8,28E-01 | 9,06E-01 |
| PRKRIP1 | 786,22 | 0,00 | 0,01 | 0,22 | 8,29E-01 | 9,07E-01 |
| ZDHHC24 | 400,54 | 0,01 | 0,03 | 0,22 | 8,29E-01 | 9,07E-01 |
| ELP3 | 1204,06 | 0,00 | 0,02 | -0,22 | 8,29E-01 | 9,07E-01 |
| LENG8 | 2078,07 | 0,02 | 0,07 | 0,22 | 8,29E-01 | 9,07E-01 |
| CENPN | 78,70 | 0,01 | 0,04 | 0,22 | 8,29E-01 | 9,07E-01 |
| TCTN1 | 383,30 | 0,01 | 0,03 | 0,22 | 8,30E-01 | 9,07E-01 |
| PTRH2 | 516,06 | 0,00 | 0,02 | -0,22 | 8,29E-01 | 9,07E-01 |
| PTGDR | 1149,63 | 0,01 | 0,05 | 0,21 | 8,30E-01 | 9,07E-01 |
| ECT2 | 307,94 | 0,01 | 0,04 | 0,21 | 8,30E-01 | 9,08E-01 |
| ATP13A3 | 3252,66 | 0,00 | 0,02 | 0,21 | 8,30E-01 | 9,08E-01 |
| CSF1R | 1307,24 | -0,01 | 0,05 | -0,21 | 8,31E-01 | 9,08E-01 |
| FOXK1 | 1235,56 | 0,01 | 0,04 | 0,21 | 8,31E-01 | 9,08E-01 |
| RCBTB1 | 997,47 | 0,00 | 0,02 | -0,21 | 8,31E-01 | 9,08E-01 |
| HOMER3 | 225,21 | 0,01 | 0,04 | 0,21 | 8,31E-01 | 9,08E-01 |
| KCTD1 | 122,94 | 0,01 | 0,05 | 0,21 | 8,32E-01 | 9,09E-01 |
| CAMP | 396,73 | 0,02 | 0,08 | 0,21 | 8,32E-01 | 9,09E-01 |
| SUMO1 | 1541,98 | -0,01 | 0,03 | -0,21 | 8,32E-01 | 9,09E-01 |
| SLC35F5 | 2944,35 | 0,01 | 0,03 | 0,21 | 8,32E-01 | 9,09E-01 |
| FBXO46 | 123,99 | -0,01 | 0,05 | -0,21 | 8,33E-01 | 9,09E-01 |
| ANKRD27 | 1260,05 | 0,00 | 0,01 | -0,21 | 8,33E-01 | 9,09E-01 |
| MYO3B | 82,01 | -0,03 | 0,12 | -0,21 | 8,33E-01 | 9,10E-01 |

| | | | | | | |
|---|---|---|---|---|---|---|
| RMC1 | 1358,30 | 0,00 | 0,02 | 0,21 | 8,34E-01 | 9,10E-01 |
| LGALS3BP | 97,52 | -0,01 | 0,07 | -0,21 | 8,34E-01 | 9,11E-01 |
| STARD9 | 729,72 | 0,01 | 0,04 | 0,21 | 8,35E-01 | 9,11E-01 |
| PAQR7 | 220,90 | 0,01 | 0,03 | 0,21 | 8,35E-01 | 9,11E-01 |
| NGLY1 | 2762,84 | 0,00 | 0,02 | -0,21 | 8,34E-01 | 9,11E-01 |
| TERF2 | 565,73 | 0,00 | 0,02 | 0,21 | 8,35E-01 | 9,11E-01 |
| LDHA | 2210,26 | 0,00 | 0,02 | -0,21 | 8,34E-01 | 9,11E-01 |
| ALKBH5 | 2130,13 | 0,00 | 0,02 | 0,21 | 8,35E-01 | 9,11E-01 |
| DNASE2 | 749,29 | -0,01 | 0,03 | -0,21 | 8,35E-01 | 9,11E-01 |
| LCLAT1 | 767,52 | -0,01 | 0,03 | -0,21 | 8,35E-01 | 9,11E-01 |
| ERCC3 | 1160,64 | 0,00 | 0,02 | 0,21 | 8,35E-01 | 9,11E-01 |
| TCFL5 | 309,58 | 0,00 | 0,02 | 0,21 | 8,36E-01 | 9,12E-01 |
| SLC3A2 | 1227,03 | 0,00 | 0,02 | -0,21 | 8,36E-01 | 9,12E-01 |
| KLHDC4 | 1331,14 | 0,01 | 0,04 | 0,21 | 8,36E-01 | 9,12E-01 |
| EIF2AK4 | 931,34 | 0,00 | 0,02 | 0,21 | 8,36E-01 | 9,12E-01 |
| DAZAP1 | 1731,58 | 0,01 | 0,03 | 0,21 | 8,36E-01 | 9,12E-01 |
| NEMF | 2761,45 | 0,00 | 0,02 | -0,21 | 8,37E-01 | 9,12E-01 |
| FAM153A | 359,15 | -0,01 | 0,06 | -0,21 | 8,37E-01 | 9,12E-01 |
| ERMAP | 658,19 | 0,01 | 0,03 | 0,21 | 8,37E-01 | 9,12E-01 |
| PPIB | 2571,65 | -0,01 | 0,03 | -0,21 | 8,37E-01 | 9,12E-01 |
| DPAGT1 | 169,40 | 0,01 | 0,03 | 0,20 | 8,38E-01 | 9,13E-01 |
| CLEC4F | 75,39 | -0,03 | 0,13 | -0,20 | 8,38E-01 | 9,13E-01 |
| KCNA2 | 121,30 | 0,01 | 0,07 | 0,20 | 8,38E-01 | 9,13E-01 |
| MKNK2 | 2631,96 | -0,01 | 0,05 | -0,20 | 8,38E-01 | 9,13E-01 |
| PTGER4 | 2148,11 | 0,00 | 0,02 | 0,20 | 8,39E-01 | 9,14E-01 |
| MRFAP1L1 | 1754,25 | -0,01 | 0,04 | -0,20 | 8,39E-01 | 9,14E-01 |
| RGPD2 | 220,86 | -0,02 | 0,11 | -0,20 | 8,39E-01 | 9,14E-01 |
| C12orf42 | 248,95 | -0,01 | 0,04 | -0,20 | 8,39E-01 | 9,14E-01 |
| CACYBP | 918,70 | 0,01 | 0,03 | 0,20 | 8,40E-01 | 9,14E-01 |
| ITFG1 | 1659,76 | 0,00 | 0,02 | -0,20 | 8,40E-01 | 9,14E-01 |
| UBAC2 | 1763,05 | 0,00 | 0,02 | 0,20 | 8,39E-01 | 9,14E-01 |
| MYO1D | 297,23 | -0,01 | 0,04 | -0,20 | 8,39E-01 | 9,14E-01 |
| IDH1 | 1251,57 | -0,01 | 0,03 | -0,20 | 8,39E-01 | 9,14E-01 |
| ZNF671 | 987,93 | 0,00 | 0,02 | -0,20 | 8,40E-01 | 9,14E-01 |
| GAS6 | 117,26 | -0,01 | 0,06 | -0,20 | 8,40E-01 | 9,14E-01 |
| CTDSP1 | 1523,98 | 0,01 | 0,03 | 0,20 | 8,41E-01 | 9,15E-01 |
| H2BC12 | 2258,24 | 0,01 | 0,03 | 0,20 | 8,41E-01 | 9,15E-01 |
| P2RY2 | 269,65 | 0,01 | 0,06 | 0,20 | 8,40E-01 | 9,15E-01 |
| SDK2 | 124,17 | -0,02 | 0,10 | -0,20 | 8,40E-01 | 9,15E-01 |
| NAPRT | 584,30 | 0,01 | 0,04 | 0,20 | 8,42E-01 | 9,16E-01 |
| TUBGCP2 | 1520,47 | 0,00 | 0,02 | -0,20 | 8,42E-01 | 9,16E-01 |
| RIN2 | 543,55 | -0,01 | 0,04 | -0,20 | 8,42E-01 | 9,16E-01 |
| TCEAL4 | 157,12 | 0,01 | 0,04 | 0,20 | 8,42E-01 | 9,16E-01 |
| AP1G2 | 1214,26 | 0,01 | 0,03 | 0,20 | 8,42E-01 | 9,16E-01 |
| MAPKAPK5 | 1497,38 | 0,00 | 0,02 | -0,20 | 8,43E-01 | 9,16E-01 |
| GET4 | 155,68 | -0,01 | 0,04 | -0,20 | 8,43E-01 | 9,17E-01 |

| | | | | | | |
|---|---|---|---|---|---|---|
| DENND1C | 457,03 | -0,01 | 0,05 | -0,20 | 8,44E-01 | 9,17E-01 |
| CDADC1 | 388,65 | 0,00 | 0,02 | -0,20 | 8,44E-01 | 9,17E-01 |
| USP33 | 3844,46 | 0,00 | 0,02 | -0,20 | 8,44E-01 | 9,17E-01 |
| TBP | 409,59 | 0,00 | 0,01 | 0,20 | 8,44E-01 | 9,17E-01 |
| CSDE1 | 15252,28 | 0,01 | 0,03 | 0,20 | 8,45E-01 | 9,18E-01 |
| CTTN | 492,62 | 0,01 | 0,07 | 0,20 | 8,45E-01 | 9,18E-01 |
| ATP1A1 | 3353,29 | 0,00 | 0,02 | 0,20 | 8,45E-01 | 9,18E-01 |
| CARMIL1 | 436,48 | -0,01 | 0,05 | -0,20 | 8,45E-01 | 9,18E-01 |
| SET | 5636,98 | -0,01 | 0,03 | -0,20 | 8,45E-01 | 9,18E-01 |
| CIT | 124,15 | -0,02 | 0,09 | -0,19 | 8,45E-01 | 9,18E-01 |
| NELFA | 252,61 | 0,01 | 0,05 | 0,19 | 8,46E-01 | 9,18E-01 |
| PPARA | 555,11 | -0,01 | 0,03 | -0,19 | 8,46E-01 | 9,18E-01 |
| LIME1 | 132,47 | -0,01 | 0,03 | -0,19 | 8,46E-01 | 9,18E-01 |
| PRRC2A | 1097,41 | -0,01 | 0,07 | -0,19 | 8,46E-01 | 9,18E-01 |
| MNT | 716,06 | -0,01 | 0,05 | -0,19 | 8,46E-01 | 9,18E-01 |
| STX17 | 1393,49 | 0,00 | 0,02 | -0,19 | 8,46E-01 | 9,19E-01 |
| SLC8B1 | 1307,03 | 0,01 | 0,03 | 0,19 | 8,47E-01 | 9,19E-01 |
| MEGF6 | 1546,77 | -0,01 | 0,04 | -0,19 | 8,47E-01 | 9,19E-01 |
| OCRL | 245,82 | -0,01 | 0,03 | -0,19 | 8,47E-01 | 9,19E-01 |
| ZKSCAN7 | 222,34 | 0,00 | 0,03 | 0,19 | 8,47E-01 | 9,19E-01 |
| CDK2 | 210,67 | 0,00 | 0,02 | 0,19 | 8,47E-01 | 9,19E-01 |
| TNFSF13 | 485,49 | 0,01 | 0,03 | 0,19 | 8,47E-01 | 9,19E-01 |
| TRAF3IP1 | 265,98 | -0,01 | 0,03 | -0,19 | 8,48E-01 | 9,19E-01 |
| PDLIM5 | 1672,29 | 0,00 | 0,02 | -0,19 | 8,48E-01 | 9,19E-01 |
| ZNF100 | 830,43 | -0,01 | 0,05 | -0,19 | 8,48E-01 | 9,19E-01 |
| ZNF79 | 483,46 | 0,00 | 0,02 | 0,19 | 8,48E-01 | 9,19E-01 |
| SOX13 | 101,80 | 0,02 | 0,08 | 0,19 | 8,48E-01 | 9,19E-01 |
| LILRB1 | 670,56 | 0,01 | 0,05 | 0,19 | 8,48E-01 | 9,19E-01 |
| ZNF672 | 745,22 | 0,00 | 0,02 | -0,19 | 8,48E-01 | 9,19E-01 |
| PRPF31 | 828,31 | 0,00 | 0,02 | 0,19 | 8,48E-01 | 9,19E-01 |
| AIFM3 | 128,00 | 0,01 | 0,06 | 0,19 | 8,49E-01 | 9,20E-01 |
| DEPDC5 | 748,87 | -0,01 | 0,03 | -0,19 | 8,49E-01 | 9,20E-01 |
| MSH4 | 93,76 | 0,01 | 0,04 | 0,19 | 8,49E-01 | 9,20E-01 |
| WDR54 | 139,17 | 0,01 | 0,03 | 0,19 | 8,49E-01 | 9,20E-01 |
| RPRD2 | 1024,88 | 0,00 | 0,02 | -0,19 | 8,49E-01 | 9,20E-01 |
| DCLRE1C | 1304,28 | 0,00 | 0,01 | 0,19 | 8,49E-01 | 9,20E-01 |
| AMBRA1 | 970,39 | -0,01 | 0,03 | -0,19 | 8,50E-01 | 9,20E-01 |
| PRPSAP1 | 388,86 | 0,00 | 0,02 | -0,19 | 8,49E-01 | 9,20E-01 |
| BCOR | 638,22 | 0,01 | 0,03 | 0,19 | 8,49E-01 | 9,20E-01 |
| MAGIX | 76,18 | -0,01 | 0,04 | -0,19 | 8,50E-01 | 9,20E-01 |
| IGHMBP2 | 412,91 | -0,01 | 0,04 | -0,19 | 8,50E-01 | 9,20E-01 |
| BET1L | 491,11 | -0,01 | 0,04 | -0,19 | 8,50E-01 | 9,20E-01 |
| ACTR10 | 1214,34 | 0,00 | 0,02 | -0,19 | 8,50E-01 | 9,20E-01 |
| RELCH | 5805,73 | 0,00 | 0,02 | 0,19 | 8,51E-01 | 9,21E-01 |
| CLEC4C | 224,25 | 0,02 | 0,08 | 0,19 | 8,51E-01 | 9,21E-01 |
| RASAL3 | 860,11 | 0,01 | 0,05 | 0,19 | 8,51E-01 | 9,21E-01 |

| | | | | | | |
|---|---|---|---|---|---|---|
| ZFP37 | 96,11 | 0,01 | 0,05 | 0,19 | 8,51E-01 | 9,21E-01 |
| BCR | 1555,54 | 0,01 | 0,04 | 0,19 | 8,51E-01 | 9,21E-01 |
| GNPDA1 | 527,41 | 0,00 | 0,02 | -0,19 | 8,51E-01 | 9,21E-01 |
| CDC42BPB | 396,35 | -0,01 | 0,04 | -0,19 | 8,52E-01 | 9,21E-01 |
| LAMTOR1 | 1867,10 | 0,00 | 0,02 | 0,19 | 8,52E-01 | 9,21E-01 |
| CCDC136 | 265,62 | 0,01 | 0,04 | 0,19 | 8,53E-01 | 9,21E-01 |
| ADGRA3 | 136,39 | -0,01 | 0,06 | -0,19 | 8,52E-01 | 9,21E-01 |
| TMEM65 | 3500,36 | 0,00 | 0,03 | 0,19 | 8,52E-01 | 9,21E-01 |
| CSNK2A1 | 3477,57 | 0,00 | 0,01 | -0,19 | 8,53E-01 | 9,21E-01 |
| PMF1 | 281,47 | 0,00 | 0,02 | -0,19 | 8,52E-01 | 9,21E-01 |
| MSI2 | 2556,70 | 0,00 | 0,02 | 0,19 | 8,53E-01 | 9,21E-01 |
| ESCO1 | 1350,52 | 0,01 | 0,05 | 0,19 | 8,52E-01 | 9,21E-01 |
| DDX39B | 1556,21 | -0,01 | 0,05 | -0,19 | 8,52E-01 | 9,21E-01 |
| USP21 | 283,19 | 0,00 | 0,03 | 0,19 | 8,53E-01 | 9,22E-01 |
| METTL14 | 1106,78 | 0,01 | 0,03 | 0,18 | 8,53E-01 | 9,22E-01 |
| VHL | 2972,88 | 0,00 | 0,01 | -0,18 | 8,53E-01 | 9,22E-01 |
| PRKCSH | 957,40 | -0,01 | 0,04 | -0,18 | 8,53E-01 | 9,22E-01 |
| SLC7A7 | 2457,95 | -0,01 | 0,03 | -0,18 | 8,54E-01 | 9,22E-01 |
| ZNF781 | 80,67 | -0,01 | 0,05 | -0,18 | 8,54E-01 | 9,23E-01 |
| PANK4 | 857,02 | 0,00 | 0,02 | 0,18 | 8,55E-01 | 9,23E-01 |
| MYLK4 | 93,16 | 0,01 | 0,05 | 0,18 | 8,55E-01 | 9,23E-01 |
| BCAR3 | 77,00 | -0,01 | 0,08 | -0,18 | 8,55E-01 | 9,23E-01 |
| TNFRSF13B | 79,17 | 0,01 | 0,08 | 0,18 | 8,54E-01 | 9,23E-01 |
| ZCCHC17 | 632,74 | 0,00 | 0,02 | 0,18 | 8,55E-01 | 9,23E-01 |
| LTBP1 | 517,97 | 0,01 | 0,06 | 0,18 | 8,55E-01 | 9,23E-01 |
| HMCES | 897,45 | 0,00 | 0,02 | -0,18 | 8,55E-01 | 9,23E-01 |
| EFEMP2 | 96,37 | 0,01 | 0,04 | 0,18 | 8,56E-01 | 9,23E-01 |
| LRRC42 | 114,81 | 0,01 | 0,03 | 0,18 | 8,56E-01 | 9,23E-01 |
| C19orf47 | 84,18 | 0,01 | 0,06 | 0,18 | 8,55E-01 | 9,23E-01 |
| SNAPC4 | 266,36 | 0,01 | 0,05 | 0,18 | 8,56E-01 | 9,23E-01 |
| DYNLRB1 | 390,20 | 0,00 | 0,02 | 0,18 | 8,56E-01 | 9,23E-01 |
| FGD6 | 368,16 | 0,01 | 0,04 | 0,18 | 8,56E-01 | 9,24E-01 |
| RGP1 | 817,00 | 0,01 | 0,03 | 0,18 | 8,57E-01 | 9,24E-01 |
| SLC35C2 | 1654,18 | 0,00 | 0,02 | 0,18 | 8,58E-01 | 9,25E-01 |
| EEF2K | 524,58 | 0,01 | 0,04 | 0,18 | 8,58E-01 | 9,25E-01 |
| CELSR2 | 87,34 | 0,01 | 0,08 | 0,18 | 8,58E-01 | 9,25E-01 |
| COG7 | 372,80 | 0,00 | 0,02 | 0,18 | 8,58E-01 | 9,25E-01 |
| SNRPB2 | 653,24 | 0,01 | 0,04 | 0,18 | 8,58E-01 | 9,25E-01 |
| VWF | 138,78 | 0,02 | 0,10 | 0,18 | 8,59E-01 | 9,25E-01 |
| RBM12 | 3554,61 | 0,00 | 0,03 | 0,18 | 8,58E-01 | 9,25E-01 |
| SLC27A3 | 594,08 | 0,01 | 0,03 | 0,18 | 8,58E-01 | 9,25E-01 |
| RNF44 | 865,59 | 0,01 | 0,05 | 0,18 | 8,59E-01 | 9,25E-01 |
| YTHDF1 | 806,55 | 0,00 | 0,01 | -0,18 | 8,59E-01 | 9,26E-01 |
| SPARC | 774,21 | 0,01 | 0,07 | 0,18 | 8,59E-01 | 9,26E-01 |
| STON1 | 85,74 | -0,01 | 0,05 | -0,18 | 8,59E-01 | 9,26E-01 |
| SNRPB | 662,86 | 0,00 | 0,02 | 0,18 | 8,59E-01 | 9,26E-01 |

| | | | | | | |
|---|---|---|---|---|---|---|
| PPARD | 465,93 | -0,01 | 0,04 | -0,18 | 8,59E-01 | 9,26E-01 |
| POP1 | 277,99 | 0,00 | 0,02 | -0,18 | 8,59E-01 | 9,26E-01 |
| AAMP | 916,68 | 0,00 | 0,02 | -0,18 | 8,59E-01 | 9,26E-01 |
| TNPO1 | 7928,04 | 0,01 | 0,03 | 0,18 | 8,59E-01 | 9,26E-01 |
| MAP4K2 | 777,02 | -0,01 | 0,04 | -0,18 | 8,60E-01 | 9,26E-01 |
| EFR3A | 3635,92 | 0,00 | 0,03 | 0,18 | 8,60E-01 | 9,26E-01 |
| TMEM115 | 323,22 | 0,00 | 0,02 | -0,18 | 8,60E-01 | 9,26E-01 |
| SERGEF | 589,62 | 0,00 | 0,02 | -0,18 | 8,60E-01 | 9,26E-01 |
| SLC30A5 | 1370,22 | 0,00 | 0,02 | 0,18 | 8,60E-01 | 9,26E-01 |
| AMPD3 | 1866,36 | -0,01 | 0,03 | -0,18 | 8,60E-01 | 9,26E-01 |
| PPHLN1 | 2181,68 | 0,00 | 0,02 | -0,18 | 8,60E-01 | 9,26E-01 |
| CRELD1 | 340,17 | 0,00 | 0,02 | 0,18 | 8,61E-01 | 9,26E-01 |
| DHFR | 440,85 | 0,00 | 0,02 | -0,17 | 8,62E-01 | 9,27E-01 |
| TTC39A | 73,89 | -0,02 | 0,10 | -0,17 | 8,62E-01 | 9,27E-01 |
| ZNF827 | 302,66 | 0,01 | 0,04 | 0,17 | 8,62E-01 | 9,27E-01 |
| ANKRD37 | 77,60 | 0,01 | 0,04 | 0,17 | 8,62E-01 | 9,28E-01 |
| MICU2 | 1479,29 | -0,01 | 0,03 | -0,17 | 8,63E-01 | 9,28E-01 |
| MAP3K1 | 12204,49 | 0,00 | 0,02 | -0,17 | 8,63E-01 | 9,28E-01 |
| GALNT1 | 1804,55 | 0,01 | 0,04 | 0,17 | 8,63E-01 | 9,28E-01 |
| ZNF607 | 206,32 | -0,01 | 0,03 | -0,17 | 8,63E-01 | 9,28E-01 |
| GARNL3 | 152,23 | -0,01 | 0,06 | -0,17 | 8,63E-01 | 9,28E-01 |
| MIER2 | 173,40 | -0,01 | 0,06 | -0,17 | 8,63E-01 | 9,28E-01 |
| ZBTB48 | 691,98 | 0,00 | 0,02 | 0,17 | 8,63E-01 | 9,28E-01 |
| ACBD5 | 1420,03 | 0,00 | 0,02 | -0,17 | 8,64E-01 | 9,28E-01 |
| STK11IP | 315,02 | 0,01 | 0,04 | 0,17 | 8,64E-01 | 9,28E-01 |
| SRF | 614,44 | 0,00 | 0,03 | -0,17 | 8,64E-01 | 9,28E-01 |
| UBE2B | 2566,20 | 0,01 | 0,03 | 0,17 | 8,64E-01 | 9,28E-01 |
| CYP1B1 | 944,10 | -0,01 | 0,05 | -0,17 | 8,64E-01 | 9,28E-01 |
| CNNM4 | 135,67 | -0,01 | 0,05 | -0,17 | 8,64E-01 | 9,28E-01 |
| CHST12 | 963,16 | -0,01 | 0,06 | -0,17 | 8,65E-01 | 9,29E-01 |
| PPP1R16B | 1389,08 | 0,01 | 0,03 | 0,17 | 8,65E-01 | 9,29E-01 |
| DCAF15 | 350,79 | 0,01 | 0,04 | 0,17 | 8,65E-01 | 9,29E-01 |
| LTA4H | 7015,08 | 0,01 | 0,04 | 0,17 | 8,66E-01 | 9,30E-01 |
| TBC1D17 | 453,38 | -0,01 | 0,04 | -0,17 | 8,66E-01 | 9,30E-01 |
| PPP4R3B | 5381,05 | 0,00 | 0,03 | 0,17 | 8,66E-01 | 9,30E-01 |
| HNRNPUL1 | 2582,04 | -0,01 | 0,04 | -0,17 | 8,66E-01 | 9,30E-01 |
| DYNC2I2 | 98,33 | 0,01 | 0,04 | 0,17 | 8,66E-01 | 9,30E-01 |
| CPNE3 | 2851,44 | 0,00 | 0,02 | -0,17 | 8,66E-01 | 9,30E-01 |
| SPRYD3 | 379,21 | -0,01 | 0,03 | -0,17 | 8,66E-01 | 9,30E-01 |
| MAP1A | 117,81 | -0,01 | 0,07 | -0,17 | 8,67E-01 | 9,30E-01 |
| ARRB1 | 1539,56 | 0,01 | 0,04 | 0,17 | 8,67E-01 | 9,30E-01 |
| RTCB | 1032,92 | 0,00 | 0,02 | -0,17 | 8,67E-01 | 9,30E-01 |
| DPH3 | 503,64 | 0,01 | 0,04 | 0,17 | 8,67E-01 | 9,30E-01 |
| TTI2 | 453,10 | 0,00 | 0,01 | 0,17 | 8,67E-01 | 9,30E-01 |
| SLC35G1 | 178,29 | -0,01 | 0,03 | -0,17 | 8,67E-01 | 9,30E-01 |
| ACAD10 | 744,56 | 0,00 | 0,03 | 0,17 | 8,68E-01 | 9,31E-01 |

| | | | | | | |
|---|---|---|---|---|---|---|
| ZKSCAN3 | 417,17 | 0,00 | 0,02 | -0,17 | 8,68E-01 | 9,31E-01 |
| CANX | 7777,09 | 0,00 | 0,02 | 0,17 | 8,68E-01 | 9,31E-01 |
| TULP4 | 1309,14 | 0,00 | 0,02 | 0,17 | 8,69E-01 | 9,32E-01 |
| ALDH2 | 486,63 | -0,01 | 0,04 | -0,16 | 8,69E-01 | 9,32E-01 |
| PCM1 | 7475,25 | 0,00 | 0,02 | -0,16 | 8,69E-01 | 9,32E-01 |
| CCNL2 | 4122,36 | 0,00 | 0,02 | -0,16 | 8,69E-01 | 9,32E-01 |
| CRIM1 | 312,14 | 0,01 | 0,03 | 0,16 | 8,69E-01 | 9,32E-01 |
| PLCD1 | 371,42 | 0,00 | 0,02 | -0,16 | 8,69E-01 | 9,32E-01 |
| CLEC1B | 114,11 | 0,01 | 0,06 | 0,16 | 8,69E-01 | 9,32E-01 |
| DUSP5 | 363,81 | 0,01 | 0,03 | 0,16 | 8,69E-01 | 9,32E-01 |
| DNTTIP2 | 922,05 | -0,01 | 0,05 | -0,16 | 8,70E-01 | 9,32E-01 |
| TGIF2 | 375,68 | 0,00 | 0,03 | -0,16 | 8,70E-01 | 9,32E-01 |
| BAG5 | 1517,28 | 0,00 | 0,02 | 0,16 | 8,70E-01 | 9,32E-01 |
| CHD3 | 5030,42 | 0,00 | 0,03 | -0,16 | 8,70E-01 | 9,32E-01 |
| ESR1 | 93,18 | 0,01 | 0,08 | 0,16 | 8,70E-01 | 9,32E-01 |
| DLG2 | 88,95 | 0,01 | 0,08 | 0,16 | 8,71E-01 | 9,32E-01 |
| SLC35B1 | 498,01 | 0,00 | 0,01 | -0,16 | 8,71E-01 | 9,32E-01 |
| FBXL18 | 114,22 | 0,01 | 0,07 | 0,16 | 8,71E-01 | 9,32E-01 |
| TMEM60 | 335,30 | 0,00 | 0,03 | -0,16 | 8,71E-01 | 9,32E-01 |
| SCD5 | 78,28 | 0,01 | 0,07 | 0,16 | 8,71E-01 | 9,32E-01 |
| POMT2 | 133,89 | -0,01 | 0,05 | -0,16 | 8,71E-01 | 9,32E-01 |
| CLCN3 | 2691,77 | 0,00 | 0,02 | 0,16 | 8,71E-01 | 9,32E-01 |
| P4HA1 | 1016,39 | 0,00 | 0,03 | -0,16 | 8,71E-01 | 9,33E-01 |
| ABLIM2 | 76,96 | -0,01 | 0,08 | -0,16 | 8,72E-01 | 9,33E-01 |
| HPS5 | 1078,10 | 0,00 | 0,02 | 0,16 | 8,72E-01 | 9,33E-01 |
| PLCXD2 | 1012,07 | 0,00 | 0,03 | 0,16 | 8,72E-01 | 9,33E-01 |
| ENPP2 | 104,61 | 0,01 | 0,06 | 0,16 | 8,73E-01 | 9,34E-01 |
| PARP1 | 2350,19 | 0,00 | 0,02 | -0,16 | 8,73E-01 | 9,34E-01 |
| MAD2L1BP | 362,96 | 0,00 | 0,02 | -0,16 | 8,73E-01 | 9,34E-01 |
| ZRANB1 | 2527,33 | 0,00 | 0,03 | -0,16 | 8,73E-01 | 9,34E-01 |
| GARS1 | 762,96 | 0,00 | 0,02 | -0,16 | 8,73E-01 | 9,34E-01 |
| SIRT6 | 187,59 | 0,00 | 0,03 | 0,16 | 8,73E-01 | 9,34E-01 |
| LDLRAD3 | 88,68 | -0,01 | 0,06 | -0,16 | 8,73E-01 | 9,34E-01 |
| MED18 | 645,76 | 0,00 | 0,02 | -0,16 | 8,73E-01 | 9,34E-01 |
| GTF3C4 | 647,66 | 0,00 | 0,01 | 0,16 | 8,74E-01 | 9,34E-01 |
| NEB | 313,79 | -0,01 | 0,07 | -0,16 | 8,74E-01 | 9,34E-01 |
| IGBP1 | 909,44 | -0,01 | 0,03 | -0,16 | 8,74E-01 | 9,34E-01 |
| PDZD2 | 76,39 | 0,02 | 0,11 | 0,16 | 8,74E-01 | 9,34E-01 |
| ZFP36L2 | 3895,14 | 0,00 | 0,03 | 0,16 | 8,75E-01 | 9,34E-01 |
| CC2D1B | 972,07 | 0,00 | 0,03 | -0,16 | 8,75E-01 | 9,34E-01 |
| PLCL2 | 5184,19 | 0,00 | 0,02 | -0,16 | 8,74E-01 | 9,34E-01 |
| MZF1 | 637,02 | 0,00 | 0,03 | -0,16 | 8,75E-01 | 9,34E-01 |
| EPN1 | 808,07 | -0,01 | 0,05 | -0,16 | 8,75E-01 | 9,34E-01 |
| MLLT1 | 315,81 | -0,01 | 0,06 | -0,16 | 8,75E-01 | 9,35E-01 |
| FANCC | 296,59 | -0,01 | 0,05 | -0,16 | 8,75E-01 | 9,35E-01 |
| PPBP | 4305,82 | -0,01 | 0,07 | -0,16 | 8,76E-01 | 9,35E-01 |

| | | | | | | |
|---|---|---|---|---|---|---|
| NFATC1 | 504,42 | -0,01 | 0,05 | -0,16 | 8,76E-01 | 9,35E-01 |
| CRYZ | 314,13 | 0,01 | 0,05 | 0,16 | 8,76E-01 | 9,35E-01 |
| RMND5A | 7655,76 | 0,00 | 0,03 | 0,16 | 8,76E-01 | 9,35E-01 |
| TICAM2 | 131,35 | 0,00 | 0,03 | -0,16 | 8,76E-01 | 9,36E-01 |
| MICALL2 | 73,46 | 0,01 | 0,09 | 0,16 | 8,77E-01 | 9,36E-01 |
| DONSON | 159,44 | 0,00 | 0,02 | -0,16 | 8,77E-01 | 9,36E-01 |
| DUSP2 | 323,18 | -0,01 | 0,06 | -0,15 | 8,77E-01 | 9,36E-01 |
| TBC1D10A | 482,85 | 0,01 | 0,04 | 0,15 | 8,78E-01 | 9,36E-01 |
| OPHN1 | 106,85 | -0,01 | 0,05 | -0,15 | 8,78E-01 | 9,37E-01 |
| MSANTD7 | 923,91 | 0,00 | 0,02 | 0,15 | 8,78E-01 | 9,37E-01 |
| PRR11 | 126,02 | 0,01 | 0,04 | 0,15 | 8,78E-01 | 9,37E-01 |
| SLC24A1 | 158,43 | 0,01 | 0,04 | 0,15 | 8,79E-01 | 9,37E-01 |
| LEPROTL1 | 3130,52 | 0,00 | 0,03 | 0,15 | 8,79E-01 | 9,37E-01 |
| SRSF1 | 1893,80 | 0,00 | 0,03 | -0,15 | 8,80E-01 | 9,38E-01 |
| B3GNT2 | 870,11 | 0,00 | 0,03 | -0,15 | 8,80E-01 | 9,38E-01 |
| FAIM | 89,79 | 0,01 | 0,04 | 0,15 | 8,81E-01 | 9,39E-01 |
| GEMIN5 | 575,25 | 0,00 | 0,02 | 0,15 | 8,81E-01 | 9,39E-01 |
| SSB | 815,26 | 0,01 | 0,05 | 0,15 | 8,81E-01 | 9,39E-01 |
| TAF13 | 305,66 | 0,01 | 0,04 | 0,15 | 8,81E-01 | 9,39E-01 |
| EFHC1 | 426,78 | 0,01 | 0,03 | 0,15 | 8,81E-01 | 9,39E-01 |
| MCM4 | 259,99 | 0,00 | 0,03 | -0,15 | 8,81E-01 | 9,39E-01 |
| ZNF24 | 5456,66 | 0,00 | 0,03 | -0,15 | 8,81E-01 | 9,39E-01 |
| CALU | 835,50 | 0,00 | 0,02 | 0,15 | 8,82E-01 | 9,39E-01 |
| TMSB4X | 74251,96 | 0,01 | 0,04 | 0,15 | 8,82E-01 | 9,40E-01 |
| CDC73 | 3779,68 | 0,00 | 0,03 | -0,15 | 8,82E-01 | 9,40E-01 |
| PTPRM | 428,27 | -0,01 | 0,06 | -0,15 | 8,82E-01 | 9,40E-01 |
| PHEX | 89,33 | 0,01 | 0,07 | 0,15 | 8,83E-01 | 9,40E-01 |
| XBP1 | 2550,19 | 0,00 | 0,02 | -0,15 | 8,83E-01 | 9,40E-01 |
| CCDC9B | 116,62 | -0,01 | 0,05 | -0,15 | 8,82E-01 | 9,40E-01 |
| CAMTA2 | 343,96 | 0,01 | 0,07 | 0,15 | 8,82E-01 | 9,40E-01 |
| PRR7 | 186,68 | -0,01 | 0,03 | -0,15 | 8,83E-01 | 9,40E-01 |
| TSC22D4 | 721,41 | -0,01 | 0,06 | -0,15 | 8,83E-01 | 9,40E-01 |
| CDC42EP4 | 115,17 | -0,01 | 0,06 | -0,15 | 8,83E-01 | 9,40E-01 |
| DAPK1 | 1673,83 | -0,01 | 0,04 | -0,15 | 8,83E-01 | 9,40E-01 |
| ALS2 | 1012,45 | 0,00 | 0,02 | -0,15 | 8,84E-01 | 9,40E-01 |
| UQCRB | 903,92 | 0,01 | 0,08 | 0,15 | 8,84E-01 | 9,40E-01 |
| HGD | 103,81 | 0,01 | 0,04 | 0,15 | 8,84E-01 | 9,40E-01 |
| ARAP2 | 5911,65 | 0,01 | 0,04 | 0,15 | 8,84E-01 | 9,41E-01 |
| FICD | 266,16 | 0,00 | 0,02 | 0,15 | 8,84E-01 | 9,41E-01 |
| HYOU1 | 575,05 | -0,01 | 0,04 | -0,15 | 8,84E-01 | 9,41E-01 |
| CAMSAP2 | 222,03 | 0,01 | 0,04 | 0,15 | 8,84E-01 | 9,41E-01 |
| KYAT1 | 151,44 | -0,01 | 0,04 | -0,14 | 8,85E-01 | 9,41E-01 |
| ATP6V1E2 | 199,00 | 0,00 | 0,03 | -0,14 | 8,85E-01 | 9,41E-01 |
| P3H4 | 76,91 | 0,01 | 0,05 | 0,14 | 8,86E-01 | 9,41E-01 |
| RRN3 | 1063,84 | 0,00 | 0,02 | 0,14 | 8,86E-01 | 9,41E-01 |
| ERMN | 434,05 | 0,00 | 0,03 | -0,14 | 8,85E-01 | 9,41E-01 |

| | | | | | | |
|---|---|---|---|---|---|---|
| VIL1 | 86,96 | 0,01 | 0,08 | 0,14 | 8,86E-01 | 9,41E-01 |
| CST3 | 1683,14 | 0,01 | 0,04 | 0,14 | 8,86E-01 | 9,41E-01 |
| GPD1L | 797,12 | 0,00 | 0,02 | -0,14 | 8,85E-01 | 9,41E-01 |
| ATP2A3 | 3697,68 | -0,01 | 0,04 | -0,14 | 8,86E-01 | 9,41E-01 |
| SLC9A6 | 914,52 | 0,00 | 0,02 | 0,14 | 8,86E-01 | 9,41E-01 |
| FLVCR2 | 352,13 | -0,01 | 0,04 | -0,14 | 8,86E-01 | 9,41E-01 |
| HPS3 | 2167,44 | 0,00 | 0,02 | -0,14 | 8,86E-01 | 9,41E-01 |
| ANO6 | 2922,74 | 0,00 | 0,02 | 0,14 | 8,87E-01 | 9,42E-01 |
| MTNAP1 | 475,06 | 0,00 | 0,02 | -0,14 | 8,87E-01 | 9,42E-01 |
| BCLAF1 | 9607,57 | 0,00 | 0,03 | 0,14 | 8,88E-01 | 9,42E-01 |
| FCRL6 | 1072,90 | 0,01 | 0,06 | 0,14 | 8,88E-01 | 9,42E-01 |
| LAX1 | 743,54 | 0,00 | 0,03 | 0,14 | 8,88E-01 | 9,42E-01 |
| MAP3K14 | 607,60 | 0,01 | 0,04 | 0,14 | 8,87E-01 | 9,42E-01 |
| TLR10 | 717,54 | 0,01 | 0,04 | 0,14 | 8,88E-01 | 9,42E-01 |
| TAF3 | 370,00 | 0,00 | 0,02 | 0,14 | 8,88E-01 | 9,42E-01 |
| CHMP1A | 681,63 | 0,00 | 0,03 | -0,14 | 8,88E-01 | 9,42E-01 |
| GTF2E1 | 754,07 | 0,00 | 0,03 | 0,14 | 8,89E-01 | 9,43E-01 |
| ABHD11 | 128,72 | 0,00 | 0,03 | 0,14 | 8,89E-01 | 9,43E-01 |
| SAMTOR | 1002,20 | 0,00 | 0,03 | -0,14 | 8,89E-01 | 9,43E-01 |
| C8orf58 | 89,70 | 0,00 | 0,03 | -0,14 | 8,89E-01 | 9,43E-01 |
| TEPSIN | 632,54 | -0,01 | 0,04 | -0,14 | 8,89E-01 | 9,44E-01 |
| ATP8A2 | 94,52 | 0,01 | 0,09 | 0,14 | 8,90E-01 | 9,44E-01 |
| TDRD9 | 244,00 | -0,01 | 0,07 | -0,14 | 8,90E-01 | 9,44E-01 |
| TENT4A | 868,97 | 0,00 | 0,03 | -0,14 | 8,90E-01 | 9,44E-01 |
| CD37 | 2008,93 | -0,01 | 0,05 | -0,14 | 8,91E-01 | 9,44E-01 |
| ITPKB | 3459,33 | 0,00 | 0,03 | 0,14 | 8,91E-01 | 9,44E-01 |
| KLHL7 | 695,37 | 0,00 | 0,02 | -0,14 | 8,91E-01 | 9,44E-01 |
| TNFRSF10D | 325,44 | 0,00 | 0,02 | 0,14 | 8,91E-01 | 9,45E-01 |
| CCDC85C | 105,22 | 0,01 | 0,08 | 0,14 | 8,91E-01 | 9,45E-01 |
| UNC5CL | 94,59 | 0,01 | 0,04 | 0,14 | 8,91E-01 | 9,45E-01 |
| SMAP1 | 839,58 | 0,00 | 0,02 | 0,14 | 8,92E-01 | 9,45E-01 |
| LDLRAD4 | 1718,55 | 0,00 | 0,02 | 0,14 | 8,92E-01 | 9,45E-01 |
| MFAP3L | 890,51 | 0,01 | 0,05 | 0,13 | 8,93E-01 | 9,45E-01 |
| KRCC1 | 727,46 | 0,01 | 0,05 | 0,13 | 8,94E-01 | 9,45E-01 |
| ENTPD7 | 331,80 | 0,00 | 0,02 | -0,13 | 8,93E-01 | 9,45E-01 |
| HMGXB4 | 465,62 | 0,00 | 0,01 | -0,13 | 8,94E-01 | 9,45E-01 |
| CLSTN3 | 547,85 | 0,01 | 0,05 | 0,13 | 8,94E-01 | 9,45E-01 |
| TRDMT1 | 1186,79 | 0,00 | 0,02 | 0,14 | 8,93E-01 | 9,45E-01 |
| ZNF814 | 711,28 | 0,00 | 0,03 | 0,13 | 8,93E-01 | 9,45E-01 |
| NECTIN2 | 87,29 | 0,02 | 0,13 | 0,13 | 8,93E-01 | 9,45E-01 |
| WNT7A | 138,98 | 0,01 | 0,05 | 0,14 | 8,93E-01 | 9,45E-01 |
| MAFG | 285,29 | -0,01 | 0,05 | -0,13 | 8,93E-01 | 9,45E-01 |
| DUSP19 | 106,34 | 0,00 | 0,03 | 0,13 | 8,93E-01 | 9,45E-01 |
| ENSG000002 | 181,78 | -0,01 | 0,05 | -0,13 | 8,93E-01 | 9,45E-01 |
| POLR3A | 835,29 | 0,00 | 0,02 | 0,14 | 8,92E-01 | 9,45E-01 |
| DTX1 | 89,62 | 0,01 | 0,07 | 0,13 | 8,93E-01 | 9,45E-01 |

| | | | | | | |
|---|---|---|---|---|---|---|
| MAP3K13 | 171,83 | 0,01 | 0,04 | 0,13 | 8,93E-01 | 9,45E-01 |
| RAD51B | 711,00 | 0,00 | 0,03 | 0,13 | 8,94E-01 | 9,45E-01 |
| LTO1 | 946,52 | 0,00 | 0,02 | -0,14 | 8,92E-01 | 9,45E-01 |
| SCFD1 | 1987,04 | 0,00 | 0,03 | 0,13 | 8,93E-01 | 9,45E-01 |
| PRDM15 | 469,67 | 0,00 | 0,03 | -0,13 | 8,94E-01 | 9,46E-01 |
| CLSTN1 | 620,96 | -0,01 | 0,05 | -0,13 | 8,94E-01 | 9,46E-01 |
| GNB1L | 153,34 | -0,01 | 0,04 | -0,13 | 8,94E-01 | 9,46E-01 |
| NINJ2 | 604,13 | -0,01 | 0,05 | -0,13 | 8,94E-01 | 9,46E-01 |
| VLDLR | 88,69 | 0,01 | 0,08 | 0,13 | 8,95E-01 | 9,46E-01 |
| MYOF | 778,11 | -0,01 | 0,05 | -0,13 | 8,95E-01 | 9,46E-01 |
| ARHGEF1 | 4070,79 | -0,01 | 0,05 | -0,13 | 8,95E-01 | 9,46E-01 |
| TAF7 | 1292,38 | -0,01 | 0,05 | -0,13 | 8,95E-01 | 9,46E-01 |
| SRRT | 489,99 | 0,01 | 0,04 | 0,13 | 8,95E-01 | 9,46E-01 |
| TMEM184C | 1271,78 | 0,00 | 0,02 | -0,13 | 8,95E-01 | 9,46E-01 |
| MBTD1 | 2241,78 | 0,00 | 0,02 | 0,13 | 8,96E-01 | 9,47E-01 |
| MAT2B | 5355,38 | 0,00 | 0,03 | 0,13 | 8,96E-01 | 9,47E-01 |
| AHI1 | 370,74 | 0,01 | 0,05 | 0,13 | 8,96E-01 | 9,47E-01 |
| RECQL5 | 514,13 | 0,00 | 0,03 | 0,13 | 8,96E-01 | 9,47E-01 |
| TBRG1 | 2399,49 | 0,00 | 0,01 | 0,13 | 8,96E-01 | 9,47E-01 |
| RRM1 | 631,40 | 0,00 | 0,02 | 0,13 | 8,97E-01 | 9,47E-01 |
| ESPN | 200,49 | 0,01 | 0,11 | 0,13 | 8,96E-01 | 9,47E-01 |
| PEX11A | 77,38 | 0,01 | 0,04 | 0,13 | 8,97E-01 | 9,47E-01 |
| DOP1A | 2686,02 | 0,00 | 0,02 | -0,13 | 8,97E-01 | 9,47E-01 |
| BAX | 808,82 | 0,00 | 0,02 | -0,13 | 8,97E-01 | 9,47E-01 |
| ECHS1 | 503,76 | 0,00 | 0,02 | 0,13 | 8,97E-01 | 9,48E-01 |
| GOLPH3L | 460,84 | 0,00 | 0,02 | -0,13 | 8,98E-01 | 9,48E-01 |
| TM9SF4 | 809,37 | 0,00 | 0,04 | 0,13 | 8,98E-01 | 9,48E-01 |
| COQ7 | 436,13 | 0,00 | 0,02 | 0,13 | 8,98E-01 | 9,48E-01 |
| ETS1 | 11662,79 | 0,00 | 0,03 | 0,13 | 8,98E-01 | 9,48E-01 |
| LRCH3 | 2254,13 | 0,00 | 0,02 | 0,13 | 8,99E-01 | 9,48E-01 |
| BORCS6 | 487,47 | 0,00 | 0,02 | -0,13 | 8,98E-01 | 9,48E-01 |
| PTGS1 | 1174,13 | -0,01 | 0,05 | -0,13 | 8,99E-01 | 9,48E-01 |
| ADD3 | 7423,49 | 0,00 | 0,03 | 0,13 | 8,98E-01 | 9,48E-01 |
| TOLLIP | 864,89 | 0,00 | 0,04 | -0,13 | 8,99E-01 | 9,48E-01 |
| NUMA1 | 5994,71 | 0,00 | 0,02 | 0,13 | 8,99E-01 | 9,48E-01 |
| UBE2V1 | 713,88 | 0,00 | 0,02 | -0,13 | 8,99E-01 | 9,49E-01 |
| LAMA2 | 121,78 | -0,01 | 0,06 | -0,13 | 9,00E-01 | 9,49E-01 |
| FOXRED1 | 287,90 | 0,00 | 0,03 | -0,13 | 9,00E-01 | 9,49E-01 |
| GIGYF1 | 1028,38 | 0,01 | 0,06 | 0,13 | 9,00E-01 | 9,49E-01 |
| DIP2C | 310,20 | 0,01 | 0,05 | 0,13 | 9,00E-01 | 9,49E-01 |
| LRMDA | 227,91 | -0,01 | 0,07 | -0,13 | 9,00E-01 | 9,49E-01 |
| CHTF8 | 1086,31 | 0,00 | 0,02 | 0,12 | 9,01E-01 | 9,50E-01 |
| OGFOD1 | 460,51 | 0,00 | 0,02 | -0,12 | 9,01E-01 | 9,50E-01 |
| RNGTT | 1475,27 | 0,00 | 0,02 | 0,12 | 9,01E-01 | 9,50E-01 |
| MRPL51 | 456,54 | 0,01 | 0,04 | 0,12 | 9,02E-01 | 9,50E-01 |
| SNX29 | 2219,18 | 0,00 | 0,03 | -0,12 | 9,02E-01 | 9,50E-01 |

| | | | | | | |
|---|---|---|---|---|---|---|
| TMEM187 | 108,25 | 0,00 | 0,03 | -0,12 | 9,02E-01 | 9,50E-01 |
| PSMA4 | 1292,06 | -0,01 | 0,05 | -0,12 | 9,02E-01 | 9,50E-01 |
| CLPP | 250,95 | 0,00 | 0,02 | 0,12 | 9,02E-01 | 9,50E-01 |
| AKAP9 | 5024,52 | 0,00 | 0,03 | 0,12 | 9,02E-01 | 9,50E-01 |
| POM121 | 727,30 | 0,00 | 0,04 | 0,12 | 9,02E-01 | 9,50E-01 |
| GSTM3 | 97,90 | -0,01 | 0,10 | -0,12 | 9,02E-01 | 9,50E-01 |
| SMARCD3 | 385,27 | 0,00 | 0,04 | 0,12 | 9,02E-01 | 9,50E-01 |
| C19orf25 | 845,25 | 0,00 | 0,03 | 0,12 | 9,02E-01 | 9,50E-01 |
| BBS2 | 1015,17 | 0,00 | 0,03 | -0,12 | 9,03E-01 | 9,50E-01 |
| CBFA2T3 | 329,99 | 0,01 | 0,06 | 0,12 | 9,03E-01 | 9,50E-01 |
| RAD9A | 173,56 | 0,01 | 0,04 | 0,12 | 9,03E-01 | 9,50E-01 |
| CEP89 | 240,77 | -0,01 | 0,04 | -0,12 | 9,03E-01 | 9,51E-01 |
| POLR1B | 760,64 | 0,00 | 0,02 | 0,12 | 9,03E-01 | 9,51E-01 |
| GAA | 984,80 | -0,01 | 0,05 | -0,12 | 9,03E-01 | 9,51E-01 |
| TPST2 | 2116,02 | 0,00 | 0,02 | 0,12 | 9,03E-01 | 9,51E-01 |
| INPPL1 | 929,78 | -0,01 | 0,05 | -0,12 | 9,03E-01 | 9,51E-01 |
| ARHGAP17 | 1193,17 | 0,00 | 0,04 | 0,12 | 9,04E-01 | 9,51E-01 |
| TM7SF3 | 1079,47 | 0,00 | 0,02 | 0,12 | 9,04E-01 | 9,51E-01 |
| C1orf74 | 105,65 | 0,00 | 0,03 | 0,12 | 9,05E-01 | 9,51E-01 |
| PIM3 | 1139,43 | 0,00 | 0,03 | 0,12 | 9,05E-01 | 9,51E-01 |
| TMEM39B | 113,03 | 0,01 | 0,05 | 0,12 | 9,05E-01 | 9,51E-01 |
| CLEC5A | 161,69 | -0,01 | 0,06 | -0,12 | 9,05E-01 | 9,51E-01 |
| PNISR | 9858,02 | 0,00 | 0,03 | -0,12 | 9,05E-01 | 9,51E-01 |
| ZGPAT | 594,54 | 0,00 | 0,02 | -0,12 | 9,05E-01 | 9,51E-01 |
| PPIL4 | 1165,64 | 0,00 | 0,03 | 0,12 | 9,05E-01 | 9,52E-01 |
| DYM | 1937,13 | 0,00 | 0,02 | 0,12 | 9,06E-01 | 9,52E-01 |
| DRG1 | 699,03 | 0,00 | 0,02 | 0,12 | 9,06E-01 | 9,52E-01 |
| PREP | 817,56 | 0,00 | 0,02 | -0,12 | 9,06E-01 | 9,52E-01 |
| ADAMTS17 | 181,01 | 0,01 | 0,06 | 0,12 | 9,06E-01 | 9,52E-01 |
| GPRASP1 | 1230,40 | 0,00 | 0,03 | 0,12 | 9,06E-01 | 9,52E-01 |
| POU2AF1 | 365,89 | 0,01 | 0,06 | 0,12 | 9,06E-01 | 9,52E-01 |
| COPS6 | 406,96 | 0,00 | 0,02 | -0,12 | 9,07E-01 | 9,53E-01 |
| REV1 | 1860,43 | 0,00 | 0,02 | 0,12 | 9,07E-01 | 9,53E-01 |
| ASB6 | 340,42 | 0,00 | 0,04 | -0,12 | 9,07E-01 | 9,53E-01 |
| PPP1R9A | 124,73 | 0,01 | 0,07 | 0,12 | 9,07E-01 | 9,53E-01 |
| CPVL | 2857,32 | 0,00 | 0,04 | 0,12 | 9,08E-01 | 9,53E-01 |
| SEC63 | 1584,26 | 0,00 | 0,02 | 0,12 | 9,07E-01 | 9,53E-01 |
| STK39 | 923,81 | 0,00 | 0,03 | -0,12 | 9,08E-01 | 9,53E-01 |
| CD99L2 | 501,27 | 0,00 | 0,02 | 0,12 | 9,07E-01 | 9,53E-01 |
| KRBOX4 | 390,16 | 0,00 | 0,02 | -0,12 | 9,08E-01 | 9,53E-01 |
| TMEM170B | 3491,40 | 0,01 | 0,05 | 0,12 | 9,08E-01 | 9,53E-01 |
| MAP7D1 | 983,85 | -0,01 | 0,05 | -0,12 | 9,08E-01 | 9,53E-01 |
| VPS72 | 498,63 | 0,00 | 0,02 | 0,12 | 9,08E-01 | 9,53E-01 |
| RAB10 | 4130,74 | 0,00 | 0,02 | 0,12 | 9,08E-01 | 9,53E-01 |
| DGKZ | 2015,07 | -0,01 | 0,05 | -0,12 | 9,07E-01 | 9,53E-01 |
| NCLN | 447,22 | 0,01 | 0,05 | 0,12 | 9,08E-01 | 9,53E-01 |

| | | | | | | |
|---|---|---|---|---|---|---|
| ALDH3B1 | 533,96 | 0,00 | 0,04 | -0,11 | 9,08E-01 | 9,53E-01 |
| ANKRD42 | 401,56 | 0,00 | 0,02 | 0,11 | 9,09E-01 | 9,53E-01 |
| DGUOK | 487,24 | 0,00 | 0,02 | -0,11 | 9,09E-01 | 9,53E-01 |
| PIP4P1 | 943,11 | 0,00 | 0,01 | 0,11 | 9,09E-01 | 9,53E-01 |
| PI4KB | 1265,72 | 0,00 | 0,03 | -0,12 | 9,08E-01 | 9,53E-01 |
| NATD1 | 682,94 | -0,01 | 0,07 | -0,12 | 9,08E-01 | 9,53E-01 |
| LARP7 | 695,10 | 0,00 | 0,04 | -0,11 | 9,09E-01 | 9,53E-01 |
| TNFRSF14 | 2166,87 | 0,00 | 0,03 | -0,12 | 9,08E-01 | 9,53E-01 |
| ILRUN | 2011,37 | 0,00 | 0,02 | 0,12 | 9,08E-01 | 9,53E-01 |
| LONP2 | 3805,21 | 0,00 | 0,02 | 0,11 | 9,09E-01 | 9,53E-01 |
| PROCA1 | 114,73 | -0,01 | 0,05 | -0,11 | 9,09E-01 | 9,53E-01 |
| CCDC57 | 995,72 | 0,00 | 0,04 | 0,11 | 9,09E-01 | 9,53E-01 |
| TSC2 | 912,20 | 0,01 | 0,06 | 0,11 | 9,09E-01 | 9,53E-01 |
| INTS11 | 803,08 | 0,00 | 0,03 | 0,11 | 9,09E-01 | 9,53E-01 |
| CCRL2 | 148,39 | 0,00 | 0,03 | -0,11 | 9,10E-01 | 9,53E-01 |
| ANKRD52 | 580,47 | 0,01 | 0,04 | 0,11 | 9,10E-01 | 9,53E-01 |
| XRCC1 | 226,31 | 0,00 | 0,04 | 0,11 | 9,10E-01 | 9,53E-01 |
| SMIM27 | 187,25 | 0,00 | 0,03 | -0,11 | 9,10E-01 | 9,53E-01 |
| PDCD6IP | 4864,81 | 0,00 | 0,02 | -0,11 | 9,10E-01 | 9,53E-01 |
| GOLIM4 | 474,72 | 0,00 | 0,03 | 0,11 | 9,10E-01 | 9,53E-01 |
| PSMB1 | 1898,40 | 0,00 | 0,03 | 0,11 | 9,10E-01 | 9,53E-01 |
| TDRP | 109,63 | 0,01 | 0,07 | 0,11 | 9,11E-01 | 9,53E-01 |
| MICALL1 | 207,33 | 0,01 | 0,06 | 0,11 | 9,11E-01 | 9,53E-01 |
| MIB1 | 3675,77 | 0,00 | 0,03 | 0,11 | 9,11E-01 | 9,53E-01 |
| NPRL2 | 671,75 | 0,00 | 0,02 | 0,11 | 9,11E-01 | 9,53E-01 |
| GXYLT1 | 1053,47 | 0,00 | 0,03 | 0,11 | 9,11E-01 | 9,54E-01 |
| SOX5 | 71,69 | 0,01 | 0,09 | 0,11 | 9,12E-01 | 9,54E-01 |
| CEP104 | 854,28 | 0,00 | 0,03 | 0,11 | 9,12E-01 | 9,54E-01 |
| SLAMF6 | 1095,70 | 0,00 | 0,03 | 0,11 | 9,11E-01 | 9,54E-01 |
| ANKRD36B | 1318,37 | 0,00 | 0,04 | 0,11 | 9,11E-01 | 9,54E-01 |
| NCK2 | 2157,35 | 0,00 | 0,02 | 0,11 | 9,12E-01 | 9,54E-01 |
| CDC14B | 363,72 | 0,00 | 0,04 | 0,11 | 9,12E-01 | 9,54E-01 |
| IRGQ | 536,57 | 0,00 | 0,02 | -0,11 | 9,13E-01 | 9,55E-01 |
| ARB2A | 1436,18 | 0,00 | 0,03 | 0,11 | 9,13E-01 | 9,55E-01 |
| EIF2S3 | 5625,02 | 0,00 | 0,02 | -0,11 | 9,13E-01 | 9,55E-01 |
| ANAPC5 | 3651,73 | 0,00 | 0,01 | -0,11 | 9,13E-01 | 9,55E-01 |
| TRPC6 | 78,61 | -0,01 | 0,08 | -0,11 | 9,14E-01 | 9,55E-01 |
| ZFPL1 | 222,35 | 0,00 | 0,02 | -0,11 | 9,14E-01 | 9,55E-01 |
| USF2 | 804,93 | 0,00 | 0,04 | 0,11 | 9,14E-01 | 9,55E-01 |
| RMDN2 | 290,16 | 0,00 | 0,03 | 0,11 | 9,14E-01 | 9,55E-01 |
| VCAN | 27462,09 | 0,00 | 0,04 | -0,11 | 9,14E-01 | 9,55E-01 |
| ADCK1 | 155,27 | 0,01 | 0,05 | 0,11 | 9,14E-01 | 9,55E-01 |
| SH3BGRL2 | 1383,61 | -0,01 | 0,06 | -0,11 | 9,14E-01 | 9,55E-01 |
| RBM48 | 825,14 | 0,00 | 0,02 | -0,11 | 9,15E-01 | 9,56E-01 |
| CLCC1 | 962,85 | 0,00 | 0,02 | 0,11 | 9,15E-01 | 9,56E-01 |
| SRGAP3 | 245,03 | -0,01 | 0,07 | -0,11 | 9,15E-01 | 9,56E-01 |

| | | | | | | |
|---|---|---|---|---|---|---|
| SYNJ2 | 569,30 | 0,00 | 0,04 | -0,11 | 9,15E-01 | 9,56E-01 |
| ZNF425 | 83,91 | 0,00 | 0,03 | 0,11 | 9,15E-01 | 9,56E-01 |
| SLC39A11 | 642,25 | 0,00 | 0,03 | 0,11 | 9,15E-01 | 9,56E-01 |
| PDGFA | 131,77 | 0,01 | 0,07 | 0,11 | 9,15E-01 | 9,56E-01 |
| RNF6 | 1416,74 | 0,00 | 0,03 | -0,11 | 9,15E-01 | 9,56E-01 |
| NUP188 | 660,33 | 0,00 | 0,03 | 0,11 | 9,16E-01 | 9,56E-01 |
| LMNB2 | 402,09 | 0,00 | 0,02 | 0,11 | 9,16E-01 | 9,56E-01 |
| FBXL19 | 322,21 | 0,00 | 0,03 | -0,10 | 9,16E-01 | 9,57E-01 |
| SLC39A14 | 117,27 | 0,00 | 0,04 | 0,10 | 9,17E-01 | 9,57E-01 |
| CDC42EP3 | 3820,91 | 0,00 | 0,03 | -0,10 | 9,17E-01 | 9,57E-01 |
| GFI1 | 325,09 | 0,00 | 0,05 | 0,10 | 9,17E-01 | 9,57E-01 |
| BRMS1 | 737,95 | 0,00 | 0,02 | -0,10 | 9,17E-01 | 9,57E-01 |
| DCAF11 | 2945,54 | 0,00 | 0,02 | 0,10 | 9,17E-01 | 9,57E-01 |
| OAT | 998,35 | 0,00 | 0,03 | -0,10 | 9,17E-01 | 9,57E-01 |
| POLR3B | 490,36 | 0,00 | 0,02 | -0,10 | 9,18E-01 | 9,57E-01 |
| XAB2 | 647,50 | 0,00 | 0,03 | 0,10 | 9,18E-01 | 9,57E-01 |
| ZNF875 | 1036,47 | 0,00 | 0,03 | -0,10 | 9,18E-01 | 9,57E-01 |
| FEM1C | 1774,08 | 0,00 | 0,03 | 0,10 | 9,18E-01 | 9,57E-01 |
| RABEP1 | 2328,23 | 0,00 | 0,02 | -0,10 | 9,18E-01 | 9,57E-01 |
| SARDH | 107,96 | -0,01 | 0,07 | -0,10 | 9,18E-01 | 9,57E-01 |
| MESD | 1090,32 | 0,00 | 0,02 | -0,10 | 9,18E-01 | 9,57E-01 |
| ICA1 | 128,18 | 0,00 | 0,04 | 0,10 | 9,19E-01 | 9,57E-01 |
| CNOT6L | 7295,59 | 0,00 | 0,03 | 0,10 | 9,19E-01 | 9,57E-01 |
| ARAF | 490,33 | 0,00 | 0,04 | 0,10 | 9,19E-01 | 9,57E-01 |
| MCF2L | 288,69 | -0,01 | 0,07 | -0,10 | 9,19E-01 | 9,58E-01 |
| RELA | 606,51 | 0,00 | 0,04 | -0,10 | 9,19E-01 | 9,58E-01 |
| PRKCQ | 1519,31 | 0,00 | 0,03 | -0,10 | 9,20E-01 | 9,58E-01 |
| PTS | 450,31 | 0,00 | 0,02 | 0,10 | 9,20E-01 | 9,58E-01 |
| MAPK9 | 1015,12 | 0,00 | 0,02 | 0,10 | 9,20E-01 | 9,59E-01 |
| PCSK6 | 192,27 | 0,01 | 0,08 | 0,10 | 9,20E-01 | 9,59E-01 |
| MGME1 | 676,15 | 0,00 | 0,02 | -0,10 | 9,21E-01 | 9,59E-01 |
| ZNF134 | 860,65 | 0,00 | 0,02 | -0,10 | 9,22E-01 | 9,60E-01 |
| NEK7 | 5797,15 | 0,00 | 0,04 | -0,10 | 9,22E-01 | 9,60E-01 |
| PSEN2 | 168,90 | 0,00 | 0,03 | -0,10 | 9,22E-01 | 9,60E-01 |
| SLC35A4 | 239,84 | 0,00 | 0,04 | 0,10 | 9,22E-01 | 9,60E-01 |
| TBCE | 233,65 | 0,00 | 0,03 | -0,10 | 9,22E-01 | 9,60E-01 |
| RHBDF2 | 1050,51 | 0,00 | 0,05 | -0,10 | 9,22E-01 | 9,60E-01 |
| SFT2D2 | 4864,92 | 0,00 | 0,03 | -0,10 | 9,22E-01 | 9,60E-01 |
| H2AC14 | 266,03 | 0,01 | 0,07 | 0,10 | 9,22E-01 | 9,60E-01 |
| ZNF630 | 151,53 | 0,00 | 0,03 | -0,10 | 9,23E-01 | 9,60E-01 |
| TM2D1 | 1447,00 | 0,00 | 0,02 | -0,10 | 9,22E-01 | 9,60E-01 |
| ZBTB11 | 2271,73 | 0,00 | 0,03 | 0,10 | 9,23E-01 | 9,60E-01 |
| HSPA8 | 12647,69 | 0,00 | 0,03 | 0,10 | 9,23E-01 | 9,60E-01 |
| TSPAN9 | 104,81 | -0,01 | 0,09 | -0,10 | 9,23E-01 | 9,60E-01 |
| UCK2 | 379,88 | 0,00 | 0,03 | -0,10 | 9,23E-01 | 9,61E-01 |
| LRRC75A | 240,05 | 0,00 | 0,05 | -0,10 | 9,24E-01 | 9,61E-01 |

| | | | | | | |
|---|---|---|---|---|---|---|
| TCP11L2 | 10250,00 | 0,00 | 0,03 | 0,10 | 9,24E-01 | 9,61E-01 |
| TIMELESS | 214,69 | 0,00 | 0,04 | 0,10 | 9,24E-01 | 9,61E-01 |
| SPATA6L | 92,89 | -0,01 | 0,06 | -0,09 | 9,25E-01 | 9,61E-01 |
| IRAK1 | 768,80 | 0,00 | 0,04 | 0,09 | 9,25E-01 | 9,62E-01 |
| GAS8 | 126,38 | 0,00 | 0,04 | -0,09 | 9,25E-01 | 9,62E-01 |
| TPI1 | 3265,66 | 0,00 | 0,02 | -0,09 | 9,25E-01 | 9,62E-01 |
| AP3D1 | 1309,22 | 0,00 | 0,04 | -0,09 | 9,25E-01 | 9,62E-01 |
| PHLDB2 | 381,09 | 0,00 | 0,04 | 0,09 | 9,26E-01 | 9,62E-01 |
| HOOK2 | 219,51 | 0,00 | 0,03 | 0,09 | 9,26E-01 | 9,62E-01 |
| MORN1 | 291,44 | 0,00 | 0,03 | 0,09 | 9,26E-01 | 9,62E-01 |
| EFCAB11 | 121,70 | 0,00 | 0,04 | -0,09 | 9,26E-01 | 9,62E-01 |
| EPB41L3 | 1075,31 | 0,00 | 0,04 | -0,09 | 9,26E-01 | 9,62E-01 |
| UBL3 | 2228,47 | 0,00 | 0,03 | -0,09 | 9,26E-01 | 9,62E-01 |
| AP4B1 | 1238,57 | 0,00 | 0,02 | 0,09 | 9,27E-01 | 9,62E-01 |
| PITRM1 | 1003,25 | 0,00 | 0,02 | 0,09 | 9,26E-01 | 9,62E-01 |
| CDKN2A | 136,64 | 0,00 | 0,05 | -0,09 | 9,27E-01 | 9,62E-01 |
| MED15 | 1859,47 | 0,00 | 0,04 | -0,09 | 9,28E-01 | 9,62E-01 |
| PSME4 | 4467,65 | 0,00 | 0,02 | 0,09 | 9,27E-01 | 9,62E-01 |
| WWOX | 456,06 | 0,00 | 0,03 | 0,09 | 9,27E-01 | 9,62E-01 |
| ZNF174 | 242,17 | 0,00 | 0,01 | -0,09 | 9,27E-01 | 9,62E-01 |
| PIAS3 | 153,51 | 0,00 | 0,05 | -0,09 | 9,28E-01 | 9,62E-01 |
| ZC4H2 | 120,06 | 0,00 | 0,03 | 0,09 | 9,28E-01 | 9,62E-01 |
| CD86 | 1043,90 | 0,00 | 0,03 | -0,09 | 9,28E-01 | 9,62E-01 |
| MARCHF6 | 6818,44 | 0,00 | 0,02 | 0,09 | 9,28E-01 | 9,62E-01 |
| LRCH1 | 987,13 | 0,00 | 0,02 | -0,09 | 9,27E-01 | 9,62E-01 |
| TRABD | 634,42 | 0,00 | 0,05 | 0,09 | 9,27E-01 | 9,62E-01 |
| ATXN7L1 | 809,97 | 0,00 | 0,03 | 0,09 | 9,27E-01 | 9,62E-01 |
| NAA16 | 1086,11 | 0,00 | 0,02 | 0,09 | 9,27E-01 | 9,62E-01 |
| SLC12A4 | 481,85 | 0,00 | 0,04 | 0,09 | 9,27E-01 | 9,62E-01 |
| KLHL25 | 108,23 | -0,01 | 0,06 | -0,09 | 9,27E-01 | 9,62E-01 |
| DPP3 | 241,38 | 0,00 | 0,03 | -0,09 | 9,27E-01 | 9,62E-01 |
| IDH3B | 949,12 | 0,00 | 0,02 | -0,09 | 9,28E-01 | 9,62E-01 |
| TADA3 | 893,08 | 0,00 | 0,03 | 0,09 | 9,28E-01 | 9,62E-01 |
| RAVER1 | 219,49 | 0,00 | 0,05 | -0,09 | 9,28E-01 | 9,62E-01 |
| TGS1 | 972,71 | 0,00 | 0,02 | 0,09 | 9,28E-01 | 9,62E-01 |
| MEF2A | 2183,18 | 0,00 | 0,02 | -0,09 | 9,29E-01 | 9,62E-01 |
| GRAMD4 | 1103,52 | 0,00 | 0,03 | -0,09 | 9,28E-01 | 9,62E-01 |
| PDCL3 | 177,75 | 0,00 | 0,03 | -0,09 | 9,28E-01 | 9,62E-01 |
| RBM14-RBM | 271,34 | 0,00 | 0,02 | -0,09 | 9,29E-01 | 9,62E-01 |
| LAMC1 | 189,64 | -0,01 | 0,06 | -0,09 | 9,29E-01 | 9,63E-01 |
| CRTC3 | 1574,72 | 0,00 | 0,03 | 0,09 | 9,30E-01 | 9,63E-01 |
| ZEB2 | 17009,29 | 0,00 | 0,03 | -0,09 | 9,30E-01 | 9,63E-01 |
| GEMIN4 | 441,17 | 0,00 | 0,02 | 0,09 | 9,30E-01 | 9,63E-01 |
| ZFP64 | 233,53 | 0,00 | 0,03 | 0,09 | 9,30E-01 | 9,63E-01 |
| FXR1 | 2971,98 | 0,00 | 0,03 | -0,09 | 9,30E-01 | 9,63E-01 |
| MANF | 451,87 | 0,00 | 0,03 | -0,09 | 9,30E-01 | 9,63E-01 |

| | | | | | | |
|---|---|---|---|---|---|---|
| TXK | 2882,97 | 0,00 | 0,04 | -0,09 | 9,30E-01 | 9,63E-01 |
| DUSP6 | 2627,35 | 0,00 | 0,03 | -0,09 | 9,31E-01 | 9,63E-01 |
| CDC37 | 2235,92 | 0,00 | 0,02 | -0,09 | 9,30E-01 | 9,63E-01 |
| CSNK2A2 | 1058,14 | 0,00 | 0,02 | -0,09 | 9,30E-01 | 9,63E-01 |
| GABPB2 | 1172,93 | 0,00 | 0,02 | 0,09 | 9,31E-01 | 9,63E-01 |
| GZMH | 1267,56 | 0,01 | 0,08 | 0,09 | 9,31E-01 | 9,63E-01 |
| ING1 | 930,18 | 0,00 | 0,02 | 0,09 | 9,31E-01 | 9,63E-01 |
| DNMT1 | 1742,79 | 0,00 | 0,03 | 0,09 | 9,31E-01 | 9,64E-01 |
| MED31 | 476,70 | 0,00 | 0,03 | -0,09 | 9,31E-01 | 9,64E-01 |
| FOCAD | 1444,53 | 0,00 | 0,03 | 0,09 | 9,32E-01 | 9,64E-01 |
| TFAP2E | 76,87 | 0,00 | 0,05 | -0,09 | 9,32E-01 | 9,64E-01 |
| LAMTOR3 | 1117,91 | 0,00 | 0,04 | 0,08 | 9,32E-01 | 9,64E-01 |
| EIF5 | 4342,88 | 0,00 | 0,02 | 0,08 | 9,33E-01 | 9,65E-01 |
| WDR59 | 900,87 | 0,00 | 0,02 | -0,08 | 9,33E-01 | 9,65E-01 |
| KIAA1671 | 162,88 | -0,01 | 0,09 | -0,08 | 9,33E-01 | 9,65E-01 |
| CAST | 5874,75 | 0,00 | 0,02 | 0,08 | 9,33E-01 | 9,65E-01 |
| PLAAT4 | 811,78 | 0,00 | 0,03 | -0,08 | 9,34E-01 | 9,66E-01 |
| PDE4DIP | 1099,52 | 0,00 | 0,02 | 0,08 | 9,34E-01 | 9,66E-01 |
| SCART1 | 381,72 | 0,00 | 0,06 | -0,08 | 9,34E-01 | 9,66E-01 |
| LARS1 | 2715,73 | 0,00 | 0,02 | -0,08 | 9,34E-01 | 9,66E-01 |
| SGSM1 | 161,36 | -0,01 | 0,07 | -0,08 | 9,34E-01 | 9,66E-01 |
| MED30 | 509,14 | 0,00 | 0,03 | -0,08 | 9,35E-01 | 9,66E-01 |
| USP40 | 579,91 | 0,00 | 0,03 | 0,08 | 9,35E-01 | 9,66E-01 |
| FAM161B | 155,23 | 0,00 | 0,02 | 0,08 | 9,35E-01 | 9,66E-01 |
| MED27 | 289,94 | 0,00 | 0,02 | 0,08 | 9,35E-01 | 9,66E-01 |
| HVCN1 | 1189,14 | 0,00 | 0,03 | 0,08 | 9,36E-01 | 9,67E-01 |
| CLPTM1 | 363,15 | 0,00 | 0,04 | 0,08 | 9,36E-01 | 9,67E-01 |
| SLC25A29 | 614,69 | 0,00 | 0,05 | -0,08 | 9,36E-01 | 9,67E-01 |
| PTP4A1 | 74,03 | 0,00 | 0,03 | -0,08 | 9,36E-01 | 9,67E-01 |
| LEMD3 | 2364,73 | 0,00 | 0,03 | 0,08 | 9,36E-01 | 9,67E-01 |
| CDK16 | 409,21 | 0,00 | 0,03 | 0,08 | 9,36E-01 | 9,67E-01 |
| RNF181 | 596,08 | 0,00 | 0,02 | 0,08 | 9,37E-01 | 9,67E-01 |
| DDRGK1 | 731,45 | 0,00 | 0,02 | -0,08 | 9,37E-01 | 9,67E-01 |
| PMEPA1 | 93,29 | 0,00 | 0,06 | -0,08 | 9,37E-01 | 9,67E-01 |
| AASS | 154,95 | 0,00 | 0,05 | -0,08 | 9,37E-01 | 9,67E-01 |
| USP35 | 181,70 | 0,00 | 0,04 | -0,08 | 9,37E-01 | 9,67E-01 |
| FBP1 | 238,92 | 0,00 | 0,03 | -0,08 | 9,37E-01 | 9,67E-01 |
| FAM199X | 2475,52 | 0,00 | 0,03 | 0,08 | 9,38E-01 | 9,68E-01 |
| RNPC3 | 2005,44 | 0,00 | 0,03 | 0,08 | 9,38E-01 | 9,68E-01 |
| ST8SIA6 | 151,65 | 0,00 | 0,04 | 0,08 | 9,38E-01 | 9,68E-01 |
| CIPC | 473,53 | 0,00 | 0,02 | 0,08 | 9,38E-01 | 9,68E-01 |
| DOCK9 | 2154,80 | 0,00 | 0,03 | 0,08 | 9,39E-01 | 9,68E-01 |
| ORMDL2 | 683,15 | 0,00 | 0,01 | 0,08 | 9,39E-01 | 9,68E-01 |
| CEP131 | 179,22 | 0,00 | 0,04 | -0,08 | 9,39E-01 | 9,68E-01 |
| MCC | 122,75 | -0,01 | 0,07 | -0,08 | 9,39E-01 | 9,68E-01 |
| PTK2 | 442,86 | 0,00 | 0,03 | -0,08 | 9,39E-01 | 9,68E-01 |

| | | | | | | |
|---|---|---|---|---|---|---|
| PGM5 | 212,67 | -0,01 | 0,10 | -0,08 | 9,39E-01 | 9,68E-01 |
| ZSCAN25 | 1143,45 | 0,00 | 0,02 | -0,08 | 9,39E-01 | 9,68E-01 |
| MIB2 | 693,50 | 0,00 | 0,03 | -0,08 | 9,40E-01 | 9,69E-01 |
| FUT11 | 1108,55 | 0,00 | 0,02 | -0,08 | 9,40E-01 | 9,69E-01 |
| NMRK1 | 976,27 | 0,00 | 0,03 | 0,08 | 9,40E-01 | 9,69E-01 |
| SUMO4 | 381,48 | 0,00 | 0,04 | -0,08 | 9,40E-01 | 9,69E-01 |
| NBPF15 | 1281,28 | 0,00 | 0,03 | 0,08 | 9,40E-01 | 9,69E-01 |
| PLAGL2 | 586,79 | 0,00 | 0,04 | 0,08 | 9,40E-01 | 9,69E-01 |
| PPIL2 | 924,92 | 0,00 | 0,03 | 0,07 | 9,41E-01 | 9,69E-01 |
| CAPRIN1 | 3963,85 | 0,00 | 0,01 | -0,07 | 9,41E-01 | 9,69E-01 |
| PIK3R3 | 109,74 | 0,00 | 0,05 | 0,07 | 9,41E-01 | 9,69E-01 |
| POLR2F | 494,09 | 0,00 | 0,02 | 0,07 | 9,41E-01 | 9,69E-01 |
| TTC28 | 142,11 | -0,01 | 0,08 | -0,07 | 9,41E-01 | 9,69E-01 |
| SLC35D1 | 1301,78 | 0,00 | 0,02 | 0,07 | 9,41E-01 | 9,69E-01 |
| SLC37A2 | 739,60 | 0,00 | 0,04 | 0,07 | 9,40E-01 | 9,69E-01 |
| PSMD7 | 1590,53 | 0,00 | 0,02 | 0,07 | 9,41E-01 | 9,69E-01 |
| ZCCHC24 | 104,30 | 0,00 | 0,05 | 0,07 | 9,42E-01 | 9,69E-01 |
| LTN1 | 3373,56 | 0,00 | 0,03 | 0,07 | 9,42E-01 | 9,69E-01 |
| INO80D | 5164,63 | 0,00 | 0,02 | 0,07 | 9,42E-01 | 9,69E-01 |
| PCNX3 | 798,68 | 0,00 | 0,05 | 0,07 | 9,42E-01 | 9,69E-01 |
| ZNF224 | 2212,84 | 0,00 | 0,02 | 0,07 | 9,42E-01 | 9,69E-01 |
| TRIQK | 714,29 | 0,00 | 0,04 | -0,07 | 9,42E-01 | 9,69E-01 |
| HDAC7 | 1731,80 | 0,00 | 0,05 | -0,07 | 9,43E-01 | 9,70E-01 |
| ZNF335 | 813,71 | 0,00 | 0,04 | 0,07 | 9,43E-01 | 9,70E-01 |
| STEEP1 | 455,21 | 0,00 | 0,02 | -0,07 | 9,43E-01 | 9,70E-01 |
| HENMT1 | 492,16 | 0,00 | 0,03 | -0,07 | 9,43E-01 | 9,70E-01 |
| PLEKHM2 | 570,57 | 0,00 | 0,04 | 0,07 | 9,44E-01 | 9,71E-01 |
| YTHDF2 | 1200,97 | 0,00 | 0,01 | 0,07 | 9,44E-01 | 9,71E-01 |
| ZDHHC14 | 174,01 | 0,00 | 0,05 | -0,07 | 9,44E-01 | 9,71E-01 |
| AKAP6 | 236,37 | 0,00 | 0,06 | 0,07 | 9,44E-01 | 9,71E-01 |
| SGSH | 366,63 | 0,00 | 0,07 | 0,07 | 9,44E-01 | 9,71E-01 |
| SLC24A4 | 1327,28 | 0,00 | 0,06 | -0,07 | 9,44E-01 | 9,71E-01 |
| MAST2 | 294,48 | 0,00 | 0,04 | 0,07 | 9,45E-01 | 9,72E-01 |
| EDC3 | 444,26 | 0,00 | 0,03 | 0,07 | 9,45E-01 | 9,72E-01 |
| SLFN11 | 2001,75 | 0,00 | 0,02 | 0,07 | 9,46E-01 | 9,72E-01 |
| CBX7 | 1820,24 | 0,00 | 0,04 | -0,07 | 9,46E-01 | 9,72E-01 |
| FRMPD3 | 85,46 | -0,01 | 0,08 | -0,07 | 9,46E-01 | 9,72E-01 |
| ZNF169 | 652,96 | 0,00 | 0,02 | -0,07 | 9,47E-01 | 9,73E-01 |
| DUS2 | 709,42 | 0,00 | 0,02 | 0,07 | 9,48E-01 | 9,74E-01 |
| AGFG1 | 4588,61 | 0,00 | 0,03 | 0,07 | 9,48E-01 | 9,74E-01 |
| SPG7 | 3706,95 | 0,00 | 0,03 | -0,07 | 9,48E-01 | 9,74E-01 |
| RNF125 | 1733,67 | 0,00 | 0,02 | 0,07 | 9,48E-01 | 9,74E-01 |
| CHP1 | 1242,39 | 0,00 | 0,04 | -0,06 | 9,49E-01 | 9,74E-01 |
| JUN | 304,15 | 0,00 | 0,04 | -0,06 | 9,49E-01 | 9,74E-01 |
| AMY2B | 703,14 | 0,00 | 0,02 | -0,06 | 9,49E-01 | 9,74E-01 |
| ZNF836 | 449,71 | 0,00 | 0,04 | 0,06 | 9,49E-01 | 9,74E-01 |

| | | | | | | |
|---|---|---|---|---|---|---|
| LCMT1 | 246,78 | 0,00 | 0,02 | 0,06 | 9,48E-01 | 9,74E-01 |
| PHKB | 3107,73 | 0,00 | 0,02 | 0,06 | 9,49E-01 | 9,74E-01 |
| TERF2IP | 3039,89 | 0,00 | 0,03 | 0,06 | 9,48E-01 | 9,74E-01 |
| FLNA | 18907,32 | 0,00 | 0,05 | 0,06 | 9,48E-01 | 9,74E-01 |
| PFDN6 | 313,39 | 0,00 | 0,03 | -0,06 | 9,49E-01 | 9,74E-01 |
| TMA7B | 228,19 | 0,00 | 0,04 | -0,06 | 9,49E-01 | 9,74E-01 |
| SPAG5 | 93,48 | 0,00 | 0,04 | -0,06 | 9,49E-01 | 9,74E-01 |
| PHC1 | 289,96 | 0,00 | 0,03 | 0,06 | 9,50E-01 | 9,74E-01 |
| GGTA1 | 144,22 | 0,00 | 0,06 | -0,06 | 9,50E-01 | 9,74E-01 |
| PRKAG1 | 902,69 | 0,00 | 0,02 | 0,06 | 9,50E-01 | 9,74E-01 |
| RAPGEF6 | 1850,67 | 0,00 | 0,03 | 0,06 | 9,50E-01 | 9,74E-01 |
| ST3GAL1 | 4695,40 | 0,00 | 0,03 | -0,06 | 9,50E-01 | 9,74E-01 |
| B2M | 119849,29 | 0,00 | 0,06 | -0,06 | 9,50E-01 | 9,75E-01 |
| RB1 | 2248,82 | 0,00 | 0,03 | -0,06 | 9,51E-01 | 9,75E-01 |
| RNF19A | 4948,63 | 0,00 | 0,03 | -0,06 | 9,51E-01 | 9,75E-01 |
| MGAT4A | 6619,32 | 0,00 | 0,03 | -0,06 | 9,51E-01 | 9,75E-01 |
| MAP3K4 | 1812,52 | 0,00 | 0,02 | 0,06 | 9,51E-01 | 9,75E-01 |
| GCC2 | 3387,48 | 0,00 | 0,04 | -0,06 | 9,51E-01 | 9,75E-01 |
| HEXD | 383,28 | 0,00 | 0,05 | 0,06 | 9,52E-01 | 9,75E-01 |
| PYGB | 1242,06 | 0,00 | 0,03 | 0,06 | 9,52E-01 | 9,75E-01 |
| SCN8A | 95,73 | -0,01 | 0,10 | -0,06 | 9,52E-01 | 9,75E-01 |
| PARP6 | 831,09 | 0,00 | 0,03 | -0,06 | 9,52E-01 | 9,75E-01 |
| SENP3 | 303,14 | 0,00 | 0,02 | 0,06 | 9,52E-01 | 9,76E-01 |
| NBPF3 | 406,30 | 0,00 | 0,04 | 0,06 | 9,53E-01 | 9,76E-01 |
| GCOM1 | 87,93 | -0,01 | 0,09 | -0,06 | 9,53E-01 | 9,76E-01 |
| RAB32 | 921,92 | 0,00 | 0,03 | 0,06 | 9,53E-01 | 9,76E-01 |
| MAPKBP1 | 602,80 | 0,00 | 0,03 | 0,06 | 9,53E-01 | 9,76E-01 |
| ZNF710 | 721,19 | 0,00 | 0,05 | -0,06 | 9,53E-01 | 9,76E-01 |
| LYSMD2 | 480,72 | 0,00 | 0,03 | 0,06 | 9,54E-01 | 9,76E-01 |
| MDN1 | 3714,65 | 0,00 | 0,03 | -0,06 | 9,54E-01 | 9,76E-01 |
| BCAS2 | 442,37 | 0,00 | 0,03 | -0,06 | 9,54E-01 | 9,76E-01 |
| TRMT10B | 351,93 | 0,00 | 0,02 | 0,06 | 9,54E-01 | 9,76E-01 |
| DTHD1 | 856,98 | 0,00 | 0,06 | 0,06 | 9,54E-01 | 9,76E-01 |
| SDCCAG8 | 1140,22 | 0,00 | 0,02 | -0,06 | 9,54E-01 | 9,76E-01 |
| NETO2 | 151,48 | 0,00 | 0,04 | 0,06 | 9,54E-01 | 9,77E-01 |
| SLC22A18 | 359,84 | 0,00 | 0,05 | 0,06 | 9,54E-01 | 9,77E-01 |
| JADE2 | 2437,89 | 0,00 | 0,03 | 0,06 | 9,54E-01 | 9,77E-01 |
| TXLNG | 626,82 | 0,00 | 0,02 | -0,06 | 9,54E-01 | 9,77E-01 |
| CALD1 | 91,89 | 0,01 | 0,09 | 0,06 | 9,55E-01 | 9,77E-01 |
| DLGAP4 | 224,60 | 0,00 | 0,08 | -0,06 | 9,55E-01 | 9,77E-01 |
| ITPRIPL1 | 224,55 | 0,00 | 0,04 | 0,06 | 9,55E-01 | 9,77E-01 |
| SLX4 | 156,90 | 0,00 | 0,07 | -0,06 | 9,55E-01 | 9,77E-01 |
| FKRP | 221,19 | 0,00 | 0,04 | 0,06 | 9,55E-01 | 9,77E-01 |
| ZBTB39 | 390,50 | 0,00 | 0,02 | 0,06 | 9,55E-01 | 9,77E-01 |
| ZNF430 | 1327,61 | 0,00 | 0,02 | 0,06 | 9,55E-01 | 9,77E-01 |
| URGCP | 655,09 | 0,00 | 0,02 | 0,06 | 9,56E-01 | 9,77E-01 |

| | | | | | | |
|---|---|---|---|---|---|---|
| FAM200B | 1009,65 | 0,00 | 0,04 | 0,06 | 9,56E-01 | 9,77E-01 |
| LDAF1 | 346,95 | 0,00 | 0,02 | -0,05 | 9,57E-01 | 9,78E-01 |
| KLRC3 | 132,82 | 0,00 | 0,08 | -0,05 | 9,57E-01 | 9,78E-01 |
| PCDHGA10 | 75,16 | 0,00 | 0,06 | -0,05 | 9,57E-01 | 9,78E-01 |
| DPM1 | 764,68 | 0,00 | 0,04 | -0,05 | 9,57E-01 | 9,78E-01 |
| CPT2 | 655,04 | 0,00 | 0,01 | -0,05 | 9,57E-01 | 9,78E-01 |
| BLVRA | 469,69 | 0,00 | 0,04 | -0,05 | 9,58E-01 | 9,79E-01 |
| CALML4 | 98,58 | 0,00 | 0,03 | 0,05 | 9,58E-01 | 9,79E-01 |
| DAGLB | 603,21 | 0,00 | 0,03 | -0,05 | 9,58E-01 | 9,79E-01 |
| LSS | 870,74 | 0,00 | 0,03 | 0,05 | 9,58E-01 | 9,79E-01 |
| ZNF768 | 183,77 | 0,00 | 0,03 | -0,05 | 9,59E-01 | 9,79E-01 |
| PARD3 | 111,09 | 0,00 | 0,07 | 0,05 | 9,59E-01 | 9,79E-01 |
| BMP2K | 4623,39 | 0,00 | 0,03 | 0,05 | 9,59E-01 | 9,79E-01 |
| ZNF784 | 141,80 | 0,00 | 0,03 | 0,05 | 9,59E-01 | 9,80E-01 |
| PTGDS | 89,77 | 0,00 | 0,07 | 0,05 | 9,60E-01 | 9,80E-01 |
| ATRN | 1249,62 | 0,00 | 0,02 | 0,05 | 9,60E-01 | 9,80E-01 |
| CORO2A | 271,23 | 0,00 | 0,03 | 0,05 | 9,60E-01 | 9,80E-01 |
| FCER1A | 477,13 | 0,00 | 0,08 | -0,05 | 9,60E-01 | 9,80E-01 |
| H2BC17 | 204,70 | 0,00 | 0,05 | -0,05 | 9,60E-01 | 9,80E-01 |
| KLRG1 | 1720,72 | 0,00 | 0,05 | 0,05 | 9,60E-01 | 9,80E-01 |
| VASH1 | 335,39 | 0,00 | 0,06 | -0,05 | 9,60E-01 | 9,80E-01 |
| MERTK | 181,48 | 0,00 | 0,06 | -0,05 | 9,61E-01 | 9,80E-01 |
| STK36 | 237,76 | 0,00 | 0,03 | 0,05 | 9,61E-01 | 9,80E-01 |
| BRD9 | 1662,42 | 0,00 | 0,02 | -0,05 | 9,61E-01 | 9,80E-01 |
| PRMT2 | 5301,53 | 0,00 | 0,02 | 0,05 | 9,61E-01 | 9,80E-01 |
| MYO18B | 76,34 | -0,01 | 0,16 | -0,05 | 9,61E-01 | 9,80E-01 |
| MAPK10 | 83,64 | -0,01 | 0,14 | -0,05 | 9,61E-01 | 9,80E-01 |
| SAMD3 | 1772,82 | 0,00 | 0,04 | -0,05 | 9,61E-01 | 9,80E-01 |
| NAV2 | 79,75 | 0,01 | 0,13 | 0,05 | 9,62E-01 | 9,81E-01 |
| ENSG000002 | 76,25 | 0,00 | 0,05 | -0,05 | 9,62E-01 | 9,81E-01 |
| ITGB8 | 169,34 | 0,00 | 0,10 | 0,05 | 9,63E-01 | 9,81E-01 |
| PER2 | 258,12 | 0,00 | 0,04 | 0,05 | 9,63E-01 | 9,82E-01 |
| MTPN | 9735,46 | 0,00 | 0,03 | 0,05 | 9,63E-01 | 9,82E-01 |
| RBMX2 | 414,83 | 0,00 | 0,02 | -0,05 | 9,63E-01 | 9,82E-01 |
| SUCLG1 | 1144,79 | 0,00 | 0,02 | -0,05 | 9,63E-01 | 9,82E-01 |
| FAR1 | 5273,53 | 0,00 | 0,04 | -0,05 | 9,63E-01 | 9,82E-01 |
| AMOT | 120,26 | 0,00 | 0,04 | -0,05 | 9,63E-01 | 9,82E-01 |
| CDC40 | 2119,44 | 0,00 | 0,03 | -0,05 | 9,64E-01 | 9,82E-01 |
| ITGB3 | 1419,38 | 0,00 | 0,09 | -0,05 | 9,64E-01 | 9,82E-01 |
| FCF1 | 938,71 | 0,00 | 0,03 | -0,04 | 9,64E-01 | 9,82E-01 |
| PCBD2 | 462,21 | 0,00 | 0,02 | -0,04 | 9,65E-01 | 9,83E-01 |
| DSC1 | 124,87 | 0,00 | 0,08 | -0,04 | 9,65E-01 | 9,83E-01 |
| CBX5 | 1738,25 | 0,00 | 0,03 | -0,04 | 9,65E-01 | 9,83E-01 |
| PIN4 | 257,76 | 0,00 | 0,03 | -0,04 | 9,65E-01 | 9,83E-01 |
| BICRA | 137,80 | 0,00 | 0,09 | 0,04 | 9,65E-01 | 9,83E-01 |
| S100A6 | 2712,52 | 0,00 | 0,03 | 0,04 | 9,66E-01 | 9,83E-01 |

| | | | | | | |
|---|---|---|---|---|---|---|
| STAT4 | 2709,30 | 0,00 | 0,03 | 0,04 | 9,66E-01 | 9,83E-01 |
| THAP4 | 178,64 | 0,00 | 0,03 | -0,04 | 9,66E-01 | 9,83E-01 |
| DSP | 96,84 | -0,01 | 0,13 | -0,04 | 9,66E-01 | 9,83E-01 |
| CCDC127 | 508,21 | 0,00 | 0,02 | -0,04 | 9,66E-01 | 9,83E-01 |
| PRKCA | 1483,11 | 0,00 | 0,04 | 0,04 | 9,66E-01 | 9,83E-01 |
| SUFU | 241,15 | 0,00 | 0,05 | 0,04 | 9,67E-01 | 9,84E-01 |
| HEXA | 1560,04 | 0,00 | 0,02 | 0,04 | 9,68E-01 | 9,84E-01 |
| ZNF844 | 926,74 | 0,00 | 0,04 | 0,04 | 9,68E-01 | 9,84E-01 |
| MED23 | 3412,58 | 0,00 | 0,02 | -0,04 | 9,68E-01 | 9,84E-01 |
| JAK2 | 6038,76 | 0,00 | 0,04 | 0,04 | 9,68E-01 | 9,84E-01 |
| OTUD3 | 789,90 | 0,00 | 0,02 | 0,04 | 9,68E-01 | 9,84E-01 |
| EIF4G1 | 2158,60 | 0,00 | 0,04 | -0,04 | 9,68E-01 | 9,85E-01 |
| STX8 | 477,74 | 0,00 | 0,02 | 0,04 | 9,69E-01 | 9,85E-01 |
| C1orf35 | 373,41 | 0,00 | 0,02 | -0,04 | 9,69E-01 | 9,85E-01 |
| CNNM2 | 476,27 | 0,00 | 0,03 | 0,04 | 9,69E-01 | 9,85E-01 |
| RFT1 | 647,55 | 0,00 | 0,02 | 0,04 | 9,69E-01 | 9,85E-01 |
| CCDC97 | 370,36 | 0,00 | 0,05 | -0,04 | 9,69E-01 | 9,85E-01 |
| PGLS | 464,35 | 0,00 | 0,04 | -0,04 | 9,69E-01 | 9,85E-01 |
| CEACAM21 | 450,70 | 0,00 | 0,02 | 0,04 | 9,69E-01 | 9,85E-01 |
| RUNX3 | 1126,43 | 0,00 | 0,04 | 0,04 | 9,70E-01 | 9,85E-01 |
| ZNF394 | 2525,91 | 0,00 | 0,02 | -0,04 | 9,70E-01 | 9,85E-01 |
| COL4A3 | 201,35 | 0,00 | 0,08 | -0,04 | 9,70E-01 | 9,85E-01 |
| DNAJA2 | 2301,26 | 0,00 | 0,03 | 0,04 | 9,70E-01 | 9,85E-01 |
| PBDC1 | 245,39 | 0,00 | 0,03 | -0,04 | 9,70E-01 | 9,85E-01 |
| HMG20A | 1110,77 | 0,00 | 0,02 | -0,04 | 9,71E-01 | 9,86E-01 |
| HSPH1 | 1112,35 | 0,00 | 0,04 | 0,04 | 9,71E-01 | 9,86E-01 |
| MVB12B | 223,08 | 0,00 | 0,06 | -0,04 | 9,72E-01 | 9,86E-01 |
| ACOT8 | 674,39 | 0,00 | 0,01 | -0,04 | 9,72E-01 | 9,86E-01 |
| DENND2C | 119,36 | 0,00 | 0,06 | 0,04 | 9,71E-01 | 9,86E-01 |
| PCYOX1 | 665,37 | 0,00 | 0,02 | 0,04 | 9,71E-01 | 9,86E-01 |
| HOOK3 | 3608,82 | 0,00 | 0,02 | 0,04 | 9,71E-01 | 9,86E-01 |
| MCM5 | 1117,41 | 0,00 | 0,02 | 0,04 | 9,71E-01 | 9,86E-01 |
| ERICH1 | 3654,12 | 0,00 | 0,03 | 0,04 | 9,71E-01 | 9,86E-01 |
| ACSL5 | 1693,57 | 0,00 | 0,02 | 0,04 | 9,71E-01 | 9,86E-01 |
| EPHA4 | 753,72 | 0,00 | 0,04 | 0,04 | 9,71E-01 | 9,86E-01 |
| SEC61B | 996,50 | 0,00 | 0,03 | -0,04 | 9,72E-01 | 9,86E-01 |
| XRCC4 | 352,35 | 0,00 | 0,06 | -0,04 | 9,72E-01 | 9,86E-01 |
| SH3TC2 | 130,02 | 0,00 | 0,08 | 0,03 | 9,72E-01 | 9,86E-01 |
| CBFA2T2 | 588,41 | 0,00 | 0,04 | -0,03 | 9,72E-01 | 9,86E-01 |
| MAP1S | 254,47 | 0,00 | 0,04 | 0,03 | 9,73E-01 | 9,86E-01 |
| RSU1 | 2010,02 | 0,00 | 0,03 | -0,03 | 9,73E-01 | 9,87E-01 |
| MDGA1 | 242,66 | 0,00 | 0,13 | 0,03 | 9,73E-01 | 9,87E-01 |
| MYOM2 | 428,62 | 0,00 | 0,14 | -0,03 | 9,73E-01 | 9,87E-01 |
| AACS | 331,87 | 0,00 | 0,05 | -0,03 | 9,73E-01 | 9,87E-01 |
| CTNS | 270,11 | 0,00 | 0,02 | 0,03 | 9,73E-01 | 9,87E-01 |
| REEP5 | 1668,60 | 0,00 | 0,03 | 0,03 | 9,73E-01 | 9,87E-01 |

| | | | | | | |
|---|---|---|---|---|---|---|
| FCRL5 | 859,03 | 0,00 | 0,10 | -0,03 | 9,74E-01 | 9,87E-01 |
| EML3 | 675,06 | 0,00 | 0,05 | 0,03 | 9,74E-01 | 9,87E-01 |
| UBAP1L | 398,46 | 0,00 | 0,05 | -0,03 | 9,74E-01 | 9,87E-01 |
| GLIS3 | 83,37 | 0,00 | 0,09 | 0,03 | 9,74E-01 | 9,87E-01 |
| H3C6 | 168,76 | 0,00 | 0,05 | -0,03 | 9,74E-01 | 9,87E-01 |
| ZNF708 | 1395,42 | 0,00 | 0,04 | 0,03 | 9,75E-01 | 9,88E-01 |
| MAGI3 | 168,73 | 0,00 | 0,04 | 0,03 | 9,76E-01 | 9,88E-01 |
| MLLT6 | 2712,90 | 0,00 | 0,05 | -0,03 | 9,76E-01 | 9,88E-01 |
| SELENOO | 680,50 | 0,00 | 0,03 | 0,03 | 9,76E-01 | 9,89E-01 |
| PDHB | 1117,62 | 0,00 | 0,02 | 0,03 | 9,76E-01 | 9,89E-01 |
| S1PR3 | 461,59 | 0,00 | 0,04 | -0,03 | 9,76E-01 | 9,89E-01 |
| VDAC3 | 1449,27 | 0,00 | 0,03 | 0,03 | 9,77E-01 | 9,89E-01 |
| TSEN54 | 318,01 | 0,00 | 0,04 | 0,03 | 9,77E-01 | 9,89E-01 |
| ARHGAP29 | 87,02 | 0,00 | 0,08 | 0,03 | 9,77E-01 | 9,89E-01 |
| EPC1 | 4441,87 | 0,00 | 0,02 | -0,03 | 9,77E-01 | 9,89E-01 |
| RRP12 | 848,22 | 0,00 | 0,05 | 0,03 | 9,77E-01 | 9,89E-01 |
| AMOTL1 | 108,82 | 0,00 | 0,07 | -0,03 | 9,77E-01 | 9,89E-01 |
| ASTN2 | 178,55 | 0,00 | 0,05 | -0,03 | 9,77E-01 | 9,89E-01 |
| C14orf119 | 964,49 | 0,00 | 0,02 | 0,03 | 9,77E-01 | 9,89E-01 |
| XPNPEP3 | 500,91 | 0,00 | 0,02 | -0,03 | 9,78E-01 | 9,89E-01 |
| RYR3 | 135,82 | 0,00 | 0,14 | -0,03 | 9,78E-01 | 9,89E-01 |
| ZNF619 | 390,44 | 0,00 | 0,02 | -0,03 | 9,78E-01 | 9,89E-01 |
| FRMD3 | 311,68 | 0,00 | 0,05 | -0,03 | 9,78E-01 | 9,89E-01 |
| TTLL7 | 81,84 | 0,00 | 0,08 | -0,03 | 9,78E-01 | 9,89E-01 |
| HPCAL1 | 827,44 | 0,00 | 0,05 | 0,03 | 9,78E-01 | 9,89E-01 |
| SYTL2 | 2364,09 | 0,00 | 0,05 | -0,03 | 9,78E-01 | 9,89E-01 |
| PHAF1 | 338,53 | 0,00 | 0,03 | -0,03 | 9,78E-01 | 9,89E-01 |
| PLEKHG5 | 91,98 | 0,00 | 0,09 | -0,03 | 9,78E-01 | 9,89E-01 |
| PI4K2A | 291,47 | 0,00 | 0,02 | 0,03 | 9,79E-01 | 9,89E-01 |
| CEP250 | 1853,69 | 0,00 | 0,02 | 0,03 | 9,79E-01 | 9,89E-01 |
| WIZ | 254,11 | 0,00 | 0,06 | -0,03 | 9,79E-01 | 9,90E-01 |
| JMJD7-PLA2( | 313,36 | 0,00 | 0,05 | 0,03 | 9,79E-01 | 9,90E-01 |
| INO80E | 1116,89 | 0,00 | 0,03 | 0,03 | 9,79E-01 | 9,90E-01 |
| ADAM23 | 236,12 | 0,00 | 0,07 | 0,03 | 9,79E-01 | 9,90E-01 |
| RTL10 | 453,29 | 0,00 | 0,03 | 0,03 | 9,79E-01 | 9,90E-01 |
| HSDL1 | 645,15 | 0,00 | 0,02 | 0,03 | 9,80E-01 | 9,90E-01 |
| ZNF346 | 496,18 | 0,00 | 0,02 | 0,02 | 9,80E-01 | 9,90E-01 |
| PITPNM1 | 856,47 | 0,00 | 0,05 | -0,02 | 9,80E-01 | 9,90E-01 |
| RPS6KA2 | 320,67 | 0,00 | 0,05 | -0,02 | 9,80E-01 | 9,90E-01 |
| ADGRG5 | 449,86 | 0,00 | 0,06 | -0,02 | 9,80E-01 | 9,90E-01 |
| ZNF609 | 2094,52 | 0,00 | 0,03 | 0,02 | 9,80E-01 | 9,90E-01 |
| COPS5 | 1492,15 | 0,00 | 0,02 | 0,02 | 9,81E-01 | 9,90E-01 |
| SNX17 | 2278,41 | 0,00 | 0,02 | 0,02 | 9,81E-01 | 9,90E-01 |
| SNX2 | 2887,40 | 0,00 | 0,03 | 0,02 | 9,81E-01 | 9,91E-01 |
| PRPF40B | 189,29 | 0,00 | 0,04 | -0,02 | 9,81E-01 | 9,91E-01 |
| AIFM2 | 117,95 | 0,00 | 0,05 | 0,02 | 9,81E-01 | 9,91E-01 |

| | | | | | | |
|---|---|---|---|---|---|---|
| NSUN5 | 512,64 | 0,00 | 0,02 | 0,02 | 9,81E-01 | 9,91E-01 |
| CLN8 | 746,65 | 0,00 | 0,02 | 0,02 | 9,82E-01 | 9,91E-01 |
| CSMD1 | 73,04 | 0,00 | 0,16 | 0,02 | 9,82E-01 | 9,91E-01 |
| KHDRBS1 | 1783,46 | 0,00 | 0,02 | 0,02 | 9,82E-01 | 9,91E-01 |
| NAT1 | 267,32 | 0,00 | 0,03 | 0,02 | 9,82E-01 | 9,91E-01 |
| SMUG1 | 768,00 | 0,00 | 0,01 | -0,02 | 9,82E-01 | 9,91E-01 |
| MICAL2 | 7468,61 | 0,00 | 0,05 | -0,02 | 9,82E-01 | 9,91E-01 |
| MAN2B1 | 1988,73 | 0,00 | 0,03 | 0,02 | 9,82E-01 | 9,91E-01 |
| DNAJC9 | 911,07 | 0,00 | 0,02 | -0,02 | 9,83E-01 | 9,91E-01 |
| ENSG000002 | 109,07 | 0,00 | 0,05 | 0,02 | 9,83E-01 | 9,91E-01 |
| ARF1 | 3885,12 | 0,00 | 0,03 | 0,02 | 9,83E-01 | 9,92E-01 |
| CORO7 | 559,55 | 0,00 | 0,04 | -0,02 | 9,84E-01 | 9,92E-01 |
| ZBED4 | 1025,89 | 0,00 | 0,02 | 0,02 | 9,84E-01 | 9,92E-01 |
| TTN | 6690,99 | 0,00 | 0,05 | -0,02 | 9,83E-01 | 9,92E-01 |
| C2CD2 | 347,42 | 0,00 | 0,03 | 0,02 | 9,84E-01 | 9,92E-01 |
| CENPF | 182,39 | 0,00 | 0,06 | 0,02 | 9,83E-01 | 9,92E-01 |
| CTNNBIP1 | 183,69 | 0,00 | 0,04 | 0,02 | 9,84E-01 | 9,92E-01 |
| TIGAR | 151,51 | 0,00 | 0,02 | 0,02 | 9,84E-01 | 9,92E-01 |
| H2BC9 | 602,18 | 0,00 | 0,04 | 0,02 | 9,84E-01 | 9,92E-01 |
| SRP68 | 649,40 | 0,00 | 0,02 | 0,02 | 9,84E-01 | 9,92E-01 |
| KIAA0586 | 1405,62 | 0,00 | 0,02 | -0,02 | 9,85E-01 | 9,92E-01 |
| RAD54L2 | 1485,19 | 0,00 | 0,02 | -0,02 | 9,85E-01 | 9,92E-01 |
| RAB1B | 722,45 | 0,00 | 0,04 | 0,02 | 9,85E-01 | 9,92E-01 |
| TXNDC11 | 812,37 | 0,00 | 0,02 | -0,02 | 9,85E-01 | 9,92E-01 |
| PKP4 | 634,65 | 0,00 | 0,03 | -0,02 | 9,86E-01 | 9,92E-01 |
| DMTF1 | 4825,01 | 0,00 | 0,02 | -0,02 | 9,86E-01 | 9,92E-01 |
| GNAO1 | 101,72 | 0,00 | 0,10 | -0,02 | 9,86E-01 | 9,92E-01 |
| INSIG2 | 859,53 | 0,00 | 0,03 | 0,02 | 9,86E-01 | 9,92E-01 |
| FSTL1 | 190,29 | 0,00 | 0,08 | -0,02 | 9,86E-01 | 9,92E-01 |
| ZNF418 | 229,37 | 0,00 | 0,04 | -0,02 | 9,86E-01 | 9,92E-01 |
| CRTC2 | 519,07 | 0,00 | 0,04 | 0,02 | 9,86E-01 | 9,92E-01 |
| UAP1 | 474,41 | 0,00 | 0,03 | 0,02 | 9,86E-01 | 9,92E-01 |
| SENP7 | 3447,46 | 0,00 | 0,04 | 0,02 | 9,86E-01 | 9,92E-01 |
| BUB1 | 111,14 | 0,00 | 0,06 | -0,02 | 9,86E-01 | 9,92E-01 |
| POLR3GL | 255,06 | 0,00 | 0,04 | -0,02 | 9,86E-01 | 9,93E-01 |
| P2RY14 | 483,28 | 0,00 | 0,07 | -0,02 | 9,87E-01 | 9,93E-01 |
| ZNF155 | 318,69 | 0,00 | 0,04 | -0,02 | 9,87E-01 | 9,93E-01 |
| HDAC8 | 871,16 | 0,00 | 0,03 | -0,02 | 9,87E-01 | 9,93E-01 |
| PKD2 | 709,70 | 0,00 | 0,02 | 0,02 | 9,87E-01 | 9,93E-01 |
| FBN1 | 152,00 | 0,00 | 0,10 | 0,02 | 9,87E-01 | 9,93E-01 |
| CALHM6 | 269,87 | 0,00 | 0,07 | -0,02 | 9,87E-01 | 9,93E-01 |
| PLK1 | 79,21 | 0,00 | 0,05 | 0,02 | 9,87E-01 | 9,93E-01 |
| TTC7B | 203,18 | 0,00 | 0,07 | -0,02 | 9,88E-01 | 9,93E-01 |
| FBXO32 | 429,60 | 0,00 | 0,03 | 0,02 | 9,88E-01 | 9,93E-01 |
| ABCF1 | 806,60 | 0,00 | 0,03 | 0,01 | 9,88E-01 | 9,93E-01 |
| NIPAL3 | 2194,61 | 0,00 | 0,02 | 0,01 | 9,88E-01 | 9,93E-01 |

| | | | | | | |
|---|---|---|---|---|---|---|
| CD244 | 1682,97 | 0,00 | 0,03 | 0,01 | 9,88E-01 | 9,93E-01 |
| OGT | 19934,51 | 0,00 | 0,02 | -0,01 | 9,88E-01 | 9,94E-01 |
| ING2 | 124,24 | 0,00 | 0,03 | -0,01 | 9,89E-01 | 9,94E-01 |
| DNASE1L1 | 771,59 | 0,00 | 0,02 | 0,01 | 9,89E-01 | 9,94E-01 |
| GSTO2 | 73,66 | 0,00 | 0,06 | 0,01 | 9,89E-01 | 9,94E-01 |
| GBGT1 | 563,50 | 0,00 | 0,03 | 0,01 | 9,89E-01 | 9,94E-01 |
| TMEM87A | 1960,55 | 0,00 | 0,02 | -0,01 | 9,89E-01 | 9,94E-01 |
| ATP10D | 1654,88 | 0,00 | 0,02 | 0,01 | 9,90E-01 | 9,94E-01 |
| SEMA5A | 95,97 | 0,00 | 0,08 | -0,01 | 9,90E-01 | 9,94E-01 |
| CHRNE | 75,83 | 0,00 | 0,06 | -0,01 | 9,90E-01 | 9,94E-01 |
| DGCR8 | 624,76 | 0,00 | 0,03 | -0,01 | 9,90E-01 | 9,95E-01 |
| AJAP1 | 87,34 | 0,00 | 0,11 | -0,01 | 9,91E-01 | 9,95E-01 |
| USP36 | 718,93 | 0,00 | 0,04 | -0,01 | 9,91E-01 | 9,95E-01 |
| MCUR1 | 419,53 | 0,00 | 0,02 | -0,01 | 9,91E-01 | 9,95E-01 |
| DDX18 | 2244,83 | 0,00 | 0,02 | 0,01 | 9,91E-01 | 9,95E-01 |
| HTT | 3047,46 | 0,00 | 0,03 | -0,01 | 9,91E-01 | 9,95E-01 |
| GINS4 | 89,14 | 0,00 | 0,04 | 0,01 | 9,91E-01 | 9,95E-01 |
| UBL7 | 990,72 | 0,00 | 0,03 | -0,01 | 9,92E-01 | 9,96E-01 |
| SLC1A4 | 162,10 | 0,00 | 0,04 | -0,01 | 9,93E-01 | 9,96E-01 |
| PPCS | 854,95 | 0,00 | 0,02 | 0,01 | 9,93E-01 | 9,96E-01 |
| IMMT | 1126,78 | 0,00 | 0,02 | -0,01 | 9,93E-01 | 9,96E-01 |
| NFKBID | 338,12 | 0,00 | 0,04 | -0,01 | 9,93E-01 | 9,96E-01 |
| CYFIP1 | 975,57 | 0,00 | 0,04 | 0,01 | 9,93E-01 | 9,96E-01 |
| ARID2 | 2817,29 | 0,00 | 0,02 | -0,01 | 9,93E-01 | 9,96E-01 |
| RC3H2 | 4387,71 | 0,00 | 0,02 | -0,01 | 9,93E-01 | 9,96E-01 |
| ZNF75D | 1435,66 | 0,00 | 0,02 | -0,01 | 9,94E-01 | 9,97E-01 |
| RBBP7 | 1290,54 | 0,00 | 0,02 | 0,01 | 9,94E-01 | 9,97E-01 |
| HNRNPA3 | 4889,39 | 0,00 | 0,02 | -0,01 | 9,94E-01 | 9,97E-01 |
| GALK1 | 337,69 | 0,00 | 0,03 | 0,01 | 9,94E-01 | 9,97E-01 |
| TIMM10 | 238,86 | 0,00 | 0,04 | 0,01 | 9,94E-01 | 9,97E-01 |
| VPS35L | 782,91 | 0,00 | 0,02 | -0,01 | 9,94E-01 | 9,97E-01 |
| ITGB1 | 7779,73 | 0,00 | 0,04 | -0,01 | 9,95E-01 | 9,97E-01 |
| STK26 | 2243,84 | 0,00 | 0,03 | 0,01 | 9,95E-01 | 9,97E-01 |
| APOL3 | 4184,42 | 0,00 | 0,03 | 0,01 | 9,95E-01 | 9,97E-01 |
| PTPN18 | 1407,05 | 0,00 | 0,04 | -0,01 | 9,96E-01 | 9,98E-01 |
| SFMBT2 | 1524,14 | 0,00 | 0,02 | -0,01 | 9,96E-01 | 9,98E-01 |
| SLC35E2B | 1013,66 | 0,00 | 0,05 | 0,00 | 9,97E-01 | 9,98E-01 |
| SLC16A6 | 1199,86 | 0,00 | 0,03 | 0,00 | 9,97E-01 | 9,98E-01 |
| FLT1 | 136,76 | 0,00 | 0,08 | 0,00 | 9,97E-01 | 9,98E-01 |
| UBE2A | 861,54 | 0,00 | 0,03 | 0,00 | 9,97E-01 | 9,99E-01 |
| SEC31B | 296,64 | 0,00 | 0,06 | 0,00 | 9,97E-01 | 9,99E-01 |
| GCH1 | 1760,44 | 0,00 | 0,04 | 0,00 | 9,97E-01 | 9,99E-01 |
| CCT8 | 2184,36 | 0,00 | 0,03 | 0,00 | 9,98E-01 | 9,99E-01 |
| TIGIT | 1323,19 | 0,00 | 0,04 | 0,00 | 9,98E-01 | 9,99E-01 |
| ITPKC | 292,07 | 0,00 | 0,02 | 0,00 | 9,98E-01 | 9,99E-01 |
| SNRNP70 | 1797,92 | 0,00 | 0,04 | 0,00 | 9,98E-01 | 9,99E-01 |

| | | | | | | |
|---|---|---|---|---|---|---|
| SYVN1 | 359,60 | 0,00 | 0,04 | 0,00 | 9,99E-01 | 9,99E-01 |
| PINX1 | 155,98 | 0,00 | 0,02 | 0,00 | 9,99E-01 | 9,99E-01 |
| ZNF595 | 364,10 | 0,00 | 0,05 | 0,00 | 9,99E-01 | 9,99E-01 |
| HS3ST3B1 | 571,90 | 0,00 | 0,03 | 0,00 | 9,99E-01 | 1,00E+00 |
| ATXN2L | 703,86 | 0,00 | 0,06 | 0,00 | 9,99E-01 | 1,00E+00 |
| PHRF1 | 976,64 | 0,00 | 0,04 | 0,00 | 9,99E-01 | 1,00E+00 |
| SCLY | 206,61 | 0,00 | 0,04 | 0,00 | 9,99E-01 | 1,00E+00 |
| DCTN5 | 2089,86 | 0,00 | 0,02 | 0,00 | 1,00E+00 | 1,00E+00 |
| PRPS1 | 771,72 | 0,00 | 0,03 | 0,00 | 1,00E+00 | 1,00E+00 |
| TIFA | 782,21 | 0,00 | 0,04 | 0,00 | 1,00E+00 | 1,00E+00 |
| SAE1 | 823,95 | 0,00 | 0,02 | 0,00 | 1,00E+00 | 1,00E+00 |
| GTF3C5 | 723,80 | 0,00 | 0,02 | 0,00 | 1,00E+00 | 1,00E+00 |
| EXOC3L1 | 57,43 | -0,14 | 0,09 | -1,50 | 1,34E-01 | |
| MELK | 54,10 | 0,06 | 0,07 | 0,85 | 3,95E-01 | |
| POC1A | 63,88 | -0,03 | 0,04 | -0,82 | 4,14E-01 | |
| ENSG000002 | 38,46 | -0,07 | 0,03 | -1,93 | 5,33E-02 | |
| DDO | 40,33 | 0,08 | 0,07 | 1,21 | 2,26E-01 | |
| ICAM5 | 49,04 | 0,09 | 0,07 | 1,39 | 1,65E-01 | |
| ZNF415 | 43,30 | 0,01 | 0,08 | 0,12 | 9,05E-01 | |
| CCR8 | 52,29 | 0,01 | 0,06 | 0,12 | 9,02E-01 | |
| DEPTOR | 45,82 | 0,03 | 0,06 | 0,53 | 5,99E-01 | |
| CUX2 | 50,92 | -0,06 | 0,11 | -0,50 | 6,17E-01 | |
| AGAP5 | 58,86 | -0,03 | 0,03 | -0,99 | 3,24E-01 | |
| CORO6 | 36,92 | 0,05 | 0,07 | 0,65 | 5,16E-01 | |
| LRGUK | 42,76 | 0,02 | 0,09 | 0,18 | 8,58E-01 | |
| ORC6 | 55,05 | -0,03 | 0,05 | -0,66 | 5,11E-01 | |
| HES1 | 44,80 | 0,02 | 0,06 | 0,24 | 8,06E-01 | |
| CACHD1 | 67,49 | -0,02 | 0,09 | -0,25 | 8,04E-01 | |
| CCR9 | 45,80 | 0,09 | 0,06 | 1,46 | 1,45E-01 | |
| RPP21 | 52,42 | 0,01 | 0,03 | 0,42 | 6,77E-01 | |
| RRH | 28,95 | 0,01 | 0,04 | 0,12 | 9,08E-01 | |
| PCDHGB2 | 31,74 | -0,05 | 0,06 | -0,76 | 4,45E-01 | |
| SAC3D1 | 55,19 | 0,05 | 0,04 | 1,44 | 1,50E-01 | |
| NRN1 | 45,34 | -0,25 | 0,11 | -2,34 | 1,94E-02 | |
| IFTAP | 36,02 | 0,05 | 0,05 | 0,97 | 3,33E-01 | |
| RPA4 | 62,99 | 0,04 | 0,03 | 1,17 | 2,43E-01 | |
| PTPRG | 68,34 | 0,11 | 0,12 | 0,87 | 3,83E-01 | |
| PPARGC1A | 41,12 | -0,02 | 0,13 | -0,15 | 8,82E-01 | |
| KIR3DL2 | 33,04 | -0,07 | 0,09 | -0,82 | 4,13E-01 | |
| TRPV1 | 69,28 | 0,01 | 0,06 | 0,10 | 9,19E-01 | |
| TMEM236 | 26,80 | 0,01 | 0,07 | 0,10 | 9,22E-01 | |
| EPS8L1 | 58,65 | -0,05 | 0,10 | -0,47 | 6,35E-01 | |
| KIF2C | 33,29 | 0,09 | 0,09 | 1,06 | 2,88E-01 | |
| LRRC56 | 47,27 | 0,15 | 0,06 | 2,41 | 1,59E-02 | |
| ZNF521 | 40,01 | -0,04 | 0,10 | -0,43 | 6,64E-01 | |
| UQCRHL | 41,76 | 0,03 | 0,04 | 0,59 | 5,56E-01 | |

| | | | | | |
|---|---|---|---|---|---|
| AMN | 57,48 | -0,01 | 0,09 | -0,10 | 9,18E-01 |
| WNT10A | 49,55 | 0,03 | 0,07 | 0,44 | 6,57E-01 |
| SIGLEC15 | 50,77 | 0,03 | 0,07 | 0,48 | 6,32E-01 |
| MOSPD3 | 48,77 | 0,02 | 0,04 | 0,63 | 5,28E-01 |
| ANKRD31 | 62,13 | 0,06 | 0,06 | 1,01 | 3,12E-01 |
| NOP2 | 57,20 | 0,02 | 0,03 | 0,59 | 5,56E-01 |
| STX1A | 38,03 | 0,05 | 0,08 | 0,64 | 5,22E-01 |
| PRSS53 | 58,89 | 0,04 | 0,05 | 0,88 | 3,80E-01 |
| LAT | 64,18 | 0,08 | 0,07 | 1,16 | 2,45E-01 |
| CLCN2 | 38,19 | 0,03 | 0,10 | 0,30 | 7,62E-01 |
| RAB15 | 42,20 | 0,00 | 0,07 | 0,06 | 9,53E-01 |
| C17orf107 | 31,59 | 0,12 | 0,08 | 1,54 | 1,24E-01 |
| APBA1 | 67,21 | 0,01 | 0,09 | 0,15 | 8,81E-01 |
| NT5DC4 | 35,27 | -0,13 | 0,08 | -1,53 | 1,27E-01 |
| NUP62CL | 38,63 | 0,18 | 0,06 | 3,02 | 2,49E-03 |
| SMAD6 | 59,17 | -0,04 | 0,12 | -0,31 | 7,59E-01 |
| PROSER2 | 36,73 | 0,06 | 0,08 | 0,70 | 4,83E-01 |
| MCOLN3 | 44,19 | 0,05 | 0,09 | 0,55 | 5,79E-01 |
| DDR1 | 68,89 | 0,05 | 0,09 | 0,58 | 5,64E-01 |
| PLSCR2 | 53,16 | -0,16 | 0,06 | -2,50 | 1,26E-02 |
| FAM171B | 27,67 | 0,09 | 0,07 | 1,22 | 2,24E-01 |
| C9orf43 | 26,48 | 0,10 | 0,07 | 1,46 | 1,45E-01 |
| PLA2G4B | 53,78 | -0,02 | 0,05 | -0,44 | 6,63E-01 |
| ZNRF3 | 51,01 | -0,02 | 0,09 | -0,21 | 8,32E-01 |
| GINS2 | 34,42 | -0,09 | 0,07 | -1,37 | 1,71E-01 |
| YPEL4 | 45,80 | 0,09 | 0,07 | 1,31 | 1,91E-01 |
| JAKMIP3 | 50,36 | -0,09 | 0,11 | -0,78 | 4,38E-01 |
| ZNF112 | 28,14 | 0,11 | 0,08 | 1,39 | 1,63E-01 |
| AK4 | 49,25 | 0,00 | 0,08 | -0,01 | 9,95E-01 |
| GNA11 | 61,55 | 0,08 | 0,07 | 1,11 | 2,66E-01 |
| RUFY4 | 63,61 | -0,07 | 0,08 | -0,94 | 3,47E-01 |
| JAKMIP1 | 46,24 | 0,06 | 0,06 | 0,88 | 3,79E-01 |
| ZFP92 | 56,58 | -0,09 | 0,06 | -1,36 | 1,73E-01 |
| NPIPA1 | 52,28 | 0,06 | 0,06 | 1,09 | 2,78E-01 |
| ACSM1 | 38,17 | 0,01 | 0,08 | 0,14 | 8,90E-01 |
| CKB | 63,72 | 0,09 | 0,06 | 1,63 | 1,03E-01 |
| MLF1 | 26,72 | 0,04 | 0,08 | 0,46 | 6,46E-01 |
| EPDR1 | 38,07 | -0,04 | 0,06 | -0,63 | 5,29E-01 |
| ALDH8A1 | 71,14 | 0,01 | 0,05 | 0,11 | 9,09E-01 |
| B3GALNT1 | 47,16 | 0,01 | 0,07 | 0,13 | 8,99E-01 |
| DRC1 | 40,64 | -0,11 | 0,09 | -1,19 | 2,34E-01 |
| KCNK13 | 36,80 | 0,01 | 0,06 | 0,11 | 9,10E-01 |
| DNAH12 | 62,54 | 0,02 | 0,13 | 0,13 | 8,96E-01 |
| CLDN23 | 40,13 | -0,08 | 0,07 | -1,18 | 2,39E-01 |
| MARVELD1 | 46,78 | 0,01 | 0,08 | 0,08 | 9,38E-01 |
| SH3PXD2B | 41,15 | -0,12 | 0,11 | -1,06 | 2,89E-01 |

| | | | | | |
|---|---|---|---|---|---|
| SERF1B | 45,06 | -0,05 | 0,05 | -1,18 | 2,38E-01 |
| AHRR | 49,74 | -0,03 | 0,13 | -0,23 | 8,21E-01 |
| SDCBP2 | 34,10 | -0,10 | 0,06 | -1,63 | 1,03E-01 |
| RNFT2 | 64,06 | 0,02 | 0,07 | 0,28 | 7,77E-01 |
| SV2A | 40,48 | 0,10 | 0,07 | 1,35 | 1,77E-01 |
| UNC45B | 51,30 | 0,03 | 0,08 | 0,39 | 6,97E-01 |
| ARG2 | 41,40 | 0,03 | 0,07 | 0,42 | 6,72E-01 |
| CNBD2 | 56,97 | 0,03 | 0,06 | 0,51 | 6,10E-01 |
| AARSD1 | 68,49 | 0,08 | 0,04 | 2,24 | 2,53E-02 |
| NTN4 | 49,15 | -0,02 | 0,09 | -0,24 | 8,13E-01 |
| SLC25A34 | 30,15 | -0,03 | 0,07 | -0,50 | 6,18E-01 |
| LRRC63 | 31,39 | 0,08 | 0,05 | 1,47 | 1,41E-01 |
| TRAPPC3L | 43,99 | -0,01 | 0,08 | -0,13 | 8,94E-01 |
| DYTN | 33,17 | 0,07 | 0,08 | 0,91 | 3,64E-01 |
| PGBD1 | 58,29 | 0,02 | 0,04 | 0,50 | 6,20E-01 |
| TRIM47 | 43,37 | 0,08 | 0,05 | 1,55 | 1,20E-01 |
| TTC23 | 50,96 | 0,09 | 0,08 | 1,18 | 2,38E-01 |
| NTRK1 | 39,51 | -0,10 | 0,12 | -0,81 | 4,20E-01 |
| KLHDC7B | 53,27 | 0,06 | 0,07 | 0,84 | 4,03E-01 |
| SAMD10 | 49,85 | 0,08 | 0,06 | 1,26 | 2,07E-01 |
| C6orf52 | 29,00 | 0,07 | 0,05 | 1,32 | 1,88E-01 |
| C3orf33 | 31,21 | 0,01 | 0,05 | 0,19 | 8,52E-01 |
| PCLAF | 27,09 | 0,12 | 0,07 | 1,88 | 6,06E-02 |
| PLLP | 31,15 | 0,07 | 0,10 | 0,72 | 4,69E-01 |
| PODXL2 | 40,17 | 0,00 | 0,06 | 0,08 | 9,39E-01 |
| RPL26L1 | 68,86 | 0,08 | 0,07 | 1,26 | 2,07E-01 |
| GOLGA8R | 58,94 | -0,10 | 0,06 | -1,72 | 8,51E-02 |
| MAP2 | 45,92 | -0,09 | 0,10 | -0,83 | 4,05E-01 |
| ACKR2 | 36,45 | 0,08 | 0,09 | 0,96 | 3,35E-01 |
| NIPAL1 | 55,24 | -0,13 | 0,08 | -1,57 | 1,17E-01 |
| RAB38 | 30,10 | 0,00 | 0,06 | 0,08 | 9,33E-01 |
| LYSMD1 | 42,87 | 0,01 | 0,04 | 0,18 | 8,57E-01 |
| PLPP7 | 34,23 | -0,03 | 0,08 | -0,37 | 7,10E-01 |
| ASNS | 68,38 | 0,01 | 0,04 | 0,22 | 8,29E-01 |
| CACNG6 | 47,83 | 0,13 | 0,09 | 1,42 | 1,55E-01 |
| RASL11A | 65,73 | 0,00 | 0,05 | 0,07 | 9,44E-01 |
| TLCD2 | 55,18 | 0,14 | 0,08 | 1,92 | 5,48E-02 |
| C1orf56 | 31,85 | 0,09 | 0,04 | 2,13 | 3,28E-02 |
| HMCN1 | 66,19 | 0,00 | 0,11 | -0,03 | 9,74E-01 |
| REDIC1 | 34,84 | 0,09 | 0,07 | 1,22 | 2,24E-01 |
| GPRASP2 | 65,46 | 0,03 | 0,04 | 0,66 | 5,11E-01 |
| PCDHGA9 | 34,37 | -0,07 | 0,07 | -1,04 | 2,99E-01 |
| ENSG000002 | 21,39 | 0,01 | 0,06 | 0,27 | 7,89E-01 |
| LY6G6F | 26,27 | 0,06 | 0,07 | 0,82 | 4,10E-01 |
| GOLGA6L10 | 45,07 | 0,09 | 0,10 | 0,94 | 3,49E-01 |
| SMIM10L2A | 37,24 | 0,01 | 0,07 | 0,16 | 8,75E-01 |

| | | | | | |
|---|---|---|---|---|---|
| ADGRA2 | 69,38 | 0,12 | 0,08 | 1,43 | 1,52E-01 |
| PRCD | 49,15 | 0,02 | 0,08 | 0,20 | 8,43E-01 |
| MMP17 | 59,27 | 0,10 | 0,10 | 1,05 | 2,92E-01 |
| BCL2L12 | 59,15 | -0,01 | 0,03 | -0,24 | 8,09E-01 |
| ANKRD34B | 69,85 | 0,06 | 0,09 | 0,71 | 4,80E-01 |
| CMYA5 | 71,18 | -0,02 | 0,08 | -0,21 | 8,33E-01 |
| SLC9A5 | 35,56 | 0,04 | 0,09 | 0,46 | 6,47E-01 |
| PRTFDC1 | 35,99 | 0,04 | 0,06 | 0,63 | 5,31E-01 |
| B3GNT7 | 34,76 | -0,05 | 0,09 | -0,62 | 5,35E-01 |
| PTK6 | 39,50 | 0,08 | 0,07 | 1,16 | 2,48E-01 |
| SGPP2 | 38,81 | 0,09 | 0,07 | 1,27 | 2,03E-01 |
| GOLGA8H | 33,00 | -0,05 | 0,06 | -0,75 | 4,54E-01 |
| BMAL2 | 66,80 | 0,12 | 0,07 | 1,75 | 8,09E-02 |
| RHCE | 57,67 | 0,05 | 0,08 | 0,65 | 5,17E-01 |
| CHST14 | 71,22 | 0,04 | 0,05 | 0,82 | 4,15E-01 |
| FGF13 | 66,20 | -0,22 | 0,12 | -1,88 | 5,95E-02 |
| GARIN1A | 59,78 | -0,10 | 0,07 | -1,29 | 1,97E-01 |
| NQO1 | 41,01 | 0,06 | 0,05 | 1,36 | 1,73E-01 |
| MINAR1 | 63,59 | 0,02 | 0,07 | 0,23 | 8,17E-01 |
| SERPINE2 | 53,58 | 0,02 | 0,09 | 0,28 | 7,78E-01 |
| CKS2 | 64,91 | 0,07 | 0,06 | 1,05 | 2,95E-01 |
| TNFRSF11A | 69,72 | 0,02 | 0,09 | 0,26 | 7,96E-01 |
| PCYT1B | 47,11 | 0,01 | 0,09 | 0,11 | 9,10E-01 |
| MAMDC4 | 46,99 | 0,08 | 0,09 | 0,84 | 3,98E-01 |
| HYAL3 | 45,03 | 0,19 | 0,05 | 3,90 | 9,42E-05 |
| RNF152 | 58,58 | 0,05 | 0,10 | 0,53 | 5,98E-01 |
| METTL1 | 45,50 | 0,05 | 0,04 | 1,23 | 2,18E-01 |
| B9D1 | 49,07 | 0,05 | 0,05 | 0,95 | 3,41E-01 |
| TRIM34 | 43,83 | -0,10 | 0,04 | -2,81 | 4,91E-03 |
| SPNS2 | 53,69 | -0,07 | 0,08 | -0,96 | 3,35E-01 |
| ZNF771 | 61,30 | 0,07 | 0,04 | 1,75 | 8,02E-02 |
| IFNG | 39,55 | 0,22 | 0,09 | 2,61 | 9,14E-03 |
| CCDC163 | 36,72 | 0,13 | 0,09 | 1,50 | 1,34E-01 |
| HCN3 | 35,56 | 0,05 | 0,06 | 0,87 | 3,83E-01 |
| AGBL2 | 64,59 | 0,05 | 0,05 | 1,05 | 2,95E-01 |
| POLR3G | 44,18 | 0,07 | 0,05 | 1,39 | 1,64E-01 |
| CLEC18A | 68,01 | -0,23 | 0,09 | -2,51 | 1,22E-02 |
| ARHGEF17 | 51,89 | 0,01 | 0,13 | 0,09 | 9,27E-01 |
| RFPL2 | 37,41 | -0,03 | 0,08 | -0,38 | 7,06E-01 |
| RIBC1 | 25,46 | -0,08 | 0,06 | -1,33 | 1,85E-01 |
| LYNX1 | 65,32 | 0,11 | 0,07 | 1,43 | 1,53E-01 |
| WNT2B | 61,09 | 0,03 | 0,10 | 0,31 | 7,57E-01 |
| LY6G5C | 30,17 | 0,02 | 0,06 | 0,38 | 7,01E-01 |
| PTX3 | 50,30 | 0,02 | 0,05 | 0,30 | 7,68E-01 |
| FBXL16 | 45,58 | 0,11 | 0,08 | 1,40 | 1,63E-01 |
| EVPL | 66,04 | 0,27 | 0,10 | 2,67 | 7,47E-03 |

| | | | | | |
|---|---|---|---|---|---|
| NAPSA | 35,10 | 0,04 | 0,08 | 0,53 | 5,99E-01 |
| LYG1 | 44,07 | -0,05 | 0,04 | -1,35 | 1,76E-01 |
| TGM1 | 47,42 | -0,05 | 0,07 | -0,69 | 4,91E-01 |
| FAM241B | 28,35 | 0,00 | 0,07 | 0,02 | 9,86E-01 |
| MPIG6B | 57,78 | 0,18 | 0,11 | 1,67 | 9,46E-02 |
| FPGT-TNNI3K | 42,27 | 0,13 | 0,06 | 2,30 | 2,14E-02 |
| SHCBP1 | 60,79 | -0,04 | 0,06 | -0,57 | 5,68E-01 |
| HEXIM2 | 69,77 | 0,01 | 0,04 | 0,35 | 7,30E-01 |
| ACE | 54,54 | 0,02 | 0,09 | 0,23 | 8,20E-01 |
| ABO | 59,22 | -0,04 | 0,11 | -0,37 | 7,08E-01 |
| WWTR1 | 50,51 | -0,01 | 0,08 | -0,11 | 9,14E-01 |
| ORC1 | 39,35 | 0,00 | 0,07 | -0,06 | 9,54E-01 |
| C1orf116 | 42,28 | 0,19 | 0,08 | 2,26 | 2,39E-02 |
| SPATA1 | 63,65 | 0,00 | 0,05 | 0,05 | 9,63E-01 |
| ITGB4 | 48,89 | 0,04 | 0,11 | 0,36 | 7,16E-01 |
| ZBED6 | 45,11 | -0,01 | 0,04 | -0,16 | 8,74E-01 |
| RAB19 | 22,58 | -0,16 | 0,05 | -3,54 | 4,02E-04 |
| ARHGEF5 | 57,70 | 0,07 | 0,07 | 1,04 | 3,00E-01 |
| ENSG000002 | 27,44 | 0,01 | 0,03 | 0,22 | 8,28E-01 |
| RNF32 | 42,44 | 0,09 | 0,08 | 1,13 | 2,59E-01 |
| NHEJ1 | 59,56 | -0,06 | 0,04 | -1,51 | 1,30E-01 |
| ATP2C2 | 53,50 | -0,04 | 0,12 | -0,34 | 7,30E-01 |
| EIF4EBP3 | 24,58 | -0,05 | 0,05 | -1,08 | 2,81E-01 |
| GPT2 | 54,92 | -0,07 | 0,10 | -0,76 | 4,45E-01 |
| DTL | 57,57 | 0,14 | 0,07 | 1,98 | 4,80E-02 |
| CYB5D1 | 44,38 | 0,00 | 0,03 | 0,08 | 9,38E-01 |
| PARS2 | 52,66 | 0,07 | 0,03 | 2,03 | 4,25E-02 |
| P3H3 | 38,45 | 0,18 | 0,08 | 2,44 | 1,46E-02 |
| FBXW9 | 54,92 | 0,09 | 0,04 | 2,43 | 1,50E-02 |
| C1QTNF9 | 39,01 | -0,04 | 0,04 | -1,19 | 2,33E-01 |
| LDHD | 36,75 | 0,03 | 0,08 | 0,43 | 6,70E-01 |
| HTD2 | 21,81 | 0,01 | 0,03 | 0,18 | 8,61E-01 |
| LIPC | 58,86 | 0,10 | 0,08 | 1,26 | 2,07E-01 |
| PLAAT5 | 43,43 | 0,00 | 0,08 | -0,04 | 9,71E-01 |
| PAQR6 | 67,31 | -0,01 | 0,07 | -0,19 | 8,50E-01 |
| COPZ2 | 30,97 | 0,00 | 0,06 | 0,08 | 9,35E-01 |
| TSSK4 | 24,40 | -0,04 | 0,05 | -0,71 | 4,80E-01 |
| UMODL1 | 35,51 | 0,07 | 0,10 | 0,70 | 4,82E-01 |
| PDCD1LG2 | 40,07 | -0,12 | 0,08 | -1,44 | 1,51E-01 |
| UTS2 | 40,56 | 0,12 | 0,11 | 1,11 | 2,68E-01 |
| SULT1A2 | 38,86 | -0,11 | 0,08 | -1,30 | 1,94E-01 |
| MACROH2A2 | 60,70 | 0,07 | 0,04 | 2,05 | 4,00E-02 |
| KIFC1 | 33,15 | 0,00 | 0,08 | -0,01 | 9,96E-01 |
| MCF2 | 60,58 | -0,04 | 0,05 | -0,87 | 3,86E-01 |
| PHKG1 | 40,18 | 0,02 | 0,05 | 0,35 | 7,23E-01 |
| PEAR1 | 53,32 | 0,08 | 0,11 | 0,70 | 4,82E-01 |

| | | | | | |
|---|---|---|---|---|---|
| INTU | 54,08 | 0,04 | 0,12 | 0,31 | 7,59E-01 |
| RGPD1 | 35,70 | -0,18 | 0,09 | -1,89 | 5,85E-02 |
| C17orf49 | 38,58 | 0,05 | 0,04 | 1,13 | 2,58E-01 |
| NR4A1 | 60,63 | 0,12 | 0,09 | 1,29 | 1,97E-01 |
| CLEC11A | 58,25 | 0,00 | 0,07 | 0,04 | 9,69E-01 |
| TNFRSF21 | 43,95 | -0,05 | 0,07 | -0,66 | 5,11E-01 |
| EEF1AKMT3 | 47,89 | 0,03 | 0,05 | 0,74 | 4,57E-01 |
| SFRP2 | 38,81 | 0,18 | 0,09 | 2,03 | 4,19E-02 |
| HMMR | 33,98 | 0,10 | 0,09 | 1,11 | 2,66E-01 |
| SEMA3G | 34,25 | 0,02 | 0,10 | 0,24 | 8,11E-01 |
| RAD9B | 35,72 | 0,06 | 0,04 | 1,38 | 1,68E-01 |
| KCNK10 | 45,25 | -0,06 | 0,12 | -0,51 | 6,10E-01 |
| OR2L2 | 25,45 | 0,11 | 0,09 | 1,15 | 2,49E-01 |
| FAM110C | 52,94 | -0,03 | 0,07 | -0,45 | 6,50E-01 |
| F12 | 33,58 | 0,00 | 0,07 | -0,03 | 9,78E-01 |
| KIF4A | 40,63 | 0,04 | 0,08 | 0,46 | 6,44E-01 |
| ZNF837 | 60,78 | 0,10 | 0,05 | 2,13 | 3,31E-02 |
| SLC13A3 | 44,37 | -0,03 | 0,10 | -0,30 | 7,61E-01 |
| THNSL2 | 67,99 | -0,03 | 0,11 | -0,32 | 7,47E-01 |
| ZNF660 | 68,77 | 0,01 | 0,05 | 0,30 | 7,63E-01 |
| GAS2 | 52,98 | 0,08 | 0,05 | 1,54 | 1,24E-01 |
| GAL3ST4 | 32,12 | 0,09 | 0,08 | 1,17 | 2,41E-01 |
| ALDOC | 60,69 | -0,01 | 0,04 | -0,20 | 8,42E-01 |
| ITGA7 | 47,35 | 0,00 | 0,10 | 0,02 | 9,85E-01 |
| TMPRSS13 | 50,67 | 0,02 | 0,07 | 0,32 | 7,49E-01 |
| WDR31 | 34,14 | 0,08 | 0,08 | 1,08 | 2,79E-01 |
| NCAPH | 64,64 | 0,08 | 0,05 | 1,56 | 1,20E-01 |
| PPP1R13L | 34,70 | 0,02 | 0,10 | 0,24 | 8,12E-01 |
| MMACHC | 58,15 | 0,04 | 0,04 | 0,86 | 3,92E-01 |
| NEURL2 | 36,69 | 0,13 | 0,04 | 3,30 | 9,76E-04 |
| STPG4 | 33,96 | -0,03 | 0,05 | -0,50 | 6,15E-01 |
| HEATR9 | 42,24 | 0,01 | 0,08 | 0,07 | 9,40E-01 |
| THSD1 | 46,29 | 0,00 | 0,06 | 0,09 | 9,32E-01 |
| ARMC3 | 52,28 | 0,04 | 0,09 | 0,44 | 6,62E-01 |
| NAGS | 58,33 | 0,01 | 0,04 | 0,20 | 8,40E-01 |
| SAXO2 | 61,46 | -0,16 | 0,12 | -1,32 | 1,88E-01 |
| PRSS57 | 33,03 | 0,02 | 0,08 | 0,21 | 8,33E-01 |
| ADGRB3 | 43,86 | 0,13 | 0,10 | 1,29 | 1,99E-01 |
| RGPD3 | 34,24 | -0,03 | 0,08 | -0,35 | 7,23E-01 |
| RGS17 | 33,52 | 0,11 | 0,10 | 1,13 | 2,59E-01 |
| BBS1 | 56,73 | 0,02 | 0,04 | 0,54 | 5,91E-01 |
| CXXC4 | 27,40 | 0,17 | 0,08 | 1,97 | 4,83E-02 |
| ELOVL4 | 55,48 | 0,07 | 0,07 | 1,07 | 2,85E-01 |
| GOLGA8O | 34,69 | -0,07 | 0,08 | -0,90 | 3,69E-01 |
| THRB | 61,05 | -0,03 | 0,10 | -0,35 | 7,26E-01 |
| EDARADD | 55,87 | 0,03 | 0,06 | 0,43 | 6,69E-01 |

| | | | | | |
|---|---|---|---|---|---|
| ELF3 | 39,94 | -0,03 | 0,08 | -0,41 | 6,82E-01 |
| MAP1LC3B2 | 53,40 | -0,10 | 0,06 | -1,59 | 1,12E-01 |
| GNMT | 31,87 | -0,09 | 0,06 | -1,38 | 1,66E-01 |
| SPICE1-CFAI | 24,66 | -0,01 | 0,05 | -0,27 | 7,85E-01 |
| RAB40B | 41,66 | 0,01 | 0,07 | 0,12 | 9,07E-01 |
| LRRC7 | 65,04 | 0,03 | 0,12 | 0,26 | 7,95E-01 |
| HINT2 | 47,37 | 0,10 | 0,04 | 2,61 | 9,10E-03 |
| FBXO15 | 63,40 | 0,07 | 0,08 | 0,78 | 4,38E-01 |
| ADSS1 | 41,85 | 0,03 | 0,08 | 0,39 | 7,00E-01 |
| UCN | 32,55 | 0,03 | 0,04 | 0,73 | 4,62E-01 |
| ZDHHC9 | 51,63 | 0,03 | 0,05 | 0,65 | 5,16E-01 |
| ZNF57 | 65,67 | 0,06 | 0,03 | 2,13 | 3,29E-02 |
| MFSD4A | 34,33 | 0,05 | 0,08 | 0,59 | 5,58E-01 |
| C19orf44 | 45,11 | 0,06 | 0,06 | 1,00 | 3,16E-01 |
| SORBS2 | 61,89 | -0,08 | 0,14 | -0,56 | 5,73E-01 |
| ASF1B | 65,27 | -0,08 | 0,06 | -1,18 | 2,37E-01 |
| ADAM32 | 40,85 | 0,07 | 0,11 | 0,66 | 5,11E-01 |
| ENAH | 46,13 | 0,05 | 0,12 | 0,40 | 6,91E-01 |
| TREX2 | 25,69 | 0,06 | 0,06 | 0,99 | 3,21E-01 |
| ASAP3 | 43,58 | 0,03 | 0,10 | 0,28 | 7,78E-01 |
| ENSG000002 | 24,86 | -0,01 | 0,04 | -0,13 | 8,96E-01 |
| MBLAC1 | 65,04 | 0,05 | 0,03 | 1,47 | 1,41E-01 |
| AMZ1 | 57,81 | 0,05 | 0,09 | 0,60 | 5,51E-01 |
| ZC3H12B | 63,76 | 0,02 | 0,05 | 0,34 | 7,35E-01 |
| PLSCR4 | 37,01 | 0,13 | 0,08 | 1,71 | 8,81E-02 |
| HSPA2 | 34,76 | 0,00 | 0,07 | 0,04 | 9,65E-01 |
| ZDHHC1 | 52,88 | 0,14 | 0,06 | 2,22 | 2,62E-02 |
| ENSG000002 | 63,62 | 0,02 | 0,04 | 0,57 | 5,68E-01 |
| LMTK3 | 63,01 | 0,09 | 0,09 | 0,97 | 3,33E-01 |
| RRM2 | 71,23 | 0,02 | 0,06 | 0,30 | 7,61E-01 |
| ENSG000002 | 40,22 | -0,06 | 0,03 | -1,79 | 7,37E-02 |
| PRRT3 | 58,42 | 0,00 | 0,06 | 0,04 | 9,67E-01 |
| C17orf113 | 33,25 | 0,08 | 0,07 | 1,15 | 2,48E-01 |
| SEMA6C | 44,31 | 0,09 | 0,09 | 1,01 | 3,13E-01 |
| ERG | 51,40 | 0,11 | 0,11 | 0,99 | 3,20E-01 |
| CTSG | 37,21 | 0,15 | 0,11 | 1,40 | 1,62E-01 |
| ENSG000002 | 22,38 | 0,14 | 0,05 | 3,07 | 2,13E-03 |
| C17orf100 | 25,45 | 0,00 | 0,06 | -0,02 | 9,84E-01 |
| LRFN2 | 35,44 | -0,16 | 0,11 | -1,46 | 1,45E-01 |
| ZNF683 | 66,26 | 0,13 | 0,12 | 1,05 | 2,91E-01 |
| KIF24 | 57,82 | 0,03 | 0,05 | 0,70 | 4,87E-01 |
| FMC1 | 51,62 | 0,05 | 0,03 | 1,57 | 1,17E-01 |
| SLC2A8 | 66,58 | 0,04 | 0,06 | 0,57 | 5,70E-01 |
| SLC16A14 | 58,32 | 0,04 | 0,08 | 0,49 | 6,27E-01 |
| CCDC89 | 57,77 | -0,01 | 0,05 | -0,25 | 8,04E-01 |
| CADM4 | 70,28 | -0,16 | 0,05 | -2,86 | 4,18E-03 |

| | | | | | |
|---|---|---|---|---|---|
| GRIN3A | 39,42 | -0,10 | 0,10 | -1,04 | 2,98E-01 |
| LRP1B | 47,23 | 0,12 | 0,11 | 1,02 | 3,05E-01 |
| TMEM97 | 69,86 | 0,01 | 0,04 | 0,13 | 8,95E-01 |
| GREM2 | 34,28 | 0,07 | 0,08 | 0,88 | 3,79E-01 |
| DOK4 | 70,64 | -0,14 | 0,07 | -1,98 | 4,72E-02 |
| AZU1 | 34,63 | 0,21 | 0,09 | 2,37 | 1,76E-02 |
| CPT1B | 54,56 | -0,03 | 0,06 | -0,43 | 6,67E-01 |
| TMEM88 | 44,50 | -0,15 | 0,07 | -2,01 | 4,40E-02 |
| RNASE3 | 35,51 | 0,31 | 0,10 | 2,96 | 3,03E-03 |
| STEAP3 | 48,62 | 0,09 | 0,08 | 1,06 | 2,90E-01 |
| SHROOM4 | 40,14 | -0,07 | 0,10 | -0,69 | 4,91E-01 |
| BCAS1 | 37,37 | 0,04 | 0,11 | 0,36 | 7,19E-01 |
| CCDC24 | 55,27 | -0,04 | 0,06 | -0,65 | 5,16E-01 |
| RAI14 | 43,08 | -0,04 | 0,12 | -0,30 | 7,65E-01 |
| ZG16B | 25,64 | 0,18 | 0,07 | 2,77 | 5,68E-03 |
| KRTCAP3 | 41,15 | -0,04 | 0,05 | -0,68 | 4,95E-01 |
| CDS1 | 31,47 | 0,10 | 0,07 | 1,40 | 1,61E-01 |
| ARHGAP33 | 53,30 | 0,10 | 0,09 | 1,15 | 2,49E-01 |
| PLGLB2 | 51,48 | 0,01 | 0,10 | 0,13 | 8,96E-01 |
| NAP1L5 | 37,82 | 0,03 | 0,05 | 0,62 | 5,34E-01 |
| MRC1 | 65,29 | 0,00 | 0,09 | -0,02 | 9,84E-01 |
| DNAI2 | 38,59 | 0,01 | 0,08 | 0,13 | 8,99E-01 |
| COL11A2 | 47,81 | 0,09 | 0,12 | 0,70 | 4,84E-01 |
| CABLES2 | 64,31 | -0,05 | 0,06 | -0,72 | 4,69E-01 |
| FXYD2 | 37,36 | 0,13 | 0,08 | 1,51 | 1,30E-01 |
| CMTM8 | 25,49 | -0,03 | 0,07 | -0,40 | 6,86E-01 |
| MROH8 | 58,93 | 0,08 | 0,05 | 1,55 | 1,20E-01 |
| KCNG1 | 61,90 | 0,02 | 0,11 | 0,23 | 8,21E-01 |
| PXDNL | 55,63 | -0,11 | 0,06 | -1,75 | 8,00E-02 |
| SGIP1 | 53,42 | 0,14 | 0,11 | 1,25 | 2,10E-01 |
| ISM1 | 53,30 | 0,14 | 0,07 | 2,03 | 4,22E-02 |
| TNNT3 | 69,38 | -0,07 | 0,07 | -0,99 | 3,20E-01 |
| SMIM35 | 46,92 | 0,08 | 0,07 | 1,04 | 2,99E-01 |
| ZNF205 | 26,91 | 0,04 | 0,09 | 0,52 | 6,06E-01 |
| TRIM46 | 52,73 | 0,14 | 0,07 | 1,93 | 5,33E-02 |
| CHST13 | 43,00 | 0,05 | 0,09 | 0,59 | 5,58E-01 |
| SEZ6L | 71,57 | 0,04 | 0,09 | 0,40 | 6,89E-01 |
| RECQL4 | 41,38 | 0,12 | 0,08 | 1,53 | 1,26E-01 |
| RGS1 | 63,56 | 0,12 | 0,08 | 1,53 | 1,27E-01 |
| S100B | 62,77 | 0,06 | 0,13 | 0,44 | 6,59E-01 |
| PRAF2 | 34,21 | -0,06 | 0,05 | -1,14 | 2,55E-01 |
| SLC7A8 | 39,21 | -0,10 | 0,10 | -0,97 | 3,32E-01 |
| KLKB1 | 67,32 | 0,05 | 0,06 | 0,83 | 4,06E-01 |
| LSM2 | 50,93 | 0,07 | 0,03 | 2,24 | 2,50E-02 |
| SHROOM1 | 63,46 | 0,02 | 0,09 | 0,25 | 8,00E-01 |
| SENP8 | 33,77 | 0,08 | 0,06 | 1,26 | 2,08E-01 |

| | | | | | |
|---|---|---|---|---|---|
| EEF1AKMT4 | 22,76 | 0,06 | 0,04 | 1,53 | 1,26E-01 |
| PNLDC1 | 38,71 | -0,02 | 0,06 | -0,31 | 7,60E-01 |
| GAMT | 43,88 | 0,11 | 0,05 | 2,20 | 2,76E-02 |
| CHRFAM7A | 24,58 | 0,00 | 0,08 | 0,04 | 9,71E-01 |
| MTCP1 | 53,83 | 0,05 | 0,03 | 1,84 | 6,59E-02 |
| ITIH4 | 27,66 | -0,01 | 0,06 | -0,14 | 8,90E-01 |
| LINGO3 | 49,00 | -0,03 | 0,05 | -0,65 | 5,15E-01 |
| ENTREP3 | 45,50 | 0,04 | 0,08 | 0,57 | 5,71E-01 |
| ZNF80 | 56,12 | 0,04 | 0,08 | 0,50 | 6,19E-01 |
| SPATA25 | 25,23 | 0,08 | 0,04 | 1,97 | 4,90E-02 |
| FAM83E | 36,36 | 0,03 | 0,06 | 0,45 | 6,55E-01 |
| KHK | 63,91 | 0,02 | 0,04 | 0,47 | 6,36E-01 |
| DERL3 | 37,15 | 0,06 | 0,08 | 0,80 | 4,26E-01 |
| ENSG000002 | 53,86 | -0,04 | 0,03 | -1,11 | 2,68E-01 |
| CRIP3 | 30,77 | 0,10 | 0,07 | 1,52 | 1,29E-01 |
| OSBPL6 | 46,61 | -0,13 | 0,11 | -1,18 | 2,36E-01 |
| FBXO2 | 37,72 | -0,01 | 0,09 | -0,06 | 9,52E-01 |
| FKBPL | 59,99 | -0,02 | 0,03 | -0,55 | 5,85E-01 |
| CCDC62 | 58,72 | 0,00 | 0,04 | -0,08 | 9,36E-01 |
| FAM209A | 27,25 | -0,14 | 0,05 | -2,72 | 6,48E-03 |
| EXO1 | 39,47 | 0,02 | 0,08 | 0,23 | 8,19E-01 |
| MAP3K15 | 41,75 | -0,16 | 0,10 | -1,69 | 9,14E-02 |
| TONSL | 70,75 | 0,06 | 0,07 | 0,83 | 4,06E-01 |
| SEMA6B | 36,84 | 0,06 | 0,10 | 0,63 | 5,28E-01 |
| DDTL | 29,92 | -0,07 | 0,11 | -0,63 | 5,29E-01 |
| RAG1 | 51,28 | 0,04 | 0,07 | 0,54 | 5,88E-01 |
| FAM53A | 40,59 | -0,03 | 0,08 | -0,32 | 7,49E-01 |
| PKP2 | 54,80 | 0,04 | 0,08 | 0,49 | 6,22E-01 |
| SLITRK5 | 46,70 | 0,18 | 0,09 | 2,06 | 3,95E-02 |
| COL5A2 | 46,78 | 0,07 | 0,08 | 0,85 | 3,98E-01 |
| MRPL38 | 35,82 | 0,02 | 0,06 | 0,37 | 7,09E-01 |
| EREG | 67,94 | -0,13 | 0,09 | -1,46 | 1,44E-01 |
| HTRA4 | 60,00 | 0,00 | 0,04 | 0,02 | 9,84E-01 |
| C2orf74 | 59,68 | 0,04 | 0,03 | 1,10 | 2,72E-01 |
| TMEM63C | 71,05 | 0,01 | 0,10 | 0,09 | 9,29E-01 |
| PARD6A | 41,65 | 0,00 | 0,03 | -0,11 | 9,16E-01 |
| FBXO36 | 33,01 | 0,02 | 0,05 | 0,38 | 7,03E-01 |
| PLOD2 | 48,38 | 0,09 | 0,09 | 1,02 | 3,06E-01 |
| MARVELD2 | 29,19 | 0,03 | 0,07 | 0,45 | 6,52E-01 |
| PCDHGB3 | 51,17 | -0,10 | 0,06 | -1,71 | 8,77E-02 |
| CENPW | 26,26 | 0,04 | 0,05 | 0,80 | 4,26E-01 |
| ENSG000002 | 23,50 | 0,02 | 0,04 | 0,57 | 5,66E-01 |
| PRICKLE2 | 60,41 | 0,06 | 0,12 | 0,49 | 6,26E-01 |
| TAS2R5 | 60,38 | -0,02 | 0,04 | -0,55 | 5,80E-01 |
| ATP8B3 | 71,11 | 0,04 | 0,07 | 0,59 | 5,55E-01 |
| DDX43 | 55,93 | 0,02 | 0,09 | 0,18 | 8,54E-01 |

| | | | | | |
|---|---|---|---|---|---|
| MYZAP | 28,57 | -0,01 | 0,09 | -0,11 | 9,09E-01 |
| OSGIN1 | 43,68 | -0,04 | 0,06 | -0,57 | 5,72E-01 |
| SNX15 | 54,31 | 0,03 | 0,04 | 0,76 | 4,48E-01 |
| KCNC3 | 56,80 | 0,09 | 0,08 | 1,07 | 2,84E-01 |
| MYEOV | 36,03 | 0,10 | 0,09 | 1,05 | 2,91E-01 |
| TRAIP | 40,18 | 0,06 | 0,05 | 1,31 | 1,92E-01 |
| PCP2 | 31,37 | -0,05 | 0,05 | -1,06 | 2,88E-01 |
| C11orf65 | 31,70 | -0,01 | 0,07 | -0,21 | 8,30E-01 |
| OCEL1 | 40,23 | 0,03 | 0,06 | 0,45 | 6,51E-01 |
| PALD1 | 40,21 | -0,12 | 0,11 | -1,08 | 2,80E-01 |
| ENSG000002 | 58,29 | -0,03 | 0,07 | -0,46 | 6,42E-01 |
| ENSG000002 | 30,26 | -0,17 | 0,05 | -3,41 | 6,44E-04 |
| ENSG000002 | 56,81 | 0,09 | 0,04 | 2,06 | 3,92E-02 |
| TUBA8 | 48,20 | 0,04 | 0,05 | 0,73 | 4,67E-01 |
| FAM72A | 31,34 | 0,05 | 0,05 | 0,90 | 3,68E-01 |
| SRR | 60,04 | 0,05 | 0,04 | 1,40 | 1,62E-01 |
| GJC2 | 45,75 | 0,08 | 0,05 | 1,47 | 1,42E-01 |
| PRKG1 | 66,02 | 0,05 | 0,09 | 0,59 | 5,55E-01 |
| THSD7A | 57,07 | -0,01 | 0,11 | -0,05 | 9,59E-01 |
| GPX3 | 28,27 | -0,22 | 0,09 | -2,57 | 1,02E-02 |
| ZNF579 | 33,33 | 0,07 | 0,09 | 0,84 | 3,99E-01 |
| ENSG000002 | 63,70 | 0,05 | 0,07 | 0,73 | 4,65E-01 |
| SNAPC2 | 59,60 | 0,11 | 0,04 | 2,58 | 9,83E-03 |
| MANEAL | 38,21 | 0,01 | 0,07 | 0,12 | 9,07E-01 |
| HILPDA | 67,96 | 0,05 | 0,03 | 1,68 | 9,33E-02 |
| RAB23 | 70,82 | 0,08 | 0,06 | 1,40 | 1,61E-01 |
| MYO1A | 35,56 | 0,06 | 0,08 | 0,68 | 4,97E-01 |
| SYP | 39,63 | -0,15 | 0,07 | -2,18 | 2,93E-02 |
| UPP2 | 25,75 | 0,09 | 0,07 | 1,34 | 1,80E-01 |
| TEKTIP1 | 38,30 | 0,00 | 0,08 | 0,05 | 9,60E-01 |
| SIRT4 | 38,34 | 0,02 | 0,04 | 0,39 | 6,94E-01 |
| TRAPPC2B | 61,04 | 0,04 | 0,04 | 0,92 | 3,56E-01 |
| TRPM4 | 45,69 | 0,07 | 0,11 | 0,62 | 5,32E-01 |
| TNFRSF17 | 35,42 | 0,02 | 0,09 | 0,28 | 7,78E-01 |
| HTR7 | 39,18 | 0,10 | 0,06 | 1,83 | 6,77E-02 |
| TMEM163 | 39,55 | -0,07 | 0,11 | -0,63 | 5,27E-01 |
| PPP2R3A | 41,27 | -0,03 | 0,09 | -0,33 | 7,44E-01 |
| INSC | 34,64 | -0,12 | 0,10 | -1,20 | 2,31E-01 |
| ZFP2 | 34,51 | 0,04 | 0,06 | 0,59 | 5,58E-01 |
| ARMH2 | 59,07 | -0,10 | 0,04 | -2,71 | 6,68E-03 |
| GINS3 | 66,29 | -0,02 | 0,04 | -0,52 | 6,04E-01 |
| IL17RC | 57,96 | 0,03 | 0,06 | 0,57 | 5,66E-01 |
| TMPRSS5 | 46,96 | 0,00 | 0,07 | -0,01 | 9,89E-01 |
| ENSG000002 | 47,34 | -0,04 | 0,03 | -1,46 | 1,45E-01 |
| ZNF229 | 53,90 | 0,00 | 0,06 | 0,07 | 9,44E-01 |
| EFCAB6 | 51,98 | -0,05 | 0,12 | -0,41 | 6,79E-01 |

| | | | | | |
|---|---|---|---|---|---|
| FSBP | 30,32 | 0,05 | 0,06 | 0,88 | 3,77E-01 |
| LIN7B | 41,69 | 0,05 | 0,04 | 1,22 | 2,21E-01 |
| COL8A2 | 38,78 | 0,17 | 0,07 | 2,23 | 2,57E-02 |
| CDHR1 | 53,17 | -0,07 | 0,09 | -0,76 | 4,45E-01 |
| HEPACAM2 | 58,66 | 0,18 | 0,08 | 2,39 | 1,69E-02 |
| TRIM7 | 59,45 | 0,06 | 0,08 | 0,76 | 4,49E-01 |
| CD163L1 | 51,02 | 0,04 | 0,11 | 0,34 | 7,34E-01 |
| PROC | 33,37 | -0,06 | 0,08 | -0,78 | 4,34E-01 |
| ADGRG6 | 46,84 | 0,09 | 0,09 | 1,00 | 3,19E-01 |
| EVC2 | 70,29 | -0,09 | 0,09 | -0,98 | 3,30E-01 |
| CDK1 | 42,36 | 0,13 | 0,07 | 1,80 | 7,21E-02 |
| LARGE2 | 57,91 | 0,06 | 0,07 | 0,85 | 3,97E-01 |
| ZNF727 | 25,81 | 0,11 | 0,09 | 1,23 | 2,20E-01 |
| USP51 | 48,56 | 0,11 | 0,05 | 2,28 | 2,24E-02 |
| RHEX | 40,76 | -0,05 | 0,09 | -0,53 | 5,96E-01 |
| CASTOR1 | 54,82 | 0,01 | 0,03 | 0,46 | 6,49E-01 |
| SLC8A3 | 54,27 | -0,01 | 0,11 | -0,08 | 9,38E-01 |
| AJUBA | 39,13 | -0,01 | 0,07 | -0,15 | 8,79E-01 |
| CDC6 | 47,44 | -0,08 | 0,06 | -1,28 | 2,02E-01 |
| ABHD14A | 51,33 | 0,05 | 0,03 | 1,65 | 9,98E-02 |
| DEPDC1B | 43,28 | 0,12 | 0,07 | 1,66 | 9,65E-02 |
| SLC2A14 | 52,88 | -0,20 | 0,11 | -1,82 | 6,90E-02 |
| ALPK2 | 52,63 | -0,06 | 0,12 | -0,52 | 6,02E-01 |
| PCDHGA6 | 61,24 | -0,05 | 0,06 | -0,79 | 4,32E-01 |
| SGCD | 67,22 | -0,01 | 0,11 | -0,05 | 9,61E-01 |
| TACSTD2 | 45,99 | 0,00 | 0,12 | 0,01 | 9,89E-01 |
| HJURP | 37,69 | 0,03 | 0,09 | 0,37 | 7,08E-01 |
| NPIPB3 | 67,36 | 0,03 | 0,07 | 0,40 | 6,90E-01 |
| ITGA2 | 67,70 | 0,10 | 0,08 | 1,36 | 1,74E-01 |
| ISOC2 | 51,12 | 0,06 | 0,05 | 1,08 | 2,78E-01 |
| IQCH | 47,74 | -0,04 | 0,10 | -0,37 | 7,12E-01 |
| TANC1 | 57,27 | 0,00 | 0,11 | 0,01 | 9,90E-01 |
| NUF2 | 54,09 | 0,07 | 0,06 | 1,16 | 2,45E-01 |
| ENSG000002 | 51,82 | -0,07 | 0,09 | -0,79 | 4,30E-01 |
| SOX12 | 49,43 | 0,10 | 0,08 | 1,26 | 2,09E-01 |
| INHBB | 37,05 | -0,17 | 0,08 | -2,09 | 3,62E-02 |
| PODXL | 39,81 | -0,04 | 0,10 | -0,39 | 6,98E-01 |
| PARP3 | 66,53 | 0,03 | 0,05 | 0,55 | 5,80E-01 |
| SPRED2 | 65,70 | -0,06 | 0,06 | -0,94 | 3,49E-01 |
| UBE2T | 37,45 | 0,00 | 0,05 | -0,08 | 9,35E-01 |
| ZNF705EP | 24,17 | 0,00 | 0,05 | 0,08 | 9,34E-01 |
| UBE2C | 33,54 | -0,07 | 0,06 | -1,30 | 1,95E-01 |
| EMID1 | 59,85 | 0,08 | 0,09 | 0,91 | 3,62E-01 |
| MAGI2 | 60,54 | 0,06 | 0,13 | 0,47 | 6,35E-01 |
| HRK | 67,08 | 0,06 | 0,11 | 0,55 | 5,84E-01 |
| RLN1 | 53,62 | 0,03 | 0,05 | 0,68 | 4,98E-01 |

| | | | | | |
|---|---|---|---|---|---|
| PLXNB1 | 45,85 | 0,01 | 0,12 | 0,11 | 9,14E-01 |
| KLHL33 | 32,55 | 0,08 | 0,09 | 0,86 | 3,92E-01 |
| APOBEC3D | 61,56 | -0,01 | 0,04 | -0,36 | 7,18E-01 |
| NUDT8 | 23,98 | 0,12 | 0,06 | 2,02 | 4,32E-02 |
| RTEL1-TNFRS | 45,30 | 0,11 | 0,08 | 1,47 | 1,40E-01 |
| LRRC2 | 35,65 | 0,10 | 0,10 | 1,03 | 3,01E-01 |
| FAHD2B | 69,06 | 0,05 | 0,04 | 1,52 | 1,28E-01 |
| CYP2E1 | 47,54 | -0,02 | 0,12 | -0,16 | 8,75E-01 |
| NR2F6 | 44,20 | 0,18 | 0,06 | 3,08 | 2,07E-03 |
| LDB2 | 43,84 | -0,06 | 0,10 | -0,66 | 5,11E-01 |
| HAUS7 | 63,13 | -0,02 | 0,04 | -0,52 | 6,06E-01 |
| FNDC11 | 29,20 | 0,05 | 0,08 | 0,61 | 5,40E-01 |
| GPAT2 | 43,52 | -0,08 | 0,10 | -0,77 | 4,44E-01 |
| DEPDC7 | 34,11 | 0,07 | 0,06 | 1,18 | 2,38E-01 |
| ENSG000002 | 41,40 | 0,00 | 0,03 | 0,05 | 9,59E-01 |
| ARHGEF4 | 55,81 | -0,08 | 0,13 | -0,63 | 5,28E-01 |
| SIGLEC6 | 47,28 | 0,12 | 0,07 | 1,83 | 6,68E-02 |
| SMTN | 61,77 | 0,07 | 0,10 | 0,67 | 5,04E-01 |
| AVPI1 | 34,83 | -0,07 | 0,05 | -1,56 | 1,18E-01 |
| CFAP53 | 35,66 | 0,00 | 0,06 | 0,00 | 9,96E-01 |
| NRIP3 | 43,94 | -0,02 | 0,08 | -0,29 | 7,75E-01 |
| DTNA | 63,77 | 0,06 | 0,10 | 0,65 | 5,14E-01 |
| SCN1B | 68,21 | -0,08 | 0,08 | -1,09 | 2,76E-01 |
| FBLN2 | 57,73 | 0,03 | 0,09 | 0,28 | 7,78E-01 |
| ACHE | 34,78 | 0,26 | 0,10 | 2,46 | 1,38E-02 |
| FAM171A1 | 67,63 | 0,03 | 0,05 | 0,49 | 6,25E-01 |
| APCDD1 | 40,49 | 0,05 | 0,11 | 0,43 | 6,67E-01 |
| PLS1 | 58,66 | 0,02 | 0,06 | 0,27 | 7,88E-01 |
| STAG3 | 43,55 | 0,04 | 0,08 | 0,56 | 5,77E-01 |
| CNTNAP3B | 43,06 | -0,14 | 0,11 | -1,24 | 2,16E-01 |
| RTKN | 32,91 | -0,02 | 0,10 | -0,25 | 8,02E-01 |
| GP6 | 42,90 | 0,07 | 0,08 | 0,88 | 3,81E-01 |
| SEMA3A | 36,67 | 0,13 | 0,09 | 1,43 | 1,53E-01 |
| SNORC | 51,56 | -0,07 | 0,07 | -1,02 | 3,07E-01 |
| ZNF703 | 49,85 | 0,07 | 0,08 | 0,90 | 3,71E-01 |
| DUSP4 | 53,97 | 0,03 | 0,09 | 0,35 | 7,25E-01 |
| IL23R | 50,30 | 0,04 | 0,09 | 0,48 | 6,30E-01 |
| SLC25A35 | 67,13 | 0,03 | 0,05 | 0,67 | 5,04E-01 |
| SPIRE2 | 40,58 | -0,04 | 0,10 | -0,34 | 7,31E-01 |
| C3 | 46,88 | 0,11 | 0,10 | 1,11 | 2,67E-01 |
| WNT10B | 67,26 | 0,10 | 0,05 | 1,94 | 5,28E-02 |
| RAB9B | 40,06 | 0,09 | 0,05 | 2,08 | 3,73E-02 |
| CCDC81 | 37,31 | 0,00 | 0,08 | 0,01 | 9,91E-01 |
| BMP8B | 52,88 | -0,04 | 0,07 | -0,59 | 5,55E-01 |
| SLFNL1 | 32,30 | -0,03 | 0,09 | -0,29 | 7,74E-01 |
| ACOT7 | 59,92 | -0,01 | 0,08 | -0,15 | 8,82E-01 |

| | | | | | |
|---|---|---|---|---|---|
| CDCA8 | 34,34 | -0,04 | 0,06 | -0,70 | 4,84E-01 |
| PORCN | 47,68 | 0,01 | 0,06 | 0,11 | 9,11E-01 |
| SLC16A13 | 47,29 | -0,02 | 0,05 | -0,29 | 7,75E-01 |
| ELANE | 32,76 | 0,12 | 0,09 | 1,28 | 2,01E-01 |
| OMG | 41,65 | 0,03 | 0,05 | 0,62 | 5,37E-01 |
| GLIPR1L2 | 33,50 | 0,08 | 0,07 | 1,06 | 2,88E-01 |
| ATF7-NPFF | 30,30 | -0,05 | 0,04 | -1,27 | 2,03E-01 |
| BCKDHA | 32,30 | -0,03 | 0,03 | -1,04 | 2,97E-01 |
| FBXO24 | 38,10 | 0,03 | 0,06 | 0,55 | 5,80E-01 |
| FAM81A | 37,77 | 0,09 | 0,10 | 0,98 | 3,28E-01 |
| GPR153 | 48,58 | -0,09 | 0,09 | -0,93 | 3,51E-01 |
| CC2D2A | 49,31 | 0,03 | 0,09 | 0,27 | 7,85E-01 |
| OXTR | 40,03 | 0,01 | 0,10 | 0,14 | 8,89E-01 |
| ATXN7L2 | 53,85 | 0,06 | 0,07 | 0,86 | 3,89E-01 |
| POLR1G | 45,94 | 0,03 | 0,04 | 0,65 | 5,15E-01 |
| STRIP2 | 53,19 | 0,05 | 0,08 | 0,65 | 5,15E-01 |
| ANKRD34A | 49,91 | 0,06 | 0,04 | 1,32 | 1,87E-01 |
| MAPK11 | 62,35 | 0,10 | 0,05 | 2,04 | 4,16E-02 |
| EGFL7 | 33,67 | 0,13 | 0,07 | 1,76 | 7,90E-02 |
| TNK1 | 31,84 | 0,10 | 0,07 | 1,44 | 1,49E-01 |
| KLHL34 | 32,54 | -0,05 | 0,07 | -0,76 | 4,47E-01 |
| CHRM3 | 37,00 | 0,06 | 0,11 | 0,53 | 5,98E-01 |
| KIR3DL1 | 43,70 | -0,15 | 0,09 | -1,60 | 1,09E-01 |
| TMEM53 | 47,19 | 0,03 | 0,04 | 0,59 | 5,56E-01 |
| ASAH2 | 36,33 | 0,09 | 0,10 | 0,94 | 3,46E-01 |
| VEPH1 | 66,53 | -0,04 | 0,08 | -0,54 | 5,92E-01 |
| WDR62 | 54,25 | 0,07 | 0,09 | 0,74 | 4,57E-01 |
| FANK1 | 45,01 | 0,04 | 0,08 | 0,48 | 6,31E-01 |
| U2AF1L4 | 32,91 | 0,07 | 0,05 | 1,57 | 1,16E-01 |
| LMNTD2 | 44,63 | 0,12 | 0,06 | 1,97 | 4,92E-02 |
| REXO5 | 70,01 | 0,01 | 0,06 | 0,12 | 9,02E-01 |
| TTK | 46,08 | 0,14 | 0,09 | 1,65 | 9,88E-02 |
| MRPS24 | 32,66 | 0,02 | 0,04 | 0,49 | 6,21E-01 |
| NMB | 33,60 | -0,01 | 0,04 | -0,21 | 8,36E-01 |
| TMIGD3 | 31,57 | 0,05 | 0,07 | 0,70 | 4,86E-01 |
| MUC20 | 30,94 | 0,06 | 0,09 | 0,73 | 4,66E-01 |
| GCSH | 22,63 | 0,12 | 0,06 | 2,24 | 2,50E-02 |
| CCND1 | 56,16 | -0,02 | 0,09 | -0,20 | 8,41E-01 |
| PCDHGA5 | 43,63 | -0,08 | 0,06 | -1,26 | 2,07E-01 |
| CD80 | 55,85 | 0,06 | 0,06 | 1,10 | 2,73E-01 |
| ADORA2B | 48,50 | 0,05 | 0,05 | 0,93 | 3,54E-01 |
| NOCT | 68,11 | 0,00 | 0,04 | 0,05 | 9,60E-01 |
| SH2D6 | 31,96 | 0,09 | 0,08 | 1,23 | 2,17E-01 |
| ME1 | 49,02 | 0,05 | 0,06 | 0,75 | 4,51E-01 |
| PLEKHH2 | 59,46 | 0,02 | 0,10 | 0,18 | 8,53E-01 |
| SLC1A3 | 46,13 | 0,04 | 0,09 | 0,41 | 6,80E-01 |

| | | | | | |
|---|---|---|---|---|---|
| PARM1 | 51,15 | 0,07 | 0,06 | 1,08 | 2,80E-01 |
| PCDHGA11 | 66,18 | -0,02 | 0,07 | -0,31 | 7,53E-01 |
| HLA-DQB2 | 67,97 | 0,00 | 0,13 | 0,02 | 9,84E-01 |
| PSD | 54,64 | 0,15 | 0,09 | 1,69 | 9,20E-02 |
| SLC35F3 | 53,89 | -0,02 | 0,07 | -0,25 | 8,00E-01 |
| SLC44A5 | 59,91 | 0,12 | 0,17 | 0,74 | 4,61E-01 |
| IKZF4 | 60,89 | 0,03 | 0,06 | 0,43 | 6,68E-01 |
| GAREM2 | 42,37 | 0,02 | 0,08 | 0,24 | 8,07E-01 |
| AEBP1 | 51,33 | 0,05 | 0,10 | 0,47 | 6,35E-01 |
| MYL6B | 41,87 | 0,12 | 0,05 | 2,32 | 2,03E-02 |
| SPAG8 | 56,01 | 0,04 | 0,04 | 1,16 | 2,47E-01 |
| PDE2A | 65,74 | -0,04 | 0,10 | -0,39 | 6,95E-01 |
| SLC22A18AS | 27,52 | 0,00 | 0,08 | 0,01 | 9,92E-01 |
| ITGAD | 48,21 | -0,13 | 0,10 | -1,27 | 2,03E-01 |
| PAPLN | 62,16 | 0,05 | 0,10 | 0,48 | 6,30E-01 |
| GLS2 | 35,66 | 0,08 | 0,06 | 1,43 | 1,52E-01 |
| PCDH12 | 40,83 | 0,01 | 0,08 | 0,11 | 9,11E-01 |
| SYCP3 | 53,75 | -0,07 | 0,05 | -1,39 | 1,65E-01 |
| LGMN | 50,07 | -0,03 | 0,06 | -0,54 | 5,88E-01 |
| LRFN4 | 58,50 | 0,06 | 0,07 | 0,83 | 4,06E-01 |
| XKRX | 68,86 | 0,00 | 0,07 | -0,01 | 9,92E-01 |
| KCNMB4 | 59,31 | 0,07 | 0,05 | 1,20 | 2,28E-01 |
| CHKB-CPT1E | 43,78 | 0,01 | 0,05 | 0,32 | 7,49E-01 |
| CRACD | 48,83 | -0,04 | 0,12 | -0,37 | 7,08E-01 |
| HMGN5 | 35,32 | 0,00 | 0,05 | 0,06 | 9,49E-01 |
| PAK3 | 46,50 | -0,02 | 0,09 | -0,18 | 8,54E-01 |
| SLC29A2 | 51,65 | 0,07 | 0,07 | 1,10 | 2,70E-01 |
| FAM83F | 71,56 | 0,07 | 0,11 | 0,70 | 4,86E-01 |
| FAM83D | 40,83 | -0,02 | 0,06 | -0,39 | 6,97E-01 |
| SATB2 | 49,11 | -0,01 | 0,10 | -0,06 | 9,51E-01 |
| PLEKHG7 | 49,13 | 0,00 | 0,08 | 0,03 | 9,75E-01 |
| TMC4 | 41,31 | 0,01 | 0,07 | 0,17 | 8,68E-01 |
| ZNF404 | 44,65 | 0,08 | 0,06 | 1,48 | 1,40E-01 |
| FREY1 | 24,40 | 0,03 | 0,04 | 0,94 | 3,49E-01 |
| GUCA1B | 64,89 | -0,01 | 0,04 | -0,31 | 7,53E-01 |
| MFSD6L | 68,21 | -0,01 | 0,06 | -0,15 | 8,84E-01 |
| CAVIN3 | 26,66 | 0,08 | 0,09 | 0,84 | 4,03E-01 |
| SAXO4 | 32,32 | -0,01 | 0,08 | -0,09 | 9,31E-01 |
| GLB1L2 | 64,90 | -0,03 | 0,11 | -0,23 | 8,19E-01 |
| IL20RB | 56,19 | -0,05 | 0,05 | -0,98 | 3,27E-01 |
| ZNF492 | 31,02 | 0,14 | 0,07 | 1,90 | 5,72E-02 |
| PIF1 | 39,62 | 0,00 | 0,07 | -0,03 | 9,75E-01 |
| STK31 | 47,06 | 0,00 | 0,08 | -0,02 | 9,83E-01 |
| SPATA24 | 30,21 | 0,00 | 0,03 | 0,13 | 9,00E-01 |
| DNAH14 | 71,19 | 0,11 | 0,13 | 0,86 | 3,87E-01 |
| LRFN3 | 32,65 | 0,09 | 0,09 | 0,96 | 3,35E-01 |

| | | | | | |
|---|---|---|---|---|---|
| NME2 | 41,33 | 0,11 | 0,07 | 1,62 | 1,06E-01 |
| KIF19 | 63,76 | -0,03 | 0,11 | -0,29 | 7,71E-01 |
| ALOX12 | 48,46 | 0,07 | 0,09 | 0,76 | 4,46E-01 |
| PIP5KL1 | 35,33 | 0,12 | 0,07 | 1,73 | 8,43E-02 |
| HBEGF | 39,88 | 0,02 | 0,07 | 0,28 | 7,79E-01 |
| PIGZ | 33,55 | 0,02 | 0,06 | 0,32 | 7,52E-01 |
| TBC1D3L | 42,25 | -0,12 | 0,10 | -1,22 | 2,23E-01 |
| ABCA9 | 56,02 | -0,01 | 0,11 | -0,10 | 9,22E-01 |
| FAM131B | 58,69 | 0,01 | 0,06 | 0,15 | 8,82E-01 |
| PODN | 48,87 | -0,10 | 0,11 | -0,93 | 3,52E-01 |
| ENSG000002 | 32,92 | -0,05 | 0,05 | -0,87 | 3,85E-01 |
| CLEC18B | 47,91 | -0,12 | 0,10 | -1,17 | 2,41E-01 |
| LPL | 37,07 | -0,07 | 0,11 | -0,65 | 5,17E-01 |
| MST1 | 55,46 | 0,01 | 0,05 | 0,16 | 8,75E-01 |
| AQP10 | 59,75 | 0,00 | 0,08 | -0,01 | 9,92E-01 |
| GFRA2 | 55,13 | -0,01 | 0,10 | -0,15 | 8,78E-01 |
| ZNF578 | 60,87 | 0,01 | 0,06 | 0,20 | 8,43E-01 |
| LRP3 | 66,53 | 0,03 | 0,07 | 0,50 | 6,18E-01 |
| MYH10 | 68,34 | 0,07 | 0,09 | 0,74 | 4,57E-01 |
| CAV2 | 59,82 | 0,00 | 0,10 | 0,00 | 9,98E-01 |
| HBQ1 | 55,62 | 0,18 | 0,08 | 2,33 | 1,95E-02 |
| FSIP1 | 54,70 | -0,03 | 0,06 | -0,49 | 6,27E-01 |
| EIF4EBP1 | 71,40 | 0,08 | 0,03 | 2,39 | 1,66E-02 |
| ABCF2 | 71,57 | 0,02 | 0,03 | 0,61 | 5,42E-01 |
| LAMP5 | 28,15 | -0,02 | 0,07 | -0,28 | 7,79E-01 |
| LRRC24 | 32,80 | 0,05 | 0,06 | 0,92 | 3,57E-01 |
| NPIPB2 | 43,28 | -0,01 | 0,06 | -0,16 | 8,71E-01 |
| MISFA | 42,18 | -0,06 | 0,04 | -1,37 | 1,72E-01 |
| DCLK2 | 49,38 | -0,12 | 0,12 | -1,03 | 3,03E-01 |
| ENSG000002 | 68,15 | -0,13 | 0,06 | -2,17 | 3,02E-02 |
| ZSCAN23 | 68,70 | 0,07 | 0,07 | 1,10 | 2,73E-01 |
| WHRN | 70,99 | 0,06 | 0,08 | 0,80 | 4,24E-01 |
| ZNF628 | 59,25 | 0,06 | 0,07 | 0,87 | 3,85E-01 |
| KCND3 | 42,31 | 0,02 | 0,11 | 0,17 | 8,63E-01 |
| DECR2 | 40,98 | 0,07 | 0,07 | 1,10 | 2,71E-01 |
| ATOH8 | 52,47 | -0,13 | 0,11 | -1,15 | 2,48E-01 |
| CDKN3 | 29,72 | 0,13 | 0,08 | 1,63 | 1,02E-01 |
| PTPRB | 56,90 | -0,03 | 0,11 | -0,30 | 7,65E-01 |
| ACOXL | 56,37 | -0,13 | 0,09 | -1,40 | 1,61E-01 |
| SPR | 33,34 | -0,02 | 0,06 | -0,36 | 7,17E-01 |
| KIF9 | 66,65 | -0,02 | 0,05 | -0,35 | 7,27E-01 |
| TMEM40 | 70,24 | 0,11 | 0,06 | 1,71 | 8,65E-02 |
| LRRC70 | 46,36 | 0,06 | 0,05 | 1,07 | 2,84E-01 |
| MMP11 | 36,31 | 0,07 | 0,09 | 0,77 | 4,38E-01 |
| INKA1 | 42,57 | 0,05 | 0,04 | 1,17 | 2,42E-01 |
| AJM1 | 70,28 | 0,06 | 0,07 | 0,98 | 3,26E-01 |

| | | | | | |
|---|---|---|---|---|---|
| FGF9 | 51,25 | 0,06 | 0,07 | 0,81 | 4,16E-01 |
| OTX1 | 46,35 | -0,13 | 0,10 | -1,30 | 1,93E-01 |
| GALNT8 | 34,46 | -0,05 | 0,09 | -0,54 | 5,87E-01 |
| ACSBG1 | 64,43 | 0,01 | 0,10 | 0,09 | 9,25E-01 |
| CABYR | 34,38 | -0,04 | 0,08 | -0,54 | 5,88E-01 |
| CBLN3 | 41,36 | 0,10 | 0,07 | 1,47 | 1,42E-01 |
| NBL1 | 57,66 | 0,05 | 0,08 | 0,71 | 4,80E-01 |
| ENSG000001 | 68,81 | -0,11 | 0,08 | -1,34 | 1,79E-01 |
| KL | 46,98 | -0,09 | 0,07 | -1,25 | 2,12E-01 |
| FBXO10 | 30,61 | 0,03 | 0,07 | 0,47 | 6,41E-01 |
| TSPAN15 | 66,17 | 0,03 | 0,07 | 0,46 | 6,44E-01 |
| ACKR1 | 36,29 | 0,04 | 0,09 | 0,40 | 6,89E-01 |
| NOL3 | 60,17 | 0,09 | 0,06 | 1,59 | 1,11E-01 |
| S100A13 | 33,84 | 0,02 | 0,04 | 0,50 | 6,18E-01 |
| DAGLA | 49,51 | 0,05 | 0,09 | 0,53 | 5,93E-01 |
| TPM2 | 63,98 | 0,01 | 0,06 | 0,09 | 9,27E-01 |
| SUCNR1 | 53,26 | 0,08 | 0,06 | 1,34 | 1,81E-01 |
| ACTA2 | 67,12 | -0,12 | 0,07 | -1,83 | 6,70E-02 |
| DNM1 | 51,17 | 0,04 | 0,11 | 0,35 | 7,25E-01 |
| CDH2 | 67,00 | -0,11 | 0,09 | -1,30 | 1,92E-01 |
| PTPN20 | 35,96 | 0,02 | 0,10 | 0,16 | 8,71E-01 |
| FCRLB | 36,78 | -0,02 | 0,06 | -0,34 | 7,33E-01 |
| DDR2 | 70,98 | -0,06 | 0,10 | -0,67 | 5,03E-01 |
| MYOCOS | 28,06 | -0,01 | 0,06 | -0,19 | 8,51E-01 |
| LGSN | 33,04 | -0,26 | 0,12 | -2,05 | 3,99E-02 |
| TAS2R10 | 31,12 | 0,07 | 0,05 | 1,49 | 1,36E-01 |
| ADAMTS4 | 55,30 | 0,14 | 0,07 | 2,02 | 4,35E-02 |
| DND1 | 48,96 | -0,04 | 0,04 | -0,91 | 3,63E-01 |
| C3orf52 | 35,19 | 0,04 | 0,09 | 0,49 | 6,27E-01 |
| RNASEH2A | 24,53 | 0,00 | 0,04 | 0,10 | 9,17E-01 |
| BUB1B | 57,35 | 0,08 | 0,06 | 1,31 | 1,91E-01 |
| LRP5 | 65,48 | 0,00 | 0,10 | 0,03 | 9,77E-01 |
| NFIB | 44,09 | 0,10 | 0,12 | 0,84 | 4,00E-01 |
| SLC39A4 | 54,86 | 0,02 | 0,06 | 0,35 | 7,29E-01 |
| HES6 | 61,10 | 0,08 | 0,06 | 1,35 | 1,76E-01 |
| CCDC78 | 28,15 | 0,09 | 0,08 | 1,10 | 2,71E-01 |
| DIAPH3 | 59,01 | 0,12 | 0,08 | 1,48 | 1,38E-01 |
| SYCP2L | 46,55 | -0,09 | 0,12 | -0,75 | 4,54E-01 |
| PXDC1 | 44,81 | -0,03 | 0,07 | -0,38 | 7,06E-01 |
| NRM | 61,15 | 0,08 | 0,04 | 2,24 | 2,52E-02 |
| ZNF385D | 38,80 | 0,08 | 0,10 | 0,76 | 4,48E-01 |
| TMC5 | 69,53 | 0,02 | 0,09 | 0,16 | 8,71E-01 |
| RTL5 | 29,23 | -0,03 | 0,07 | -0,41 | 6,81E-01 |
| HSD3B7 | 31,23 | 0,12 | 0,07 | 1,80 | 7,26E-02 |
| SPTSSB | 54,51 | 0,05 | 0,07 | 0,80 | 4,26E-01 |
| ENSG000002 | 49,21 | -0,09 | 0,03 | -3,05 | 2,30E-03 |

| | | | | | |
|---|---|---|---|---|---|
| SLC6A4 | 48,80 | -0,02 | 0,10 | -0,21 | 8,33E-01 |
| INO80B | 44,69 | 0,07 | 0,03 | 2,13 | 3,29E-02 |
| CLBA1 | 47,44 | 0,04 | 0,06 | 0,65 | 5,17E-01 |
| TMEM25 | 69,69 | 0,05 | 0,05 | 1,07 | 2,83E-01 |
| C20orf96 | 28,54 | 0,04 | 0,07 | 0,62 | 5,36E-01 |
| FOLR2 | 49,72 | 0,03 | 0,05 | 0,66 | 5,09E-01 |
| ZNF391 | 44,46 | 0,01 | 0,06 | 0,16 | 8,73E-01 |
| RBMS3 | 52,45 | 0,06 | 0,10 | 0,54 | 5,88E-01 |
| PTH2R | 54,29 | 0,09 | 0,06 | 1,60 | 1,10E-01 |
| ALDH4A1 | 31,85 | -0,01 | 0,07 | -0,19 | 8,47E-01 |
| L1TD1 | 40,08 | -0,01 | 0,11 | -0,06 | 9,53E-01 |
| MCM10 | 45,34 | 0,02 | 0,10 | 0,21 | 8,38E-01 |
| GP5 | 54,82 | 0,08 | 0,07 | 1,22 | 2,23E-01 |
| BEND3 | 54,78 | -0,03 | 0,07 | -0,45 | 6,54E-01 |
| BIRC5 | 31,91 | 0,02 | 0,08 | 0,28 | 7,81E-01 |
| KLHL29 | 71,47 | -0,01 | 0,11 | -0,11 | 9,15E-01 |
| ERRFI1 | 37,43 | -0,10 | 0,06 | -1,65 | 9,80E-02 |
| MDK | 27,35 | -0,10 | 0,08 | -1,37 | 1,70E-01 |
| PROCR | 34,12 | 0,03 | 0,05 | 0,51 | 6,10E-01 |
| SYCE1L | 42,91 | 0,04 | 0,05 | 0,91 | 3,65E-01 |
| ENSG00000Z | 33,62 | -0,04 | 0,06 | -0,69 | 4,91E-01 |
| NPHP1 | 59,10 | 0,01 | 0,11 | 0,09 | 9,31E-01 |
| FAM229B | 32,61 | 0,13 | 0,06 | 1,96 | 5,05E-02 |
| HS3ST1 | 57,88 | 0,06 | 0,08 | 0,73 | 4,63E-01 |
| WASF1 | 45,18 | 0,06 | 0,06 | 1,17 | 2,43E-01 |
| PRSS36 | 61,43 | 0,02 | 0,05 | 0,40 | 6,89E-01 |
| ITGB1BP2 | 47,75 | -0,07 | 0,04 | -1,50 | 1,35E-01 |
| MACROD1 | 58,28 | 0,05 | 0,05 | 1,07 | 2,87E-01 |
| STIMATE-MUS | 31,76 | -0,02 | 0,06 | -0,37 | 7,09E-01 |
| DLL1 | 57,27 | 0,04 | 0,08 | 0,55 | 5,81E-01 |
| PMP22 | 42,61 | -0,10 | 0,10 | -1,00 | 3,16E-01 |
| E2F1 | 59,70 | 0,02 | 0,05 | 0,28 | 7,76E-01 |
| SOX8 | 46,03 | -0,04 | 0,09 | -0,46 | 6,48E-01 |
| RNF26 | 68,99 | 0,09 | 0,05 | 1,65 | 9,87E-02 |
| CCNB2 | 30,07 | 0,04 | 0,07 | 0,54 | 5,91E-01 |
| CCDC34 | 49,22 | 0,04 | 0,06 | 0,71 | 4,78E-01 |
| ENSG00000Z | 27,26 | -0,11 | 0,06 | -1,69 | 9,09E-02 |
| NLGN3 | 61,35 | -0,10 | 0,07 | -1,38 | 1,68E-01 |
| GINS1 | 43,57 | 0,03 | 0,07 | 0,43 | 6,71E-01 |
| SLCO5A1 | 47,73 | 0,02 | 0,11 | 0,22 | 8,24E-01 |
| CCDC120 | 32,06 | -0,02 | 0,08 | -0,19 | 8,46E-01 |
| CFAP161 | 66,21 | -0,01 | 0,05 | -0,12 | 9,08E-01 |
| PEX11G | 29,77 | -0,03 | 0,08 | -0,41 | 6,78E-01 |
| LGALS9C | 42,44 | -0,22 | 0,10 | -2,32 | 2,03E-02 |
| G0S2 | 45,36 | -0,02 | 0,08 | -0,22 | 8,25E-01 |
| IGFBP4 | 61,64 | 0,11 | 0,07 | 1,57 | 1,17E-01 |

| | | | | | |
|---|---|---|---|---|---|
| DRICH1 | 50,20 | -0,07 | 0,05 | -1,40 | 1,61E-01 |
| ANKLE1 | 35,06 | 0,01 | 0,06 | 0,11 | 9,16E-01 |
| DPPA4 | 28,70 | 0,04 | 0,08 | 0,55 | 5,84E-01 |
| PTGFRN | 70,06 | 0,04 | 0,07 | 0,53 | 5,99E-01 |
| MTFR2 | 39,67 | 0,01 | 0,05 | 0,24 | 8,08E-01 |
| LNP1 | 49,88 | 0,11 | 0,05 | 2,45 | 1,44E-02 |
| SEC14L2 | 70,66 | 0,03 | 0,06 | 0,56 | 5,75E-01 |
| GRK4 | 71,36 | 0,01 | 0,05 | 0,24 | 8,10E-01 |
| PYCR3 | 69,46 | 0,11 | 0,04 | 2,57 | 1,02E-02 |
| NPAS2 | 52,17 | 0,05 | 0,12 | 0,44 | 6,60E-01 |
| TCN2 | 68,55 | -0,09 | 0,06 | -1,46 | 1,45E-01 |
| GP1BA | 28,55 | 0,15 | 0,10 | 1,52 | 1,28E-01 |
| CTH | 53,02 | 0,05 | 0,05 | 1,08 | 2,80E-01 |
| ANKRD20A4I | 35,05 | -0,03 | 0,13 | -0,21 | 8,37E-01 |
| ZNF414 | 51,28 | 0,11 | 0,07 | 1,62 | 1,06E-01 |
| IRF6 | 35,63 | 0,01 | 0,10 | 0,11 | 9,16E-01 |
| IRGM | 41,54 | 0,04 | 0,04 | 1,06 | 2,90E-01 |
| ADORA3 | 67,90 | 0,11 | 0,07 | 1,60 | 1,10E-01 |
| SPOCK1 | 49,40 | 0,01 | 0,09 | 0,09 | 9,26E-01 |
| PCDHGB7 | 59,01 | -0,03 | 0,06 | -0,42 | 6,77E-01 |
| EFNA5 | 41,65 | -0,11 | 0,09 | -1,25 | 2,10E-01 |
| DLGAP3 | 49,86 | -0,07 | 0,12 | -0,62 | 5,38E-01 |
| F2RL3 | 37,15 | 0,01 | 0,08 | 0,12 | 9,05E-01 |
| RGS7 | 30,38 | 0,00 | 0,08 | -0,04 | 9,68E-01 |
| CABP4 | 42,45 | 0,05 | 0,07 | 0,75 | 4,54E-01 |
| PSMC3IP | 64,48 | -0,07 | 0,03 | -2,24 | 2,52E-02 |
| TIMM23B-AG | 67,88 | -0,01 | 0,05 | -0,27 | 7,89E-01 |
| E2F7 | 35,04 | 0,02 | 0,10 | 0,17 | 8,65E-01 |
| RPL39L | 35,42 | 0,10 | 0,05 | 1,92 | 5,47E-02 |
| SARS2 | 57,27 | 0,09 | 0,04 | 2,30 | 2,15E-02 |
| ZMAT4 | 41,26 | -0,01 | 0,11 | -0,07 | 9,47E-01 |
| LINGO2 | 51,07 | -0,05 | 0,09 | -0,61 | 5,39E-01 |
| FILIP1L | 38,73 | 0,00 | 0,11 | -0,03 | 9,76E-01 |
| NRL | 70,65 | 0,08 | 0,03 | 2,41 | 1,58E-02 |
| NAP1L3 | 58,90 | 0,05 | 0,06 | 0,98 | 3,30E-01 |
| CD1E | 34,01 | 0,08 | 0,08 | 0,98 | 3,29E-01 |
| ZNF280B | 66,50 | 0,02 | 0,05 | 0,38 | 7,01E-01 |
| KNDC1 | 66,07 | 0,17 | 0,12 | 1,45 | 1,46E-01 |
| TMEM232 | 47,35 | 0,15 | 0,07 | 2,20 | 2,76E-02 |
| ZSCAN31 | 35,96 | 0,04 | 0,07 | 0,48 | 6,31E-01 |
| NPTXR | 36,64 | 0,08 | 0,10 | 0,88 | 3,78E-01 |
| LIPH | 38,11 | -0,04 | 0,09 | -0,49 | 6,26E-01 |
| CPLANE2 | 61,58 | 0,02 | 0,03 | 0,66 | 5,12E-01 |
| KPTN | 66,82 | -0,07 | 0,05 | -1,28 | 2,01E-01 |
| SLC27A2 | 36,16 | 0,01 | 0,07 | 0,10 | 9,21E-01 |
| HSD17B13 | 57,45 | -0,13 | 0,05 | -2,99 | 2,81E-03 |

| | | | | | |
|---|---|---|---|---|---|
| CAPN14 | 59,91 | -0,01 | 0,07 | -0,16 | 8,74E-01 |
| PFKFB1 | 28,94 | 0,01 | 0,06 | 0,12 | 9,05E-01 |
| PCDHGA7 | 38,76 | -0,15 | 0,07 | -2,30 | 2,17E-02 |
| SMIM38 | 38,78 | -0,16 | 0,09 | -1,84 | 6,54E-02 |
| LRRIQ3 | 44,48 | 0,19 | 0,09 | 2,26 | 2,37E-02 |
| VMO1 | 30,51 | 0,00 | 0,08 | 0,00 | 9,96E-01 |
| NR1D1 | 33,23 | 0,14 | 0,07 | 2,06 | 3,94E-02 |
| PPP4R4 | 70,38 | 0,12 | 0,08 | 1,49 | 1,37E-01 |
| EMILIN1 | 54,53 | 0,07 | 0,07 | 1,02 | 3,10E-01 |
| ZNF462 | 55,31 | -0,06 | 0,13 | -0,44 | 6,58E-01 |
| BCAM | 42,97 | 0,14 | 0,12 | 1,14 | 2,53E-01 |
| ZNF433 | 35,06 | 0,06 | 0,06 | 1,06 | 2,89E-01 |
| PDCD2L | 50,19 | 0,08 | 0,03 | 2,27 | 2,32E-02 |
| KCNAB3 | 53,09 | 0,02 | 0,07 | 0,29 | 7,72E-01 |
| XRCC2 | 67,15 | 0,06 | 0,06 | 1,06 | 2,90E-01 |
| KATNAL2 | 42,15 | -0,11 | 0,10 | -1,14 | 2,54E-01 |
| NKAPL | 50,04 | -0,21 | 0,05 | -3,94 | 8,05E-05 |
| CCDC183 | 28,51 | 0,01 | 0,05 | 0,14 | 8,86E-01 |
| CBR3 | 57,03 | 0,09 | 0,06 | 1,48 | 1,39E-01 |
| TMEM201 | 50,77 | 0,06 | 0,09 | 0,67 | 5,05E-01 |
| ST18 | 54,14 | 0,00 | 0,13 | -0,01 | 9,88E-01 |
| CHL1 | 66,68 | 0,00 | 0,09 | 0,05 | 9,62E-01 |
| CDK15 | 42,32 | -0,09 | 0,11 | -0,81 | 4,20E-01 |
| DTX3 | 57,71 | 0,06 | 0,07 | 0,89 | 3,72E-01 |
| SLC25A27 | 36,72 | -0,02 | 0,07 | -0,25 | 8,02E-01 |
| SLC35E4 | 61,10 | 0,01 | 0,07 | 0,23 | 8,19E-01 |
| CHDH | 50,47 | -0,12 | 0,12 | -0,98 | 3,25E-01 |
| PCDHGC3 | 29,74 | -0,04 | 0,07 | -0,47 | 6,38E-01 |
| ESM1 | 30,31 | 0,07 | 0,09 | 0,79 | 4,30E-01 |
| CHCHD5 | 44,24 | 0,01 | 0,04 | 0,18 | 8,57E-01 |
| ANLN | 68,39 | 0,15 | 0,07 | 2,03 | 4,22E-02 |
| ATP6V1G2 | 43,81 | -0,01 | 0,04 | -0,35 | 7,26E-01 |
| WASF3 | 52,90 | -0,05 | 0,09 | -0,52 | 6,01E-01 |
| GRM2 | 41,06 | -0,04 | 0,11 | -0,39 | 6,95E-01 |
| HYKK | 38,87 | 0,06 | 0,07 | 0,87 | 3,84E-01 |
| BFSP1 | 58,46 | -0,02 | 0,09 | -0,24 | 8,10E-01 |
| HSD17B1 | 30,94 | 0,08 | 0,07 | 1,22 | 2,21E-01 |
| GLB1L | 49,59 | 0,05 | 0,06 | 0,75 | 4,51E-01 |
| WNT16 | 53,05 | 0,09 | 0,09 | 0,95 | 3,40E-01 |
| TGFB3 | 54,68 | 0,03 | 0,07 | 0,41 | 6,85E-01 |
| DNLZ | 29,34 | 0,10 | 0,06 | 1,69 | 9,10E-02 |
| PKD1L3 | 32,98 | 0,16 | 0,10 | 1,56 | 1,18E-01 |
| CEP15 | 40,73 | 0,11 | 0,07 | 1,61 | 1,08E-01 |
| TBX6 | 25,78 | 0,07 | 0,06 | 1,21 | 2,28E-01 |
| SAPCD2 | 45,62 | 0,10 | 0,09 | 1,11 | 2,67E-01 |
| KLHL23 | 41,25 | 0,04 | 0,06 | 0,67 | 5,05E-01 |

| | | | | | |
|---|---|---|---|---|---|
| IZUMO4 | 45,81 | 0,04 | 0,05 | 0,99 | 3,23E-01 |
| LAMB3 | 38,98 | -0,07 | 0,10 | -0,72 | 4,70E-01 |
| ARL6IP4 | 54,91 | 0,02 | 0,03 | 0,71 | 4,80E-01 |
| AKR1C8 | 31,50 | 0,01 | 0,08 | 0,12 | 9,07E-01 |
| FOXD4L1 | 24,62 | -0,02 | 0,05 | -0,41 | 6,84E-01 |
| OXER1 | 58,21 | -0,05 | 0,08 | -0,63 | 5,26E-01 |
| DZIP1L | 54,65 | 0,00 | 0,09 | 0,04 | 9,70E-01 |
| TPPP3 | 61,73 | 0,16 | 0,06 | 2,91 | 3,58E-03 |
| FRMD6 | 54,95 | -0,04 | 0,10 | -0,43 | 6,65E-01 |
| TNNC2 | 29,55 | -0,03 | 0,08 | -0,41 | 6,81E-01 |
| CRYBG2 | 55,50 | -0,01 | 0,07 | -0,13 | 8,93E-01 |
| A3GALT2 | 28,92 | -0,16 | 0,08 | -2,08 | 3,77E-02 |
| PET117 | 64,36 | 0,06 | 0,04 | 1,64 | 1,01E-01 |
| OCLN | 36,90 | 0,02 | 0,11 | 0,17 | 8,68E-01 |
| TRPV3 | 64,39 | -0,10 | 0,11 | -0,90 | 3,67E-01 |
| CCNE2 | 56,30 | 0,07 | 0,05 | 1,23 | 2,18E-01 |
| HMGB3 | 50,61 | -0,02 | 0,06 | -0,42 | 6,77E-01 |
| TRIP10 | 39,86 | 0,09 | 0,07 | 1,33 | 1,83E-01 |
| CDNF | 36,29 | 0,06 | 0,05 | 1,27 | 2,06E-01 |
| ATF3 | 42,15 | -0,04 | 0,09 | -0,43 | 6,67E-01 |
| GCM1 | 35,25 | -0,12 | 0,06 | -2,00 | 4,60E-02 |
| TSPEAR | 36,35 | -0,01 | 0,12 | -0,10 | 9,17E-01 |
| NECAB2 | 50,71 | -0,18 | 0,08 | -2,23 | 2,58E-02 |
| RDH16 | 50,44 | -0,01 | 0,06 | -0,21 | 8,37E-01 |
| STARD13 | 51,96 | -0,01 | 0,10 | -0,10 | 9,17E-01 |
| ACTMAP | 39,96 | 0,05 | 0,06 | 0,74 | 4,58E-01 |
| P2RY6 | 33,56 | 0,03 | 0,09 | 0,31 | 7,53E-01 |
| SLC35G5 | 25,07 | -0,01 | 0,05 | -0,15 | 8,77E-01 |
| NHLRC1 | 38,23 | 0,08 | 0,04 | 1,99 | 4,67E-02 |
| ADCY10 | 60,23 | -0,07 | 0,07 | -1,03 | 3,04E-01 |
| RAPSN | 38,61 | -0,02 | 0,07 | -0,25 | 7,99E-01 |
| ADGRB2 | 51,89 | -0,08 | 0,13 | -0,61 | 5,41E-01 |
| ACVRL1 | 44,79 | -0,02 | 0,09 | -0,25 | 8,04E-01 |
| POMC | 65,26 | 0,18 | 0,06 | 2,80 | 5,15E-03 |
| FITM1 | 26,54 | 0,04 | 0,04 | 0,91 | 3,64E-01 |
| SLC6A13 | 35,35 | -0,13 | 0,10 | -1,34 | 1,81E-01 |
| CALHM5 | 39,58 | 0,13 | 0,08 | 1,63 | 1,03E-01 |
| LYVE1 | 61,80 | -0,18 | 0,06 | -3,27 | 1,06E-03 |
| TAS2R60 | 47,59 | -0,07 | 0,08 | -0,85 | 3,97E-01 |
| CLCF1 | 54,26 | 0,06 | 0,05 | 1,22 | 2,21E-01 |
| BTNL9 | 45,05 | 0,08 | 0,12 | 0,68 | 4,98E-01 |
| OLFM1 | 62,68 | 0,07 | 0,11 | 0,61 | 5,42E-01 |
| HSPA12A | 68,93 | -0,06 | 0,11 | -0,53 | 5,96E-01 |
| GSDME | 48,01 | 0,03 | 0,11 | 0,32 | 7,50E-01 |
| KCNK5 | 33,46 | -0,05 | 0,09 | -0,51 | 6,13E-01 |
| FAM167A | 43,13 | -0,12 | 0,10 | -1,18 | 2,39E-01 |

| | | | | | |
|---|---|---|---|---|---|
| CMBL | 48,42 | 0,11 | 0,12 | 0,94 | 3,48E-01 |
| ESCO2 | 49,49 | 0,06 | 0,08 | 0,80 | 4,24E-01 |
| ZMYND10 | 48,14 | -0,03 | 0,05 | -0,67 | 5,00E-01 |
| SERINC2 | 43,89 | 0,03 | 0,12 | 0,27 | 7,90E-01 |
| DNAI3 | 38,47 | 0,01 | 0,09 | 0,09 | 9,25E-01 |
| FHL2 | 54,13 | 0,13 | 0,08 | 1,59 | 1,12E-01 |
| PHF24 | 32,66 | -0,18 | 0,11 | -1,67 | 9,40E-02 |
| ABHD17C | 55,28 | -0,06 | 0,06 | -0,91 | 3,62E-01 |
| GGH | 53,56 | -0,02 | 0,06 | -0,31 | 7,57E-01 |
| ENSG000002 | 25,23 | -0,06 | 0,08 | -0,75 | 4,54E-01 |
| APOBEC3H | 38,90 | 0,02 | 0,06 | 0,33 | 7,40E-01 |
| NPR3 | 39,76 | 0,07 | 0,11 | 0,63 | 5,28E-01 |
| GTSF1 | 57,43 | 0,02 | 0,06 | 0,43 | 6,64E-01 |
| RBM11 | 58,93 | 0,12 | 0,06 | 2,05 | 3,99E-02 |
| XCL1 | 52,60 | 0,14 | 0,07 | 2,04 | 4,17E-02 |
| ENSG000002 | 41,68 | 0,03 | 0,04 | 0,88 | 3,79E-01 |
| CCNA2 | 69,47 | -0,05 | 0,05 | -1,04 | 3,00E-01 |
| POMZP3 | 60,12 | 0,04 | 0,04 | 0,99 | 3,20E-01 |
| SLC18A1 | 34,35 | 0,00 | 0,11 | 0,04 | 9,71E-01 |
| TIMD4 | 35,78 | 0,01 | 0,06 | 0,17 | 8,63E-01 |
| TMPRSS3 | 50,56 | 0,01 | 0,08 | 0,07 | 9,44E-01 |
| UPK3A | 37,83 | 0,11 | 0,06 | 2,01 | 4,45E-02 |
| ZNF503 | 35,43 | -0,01 | 0,10 | -0,12 | 9,02E-01 |
| ZNRD2 | 68,08 | 0,06 | 0,03 | 2,04 | 4,18E-02 |
| ENSG000002 | 35,03 | -0,07 | 0,06 | -1,12 | 2,64E-01 |
| IL31RA | 55,30 | -0,02 | 0,09 | -0,24 | 8,11E-01 |
| DBNDD2 | 50,30 | -0,02 | 0,05 | -0,30 | 7,67E-01 |
| TTC24 | 53,77 | 0,17 | 0,06 | 2,68 | 7,36E-03 |
| CABP5 | 47,76 | 0,01 | 0,08 | 0,09 | 9,26E-01 |
| MAMLD1 | 40,62 | 0,08 | 0,08 | 1,12 | 2,63E-01 |
| SMAGP | 62,73 | -0,01 | 0,04 | -0,22 | 8,24E-01 |
| ZNF775 | 63,38 | 0,00 | 0,03 | 0,10 | 9,17E-01 |
| C17orf58 | 31,07 | 0,00 | 0,06 | -0,08 | 9,37E-01 |
| HAAO | 48,98 | 0,03 | 0,06 | 0,50 | 6,18E-01 |
| MMEL1 | 34,27 | -0,02 | 0,10 | -0,19 | 8,49E-01 |
| SCML2 | 45,10 | 0,02 | 0,05 | 0,46 | 6,44E-01 |
| PITPNM3 | 58,04 | -0,11 | 0,13 | -0,84 | 4,00E-01 |
| LONRF2 | 57,10 | -0,05 | 0,14 | -0,39 | 6,93E-01 |
| PNOC | 35,37 | 0,08 | 0,06 | 1,31 | 1,91E-01 |
| ZNF670 | 45,28 | 0,08 | 0,05 | 1,63 | 1,03E-01 |
| SERPINF2 | 45,51 | 0,06 | 0,06 | 0,97 | 3,34E-01 |
| PTK7 | 65,29 | 0,06 | 0,08 | 0,82 | 4,10E-01 |
| RAPGEF5 | 68,41 | 0,01 | 0,10 | 0,11 | 9,09E-01 |
| CENPM | 28,18 | 0,05 | 0,07 | 0,74 | 4,59E-01 |
| PRR4 | 25,67 | 0,00 | 0,06 | -0,01 | 9,94E-01 |
| BMERB1 | 47,20 | 0,01 | 0,06 | 0,14 | 8,88E-01 |

| | | | | | |
|---|---|---|---|---|---|
| APOO | 70,72 | 0,06 | 0,03 | 2,17 | 3,01E-02 |
| THBS4 | 67,66 | 0,08 | 0,07 | 1,16 | 2,44E-01 |
| REELD1 | 48,46 | -0,04 | 0,07 | -0,56 | 5,74E-01 |
| LAMB2 | 52,20 | 0,06 | 0,09 | 0,75 | 4,55E-01 |
| TMEM44 | 62,20 | 0,01 | 0,07 | 0,15 | 8,81E-01 |
| PTGES | 31,03 | -0,05 | 0,09 | -0,49 | 6,26E-01 |
| TMEM150A | 57,61 | 0,04 | 0,05 | 0,70 | 4,83E-01 |
| ARHGEF10 | 44,73 | -0,04 | 0,09 | -0,47 | 6,39E-01 |
| CATIP | 27,21 | -0,01 | 0,06 | -0,18 | 8,55E-01 |
| SUGCT | 67,85 | -0,04 | 0,06 | -0,74 | 4,58E-01 |
| WNT5B | 45,41 | 0,02 | 0,09 | 0,18 | 8,60E-01 |
| LRRC34 | 41,02 | 0,14 | 0,06 | 2,46 | 1,39E-02 |
| C3orf20 | 57,91 | -0,11 | 0,07 | -1,58 | 1,14E-01 |
| FPR3 | 60,07 | -0,09 | 0,06 | -1,44 | 1,50E-01 |
| FAM72B | 24,87 | 0,07 | 0,05 | 1,33 | 1,82E-01 |
| ALDH7A1 | 54,59 | 0,03 | 0,11 | 0,30 | 7,65E-01 |
| FDXACB1 | 52,78 | 0,04 | 0,03 | 1,50 | 1,34E-01 |
| SLC1A7 | 60,75 | -0,15 | 0,10 | -1,50 | 1,32E-01 |
| ST6GALNAC1 | 43,22 | 0,01 | 0,07 | 0,19 | 8,52E-01 |
| CAND2 | 34,88 | 0,00 | 0,10 | 0,04 | 9,65E-01 |
| ZDHHC11B | 47,44 | 0,14 | 0,09 | 1,55 | 1,21E-01 |
| FAM161A | 45,35 | 0,05 | 0,06 | 0,83 | 4,06E-01 |
| PJVK | 47,09 | 0,06 | 0,05 | 1,19 | 2,33E-01 |
| RAD51AP1 | 51,73 | 0,09 | 0,06 | 1,56 | 1,18E-01 |
| FAM83H | 53,33 | 0,09 | 0,08 | 1,19 | 2,35E-01 |
| RPH3A | 57,06 | -0,12 | 0,13 | -0,89 | 3,73E-01 |
| STK33 | 65,17 | -0,02 | 0,09 | -0,25 | 8,01E-01 |
| SMPDL3B | 28,75 | -0,10 | 0,07 | -1,41 | 1,58E-01 |
| NOS3 | 51,56 | 0,07 | 0,09 | 0,84 | 4,00E-01 |
| BEX5 | 31,74 | 0,01 | 0,06 | 0,24 | 8,10E-01 |
| P2RX5-TAX1E | 35,04 | 0,06 | 0,07 | 0,86 | 3,89E-01 |
| USE1 | 67,92 | 0,07 | 0,04 | 1,77 | 7,71E-02 |
| YY2 | 53,79 | 0,02 | 0,03 | 0,60 | 5,51E-01 |
| TNFSF15 | 53,28 | 0,08 | 0,07 | 1,14 | 2,56E-01 |
| TPPP | 66,95 | -0,06 | 0,08 | -0,69 | 4,91E-01 |
| RAB3C | 33,64 | -0,01 | 0,09 | -0,13 | 8,98E-01 |
| TMEM30B | 46,54 | 0,00 | 0,06 | 0,02 | 9,86E-01 |
| SAXO1 | 59,05 | 0,06 | 0,08 | 0,68 | 4,99E-01 |
| PLK2 | 46,01 | -0,06 | 0,06 | -1,06 | 2,90E-01 |
| TAS2R41 | 50,46 | -0,08 | 0,08 | -1,10 | 2,71E-01 |
| TECTA | 66,92 | 0,05 | 0,09 | 0,56 | 5,73E-01 |
| CHPF | 50,76 | -0,01 | 0,06 | -0,23 | 8,18E-01 |
| ENSG000001 | 28,33 | 0,05 | 0,04 | 1,14 | 2,55E-01 |
| SH3BP4 | 66,10 | 0,01 | 0,09 | 0,16 | 8,70E-01 |
| RHEBL1 | 43,57 | 0,01 | 0,04 | 0,15 | 8,79E-01 |
| ACOT4 | 36,48 | 0,07 | 0,06 | 1,12 | 2,63E-01 |

| | | | | | |
|---|---|---|---|---|---|
| NT5DC2 | 35,64 | 0,02 | 0,09 | 0,19 | 8,47E-01 |
| KANK3 | 53,99 | 0,04 | 0,07 | 0,60 | 5,51E-01 |
| RNF212 | 41,58 | 0,13 | 0,08 | 1,58 | 1,14E-01 |
| XKR9 | 40,19 | 0,07 | 0,08 | 0,87 | 3,82E-01 |
| RASSF6 | 44,19 | 0,07 | 0,08 | 0,85 | 3,94E-01 |
| ATAT1 | 43,94 | 0,04 | 0,05 | 0,84 | 4,01E-01 |
| MPV17L2 | 62,95 | 0,03 | 0,03 | 0,92 | 3,58E-01 |
| MYPOP | 59,82 | 0,05 | 0,06 | 0,72 | 4,72E-01 |
| C12orf60 | 28,47 | 0,01 | 0,07 | 0,18 | 8,59E-01 |
| DNASE1L3 | 32,26 | 0,02 | 0,07 | 0,32 | 7,52E-01 |
| ENG | 46,40 | 0,08 | 0,09 | 0,84 | 3,99E-01 |
| NDUFA13 | 64,11 | 0,00 | 0,05 | 0,07 | 9,44E-01 |
| CD1A | 41,14 | 0,06 | 0,06 | 0,97 | 3,32E-01 |
| RSPH14 | 45,07 | -0,20 | 0,07 | -2,98 | 2,85E-03 |
| TSPO2 | 61,24 | 0,12 | 0,08 | 1,48 | 1,38E-01 |
| ZCWPW2 | 41,78 | 0,04 | 0,04 | 1,07 | 2,85E-01 |
| KY | 37,79 | -0,03 | 0,08 | -0,45 | 6,54E-01 |
| ZNF223 | 58,78 | 0,03 | 0,03 | 0,85 | 3,97E-01 |
| OGFOD2 | 66,26 | 0,02 | 0,03 | 0,75 | 4,55E-01 |
| LEKR1 | 54,10 | 0,14 | 0,07 | 1,94 | 5,26E-02 |
| C1R | 34,93 | -0,06 | 0,08 | -0,75 | 4,52E-01 |
| SLC5A9 | 32,26 | -0,13 | 0,10 | -1,34 | 1,80E-01 |
| ARMCX1 | 52,34 | 0,02 | 0,04 | 0,48 | 6,34E-01 |
| SPTLC3 | 54,43 | 0,08 | 0,09 | 0,90 | 3,67E-01 |
| CALCB | 53,51 | -0,08 | 0,07 | -1,23 | 2,19E-01 |
| AMDHD1 | 53,92 | 0,07 | 0,05 | 1,51 | 1,32E-01 |
| TAF11L2 | 33,10 | -0,30 | 0,07 | -4,15 | 3,31E-05 |
| DOCK1 | 69,28 | -0,01 | 0,10 | -0,08 | 9,36E-01 |
| MYCBPAP | 52,74 | -0,01 | 0,10 | -0,11 | 9,14E-01 |
| TBCEL-TECT/ | 27,20 | 0,06 | 0,07 | 0,83 | 4,08E-01 |
| MPPED2 | 39,32 | 0,07 | 0,12 | 0,59 | 5,57E-01 |
| ZNF366 | 52,15 | -0,02 | 0,06 | -0,30 | 7,63E-01 |
| ANG | 27,55 | -0,03 | 0,05 | -0,60 | 5,51E-01 |
| CATSPERE | 67,81 | 0,07 | 0,05 | 1,43 | 1,52E-01 |
| EFCAB5 | 71,62 | 0,06 | 0,06 | 0,94 | 3,48E-01 |
| RAMACL | 57,40 | -0,05 | 0,05 | -0,96 | 3,39E-01 |
| H2BK1 | 22,97 | 0,00 | 0,04 | 0,03 | 9,76E-01 |
| RDH5 | 68,45 | -0,03 | 0,06 | -0,53 | 5,97E-01 |
| TDRD15 | 34,12 | 0,15 | 0,11 | 1,37 | 1,72E-01 |
| MMP28 | 46,06 | 0,01 | 0,09 | 0,17 | 8,66E-01 |
| SNTG2 | 65,47 | 0,10 | 0,09 | 1,06 | 2,88E-01 |
| GPR63 | 45,58 | 0,08 | 0,07 | 1,12 | 2,63E-01 |
| ZNF219 | 68,70 | 0,04 | 0,07 | 0,59 | 5,55E-01 |
| S1PR2 | 52,43 | 0,03 | 0,05 | 0,63 | 5,29E-01 |
| CTSE | 67,63 | 0,08 | 0,09 | 0,94 | 3,48E-01 |
| PCDHGA8 | 36,37 | -0,10 | 0,07 | -1,39 | 1,63E-01 |

| | | | | | |
|---|---|---|---|---|---|
| GRIP1 | 70,07 | 0,10 | 0,08 | 1,33 | 1,84E-01 |
| NUAK1 | 66,72 | -0,10 | 0,10 | -0,96 | 3,37E-01 |
| DLC1 | 45,68 | -0,02 | 0,12 | -0,16 | 8,71E-01 |
| CNN3 | 33,28 | 0,08 | 0,07 | 1,13 | 2,59E-01 |
| KCNJ5-AS1 | 28,68 | 0,07 | 0,08 | 0,88 | 3,77E-01 |
| CERS3 | 58,67 | -0,05 | 0,08 | -0,64 | 5,22E-01 |
| DIPK1B | 42,02 | 0,13 | 0,08 | 1,70 | 8,83E-02 |
| ZMYND15 | 33,26 | 0,03 | 0,09 | 0,36 | 7,17E-01 |
| EME1 | 26,05 | 0,01 | 0,07 | 0,18 | 8,54E-01 |
| XKR3 | 58,14 | -0,22 | 0,12 | -1,84 | 6,65E-02 |
| CCNE1 | 38,68 | 0,02 | 0,06 | 0,30 | 7,67E-01 |
| CXCL6 | 31,20 | 0,00 | 0,06 | -0,07 | 9,43E-01 |
| SCUBE2 | 66,33 | -0,09 | 0,08 | -1,03 | 3,04E-01 |
| PTGER3 | 35,80 | 0,08 | 0,11 | 0,74 | 4,57E-01 |
| FAM156A | 46,29 | -0,05 | 0,03 | -1,73 | 8,33E-02 |
| JMJD7 | 44,28 | -0,01 | 0,05 | -0,24 | 8,13E-01 |
| PXMP2 | 25,60 | 0,07 | 0,04 | 1,67 | 9,49E-02 |
| GSTA4 | 31,73 | 0,11 | 0,06 | 1,78 | 7,52E-02 |
| FSCN3 | 41,45 | -0,02 | 0,05 | -0,38 | 7,07E-01 |
| PIWIL2 | 48,99 | -0,02 | 0,09 | -0,20 | 8,43E-01 |
| CYP7B1 | 31,48 | 0,10 | 0,08 | 1,25 | 2,11E-01 |
| KLHL13 | 35,18 | 0,02 | 0,07 | 0,23 | 8,18E-01 |
| VSIG4 | 54,15 | -0,06 | 0,06 | -0,94 | 3,49E-01 |
| MOXD1 | 28,80 | -0,01 | 0,09 | -0,11 | 9,16E-01 |
| PAQR9 | 34,71 | 0,13 | 0,11 | 1,17 | 2,41E-01 |
| ESR2 | 54,87 | -0,02 | 0,10 | -0,22 | 8,30E-01 |
| DNMT3B | 41,20 | 0,00 | 0,10 | 0,00 | 1,00E+00 |
| IQCC | 27,51 | 0,06 | 0,05 | 1,27 | 2,03E-01 |
| CDC42EP2 | 54,19 | -0,17 | 0,05 | -3,27 | 1,08E-03 |
| KRT18 | 56,52 | 0,08 | 0,06 | 1,40 | 1,61E-01 |
| ZWINT | 70,94 | -0,03 | 0,05 | -0,57 | 5,66E-01 |
| FAM200C | 65,92 | 0,07 | 0,04 | 1,73 | 8,43E-02 |
| DLEU7 | 48,78 | 0,03 | 0,06 | 0,47 | 6,38E-01 |
| ENSG000002 | 50,63 | 0,06 | 0,04 | 1,64 | 1,01E-01 |
| SERPINB2 | 61,88 | -0,05 | 0,06 | -0,84 | 4,03E-01 |
| NUTM2D | 38,18 | 0,00 | 0,06 | -0,04 | 9,67E-01 |
| PPFIA3 | 52,43 | 0,03 | 0,10 | 0,31 | 7,57E-01 |
| IFT22 | 36,94 | 0,10 | 0,06 | 1,72 | 8,63E-02 |
| SPATA9 | 32,99 | 0,02 | 0,08 | 0,28 | 7,79E-01 |
| SCNN1D | 59,67 | 0,04 | 0,07 | 0,60 | 5,48E-01 |
| NHSL3 | 36,21 | 0,03 | 0,10 | 0,31 | 7,56E-01 |
| HDAC10 | 55,91 | -0,01 | 0,06 | -0,15 | 8,79E-01 |
| ENSG000002 | 64,09 | -0,01 | 0,03 | -0,39 | 6,93E-01 |
| ALG1L2 | 64,67 | 0,07 | 0,07 | 1,13 | 2,58E-01 |
| CD70 | 27,16 | 0,05 | 0,06 | 0,73 | 4,67E-01 |
| PCDHGB5 | 35,74 | -0,06 | 0,07 | -0,98 | 3,28E-01 |

| | | | | | |
|---|---|---|---|---|---|
| SLC16A4 | 36,72 | 0,05 | 0,06 | 0,88 | 3,80E-01 |
| PERP | 57,01 | 0,02 | 0,06 | 0,33 | 7,44E-01 |
| RMI2 | 57,00 | -0,12 | 0,08 | -1,53 | 1,26E-01 |
| KISS1R | 26,85 | 0,09 | 0,07 | 1,34 | 1,79E-01 |
| ZNF497 | 27,41 | 0,03 | 0,07 | 0,43 | 6,66E-01 |
| OR4D1 | 49,43 | 0,00 | 0,05 | -0,08 | 9,38E-01 |
| GPR84 | 29,61 | -0,01 | 0,07 | -0,10 | 9,16E-01 |
| ENSG000002 | 38,94 | 0,02 | 0,04 | 0,58 | 5,65E-01 |
| SPSB1 | 44,75 | 0,08 | 0,07 | 1,26 | 2,08E-01 |
| H1-6 | 50,04 | -0,25 | 0,05 | -4,79 | 1,65E-06 |
| ZBTB46 | 53,67 | 0,04 | 0,07 | 0,50 | 6,14E-01 |
| IGSF22 | 41,18 | -0,05 | 0,10 | -0,47 | 6,38E-01 |
| NUSAP1 | 35,55 | 0,07 | 0,04 | 1,64 | 1,00E-01 |
| LRP2BP | 58,86 | 0,09 | 0,06 | 1,47 | 1,43E-01 |
| STBD1 | 42,87 | 0,05 | 0,05 | 0,96 | 3,35E-01 |
| LCAT | 36,97 | 0,04 | 0,06 | 0,67 | 5,00E-01 |
| ZDHHC11 | 57,78 | 0,00 | 0,10 | 0,04 | 9,68E-01 |
| CCDC154 | 55,72 | 0,03 | 0,09 | 0,37 | 7,09E-01 |
| HAS3 | 41,28 | -0,04 | 0,08 | -0,51 | 6,07E-01 |
| EFCAB12 | 65,05 | -0,18 | 0,07 | -2,63 | 8,66E-03 |
| LILRA4 | 58,32 | 0,03 | 0,07 | 0,44 | 6,63E-01 |
| TMEM191C | 24,96 | -0,04 | 0,06 | -0,67 | 5,01E-01 |
| MSH5 | 57,54 | 0,08 | 0,05 | 1,59 | 1,13E-01 |
| POU5F1B | 45,85 | 0,05 | 0,06 | 0,75 | 4,52E-01 |
| AKR1C1 | 62,32 | 0,13 | 0,08 | 1,73 | 8,41E-02 |
| APBB2 | 52,32 | 0,09 | 0,12 | 0,75 | 4,55E-01 |
| HLF | 52,79 | 0,02 | 0,08 | 0,26 | 7,92E-01 |
| ZNF853 | 64,16 | 0,10 | 0,05 | 1,87 | 6,09E-02 |
| SPRY1 | 53,75 | 0,08 | 0,07 | 1,19 | 2,34E-01 |
| MMP14 | 50,31 | 0,12 | 0,07 | 1,73 | 8,37E-02 |
| USP6 | 51,91 | 0,03 | 0,12 | 0,27 | 7,88E-01 |
| SLC23A3 | 25,66 | -0,09 | 0,07 | -1,29 | 1,98E-01 |
| EVC | 68,43 | -0,07 | 0,10 | -0,67 | 5,00E-01 |
| TMSB15B | 45,40 | 0,04 | 0,04 | 0,97 | 3,32E-01 |
| SYNGR1 | 51,09 | 0,04 | 0,08 | 0,50 | 6,17E-01 |
| ZNF610 | 36,43 | -0,01 | 0,06 | -0,19 | 8,52E-01 |
| PCDHGB6 | 67,82 | -0,04 | 0,06 | -0,56 | 5,73E-01 |
| RAB36 | 65,37 | -0,11 | 0,07 | -1,51 | 1,30E-01 |
| TEK | 67,27 | -0,19 | 0,10 | -1,88 | 6,03E-02 |
| FN3K | 45,85 | 0,09 | 0,07 | 1,26 | 2,09E-01 |
| ROM1 | 29,56 | 0,06 | 0,06 | 1,02 | 3,06E-01 |
| SLC25A23 | 61,71 | 0,03 | 0,06 | 0,52 | 6,02E-01 |
| C9orf153 | 41,78 | -0,04 | 0,05 | -0,83 | 4,06E-01 |
| DLGAP5 | 58,93 | 0,09 | 0,08 | 1,03 | 3,01E-01 |
| MIS18A | 65,56 | 0,01 | 0,03 | 0,43 | 6,67E-01 |
| ODAD4 | 54,95 | 0,05 | 0,08 | 0,67 | 5,03E-01 |

| | | | | | |
|---|---|---|---|---|---|
| NLGN4Y | 30,27 | 0,07 | 0,07 | 0,95 | 3,40E-01 |
| PLCH1 | 43,19 | 0,01 | 0,10 | 0,05 | 9,58E-01 |
| APOM | 31,40 | 0,01 | 0,03 | 0,41 | 6,81E-01 |
| CDCA2 | 30,89 | 0,03 | 0,08 | 0,40 | 6,91E-01 |
| SERPINI2 | 58,27 | 0,05 | 0,06 | 0,84 | 4,01E-01 |
| HACD1 | 59,22 | 0,17 | 0,06 | 2,76 | 5,83E-03 |
| PMFBP1 | 60,38 | 0,04 | 0,08 | 0,50 | 6,16E-01 |
| DMGDH | 38,65 | 0,02 | 0,10 | 0,18 | 8,56E-01 |
| COX16 | 69,27 | 0,07 | 0,03 | 2,36 | 1,85E-02 |
| PSAT1 | 65,78 | -0,04 | 0,05 | -0,87 | 3,83E-01 |
| SGO1 | 33,11 | 0,13 | 0,06 | 2,18 | 2,94E-02 |
| DENND6B | 46,90 | 0,13 | 0,08 | 1,67 | 9,44E-02 |
| FHAD1 | 58,59 | 0,04 | 0,14 | 0,31 | 7,56E-01 |
| TEX22 | 64,90 | 0,06 | 0,04 | 1,26 | 2,08E-01 |
| PDE9A | 63,80 | 0,06 | 0,07 | 0,84 | 3,98E-01 |
| CDKL3 | 39,66 | 0,05 | 0,04 | 1,24 | 2,13E-01 |
| MISP3 | 27,60 | 0,06 | 0,06 | 1,01 | 3,10E-01 |
| MTFP1 | 34,27 | -0,01 | 0,04 | -0,19 | 8,49E-01 |
| RPS6KL1 | 37,83 | 0,09 | 0,09 | 0,98 | 3,30E-01 |
| MAOA | 54,45 | 0,14 | 0,11 | 1,33 | 1,84E-01 |
| SPRY2 | 64,41 | 0,07 | 0,05 | 1,40 | 1,61E-01 |
| CCDC150 | 44,70 | 0,05 | 0,11 | 0,42 | 6,74E-01 |
| TIFAB | 47,19 | 0,08 | 0,06 | 1,36 | 1,74E-01 |
| SLC47A1 | 53,82 | -0,01 | 0,10 | -0,12 | 9,06E-01 |
| ENSG000002 | 46,49 | -0,08 | 0,04 | -1,87 | 6,08E-02 |
| SLC22A17 | 37,50 | 0,04 | 0,08 | 0,45 | 6,54E-01 |
| SOCS1 | 62,91 | -0,09 | 0,07 | -1,41 | 1,60E-01 |
| APOL4 | 43,69 | -0,06 | 0,10 | -0,60 | 5,52E-01 |
| DAB1 | 56,63 | 0,01 | 0,08 | 0,08 | 9,39E-01 |
| CLDN7 | 29,22 | 0,06 | 0,07 | 0,96 | 3,37E-01 |
| NPIPB11 | 37,00 | 0,00 | 0,08 | -0,05 | 9,61E-01 |
| TFCP2L1 | 63,92 | -0,05 | 0,12 | -0,46 | 6,43E-01 |
| TMEM200A | 56,26 | 0,02 | 0,07 | 0,33 | 7,45E-01 |
| FBXO39 | 31,79 | -0,14 | 0,09 | -1,67 | 9,50E-02 |
| RIMBP3 | 64,09 | 0,01 | 0,05 | 0,21 | 8,31E-01 |
| SLC25A2 | 27,58 | 0,02 | 0,05 | 0,45 | 6,49E-01 |
| ZNF713 | 54,25 | 0,02 | 0,05 | 0,44 | 6,62E-01 |
| FIGNL2 | 60,68 | 0,02 | 0,08 | 0,23 | 8,15E-01 |
| PARD3B | 62,57 | 0,00 | 0,10 | -0,02 | 9,87E-01 |
| FABP5 | 41,52 | 0,03 | 0,06 | 0,56 | 5,77E-01 |
| LRRC20 | 61,06 | 0,05 | 0,05 | 1,01 | 3,14E-01 |
| ENSG000002 | 36,37 | -0,11 | 0,06 | -1,99 | 4,63E-02 |
| LIMS2 | 59,93 | 0,03 | 0,09 | 0,37 | 7,14E-01 |
| LRRC39 | 26,26 | 0,10 | 0,05 | 1,89 | 5,82E-02 |
| FSTL3 | 65,75 | 0,06 | 0,05 | 1,28 | 2,02E-01 |
| CHAC2 | 54,65 | 0,06 | 0,05 | 1,10 | 2,71E-01 |

| | | | | | |
|---|---|---|---|---|---|
| CILP | 37,97 | -0,07 | 0,08 | -0,90 | 3,68E-01 |
| SHPK | 33,19 | 0,03 | 0,04 | 0,79 | 4,27E-01 |
| NETO1 | 54,92 | 0,02 | 0,08 | 0,23 | 8,15E-01 |
