## Supplementary material for "Longitudinal Assessment of DNA Repair Signature Trajectory in Prodromal versus Established Parkinson’s Disease": Table S3

| | baseMean | log2FoldCha | lfcSE | stat | pvalue | padj |
|---|---|---|---|---|---|---|
| CACNA1A | 124,99 | -1,56 | 0,35 | -4,48 | 7,58E-06 | 9,36E-02 |
| FHDC1 | 92,26 | -0,49 | 0,12 | -3,94 | 8,12E-05 | 1,28E-01 |
| COL4A1 | 64,51 | -1,14 | 0,29 | -3,94 | 8,05E-05 | 1,28E-01 |
| NRXN2 | 56,36 | -1,08 | 0,27 | -3,96 | 7,60E-05 | 1,28E-01 |
| FARP1 | 108,20 | -1,32 | 0,32 | -4,05 | 5,14E-05 | 1,28E-01 |
| IFI27 | 408,75 | -1,11 | 0,27 | -4,11 | 3,95E-05 | 1,28E-01 |
| RTN2 | 95,02 | -0,44 | 0,10 | -4,17 | 3,06E-05 | 1,28E-01 |
| ARHGEF4 | 61,27 | -1,00 | 0,25 | -3,94 | 8,32E-05 | 1,28E-01 |
| RHCE | 64,63 | -0,56 | 0,15 | -3,84 | 1,23E-04 | 1,42E-01 |
| PITPNM3 | 62,58 | -0,96 | 0,25 | -3,83 | 1,27E-04 | 1,42E-01 |
| HBM | 228,26 | -0,65 | 0,17 | -3,85 | 1,18E-04 | 1,42E-01 |
| PDZD7 | 41,10 | -0,86 | 0,23 | -3,71 | 2,07E-04 | 1,46E-01 |
| ENSG000002 | 278,79 | -0,34 | 0,09 | -3,74 | 1,81E-04 | 1,46E-01 |
| E2F1 | 63,46 | -0,40 | 0,11 | -3,64 | 2,71E-04 | 1,46E-01 |
| PLCD3 | 35,38 | -0,82 | 0,23 | -3,66 | 2,53E-04 | 1,46E-01 |
| ADGRB2 | 57,40 | -0,95 | 0,25 | -3,73 | 1,92E-04 | 1,46E-01 |
| FAM89B | 102,45 | -0,24 | 0,07 | -3,65 | 2,62E-04 | 1,46E-01 |
| MST1 | 58,15 | -0,38 | 0,10 | -3,76 | 1,69E-04 | 1,46E-01 |
| FOXD2 | 40,06 | -0,84 | 0,23 | -3,72 | 1,98E-04 | 1,46E-01 |
| RAPGEF3 | 52,46 | -1,02 | 0,28 | -3,64 | 2,71E-04 | 1,46E-01 |
| PF4V1 | 96,71 | -0,71 | 0,19 | -3,71 | 2,06E-04 | 1,46E-01 |
| LRRC2 | 40,24 | -0,70 | 0,19 | -3,65 | 2,65E-04 | 1,46E-01 |
| TNNC2 | 30,15 | -0,55 | 0,15 | -3,67 | 2,42E-04 | 1,46E-01 |
| RYR1 | 123,22 | -0,85 | 0,24 | -3,54 | 3,97E-04 | 1,46E-01 |
| LSG1 | 982,80 | 0,16 | 0,04 | 3,46 | 5,45E-04 | 1,46E-01 |
| WDR11 | 2925,85 | 0,17 | 0,05 | 3,59 | 3,26E-04 | 1,46E-01 |
| SKOR1 | 47,70 | -0,81 | 0,23 | -3,47 | 5,13E-04 | 1,46E-01 |
| YPEL4 | 49,20 | -0,47 | 0,13 | -3,57 | 3,59E-04 | 1,46E-01 |
| RPL36A | 170,62 | 0,17 | 0,05 | 3,58 | 3,41E-04 | 1,46E-01 |
| DNAH17 | 67,31 | -1,04 | 0,30 | -3,48 | 5,02E-04 | 1,46E-01 |
| DLGAP3 | 52,46 | -0,84 | 0,24 | -3,47 | 5,29E-04 | 1,46E-01 |
| MAPK10 | 94,52 | -1,00 | 0,28 | -3,53 | 4,16E-04 | 1,46E-01 |
| LRP5 | 71,61 | -0,68 | 0,19 | -3,51 | 4,50E-04 | 1,46E-01 |
| GLB1L3 | 45,49 | -0,92 | 0,26 | -3,60 | 3,18E-04 | 1,46E-01 |
| VSIG10 | 168,18 | -0,44 | 0,12 | -3,63 | 2,88E-04 | 1,46E-01 |
| TF | 72,00 | -1,00 | 0,28 | -3,54 | 3,98E-04 | 1,46E-01 |
| TMEM179B | 261,36 | -0,18 | 0,05 | -3,54 | 4,05E-04 | 1,46E-01 |
| ITGA7 | 52,72 | -0,71 | 0,20 | -3,49 | 4,87E-04 | 1,46E-01 |
| TSPO2 | 70,66 | -0,54 | 0,15 | -3,53 | 4,20E-04 | 1,46E-01 |
| PCDH9 | 1232,67 | 0,49 | 0,14 | 3,50 | 4,58E-04 | 1,46E-01 |
| ARMCX5 | 345,49 | 0,17 | 0,05 | 3,50 | 4,59E-04 | 1,46E-01 |
| ZNF121 | 1935,49 | 0,19 | 0,05 | 3,59 | 3,34E-04 | 1,46E-01 |
| DLGAP1 | 50,55 | -0,86 | 0,25 | -3,46 | 5,38E-04 | 1,46E-01 |
| DLC1 | 49,37 | -0,81 | 0,23 | -3,53 | 4,14E-04 | 1,46E-01 |
| EXOSC6 | 1219,63 | 0,15 | 0,04 | 3,49 | 4,78E-04 | 1,46E-01 |

| | | | | | | |
|---|---|---|---|---|---|---|
| DNAH10 | 122,61 | -0,81 | 0,23 | -3,48 | 4,94E-04 | 1,46E-01 |
| PLXNB1 | 52,71 | -0,80 | 0,23 | -3,45 | 5,59E-04 | 1,47E-01 |
| ADGRB1 | 95,35 | -0,79 | 0,23 | -3,41 | 6,60E-04 | 1,50E-01 |
| AP3B2 | 53,79 | -0,97 | 0,29 | -3,41 | 6,41E-04 | 1,50E-01 |
| TRMT9B | 46,43 | -0,88 | 0,26 | -3,38 | 7,15E-04 | 1,50E-01 |
| TRIB3 | 106,69 | -0,34 | 0,10 | -3,43 | 5,96E-04 | 1,50E-01 |
| CLCN2 | 42,46 | -0,66 | 0,20 | -3,38 | 7,15E-04 | 1,50E-01 |
| BCAM | 49,45 | -0,79 | 0,23 | -3,41 | 6,59E-04 | 1,50E-01 |
| CTSE | 72,63 | -0,60 | 0,18 | -3,42 | 6,33E-04 | 1,50E-01 |
| UBE2C | 33,80 | -0,38 | 0,11 | -3,42 | 6,33E-04 | 1,50E-01 |
| CERCAM | 36,91 | -0,81 | 0,24 | -3,40 | 6,69E-04 | 1,50E-01 |
| PRDM8 | 353,13 | -0,21 | 0,06 | -3,39 | 6,94E-04 | 1,50E-01 |
| FAM83F | 80,95 | -0,72 | 0,21 | -3,42 | 6,15E-04 | 1,50E-01 |
| CAND2 | 37,90 | -0,67 | 0,20 | -3,38 | 7,14E-04 | 1,50E-01 |
| TMC5 | 73,30 | -0,59 | 0,17 | -3,37 | 7,47E-04 | 1,51E-01 |
| ZNF551 | 494,57 | 0,14 | 0,04 | 3,37 | 7,40E-04 | 1,51E-01 |
| FAM241B | 29,59 | -0,45 | 0,13 | -3,36 | 7,66E-04 | 1,53E-01 |
| JAKMIP3 | 54,56 | -0,73 | 0,22 | -3,28 | 1,03E-03 | 1,55E-01 |
| GPRC5C | 38,02 | -0,78 | 0,24 | -3,32 | 9,12E-04 | 1,55E-01 |
| CLNS1A | 1332,56 | 0,16 | 0,05 | 3,25 | 1,17E-03 | 1,55E-01 |
| ATMIN | 1228,48 | 0,11 | 0,04 | 3,26 | 1,12E-03 | 1,55E-01 |
| RAB3IL1 | 85,54 | -0,55 | 0,17 | -3,24 | 1,22E-03 | 1,55E-01 |
| PSMD9 | 309,92 | -0,14 | 0,04 | -3,23 | 1,22E-03 | 1,55E-01 |
| NT5DC4 | 36,31 | -0,55 | 0,17 | -3,28 | 1,04E-03 | 1,55E-01 |
| LMTK3 | 70,95 | -0,62 | 0,19 | -3,24 | 1,18E-03 | 1,55E-01 |
| MEF2C | 3293,77 | 0,24 | 0,07 | 3,29 | 9,97E-04 | 1,55E-01 |
| SLC29A1 | 141,58 | -0,50 | 0,15 | -3,23 | 1,22E-03 | 1,55E-01 |
| KIFBP | 360,22 | 0,12 | 0,04 | 3,28 | 1,03E-03 | 1,55E-01 |
| PARVA | 48,96 | -0,81 | 0,25 | -3,29 | 9,92E-04 | 1,55E-01 |
| PIP5KL1 | 39,15 | -0,49 | 0,15 | -3,33 | 8,69E-04 | 1,55E-01 |
| CACNA1E | 378,18 | -0,46 | 0,14 | -3,26 | 1,13E-03 | 1,55E-01 |
| AGBL5 | 119,62 | -0,30 | 0,09 | -3,26 | 1,11E-03 | 1,55E-01 |
| PLSCR4 | 40,72 | -0,46 | 0,14 | -3,35 | 8,21E-04 | 1,55E-01 |
| PTS | 425,51 | 0,13 | 0,04 | 3,32 | 9,04E-04 | 1,55E-01 |
| KIF19 | 67,21 | -0,67 | 0,21 | -3,26 | 1,12E-03 | 1,55E-01 |
| GPR153 | 49,86 | -0,60 | 0,19 | -3,26 | 1,11E-03 | 1,55E-01 |
| MPST | 446,41 | -0,22 | 0,07 | -3,25 | 1,15E-03 | 1,55E-01 |
| FASTKD3 | 269,20 | 0,17 | 0,05 | 3,33 | 8,64E-04 | 1,55E-01 |
| MICALL2 | 80,05 | -0,61 | 0,18 | -3,34 | 8,37E-04 | 1,55E-01 |
| SNAPC5 | 419,47 | 0,13 | 0,04 | 3,26 | 1,12E-03 | 1,55E-01 |
| MATN2 | 40,60 | -0,80 | 0,25 | -3,23 | 1,23E-03 | 1,55E-01 |
| RABEP1 | 2175,94 | 0,14 | 0,04 | 3,24 | 1,18E-03 | 1,55E-01 |
| ESPN | 210,97 | -0,68 | 0,21 | -3,24 | 1,22E-03 | 1,55E-01 |
| GGT5 | 30,95 | -0,78 | 0,23 | -3,34 | 8,25E-04 | 1,55E-01 |
| CYP4F22 | 25,96 | -0,64 | 0,19 | -3,30 | 9,57E-04 | 1,55E-01 |
| RIOX2 | 1040,10 | 0,16 | 0,05 | 3,27 | 1,06E-03 | 1,55E-01 |

| | | | | | | |
|---|---|---|---|---|---|---|
| C17orf99 | 25,79 | -0,55 | 0,17 | -3,24 | 1,21E-03 | 1,55E-01 |
| ZNF44 | 1184,05 | 0,14 | 0,04 | 3,24 | 1,18E-03 | 1,55E-01 |
| TTC39A | 78,88 | -0,68 | 0,20 | -3,30 | 9,56E-04 | 1,55E-01 |
| MYO18B | 84,05 | -1,03 | 0,31 | -3,32 | 8,90E-04 | 1,55E-01 |
| TIMM9 | 82,88 | 0,20 | 0,06 | 3,26 | 1,10E-03 | 1,55E-01 |
| LONRF2 | 62,63 | -0,86 | 0,26 | -3,28 | 1,04E-03 | 1,55E-01 |
| PKD1L3 | 37,53 | -0,60 | 0,18 | -3,26 | 1,12E-03 | 1,55E-01 |
| NRN1 | 42,62 | -0,63 | 0,19 | -3,22 | 1,28E-03 | 1,55E-01 |
| ALKBH3 | 516,94 | 0,14 | 0,04 | 3,22 | 1,27E-03 | 1,55E-01 |
| PKN3 | 32,55 | -0,69 | 0,22 | -3,22 | 1,29E-03 | 1,55E-01 |
| DYNLRB1 | 386,40 | -0,14 | 0,05 | -3,22 | 1,28E-03 | 1,55E-01 |
| CRYBG2 | 58,06 | -0,45 | 0,14 | -3,22 | 1,29E-03 | 1,55E-01 |
| SMIM10L2A | 37,52 | -0,41 | 0,13 | -3,16 | 1,59E-03 | 1,56E-01 |
| GPRC5B | 47,10 | -0,82 | 0,26 | -3,16 | 1,56E-03 | 1,56E-01 |
| MAMLD1 | 42,60 | -0,45 | 0,14 | -3,20 | 1,38E-03 | 1,56E-01 |
| EPOR | 129,41 | -0,32 | 0,10 | -3,10 | 1,93E-03 | 1,56E-01 |
| TYMS | 33,69 | -0,58 | 0,18 | -3,12 | 1,83E-03 | 1,56E-01 |
| CYP26C1 | 83,45 | -0,37 | 0,12 | -3,14 | 1,68E-03 | 1,56E-01 |
| ASAP3 | 47,63 | -0,61 | 0,20 | -3,11 | 1,87E-03 | 1,56E-01 |
| SLC5A9 | 33,80 | -0,61 | 0,20 | -3,12 | 1,80E-03 | 1,56E-01 |
| KCNH2 | 100,87 | -0,48 | 0,15 | -3,12 | 1,83E-03 | 1,56E-01 |
| HEATR3 | 977,79 | 0,15 | 0,05 | 3,17 | 1,55E-03 | 1,56E-01 |
| MMEL1 | 37,63 | -0,64 | 0,20 | -3,16 | 1,56E-03 | 1,56E-01 |
| ATP8B3 | 78,26 | -0,44 | 0,14 | -3,12 | 1,83E-03 | 1,56E-01 |
| DARS2 | 412,68 | 0,09 | 0,03 | 3,16 | 1,55E-03 | 1,56E-01 |
| AGRN | 114,81 | -0,70 | 0,22 | -3,13 | 1,73E-03 | 1,56E-01 |
| SEMA6B | 42,26 | -0,66 | 0,21 | -3,20 | 1,35E-03 | 1,56E-01 |
| SDK1 | 85,43 | -0,78 | 0,25 | -3,13 | 1,77E-03 | 1,56E-01 |
| GPR146 | 740,37 | -0,48 | 0,15 | -3,17 | 1,54E-03 | 1,56E-01 |
| APPL1 | 2135,50 | 0,18 | 0,06 | 3,16 | 1,58E-03 | 1,56E-01 |
| DTD2 | 226,77 | 0,16 | 0,05 | 3,13 | 1,75E-03 | 1,56E-01 |
| DNAJC21 | 1180,91 | 0,15 | 0,05 | 3,10 | 1,92E-03 | 1,56E-01 |
| EEF1B2 | 1607,04 | 0,44 | 0,14 | 3,13 | 1,77E-03 | 1,56E-01 |
| GPAT2 | 44,75 | -0,56 | 0,18 | -3,10 | 1,95E-03 | 1,56E-01 |
| PROB1 | 32,09 | -0,68 | 0,21 | -3,19 | 1,40E-03 | 1,56E-01 |
| SYNJ2BP | 1001,86 | 0,16 | 0,05 | 3,10 | 1,97E-03 | 1,56E-01 |
| GFM1 | 1331,38 | 0,14 | 0,04 | 3,16 | 1,58E-03 | 1,56E-01 |
| CTSO | 808,62 | 0,18 | 0,06 | 3,20 | 1,39E-03 | 1,56E-01 |
| B4GALT3 | 396,71 | -0,27 | 0,09 | -3,15 | 1,64E-03 | 1,56E-01 |
| TMEM163 | 42,03 | -0,71 | 0,22 | -3,18 | 1,48E-03 | 1,56E-01 |
| SPIRE2 | 44,61 | -0,65 | 0,21 | -3,11 | 1,89E-03 | 1,56E-01 |
| LRPPRC | 2073,57 | 0,17 | 0,05 | 3,14 | 1,71E-03 | 1,56E-01 |
| ANKLE1 | 36,97 | -0,36 | 0,12 | -3,12 | 1,82E-03 | 1,56E-01 |
| ILDR2 | 51,28 | -0,77 | 0,25 | -3,12 | 1,83E-03 | 1,56E-01 |
| MYRF | 47,21 | -0,90 | 0,28 | -3,19 | 1,40E-03 | 1,56E-01 |
| ZNF136 | 897,08 | 0,13 | 0,04 | 3,14 | 1,67E-03 | 1,56E-01 |

| | | | | | | |
|---|---|---|---|---|---|---|
| COL11A2 | 56,12 | -0,77 | 0,25 | -3,12 | 1,83E-03 | 1,56E-01 |
| SH3PXD2B | 45,05 | -0,70 | 0,23 | -3,10 | 1,92E-03 | 1,56E-01 |
| CORO6 | 40,19 | -0,46 | 0,15 | -3,10 | 1,91E-03 | 1,56E-01 |
| BDH2 | 161,70 | 0,26 | 0,08 | 3,10 | 1,92E-03 | 1,56E-01 |
| SLC5A2 | 35,39 | -0,66 | 0,21 | -3,17 | 1,51E-03 | 1,56E-01 |
| GLB1L2 | 67,84 | -0,69 | 0,22 | -3,12 | 1,78E-03 | 1,56E-01 |
| COX6C | 439,45 | 0,26 | 0,08 | 3,16 | 1,59E-03 | 1,56E-01 |
| CLEC18B | 49,35 | -0,60 | 0,19 | -3,10 | 1,96E-03 | 1,56E-01 |
| ACHE | 41,67 | -0,66 | 0,21 | -3,17 | 1,54E-03 | 1,56E-01 |
| CHPF | 52,45 | -0,40 | 0,13 | -3,18 | 1,47E-03 | 1,56E-01 |
| CCNE1 | 40,59 | -0,43 | 0,14 | -3,16 | 1,60E-03 | 1,56E-01 |
| ATP8A2 | 99,39 | -0,56 | 0,17 | -3,19 | 1,41E-03 | 1,56E-01 |
| ALDH7A1 | 60,82 | -0,73 | 0,23 | -3,18 | 1,48E-03 | 1,56E-01 |
| PPFIA3 | 57,66 | -0,64 | 0,20 | -3,14 | 1,71E-03 | 1,56E-01 |
| DNAH8 | 102,84 | -0,76 | 0,24 | -3,21 | 1,34E-03 | 1,56E-01 |
| ABCB7 | 757,92 | 0,16 | 0,05 | 3,10 | 1,91E-03 | 1,56E-01 |
| PDZK1IP1 | 709,46 | -0,43 | 0,14 | -3,11 | 1,87E-03 | 1,56E-01 |
| HLA-DMB | 2879,61 | 0,19 | 0,06 | 3,18 | 1,48E-03 | 1,56E-01 |
| OLFM2 | 96,24 | -0,28 | 0,09 | -3,12 | 1,82E-03 | 1,56E-01 |
| HMGCS1 | 717,44 | 0,17 | 0,05 | 3,09 | 2,01E-03 | 1,57E-01 |
| SLC7A8 | 40,95 | -0,59 | 0,19 | -3,09 | 2,03E-03 | 1,57E-01 |
| ATXN3 | 1843,44 | 0,11 | 0,03 | 3,09 | 2,03E-03 | 1,57E-01 |
| SDC3 | 39,13 | -0,77 | 0,25 | -3,09 | 2,01E-03 | 1,57E-01 |
| HID1 | 36,87 | -0,66 | 0,21 | -3,08 | 2,05E-03 | 1,57E-01 |
| SLIT1 | 84,92 | -0,60 | 0,19 | -3,08 | 2,07E-03 | 1,58E-01 |
| SRGAP1 | 79,81 | -0,84 | 0,27 | -3,07 | 2,17E-03 | 1,63E-01 |
| KCNK12 | 36,66 | -0,61 | 0,20 | -3,07 | 2,17E-03 | 1,63E-01 |
| DNMT3B | 44,81 | -0,64 | 0,21 | -3,07 | 2,17E-03 | 1,63E-01 |
| RRP15 | 444,51 | 0,19 | 0,06 | 3,06 | 2,19E-03 | 1,63E-01 |
| HBB | 1905,85 | -0,75 | 0,25 | -3,05 | 2,32E-03 | 1,63E-01 |
| IPO5 | 1418,23 | 0,12 | 0,04 | 3,02 | 2,50E-03 | 1,63E-01 |
| TMEM19 | 830,43 | 0,17 | 0,05 | 3,05 | 2,29E-03 | 1,63E-01 |
| MAGI2 | 67,37 | -0,77 | 0,26 | -3,03 | 2,48E-03 | 1,63E-01 |
| DNAJC2 | 539,38 | 0,14 | 0,04 | 3,03 | 2,45E-03 | 1,63E-01 |
| IQCK | 35,04 | -0,57 | 0,19 | -3,03 | 2,49E-03 | 1,63E-01 |
| LGMN | 50,77 | -0,38 | 0,13 | -3,05 | 2,33E-03 | 1,63E-01 |
| SYPL1 | 721,15 | 0,19 | 0,06 | 3,06 | 2,22E-03 | 1,63E-01 |
| CABYR | 35,71 | -0,51 | 0,17 | -3,03 | 2,46E-03 | 1,63E-01 |
| ZNF37A | 1045,59 | 0,24 | 0,08 | 3,05 | 2,26E-03 | 1,63E-01 |
| SLC8A3 | 58,56 | -0,66 | 0,22 | -3,04 | 2,40E-03 | 1,63E-01 |
| MTIF2 | 640,35 | 0,15 | 0,05 | 3,04 | 2,36E-03 | 1,63E-01 |
| SNRPD3 | 920,14 | 0,10 | 0,03 | 3,04 | 2,40E-03 | 1,63E-01 |
| GFI1B | 298,12 | -0,34 | 0,11 | -3,05 | 2,27E-03 | 1,63E-01 |
| ZNF124 | 400,83 | 0,21 | 0,07 | 3,04 | 2,35E-03 | 1,63E-01 |
| FAXDC2 | 1427,58 | -0,44 | 0,14 | -3,05 | 2,28E-03 | 1,63E-01 |
| SORBS1 | 85,80 | -0,51 | 0,17 | -3,03 | 2,45E-03 | 1,63E-01 |

| | | | | | | |
|---|---|---|---|---|---|---|
| EPRS1 | 2090,95 | 0,17 | 0,06 | 3,06 | 2,25E-03 | 1,63E-01 |
| COL17A1 | 58,01 | -0,82 | 0,27 | -3,03 | 2,45E-03 | 1,63E-01 |
| SMTN | 69,68 | -0,63 | 0,21 | -3,02 | 2,50E-03 | 1,63E-01 |
| LSM12 | 199,70 | -0,12 | 0,04 | -3,03 | 2,46E-03 | 1,63E-01 |
| HLA-A | 39739,42 | -0,28 | 0,09 | -3,02 | 2,49E-03 | 1,63E-01 |
| COL7A1 | 99,05 | -0,60 | 0,20 | -3,03 | 2,41E-03 | 1,63E-01 |
| SEMA3G | 38,10 | -0,59 | 0,19 | -3,02 | 2,52E-03 | 1,64E-01 |
| LZTS3 | 35,13 | -0,71 | 0,24 | -3,02 | 2,55E-03 | 1,64E-01 |
| AOC1 | 24,95 | -0,72 | 0,24 | -3,02 | 2,55E-03 | 1,64E-01 |
| ADPRHL1 | 79,28 | -0,54 | 0,18 | -2,98 | 2,91E-03 | 1,65E-01 |
| EPS8L2 | 91,60 | -0,31 | 0,10 | -3,00 | 2,73E-03 | 1,65E-01 |
| CDH1 | 158,39 | -0,42 | 0,14 | -2,97 | 2,94E-03 | 1,65E-01 |
| KLHDC8A | 31,45 | -0,61 | 0,20 | -2,97 | 2,96E-03 | 1,65E-01 |
| LIMS2 | 63,99 | -0,50 | 0,17 | -2,98 | 2,88E-03 | 1,65E-01 |
| SMARCA5 | 2866,60 | 0,22 | 0,07 | 2,98 | 2,90E-03 | 1,65E-01 |
| TBC1D20 | 1278,17 | -0,11 | 0,04 | -2,97 | 2,93E-03 | 1,65E-01 |
| HELQ | 841,95 | 0,16 | 0,05 | 2,99 | 2,80E-03 | 1,65E-01 |
| ZNF28 | 549,37 | 0,23 | 0,08 | 2,97 | 2,97E-03 | 1,65E-01 |
| KLHL35 | 35,61 | -0,57 | 0,19 | -2,97 | 3,02E-03 | 1,65E-01 |
| ZNF468 | 638,60 | 0,21 | 0,07 | 2,98 | 2,87E-03 | 1,65E-01 |
| NHSL3 | 40,78 | -0,62 | 0,21 | -2,99 | 2,76E-03 | 1,65E-01 |
| IDH3A | 778,43 | 0,09 | 0,03 | 2,97 | 2,97E-03 | 1,65E-01 |
| NRXN3 | 102,59 | -0,62 | 0,21 | -2,98 | 2,87E-03 | 1,65E-01 |
| SYN3 | 45,54 | -0,78 | 0,26 | -3,01 | 2,64E-03 | 1,65E-01 |
| EVC | 73,43 | -0,59 | 0,20 | -2,98 | 2,84E-03 | 1,65E-01 |
| LSM10 | 419,54 | -0,14 | 0,05 | -3,00 | 2,69E-03 | 1,65E-01 |
| KCNH3 | 79,70 | -0,48 | 0,16 | -3,00 | 2,74E-03 | 1,65E-01 |
| TRPV3 | 68,21 | -0,63 | 0,21 | -3,01 | 2,62E-03 | 1,65E-01 |
| NRIP3 | 46,71 | -0,47 | 0,16 | -3,01 | 2,65E-03 | 1,65E-01 |
| ZNF134 | 804,83 | 0,14 | 0,05 | 2,97 | 2,99E-03 | 1,65E-01 |
| ZNF3 | 804,91 | 0,07 | 0,03 | 2,99 | 2,83E-03 | 1,65E-01 |
| PMP22 | 43,96 | -0,57 | 0,19 | -2,98 | 2,86E-03 | 1,65E-01 |
| MCOLN1 | 360,93 | -0,34 | 0,11 | -2,97 | 2,97E-03 | 1,65E-01 |
| RRP36 | 682,87 | 0,09 | 0,03 | 3,00 | 2,74E-03 | 1,65E-01 |
| EFCAB6 | 55,01 | -0,70 | 0,23 | -3,00 | 2,73E-03 | 1,65E-01 |
| LRFN2 | 35,72 | -0,64 | 0,21 | -2,97 | 3,01E-03 | 1,65E-01 |
| PLEKHA7 | 107,25 | -0,48 | 0,16 | -2,96 | 3,03E-03 | 1,65E-01 |
| PODN | 50,96 | -0,64 | 0,21 | -2,99 | 2,75E-03 | 1,65E-01 |
| STARD10 | 126,67 | -0,31 | 0,10 | -2,99 | 2,83E-03 | 1,65E-01 |
| CWF19L1 | 1263,34 | 0,09 | 0,03 | 2,97 | 2,98E-03 | 1,65E-01 |
| PTPN2 | 1996,96 | 0,13 | 0,04 | 2,99 | 2,74E-03 | 1,65E-01 |
| RAPGEF4 | 45,42 | -0,73 | 0,25 | -2,96 | 3,03E-03 | 1,65E-01 |
| ADGRV1 | 112,63 | -0,82 | 0,27 | -3,00 | 2,73E-03 | 1,65E-01 |
| PTPRF | 103,05 | -0,63 | 0,21 | -2,99 | 2,80E-03 | 1,65E-01 |
| PODXL | 42,45 | -0,55 | 0,19 | -2,86 | 4,21E-03 | 1,65E-01 |
| ZNF175 | 377,51 | 0,12 | 0,04 | 2,92 | 3,51E-03 | 1,65E-01 |

| | | | | | | |
|---|---|---|---|---|---|---|
| BAHCC1 | 91,62 | -0,65 | 0,22 | -2,89 | 3,81E-03 | 1,65E-01 |
| GNAO1 | 109,15 | -0,58 | 0,20 | -2,90 | 3,73E-03 | 1,65E-01 |
| RPS24 | 3209,78 | 0,32 | 0,11 | 2,91 | 3,58E-03 | 1,65E-01 |
| GPALPP1 | 872,26 | 0,12 | 0,04 | 2,94 | 3,32E-03 | 1,65E-01 |
| PDE4A | 102,48 | -0,41 | 0,14 | -2,86 | 4,24E-03 | 1,65E-01 |
| PDE2A | 71,11 | -0,56 | 0,19 | -2,87 | 4,15E-03 | 1,65E-01 |
| CSTF2T | 1313,67 | 0,15 | 0,05 | 2,88 | 4,00E-03 | 1,65E-01 |
| PTMA | 6828,21 | 0,16 | 0,05 | 2,92 | 3,53E-03 | 1,65E-01 |
| PHAX | 658,41 | 0,17 | 0,06 | 2,88 | 3,94E-03 | 1,65E-01 |
| TPRKB | 299,95 | 0,16 | 0,06 | 2,86 | 4,21E-03 | 1,65E-01 |
| CYSTM1 | 512,55 | -0,26 | 0,09 | -2,87 | 4,05E-03 | 1,65E-01 |
| WWTR1 | 53,10 | -0,46 | 0,16 | -2,95 | 3,14E-03 | 1,65E-01 |
| GRSF1 | 1682,28 | 0,11 | 0,04 | 2,91 | 3,66E-03 | 1,65E-01 |
| CLBA1 | 49,36 | -0,39 | 0,13 | -2,94 | 3,24E-03 | 1,65E-01 |
| ZNF75A | 1080,90 | 0,14 | 0,05 | 2,92 | 3,45E-03 | 1,65E-01 |
| DNAH12 | 69,24 | -0,71 | 0,25 | -2,86 | 4,23E-03 | 1,65E-01 |
| NOP58 | 1351,70 | 0,21 | 0,07 | 2,89 | 3,91E-03 | 1,65E-01 |
| ZNF326 | 1261,22 | 0,18 | 0,06 | 2,95 | 3,15E-03 | 1,65E-01 |
| MIGA1 | 1041,32 | 0,14 | 0,05 | 2,87 | 4,11E-03 | 1,65E-01 |
| ABCB6 | 22,84 | -0,30 | 0,11 | -2,86 | 4,24E-03 | 1,65E-01 |
| WDR43 | 1311,42 | 0,17 | 0,06 | 2,90 | 3,74E-03 | 1,65E-01 |
| CHDH | 52,31 | -0,68 | 0,23 | -2,95 | 3,14E-03 | 1,65E-01 |
| SDAD1 | 909,20 | 0,16 | 0,06 | 2,94 | 3,32E-03 | 1,65E-01 |
| OXTR | 42,43 | -0,57 | 0,20 | -2,90 | 3,74E-03 | 1,65E-01 |
| EXOSC2 | 886,64 | 0,10 | 0,03 | 2,90 | 3,68E-03 | 1,65E-01 |
| CDC14B | 362,34 | -0,21 | 0,07 | -2,93 | 3,42E-03 | 1,65E-01 |
| KIFC1 | 36,19 | -0,48 | 0,16 | -2,88 | 3,97E-03 | 1,65E-01 |
| KLHL29 | 76,52 | -0,62 | 0,22 | -2,86 | 4,21E-03 | 1,65E-01 |
| INTU | 59,52 | -0,69 | 0,24 | -2,91 | 3,60E-03 | 1,65E-01 |
| NUCKS1 | 3953,67 | 0,17 | 0,06 | 2,89 | 3,91E-03 | 1,65E-01 |
| UHRF1 | 75,12 | -0,42 | 0,14 | -2,95 | 3,23E-03 | 1,65E-01 |
| HMG20A | 1051,69 | 0,10 | 0,03 | 2,93 | 3,43E-03 | 1,65E-01 |
| SCARF2 | 34,94 | -0,59 | 0,20 | -2,92 | 3,54E-03 | 1,65E-01 |
| SLC2A1 | 2123,27 | -0,29 | 0,10 | -2,88 | 4,03E-03 | 1,65E-01 |
| IRF8 | 2148,25 | 0,17 | 0,06 | 2,90 | 3,72E-03 | 1,65E-01 |
| FRMD4A | 188,35 | -0,50 | 0,17 | -2,90 | 3,71E-03 | 1,65E-01 |
| ENOX2 | 431,83 | 0,10 | 0,03 | 2,88 | 4,00E-03 | 1,65E-01 |
| UTP14A | 360,10 | 0,13 | 0,04 | 2,95 | 3,19E-03 | 1,65E-01 |
| PDZD2 | 82,15 | -0,67 | 0,23 | -2,96 | 3,11E-03 | 1,65E-01 |
| STAP1 | 257,38 | 0,31 | 0,11 | 2,94 | 3,25E-03 | 1,65E-01 |
| BBLN | 288,59 | -0,16 | 0,06 | -2,87 | 4,07E-03 | 1,65E-01 |
| TATDN1 | 281,14 | 0,15 | 0,05 | 2,90 | 3,70E-03 | 1,65E-01 |
| CCDC78 | 30,40 | -0,48 | 0,16 | -2,92 | 3,55E-03 | 1,65E-01 |
| FAM133B | 688,51 | 0,20 | 0,07 | 2,90 | 3,75E-03 | 1,65E-01 |
| MDK | 27,63 | -0,42 | 0,15 | -2,86 | 4,22E-03 | 1,65E-01 |
| ST18 | 58,31 | -0,74 | 0,26 | -2,89 | 3,89E-03 | 1,65E-01 |

| | | | | | | |
|---|---|---|---|---|---|---|
| ZNF462 | 58,41 | -0,74 | 0,25 | -2,92 | 3,47E-03 | 1,65E-01 |
| TMEM40 | 73,67 | -0,37 | 0,13 | -2,89 | 3,86E-03 | 1,65E-01 |
| CD83 | 294,32 | 0,22 | 0,08 | 2,92 | 3,50E-03 | 1,65E-01 |
| WDR70 | 1097,14 | 0,13 | 0,05 | 2,86 | 4,25E-03 | 1,65E-01 |
| NAV2 | 87,49 | -0,71 | 0,25 | -2,88 | 3,96E-03 | 1,65E-01 |
| RTL5 | 29,69 | -0,42 | 0,14 | -2,94 | 3,31E-03 | 1,65E-01 |
| PIF1 | 41,64 | -0,43 | 0,15 | -2,91 | 3,56E-03 | 1,65E-01 |
| MGAT3 | 111,70 | -0,35 | 0,12 | -2,89 | 3,82E-03 | 1,65E-01 |
| RSL1D1 | 1592,52 | 0,16 | 0,06 | 2,93 | 3,36E-03 | 1,65E-01 |
| TMEM178B | 42,89 | -0,76 | 0,26 | -2,93 | 3,37E-03 | 1,65E-01 |
| RIPOR3 | 113,79 | -0,41 | 0,14 | -2,91 | 3,57E-03 | 1,65E-01 |
| GNG5 | 304,75 | -0,14 | 0,05 | -2,88 | 3,93E-03 | 1,65E-01 |
| DBNDD2 | 51,26 | -0,31 | 0,11 | -2,88 | 3,98E-03 | 1,65E-01 |
| WDR62 | 60,46 | -0,53 | 0,19 | -2,88 | 3,92E-03 | 1,65E-01 |
| NOC3L | 723,76 | 0,23 | 0,08 | 2,91 | 3,60E-03 | 1,65E-01 |
| ETV5 | 37,42 | -0,62 | 0,22 | -2,88 | 3,92E-03 | 1,65E-01 |
| HDAC9 | 1621,69 | 0,23 | 0,08 | 2,93 | 3,42E-03 | 1,65E-01 |
| OSBPL6 | 48,34 | -0,59 | 0,21 | -2,86 | 4,26E-03 | 1,65E-01 |
| H2AX | 162,39 | -0,27 | 0,09 | -2,89 | 3,84E-03 | 1,65E-01 |
| ADCY6 | 46,06 | -0,72 | 0,25 | -2,96 | 3,12E-03 | 1,65E-01 |
| RPL34 | 2057,29 | 0,53 | 0,18 | 2,96 | 3,11E-03 | 1,65E-01 |
| EFCAB12 | 63,36 | -0,40 | 0,14 | -2,92 | 3,51E-03 | 1,65E-01 |
| UMODL1 | 39,18 | -0,60 | 0,21 | -2,88 | 4,01E-03 | 1,65E-01 |
| SCN1B | 69,76 | -0,44 | 0,15 | -2,87 | 4,09E-03 | 1,65E-01 |
| CDCA3 | 27,56 | -0,44 | 0,15 | -2,86 | 4,19E-03 | 1,65E-01 |
| ACRBP | 246,75 | -0,29 | 0,10 | -2,93 | 3,42E-03 | 1,65E-01 |
| PLLP | 34,16 | -0,57 | 0,20 | -2,87 | 4,10E-03 | 1,65E-01 |
| ABHD4 | 584,59 | -0,18 | 0,06 | -2,90 | 3,77E-03 | 1,65E-01 |
| EIF4B | 13923,10 | 0,16 | 0,06 | 2,87 | 4,16E-03 | 1,65E-01 |
| NUP42 | 897,89 | 0,12 | 0,04 | 2,95 | 3,18E-03 | 1,65E-01 |
| RBFOX2 | 84,31 | -0,59 | 0,21 | -2,89 | 3,91E-03 | 1,65E-01 |
| C3orf33 | 31,43 | -0,27 | 0,09 | -2,94 | 3,30E-03 | 1,65E-01 |
| POLR1F | 352,54 | 0,25 | 0,09 | 2,95 | 3,21E-03 | 1,65E-01 |
| HPS5 | 1024,02 | 0,10 | 0,03 | 2,91 | 3,60E-03 | 1,65E-01 |
| GTPBP4 | 787,86 | 0,10 | 0,03 | 2,86 | 4,24E-03 | 1,65E-01 |
| MFSD6L | 71,68 | -0,37 | 0,13 | -2,91 | 3,63E-03 | 1,65E-01 |
| ZBTB5 | 500,66 | 0,12 | 0,04 | 2,87 | 4,07E-03 | 1,65E-01 |
| B4GALNT3 | 180,08 | -0,51 | 0,17 | -2,94 | 3,28E-03 | 1,65E-01 |
| GFUS | 844,06 | -0,35 | 0,12 | -2,95 | 3,19E-03 | 1,65E-01 |
| MRPS35 | 527,62 | 0,17 | 0,06 | 2,93 | 3,44E-03 | 1,65E-01 |
| TANC1 | 62,38 | -0,65 | 0,23 | -2,88 | 3,94E-03 | 1,65E-01 |
| INTS10 | 1590,78 | 0,09 | 0,03 | 2,89 | 3,87E-03 | 1,65E-01 |
| EMID1 | 64,81 | -0,49 | 0,17 | -2,90 | 3,74E-03 | 1,65E-01 |
| NBL1 | 60,96 | -0,44 | 0,15 | -2,85 | 4,32E-03 | 1,66E-01 |
| FTL | 45405,84 | -0,25 | 0,09 | -2,85 | 4,31E-03 | 1,66E-01 |
| ZNF385D | 42,28 | -0,57 | 0,20 | -2,86 | 4,30E-03 | 1,66E-01 |

| | | | | | | |
|---|---|---|---|---|---|---|
| CAV2 | 64,50 | -0,54 | 0,19 | -2,85 | 4,31E-03 | 1,66E-01 |
| WDR90 | 122,92 | -0,45 | 0,16 | -2,84 | 4,49E-03 | 1,66E-01 |
| SLC25A38 | 810,64 | -0,13 | 0,05 | -2,85 | 4,39E-03 | 1,66E-01 |
| TMEM88 | 44,23 | -0,40 | 0,14 | -2,84 | 4,56E-03 | 1,66E-01 |
| NUP107 | 1140,49 | 0,16 | 0,06 | 2,85 | 4,37E-03 | 1,66E-01 |
| ALPK2 | 54,78 | -0,66 | 0,23 | -2,84 | 4,51E-03 | 1,66E-01 |
| RNF152 | 62,61 | -0,55 | 0,19 | -2,85 | 4,37E-03 | 1,66E-01 |
| ZMYND10 | 48,71 | -0,28 | 0,10 | -2,84 | 4,45E-03 | 1,66E-01 |
| SCAND1 | 461,24 | -0,12 | 0,04 | -2,84 | 4,50E-03 | 1,66E-01 |
| ZNF525 | 516,99 | 0,31 | 0,11 | 2,84 | 4,52E-03 | 1,66E-01 |
| EXOSC9 | 674,78 | 0,17 | 0,06 | 2,84 | 4,55E-03 | 1,66E-01 |
| GBA1 | 263,29 | -0,22 | 0,08 | -2,84 | 4,46E-03 | 1,66E-01 |
| TRIP13 | 33,90 | -0,54 | 0,19 | -2,84 | 4,56E-03 | 1,66E-01 |
| CFAP251 | 77,51 | -0,48 | 0,17 | -2,84 | 4,53E-03 | 1,66E-01 |
| ZNF737 | 782,44 | 0,23 | 0,08 | 2,84 | 4,48E-03 | 1,66E-01 |
| WASF3 | 55,13 | -0,50 | 0,18 | -2,84 | 4,46E-03 | 1,66E-01 |
| SLC16A13 | 48,68 | -0,33 | 0,11 | -2,85 | 4,36E-03 | 1,66E-01 |
| ALKBH8 | 536,65 | 0,17 | 0,06 | 2,85 | 4,43E-03 | 1,66E-01 |
| SRXN1 | 599,48 | -0,32 | 0,12 | -2,78 | 5,51E-03 | 1,66E-01 |
| MAK16 | 527,90 | 0,16 | 0,06 | 2,75 | 6,01E-03 | 1,66E-01 |
| ZNG1A | 828,48 | 0,18 | 0,07 | 2,74 | 6,06E-03 | 1,66E-01 |
| UBAP1 | 1460,04 | -0,20 | 0,07 | -2,76 | 5,71E-03 | 1,66E-01 |
| RIOK1 | 500,38 | 0,12 | 0,04 | 2,75 | 5,88E-03 | 1,66E-01 |
| ARHGEF28 | 102,12 | -0,55 | 0,20 | -2,80 | 5,07E-03 | 1,66E-01 |
| TNXB | 132,29 | -0,48 | 0,18 | -2,72 | 6,43E-03 | 1,66E-01 |
| CUX2 | 54,39 | -0,64 | 0,23 | -2,82 | 4,76E-03 | 1,66E-01 |
| TRMT61B | 311,79 | 0,12 | 0,04 | 2,76 | 5,81E-03 | 1,66E-01 |
| NLRP6 | 1035,13 | -0,27 | 0,10 | -2,74 | 6,12E-03 | 1,66E-01 |
| SLC6A13 | 36,33 | -0,51 | 0,19 | -2,71 | 6,65E-03 | 1,66E-01 |
| MRPS25 | 1615,33 | 0,10 | 0,04 | 2,77 | 5,57E-03 | 1,66E-01 |
| SLC2A14 | 48,47 | -0,56 | 0,20 | -2,80 | 5,14E-03 | 1,66E-01 |
| SLC12A3 | 36,33 | -0,59 | 0,22 | -2,75 | 5,87E-03 | 1,66E-01 |
| CHML | 999,77 | 0,30 | 0,11 | 2,81 | 4,97E-03 | 1,66E-01 |
| ITGB1BP2 | 46,51 | -0,23 | 0,08 | -2,78 | 5,38E-03 | 1,66E-01 |
| ZNF485 | 140,77 | 0,16 | 0,06 | 2,73 | 6,40E-03 | 1,66E-01 |
| ZNRF1 | 154,21 | -0,28 | 0,10 | -2,79 | 5,33E-03 | 1,66E-01 |
| HTRA2 | 721,63 | -0,09 | 0,03 | -2,72 | 6,53E-03 | 1,66E-01 |
| EIF2AK3 | 1107,03 | 0,12 | 0,04 | 2,78 | 5,48E-03 | 1,66E-01 |
| COMMD6 | 295,87 | 0,46 | 0,17 | 2,72 | 6,53E-03 | 1,66E-01 |
| ELF3 | 41,99 | -0,45 | 0,17 | -2,73 | 6,39E-03 | 1,66E-01 |
| FBXO24 | 40,17 | -0,36 | 0,13 | -2,76 | 5,71E-03 | 1,66E-01 |
| EEA1 | 1198,65 | 0,20 | 0,07 | 2,73 | 6,28E-03 | 1,66E-01 |
| ARHGEF37 | 133,31 | -0,39 | 0,14 | -2,78 | 5,37E-03 | 1,66E-01 |
| RSPH9 | 24,12 | -0,48 | 0,17 | -2,76 | 5,73E-03 | 1,66E-01 |
| ZNF253 | 416,40 | 0,21 | 0,08 | 2,72 | 6,62E-03 | 1,66E-01 |
| GAREM2 | 45,04 | -0,43 | 0,16 | -2,73 | 6,25E-03 | 1,66E-01 |

| | | | | | | |
|---|---|---|---|---|---|---|
| SPNS2 | 55,56 | -0,42 | 0,15 | -2,81 | 4,99E-03 | 1,66E-01 |
| NFIB | 49,27 | -0,63 | 0,23 | -2,74 | 6,18E-03 | 1,66E-01 |
| ZNF528 | 783,27 | 0,17 | 0,06 | 2,81 | 4,90E-03 | 1,66E-01 |
| TCERG1 | 2124,10 | 0,15 | 0,05 | 2,82 | 4,80E-03 | 1,66E-01 |
| HEPACAM2 | 63,71 | -0,39 | 0,14 | -2,82 | 4,79E-03 | 1,66E-01 |
| USP6 | 56,15 | -0,66 | 0,24 | -2,79 | 5,21E-03 | 1,66E-01 |
| SRFBP1 | 371,71 | 0,17 | 0,06 | 2,76 | 5,78E-03 | 1,66E-01 |
| AJAP1 | 93,65 | -0,59 | 0,21 | -2,76 | 5,70E-03 | 1,66E-01 |
| SCNN1D | 62,68 | -0,38 | 0,14 | -2,74 | 6,07E-03 | 1,66E-01 |
| SPTBN5 | 157,46 | -0,46 | 0,17 | -2,71 | 6,67E-03 | 1,66E-01 |
| BIRC3 | 5969,80 | 0,28 | 0,10 | 2,76 | 5,72E-03 | 1,66E-01 |
| SDCBP2 | 34,20 | -0,37 | 0,13 | -2,76 | 5,81E-03 | 1,66E-01 |
| CBX8 | 137,90 | -0,22 | 0,08 | -2,79 | 5,25E-03 | 1,66E-01 |
| ZNF766 | 700,25 | 0,12 | 0,04 | 2,76 | 5,84E-03 | 1,66E-01 |
| EAF2 | 409,87 | 0,23 | 0,08 | 2,77 | 5,63E-03 | 1,66E-01 |
| CCDC88A | 2891,99 | 0,21 | 0,08 | 2,72 | 6,57E-03 | 1,66E-01 |
| EVC2 | 72,97 | -0,48 | 0,17 | -2,79 | 5,29E-03 | 1,66E-01 |
| RASL11A | 66,72 | -0,25 | 0,09 | -2,72 | 6,50E-03 | 1,66E-01 |
| IPO7 | 1903,30 | 0,16 | 0,06 | 2,80 | 5,06E-03 | 1,66E-01 |
| PPIL4 | 1085,23 | 0,18 | 0,06 | 2,83 | 4,66E-03 | 1,66E-01 |
| B3GNT7 | 36,26 | -0,46 | 0,17 | -2,72 | 6,51E-03 | 1,66E-01 |
| PTK6 | 42,82 | -0,42 | 0,15 | -2,80 | 5,11E-03 | 1,66E-01 |
| KATNA1 | 367,83 | 0,10 | 0,04 | 2,73 | 6,27E-03 | 1,66E-01 |
| STK32B | 48,06 | -0,64 | 0,23 | -2,78 | 5,49E-03 | 1,66E-01 |
| ABLIM2 | 79,76 | -0,44 | 0,16 | -2,78 | 5,36E-03 | 1,66E-01 |
| CIT | 130,11 | -0,49 | 0,17 | -2,81 | 5,02E-03 | 1,66E-01 |
| DNM1 | 57,99 | -0,60 | 0,22 | -2,74 | 6,12E-03 | 1,66E-01 |
| HSPA9 | 2594,26 | 0,12 | 0,05 | 2,77 | 5,66E-03 | 1,66E-01 |
| FANCM | 542,24 | 0,20 | 0,07 | 2,80 | 5,08E-03 | 1,66E-01 |
| DCLRE1C | 1241,32 | 0,09 | 0,03 | 2,77 | 5,68E-03 | 1,66E-01 |
| HSD17B1 | 33,70 | -0,41 | 0,15 | -2,73 | 6,39E-03 | 1,66E-01 |
| POLG2 | 637,23 | 0,10 | 0,04 | 2,75 | 5,96E-03 | 1,66E-01 |
| ELOF1 | 594,69 | -0,29 | 0,11 | -2,72 | 6,47E-03 | 1,66E-01 |
| GAS6 | 123,23 | -0,35 | 0,13 | -2,74 | 6,08E-03 | 1,66E-01 |
| NRIP1 | 1648,31 | 0,22 | 0,08 | 2,82 | 4,75E-03 | 1,66E-01 |
| IMPACT | 513,91 | 0,15 | 0,06 | 2,73 | 6,35E-03 | 1,66E-01 |
| CAND1 | 3260,09 | 0,16 | 0,06 | 2,81 | 5,01E-03 | 1,66E-01 |
| SPC24 | 26,40 | -0,39 | 0,14 | -2,75 | 5,89E-03 | 1,66E-01 |
| ZNF25 | 531,05 | 0,17 | 0,06 | 2,79 | 5,32E-03 | 1,66E-01 |
| CDHR1 | 55,72 | -0,50 | 0,18 | -2,80 | 5,09E-03 | 1,66E-01 |
| AFDN | 770,93 | -0,27 | 0,10 | -2,71 | 6,66E-03 | 1,66E-01 |
| TOMM70 | 1213,96 | 0,17 | 0,06 | 2,83 | 4,59E-03 | 1,66E-01 |
| SNAPC3 | 2158,68 | 0,13 | 0,05 | 2,76 | 5,85E-03 | 1,66E-01 |
| RNF212 | 44,86 | -0,46 | 0,17 | -2,72 | 6,46E-03 | 1,66E-01 |
| TRPM4 | 51,64 | -0,61 | 0,22 | -2,79 | 5,19E-03 | 1,66E-01 |
| EPS8L1 | 62,99 | -0,56 | 0,20 | -2,79 | 5,22E-03 | 1,66E-01 |

| | | | | | | |
|---|---|---|---|---|---|---|
| KCNK5 | 34,60 | -0,50 | 0,18 | -2,77 | 5,64E-03 | 1,66E-01 |
| FAM200A | 294,56 | 0,18 | 0,06 | 2,73 | 6,38E-03 | 1,66E-01 |
| TGM2 | 180,65 | -0,43 | 0,15 | -2,81 | 4,96E-03 | 1,66E-01 |
| YPEL3 | 2438,65 | -0,32 | 0,11 | -2,76 | 5,74E-03 | 1,66E-01 |
| EEF1E1 | 117,01 | 0,23 | 0,08 | 2,74 | 6,12E-03 | 1,66E-01 |
| MYO10 | 101,11 | -0,52 | 0,19 | -2,72 | 6,56E-03 | 1,66E-01 |
| C3 | 52,65 | -0,54 | 0,20 | -2,77 | 5,64E-03 | 1,66E-01 |
| ZNF627 | 345,59 | 0,18 | 0,06 | 2,72 | 6,59E-03 | 1,66E-01 |
| CRYBG3 | 1184,25 | 0,18 | 0,06 | 2,83 | 4,71E-03 | 1,66E-01 |
| ZNF146 | 1152,10 | 0,22 | 0,08 | 2,78 | 5,47E-03 | 1,66E-01 |
| PCYT1B | 48,91 | -0,46 | 0,17 | -2,73 | 6,36E-03 | 1,66E-01 |
| PHOSPHO2 | 93,71 | 0,21 | 0,08 | 2,75 | 5,93E-03 | 1,66E-01 |
| FAM177B | 111,54 | 0,26 | 0,10 | 2,79 | 5,30E-03 | 1,66E-01 |
| GNL2 | 690,18 | 0,10 | 0,04 | 2,80 | 5,17E-03 | 1,66E-01 |
| BMP8B | 54,64 | -0,38 | 0,13 | -2,78 | 5,39E-03 | 1,66E-01 |
| ACE | 58,02 | -0,50 | 0,18 | -2,74 | 6,20E-03 | 1,66E-01 |
| MMUT | 520,92 | 0,18 | 0,07 | 2,75 | 5,97E-03 | 1,66E-01 |
| COL5A2 | 50,74 | -0,46 | 0,17 | -2,76 | 5,79E-03 | 1,66E-01 |
| ZNRF3 | 53,44 | -0,46 | 0,17 | -2,75 | 5,90E-03 | 1,66E-01 |
| CEP83 | 541,62 | 0,15 | 0,05 | 2,79 | 5,21E-03 | 1,66E-01 |
| ARF1 | 3861,03 | -0,18 | 0,07 | -2,74 | 6,23E-03 | 1,66E-01 |
| ZNF703 | 54,95 | -0,45 | 0,17 | -2,72 | 6,52E-03 | 1,66E-01 |
| SUPT7L | 1182,44 | 0,13 | 0,05 | 2,73 | 6,30E-03 | 1,66E-01 |
| USP16 | 1675,81 | 0,19 | 0,07 | 2,77 | 5,61E-03 | 1,66E-01 |
| SGK3 | 1106,29 | 0,12 | 0,04 | 2,83 | 4,63E-03 | 1,66E-01 |
| LAMB3 | 40,79 | -0,58 | 0,21 | -2,80 | 5,12E-03 | 1,66E-01 |
| LYG1 | 43,35 | -0,20 | 0,07 | -2,73 | 6,35E-03 | 1,66E-01 |
| IGF2BP3 | 113,00 | -0,36 | 0,13 | -2,75 | 5,93E-03 | 1,66E-01 |
| FAM53A | 42,79 | -0,46 | 0,17 | -2,77 | 5,61E-03 | 1,66E-01 |
| SLC43A1 | 222,49 | -0,20 | 0,07 | -2,83 | 4,70E-03 | 1,66E-01 |
| PARG | 1166,27 | 0,12 | 0,04 | 2,77 | 5,68E-03 | 1,66E-01 |
| DNAJC15 | 688,27 | 0,24 | 0,09 | 2,75 | 5,97E-03 | 1,66E-01 |
| IBTK | 1862,21 | 0,16 | 0,06 | 2,81 | 5,01E-03 | 1,66E-01 |
| IL4I1 | 29,68 | -0,42 | 0,15 | -2,73 | 6,38E-03 | 1,66E-01 |
| CRACD | 52,51 | -0,65 | 0,23 | -2,80 | 5,04E-03 | 1,66E-01 |
| DNAH14 | 80,38 | -0,72 | 0,26 | -2,82 | 4,86E-03 | 1,66E-01 |
| GPT2 | 56,82 | -0,54 | 0,19 | -2,79 | 5,20E-03 | 1,66E-01 |
| PRKAB2 | 1024,18 | 0,11 | 0,04 | 2,80 | 5,05E-03 | 1,66E-01 |
| C1R | 36,28 | -0,46 | 0,16 | -2,82 | 4,83E-03 | 1,66E-01 |
| ZNF860 | 190,56 | 0,35 | 0,13 | 2,71 | 6,67E-03 | 1,66E-01 |
| SERINC2 | 48,98 | -0,62 | 0,23 | -2,76 | 5,86E-03 | 1,66E-01 |
| NT5DC2 | 38,71 | -0,50 | 0,18 | -2,72 | 6,55E-03 | 1,66E-01 |
| NFE2L3 | 280,42 | 0,20 | 0,07 | 2,81 | 5,00E-03 | 1,66E-01 |
| PRRT3 | 61,85 | -0,37 | 0,13 | -2,76 | 5,72E-03 | 1,66E-01 |
| DNAH6 | 152,86 | -0,43 | 0,16 | -2,78 | 5,42E-03 | 1,66E-01 |
| MYO16 | 89,72 | -0,62 | 0,22 | -2,79 | 5,20E-03 | 1,66E-01 |

| | | | | | | |
|---|---|---|---|---|---|---|
| FBXO2 | 40,01 | -0,48 | 0,18 | -2,71 | 6,68E-03 | 1,66E-01 |
| KIF18B | 29,18 | -0,55 | 0,20 | -2,72 | 6,53E-03 | 1,66E-01 |
| ZNF808 | 1004,74 | 0,14 | 0,05 | 2,75 | 6,01E-03 | 1,66E-01 |
| DTHD1 | 809,35 | 0,31 | 0,11 | 2,72 | 6,44E-03 | 1,66E-01 |
| BRIX1 | 351,98 | 0,21 | 0,07 | 2,78 | 5,45E-03 | 1,66E-01 |
| PXDC1 | 46,03 | -0,39 | 0,14 | -2,72 | 6,55E-03 | 1,66E-01 |
| BCCIP | 588,27 | 0,14 | 0,05 | 2,82 | 4,82E-03 | 1,66E-01 |
| NUP133 | 1398,32 | 0,08 | 0,03 | 2,74 | 6,19E-03 | 1,66E-01 |
| MFSD13A | 111,51 | -0,27 | 0,10 | -2,77 | 5,58E-03 | 1,66E-01 |
| GPX1 | 7606,08 | -0,34 | 0,12 | -2,82 | 4,82E-03 | 1,66E-01 |
| SMAD6 | 62,76 | -0,64 | 0,23 | -2,83 | 4,67E-03 | 1,66E-01 |
| L3MBTL3 | 1112,37 | 0,14 | 0,05 | 2,81 | 4,96E-03 | 1,66E-01 |
| SLC1A7 | 60,90 | -0,53 | 0,19 | -2,79 | 5,31E-03 | 1,66E-01 |
| MYLK3 | 42,25 | -0,62 | 0,22 | -2,78 | 5,38E-03 | 1,66E-01 |
| ZSCAN12 | 395,66 | 0,21 | 0,08 | 2,81 | 4,89E-03 | 1,66E-01 |
| KIFC3 | 226,37 | -0,32 | 0,11 | -2,81 | 4,96E-03 | 1,66E-01 |
| BTLA | 1019,71 | 0,30 | 0,11 | 2,82 | 4,85E-03 | 1,66E-01 |
| TECTA | 71,73 | -0,47 | 0,17 | -2,78 | 5,47E-03 | 1,66E-01 |
| MTDH | 2820,39 | 0,13 | 0,05 | 2,73 | 6,34E-03 | 1,66E-01 |
| NEDD4L | 382,53 | -0,29 | 0,10 | -2,82 | 4,74E-03 | 1,66E-01 |
| DECR2 | 43,93 | -0,38 | 0,14 | -2,73 | 6,30E-03 | 1,66E-01 |
| ITGB4 | 54,03 | -0,62 | 0,23 | -2,73 | 6,28E-03 | 1,66E-01 |
| ZNF675 | 584,21 | 0,18 | 0,07 | 2,77 | 5,53E-03 | 1,66E-01 |
| MEIS2 | 48,62 | -0,70 | 0,25 | -2,81 | 4,98E-03 | 1,66E-01 |
| MTCL1 | 102,69 | -0,63 | 0,23 | -2,76 | 5,81E-03 | 1,66E-01 |
| POP5 | 340,33 | 0,13 | 0,05 | 2,74 | 6,08E-03 | 1,66E-01 |
| FGF13 | 63,95 | -0,59 | 0,21 | -2,76 | 5,84E-03 | 1,66E-01 |
| RNF170 | 650,17 | 0,12 | 0,04 | 2,81 | 4,98E-03 | 1,66E-01 |
| EPHB2 | 78,70 | -0,63 | 0,23 | -2,81 | 4,98E-03 | 1,66E-01 |
| RPH3A | 58,85 | -0,67 | 0,25 | -2,71 | 6,63E-03 | 1,66E-01 |
| RUFY3 | 1449,83 | 0,11 | 0,04 | 2,78 | 5,44E-03 | 1,66E-01 |
| SFPQ | 9782,44 | 0,12 | 0,04 | 2,71 | 6,68E-03 | 1,66E-01 |
| PPM1K | 3299,20 | 0,15 | 0,06 | 2,71 | 6,64E-03 | 1,66E-01 |
| MARCHF1 | 3825,02 | 0,24 | 0,09 | 2,73 | 6,36E-03 | 1,66E-01 |
| CENPM | 29,68 | -0,38 | 0,14 | -2,80 | 5,08E-03 | 1,66E-01 |
| ZNF347 | 503,29 | 0,18 | 0,06 | 2,75 | 5,91E-03 | 1,66E-01 |
| CPSF3 | 684,31 | 0,11 | 0,04 | 2,72 | 6,57E-03 | 1,66E-01 |
| NTRK1 | 41,18 | -0,68 | 0,25 | -2,71 | 6,70E-03 | 1,66E-01 |
| ICE2 | 1941,66 | 0,15 | 0,05 | 2,71 | 6,72E-03 | 1,67E-01 |
| FBXO4 | 271,02 | 0,19 | 0,07 | 2,70 | 6,84E-03 | 1,67E-01 |
| SRPRB | 822,04 | 0,13 | 0,05 | 2,71 | 6,78E-03 | 1,67E-01 |
| DCLK2 | 49,25 | -0,59 | 0,22 | -2,70 | 6,86E-03 | 1,67E-01 |
| SLC39A4 | 57,94 | -0,35 | 0,13 | -2,71 | 6,77E-03 | 1,67E-01 |
| NUAK1 | 66,88 | -0,56 | 0,21 | -2,71 | 6,79E-03 | 1,67E-01 |
| TGS1 | 916,21 | 0,10 | 0,04 | 2,70 | 6,86E-03 | 1,67E-01 |
| CABP5 | 48,00 | -0,41 | 0,15 | -2,70 | 6,85E-03 | 1,67E-01 |

| | | | | | | |
|---|---|---|---|---|---|---|
| CFAP119 | 113,80 | -0,18 | 0,07 | -2,70 | 6,85E-03 | 1,67E-01 |
| C3orf20 | 57,56 | -0,37 | 0,14 | -2,71 | 6,82E-03 | 1,67E-01 |
| TFR2 | 105,68 | -0,37 | 0,14 | -2,70 | 6,88E-03 | 1,67E-01 |
| SOX8 | 47,98 | -0,52 | 0,19 | -2,70 | 6,92E-03 | 1,67E-01 |
| ACSBG1 | 67,44 | -0,51 | 0,19 | -2,70 | 6,96E-03 | 1,67E-01 |
| METAP2 | 763,00 | 0,19 | 0,07 | 2,70 | 6,94E-03 | 1,67E-01 |
| CAMK2D | 2119,85 | 0,17 | 0,06 | 2,70 | 7,00E-03 | 1,67E-01 |
| UMPS | 635,84 | 0,10 | 0,04 | 2,70 | 6,98E-03 | 1,67E-01 |
| COL19A1 | 685,09 | 0,32 | 0,12 | 2,70 | 6,99E-03 | 1,67E-01 |
| ZNF426 | 770,41 | 0,15 | 0,05 | 2,70 | 6,92E-03 | 1,67E-01 |
| ZNF555 | 403,87 | 0,16 | 0,06 | 2,70 | 7,01E-03 | 1,67E-01 |
| CDHR2 | 120,33 | -0,29 | 0,11 | -2,70 | 6,98E-03 | 1,67E-01 |
| WDR31 | 37,11 | -0,44 | 0,17 | -2,69 | 7,11E-03 | 1,68E-01 |
| SLC22A17 | 39,79 | -0,42 | 0,16 | -2,69 | 7,11E-03 | 1,68E-01 |
| PRDX2 | 1149,38 | -0,31 | 0,12 | -2,69 | 7,06E-03 | 1,68E-01 |
| ZZZ3 | 1829,49 | 0,18 | 0,07 | 2,69 | 7,08E-03 | 1,68E-01 |
| ZFAND2B | 505,59 | -0,11 | 0,04 | -2,69 | 7,07E-03 | 1,68E-01 |
| BIRC5 | 33,92 | -0,40 | 0,15 | -2,69 | 7,11E-03 | 1,68E-01 |
| WDR45 | 2536,10 | -0,22 | 0,08 | -2,69 | 7,13E-03 | 1,68E-01 |
| PROSER2 | 39,66 | -0,44 | 0,16 | -2,69 | 7,15E-03 | 1,68E-01 |
| SMC6 | 1396,52 | 0,19 | 0,07 | 2,69 | 7,18E-03 | 1,68E-01 |
| MPHOSPH10 | 621,40 | 0,15 | 0,06 | 2,69 | 7,19E-03 | 1,68E-01 |
| TAF1A | 297,62 | 0,16 | 0,06 | 2,69 | 7,22E-03 | 1,69E-01 |
| KLHDC7B | 57,47 | -0,38 | 0,14 | -2,69 | 7,22E-03 | 1,69E-01 |
| ABCA3 | 107,26 | -0,39 | 0,15 | -2,68 | 7,30E-03 | 1,70E-01 |
| FRMD6 | 57,50 | -0,56 | 0,21 | -2,68 | 7,32E-03 | 1,70E-01 |
| AGPAT5 | 741,23 | 0,20 | 0,07 | 2,68 | 7,33E-03 | 1,70E-01 |
| RYR3 | 148,16 | -0,74 | 0,28 | -2,68 | 7,31E-03 | 1,70E-01 |
| ARG2 | 47,66 | -0,40 | 0,15 | -2,68 | 7,36E-03 | 1,70E-01 |
| ZNF131 | 1279,07 | 0,11 | 0,04 | 2,68 | 7,36E-03 | 1,70E-01 |
| MANEAL | 39,77 | -0,38 | 0,14 | -2,68 | 7,37E-03 | 1,70E-01 |
| RPAIN | 1426,68 | 0,11 | 0,04 | 2,68 | 7,44E-03 | 1,70E-01 |
| LANCL3 | 144,51 | -0,22 | 0,08 | -2,68 | 7,45E-03 | 1,70E-01 |
| IGSF22 | 42,90 | -0,51 | 0,19 | -2,68 | 7,44E-03 | 1,70E-01 |
| BAG5 | 1426,28 | 0,12 | 0,05 | 2,67 | 7,49E-03 | 1,70E-01 |
| ZNF714 | 535,91 | 0,15 | 0,06 | 2,67 | 7,48E-03 | 1,70E-01 |
| SRSF10 | 3912,96 | 0,19 | 0,07 | 2,68 | 7,47E-03 | 1,70E-01 |
| CMC1 | 953,00 | 0,25 | 0,09 | 2,68 | 7,42E-03 | 1,70E-01 |
| DNAJC10 | 2629,53 | 0,19 | 0,07 | 2,68 | 7,40E-03 | 1,70E-01 |
| TYW3 | 503,11 | 0,21 | 0,08 | 2,67 | 7,53E-03 | 1,70E-01 |
| SET | 5206,85 | 0,17 | 0,06 | 2,67 | 7,54E-03 | 1,70E-01 |
| PHLDA3 | 27,93 | -0,46 | 0,17 | -2,67 | 7,54E-03 | 1,70E-01 |
| PPOX | 411,01 | -0,12 | 0,04 | -2,67 | 7,58E-03 | 1,71E-01 |
| SERPINE2 | 56,04 | -0,49 | 0,18 | -2,67 | 7,65E-03 | 1,71E-01 |
| CUL2 | 695,04 | 0,13 | 0,05 | 2,67 | 7,65E-03 | 1,71E-01 |
| NCBP2 | 1296,15 | 0,13 | 0,05 | 2,67 | 7,65E-03 | 1,71E-01 |

| | | | | | | |
|---|---|---|---|---|---|---|
| ZNF503 | 37,94 | -0,53 | 0,20 | -2,67 | 7,63E-03 | 1,71E-01 |
| NECAB2 | 49,04 | -0,41 | 0,15 | -2,67 | 7,64E-03 | 1,71E-01 |
| ZNF33B | 1428,49 | 0,17 | 0,06 | 2,66 | 7,86E-03 | 1,71E-01 |
| DPF3 | 109,74 | -0,38 | 0,14 | -2,66 | 7,78E-03 | 1,71E-01 |
| CXCL8 | 598,85 | 0,44 | 0,16 | 2,66 | 7,74E-03 | 1,71E-01 |
| ZNF681 | 248,65 | 0,28 | 0,10 | 2,66 | 7,77E-03 | 1,71E-01 |
| ZFP64 | 236,26 | -0,19 | 0,07 | -2,66 | 7,88E-03 | 1,71E-01 |
| ZNF283 | 516,69 | 0,15 | 0,05 | 2,66 | 7,86E-03 | 1,71E-01 |
| LARP7 | 636,17 | 0,22 | 0,08 | 2,66 | 7,79E-03 | 1,71E-01 |
| FAM3C | 722,99 | 0,24 | 0,09 | 2,66 | 7,84E-03 | 1,71E-01 |
| PIWIL2 | 52,35 | -0,49 | 0,18 | -2,67 | 7,70E-03 | 1,71E-01 |
| PPIL3 | 330,25 | 0,19 | 0,07 | 2,66 | 7,87E-03 | 1,71E-01 |
| SH3BP5 | 2889,39 | 0,12 | 0,05 | 2,66 | 7,72E-03 | 1,71E-01 |
| NOS3 | 56,01 | -0,47 | 0,18 | -2,66 | 7,85E-03 | 1,71E-01 |
| CSKMT | 135,09 | 0,15 | 0,06 | 2,66 | 7,77E-03 | 1,71E-01 |
| SHLD2 | 750,07 | 0,22 | 0,08 | 2,66 | 7,73E-03 | 1,71E-01 |
| PRTFDC1 | 36,59 | -0,32 | 0,12 | -2,66 | 7,70E-03 | 1,71E-01 |
| TRPV1 | 72,25 | -0,33 | 0,12 | -2,66 | 7,89E-03 | 1,71E-01 |
| QRSL1 | 679,79 | 0,15 | 0,06 | 2,65 | 8,00E-03 | 1,71E-01 |
| DNAI2 | 40,08 | -0,40 | 0,15 | -2,65 | 7,99E-03 | 1,71E-01 |
| NPHP1 | 63,00 | -0,56 | 0,21 | -2,66 | 7,93E-03 | 1,71E-01 |
| EXOSC8 | 554,86 | 0,17 | 0,06 | 2,65 | 7,98E-03 | 1,71E-01 |
| SLC7A5 | 223,90 | -0,41 | 0,15 | -2,65 | 7,95E-03 | 1,71E-01 |
| E2F7 | 37,59 | -0,49 | 0,19 | -2,66 | 7,93E-03 | 1,71E-01 |
| KAT14 | 499,85 | 0,11 | 0,04 | 2,65 | 8,03E-03 | 1,71E-01 |
| ACTR6 | 494,45 | 0,25 | 0,09 | 2,65 | 8,03E-03 | 1,71E-01 |
| SMC3 | 1455,79 | 0,20 | 0,07 | 2,65 | 8,03E-03 | 1,71E-01 |
| UTP15 | 520,99 | 0,16 | 0,06 | 2,65 | 7,97E-03 | 1,71E-01 |
| LRFN3 | 36,90 | -0,52 | 0,20 | -2,65 | 8,05E-03 | 1,71E-01 |
| METTL14 | 1034,64 | 0,16 | 0,06 | 2,65 | 8,15E-03 | 1,72E-01 |
| RABGGTB | 1711,51 | 0,17 | 0,07 | 2,65 | 8,13E-03 | 1,72E-01 |
| ZNF557 | 1231,23 | 0,13 | 0,05 | 2,65 | 8,12E-03 | 1,72E-01 |
| SKIC8 | 811,72 | 0,09 | 0,03 | 2,65 | 8,10E-03 | 1,72E-01 |
| FAM220A | 306,39 | 0,11 | 0,04 | 2,65 | 8,12E-03 | 1,72E-01 |
| MYL5 | 90,67 | -0,26 | 0,10 | -2,65 | 8,15E-03 | 1,72E-01 |
| GTF3C3 | 1144,69 | 0,15 | 0,06 | 2,65 | 8,08E-03 | 1,72E-01 |
| CRHBP | 26,17 | -0,39 | 0,15 | -2,64 | 8,22E-03 | 1,72E-01 |
| CTSK | 151,56 | -0,17 | 0,07 | -2,64 | 8,21E-03 | 1,72E-01 |
| RANBP6 | 989,64 | 0,29 | 0,11 | 2,64 | 8,22E-03 | 1,72E-01 |
| ZNF439 | 698,46 | 0,16 | 0,06 | 2,64 | 8,22E-03 | 1,72E-01 |
| ZNF736 | 1986,67 | 0,15 | 0,06 | 2,64 | 8,27E-03 | 1,72E-01 |
| TFB2M | 202,40 | 0,15 | 0,06 | 2,64 | 8,26E-03 | 1,72E-01 |
| VEZT | 1226,17 | 0,13 | 0,05 | 2,64 | 8,26E-03 | 1,72E-01 |
| ABLIM3 | 93,35 | -0,49 | 0,18 | -2,64 | 8,29E-03 | 1,72E-01 |
| CYP2E1 | 51,30 | -0,62 | 0,24 | -2,64 | 8,32E-03 | 1,72E-01 |
| NAA35 | 835,12 | 0,11 | 0,04 | 2,64 | 8,32E-03 | 1,72E-01 |

| | | | | | | |
|---|---|---|---|---|---|---|
| CCDC43 | 437,41 | 0,16 | 0,06 | 2,64 | 8,37E-03 | 1,73E-01 |
| PNO1 | 295,65 | 0,11 | 0,04 | 2,64 | 8,38E-03 | 1,73E-01 |
| KIF2C | 36,94 | -0,45 | 0,17 | -2,64 | 8,41E-03 | 1,73E-01 |
| TG | 320,82 | -0,29 | 0,11 | -2,64 | 8,41E-03 | 1,73E-01 |
| FHAD1 | 64,15 | -0,72 | 0,27 | -2,63 | 8,42E-03 | 1,73E-01 |
| KRBA2 | 126,31 | 0,15 | 0,06 | 2,63 | 8,63E-03 | 1,74E-01 |
| LRRC61 | 120,72 | -0,19 | 0,07 | -2,63 | 8,60E-03 | 1,74E-01 |
| ZNG1B | 1360,72 | 0,17 | 0,06 | 2,63 | 8,63E-03 | 1,74E-01 |
| NDUFAF2 | 104,11 | 0,16 | 0,06 | 2,63 | 8,53E-03 | 1,74E-01 |
| AJUBA | 40,24 | -0,38 | 0,15 | -2,63 | 8,64E-03 | 1,74E-01 |
| POLK | 1746,43 | 0,19 | 0,07 | 2,63 | 8,51E-03 | 1,74E-01 |
| GPATCH4 | 331,82 | 0,12 | 0,05 | 2,63 | 8,59E-03 | 1,74E-01 |
| LSM7 | 516,47 | 0,19 | 0,07 | 2,63 | 8,61E-03 | 1,74E-01 |
| RPS6 | 11856,97 | 0,29 | 0,11 | 2,63 | 8,64E-03 | 1,74E-01 |
| FAM209A | 26,32 | -0,26 | 0,10 | -2,63 | 8,59E-03 | 1,74E-01 |
| CEBPZ | 963,52 | 0,25 | 0,10 | 2,63 | 8,57E-03 | 1,74E-01 |
| MAP3K1 | 11519,85 | 0,12 | 0,05 | 2,63 | 8,62E-03 | 1,74E-01 |
| GJC2 | 47,91 | -0,29 | 0,11 | -2,62 | 8,67E-03 | 1,74E-01 |
| CCDC120 | 33,09 | -0,46 | 0,18 | -2,62 | 8,77E-03 | 1,74E-01 |
| TTC27 | 633,51 | 0,13 | 0,05 | 2,62 | 8,75E-03 | 1,74E-01 |
| MS4A4E | 418,46 | 0,21 | 0,08 | 2,62 | 8,75E-03 | 1,74E-01 |
| ZNF649 | 579,63 | 0,15 | 0,06 | 2,62 | 8,72E-03 | 1,74E-01 |
| SSB | 747,30 | 0,22 | 0,09 | 2,62 | 8,70E-03 | 1,74E-01 |
| SSR3 | 2628,33 | 0,16 | 0,06 | 2,62 | 8,79E-03 | 1,74E-01 |
| MYO7A | 116,95 | -0,56 | 0,22 | -2,62 | 8,71E-03 | 1,74E-01 |
| PLAU | 27,98 | -0,40 | 0,15 | -2,62 | 8,79E-03 | 1,74E-01 |
| TLN2 | 171,13 | -0,51 | 0,20 | -2,60 | 9,37E-03 | 1,75E-01 |
| UFL1 | 1448,80 | 0,22 | 0,09 | 2,59 | 9,55E-03 | 1,75E-01 |
| NAA15 | 1785,44 | 0,16 | 0,06 | 2,59 | 9,49E-03 | 1,75E-01 |
| ATXN10 | 859,53 | 0,11 | 0,04 | 2,60 | 9,35E-03 | 1,75E-01 |
| PUS7L | 1841,53 | 0,21 | 0,08 | 2,61 | 9,03E-03 | 1,75E-01 |
| ENSG000002 | 26,38 | -0,46 | 0,18 | -2,59 | 9,72E-03 | 1,75E-01 |
| AIMP1 | 486,71 | 0,22 | 0,09 | 2,58 | 9,92E-03 | 1,75E-01 |
| ZC3H8 | 519,60 | 0,12 | 0,05 | 2,57 | 1,02E-02 | 1,75E-01 |
| CELSR2 | 92,97 | -0,40 | 0,16 | -2,57 | 1,03E-02 | 1,75E-01 |
| ABRAXAS1 | 822,52 | 0,17 | 0,06 | 2,60 | 9,41E-03 | 1,75E-01 |
| DCAF13 | 794,61 | 0,18 | 0,07 | 2,61 | 9,02E-03 | 1,75E-01 |
| ZNF182 | 339,85 | 0,10 | 0,04 | 2,56 | 1,04E-02 | 1,75E-01 |
| ZUP1 | 443,04 | 0,19 | 0,07 | 2,59 | 9,69E-03 | 1,75E-01 |
| ROM1 | 31,23 | -0,33 | 0,13 | -2,60 | 9,40E-03 | 1,75E-01 |
| PRPS2 | 261,44 | 0,14 | 0,05 | 2,61 | 9,17E-03 | 1,75E-01 |
| GSE1 | 1614,16 | -0,20 | 0,08 | -2,57 | 1,02E-02 | 1,75E-01 |
| PIGA | 567,29 | 0,15 | 0,06 | 2,58 | 9,86E-03 | 1,75E-01 |
| MARVELD2 | 31,00 | -0,38 | 0,15 | -2,56 | 1,04E-02 | 1,75E-01 |
| CC2D2A | 53,63 | -0,48 | 0,19 | -2,58 | 9,85E-03 | 1,75E-01 |
| SH2D1A | 903,81 | 0,28 | 0,11 | 2,57 | 1,02E-02 | 1,75E-01 |

| | | | | | | |
|---|---|---|---|---|---|---|
| MCM10 | 48,27 | -0,50 | 0,19 | -2,60 | 9,41E-03 | 1,75E-01 |
| ZNF888 | 271,95 | 0,20 | 0,08 | 2,60 | 9,39E-03 | 1,75E-01 |
| PLEKHG5 | 97,82 | -0,48 | 0,18 | -2,59 | 9,63E-03 | 1,75E-01 |
| MFSD2B | 283,02 | -0,35 | 0,13 | -2,61 | 9,02E-03 | 1,75E-01 |
| TUBB4B | 1439,70 | -0,16 | 0,06 | -2,57 | 1,03E-02 | 1,75E-01 |
| PLEKHD1 | 43,67 | -0,64 | 0,25 | -2,60 | 9,45E-03 | 1,75E-01 |
| BCL2L12 | 59,48 | -0,18 | 0,07 | -2,58 | 1,00E-02 | 1,75E-01 |
| ZRSR2 | 666,59 | 0,10 | 0,04 | 2,59 | 9,72E-03 | 1,75E-01 |
| DCTN5 | 1977,73 | 0,11 | 0,04 | 2,56 | 1,04E-02 | 1,75E-01 |
| INIP | 907,63 | 0,09 | 0,04 | 2,56 | 1,05E-02 | 1,75E-01 |
| SYP | 37,32 | -0,35 | 0,13 | -2,61 | 9,18E-03 | 1,75E-01 |
| ZSCAN9 | 373,95 | 0,13 | 0,05 | 2,59 | 9,73E-03 | 1,75E-01 |
| MSANTD4 | 758,18 | 0,19 | 0,07 | 2,56 | 1,04E-02 | 1,75E-01 |
| TARBP1 | 1540,37 | 0,11 | 0,04 | 2,57 | 1,01E-02 | 1,75E-01 |
| YEATS4 | 222,59 | 0,23 | 0,09 | 2,58 | 9,87E-03 | 1,75E-01 |
| ACER2 | 99,68 | -0,20 | 0,08 | -2,61 | 9,02E-03 | 1,75E-01 |
| UBE3A | 3332,00 | 0,15 | 0,06 | 2,59 | 9,70E-03 | 1,75E-01 |
| GINS2 | 34,49 | -0,36 | 0,14 | -2,59 | 9,56E-03 | 1,75E-01 |
| SSPN | 88,68 | -0,31 | 0,12 | -2,56 | 1,05E-02 | 1,75E-01 |
| OCLN | 39,04 | -0,56 | 0,22 | -2,60 | 9,27E-03 | 1,75E-01 |
| PNLDC1 | 39,56 | -0,31 | 0,12 | -2,58 | 9,87E-03 | 1,75E-01 |
| SHARPIN | 604,37 | -0,25 | 0,10 | -2,60 | 9,38E-03 | 1,75E-01 |
| ATL2 | 1151,93 | 0,16 | 0,06 | 2,60 | 9,27E-03 | 1,75E-01 |
| AQR | 3217,32 | 0,12 | 0,05 | 2,59 | 9,56E-03 | 1,75E-01 |
| MCUR1 | 408,22 | -0,11 | 0,04 | -2,60 | 9,21E-03 | 1,75E-01 |
| WDR75 | 1342,11 | 0,16 | 0,06 | 2,56 | 1,04E-02 | 1,75E-01 |
| ZNF180 | 579,38 | 0,16 | 0,06 | 2,62 | 8,87E-03 | 1,75E-01 |
| HYAL3 | 49,46 | -0,25 | 0,10 | -2,57 | 1,00E-02 | 1,75E-01 |
| AFAP1L2 | 78,74 | -0,33 | 0,13 | -2,57 | 1,02E-02 | 1,75E-01 |
| POLR2F | 481,50 | -0,08 | 0,03 | -2,57 | 1,03E-02 | 1,75E-01 |
| TOP2B | 5356,41 | 0,17 | 0,07 | 2,58 | 9,85E-03 | 1,75E-01 |
| ACOXL | 57,22 | -0,48 | 0,19 | -2,56 | 1,03E-02 | 1,75E-01 |
| ZNF189 | 809,43 | 0,18 | 0,07 | 2,60 | 9,41E-03 | 1,75E-01 |
| TRPS1 | 2463,05 | 0,12 | 0,05 | 2,62 | 8,91E-03 | 1,75E-01 |
| NUMBL | 79,56 | -0,36 | 0,14 | -2,56 | 1,04E-02 | 1,75E-01 |
| CNTNAP3B | 42,60 | -0,60 | 0,23 | -2,62 | 8,88E-03 | 1,75E-01 |
| NMB | 33,44 | -0,22 | 0,08 | -2,57 | 1,01E-02 | 1,75E-01 |
| RAB30 | 757,21 | 0,19 | 0,07 | 2,56 | 1,04E-02 | 1,75E-01 |
| RALGPS2 | 3104,47 | 0,25 | 0,10 | 2,57 | 1,01E-02 | 1,75E-01 |
| P2RY10 | 1050,16 | 0,18 | 0,07 | 2,58 | 9,78E-03 | 1,75E-01 |
| KCNG1 | 64,93 | -0,56 | 0,21 | -2,61 | 8,94E-03 | 1,75E-01 |
| PAK3 | 46,22 | -0,44 | 0,17 | -2,59 | 9,52E-03 | 1,75E-01 |
| PKD1L1 | 90,76 | -0,45 | 0,17 | -2,61 | 9,00E-03 | 1,75E-01 |
| SGIP1 | 58,87 | -0,54 | 0,21 | -2,56 | 1,04E-02 | 1,75E-01 |
| PIK3C2A | 2336,66 | 0,13 | 0,05 | 2,57 | 1,02E-02 | 1,75E-01 |
| OCIAD1 | 1518,96 | 0,11 | 0,04 | 2,56 | 1,04E-02 | 1,75E-01 |

| | | | | | | |
|---|---|---|---|---|---|---|
| NEMF | 2595,70 | 0,12 | 0,05 | 2,56 | 1,04E-02 | 1,75E-01 |
| ABCE1 | 1145,92 | 0,21 | 0,08 | 2,58 | 9,74E-03 | 1,75E-01 |
| KIAA1671 | 168,81 | -0,47 | 0,18 | -2,58 | 9,96E-03 | 1,75E-01 |
| KPTN | 67,00 | -0,27 | 0,10 | -2,62 | 8,89E-03 | 1,75E-01 |
| TADA3 | 890,80 | -0,16 | 0,06 | -2,59 | 9,64E-03 | 1,75E-01 |
| EIF5B | 1002,17 | 0,11 | 0,04 | 2,61 | 9,13E-03 | 1,75E-01 |
| HMCN1 | 72,36 | -0,61 | 0,24 | -2,58 | 1,00E-02 | 1,75E-01 |
| ZNF92 | 382,32 | 0,32 | 0,12 | 2,58 | 9,99E-03 | 1,75E-01 |
| UTP23 | 780,34 | 0,17 | 0,06 | 2,61 | 9,12E-03 | 1,75E-01 |
| NVL | 716,48 | 0,09 | 0,04 | 2,58 | 9,93E-03 | 1,75E-01 |
| ZNF579 | 36,74 | -0,47 | 0,18 | -2,58 | 9,86E-03 | 1,75E-01 |
| ARHGAP6 | 130,26 | -0,31 | 0,12 | -2,56 | 1,04E-02 | 1,75E-01 |
| ANKRD33B | 338,88 | -0,24 | 0,09 | -2,60 | 9,30E-03 | 1,75E-01 |
| HTATSF1 | 769,91 | 0,17 | 0,06 | 2,60 | 9,31E-03 | 1,75E-01 |
| EPDR1 | 38,36 | -0,30 | 0,11 | -2,59 | 9,50E-03 | 1,75E-01 |
| UTP18 | 901,46 | 0,12 | 0,05 | 2,58 | 9,99E-03 | 1,75E-01 |
| OSGIN1 | 44,29 | -0,34 | 0,13 | -2,57 | 1,02E-02 | 1,75E-01 |
| DBT | 810,49 | 0,08 | 0,03 | 2,60 | 9,19E-03 | 1,75E-01 |
| TTLL7 | 83,94 | -0,41 | 0,16 | -2,59 | 9,69E-03 | 1,75E-01 |
| ZNF35 | 222,05 | 0,15 | 0,06 | 2,58 | 9,81E-03 | 1,75E-01 |
| TERF1 | 907,01 | 0,10 | 0,04 | 2,60 | 9,25E-03 | 1,75E-01 |
| ZNF101 | 2718,44 | 0,11 | 0,04 | 2,58 | 9,92E-03 | 1,75E-01 |
| ODAD4 | 57,02 | -0,38 | 0,15 | -2,56 | 1,04E-02 | 1,75E-01 |
| RAB15 | 44,17 | -0,41 | 0,16 | -2,58 | 9,84E-03 | 1,75E-01 |
| SMARCAD1 | 1180,25 | 0,20 | 0,08 | 2,59 | 9,56E-03 | 1,75E-01 |
| PLEKHF2 | 1583,78 | 0,22 | 0,08 | 2,59 | 9,69E-03 | 1,75E-01 |
| KLLN | 76,92 | -0,21 | 0,08 | -2,58 | 9,97E-03 | 1,75E-01 |
| UBA2 | 1569,89 | 0,15 | 0,06 | 2,56 | 1,04E-02 | 1,75E-01 |
| RPL23 | 3134,32 | 0,44 | 0,17 | 2,57 | 1,03E-02 | 1,75E-01 |
| MRPS27 | 796,75 | 0,09 | 0,03 | 2,61 | 9,15E-03 | 1,75E-01 |
| RIBC1 | 24,13 | -0,30 | 0,12 | -2,61 | 9,00E-03 | 1,75E-01 |
| CCDC50 | 1305,17 | 0,19 | 0,07 | 2,57 | 1,03E-02 | 1,75E-01 |
| MOK | 79,66 | -0,46 | 0,18 | -2,59 | 9,70E-03 | 1,75E-01 |
| SBDS | 437,94 | 0,19 | 0,07 | 2,58 | 9,82E-03 | 1,75E-01 |
| SKIC3 | 2859,05 | 0,18 | 0,07 | 2,59 | 9,71E-03 | 1,75E-01 |
| ZNF219 | 73,24 | -0,38 | 0,15 | -2,59 | 9,59E-03 | 1,75E-01 |
| SLC48A1 | 384,35 | -0,34 | 0,13 | -2,60 | 9,38E-03 | 1,75E-01 |
| DIPK1B | 45,20 | -0,38 | 0,15 | -2,56 | 1,05E-02 | 1,75E-01 |
| ANK2 | 55,04 | -0,77 | 0,29 | -2,61 | 8,96E-03 | 1,75E-01 |
| TFAM | 1449,14 | 0,19 | 0,07 | 2,60 | 9,39E-03 | 1,75E-01 |
| PCDH1 | 89,24 | -0,36 | 0,14 | -2,57 | 1,01E-02 | 1,75E-01 |
| MYH10 | 73,98 | -0,49 | 0,19 | -2,60 | 9,20E-03 | 1,75E-01 |
| TYW5 | 776,97 | 0,12 | 0,05 | 2,57 | 1,02E-02 | 1,75E-01 |
| PLAAT5 | 45,78 | -0,42 | 0,16 | -2,58 | 1,00E-02 | 1,75E-01 |
| HSP90B1 | 2107,58 | 0,13 | 0,05 | 2,57 | 1,03E-02 | 1,75E-01 |
| LRRC24 | 34,45 | -0,32 | 0,12 | -2,60 | 9,45E-03 | 1,75E-01 |

| | | | | | | |
|---|---|---|---|---|---|---|
| TRIP6 | 123,77 | -0,20 | 0,08 | -2,58 | 9,87E-03 | 1,75E-01 |
| SLC24A3 | 132,52 | -0,35 | 0,14 | -2,61 | 9,02E-03 | 1,75E-01 |
| HES6 | 65,80 | -0,31 | 0,12 | -2,60 | 9,21E-03 | 1,75E-01 |
| PTPRO | 487,85 | 0,21 | 0,08 | 2,57 | 1,01E-02 | 1,75E-01 |
| FAH | 234,62 | -0,19 | 0,07 | -2,61 | 8,98E-03 | 1,75E-01 |
| CANX | 7369,93 | 0,12 | 0,04 | 2,61 | 9,07E-03 | 1,75E-01 |
| MYCBPAP | 56,01 | -0,50 | 0,19 | -2,56 | 1,05E-02 | 1,75E-01 |
| MAX | 3507,33 | -0,11 | 0,04 | -2,56 | 1,05E-02 | 1,75E-01 |
| WSB2 | 956,32 | -0,11 | 0,04 | -2,54 | 1,11E-02 | 1,75E-01 |
| EEIG2 | 1050,25 | 0,13 | 0,05 | 2,55 | 1,08E-02 | 1,75E-01 |
| DARS1 | 1281,29 | 0,16 | 0,06 | 2,54 | 1,10E-02 | 1,75E-01 |
| CCDC154 | 60,97 | -0,47 | 0,19 | -2,55 | 1,07E-02 | 1,75E-01 |
| GEMIN6 | 272,49 | 0,14 | 0,05 | 2,54 | 1,10E-02 | 1,75E-01 |
| OTX1 | 46,98 | -0,51 | 0,20 | -2,55 | 1,08E-02 | 1,75E-01 |
| LTV1 | 212,49 | 0,15 | 0,06 | 2,54 | 1,11E-02 | 1,75E-01 |
| SMC5 | 2259,69 | 0,18 | 0,07 | 2,54 | 1,12E-02 | 1,75E-01 |
| PC | 85,68 | -0,38 | 0,15 | -2,54 | 1,11E-02 | 1,75E-01 |
| SLC39A6 | 660,13 | 0,12 | 0,05 | 2,54 | 1,12E-02 | 1,75E-01 |
| MS4A1 | 2153,43 | 0,29 | 0,11 | 2,54 | 1,09E-02 | 1,75E-01 |
| PRR3 | 351,09 | 0,09 | 0,04 | 2,54 | 1,12E-02 | 1,75E-01 |
| RPL7 | 6889,55 | 0,53 | 0,21 | 2,54 | 1,10E-02 | 1,75E-01 |
| ARL1 | 740,30 | 0,15 | 0,06 | 2,55 | 1,07E-02 | 1,75E-01 |
| MRPL32 | 480,02 | 0,24 | 0,10 | 2,55 | 1,09E-02 | 1,75E-01 |
| ACOT7 | 62,52 | -0,44 | 0,17 | -2,55 | 1,09E-02 | 1,75E-01 |
| IQCB1 | 1020,46 | 0,17 | 0,07 | 2,54 | 1,12E-02 | 1,75E-01 |
| AHRR | 54,11 | -0,64 | 0,25 | -2,54 | 1,11E-02 | 1,75E-01 |
| SMIM15 | 697,29 | 0,20 | 0,08 | 2,55 | 1,08E-02 | 1,75E-01 |
| KIF3C | 168,74 | -0,25 | 0,10 | -2,54 | 1,11E-02 | 1,75E-01 |
| BSG | 15650,79 | -0,34 | 0,13 | -2,55 | 1,07E-02 | 1,75E-01 |
| SUPT16H | 1900,94 | 0,10 | 0,04 | 2,55 | 1,09E-02 | 1,75E-01 |
| TUBA8 | 50,02 | -0,29 | 0,11 | -2,54 | 1,10E-02 | 1,75E-01 |
| TMC4 | 43,32 | -0,39 | 0,15 | -2,56 | 1,06E-02 | 1,75E-01 |
| RARS1 | 1238,44 | 0,17 | 0,07 | 2,54 | 1,10E-02 | 1,75E-01 |
| MTAP | 1160,03 | 0,11 | 0,04 | 2,55 | 1,07E-02 | 1,75E-01 |
| DNM1L | 1550,98 | 0,12 | 0,05 | 2,54 | 1,10E-02 | 1,75E-01 |
| DENR | 1085,48 | 0,12 | 0,05 | 2,56 | 1,06E-02 | 1,75E-01 |
| MTREX | 2193,35 | 0,18 | 0,07 | 2,55 | 1,07E-02 | 1,75E-01 |
| EVA1C | 201,69 | -0,21 | 0,08 | -2,55 | 1,09E-02 | 1,75E-01 |
| ZNF45 | 693,58 | 0,10 | 0,04 | 2,54 | 1,10E-02 | 1,75E-01 |
| RPL6 | 5189,40 | 0,27 | 0,11 | 2,55 | 1,09E-02 | 1,75E-01 |
| TAF11 | 668,53 | 0,09 | 0,04 | 2,54 | 1,10E-02 | 1,75E-01 |
| GRB10 | 369,98 | -0,29 | 0,11 | -2,54 | 1,09E-02 | 1,75E-01 |
| COX7B | 424,33 | 0,24 | 0,09 | 2,54 | 1,11E-02 | 1,75E-01 |
| CATIP | 27,55 | -0,33 | 0,13 | -2,55 | 1,07E-02 | 1,75E-01 |
| DDR1 | 74,41 | -0,46 | 0,18 | -2,54 | 1,12E-02 | 1,75E-01 |
| PLD5P1 | 161,08 | 0,17 | 0,07 | 2,55 | 1,08E-02 | 1,75E-01 |

| | | | | | | |
|---|---|---|---|---|---|---|
| HBQ1 | 64,29 | -0,41 | 0,16 | -2,54 | 1,10E-02 | 1,75E-01 |
| GPATCH2 | 923,74 | 0,16 | 0,06 | 2,54 | 1,10E-02 | 1,75E-01 |
| PLOD2 | 52,93 | -0,43 | 0,17 | -2,54 | 1,11E-02 | 1,75E-01 |
| MTCH2 | 693,43 | 0,09 | 0,03 | 2,56 | 1,06E-02 | 1,75E-01 |
| OSTC | 697,27 | 0,22 | 0,09 | 2,55 | 1,09E-02 | 1,75E-01 |
| AMN | 60,34 | -0,45 | 0,18 | -2,55 | 1,07E-02 | 1,75E-01 |
| TPM1 | 892,80 | -0,20 | 0,08 | -2,54 | 1,12E-02 | 1,75E-01 |
| ARMC5 | 101,68 | -0,26 | 0,10 | -2,54 | 1,12E-02 | 1,75E-01 |
| RPL31 | 3212,25 | 0,47 | 0,19 | 2,54 | 1,12E-02 | 1,75E-01 |
| COL18A1 | 1570,55 | -0,30 | 0,12 | -2,54 | 1,12E-02 | 1,75E-01 |
| RPL7L1 | 2154,03 | 0,10 | 0,04 | 2,53 | 1,13E-02 | 1,76E-01 |
| PBRM1 | 3297,14 | 0,09 | 0,04 | 2,53 | 1,13E-02 | 1,76E-01 |
| KATNAL2 | 43,22 | -0,48 | 0,19 | -2,53 | 1,13E-02 | 1,76E-01 |
| H2AJ | 908,46 | -0,24 | 0,09 | -2,53 | 1,14E-02 | 1,76E-01 |
| EFNA5 | 41,27 | -0,43 | 0,17 | -2,53 | 1,14E-02 | 1,76E-01 |
| GOLGA4 | 3552,52 | 0,17 | 0,07 | 2,53 | 1,14E-02 | 1,76E-01 |
| NPTXR | 40,03 | -0,51 | 0,20 | -2,53 | 1,14E-02 | 1,76E-01 |
| AP3M1 | 1366,97 | 0,10 | 0,04 | 2,53 | 1,14E-02 | 1,76E-01 |
| MTX3 | 908,87 | 0,15 | 0,06 | 2,53 | 1,14E-02 | 1,76E-01 |
| SORBS2 | 66,97 | -0,70 | 0,28 | -2,53 | 1,14E-02 | 1,76E-01 |
| MAMDC4 | 51,86 | -0,46 | 0,18 | -2,53 | 1,15E-02 | 1,76E-01 |
| GATA1 | 593,91 | -0,34 | 0,13 | -2,53 | 1,15E-02 | 1,77E-01 |
| FAM20A | 37,17 | -0,52 | 0,21 | -2,53 | 1,15E-02 | 1,77E-01 |
| CMPK1 | 2382,40 | 0,20 | 0,08 | 2,53 | 1,15E-02 | 1,77E-01 |
| PHF6 | 1040,99 | 0,19 | 0,08 | 2,52 | 1,16E-02 | 1,77E-01 |
| SNX2 | 2685,33 | 0,18 | 0,07 | 2,52 | 1,16E-02 | 1,77E-01 |
| LYRM2 | 657,49 | 0,21 | 0,08 | 2,52 | 1,17E-02 | 1,79E-01 |
| LIPE | 83,61 | -0,29 | 0,11 | -2,52 | 1,17E-02 | 1,79E-01 |
| NPM1 | 4555,54 | 0,22 | 0,09 | 2,52 | 1,18E-02 | 1,79E-01 |
| RSRC2 | 3166,83 | 0,13 | 0,05 | 2,52 | 1,17E-02 | 1,79E-01 |
| USP8 | 3365,39 | 0,09 | 0,03 | 2,52 | 1,18E-02 | 1,79E-01 |
| ZNF845 | 584,98 | 0,28 | 0,11 | 2,52 | 1,18E-02 | 1,80E-01 |
| WDR36 | 1669,84 | 0,17 | 0,07 | 2,52 | 1,19E-02 | 1,80E-01 |
| METTL6 | 453,18 | 0,07 | 0,03 | 2,51 | 1,19E-02 | 1,80E-01 |
| COA5 | 1093,49 | 0,13 | 0,05 | 2,51 | 1,19E-02 | 1,80E-01 |
| SMPD3 | 278,15 | -0,41 | 0,16 | -2,52 | 1,19E-02 | 1,80E-01 |
| ZNF26 | 1425,03 | 0,09 | 0,03 | 2,51 | 1,19E-02 | 1,80E-01 |
| PPP1R13L | 37,91 | -0,51 | 0,20 | -2,51 | 1,20E-02 | 1,80E-01 |
| CHRM3 | 40,11 | -0,54 | 0,21 | -2,51 | 1,20E-02 | 1,80E-01 |
| WDR3 | 866,87 | 0,11 | 0,05 | 2,51 | 1,20E-02 | 1,80E-01 |
| SNRPD1 | 400,43 | 0,15 | 0,06 | 2,51 | 1,20E-02 | 1,80E-01 |
| DAP | 636,40 | -0,22 | 0,09 | -2,51 | 1,20E-02 | 1,80E-01 |
| FHL2 | 58,53 | -0,41 | 0,16 | -2,51 | 1,20E-02 | 1,80E-01 |
| DIS3 | 2189,39 | 0,15 | 0,06 | 2,51 | 1,20E-02 | 1,80E-01 |
| BMP1 | 93,47 | -0,34 | 0,14 | -2,51 | 1,20E-02 | 1,80E-01 |
| TREX2 | 27,04 | -0,32 | 0,13 | -2,51 | 1,21E-02 | 1,81E-01 |

| | | | | | | |
|---|---|---|---|---|---|---|
| AASDHPPT | 1008,04 | 0,17 | 0,07 | 2,50 | 1,23E-02 | 1,81E-01 |
| ARMC3 | 53,21 | -0,43 | 0,17 | -2,51 | 1,22E-02 | 1,81E-01 |
| SUCLA2 | 673,44 | 0,11 | 0,04 | 2,50 | 1,23E-02 | 1,81E-01 |
| HNRNPA0 | 1409,07 | 0,12 | 0,05 | 2,50 | 1,23E-02 | 1,81E-01 |
| KBTBD8 | 367,10 | 0,31 | 0,13 | 2,51 | 1,22E-02 | 1,81E-01 |
| SUCLG2 | 1476,49 | 0,12 | 0,05 | 2,50 | 1,23E-02 | 1,81E-01 |
| MYEOV | 38,85 | -0,47 | 0,19 | -2,50 | 1,23E-02 | 1,81E-01 |
| TDRD12 | 46,71 | -0,67 | 0,27 | -2,50 | 1,23E-02 | 1,81E-01 |
| SH3GL1 | 523,47 | -0,22 | 0,09 | -2,51 | 1,22E-02 | 1,81E-01 |
| ALDH3A2 | 1078,70 | 0,08 | 0,03 | 2,50 | 1,23E-02 | 1,81E-01 |
| ENAH | 50,97 | -0,61 | 0,25 | -2,50 | 1,23E-02 | 1,82E-01 |
| MAP2K7 | 957,35 | -0,21 | 0,08 | -2,50 | 1,24E-02 | 1,82E-01 |
| CCT6A | 1713,13 | 0,13 | 0,05 | 2,50 | 1,23E-02 | 1,82E-01 |
| TFPI | 115,15 | -0,26 | 0,10 | -2,50 | 1,24E-02 | 1,82E-01 |
| TMPRSS3 | 52,62 | -0,40 | 0,16 | -2,50 | 1,24E-02 | 1,82E-01 |
| HNRNPA3 | 4651,88 | 0,11 | 0,04 | 2,50 | 1,24E-02 | 1,82E-01 |
| KANK3 | 56,46 | -0,35 | 0,14 | -2,50 | 1,24E-02 | 1,82E-01 |
| CHD9 | 3448,25 | 0,14 | 0,05 | 2,50 | 1,24E-02 | 1,82E-01 |
| TMEM18 | 441,29 | 0,08 | 0,03 | 2,50 | 1,25E-02 | 1,82E-01 |
| ZNF43 | 771,86 | 0,13 | 0,05 | 2,50 | 1,26E-02 | 1,82E-01 |
| ATP7B | 127,13 | -0,30 | 0,12 | -2,50 | 1,25E-02 | 1,82E-01 |
| C9orf85 | 317,43 | 0,08 | 0,03 | 2,50 | 1,26E-02 | 1,82E-01 |
| ZNF227 | 942,77 | 0,14 | 0,06 | 2,50 | 1,26E-02 | 1,82E-01 |
| PTPRS | 143,94 | -0,47 | 0,19 | -2,50 | 1,25E-02 | 1,82E-01 |
| TTC7B | 214,97 | -0,37 | 0,15 | -2,50 | 1,26E-02 | 1,82E-01 |
| ACBD3 | 1587,77 | 0,13 | 0,05 | 2,49 | 1,26E-02 | 1,83E-01 |
| ZNF197 | 695,90 | 0,16 | 0,06 | 2,49 | 1,26E-02 | 1,83E-01 |
| BEND3 | 56,33 | -0,36 | 0,15 | -2,49 | 1,27E-02 | 1,83E-01 |
| HAUS6 | 1169,51 | 0,13 | 0,05 | 2,49 | 1,27E-02 | 1,83E-01 |
| DTX1 | 92,17 | -0,35 | 0,14 | -2,49 | 1,27E-02 | 1,83E-01 |
| PCBP3 | 102,85 | -0,42 | 0,17 | -2,49 | 1,27E-02 | 1,83E-01 |
| MBTPS2 | 343,38 | 0,13 | 0,05 | 2,49 | 1,28E-02 | 1,84E-01 |
| SLC6A9 | 130,60 | -0,42 | 0,17 | -2,49 | 1,28E-02 | 1,84E-01 |
| ENSG000002 | 35,53 | -0,27 | 0,11 | -2,49 | 1,29E-02 | 1,84E-01 |
| DPM2 | 1357,88 | -0,30 | 0,12 | -2,49 | 1,29E-02 | 1,84E-01 |
| SCRN3 | 490,49 | 0,13 | 0,05 | 2,49 | 1,29E-02 | 1,84E-01 |
| DYRK3 | 147,19 | -0,32 | 0,13 | -2,49 | 1,29E-02 | 1,85E-01 |
| ARMC1 | 672,58 | 0,15 | 0,06 | 2,49 | 1,30E-02 | 1,85E-01 |
| SINHCAF | 1549,32 | 0,14 | 0,06 | 2,48 | 1,30E-02 | 1,85E-01 |
| CFAP97 | 1071,44 | 0,18 | 0,07 | 2,48 | 1,30E-02 | 1,85E-01 |
| RBM48 | 774,58 | 0,12 | 0,05 | 2,48 | 1,31E-02 | 1,85E-01 |
| PIK3R6 | 341,10 | -0,29 | 0,12 | -2,48 | 1,30E-02 | 1,85E-01 |
| LIG4 | 552,56 | 0,25 | 0,10 | 2,48 | 1,31E-02 | 1,85E-01 |
| FAM20C | 104,02 | -0,36 | 0,15 | -2,48 | 1,31E-02 | 1,85E-01 |
| DHX57 | 812,53 | 0,09 | 0,04 | 2,48 | 1,30E-02 | 1,85E-01 |
| ZNF461 | 303,02 | 0,13 | 0,05 | 2,48 | 1,31E-02 | 1,85E-01 |

| | | | | | | |
|---|---|---|---|---|---|---|
| RGS6 | 195,21 | -0,34 | 0,14 | -2,48 | 1,31E-02 | 1,85E-01 |
| PDCD5 | 417,25 | 0,11 | 0,05 | 2,48 | 1,33E-02 | 1,85E-01 |
| WRAP73 | 570,49 | -0,13 | 0,05 | -2,48 | 1,33E-02 | 1,85E-01 |
| TRPM7 | 3685,44 | 0,14 | 0,06 | 2,48 | 1,33E-02 | 1,85E-01 |
| LRRC40 | 554,18 | 0,22 | 0,09 | 2,48 | 1,32E-02 | 1,85E-01 |
| RBBP9 | 303,26 | 0,11 | 0,04 | 2,48 | 1,32E-02 | 1,85E-01 |
| GNL3 | 785,97 | 0,20 | 0,08 | 2,48 | 1,32E-02 | 1,85E-01 |
| TONSL | 76,65 | -0,38 | 0,15 | -2,48 | 1,32E-02 | 1,85E-01 |
| LYRM7 | 881,47 | 0,15 | 0,06 | 2,48 | 1,32E-02 | 1,85E-01 |
| IQCH | 49,51 | -0,48 | 0,19 | -2,48 | 1,33E-02 | 1,85E-01 |
| UBE2Q2 | 775,57 | 0,14 | 0,06 | 2,48 | 1,33E-02 | 1,85E-01 |
| PHKG1 | 42,06 | -0,29 | 0,12 | -2,48 | 1,32E-02 | 1,85E-01 |
| KCTD18 | 805,33 | 0,08 | 0,03 | 2,48 | 1,33E-02 | 1,85E-01 |
| ETAA1 | 468,50 | 0,28 | 0,11 | 2,48 | 1,33E-02 | 1,85E-01 |
| HECTD1 | 5758,11 | 0,11 | 0,04 | 2,47 | 1,33E-02 | 1,85E-01 |
| DMGDH | 40,97 | -0,47 | 0,19 | -2,47 | 1,34E-02 | 1,86E-01 |
| AKAP9 | 4694,48 | 0,15 | 0,06 | 2,47 | 1,34E-02 | 1,86E-01 |
| KIF20B | 445,52 | 0,28 | 0,11 | 2,47 | 1,34E-02 | 1,86E-01 |
| MRPS10 | 684,12 | 0,13 | 0,05 | 2,47 | 1,34E-02 | 1,86E-01 |
| MIEN1 | 95,28 | -0,20 | 0,08 | -2,47 | 1,35E-02 | 1,86E-01 |
| ZNF346 | 493,09 | -0,13 | 0,05 | -2,47 | 1,34E-02 | 1,86E-01 |
| CORO2A | 273,60 | -0,17 | 0,07 | -2,47 | 1,35E-02 | 1,86E-01 |
| BPNT2 | 1801,03 | 0,11 | 0,05 | 2,47 | 1,35E-02 | 1,86E-01 |
| TRMT6 | 433,27 | 0,10 | 0,04 | 2,47 | 1,35E-02 | 1,86E-01 |
| CD69 | 384,16 | 0,33 | 0,13 | 2,47 | 1,35E-02 | 1,86E-01 |
| STRBP | 1050,87 | 0,19 | 0,08 | 2,47 | 1,36E-02 | 1,86E-01 |
| RFC1 | 1681,17 | 0,11 | 0,04 | 2,47 | 1,36E-02 | 1,86E-01 |
| VARS2 | 230,54 | -0,22 | 0,09 | -2,47 | 1,37E-02 | 1,86E-01 |
| PRIM2 | 592,11 | 0,13 | 0,05 | 2,46 | 1,38E-02 | 1,86E-01 |
| NPR3 | 43,36 | -0,53 | 0,22 | -2,46 | 1,39E-02 | 1,86E-01 |
| CCNC | 1135,56 | 0,21 | 0,09 | 2,46 | 1,39E-02 | 1,86E-01 |
| SETDB2 | 1336,65 | 0,12 | 0,05 | 2,46 | 1,38E-02 | 1,86E-01 |
| TSPAN3 | 681,23 | 0,14 | 0,06 | 2,46 | 1,38E-02 | 1,86E-01 |
| CCDC6 | 1132,51 | 0,10 | 0,04 | 2,47 | 1,37E-02 | 1,86E-01 |
| FAM83H | 59,30 | -0,41 | 0,17 | -2,47 | 1,36E-02 | 1,86E-01 |
| SLCO5A1 | 50,28 | -0,56 | 0,23 | -2,47 | 1,36E-02 | 1,86E-01 |
| GMPS | 1105,40 | 0,11 | 0,05 | 2,47 | 1,37E-02 | 1,86E-01 |
| FYTTD1 | 1646,39 | 0,14 | 0,06 | 2,46 | 1,38E-02 | 1,86E-01 |
| ZFP14 | 808,31 | 0,15 | 0,06 | 2,47 | 1,37E-02 | 1,86E-01 |
| ENTPD5 | 487,90 | -0,17 | 0,07 | -2,46 | 1,38E-02 | 1,86E-01 |
| MYO7B | 131,40 | -0,32 | 0,13 | -2,46 | 1,38E-02 | 1,86E-01 |
| ZNF567 | 315,94 | 0,18 | 0,07 | 2,46 | 1,39E-02 | 1,86E-01 |
| BORCS7 | 365,39 | 0,23 | 0,09 | 2,46 | 1,39E-02 | 1,86E-01 |
| CDC42BPG | 92,73 | -0,35 | 0,14 | -2,46 | 1,38E-02 | 1,86E-01 |
| NIFK | 492,35 | 0,17 | 0,07 | 2,46 | 1,38E-02 | 1,86E-01 |
| BTAF1 | 5544,74 | 0,13 | 0,05 | 2,47 | 1,37E-02 | 1,86E-01 |

| | | | | | | |
|---|---|---|---|---|---|---|
| ALG5 | 368,60 | 0,15 | 0,06 | 2,46 | 1,37E-02 | 1,86E-01 |
| RIF1 | 4730,02 | 0,16 | 0,07 | 2,46 | 1,38E-02 | 1,86E-01 |
| C21orf58 | 86,89 | -0,26 | 0,11 | -2,46 | 1,39E-02 | 1,86E-01 |
| IARS2 | 1328,67 | 0,09 | 0,04 | 2,46 | 1,38E-02 | 1,86E-01 |
| PLXNA1 | 217,13 | -0,35 | 0,14 | -2,46 | 1,39E-02 | 1,86E-01 |
| BCAS1 | 40,78 | -0,54 | 0,22 | -2,46 | 1,40E-02 | 1,86E-01 |
| TMUB2 | 637,65 | -0,14 | 0,06 | -2,46 | 1,41E-02 | 1,86E-01 |
| CDPF1 | 108,08 | -0,11 | 0,05 | -2,46 | 1,41E-02 | 1,86E-01 |
| CFAP92 | 366,99 | -0,25 | 0,10 | -2,46 | 1,40E-02 | 1,86E-01 |
| PDCD4 | 6547,28 | 0,14 | 0,06 | 2,46 | 1,40E-02 | 1,86E-01 |
| ZNF226 | 1269,10 | 0,19 | 0,08 | 2,46 | 1,39E-02 | 1,86E-01 |
| NARS2 | 589,18 | 0,11 | 0,04 | 2,45 | 1,41E-02 | 1,86E-01 |
| RAI14 | 46,66 | -0,61 | 0,25 | -2,46 | 1,40E-02 | 1,86E-01 |
| PAK1IP1 | 160,62 | 0,16 | 0,07 | 2,46 | 1,41E-02 | 1,86E-01 |
| ZNF510 | 854,27 | 0,19 | 0,08 | 2,46 | 1,41E-02 | 1,86E-01 |
| EDARADD | 57,09 | -0,30 | 0,12 | -2,46 | 1,41E-02 | 1,86E-01 |
| SMC2 | 470,49 | 0,19 | 0,08 | 2,45 | 1,41E-02 | 1,86E-01 |
| RTCB | 969,65 | 0,11 | 0,05 | 2,45 | 1,42E-02 | 1,87E-01 |
| ENY2 | 762,12 | 0,15 | 0,06 | 2,45 | 1,42E-02 | 1,87E-01 |
| DEK | 2753,81 | 0,23 | 0,09 | 2,45 | 1,42E-02 | 1,87E-01 |
| DRC1 | 39,93 | -0,47 | 0,19 | -2,45 | 1,43E-02 | 1,87E-01 |
| RANBP2 | 5318,91 | 0,14 | 0,06 | 2,45 | 1,43E-02 | 1,87E-01 |
| TAF2 | 1801,47 | 0,12 | 0,05 | 2,45 | 1,43E-02 | 1,87E-01 |
| LSS | 866,73 | -0,15 | 0,06 | -2,45 | 1,43E-02 | 1,88E-01 |
| ROPN1L | 377,69 | -0,20 | 0,08 | -2,45 | 1,43E-02 | 1,88E-01 |
| ZNF844 | 856,00 | 0,19 | 0,08 | 2,45 | 1,44E-02 | 1,88E-01 |
| DDX11 | 508,28 | 0,31 | 0,13 | 2,45 | 1,44E-02 | 1,88E-01 |
| TPRG1L | 1181,34 | -0,14 | 0,06 | -2,45 | 1,45E-02 | 1,88E-01 |
| ZNF772 | 412,75 | 0,19 | 0,08 | 2,45 | 1,45E-02 | 1,88E-01 |
| THUMPD1 | 2169,43 | 0,16 | 0,07 | 2,44 | 1,45E-02 | 1,88E-01 |
| ZFYVE19 | 375,29 | -0,10 | 0,04 | -2,44 | 1,45E-02 | 1,88E-01 |
| ATP10D | 1565,22 | 0,13 | 0,05 | 2,44 | 1,45E-02 | 1,88E-01 |
| USP14 | 838,29 | 0,14 | 0,06 | 2,45 | 1,45E-02 | 1,88E-01 |
| ZNF417 | 1235,72 | 0,11 | 0,04 | 2,44 | 1,45E-02 | 1,88E-01 |
| SLTM | 4064,96 | 0,14 | 0,06 | 2,44 | 1,46E-02 | 1,88E-01 |
| ZNF430 | 1250,94 | 0,11 | 0,04 | 2,44 | 1,46E-02 | 1,88E-01 |
| THOC2 | 2701,95 | 0,15 | 0,06 | 2,44 | 1,45E-02 | 1,88E-01 |
| GSPT2 | 126,04 | 0,16 | 0,07 | 2,44 | 1,48E-02 | 1,88E-01 |
| GNA11 | 67,24 | -0,37 | 0,15 | -2,43 | 1,50E-02 | 1,88E-01 |
| G3BP1 | 5057,35 | 0,09 | 0,04 | 2,43 | 1,51E-02 | 1,88E-01 |
| CUL5 | 1610,34 | 0,18 | 0,07 | 2,43 | 1,51E-02 | 1,88E-01 |
| MAOA | 58,85 | -0,51 | 0,21 | -2,44 | 1,48E-02 | 1,88E-01 |
| DENND6B | 52,55 | -0,41 | 0,17 | -2,44 | 1,48E-02 | 1,88E-01 |
| SYNCRIP | 3190,48 | 0,15 | 0,06 | 2,44 | 1,47E-02 | 1,88E-01 |
| KIAA1191 | 1064,37 | -0,09 | 0,04 | -2,43 | 1,51E-02 | 1,88E-01 |
| TMEM192 | 490,21 | 0,07 | 0,03 | 2,43 | 1,50E-02 | 1,88E-01 |

| | | | | | | |
|---|---|---|---|---|---|---|
| DNAI3 | 40,33 | -0,42 | 0,17 | -2,43 | 1,50E-02 | 1,88E-01 |
| USP37 | 1206,94 | 0,10 | 0,04 | 2,43 | 1,49E-02 | 1,88E-01 |
| ZNF561 | 741,06 | 0,11 | 0,05 | 2,43 | 1,51E-02 | 1,88E-01 |
| SLC9A5 | 39,17 | -0,46 | 0,19 | -2,43 | 1,52E-02 | 1,88E-01 |
| ZPR1 | 845,78 | 0,08 | 0,03 | 2,44 | 1,46E-02 | 1,88E-01 |
| HAS3 | 42,55 | -0,41 | 0,17 | -2,43 | 1,50E-02 | 1,88E-01 |
| ZDHHC6 | 1334,65 | 0,10 | 0,04 | 2,44 | 1,46E-02 | 1,88E-01 |
| ZNF107 | 1312,32 | 0,21 | 0,09 | 2,44 | 1,47E-02 | 1,88E-01 |
| VEPH1 | 66,66 | -0,38 | 0,16 | -2,43 | 1,49E-02 | 1,88E-01 |
| ANKRD27 | 1231,19 | -0,07 | 0,03 | -2,43 | 1,51E-02 | 1,88E-01 |
| RASA1 | 2367,04 | 0,13 | 0,05 | 2,43 | 1,52E-02 | 1,88E-01 |
| DNTTIP2 | 839,96 | 0,21 | 0,09 | 2,43 | 1,49E-02 | 1,88E-01 |
| GPATCH11 | 570,60 | 0,20 | 0,08 | 2,44 | 1,48E-02 | 1,88E-01 |
| UBL7 | 976,26 | -0,16 | 0,07 | -2,44 | 1,47E-02 | 1,88E-01 |
| BLVRB | 9110,95 | -0,30 | 0,12 | -2,43 | 1,50E-02 | 1,88E-01 |
| TRA2B | 6404,80 | 0,13 | 0,05 | 2,44 | 1,48E-02 | 1,88E-01 |
| ADCY10 | 60,15 | -0,32 | 0,13 | -2,43 | 1,51E-02 | 1,88E-01 |
| DDX18 | 2101,42 | 0,13 | 0,05 | 2,44 | 1,46E-02 | 1,88E-01 |
| MSH3 | 819,48 | 0,11 | 0,04 | 2,44 | 1,48E-02 | 1,88E-01 |
| MRPS18C | 687,63 | 0,20 | 0,08 | 2,43 | 1,49E-02 | 1,88E-01 |
| MORF4L2 | 1178,85 | 0,14 | 0,06 | 2,44 | 1,47E-02 | 1,88E-01 |
| CYB5R3 | 315,86 | -0,27 | 0,11 | -2,43 | 1,52E-02 | 1,88E-01 |
| GIN1 | 336,80 | 0,20 | 0,08 | 2,43 | 1,50E-02 | 1,88E-01 |
| KRR1 | 1305,73 | 0,20 | 0,08 | 2,43 | 1,51E-02 | 1,88E-01 |
| EIF3A | 5870,65 | 0,11 | 0,05 | 2,44 | 1,48E-02 | 1,88E-01 |
| ZNF337 | 1056,86 | 0,10 | 0,04 | 2,43 | 1,50E-02 | 1,88E-01 |
| MCUB | 1456,63 | 0,11 | 0,05 | 2,43 | 1,51E-02 | 1,88E-01 |
| CIP2A | 268,93 | 0,20 | 0,08 | 2,43 | 1,50E-02 | 1,88E-01 |
| ZNF880 | 224,10 | 0,15 | 0,06 | 2,43 | 1,49E-02 | 1,88E-01 |
| KMT5A | 331,62 | -0,13 | 0,05 | -2,43 | 1,52E-02 | 1,89E-01 |
| SECISBP2L | 2268,02 | 0,11 | 0,05 | 2,43 | 1,52E-02 | 1,89E-01 |
| ENTREP3 | 49,14 | -0,40 | 0,16 | -2,43 | 1,52E-02 | 1,89E-01 |
| WBP11 | 413,70 | 0,13 | 0,05 | 2,43 | 1,53E-02 | 1,89E-01 |
| TAS2R19 | 236,84 | 0,19 | 0,08 | 2,43 | 1,53E-02 | 1,89E-01 |
| MRPL27 | 413,04 | 0,10 | 0,04 | 2,43 | 1,53E-02 | 1,89E-01 |
| UBFD1 | 693,78 | -0,11 | 0,04 | -2,42 | 1,53E-02 | 1,89E-01 |
| SLC13A3 | 46,06 | -0,47 | 0,19 | -2,42 | 1,53E-02 | 1,89E-01 |
| CARM1 | 500,27 | -0,34 | 0,14 | -2,42 | 1,54E-02 | 1,90E-01 |
| ZNF235 | 562,55 | 0,09 | 0,04 | 2,42 | 1,54E-02 | 1,90E-01 |
| HDHD2 | 585,23 | 0,12 | 0,05 | 2,42 | 1,54E-02 | 1,90E-01 |
| C1S | 36,73 | -0,53 | 0,22 | -2,42 | 1,55E-02 | 1,90E-01 |
| RGS1 | 61,30 | 0,37 | 0,15 | 2,42 | 1,55E-02 | 1,90E-01 |
| AJM1 | 75,95 | -0,34 | 0,14 | -2,42 | 1,56E-02 | 1,91E-01 |
| CBR4 | 1491,08 | 0,13 | 0,05 | 2,42 | 1,56E-02 | 1,91E-01 |
| SEPHS2 | 552,98 | -0,12 | 0,05 | -2,42 | 1,56E-02 | 1,91E-01 |
| PPAT | 340,92 | 0,14 | 0,06 | 2,42 | 1,57E-02 | 1,91E-01 |

| | | | | | | |
|---|---|---|---|---|---|---|
| BAIAP2 | 114,47 | -0,44 | 0,18 | -2,42 | 1,57E-02 | 1,91E-01 |
| HLTF | 653,66 | 0,18 | 0,08 | 2,42 | 1,56E-02 | 1,91E-01 |
| RAB3C | 35,33 | -0,45 | 0,19 | -2,42 | 1,57E-02 | 1,91E-01 |
| CCDC85C | 109,82 | -0,41 | 0,17 | -2,42 | 1,57E-02 | 1,91E-01 |
| ZNF644 | 2735,07 | 0,22 | 0,09 | 2,42 | 1,56E-02 | 1,91E-01 |
| BDP1 | 3485,80 | 0,14 | 0,06 | 2,42 | 1,56E-02 | 1,91E-01 |
| THAP9 | 145,48 | 0,21 | 0,09 | 2,42 | 1,57E-02 | 1,91E-01 |
| PMPCB | 1381,12 | 0,12 | 0,05 | 2,42 | 1,56E-02 | 1,91E-01 |
| INCENP | 278,77 | -0,15 | 0,06 | -2,41 | 1,59E-02 | 1,91E-01 |
| DUSP4 | 57,55 | -0,44 | 0,18 | -2,41 | 1,59E-02 | 1,91E-01 |
| MGA | 4241,23 | 0,09 | 0,04 | 2,41 | 1,59E-02 | 1,91E-01 |
| IARS1 | 1980,98 | 0,10 | 0,04 | 2,41 | 1,58E-02 | 1,91E-01 |
| DUSP12 | 369,12 | 0,11 | 0,05 | 2,41 | 1,58E-02 | 1,91E-01 |
| PGM3 | 718,13 | 0,14 | 0,06 | 2,41 | 1,58E-02 | 1,91E-01 |
| KANK2 | 193,37 | -0,42 | 0,17 | -2,41 | 1,58E-02 | 1,91E-01 |
| ALG11 | 949,06 | 0,10 | 0,04 | 2,41 | 1,60E-02 | 1,91E-01 |
| IFT74 | 268,34 | 0,15 | 0,06 | 2,41 | 1,59E-02 | 1,91E-01 |
| LRFN4 | 62,39 | -0,34 | 0,14 | -2,41 | 1,60E-02 | 1,91E-01 |
| CDCA7L | 628,94 | 0,19 | 0,08 | 2,41 | 1,59E-02 | 1,91E-01 |
| DMAC2 | 106,07 | -0,17 | 0,07 | -2,41 | 1,59E-02 | 1,91E-01 |
| NEPRO | 1365,85 | 0,12 | 0,05 | 2,41 | 1,59E-02 | 1,91E-01 |
| TGM1 | 49,10 | -0,33 | 0,14 | -2,41 | 1,60E-02 | 1,91E-01 |
| PYGM | 149,05 | -0,26 | 0,11 | -2,41 | 1,58E-02 | 1,91E-01 |
| ZNF511 | 99,33 | -0,15 | 0,06 | -2,41 | 1,58E-02 | 1,91E-01 |
| IFT70B | 141,41 | 0,18 | 0,08 | 2,41 | 1,61E-02 | 1,91E-01 |
| COMTD1 | 102,20 | -0,16 | 0,07 | -2,41 | 1,60E-02 | 1,91E-01 |
| SEMA6C | 49,24 | -0,48 | 0,20 | -2,41 | 1,61E-02 | 1,91E-01 |
| ZNF234 | 627,95 | 0,09 | 0,04 | 2,41 | 1,61E-02 | 1,91E-01 |
| NGDN | 521,24 | 0,12 | 0,05 | 2,41 | 1,61E-02 | 1,91E-01 |
| NFYB | 439,68 | 0,22 | 0,09 | 2,41 | 1,61E-02 | 1,91E-01 |
| HSPA12A | 71,41 | -0,52 | 0,21 | -2,41 | 1,61E-02 | 1,91E-01 |
| ITGA10 | 92,09 | -0,28 | 0,12 | -2,41 | 1,61E-02 | 1,91E-01 |
| OPRL1 | 240,57 | -0,19 | 0,08 | -2,41 | 1,61E-02 | 1,91E-01 |
| MDFIC | 2630,19 | 0,16 | 0,07 | 2,41 | 1,61E-02 | 1,91E-01 |
| GCNT2 | 497,91 | 0,15 | 0,06 | 2,41 | 1,62E-02 | 1,91E-01 |
| ARHGEF17 | 56,71 | -0,61 | 0,25 | -2,40 | 1,65E-02 | 1,91E-01 |
| FUCA2 | 495,63 | 0,12 | 0,05 | 2,40 | 1,66E-02 | 1,91E-01 |
| DOCK1 | 74,00 | -0,49 | 0,21 | -2,40 | 1,66E-02 | 1,91E-01 |
| ZMYND12 | 24,08 | -0,32 | 0,14 | -2,40 | 1,62E-02 | 1,91E-01 |
| MRPL30 | 493,91 | 0,11 | 0,05 | 2,40 | 1,62E-02 | 1,91E-01 |
| KCNA2 | 127,56 | -0,35 | 0,15 | -2,40 | 1,66E-02 | 1,91E-01 |
| PRMT9 | 484,00 | 0,12 | 0,05 | 2,40 | 1,65E-02 | 1,91E-01 |
| TAF1B | 572,61 | 0,12 | 0,05 | 2,40 | 1,64E-02 | 1,91E-01 |
| PRRC1 | 1199,51 | 0,11 | 0,05 | 2,40 | 1,63E-02 | 1,91E-01 |
| NOL10 | 690,98 | 0,08 | 0,03 | 2,39 | 1,68E-02 | 1,91E-01 |
| ZSCAN26 | 409,91 | 0,12 | 0,05 | 2,40 | 1,63E-02 | 1,91E-01 |

| | | | | | | |
|---|---|---|---|---|---|---|
| CASZ1 | 236,44 | -0,32 | 0,13 | -2,39 | 1,67E-02 | 1,91E-01 |
| RBIS | 466,94 | 0,21 | 0,09 | 2,40 | 1,64E-02 | 1,91E-01 |
| HSD3B7 | 34,61 | -0,34 | 0,14 | -2,39 | 1,68E-02 | 1,91E-01 |
| WDR89 | 466,91 | 0,27 | 0,11 | 2,39 | 1,69E-02 | 1,91E-01 |
| MISP3 | 29,04 | -0,31 | 0,13 | -2,40 | 1,65E-02 | 1,91E-01 |
| CMTR2 | 1360,12 | 0,19 | 0,08 | 2,40 | 1,63E-02 | 1,91E-01 |
| ERCC6L2 | 2082,34 | 0,14 | 0,06 | 2,40 | 1,66E-02 | 1,91E-01 |
| KY | 39,17 | -0,39 | 0,16 | -2,40 | 1,66E-02 | 1,91E-01 |
| TMPRSS5 | 48,77 | -0,34 | 0,14 | -2,39 | 1,69E-02 | 1,91E-01 |
| PDE12 | 1146,16 | 0,07 | 0,03 | 2,40 | 1,63E-02 | 1,91E-01 |
| S1PR2 | 54,54 | -0,27 | 0,11 | -2,39 | 1,67E-02 | 1,91E-01 |
| NECAB3 | 125,00 | -0,21 | 0,09 | -2,40 | 1,65E-02 | 1,91E-01 |
| ZNF160 | 1379,16 | 0,08 | 0,03 | 2,39 | 1,68E-02 | 1,91E-01 |
| SLC23A3 | 25,92 | -0,35 | 0,15 | -2,39 | 1,68E-02 | 1,91E-01 |
| CD86 | 978,74 | 0,15 | 0,06 | 2,39 | 1,67E-02 | 1,91E-01 |
| HES1 | 46,33 | -0,30 | 0,13 | -2,40 | 1,64E-02 | 1,91E-01 |
| FANCA | 305,15 | -0,25 | 0,10 | -2,40 | 1,65E-02 | 1,91E-01 |
| NME4 | 280,74 | -0,19 | 0,08 | -2,39 | 1,68E-02 | 1,91E-01 |
| MASTL | 455,69 | 0,16 | 0,07 | 2,39 | 1,69E-02 | 1,91E-01 |
| PDP1 | 1589,13 | 0,15 | 0,06 | 2,39 | 1,67E-02 | 1,91E-01 |
| SHQ1 | 573,41 | 0,10 | 0,04 | 2,40 | 1,65E-02 | 1,91E-01 |
| CPS1 | 44,84 | -0,56 | 0,23 | -2,39 | 1,69E-02 | 1,91E-01 |
| PHF10 | 1347,48 | 0,15 | 0,06 | 2,39 | 1,68E-02 | 1,91E-01 |
| POC5 | 571,14 | 0,12 | 0,05 | 2,39 | 1,68E-02 | 1,91E-01 |
| PIN4 | 239,95 | 0,13 | 0,06 | 2,40 | 1,66E-02 | 1,91E-01 |
| NOD2 | 1611,13 | -0,20 | 0,08 | -2,40 | 1,65E-02 | 1,91E-01 |
| RTRAF | 1029,69 | 0,16 | 0,07 | 2,40 | 1,65E-02 | 1,91E-01 |
| SLC47A1 | 57,83 | -0,46 | 0,19 | -2,40 | 1,64E-02 | 1,91E-01 |
| FKBP9 | 126,14 | -0,26 | 0,11 | -2,39 | 1,68E-02 | 1,91E-01 |
| PTPRN2 | 288,92 | -0,23 | 0,10 | -2,39 | 1,67E-02 | 1,91E-01 |
| METTL5 | 239,73 | 0,14 | 0,06 | 2,40 | 1,64E-02 | 1,91E-01 |
| GPLD1 | 92,25 | -0,32 | 0,13 | -2,40 | 1,65E-02 | 1,91E-01 |
| TXN2 | 158,03 | -0,16 | 0,06 | -2,40 | 1,66E-02 | 1,91E-01 |
| TMA16 | 550,63 | 0,16 | 0,07 | 2,40 | 1,66E-02 | 1,91E-01 |
| C1orf198 | 254,06 | -0,28 | 0,12 | -2,39 | 1,69E-02 | 1,91E-01 |
| BBS5 | 28,16 | -0,42 | 0,18 | -2,39 | 1,69E-02 | 1,91E-01 |
| ZNF329 | 352,34 | 0,12 | 0,05 | 2,39 | 1,69E-02 | 1,91E-01 |
| KCNMA1 | 173,35 | -0,64 | 0,27 | -2,39 | 1,70E-02 | 1,91E-01 |
| UBA5 | 842,37 | 0,18 | 0,08 | 2,39 | 1,70E-02 | 1,92E-01 |
| MRPL3 | 807,01 | 0,19 | 0,08 | 2,38 | 1,71E-02 | 1,92E-01 |
| MTFR1L | 722,04 | -0,08 | 0,03 | -2,38 | 1,71E-02 | 1,92E-01 |
| GALNT14 | 159,48 | -0,34 | 0,14 | -2,38 | 1,71E-02 | 1,92E-01 |
| CCND1 | 57,93 | -0,43 | 0,18 | -2,39 | 1,71E-02 | 1,92E-01 |
| NDUFAF3 | 602,25 | -0,12 | 0,05 | -2,39 | 1,70E-02 | 1,92E-01 |
| MTERF1 | 701,70 | 0,19 | 0,08 | 2,39 | 1,71E-02 | 1,92E-01 |
| ZNF397 | 2237,92 | 0,08 | 0,03 | 2,39 | 1,71E-02 | 1,92E-01 |

| | | | | | | |
|---|---|---|---|---|---|---|
| ZNF14 | 499,43 | 0,23 | 0,10 | 2,39 | 1,71E-02 | 1,92E-01 |
| ZCCHC7 | 1786,65 | 0,16 | 0,07 | 2,38 | 1,73E-02 | 1,92E-01 |
| CCDC73 | 32,45 | -0,47 | 0,20 | -2,37 | 1,77E-02 | 1,92E-01 |
| ZC3H13 | 2592,51 | 0,09 | 0,04 | 2,36 | 1,80E-02 | 1,92E-01 |
| BNIPL | 72,97 | -0,19 | 0,08 | -2,38 | 1,75E-02 | 1,92E-01 |
| CMYA5 | 73,90 | -0,39 | 0,16 | -2,38 | 1,75E-02 | 1,92E-01 |
| CHCHD7 | 818,70 | 0,12 | 0,05 | 2,36 | 1,80E-02 | 1,92E-01 |
| PUM3 | 706,68 | 0,14 | 0,06 | 2,37 | 1,78E-02 | 1,92E-01 |
| ZNF791 | 1391,22 | 0,14 | 0,06 | 2,37 | 1,76E-02 | 1,92E-01 |
| MIER2 | 178,70 | -0,31 | 0,13 | -2,38 | 1,74E-02 | 1,92E-01 |
| PRP4K | 3376,49 | 0,12 | 0,05 | 2,37 | 1,78E-02 | 1,92E-01 |
| CDC42EP2 | 50,43 | -0,23 | 0,10 | -2,36 | 1,81E-02 | 1,92E-01 |
| JMY | 1311,30 | 0,11 | 0,05 | 2,37 | 1,76E-02 | 1,92E-01 |
| CEP57 | 1421,85 | 0,19 | 0,08 | 2,37 | 1,79E-02 | 1,92E-01 |
| DCAF16 | 1865,97 | 0,09 | 0,04 | 2,36 | 1,81E-02 | 1,92E-01 |
| COA8 | 264,75 | -0,09 | 0,04 | -2,37 | 1,76E-02 | 1,92E-01 |
| ZSWIM4 | 82,55 | -0,32 | 0,13 | -2,37 | 1,78E-02 | 1,92E-01 |
| ZNF816 | 312,00 | 0,23 | 0,10 | 2,38 | 1,75E-02 | 1,92E-01 |
| PEAR1 | 58,95 | -0,53 | 0,22 | -2,38 | 1,72E-02 | 1,92E-01 |
| CCAR1 | 1866,76 | 0,08 | 0,03 | 2,37 | 1,77E-02 | 1,92E-01 |
| COIL | 482,14 | 0,12 | 0,05 | 2,36 | 1,81E-02 | 1,92E-01 |
| SCUBE2 | 67,58 | -0,41 | 0,17 | -2,37 | 1,77E-02 | 1,92E-01 |
| KLHL11 | 408,35 | 0,10 | 0,04 | 2,38 | 1,75E-02 | 1,92E-01 |
| MMP11 | 38,67 | -0,42 | 0,18 | -2,36 | 1,81E-02 | 1,92E-01 |
| C19orf47 | 87,12 | -0,30 | 0,13 | -2,36 | 1,80E-02 | 1,92E-01 |
| CCSER2 | 2773,53 | 0,15 | 0,06 | 2,37 | 1,76E-02 | 1,92E-01 |
| CCDC25 | 460,22 | 0,12 | 0,05 | 2,37 | 1,78E-02 | 1,92E-01 |
| MYO3B | 83,54 | -0,54 | 0,23 | -2,36 | 1,80E-02 | 1,92E-01 |
| YIPF5 | 914,28 | 0,16 | 0,07 | 2,37 | 1,76E-02 | 1,92E-01 |
| RUSC1 | 84,22 | -0,25 | 0,11 | -2,37 | 1,77E-02 | 1,92E-01 |
| FHL1 | 352,12 | -0,20 | 0,09 | -2,38 | 1,74E-02 | 1,92E-01 |
| POLR2B | 4036,25 | 0,15 | 0,06 | 2,38 | 1,74E-02 | 1,92E-01 |
| CDC5L | 1977,09 | 0,15 | 0,06 | 2,37 | 1,79E-02 | 1,92E-01 |
| FASTKD2 | 840,61 | 0,16 | 0,07 | 2,37 | 1,77E-02 | 1,92E-01 |
| MRPL10 | 224,39 | -0,11 | 0,05 | -2,38 | 1,75E-02 | 1,92E-01 |
| ZNF700 | 819,05 | 0,11 | 0,05 | 2,38 | 1,74E-02 | 1,92E-01 |
| DHX15 | 4936,63 | 0,15 | 0,06 | 2,38 | 1,74E-02 | 1,92E-01 |
| PAPLN | 67,90 | -0,46 | 0,19 | -2,37 | 1,79E-02 | 1,92E-01 |
| IL17RC | 60,68 | -0,29 | 0,12 | -2,37 | 1,78E-02 | 1,92E-01 |
| ZMAT5 | 280,90 | -0,10 | 0,04 | -2,37 | 1,80E-02 | 1,92E-01 |
| ITGAD | 48,68 | -0,46 | 0,19 | -2,37 | 1,79E-02 | 1,92E-01 |
| KTN1 | 2870,37 | 0,17 | 0,07 | 2,38 | 1,73E-02 | 1,92E-01 |
| PTDSS2 | 195,81 | -0,20 | 0,08 | -2,38 | 1,74E-02 | 1,92E-01 |
| AEBP1 | 56,41 | -0,48 | 0,20 | -2,37 | 1,77E-02 | 1,92E-01 |
| SLC6A4 | 50,78 | -0,45 | 0,19 | -2,37 | 1,79E-02 | 1,92E-01 |
| MAP3K6 | 131,82 | -0,24 | 0,10 | -2,38 | 1,71E-02 | 1,92E-01 |

| | | | | | | |
|---|---|---|---|---|---|---|
| IFITM10 | 76,15 | -0,26 | 0,11 | -2,37 | 1,76E-02 | 1,92E-01 |
| RNMT | 1100,63 | 0,10 | 0,04 | 2,37 | 1,80E-02 | 1,92E-01 |
| CWC22 | 1032,48 | 0,17 | 0,07 | 2,38 | 1,74E-02 | 1,92E-01 |
| DHX36 | 2613,92 | 0,12 | 0,05 | 2,38 | 1,75E-02 | 1,92E-01 |
| LSM3 | 258,56 | 0,19 | 0,08 | 2,38 | 1,74E-02 | 1,92E-01 |
| GNPDA2 | 489,84 | 0,21 | 0,09 | 2,37 | 1,79E-02 | 1,92E-01 |
| DDX21 | 2669,71 | 0,13 | 0,05 | 2,37 | 1,80E-02 | 1,92E-01 |
| MYD88 | 1841,92 | -0,19 | 0,08 | -2,38 | 1,72E-02 | 1,92E-01 |
| RPP25 | 27,08 | -0,34 | 0,14 | -2,38 | 1,73E-02 | 1,92E-01 |
| FSCN3 | 42,04 | -0,26 | 0,11 | -2,37 | 1,79E-02 | 1,92E-01 |
| HDHD3 | 152,49 | -0,15 | 0,06 | -2,38 | 1,74E-02 | 1,92E-01 |
| FCRLB | 37,19 | -0,28 | 0,12 | -2,37 | 1,78E-02 | 1,92E-01 |
| HMBS | 226,13 | -0,25 | 0,10 | -2,38 | 1,74E-02 | 1,92E-01 |
| PPP2R5B | 604,73 | -0,24 | 0,10 | -2,37 | 1,80E-02 | 1,92E-01 |
| ACTA2 | 65,87 | -0,30 | 0,13 | -2,38 | 1,75E-02 | 1,92E-01 |
| DDX20 | 671,14 | 0,13 | 0,05 | 2,38 | 1,73E-02 | 1,92E-01 |
| ZNF683 | 69,43 | -0,53 | 0,22 | -2,37 | 1,79E-02 | 1,92E-01 |
| ZBTB26 | 423,80 | 0,14 | 0,06 | 2,36 | 1,81E-02 | 1,92E-01 |
| SLFNL1 | 33,43 | -0,42 | 0,18 | -2,36 | 1,81E-02 | 1,92E-01 |
| THOC1 | 1167,95 | 0,13 | 0,06 | 2,36 | 1,81E-02 | 1,92E-01 |
| FAM114A1 | 106,62 | -0,18 | 0,08 | -2,36 | 1,82E-02 | 1,92E-01 |
| MCEE | 306,54 | 0,10 | 0,04 | 2,36 | 1,82E-02 | 1,92E-01 |
| LANCL1 | 1049,52 | 0,13 | 0,05 | 2,36 | 1,83E-02 | 1,92E-01 |
| AMZ1 | 61,46 | -0,40 | 0,17 | -2,36 | 1,83E-02 | 1,92E-01 |
| DDX50 | 845,59 | 0,16 | 0,07 | 2,36 | 1,83E-02 | 1,92E-01 |
| PANX1 | 521,29 | 0,11 | 0,05 | 2,36 | 1,82E-02 | 1,92E-01 |
| SLC33A1 | 693,47 | 0,11 | 0,05 | 2,36 | 1,83E-02 | 1,92E-01 |
| TFE3 | 893,07 | -0,20 | 0,08 | -2,36 | 1,83E-02 | 1,92E-01 |
| SENP5 | 2990,78 | 0,08 | 0,03 | 2,36 | 1,83E-02 | 1,92E-01 |
| DCTN3 | 658,37 | -0,09 | 0,04 | -2,36 | 1,82E-02 | 1,92E-01 |
| ZNF724 | 74,22 | 0,15 | 0,06 | 2,36 | 1,83E-02 | 1,92E-01 |
| NCL | 3603,10 | 0,12 | 0,05 | 2,36 | 1,84E-02 | 1,92E-01 |
| ZNF432 | 454,04 | 0,19 | 0,08 | 2,36 | 1,84E-02 | 1,92E-01 |
| OMA1 | 1250,76 | 0,15 | 0,06 | 2,36 | 1,84E-02 | 1,92E-01 |
| SELENOS | 474,10 | 0,09 | 0,04 | 2,36 | 1,84E-02 | 1,92E-01 |
| COL6A3 | 163,57 | -0,42 | 0,18 | -2,36 | 1,84E-02 | 1,92E-01 |
| PRKCZ | 541,52 | -0,14 | 0,06 | -2,36 | 1,84E-02 | 1,92E-01 |
| ADAM32 | 44,53 | -0,50 | 0,21 | -2,35 | 1,86E-02 | 1,92E-01 |
| SEC63 | 1506,48 | 0,09 | 0,04 | 2,35 | 1,89E-02 | 1,92E-01 |
| TMEM191C | 25,29 | -0,27 | 0,12 | -2,35 | 1,88E-02 | 1,92E-01 |
| C10orf88 | 327,53 | 0,13 | 0,06 | 2,35 | 1,89E-02 | 1,92E-01 |
| ABTB2 | 142,83 | -0,32 | 0,14 | -2,35 | 1,87E-02 | 1,92E-01 |
| CERS4 | 361,73 | -0,17 | 0,07 | -2,36 | 1,85E-02 | 1,92E-01 |
| TXLNG | 583,46 | 0,11 | 0,04 | 2,35 | 1,87E-02 | 1,92E-01 |
| WBP2 | 1189,20 | -0,25 | 0,11 | -2,35 | 1,86E-02 | 1,92E-01 |
| AFG3L2 | 1080,80 | 0,07 | 0,03 | 2,35 | 1,88E-02 | 1,92E-01 |

| | | | | | | |
|---|---|---|---|---|---|---|
| NIF3L1 | 420,30 | 0,11 | 0,05 | 2,35 | 1,89E-02 | 1,92E-01 |
| SLC25A24 | 1476,24 | 0,15 | 0,06 | 2,35 | 1,89E-02 | 1,92E-01 |
| NOL11 | 1047,03 | 0,13 | 0,05 | 2,35 | 1,89E-02 | 1,92E-01 |
| HAUS2 | 477,31 | 0,09 | 0,04 | 2,35 | 1,86E-02 | 1,92E-01 |
| RAB40B | 43,23 | -0,36 | 0,15 | -2,35 | 1,85E-02 | 1,92E-01 |
| MAIP1 | 117,48 | 0,14 | 0,06 | 2,35 | 1,87E-02 | 1,92E-01 |
| SACS | 2111,33 | 0,24 | 0,10 | 2,35 | 1,89E-02 | 1,92E-01 |
| PSMC6 | 1340,50 | 0,18 | 0,08 | 2,36 | 1,85E-02 | 1,92E-01 |
| UNC45B | 53,52 | -0,38 | 0,16 | -2,35 | 1,88E-02 | 1,92E-01 |
| FN3K | 49,49 | -0,36 | 0,15 | -2,35 | 1,89E-02 | 1,92E-01 |
| KDM1A | 1183,94 | 0,08 | 0,04 | 2,35 | 1,87E-02 | 1,92E-01 |
| MTRF1L | 523,43 | 0,16 | 0,07 | 2,35 | 1,89E-02 | 1,92E-01 |
| GALNT5 | 91,35 | -0,31 | 0,13 | -2,35 | 1,89E-02 | 1,92E-01 |
| UHRF2 | 3436,94 | 0,12 | 0,05 | 2,35 | 1,89E-02 | 1,92E-01 |
| BANK1 | 1573,02 | 0,24 | 0,10 | 2,35 | 1,88E-02 | 1,92E-01 |
| UQCRHL | 42,10 | -0,21 | 0,09 | -2,35 | 1,88E-02 | 1,92E-01 |
| MNAT1 | 386,17 | 0,15 | 0,06 | 2,36 | 1,85E-02 | 1,92E-01 |
| CNIH1 | 688,47 | 0,18 | 0,08 | 2,35 | 1,89E-02 | 1,92E-01 |
| ALPK3 | 104,35 | -0,37 | 0,16 | -2,35 | 1,88E-02 | 1,92E-01 |
| NUP54 | 805,73 | 0,16 | 0,07 | 2,35 | 1,87E-02 | 1,92E-01 |
| SNRNP40 | 1002,64 | 0,09 | 0,04 | 2,35 | 1,86E-02 | 1,92E-01 |
| CWC15 | 559,76 | 0,16 | 0,07 | 2,35 | 1,86E-02 | 1,92E-01 |
| DYNC1I2 | 985,25 | 0,12 | 0,05 | 2,35 | 1,89E-02 | 1,92E-01 |
| PRX | 95,62 | -0,25 | 0,11 | -2,35 | 1,89E-02 | 1,92E-01 |
| COPZ1 | 918,00 | -0,09 | 0,04 | -2,35 | 1,90E-02 | 1,92E-01 |
| CDCA2 | 33,12 | -0,40 | 0,17 | -2,34 | 1,90E-02 | 1,92E-01 |
| FAM136A | 383,17 | 0,10 | 0,04 | 2,35 | 1,90E-02 | 1,92E-01 |
| HLF | 55,15 | -0,38 | 0,16 | -2,35 | 1,90E-02 | 1,92E-01 |
| SCD5 | 79,11 | -0,32 | 0,14 | -2,34 | 1,91E-02 | 1,93E-01 |
| PINLYP | 26,51 | -0,44 | 0,19 | -2,34 | 1,91E-02 | 1,93E-01 |
| LRRC75A | 234,68 | -0,21 | 0,09 | -2,34 | 1,91E-02 | 1,93E-01 |
| ZC3H15 | 976,02 | 0,22 | 0,09 | 2,34 | 1,91E-02 | 1,93E-01 |
| GLRX2 | 600,48 | 0,13 | 0,05 | 2,34 | 1,91E-02 | 1,93E-01 |
| KNDC1 | 77,73 | -0,56 | 0,24 | -2,34 | 1,92E-02 | 1,93E-01 |
| GPATCH2L | 5795,61 | 0,09 | 0,04 | 2,34 | 1,92E-02 | 1,93E-01 |
| C2orf49 | 927,64 | 0,13 | 0,06 | 2,33 | 1,96E-02 | 1,93E-01 |
| CCDC65 | 136,15 | 0,22 | 0,09 | 2,34 | 1,92E-02 | 1,93E-01 |
| STEEP1 | 426,97 | 0,11 | 0,05 | 2,33 | 1,97E-02 | 1,93E-01 |
| ALOX12 | 51,57 | -0,44 | 0,19 | -2,34 | 1,95E-02 | 1,93E-01 |
| QTRT2 | 1090,08 | 0,07 | 0,03 | 2,33 | 1,97E-02 | 1,93E-01 |
| RFFL | 2547,98 | -0,12 | 0,05 | -2,34 | 1,95E-02 | 1,93E-01 |
| PBX1 | 373,33 | -0,30 | 0,13 | -2,33 | 1,97E-02 | 1,93E-01 |
| FAM83E | 37,81 | -0,31 | 0,13 | -2,34 | 1,94E-02 | 1,93E-01 |
| LYSET | 154,89 | 0,15 | 0,06 | 2,33 | 1,96E-02 | 1,93E-01 |
| LSM8 | 1371,38 | 0,16 | 0,07 | 2,34 | 1,95E-02 | 1,93E-01 |
| RBBP4 | 2826,32 | 0,08 | 0,03 | 2,34 | 1,93E-02 | 1,93E-01 |

| | | | | | | |
|---|---|---|---|---|---|---|
| STRIP2 | 56,22 | -0,37 | 0,16 | -2,33 | 1,97E-02 | 1,93E-01 |
| LANCL2 | 297,30 | 0,08 | 0,03 | 2,33 | 1,96E-02 | 1,93E-01 |
| TMPRSS13 | 53,33 | -0,35 | 0,15 | -2,34 | 1,94E-02 | 1,93E-01 |
| TEX10 | 877,23 | 0,10 | 0,04 | 2,34 | 1,95E-02 | 1,93E-01 |
| PMS1 | 771,31 | 0,17 | 0,07 | 2,34 | 1,93E-02 | 1,93E-01 |
| NKAPD1 | 749,71 | 0,15 | 0,07 | 2,34 | 1,93E-02 | 1,93E-01 |
| B3GNT9 | 107,33 | -0,17 | 0,07 | -2,33 | 1,97E-02 | 1,93E-01 |
| ATP9B | 2913,06 | 0,10 | 0,04 | 2,34 | 1,93E-02 | 1,93E-01 |
| GABRR2 | 74,13 | -0,24 | 0,10 | -2,34 | 1,95E-02 | 1,93E-01 |
| TESMIN | 190,94 | -0,17 | 0,07 | -2,34 | 1,92E-02 | 1,93E-01 |
| MGLL | 202,01 | -0,30 | 0,13 | -2,33 | 1,97E-02 | 1,93E-01 |
| URI1 | 1681,73 | 0,14 | 0,06 | 2,33 | 1,97E-02 | 1,93E-01 |
| ATP8A1 | 5030,19 | 0,17 | 0,07 | 2,33 | 1,97E-02 | 1,93E-01 |
| DMPK | 115,05 | -0,35 | 0,15 | -2,34 | 1,93E-02 | 1,93E-01 |
| SLC66A2 | 1510,93 | -0,18 | 0,08 | -2,33 | 1,97E-02 | 1,93E-01 |
| EYA3 | 1028,21 | -0,09 | 0,04 | -2,33 | 1,96E-02 | 1,93E-01 |
| RPAP2 | 1246,00 | 0,13 | 0,06 | 2,33 | 1,96E-02 | 1,93E-01 |
| CAAP1 | 649,21 | 0,13 | 0,05 | 2,33 | 1,96E-02 | 1,93E-01 |
| IFT25 | 526,28 | 0,12 | 0,05 | 2,33 | 1,97E-02 | 1,93E-01 |
| DRAXIN | 132,43 | -0,23 | 0,10 | -2,34 | 1,94E-02 | 1,93E-01 |
| PTCD3 | 2509,32 | 0,11 | 0,05 | 2,34 | 1,94E-02 | 1,93E-01 |
| COPS8 | 759,57 | 0,15 | 0,06 | 2,34 | 1,93E-02 | 1,93E-01 |
| ZSWIM7 | 342,34 | 0,08 | 0,03 | 2,33 | 1,97E-02 | 1,93E-01 |
| TGFB3 | 56,60 | -0,34 | 0,15 | -2,33 | 1,98E-02 | 1,93E-01 |
| NR1D2 | 1605,85 | 0,16 | 0,07 | 2,33 | 1,98E-02 | 1,93E-01 |
| NAA20 | 467,45 | 0,10 | 0,04 | 2,33 | 1,98E-02 | 1,93E-01 |
| CDC42EP4 | 117,51 | -0,28 | 0,12 | -2,33 | 1,99E-02 | 1,94E-01 |
| DFFA | 1085,89 | 0,08 | 0,04 | 2,33 | 1,99E-02 | 1,94E-01 |
| OSGEPL1 | 303,16 | 0,16 | 0,07 | 2,33 | 1,99E-02 | 1,94E-01 |
| UBE2S | 345,24 | -0,14 | 0,06 | -2,33 | 2,00E-02 | 1,94E-01 |
| UQCRB | 800,09 | 0,31 | 0,13 | 2,33 | 2,00E-02 | 1,94E-01 |
| PTK7 | 68,88 | -0,36 | 0,15 | -2,33 | 2,00E-02 | 1,94E-01 |
| KLF12 | 3861,46 | 0,17 | 0,07 | 2,33 | 2,00E-02 | 1,94E-01 |
| TMEM45B | 212,92 | -0,24 | 0,10 | -2,33 | 2,00E-02 | 1,94E-01 |
| CCDC59 | 636,06 | 0,23 | 0,10 | 2,32 | 2,01E-02 | 1,94E-01 |
| CTDSPL2 | 2359,70 | 0,15 | 0,07 | 2,32 | 2,01E-02 | 1,95E-01 |
| MRPL19 | 634,26 | 0,11 | 0,05 | 2,32 | 2,02E-02 | 1,95E-01 |
| SLC30A9 | 1723,47 | 0,15 | 0,06 | 2,32 | 2,02E-02 | 1,95E-01 |
| RPL11 | 5176,72 | 0,30 | 0,13 | 2,32 | 2,02E-02 | 1,95E-01 |
| RNF175 | 82,73 | -0,28 | 0,12 | -2,32 | 2,05E-02 | 1,95E-01 |
| SAV1 | 389,39 | 0,12 | 0,05 | 2,32 | 2,03E-02 | 1,95E-01 |
| VIRMA | 1968,62 | 0,08 | 0,04 | 2,32 | 2,04E-02 | 1,95E-01 |
| ADGRG5 | 458,89 | -0,25 | 0,11 | -2,32 | 2,03E-02 | 1,95E-01 |
| INTS14 | 668,34 | 0,08 | 0,03 | 2,32 | 2,03E-02 | 1,95E-01 |
| SNW1 | 1402,94 | 0,12 | 0,05 | 2,32 | 2,03E-02 | 1,95E-01 |
| MLH1 | 959,30 | 0,08 | 0,04 | 2,32 | 2,03E-02 | 1,95E-01 |

| | | | | | | |
|---|---|---|---|---|---|---|
| RTKN | 35,14 | -0,46 | 0,20 | -2,32 | 2,04E-02 | 1,95E-01 |
| CABLES2 | 66,42 | -0,29 | 0,13 | -2,32 | 2,05E-02 | 1,95E-01 |
| POP7 | 191,45 | -0,13 | 0,06 | -2,32 | 2,05E-02 | 1,95E-01 |
| STT3B | 5242,29 | 0,16 | 0,07 | 2,32 | 2,04E-02 | 1,95E-01 |
| ADSS1 | 43,89 | -0,37 | 0,16 | -2,32 | 2,03E-02 | 1,95E-01 |
| IRAK1BP1 | 150,81 | 0,19 | 0,08 | 2,32 | 2,04E-02 | 1,95E-01 |
| MRPL38 | 37,30 | -0,29 | 0,13 | -2,32 | 2,04E-02 | 1,95E-01 |
| CEP112 | 79,43 | -0,41 | 0,18 | -2,32 | 2,03E-02 | 1,95E-01 |
| STARD4 | 624,23 | 0,21 | 0,09 | 2,32 | 2,05E-02 | 1,95E-01 |
| OBI1 | 530,40 | 0,23 | 0,10 | 2,32 | 2,02E-02 | 1,95E-01 |
| DIAPH2 | 1532,28 | 0,13 | 0,06 | 2,32 | 2,05E-02 | 1,95E-01 |
| WDCP | 306,08 | 0,10 | 0,04 | 2,31 | 2,06E-02 | 1,95E-01 |
| SPX | 90,32 | -0,30 | 0,13 | -2,32 | 2,06E-02 | 1,95E-01 |
| LRP11 | 29,29 | -0,40 | 0,17 | -2,32 | 2,06E-02 | 1,95E-01 |
| P3H4 | 77,89 | -0,24 | 0,10 | -2,31 | 2,07E-02 | 1,95E-01 |
| FAF1 | 1125,00 | 0,08 | 0,03 | 2,31 | 2,07E-02 | 1,95E-01 |
| EIF3J | 986,95 | 0,14 | 0,06 | 2,31 | 2,07E-02 | 1,95E-01 |
| ZNF304 | 470,69 | 0,11 | 0,05 | 2,31 | 2,08E-02 | 1,95E-01 |
| TAS2R20 | 179,94 | 0,22 | 0,10 | 2,31 | 2,07E-02 | 1,95E-01 |
| LARP4 | 1083,66 | 0,14 | 0,06 | 2,31 | 2,08E-02 | 1,95E-01 |
| GBA2 | 1228,29 | -0,13 | 0,06 | -2,31 | 2,07E-02 | 1,95E-01 |
| MCCC2 | 675,28 | 0,08 | 0,03 | 2,31 | 2,08E-02 | 1,95E-01 |
| LYAR | 654,13 | 0,15 | 0,06 | 2,31 | 2,07E-02 | 1,95E-01 |
| RILPL1 | 134,93 | -0,23 | 0,10 | -2,31 | 2,08E-02 | 1,95E-01 |
| RGS9 | 130,91 | -0,31 | 0,13 | -2,32 | 2,06E-02 | 1,95E-01 |
| PRKRA | 605,77 | 0,10 | 0,04 | 2,31 | 2,07E-02 | 1,95E-01 |
| SLIRP | 212,87 | 0,13 | 0,06 | 2,31 | 2,08E-02 | 1,95E-01 |
| TSN | 1302,83 | 0,13 | 0,06 | 2,31 | 2,07E-02 | 1,95E-01 |
| PIBF1 | 1039,72 | 0,19 | 0,08 | 2,31 | 2,07E-02 | 1,95E-01 |
| TMEM201 | 54,87 | -0,43 | 0,19 | -2,31 | 2,08E-02 | 1,95E-01 |
| ERAL1 | 181,71 | -0,10 | 0,04 | -2,31 | 2,09E-02 | 1,96E-01 |
| ZNF17 | 374,26 | 0,17 | 0,07 | 2,31 | 2,09E-02 | 1,96E-01 |
| ZNF83 | 1016,88 | 0,18 | 0,08 | 2,31 | 2,11E-02 | 1,96E-01 |
| DEDD | 281,38 | -0,13 | 0,06 | -2,31 | 2,09E-02 | 1,96E-01 |
| DNAJC24 | 742,63 | 0,14 | 0,06 | 2,31 | 2,10E-02 | 1,96E-01 |
| MMACHC | 59,42 | -0,20 | 0,09 | -2,31 | 2,09E-02 | 1,96E-01 |
| PAQR9 | 37,60 | -0,50 | 0,21 | -2,31 | 2,10E-02 | 1,96E-01 |
| ZFP92 | 56,13 | -0,28 | 0,12 | -2,31 | 2,11E-02 | 1,96E-01 |
| ATOH8 | 54,43 | -0,52 | 0,23 | -2,31 | 2,10E-02 | 1,96E-01 |
| AP4E1 | 1238,06 | 0,11 | 0,05 | 2,31 | 2,10E-02 | 1,96E-01 |
| TSPAN17 | 541,46 | -0,11 | 0,05 | -2,31 | 2,10E-02 | 1,96E-01 |
| MSH2 | 732,16 | 0,14 | 0,06 | 2,31 | 2,10E-02 | 1,96E-01 |
| CEP135 | 1068,79 | 0,12 | 0,05 | 2,31 | 2,11E-02 | 1,96E-01 |
| TPP2 | 4409,02 | 0,12 | 0,05 | 2,31 | 2,11E-02 | 1,96E-01 |
| ATM | 26475,10 | 0,13 | 0,06 | 2,31 | 2,11E-02 | 1,96E-01 |
| RFK | 424,91 | 0,15 | 0,07 | 2,31 | 2,12E-02 | 1,96E-01 |

| | | | | | | |
|---|---|---|---|---|---|---|
| NEMP1 | 868,22 | 0,11 | 0,05 | 2,30 | 2,12E-02 | 1,96E-01 |
| GIPR | 97,77 | -0,27 | 0,12 | -2,30 | 2,12E-02 | 1,96E-01 |
| NMD3 | 785,54 | 0,22 | 0,10 | 2,30 | 2,12E-02 | 1,96E-01 |
| HNRNPR | 4501,78 | 0,14 | 0,06 | 2,30 | 2,12E-02 | 1,96E-01 |
| ZNF84 | 955,58 | 0,16 | 0,07 | 2,30 | 2,13E-02 | 1,96E-01 |
| UPF3B | 391,20 | 0,09 | 0,04 | 2,30 | 2,13E-02 | 1,96E-01 |
| UCK1 | 393,23 | -0,08 | 0,03 | -2,30 | 2,14E-02 | 1,97E-01 |
| PTGER3 | 38,38 | -0,47 | 0,21 | -2,30 | 2,14E-02 | 1,97E-01 |
| RGPD8 | 277,96 | 0,22 | 0,09 | 2,30 | 2,14E-02 | 1,97E-01 |
| PRKD3 | 1730,99 | 0,11 | 0,05 | 2,30 | 2,14E-02 | 1,97E-01 |
| CD36 | 5057,83 | 0,22 | 0,09 | 2,30 | 2,14E-02 | 1,97E-01 |
| WDR86 | 78,42 | -0,32 | 0,14 | -2,30 | 2,14E-02 | 1,97E-01 |
| GOLGA6L10 | 48,49 | -0,45 | 0,20 | -2,30 | 2,15E-02 | 1,97E-01 |
| MFSD4A | 35,66 | -0,38 | 0,16 | -2,30 | 2,15E-02 | 1,97E-01 |
| KLHL25 | 109,74 | -0,27 | 0,12 | -2,30 | 2,15E-02 | 1,97E-01 |
| APBA1 | 71,57 | -0,42 | 0,18 | -2,30 | 2,15E-02 | 1,97E-01 |
| CACNB3 | 77,50 | -0,30 | 0,13 | -2,30 | 2,17E-02 | 1,97E-01 |
| GLUD1 | 3090,33 | 0,10 | 0,04 | 2,30 | 2,16E-02 | 1,97E-01 |
| CLOCK | 955,09 | 0,12 | 0,05 | 2,30 | 2,16E-02 | 1,97E-01 |
| DAGLA | 54,08 | -0,42 | 0,18 | -2,30 | 2,16E-02 | 1,97E-01 |
| DPH5 | 423,52 | 0,15 | 0,07 | 2,30 | 2,16E-02 | 1,97E-01 |
| ZSCAN30 | 651,74 | 0,10 | 0,04 | 2,30 | 2,17E-02 | 1,98E-01 |
| PDIK1L | 404,73 | 0,22 | 0,10 | 2,29 | 2,18E-02 | 1,98E-01 |
| NUP43 | 1353,36 | 0,10 | 0,04 | 2,29 | 2,18E-02 | 1,98E-01 |
| EDRF1 | 1805,77 | 0,08 | 0,03 | 2,30 | 2,17E-02 | 1,98E-01 |
| EAPP | 938,24 | 0,15 | 0,07 | 2,30 | 2,17E-02 | 1,98E-01 |
| APCDD1 | 43,59 | -0,49 | 0,21 | -2,29 | 2,19E-02 | 1,98E-01 |
| ZNF383 | 772,29 | 0,09 | 0,04 | 2,29 | 2,21E-02 | 1,98E-01 |
| RHAG | 141,14 | -0,32 | 0,14 | -2,29 | 2,20E-02 | 1,98E-01 |
| SNRPG | 403,92 | 0,21 | 0,09 | 2,29 | 2,20E-02 | 1,98E-01 |
| ZNF91 | 1732,66 | 0,19 | 0,08 | 2,29 | 2,19E-02 | 1,98E-01 |
| CSNK2A1 | 3321,71 | 0,06 | 0,03 | 2,29 | 2,18E-02 | 1,98E-01 |
| NAA30 | 1118,78 | 0,14 | 0,06 | 2,29 | 2,20E-02 | 1,98E-01 |
| ZNF140 | 641,52 | 0,10 | 0,04 | 2,29 | 2,18E-02 | 1,98E-01 |
| ACADM | 778,09 | 0,16 | 0,07 | 2,29 | 2,19E-02 | 1,98E-01 |
| ASF1A | 803,34 | 0,18 | 0,08 | 2,29 | 2,19E-02 | 1,98E-01 |
| CTIF | 151,48 | -0,32 | 0,14 | -2,29 | 2,19E-02 | 1,98E-01 |
| SLC44A2 | 3212,73 | -0,21 | 0,09 | -2,29 | 2,20E-02 | 1,98E-01 |
| TBC1D24 | 117,23 | -0,30 | 0,13 | -2,29 | 2,21E-02 | 1,98E-01 |
| CMTM5 | 182,63 | -0,32 | 0,14 | -2,29 | 2,19E-02 | 1,98E-01 |
| PXDNL | 54,76 | -0,26 | 0,11 | -2,29 | 2,20E-02 | 1,98E-01 |
| DMXL1 | 4197,95 | 0,13 | 0,06 | 2,29 | 2,21E-02 | 1,98E-01 |
| CELSR3 | 120,58 | -0,37 | 0,16 | -2,29 | 2,21E-02 | 1,98E-01 |
| EPM2AIP1 | 2199,91 | 0,16 | 0,07 | 2,29 | 2,18E-02 | 1,98E-01 |
| ATP9A | 305,65 | -0,23 | 0,10 | -2,29 | 2,21E-02 | 1,98E-01 |
| DAB1 | 59,24 | -0,39 | 0,17 | -2,29 | 2,19E-02 | 1,98E-01 |

| | | | | | | |
|---|---|---|---|---|---|---|
| INPP5K | 1303,48 | -0,15 | 0,06 | -2,28 | 2,24E-02 | 1,98E-01 |
| ZNF548 | 835,98 | 0,10 | 0,04 | 2,28 | 2,24E-02 | 1,98E-01 |
| TMEM63C | 75,05 | -0,42 | 0,18 | -2,29 | 2,22E-02 | 1,98E-01 |
| CEACAM1 | 498,82 | -0,21 | 0,09 | -2,29 | 2,23E-02 | 1,98E-01 |
| SLC9A7 | 865,25 | 0,17 | 0,07 | 2,28 | 2,24E-02 | 1,98E-01 |
| NOL8 | 1437,59 | 0,11 | 0,05 | 2,28 | 2,23E-02 | 1,98E-01 |
| RGS19 | 1685,40 | -0,15 | 0,07 | -2,29 | 2,22E-02 | 1,98E-01 |
| CROT | 425,29 | 0,14 | 0,06 | 2,29 | 2,22E-02 | 1,98E-01 |
| GRIN3A | 40,25 | -0,45 | 0,20 | -2,28 | 2,24E-02 | 1,98E-01 |
| EXO1 | 41,54 | -0,37 | 0,16 | -2,29 | 2,22E-02 | 1,98E-01 |
| TNFAIP1 | 384,55 | -0,09 | 0,04 | -2,29 | 2,23E-02 | 1,98E-01 |
| ALG10B | 662,71 | 0,20 | 0,09 | 2,28 | 2,23E-02 | 1,98E-01 |
| SMPDL3B | 28,52 | -0,32 | 0,14 | -2,29 | 2,22E-02 | 1,98E-01 |
| RHD | 104,02 | -0,58 | 0,25 | -2,29 | 2,22E-02 | 1,98E-01 |
| MT-ND6 | 224,76 | -0,55 | 0,24 | -2,28 | 2,24E-02 | 1,98E-01 |
| FBXO10 | 32,25 | -0,35 | 0,15 | -2,28 | 2,24E-02 | 1,98E-01 |
| ZNF155 | 298,00 | 0,17 | 0,07 | 2,28 | 2,24E-02 | 1,98E-01 |
| TRAF5 | 2255,61 | 0,12 | 0,05 | 2,28 | 2,25E-02 | 1,98E-01 |
| TBC1D16 | 109,34 | -0,34 | 0,15 | -2,28 | 2,25E-02 | 1,99E-01 |
| ZNF615 | 437,31 | 0,17 | 0,07 | 2,28 | 2,25E-02 | 1,99E-01 |
| SEC22A | 507,21 | 0,09 | 0,04 | 2,28 | 2,25E-02 | 1,99E-01 |
| FKTN | 674,21 | 0,12 | 0,05 | 2,28 | 2,27E-02 | 1,99E-01 |
| HCAR3 | 737,33 | -0,22 | 0,10 | -2,28 | 2,26E-02 | 1,99E-01 |
| EXOC5 | 2334,71 | 0,17 | 0,07 | 2,28 | 2,28E-02 | 1,99E-01 |
| CYREN | 852,84 | 0,05 | 0,02 | 2,28 | 2,26E-02 | 1,99E-01 |
| TMEM161A | 116,00 | -0,17 | 0,07 | -2,28 | 2,27E-02 | 1,99E-01 |
| ARL14EP | 1288,80 | 0,20 | 0,09 | 2,28 | 2,27E-02 | 1,99E-01 |
| B4GALNT4 | 37,70 | -0,53 | 0,23 | -2,28 | 2,28E-02 | 1,99E-01 |
| ZNF830 | 692,22 | 0,16 | 0,07 | 2,28 | 2,27E-02 | 1,99E-01 |
| PAXBP1 | 1944,32 | 0,14 | 0,06 | 2,28 | 2,26E-02 | 1,99E-01 |
| SLC4A1 | 14251,01 | -0,37 | 0,16 | -2,28 | 2,27E-02 | 1,99E-01 |
| TGFA | 772,04 | -0,17 | 0,08 | -2,28 | 2,27E-02 | 1,99E-01 |
| N4BP2L1 | 1287,77 | 0,09 | 0,04 | 2,28 | 2,28E-02 | 1,99E-01 |
| FXYD6 | 31,16 | -0,45 | 0,20 | -2,28 | 2,27E-02 | 1,99E-01 |
| CWF19L2 | 1519,52 | 0,17 | 0,07 | 2,28 | 2,27E-02 | 1,99E-01 |
| ZNF670 | 43,07 | 0,23 | 0,10 | 2,28 | 2,26E-02 | 1,99E-01 |
| PPP1R2 | 3247,14 | 0,13 | 0,06 | 2,28 | 2,28E-02 | 1,99E-01 |
| GCC2 | 3133,66 | 0,17 | 0,07 | 2,28 | 2,28E-02 | 1,99E-01 |
| ARHGAP12 | 1404,09 | 0,16 | 0,07 | 2,28 | 2,29E-02 | 1,99E-01 |
| DYNLT2 | 94,81 | 0,22 | 0,09 | 2,28 | 2,29E-02 | 1,99E-01 |
| SLC7A1 | 521,96 | -0,16 | 0,07 | -2,28 | 2,29E-02 | 1,99E-01 |
| CRYZL1 | 817,89 | 0,09 | 0,04 | 2,27 | 2,29E-02 | 1,99E-01 |
| STX1A | 40,72 | -0,36 | 0,16 | -2,27 | 2,32E-02 | 1,99E-01 |
| RIPK3 | 226,50 | -0,18 | 0,08 | -2,27 | 2,32E-02 | 1,99E-01 |
| LARS1 | 2558,61 | 0,11 | 0,05 | 2,27 | 2,32E-02 | 1,99E-01 |
| ENSG000002 | 128,55 | -0,51 | 0,23 | -2,27 | 2,32E-02 | 1,99E-01 |

| | | | | | | |
|---|---|---|---|---|---|---|
| POMT2 | 136,72 | -0,24 | 0,11 | -2,27 | 2,33E-02 | 1,99E-01 |
| PRPF39 | 965,02 | 0,17 | 0,07 | 2,27 | 2,32E-02 | 1,99E-01 |
| MEX3C | 2056,89 | 0,15 | 0,06 | 2,27 | 2,32E-02 | 1,99E-01 |
| TDG | 961,02 | 0,10 | 0,04 | 2,27 | 2,31E-02 | 1,99E-01 |
| ATR | 3245,03 | 0,13 | 0,06 | 2,27 | 2,31E-02 | 1,99E-01 |
| ZFYVE21 | 417,68 | -0,12 | 0,05 | -2,27 | 2,33E-02 | 1,99E-01 |
| THUMPD2 | 381,06 | 0,14 | 0,06 | 2,27 | 2,31E-02 | 1,99E-01 |
| RPS3A | 6103,45 | 0,45 | 0,20 | 2,27 | 2,31E-02 | 1,99E-01 |
| CACNA1F | 27,81 | -0,44 | 0,19 | -2,27 | 2,33E-02 | 1,99E-01 |
| ZCCHC4 | 458,77 | 0,10 | 0,05 | 2,27 | 2,32E-02 | 1,99E-01 |
| TOGARAM1 | 864,23 | 0,09 | 0,04 | 2,27 | 2,30E-02 | 1,99E-01 |
| DDR2 | 72,47 | -0,43 | 0,19 | -2,27 | 2,32E-02 | 1,99E-01 |
| SLC25A34 | 31,15 | -0,33 | 0,14 | -2,27 | 2,30E-02 | 1,99E-01 |
| PWP1 | 798,89 | 0,12 | 0,05 | 2,27 | 2,30E-02 | 1,99E-01 |
| INPP5A | 943,02 | -0,13 | 0,06 | -2,27 | 2,33E-02 | 1,99E-01 |
| MBNL2 | 1347,85 | 0,15 | 0,07 | 2,27 | 2,33E-02 | 1,99E-01 |
| TRMT10C | 328,38 | 0,25 | 0,11 | 2,27 | 2,31E-02 | 1,99E-01 |
| UNC119B | 668,26 | -0,10 | 0,04 | -2,27 | 2,32E-02 | 1,99E-01 |
| RMND1 | 362,63 | 0,10 | 0,04 | 2,27 | 2,33E-02 | 2,00E-01 |
| JOSD2 | 133,35 | -0,14 | 0,06 | -2,26 | 2,36E-02 | 2,00E-01 |
| ZFX | 3314,40 | 0,10 | 0,04 | 2,26 | 2,37E-02 | 2,00E-01 |
| ZSCAN32 | 732,09 | 0,06 | 0,03 | 2,26 | 2,36E-02 | 2,00E-01 |
| PTPN11 | 2129,01 | 0,08 | 0,04 | 2,27 | 2,35E-02 | 2,00E-01 |
| SLF2 | 1928,12 | 0,09 | 0,04 | 2,26 | 2,37E-02 | 2,00E-01 |
| ZBTB46 | 56,51 | -0,34 | 0,15 | -2,27 | 2,35E-02 | 2,00E-01 |
| SLC22A23 | 258,58 | -0,27 | 0,12 | -2,27 | 2,34E-02 | 2,00E-01 |
| GTF2H3 | 489,79 | 0,11 | 0,05 | 2,26 | 2,37E-02 | 2,00E-01 |
| UBE2V2 | 584,92 | 0,13 | 0,06 | 2,26 | 2,37E-02 | 2,00E-01 |
| TMEM263 | 638,75 | 0,16 | 0,07 | 2,27 | 2,35E-02 | 2,00E-01 |
| IGSF11 | 37,01 | -0,50 | 0,22 | -2,27 | 2,34E-02 | 2,00E-01 |
| ATG9B | 76,61 | -0,33 | 0,15 | -2,26 | 2,36E-02 | 2,00E-01 |
| ASAP2 | 171,84 | -0,27 | 0,12 | -2,26 | 2,37E-02 | 2,00E-01 |
| DNAJC7 | 2094,77 | 0,05 | 0,02 | 2,27 | 2,34E-02 | 2,00E-01 |
| SNORC | 52,17 | -0,31 | 0,14 | -2,26 | 2,36E-02 | 2,00E-01 |
| EXOC2 | 1893,45 | 0,10 | 0,04 | 2,27 | 2,34E-02 | 2,00E-01 |
| EIF3E | 3787,21 | 0,25 | 0,11 | 2,27 | 2,35E-02 | 2,00E-01 |
| UTP20 | 983,56 | 0,10 | 0,04 | 2,26 | 2,37E-02 | 2,00E-01 |
| TBCEL-TECTA | 27,40 | -0,28 | 0,12 | -2,27 | 2,34E-02 | 2,00E-01 |
| EIF1AX | 1060,63 | 0,20 | 0,09 | 2,27 | 2,35E-02 | 2,00E-01 |
| MIX23 | 130,27 | 0,14 | 0,06 | 2,26 | 2,37E-02 | 2,00E-01 |
| MOAP1 | 941,93 | 0,10 | 0,04 | 2,26 | 2,37E-02 | 2,00E-01 |
| CCNB1IP1 | 363,27 | 0,13 | 0,06 | 2,25 | 2,44E-02 | 2,00E-01 |
| RBMX | 3365,13 | 0,10 | 0,05 | 2,25 | 2,42E-02 | 2,00E-01 |
| PTCD1 | 102,82 | -0,14 | 0,06 | -2,25 | 2,43E-02 | 2,00E-01 |
| ABCB9 | 96,44 | -0,34 | 0,15 | -2,25 | 2,44E-02 | 2,00E-01 |
| NOTCH4 | 79,98 | -0,35 | 0,16 | -2,25 | 2,41E-02 | 2,00E-01 |

| | | | | | | |
|---|---|---|---|---|---|---|
| TMEM44 | 65,54 | -0,34 | 0,15 | -2,25 | 2,44E-02 | 2,00E-01 |
| ANKRD46 | 605,08 | 0,13 | 0,06 | 2,26 | 2,40E-02 | 2,00E-01 |
| PSMD12 | 1375,21 | 0,09 | 0,04 | 2,26 | 2,39E-02 | 2,00E-01 |
| CRTAP | 1735,42 | 0,09 | 0,04 | 2,26 | 2,41E-02 | 2,00E-01 |
| R3HDM4 | 10144,09 | -0,30 | 0,13 | -2,26 | 2,39E-02 | 2,00E-01 |
| SLC2A8 | 69,34 | -0,29 | 0,13 | -2,26 | 2,40E-02 | 2,00E-01 |
| TMCC2 | 500,45 | -0,35 | 0,15 | -2,26 | 2,40E-02 | 2,00E-01 |
| CYP20A1 | 873,51 | 0,07 | 0,03 | 2,25 | 2,43E-02 | 2,00E-01 |
| RFPL2 | 38,43 | -0,36 | 0,16 | -2,25 | 2,43E-02 | 2,00E-01 |
| RNASEH2B | 4165,47 | 0,10 | 0,05 | 2,26 | 2,40E-02 | 2,00E-01 |
| MRPS22 | 552,05 | 0,13 | 0,06 | 2,25 | 2,43E-02 | 2,00E-01 |
| ZNF780B | 2270,95 | 0,17 | 0,08 | 2,26 | 2,40E-02 | 2,00E-01 |
| CEP350 | 6808,95 | 0,10 | 0,04 | 2,25 | 2,44E-02 | 2,00E-01 |
| CCT2 | 1195,54 | 0,19 | 0,09 | 2,25 | 2,43E-02 | 2,00E-01 |
| AP1S2 | 3187,40 | 0,25 | 0,11 | 2,25 | 2,43E-02 | 2,00E-01 |
| RNF215 | 152,18 | -0,21 | 0,09 | -2,26 | 2,41E-02 | 2,00E-01 |
| DCAF4L1 | 78,99 | -0,17 | 0,08 | -2,25 | 2,43E-02 | 2,00E-01 |
| ODR4 | 801,52 | 0,16 | 0,07 | 2,25 | 2,42E-02 | 2,00E-01 |
| BOD1 | 479,05 | 0,10 | 0,05 | 2,25 | 2,43E-02 | 2,00E-01 |
| CARNS1 | 90,40 | -0,35 | 0,16 | -2,26 | 2,41E-02 | 2,00E-01 |
| HSF2 | 385,16 | 0,16 | 0,07 | 2,26 | 2,39E-02 | 2,00E-01 |
| TMPO | 5685,10 | 0,13 | 0,06 | 2,26 | 2,39E-02 | 2,00E-01 |
| SAXO1 | 62,52 | -0,38 | 0,17 | -2,26 | 2,40E-02 | 2,00E-01 |
| HLA-DOA | 879,20 | 0,19 | 0,08 | 2,26 | 2,40E-02 | 2,00E-01 |
| FAM98B | 677,92 | 0,14 | 0,06 | 2,26 | 2,41E-02 | 2,00E-01 |
| ARMC10 | 757,50 | 0,09 | 0,04 | 2,25 | 2,43E-02 | 2,00E-01 |
| C17orf49 | 39,41 | -0,17 | 0,08 | -2,26 | 2,40E-02 | 2,00E-01 |
| GRM2 | 43,57 | -0,50 | 0,22 | -2,25 | 2,44E-02 | 2,00E-01 |
| HJURP | 40,22 | -0,40 | 0,18 | -2,26 | 2,41E-02 | 2,00E-01 |
| RPP30 | 544,34 | 0,11 | 0,05 | 2,26 | 2,40E-02 | 2,00E-01 |
| BCLAF1 | 8962,88 | 0,17 | 0,07 | 2,25 | 2,42E-02 | 2,00E-01 |
| RAD17 | 640,54 | 0,12 | 0,05 | 2,26 | 2,40E-02 | 2,00E-01 |
| RAB37 | 1717,04 | -0,13 | 0,06 | -2,25 | 2,44E-02 | 2,00E-01 |
| PPIL1 | 180,23 | 0,11 | 0,05 | 2,25 | 2,45E-02 | 2,01E-01 |
| EIF4A2 | 8642,71 | 0,19 | 0,08 | 2,25 | 2,45E-02 | 2,01E-01 |
| PDAP1 | 749,16 | -0,09 | 0,04 | -2,25 | 2,45E-02 | 2,01E-01 |
| ZNF256 | 165,29 | 0,14 | 0,06 | 2,25 | 2,45E-02 | 2,01E-01 |
| TPR | 7796,68 | 0,12 | 0,05 | 2,25 | 2,45E-02 | 2,01E-01 |
| ZNF254 | 617,29 | 0,20 | 0,09 | 2,25 | 2,46E-02 | 2,01E-01 |
| PUS3 | 170,32 | 0,14 | 0,06 | 2,25 | 2,45E-02 | 2,01E-01 |
| ZNF639 | 736,01 | 0,12 | 0,05 | 2,25 | 2,46E-02 | 2,01E-01 |
| CDKN1A | 252,27 | -0,26 | 0,12 | -2,25 | 2,46E-02 | 2,01E-01 |
| TAPT1 | 1824,98 | 0,11 | 0,05 | 2,25 | 2,47E-02 | 2,01E-01 |
| MTERF2 | 363,50 | 0,11 | 0,05 | 2,25 | 2,47E-02 | 2,01E-01 |
| TM7SF2 | 205,63 | -0,11 | 0,05 | -2,25 | 2,47E-02 | 2,01E-01 |
| WNT2B | 64,99 | -0,45 | 0,20 | -2,25 | 2,47E-02 | 2,01E-01 |

| | | | | | | |
|---|---|---|---|---|---|---|
| LRRC58 | 2024,85 | 0,14 | 0,06 | 2,25 | 2,47E-02 | 2,01E-01 |
| LSM6 | 1034,61 | 0,13 | 0,06 | 2,25 | 2,47E-02 | 2,01E-01 |
| ZBTB21 | 678,77 | 0,11 | 0,05 | 2,24 | 2,48E-02 | 2,01E-01 |
| RMDN1 | 1273,28 | 0,12 | 0,05 | 2,24 | 2,48E-02 | 2,01E-01 |
| GCM1 | 34,61 | -0,25 | 0,11 | -2,24 | 2,48E-02 | 2,01E-01 |
| PLPP7 | 35,87 | -0,36 | 0,16 | -2,24 | 2,48E-02 | 2,01E-01 |
| MRPS23 | 430,36 | 0,11 | 0,05 | 2,24 | 2,48E-02 | 2,01E-01 |
| GFPT2 | 39,29 | -0,61 | 0,27 | -2,24 | 2,49E-02 | 2,01E-01 |
| WNT5B | 49,13 | -0,41 | 0,18 | -2,24 | 2,49E-02 | 2,01E-01 |
| TLK1 | 2245,60 | 0,13 | 0,06 | 2,24 | 2,50E-02 | 2,01E-01 |
| SMS | 818,69 | 0,12 | 0,05 | 2,24 | 2,49E-02 | 2,01E-01 |
| HSPA14 | 339,49 | 0,18 | 0,08 | 2,24 | 2,50E-02 | 2,01E-01 |
| TRIM52 | 1641,88 | 0,10 | 0,04 | 2,24 | 2,49E-02 | 2,01E-01 |
| METTL21A | 484,53 | 0,07 | 0,03 | 2,24 | 2,50E-02 | 2,01E-01 |
| TTF1 | 707,07 | 0,10 | 0,04 | 2,24 | 2,49E-02 | 2,01E-01 |
| TESK2 | 412,83 | -0,11 | 0,05 | -2,24 | 2,50E-02 | 2,01E-01 |
| GRINA | 752,01 | -0,39 | 0,17 | -2,24 | 2,51E-02 | 2,02E-01 |
| ADSS2 | 2341,77 | 0,12 | 0,05 | 2,24 | 2,51E-02 | 2,02E-01 |
| ZNF141 | 1071,12 | 0,21 | 0,10 | 2,24 | 2,51E-02 | 2,02E-01 |
| MRPS28 | 239,29 | 0,13 | 0,06 | 2,24 | 2,52E-02 | 2,02E-01 |
| COQ7 | 414,64 | 0,08 | 0,03 | 2,24 | 2,52E-02 | 2,02E-01 |
| MAN1A2 | 3028,02 | 0,15 | 0,07 | 2,24 | 2,52E-02 | 2,02E-01 |
| DMWD | 204,03 | -0,23 | 0,10 | -2,24 | 2,53E-02 | 2,02E-01 |
| TTC34 | 97,41 | -0,37 | 0,17 | -2,24 | 2,53E-02 | 2,02E-01 |
| GSDME | 51,33 | -0,50 | 0,22 | -2,24 | 2,52E-02 | 2,02E-01 |
| OPN3 | 260,48 | 0,11 | 0,05 | 2,24 | 2,53E-02 | 2,02E-01 |
| CNOT6L | 6814,41 | 0,15 | 0,07 | 2,24 | 2,52E-02 | 2,02E-01 |
| PRPF40A | 4261,20 | 0,16 | 0,07 | 2,24 | 2,53E-02 | 2,02E-01 |
| ANKIB1 | 2088,38 | 0,10 | 0,04 | 2,24 | 2,53E-02 | 2,02E-01 |
| SNRPE | 327,57 | 0,20 | 0,09 | 2,24 | 2,54E-02 | 2,03E-01 |
| SGSM1 | 163,72 | -0,31 | 0,14 | -2,23 | 2,55E-02 | 2,03E-01 |
| MAGIX | 74,65 | -0,15 | 0,07 | -2,23 | 2,55E-02 | 2,03E-01 |
| RPS18 | 14337,47 | 0,29 | 0,13 | 2,23 | 2,55E-02 | 2,03E-01 |
| NTAN1 | 441,81 | -0,16 | 0,07 | -2,23 | 2,55E-02 | 2,03E-01 |
| CNOT7 | 2488,60 | 0,16 | 0,07 | 2,23 | 2,55E-02 | 2,03E-01 |
| ISOC2 | 53,73 | -0,26 | 0,12 | -2,23 | 2,56E-02 | 2,03E-01 |
| RASAL2 | 84,92 | -0,40 | 0,18 | -2,23 | 2,56E-02 | 2,03E-01 |
| GOPC | 1620,19 | 0,16 | 0,07 | 2,23 | 2,56E-02 | 2,03E-01 |
| ADAM28 | 1583,76 | 0,19 | 0,09 | 2,23 | 2,56E-02 | 2,03E-01 |
| DDHD2 | 1497,97 | 0,11 | 0,05 | 2,23 | 2,56E-02 | 2,03E-01 |
| MIS18BP1 | 2418,83 | 0,18 | 0,08 | 2,23 | 2,57E-02 | 2,03E-01 |
| SHLD3 | 130,38 | 0,24 | 0,11 | 2,23 | 2,57E-02 | 2,04E-01 |
| ZNF330 | 838,52 | 0,11 | 0,05 | 2,23 | 2,58E-02 | 2,04E-01 |
| SMIM5 | 627,51 | -0,28 | 0,12 | -2,23 | 2,58E-02 | 2,04E-01 |
| ELK4 | 4389,64 | 0,10 | 0,04 | 2,23 | 2,58E-02 | 2,04E-01 |
| SH2D6 | 34,48 | -0,36 | 0,16 | -2,23 | 2,58E-02 | 2,04E-01 |

| | | | | | | |
|---|---|---|---|---|---|---|
| BICRA | 146,69 | -0,41 | 0,18 | -2,23 | 2,58E-02 | 2,04E-01 |
| CHID1 | 485,80 | -0,14 | 0,06 | -2,23 | 2,58E-02 | 2,04E-01 |
| GUF1 | 1014,88 | 0,15 | 0,07 | 2,23 | 2,59E-02 | 2,04E-01 |
| OPA1 | 3268,87 | 0,14 | 0,06 | 2,23 | 2,59E-02 | 2,04E-01 |
| CERS3 | 59,07 | -0,34 | 0,15 | -2,23 | 2,60E-02 | 2,05E-01 |
| FBXO44 | 228,91 | -0,15 | 0,07 | -2,23 | 2,60E-02 | 2,05E-01 |
| ARHGAP5 | 1913,15 | 0,21 | 0,09 | 2,23 | 2,60E-02 | 2,05E-01 |
| STARD7 | 3010,29 | 0,10 | 0,04 | 2,23 | 2,61E-02 | 2,05E-01 |
| CKB | 68,25 | -0,27 | 0,12 | -2,23 | 2,61E-02 | 2,05E-01 |
| HPS1 | 1877,61 | -0,20 | 0,09 | -2,23 | 2,61E-02 | 2,05E-01 |
| TMEM87A | 1857,10 | 0,09 | 0,04 | 2,22 | 2,61E-02 | 2,05E-01 |
| CDC23 | 605,26 | 0,08 | 0,04 | 2,22 | 2,61E-02 | 2,05E-01 |
| SSNA1 | 490,88 | -0,11 | 0,05 | -2,22 | 2,61E-02 | 2,05E-01 |
| SRSF6 | 3938,15 | 0,11 | 0,05 | 2,22 | 2,62E-02 | 2,05E-01 |
| DESI2 | 1038,82 | 0,10 | 0,05 | 2,22 | 2,62E-02 | 2,05E-01 |
| KEL | 178,77 | -0,32 | 0,14 | -2,22 | 2,62E-02 | 2,05E-01 |
| ECSIT | 253,96 | -0,18 | 0,08 | -2,22 | 2,62E-02 | 2,05E-01 |
| RECQL | 1753,01 | 0,21 | 0,09 | 2,22 | 2,62E-02 | 2,05E-01 |
| ANKRD36C | 2114,68 | 0,15 | 0,07 | 2,22 | 2,63E-02 | 2,05E-01 |
| IL12RB2 | 160,97 | -0,23 | 0,10 | -2,22 | 2,63E-02 | 2,05E-01 |
| SEC14L4 | 89,58 | -0,34 | 0,15 | -2,22 | 2,63E-02 | 2,05E-01 |
| ANKRD6 | 292,16 | -0,12 | 0,05 | -2,22 | 2,66E-02 | 2,05E-01 |
| EMP3 | 2638,11 | 0,12 | 0,06 | 2,22 | 2,63E-02 | 2,05E-01 |
| SOCS4 | 1383,36 | 0,19 | 0,09 | 2,22 | 2,66E-02 | 2,05E-01 |
| BLOC1S5 | 465,03 | 0,11 | 0,05 | 2,22 | 2,65E-02 | 2,05E-01 |
| KLHL42 | 875,79 | 0,08 | 0,04 | 2,22 | 2,66E-02 | 2,05E-01 |
| RNF138 | 1192,08 | 0,19 | 0,09 | 2,22 | 2,66E-02 | 2,05E-01 |
| ADPGK | 5322,80 | 0,08 | 0,03 | 2,22 | 2,65E-02 | 2,05E-01 |
| SIAE | 183,87 | -0,17 | 0,08 | -2,22 | 2,64E-02 | 2,05E-01 |
| DRAM2 | 1728,62 | 0,14 | 0,07 | 2,22 | 2,65E-02 | 2,05E-01 |
| AFG2A | 1007,71 | 0,12 | 0,05 | 2,22 | 2,65E-02 | 2,05E-01 |
| RRAGB | 530,87 | 0,09 | 0,04 | 2,22 | 2,65E-02 | 2,05E-01 |
| MXD3 | 981,16 | -0,17 | 0,08 | -2,22 | 2,66E-02 | 2,05E-01 |
| MORC3 | 3980,64 | 0,20 | 0,09 | 2,22 | 2,65E-02 | 2,05E-01 |
| TMEM243 | 1260,96 | 0,12 | 0,05 | 2,22 | 2,65E-02 | 2,05E-01 |
| MOSMO | 797,96 | 0,13 | 0,06 | 2,22 | 2,66E-02 | 2,05E-01 |
| ZNF789 | 479,89 | 0,08 | 0,04 | 2,22 | 2,64E-02 | 2,05E-01 |
| CA2 | 477,24 | -0,25 | 0,11 | -2,22 | 2,66E-02 | 2,05E-01 |
| NAF1 | 441,14 | 0,11 | 0,05 | 2,22 | 2,64E-02 | 2,05E-01 |
| CHM | 851,09 | 0,15 | 0,07 | 2,22 | 2,66E-02 | 2,05E-01 |
| SRSF7 | 3727,14 | 0,14 | 0,06 | 2,22 | 2,64E-02 | 2,05E-01 |
| G2E3 | 1424,00 | 0,13 | 0,06 | 2,22 | 2,67E-02 | 2,06E-01 |
| CDC6 | 47,29 | -0,28 | 0,13 | -2,22 | 2,68E-02 | 2,06E-01 |
| EIF5 | 4089,07 | 0,11 | 0,05 | 2,21 | 2,68E-02 | 2,06E-01 |
| AP1AR | 610,90 | 0,19 | 0,09 | 2,21 | 2,68E-02 | 2,06E-01 |
| TMEM79 | 80,33 | -0,16 | 0,07 | -2,21 | 2,68E-02 | 2,06E-01 |

| | | | | | | |
|---|---|---|---|---|---|---|
| PDS5A | 3819,29 | 0,10 | 0,05 | 2,21 | 2,69E-02 | 2,06E-01 |
| PURA | 2863,13 | 0,15 | 0,07 | 2,21 | 2,69E-02 | 2,06E-01 |
| ZBTB33 | 1110,50 | 0,20 | 0,09 | 2,21 | 2,69E-02 | 2,06E-01 |
| LCN2 | 118,28 | -0,35 | 0,16 | -2,21 | 2,69E-02 | 2,06E-01 |
| CHAF1B | 75,34 | -0,25 | 0,11 | -2,21 | 2,69E-02 | 2,06E-01 |
| FIGNL2 | 63,61 | -0,34 | 0,15 | -2,21 | 2,70E-02 | 2,06E-01 |
| SIGLEC10 | 1973,78 | -0,20 | 0,09 | -2,21 | 2,71E-02 | 2,07E-01 |
| MSANTD1 | 83,63 | -0,37 | 0,17 | -2,21 | 2,71E-02 | 2,07E-01 |
| SAMD9 | 4576,84 | 0,35 | 0,16 | 2,21 | 2,71E-02 | 2,07E-01 |
| MBD2 | 2069,76 | 0,10 | 0,04 | 2,21 | 2,71E-02 | 2,07E-01 |
| SMIM38 | 37,99 | -0,39 | 0,18 | -2,21 | 2,72E-02 | 2,07E-01 |
| TLR7 | 483,49 | 0,17 | 0,08 | 2,21 | 2,72E-02 | 2,07E-01 |
| ETFRF1 | 309,98 | 0,18 | 0,08 | 2,21 | 2,72E-02 | 2,07E-01 |
| C17orf113 | 35,23 | -0,31 | 0,14 | -2,21 | 2,72E-02 | 2,07E-01 |
| HSPA6 | 3607,38 | -0,19 | 0,09 | -2,21 | 2,74E-02 | 2,07E-01 |
| TRIM13 | 1117,18 | 0,13 | 0,06 | 2,21 | 2,73E-02 | 2,07E-01 |
| DND1 | 48,50 | -0,20 | 0,09 | -2,21 | 2,74E-02 | 2,07E-01 |
| SMPD1 | 160,71 | -0,20 | 0,09 | -2,21 | 2,73E-02 | 2,07E-01 |
| CALD1 | 95,82 | -0,38 | 0,17 | -2,21 | 2,73E-02 | 2,07E-01 |
| CFAP54 | 128,92 | -0,29 | 0,13 | -2,21 | 2,73E-02 | 2,07E-01 |
| DOCK3 | 129,73 | -0,26 | 0,12 | -2,21 | 2,74E-02 | 2,07E-01 |
| SYNGR1 | 54,00 | -0,36 | 0,16 | -2,21 | 2,74E-02 | 2,07E-01 |
| PRUNE1 | 651,88 | -0,11 | 0,05 | -2,21 | 2,74E-02 | 2,08E-01 |
| TAF9B | 160,96 | 0,20 | 0,09 | 2,20 | 2,75E-02 | 2,08E-01 |
| COMMD2 | 885,59 | 0,14 | 0,06 | 2,20 | 2,75E-02 | 2,08E-01 |
| POLA1 | 608,69 | 0,11 | 0,05 | 2,20 | 2,75E-02 | 2,08E-01 |
| RSL24D1 | 1038,71 | 0,35 | 0,16 | 2,20 | 2,80E-02 | 2,08E-01 |
| SATB2 | 51,99 | -0,43 | 0,19 | -2,20 | 2,77E-02 | 2,08E-01 |
| SH3D21 | 110,83 | -0,23 | 0,11 | -2,20 | 2,80E-02 | 2,08E-01 |
| HACL1 | 806,03 | 0,10 | 0,05 | 2,20 | 2,76E-02 | 2,08E-01 |
| ATAT1 | 46,05 | -0,24 | 0,11 | -2,20 | 2,78E-02 | 2,08E-01 |
| SLC25A37 | 76097,56 | -0,24 | 0,11 | -2,20 | 2,78E-02 | 2,08E-01 |
| SCN8A | 101,76 | -0,45 | 0,21 | -2,20 | 2,79E-02 | 2,08E-01 |
| ACADSB | 595,26 | 0,09 | 0,04 | 2,20 | 2,77E-02 | 2,08E-01 |
| ALAS2 | 43441,65 | -0,38 | 0,18 | -2,20 | 2,80E-02 | 2,08E-01 |
| C4BPA | 117,66 | -0,69 | 0,31 | -2,20 | 2,80E-02 | 2,08E-01 |
| SLC1A5 | 1477,26 | -0,29 | 0,13 | -2,20 | 2,76E-02 | 2,08E-01 |
| FNDC11 | 31,11 | -0,38 | 0,17 | -2,20 | 2,76E-02 | 2,08E-01 |
| KALRN | 206,88 | -0,38 | 0,17 | -2,20 | 2,80E-02 | 2,08E-01 |
| TRUB1 | 523,90 | 0,17 | 0,08 | 2,20 | 2,80E-02 | 2,08E-01 |
| CSE1L | 1254,28 | 0,12 | 0,05 | 2,20 | 2,78E-02 | 2,08E-01 |
| PDIA6 | 2068,28 | 0,12 | 0,05 | 2,20 | 2,80E-02 | 2,08E-01 |
| GOLGB1 | 4275,51 | 0,11 | 0,05 | 2,20 | 2,80E-02 | 2,08E-01 |
| COX18 | 870,12 | 0,09 | 0,04 | 2,20 | 2,76E-02 | 2,08E-01 |
| MRM3 | 310,76 | 0,07 | 0,03 | 2,20 | 2,79E-02 | 2,08E-01 |
| CCDC66 | 1190,65 | 0,13 | 0,06 | 2,20 | 2,80E-02 | 2,08E-01 |

| | | | | | | |
|---|---|---|---|---|---|---|
| NHLRC3 | 1178,10 | 0,10 | 0,05 | 2,20 | 2,78E-02 | 2,08E-01 |
| ZNF441 | 566,36 | 0,28 | 0,13 | 2,20 | 2,79E-02 | 2,08E-01 |
| SENP7 | 3203,30 | 0,17 | 0,08 | 2,20 | 2,77E-02 | 2,08E-01 |
| CSNK1G3 | 1176,57 | 0,21 | 0,09 | 2,20 | 2,79E-02 | 2,08E-01 |
| P2RY6 | 36,41 | -0,40 | 0,18 | -2,20 | 2,79E-02 | 2,08E-01 |
| ATG4C | 624,52 | 0,18 | 0,08 | 2,20 | 2,77E-02 | 2,08E-01 |
| PBLD | 97,10 | -0,13 | 0,06 | -2,20 | 2,76E-02 | 2,08E-01 |
| USO1 | 2316,89 | 0,13 | 0,06 | 2,20 | 2,80E-02 | 2,08E-01 |
| LAPTM4B | 108,34 | -0,19 | 0,09 | -2,20 | 2,76E-02 | 2,08E-01 |
| ZNF224 | 2085,30 | 0,10 | 0,05 | 2,20 | 2,80E-02 | 2,08E-01 |
| SC5D | 400,29 | 0,16 | 0,07 | 2,20 | 2,81E-02 | 2,08E-01 |
| FLT4 | 119,29 | -0,39 | 0,18 | -2,20 | 2,81E-02 | 2,08E-01 |
| RMI2 | 54,24 | -0,28 | 0,13 | -2,20 | 2,81E-02 | 2,08E-01 |
| SNAI3 | 173,62 | -0,15 | 0,07 | -2,20 | 2,81E-02 | 2,08E-01 |
| BLOC1S2 | 612,83 | 0,20 | 0,09 | 2,20 | 2,81E-02 | 2,08E-01 |
| RPL5 | 7638,60 | 0,22 | 0,10 | 2,20 | 2,81E-02 | 2,08E-01 |
| CCNF | 94,54 | -0,19 | 0,09 | -2,19 | 2,82E-02 | 2,08E-01 |
| PIGQ | 221,37 | -0,24 | 0,11 | -2,19 | 2,82E-02 | 2,08E-01 |
| PWP2 | 84,40 | 0,52 | 0,24 | 2,19 | 2,84E-02 | 2,08E-01 |
| ACACB | 557,04 | -0,20 | 0,09 | -2,19 | 2,84E-02 | 2,08E-01 |
| IER3IP1 | 279,23 | 0,21 | 0,09 | 2,19 | 2,83E-02 | 2,08E-01 |
| DHCR7 | 88,72 | -0,23 | 0,11 | -2,19 | 2,84E-02 | 2,08E-01 |
| UAP1 | 443,10 | 0,12 | 0,06 | 2,19 | 2,84E-02 | 2,08E-01 |
| HPS3 | 2048,26 | 0,10 | 0,04 | 2,19 | 2,83E-02 | 2,08E-01 |
| SHROOM1 | 67,44 | -0,38 | 0,17 | -2,19 | 2,84E-02 | 2,08E-01 |
| PRIM1 | 215,33 | 0,14 | 0,06 | 2,19 | 2,84E-02 | 2,08E-01 |
| CENPQ | 108,05 | 0,27 | 0,12 | 2,19 | 2,84E-02 | 2,08E-01 |
| PCSK6 | 201,10 | -0,35 | 0,16 | -2,19 | 2,84E-02 | 2,08E-01 |
| ZMYM1 | 567,65 | 0,15 | 0,07 | 2,19 | 2,83E-02 | 2,08E-01 |
| KLC3 | 404,66 | -0,35 | 0,16 | -2,19 | 2,85E-02 | 2,09E-01 |
| KLHL20 | 1009,84 | 0,11 | 0,05 | 2,19 | 2,86E-02 | 2,09E-01 |
| NPAS2 | 56,24 | -0,53 | 0,24 | -2,19 | 2,86E-02 | 2,09E-01 |
| GSN | 2028,32 | -0,19 | 0,08 | -2,19 | 2,85E-02 | 2,09E-01 |
| TBC1D30 | 213,13 | -0,18 | 0,08 | -2,19 | 2,85E-02 | 2,09E-01 |
| SSR1 | 3865,92 | 0,08 | 0,04 | 2,19 | 2,86E-02 | 2,09E-01 |
| NUP155 | 1212,91 | 0,06 | 0,03 | 2,19 | 2,86E-02 | 2,09E-01 |
| HGD | 102,70 | -0,16 | 0,08 | -2,19 | 2,86E-02 | 2,09E-01 |
| ABHD10 | 820,39 | 0,18 | 0,08 | 2,19 | 2,86E-02 | 2,09E-01 |
| ZNF268 | 651,83 | 0,20 | 0,09 | 2,19 | 2,87E-02 | 2,09E-01 |
| NARS1 | 2200,44 | 0,11 | 0,05 | 2,19 | 2,87E-02 | 2,09E-01 |
| PNPLA8 | 1720,11 | 0,21 | 0,09 | 2,19 | 2,87E-02 | 2,09E-01 |
| RHPN1 | 112,64 | -0,27 | 0,12 | -2,19 | 2,87E-02 | 2,09E-01 |
| CCDC150 | 47,97 | -0,48 | 0,22 | -2,19 | 2,87E-02 | 2,09E-01 |
| RBMS3 | 56,25 | -0,45 | 0,21 | -2,19 | 2,88E-02 | 2,09E-01 |
| ACKR2 | 38,73 | -0,39 | 0,18 | -2,19 | 2,88E-02 | 2,09E-01 |
| MACROD1 | 59,73 | -0,23 | 0,11 | -2,19 | 2,88E-02 | 2,09E-01 |

| | | | | | | |
|---|---|---|---|---|---|---|
| USP42 | 386,56 | -0,13 | 0,06 | -2,19 | 2,88E-02 | 2,09E-01 |
| ZNF302 | 943,39 | 0,18 | 0,08 | 2,19 | 2,88E-02 | 2,09E-01 |
| EGFL7 | 36,89 | -0,34 | 0,15 | -2,18 | 2,90E-02 | 2,09E-01 |
| TRMT11 | 581,22 | 0,14 | 0,06 | 2,18 | 2,91E-02 | 2,09E-01 |
| ST6GALNAC4 | 388,46 | -0,32 | 0,15 | -2,18 | 2,94E-02 | 2,09E-01 |
| RAD23A | 355,89 | -0,31 | 0,14 | -2,18 | 2,93E-02 | 2,09E-01 |
| SREK1 | 4239,93 | 0,15 | 0,07 | 2,18 | 2,94E-02 | 2,09E-01 |
| SLC25A36 | 1928,90 | 0,19 | 0,09 | 2,18 | 2,93E-02 | 2,09E-01 |
| NCBP1 | 1744,45 | 0,10 | 0,05 | 2,18 | 2,93E-02 | 2,09E-01 |
| ADAT2 | 451,51 | 0,10 | 0,05 | 2,18 | 2,92E-02 | 2,09E-01 |
| SRP9 | 1854,64 | 0,22 | 0,10 | 2,18 | 2,90E-02 | 2,09E-01 |
| SEMA7A | 299,76 | -0,21 | 0,09 | -2,18 | 2,89E-02 | 2,09E-01 |
| ZNF549 | 629,94 | 0,09 | 0,04 | 2,18 | 2,92E-02 | 2,09E-01 |
| PDE9A | 66,91 | -0,33 | 0,15 | -2,18 | 2,91E-02 | 2,09E-01 |
| CACNA1D | 226,22 | -0,34 | 0,16 | -2,18 | 2,90E-02 | 2,09E-01 |
| HSPD1 | 1399,17 | 0,11 | 0,05 | 2,18 | 2,90E-02 | 2,09E-01 |
| LRRN1 | 89,34 | -0,25 | 0,12 | -2,18 | 2,91E-02 | 2,09E-01 |
| POLR3GL | 236,20 | 0,16 | 0,07 | 2,18 | 2,93E-02 | 2,09E-01 |
| PDHB | 1055,41 | 0,08 | 0,04 | 2,18 | 2,89E-02 | 2,09E-01 |
| YTHDC1 | 5135,45 | 0,14 | 0,06 | 2,18 | 2,93E-02 | 2,09E-01 |
| ADCY4 | 117,58 | -0,28 | 0,13 | -2,18 | 2,93E-02 | 2,09E-01 |
| AK4 | 52,31 | -0,35 | 0,16 | -2,18 | 2,90E-02 | 2,09E-01 |
| PRNP | 1257,49 | 0,11 | 0,05 | 2,18 | 2,94E-02 | 2,09E-01 |
| TMEM218 | 653,24 | 0,07 | 0,03 | 2,18 | 2,90E-02 | 2,09E-01 |
| ATP2C2 | 56,59 | -0,51 | 0,23 | -2,18 | 2,89E-02 | 2,09E-01 |
| TFAP2E | 78,19 | -0,22 | 0,10 | -2,18 | 2,89E-02 | 2,09E-01 |
| SPATA9 | 34,37 | -0,33 | 0,15 | -2,18 | 2,92E-02 | 2,09E-01 |
| ADO | 566,29 | 0,11 | 0,05 | 2,18 | 2,89E-02 | 2,09E-01 |
| DDX52 | 1017,59 | 0,07 | 0,03 | 2,18 | 2,93E-02 | 2,09E-01 |
| NFIX | 352,02 | -0,32 | 0,15 | -2,18 | 2,92E-02 | 2,09E-01 |
| FERRY3 | 546,55 | 0,13 | 0,06 | 2,18 | 2,93E-02 | 2,09E-01 |
| ZDHHC9 | 52,09 | -0,20 | 0,09 | -2,18 | 2,92E-02 | 2,09E-01 |
| DHFR2 | 407,13 | 0,15 | 0,07 | 2,18 | 2,94E-02 | 2,09E-01 |
| RAN | 2066,99 | 0,13 | 0,06 | 2,18 | 2,91E-02 | 2,09E-01 |
| GGCT | 120,14 | 0,15 | 0,07 | 2,18 | 2,94E-02 | 2,09E-01 |
| CTSA | 769,78 | -0,22 | 0,10 | -2,18 | 2,94E-02 | 2,09E-01 |
| CYP2R1 | 619,23 | 0,11 | 0,05 | 2,18 | 2,95E-02 | 2,09E-01 |
| RPL9 | 5294,21 | 0,37 | 0,17 | 2,18 | 2,95E-02 | 2,09E-01 |
| CDADC1 | 365,54 | 0,11 | 0,05 | 2,18 | 2,95E-02 | 2,09E-01 |
| SAR1B | 1430,26 | 0,09 | 0,04 | 2,18 | 2,95E-02 | 2,09E-01 |
| CIPC | 452,01 | 0,07 | 0,03 | 2,17 | 2,97E-02 | 2,09E-01 |
| UBR1 | 2847,15 | 0,09 | 0,04 | 2,18 | 2,96E-02 | 2,09E-01 |
| LSM5 | 274,65 | 0,19 | 0,09 | 2,18 | 2,96E-02 | 2,09E-01 |
| LARGE2 | 59,78 | -0,32 | 0,15 | -2,18 | 2,96E-02 | 2,09E-01 |
| CHCHD3 | 953,27 | 0,09 | 0,04 | 2,17 | 2,97E-02 | 2,09E-01 |
| R3HCC1L | 1365,53 | 0,12 | 0,05 | 2,18 | 2,96E-02 | 2,09E-01 |

| | | | | | | |
|---|---|---|---|---|---|---|
| ATAD1 | 1411,33 | 0,18 | 0,08 | 2,17 | 2,96E-02 | 2,09E-01 |
| TTLL11 | 146,50 | -0,15 | 0,07 | -2,18 | 2,96E-02 | 2,09E-01 |
| CETN3 | 169,11 | 0,25 | 0,11 | 2,17 | 2,96E-02 | 2,09E-01 |
| SLC35G5 | 25,52 | -0,25 | 0,12 | -2,17 | 2,97E-02 | 2,10E-01 |
| KYAT1 | 152,08 | -0,19 | 0,09 | -2,17 | 2,98E-02 | 2,10E-01 |
| HYCC1 | 1943,03 | 0,15 | 0,07 | 2,17 | 2,98E-02 | 2,10E-01 |
| ZFP62 | 1213,35 | 0,15 | 0,07 | 2,17 | 3,00E-02 | 2,10E-01 |
| PHTF2 | 1948,22 | 0,16 | 0,07 | 2,17 | 3,00E-02 | 2,10E-01 |
| GTF2E1 | 704,90 | 0,14 | 0,06 | 2,17 | 3,00E-02 | 2,10E-01 |
| BRD3OS | 115,97 | -0,17 | 0,08 | -2,17 | 2,98E-02 | 2,10E-01 |
| LZIC | 624,52 | 0,10 | 0,05 | 2,17 | 3,00E-02 | 2,10E-01 |
| KMO | 339,22 | 0,16 | 0,07 | 2,17 | 3,00E-02 | 2,10E-01 |
| ZNF521 | 41,16 | -0,42 | 0,19 | -2,17 | 3,00E-02 | 2,10E-01 |
| MCTS1 | 405,47 | 0,13 | 0,06 | 2,17 | 2,99E-02 | 2,10E-01 |
| ICE1 | 1920,44 | 0,09 | 0,04 | 2,17 | 2,99E-02 | 2,10E-01 |
| RBM41 | 756,09 | 0,13 | 0,06 | 2,17 | 2,99E-02 | 2,10E-01 |
| FASN | 201,23 | -0,37 | 0,17 | -2,17 | 3,00E-02 | 2,10E-01 |
| ZNF776 | 1187,94 | 0,13 | 0,06 | 2,17 | 3,03E-02 | 2,10E-01 |
| KCTD12 | 3490,47 | 0,16 | 0,07 | 2,17 | 3,03E-02 | 2,10E-01 |
| AKAP11 | 2990,92 | 0,13 | 0,06 | 2,17 | 3,01E-02 | 2,10E-01 |
| ITGA3 | 101,03 | -0,32 | 0,15 | -2,17 | 3,01E-02 | 2,10E-01 |
| TRMT13 | 878,15 | 0,19 | 0,09 | 2,17 | 3,03E-02 | 2,10E-01 |
| VBP1 | 632,17 | 0,24 | 0,11 | 2,17 | 3,02E-02 | 2,10E-01 |
| ZNF570 | 381,69 | 0,14 | 0,06 | 2,17 | 3,01E-02 | 2,10E-01 |
| CASP8AP2 | 1384,99 | 0,20 | 0,09 | 2,17 | 3,02E-02 | 2,10E-01 |
| P2RY2 | 276,87 | -0,23 | 0,11 | -2,17 | 3,02E-02 | 2,10E-01 |
| STPG1 | 101,92 | -0,22 | 0,10 | -2,17 | 3,02E-02 | 2,10E-01 |
| ABCG2 | 158,71 | -0,28 | 0,13 | -2,17 | 3,02E-02 | 2,10E-01 |
| FGFRL1 | 172,59 | -0,19 | 0,09 | -2,17 | 3,02E-02 | 2,10E-01 |
| EIF3M | 2200,66 | 0,13 | 0,06 | 2,17 | 3,02E-02 | 2,10E-01 |
| VPS45 | 865,46 | 0,10 | 0,05 | 2,17 | 3,01E-02 | 2,10E-01 |
| CAPN5 | 167,62 | -0,30 | 0,14 | -2,17 | 3,01E-02 | 2,10E-01 |
| GADD45A | 285,09 | -0,16 | 0,08 | -2,17 | 3,04E-02 | 2,10E-01 |
| ZMYND11 | 1365,12 | 0,12 | 0,05 | 2,17 | 3,04E-02 | 2,10E-01 |
| CLASP2 | 2997,41 | 0,09 | 0,04 | 2,17 | 3,04E-02 | 2,10E-01 |
| ECD | 1029,16 | 0,07 | 0,03 | 2,17 | 3,04E-02 | 2,10E-01 |
| ZNF22 | 512,05 | 0,24 | 0,11 | 2,16 | 3,04E-02 | 2,11E-01 |
| HNRNPU | 22951,92 | 0,10 | 0,05 | 2,16 | 3,07E-02 | 2,11E-01 |
| DNAJB2 | 2688,79 | -0,21 | 0,10 | -2,16 | 3,07E-02 | 2,11E-01 |
| ZNF574 | 198,97 | -0,14 | 0,06 | -2,16 | 3,05E-02 | 2,11E-01 |
| CCDC157 | 80,24 | -0,23 | 0,11 | -2,16 | 3,07E-02 | 2,11E-01 |
| EIF5A | 1824,00 | -0,11 | 0,05 | -2,16 | 3,07E-02 | 2,11E-01 |
| NUS1 | 762,79 | 0,17 | 0,08 | 2,16 | 3,07E-02 | 2,11E-01 |
| PYROXD1 | 1096,95 | 0,17 | 0,08 | 2,16 | 3,07E-02 | 2,11E-01 |
| ZNF611 | 1019,04 | 0,08 | 0,04 | 2,16 | 3,05E-02 | 2,11E-01 |
| FRS3 | 75,55 | -0,20 | 0,09 | -2,16 | 3,07E-02 | 2,11E-01 |

| | | | | | | |
|---|---|---|---|---|---|---|
| ARVCF | 129,01 | -0,28 | 0,13 | -2,16 | 3,06E-02 | 2,11E-01 |
| TRAPPC11 | 2055,92 | 0,08 | 0,04 | 2,16 | 3,06E-02 | 2,11E-01 |
| ACSM1 | 40,03 | -0,36 | 0,17 | -2,16 | 3,06E-02 | 2,11E-01 |
| ETV3 | 1767,13 | 0,07 | 0,03 | 2,16 | 3,07E-02 | 2,11E-01 |
| KIF4A | 41,71 | -0,34 | 0,16 | -2,16 | 3,07E-02 | 2,11E-01 |
| PPP2R5E | 1327,33 | 0,11 | 0,05 | 2,16 | 3,06E-02 | 2,11E-01 |
| ZBTB1 | 2027,26 | 0,18 | 0,08 | 2,16 | 3,07E-02 | 2,11E-01 |
| ZNF263 | 991,99 | 0,06 | 0,03 | 2,16 | 3,09E-02 | 2,11E-01 |
| TMEM168 | 1812,36 | 0,14 | 0,07 | 2,16 | 3,10E-02 | 2,11E-01 |
| FBXL13 | 730,29 | -0,22 | 0,10 | -2,16 | 3,09E-02 | 2,11E-01 |
| N4BP2 | 1562,55 | 0,17 | 0,08 | 2,16 | 3,10E-02 | 2,11E-01 |
| FEZ1 | 108,13 | -0,30 | 0,14 | -2,16 | 3,09E-02 | 2,11E-01 |
| TIPRL | 964,04 | 0,14 | 0,07 | 2,16 | 3,08E-02 | 2,11E-01 |
| PEX3 | 408,37 | 0,16 | 0,07 | 2,16 | 3,10E-02 | 2,11E-01 |
| XPA | 568,62 | 0,14 | 0,07 | 2,16 | 3,09E-02 | 2,11E-01 |
| PAICS | 380,20 | 0,09 | 0,04 | 2,16 | 3,09E-02 | 2,11E-01 |
| HMGN3 | 453,94 | 0,13 | 0,06 | 2,16 | 3,09E-02 | 2,11E-01 |
| RPE | 2375,01 | 0,14 | 0,06 | 2,16 | 3,09E-02 | 2,11E-01 |
| APPBP2 | 2025,49 | 0,09 | 0,04 | 2,16 | 3,08E-02 | 2,11E-01 |
| SFRP2 | 42,86 | -0,39 | 0,18 | -2,16 | 3,09E-02 | 2,11E-01 |
| PNP | 1650,19 | -0,15 | 0,07 | -2,16 | 3,10E-02 | 2,11E-01 |
| CACNA1I | 327,68 | -0,31 | 0,14 | -2,16 | 3,10E-02 | 2,11E-01 |
| LYPD6B | 30,44 | -0,51 | 0,24 | -2,16 | 3,11E-02 | 2,11E-01 |
| ZNF341 | 112,55 | -0,28 | 0,13 | -2,15 | 3,13E-02 | 2,11E-01 |
| UFM1 | 1763,22 | 0,23 | 0,11 | 2,15 | 3,12E-02 | 2,11E-01 |
| MRPS31 | 602,14 | 0,14 | 0,06 | 2,15 | 3,13E-02 | 2,11E-01 |
| AGPS | 1850,58 | 0,14 | 0,07 | 2,15 | 3,13E-02 | 2,11E-01 |
| DNAJC25 | 145,65 | 0,13 | 0,06 | 2,15 | 3,13E-02 | 2,11E-01 |
| TMEM156 | 369,60 | 0,14 | 0,07 | 2,16 | 3,11E-02 | 2,11E-01 |
| PROSER1 | 1049,62 | 0,07 | 0,03 | 2,15 | 3,12E-02 | 2,11E-01 |
| XPO4 | 1957,87 | 0,11 | 0,05 | 2,15 | 3,13E-02 | 2,11E-01 |
| TRIP11 | 2269,82 | 0,14 | 0,07 | 2,15 | 3,12E-02 | 2,11E-01 |
| GARIN1A | 60,50 | -0,32 | 0,15 | -2,16 | 3,11E-02 | 2,11E-01 |
| FIGNL1 | 387,94 | 0,16 | 0,07 | 2,15 | 3,13E-02 | 2,11E-01 |
| TSFM | 247,51 | 0,08 | 0,04 | 2,15 | 3,12E-02 | 2,11E-01 |
| RAB21 | 3247,78 | 0,09 | 0,04 | 2,15 | 3,12E-02 | 2,11E-01 |
| ECHDC1 | 1427,56 | 0,16 | 0,07 | 2,15 | 3,12E-02 | 2,11E-01 |
| LTBP2 | 223,77 | -0,25 | 0,12 | -2,15 | 3,14E-02 | 2,12E-01 |
| SHPRH | 3167,84 | 0,12 | 0,06 | 2,15 | 3,14E-02 | 2,12E-01 |
| THOP1 | 221,27 | -0,18 | 0,08 | -2,15 | 3,14E-02 | 2,12E-01 |
| TBC1D25 | 296,63 | -0,18 | 0,08 | -2,15 | 3,15E-02 | 2,12E-01 |
| RDH14 | 234,79 | 0,18 | 0,08 | 2,15 | 3,14E-02 | 2,12E-01 |
| ADPRM | 444,98 | 0,18 | 0,08 | 2,15 | 3,14E-02 | 2,12E-01 |
| DDX1 | 1027,45 | 0,12 | 0,06 | 2,15 | 3,16E-02 | 2,12E-01 |
| RNF6 | 1317,30 | 0,14 | 0,07 | 2,15 | 3,16E-02 | 2,12E-01 |
| WDR18 | 167,12 | -0,14 | 0,06 | -2,15 | 3,16E-02 | 2,12E-01 |

| | | | | | | |
|---|---|---|---|---|---|---|
| FAT1 | 88,37 | -0,46 | 0,21 | -2,15 | 3,16E-02 | 2,12E-01 |
| ASCC2 | 4898,88 | -0,27 | 0,12 | -2,15 | 3,15E-02 | 2,12E-01 |
| RAB3GAP2 | 2843,54 | 0,09 | 0,04 | 2,15 | 3,16E-02 | 2,12E-01 |
| PKD2 | 678,13 | 0,09 | 0,04 | 2,15 | 3,17E-02 | 2,12E-01 |
| CHD1L | 989,89 | 0,08 | 0,04 | 2,15 | 3,17E-02 | 2,12E-01 |
| CCDC124 | 142,05 | -0,19 | 0,09 | -2,15 | 3,17E-02 | 2,12E-01 |
| CENPC | 2515,65 | 0,13 | 0,06 | 2,15 | 3,17E-02 | 2,12E-01 |
| HSPA4 | 1942,01 | 0,11 | 0,05 | 2,15 | 3,17E-02 | 2,12E-01 |
| RECQL4 | 45,60 | -0,36 | 0,17 | -2,15 | 3,17E-02 | 2,12E-01 |
| INPP5E | 174,26 | -0,25 | 0,12 | -2,15 | 3,18E-02 | 2,12E-01 |
| AVIL | 665,98 | -0,18 | 0,09 | -2,15 | 3,17E-02 | 2,12E-01 |
| RBM27 | 904,80 | 0,15 | 0,07 | 2,15 | 3,18E-02 | 2,12E-01 |
| CLEC2B | 1521,49 | 0,24 | 0,11 | 2,15 | 3,18E-02 | 2,12E-01 |
| TRMO | 1454,01 | 0,10 | 0,05 | 2,15 | 3,18E-02 | 2,12E-01 |
| CYCS | 1463,38 | 0,14 | 0,07 | 2,15 | 3,18E-02 | 2,12E-01 |
| PPP4R3B | 5053,66 | 0,14 | 0,06 | 2,15 | 3,19E-02 | 2,12E-01 |
| CTR9 | 1573,73 | 0,09 | 0,04 | 2,15 | 3,19E-02 | 2,12E-01 |
| MPP1 | 9984,81 | -0,25 | 0,12 | -2,15 | 3,19E-02 | 2,12E-01 |
| PGGT1B | 2948,27 | 0,16 | 0,07 | 2,15 | 3,18E-02 | 2,12E-01 |
| SIKE1 | 1392,78 | 0,16 | 0,07 | 2,14 | 3,20E-02 | 2,12E-01 |
| VPS50 | 1669,53 | 0,18 | 0,09 | 2,14 | 3,20E-02 | 2,12E-01 |
| SEMA3C | 436,06 | 0,16 | 0,08 | 2,15 | 3,19E-02 | 2,12E-01 |
| LGALS12 | 183,35 | -0,26 | 0,12 | -2,15 | 3,19E-02 | 2,12E-01 |
| DOK4 | 68,67 | -0,31 | 0,15 | -2,14 | 3,20E-02 | 2,12E-01 |
| SLFN5 | 6650,67 | 0,13 | 0,06 | 2,14 | 3,21E-02 | 2,12E-01 |
| EVA1B | 162,63 | -0,17 | 0,08 | -2,14 | 3,21E-02 | 2,12E-01 |
| WDR5B | 577,35 | 0,17 | 0,08 | 2,14 | 3,22E-02 | 2,13E-01 |
| LY6G5C | 31,76 | -0,29 | 0,13 | -2,14 | 3,22E-02 | 2,13E-01 |
| PLD6 | 160,41 | -0,14 | 0,07 | -2,14 | 3,22E-02 | 2,13E-01 |
| CCDC61 | 117,08 | -0,21 | 0,10 | -2,14 | 3,21E-02 | 2,13E-01 |
| TMEM106B | 2170,19 | 0,18 | 0,09 | 2,14 | 3,22E-02 | 2,13E-01 |
| SUB1 | 2406,01 | 0,18 | 0,09 | 2,14 | 3,22E-02 | 2,13E-01 |
| RALGAPA1 | 2833,15 | 0,11 | 0,05 | 2,14 | 3,26E-02 | 2,13E-01 |
| NT5E | 228,23 | 0,24 | 0,11 | 2,14 | 3,25E-02 | 2,13E-01 |
| NR2F6 | 48,97 | -0,27 | 0,13 | -2,14 | 3,27E-02 | 2,13E-01 |
| PIK3C3 | 3820,91 | 0,12 | 0,05 | 2,14 | 3,24E-02 | 2,13E-01 |
| AHNAK | 59926,05 | 0,10 | 0,05 | 2,14 | 3,25E-02 | 2,13E-01 |
| SLC12A7 | 718,54 | -0,27 | 0,13 | -2,14 | 3,25E-02 | 2,13E-01 |
| GLMN | 418,76 | 0,14 | 0,07 | 2,14 | 3,23E-02 | 2,13E-01 |
| EXOSC7 | 622,48 | 0,08 | 0,04 | 2,14 | 3,26E-02 | 2,13E-01 |
| ZNF431 | 2207,65 | 0,09 | 0,04 | 2,14 | 3,26E-02 | 2,13E-01 |
| EID1 | 1898,21 | 0,18 | 0,09 | 2,14 | 3,26E-02 | 2,13E-01 |
| RPS8 | 9852,37 | 0,21 | 0,10 | 2,14 | 3,26E-02 | 2,13E-01 |
| RPL22L1 | 234,39 | 0,25 | 0,12 | 2,14 | 3,23E-02 | 2,13E-01 |
| GRAMD1B | 448,33 | -0,22 | 0,10 | -2,14 | 3,27E-02 | 2,13E-01 |
| UBB | 57353,89 | -0,27 | 0,13 | -2,14 | 3,24E-02 | 2,13E-01 |

| | | | | | | |
|---|---|---|---|---|---|---|
| SLC38A5 | 306,97 | -0,32 | 0,15 | -2,14 | 3,23E-02 | 2,13E-01 |
| ABCD3 | 1132,41 | 0,12 | 0,05 | 2,14 | 3,26E-02 | 2,13E-01 |
| COMMD10 | 787,78 | 0,13 | 0,06 | 2,14 | 3,25E-02 | 2,13E-01 |
| UROD | 704,74 | -0,13 | 0,06 | -2,14 | 3,26E-02 | 2,13E-01 |
| TMED10 | 3554,78 | 0,10 | 0,05 | 2,14 | 3,26E-02 | 2,13E-01 |
| GALNT8 | 35,47 | -0,38 | 0,18 | -2,14 | 3,25E-02 | 2,13E-01 |
| MYO9A | 2250,52 | 0,10 | 0,05 | 2,14 | 3,26E-02 | 2,13E-01 |
| TRAPPC13 | 687,10 | 0,15 | 0,07 | 2,14 | 3,27E-02 | 2,13E-01 |
| ANKAR | 628,86 | 0,12 | 0,06 | 2,14 | 3,24E-02 | 2,13E-01 |
| FARSB | 578,25 | 0,10 | 0,04 | 2,14 | 3,27E-02 | 2,13E-01 |
| STAG3 | 46,79 | -0,35 | 0,16 | -2,14 | 3,27E-02 | 2,13E-01 |
| YBX3 | 14786,27 | -0,28 | 0,13 | -2,14 | 3,23E-02 | 2,13E-01 |
| CISD1 | 225,96 | 0,17 | 0,08 | 2,14 | 3,24E-02 | 2,13E-01 |
| SLC26A8 | 210,51 | -0,27 | 0,13 | -2,14 | 3,28E-02 | 2,13E-01 |
| COPS4 | 432,97 | 0,18 | 0,08 | 2,13 | 3,28E-02 | 2,13E-01 |
| FKBP3 | 454,38 | 0,16 | 0,08 | 2,13 | 3,28E-02 | 2,13E-01 |
| VPS13C | 18806,07 | 0,14 | 0,07 | 2,13 | 3,29E-02 | 2,14E-01 |
| ADIPOR1 | 30706,01 | -0,27 | 0,13 | -2,13 | 3,29E-02 | 2,14E-01 |
| TPPP | 67,52 | -0,36 | 0,17 | -2,13 | 3,30E-02 | 2,14E-01 |
| KCND3 | 44,73 | -0,46 | 0,21 | -2,13 | 3,29E-02 | 2,14E-01 |
| ZFR | 3088,44 | 0,10 | 0,05 | 2,13 | 3,29E-02 | 2,14E-01 |
| FANCF | 315,55 | 0,12 | 0,06 | 2,13 | 3,30E-02 | 2,14E-01 |
| NAPA | 1010,84 | -0,20 | 0,09 | -2,13 | 3,30E-02 | 2,14E-01 |
| GFM2 | 805,87 | 0,13 | 0,06 | 2,13 | 3,30E-02 | 2,14E-01 |
| TREML1 | 176,57 | -0,33 | 0,16 | -2,13 | 3,32E-02 | 2,14E-01 |
| ERMAP | 649,53 | -0,11 | 0,05 | -2,13 | 3,31E-02 | 2,14E-01 |
| AP2S1 | 1093,32 | -0,15 | 0,07 | -2,13 | 3,32E-02 | 2,14E-01 |
| NBEAL1 | 1178,84 | 0,10 | 0,05 | 2,13 | 3,32E-02 | 2,14E-01 |
| ZBED5 | 1792,28 | 0,18 | 0,09 | 2,13 | 3,32E-02 | 2,14E-01 |
| LYL1 | 2677,97 | -0,21 | 0,10 | -2,13 | 3,32E-02 | 2,14E-01 |
| TMEM187 | 105,61 | -0,11 | 0,05 | -2,13 | 3,31E-02 | 2,14E-01 |
| TMEM126B | 516,66 | 0,16 | 0,08 | 2,13 | 3,32E-02 | 2,14E-01 |
| FBXO22 | 1289,51 | 0,13 | 0,06 | 2,13 | 3,31E-02 | 2,14E-01 |
| ARHGEF10 | 46,82 | -0,35 | 0,17 | -2,13 | 3,32E-02 | 2,14E-01 |
| WASL | 632,40 | 0,14 | 0,07 | 2,13 | 3,31E-02 | 2,14E-01 |
| SRP72 | 1446,61 | 0,14 | 0,07 | 2,13 | 3,33E-02 | 2,14E-01 |
| PTP4A3 | 512,05 | -0,16 | 0,07 | -2,13 | 3,33E-02 | 2,14E-01 |
| ICAM5 | 53,06 | -0,29 | 0,14 | -2,13 | 3,34E-02 | 2,14E-01 |
| RPH3AL | 86,26 | -0,33 | 0,16 | -2,13 | 3,35E-02 | 2,14E-01 |
| GMFB | 1958,16 | 0,23 | 0,11 | 2,13 | 3,35E-02 | 2,14E-01 |
| RASGEF1B | 707,89 | 0,11 | 0,05 | 2,13 | 3,35E-02 | 2,14E-01 |
| LMNTD2 | 48,36 | -0,29 | 0,13 | -2,13 | 3,34E-02 | 2,14E-01 |
| CTTNBP2 | 38,30 | -0,50 | 0,23 | -2,13 | 3,34E-02 | 2,14E-01 |
| TTC19 | 1094,67 | 0,08 | 0,04 | 2,13 | 3,35E-02 | 2,14E-01 |
| ZNF277 | 605,80 | 0,15 | 0,07 | 2,13 | 3,35E-02 | 2,14E-01 |
| VRK1 | 786,09 | 0,22 | 0,11 | 2,12 | 3,36E-02 | 2,15E-01 |

| | | | | | | |
|---|---|---|---|---|---|---|
| CDV3 | 5736,47 | 0,11 | 0,05 | 2,13 | 3,36E-02 | 2,15E-01 |
| NDUFS4 | 231,54 | 0,17 | 0,08 | 2,12 | 3,36E-02 | 2,15E-01 |
| CSRNP2 | 519,18 | -0,09 | 0,04 | -2,12 | 3,36E-02 | 2,15E-01 |
| THUMPD3 | 523,17 | 0,09 | 0,04 | 2,12 | 3,37E-02 | 2,15E-01 |
| ZNHIT6 | 709,61 | 0,12 | 0,06 | 2,12 | 3,37E-02 | 2,15E-01 |
| ZNF628 | 63,38 | -0,33 | 0,16 | -2,12 | 3,37E-02 | 2,15E-01 |
| ANKRD12 | 6058,04 | 0,18 | 0,09 | 2,12 | 3,38E-02 | 2,15E-01 |
| TENM4 | 113,09 | -0,62 | 0,29 | -2,12 | 3,37E-02 | 2,15E-01 |
| CPSF2 | 2606,84 | 0,08 | 0,04 | 2,12 | 3,39E-02 | 2,15E-01 |
| TCF4 | 1369,55 | 0,19 | 0,09 | 2,12 | 3,39E-02 | 2,15E-01 |
| GIMAP6 | 6707,58 | 0,11 | 0,05 | 2,12 | 3,38E-02 | 2,15E-01 |
| MYO5C | 86,79 | -0,37 | 0,17 | -2,12 | 3,39E-02 | 2,15E-01 |
| TIMM23B | 551,54 | 0,08 | 0,04 | 2,12 | 3,38E-02 | 2,15E-01 |
| PIAS3 | 157,79 | -0,22 | 0,10 | -2,12 | 3,39E-02 | 2,15E-01 |
| NAP1L1 | 3731,91 | 0,14 | 0,07 | 2,12 | 3,39E-02 | 2,15E-01 |
| ORC1 | 40,39 | -0,28 | 0,13 | -2,12 | 3,39E-02 | 2,15E-01 |
| MOXD1 | 29,48 | -0,40 | 0,19 | -2,12 | 3,39E-02 | 2,15E-01 |
| CEP85L | 3347,47 | 0,14 | 0,07 | 2,12 | 3,40E-02 | 2,15E-01 |
| BRWD1 | 7599,92 | 0,14 | 0,07 | 2,11 | 3,45E-02 | 2,15E-01 |
| LTC4S | 127,95 | -0,17 | 0,08 | -2,11 | 3,45E-02 | 2,15E-01 |
| TMEM160 | 179,03 | -0,10 | 0,05 | -2,11 | 3,49E-02 | 2,15E-01 |
| SARDH | 111,52 | -0,32 | 0,15 | -2,11 | 3,45E-02 | 2,15E-01 |
| TRPT1 | 99,67 | -0,14 | 0,07 | -2,11 | 3,45E-02 | 2,15E-01 |
| ZNF573 | 388,62 | 0,10 | 0,05 | 2,12 | 3,42E-02 | 2,15E-01 |
| MYSM1 | 3322,64 | 0,13 | 0,06 | 2,11 | 3,49E-02 | 2,15E-01 |
| ALDH4A1 | 32,65 | -0,31 | 0,15 | -2,11 | 3,47E-02 | 2,15E-01 |
| SNX14 | 2341,17 | 0,13 | 0,06 | 2,12 | 3,41E-02 | 2,15E-01 |
| SPHK1 | 152,33 | -0,18 | 0,09 | -2,11 | 3,50E-02 | 2,15E-01 |
| SLC9A9 | 948,82 | 0,10 | 0,05 | 2,11 | 3,49E-02 | 2,15E-01 |
| MAPK8 | 1563,39 | 0,12 | 0,06 | 2,11 | 3,45E-02 | 2,15E-01 |
| PRMT3 | 799,60 | 0,10 | 0,05 | 2,11 | 3,47E-02 | 2,15E-01 |
| TMEM39B | 115,64 | -0,20 | 0,09 | -2,12 | 3,41E-02 | 2,15E-01 |
| ZNF484 | 906,91 | 0,20 | 0,09 | 2,12 | 3,44E-02 | 2,15E-01 |
| F2RL3 | 38,19 | -0,32 | 0,15 | -2,11 | 3,49E-02 | 2,15E-01 |
| OTUD6B | 706,78 | 0,18 | 0,09 | 2,11 | 3,47E-02 | 2,15E-01 |
| RTEL1 | 79,15 | -0,18 | 0,09 | -2,11 | 3,50E-02 | 2,15E-01 |
| PSD | 60,34 | -0,39 | 0,18 | -2,12 | 3,42E-02 | 2,15E-01 |
| KCTD1 | 124,45 | -0,23 | 0,11 | -2,11 | 3,46E-02 | 2,15E-01 |
| KCTD3 | 237,61 | 0,13 | 0,06 | 2,11 | 3,45E-02 | 2,15E-01 |
| BMS1 | 1276,73 | 0,09 | 0,04 | 2,12 | 3,40E-02 | 2,15E-01 |
| BEND2 | 249,23 | -0,27 | 0,13 | -2,11 | 3,50E-02 | 2,15E-01 |
| CEBPG | 987,94 | 0,14 | 0,07 | 2,12 | 3,43E-02 | 2,15E-01 |
| FBXO39 | 30,80 | -0,34 | 0,16 | -2,11 | 3,49E-02 | 2,15E-01 |
| REC8 | 176,80 | -0,29 | 0,14 | -2,12 | 3,44E-02 | 2,15E-01 |
| KLHL17 | 126,28 | -0,21 | 0,10 | -2,12 | 3,40E-02 | 2,15E-01 |
| TTI1 | 965,21 | 0,06 | 0,03 | 2,11 | 3,48E-02 | 2,15E-01 |

| | | | | | | |
|---|---|---|---|---|---|---|
| RRAS2 | 754,60 | 0,17 | 0,08 | 2,12 | 3,42E-02 | 2,15E-01 |
| GPAM | 455,35 | 0,11 | 0,05 | 2,11 | 3,45E-02 | 2,15E-01 |
| PPWD1 | 1923,67 | 0,15 | 0,07 | 2,11 | 3,45E-02 | 2,15E-01 |
| FIP1L1 | 1241,67 | 0,11 | 0,05 | 2,11 | 3,50E-02 | 2,15E-01 |
| UBTD2 | 240,55 | 0,10 | 0,05 | 2,11 | 3,48E-02 | 2,15E-01 |
| RCN2 | 450,65 | 0,13 | 0,06 | 2,11 | 3,47E-02 | 2,15E-01 |
| SLC6A12 | 118,84 | -0,27 | 0,13 | -2,11 | 3,45E-02 | 2,15E-01 |
| ZNF761 | 316,46 | 0,16 | 0,08 | 2,11 | 3,48E-02 | 2,15E-01 |
| FAM171A1 | 68,51 | -0,23 | 0,11 | -2,12 | 3,43E-02 | 2,15E-01 |
| TDRD3 | 833,44 | 0,12 | 0,06 | 2,11 | 3,50E-02 | 2,15E-01 |
| TRIM10 | 552,46 | -0,26 | 0,13 | -2,11 | 3,47E-02 | 2,15E-01 |
| LTN1 | 3162,16 | 0,14 | 0,07 | 2,11 | 3,47E-02 | 2,15E-01 |
| SESN1 | 1361,57 | 0,14 | 0,07 | 2,11 | 3,49E-02 | 2,15E-01 |
| CHST13 | 46,68 | -0,35 | 0,17 | -2,12 | 3,43E-02 | 2,15E-01 |
| ATRX | 6144,08 | 0,14 | 0,07 | 2,12 | 3,42E-02 | 2,15E-01 |
| GZF1 | 608,50 | 0,10 | 0,05 | 2,11 | 3,47E-02 | 2,15E-01 |
| MAGOH | 399,99 | 0,14 | 0,07 | 2,11 | 3,46E-02 | 2,15E-01 |
| ECH1 | 298,60 | -0,13 | 0,06 | -2,11 | 3,47E-02 | 2,15E-01 |
| SUZ12 | 1243,34 | 0,17 | 0,08 | 2,11 | 3,48E-02 | 2,15E-01 |
| ZBTB10 | 802,28 | 0,17 | 0,08 | 2,11 | 3,45E-02 | 2,15E-01 |
| HRK | 71,61 | -0,47 | 0,22 | -2,11 | 3,46E-02 | 2,15E-01 |
| FAM167A | 44,52 | -0,41 | 0,19 | -2,12 | 3,42E-02 | 2,15E-01 |
| UCHL5 | 1229,10 | 0,18 | 0,09 | 2,11 | 3,47E-02 | 2,15E-01 |
| ZNF449 | 338,66 | 0,12 | 0,06 | 2,11 | 3,47E-02 | 2,15E-01 |
| IGIP | 652,09 | 0,14 | 0,06 | 2,11 | 3,50E-02 | 2,15E-01 |
| FH | 584,60 | 0,09 | 0,04 | 2,11 | 3,48E-02 | 2,15E-01 |
| CPNE8 | 621,40 | 0,16 | 0,08 | 2,11 | 3,48E-02 | 2,15E-01 |
| KREMEN1 | 600,72 | -0,31 | 0,14 | -2,11 | 3,46E-02 | 2,15E-01 |
| THAP12 | 1706,52 | 0,17 | 0,08 | 2,12 | 3,41E-02 | 2,15E-01 |
| ZSCAN29 | 944,77 | 0,07 | 0,03 | 2,11 | 3,51E-02 | 2,15E-01 |
| NCDN | 106,66 | -0,28 | 0,13 | -2,11 | 3,51E-02 | 2,15E-01 |
| ZCCHC9 | 718,56 | 0,12 | 0,06 | 2,11 | 3,51E-02 | 2,15E-01 |
| SMIM3 | 317,73 | -0,16 | 0,08 | -2,11 | 3,50E-02 | 2,15E-01 |
| CTDSPL | 225,92 | -0,29 | 0,14 | -2,11 | 3,51E-02 | 2,15E-01 |
| ZFAND1 | 613,49 | 0,20 | 0,09 | 2,11 | 3,51E-02 | 2,15E-01 |
| MYNN | 1027,92 | 0,18 | 0,09 | 2,11 | 3,51E-02 | 2,15E-01 |
| LRRC20 | 62,64 | -0,22 | 0,11 | -2,11 | 3,52E-02 | 2,16E-01 |
| TMTC3 | 804,75 | 0,18 | 0,09 | 2,10 | 3,53E-02 | 2,16E-01 |
| SUPV3L1 | 550,35 | 0,08 | 0,04 | 2,10 | 3,54E-02 | 2,16E-01 |
| MIA3 | 2654,45 | 0,08 | 0,04 | 2,10 | 3,53E-02 | 2,16E-01 |
| MIB1 | 3441,60 | 0,13 | 0,06 | 2,10 | 3,54E-02 | 2,16E-01 |
| ZNF669 | 269,52 | 0,15 | 0,07 | 2,10 | 3,54E-02 | 2,16E-01 |
| PDLIM1 | 378,93 | -0,18 | 0,09 | -2,10 | 3,54E-02 | 2,16E-01 |
| APOLD1 | 251,52 | -0,15 | 0,07 | -2,10 | 3,54E-02 | 2,16E-01 |
| TMCO1 | 1164,95 | 0,20 | 0,10 | 2,11 | 3,52E-02 | 2,16E-01 |
| CCDC9B | 118,70 | -0,24 | 0,11 | -2,11 | 3,53E-02 | 2,16E-01 |

| | | | | | | |
|---|---|---|---|---|---|---|
| NELFA | 257,73 | -0,21 | 0,10 | -2,10 | 3,53E-02 | 2,16E-01 |
| OR4D1 | 49,00 | -0,20 | 0,10 | -2,11 | 3,53E-02 | 2,16E-01 |
| POLR2K | 242,72 | 0,32 | 0,15 | 2,10 | 3,53E-02 | 2,16E-01 |
| IPO11 | 586,26 | 0,12 | 0,06 | 2,10 | 3,54E-02 | 2,16E-01 |
| ARMCX3 | 817,26 | 0,09 | 0,04 | 2,10 | 3,54E-02 | 2,16E-01 |
| SEC61G | 618,04 | 0,16 | 0,07 | 2,10 | 3,55E-02 | 2,16E-01 |
| AP2A1 | 1796,93 | -0,27 | 0,13 | -2,10 | 3,55E-02 | 2,16E-01 |
| OSBP2 | 1419,25 | -0,33 | 0,16 | -2,10 | 3,55E-02 | 2,16E-01 |
| GALNTL6 | 40,32 | -0,50 | 0,24 | -2,10 | 3,55E-02 | 2,16E-01 |
| SHROOM4 | 39,38 | -0,39 | 0,19 | -2,10 | 3,55E-02 | 2,16E-01 |
| FAM131B | 60,07 | -0,26 | 0,12 | -2,10 | 3,56E-02 | 2,16E-01 |
| GYPC | 19489,95 | -0,27 | 0,13 | -2,10 | 3,56E-02 | 2,16E-01 |
| ACTL6A | 686,84 | 0,11 | 0,05 | 2,10 | 3,56E-02 | 2,16E-01 |
| FEM1B | 2813,58 | 0,13 | 0,06 | 2,10 | 3,56E-02 | 2,16E-01 |
| NUP205 | 2180,15 | 0,09 | 0,04 | 2,10 | 3,56E-02 | 2,16E-01 |
| PPIE | 751,60 | 0,13 | 0,06 | 2,10 | 3,57E-02 | 2,16E-01 |
| SEPHS1 | 614,19 | 0,06 | 0,03 | 2,10 | 3,57E-02 | 2,16E-01 |
| UTP25 | 822,97 | 0,09 | 0,04 | 2,10 | 3,57E-02 | 2,16E-01 |
| BCAS2 | 410,41 | 0,14 | 0,07 | 2,10 | 3,57E-02 | 2,16E-01 |
| GRPEL1 | 684,63 | 0,08 | 0,04 | 2,10 | 3,58E-02 | 2,16E-01 |
| RRM2 | 73,72 | -0,24 | 0,12 | -2,10 | 3,58E-02 | 2,17E-01 |
| LGALS9C | 39,80 | -0,38 | 0,18 | -2,10 | 3,59E-02 | 2,17E-01 |
| DDX24 | 2854,55 | 0,09 | 0,04 | 2,10 | 3,60E-02 | 2,17E-01 |
| TFCP2L1 | 66,79 | -0,48 | 0,23 | -2,10 | 3,60E-02 | 2,17E-01 |
| DCXR | 328,45 | -0,11 | 0,05 | -2,10 | 3,59E-02 | 2,17E-01 |
| TMEM267 | 372,85 | 0,14 | 0,07 | 2,10 | 3,59E-02 | 2,17E-01 |
| RAB5C | 668,27 | -0,13 | 0,06 | -2,10 | 3,59E-02 | 2,17E-01 |
| SLC38A1 | 10075,20 | 0,12 | 0,06 | 2,10 | 3,59E-02 | 2,17E-01 |
| UBE2G2 | 2950,34 | 0,07 | 0,03 | 2,10 | 3,61E-02 | 2,17E-01 |
| SNX20 | 1521,87 | -0,13 | 0,06 | -2,10 | 3,61E-02 | 2,17E-01 |
| RPF1 | 629,66 | 0,16 | 0,07 | 2,10 | 3,60E-02 | 2,17E-01 |
| RNF103 | 1297,52 | 0,09 | 0,04 | 2,10 | 3,61E-02 | 2,17E-01 |
| SEPTIN7 | 6377,80 | 0,14 | 0,07 | 2,10 | 3,61E-02 | 2,17E-01 |
| PRKAR1B | 147,94 | -0,24 | 0,12 | -2,10 | 3,61E-02 | 2,17E-01 |
| RNFT2 | 66,21 | -0,29 | 0,14 | -2,10 | 3,62E-02 | 2,17E-01 |
| SPARC | 784,51 | -0,31 | 0,15 | -2,10 | 3,62E-02 | 2,17E-01 |
| FAM83D | 40,53 | -0,25 | 0,12 | -2,09 | 3,62E-02 | 2,17E-01 |
| POT1 | 666,25 | 0,19 | 0,09 | 2,09 | 3,63E-02 | 2,17E-01 |
| YTHDC2 | 2416,82 | 0,12 | 0,06 | 2,09 | 3,64E-02 | 2,17E-01 |
| EIF2A | 1512,33 | 0,18 | 0,09 | 2,09 | 3,64E-02 | 2,17E-01 |
| ATG5 | 943,46 | 0,18 | 0,09 | 2,09 | 3,64E-02 | 2,17E-01 |
| FBN1 | 161,81 | -0,40 | 0,19 | -2,09 | 3,64E-02 | 2,17E-01 |
| PTPRB | 60,60 | -0,44 | 0,21 | -2,09 | 3,65E-02 | 2,17E-01 |
| USPL1 | 1198,39 | 0,11 | 0,05 | 2,09 | 3,63E-02 | 2,17E-01 |
| RBM26 | 3871,89 | 0,11 | 0,05 | 2,09 | 3,64E-02 | 2,17E-01 |
| ENSG000002 | 23,94 | -0,34 | 0,16 | -2,09 | 3,64E-02 | 2,17E-01 |

| | | | | | | |
|---|---|---|---|---|---|---|
| PRPF38A | 1367,19 | 0,08 | 0,04 | 2,09 | 3,64E-02 | 2,17E-01 |
| VCF1 | 5009,20 | -0,22 | 0,10 | -2,09 | 3,65E-02 | 2,17E-01 |
| GFPT1 | 1095,32 | 0,09 | 0,04 | 2,09 | 3,64E-02 | 2,17E-01 |
| SMURF2 | 1801,87 | 0,08 | 0,04 | 2,09 | 3,66E-02 | 2,17E-01 |
| RUNDC3A | 717,24 | -0,32 | 0,15 | -2,09 | 3,65E-02 | 2,17E-01 |
| ANKRD13D | 2741,61 | -0,16 | 0,08 | -2,09 | 3,66E-02 | 2,17E-01 |
| SLC25A23 | 63,91 | -0,27 | 0,13 | -2,09 | 3,65E-02 | 2,17E-01 |
| ABAT | 611,98 | -0,17 | 0,08 | -2,09 | 3,65E-02 | 2,17E-01 |
| PHF3 | 6324,81 | 0,13 | 0,06 | 2,09 | 3,66E-02 | 2,17E-01 |
| STRAP | 516,60 | 0,13 | 0,06 | 2,09 | 3,66E-02 | 2,17E-01 |
| KCTD21 | 253,50 | -0,14 | 0,07 | -2,09 | 3,66E-02 | 2,17E-01 |
| MED7 | 486,34 | 0,16 | 0,07 | 2,09 | 3,67E-02 | 2,18E-01 |
| LTBP4 | 275,20 | -0,27 | 0,13 | -2,09 | 3,67E-02 | 2,18E-01 |
| EIF2S1 | 1354,53 | 0,15 | 0,07 | 2,09 | 3,68E-02 | 2,18E-01 |
| SLC39A9 | 1742,76 | 0,06 | 0,03 | 2,09 | 3,68E-02 | 2,18E-01 |
| ZBTB42 | 131,81 | -0,20 | 0,09 | -2,09 | 3,68E-02 | 2,18E-01 |
| C3orf52 | 36,66 | -0,37 | 0,18 | -2,09 | 3,68E-02 | 2,18E-01 |
| ZNF12 | 1712,75 | 0,15 | 0,07 | 2,09 | 3,69E-02 | 2,18E-01 |
| TMEM161B | 1748,34 | 0,14 | 0,07 | 2,09 | 3,69E-02 | 2,18E-01 |
| MRFAP1L2 | 646,30 | 0,10 | 0,05 | 2,09 | 3,70E-02 | 2,18E-01 |
| RPS6KA2 | 326,64 | -0,22 | 0,11 | -2,09 | 3,70E-02 | 2,18E-01 |
| BTBD1 | 1563,48 | 0,11 | 0,05 | 2,09 | 3,70E-02 | 2,18E-01 |
| ZKSCAN8 | 1690,81 | 0,08 | 0,04 | 2,09 | 3,70E-02 | 2,19E-01 |
| MRPS30 | 1440,39 | 0,13 | 0,06 | 2,09 | 3,70E-02 | 2,19E-01 |
| ADK | 899,63 | 0,14 | 0,07 | 2,09 | 3,71E-02 | 2,19E-01 |
| PIGZ | 34,68 | -0,27 | 0,13 | -2,08 | 3,71E-02 | 2,19E-01 |
| ANKMY2 | 441,97 | 0,11 | 0,05 | 2,08 | 3,72E-02 | 2,19E-01 |
| RPS6KC1 | 1137,10 | 0,11 | 0,05 | 2,08 | 3,72E-02 | 2,19E-01 |
| FER | 778,26 | 0,12 | 0,06 | 2,08 | 3,72E-02 | 2,19E-01 |
| EEF1A1 | 125486,52 | 0,22 | 0,11 | 2,08 | 3,72E-02 | 2,19E-01 |
| C2orf69 | 408,91 | 0,15 | 0,07 | 2,08 | 3,73E-02 | 2,19E-01 |
| PFKFB1 | 28,85 | -0,26 | 0,12 | -2,08 | 3,73E-02 | 2,19E-01 |
| EVPL | 77,06 | -0,42 | 0,20 | -2,08 | 3,73E-02 | 2,19E-01 |
| ZBTB11 | 2129,83 | 0,12 | 0,06 | 2,08 | 3,73E-02 | 2,19E-01 |
| ZNF250 | 394,14 | 0,08 | 0,04 | 2,08 | 3,75E-02 | 2,19E-01 |
| CFAP36 | 306,41 | 0,13 | 0,06 | 2,08 | 3,75E-02 | 2,19E-01 |
| TIMM21 | 330,75 | 0,10 | 0,05 | 2,08 | 3,74E-02 | 2,19E-01 |
| NFE2 | 5451,24 | -0,17 | 0,08 | -2,08 | 3,74E-02 | 2,19E-01 |
| RIMS3 | 90,45 | -0,23 | 0,11 | -2,08 | 3,75E-02 | 2,19E-01 |
| TMEM236 | 28,10 | -0,30 | 0,14 | -2,08 | 3,75E-02 | 2,19E-01 |
| SLC35E1 | 758,36 | -0,14 | 0,07 | -2,08 | 3,75E-02 | 2,19E-01 |
| CWC27 | 476,06 | 0,12 | 0,06 | 2,08 | 3,75E-02 | 2,19E-01 |
| PDCD2 | 1579,24 | 0,11 | 0,05 | 2,08 | 3,76E-02 | 2,20E-01 |
| CRNKL1 | 1920,22 | 0,11 | 0,06 | 2,08 | 3,77E-02 | 2,20E-01 |
| RBAK | 827,37 | 0,22 | 0,11 | 2,08 | 3,76E-02 | 2,20E-01 |
| ABCC2 | 204,49 | -0,16 | 0,08 | -2,08 | 3,77E-02 | 2,20E-01 |

| | | | | | | |
|---|---|---|---|---|---|---|
| TM9SF3 | 4243,09 | 0,15 | 0,07 | 2,08 | 3,76E-02 | 2,20E-01 |
| UGP2 | 2290,94 | 0,09 | 0,04 | 2,08 | 3,78E-02 | 2,20E-01 |
| ZNF527 | 310,04 | 0,12 | 0,06 | 2,08 | 3,78E-02 | 2,20E-01 |
| SLC14A1 | 1434,18 | -0,22 | 0,11 | -2,08 | 3,77E-02 | 2,20E-01 |
| IDNK | 276,51 | 0,09 | 0,04 | 2,08 | 3,78E-02 | 2,20E-01 |
| ARHGAP33 | 58,81 | -0,38 | 0,18 | -2,08 | 3,78E-02 | 2,20E-01 |
| FBXO15 | 66,56 | -0,36 | 0,17 | -2,08 | 3,78E-02 | 2,20E-01 |
| AIFM2 | 118,83 | -0,20 | 0,10 | -2,08 | 3,78E-02 | 2,20E-01 |
| PHF14 | 2147,56 | 0,15 | 0,07 | 2,08 | 3,78E-02 | 2,20E-01 |
| TXNDC9 | 483,64 | 0,20 | 0,10 | 2,07 | 3,81E-02 | 2,20E-01 |
| CHERP | 227,49 | -0,20 | 0,10 | -2,07 | 3,80E-02 | 2,20E-01 |
| WDR59 | 889,65 | -0,09 | 0,05 | -2,07 | 3,81E-02 | 2,20E-01 |
| PDGFRB | 100,14 | -0,32 | 0,15 | -2,07 | 3,81E-02 | 2,20E-01 |
| GDPD3 | 102,26 | -0,25 | 0,12 | -2,07 | 3,81E-02 | 2,20E-01 |
| MRPL13 | 273,23 | 0,19 | 0,09 | 2,07 | 3,81E-02 | 2,20E-01 |
| CUL3 | 3456,89 | 0,11 | 0,05 | 2,08 | 3,79E-02 | 2,20E-01 |
| EPN2 | 285,61 | -0,19 | 0,09 | -2,08 | 3,79E-02 | 2,20E-01 |
| HSPA13 | 929,50 | 0,22 | 0,10 | 2,07 | 3,81E-02 | 2,20E-01 |
| EIF3H | 3346,42 | 0,12 | 0,06 | 2,07 | 3,80E-02 | 2,20E-01 |
| CDKAL1 | 1181,10 | 0,10 | 0,05 | 2,07 | 3,81E-02 | 2,20E-01 |
| SETD7 | 1134,28 | 0,09 | 0,04 | 2,07 | 3,81E-02 | 2,20E-01 |
| ZNF260 | 376,61 | 0,21 | 0,10 | 2,07 | 3,80E-02 | 2,20E-01 |
| SOCS6 | 229,26 | 0,15 | 0,07 | 2,07 | 3,81E-02 | 2,20E-01 |
| FOXD4L1 | 24,80 | -0,23 | 0,11 | -2,08 | 3,80E-02 | 2,20E-01 |
| ADGRL1 | 119,64 | -0,32 | 0,16 | -2,07 | 3,81E-02 | 2,20E-01 |
| SRRM1 | 1927,10 | 0,08 | 0,04 | 2,07 | 3,82E-02 | 2,20E-01 |
| FBL | 960,60 | 0,13 | 0,06 | 2,07 | 3,83E-02 | 2,20E-01 |
| TSPAN13 | 173,26 | 0,18 | 0,09 | 2,07 | 3,82E-02 | 2,20E-01 |
| CENPV | 90,15 | -0,15 | 0,07 | -2,07 | 3,83E-02 | 2,20E-01 |
| GINS3 | 66,06 | -0,19 | 0,09 | -2,07 | 3,82E-02 | 2,20E-01 |
| TRIM37 | 900,61 | 0,09 | 0,05 | 2,07 | 3,83E-02 | 2,20E-01 |
| USP47 | 3230,75 | 0,15 | 0,07 | 2,07 | 3,83E-02 | 2,20E-01 |
| NHLRC2 | 2039,68 | 0,12 | 0,06 | 2,07 | 3,86E-02 | 2,21E-01 |
| SNX21 | 145,76 | -0,16 | 0,08 | -2,07 | 3,85E-02 | 2,21E-01 |
| ZNF770 | 2436,50 | 0,14 | 0,07 | 2,07 | 3,85E-02 | 2,21E-01 |
| PIK3R1 | 5045,89 | 0,11 | 0,05 | 2,07 | 3,85E-02 | 2,21E-01 |
| CEP120 | 1968,76 | 0,12 | 0,06 | 2,07 | 3,85E-02 | 2,21E-01 |
| VPS26B | 1270,16 | -0,09 | 0,04 | -2,07 | 3,85E-02 | 2,21E-01 |
| FAM135A | 808,62 | 0,14 | 0,07 | 2,07 | 3,85E-02 | 2,21E-01 |
| NRAS | 1557,59 | 0,13 | 0,06 | 2,07 | 3,86E-02 | 2,21E-01 |
| CDK16 | 403,47 | -0,12 | 0,06 | -2,07 | 3,86E-02 | 2,21E-01 |
| COMMD8 | 313,81 | 0,30 | 0,15 | 2,07 | 3,85E-02 | 2,21E-01 |
| ALG1L2 | 66,21 | -0,27 | 0,13 | -2,07 | 3,85E-02 | 2,21E-01 |
| CD8B | 772,23 | 0,26 | 0,12 | 2,07 | 3,85E-02 | 2,21E-01 |
| CENPH | 85,18 | 0,16 | 0,08 | 2,07 | 3,85E-02 | 2,21E-01 |
| TNK1 | 34,15 | -0,32 | 0,15 | -2,07 | 3,87E-02 | 2,21E-01 |

| | | | | | | |
|---|---|---|---|---|---|---|
| EPAS1 | 134,59 | -0,25 | 0,12 | -2,07 | 3,88E-02 | 2,21E-01 |
| WDR97 | 80,48 | -0,32 | 0,15 | -2,07 | 3,88E-02 | 2,21E-01 |
| DBF4 | 377,38 | 0,14 | 0,07 | 2,07 | 3,87E-02 | 2,21E-01 |
| FLCN | 1305,80 | -0,13 | 0,06 | -2,07 | 3,87E-02 | 2,21E-01 |
| SPPL3 | 827,62 | -0,11 | 0,05 | -2,07 | 3,87E-02 | 2,21E-01 |
| ZDHHC12 | 93,34 | -0,20 | 0,10 | -2,07 | 3,88E-02 | 2,21E-01 |
| ZDHHC1 | 57,07 | -0,27 | 0,13 | -2,07 | 3,87E-02 | 2,21E-01 |
| UGCG | 1521,86 | 0,17 | 0,08 | 2,07 | 3,87E-02 | 2,21E-01 |
| RNGTT | 1394,36 | 0,10 | 0,05 | 2,07 | 3,88E-02 | 2,21E-01 |
| SLC16A7 | 3027,32 | 0,14 | 0,07 | 2,07 | 3,88E-02 | 2,21E-01 |
| GTF2E2 | 822,59 | 0,07 | 0,03 | 2,07 | 3,88E-02 | 2,21E-01 |
| PPHLN1 | 2074,22 | 0,08 | 0,04 | 2,07 | 3,89E-02 | 2,21E-01 |
| PSMD14 | 649,32 | 0,17 | 0,08 | 2,07 | 3,89E-02 | 2,21E-01 |
| PYURF | 706,93 | 0,17 | 0,08 | 2,07 | 3,88E-02 | 2,21E-01 |
| RNF185 | 804,20 | -0,09 | 0,04 | -2,07 | 3,89E-02 | 2,21E-01 |
| SRSF3 | 7379,31 | 0,13 | 0,06 | 2,07 | 3,89E-02 | 2,21E-01 |
| MAPK6 | 692,28 | 0,12 | 0,06 | 2,07 | 3,89E-02 | 2,21E-01 |
| PPP4R3A | 4825,15 | 0,09 | 0,04 | 2,06 | 3,89E-02 | 2,21E-01 |
| KIAA0586 | 1322,49 | 0,10 | 0,05 | 2,06 | 3,90E-02 | 2,21E-01 |
| LONP2 | 3628,51 | 0,08 | 0,04 | 2,06 | 3,90E-02 | 2,21E-01 |
| IP6K1 | 1449,32 | -0,16 | 0,08 | -2,06 | 3,91E-02 | 2,21E-01 |
| MEAF6 | 1110,87 | 0,09 | 0,04 | 2,06 | 3,91E-02 | 2,21E-01 |
| KCNC3 | 62,64 | -0,36 | 0,17 | -2,06 | 3,91E-02 | 2,21E-01 |
| IPO8 | 2322,77 | 0,09 | 0,04 | 2,06 | 3,92E-02 | 2,21E-01 |
| PTER | 587,85 | 0,13 | 0,06 | 2,06 | 3,92E-02 | 2,21E-01 |
| TNFAIP8 | 5064,98 | 0,11 | 0,05 | 2,06 | 3,92E-02 | 2,21E-01 |
| DYNLT3 | 385,04 | 0,26 | 0,12 | 2,06 | 3,92E-02 | 2,21E-01 |
| FBXO3 | 2881,29 | 0,11 | 0,05 | 2,06 | 3,93E-02 | 2,22E-01 |
| ALG12 | 389,85 | -0,10 | 0,05 | -2,06 | 3,94E-02 | 2,22E-01 |
| CBL | 4416,19 | -0,19 | 0,09 | -2,06 | 3,93E-02 | 2,22E-01 |
| TRIM46 | 58,33 | -0,31 | 0,15 | -2,06 | 3,94E-02 | 2,22E-01 |
| TMEM123 | 7039,70 | 0,20 | 0,10 | 2,06 | 3,94E-02 | 2,22E-01 |
| BBS10 | 628,46 | 0,17 | 0,08 | 2,06 | 3,95E-02 | 2,22E-01 |
| MAP2 | 45,08 | -0,40 | 0,19 | -2,06 | 3,95E-02 | 2,22E-01 |
| DHRS1 | 194,28 | -0,13 | 0,06 | -2,06 | 3,95E-02 | 2,22E-01 |
| GBGT1 | 562,93 | -0,14 | 0,07 | -2,06 | 3,95E-02 | 2,22E-01 |
| API5 | 2581,50 | 0,12 | 0,06 | 2,06 | 3,95E-02 | 2,22E-01 |
| KRBA1 | 131,51 | -0,24 | 0,12 | -2,06 | 3,95E-02 | 2,22E-01 |
| PHF13 | 230,43 | -0,11 | 0,05 | -2,06 | 3,96E-02 | 2,22E-01 |
| ZNF805 | 674,11 | 0,08 | 0,04 | 2,06 | 3,96E-02 | 2,22E-01 |
| DMAC1 | 582,07 | 0,17 | 0,08 | 2,06 | 3,97E-02 | 2,22E-01 |
| INSC | 34,47 | -0,39 | 0,19 | -2,05 | 3,99E-02 | 2,22E-01 |
| PDGFA | 133,16 | -0,28 | 0,14 | -2,06 | 3,97E-02 | 2,22E-01 |
| FBLN2 | 60,20 | -0,39 | 0,19 | -2,05 | 4,00E-02 | 2,22E-01 |
| ORC5 | 645,98 | 0,14 | 0,07 | 2,05 | 4,00E-02 | 2,22E-01 |
| PRUNE2 | 96,14 | -0,42 | 0,21 | -2,06 | 3,98E-02 | 2,22E-01 |

| | | | | | | |
|---|---|---|---|---|---|---|
| ALG10 | 563,74 | 0,12 | 0,06 | 2,06 | 3,97E-02 | 2,22E-01 |
| GABPA | 1776,54 | 0,17 | 0,08 | 2,06 | 3,97E-02 | 2,22E-01 |
| HYKK | 40,27 | -0,30 | 0,15 | -2,06 | 3,97E-02 | 2,22E-01 |
| EVI2A | 1893,70 | 0,36 | 0,18 | 2,05 | 4,00E-02 | 2,22E-01 |
| ZNF319 | 327,62 | -0,18 | 0,09 | -2,05 | 3,99E-02 | 2,22E-01 |
| MAPRE3 | 73,38 | -0,26 | 0,13 | -2,06 | 3,98E-02 | 2,22E-01 |
| SLC35E4 | 62,81 | -0,28 | 0,14 | -2,05 | 4,00E-02 | 2,22E-01 |
| ATXN7L2 | 57,56 | -0,31 | 0,15 | -2,06 | 3,97E-02 | 2,22E-01 |
| ZC3HAV1L | 83,02 | -0,21 | 0,10 | -2,05 | 4,00E-02 | 2,22E-01 |
| PREPL | 1375,55 | 0,10 | 0,05 | 2,05 | 3,99E-02 | 2,22E-01 |
| EMB | 7356,96 | 0,15 | 0,07 | 2,05 | 3,99E-02 | 2,22E-01 |
| TSPAN9 | 108,61 | -0,38 | 0,19 | -2,06 | 3,98E-02 | 2,22E-01 |
| NAV3 | 99,53 | -0,35 | 0,17 | -2,05 | 3,99E-02 | 2,22E-01 |
| PNPLA2 | 878,60 | -0,24 | 0,12 | -2,05 | 3,99E-02 | 2,22E-01 |
| PANX2 | 121,86 | -0,34 | 0,17 | -2,06 | 3,98E-02 | 2,22E-01 |
| ITLN1 | 104,22 | -0,36 | 0,18 | -2,05 | 3,99E-02 | 2,22E-01 |
| TRIM4 | 1319,28 | 0,06 | 0,03 | 2,05 | 4,00E-02 | 2,22E-01 |
| THOC5 | 1641,81 | -0,14 | 0,07 | -2,05 | 4,01E-02 | 2,22E-01 |
| KMT5C | 129,97 | -0,21 | 0,10 | -2,05 | 4,01E-02 | 2,22E-01 |
| PTPRG | 76,82 | -0,50 | 0,25 | -2,05 | 4,01E-02 | 2,23E-01 |
| NRL | 72,87 | -0,14 | 0,07 | -2,05 | 4,02E-02 | 2,23E-01 |
| SZRD1 | 1450,14 | -0,10 | 0,05 | -2,05 | 4,02E-02 | 2,23E-01 |
| ERG | 58,58 | -0,44 | 0,21 | -2,05 | 4,03E-02 | 2,23E-01 |
| FCF1 | 879,26 | 0,11 | 0,05 | 2,05 | 4,03E-02 | 2,23E-01 |
| CHD1 | 5930,94 | 0,14 | 0,07 | 2,05 | 4,03E-02 | 2,23E-01 |
| CDK15 | 43,81 | -0,42 | 0,21 | -2,05 | 4,03E-02 | 2,23E-01 |
| NIPAL2 | 536,32 | 0,19 | 0,09 | 2,05 | 4,04E-02 | 2,23E-01 |
| HEXD | 388,18 | -0,18 | 0,09 | -2,05 | 4,04E-02 | 2,23E-01 |
| CYSLTR1 | 956,01 | 0,14 | 0,07 | 2,05 | 4,04E-02 | 2,23E-01 |
| ANKRD49 | 741,09 | 0,21 | 0,10 | 2,05 | 4,04E-02 | 2,23E-01 |
| TNKS1BP1 | 119,00 | -0,28 | 0,14 | -2,05 | 4,04E-02 | 2,23E-01 |
| SAP30L | 1811,36 | -0,06 | 0,03 | -2,05 | 4,05E-02 | 2,23E-01 |
| GIMAP7 | 2768,56 | 0,18 | 0,09 | 2,05 | 4,05E-02 | 2,23E-01 |
| MPHOSPH6 | 423,71 | 0,14 | 0,07 | 2,05 | 4,05E-02 | 2,23E-01 |
| ATG9A | 453,99 | -0,15 | 0,07 | -2,05 | 4,05E-02 | 2,23E-01 |
| UNC13B | 111,89 | -0,39 | 0,19 | -2,05 | 4,06E-02 | 2,23E-01 |
| PCDHGB2 | 31,89 | -0,26 | 0,13 | -2,05 | 4,06E-02 | 2,23E-01 |
| RSF1 | 3269,65 | 0,10 | 0,05 | 2,05 | 4,06E-02 | 2,23E-01 |
| VMA21 | 1328,69 | 0,12 | 0,06 | 2,05 | 4,06E-02 | 2,23E-01 |
| C17orf58 | 31,54 | -0,24 | 0,11 | -2,05 | 4,06E-02 | 2,23E-01 |
| PAIP1 | 746,43 | 0,12 | 0,06 | 2,05 | 4,06E-02 | 2,23E-01 |
| PIGS | 547,93 | -0,10 | 0,05 | -2,05 | 4,07E-02 | 2,23E-01 |
| PLRG1 | 1053,57 | 0,15 | 0,08 | 2,05 | 4,07E-02 | 2,23E-01 |
| TATDN3 | 532,23 | 0,10 | 0,05 | 2,05 | 4,07E-02 | 2,23E-01 |
| HMGN1 | 2811,34 | 0,11 | 0,05 | 2,05 | 4,08E-02 | 2,23E-01 |
| POLR2J3.1 | 118,31 | -0,30 | 0,15 | -2,05 | 4,08E-02 | 2,23E-01 |

| | | | | | | |
|---|---|---|---|---|---|---|
| BMPR2 | 985,29 | 0,11 | 0,06 | 2,04 | 4,09E-02 | 2,24E-01 |
| TCEAL8 | 248,78 | 0,21 | 0,10 | 2,04 | 4,09E-02 | 2,24E-01 |
| ESD | 901,93 | 0,12 | 0,06 | 2,05 | 4,08E-02 | 2,24E-01 |
| ESCO1 | 1243,50 | 0,20 | 0,10 | 2,05 | 4,08E-02 | 2,24E-01 |
| NIPSNAP2 | 796,58 | 0,08 | 0,04 | 2,04 | 4,09E-02 | 2,24E-01 |
| FAM222B | 111,86 | -0,19 | 0,09 | -2,04 | 4,09E-02 | 2,24E-01 |
| GANC | 1668,36 | 0,08 | 0,04 | 2,04 | 4,09E-02 | 2,24E-01 |
| NQO2 | 1434,56 | -0,19 | 0,09 | -2,04 | 4,09E-02 | 2,24E-01 |
| USP1 | 1483,13 | 0,19 | 0,09 | 2,04 | 4,10E-02 | 2,24E-01 |
| CARNMT1 | 777,55 | 0,15 | 0,07 | 2,04 | 4,10E-02 | 2,24E-01 |
| ZNF530 | 261,90 | 0,08 | 0,04 | 2,04 | 4,12E-02 | 2,25E-01 |
| TMEM242 | 426,48 | 0,08 | 0,04 | 2,04 | 4,12E-02 | 2,25E-01 |
| MAP1LC3A | 77,34 | -0,23 | 0,11 | -2,04 | 4,12E-02 | 2,25E-01 |
| RHOBTB1 | 205,81 | -0,24 | 0,12 | -2,04 | 4,12E-02 | 2,25E-01 |
| CCNA2 | 67,28 | -0,18 | 0,09 | -2,04 | 4,12E-02 | 2,25E-01 |
| SRSF11 | 6022,96 | 0,12 | 0,06 | 2,04 | 4,13E-02 | 2,25E-01 |
| ENSG000002 | 395,52 | -0,14 | 0,07 | -2,04 | 4,13E-02 | 2,25E-01 |
| PARD3 | 111,80 | -0,28 | 0,14 | -2,04 | 4,14E-02 | 2,25E-01 |
| ZDHHC18 | 5264,22 | -0,17 | 0,08 | -2,04 | 4,14E-02 | 2,25E-01 |
| RGS12 | 289,32 | -0,22 | 0,11 | -2,04 | 4,14E-02 | 2,25E-01 |
| CERS2 | 2225,92 | -0,09 | 0,05 | -2,04 | 4,14E-02 | 2,25E-01 |
| RBBP7 | 1219,29 | 0,08 | 0,04 | 2,04 | 4,15E-02 | 2,25E-01 |
| RPS20 | 8996,88 | 0,24 | 0,12 | 2,04 | 4,15E-02 | 2,25E-01 |
| RAB11FIP5 | 145,24 | -0,26 | 0,13 | -2,04 | 4,15E-02 | 2,25E-01 |
| RAB6B | 105,99 | -0,31 | 0,15 | -2,04 | 4,15E-02 | 2,25E-01 |
| LRSAM1 | 465,71 | -0,15 | 0,08 | -2,04 | 4,15E-02 | 2,25E-01 |
| LGALS2 | 504,69 | 0,32 | 0,16 | 2,04 | 4,16E-02 | 2,25E-01 |
| ETF1 | 1864,85 | 0,09 | 0,05 | 2,04 | 4,16E-02 | 2,25E-01 |
| UBA6 | 2699,49 | 0,14 | 0,07 | 2,04 | 4,17E-02 | 2,25E-01 |
| MYOM1 | 222,45 | -0,15 | 0,07 | -2,04 | 4,17E-02 | 2,25E-01 |
| MALT1 | 3630,03 | 0,11 | 0,05 | 2,04 | 4,17E-02 | 2,25E-01 |
| SMAD4 | 4316,22 | 0,08 | 0,04 | 2,04 | 4,17E-02 | 2,25E-01 |
| DNAJC8 | 1128,91 | 0,13 | 0,06 | 2,03 | 4,19E-02 | 2,25E-01 |
| NAPSA | 35,99 | -0,32 | 0,16 | -2,04 | 4,18E-02 | 2,25E-01 |
| TSEN15 | 921,17 | 0,12 | 0,06 | 2,03 | 4,19E-02 | 2,25E-01 |
| POLR2M | 666,40 | 0,17 | 0,08 | 2,04 | 4,18E-02 | 2,25E-01 |
| INSIG2 | 804,09 | 0,14 | 0,07 | 2,03 | 4,19E-02 | 2,25E-01 |
| PPIG | 2150,00 | 0,21 | 0,10 | 2,04 | 4,18E-02 | 2,25E-01 |
| PPP2R3A | 42,66 | -0,35 | 0,17 | -2,04 | 4,18E-02 | 2,25E-01 |
| MICU2 | 1377,22 | 0,14 | 0,07 | 2,04 | 4,18E-02 | 2,25E-01 |
| RINT1 | 544,44 | 0,07 | 0,03 | 2,04 | 4,18E-02 | 2,25E-01 |
| RNF167 | 374,81 | -0,15 | 0,08 | -2,03 | 4,19E-02 | 2,25E-01 |
| DLL1 | 59,82 | -0,31 | 0,15 | -2,03 | 4,19E-02 | 2,25E-01 |
| GUK1 | 25649,81 | -0,25 | 0,12 | -2,03 | 4,19E-02 | 2,25E-01 |
| SCOC | 348,05 | 0,24 | 0,12 | 2,03 | 4,20E-02 | 2,26E-01 |
| GINM1 | 650,25 | 0,08 | 0,04 | 2,03 | 4,20E-02 | 2,26E-01 |

| | | | | | | |
|---|---|---|---|---|---|---|
| BICRAL | 1725,09 | -0,10 | 0,05 | -2,03 | 4,20E-02 | 2,26E-01 |
| ZNF497 | 28,17 | -0,28 | 0,14 | -2,03 | 4,20E-02 | 2,26E-01 |
| POC1A | 62,99 | -0,17 | 0,08 | -2,03 | 4,20E-02 | 2,26E-01 |
| DDX46 | 2533,05 | 0,08 | 0,04 | 2,03 | 4,23E-02 | 2,26E-01 |
| CILP | 38,17 | -0,30 | 0,15 | -2,03 | 4,22E-02 | 2,26E-01 |
| VAMP4 | 915,06 | 0,17 | 0,08 | 2,03 | 4,23E-02 | 2,26E-01 |
| NAA25 | 1186,78 | 0,08 | 0,04 | 2,03 | 4,24E-02 | 2,26E-01 |
| PRIMPOL | 436,10 | 0,10 | 0,05 | 2,03 | 4,22E-02 | 2,26E-01 |
| RABL3 | 552,69 | 0,12 | 0,06 | 2,03 | 4,22E-02 | 2,26E-01 |
| NUDT21 | 1835,01 | 0,16 | 0,08 | 2,03 | 4,22E-02 | 2,26E-01 |
| MSRB3 | 193,30 | -0,19 | 0,09 | -2,03 | 4,22E-02 | 2,26E-01 |
| PDHX | 454,17 | 0,10 | 0,05 | 2,03 | 4,23E-02 | 2,26E-01 |
| CFAP161 | 65,63 | -0,20 | 0,10 | -2,03 | 4,23E-02 | 2,26E-01 |
| ZCCHC10 | 215,14 | 0,21 | 0,11 | 2,03 | 4,23E-02 | 2,26E-01 |
| PMM1 | 162,56 | -0,12 | 0,06 | -2,03 | 4,22E-02 | 2,26E-01 |
| SNAPC2 | 62,80 | -0,19 | 0,09 | -2,03 | 4,24E-02 | 2,26E-01 |
| LPIN2 | 4448,07 | -0,13 | 0,07 | -2,03 | 4,23E-02 | 2,26E-01 |
| EBAG9 | 446,88 | 0,14 | 0,07 | 2,03 | 4,24E-02 | 2,26E-01 |
| TSC22D1 | 1189,33 | -0,12 | 0,06 | -2,03 | 4,24E-02 | 2,26E-01 |
| SELENBP1 | 3897,78 | -0,31 | 0,15 | -2,03 | 4,21E-02 | 2,26E-01 |
| PARP3 | 68,57 | -0,20 | 0,10 | -2,02 | 4,29E-02 | 2,26E-01 |
| EIF3D | 2915,91 | 0,09 | 0,04 | 2,03 | 4,29E-02 | 2,26E-01 |
| NSF | 757,03 | 0,08 | 0,04 | 2,03 | 4,27E-02 | 2,26E-01 |
| TMEM252 | 110,07 | -0,20 | 0,10 | -2,03 | 4,25E-02 | 2,26E-01 |
| REV1 | 1769,52 | 0,08 | 0,04 | 2,03 | 4,29E-02 | 2,26E-01 |
| ATP13A4 | 111,81 | -0,26 | 0,13 | -2,03 | 4,27E-02 | 2,26E-01 |
| DUSP11 | 1135,57 | 0,08 | 0,04 | 2,03 | 4,26E-02 | 2,26E-01 |
| SLC25A46 | 2029,92 | 0,16 | 0,08 | 2,03 | 4,28E-02 | 2,26E-01 |
| ALAS1 | 447,68 | -0,12 | 0,06 | -2,03 | 4,26E-02 | 2,26E-01 |
| CYB5B | 1061,48 | 0,08 | 0,04 | 2,03 | 4,25E-02 | 2,26E-01 |
| PEX2 | 1359,01 | 0,14 | 0,07 | 2,03 | 4,28E-02 | 2,26E-01 |
| ANKRD9 | 487,22 | -0,28 | 0,14 | -2,03 | 4,24E-02 | 2,26E-01 |
| FLT3LG | 87,75 | -0,28 | 0,14 | -2,03 | 4,27E-02 | 2,26E-01 |
| BAMBI | 26,16 | -0,35 | 0,17 | -2,03 | 4,28E-02 | 2,26E-01 |
| TTC14 | 3661,70 | 0,16 | 0,08 | 2,03 | 4,26E-02 | 2,26E-01 |
| ZFPM1 | 169,03 | -0,29 | 0,14 | -2,03 | 4,27E-02 | 2,26E-01 |
| TAF11L2 | 29,29 | -0,26 | 0,13 | -2,03 | 4,25E-02 | 2,26E-01 |
| TRMT5 | 218,27 | 0,12 | 0,06 | 2,03 | 4,26E-02 | 2,26E-01 |
| XPO1 | 5175,93 | 0,11 | 0,05 | 2,03 | 4,25E-02 | 2,26E-01 |
| ABCF2 | 72,00 | -0,14 | 0,07 | -2,03 | 4,24E-02 | 2,26E-01 |
| GSKIP | 601,86 | 0,19 | 0,09 | 2,03 | 4,28E-02 | 2,26E-01 |
| AEBP2 | 1436,61 | 0,12 | 0,06 | 2,03 | 4,26E-02 | 2,26E-01 |
| SMOX | 337,76 | -0,27 | 0,13 | -2,03 | 4,27E-02 | 2,26E-01 |
| SH3TC2 | 132,66 | -0,33 | 0,17 | -2,03 | 4,27E-02 | 2,26E-01 |
| UBLCP1 | 945,08 | 0,15 | 0,07 | 2,03 | 4,28E-02 | 2,26E-01 |
| NT5C3B | 180,92 | -0,18 | 0,09 | -2,03 | 4,26E-02 | 2,26E-01 |

| | | | | | | |
|---|---|---|---|---|---|---|
| ST6GALNAC1 | 44,16 | -0,29 | 0,14 | -2,03 | 4,28E-02 | 2,26E-01 |
| NAE1 | 673,00 | 0,12 | 0,06 | 2,02 | 4,30E-02 | 2,26E-01 |
| CLECL1P | 128,96 | 0,26 | 0,13 | 2,02 | 4,30E-02 | 2,26E-01 |
| SMIM29 | 207,53 | -0,09 | 0,05 | -2,02 | 4,30E-02 | 2,26E-01 |
| ALG2 | 604,52 | 0,07 | 0,04 | 2,02 | 4,30E-02 | 2,26E-01 |
| ZNF594 | 579,94 | 0,19 | 0,09 | 2,02 | 4,30E-02 | 2,26E-01 |
| ERMARD | 923,76 | 0,07 | 0,04 | 2,02 | 4,30E-02 | 2,26E-01 |
| TAS2R50 | 92,97 | 0,19 | 0,09 | 2,02 | 4,31E-02 | 2,26E-01 |
| FAM210B | 8820,76 | -0,28 | 0,14 | -2,02 | 4,31E-02 | 2,26E-01 |
| ABRACL | 638,23 | 0,16 | 0,08 | 2,02 | 4,31E-02 | 2,26E-01 |
| RGS14 | 1400,39 | -0,16 | 0,08 | -2,02 | 4,31E-02 | 2,26E-01 |
| FAM43A | 425,47 | 0,11 | 0,06 | 2,02 | 4,32E-02 | 2,26E-01 |
| USP35 | 183,67 | -0,18 | 0,09 | -2,02 | 4,32E-02 | 2,26E-01 |
| BBX | 4138,41 | 0,09 | 0,05 | 2,02 | 4,33E-02 | 2,26E-01 |
| NUTM2D | 38,91 | -0,25 | 0,12 | -2,02 | 4,33E-02 | 2,27E-01 |
| NUP35 | 435,55 | 0,12 | 0,06 | 2,02 | 4,33E-02 | 2,27E-01 |
| ITGA1 | 264,89 | -0,19 | 0,09 | -2,02 | 4,34E-02 | 2,27E-01 |
| GCFC2 | 601,54 | 0,13 | 0,06 | 2,02 | 4,34E-02 | 2,27E-01 |
| RNASE3 | 44,65 | -0,42 | 0,21 | -2,02 | 4,35E-02 | 2,27E-01 |
| BTG1 | 5092,92 | 0,13 | 0,06 | 2,02 | 4,34E-02 | 2,27E-01 |
| CCDC91 | 1533,86 | 0,13 | 0,06 | 2,02 | 4,35E-02 | 2,27E-01 |
| TMEM184C | 1197,45 | 0,10 | 0,05 | 2,02 | 4,35E-02 | 2,27E-01 |
| ILF2 | 1325,40 | 0,10 | 0,05 | 2,02 | 4,36E-02 | 2,27E-01 |
| TIMM17A | 574,68 | 0,11 | 0,06 | 2,02 | 4,36E-02 | 2,27E-01 |
| FAM118B | 553,94 | 0,10 | 0,05 | 2,02 | 4,37E-02 | 2,27E-01 |
| MOSPD3 | 49,55 | -0,16 | 0,08 | -2,02 | 4,37E-02 | 2,27E-01 |
| PABPC1L | 335,66 | -0,22 | 0,11 | -2,02 | 4,37E-02 | 2,27E-01 |
| TSR1 | 751,94 | 0,07 | 0,03 | 2,02 | 4,36E-02 | 2,27E-01 |
| MAST2 | 296,33 | -0,16 | 0,08 | -2,02 | 4,37E-02 | 2,27E-01 |
| SIGLEC5 | 76,31 | -0,23 | 0,11 | -2,02 | 4,37E-02 | 2,27E-01 |
| KLHL15 | 622,20 | 0,11 | 0,06 | 2,02 | 4,37E-02 | 2,27E-01 |
| TSPAN5 | 3841,22 | -0,24 | 0,12 | -2,02 | 4,36E-02 | 2,27E-01 |
| ZNF416 | 228,11 | 0,11 | 0,06 | 2,02 | 4,39E-02 | 2,27E-01 |
| MRPS18B | 563,93 | 0,09 | 0,05 | 2,01 | 4,39E-02 | 2,27E-01 |
| C5orf24 | 1193,60 | 0,11 | 0,06 | 2,02 | 4,39E-02 | 2,27E-01 |
| TTLL12 | 228,95 | -0,17 | 0,09 | -2,01 | 4,40E-02 | 2,27E-01 |
| OCM | 21,41 | -0,23 | 0,11 | -2,02 | 4,38E-02 | 2,27E-01 |
| HSPA2 | 35,57 | -0,27 | 0,13 | -2,01 | 4,39E-02 | 2,27E-01 |
| PNPLA6 | 939,83 | -0,21 | 0,11 | -2,02 | 4,38E-02 | 2,27E-01 |
| MMGT1 | 756,22 | 0,09 | 0,05 | 2,01 | 4,39E-02 | 2,27E-01 |
| SETD3 | 1537,42 | 0,09 | 0,04 | 2,01 | 4,39E-02 | 2,27E-01 |
| STXBP1 | 103,91 | -0,27 | 0,14 | -2,01 | 4,39E-02 | 2,27E-01 |
| TNIP1 | 3835,13 | -0,14 | 0,07 | -2,01 | 4,40E-02 | 2,27E-01 |
| ADA | 319,23 | -0,11 | 0,05 | -2,01 | 4,40E-02 | 2,27E-01 |
| CEP44 | 958,26 | 0,13 | 0,06 | 2,01 | 4,41E-02 | 2,28E-01 |
| DLAT | 370,16 | 0,07 | 0,04 | 2,01 | 4,41E-02 | 2,28E-01 |

| | | | | | | |
|---|---|---|---|---|---|---|
| MLST8 | 149,90 | -0,13 | 0,06 | -2,01 | 4,43E-02 | 2,29E-01 |
| ZNF658 | 165,17 | 0,20 | 0,10 | 2,01 | 4,43E-02 | 2,29E-01 |
| PLCB3 | 164,65 | -0,21 | 0,10 | -2,01 | 4,43E-02 | 2,29E-01 |
| HEATR1 | 2082,83 | 0,09 | 0,05 | 2,01 | 4,48E-02 | 2,29E-01 |
| C1orf159 | 183,02 | -0,20 | 0,10 | -2,01 | 4,48E-02 | 2,29E-01 |
| HAUS3 | 964,07 | 0,12 | 0,06 | 2,01 | 4,45E-02 | 2,29E-01 |
| TUBG1 | 79,46 | -0,12 | 0,06 | -2,01 | 4,46E-02 | 2,29E-01 |
| CASD1 | 1547,56 | 0,17 | 0,08 | 2,01 | 4,49E-02 | 2,29E-01 |
| NAGS | 58,63 | -0,18 | 0,09 | -2,01 | 4,49E-02 | 2,29E-01 |
| ZHX1 | 819,84 | 0,16 | 0,08 | 2,01 | 4,46E-02 | 2,29E-01 |
| AQP10 | 61,09 | -0,34 | 0,17 | -2,01 | 4,46E-02 | 2,29E-01 |
| PGBD2 | 479,67 | 0,08 | 0,04 | 2,01 | 4,44E-02 | 2,29E-01 |
| ANKRA2 | 918,72 | 0,11 | 0,06 | 2,00 | 4,50E-02 | 2,29E-01 |
| ADD3 | 6973,02 | 0,11 | 0,06 | 2,01 | 4,48E-02 | 2,29E-01 |
| EMG1 | 982,75 | 0,08 | 0,04 | 2,01 | 4,48E-02 | 2,29E-01 |
| MINDY3 | 1115,84 | 0,14 | 0,07 | 2,01 | 4,50E-02 | 2,29E-01 |
| USP27X | 102,74 | 0,10 | 0,05 | 2,01 | 4,45E-02 | 2,29E-01 |
| USP45 | 571,63 | 0,15 | 0,08 | 2,01 | 4,47E-02 | 2,29E-01 |
| UBE2L3 | 1016,58 | -0,06 | 0,03 | -2,01 | 4,48E-02 | 2,29E-01 |
| VPS54 | 1125,91 | 0,11 | 0,06 | 2,01 | 4,49E-02 | 2,29E-01 |
| ZBTB24 | 1226,72 | 0,09 | 0,05 | 2,00 | 4,50E-02 | 2,29E-01 |
| MIOS | 1097,11 | 0,14 | 0,07 | 2,01 | 4,49E-02 | 2,29E-01 |
| ZNF813 | 304,42 | 0,18 | 0,09 | 2,01 | 4,44E-02 | 2,29E-01 |
| GLO1 | 650,14 | 0,14 | 0,07 | 2,01 | 4,49E-02 | 2,29E-01 |
| PAIP2B | 388,51 | 0,12 | 0,06 | 2,01 | 4,46E-02 | 2,29E-01 |
| ALKBH6 | 102,03 | -0,12 | 0,06 | -2,00 | 4,50E-02 | 2,29E-01 |
| EGF | 186,90 | -0,27 | 0,14 | -2,01 | 4,47E-02 | 2,29E-01 |
| TRIM44 | 2532,61 | 0,07 | 0,04 | 2,01 | 4,48E-02 | 2,29E-01 |
| CHAMP1 | 255,85 | 0,08 | 0,04 | 2,00 | 4,50E-02 | 2,29E-01 |
| HAVCR2 | 737,22 | 0,12 | 0,06 | 2,00 | 4,50E-02 | 2,29E-01 |
| ESF1 | 332,13 | 0,22 | 0,11 | 2,01 | 4,47E-02 | 2,29E-01 |
| CCDC77 | 250,97 | 0,09 | 0,04 | 2,01 | 4,46E-02 | 2,29E-01 |
| C14orf28 | 199,71 | 0,15 | 0,08 | 2,01 | 4,46E-02 | 2,29E-01 |
| PCIF1 | 936,50 | -0,12 | 0,06 | -2,01 | 4,46E-02 | 2,29E-01 |
| UTP11 | 477,73 | 0,09 | 0,04 | 2,01 | 4,47E-02 | 2,29E-01 |
| ISL2 | 108,66 | -0,26 | 0,13 | -2,01 | 4,44E-02 | 2,29E-01 |
| DNAH7 | 95,66 | -0,36 | 0,18 | -2,01 | 4,48E-02 | 2,29E-01 |
| MSANTD7 | 876,03 | 0,08 | 0,04 | 2,00 | 4,50E-02 | 2,29E-01 |
| TTC21B | 1437,19 | 0,12 | 0,06 | 2,00 | 4,51E-02 | 2,29E-01 |
| IPO13 | 212,63 | -0,17 | 0,09 | -2,00 | 4,51E-02 | 2,29E-01 |
| CD200R1 | 693,74 | 0,16 | 0,08 | 2,00 | 4,53E-02 | 2,29E-01 |
| CTSB | 10081,08 | -0,17 | 0,08 | -2,00 | 4,53E-02 | 2,29E-01 |
| LLGL1 | 296,30 | -0,16 | 0,08 | -2,00 | 4,52E-02 | 2,29E-01 |
| BLM | 317,64 | 0,10 | 0,05 | 2,00 | 4,52E-02 | 2,29E-01 |
| YIPF2 | 339,87 | -0,10 | 0,05 | -2,00 | 4,53E-02 | 2,29E-01 |
| NEK6 | 1683,32 | -0,14 | 0,07 | -2,00 | 4,53E-02 | 2,29E-01 |

| | | | | | | |
|---|---|---|---|---|---|---|
| MRPL35 | 506,84 | 0,12 | 0,06 | 2,00 | 4,52E-02 | 2,29E-01 |
| SMU1 | 2386,37 | 0,07 | 0,04 | 2,00 | 4,52E-02 | 2,29E-01 |
| PARD6A | 41,08 | -0,15 | 0,07 | -2,00 | 4,51E-02 | 2,29E-01 |
| IMPDH2 | 817,06 | 0,09 | 0,05 | 2,00 | 4,53E-02 | 2,29E-01 |
| FAM234A | 75,67 | -0,22 | 0,11 | -2,00 | 4,53E-02 | 2,29E-01 |
| NSUN6 | 706,22 | 0,09 | 0,04 | 2,00 | 4,54E-02 | 2,30E-01 |
| TBC1D15 | 1985,36 | 0,14 | 0,07 | 2,00 | 4,55E-02 | 2,30E-01 |
| UBE2E2 | 220,13 | 0,12 | 0,06 | 2,00 | 4,55E-02 | 2,30E-01 |
| TAF13 | 284,78 | 0,18 | 0,09 | 2,00 | 4,55E-02 | 2,30E-01 |
| RBM7 | 679,34 | 0,12 | 0,06 | 2,00 | 4,56E-02 | 2,30E-01 |
| SPSB3 | 1461,94 | -0,10 | 0,05 | -2,00 | 4,56E-02 | 2,30E-01 |
| SRGAP3 | 251,63 | -0,28 | 0,14 | -2,00 | 4,56E-02 | 2,30E-01 |
| CCR2 | 3660,15 | 0,16 | 0,08 | 2,00 | 4,56E-02 | 2,30E-01 |
| SREK1IP1 | 911,33 | 0,14 | 0,07 | 2,00 | 4,56E-02 | 2,30E-01 |
| DLG1 | 2093,86 | 0,09 | 0,05 | 2,00 | 4,57E-02 | 2,30E-01 |
| CD180 | 619,18 | 0,16 | 0,08 | 2,00 | 4,57E-02 | 2,30E-01 |
| ALKBH1 | 363,71 | 0,05 | 0,03 | 2,00 | 4,57E-02 | 2,30E-01 |
| GDI2 | 4541,20 | 0,13 | 0,06 | 2,00 | 4,57E-02 | 2,30E-01 |
| THRB | 62,42 | -0,38 | 0,19 | -2,00 | 4,58E-02 | 2,30E-01 |
| FAM89A | 101,73 | -0,15 | 0,07 | -2,00 | 4,58E-02 | 2,30E-01 |
| LIN52 | 292,13 | 0,07 | 0,03 | 2,00 | 4,58E-02 | 2,30E-01 |
| COPS5 | 1416,14 | 0,07 | 0,04 | 2,00 | 4,59E-02 | 2,30E-01 |
| ZSWIM3 | 91,70 | -0,11 | 0,05 | -2,00 | 4,58E-02 | 2,30E-01 |
| ESAM | 106,03 | -0,26 | 0,13 | -2,00 | 4,59E-02 | 2,30E-01 |
| LEMD3 | 2222,12 | 0,11 | 0,05 | 2,00 | 4,58E-02 | 2,30E-01 |
| BCKDHB | 388,34 | 0,10 | 0,05 | 2,00 | 4,59E-02 | 2,30E-01 |
| DHX9 | 4700,07 | 0,10 | 0,05 | 2,00 | 4,59E-02 | 2,30E-01 |
| ADNP | 3070,61 | 0,07 | 0,03 | 2,00 | 4,59E-02 | 2,30E-01 |
| DNAJC13 | 3709,99 | 0,10 | 0,05 | 2,00 | 4,60E-02 | 2,30E-01 |
| PRKACB | 3365,96 | 0,20 | 0,10 | 2,00 | 4,60E-02 | 2,30E-01 |
| SASS6 | 227,62 | 0,13 | 0,07 | 1,99 | 4,61E-02 | 2,30E-01 |
| ZNF771 | 63,28 | -0,19 | 0,10 | -1,99 | 4,61E-02 | 2,31E-01 |
| NUP160 | 1549,12 | 0,07 | 0,03 | 1,99 | 4,62E-02 | 2,31E-01 |
| SLA2 | 700,15 | -0,13 | 0,07 | -1,99 | 4,61E-02 | 2,31E-01 |
| ZNF480 | 429,38 | 0,19 | 0,09 | 1,99 | 4,61E-02 | 2,31E-01 |
| CEP290 | 910,67 | 0,18 | 0,09 | 1,99 | 4,62E-02 | 2,31E-01 |
| TESC | 2081,59 | -0,24 | 0,12 | -1,99 | 4,63E-02 | 2,31E-01 |
| PLBD2 | 201,10 | -0,23 | 0,11 | -1,99 | 4,63E-02 | 2,31E-01 |
| BCAP29 | 605,77 | 0,12 | 0,06 | 1,99 | 4,63E-02 | 2,31E-01 |
| BCAR3 | 79,02 | -0,31 | 0,16 | -1,99 | 4,63E-02 | 2,31E-01 |
| ZNF273 | 324,88 | 0,17 | 0,08 | 1,99 | 4,63E-02 | 2,31E-01 |
| TMEM68 | 427,93 | 0,09 | 0,05 | 1,99 | 4,64E-02 | 2,31E-01 |
| CASP2 | 1576,18 | -0,10 | 0,05 | -1,99 | 4,65E-02 | 2,31E-01 |
| TMA7 | 752,62 | 0,29 | 0,14 | 1,99 | 4,64E-02 | 2,31E-01 |
| COG6 | 821,04 | 0,16 | 0,08 | 1,99 | 4,64E-02 | 2,31E-01 |
| RPL17 | 449,74 | 0,16 | 0,08 | 1,99 | 4,65E-02 | 2,31E-01 |

| | | | | | | |
|---|---|---|---|---|---|---|
| BCL2A1 | 540,49 | 0,30 | 0,15 | 1,99 | 4,65E-02 | 2,31E-01 |
| EFCAB2 | 210,60 | -0,15 | 0,08 | -1,99 | 4,66E-02 | 2,31E-01 |
| RIOK2 | 533,82 | 0,17 | 0,08 | 1,99 | 4,66E-02 | 2,31E-01 |
| SLFN12 | 742,93 | 0,15 | 0,07 | 1,99 | 4,66E-02 | 2,31E-01 |
| ESM1 | 30,67 | -0,35 | 0,17 | -1,99 | 4,67E-02 | 2,32E-01 |
| GZMK | 706,55 | 0,26 | 0,13 | 1,99 | 4,67E-02 | 2,32E-01 |
| CAPN1 | 1335,32 | -0,15 | 0,08 | -1,99 | 4,67E-02 | 2,32E-01 |
| U2SURP | 4211,28 | 0,14 | 0,07 | 1,99 | 4,67E-02 | 2,32E-01 |
| WARS2 | 264,33 | 0,09 | 0,05 | 1,99 | 4,68E-02 | 2,32E-01 |
| STOM | 4658,72 | -0,18 | 0,09 | -1,99 | 4,68E-02 | 2,32E-01 |
| INHBB | 35,57 | -0,31 | 0,16 | -1,99 | 4,68E-02 | 2,32E-01 |
| EXOSC3 | 815,04 | 0,10 | 0,05 | 1,99 | 4,68E-02 | 2,32E-01 |
| PNPLA7 | 121,94 | -0,38 | 0,19 | -1,99 | 4,68E-02 | 2,32E-01 |
| ZNF550 | 647,38 | 0,12 | 0,06 | 1,99 | 4,69E-02 | 2,32E-01 |
| ABO | 61,98 | -0,41 | 0,20 | -1,99 | 4,70E-02 | 2,32E-01 |
| CMTM8 | 26,29 | -0,29 | 0,14 | -1,99 | 4,71E-02 | 2,32E-01 |
| EMC2 | 1004,70 | 0,18 | 0,09 | 1,99 | 4,71E-02 | 2,32E-01 |
| ATP5F1D | 97,15 | -0,24 | 0,12 | -1,99 | 4,71E-02 | 2,32E-01 |
| CFL2 | 235,17 | 0,16 | 0,08 | 1,99 | 4,71E-02 | 2,32E-01 |
| TOMM20 | 1856,11 | 0,14 | 0,07 | 1,99 | 4,70E-02 | 2,32E-01 |
| SLC41A1 | 185,00 | -0,19 | 0,10 | -1,99 | 4,70E-02 | 2,32E-01 |
| NRP1 | 113,08 | -0,31 | 0,16 | -1,98 | 4,72E-02 | 2,32E-01 |
| MCM9 | 1804,13 | 0,07 | 0,04 | 1,98 | 4,71E-02 | 2,32E-01 |
| DOT1L | 343,01 | -0,28 | 0,14 | -1,99 | 4,71E-02 | 2,32E-01 |
| SLC29A2 | 54,11 | -0,28 | 0,14 | -1,98 | 4,72E-02 | 2,32E-01 |
| FBXO45 | 377,90 | 0,13 | 0,06 | 1,98 | 4,72E-02 | 2,32E-01 |
| OTULINL | 1632,60 | 0,12 | 0,06 | 1,98 | 4,72E-02 | 2,32E-01 |
| ATP2B1 | 6997,82 | 0,11 | 0,06 | 1,98 | 4,74E-02 | 2,32E-01 |
| SIRT1 | 1386,42 | 0,10 | 0,05 | 1,98 | 4,73E-02 | 2,32E-01 |
| MRPS11 | 603,79 | 0,05 | 0,03 | 1,98 | 4,74E-02 | 2,32E-01 |
| NSA2 | 986,87 | 0,19 | 0,10 | 1,98 | 4,73E-02 | 2,32E-01 |
| ZGRF1 | 450,26 | 0,14 | 0,07 | 1,98 | 4,73E-02 | 2,32E-01 |
| TMEM63B | 954,80 | -0,28 | 0,14 | -1,98 | 4,73E-02 | 2,32E-01 |
| SOX13 | 106,52 | -0,32 | 0,16 | -1,98 | 4,73E-02 | 2,32E-01 |
| MRPS18A | 310,06 | 0,07 | 0,03 | 1,98 | 4,74E-02 | 2,32E-01 |
| C1QTNF9 | 38,20 | -0,15 | 0,08 | -1,98 | 4,75E-02 | 2,33E-01 |
| CAPN7 | 2435,48 | 0,11 | 0,06 | 1,98 | 4,75E-02 | 2,33E-01 |
| ZNF841 | 841,36 | 0,09 | 0,05 | 1,98 | 4,74E-02 | 2,33E-01 |
| CDON | 75,45 | -0,34 | 0,17 | -1,98 | 4,75E-02 | 2,33E-01 |
| RBMX2 | 389,32 | 0,09 | 0,05 | 1,98 | 4,75E-02 | 2,33E-01 |
| CCDC90B | 865,14 | 0,11 | 0,05 | 1,98 | 4,76E-02 | 2,33E-01 |
| ATP13A3 | 3101,24 | 0,10 | 0,05 | 1,98 | 4,77E-02 | 2,33E-01 |
| SLC39A14 | 117,44 | -0,17 | 0,08 | -1,98 | 4,77E-02 | 2,33E-01 |
| ZNF606 | 581,40 | 0,08 | 0,04 | 1,98 | 4,76E-02 | 2,33E-01 |
| SEL1L3 | 2413,36 | 0,12 | 0,06 | 1,98 | 4,77E-02 | 2,33E-01 |
| CD151 | 486,04 | -0,20 | 0,10 | -1,98 | 4,77E-02 | 2,33E-01 |

| | | | | | | |
|---|---|---|---|---|---|---|
| ATG14 | 997,05 | 0,06 | 0,03 | 1,98 | 4,76E-02 | 2,33E-01 |
| CDK2AP1 | 297,44 | -0,13 | 0,07 | -1,98 | 4,76E-02 | 2,33E-01 |
| PIAS2 | 1988,20 | 0,11 | 0,06 | 1,98 | 4,78E-02 | 2,33E-01 |
| CDIPT | 1522,67 | -0,14 | 0,07 | -1,98 | 4,78E-02 | 2,33E-01 |
| ACOT11 | 116,57 | -0,32 | 0,16 | -1,98 | 4,78E-02 | 2,33E-01 |
| COX20 | 735,06 | 0,10 | 0,05 | 1,98 | 4,78E-02 | 2,33E-01 |
| RFX2 | 369,13 | -0,22 | 0,11 | -1,98 | 4,78E-02 | 2,33E-01 |
| KANSL2 | 1119,16 | 0,08 | 0,04 | 1,98 | 4,78E-02 | 2,33E-01 |
| RARS2 | 1178,26 | 0,09 | 0,05 | 1,98 | 4,79E-02 | 2,33E-01 |
| APBA3 | 397,99 | -0,11 | 0,06 | -1,98 | 4,79E-02 | 2,33E-01 |
| TAF9 | 388,54 | 0,14 | 0,07 | 1,98 | 4,80E-02 | 2,33E-01 |
| XRCC4 | 322,65 | 0,22 | 0,11 | 1,98 | 4,80E-02 | 2,33E-01 |
| ALDH1A2 | 178,49 | -0,28 | 0,14 | -1,98 | 4,80E-02 | 2,33E-01 |
| PTPN22 | 1372,80 | 0,15 | 0,08 | 1,98 | 4,82E-02 | 2,33E-01 |
| LIN7C | 1778,24 | 0,18 | 0,09 | 1,98 | 4,81E-02 | 2,33E-01 |
| USP54 | 143,41 | -0,22 | 0,11 | -1,98 | 4,82E-02 | 2,33E-01 |
| PDE6B | 155,95 | -0,17 | 0,09 | -1,98 | 4,81E-02 | 2,33E-01 |
| OSBPL9 | 2257,71 | 0,08 | 0,04 | 1,98 | 4,82E-02 | 2,33E-01 |
| DNAJC19 | 723,98 | 0,14 | 0,07 | 1,98 | 4,81E-02 | 2,33E-01 |
| SIGLEC15 | 53,53 | -0,27 | 0,13 | -1,98 | 4,82E-02 | 2,34E-01 |
| PALS2 | 681,01 | 0,16 | 0,08 | 1,98 | 4,82E-02 | 2,34E-01 |
| NTNG2 | 846,41 | -0,23 | 0,11 | -1,98 | 4,83E-02 | 2,34E-01 |
| SLC12A2 | 746,55 | 0,11 | 0,06 | 1,97 | 4,83E-02 | 2,34E-01 |
| CACHD1 | 66,95 | -0,37 | 0,19 | -1,97 | 4,84E-02 | 2,34E-01 |
| PIP5K1C | 344,26 | -0,26 | 0,13 | -1,97 | 4,83E-02 | 2,34E-01 |
| RPL10A | 8051,93 | 0,17 | 0,09 | 1,97 | 4,84E-02 | 2,34E-01 |
| PSEN2 | 169,01 | -0,14 | 0,07 | -1,97 | 4,84E-02 | 2,34E-01 |
| VIL1 | 90,00 | -0,32 | 0,16 | -1,97 | 4,85E-02 | 2,34E-01 |
| TRIM47 | 45,75 | -0,22 | 0,11 | -1,97 | 4,85E-02 | 2,34E-01 |
| CAPRIN1 | 3787,96 | 0,06 | 0,03 | 1,97 | 4,86E-02 | 2,34E-01 |
| ZNF597 | 401,24 | 0,11 | 0,06 | 1,97 | 4,86E-02 | 2,35E-01 |
| IFT57 | 461,04 | 0,17 | 0,09 | 1,97 | 4,87E-02 | 2,35E-01 |
| RFX1 | 206,13 | -0,28 | 0,14 | -1,97 | 4,87E-02 | 2,35E-01 |
| MRPL47 | 191,60 | 0,20 | 0,10 | 1,97 | 4,87E-02 | 2,35E-01 |
| DPH6 | 430,22 | 0,15 | 0,07 | 1,97 | 4,87E-02 | 2,35E-01 |
| AGPAT1 | 187,79 | -0,21 | 0,11 | -1,97 | 4,88E-02 | 2,35E-01 |
| PLCH1 | 45,05 | -0,36 | 0,18 | -1,97 | 4,88E-02 | 2,35E-01 |
| CC2D1A | 142,23 | -0,28 | 0,14 | -1,97 | 4,89E-02 | 2,35E-01 |
| VPS41 | 3670,16 | 0,10 | 0,05 | 1,97 | 4,90E-02 | 2,35E-01 |
| CPT1A | 1241,82 | -0,17 | 0,09 | -1,97 | 4,90E-02 | 2,35E-01 |
| DTWD1 | 795,69 | 0,11 | 0,06 | 1,97 | 4,89E-02 | 2,35E-01 |
| GRPEL2 | 757,90 | 0,12 | 0,06 | 1,97 | 4,89E-02 | 2,35E-01 |
| TASOR | 4676,02 | 0,12 | 0,06 | 1,97 | 4,90E-02 | 2,35E-01 |
| CRBN | 3174,28 | 0,13 | 0,07 | 1,97 | 4,90E-02 | 2,35E-01 |
| STEAP3 | 52,74 | -0,32 | 0,16 | -1,97 | 4,90E-02 | 2,35E-01 |
| ZNF614 | 366,65 | 0,14 | 0,07 | 1,97 | 4,91E-02 | 2,35E-01 |

| | | | | | | |
|---|---|---|---|---|---|---|
| E2F5 | 253,65 | 0,19 | 0,09 | 1,97 | 4,91E-02 | 2,35E-01 |
| SLC25A27 | 37,49 | -0,27 | 0,14 | -1,97 | 4,91E-02 | 2,35E-01 |
| TTPAL | 843,29 | -0,07 | 0,04 | -1,97 | 4,91E-02 | 2,35E-01 |
| SWAP70 | 1573,63 | 0,13 | 0,07 | 1,97 | 4,92E-02 | 2,35E-01 |
| WBP4 | 424,70 | 0,13 | 0,06 | 1,97 | 4,92E-02 | 2,36E-01 |
| ARF6 | 4381,29 | 0,09 | 0,05 | 1,97 | 4,93E-02 | 2,36E-01 |
| CDKN2C | 136,57 | -0,16 | 0,08 | -1,97 | 4,92E-02 | 2,36E-01 |
| ADGRE2 | 3072,38 | -0,18 | 0,09 | -1,97 | 4,93E-02 | 2,36E-01 |
| A3GALT2 | 28,38 | -0,31 | 0,16 | -1,96 | 4,94E-02 | 2,36E-01 |
| NINL | 118,70 | -0,32 | 0,16 | -1,97 | 4,94E-02 | 2,36E-01 |
| C8orf33 | 546,57 | 0,06 | 0,03 | 1,96 | 4,95E-02 | 2,36E-01 |
| BFSP1 | 58,62 | -0,32 | 0,17 | -1,96 | 4,94E-02 | 2,36E-01 |
| ZMAT1 | 1098,69 | 0,12 | 0,06 | 1,96 | 4,94E-02 | 2,36E-01 |
| NSMCE4A | 900,89 | 0,08 | 0,04 | 1,97 | 4,94E-02 | 2,36E-01 |
| TBC1D22B | 638,11 | -0,17 | 0,09 | -1,96 | 4,95E-02 | 2,36E-01 |
| CNIH3 | 190,14 | -0,15 | 0,08 | -1,96 | 4,95E-02 | 2,36E-01 |
| MTO1 | 1235,43 | 0,08 | 0,04 | 1,96 | 4,95E-02 | 2,36E-01 |
| EFCAB8 | 102,10 | -0,27 | 0,14 | -1,96 | 4,95E-02 | 2,36E-01 |
| TDRP | 109,50 | -0,26 | 0,13 | -1,96 | 4,96E-02 | 2,36E-01 |
| GCNT4 | 724,36 | 0,18 | 0,09 | 1,96 | 4,96E-02 | 2,36E-01 |
| ZNF184 | 394,05 | 0,16 | 0,08 | 1,96 | 4,98E-02 | 2,36E-01 |
| MIER3 | 775,76 | 0,18 | 0,09 | 1,96 | 4,97E-02 | 2,36E-01 |
| APIP | 388,41 | 0,13 | 0,07 | 1,96 | 4,97E-02 | 2,36E-01 |
| TMEM216 | 215,84 | -0,08 | 0,04 | -1,96 | 4,97E-02 | 2,36E-01 |
| MTF2 | 1663,85 | 0,11 | 0,06 | 1,96 | 4,98E-02 | 2,36E-01 |
| DSN1 | 253,34 | 0,08 | 0,04 | 1,96 | 4,98E-02 | 2,36E-01 |
| NDUFA10 | 2164,58 | 0,06 | 0,03 | 1,96 | 4,98E-02 | 2,36E-01 |
| FOLR3 | 703,00 | -0,35 | 0,18 | -1,96 | 4,98E-02 | 2,36E-01 |
| CELSR1 | 170,20 | -0,37 | 0,19 | -1,96 | 4,99E-02 | 2,37E-01 |
| RAB3GAP1 | 2231,94 | 0,09 | 0,05 | 1,96 | 5,00E-02 | 2,37E-01 |
| XRCC5 | 7922,24 | 0,11 | 0,05 | 1,96 | 4,99E-02 | 2,37E-01 |
| TANGO2 | 2446,19 | -0,19 | 0,10 | -1,96 | 5,00E-02 | 2,37E-01 |
| PWWP3A | 377,75 | -0,14 | 0,07 | -1,96 | 4,99E-02 | 2,37E-01 |
| PCF11 | 6578,11 | 0,09 | 0,05 | 1,96 | 5,00E-02 | 2,37E-01 |
| BLOC1S4 | 599,76 | 0,08 | 0,04 | 1,96 | 5,00E-02 | 2,37E-01 |
| NBPF14 | 1648,25 | -0,14 | 0,07 | -1,96 | 5,00E-02 | 2,37E-01 |
| KAZN | 220,78 | -0,34 | 0,17 | -1,96 | 5,00E-02 | 2,37E-01 |
| FBXO28 | 1533,30 | 0,11 | 0,05 | 1,96 | 5,01E-02 | 2,37E-01 |
| NDUFAF4 | 182,22 | 0,19 | 0,10 | 1,96 | 5,01E-02 | 2,37E-01 |
| TPX2 | 74,73 | -0,22 | 0,11 | -1,96 | 5,01E-02 | 2,37E-01 |
| NRXN1 | 160,11 | -0,30 | 0,15 | -1,96 | 5,02E-02 | 2,37E-01 |
| UBTD1 | 79,50 | -0,19 | 0,09 | -1,96 | 5,02E-02 | 2,37E-01 |
| MRPL50 | 466,77 | 0,23 | 0,12 | 1,96 | 5,01E-02 | 2,37E-01 |
| ACBD5 | 1336,13 | 0,10 | 0,05 | 1,96 | 5,02E-02 | 2,37E-01 |
| CD109 | 99,03 | -0,26 | 0,13 | -1,96 | 5,02E-02 | 2,37E-01 |
| MROH6 | 83,16 | -0,28 | 0,14 | -1,96 | 5,03E-02 | 2,37E-01 |

| | | | | | | |
|---|---|---|---|---|---|---|
| CNOT11 | 602,66 | 0,05 | 0,02 | 1,96 | 5,02E-02 | 2,37E-01 |
| E2F6 | 237,81 | 0,08 | 0,04 | 1,96 | 5,03E-02 | 2,37E-01 |
| GNPTAB | 2406,21 | 0,09 | 0,05 | 1,96 | 5,03E-02 | 2,37E-01 |
| WRN | 853,63 | 0,11 | 0,06 | 1,96 | 5,03E-02 | 2,37E-01 |
| RAP1GDS1 | 1214,86 | 0,10 | 0,05 | 1,96 | 5,04E-02 | 2,37E-01 |
| ANKRD10 | 5625,85 | 0,11 | 0,06 | 1,96 | 5,04E-02 | 2,37E-01 |
| JAK3 | 3090,01 | -0,22 | 0,11 | -1,96 | 5,04E-02 | 2,37E-01 |
| ZNF701 | 988,40 | 0,11 | 0,06 | 1,96 | 5,05E-02 | 2,37E-01 |
| SYTL1 | 1814,26 | -0,12 | 0,06 | -1,96 | 5,05E-02 | 2,37E-01 |
| PA2G4 | 807,34 | -0,11 | 0,05 | -1,95 | 5,07E-02 | 2,37E-01 |
| SFT2D3 | 564,10 | 0,12 | 0,06 | 1,95 | 5,07E-02 | 2,37E-01 |
| FAM114A2 | 736,91 | 0,09 | 0,05 | 1,95 | 5,07E-02 | 2,37E-01 |
| CORIN | 87,50 | -0,40 | 0,20 | -1,96 | 5,06E-02 | 2,37E-01 |
| PEX1 | 1064,83 | 0,08 | 0,04 | 1,95 | 5,06E-02 | 2,37E-01 |
| MSMO1 | 457,98 | 0,13 | 0,07 | 1,95 | 5,07E-02 | 2,37E-01 |
| AGL | 1109,64 | 0,11 | 0,06 | 1,95 | 5,07E-02 | 2,37E-01 |
| RFXANK | 646,87 | -0,09 | 0,05 | -1,95 | 5,06E-02 | 2,37E-01 |
| RBM3 | 4361,31 | 0,12 | 0,06 | 1,95 | 5,08E-02 | 2,37E-01 |
| GNAS | 5667,36 | -0,16 | 0,08 | -1,95 | 5,06E-02 | 2,37E-01 |
| TRAF3IP2 | 140,76 | -0,14 | 0,07 | -1,95 | 5,07E-02 | 2,37E-01 |
| GTF2F2 | 586,15 | 0,10 | 0,05 | 1,95 | 5,07E-02 | 2,37E-01 |
| HIBADH | 334,67 | 0,10 | 0,05 | 1,95 | 5,07E-02 | 2,37E-01 |
| ST7 | 193,54 | -0,11 | 0,06 | -1,95 | 5,08E-02 | 2,37E-01 |
| NT5M | 239,05 | -0,27 | 0,14 | -1,95 | 5,08E-02 | 2,37E-01 |
| FANK1 | 47,33 | -0,31 | 0,16 | -1,95 | 5,08E-02 | 2,37E-01 |
| MED12L | 144,15 | -0,21 | 0,11 | -1,95 | 5,09E-02 | 2,37E-01 |
| C1orf56 | 33,13 | -0,19 | 0,10 | -1,95 | 5,09E-02 | 2,37E-01 |
| SECTM1 | 1645,87 | -0,20 | 0,10 | -1,95 | 5,10E-02 | 2,38E-01 |
| TUFT1 | 148,54 | -0,14 | 0,07 | -1,95 | 5,10E-02 | 2,38E-01 |
| TJP3 | 86,33 | -0,24 | 0,12 | -1,95 | 5,10E-02 | 2,38E-01 |
| PRDX6 | 5705,24 | -0,22 | 0,11 | -1,95 | 5,11E-02 | 2,38E-01 |
| ING3 | 923,82 | 0,12 | 0,06 | 1,95 | 5,12E-02 | 2,38E-01 |
| AGGF1 | 648,33 | 0,12 | 0,06 | 1,95 | 5,12E-02 | 2,38E-01 |
| DLG2 | 92,33 | -0,32 | 0,16 | -1,95 | 5,12E-02 | 2,38E-01 |
| CSAD | 774,90 | -0,19 | 0,10 | -1,95 | 5,12E-02 | 2,38E-01 |
| RAD1 | 630,17 | 0,08 | 0,04 | 1,95 | 5,13E-02 | 2,38E-01 |
| PIGK | 749,88 | 0,18 | 0,09 | 1,95 | 5,13E-02 | 2,38E-01 |
| SEPSECS | 952,62 | 0,11 | 0,06 | 1,95 | 5,13E-02 | 2,38E-01 |
| OXR1 | 2199,95 | 0,17 | 0,09 | 1,95 | 5,13E-02 | 2,38E-01 |
| TAS2R14 | 302,05 | 0,17 | 0,09 | 1,95 | 5,14E-02 | 2,38E-01 |
| UBXN4 | 3405,14 | 0,11 | 0,06 | 1,95 | 5,14E-02 | 2,39E-01 |
| SFR1 | 154,79 | 0,16 | 0,08 | 1,95 | 5,15E-02 | 2,39E-01 |
| ELMO3 | 97,80 | -0,19 | 0,10 | -1,95 | 5,15E-02 | 2,39E-01 |
| KANTR | 861,40 | 0,10 | 0,05 | 1,95 | 5,16E-02 | 2,39E-01 |
| FKBP14 | 201,53 | 0,11 | 0,06 | 1,95 | 5,16E-02 | 2,39E-01 |
| YAE1 | 183,16 | 0,16 | 0,08 | 1,95 | 5,16E-02 | 2,39E-01 |

| | | | | | | |
|---|---|---|---|---|---|---|
| UCP3 | 101,42 | -0,15 | 0,08 | -1,95 | 5,17E-02 | 2,39E-01 |
| HNRNPDL | 3615,37 | 0,09 | 0,05 | 1,95 | 5,17E-02 | 2,39E-01 |
| ELOVL7 | 439,72 | -0,23 | 0,12 | -1,95 | 5,18E-02 | 2,39E-01 |
| ADGRE1 | 1294,52 | -0,18 | 0,09 | -1,95 | 5,18E-02 | 2,39E-01 |
| CCNT1 | 2278,58 | 0,09 | 0,04 | 1,95 | 5,17E-02 | 2,39E-01 |
| MAGT1 | 2351,63 | 0,11 | 0,06 | 1,95 | 5,18E-02 | 2,39E-01 |
| REST | 2825,67 | 0,10 | 0,05 | 1,95 | 5,17E-02 | 2,39E-01 |
| RELL2 | 245,39 | -0,12 | 0,06 | -1,94 | 5,19E-02 | 2,39E-01 |
| FBXO25 | 745,95 | 0,08 | 0,04 | 1,94 | 5,19E-02 | 2,39E-01 |
| ZNF680 | 537,61 | 0,18 | 0,09 | 1,94 | 5,19E-02 | 2,39E-01 |
| FBXO11 | 3167,64 | 0,11 | 0,05 | 1,94 | 5,19E-02 | 2,39E-01 |
| FCER2 | 111,99 | 0,28 | 0,14 | 1,94 | 5,19E-02 | 2,39E-01 |
| MAP9 | 228,35 | 0,20 | 0,10 | 1,94 | 5,20E-02 | 2,39E-01 |
| RPS27A | 6109,98 | 0,24 | 0,12 | 1,94 | 5,20E-02 | 2,39E-01 |
| ITGA2 | 71,73 | -0,29 | 0,15 | -1,94 | 5,20E-02 | 2,39E-01 |
| PSMA4 | 1194,10 | 0,18 | 0,09 | 1,94 | 5,21E-02 | 2,40E-01 |
| SMIM14 | 1793,77 | 0,10 | 0,05 | 1,94 | 5,21E-02 | 2,40E-01 |
| RLIM | 5141,64 | 0,12 | 0,06 | 1,94 | 5,21E-02 | 2,40E-01 |
| RGS5 | 28,89 | -0,34 | 0,17 | -1,94 | 5,21E-02 | 2,40E-01 |
| CCDC47 | 1275,49 | 0,08 | 0,04 | 1,94 | 5,24E-02 | 2,40E-01 |
| PSMA1 | 833,07 | 0,09 | 0,05 | 1,94 | 5,25E-02 | 2,40E-01 |
| ZNF211 | 732,11 | 0,07 | 0,04 | 1,94 | 5,23E-02 | 2,40E-01 |
| RWDD4 | 309,06 | 0,11 | 0,05 | 1,94 | 5,24E-02 | 2,40E-01 |
| EEF2K | 527,18 | -0,15 | 0,08 | -1,94 | 5,23E-02 | 2,40E-01 |
| DOHH | 191,65 | -0,13 | 0,07 | -1,94 | 5,24E-02 | 2,40E-01 |
| ZNF239 | 101,27 | 0,15 | 0,08 | 1,94 | 5,23E-02 | 2,40E-01 |
| ZBTB41 | 1064,00 | 0,24 | 0,12 | 1,94 | 5,24E-02 | 2,40E-01 |
| MYO1C | 201,70 | -0,23 | 0,12 | -1,94 | 5,24E-02 | 2,40E-01 |
| ATP5F1A | 5246,65 | 0,09 | 0,05 | 1,94 | 5,24E-02 | 2,40E-01 |
| NSG1 | 132,02 | -0,26 | 0,13 | -1,94 | 5,24E-02 | 2,40E-01 |
| CSF1 | 179,56 | -0,22 | 0,11 | -1,94 | 5,23E-02 | 2,40E-01 |
| ENPP4 | 754,87 | 0,22 | 0,11 | 1,94 | 5,24E-02 | 2,40E-01 |
| APBB2 | 57,56 | -0,45 | 0,23 | -1,94 | 5,24E-02 | 2,40E-01 |
| RGPD3 | 34,53 | -0,33 | 0,17 | -1,94 | 5,25E-02 | 2,40E-01 |
| TPD52 | 642,55 | 0,15 | 0,08 | 1,94 | 5,26E-02 | 2,40E-01 |
| SLC25A22 | 161,05 | -0,17 | 0,09 | -1,93 | 5,33E-02 | 2,41E-01 |
| PIK3CA | 2792,27 | 0,14 | 0,07 | 1,94 | 5,30E-02 | 2,41E-01 |
| APOBEC3B | 51,75 | -0,57 | 0,30 | -1,93 | 5,33E-02 | 2,41E-01 |
| KLF9 | 980,18 | 0,12 | 0,06 | 1,93 | 5,34E-02 | 2,41E-01 |
| GNPAT | 1214,29 | 0,07 | 0,04 | 1,94 | 5,29E-02 | 2,41E-01 |
| KBTBD6 | 506,13 | 0,13 | 0,06 | 1,94 | 5,29E-02 | 2,41E-01 |
| SCFD1 | 1869,88 | 0,12 | 0,06 | 1,94 | 5,29E-02 | 2,41E-01 |
| PCBP2 | 4635,69 | -0,12 | 0,06 | -1,93 | 5,32E-02 | 2,41E-01 |
| CBX3 | 1997,65 | 0,16 | 0,08 | 1,94 | 5,28E-02 | 2,41E-01 |
| FMR1 | 3089,72 | 0,12 | 0,06 | 1,93 | 5,32E-02 | 2,41E-01 |
| NDUFS5 | 303,16 | 0,20 | 0,10 | 1,93 | 5,31E-02 | 2,41E-01 |

| | | | | | | |
|---|---|---|---|---|---|---|
| ARFGEF1 | 5522,77 | 0,09 | 0,05 | 1,93 | 5,33E-02 | 2,41E-01 |
| SSBP3 | 570,45 | -0,18 | 0,09 | -1,93 | 5,32E-02 | 2,41E-01 |
| FZD5 | 142,87 | -0,18 | 0,10 | -1,93 | 5,32E-02 | 2,41E-01 |
| RCHY1 | 1246,13 | 0,14 | 0,07 | 1,93 | 5,33E-02 | 2,41E-01 |
| TWF1 | 1067,11 | 0,20 | 0,10 | 1,93 | 5,31E-02 | 2,41E-01 |
| HEXIM2 | 69,27 | -0,15 | 0,08 | -1,93 | 5,31E-02 | 2,41E-01 |
| PNRC2 | 3615,25 | 0,16 | 0,08 | 1,93 | 5,30E-02 | 2,41E-01 |
| CRACDL | 80,95 | -0,27 | 0,14 | -1,94 | 5,27E-02 | 2,41E-01 |
| PHIP | 8651,46 | 0,14 | 0,07 | 1,93 | 5,32E-02 | 2,41E-01 |
| VTA1 | 910,70 | 0,13 | 0,07 | 1,93 | 5,32E-02 | 2,41E-01 |
| SLC16A14 | 59,46 | -0,28 | 0,14 | -1,94 | 5,30E-02 | 2,41E-01 |
| MCCC1 | 785,98 | 0,07 | 0,04 | 1,94 | 5,30E-02 | 2,41E-01 |
| TJP2 | 178,55 | -0,19 | 0,10 | -1,93 | 5,31E-02 | 2,41E-01 |
| NOCT | 68,79 | -0,16 | 0,08 | -1,93 | 5,32E-02 | 2,41E-01 |
| ABHD8 | 132,59 | -0,12 | 0,06 | -1,93 | 5,31E-02 | 2,41E-01 |
| HIVEP2 | 3200,35 | 0,08 | 0,04 | 1,93 | 5,30E-02 | 2,41E-01 |
| CEP57L1 | 256,23 | 0,14 | 0,07 | 1,93 | 5,31E-02 | 2,41E-01 |
| PRPF38B | 3017,56 | 0,12 | 0,06 | 1,94 | 5,30E-02 | 2,41E-01 |
| TMEM38B | 306,03 | 0,12 | 0,06 | 1,93 | 5,32E-02 | 2,41E-01 |
| FIG4 | 1392,82 | 0,10 | 0,05 | 1,94 | 5,28E-02 | 2,41E-01 |
| GLS | 7497,39 | 0,14 | 0,07 | 1,93 | 5,33E-02 | 2,41E-01 |
| DDHD1 | 3585,41 | 0,10 | 0,05 | 1,94 | 5,28E-02 | 2,41E-01 |
| ZFP91 | 1379,99 | 0,07 | 0,03 | 1,93 | 5,35E-02 | 2,41E-01 |
| MAP3K7 | 2282,49 | 0,11 | 0,06 | 1,93 | 5,36E-02 | 2,41E-01 |
| IDE | 961,68 | 0,07 | 0,03 | 1,93 | 5,36E-02 | 2,41E-01 |
| VWDE | 169,90 | 0,32 | 0,17 | 1,93 | 5,35E-02 | 2,41E-01 |
| HBS1L | 1956,16 | 0,13 | 0,07 | 1,93 | 5,36E-02 | 2,41E-01 |
| PGBD4 | 299,20 | 0,14 | 0,07 | 1,93 | 5,35E-02 | 2,41E-01 |
| TASP1 | 514,06 | 0,10 | 0,05 | 1,93 | 5,35E-02 | 2,41E-01 |
| KPNA5 | 1264,39 | 0,19 | 0,10 | 1,93 | 5,35E-02 | 2,41E-01 |
| NOC4L | 88,59 | -0,22 | 0,11 | -1,93 | 5,36E-02 | 2,41E-01 |
| TMBIM1 | 3599,74 | -0,10 | 0,05 | -1,93 | 5,36E-02 | 2,41E-01 |
| ANK1 | 1510,37 | -0,29 | 0,15 | -1,93 | 5,35E-02 | 2,41E-01 |
| OARD1 | 1061,50 | 0,09 | 0,05 | 1,93 | 5,37E-02 | 2,41E-01 |
| ABHD6 | 247,28 | -0,08 | 0,04 | -1,93 | 5,37E-02 | 2,41E-01 |
| NSMCE1 | 513,08 | 0,08 | 0,04 | 1,93 | 5,37E-02 | 2,41E-01 |
| MLX | 1337,69 | -0,07 | 0,04 | -1,93 | 5,38E-02 | 2,41E-01 |
| CYBC1 | 2859,59 | -0,18 | 0,09 | -1,93 | 5,38E-02 | 2,42E-01 |
| MIIP | 921,92 | -0,12 | 0,06 | -1,93 | 5,38E-02 | 2,42E-01 |
| MMD | 1218,26 | -0,19 | 0,10 | -1,93 | 5,39E-02 | 2,42E-01 |
| CD79B | 855,72 | 0,17 | 0,09 | 1,93 | 5,39E-02 | 2,42E-01 |
| AGAP3 | 293,76 | -0,23 | 0,12 | -1,93 | 5,41E-02 | 2,42E-01 |
| POLR3F | 257,34 | 0,10 | 0,05 | 1,93 | 5,40E-02 | 2,42E-01 |
| PAK4 | 83,28 | -0,31 | 0,16 | -1,93 | 5,41E-02 | 2,42E-01 |
| EED | 1085,11 | 0,13 | 0,07 | 1,93 | 5,41E-02 | 2,42E-01 |
| BRCC3 | 553,49 | 0,08 | 0,04 | 1,93 | 5,42E-02 | 2,42E-01 |

| | | | | | | |
|---|---|---|---|---|---|---|
| MKI67 | 297,01 | -0,21 | 0,11 | -1,93 | 5,42E-02 | 2,42E-01 |
| MRPL42 | 1361,79 | 0,09 | 0,05 | 1,93 | 5,41E-02 | 2,42E-01 |
| HS2ST1 | 945,20 | 0,11 | 0,06 | 1,93 | 5,40E-02 | 2,42E-01 |
| LUC7L3 | 5499,80 | 0,12 | 0,06 | 1,93 | 5,41E-02 | 2,42E-01 |
| ATP6V1G2 | 43,60 | -0,15 | 0,08 | -1,93 | 5,41E-02 | 2,42E-01 |
| NKD1 | 114,13 | -0,32 | 0,17 | -1,93 | 5,41E-02 | 2,42E-01 |
| THNSL1 | 152,30 | 0,18 | 0,09 | 1,93 | 5,42E-02 | 2,42E-01 |
| MEST | 137,00 | -0,20 | 0,10 | -1,92 | 5,45E-02 | 2,42E-01 |
| POMGNT2 | 83,04 | -0,15 | 0,08 | -1,92 | 5,45E-02 | 2,42E-01 |
| COG3 | 2129,58 | 0,08 | 0,04 | 1,92 | 5,45E-02 | 2,42E-01 |
| MED1 | 1569,40 | 0,04 | 0,02 | 1,92 | 5,45E-02 | 2,42E-01 |
| KIAA1586 | 599,91 | 0,22 | 0,11 | 1,92 | 5,44E-02 | 2,42E-01 |
| NFKBIB | 293,05 | -0,09 | 0,05 | -1,92 | 5,44E-02 | 2,42E-01 |
| HDAC4 | 2916,41 | -0,14 | 0,07 | -1,92 | 5,44E-02 | 2,42E-01 |
| ATP6AP2 | 2747,60 | 0,10 | 0,05 | 1,92 | 5,43E-02 | 2,42E-01 |
| GORAB | 367,12 | 0,13 | 0,07 | 1,92 | 5,44E-02 | 2,42E-01 |
| FNTA | 2510,59 | 0,10 | 0,05 | 1,92 | 5,45E-02 | 2,42E-01 |
| ZNF322 | 499,83 | 0,17 | 0,09 | 1,92 | 5,42E-02 | 2,42E-01 |
| KHK | 63,76 | -0,17 | 0,09 | -1,92 | 5,44E-02 | 2,42E-01 |
| SERBP1 | 5546,44 | 0,10 | 0,05 | 1,92 | 5,45E-02 | 2,42E-01 |
| TADA1 | 311,49 | 0,10 | 0,05 | 1,92 | 5,43E-02 | 2,42E-01 |
| ZC2HC1A | 301,22 | 0,21 | 0,11 | 1,92 | 5,43E-02 | 2,42E-01 |
| ADAMTSL4 | 197,36 | -0,32 | 0,17 | -1,92 | 5,45E-02 | 2,42E-01 |
| RPL23A | 11999,50 | 0,14 | 0,07 | 1,92 | 5,44E-02 | 2,42E-01 |
| NHSL1 | 85,02 | -0,29 | 0,15 | -1,92 | 5,46E-02 | 2,42E-01 |
| VPS72 | 475,26 | 0,06 | 0,03 | 1,92 | 5,47E-02 | 2,42E-01 |
| KSR1 | 1299,73 | -0,19 | 0,10 | -1,92 | 5,46E-02 | 2,42E-01 |
| FPR1 | 11607,68 | -0,16 | 0,08 | -1,92 | 5,47E-02 | 2,42E-01 |
| CPSF6 | 3166,43 | 0,16 | 0,08 | 1,92 | 5,47E-02 | 2,42E-01 |
| NUDT17 | 74,57 | -0,11 | 0,06 | -1,92 | 5,47E-02 | 2,42E-01 |
| LPCAT4 | 350,84 | -0,13 | 0,07 | -1,92 | 5,48E-02 | 2,42E-01 |
| DKC1 | 801,16 | 0,07 | 0,04 | 1,92 | 5,48E-02 | 2,42E-01 |
| RNF20 | 2037,87 | 0,10 | 0,05 | 1,92 | 5,48E-02 | 2,43E-01 |
| ZW10 | 438,69 | 0,08 | 0,04 | 1,92 | 5,49E-02 | 2,43E-01 |
| ESR1 | 96,14 | -0,30 | 0,15 | -1,92 | 5,49E-02 | 2,43E-01 |
| ARHGEF11 | 1117,25 | -0,17 | 0,09 | -1,92 | 5,49E-02 | 2,43E-01 |
| TOPBP1 | 2131,12 | 0,08 | 0,04 | 1,92 | 5,49E-02 | 2,43E-01 |
| SUMO1 | 1431,68 | 0,13 | 0,07 | 1,92 | 5,51E-02 | 2,43E-01 |
| AKT3 | 2158,20 | 0,11 | 0,06 | 1,92 | 5,51E-02 | 2,43E-01 |
| PIM1 | 12802,49 | -0,19 | 0,10 | -1,92 | 5,50E-02 | 2,43E-01 |
| TMX3 | 2558,11 | 0,19 | 0,10 | 1,92 | 5,50E-02 | 2,43E-01 |
| ABCA9 | 56,83 | -0,41 | 0,22 | -1,92 | 5,51E-02 | 2,43E-01 |
| NAA50 | 3710,72 | 0,11 | 0,06 | 1,92 | 5,50E-02 | 2,43E-01 |
| IRAK2 | 217,07 | -0,11 | 0,06 | -1,92 | 5,50E-02 | 2,43E-01 |
| ZNF764 | 128,02 | -0,17 | 0,09 | -1,92 | 5,50E-02 | 2,43E-01 |
| THSD1 | 47,12 | -0,21 | 0,11 | -1,92 | 5,51E-02 | 2,43E-01 |

| | | | | | | |
|---|---|---|---|---|---|---|
| ZNF90 | 129,38 | 0,15 | 0,08 | 1,92 | 5,51E-02 | 2,43E-01 |
| PODXL2 | 40,70 | -0,25 | 0,13 | -1,92 | 5,52E-02 | 2,43E-01 |
| ZNF773 | 311,23 | 0,09 | 0,05 | 1,92 | 5,52E-02 | 2,43E-01 |
| DDX10 | 781,02 | 0,10 | 0,05 | 1,92 | 5,52E-02 | 2,43E-01 |
| ZNF721 | 706,96 | 0,21 | 0,11 | 1,92 | 5,52E-02 | 2,43E-01 |
| SLC25A39 | 11302,83 | -0,29 | 0,15 | -1,92 | 5,52E-02 | 2,43E-01 |
| F11R | 670,68 | -0,19 | 0,10 | -1,92 | 5,52E-02 | 2,43E-01 |
| ZNF799 | 313,64 | 0,09 | 0,05 | 1,92 | 5,53E-02 | 2,43E-01 |
| NAPG | 1437,49 | 0,10 | 0,05 | 1,92 | 5,54E-02 | 2,43E-01 |
| PSMD7 | 1503,33 | 0,09 | 0,05 | 1,92 | 5,54E-02 | 2,43E-01 |
| CCNDBP1 | 5422,65 | -0,12 | 0,06 | -1,92 | 5,55E-02 | 2,43E-01 |
| SNRPF | 280,84 | 0,15 | 0,08 | 1,92 | 5,55E-02 | 2,43E-01 |
| ZNF682 | 192,80 | 0,11 | 0,06 | 1,91 | 5,55E-02 | 2,43E-01 |
| ZNF10 | 262,20 | 0,13 | 0,07 | 1,91 | 5,56E-02 | 2,43E-01 |
| EID2 | 234,15 | 0,10 | 0,05 | 1,91 | 5,56E-02 | 2,43E-01 |
| FKBP4 | 366,02 | -0,09 | 0,05 | -1,91 | 5,56E-02 | 2,43E-01 |
| RPS7 | 2743,52 | 0,32 | 0,17 | 1,91 | 5,56E-02 | 2,43E-01 |
| DCAF17 | 862,41 | 0,10 | 0,05 | 1,91 | 5,57E-02 | 2,43E-01 |
| MBLAC2 | 251,95 | 0,17 | 0,09 | 1,91 | 5,57E-02 | 2,43E-01 |
| PLEK2 | 488,34 | -0,26 | 0,13 | -1,91 | 5,57E-02 | 2,43E-01 |
| MRS2 | 602,40 | 0,08 | 0,04 | 1,91 | 5,57E-02 | 2,44E-01 |
| ANKRD37 | 76,66 | -0,14 | 0,07 | -1,91 | 5,58E-02 | 2,44E-01 |
| DTX2 | 272,82 | -0,22 | 0,12 | -1,91 | 5,58E-02 | 2,44E-01 |
| CPTP | 79,83 | -0,24 | 0,12 | -1,91 | 5,59E-02 | 2,44E-01 |
| ZMYM6 | 897,20 | 0,09 | 0,04 | 1,91 | 5,59E-02 | 2,44E-01 |
| UFD1 | 875,18 | -0,06 | 0,03 | -1,91 | 5,59E-02 | 2,44E-01 |
| FAM76A | 449,69 | 0,07 | 0,04 | 1,91 | 5,59E-02 | 2,44E-01 |
| FAM83G | 122,14 | -0,22 | 0,12 | -1,91 | 5,60E-02 | 2,44E-01 |
| DENND4C | 3043,06 | 0,10 | 0,05 | 1,91 | 5,60E-02 | 2,44E-01 |
| CCDC17 | 229,73 | -0,18 | 0,09 | -1,91 | 5,59E-02 | 2,44E-01 |
| CDC40 | 1986,20 | 0,12 | 0,06 | 1,91 | 5,60E-02 | 2,44E-01 |
| COX15 | 695,96 | 0,06 | 0,03 | 1,91 | 5,61E-02 | 2,44E-01 |
| POLR1B | 725,83 | 0,07 | 0,04 | 1,91 | 5,61E-02 | 2,44E-01 |
| FEN1 | 205,11 | 0,09 | 0,05 | 1,91 | 5,61E-02 | 2,44E-01 |
| GOLGA5 | 985,93 | 0,08 | 0,04 | 1,91 | 5,63E-02 | 2,44E-01 |
| PSMF1 | 2650,68 | -0,21 | 0,11 | -1,91 | 5,63E-02 | 2,44E-01 |
| EFR3A | 3436,15 | 0,12 | 0,06 | 1,91 | 5,63E-02 | 2,44E-01 |
| PEBP1 | 973,21 | 0,11 | 0,06 | 1,91 | 5,62E-02 | 2,44E-01 |
| PDCD1LG2 | 38,36 | -0,26 | 0,14 | -1,91 | 5,63E-02 | 2,44E-01 |
| ANKRD20A4I | 35,49 | -0,49 | 0,26 | -1,91 | 5,63E-02 | 2,44E-01 |
| CALCB | 51,84 | -0,25 | 0,13 | -1,91 | 5,63E-02 | 2,44E-01 |
| VLDLR | 91,99 | -0,30 | 0,15 | -1,91 | 5,62E-02 | 2,44E-01 |
| SNX5 | 2229,59 | 0,07 | 0,04 | 1,91 | 5,63E-02 | 2,44E-01 |
| ZNF138 | 394,59 | 0,17 | 0,09 | 1,91 | 5,65E-02 | 2,45E-01 |
| ZNF624 | 298,36 | 0,16 | 0,08 | 1,91 | 5,65E-02 | 2,45E-01 |
| RPS6KL1 | 40,89 | -0,37 | 0,19 | -1,91 | 5,65E-02 | 2,45E-01 |

| | | | | | | |
|---|---|---|---|---|---|---|
| WASHC4 | 9814,68 | 0,16 | 0,08 | 1,91 | 5,66E-02 | 2,45E-01 |
| NDUFAF5 | 489,66 | 0,08 | 0,04 | 1,91 | 5,67E-02 | 2,45E-01 |
| TRIM3 | 139,28 | -0,20 | 0,10 | -1,91 | 5,67E-02 | 2,45E-01 |
| PARP11 | 1108,15 | 0,09 | 0,05 | 1,91 | 5,67E-02 | 2,45E-01 |
| PHKG2 | 348,19 | -0,10 | 0,05 | -1,90 | 5,68E-02 | 2,46E-01 |
| SESN2 | 181,09 | -0,16 | 0,08 | -1,90 | 5,69E-02 | 2,46E-01 |
| PEX13 | 1127,13 | 0,09 | 0,05 | 1,90 | 5,69E-02 | 2,46E-01 |
| ZMYM4 | 1216,74 | 0,07 | 0,04 | 1,90 | 5,70E-02 | 2,46E-01 |
| NSMAF | 1852,22 | 0,08 | 0,04 | 1,90 | 5,69E-02 | 2,46E-01 |
| CCDC14 | 1835,62 | 0,12 | 0,07 | 1,90 | 5,70E-02 | 2,46E-01 |
| TRAT1 | 1350,97 | 0,21 | 0,11 | 1,90 | 5,70E-02 | 2,46E-01 |
| DNAJB14 | 4576,67 | 0,18 | 0,09 | 1,90 | 5,71E-02 | 2,46E-01 |
| ITGA4 | 12362,10 | 0,13 | 0,07 | 1,90 | 5,71E-02 | 2,46E-01 |
| EXOC1 | 2427,66 | 0,12 | 0,06 | 1,90 | 5,71E-02 | 2,46E-01 |
| EPHX1 | 27,13 | -0,32 | 0,17 | -1,90 | 5,71E-02 | 2,46E-01 |
| TBK1 | 1419,75 | 0,14 | 0,08 | 1,90 | 5,72E-02 | 2,46E-01 |
| RASSF5 | 5418,55 | -0,10 | 0,05 | -1,90 | 5,72E-02 | 2,46E-01 |
| PCDHGA5 | 42,62 | -0,23 | 0,12 | -1,90 | 5,72E-02 | 2,46E-01 |
| ERGIC2 | 1614,04 | 0,16 | 0,09 | 1,90 | 5,72E-02 | 2,46E-01 |
| MGAT4A | 6203,57 | 0,12 | 0,06 | 1,90 | 5,73E-02 | 2,46E-01 |
| ATG4D | 322,00 | -0,11 | 0,06 | -1,90 | 5,73E-02 | 2,46E-01 |
| P4HA2 | 194,86 | -0,17 | 0,09 | -1,90 | 5,73E-02 | 2,46E-01 |
| CDK6 | 2334,13 | 0,10 | 0,05 | 1,90 | 5,74E-02 | 2,46E-01 |
| AP5M1 | 2535,28 | 0,15 | 0,08 | 1,90 | 5,74E-02 | 2,46E-01 |
| RPAP3 | 1304,44 | 0,13 | 0,07 | 1,90 | 5,74E-02 | 2,46E-01 |
| GAMT | 45,88 | -0,19 | 0,10 | -1,90 | 5,74E-02 | 2,47E-01 |
| GINS1 | 44,90 | -0,26 | 0,13 | -1,90 | 5,75E-02 | 2,47E-01 |
| RNF123 | 1723,16 | -0,17 | 0,09 | -1,90 | 5,75E-02 | 2,47E-01 |
| JAM3 | 412,10 | -0,21 | 0,11 | -1,90 | 5,75E-02 | 2,47E-01 |
| CYB561 | 364,92 | -0,16 | 0,08 | -1,90 | 5,76E-02 | 2,47E-01 |
| GP5 | 58,26 | -0,26 | 0,14 | -1,90 | 5,77E-02 | 2,47E-01 |
| UFSP2 | 365,89 | 0,11 | 0,06 | 1,90 | 5,78E-02 | 2,47E-01 |
| UNC5CL | 95,20 | -0,16 | 0,08 | -1,90 | 5,77E-02 | 2,47E-01 |
| CCR5 | 683,31 | 0,19 | 0,10 | 1,90 | 5,77E-02 | 2,47E-01 |
| NPR2 | 86,78 | -0,20 | 0,10 | -1,90 | 5,77E-02 | 2,47E-01 |
| PGRMC2 | 488,79 | 0,07 | 0,04 | 1,90 | 5,77E-02 | 2,47E-01 |
| ZNF678 | 838,42 | 0,18 | 0,09 | 1,90 | 5,77E-02 | 2,47E-01 |
| CKAP2L | 24,82 | -0,28 | 0,15 | -1,90 | 5,77E-02 | 2,47E-01 |
| RNF32 | 44,62 | -0,31 | 0,16 | -1,90 | 5,77E-02 | 2,47E-01 |
| HOXB3 | 116,46 | -0,25 | 0,13 | -1,90 | 5,78E-02 | 2,47E-01 |
| MCC | 123,25 | -0,27 | 0,14 | -1,90 | 5,78E-02 | 2,47E-01 |
| TMEM87B | 1215,67 | 0,09 | 0,05 | 1,90 | 5,78E-02 | 2,47E-01 |
| ELMOD2 | 1387,02 | 0,18 | 0,09 | 1,90 | 5,79E-02 | 2,47E-01 |
| SLU7 | 1763,44 | 0,13 | 0,07 | 1,90 | 5,79E-02 | 2,47E-01 |
| ARL5A | 1905,99 | 0,14 | 0,07 | 1,90 | 5,80E-02 | 2,47E-01 |
| MIS12 | 602,78 | 0,13 | 0,07 | 1,90 | 5,80E-02 | 2,47E-01 |

| | | | | | | |
|---|---|---|---|---|---|---|
| CAPN14 | 60,10 | -0,26 | 0,14 | -1,89 | 5,81E-02 | 2,48E-01 |
| CTH | 53,46 | -0,17 | 0,09 | -1,89 | 5,81E-02 | 2,48E-01 |
| C3orf38 | 1069,20 | 0,16 | 0,08 | 1,89 | 5,82E-02 | 2,48E-01 |
| PCDHGA7 | 37,24 | -0,25 | 0,13 | -1,89 | 5,82E-02 | 2,48E-01 |
| WDR12 | 440,13 | 0,07 | 0,04 | 1,89 | 5,82E-02 | 2,48E-01 |
| NEDD1 | 976,79 | 0,14 | 0,07 | 1,89 | 5,82E-02 | 2,48E-01 |
| MYBL1 | 2305,74 | 0,20 | 0,11 | 1,89 | 5,83E-02 | 2,48E-01 |
| COPS2 | 1278,17 | 0,20 | 0,10 | 1,89 | 5,83E-02 | 2,48E-01 |
| ZNF765 | 536,50 | 0,19 | 0,10 | 1,89 | 5,83E-02 | 2,48E-01 |
| SYF2 | 1501,47 | 0,13 | 0,07 | 1,89 | 5,83E-02 | 2,48E-01 |
| WIZ | 262,72 | -0,25 | 0,13 | -1,89 | 5,85E-02 | 2,48E-01 |
| THAP6 | 748,17 | 0,11 | 0,06 | 1,89 | 5,85E-02 | 2,48E-01 |
| MAGOHB | 446,58 | 0,13 | 0,07 | 1,89 | 5,85E-02 | 2,48E-01 |
| GTPBP2 | 895,97 | -0,13 | 0,07 | -1,89 | 5,85E-02 | 2,48E-01 |
| ASPRV1 | 273,33 | -0,21 | 0,11 | -1,89 | 5,86E-02 | 2,48E-01 |
| MRPL53 | 156,04 | 0,08 | 0,04 | 1,89 | 5,86E-02 | 2,48E-01 |
| ZDHHC21 | 1669,95 | 0,18 | 0,10 | 1,89 | 5,86E-02 | 2,48E-01 |
| DYRK1B | 265,33 | -0,15 | 0,08 | -1,89 | 5,87E-02 | 2,49E-01 |
| RALGPS1 | 581,77 | -0,11 | 0,06 | -1,89 | 5,87E-02 | 2,49E-01 |
| ACVRL1 | 46,11 | -0,35 | 0,18 | -1,89 | 5,87E-02 | 2,49E-01 |
| YME1L1 | 4219,30 | 0,09 | 0,05 | 1,89 | 5,87E-02 | 2,49E-01 |
| HIKESHI | 511,73 | 0,10 | 0,05 | 1,89 | 5,88E-02 | 2,49E-01 |
| PANK3 | 3531,74 | 0,11 | 0,06 | 1,89 | 5,88E-02 | 2,49E-01 |
| NCKAP1 | 347,97 | -0,15 | 0,08 | -1,89 | 5,88E-02 | 2,49E-01 |
| DPAGT1 | 169,93 | -0,10 | 0,05 | -1,89 | 5,88E-02 | 2,49E-01 |
| PLXNA3 | 338,13 | -0,23 | 0,12 | -1,89 | 5,89E-02 | 2,49E-01 |
| RAB39B | 349,18 | 0,11 | 0,06 | 1,89 | 5,90E-02 | 2,49E-01 |
| GOLGA7B | 242,00 | -0,14 | 0,08 | -1,89 | 5,89E-02 | 2,49E-01 |
| C6orf136 | 121,11 | -0,11 | 0,06 | -1,89 | 5,90E-02 | 2,49E-01 |
| PCDHGB3 | 49,55 | -0,22 | 0,11 | -1,89 | 5,89E-02 | 2,49E-01 |
| NT5DC1 | 691,89 | 0,10 | 0,05 | 1,89 | 5,90E-02 | 2,49E-01 |
| REXO5 | 70,37 | -0,22 | 0,12 | -1,89 | 5,90E-02 | 2,49E-01 |
| TLR5 | 971,96 | -0,16 | 0,09 | -1,89 | 5,89E-02 | 2,49E-01 |
| ENOPH1 | 469,74 | 0,10 | 0,05 | 1,89 | 5,90E-02 | 2,49E-01 |
| ENSG000002 | 141,10 | 0,18 | 0,09 | 1,89 | 5,91E-02 | 2,49E-01 |
| VPS35 | 3190,30 | 0,08 | 0,04 | 1,89 | 5,91E-02 | 2,49E-01 |
| RUBCN | 1084,68 | -0,16 | 0,08 | -1,89 | 5,91E-02 | 2,49E-01 |
| NKIRAS2 | 440,13 | -0,09 | 0,05 | -1,89 | 5,91E-02 | 2,49E-01 |
| PNISR | 9243,71 | 0,12 | 0,06 | 1,89 | 5,92E-02 | 2,49E-01 |
| LYSMD3 | 1440,34 | 0,20 | 0,11 | 1,89 | 5,93E-02 | 2,49E-01 |
| PPP1R37 | 194,37 | -0,20 | 0,11 | -1,89 | 5,93E-02 | 2,49E-01 |
| MARCKSL1 | 100,99 | -0,21 | 0,11 | -1,89 | 5,93E-02 | 2,49E-01 |
| EIF4E | 1343,13 | 0,10 | 0,06 | 1,89 | 5,93E-02 | 2,49E-01 |
| DDX27 | 983,04 | 0,07 | 0,04 | 1,89 | 5,93E-02 | 2,49E-01 |
| TAP1 | 6111,04 | -0,13 | 0,07 | -1,88 | 5,98E-02 | 2,49E-01 |
| GALNT1 | 1677,84 | 0,16 | 0,08 | 1,88 | 5,99E-02 | 2,49E-01 |

| | | | | | | |
|---|---|---|---|---|---|---|
| PNPT1 | 657,17 | 0,12 | 0,07 | 1,88 | 5,98E-02 | 2,49E-01 |
| KICS2 | 393,16 | 0,08 | 0,04 | 1,88 | 5,95E-02 | 2,49E-01 |
| MRC2 | 806,72 | -0,25 | 0,13 | -1,88 | 5,96E-02 | 2,49E-01 |
| RUFY4 | 65,12 | -0,28 | 0,15 | -1,88 | 5,95E-02 | 2,49E-01 |
| ZNF501 | 137,01 | 0,18 | 0,09 | 1,88 | 5,99E-02 | 2,49E-01 |
| KLHL9 | 2179,42 | 0,15 | 0,08 | 1,88 | 5,96E-02 | 2,49E-01 |
| GMPR | 2293,65 | -0,28 | 0,15 | -1,88 | 5,96E-02 | 2,49E-01 |
| GGNBP2 | 2786,27 | 0,07 | 0,04 | 1,88 | 5,99E-02 | 2,49E-01 |
| COX11 | 1078,84 | 0,09 | 0,05 | 1,88 | 5,94E-02 | 2,49E-01 |
| SMG8 | 600,92 | 0,07 | 0,04 | 1,88 | 5,97E-02 | 2,49E-01 |
| LUC7L2 | 1527,25 | 0,09 | 0,05 | 1,88 | 5,96E-02 | 2,49E-01 |
| RPL24 | 4607,99 | 0,22 | 0,12 | 1,89 | 5,94E-02 | 2,49E-01 |
| MVB12A | 81,94 | -0,24 | 0,13 | -1,88 | 5,98E-02 | 2,49E-01 |
| TAL1 | 1588,95 | -0,23 | 0,12 | -1,88 | 5,99E-02 | 2,49E-01 |
| RPSA | 9767,72 | 0,17 | 0,09 | 1,88 | 5,98E-02 | 2,49E-01 |
| MCF2L | 293,14 | -0,27 | 0,15 | -1,88 | 5,99E-02 | 2,49E-01 |
| ZMYM2 | 5253,89 | 0,11 | 0,06 | 1,88 | 5,99E-02 | 2,49E-01 |
| DOCK10 | 7638,32 | 0,10 | 0,05 | 1,88 | 5,97E-02 | 2,49E-01 |
| MED21 | 872,67 | 0,11 | 0,06 | 1,88 | 5,97E-02 | 2,49E-01 |
| CHRAC1 | 234,54 | -0,07 | 0,04 | -1,88 | 5,95E-02 | 2,49E-01 |
| SPACA6 | 235,81 | -0,18 | 0,10 | -1,88 | 5,97E-02 | 2,49E-01 |
| RNF24 | 4054,04 | -0,21 | 0,11 | -1,88 | 5,95E-02 | 2,49E-01 |
| ZNF711 | 74,95 | 0,21 | 0,11 | 1,88 | 5,96E-02 | 2,49E-01 |
| YWHAQ | 2014,18 | 0,09 | 0,05 | 1,88 | 5,99E-02 | 2,49E-01 |
| FUBP1 | 5324,83 | 0,12 | 0,06 | 1,88 | 5,99E-02 | 2,49E-01 |
| PLB1 | 817,37 | -0,24 | 0,13 | -1,88 | 6,00E-02 | 2,49E-01 |
| HK1 | 2270,45 | -0,18 | 0,09 | -1,88 | 6,02E-02 | 2,50E-01 |
| ZNF518A | 2775,07 | 0,16 | 0,09 | 1,88 | 6,02E-02 | 2,50E-01 |
| CDCA7 | 74,20 | -0,19 | 0,10 | -1,88 | 6,02E-02 | 2,50E-01 |
| IFT80 | 440,92 | 0,12 | 0,06 | 1,88 | 6,03E-02 | 2,50E-01 |
| TMEM14A | 128,86 | 0,14 | 0,07 | 1,88 | 6,03E-02 | 2,50E-01 |
| ANKRD36 | 3144,88 | 0,12 | 0,06 | 1,88 | 6,03E-02 | 2,50E-01 |
| PEX11G | 30,26 | -0,30 | 0,16 | -1,88 | 6,03E-02 | 2,50E-01 |
| ZNF350 | 958,26 | 0,10 | 0,06 | 1,88 | 6,03E-02 | 2,50E-01 |
| ACTR1A | 1440,81 | -0,08 | 0,04 | -1,88 | 6,03E-02 | 2,50E-01 |
| WDR48 | 2046,98 | 0,08 | 0,04 | 1,88 | 6,04E-02 | 2,50E-01 |
| PCGF3 | 1571,90 | -0,09 | 0,05 | -1,88 | 6,04E-02 | 2,50E-01 |
| NDST2 | 92,99 | -0,17 | 0,09 | -1,88 | 6,04E-02 | 2,50E-01 |
| RPL35A | 4025,01 | 0,23 | 0,12 | 1,88 | 6,04E-02 | 2,50E-01 |
| NR1D1 | 36,15 | -0,26 | 0,14 | -1,88 | 6,05E-02 | 2,50E-01 |
| INHBA | 49,02 | -0,45 | 0,24 | -1,88 | 6,05E-02 | 2,50E-01 |
| SPIN1 | 738,48 | 0,06 | 0,03 | 1,88 | 6,05E-02 | 2,50E-01 |
| KLC4 | 280,42 | -0,12 | 0,07 | -1,88 | 6,06E-02 | 2,51E-01 |
| POLL | 840,29 | -0,17 | 0,09 | -1,88 | 6,06E-02 | 2,51E-01 |
| KLHL34 | 31,43 | -0,25 | 0,13 | -1,88 | 6,08E-02 | 2,51E-01 |
| MRPS2 | 176,85 | -0,12 | 0,06 | -1,87 | 6,08E-02 | 2,51E-01 |

| | | | | | | |
|---|---|---|---|---|---|---|
| SLC30A1 | 1255,21 | 0,19 | 0,10 | 1,87 | 6,08E-02 | 2,51E-01 |
| AHDC1 | 125,51 | -0,28 | 0,15 | -1,87 | 6,08E-02 | 2,51E-01 |
| TSTD2 | 598,93 | 0,06 | 0,03 | 1,87 | 6,09E-02 | 2,51E-01 |
| CBLL1 | 1078,81 | 0,12 | 0,07 | 1,87 | 6,09E-02 | 2,51E-01 |
| OXER1 | 58,90 | -0,29 | 0,15 | -1,87 | 6,10E-02 | 2,51E-01 |
| PSMD1 | 1783,06 | 0,08 | 0,04 | 1,87 | 6,10E-02 | 2,51E-01 |
| FCGBP | 261,96 | -0,30 | 0,16 | -1,87 | 6,10E-02 | 2,51E-01 |
| ERLEC1 | 658,28 | 0,13 | 0,07 | 1,87 | 6,09E-02 | 2,51E-01 |
| RBM12B | 3651,90 | 0,15 | 0,08 | 1,87 | 6,10E-02 | 2,51E-01 |
| BUB3 | 2700,84 | 0,09 | 0,05 | 1,87 | 6,10E-02 | 2,51E-01 |
| SNX4 | 489,56 | 0,18 | 0,10 | 1,87 | 6,11E-02 | 2,51E-01 |
| HMGB1 | 5217,07 | 0,19 | 0,10 | 1,87 | 6,10E-02 | 2,51E-01 |
| MOCS2 | 308,03 | 0,13 | 0,07 | 1,87 | 6,11E-02 | 2,51E-01 |
| SLC35A2 | 192,42 | -0,09 | 0,05 | -1,87 | 6,11E-02 | 2,51E-01 |
| MARCHF2 | 2568,46 | -0,21 | 0,11 | -1,87 | 6,11E-02 | 2,51E-01 |
| SMAD5 | 1492,97 | 0,14 | 0,07 | 1,87 | 6,12E-02 | 2,51E-01 |
| SKIL | 1690,70 | 0,16 | 0,08 | 1,87 | 6,12E-02 | 2,52E-01 |
| TCL1A | 563,09 | 0,27 | 0,14 | 1,87 | 6,13E-02 | 2,52E-01 |
| CLCC1 | 924,45 | 0,07 | 0,04 | 1,87 | 6,13E-02 | 2,52E-01 |
| DIP2A | 2865,62 | -0,13 | 0,07 | -1,87 | 6,17E-02 | 2,52E-01 |
| HDDC2 | 546,57 | 0,12 | 0,06 | 1,87 | 6,18E-02 | 2,52E-01 |
| XRCC6 | 4440,32 | 0,11 | 0,06 | 1,87 | 6,17E-02 | 2,52E-01 |
| THAP5 | 1229,39 | 0,16 | 0,09 | 1,87 | 6,18E-02 | 2,52E-01 |
| ENO2 | 306,81 | -0,14 | 0,08 | -1,87 | 6,19E-02 | 2,52E-01 |
| MEGF8 | 240,37 | -0,23 | 0,12 | -1,87 | 6,15E-02 | 2,52E-01 |
| KLF1 | 1289,01 | -0,27 | 0,15 | -1,87 | 6,13E-02 | 2,52E-01 |
| USB1 | 919,22 | -0,14 | 0,07 | -1,87 | 6,16E-02 | 2,52E-01 |
| SORBS3 | 171,77 | -0,22 | 0,12 | -1,87 | 6,16E-02 | 2,52E-01 |
| TNFRSF11A | 74,04 | -0,33 | 0,18 | -1,87 | 6,16E-02 | 2,52E-01 |
| CRLS1 | 927,60 | 0,09 | 0,05 | 1,87 | 6,15E-02 | 2,52E-01 |
| ZNF836 | 417,78 | 0,12 | 0,07 | 1,87 | 6,15E-02 | 2,52E-01 |
| PHOSPHO1 | 3177,20 | -0,23 | 0,12 | -1,87 | 6,17E-02 | 2,52E-01 |
| ATG12 | 2953,99 | 0,12 | 0,07 | 1,87 | 6,18E-02 | 2,52E-01 |
| AP4M1 | 122,98 | -0,14 | 0,07 | -1,87 | 6,19E-02 | 2,52E-01 |
| DZIP3 | 718,46 | 0,17 | 0,09 | 1,87 | 6,18E-02 | 2,52E-01 |
| RFC3 | 289,70 | 0,11 | 0,06 | 1,87 | 6,14E-02 | 2,52E-01 |
| VPS28 | 1418,32 | -0,12 | 0,06 | -1,87 | 6,14E-02 | 2,52E-01 |
| ACSF2 | 306,42 | -0,13 | 0,07 | -1,87 | 6,14E-02 | 2,52E-01 |
| EID2B | 156,78 | 0,10 | 0,06 | 1,87 | 6,16E-02 | 2,52E-01 |
| MCAT | 139,95 | -0,08 | 0,04 | -1,87 | 6,18E-02 | 2,52E-01 |
| TTC13 | 2041,40 | 0,09 | 0,05 | 1,87 | 6,16E-02 | 2,52E-01 |
| SIRT7 | 1564,90 | -0,08 | 0,05 | -1,87 | 6,19E-02 | 2,52E-01 |
| RAB7A | 1026,68 | -0,13 | 0,07 | -1,87 | 6,18E-02 | 2,52E-01 |
| STYX | 1764,22 | 0,13 | 0,07 | 1,87 | 6,18E-02 | 2,52E-01 |
| CD163 | 1022,10 | 0,14 | 0,08 | 1,87 | 6,18E-02 | 2,52E-01 |
| RAP1GAP | 239,81 | -0,56 | 0,30 | -1,87 | 6,15E-02 | 2,52E-01 |

| | | | | | | |
|---|---|---|---|---|---|---|
| TRMT12 | 529,54 | 0,13 | 0,07 | 1,87 | 6,18E-02 | 2,52E-01 |
| NUP214 | 3201,99 | -0,07 | 0,04 | -1,87 | 6,21E-02 | 2,52E-01 |
| LZTR1 | 444,88 | -0,12 | 0,06 | -1,87 | 6,20E-02 | 2,52E-01 |
| PSMA3 | 737,65 | 0,12 | 0,06 | 1,87 | 6,21E-02 | 2,52E-01 |
| MRPL39 | 280,41 | 0,12 | 0,06 | 1,87 | 6,21E-02 | 2,52E-01 |
| TMEM128 | 387,42 | 0,11 | 0,06 | 1,87 | 6,20E-02 | 2,52E-01 |
| ZBTB6 | 990,82 | 0,18 | 0,09 | 1,87 | 6,19E-02 | 2,52E-01 |
| RPL14 | 7904,78 | 0,14 | 0,07 | 1,87 | 6,21E-02 | 2,52E-01 |
| METTL15 | 733,42 | 0,11 | 0,06 | 1,87 | 6,21E-02 | 2,52E-01 |
| CD164 | 8520,03 | 0,13 | 0,07 | 1,87 | 6,20E-02 | 2,52E-01 |
| APOOL | 297,71 | 0,08 | 0,04 | 1,87 | 6,21E-02 | 2,52E-01 |
| TMEM53 | 47,74 | -0,18 | 0,10 | -1,87 | 6,20E-02 | 2,52E-01 |
| PLCB1 | 1376,08 | 0,12 | 0,06 | 1,86 | 6,22E-02 | 2,52E-01 |
| NUDCD2 | 407,48 | 0,11 | 0,06 | 1,86 | 6,23E-02 | 2,52E-01 |
| ENO3 | 130,53 | -0,11 | 0,06 | -1,86 | 6,23E-02 | 2,52E-01 |
| CHST14 | 73,00 | -0,18 | 0,10 | -1,86 | 6,24E-02 | 2,53E-01 |
| CDK20 | 87,69 | -0,18 | 0,10 | -1,86 | 6,25E-02 | 2,53E-01 |
| MXD1 | 22857,83 | -0,14 | 0,08 | -1,86 | 6,25E-02 | 2,53E-01 |
| RCCD1 | 189,45 | -0,18 | 0,10 | -1,86 | 6,25E-02 | 2,53E-01 |
| FKBP1A | 1704,51 | -0,14 | 0,07 | -1,86 | 6,26E-02 | 2,53E-01 |
| ERCC8 | 468,91 | 0,13 | 0,07 | 1,86 | 6,26E-02 | 2,53E-01 |
| ZFPL1 | 217,82 | -0,08 | 0,05 | -1,86 | 6,27E-02 | 2,53E-01 |
| F12 | 34,77 | -0,27 | 0,14 | -1,86 | 6,27E-02 | 2,53E-01 |
| C1RL | 2213,52 | -0,13 | 0,07 | -1,86 | 6,28E-02 | 2,53E-01 |
| ENSG000001 | 66,52 | -0,27 | 0,14 | -1,86 | 6,28E-02 | 2,53E-01 |
| OLIG2 | 122,37 | -0,30 | 0,16 | -1,86 | 6,28E-02 | 2,54E-01 |
| ASCC3 | 3094,45 | 0,14 | 0,08 | 1,86 | 6,29E-02 | 2,54E-01 |
| QDPR | 118,47 | -0,15 | 0,08 | -1,86 | 6,29E-02 | 2,54E-01 |
| PRAF2 | 33,28 | -0,19 | 0,10 | -1,86 | 6,29E-02 | 2,54E-01 |
| CBFB | 2154,52 | 0,09 | 0,05 | 1,86 | 6,30E-02 | 2,54E-01 |
| ERP44 | 1555,66 | 0,10 | 0,06 | 1,86 | 6,30E-02 | 2,54E-01 |
| KCTD6 | 254,87 | 0,08 | 0,04 | 1,86 | 6,32E-02 | 2,54E-01 |
| POLE3 | 817,95 | 0,09 | 0,05 | 1,86 | 6,33E-02 | 2,54E-01 |
| ALOX15 | 315,14 | -0,35 | 0,19 | -1,86 | 6,33E-02 | 2,54E-01 |
| FAM76B | 1595,49 | 0,16 | 0,08 | 1,86 | 6,32E-02 | 2,54E-01 |
| KPNA3 | 849,31 | 0,10 | 0,06 | 1,86 | 6,31E-02 | 2,54E-01 |
| EEF1AKMT2 | 346,46 | 0,12 | 0,07 | 1,86 | 6,31E-02 | 2,54E-01 |
| TIGD7 | 443,43 | 0,17 | 0,09 | 1,86 | 6,31E-02 | 2,54E-01 |
| VPS36 | 2141,33 | 0,08 | 0,04 | 1,86 | 6,32E-02 | 2,54E-01 |
| RRM1 | 595,86 | 0,09 | 0,05 | 1,86 | 6,33E-02 | 2,54E-01 |
| NDUFV2 | 450,42 | 0,08 | 0,04 | 1,86 | 6,32E-02 | 2,54E-01 |
| WIPI2 | 2028,89 | -0,11 | 0,06 | -1,86 | 6,33E-02 | 2,54E-01 |
| FRA10AC1 | 952,34 | 0,13 | 0,07 | 1,86 | 6,34E-02 | 2,54E-01 |
| MAPK3 | 503,21 | -0,14 | 0,07 | -1,86 | 6,34E-02 | 2,54E-01 |
| SLC26A2 | 901,90 | 0,08 | 0,04 | 1,86 | 6,34E-02 | 2,54E-01 |
| DNAJC9 | 856,77 | 0,09 | 0,05 | 1,86 | 6,35E-02 | 2,54E-01 |

| | | | | | | |
|---|---|---|---|---|---|---|
| MRFAP1L1 | 1618,99 | 0,14 | 0,07 | 1,86 | 6,35E-02 | 2,54E-01 |
| MAP1LC3B2 | 51,68 | -0,22 | 0,12 | -1,85 | 6,36E-02 | 2,55E-01 |
| CLEC1A | 93,07 | -0,16 | 0,09 | -1,85 | 6,36E-02 | 2,55E-01 |
| MBIP | 307,06 | 0,15 | 0,08 | 1,85 | 6,37E-02 | 2,55E-01 |
| CDS1 | 33,42 | -0,28 | 0,15 | -1,85 | 6,37E-02 | 2,55E-01 |
| KIF3A | 533,90 | 0,11 | 0,06 | 1,85 | 6,39E-02 | 2,55E-01 |
| DYRK2 | 5568,53 | 0,11 | 0,06 | 1,85 | 6,39E-02 | 2,55E-01 |
| FAM107B | 4396,43 | 0,07 | 0,04 | 1,85 | 6,38E-02 | 2,55E-01 |
| SRI | 1095,93 | 0,10 | 0,05 | 1,85 | 6,39E-02 | 2,55E-01 |
| MAP2K2 | 1061,03 | -0,16 | 0,09 | -1,85 | 6,39E-02 | 2,55E-01 |
| AP2A2 | 742,59 | -0,16 | 0,09 | -1,85 | 6,39E-02 | 2,55E-01 |
| NHP2 | 506,21 | 0,09 | 0,05 | 1,85 | 6,38E-02 | 2,55E-01 |
| POLR2J | 532,94 | -0,09 | 0,05 | -1,85 | 6,40E-02 | 2,55E-01 |
| DNAJB9 | 519,22 | 0,18 | 0,10 | 1,85 | 6,40E-02 | 2,55E-01 |
| THEMIS | 2144,67 | 0,17 | 0,09 | 1,85 | 6,41E-02 | 2,56E-01 |
| PRRG4 | 1136,46 | -0,17 | 0,09 | -1,85 | 6,41E-02 | 2,56E-01 |
| HSP90AA1 | 2927,99 | 0,18 | 0,10 | 1,85 | 6,42E-02 | 2,56E-01 |
| GDPD5 | 425,34 | -0,21 | 0,11 | -1,85 | 6,42E-02 | 2,56E-01 |
| AK2 | 2770,14 | 0,07 | 0,04 | 1,85 | 6,43E-02 | 2,56E-01 |
| FLOT1 | 2184,57 | -0,12 | 0,06 | -1,85 | 6,44E-02 | 2,56E-01 |
| NDUFAB1 | 491,66 | 0,09 | 0,05 | 1,85 | 6,44E-02 | 2,56E-01 |
| ADCY3 | 217,07 | -0,18 | 0,10 | -1,85 | 6,44E-02 | 2,56E-01 |
| RMDN2 | 274,71 | 0,11 | 0,06 | 1,85 | 6,43E-02 | 2,56E-01 |
| MRPS9 | 427,65 | 0,11 | 0,06 | 1,85 | 6,44E-02 | 2,56E-01 |
| GOLGA8K | 21,48 | -0,32 | 0,17 | -1,85 | 6,43E-02 | 2,56E-01 |
| FAM117A | 1609,17 | -0,20 | 0,11 | -1,85 | 6,44E-02 | 2,56E-01 |
| ACTR8 | 1181,59 | 0,07 | 0,04 | 1,85 | 6,43E-02 | 2,56E-01 |
| PHF1 | 623,07 | -0,12 | 0,07 | -1,85 | 6,44E-02 | 2,56E-01 |
| FXR1 | 2772,23 | 0,12 | 0,07 | 1,85 | 6,45E-02 | 2,56E-01 |
| TRMT10A | 155,95 | 0,17 | 0,09 | 1,85 | 6,45E-02 | 2,56E-01 |
| PRKG1 | 69,09 | -0,34 | 0,18 | -1,85 | 6,46E-02 | 2,56E-01 |
| TSNAX | 1294,13 | 0,15 | 0,08 | 1,85 | 6,46E-02 | 2,56E-01 |
| METTL22 | 968,89 | -0,10 | 0,05 | -1,84 | 6,51E-02 | 2,56E-01 |
| CD1D | 1262,98 | 0,12 | 0,06 | 1,84 | 6,51E-02 | 2,56E-01 |
| SRSF1 | 1771,95 | 0,13 | 0,07 | 1,85 | 6,49E-02 | 2,56E-01 |
| CNOT2 | 4304,46 | 0,10 | 0,06 | 1,84 | 6,51E-02 | 2,56E-01 |
| SLC4A8 | 533,58 | -0,14 | 0,08 | -1,84 | 6,51E-02 | 2,56E-01 |
| DPM1 | 707,60 | 0,16 | 0,09 | 1,85 | 6,50E-02 | 2,56E-01 |
| NFS1 | 384,95 | -0,06 | 0,03 | -1,85 | 6,49E-02 | 2,56E-01 |
| ADAM12 | 104,75 | -0,33 | 0,18 | -1,85 | 6,48E-02 | 2,56E-01 |
| FUBP3 | 1293,80 | 0,05 | 0,03 | 1,84 | 6,51E-02 | 2,56E-01 |
| NOMO1 | 348,39 | -0,13 | 0,07 | -1,84 | 6,51E-02 | 2,56E-01 |
| BZW1 | 3497,30 | 0,12 | 0,07 | 1,85 | 6,47E-02 | 2,56E-01 |
| ORC3 | 607,54 | 0,10 | 0,06 | 1,85 | 6,50E-02 | 2,56E-01 |
| SLC66A1 | 121,67 | -0,14 | 0,08 | -1,85 | 6,47E-02 | 2,56E-01 |
| STK33 | 66,98 | -0,35 | 0,19 | -1,85 | 6,50E-02 | 2,56E-01 |

| | | | | | | |
|---|---|---|---|---|---|---|
| IL5RA | 779,94 | -0,24 | 0,13 | -1,85 | 6,48E-02 | 2,56E-01 |
| FBXL18 | 118,15 | -0,25 | 0,14 | -1,85 | 6,47E-02 | 2,56E-01 |
| DUSP18 | 353,65 | -0,12 | 0,07 | -1,84 | 6,51E-02 | 2,56E-01 |
| CLPTM1 | 367,77 | -0,17 | 0,09 | -1,85 | 6,47E-02 | 2,56E-01 |
| TBC1D23 | 2179,50 | 0,13 | 0,07 | 1,85 | 6,49E-02 | 2,56E-01 |
| TMF1 | 3546,28 | 0,13 | 0,07 | 1,85 | 6,49E-02 | 2,56E-01 |
| GAPT | 1263,58 | 0,19 | 0,10 | 1,85 | 6,50E-02 | 2,56E-01 |
| SYTL2 | 2178,37 | 0,19 | 0,10 | 1,85 | 6,48E-02 | 2,56E-01 |
| ZNF655 | 4201,23 | 0,06 | 0,03 | 1,85 | 6,48E-02 | 2,56E-01 |
| PRR7 | 185,73 | -0,13 | 0,07 | -1,85 | 6,49E-02 | 2,56E-01 |
| HDX | 110,00 | 0,14 | 0,08 | 1,84 | 6,52E-02 | 2,57E-01 |
| MTIF3 | 594,90 | 0,09 | 0,05 | 1,84 | 6,52E-02 | 2,57E-01 |
| PRSS36 | 63,26 | -0,20 | 0,11 | -1,84 | 6,52E-02 | 2,57E-01 |
| BCL2 | 5452,60 | 0,13 | 0,07 | 1,84 | 6,53E-02 | 2,57E-01 |
| MPC1 | 624,95 | 0,10 | 0,05 | 1,84 | 6,54E-02 | 2,57E-01 |
| RILP | 1432,63 | -0,24 | 0,13 | -1,84 | 6,54E-02 | 2,57E-01 |
| NUCB1 | 1106,56 | -0,20 | 0,11 | -1,84 | 6,57E-02 | 2,57E-01 |
| TMEM167A | 2804,76 | 0,17 | 0,09 | 1,84 | 6,57E-02 | 2,57E-01 |
| GTF2H1 | 1372,66 | 0,08 | 0,04 | 1,84 | 6,54E-02 | 2,57E-01 |
| SNX25 | 488,97 | 0,11 | 0,06 | 1,84 | 6,57E-02 | 2,57E-01 |
| ARID3B | 579,61 | -0,17 | 0,09 | -1,84 | 6,55E-02 | 2,57E-01 |
| PLEKHH2 | 61,29 | -0,37 | 0,20 | -1,84 | 6,57E-02 | 2,57E-01 |
| GSTK1 | 2567,93 | -0,08 | 0,04 | -1,84 | 6,55E-02 | 2,57E-01 |
| NFIA | 463,96 | -0,13 | 0,07 | -1,84 | 6,57E-02 | 2,57E-01 |
| DLGAP4 | 237,17 | -0,30 | 0,16 | -1,84 | 6,56E-02 | 2,57E-01 |
| MED6 | 927,93 | 0,08 | 0,04 | 1,84 | 6,55E-02 | 2,57E-01 |
| PFKM | 292,66 | -0,10 | 0,05 | -1,84 | 6,55E-02 | 2,57E-01 |
| FAN1 | 1038,77 | 0,09 | 0,05 | 1,84 | 6,57E-02 | 2,57E-01 |
| PPARA | 550,36 | -0,10 | 0,06 | -1,84 | 6,57E-02 | 2,57E-01 |
| SPAG5 | 93,77 | -0,15 | 0,08 | -1,84 | 6,57E-02 | 2,57E-01 |
| SOCS1 | 61,06 | -0,22 | 0,12 | -1,84 | 6,57E-02 | 2,57E-01 |
| TMPRSS9 | 417,53 | -0,30 | 0,16 | -1,84 | 6,56E-02 | 2,57E-01 |
| AGTRAP | 850,00 | -0,14 | 0,07 | -1,84 | 6,57E-02 | 2,57E-01 |
| GPR180 | 529,69 | 0,14 | 0,08 | 1,84 | 6,58E-02 | 2,57E-01 |
| GPATCH8 | 2527,95 | 0,06 | 0,03 | 1,84 | 6,58E-02 | 2,57E-01 |
| DNAAF5 | 202,64 | -0,18 | 0,10 | -1,84 | 6,58E-02 | 2,57E-01 |
| OAZ1 | 44328,78 | -0,17 | 0,09 | -1,84 | 6,59E-02 | 2,57E-01 |
| ERCC4 | 530,79 | 0,06 | 0,03 | 1,84 | 6,59E-02 | 2,57E-01 |
| TMEM169 | 124,36 | -0,09 | 0,05 | -1,84 | 6,59E-02 | 2,57E-01 |
| HAGHL | 109,12 | -0,20 | 0,11 | -1,84 | 6,62E-02 | 2,57E-01 |
| SLC25A40 | 1269,17 | 0,13 | 0,07 | 1,84 | 6,62E-02 | 2,57E-01 |
| NDUFA5 | 569,04 | 0,19 | 0,10 | 1,84 | 6,61E-02 | 2,57E-01 |
| WRAP53 | 129,68 | -0,10 | 0,05 | -1,84 | 6,60E-02 | 2,57E-01 |
| KRCC1 | 674,33 | 0,17 | 0,09 | 1,84 | 6,62E-02 | 2,57E-01 |
| DPY19L4 | 622,85 | 0,13 | 0,07 | 1,84 | 6,63E-02 | 2,57E-01 |
| LACTB2 | 268,81 | 0,10 | 0,06 | 1,84 | 6,61E-02 | 2,57E-01 |

| | | | | | | |
|---|---|---|---|---|---|---|
| RBM25 | 7091,28 | 0,10 | 0,06 | 1,84 | 6,63E-02 | 2,57E-01 |
| SCAMP1 | 1226,96 | 0,13 | 0,07 | 1,84 | 6,63E-02 | 2,57E-01 |
| RNF144A | 1204,92 | -0,13 | 0,07 | -1,84 | 6,61E-02 | 2,57E-01 |
| SBK1 | 143,50 | -0,29 | 0,16 | -1,84 | 6,62E-02 | 2,57E-01 |
| AP3S2 | 558,04 | -0,11 | 0,06 | -1,84 | 6,61E-02 | 2,57E-01 |
| GTPBP8 | 452,09 | 0,12 | 0,07 | 1,84 | 6,63E-02 | 2,57E-01 |
| MCOLN3 | 46,02 | -0,31 | 0,17 | -1,84 | 6,63E-02 | 2,57E-01 |
| SHISA5 | 1984,53 | -0,14 | 0,08 | -1,84 | 6,63E-02 | 2,57E-01 |
| MANEA | 511,81 | 0,19 | 0,10 | 1,84 | 6,64E-02 | 2,58E-01 |
| ASTE1 | 664,16 | 0,10 | 0,05 | 1,84 | 6,64E-02 | 2,58E-01 |
| FRG1 | 509,81 | 0,13 | 0,07 | 1,83 | 6,66E-02 | 2,58E-01 |
| ARSA | 225,49 | -0,27 | 0,15 | -1,84 | 6,65E-02 | 2,58E-01 |
| TMEM150A | 59,60 | -0,21 | 0,11 | -1,83 | 6,66E-02 | 2,58E-01 |
| SPCS3 | 7238,17 | 0,13 | 0,07 | 1,83 | 6,66E-02 | 2,58E-01 |
| FKBP8 | 24037,65 | -0,26 | 0,14 | -1,84 | 6,65E-02 | 2,58E-01 |
| TPT1 | 11996,27 | 0,24 | 0,13 | 1,83 | 6,66E-02 | 2,58E-01 |
| MRPS33 | 282,37 | 0,15 | 0,08 | 1,83 | 6,66E-02 | 2,58E-01 |
| RADX | 228,42 | 0,15 | 0,08 | 1,83 | 6,65E-02 | 2,58E-01 |
| PRKAG1 | 858,54 | 0,07 | 0,04 | 1,83 | 6,66E-02 | 2,58E-01 |
| ETNK1 | 4144,27 | 0,16 | 0,09 | 1,83 | 6,66E-02 | 2,58E-01 |
| DNAJC4 | 141,55 | -0,15 | 0,08 | -1,84 | 6,65E-02 | 2,58E-01 |
| CCDC81 | 38,53 | -0,30 | 0,16 | -1,83 | 6,68E-02 | 2,58E-01 |
| DDX42 | 2660,30 | 0,06 | 0,03 | 1,83 | 6,68E-02 | 2,58E-01 |
| EPC1 | 4211,12 | 0,08 | 0,05 | 1,83 | 6,67E-02 | 2,58E-01 |
| STAT5B | 5599,48 | -0,13 | 0,07 | -1,83 | 6,68E-02 | 2,58E-01 |
| ATPSCKMT | 410,30 | 0,10 | 0,05 | 1,83 | 6,68E-02 | 2,58E-01 |
| GPN1 | 411,91 | 0,08 | 0,04 | 1,83 | 6,69E-02 | 2,58E-01 |
| PROC | 33,12 | -0,31 | 0,17 | -1,83 | 6,69E-02 | 2,58E-01 |
| ZNF566 | 336,30 | 0,10 | 0,06 | 1,83 | 6,70E-02 | 2,58E-01 |
| PDE1B | 225,58 | -0,18 | 0,10 | -1,83 | 6,71E-02 | 2,59E-01 |
| BCL7B | 297,67 | -0,08 | 0,04 | -1,83 | 6,72E-02 | 2,59E-01 |
| BMI1 | 734,47 | 0,17 | 0,09 | 1,83 | 6,72E-02 | 2,59E-01 |
| MTERF3 | 266,71 | 0,09 | 0,05 | 1,83 | 6,71E-02 | 2,59E-01 |
| MTUS1 | 138,71 | -0,30 | 0,16 | -1,83 | 6,71E-02 | 2,59E-01 |
| TAPBP | 2722,07 | -0,20 | 0,11 | -1,83 | 6,71E-02 | 2,59E-01 |
| PCNA | 557,68 | 0,10 | 0,06 | 1,83 | 6,71E-02 | 2,59E-01 |
| SRP54 | 1446,19 | 0,09 | 0,05 | 1,83 | 6,72E-02 | 2,59E-01 |
| TBL2 | 550,79 | 0,06 | 0,03 | 1,83 | 6,73E-02 | 2,59E-01 |
| PMFBP1 | 62,65 | -0,28 | 0,15 | -1,83 | 6,73E-02 | 2,59E-01 |
| TFEC | 1786,83 | 0,23 | 0,13 | 1,83 | 6,73E-02 | 2,59E-01 |
| ATOSB | 1364,51 | -0,15 | 0,08 | -1,83 | 6,73E-02 | 2,59E-01 |
| TBKBP1 | 266,69 | -0,21 | 0,11 | -1,83 | 6,74E-02 | 2,59E-01 |
| RBM12 | 3350,36 | 0,10 | 0,06 | 1,83 | 6,75E-02 | 2,59E-01 |
| KATNB1 | 264,11 | -0,14 | 0,08 | -1,83 | 6,75E-02 | 2,59E-01 |
| EFEMP2 | 96,65 | -0,14 | 0,08 | -1,83 | 6,75E-02 | 2,59E-01 |
| MPEG1 | 6957,67 | 0,11 | 0,06 | 1,83 | 6,75E-02 | 2,59E-01 |

| | | | | | | |
|---|---|---|---|---|---|---|
| PLK2 | 45,45 | -0,21 | 0,12 | -1,83 | 6,76E-02 | 2,59E-01 |
| IFITM1 | 2995,47 | -0,15 | 0,08 | -1,83 | 6,76E-02 | 2,59E-01 |
| SURF1 | 487,19 | -0,07 | 0,04 | -1,83 | 6,77E-02 | 2,59E-01 |
| MED4 | 979,79 | 0,10 | 0,05 | 1,83 | 6,78E-02 | 2,59E-01 |
| FNBP4 | 3750,46 | 0,07 | 0,04 | 1,83 | 6,78E-02 | 2,59E-01 |
| NLGN3 | 60,17 | -0,26 | 0,14 | -1,83 | 6,78E-02 | 2,59E-01 |
| DGKE | 1218,27 | 0,10 | 0,06 | 1,83 | 6,78E-02 | 2,59E-01 |
| ETFDH | 611,16 | 0,09 | 0,05 | 1,83 | 6,79E-02 | 2,60E-01 |
| ZNF436 | 435,20 | 0,08 | 0,04 | 1,83 | 6,78E-02 | 2,60E-01 |
| LRRC8A | 614,17 | -0,14 | 0,07 | -1,83 | 6,79E-02 | 2,60E-01 |
| LOXL2 | 145,21 | -0,17 | 0,09 | -1,83 | 6,79E-02 | 2,60E-01 |
| DEAF1 | 305,54 | -0,14 | 0,08 | -1,82 | 6,80E-02 | 2,60E-01 |
| CLEC2D | 7643,25 | 0,12 | 0,07 | 1,83 | 6,80E-02 | 2,60E-01 |
| CCDC174 | 1089,17 | 0,06 | 0,03 | 1,82 | 6,80E-02 | 2,60E-01 |
| C1orf131 | 491,91 | 0,07 | 0,04 | 1,82 | 6,81E-02 | 2,60E-01 |
| CA4 | 304,70 | -0,20 | 0,11 | -1,82 | 6,81E-02 | 2,60E-01 |
| LIPH | 37,96 | -0,31 | 0,17 | -1,82 | 6,82E-02 | 2,60E-01 |
| LMBRD1 | 2884,81 | 0,13 | 0,07 | 1,82 | 6,83E-02 | 2,60E-01 |
| GP6 | 45,02 | -0,29 | 0,16 | -1,82 | 6,83E-02 | 2,60E-01 |
| DALRD3 | 390,91 | -0,07 | 0,04 | -1,82 | 6,84E-02 | 2,61E-01 |
| PUSL1 | 99,20 | -0,14 | 0,08 | -1,82 | 6,84E-02 | 2,61E-01 |
| IRF2BP2 | 3300,09 | 0,06 | 0,04 | 1,82 | 6,85E-02 | 2,61E-01 |
| HNRNPH1 | 16446,41 | 0,10 | 0,05 | 1,82 | 6,85E-02 | 2,61E-01 |
| CREBZF | 4109,46 | 0,12 | 0,07 | 1,82 | 6,85E-02 | 2,61E-01 |
| RNF139 | 1323,11 | 0,14 | 0,08 | 1,82 | 6,86E-02 | 2,61E-01 |
| ELF4 | 837,79 | -0,20 | 0,11 | -1,82 | 6,86E-02 | 2,61E-01 |
| CCT3 | 1956,74 | 0,08 | 0,04 | 1,82 | 6,88E-02 | 2,62E-01 |
| TRIP10 | 42,73 | -0,27 | 0,15 | -1,82 | 6,88E-02 | 2,62E-01 |
| METTL8 | 728,03 | 0,13 | 0,07 | 1,82 | 6,89E-02 | 2,62E-01 |
| DEDD2 | 2333,12 | -0,10 | 0,06 | -1,82 | 6,88E-02 | 2,62E-01 |
| ATOSA | 3665,71 | 0,10 | 0,05 | 1,82 | 6,88E-02 | 2,62E-01 |
| ZNF671 | 928,67 | 0,09 | 0,05 | 1,82 | 6,89E-02 | 2,62E-01 |
| CCNH | 3935,41 | 0,12 | 0,06 | 1,82 | 6,89E-02 | 2,62E-01 |
| GTPBP10 | 1232,81 | 0,11 | 0,06 | 1,82 | 6,90E-02 | 2,62E-01 |
| PPP6R3 | 6369,96 | 0,07 | 0,04 | 1,82 | 6,91E-02 | 2,62E-01 |
| CNTF | 98,02 | 0,10 | 0,05 | 1,82 | 6,90E-02 | 2,62E-01 |
| RYK | 1351,37 | 0,09 | 0,05 | 1,82 | 6,90E-02 | 2,62E-01 |
| SIAH2 | 3789,08 | -0,23 | 0,13 | -1,82 | 6,92E-02 | 2,62E-01 |
| ERCC2 | 165,73 | -0,16 | 0,09 | -1,82 | 6,91E-02 | 2,62E-01 |
| NARF | 1377,23 | -0,14 | 0,08 | -1,82 | 6,91E-02 | 2,62E-01 |
| RAD21 | 4078,73 | 0,10 | 0,05 | 1,82 | 6,91E-02 | 2,62E-01 |
| TST | 422,56 | -0,13 | 0,07 | -1,82 | 6,92E-02 | 2,62E-01 |
| SMNDC1 | 1031,91 | 0,13 | 0,07 | 1,82 | 6,92E-02 | 2,62E-01 |
| HAT1 | 608,06 | 0,16 | 0,09 | 1,82 | 6,91E-02 | 2,62E-01 |
| COPZ2 | 31,81 | -0,23 | 0,13 | -1,82 | 6,93E-02 | 2,62E-01 |
| KIAA0319 | 267,43 | -0,21 | 0,12 | -1,82 | 6,92E-02 | 2,62E-01 |

| | | | | | | |
|---|---|---|---|---|---|---|
| NPAT | 2613,95 | 0,10 | 0,05 | 1,82 | 6,92E-02 | 2,62E-01 |
| MRPS14 | 547,84 | 0,11 | 0,06 | 1,82 | 6,92E-02 | 2,62E-01 |
| CEP170 | 2988,14 | 0,10 | 0,05 | 1,82 | 6,93E-02 | 2,62E-01 |
| NDUFA9 | 873,81 | 0,08 | 0,04 | 1,82 | 6,94E-02 | 2,62E-01 |
| RPUSD4 | 499,28 | 0,06 | 0,03 | 1,82 | 6,94E-02 | 2,62E-01 |
| ILK | 907,17 | -0,12 | 0,07 | -1,82 | 6,95E-02 | 2,62E-01 |
| MRE11 | 1961,46 | 0,08 | 0,05 | 1,82 | 6,95E-02 | 2,62E-01 |
| RAB11FIP2 | 1634,13 | 0,12 | 0,07 | 1,81 | 6,96E-02 | 2,62E-01 |
| PRKCI | 681,08 | 0,11 | 0,06 | 1,81 | 6,95E-02 | 2,62E-01 |
| THAP1 | 303,65 | 0,13 | 0,07 | 1,81 | 6,97E-02 | 2,63E-01 |
| TIGD1 | 452,99 | 0,16 | 0,09 | 1,81 | 6,97E-02 | 2,63E-01 |
| DLEC1 | 165,40 | -0,20 | 0,11 | -1,81 | 6,97E-02 | 2,63E-01 |
| IDUA | 223,42 | -0,19 | 0,11 | -1,81 | 6,97E-02 | 2,63E-01 |
| RAB3D | 2834,11 | -0,18 | 0,10 | -1,81 | 6,98E-02 | 2,63E-01 |
| SLC18B1 | 684,91 | 0,10 | 0,05 | 1,81 | 6,99E-02 | 2,63E-01 |
| PTMS | 184,05 | -0,25 | 0,14 | -1,81 | 6,99E-02 | 2,63E-01 |
| SLC35B3 | 1002,90 | 0,13 | 0,07 | 1,81 | 7,00E-02 | 2,63E-01 |
| MRPL37 | 378,31 | -0,06 | 0,03 | -1,81 | 7,01E-02 | 2,63E-01 |
| TMEM164 | 2264,89 | -0,22 | 0,12 | -1,81 | 7,00E-02 | 2,63E-01 |
| AQP1 | 86,54 | -0,28 | 0,15 | -1,81 | 7,01E-02 | 2,63E-01 |
| TOMM5 | 197,25 | 0,10 | 0,06 | 1,81 | 7,01E-02 | 2,63E-01 |
| PLOD3 | 271,06 | -0,14 | 0,08 | -1,81 | 7,01E-02 | 2,64E-01 |
| KLHL28 | 1998,28 | 0,13 | 0,07 | 1,81 | 7,03E-02 | 2,64E-01 |
| ZDHHC23 | 277,38 | 0,11 | 0,06 | 1,81 | 7,05E-02 | 2,64E-01 |
| NUDCD1 | 448,07 | 0,20 | 0,11 | 1,81 | 7,03E-02 | 2,64E-01 |
| CLINT1 | 2965,87 | 0,08 | 0,05 | 1,81 | 7,04E-02 | 2,64E-01 |
| MSH4 | 91,94 | -0,13 | 0,07 | -1,81 | 7,05E-02 | 2,64E-01 |
| RAMAC | 469,36 | 0,12 | 0,07 | 1,81 | 7,04E-02 | 2,64E-01 |
| OST4 | 3078,26 | -0,14 | 0,08 | -1,81 | 7,05E-02 | 2,64E-01 |
| ZNF77 | 106,27 | 0,08 | 0,05 | 1,81 | 7,04E-02 | 2,64E-01 |
| UTP14C | 177,62 | 0,13 | 0,07 | 1,81 | 7,03E-02 | 2,64E-01 |
| DENND3 | 11794,89 | -0,17 | 0,09 | -1,81 | 7,03E-02 | 2,64E-01 |
| TRERF1 | 1644,46 | -0,11 | 0,06 | -1,81 | 7,04E-02 | 2,64E-01 |
| FGFR1 | 151,45 | -0,29 | 0,16 | -1,81 | 7,05E-02 | 2,64E-01 |
| ICAM1 | 619,03 | -0,14 | 0,08 | -1,81 | 7,04E-02 | 2,64E-01 |
| MSH6 | 1269,86 | 0,07 | 0,04 | 1,81 | 7,05E-02 | 2,64E-01 |
| FAM204A | 1977,67 | 0,09 | 0,05 | 1,81 | 7,07E-02 | 2,64E-01 |
| CHORDC1 | 2566,21 | 0,13 | 0,07 | 1,81 | 7,07E-02 | 2,64E-01 |
| SMARCE1 | 1104,25 | 0,10 | 0,06 | 1,81 | 7,07E-02 | 2,64E-01 |
| TNNT3 | 67,49 | -0,25 | 0,14 | -1,81 | 7,08E-02 | 2,64E-01 |
| ZFAND3 | 2175,57 | -0,12 | 0,07 | -1,81 | 7,08E-02 | 2,64E-01 |
| NUDT9 | 272,08 | 0,07 | 0,04 | 1,81 | 7,10E-02 | 2,65E-01 |
| RBM17 | 1591,69 | 0,06 | 0,03 | 1,81 | 7,10E-02 | 2,65E-01 |
| AASDH | 625,51 | 0,11 | 0,06 | 1,81 | 7,09E-02 | 2,65E-01 |
| EIF2S2 | 1440,46 | 0,11 | 0,06 | 1,81 | 7,09E-02 | 2,65E-01 |
| NR2C1 | 676,84 | 0,09 | 0,05 | 1,81 | 7,09E-02 | 2,65E-01 |

| | | | | | | |
|---|---|---|---|---|---|---|
| SLC30A6 | 1260,64 | 0,09 | 0,05 | 1,81 | 7,10E-02 | 2,65E-01 |
| MMP24OS | 515,94 | -0,14 | 0,08 | -1,81 | 7,10E-02 | 2,65E-01 |
| CADM4 | 65,81 | -0,20 | 0,11 | -1,81 | 7,10E-02 | 2,65E-01 |
| ZBTB38 | 2312,94 | 0,11 | 0,06 | 1,80 | 7,14E-02 | 2,65E-01 |
| ANPEP | 1194,75 | -0,27 | 0,15 | -1,80 | 7,14E-02 | 2,65E-01 |
| CEP295 | 1541,88 | 0,09 | 0,05 | 1,80 | 7,14E-02 | 2,65E-01 |
| STK31 | 48,20 | -0,31 | 0,17 | -1,80 | 7,13E-02 | 2,65E-01 |
| DPP8 | 3703,42 | 0,07 | 0,04 | 1,80 | 7,13E-02 | 2,65E-01 |
| PCBP4 | 124,27 | -0,14 | 0,08 | -1,80 | 7,13E-02 | 2,65E-01 |
| SLC5A6 | 214,78 | -0,14 | 0,08 | -1,80 | 7,13E-02 | 2,65E-01 |
| PDCD10 | 1794,14 | 0,16 | 0,09 | 1,80 | 7,14E-02 | 2,65E-01 |
| TMEM255B | 140,80 | -0,20 | 0,11 | -1,80 | 7,13E-02 | 2,65E-01 |
| BNIP2 | 5696,96 | 0,12 | 0,07 | 1,80 | 7,14E-02 | 2,65E-01 |
| SLC10A3 | 181,15 | -0,14 | 0,08 | -1,80 | 7,12E-02 | 2,65E-01 |
| SLITRK4 | 157,67 | 0,18 | 0,10 | 1,80 | 7,14E-02 | 2,65E-01 |
| CSTF3 | 797,79 | 0,11 | 0,06 | 1,80 | 7,13E-02 | 2,65E-01 |
| ZNF181 | 417,19 | 0,18 | 0,10 | 1,80 | 7,15E-02 | 2,65E-01 |
| PRR11 | 127,01 | -0,13 | 0,07 | -1,80 | 7,16E-02 | 2,65E-01 |
| TCP1 | 2420,55 | 0,08 | 0,04 | 1,80 | 7,16E-02 | 2,65E-01 |
| ROCK1 | 6615,43 | 0,11 | 0,06 | 1,80 | 7,16E-02 | 2,65E-01 |
| SCNM1 | 238,91 | -0,11 | 0,06 | -1,80 | 7,17E-02 | 2,66E-01 |
| SIRPA | 2492,79 | -0,16 | 0,09 | -1,80 | 7,17E-02 | 2,66E-01 |
| CHIT1 | 167,56 | -0,16 | 0,09 | -1,80 | 7,17E-02 | 2,66E-01 |
| AACS | 335,08 | -0,18 | 0,10 | -1,80 | 7,17E-02 | 2,66E-01 |
| PSAT1 | 64,27 | -0,17 | 0,09 | -1,80 | 7,17E-02 | 2,66E-01 |
| ADAM8 | 6288,54 | -0,15 | 0,09 | -1,80 | 7,17E-02 | 2,66E-01 |
| LRIT3 | 24,21 | -0,23 | 0,13 | -1,80 | 7,18E-02 | 2,66E-01 |
| TTC5 | 1023,64 | 0,06 | 0,03 | 1,80 | 7,19E-02 | 2,66E-01 |
| TUT4 | 2970,38 | 0,08 | 0,05 | 1,80 | 7,18E-02 | 2,66E-01 |
| RIN1 | 83,76 | -0,20 | 0,11 | -1,80 | 7,19E-02 | 2,66E-01 |
| KLRB1 | 635,78 | 0,24 | 0,13 | 1,80 | 7,18E-02 | 2,66E-01 |
| MTURN | 3029,05 | -0,13 | 0,07 | -1,80 | 7,19E-02 | 2,66E-01 |
| FBXW7 | 3791,26 | 0,08 | 0,04 | 1,80 | 7,19E-02 | 2,66E-01 |
| TMEM184B | 490,61 | -0,20 | 0,11 | -1,80 | 7,20E-02 | 2,66E-01 |
| EGLN3 | 133,39 | -0,21 | 0,12 | -1,80 | 7,20E-02 | 2,66E-01 |
| RNASE6 | 697,66 | 0,17 | 0,09 | 1,80 | 7,21E-02 | 2,66E-01 |
| CSTB | 431,74 | -0,10 | 0,05 | -1,80 | 7,21E-02 | 2,66E-01 |
| AP2M1 | 2056,80 | -0,16 | 0,09 | -1,80 | 7,22E-02 | 2,66E-01 |
| ELP4 | 1694,58 | 0,12 | 0,07 | 1,80 | 7,22E-02 | 2,66E-01 |
| C18orf21 | 197,06 | 0,13 | 0,07 | 1,80 | 7,22E-02 | 2,66E-01 |
| ZNF572 | 136,79 | 0,15 | 0,08 | 1,80 | 7,23E-02 | 2,66E-01 |
| GRHL1 | 112,26 | -0,21 | 0,12 | -1,80 | 7,23E-02 | 2,66E-01 |
| CR1L | 386,17 | -0,22 | 0,12 | -1,80 | 7,23E-02 | 2,66E-01 |
| INTS12 | 335,88 | 0,09 | 0,05 | 1,80 | 7,24E-02 | 2,66E-01 |
| SRC | 148,64 | -0,28 | 0,16 | -1,80 | 7,24E-02 | 2,66E-01 |
| MANSC1 | 509,77 | -0,16 | 0,09 | -1,80 | 7,24E-02 | 2,66E-01 |

| | | | | | | |
|---|---|---|---|---|---|---|
| EIF3G | 3108,33 | 0,09 | 0,05 | 1,80 | 7,25E-02 | 2,67E-01 |
| CEBPZOS | 693,87 | 0,07 | 0,04 | 1,80 | 7,26E-02 | 2,67E-01 |
| AFTPH | 3285,26 | 0,11 | 0,06 | 1,80 | 7,25E-02 | 2,67E-01 |
| SASH3 | 2970,64 | -0,09 | 0,05 | -1,80 | 7,25E-02 | 2,67E-01 |
| HSPA8 | 11831,18 | 0,11 | 0,06 | 1,80 | 7,25E-02 | 2,67E-01 |
| CNBD2 | 58,66 | -0,23 | 0,13 | -1,79 | 7,27E-02 | 2,67E-01 |
| ZNF738 | 281,81 | 0,08 | 0,05 | 1,79 | 7,27E-02 | 2,67E-01 |
| LRRC42 | 113,69 | -0,10 | 0,06 | -1,79 | 7,28E-02 | 2,67E-01 |
| ULK3 | 299,19 | -0,15 | 0,08 | -1,79 | 7,28E-02 | 2,67E-01 |
| ZNF529 | 672,14 | 0,10 | 0,05 | 1,79 | 7,28E-02 | 2,67E-01 |
| ARL5B | 1219,24 | 0,15 | 0,08 | 1,79 | 7,28E-02 | 2,67E-01 |
| MYLK | 562,42 | -0,22 | 0,12 | -1,79 | 7,28E-02 | 2,67E-01 |
| PMS2 | 434,42 | 0,05 | 0,03 | 1,79 | 7,31E-02 | 2,67E-01 |
| CD48 | 5614,36 | 0,13 | 0,07 | 1,79 | 7,31E-02 | 2,67E-01 |
| BEND7 | 91,61 | -0,28 | 0,15 | -1,79 | 7,31E-02 | 2,67E-01 |
| GEMIN2 | 155,81 | 0,12 | 0,07 | 1,79 | 7,30E-02 | 2,67E-01 |
| DAG1 | 275,12 | -0,12 | 0,07 | -1,79 | 7,30E-02 | 2,67E-01 |
| BAG4 | 834,03 | 0,11 | 0,06 | 1,79 | 7,30E-02 | 2,67E-01 |
| ZC3H6 | 1640,73 | 0,09 | 0,05 | 1,79 | 7,31E-02 | 2,67E-01 |
| CHMP6 | 253,12 | -0,13 | 0,07 | -1,79 | 7,32E-02 | 2,68E-01 |
| ZNF286A | 269,86 | 0,10 | 0,05 | 1,79 | 7,32E-02 | 2,68E-01 |
| DPEP3 | 119,38 | -0,21 | 0,12 | -1,79 | 7,33E-02 | 2,68E-01 |
| SEPTIN8 | 174,51 | -0,15 | 0,09 | -1,79 | 7,33E-02 | 2,68E-01 |
| ZBTB14 | 953,03 | 0,11 | 0,06 | 1,79 | 7,33E-02 | 2,68E-01 |
| TRAPPC6B | 1313,57 | 0,12 | 0,07 | 1,79 | 7,34E-02 | 2,68E-01 |
| SRSF5 | 10598,21 | 0,09 | 0,05 | 1,79 | 7,34E-02 | 2,68E-01 |
| CNOT4 | 1174,18 | 0,08 | 0,04 | 1,79 | 7,34E-02 | 2,68E-01 |
| HINT1 | 893,76 | 0,21 | 0,12 | 1,79 | 7,35E-02 | 2,68E-01 |
| PHGDH | 83,49 | -0,33 | 0,19 | -1,79 | 7,35E-02 | 2,68E-01 |
| HAGH | 4995,73 | -0,22 | 0,12 | -1,79 | 7,37E-02 | 2,68E-01 |
| HSPE1 | 200,32 | 0,12 | 0,07 | 1,79 | 7,37E-02 | 2,68E-01 |
| ELP2 | 2450,90 | 0,08 | 0,04 | 1,79 | 7,36E-02 | 2,68E-01 |
| RBM45 | 654,02 | 0,11 | 0,06 | 1,79 | 7,36E-02 | 2,68E-01 |
| PFN2 | 95,04 | -0,19 | 0,11 | -1,79 | 7,36E-02 | 2,68E-01 |
| ATPAF1 | 674,23 | 0,05 | 0,03 | 1,79 | 7,37E-02 | 2,68E-01 |
| GGCX | 642,97 | -0,08 | 0,05 | -1,79 | 7,38E-02 | 2,68E-01 |
| SLC18A1 | 36,29 | -0,37 | 0,20 | -1,79 | 7,38E-02 | 2,69E-01 |
| STRADB | 18948,28 | -0,24 | 0,13 | -1,79 | 7,39E-02 | 2,69E-01 |
| B9D1 | 50,68 | -0,20 | 0,11 | -1,79 | 7,40E-02 | 2,69E-01 |
| CHIC1 | 712,72 | 0,16 | 0,09 | 1,79 | 7,40E-02 | 2,69E-01 |
| ZNF564 | 134,13 | -0,10 | 0,06 | -1,79 | 7,40E-02 | 2,69E-01 |
| SF3B1 | 19728,14 | 0,12 | 0,07 | 1,79 | 7,40E-02 | 2,69E-01 |
| MAP1A | 119,33 | -0,27 | 0,15 | -1,79 | 7,39E-02 | 2,69E-01 |
| CSTF2 | 296,90 | 0,06 | 0,03 | 1,79 | 7,40E-02 | 2,69E-01 |
| RAB5B | 1421,33 | -0,17 | 0,10 | -1,79 | 7,41E-02 | 2,69E-01 |
| HPCAL1 | 828,87 | -0,17 | 0,09 | -1,79 | 7,41E-02 | 2,69E-01 |

| | | | | | | |
|---|---|---|---|---|---|---|
| CLSPN | 82,49 | -0,22 | 0,12 | -1,79 | 7,41E-02 | 2,69E-01 |
| RAPSN | 40,06 | -0,26 | 0,14 | -1,79 | 7,41E-02 | 2,69E-01 |
| DMTF1 | 4588,12 | 0,08 | 0,05 | 1,78 | 7,43E-02 | 2,69E-01 |
| STIL | 455,16 | -0,20 | 0,11 | -1,78 | 7,43E-02 | 2,69E-01 |
| CHCHD4 | 173,38 | 0,08 | 0,04 | 1,78 | 7,43E-02 | 2,69E-01 |
| SLC22A18AS | 28,04 | -0,28 | 0,16 | -1,78 | 7,43E-02 | 2,69E-01 |
| HYAL2 | 162,18 | -0,09 | 0,05 | -1,78 | 7,43E-02 | 2,69E-01 |
| STK26 | 2100,77 | 0,11 | 0,06 | 1,79 | 7,42E-02 | 2,69E-01 |
| IDI1 | 1143,14 | 0,17 | 0,10 | 1,78 | 7,44E-02 | 2,69E-01 |
| MGAT2 | 1659,87 | 0,10 | 0,05 | 1,78 | 7,44E-02 | 2,69E-01 |
| ORMDL1 | 1895,87 | 0,12 | 0,07 | 1,78 | 7,44E-02 | 2,69E-01 |
| RAVER2 | 161,18 | -0,24 | 0,13 | -1,78 | 7,44E-02 | 2,69E-01 |
| DHX29 | 1019,03 | 0,11 | 0,06 | 1,78 | 7,45E-02 | 2,69E-01 |
| ABHD12 | 333,83 | -0,10 | 0,06 | -1,78 | 7,45E-02 | 2,69E-01 |
| SUMF2 | 1575,45 | 0,08 | 0,04 | 1,78 | 7,46E-02 | 2,69E-01 |
| CIZ1 | 352,23 | -0,18 | 0,10 | -1,78 | 7,46E-02 | 2,70E-01 |
| SLC35A3 | 1398,82 | 0,08 | 0,04 | 1,78 | 7,47E-02 | 2,70E-01 |
| NUBP1 | 650,14 | 0,07 | 0,04 | 1,78 | 7,47E-02 | 2,70E-01 |
| SCAF11 | 9527,48 | 0,10 | 0,06 | 1,78 | 7,48E-02 | 2,70E-01 |
| EHBP1 | 712,78 | 0,08 | 0,05 | 1,78 | 7,48E-02 | 2,70E-01 |
| CTU2 | 93,67 | -0,15 | 0,08 | -1,78 | 7,49E-02 | 2,70E-01 |
| SLC25A3 | 3052,71 | 0,05 | 0,03 | 1,78 | 7,49E-02 | 2,70E-01 |
| SLC25A26 | 628,00 | 0,07 | 0,04 | 1,78 | 7,50E-02 | 2,70E-01 |
| PLAA | 716,24 | 0,07 | 0,04 | 1,78 | 7,49E-02 | 2,70E-01 |
| NUTF2 | 651,24 | -0,08 | 0,04 | -1,78 | 7,50E-02 | 2,70E-01 |
| APOM | 31,43 | -0,13 | 0,07 | -1,78 | 7,50E-02 | 2,70E-01 |
| TGDS | 473,73 | 0,13 | 0,07 | 1,78 | 7,51E-02 | 2,70E-01 |
| SART3 | 2169,46 | 0,07 | 0,04 | 1,78 | 7,52E-02 | 2,70E-01 |
| GRIK4 | 84,14 | -0,52 | 0,29 | -1,78 | 7,51E-02 | 2,70E-01 |
| NUP37 | 304,78 | 0,11 | 0,06 | 1,78 | 7,51E-02 | 2,70E-01 |
| GDAP1 | 267,14 | 0,10 | 0,06 | 1,78 | 7,51E-02 | 2,70E-01 |
| FGF2 | 29,15 | -0,38 | 0,21 | -1,78 | 7,51E-02 | 2,70E-01 |
| RO60 | 4402,09 | 0,13 | 0,07 | 1,78 | 7,53E-02 | 2,70E-01 |
| LRP3 | 69,72 | -0,26 | 0,15 | -1,78 | 7,52E-02 | 2,70E-01 |
| KIAA1143 | 1333,19 | 0,10 | 0,06 | 1,78 | 7,53E-02 | 2,70E-01 |
| TPGS2 | 1710,85 | -0,11 | 0,06 | -1,78 | 7,53E-02 | 2,70E-01 |
| CCDC112 | 252,25 | 0,13 | 0,07 | 1,78 | 7,53E-02 | 2,70E-01 |
| MZT1 | 228,03 | 0,18 | 0,10 | 1,78 | 7,53E-02 | 2,70E-01 |
| KCNAB3 | 55,66 | -0,26 | 0,15 | -1,78 | 7,53E-02 | 2,70E-01 |
| UQCC4 | 107,48 | 0,09 | 0,05 | 1,78 | 7,54E-02 | 2,70E-01 |
| VDAC2 | 886,02 | 0,05 | 0,03 | 1,78 | 7,54E-02 | 2,70E-01 |
| TIA1 | 4001,28 | 0,12 | 0,07 | 1,78 | 7,55E-02 | 2,70E-01 |
| CCDC82 | 1894,25 | 0,13 | 0,07 | 1,78 | 7,55E-02 | 2,70E-01 |
| PCM1 | 7027,67 | 0,10 | 0,05 | 1,78 | 7,55E-02 | 2,70E-01 |
| PGM5 | 214,65 | -0,33 | 0,18 | -1,78 | 7,55E-02 | 2,71E-01 |
| FOXO3B | 92,92 | -0,13 | 0,07 | -1,78 | 7,56E-02 | 2,71E-01 |

| | | | | | | |
|---|---|---|---|---|---|---|
| ARMH1 | 113,69 | -0,16 | 0,09 | -1,78 | 7,56E-02 | 2,71E-01 |
| PLA2G15 | 92,11 | -0,19 | 0,11 | -1,78 | 7,56E-02 | 2,71E-01 |
| HARBI1 | 141,23 | -0,07 | 0,04 | -1,78 | 7,57E-02 | 2,71E-01 |
| HOMER2 | 201,11 | -0,16 | 0,09 | -1,78 | 7,57E-02 | 2,71E-01 |
| HDAC10 | 56,77 | -0,20 | 0,11 | -1,78 | 7,58E-02 | 2,71E-01 |
| FNIP2 | 906,03 | 0,08 | 0,04 | 1,78 | 7,58E-02 | 2,71E-01 |
| RRN3 | 1002,73 | 0,09 | 0,05 | 1,78 | 7,58E-02 | 2,71E-01 |
| RPP14 | 661,61 | 0,05 | 0,03 | 1,78 | 7,58E-02 | 2,71E-01 |
| OTUB1 | 585,82 | -0,06 | 0,04 | -1,78 | 7,59E-02 | 2,71E-01 |
| USE1 | 69,97 | -0,14 | 0,08 | -1,78 | 7,59E-02 | 2,71E-01 |
| ATF2 | 2449,85 | 0,12 | 0,07 | 1,77 | 7,60E-02 | 2,71E-01 |
| CAMKK2 | 2743,32 | -0,13 | 0,07 | -1,77 | 7,60E-02 | 2,71E-01 |
| FRMD8 | 825,84 | -0,14 | 0,08 | -1,77 | 7,61E-02 | 2,71E-01 |
| LPCAT3 | 489,15 | -0,10 | 0,06 | -1,77 | 7,61E-02 | 2,71E-01 |
| TBX6 | 27,23 | -0,23 | 0,13 | -1,77 | 7,61E-02 | 2,71E-01 |
| IMP3 | 556,04 | 0,09 | 0,05 | 1,77 | 7,61E-02 | 2,71E-01 |
| FFAR2 | 2699,59 | -0,15 | 0,09 | -1,77 | 7,63E-02 | 2,71E-01 |
| LCORL | 1470,63 | 0,17 | 0,10 | 1,77 | 7,62E-02 | 2,71E-01 |
| ARSB | 849,26 | -0,10 | 0,05 | -1,77 | 7,63E-02 | 2,71E-01 |
| EXO5 | 267,87 | 0,08 | 0,05 | 1,77 | 7,62E-02 | 2,71E-01 |
| ABCG1 | 1011,72 | -0,17 | 0,10 | -1,77 | 7,62E-02 | 2,71E-01 |
| LIN7B | 42,80 | -0,17 | 0,10 | -1,77 | 7,63E-02 | 2,71E-01 |
| PRPF4 | 748,09 | 0,07 | 0,04 | 1,77 | 7,64E-02 | 2,72E-01 |
| RPL19 | 10269,78 | 0,12 | 0,07 | 1,77 | 7,64E-02 | 2,72E-01 |
| SIRPD | 158,25 | -0,16 | 0,09 | -1,77 | 7,64E-02 | 2,72E-01 |
| ZNF780A | 1927,51 | 0,13 | 0,08 | 1,77 | 7,65E-02 | 2,72E-01 |
| YTHDF2 | 1152,23 | 0,04 | 0,02 | 1,77 | 7,65E-02 | 2,72E-01 |
| SLC25A35 | 69,48 | -0,18 | 0,10 | -1,77 | 7,65E-02 | 2,72E-01 |
| RB1CC1 | 4649,97 | 0,14 | 0,08 | 1,77 | 7,66E-02 | 2,72E-01 |
| CCNI | 6438,15 | -0,15 | 0,09 | -1,77 | 7,66E-02 | 2,72E-01 |
| AVEN | 161,11 | 0,09 | 0,05 | 1,77 | 7,67E-02 | 2,72E-01 |
| INTS13 | 577,73 | 0,10 | 0,05 | 1,77 | 7,67E-02 | 2,72E-01 |
| JUP | 107,94 | -0,33 | 0,19 | -1,77 | 7,67E-02 | 2,72E-01 |
| CHST11 | 4756,12 | -0,10 | 0,06 | -1,77 | 7,68E-02 | 2,72E-01 |
| COPB2 | 3102,36 | 0,08 | 0,05 | 1,77 | 7,69E-02 | 2,72E-01 |
| ABCD2 | 1286,67 | 0,18 | 0,10 | 1,77 | 7,69E-02 | 2,72E-01 |
| AP3B1 | 3935,69 | 0,10 | 0,06 | 1,77 | 7,68E-02 | 2,72E-01 |
| FDX1 | 481,96 | 0,10 | 0,05 | 1,77 | 7,69E-02 | 2,72E-01 |
| SELENON | 115,71 | -0,26 | 0,15 | -1,77 | 7,69E-02 | 2,72E-01 |
| ZXDA | 285,56 | 0,10 | 0,06 | 1,77 | 7,68E-02 | 2,72E-01 |
| SLC4A7 | 3186,53 | 0,12 | 0,07 | 1,77 | 7,69E-02 | 2,72E-01 |
| ECI2 | 537,64 | 0,07 | 0,04 | 1,77 | 7,69E-02 | 2,72E-01 |
| SUGCT | 66,72 | -0,21 | 0,12 | -1,77 | 7,69E-02 | 2,72E-01 |
| G3BP2 | 4021,46 | 0,09 | 0,05 | 1,77 | 7,70E-02 | 2,72E-01 |
| RAB24 | 720,57 | -0,12 | 0,07 | -1,77 | 7,70E-02 | 2,72E-01 |
| ZNF408 | 249,91 | -0,07 | 0,04 | -1,77 | 7,70E-02 | 2,72E-01 |

| | | | | | | |
|---|---|---|---|---|---|---|
| NIPBL | 7684,02 | 0,09 | 0,05 | 1,77 | 7,70E-02 | 2,72E-01 |
| TRIM62 | 166,18 | -0,22 | 0,12 | -1,77 | 7,71E-02 | 2,72E-01 |
| ATN1 | 77,53 | -0,30 | 0,17 | -1,77 | 7,72E-02 | 2,73E-01 |
| STARD13 | 54,47 | -0,36 | 0,20 | -1,77 | 7,72E-02 | 2,73E-01 |
| SMARCD3 | 385,84 | -0,13 | 0,07 | -1,77 | 7,73E-02 | 2,73E-01 |
| ZDHHC11 | 62,28 | -0,36 | 0,20 | -1,77 | 7,73E-02 | 2,73E-01 |
| ENSG00000(| 64,72 | -0,20 | 0,11 | -1,77 | 7,74E-02 | 2,73E-01 |
| ZNF587B | 960,21 | 0,06 | 0,04 | 1,77 | 7,74E-02 | 2,73E-01 |
| XPO7 | 3206,66 | -0,12 | 0,07 | -1,77 | 7,74E-02 | 2,73E-01 |
| ZNF782 | 796,07 | 0,08 | 0,05 | 1,77 | 7,74E-02 | 2,73E-01 |
| SCAMP4 | 111,49 | -0,27 | 0,15 | -1,77 | 7,74E-02 | 2,73E-01 |
| DOP1A | 2541,81 | 0,10 | 0,05 | 1,76 | 7,77E-02 | 2,73E-01 |
| ZBTB44 | 4043,25 | 0,09 | 0,05 | 1,76 | 7,77E-02 | 2,73E-01 |
| ASB2 | 130,12 | -0,20 | 0,11 | -1,77 | 7,76E-02 | 2,73E-01 |
| RAC2 | 2373,54 | -0,16 | 0,09 | -1,76 | 7,77E-02 | 2,73E-01 |
| ARAP2 | 5550,41 | 0,14 | 0,08 | 1,76 | 7,77E-02 | 2,73E-01 |
| TUBA4A | 2175,03 | -0,10 | 0,06 | -1,76 | 7,77E-02 | 2,73E-01 |
| ZNF33A | 4855,51 | 0,09 | 0,05 | 1,77 | 7,76E-02 | 2,73E-01 |
| RFX5 | 1640,48 | 0,06 | 0,03 | 1,77 | 7,75E-02 | 2,73E-01 |
| ZNF626 | 371,24 | 0,14 | 0,08 | 1,76 | 7,76E-02 | 2,73E-01 |
| LRRC56 | 51,29 | -0,24 | 0,13 | -1,76 | 7,76E-02 | 2,73E-01 |
| STAMBPL1 | 1128,51 | 0,11 | 0,06 | 1,77 | 7,75E-02 | 2,73E-01 |
| SDC4 | 76,36 | -0,20 | 0,11 | -1,76 | 7,77E-02 | 2,73E-01 |
| E2F4 | 888,43 | -0,12 | 0,07 | -1,76 | 7,77E-02 | 2,73E-01 |
| CCT7 | 2014,39 | 0,07 | 0,04 | 1,76 | 7,81E-02 | 2,73E-01 |
| MRGBP | 361,48 | 0,07 | 0,04 | 1,76 | 7,79E-02 | 2,73E-01 |
| PTP4A2 | 6401,72 | 0,08 | 0,05 | 1,76 | 7,80E-02 | 2,73E-01 |
| ZNF229 | 52,32 | -0,23 | 0,13 | -1,76 | 7,81E-02 | 2,73E-01 |
| PSMA2 | 379,40 | 0,07 | 0,04 | 1,76 | 7,78E-02 | 2,73E-01 |
| ITGA9 | 106,63 | -0,30 | 0,17 | -1,76 | 7,80E-02 | 2,73E-01 |
| IFRD2 | 412,69 | -0,12 | 0,07 | -1,76 | 7,80E-02 | 2,73E-01 |
| PCDHGA8 | 35,46 | -0,26 | 0,15 | -1,76 | 7,79E-02 | 2,73E-01 |
| UTP6 | 1740,44 | 0,07 | 0,04 | 1,76 | 7,80E-02 | 2,73E-01 |
| C6orf120 | 917,98 | 0,12 | 0,07 | 1,76 | 7,80E-02 | 2,73E-01 |
| THAP3 | 271,15 | -0,07 | 0,04 | -1,76 | 7,78E-02 | 2,73E-01 |
| LSM4 | 669,78 | 0,07 | 0,04 | 1,76 | 7,81E-02 | 2,73E-01 |
| MOB3A | 2472,72 | -0,19 | 0,11 | -1,76 | 7,81E-02 | 2,73E-01 |
| TIGD2 | 153,84 | 0,17 | 0,10 | 1,76 | 7,78E-02 | 2,73E-01 |
| FMO5 | 327,92 | 0,09 | 0,05 | 1,76 | 7,80E-02 | 2,73E-01 |
| AGAP5 | 57,32 | -0,12 | 0,07 | -1,76 | 7,81E-02 | 2,73E-01 |
| LLPH | 830,51 | 0,07 | 0,04 | 1,76 | 7,81E-02 | 2,73E-01 |
| ZNF613 | 264,79 | 0,15 | 0,08 | 1,76 | 7,82E-02 | 2,73E-01 |
| AKR1C1 | 67,29 | -0,26 | 0,15 | -1,76 | 7,82E-02 | 2,73E-01 |
| ABCA2 | 806,97 | -0,22 | 0,13 | -1,76 | 7,83E-02 | 2,73E-01 |
| PPP2R5D | 199,16 | -0,15 | 0,08 | -1,76 | 7,83E-02 | 2,73E-01 |
| UNC119 | 1478,58 | -0,10 | 0,06 | -1,76 | 7,83E-02 | 2,73E-01 |

| | | | | | | |
|---|---|---|---|---|---|---|
| UPP1 | 1144,05 | -0,09 | 0,05 | -1,76 | 7,83E-02 | 2,73E-01 |
| RNF207 | 81,07 | -0,28 | 0,16 | -1,76 | 7,84E-02 | 2,73E-01 |
| TBL1XR1 | 8324,17 | 0,11 | 0,07 | 1,76 | 7,84E-02 | 2,73E-01 |
| IQCE | 713,77 | -0,13 | 0,07 | -1,76 | 7,84E-02 | 2,73E-01 |
| TMEM8B | 154,99 | -0,23 | 0,13 | -1,76 | 7,85E-02 | 2,73E-01 |
| TMEM143 | 105,57 | -0,11 | 0,06 | -1,76 | 7,85E-02 | 2,73E-01 |
| AKR1C8 | 32,71 | -0,28 | 0,16 | -1,76 | 7,86E-02 | 2,73E-01 |
| NSMCE3 | 471,16 | 0,08 | 0,05 | 1,76 | 7,85E-02 | 2,73E-01 |
| SMC4 | 1289,67 | 0,12 | 0,07 | 1,76 | 7,85E-02 | 2,73E-01 |
| PDE7A | 6753,32 | 0,09 | 0,05 | 1,76 | 7,86E-02 | 2,73E-01 |
| DHCR24 | 123,85 | -0,21 | 0,12 | -1,76 | 7,86E-02 | 2,73E-01 |
| MPHOSPH9 | 1336,06 | 0,08 | 0,04 | 1,76 | 7,86E-02 | 2,73E-01 |
| CEACAM6 | 34,44 | -0,36 | 0,21 | -1,76 | 7,87E-02 | 2,74E-01 |
| KIF5B | 4278,37 | 0,12 | 0,07 | 1,76 | 7,89E-02 | 2,74E-01 |
| ZNF518B | 1694,67 | 0,10 | 0,06 | 1,76 | 7,88E-02 | 2,74E-01 |
| RELCH | 5505,25 | 0,09 | 0,05 | 1,76 | 7,88E-02 | 2,74E-01 |
| PSMB1 | 1787,12 | 0,10 | 0,06 | 1,76 | 7,88E-02 | 2,74E-01 |
| SUOX | 143,72 | -0,14 | 0,08 | -1,76 | 7,92E-02 | 2,74E-01 |
| GGT1 | 236,18 | -0,14 | 0,08 | -1,76 | 7,90E-02 | 2,74E-01 |
| NHSL2 | 4295,46 | -0,15 | 0,09 | -1,76 | 7,92E-02 | 2,74E-01 |
| RANBP10 | 1254,79 | -0,21 | 0,12 | -1,76 | 7,92E-02 | 2,74E-01 |
| PJA1 | 347,96 | 0,10 | 0,05 | 1,76 | 7,90E-02 | 2,74E-01 |
| TM2D2 | 502,86 | 0,08 | 0,05 | 1,76 | 7,90E-02 | 2,74E-01 |
| CYBB | 19440,93 | 0,13 | 0,07 | 1,76 | 7,89E-02 | 2,74E-01 |
| NMT2 | 673,60 | 0,10 | 0,06 | 1,76 | 7,91E-02 | 2,74E-01 |
| WAPL | 4682,28 | 0,09 | 0,05 | 1,76 | 7,90E-02 | 2,74E-01 |
| ZNF706 | 1612,39 | 0,09 | 0,05 | 1,76 | 7,89E-02 | 2,74E-01 |
| ADAMTS17 | 183,79 | -0,21 | 0,12 | -1,76 | 7,91E-02 | 2,74E-01 |
| EHD1 | 2401,02 | -0,13 | 0,08 | -1,76 | 7,90E-02 | 2,74E-01 |
| TCF12 | 1966,72 | 0,07 | 0,04 | 1,76 | 7,92E-02 | 2,74E-01 |
| CEP162 | 583,63 | 0,13 | 0,07 | 1,76 | 7,91E-02 | 2,74E-01 |
| ANKRD26 | 606,79 | 0,12 | 0,07 | 1,76 | 7,92E-02 | 2,74E-01 |
| C4orf50 | 178,24 | -0,32 | 0,18 | -1,75 | 7,93E-02 | 2,74E-01 |
| PPBP | 4213,90 | -0,25 | 0,15 | -1,75 | 7,93E-02 | 2,74E-01 |
| COPB1 | 2637,60 | 0,10 | 0,06 | 1,75 | 7,93E-02 | 2,74E-01 |
| SNRPA | 196,34 | -0,13 | 0,07 | -1,75 | 7,93E-02 | 2,74E-01 |
| WBP1L | 1921,32 | -0,07 | 0,04 | -1,75 | 7,94E-02 | 2,74E-01 |
| FBRSL1 | 336,65 | -0,29 | 0,16 | -1,75 | 7,95E-02 | 2,74E-01 |
| TGM3 | 82,39 | -0,22 | 0,13 | -1,75 | 7,96E-02 | 2,75E-01 |
| ZNF664 | 1152,40 | 0,07 | 0,04 | 1,75 | 7,97E-02 | 2,75E-01 |
| B3GAT1 | 241,27 | -0,30 | 0,17 | -1,75 | 7,98E-02 | 2,75E-01 |
| IRF6 | 37,37 | -0,34 | 0,19 | -1,75 | 7,98E-02 | 2,75E-01 |
| C17orf75 | 349,88 | 0,07 | 0,04 | 1,75 | 7,99E-02 | 2,75E-01 |
| ATF3 | 41,97 | -0,28 | 0,16 | -1,75 | 8,00E-02 | 2,75E-01 |
| OTUD4 | 2112,57 | 0,07 | 0,04 | 1,75 | 7,99E-02 | 2,75E-01 |
| ANGEL2 | 2749,54 | 0,10 | 0,06 | 1,75 | 8,00E-02 | 2,75E-01 |

| | | | | | | |
|---|---|---|---|---|---|---|
| OLA1 | 488,73 | 0,10 | 0,06 | 1,75 | 8,00E-02 | 2,75E-01 |
| GRK4 | 72,66 | -0,19 | 0,11 | -1,75 | 8,01E-02 | 2,75E-01 |
| IKZF5 | 976,21 | 0,11 | 0,06 | 1,75 | 8,01E-02 | 2,75E-01 |
| GOLT1B | 1023,74 | 0,16 | 0,09 | 1,75 | 8,01E-02 | 2,76E-01 |
| NTN4 | 48,80 | -0,31 | 0,18 | -1,75 | 8,02E-02 | 2,76E-01 |
| OLIG1 | 150,96 | -0,19 | 0,11 | -1,75 | 8,03E-02 | 2,76E-01 |
| PNN | 2437,11 | 0,10 | 0,05 | 1,75 | 8,03E-02 | 2,76E-01 |
| TRNT1 | 1117,34 | 0,10 | 0,06 | 1,75 | 8,04E-02 | 2,76E-01 |
| ORAI3 | 639,03 | -0,09 | 0,05 | -1,75 | 8,05E-02 | 2,76E-01 |
| TOMM22 | 1064,44 | 0,10 | 0,06 | 1,75 | 8,05E-02 | 2,76E-01 |
| P2RX5-TAX1E | 36,60 | -0,24 | 0,14 | -1,75 | 8,05E-02 | 2,76E-01 |
| RFX7 | 1161,81 | 0,11 | 0,06 | 1,75 | 8,05E-02 | 2,76E-01 |
| LILRA6 | 706,12 | -0,18 | 0,11 | -1,75 | 8,06E-02 | 2,76E-01 |
| RWDD1 | 353,88 | 0,13 | 0,08 | 1,75 | 8,06E-02 | 2,77E-01 |
| ENSG000002 | 125,28 | -0,20 | 0,11 | -1,75 | 8,07E-02 | 2,77E-01 |
| MRPL22 | 343,36 | 0,09 | 0,05 | 1,75 | 8,09E-02 | 2,77E-01 |
| LNPEP | 11753,81 | 0,09 | 0,05 | 1,75 | 8,08E-02 | 2,77E-01 |
| GPBP1 | 4295,49 | 0,14 | 0,08 | 1,75 | 8,08E-02 | 2,77E-01 |
| GTF2A2 | 731,79 | 0,09 | 0,05 | 1,75 | 8,08E-02 | 2,77E-01 |
| RSRC1 | 961,05 | 0,10 | 0,06 | 1,75 | 8,08E-02 | 2,77E-01 |
| UQCRC2 | 2532,17 | 0,09 | 0,05 | 1,75 | 8,08E-02 | 2,77E-01 |
| ZDHHC3 | 1600,79 | -0,07 | 0,04 | -1,75 | 8,09E-02 | 2,77E-01 |
| LTBP1 | 522,14 | -0,22 | 0,13 | -1,75 | 8,07E-02 | 2,77E-01 |
| ZNF684 | 138,18 | 0,10 | 0,06 | 1,75 | 8,07E-02 | 2,77E-01 |
| MPIG6B | 64,77 | -0,38 | 0,22 | -1,75 | 8,09E-02 | 2,77E-01 |
| SPAG16 | 151,94 | 0,13 | 0,07 | 1,75 | 8,10E-02 | 2,77E-01 |
| ZNF543 | 375,56 | 0,08 | 0,04 | 1,74 | 8,11E-02 | 2,77E-01 |
| ZNF117 | 3012,52 | 0,20 | 0,12 | 1,74 | 8,12E-02 | 2,77E-01 |
| PTAR1 | 6288,41 | 0,14 | 0,08 | 1,74 | 8,11E-02 | 2,77E-01 |
| NEK1 | 757,89 | 0,10 | 0,06 | 1,74 | 8,11E-02 | 2,77E-01 |
| P4HA1 | 952,42 | 0,11 | 0,06 | 1,74 | 8,11E-02 | 2,77E-01 |
| CNN2 | 4273,07 | -0,17 | 0,10 | -1,74 | 8,11E-02 | 2,77E-01 |
| MYLK4 | 93,74 | -0,17 | 0,10 | -1,74 | 8,17E-02 | 2,77E-01 |
| PACRGL | 424,26 | 0,09 | 0,05 | 1,74 | 8,14E-02 | 2,77E-01 |
| RPS6KB1 | 1470,97 | 0,06 | 0,03 | 1,74 | 8,15E-02 | 2,77E-01 |
| N6AMT1 | 495,05 | 0,10 | 0,06 | 1,74 | 8,14E-02 | 2,77E-01 |
| SNX6 | 2266,34 | 0,12 | 0,07 | 1,74 | 8,13E-02 | 2,77E-01 |
| RAB11B | 467,42 | -0,20 | 0,11 | -1,74 | 8,14E-02 | 2,77E-01 |
| CASC3 | 2355,10 | -0,13 | 0,08 | -1,74 | 8,14E-02 | 2,77E-01 |
| LMBRD2 | 1612,93 | 0,13 | 0,08 | 1,74 | 8,16E-02 | 2,77E-01 |
| WWP1 | 2186,71 | 0,10 | 0,06 | 1,74 | 8,16E-02 | 2,77E-01 |
| MDH1 | 1113,83 | 0,10 | 0,06 | 1,74 | 8,14E-02 | 2,77E-01 |
| NOLC1 | 562,88 | 0,07 | 0,04 | 1,74 | 8,13E-02 | 2,77E-01 |
| HECTD3 | 624,87 | -0,11 | 0,07 | -1,74 | 8,14E-02 | 2,77E-01 |
| RAB12 | 665,71 | 0,08 | 0,05 | 1,74 | 8,16E-02 | 2,77E-01 |
| RHBDL1 | 25,96 | -0,28 | 0,16 | -1,74 | 8,12E-02 | 2,77E-01 |

| | | | | | | |
|---|---|---|---|---|---|---|
| STK35 | 813,24 | -0,11 | 0,07 | -1,74 | 8,15E-02 | 2,77E-01 |
| GNAI1 | 93,35 | -0,30 | 0,17 | -1,74 | 8,15E-02 | 2,77E-01 |
| IGSF9B | 145,10 | -0,26 | 0,15 | -1,74 | 8,12E-02 | 2,77E-01 |
| FSTL3 | 68,15 | -0,19 | 0,11 | -1,74 | 8,14E-02 | 2,77E-01 |
| PTGIR | 180,13 | -0,15 | 0,09 | -1,74 | 8,17E-02 | 2,77E-01 |
| RPUSD1 | 150,78 | -0,14 | 0,08 | -1,74 | 8,15E-02 | 2,77E-01 |
| TMEM203 | 559,01 | 0,07 | 0,04 | 1,74 | 8,13E-02 | 2,77E-01 |
| ITGAV | 913,56 | 0,11 | 0,06 | 1,74 | 8,16E-02 | 2,77E-01 |
| AP5Z1 | 738,46 | -0,13 | 0,08 | -1,74 | 8,16E-02 | 2,77E-01 |
| ATP5MC3 | 1147,67 | 0,10 | 0,06 | 1,74 | 8,17E-02 | 2,77E-01 |
| BZW2 | 537,60 | 0,07 | 0,04 | 1,74 | 8,18E-02 | 2,77E-01 |
| WDSUB1 | 252,57 | 0,08 | 0,05 | 1,74 | 8,18E-02 | 2,77E-01 |
| RTN4IP1 | 221,06 | 0,08 | 0,05 | 1,74 | 8,19E-02 | 2,77E-01 |
| AGO2 | 4608,56 | -0,13 | 0,08 | -1,74 | 8,20E-02 | 2,77E-01 |
| FBXL16 | 48,70 | -0,27 | 0,16 | -1,74 | 8,19E-02 | 2,77E-01 |
| RAB4B | 151,12 | -0,13 | 0,08 | -1,74 | 8,20E-02 | 2,77E-01 |
| ASPSCR1 | 315,16 | -0,22 | 0,13 | -1,74 | 8,22E-02 | 2,77E-01 |
| DESI1 | 674,43 | -0,10 | 0,06 | -1,74 | 8,20E-02 | 2,77E-01 |
| MRPL57 | 418,02 | 0,10 | 0,06 | 1,74 | 8,21E-02 | 2,77E-01 |
| CCNT2 | 4181,64 | 0,13 | 0,07 | 1,74 | 8,21E-02 | 2,77E-01 |
| PRPSAP1 | 380,97 | -0,07 | 0,04 | -1,74 | 8,21E-02 | 2,77E-01 |
| EXD3 | 245,10 | -0,17 | 0,10 | -1,74 | 8,21E-02 | 2,77E-01 |
| JRKL | 312,60 | 0,16 | 0,09 | 1,74 | 8,21E-02 | 2,77E-01 |
| NME2 | 43,62 | -0,23 | 0,13 | -1,74 | 8,23E-02 | 2,78E-01 |
| ASPHD2 | 125,90 | -0,13 | 0,08 | -1,74 | 8,22E-02 | 2,78E-01 |
| RPL3L | 30,19 | -0,47 | 0,27 | -1,74 | 8,23E-02 | 2,78E-01 |
| SCCPDH | 315,12 | 0,12 | 0,07 | 1,74 | 8,24E-02 | 2,78E-01 |
| SLC25A30 | 621,85 | 0,05 | 0,03 | 1,74 | 8,24E-02 | 2,78E-01 |
| FBXO8 | 691,66 | 0,13 | 0,07 | 1,74 | 8,25E-02 | 2,78E-01 |
| CCDC18 | 576,79 | 0,15 | 0,08 | 1,74 | 8,25E-02 | 2,78E-01 |
| ELF1 | 6682,03 | 0,09 | 0,05 | 1,74 | 8,26E-02 | 2,78E-01 |
| INO80B | 43,64 | 0,12 | 0,07 | 1,74 | 8,26E-02 | 2,78E-01 |
| CALM3 | 1213,22 | -0,13 | 0,08 | -1,74 | 8,26E-02 | 2,78E-01 |
| EMC3 | 2952,72 | -0,15 | 0,09 | -1,74 | 8,26E-02 | 2,78E-01 |
| TIMM8A | 118,71 | 0,08 | 0,05 | 1,74 | 8,26E-02 | 2,78E-01 |
| SLC1A3 | 49,29 | -0,29 | 0,17 | -1,73 | 8,29E-02 | 2,78E-01 |
| SPNS3 | 201,00 | -0,17 | 0,10 | -1,73 | 8,28E-02 | 2,78E-01 |
| LOXHD1 | 388,49 | -0,25 | 0,15 | -1,73 | 8,29E-02 | 2,78E-01 |
| MLH3 | 1222,27 | 0,10 | 0,06 | 1,73 | 8,28E-02 | 2,78E-01 |
| CCT5 | 2796,57 | 0,08 | 0,05 | 1,74 | 8,27E-02 | 2,78E-01 |
| TAF7 | 1183,45 | 0,18 | 0,10 | 1,73 | 8,29E-02 | 2,78E-01 |
| PDS5B | 3072,79 | 0,11 | 0,06 | 1,73 | 8,29E-02 | 2,78E-01 |
| ABITRAM | 194,78 | 0,07 | 0,04 | 1,73 | 8,29E-02 | 2,78E-01 |
| CACYBP | 860,43 | 0,11 | 0,06 | 1,74 | 8,27E-02 | 2,78E-01 |
| NAALADL2 | 32,57 | -0,33 | 0,19 | -1,73 | 8,28E-02 | 2,78E-01 |
| POLR2E | 510,67 | -0,11 | 0,06 | -1,73 | 8,28E-02 | 2,78E-01 |

| | | | | | | |
|---|---|---|---|---|---|---|
| ZNF800 | 2240,72 | 0,11 | 0,07 | 1,73 | 8,29E-02 | 2,78E-01 |
| MMS22L | 965,88 | 0,09 | 0,05 | 1,73 | 8,30E-02 | 2,79E-01 |
| SIRT2 | 335,09 | -0,15 | 0,08 | -1,73 | 8,31E-02 | 2,79E-01 |
| CDC37L1 | 598,91 | 0,14 | 0,08 | 1,73 | 8,31E-02 | 2,79E-01 |
| ERCC6 | 2167,35 | 0,07 | 0,04 | 1,73 | 8,32E-02 | 2,79E-01 |
| GNLY | 2295,96 | -0,22 | 0,13 | -1,73 | 8,32E-02 | 2,79E-01 |
| ENSG000002 | 214,74 | 0,08 | 0,05 | 1,73 | 8,32E-02 | 2,79E-01 |
| PPP1R16A | 104,26 | -0,20 | 0,12 | -1,73 | 8,33E-02 | 2,79E-01 |
| PSD3 | 148,15 | -0,23 | 0,13 | -1,73 | 8,33E-02 | 2,79E-01 |
| PRELID3B | 1251,22 | 0,13 | 0,07 | 1,73 | 8,33E-02 | 2,79E-01 |
| MTHFD2L | 488,66 | 0,13 | 0,07 | 1,73 | 8,33E-02 | 2,79E-01 |
| CLDN15 | 159,04 | -0,19 | 0,11 | -1,73 | 8,33E-02 | 2,79E-01 |
| TTC17 | 4896,93 | 0,07 | 0,04 | 1,73 | 8,34E-02 | 2,79E-01 |
| ZNF691 | 279,00 | 0,06 | 0,03 | 1,73 | 8,36E-02 | 2,79E-01 |
| DIMT1 | 498,03 | 0,10 | 0,06 | 1,73 | 8,35E-02 | 2,79E-01 |
| DCTN6 | 183,64 | 0,11 | 0,06 | 1,73 | 8,35E-02 | 2,79E-01 |
| PAPSS2 | 137,61 | -0,15 | 0,09 | -1,73 | 8,34E-02 | 2,79E-01 |
| RTCA | 1028,60 | 0,06 | 0,03 | 1,73 | 8,35E-02 | 2,79E-01 |
| ACTR2 | 16186,84 | 0,10 | 0,06 | 1,73 | 8,36E-02 | 2,79E-01 |
| CIAO2A | 676,44 | 0,14 | 0,08 | 1,73 | 8,36E-02 | 2,79E-01 |
| IL12RB1 | 592,22 | -0,11 | 0,07 | -1,73 | 8,35E-02 | 2,79E-01 |
| H2BC15 | 341,34 | -0,10 | 0,06 | -1,73 | 8,36E-02 | 2,79E-01 |
| ZNF777 | 85,79 | -0,21 | 0,12 | -1,73 | 8,37E-02 | 2,79E-01 |
| PSMD6 | 1834,84 | 0,07 | 0,04 | 1,73 | 8,37E-02 | 2,79E-01 |
| FIS1 | 2342,06 | -0,16 | 0,09 | -1,73 | 8,37E-02 | 2,79E-01 |
| BPNT1 | 433,27 | 0,09 | 0,05 | 1,73 | 8,37E-02 | 2,79E-01 |
| BAG6 | 2254,69 | -0,20 | 0,12 | -1,73 | 8,38E-02 | 2,79E-01 |
| B4GALT1 | 1784,62 | -0,11 | 0,06 | -1,73 | 8,38E-02 | 2,79E-01 |
| EIF2S3 | 5274,02 | 0,08 | 0,05 | 1,73 | 8,38E-02 | 2,79E-01 |
| AMFR | 2686,53 | -0,12 | 0,07 | -1,73 | 8,38E-02 | 2,79E-01 |
| ZNF425 | 82,30 | -0,11 | 0,07 | -1,73 | 8,39E-02 | 2,79E-01 |
| MYO1E | 433,13 | 0,15 | 0,09 | 1,73 | 8,40E-02 | 2,79E-01 |
| CNNM4 | 138,20 | -0,18 | 0,10 | -1,73 | 8,39E-02 | 2,79E-01 |
| SYNPO2 | 79,28 | -0,26 | 0,15 | -1,73 | 8,40E-02 | 2,79E-01 |
| PPIP5K2 | 4519,89 | 0,13 | 0,08 | 1,73 | 8,39E-02 | 2,79E-01 |
| TEFM | 413,53 | 0,09 | 0,05 | 1,73 | 8,41E-02 | 2,79E-01 |
| KBTBD3 | 224,91 | 0,13 | 0,08 | 1,73 | 8,41E-02 | 2,79E-01 |
| ALCAM | 951,74 | 0,13 | 0,08 | 1,73 | 8,40E-02 | 2,79E-01 |
| TAS2R31 | 281,17 | 0,13 | 0,08 | 1,73 | 8,41E-02 | 2,79E-01 |
| MS4A6A | 4792,96 | 0,12 | 0,07 | 1,73 | 8,42E-02 | 2,79E-01 |
| HDAC2 | 885,69 | 0,11 | 0,06 | 1,73 | 8,42E-02 | 2,79E-01 |
| ACSS2 | 569,84 | -0,11 | 0,06 | -1,73 | 8,42E-02 | 2,79E-01 |
| FAM53C | 1959,67 | -0,14 | 0,08 | -1,73 | 8,43E-02 | 2,79E-01 |
| NINJ2 | 596,81 | -0,17 | 0,10 | -1,73 | 8,42E-02 | 2,79E-01 |
| SHCBP1 | 60,67 | -0,22 | 0,13 | -1,73 | 8,43E-02 | 2,79E-01 |
| ZNF32 | 262,75 | 0,15 | 0,09 | 1,73 | 8,44E-02 | 2,80E-01 |

| | | | | | | |
|---|---|---|---|---|---|---|
| CALCRL | 109,47 | 0,18 | 0,10 | 1,73 | 8,44E-02 | 2,80E-01 |
| TFB1M | 589,03 | 0,08 | 0,05 | 1,73 | 8,45E-02 | 2,80E-01 |
| PEX11B | 404,14 | 0,06 | 0,03 | 1,73 | 8,44E-02 | 2,80E-01 |
| MRAS | 96,46 | -0,23 | 0,14 | -1,72 | 8,45E-02 | 2,80E-01 |
| ENG | 50,34 | -0,31 | 0,18 | -1,72 | 8,46E-02 | 2,80E-01 |
| SLC12A4 | 483,17 | -0,13 | 0,08 | -1,72 | 8,46E-02 | 2,80E-01 |
| DBN1 | 123,30 | -0,22 | 0,13 | -1,72 | 8,46E-02 | 2,80E-01 |
| PURB | 3768,72 | 0,10 | 0,06 | 1,72 | 8,47E-02 | 2,80E-01 |
| CBLB | 2778,65 | 0,11 | 0,06 | 1,72 | 8,47E-02 | 2,80E-01 |
| ERICH1 | 3493,02 | 0,11 | 0,07 | 1,72 | 8,47E-02 | 2,80E-01 |
| LIMK1 | 256,05 | -0,19 | 0,11 | -1,72 | 8,48E-02 | 2,80E-01 |
| TTK | 48,69 | -0,30 | 0,17 | -1,72 | 8,49E-02 | 2,80E-01 |
| NAP1L3 | 55,49 | 0,18 | 0,10 | 1,72 | 8,49E-02 | 2,80E-01 |
| TBC1D32 | 612,99 | 0,13 | 0,07 | 1,72 | 8,49E-02 | 2,80E-01 |
| TSPAN18 | 199,10 | -0,19 | 0,11 | -1,72 | 8,50E-02 | 2,80E-01 |
| RAB19 | 21,08 | -0,16 | 0,10 | -1,72 | 8,50E-02 | 2,80E-01 |
| UNC13D | 2084,98 | -0,19 | 0,11 | -1,72 | 8,50E-02 | 2,80E-01 |
| CHP1 | 1225,43 | -0,13 | 0,08 | -1,72 | 8,50E-02 | 2,80E-01 |
| ASAH2 | 38,37 | -0,33 | 0,19 | -1,72 | 8,50E-02 | 2,80E-01 |
| CCT8 | 2044,69 | 0,11 | 0,06 | 1,72 | 8,51E-02 | 2,81E-01 |
| PALB2 | 312,94 | 0,07 | 0,04 | 1,72 | 8,52E-02 | 2,81E-01 |
| L3HYPDH | 546,16 | 0,09 | 0,05 | 1,72 | 8,51E-02 | 2,81E-01 |
| NNT | 2036,69 | 0,06 | 0,04 | 1,72 | 8,52E-02 | 2,81E-01 |
| ATP2C1 | 2164,40 | 0,08 | 0,05 | 1,72 | 8,51E-02 | 2,81E-01 |
| KLHL13 | 35,22 | -0,22 | 0,13 | -1,72 | 8,52E-02 | 2,81E-01 |
| LDAH | 525,56 | 0,09 | 0,05 | 1,72 | 8,52E-02 | 2,81E-01 |
| C11orf58 | 4409,26 | 0,12 | 0,07 | 1,72 | 8,53E-02 | 2,81E-01 |
| GLOD4 | 1117,90 | 0,07 | 0,04 | 1,72 | 8,53E-02 | 2,81E-01 |
| INTS2 | 931,79 | 0,07 | 0,04 | 1,72 | 8,54E-02 | 2,81E-01 |
| TC2N | 5713,96 | 0,14 | 0,08 | 1,72 | 8,53E-02 | 2,81E-01 |
| DGLUCY | 4826,88 | -0,12 | 0,07 | -1,72 | 8,53E-02 | 2,81E-01 |
| CD163L1 | 54,34 | -0,37 | 0,22 | -1,72 | 8,54E-02 | 2,81E-01 |
| PINK1 | 3773,75 | -0,16 | 0,10 | -1,72 | 8,54E-02 | 2,81E-01 |
| RELB | 340,59 | -0,14 | 0,08 | -1,72 | 8,55E-02 | 2,81E-01 |
| ZNF846 | 336,06 | 0,08 | 0,05 | 1,72 | 8,54E-02 | 2,81E-01 |
| CANT1 | 1325,08 | -0,13 | 0,08 | -1,72 | 8,56E-02 | 2,81E-01 |
| ZFYVE1 | 680,50 | -0,08 | 0,05 | -1,72 | 8,57E-02 | 2,81E-01 |
| WHRN | 74,99 | -0,29 | 0,17 | -1,72 | 8,56E-02 | 2,81E-01 |
| CHEK1 | 86,27 | -0,21 | 0,12 | -1,72 | 8,57E-02 | 2,81E-01 |
| RNASEH1 | 359,32 | 0,04 | 0,02 | 1,72 | 8,57E-02 | 2,81E-01 |
| METTL25 | 582,15 | 0,11 | 0,07 | 1,72 | 8,58E-02 | 2,81E-01 |
| SLC25A12 | 757,91 | 0,07 | 0,04 | 1,72 | 8,58E-02 | 2,81E-01 |
| CRAT | 135,85 | -0,24 | 0,14 | -1,72 | 8,59E-02 | 2,81E-01 |
| TMOD1 | 1009,59 | -0,23 | 0,13 | -1,72 | 8,59E-02 | 2,82E-01 |
| GPR137 | 189,24 | -0,11 | 0,06 | -1,72 | 8,60E-02 | 2,82E-01 |
| SLC35B4 | 618,48 | 0,05 | 0,03 | 1,72 | 8,60E-02 | 2,82E-01 |

| | | | | | | |
|---|---|---|---|---|---|---|
| ZNF41 | 524,45 | 0,05 | 0,03 | 1,72 | 8,61E-02 | 2,82E-01 |
| PF4 | 2295,45 | -0,23 | 0,14 | -1,72 | 8,62E-02 | 2,82E-01 |
| TAF6 | 183,72 | -0,17 | 0,10 | -1,72 | 8,63E-02 | 2,82E-01 |
| CHTF18 | 188,84 | -0,17 | 0,10 | -1,72 | 8,63E-02 | 2,82E-01 |
| ETV6 | 1216,37 | -0,14 | 0,08 | -1,71 | 8,64E-02 | 2,83E-01 |
| BCL2L13 | 1376,86 | -0,09 | 0,05 | -1,71 | 8,65E-02 | 2,83E-01 |
| LAIR2 | 68,48 | 0,21 | 0,12 | 1,71 | 8,65E-02 | 2,83E-01 |
| NR6A1 | 252,29 | -0,16 | 0,09 | -1,71 | 8,66E-02 | 2,83E-01 |
| CNPPD1 | 887,71 | -0,18 | 0,11 | -1,71 | 8,66E-02 | 2,83E-01 |
| PNPLA1 | 111,51 | -0,16 | 0,09 | -1,71 | 8,66E-02 | 2,83E-01 |
| CHRNA10 | 109,89 | -0,14 | 0,08 | -1,71 | 8,67E-02 | 2,83E-01 |
| CYLD | 12011,68 | 0,09 | 0,06 | 1,71 | 8,67E-02 | 2,83E-01 |
| TSPAN33 | 379,90 | -0,17 | 0,10 | -1,71 | 8,68E-02 | 2,83E-01 |
| AZU1 | 39,80 | -0,31 | 0,18 | -1,71 | 8,69E-02 | 2,83E-01 |
| SEC11C | 481,65 | 0,09 | 0,05 | 1,71 | 8,69E-02 | 2,83E-01 |
| UBXN6 | 9274,72 | -0,23 | 0,13 | -1,71 | 8,69E-02 | 2,83E-01 |
| OPLAH | 120,09 | -0,23 | 0,14 | -1,71 | 8,69E-02 | 2,83E-01 |
| TIMM17B | 506,80 | -0,09 | 0,05 | -1,71 | 8,70E-02 | 2,83E-01 |
| H1-6 | 43,89 | -0,16 | 0,09 | -1,71 | 8,70E-02 | 2,84E-01 |
| GZMA | 824,77 | 0,24 | 0,14 | 1,71 | 8,71E-02 | 2,84E-01 |
| PI4K2B | 497,48 | 0,15 | 0,09 | 1,71 | 8,71E-02 | 2,84E-01 |
| MELK | 55,75 | -0,24 | 0,14 | -1,71 | 8,72E-02 | 2,84E-01 |
| SERF2 | 21801,87 | -0,17 | 0,10 | -1,71 | 8,72E-02 | 2,84E-01 |
| TIMMDC1 | 1080,78 | 0,08 | 0,05 | 1,71 | 8,75E-02 | 2,84E-01 |
| ESR2 | 56,97 | -0,32 | 0,19 | -1,71 | 8,74E-02 | 2,84E-01 |
| NDUFS1 | 2555,55 | 0,05 | 0,03 | 1,71 | 8,75E-02 | 2,84E-01 |
| SELENOF | 3646,76 | 0,13 | 0,08 | 1,71 | 8,76E-02 | 2,84E-01 |
| RPL4 | 14207,17 | 0,15 | 0,09 | 1,71 | 8,73E-02 | 2,84E-01 |
| FSCN2 | 26,74 | -0,26 | 0,15 | -1,71 | 8,75E-02 | 2,84E-01 |
| B3GALT9 | 117,62 | 0,09 | 0,05 | 1,71 | 8,75E-02 | 2,84E-01 |
| AIFM3 | 131,09 | -0,22 | 0,13 | -1,71 | 8,74E-02 | 2,84E-01 |
| PADI4 | 1550,94 | -0,19 | 0,11 | -1,71 | 8,74E-02 | 2,84E-01 |
| ACKR1 | 38,38 | -0,29 | 0,17 | -1,71 | 8,73E-02 | 2,84E-01 |
| FURIN | 1453,19 | -0,23 | 0,14 | -1,71 | 8,73E-02 | 2,84E-01 |
| STAG2 | 9334,68 | 0,14 | 0,08 | 1,71 | 8,74E-02 | 2,84E-01 |
| TMED2 | 2374,24 | 0,12 | 0,07 | 1,71 | 8,75E-02 | 2,84E-01 |
| MRM2 | 443,87 | 0,06 | 0,03 | 1,71 | 8,76E-02 | 2,84E-01 |
| AAGAB | 1018,35 | 0,07 | 0,04 | 1,71 | 8,77E-02 | 2,84E-01 |
| KHDRBS1 | 1709,64 | 0,05 | 0,03 | 1,71 | 8,77E-02 | 2,84E-01 |
| WASHC3 | 863,31 | 0,08 | 0,05 | 1,71 | 8,77E-02 | 2,84E-01 |
| CENPX | 265,76 | 0,08 | 0,05 | 1,71 | 8,77E-02 | 2,84E-01 |
| RFXAP | 271,06 | 0,09 | 0,05 | 1,71 | 8,78E-02 | 2,84E-01 |
| PPEF1 | 24,93 | -0,24 | 0,14 | -1,71 | 8,78E-02 | 2,84E-01 |
| TCF3 | 541,45 | -0,16 | 0,09 | -1,71 | 8,79E-02 | 2,85E-01 |
| NUDT15 | 108,14 | 0,12 | 0,07 | 1,71 | 8,80E-02 | 2,85E-01 |
| TMEM101 | 663,68 | 0,07 | 0,04 | 1,71 | 8,80E-02 | 2,85E-01 |

| | | | | | | |
|---|---|---|---|---|---|---|
| C1D | 472,45 | 0,15 | 0,09 | 1,71 | 8,81E-02 | 2,85E-01 |
| GIMAP8 | 1947,24 | 0,09 | 0,05 | 1,71 | 8,82E-02 | 2,85E-01 |
| EYS | 84,69 | -0,20 | 0,11 | -1,70 | 8,83E-02 | 2,85E-01 |
| NATD1 | 694,87 | -0,24 | 0,14 | -1,70 | 8,84E-02 | 2,86E-01 |
| PPP6R1 | 1145,72 | -0,20 | 0,12 | -1,70 | 8,85E-02 | 2,86E-01 |
| TENT5A | 1866,58 | 0,12 | 0,07 | 1,70 | 8,85E-02 | 2,86E-01 |
| IFT140 | 161,09 | -0,18 | 0,11 | -1,70 | 8,84E-02 | 2,86E-01 |
| TMX1 | 1659,17 | 0,18 | 0,11 | 1,70 | 8,85E-02 | 2,86E-01 |
| UPP2 | 26,98 | -0,24 | 0,14 | -1,70 | 8,86E-02 | 2,86E-01 |
| METTL2B | 677,79 | 0,06 | 0,03 | 1,70 | 8,87E-02 | 2,86E-01 |
| PROSER3 | 101,19 | -0,23 | 0,13 | -1,70 | 8,87E-02 | 2,86E-01 |
| ACAA1 | 1694,21 | -0,09 | 0,05 | -1,70 | 8,88E-02 | 2,86E-01 |
| PHF5A | 257,56 | 0,13 | 0,08 | 1,70 | 8,87E-02 | 2,86E-01 |
| PROS1 | 186,43 | -0,25 | 0,15 | -1,70 | 8,88E-02 | 2,86E-01 |
| TNFRSF1A | 2623,65 | -0,15 | 0,09 | -1,70 | 8,88E-02 | 2,86E-01 |
| SSH3 | 468,03 | -0,13 | 0,08 | -1,70 | 8,90E-02 | 2,86E-01 |
| C3orf62 | 1106,38 | -0,10 | 0,06 | -1,70 | 8,93E-02 | 2,86E-01 |
| TASL | 843,17 | 0,13 | 0,08 | 1,70 | 8,90E-02 | 2,86E-01 |
| AP2B1 | 3864,02 | -0,08 | 0,05 | -1,70 | 8,93E-02 | 2,86E-01 |
| PIGF | 660,81 | 0,12 | 0,07 | 1,70 | 8,92E-02 | 2,86E-01 |
| ANAPC1 | 1524,18 | 0,07 | 0,04 | 1,70 | 8,89E-02 | 2,86E-01 |
| CMBL | 48,28 | -0,36 | 0,21 | -1,70 | 8,91E-02 | 2,86E-01 |
| TOLLIP | 861,92 | -0,13 | 0,08 | -1,70 | 8,92E-02 | 2,86E-01 |
| HCN3 | 37,47 | -0,23 | 0,13 | -1,70 | 8,91E-02 | 2,86E-01 |
| PXMP2 | 26,34 | -0,16 | 0,10 | -1,70 | 8,93E-02 | 2,86E-01 |
| DBI | 405,84 | 0,15 | 0,09 | 1,70 | 8,90E-02 | 2,86E-01 |
| CTDSP2 | 4330,60 | -0,14 | 0,08 | -1,70 | 8,92E-02 | 2,86E-01 |
| NIPAL1 | 52,64 | -0,28 | 0,17 | -1,70 | 8,91E-02 | 2,86E-01 |
| ZNF414 | 55,65 | -0,25 | 0,15 | -1,70 | 8,92E-02 | 2,86E-01 |
| MED25 | 1409,08 | -0,13 | 0,08 | -1,70 | 8,91E-02 | 2,86E-01 |
| LIX1L | 1140,75 | -0,07 | 0,04 | -1,70 | 8,89E-02 | 2,86E-01 |
| SLC2A4RG | 189,25 | -0,17 | 0,10 | -1,70 | 8,90E-02 | 2,86E-01 |
| KRT1 | 1395,83 | -0,33 | 0,19 | -1,70 | 8,92E-02 | 2,86E-01 |
| RPS6KA1 | 4722,66 | -0,11 | 0,06 | -1,70 | 8,94E-02 | 2,86E-01 |
| DEPDC1B | 45,82 | -0,24 | 0,14 | -1,70 | 8,94E-02 | 2,86E-01 |
| ARMC7 | 213,05 | -0,11 | 0,07 | -1,70 | 8,94E-02 | 2,86E-01 |
| CARF | 701,37 | 0,10 | 0,06 | 1,70 | 8,94E-02 | 2,86E-01 |
| OCEL1 | 42,02 | -0,21 | 0,12 | -1,70 | 8,95E-02 | 2,87E-01 |
| HOOK2 | 217,70 | -0,11 | 0,07 | -1,70 | 8,95E-02 | 2,87E-01 |
| MT1F | 76,20 | -0,12 | 0,07 | -1,70 | 8,96E-02 | 2,87E-01 |
| GMDS | 342,21 | -0,12 | 0,07 | -1,70 | 8,96E-02 | 2,87E-01 |
| SEMA4D | 6436,39 | -0,17 | 0,10 | -1,70 | 8,97E-02 | 2,87E-01 |
| LRRCC1 | 127,56 | 0,17 | 0,10 | 1,70 | 8,97E-02 | 2,87E-01 |
| DZIP1L | 58,12 | -0,30 | 0,18 | -1,70 | 8,97E-02 | 2,87E-01 |
| DNAI4 | 89,66 | -0,19 | 0,11 | -1,70 | 8,98E-02 | 2,87E-01 |
| MBP | 9104,15 | -0,09 | 0,05 | -1,70 | 8,98E-02 | 2,87E-01 |

| | | | | | | |
|---|---|---|---|---|---|---|
| TRIM25 | 6773,49 | -0,14 | 0,08 | -1,70 | 8,99E-02 | 2,87E-01 |
| RIMBP3 | 64,53 | -0,16 | 0,10 | -1,70 | 8,99E-02 | 2,87E-01 |
| IFT122 | 389,39 | -0,10 | 0,06 | -1,70 | 8,99E-02 | 2,87E-01 |
| CPNE2 | 334,36 | -0,15 | 0,09 | -1,70 | 9,00E-02 | 2,87E-01 |
| MINAR1 | 66,73 | -0,23 | 0,14 | -1,69 | 9,01E-02 | 2,87E-01 |
| RB1 | 2123,57 | 0,10 | 0,06 | 1,70 | 9,00E-02 | 2,87E-01 |
| CTSS | 46828,69 | 0,12 | 0,07 | 1,70 | 9,00E-02 | 2,87E-01 |
| SNIP1 | 1172,84 | 0,06 | 0,03 | 1,70 | 9,01E-02 | 2,87E-01 |
| TRIM23 | 1657,25 | 0,14 | 0,08 | 1,69 | 9,01E-02 | 2,87E-01 |
| NDUFB5 | 855,11 | 0,10 | 0,06 | 1,69 | 9,02E-02 | 2,87E-01 |
| NIP7 | 603,04 | 0,08 | 0,05 | 1,69 | 9,03E-02 | 2,88E-01 |
| SSBP1 | 1292,93 | 0,09 | 0,05 | 1,69 | 9,02E-02 | 2,88E-01 |
| CCDC191 | 449,08 | 0,09 | 0,05 | 1,69 | 9,03E-02 | 2,88E-01 |
| MRRF | 539,06 | 0,06 | 0,04 | 1,69 | 9,03E-02 | 2,88E-01 |
| APOL4 | 42,62 | -0,32 | 0,19 | -1,69 | 9,03E-02 | 2,88E-01 |
| CDC73 | 3571,50 | 0,09 | 0,06 | 1,69 | 9,02E-02 | 2,88E-01 |
| RHBDD2 | 248,92 | -0,12 | 0,07 | -1,69 | 9,04E-02 | 2,88E-01 |
| PTGDR2 | 444,32 | -0,20 | 0,12 | -1,69 | 9,04E-02 | 2,88E-01 |
| SGTB | 1681,83 | 0,10 | 0,06 | 1,69 | 9,05E-02 | 2,88E-01 |
| MITD1 | 860,03 | 0,08 | 0,05 | 1,69 | 9,06E-02 | 2,88E-01 |
| RNF214 | 307,68 | -0,10 | 0,06 | -1,69 | 9,06E-02 | 2,88E-01 |
| FUNDC2 | 800,39 | -0,11 | 0,07 | -1,69 | 9,06E-02 | 2,88E-01 |
| ZNF667 | 85,26 | -0,28 | 0,16 | -1,69 | 9,07E-02 | 2,88E-01 |
| ZFTA | 145,66 | -0,15 | 0,09 | -1,69 | 9,06E-02 | 2,88E-01 |
| CLN3 | 362,58 | -0,12 | 0,07 | -1,69 | 9,07E-02 | 2,88E-01 |
| NIBAN3 | 1224,78 | 0,17 | 0,10 | 1,69 | 9,07E-02 | 2,88E-01 |
| ENSG000001 | 2270,35 | -0,25 | 0,15 | -1,69 | 9,08E-02 | 2,88E-01 |
| TMEM219 | 432,66 | -0,07 | 0,04 | -1,69 | 9,08E-02 | 2,88E-01 |
| SGTA | 182,88 | -0,17 | 0,10 | -1,69 | 9,09E-02 | 2,88E-01 |
| C1orf52 | 558,66 | 0,09 | 0,05 | 1,69 | 9,11E-02 | 2,89E-01 |
| LRBA | 7445,96 | 0,09 | 0,05 | 1,69 | 9,11E-02 | 2,89E-01 |
| SPIDR | 1862,37 | 0,05 | 0,03 | 1,69 | 9,11E-02 | 2,89E-01 |
| MTMR9 | 1580,50 | 0,07 | 0,04 | 1,69 | 9,13E-02 | 2,89E-01 |
| C2CD3 | 1382,48 | -0,08 | 0,04 | -1,69 | 9,13E-02 | 2,89E-01 |
| CUL1 | 1437,12 | 0,07 | 0,04 | 1,69 | 9,14E-02 | 2,89E-01 |
| MTBP | 380,21 | 0,11 | 0,06 | 1,69 | 9,14E-02 | 2,89E-01 |
| ADCY9 | 392,21 | -0,15 | 0,09 | -1,69 | 9,14E-02 | 2,90E-01 |
| RIOX1 | 588,57 | 0,06 | 0,04 | 1,69 | 9,15E-02 | 2,90E-01 |
| COG7 | 368,51 | -0,08 | 0,05 | -1,69 | 9,15E-02 | 2,90E-01 |
| HPRT1 | 117,92 | 0,13 | 0,08 | 1,69 | 9,16E-02 | 2,90E-01 |
| DNM3 | 465,30 | -0,15 | 0,09 | -1,69 | 9,16E-02 | 2,90E-01 |
| NFX1 | 1326,77 | 0,06 | 0,03 | 1,69 | 9,17E-02 | 2,90E-01 |
| ZNF618 | 134,15 | -0,22 | 0,13 | -1,69 | 9,17E-02 | 2,90E-01 |
| SLC30A5 | 1302,98 | 0,07 | 0,04 | 1,69 | 9,17E-02 | 2,90E-01 |
| AK6 | 79,16 | 0,15 | 0,09 | 1,69 | 9,18E-02 | 2,90E-01 |
| CCR3 | 1012,29 | -0,19 | 0,11 | -1,69 | 9,19E-02 | 2,90E-01 |

| | | | | | | |
|---|---|---|---|---|---|---|
| POMZP3 | 62,17 | -0,15 | 0,09 | -1,69 | 9,19E-02 | 2,90E-01 |
| C5 | 227,44 | -0,10 | 0,06 | -1,69 | 9,19E-02 | 2,90E-01 |
| SEC23IP | 935,23 | 0,05 | 0,03 | 1,69 | 9,19E-02 | 2,90E-01 |
| TRIM7 | 63,08 | -0,26 | 0,15 | -1,68 | 9,20E-02 | 2,90E-01 |
| SPRTN | 823,69 | 0,08 | 0,05 | 1,68 | 9,20E-02 | 2,90E-01 |
| TTC3 | 4992,44 | 0,09 | 0,05 | 1,68 | 9,21E-02 | 2,90E-01 |
| C4orf33 | 569,82 | 0,11 | 0,07 | 1,68 | 9,21E-02 | 2,90E-01 |
| NCAPH | 66,99 | -0,17 | 0,10 | -1,68 | 9,20E-02 | 2,90E-01 |
| MICU3 | 226,64 | 0,19 | 0,11 | 1,68 | 9,22E-02 | 2,90E-01 |
| SH3YL1 | 1548,23 | 0,09 | 0,05 | 1,68 | 9,22E-02 | 2,90E-01 |
| ABL1 | 354,13 | -0,13 | 0,08 | -1,68 | 9,22E-02 | 2,90E-01 |
| NDUFS7 | 377,43 | -0,09 | 0,05 | -1,68 | 9,22E-02 | 2,90E-01 |
| SERPINB8 | 558,25 | 0,08 | 0,05 | 1,68 | 9,22E-02 | 2,90E-01 |
| PHC3 | 8282,30 | 0,07 | 0,04 | 1,68 | 9,23E-02 | 2,91E-01 |
| GFRA2 | 57,47 | -0,32 | 0,19 | -1,68 | 9,24E-02 | 2,91E-01 |
| RPL13A | 30122,80 | 0,15 | 0,09 | 1,68 | 9,24E-02 | 2,91E-01 |
| KIFAP3 | 905,06 | 0,11 | 0,07 | 1,68 | 9,24E-02 | 2,91E-01 |
| BARD1 | 547,29 | 0,12 | 0,07 | 1,68 | 9,27E-02 | 2,91E-01 |
| ZNF654 | 1361,48 | 0,17 | 0,10 | 1,68 | 9,27E-02 | 2,91E-01 |
| C16orf87 | 313,84 | 0,08 | 0,05 | 1,68 | 9,26E-02 | 2,91E-01 |
| UBAC1 | 876,37 | -0,08 | 0,05 | -1,68 | 9,27E-02 | 2,91E-01 |
| ZNF451 | 4748,30 | 0,10 | 0,06 | 1,68 | 9,28E-02 | 2,91E-01 |
| FBXO5 | 189,78 | 0,12 | 0,07 | 1,68 | 9,27E-02 | 2,91E-01 |
| HCFC2 | 994,72 | 0,12 | 0,07 | 1,68 | 9,28E-02 | 2,91E-01 |
| CTNS | 265,49 | -0,08 | 0,05 | -1,68 | 9,26E-02 | 2,91E-01 |
| MFSD6 | 1264,26 | 0,07 | 0,04 | 1,68 | 9,28E-02 | 2,91E-01 |
| DNAJC30 | 92,03 | -0,13 | 0,08 | -1,68 | 9,29E-02 | 2,91E-01 |
| ZNF516 | 4836,65 | -0,13 | 0,08 | -1,68 | 9,29E-02 | 2,91E-01 |
| SMAGP | 62,67 | -0,15 | 0,09 | -1,68 | 9,29E-02 | 2,91E-01 |
| GON7 | 172,68 | 0,15 | 0,09 | 1,68 | 9,29E-02 | 2,91E-01 |
| RNF8 | 423,27 | 0,07 | 0,04 | 1,68 | 9,29E-02 | 2,91E-01 |
| SERPINA1 | 5316,41 | -0,15 | 0,09 | -1,68 | 9,32E-02 | 2,92E-01 |
| CDK2AP2 | 90,37 | -0,18 | 0,11 | -1,68 | 9,32E-02 | 2,92E-01 |
| RASGRP4 | 838,85 | -0,24 | 0,14 | -1,68 | 9,32E-02 | 2,92E-01 |
| CBX1 | 1331,36 | 0,10 | 0,06 | 1,68 | 9,32E-02 | 2,92E-01 |
| HYI | 142,86 | -0,11 | 0,07 | -1,68 | 9,31E-02 | 2,92E-01 |
| PLEKHA8 | 768,56 | 0,07 | 0,04 | 1,68 | 9,32E-02 | 2,92E-01 |
| EXOC3L1 | 52,90 | -0,28 | 0,17 | -1,68 | 9,32E-02 | 2,92E-01 |
| CNOT8 | 2956,34 | 0,08 | 0,05 | 1,68 | 9,31E-02 | 2,92E-01 |
| ZDBF2 | 389,72 | 0,18 | 0,11 | 1,68 | 9,34E-02 | 2,92E-01 |
| WDR25 | 209,51 | -0,09 | 0,05 | -1,68 | 9,34E-02 | 2,92E-01 |
| USP25 | 5060,73 | 0,10 | 0,06 | 1,68 | 9,35E-02 | 2,92E-01 |
| SNRNP48 | 1043,55 | 0,12 | 0,07 | 1,68 | 9,34E-02 | 2,92E-01 |
| ARID4B | 6083,26 | 0,10 | 0,06 | 1,68 | 9,34E-02 | 2,92E-01 |
| RNASET2 | 8530,75 | -0,13 | 0,08 | -1,68 | 9,35E-02 | 2,92E-01 |
| TOR2A | 115,94 | -0,17 | 0,10 | -1,68 | 9,35E-02 | 2,92E-01 |

| | | | | | | |
|---|---|---|---|---|---|---|
| HACD1 | 63,55 | -0,21 | 0,13 | -1,68 | 9,33E-02 | 2,92E-01 |
| IQSEC1 | 5582,71 | -0,14 | 0,08 | -1,68 | 9,34E-02 | 2,92E-01 |
| SARAF | 9415,91 | 0,09 | 0,05 | 1,68 | 9,33E-02 | 2,92E-01 |
| MAP10 | 114,39 | 0,15 | 0,09 | 1,68 | 9,35E-02 | 2,92E-01 |
| TCTN2 | 91,20 | -0,16 | 0,10 | -1,68 | 9,36E-02 | 2,92E-01 |
| PLEKHB1 | 165,70 | -0,15 | 0,09 | -1,68 | 9,36E-02 | 2,92E-01 |
| AKR1A1 | 666,25 | 0,06 | 0,04 | 1,68 | 9,36E-02 | 2,92E-01 |
| EXOSC1 | 670,90 | 0,07 | 0,04 | 1,68 | 9,37E-02 | 2,92E-01 |
| MAP3K13 | 169,81 | -0,14 | 0,09 | -1,68 | 9,38E-02 | 2,92E-01 |
| WNT10A | 50,56 | -0,23 | 0,14 | -1,67 | 9,40E-02 | 2,92E-01 |
| TMEM70 | 525,47 | 0,11 | 0,07 | 1,67 | 9,40E-02 | 2,92E-01 |
| PRPF19 | 1286,99 | 0,07 | 0,04 | 1,68 | 9,39E-02 | 2,92E-01 |
| SPINDOC | 83,69 | -0,14 | 0,08 | -1,67 | 9,40E-02 | 2,92E-01 |
| IFT22 | 38,09 | -0,19 | 0,11 | -1,67 | 9,40E-02 | 2,92E-01 |
| FBXL3 | 3748,54 | 0,11 | 0,06 | 1,67 | 9,40E-02 | 2,92E-01 |
| UBE2A | 816,24 | 0,09 | 0,05 | 1,67 | 9,41E-02 | 2,93E-01 |
| C10orf143 | 173,60 | -0,11 | 0,06 | -1,67 | 9,41E-02 | 2,93E-01 |
| PRR5 | 519,00 | -0,15 | 0,09 | -1,67 | 9,42E-02 | 2,93E-01 |
| EXTL2 | 147,50 | 0,11 | 0,07 | 1,67 | 9,43E-02 | 2,93E-01 |
| DIAPH3 | 62,57 | -0,27 | 0,16 | -1,67 | 9,43E-02 | 2,93E-01 |
| ZNRF2 | 1783,44 | 0,10 | 0,06 | 1,67 | 9,42E-02 | 2,93E-01 |
| TOR1A | 876,99 | -0,06 | 0,03 | -1,67 | 9,43E-02 | 2,93E-01 |
| SCIMP | 846,66 | 0,11 | 0,06 | 1,67 | 9,42E-02 | 2,93E-01 |
| FAM174C | 187,70 | -0,10 | 0,06 | -1,67 | 9,42E-02 | 2,93E-01 |
| PARD3B | 65,99 | -0,31 | 0,19 | -1,67 | 9,44E-02 | 2,93E-01 |
| UBE2J2 | 1289,58 | -0,07 | 0,04 | -1,67 | 9,45E-02 | 2,93E-01 |
| NDUFAF7 | 731,28 | 0,08 | 0,05 | 1,67 | 9,45E-02 | 2,93E-01 |
| ABCB1 | 611,74 | 0,13 | 0,08 | 1,67 | 9,46E-02 | 2,93E-01 |
| ZNF34 | 157,21 | 0,07 | 0,04 | 1,67 | 9,46E-02 | 2,93E-01 |
| PTTG1IP | 4784,34 | -0,08 | 0,05 | -1,67 | 9,47E-02 | 2,93E-01 |
| ATP11C | 1887,01 | 0,09 | 0,05 | 1,67 | 9,46E-02 | 2,93E-01 |
| TMEM50B | 1168,21 | 0,11 | 0,06 | 1,67 | 9,47E-02 | 2,93E-01 |
| EBLN2 | 291,18 | 0,07 | 0,04 | 1,67 | 9,47E-02 | 2,93E-01 |
| SIGLEC1 | 140,86 | -0,37 | 0,22 | -1,67 | 9,48E-02 | 2,93E-01 |
| PPP1CC | 3717,44 | 0,09 | 0,05 | 1,67 | 9,48E-02 | 2,93E-01 |
| ANXA1 | 4696,00 | 0,17 | 0,10 | 1,67 | 9,48E-02 | 2,93E-01 |
| NTPCR | 726,45 | 0,10 | 0,06 | 1,67 | 9,49E-02 | 2,93E-01 |
| SLC39A10 | 2578,39 | 0,12 | 0,07 | 1,67 | 9,50E-02 | 2,93E-01 |
| E2F3 | 1122,01 | -0,09 | 0,05 | -1,67 | 9,50E-02 | 2,93E-01 |
| DNAJB11 | 688,35 | 0,07 | 0,04 | 1,67 | 9,53E-02 | 2,93E-01 |
| RAB9A | 582,03 | 0,08 | 0,05 | 1,67 | 9,53E-02 | 2,93E-01 |
| MFAP3 | 1219,00 | 0,08 | 0,05 | 1,67 | 9,50E-02 | 2,93E-01 |
| AKR1E2 | 186,04 | -0,21 | 0,13 | -1,67 | 9,54E-02 | 2,93E-01 |
| RSBN1 | 2089,34 | 0,09 | 0,05 | 1,67 | 9,51E-02 | 2,93E-01 |
| ZDHHC5 | 1864,62 | -0,05 | 0,03 | -1,67 | 9,50E-02 | 2,93E-01 |
| SIGLEC8 | 141,87 | -0,26 | 0,16 | -1,67 | 9,54E-02 | 2,93E-01 |

| | | | | | | |
|---|---|---|---|---|---|---|
| HSD17B8 | 72,71 | 0,11 | 0,06 | 1,67 | 9,52E-02 | 2,93E-01 |
| CCNB2 | 31,07 | -0,23 | 0,14 | -1,67 | 9,52E-02 | 2,93E-01 |
| SCARB2 | 1550,36 | 0,07 | 0,04 | 1,67 | 9,51E-02 | 2,93E-01 |
| SRM | 81,65 | -0,19 | 0,11 | -1,67 | 9,49E-02 | 2,93E-01 |
| IKZF4 | 64,03 | -0,22 | 0,13 | -1,67 | 9,54E-02 | 2,93E-01 |
| SELENOI | 1065,13 | 0,08 | 0,05 | 1,67 | 9,54E-02 | 2,93E-01 |
| NUCB2 | 660,40 | 0,09 | 0,05 | 1,67 | 9,53E-02 | 2,93E-01 |
| GARNL3 | 152,91 | -0,21 | 0,12 | -1,67 | 9,54E-02 | 2,93E-01 |
| MAPKAPK2 | 1322,83 | -0,13 | 0,08 | -1,67 | 9,54E-02 | 2,93E-01 |
| CUL4B | 2097,98 | 0,10 | 0,06 | 1,67 | 9,50E-02 | 2,93E-01 |
| CHL1 | 68,67 | -0,32 | 0,19 | -1,67 | 9,53E-02 | 2,93E-01 |
| FAM153A | 351,64 | -0,19 | 0,11 | -1,67 | 9,54E-02 | 2,93E-01 |
| SBNO2 | 2622,20 | -0,19 | 0,12 | -1,67 | 9,54E-02 | 2,93E-01 |
| ALAD | 706,36 | -0,08 | 0,05 | -1,67 | 9,54E-02 | 2,93E-01 |
| TPK1 | 1087,84 | 0,08 | 0,05 | 1,67 | 9,56E-02 | 2,94E-01 |
| ZDHHC8 | 229,33 | -0,20 | 0,12 | -1,67 | 9,58E-02 | 2,94E-01 |
| UBE2M | 687,70 | -0,10 | 0,06 | -1,67 | 9,58E-02 | 2,94E-01 |
| MAPK1IP1L | 1570,40 | 0,06 | 0,04 | 1,67 | 9,58E-02 | 2,94E-01 |
| EIF2B3 | 342,07 | 0,06 | 0,03 | 1,67 | 9,59E-02 | 2,94E-01 |
| CLIP4 | 2106,65 | 0,08 | 0,05 | 1,66 | 9,59E-02 | 2,95E-01 |
| AGBL2 | 65,96 | -0,16 | 0,10 | -1,66 | 9,59E-02 | 2,95E-01 |
| AGER | 240,31 | -0,20 | 0,12 | -1,66 | 9,60E-02 | 2,95E-01 |
| SLC28A3 | 32,22 | -0,35 | 0,21 | -1,66 | 9,62E-02 | 2,95E-01 |
| FSTL1 | 188,82 | -0,26 | 0,16 | -1,66 | 9,62E-02 | 2,95E-01 |
| FBXL20 | 2257,11 | -0,09 | 0,06 | -1,66 | 9,63E-02 | 2,95E-01 |
| TRMT112 | 1349,13 | 0,10 | 0,06 | 1,66 | 9,63E-02 | 2,95E-01 |
| C20orf204 | 85,59 | -0,19 | 0,12 | -1,66 | 9,63E-02 | 2,95E-01 |
| ZNF506 | 2492,21 | 0,07 | 0,04 | 1,66 | 9,64E-02 | 2,96E-01 |
| LINS1 | 2032,60 | 0,09 | 0,05 | 1,66 | 9,67E-02 | 2,96E-01 |
| ZDHHC4 | 240,51 | -0,05 | 0,03 | -1,66 | 9,66E-02 | 2,96E-01 |
| HNMT | 475,15 | 0,15 | 0,09 | 1,66 | 9,66E-02 | 2,96E-01 |
| NR4A1 | 66,73 | -0,32 | 0,19 | -1,66 | 9,67E-02 | 2,96E-01 |
| DNASE1L3 | 32,36 | -0,24 | 0,15 | -1,66 | 9,67E-02 | 2,96E-01 |
| ARRDC1 | 609,98 | -0,13 | 0,08 | -1,66 | 9,66E-02 | 2,96E-01 |
| SYNGAP1 | 233,82 | -0,24 | 0,14 | -1,66 | 9,68E-02 | 2,96E-01 |
| DPP9 | 685,80 | -0,14 | 0,09 | -1,66 | 9,68E-02 | 2,96E-01 |
| SS18 | 2411,82 | 0,09 | 0,05 | 1,66 | 9,68E-02 | 2,96E-01 |
| LARS2 | 804,50 | 0,06 | 0,03 | 1,66 | 9,69E-02 | 2,96E-01 |
| STON1 | 85,31 | -0,15 | 0,09 | -1,66 | 9,70E-02 | 2,97E-01 |
| CHURC1 | 2006,62 | 0,12 | 0,07 | 1,66 | 9,71E-02 | 2,97E-01 |
| HACD2 | 772,84 | 0,07 | 0,04 | 1,66 | 9,71E-02 | 2,97E-01 |
| GEN1 | 592,85 | 0,11 | 0,07 | 1,66 | 9,71E-02 | 2,97E-01 |
| SLFN14 | 227,33 | -0,20 | 0,12 | -1,66 | 9,71E-02 | 2,97E-01 |
| GAS8 | 126,96 | -0,15 | 0,09 | -1,66 | 9,72E-02 | 2,97E-01 |
| RBMXL1 | 988,32 | 0,10 | 0,06 | 1,66 | 9,72E-02 | 2,97E-01 |
| UBALD1 | 235,22 | -0,15 | 0,09 | -1,66 | 9,72E-02 | 2,97E-01 |

| | | | | | | |
|---|---|---|---|---|---|---|
| JCHAIN | 1595,27 | 0,30 | 0,18 | 1,66 | 9,72E-02 | 2,97E-01 |
| RDH5 | 68,47 | -0,18 | 0,11 | -1,66 | 9,73E-02 | 2,97E-01 |
| DTX3 | 62,24 | -0,24 | 0,14 | -1,66 | 9,73E-02 | 2,97E-01 |
| DLG4 | 172,69 | -0,21 | 0,13 | -1,66 | 9,74E-02 | 2,97E-01 |
| BCAT2 | 103,11 | -0,16 | 0,09 | -1,66 | 9,76E-02 | 2,97E-01 |
| RASA2 | 4409,04 | 0,10 | 0,06 | 1,66 | 9,78E-02 | 2,97E-01 |
| KCNIP2 | 98,02 | -0,19 | 0,12 | -1,66 | 9,79E-02 | 2,97E-01 |
| ADORA2B | 50,27 | -0,19 | 0,11 | -1,66 | 9,75E-02 | 2,97E-01 |
| KRTCAP3 | 41,39 | -0,19 | 0,11 | -1,66 | 9,77E-02 | 2,97E-01 |
| NDUFA13 | 64,01 | -0,15 | 0,09 | -1,66 | 9,78E-02 | 2,97E-01 |
| ITGB5 | 379,95 | -0,23 | 0,14 | -1,66 | 9,77E-02 | 2,97E-01 |
| ITFG1 | 1574,70 | 0,07 | 0,04 | 1,66 | 9,76E-02 | 2,97E-01 |
| IFI27L1 | 75,57 | -0,12 | 0,07 | -1,66 | 9,75E-02 | 2,97E-01 |
| SDHD | 1271,71 | 0,09 | 0,05 | 1,66 | 9,78E-02 | 2,97E-01 |
| CACNA2D2 | 112,81 | -0,28 | 0,17 | -1,66 | 9,77E-02 | 2,97E-01 |
| ABTB1 | 4120,81 | -0,18 | 0,11 | -1,66 | 9,77E-02 | 2,97E-01 |
| ZNF354A | 279,73 | 0,15 | 0,09 | 1,66 | 9,75E-02 | 2,97E-01 |
| NAA16 | 1027,03 | 0,09 | 0,05 | 1,66 | 9,75E-02 | 2,97E-01 |
| SH3BP4 | 68,87 | -0,28 | 0,17 | -1,66 | 9,76E-02 | 2,97E-01 |
| IQCC | 27,92 | -0,16 | 0,10 | -1,66 | 9,79E-02 | 2,97E-01 |
| OLFM1 | 68,18 | -0,36 | 0,22 | -1,66 | 9,78E-02 | 2,97E-01 |
| ANKRD54 | 210,78 | -0,10 | 0,06 | -1,66 | 9,78E-02 | 2,97E-01 |
| FNDC3A | 2692,38 | 0,08 | 0,05 | 1,66 | 9,79E-02 | 2,97E-01 |
| RSPH14 | 42,18 | -0,22 | 0,13 | -1,65 | 9,80E-02 | 2,97E-01 |
| ARFGEF2 | 2222,54 | 0,05 | 0,03 | 1,65 | 9,81E-02 | 2,97E-01 |
| WDR33 | 3239,75 | 0,06 | 0,04 | 1,65 | 9,81E-02 | 2,97E-01 |
| WASF2 | 3465,01 | -0,17 | 0,10 | -1,65 | 9,80E-02 | 2,97E-01 |
| PTPN20 | 36,76 | -0,29 | 0,18 | -1,65 | 9,81E-02 | 2,97E-01 |
| SAR1A | 3431,15 | 0,07 | 0,04 | 1,65 | 9,80E-02 | 2,97E-01 |
| ENSG000002 | 76,47 | -0,15 | 0,09 | -1,65 | 9,81E-02 | 2,97E-01 |
| MYO1A | 38,48 | -0,29 | 0,18 | -1,65 | 9,81E-02 | 2,97E-01 |
| GMPPA | 192,52 | -0,12 | 0,07 | -1,65 | 9,82E-02 | 2,97E-01 |
| LRRC4 | 1170,50 | -0,14 | 0,09 | -1,65 | 9,82E-02 | 2,97E-01 |
| CFL1 | 2503,19 | -0,12 | 0,07 | -1,65 | 9,83E-02 | 2,97E-01 |
| TUBGCP5 | 580,21 | 0,07 | 0,04 | 1,65 | 9,83E-02 | 2,97E-01 |
| GNMT | 31,46 | -0,20 | 0,12 | -1,65 | 9,83E-02 | 2,97E-01 |
| SH3BGRL2 | 1341,34 | -0,21 | 0,13 | -1,65 | 9,83E-02 | 2,97E-01 |
| FAR1 | 4931,79 | 0,15 | 0,09 | 1,65 | 9,87E-02 | 2,97E-01 |
| OXCT1 | 850,33 | 0,10 | 0,06 | 1,65 | 9,86E-02 | 2,97E-01 |
| AGK | 1160,03 | 0,08 | 0,05 | 1,65 | 9,87E-02 | 2,97E-01 |
| RTEL1-TNFRS | 49,56 | -0,26 | 0,16 | -1,65 | 9,87E-02 | 2,97E-01 |
| SEM1 | 781,69 | 0,10 | 0,06 | 1,65 | 9,85E-02 | 2,97E-01 |
| CAD | 483,24 | -0,14 | 0,09 | -1,65 | 9,85E-02 | 2,97E-01 |
| IFIT5 | 1713,15 | 0,22 | 0,13 | 1,65 | 9,86E-02 | 2,97E-01 |
| SGSM3 | 85,66 | -0,12 | 0,07 | -1,65 | 9,87E-02 | 2,97E-01 |
| CMTM1 | 162,51 | -0,11 | 0,07 | -1,65 | 9,84E-02 | 2,97E-01 |

| | | | | | | |
|---|---|---|---|---|---|---|
| CCND3 | 3087,02 | -0,11 | 0,06 | -1,65 | 9,86E-02 | 2,97E-01 |
| TSR2 | 438,29 | 0,07 | 0,04 | 1,65 | 9,87E-02 | 2,97E-01 |
| PLPPR2 | 363,98 | -0,20 | 0,12 | -1,65 | 9,87E-02 | 2,97E-01 |
| INPP5F | 692,21 | 0,11 | 0,07 | 1,65 | 9,88E-02 | 2,97E-01 |
| PIM2 | 1493,20 | -0,11 | 0,07 | -1,65 | 9,85E-02 | 2,97E-01 |
| ARL8B | 1562,49 | 0,08 | 0,05 | 1,65 | 9,88E-02 | 2,97E-01 |
| ZNF24 | 5101,55 | 0,11 | 0,07 | 1,65 | 9,86E-02 | 2,97E-01 |
| EMILIN1 | 57,17 | -0,23 | 0,14 | -1,65 | 9,86E-02 | 2,97E-01 |
| STOML2 | 997,05 | -0,11 | 0,06 | -1,65 | 9,86E-02 | 2,97E-01 |
| ZKSCAN7 | 210,26 | 0,09 | 0,05 | 1,65 | 9,88E-02 | 2,97E-01 |
| FAM3A | 262,94 | -0,10 | 0,06 | -1,65 | 9,89E-02 | 2,97E-01 |
| GSAP | 3274,29 | 0,10 | 0,06 | 1,65 | 9,89E-02 | 2,97E-01 |
| PLEKHO2 | 984,62 | -0,18 | 0,11 | -1,65 | 9,89E-02 | 2,98E-01 |
| KIF2A | 2584,12 | 0,09 | 0,06 | 1,65 | 9,90E-02 | 2,98E-01 |
| RCL1 | 612,46 | -0,11 | 0,07 | -1,65 | 9,90E-02 | 2,98E-01 |
| SLC35G1 | 173,73 | -0,11 | 0,07 | -1,65 | 9,91E-02 | 2,98E-01 |
| C20orf96 | 29,40 | -0,22 | 0,13 | -1,65 | 9,93E-02 | 2,98E-01 |
| PRICKLE3 | 95,58 | -0,15 | 0,09 | -1,65 | 9,93E-02 | 2,98E-01 |
| CCDC152 | 153,77 | 0,18 | 0,11 | 1,65 | 9,93E-02 | 2,98E-01 |
| GTF2A1 | 2399,79 | 0,10 | 0,06 | 1,65 | 9,93E-02 | 2,98E-01 |
| FCSK | 208,23 | -0,20 | 0,12 | -1,65 | 9,94E-02 | 2,98E-01 |
| PML | 633,14 | -0,21 | 0,13 | -1,65 | 9,93E-02 | 2,98E-01 |
| PPP1R8 | 1032,34 | 0,08 | 0,05 | 1,65 | 9,93E-02 | 2,98E-01 |
| CEPT1 | 2516,99 | 0,10 | 0,06 | 1,65 | 9,94E-02 | 2,98E-01 |
| RNF2 | 734,37 | 0,11 | 0,06 | 1,65 | 9,93E-02 | 2,98E-01 |
| SAC3D1 | 56,37 | -0,13 | 0,08 | -1,65 | 9,94E-02 | 2,98E-01 |
| TTC28 | 144,47 | -0,27 | 0,16 | -1,65 | 9,94E-02 | 2,98E-01 |
| MINDY2 | 1710,25 | 0,11 | 0,07 | 1,65 | 9,95E-02 | 2,98E-01 |
| MAN2A1 | 4599,62 | 0,08 | 0,05 | 1,65 | 9,95E-02 | 2,98E-01 |
| RGL2 | 1756,79 | -0,13 | 0,08 | -1,65 | 9,95E-02 | 2,98E-01 |
| DCAF12 | 31063,03 | -0,23 | 0,14 | -1,65 | 9,97E-02 | 2,99E-01 |
| SEZ6L | 75,24 | -0,29 | 0,18 | -1,65 | 9,97E-02 | 2,99E-01 |
| KLHL24 | 3964,57 | 0,07 | 0,05 | 1,65 | 9,97E-02 | 2,99E-01 |
| PCNP | 1966,77 | 0,13 | 0,08 | 1,65 | 9,98E-02 | 2,99E-01 |
| MYG1 | 417,27 | -0,08 | 0,05 | -1,65 | 9,99E-02 | 2,99E-01 |
| METAP1 | 898,07 | 0,07 | 0,04 | 1,65 | 9,99E-02 | 2,99E-01 |
| SLC41A3 | 409,55 | -0,08 | 0,05 | -1,65 | 9,99E-02 | 2,99E-01 |
| ZNF354C | 494,58 | 0,15 | 0,09 | 1,64 | 1,00E-01 | 2,99E-01 |
| OTULIN | 3796,87 | 0,07 | 0,04 | 1,64 | 1,00E-01 | 2,99E-01 |
| ELAC1 | 203,64 | 0,08 | 0,05 | 1,64 | 1,00E-01 | 2,99E-01 |
| VRK2 | 937,85 | 0,13 | 0,08 | 1,64 | 1,00E-01 | 2,99E-01 |
| IREB2 | 2755,47 | 0,07 | 0,04 | 1,64 | 1,00E-01 | 2,99E-01 |
| SLX4 | 161,88 | -0,23 | 0,14 | -1,64 | 1,00E-01 | 2,99E-01 |
| OTUD1 | 690,01 | 0,10 | 0,06 | 1,64 | 1,00E-01 | 2,99E-01 |
| MRPL4 | 94,22 | -0,16 | 0,10 | -1,64 | 1,00E-01 | 2,99E-01 |
| LDHD | 38,87 | -0,26 | 0,16 | -1,64 | 1,00E-01 | 2,99E-01 |

| | | | | | | |
|---|---|---|---|---|---|---|
| HAUS8 | 181,17 | -0,08 | 0,05 | -1,64 | 1,00E-01 | 2,99E-01 |
| OAZ2 | 4058,03 | -0,12 | 0,07 | -1,64 | 1,00E-01 | 2,99E-01 |
| TNPO1 | 7472,48 | 0,11 | 0,07 | 1,64 | 1,00E-01 | 2,99E-01 |
| HCAR2 | 642,67 | -0,14 | 0,09 | -1,64 | 1,00E-01 | 2,99E-01 |
| RNF213 | 27829,83 | -0,16 | 0,09 | -1,64 | 1,00E-01 | 2,99E-01 |
| RAB14 | 2614,24 | 0,08 | 0,05 | 1,64 | 1,00E-01 | 2,99E-01 |
| HDAC11 | 139,78 | -0,13 | 0,08 | -1,64 | 1,00E-01 | 2,99E-01 |
| MYPOP | 62,17 | -0,21 | 0,13 | -1,64 | 1,01E-01 | 2,99E-01 |
| FAM110C | 53,33 | -0,23 | 0,14 | -1,64 | 1,01E-01 | 3,00E-01 |
| VDAC1 | 767,27 | 0,05 | 0,03 | 1,64 | 1,01E-01 | 3,00E-01 |
| MUC20 | 32,76 | -0,29 | 0,18 | -1,64 | 1,01E-01 | 3,00E-01 |
| TMED5 | 2921,59 | 0,11 | 0,07 | 1,64 | 1,01E-01 | 3,00E-01 |
| RAP2B | 4091,04 | 0,08 | 0,05 | 1,64 | 1,01E-01 | 3,00E-01 |
| USP20 | 1273,88 | -0,08 | 0,05 | -1,64 | 1,01E-01 | 3,00E-01 |
| ACTG1 | 20391,79 | -0,08 | 0,05 | -1,64 | 1,01E-01 | 3,01E-01 |
| SDK2 | 125,93 | -0,33 | 0,20 | -1,64 | 1,01E-01 | 3,01E-01 |
| ERBB2 | 211,11 | -0,18 | 0,11 | -1,64 | 1,01E-01 | 3,01E-01 |
| SLC44A1 | 1405,99 | 0,07 | 0,04 | 1,64 | 1,01E-01 | 3,01E-01 |
| REEP4 | 115,34 | -0,19 | 0,11 | -1,64 | 1,01E-01 | 3,01E-01 |
| CD1C | 281,75 | 0,13 | 0,08 | 1,64 | 1,01E-01 | 3,01E-01 |
| IL1RN | 558,44 | -0,13 | 0,08 | -1,64 | 1,02E-01 | 3,02E-01 |
| RBCK1 | 1261,33 | -0,13 | 0,08 | -1,64 | 1,02E-01 | 3,02E-01 |
| MINDY1 | 1695,38 | -0,15 | 0,09 | -1,64 | 1,02E-01 | 3,02E-01 |
| MRPL1 | 219,46 | 0,16 | 0,10 | 1,64 | 1,02E-01 | 3,02E-01 |
| SORD | 324,35 | -0,12 | 0,08 | -1,64 | 1,02E-01 | 3,02E-01 |
| NXT2 | 305,35 | 0,18 | 0,11 | 1,64 | 1,02E-01 | 3,02E-01 |
| AMZ2 | 820,51 | 0,06 | 0,04 | 1,64 | 1,02E-01 | 3,02E-01 |
| STARD5 | 84,75 | -0,14 | 0,08 | -1,64 | 1,02E-01 | 3,02E-01 |
| GIMAP2 | 2212,71 | 0,17 | 0,10 | 1,64 | 1,02E-01 | 3,02E-01 |
| ZNF502 | 138,62 | 0,18 | 0,11 | 1,64 | 1,02E-01 | 3,02E-01 |
| ORC4 | 1635,04 | 0,11 | 0,07 | 1,64 | 1,02E-01 | 3,02E-01 |
| WDR55 | 1145,45 | -0,05 | 0,03 | -1,64 | 1,02E-01 | 3,02E-01 |
| FAM199X | 2344,01 | 0,10 | 0,06 | 1,64 | 1,02E-01 | 3,02E-01 |
| ETFB | 318,71 | -0,06 | 0,04 | -1,64 | 1,02E-01 | 3,02E-01 |
| FKBP5 | 3509,52 | -0,14 | 0,09 | -1,64 | 1,02E-01 | 3,02E-01 |
| LRPAP1 | 2793,21 | -0,07 | 0,04 | -1,64 | 1,02E-01 | 3,02E-01 |
| CCR8 | 50,44 | 0,18 | 0,11 | 1,64 | 1,02E-01 | 3,02E-01 |
| SP2 | 185,51 | -0,13 | 0,08 | -1,63 | 1,02E-01 | 3,02E-01 |
| EIF4E2 | 2390,58 | 0,06 | 0,04 | 1,63 | 1,02E-01 | 3,02E-01 |
| PGAP2 | 167,53 | -0,11 | 0,06 | -1,63 | 1,02E-01 | 3,02E-01 |
| ARMCX4 | 293,29 | -0,11 | 0,07 | -1,63 | 1,02E-01 | 3,02E-01 |
| AZIN1 | 3959,14 | 0,10 | 0,06 | 1,63 | 1,02E-01 | 3,02E-01 |
| FZR1 | 1343,48 | -0,11 | 0,07 | -1,63 | 1,02E-01 | 3,02E-01 |
| RING1 | 431,84 | 0,06 | 0,04 | 1,63 | 1,02E-01 | 3,02E-01 |
| CLPX | 1760,30 | 0,06 | 0,04 | 1,63 | 1,02E-01 | 3,02E-01 |
| FAM117B | 1143,86 | 0,09 | 0,06 | 1,63 | 1,03E-01 | 3,03E-01 |

| | | | | | | |
|---|---|---|---|---|---|---|
| ADD2 | 286,39 | -0,17 | 0,10 | -1,63 | 1,03E-01 | 3,03E-01 |
| TNFRSF10A | 953,91 | 0,07 | 0,04 | 1,63 | 1,03E-01 | 3,03E-01 |
| H2AC25 | 330,31 | -0,15 | 0,09 | -1,63 | 1,03E-01 | 3,03E-01 |
| IDH3G | 604,16 | -0,07 | 0,04 | -1,63 | 1,03E-01 | 3,03E-01 |
| MAPK11 | 65,01 | -0,17 | 0,11 | -1,63 | 1,03E-01 | 3,03E-01 |
| DLD | 1602,54 | 0,10 | 0,06 | 1,63 | 1,03E-01 | 3,03E-01 |
| LDB2 | 44,48 | -0,30 | 0,18 | -1,63 | 1,03E-01 | 3,03E-01 |
| FBXL8 | 127,35 | -0,11 | 0,07 | -1,63 | 1,03E-01 | 3,03E-01 |
| DMTN | 5786,81 | -0,28 | 0,17 | -1,63 | 1,03E-01 | 3,03E-01 |
| ABCC6 | 96,75 | -0,26 | 0,16 | -1,63 | 1,03E-01 | 3,03E-01 |
| NR3C1 | 7944,41 | 0,09 | 0,05 | 1,63 | 1,03E-01 | 3,03E-01 |
| RNF26 | 72,69 | -0,19 | 0,12 | -1,63 | 1,03E-01 | 3,03E-01 |
| SNRPA1 | 780,33 | 0,07 | 0,04 | 1,63 | 1,03E-01 | 3,03E-01 |
| PKP2 | 58,37 | -0,26 | 0,16 | -1,63 | 1,03E-01 | 3,03E-01 |
| TOP1MT | 230,31 | -0,14 | 0,08 | -1,63 | 1,03E-01 | 3,03E-01 |
| SLC37A1 | 656,61 | -0,07 | 0,04 | -1,63 | 1,03E-01 | 3,03E-01 |
| STAU1 | 1339,46 | -0,11 | 0,07 | -1,63 | 1,03E-01 | 3,03E-01 |
| PCDHGA6 | 60,69 | -0,21 | 0,13 | -1,63 | 1,03E-01 | 3,03E-01 |
| FCGR2A | 17085,26 | -0,12 | 0,07 | -1,63 | 1,03E-01 | 3,03E-01 |
| DHTKD1 | 1409,09 | -0,07 | 0,04 | -1,63 | 1,03E-01 | 3,03E-01 |
| UBAP2 | 936,41 | -0,10 | 0,06 | -1,63 | 1,03E-01 | 3,03E-01 |
| AKAP7 | 442,72 | 0,14 | 0,08 | 1,63 | 1,03E-01 | 3,03E-01 |
| FASTKD1 | 688,20 | 0,11 | 0,06 | 1,63 | 1,03E-01 | 3,03E-01 |
| COA3 | 359,80 | 0,08 | 0,05 | 1,63 | 1,03E-01 | 3,03E-01 |
| TPRA1 | 435,27 | -0,07 | 0,04 | -1,63 | 1,03E-01 | 3,03E-01 |
| MROH1 | 494,43 | -0,21 | 0,13 | -1,63 | 1,04E-01 | 3,04E-01 |
| SMAD1 | 141,18 | -0,18 | 0,11 | -1,63 | 1,04E-01 | 3,04E-01 |
| ZNF202 | 384,08 | 0,06 | 0,04 | 1,63 | 1,04E-01 | 3,04E-01 |
| ALDH1A1 | 713,16 | 0,15 | 0,09 | 1,63 | 1,04E-01 | 3,04E-01 |
| CLUH | 260,75 | -0,19 | 0,11 | -1,63 | 1,04E-01 | 3,04E-01 |
| C6orf47 | 1008,25 | 0,06 | 0,04 | 1,63 | 1,04E-01 | 3,04E-01 |
| DYSF | 5085,11 | -0,21 | 0,13 | -1,63 | 1,04E-01 | 3,04E-01 |
| EPN1 | 819,17 | -0,18 | 0,11 | -1,63 | 1,04E-01 | 3,04E-01 |
| SCP2 | 1620,54 | 0,10 | 0,06 | 1,63 | 1,04E-01 | 3,04E-01 |
| ALG13 | 2287,79 | 0,09 | 0,06 | 1,63 | 1,04E-01 | 3,04E-01 |
| TUG1 | 7718,59 | 0,08 | 0,05 | 1,63 | 1,04E-01 | 3,04E-01 |
| TMIGD3 | 33,84 | -0,25 | 0,16 | -1,63 | 1,04E-01 | 3,04E-01 |
| NSFL1C | 2752,64 | -0,10 | 0,06 | -1,63 | 1,04E-01 | 3,04E-01 |
| DNAAF10 | 167,67 | 0,07 | 0,04 | 1,63 | 1,04E-01 | 3,04E-01 |
| RHEB | 816,39 | 0,07 | 0,04 | 1,63 | 1,04E-01 | 3,04E-01 |
| FANCE | 148,72 | -0,08 | 0,05 | -1,63 | 1,04E-01 | 3,04E-01 |
| EPB41L4A | 191,94 | -0,19 | 0,12 | -1,63 | 1,04E-01 | 3,04E-01 |
| EPB42 | 3299,37 | -0,24 | 0,15 | -1,63 | 1,04E-01 | 3,04E-01 |
| CLEC16A | 1346,19 | -0,11 | 0,07 | -1,62 | 1,04E-01 | 3,04E-01 |
| NPEPL1 | 335,58 | -0,16 | 0,10 | -1,62 | 1,04E-01 | 3,04E-01 |
| BSDC1 | 2600,51 | -0,06 | 0,03 | -1,62 | 1,04E-01 | 3,04E-01 |

| | | | | | | |
|---|---|---|---|---|---|---|
| SP4 | 1793,15 | 0,08 | 0,05 | 1,62 | 1,04E-01 | 3,04E-01 |
| SNPH | 161,30 | -0,18 | 0,11 | -1,62 | 1,04E-01 | 3,04E-01 |
| SEC22B | 3404,99 | 0,10 | 0,06 | 1,62 | 1,04E-01 | 3,04E-01 |
| ARRDC3 | 9419,71 | 0,16 | 0,10 | 1,62 | 1,05E-01 | 3,05E-01 |
| STAG1 | 3247,47 | 0,08 | 0,05 | 1,62 | 1,05E-01 | 3,05E-01 |
| DDA1 | 253,79 | -0,11 | 0,07 | -1,62 | 1,05E-01 | 3,05E-01 |
| BLOC1S6 | 2367,44 | 0,11 | 0,07 | 1,62 | 1,05E-01 | 3,05E-01 |
| PIK3R4 | 1619,06 | 0,06 | 0,04 | 1,62 | 1,05E-01 | 3,05E-01 |
| DTYMK | 180,01 | -0,08 | 0,05 | -1,62 | 1,05E-01 | 3,05E-01 |
| SAMTOR | 935,15 | 0,12 | 0,07 | 1,62 | 1,04E-01 | 3,05E-01 |
| TBRG1 | 2305,29 | 0,05 | 0,03 | 1,62 | 1,05E-01 | 3,05E-01 |
| USP38 | 1733,02 | 0,07 | 0,04 | 1,62 | 1,05E-01 | 3,05E-01 |
| ECPAS | 3063,24 | 0,06 | 0,03 | 1,62 | 1,05E-01 | 3,05E-01 |
| CLEC11A | 59,29 | -0,23 | 0,14 | -1,62 | 1,05E-01 | 3,05E-01 |
| SSBP2 | 1933,23 | 0,09 | 0,06 | 1,62 | 1,05E-01 | 3,05E-01 |
| CEP131 | 178,49 | -0,15 | 0,09 | -1,62 | 1,05E-01 | 3,05E-01 |
| MAP3K10 | 141,02 | -0,19 | 0,12 | -1,62 | 1,05E-01 | 3,05E-01 |
| NECTIN1 | 133,71 | -0,22 | 0,13 | -1,62 | 1,05E-01 | 3,05E-01 |
| ASTN2 | 177,83 | -0,17 | 0,10 | -1,62 | 1,05E-01 | 3,05E-01 |
| PSIP1 | 3125,88 | 0,10 | 0,06 | 1,62 | 1,05E-01 | 3,05E-01 |
| BORA | 288,07 | 0,11 | 0,07 | 1,62 | 1,05E-01 | 3,05E-01 |
| PEMT | 98,62 | -0,12 | 0,08 | -1,62 | 1,05E-01 | 3,05E-01 |
| ARL6 | 186,11 | 0,15 | 0,09 | 1,62 | 1,05E-01 | 3,05E-01 |
| SLC2A5 | 94,33 | -0,24 | 0,15 | -1,62 | 1,05E-01 | 3,05E-01 |
| ZNF420 | 354,60 | 0,12 | 0,08 | 1,62 | 1,05E-01 | 3,05E-01 |
| PPP2CA | 2432,09 | 0,08 | 0,05 | 1,62 | 1,05E-01 | 3,05E-01 |
| GAS2 | 54,29 | -0,17 | 0,11 | -1,62 | 1,05E-01 | 3,05E-01 |
| GPR174 | 2566,71 | 0,14 | 0,09 | 1,62 | 1,05E-01 | 3,05E-01 |
| PCNX4 | 3065,03 | 0,07 | 0,04 | 1,62 | 1,05E-01 | 3,05E-01 |
| RBM39 | 17075,18 | 0,10 | 0,06 | 1,62 | 1,05E-01 | 3,05E-01 |
| EIF4EBP3 | 24,35 | -0,16 | 0,10 | -1,62 | 1,05E-01 | 3,05E-01 |
| PEX12 | 136,37 | 0,09 | 0,05 | 1,62 | 1,05E-01 | 3,05E-01 |
| CEP95 | 1249,36 | 0,07 | 0,04 | 1,62 | 1,05E-01 | 3,05E-01 |
| CEP104 | 850,63 | -0,09 | 0,06 | -1,62 | 1,05E-01 | 3,05E-01 |
| ZNF70 | 203,02 | -0,13 | 0,08 | -1,62 | 1,06E-01 | 3,05E-01 |
| ANAPC16 | 3379,07 | 0,08 | 0,05 | 1,62 | 1,06E-01 | 3,05E-01 |
| ASNSD1 | 891,62 | 0,18 | 0,11 | 1,62 | 1,06E-01 | 3,05E-01 |
| RPS23 | 7823,46 | 0,19 | 0,12 | 1,62 | 1,06E-01 | 3,06E-01 |
| OXSM | 176,57 | 0,09 | 0,06 | 1,62 | 1,06E-01 | 3,06E-01 |
| BLZF1 | 820,99 | 0,13 | 0,08 | 1,62 | 1,06E-01 | 3,06E-01 |
| TMED7 | 1813,44 | 0,14 | 0,09 | 1,62 | 1,06E-01 | 3,06E-01 |
| PTRH1 | 137,41 | -0,11 | 0,07 | -1,62 | 1,06E-01 | 3,06E-01 |
| STAM2 | 2308,61 | 0,11 | 0,07 | 1,62 | 1,06E-01 | 3,06E-01 |
| SCYL3 | 1430,20 | 0,07 | 0,04 | 1,62 | 1,06E-01 | 3,06E-01 |
| FUNDC1 | 134,40 | 0,11 | 0,07 | 1,62 | 1,06E-01 | 3,06E-01 |
| TLR10 | 678,21 | 0,12 | 0,07 | 1,62 | 1,06E-01 | 3,06E-01 |

| | | | | | | |
|---|---|---|---|---|---|---|
| MAB21L3 | 67,00 | -0,21 | 0,13 | -1,62 | 1,06E-01 | 3,06E-01 |
| ABHD18 | 1468,65 | 0,05 | 0,03 | 1,62 | 1,06E-01 | 3,06E-01 |
| NCF4 | 1564,07 | -0,15 | 0,09 | -1,61 | 1,06E-01 | 3,07E-01 |
| COMMD3 | 293,99 | 0,14 | 0,09 | 1,61 | 1,06E-01 | 3,07E-01 |
| PRCD | 50,44 | -0,27 | 0,17 | -1,61 | 1,06E-01 | 3,07E-01 |
| FRS2 | 1147,96 | 0,08 | 0,05 | 1,61 | 1,07E-01 | 3,07E-01 |
| ZNF571 | 208,61 | 0,13 | 0,08 | 1,61 | 1,07E-01 | 3,07E-01 |
| STUB1 | 523,51 | -0,06 | 0,04 | -1,61 | 1,06E-01 | 3,07E-01 |
| C21orf91 | 2061,37 | 0,16 | 0,10 | 1,61 | 1,06E-01 | 3,07E-01 |
| MSANTD2 | 595,14 | 0,07 | 0,04 | 1,61 | 1,06E-01 | 3,07E-01 |
| DYNC1LI2 | 4334,75 | 0,06 | 0,04 | 1,61 | 1,07E-01 | 3,07E-01 |
| NFU1 | 237,52 | 0,08 | 0,05 | 1,61 | 1,07E-01 | 3,07E-01 |
| GABPB1 | 729,25 | 0,09 | 0,06 | 1,61 | 1,07E-01 | 3,07E-01 |
| PLEKHA1 | 2825,97 | 0,10 | 0,06 | 1,61 | 1,07E-01 | 3,07E-01 |
| SOX6 | 894,55 | -0,20 | 0,12 | -1,61 | 1,07E-01 | 3,07E-01 |
| FBXO7 | 19791,95 | -0,18 | 0,11 | -1,61 | 1,07E-01 | 3,07E-01 |
| BCL11A | 2272,50 | 0,13 | 0,08 | 1,61 | 1,07E-01 | 3,07E-01 |
| WDR53 | 212,25 | 0,07 | 0,04 | 1,61 | 1,07E-01 | 3,07E-01 |
| SOCS5 | 723,04 | 0,09 | 0,06 | 1,61 | 1,07E-01 | 3,07E-01 |
| KRBOX5 | 872,17 | 0,10 | 0,06 | 1,61 | 1,07E-01 | 3,07E-01 |
| TPRN | 95,55 | -0,17 | 0,11 | -1,61 | 1,07E-01 | 3,07E-01 |
| EIF3I | 1133,58 | 0,08 | 0,05 | 1,61 | 1,07E-01 | 3,07E-01 |
| ITSN2 | 8735,21 | 0,08 | 0,05 | 1,61 | 1,07E-01 | 3,08E-01 |
| INKA1 | 42,50 | -0,13 | 0,08 | -1,61 | 1,07E-01 | 3,08E-01 |
| CILK1 | 688,66 | 0,05 | 0,03 | 1,61 | 1,07E-01 | 3,08E-01 |
| CCDC186 | 2474,09 | 0,12 | 0,08 | 1,61 | 1,07E-01 | 3,08E-01 |
| RNPEP | 831,46 | -0,13 | 0,08 | -1,61 | 1,08E-01 | 3,08E-01 |
| ANKRD13C | 804,18 | 0,09 | 0,06 | 1,61 | 1,08E-01 | 3,08E-01 |
| TAS2R13 | 132,47 | 0,15 | 0,10 | 1,61 | 1,08E-01 | 3,08E-01 |
| TUBA1C | 604,57 | -0,09 | 0,05 | -1,61 | 1,07E-01 | 3,08E-01 |
| TMEM120B | 136,76 | -0,16 | 0,10 | -1,61 | 1,08E-01 | 3,08E-01 |
| PPP4R2 | 2990,05 | 0,15 | 0,09 | 1,61 | 1,08E-01 | 3,08E-01 |
| NPHP4 | 261,44 | -0,22 | 0,14 | -1,61 | 1,07E-01 | 3,08E-01 |
| GTF3A | 1458,08 | 0,08 | 0,05 | 1,61 | 1,07E-01 | 3,08E-01 |
| MSH5 | 61,02 | -0,16 | 0,10 | -1,61 | 1,07E-01 | 3,08E-01 |
| IMPA1 | 903,76 | 0,10 | 0,06 | 1,61 | 1,07E-01 | 3,08E-01 |
| CNOT3 | 270,53 | -0,21 | 0,13 | -1,61 | 1,08E-01 | 3,08E-01 |
| CUTC | 635,78 | 0,09 | 0,06 | 1,61 | 1,07E-01 | 3,08E-01 |
| FAM81A | 40,78 | -0,32 | 0,20 | -1,61 | 1,08E-01 | 3,08E-01 |
| MOCS3 | 465,41 | 0,07 | 0,04 | 1,61 | 1,07E-01 | 3,08E-01 |
| SH2B3 | 2474,63 | -0,11 | 0,07 | -1,61 | 1,08E-01 | 3,08E-01 |
| RDX | 600,91 | 0,10 | 0,06 | 1,61 | 1,08E-01 | 3,08E-01 |
| STAT4 | 2559,19 | 0,11 | 0,07 | 1,61 | 1,08E-01 | 3,08E-01 |
| TXNRD3 | 78,60 | -0,17 | 0,11 | -1,61 | 1,08E-01 | 3,08E-01 |
| PBX4 | 105,61 | -0,16 | 0,10 | -1,61 | 1,08E-01 | 3,08E-01 |
| TLCD4 | 587,20 | -0,19 | 0,12 | -1,61 | 1,08E-01 | 3,08E-01 |

| | | | | | | |
|---|---|---|---|---|---|---|
| DCAF11 | 2867,96 | -0,09 | 0,05 | -1,61 | 1,08E-01 | 3,08E-01 |
| PTGS1 | 1157,66 | -0,17 | 0,11 | -1,61 | 1,08E-01 | 3,08E-01 |
| FBXO46 | 126,12 | -0,19 | 0,12 | -1,61 | 1,08E-01 | 3,08E-01 |
| ZNF696 | 275,35 | -0,09 | 0,06 | -1,61 | 1,08E-01 | 3,08E-01 |
| H3C10 | 2587,52 | -0,17 | 0,10 | -1,61 | 1,08E-01 | 3,08E-01 |
| MAN2B2 | 1167,14 | -0,11 | 0,07 | -1,61 | 1,08E-01 | 3,08E-01 |
| ARPP19 | 4575,55 | 0,09 | 0,06 | 1,61 | 1,08E-01 | 3,08E-01 |
| SLC38A6 | 520,10 | 0,09 | 0,05 | 1,61 | 1,08E-01 | 3,08E-01 |
| SETX | 17154,23 | 0,09 | 0,06 | 1,61 | 1,08E-01 | 3,08E-01 |
| CSMD1 | 79,59 | -0,50 | 0,31 | -1,61 | 1,08E-01 | 3,08E-01 |
| FAM229A | 191,44 | -0,17 | 0,11 | -1,61 | 1,08E-01 | 3,08E-01 |
| RANBP3 | 727,71 | -0,13 | 0,08 | -1,61 | 1,08E-01 | 3,08E-01 |
| ERLIN2 | 891,16 | 0,04 | 0,03 | 1,61 | 1,08E-01 | 3,08E-01 |
| TMEM62 | 765,44 | 0,06 | 0,04 | 1,61 | 1,08E-01 | 3,08E-01 |
| IMPDH1 | 1654,84 | -0,14 | 0,09 | -1,61 | 1,08E-01 | 3,08E-01 |
| PLEKHA3 | 2169,64 | 0,06 | 0,04 | 1,61 | 1,08E-01 | 3,08E-01 |
| SGPP2 | 41,21 | -0,23 | 0,14 | -1,61 | 1,08E-01 | 3,08E-01 |
| BEGAIN | 86,99 | -0,37 | 0,23 | -1,61 | 1,08E-01 | 3,08E-01 |
| ARID5B | 1997,60 | 0,09 | 0,06 | 1,60 | 1,09E-01 | 3,09E-01 |
| RIC8B | 806,74 | 0,08 | 0,05 | 1,60 | 1,09E-01 | 3,09E-01 |
| PPP1CA | 1918,12 | -0,08 | 0,05 | -1,60 | 1,09E-01 | 3,09E-01 |
| CSPP1 | 645,91 | 0,08 | 0,05 | 1,60 | 1,09E-01 | 3,09E-01 |
| MRPL33 | 488,31 | 0,10 | 0,07 | 1,60 | 1,09E-01 | 3,09E-01 |
| CLU | 389,66 | -0,25 | 0,16 | -1,60 | 1,09E-01 | 3,09E-01 |
| EVI5L | 151,76 | -0,19 | 0,12 | -1,60 | 1,09E-01 | 3,09E-01 |
| FANCD2 | 718,56 | 0,06 | 0,04 | 1,60 | 1,09E-01 | 3,09E-01 |
| CA1 | 2555,22 | -0,29 | 0,18 | -1,60 | 1,09E-01 | 3,09E-01 |
| RLF | 2103,11 | 0,12 | 0,07 | 1,60 | 1,09E-01 | 3,09E-01 |
| PRPF18 | 736,28 | 0,10 | 0,06 | 1,60 | 1,09E-01 | 3,09E-01 |
| OSBP | 1297,15 | 0,04 | 0,03 | 1,60 | 1,09E-01 | 3,09E-01 |
| CCDC24 | 54,49 | -0,20 | 0,12 | -1,60 | 1,09E-01 | 3,09E-01 |
| TASOR2 | 3474,30 | 0,06 | 0,04 | 1,60 | 1,09E-01 | 3,10E-01 |
| CAST | 5672,58 | 0,06 | 0,04 | 1,60 | 1,09E-01 | 3,10E-01 |
| SUGT1 | 2273,33 | 0,06 | 0,04 | 1,60 | 1,09E-01 | 3,10E-01 |
| GNB1L | 151,48 | -0,13 | 0,08 | -1,60 | 1,09E-01 | 3,10E-01 |
| OGDH | 1480,10 | -0,11 | 0,07 | -1,60 | 1,09E-01 | 3,10E-01 |
| YBX1 | 12563,84 | -0,16 | 0,10 | -1,60 | 1,10E-01 | 3,10E-01 |
| C19orf44 | 47,04 | -0,20 | 0,12 | -1,60 | 1,10E-01 | 3,11E-01 |
| DNAJB4 | 397,76 | 0,09 | 0,06 | 1,60 | 1,10E-01 | 3,11E-01 |
| RPTOR | 863,46 | -0,14 | 0,09 | -1,60 | 1,10E-01 | 3,11E-01 |
| SKP2 | 737,24 | 0,07 | 0,04 | 1,60 | 1,10E-01 | 3,11E-01 |
| TDP1 | 881,24 | 0,06 | 0,03 | 1,60 | 1,10E-01 | 3,11E-01 |
| NOTCH1 | 3714,76 | -0,21 | 0,13 | -1,60 | 1,10E-01 | 3,11E-01 |
| CRYBG1 | 5107,16 | 0,06 | 0,04 | 1,60 | 1,10E-01 | 3,11E-01 |
| ZNF653 | 127,98 | -0,11 | 0,07 | -1,60 | 1,10E-01 | 3,11E-01 |
| KLF10 | 325,23 | 0,12 | 0,07 | 1,60 | 1,10E-01 | 3,11E-01 |

| | | | | | | |
|---|---|---|---|---|---|---|
| EFHD2 | 3642,80 | -0,14 | 0,09 | -1,60 | 1,10E-01 | 3,11E-01 |
| SCN9A | 194,88 | -0,17 | 0,11 | -1,60 | 1,10E-01 | 3,11E-01 |
| LY6E | 429,08 | -0,22 | 0,14 | -1,60 | 1,10E-01 | 3,12E-01 |
| UGT8 | 89,85 | 0,19 | 0,12 | 1,60 | 1,10E-01 | 3,12E-01 |
| AP1G2 | 1211,91 | -0,11 | 0,07 | -1,60 | 1,10E-01 | 3,12E-01 |
| TVP23B | 912,33 | 0,10 | 0,07 | 1,60 | 1,11E-01 | 3,12E-01 |
| TSEN34 | 1953,60 | -0,11 | 0,07 | -1,60 | 1,11E-01 | 3,12E-01 |
| MOV10 | 512,64 | -0,13 | 0,08 | -1,60 | 1,11E-01 | 3,12E-01 |
| HPF1 | 180,77 | 0,10 | 0,06 | 1,60 | 1,11E-01 | 3,12E-01 |
| PKHD1L1 | 242,62 | -0,21 | 0,13 | -1,60 | 1,11E-01 | 3,12E-01 |
| TAF5 | 269,98 | 0,06 | 0,04 | 1,60 | 1,11E-01 | 3,12E-01 |
| RPL3 | 11827,78 | 0,12 | 0,08 | 1,59 | 1,11E-01 | 3,12E-01 |
| FIZ1 | 110,77 | -0,18 | 0,11 | -1,59 | 1,11E-01 | 3,12E-01 |
| FANCB | 217,84 | 0,13 | 0,08 | 1,59 | 1,11E-01 | 3,12E-01 |
| CD5 | 639,80 | -0,14 | 0,09 | -1,59 | 1,11E-01 | 3,12E-01 |
| KCNJ5-AS1 | 30,19 | -0,25 | 0,16 | -1,59 | 1,11E-01 | 3,12E-01 |
| PRKX | 1851,55 | 0,05 | 0,03 | 1,59 | 1,11E-01 | 3,12E-01 |
| ENOSF1 | 935,15 | -0,12 | 0,08 | -1,59 | 1,11E-01 | 3,13E-01 |
| MAP3K12 | 230,91 | -0,13 | 0,08 | -1,59 | 1,11E-01 | 3,13E-01 |
| SEC23A | 2106,70 | 0,07 | 0,04 | 1,59 | 1,11E-01 | 3,13E-01 |
| ATAD2 | 773,32 | 0,11 | 0,07 | 1,59 | 1,11E-01 | 3,13E-01 |
| STX7 | 10163,36 | 0,10 | 0,06 | 1,59 | 1,11E-01 | 3,13E-01 |
| NUP85 | 587,10 | -0,08 | 0,05 | -1,59 | 1,11E-01 | 3,13E-01 |
| CEP89 | 240,31 | -0,14 | 0,09 | -1,59 | 1,11E-01 | 3,13E-01 |
| EOMES | 739,28 | 0,16 | 0,10 | 1,59 | 1,11E-01 | 3,13E-01 |
| MYO19 | 777,44 | -0,09 | 0,06 | -1,59 | 1,11E-01 | 3,13E-01 |
| PDE4DIP | 1048,76 | 0,07 | 0,05 | 1,59 | 1,11E-01 | 3,13E-01 |
| GLT8D1 | 618,13 | 0,05 | 0,03 | 1,59 | 1,12E-01 | 3,13E-01 |
| GKAP1 | 201,19 | 0,10 | 0,06 | 1,59 | 1,11E-01 | 3,13E-01 |
| RBM11 | 57,62 | 0,18 | 0,11 | 1,59 | 1,11E-01 | 3,13E-01 |
| C10orf105 | 149,15 | -0,21 | 0,13 | -1,59 | 1,12E-01 | 3,13E-01 |
| CHCHD5 | 44,65 | -0,15 | 0,09 | -1,59 | 1,12E-01 | 3,13E-01 |
| RAD9A | 174,80 | -0,15 | 0,09 | -1,59 | 1,12E-01 | 3,13E-01 |
| RUFY2 | 701,13 | 0,06 | 0,04 | 1,59 | 1,12E-01 | 3,13E-01 |
| RNF5 | 704,07 | -0,11 | 0,07 | -1,59 | 1,12E-01 | 3,13E-01 |
| HECTD4 | 3700,05 | -0,09 | 0,06 | -1,59 | 1,12E-01 | 3,13E-01 |
| PPP2R2A | 1807,96 | 0,08 | 0,05 | 1,59 | 1,12E-01 | 3,13E-01 |
| HEATR9 | 43,54 | -0,27 | 0,17 | -1,59 | 1,12E-01 | 3,13E-01 |
| PBDC1 | 230,69 | 0,08 | 0,05 | 1,59 | 1,12E-01 | 3,13E-01 |
| PRDM15 | 464,65 | -0,12 | 0,07 | -1,59 | 1,12E-01 | 3,13E-01 |
| CPNE1 | 2209,97 | -0,07 | 0,04 | -1,59 | 1,11E-01 | 3,13E-01 |
| CENPB | 1417,20 | 0,08 | 0,05 | 1,59 | 1,12E-01 | 3,13E-01 |
| TIFA | 734,30 | 0,13 | 0,08 | 1,59 | 1,12E-01 | 3,13E-01 |
| CPLANE1 | 975,96 | 0,09 | 0,05 | 1,59 | 1,12E-01 | 3,13E-01 |
| SP3 | 9184,59 | 0,12 | 0,08 | 1,59 | 1,12E-01 | 3,13E-01 |
| DUT | 1471,79 | 0,10 | 0,06 | 1,59 | 1,12E-01 | 3,13E-01 |

| | | | | | | |
|---|---|---|---|---|---|---|
| VPS29 | 2071,63 | 0,10 | 0,06 | 1,59 | 1,12E-01 | 3,14E-01 |
| ALDOA | 245,01 | -0,09 | 0,06 | -1,59 | 1,12E-01 | 3,14E-01 |
| NFRKB | 893,25 | -0,08 | 0,05 | -1,59 | 1,12E-01 | 3,14E-01 |
| USP30 | 278,22 | -0,06 | 0,04 | -1,59 | 1,12E-01 | 3,14E-01 |
| PMM2 | 719,56 | 0,07 | 0,04 | 1,59 | 1,12E-01 | 3,14E-01 |
| HMGN4 | 1529,78 | 0,08 | 0,05 | 1,59 | 1,13E-01 | 3,14E-01 |
| EP300 | 5333,26 | -0,09 | 0,06 | -1,59 | 1,13E-01 | 3,14E-01 |
| SPIN4 | 170,14 | 0,10 | 0,06 | 1,59 | 1,13E-01 | 3,14E-01 |
| SPRED2 | 65,57 | -0,19 | 0,12 | -1,59 | 1,13E-01 | 3,14E-01 |
| VDR | 315,80 | -0,15 | 0,09 | -1,59 | 1,13E-01 | 3,14E-01 |
| MED16 | 386,74 | -0,18 | 0,11 | -1,59 | 1,12E-01 | 3,14E-01 |
| FPGT | 602,37 | 0,18 | 0,11 | 1,59 | 1,13E-01 | 3,14E-01 |
| PDCD6IP | 4629,98 | 0,08 | 0,05 | 1,59 | 1,13E-01 | 3,14E-01 |
| SPOCK1 | 52,13 | -0,27 | 0,17 | -1,59 | 1,13E-01 | 3,15E-01 |
| KNSTRN | 297,01 | 0,09 | 0,06 | 1,59 | 1,13E-01 | 3,15E-01 |
| ERCC5 | 625,24 | 0,06 | 0,04 | 1,58 | 1,13E-01 | 3,15E-01 |
| CPT1B | 54,89 | -0,20 | 0,12 | -1,58 | 1,13E-01 | 3,15E-01 |
| DYNC2I2 | 96,78 | -0,14 | 0,09 | -1,58 | 1,13E-01 | 3,15E-01 |
| C9orf72 | 7136,02 | 0,14 | 0,09 | 1,59 | 1,13E-01 | 3,15E-01 |
| FLACC1 | 342,61 | -0,14 | 0,09 | -1,59 | 1,13E-01 | 3,15E-01 |
| TOB2 | 326,81 | -0,12 | 0,08 | -1,58 | 1,13E-01 | 3,15E-01 |
| GID4 | 560,77 | -0,13 | 0,08 | -1,58 | 1,13E-01 | 3,15E-01 |
| LEO1 | 484,79 | 0,07 | 0,05 | 1,58 | 1,13E-01 | 3,15E-01 |
| HCCS | 279,21 | 0,07 | 0,05 | 1,58 | 1,13E-01 | 3,15E-01 |
| PMEPA1 | 92,10 | -0,19 | 0,12 | -1,58 | 1,13E-01 | 3,15E-01 |
| CEP126 | 169,28 | 0,12 | 0,08 | 1,58 | 1,13E-01 | 3,15E-01 |
| COA6 | 166,92 | 0,12 | 0,07 | 1,58 | 1,13E-01 | 3,15E-01 |
| STK11IP | 316,88 | -0,14 | 0,09 | -1,58 | 1,13E-01 | 3,15E-01 |
| CR1 | 5709,08 | -0,13 | 0,08 | -1,58 | 1,13E-01 | 3,15E-01 |
| GPD1L | 759,21 | 0,06 | 0,04 | 1,58 | 1,14E-01 | 3,15E-01 |
| LMAN1 | 1705,36 | 0,10 | 0,06 | 1,58 | 1,14E-01 | 3,15E-01 |
| NSMCE2 | 613,13 | 0,08 | 0,05 | 1,58 | 1,13E-01 | 3,15E-01 |
| IL1R1 | 845,36 | -0,18 | 0,12 | -1,58 | 1,14E-01 | 3,15E-01 |
| GET3 | 913,74 | -0,12 | 0,07 | -1,58 | 1,13E-01 | 3,15E-01 |
| STX3 | 4994,70 | -0,12 | 0,07 | -1,58 | 1,14E-01 | 3,15E-01 |
| ADARB2 | 120,22 | -0,52 | 0,33 | -1,58 | 1,13E-01 | 3,15E-01 |
| TRAM2 | 503,44 | -0,10 | 0,06 | -1,58 | 1,14E-01 | 3,15E-01 |
| MFSD12 | 474,22 | -0,15 | 0,10 | -1,58 | 1,14E-01 | 3,15E-01 |
| KBTBD11 | 1007,90 | 0,09 | 0,05 | 1,58 | 1,14E-01 | 3,15E-01 |
| MAP4K3 | 680,11 | 0,09 | 0,06 | 1,58 | 1,14E-01 | 3,15E-01 |
| LRIF1 | 741,54 | 0,10 | 0,07 | 1,58 | 1,14E-01 | 3,15E-01 |
| LRRC37B | 692,65 | -0,08 | 0,05 | -1,58 | 1,14E-01 | 3,15E-01 |
| ISYNA1 | 79,97 | -0,18 | 0,12 | -1,58 | 1,14E-01 | 3,15E-01 |
| TRAM1 | 4164,87 | 0,10 | 0,07 | 1,58 | 1,14E-01 | 3,15E-01 |
| UBE2N | 1822,10 | 0,08 | 0,05 | 1,58 | 1,14E-01 | 3,15E-01 |
| RALA | 435,12 | 0,07 | 0,05 | 1,58 | 1,14E-01 | 3,16E-01 |

| | | | | | | |
|---|---|---|---|---|---|---|
| STXBP3 | 4043,34 | 0,11 | 0,07 | 1,58 | 1,14E-01 | 3,16E-01 |
| NRP2 | 101,32 | -0,29 | 0,19 | -1,58 | 1,14E-01 | 3,16E-01 |
| SPATA33 | 128,38 | -0,10 | 0,06 | -1,58 | 1,14E-01 | 3,16E-01 |
| MYO18A | 1756,80 | -0,12 | 0,08 | -1,58 | 1,14E-01 | 3,16E-01 |
| FHIP1B | 784,92 | -0,12 | 0,07 | -1,58 | 1,14E-01 | 3,16E-01 |
| MPPED2 | 41,91 | -0,37 | 0,24 | -1,58 | 1,14E-01 | 3,16E-01 |
| NOA1 | 475,82 | 0,05 | 0,03 | 1,58 | 1,14E-01 | 3,16E-01 |
| TSPEAR | 36,98 | -0,37 | 0,23 | -1,58 | 1,14E-01 | 3,16E-01 |
| EFL1 | 589,16 | 0,07 | 0,04 | 1,58 | 1,14E-01 | 3,16E-01 |
| DCUN1D5 | 271,80 | 0,12 | 0,07 | 1,58 | 1,14E-01 | 3,16E-01 |
| PHC1 | 286,91 | -0,10 | 0,06 | -1,58 | 1,14E-01 | 3,16E-01 |
| MTF1 | 1225,57 | -0,09 | 0,06 | -1,58 | 1,14E-01 | 3,16E-01 |
| C1orf116 | 46,78 | -0,26 | 0,16 | -1,58 | 1,15E-01 | 3,16E-01 |
| BET1 | 659,89 | 0,12 | 0,08 | 1,58 | 1,15E-01 | 3,16E-01 |
| TIMM10B | 564,11 | 0,06 | 0,04 | 1,58 | 1,15E-01 | 3,16E-01 |
| PADI2 | 1820,26 | -0,23 | 0,14 | -1,58 | 1,15E-01 | 3,16E-01 |
| SHFL | 521,11 | -0,10 | 0,06 | -1,58 | 1,15E-01 | 3,17E-01 |
| TROAP | 194,66 | -0,21 | 0,13 | -1,58 | 1,15E-01 | 3,17E-01 |
| ANAPC4 | 1494,77 | 0,08 | 0,05 | 1,58 | 1,15E-01 | 3,17E-01 |
| RNF122 | 219,28 | -0,12 | 0,08 | -1,58 | 1,15E-01 | 3,17E-01 |
| POLR2D | 750,02 | 0,05 | 0,03 | 1,58 | 1,15E-01 | 3,17E-01 |
| DIPK1A | 275,06 | 0,11 | 0,07 | 1,58 | 1,15E-01 | 3,17E-01 |
| MSRB1 | 3128,84 | -0,12 | 0,08 | -1,58 | 1,15E-01 | 3,17E-01 |
| PSMA6 | 600,86 | 0,10 | 0,06 | 1,58 | 1,15E-01 | 3,17E-01 |
| POM121C | 523,44 | -0,14 | 0,09 | -1,58 | 1,15E-01 | 3,17E-01 |
| AOC3 | 96,28 | -0,24 | 0,15 | -1,58 | 1,15E-01 | 3,17E-01 |
| PITPNM1 | 861,23 | -0,15 | 0,10 | -1,58 | 1,15E-01 | 3,17E-01 |
| PDLIM7 | 564,04 | -0,21 | 0,13 | -1,57 | 1,15E-01 | 3,17E-01 |
| ISCA2 | 536,31 | 0,08 | 0,05 | 1,57 | 1,15E-01 | 3,17E-01 |
| LRFN1 | 120,30 | -0,22 | 0,14 | -1,57 | 1,15E-01 | 3,17E-01 |
| HMG20B | 367,20 | -0,10 | 0,06 | -1,57 | 1,15E-01 | 3,17E-01 |
| MORF4L1 | 4055,21 | 0,07 | 0,04 | 1,57 | 1,15E-01 | 3,17E-01 |
| ICAM4 | 210,23 | -0,16 | 0,10 | -1,57 | 1,16E-01 | 3,17E-01 |
| RPS17 | 295,92 | 0,06 | 0,04 | 1,57 | 1,16E-01 | 3,17E-01 |
| PSENEN | 316,45 | -0,10 | 0,06 | -1,57 | 1,16E-01 | 3,17E-01 |
| SLC35A4 | 242,05 | -0,13 | 0,08 | -1,57 | 1,16E-01 | 3,17E-01 |
| SMN1 | 105,89 | 0,11 | 0,07 | 1,57 | 1,16E-01 | 3,17E-01 |
| MRPL15 | 199,20 | 0,12 | 0,07 | 1,57 | 1,16E-01 | 3,18E-01 |
| TULP3 | 86,04 | -0,11 | 0,07 | -1,57 | 1,16E-01 | 3,18E-01 |
| ZNF460 | 2685,94 | 0,06 | 0,04 | 1,57 | 1,16E-01 | 3,18E-01 |
| WNK1 | 8642,95 | -0,14 | 0,09 | -1,57 | 1,16E-01 | 3,18E-01 |
| HEATR5B | 3623,02 | 0,07 | 0,04 | 1,57 | 1,16E-01 | 3,18E-01 |
| CCDC138 | 207,97 | 0,10 | 0,06 | 1,57 | 1,16E-01 | 3,18E-01 |
| GGT7 | 227,73 | -0,11 | 0,07 | -1,57 | 1,16E-01 | 3,18E-01 |
| EOLA2 | 319,65 | -0,05 | 0,03 | -1,57 | 1,16E-01 | 3,18E-01 |
| WDR35 | 507,31 | 0,10 | 0,06 | 1,57 | 1,16E-01 | 3,18E-01 |

| | | | | | | |
|---|---|---|---|---|---|---|
| TMEM30B | 46,98 | -0,20 | 0,12 | -1,57 | 1,16E-01 | 3,18E-01 |
| DTWD2 | 328,75 | 0,08 | 0,05 | 1,57 | 1,16E-01 | 3,18E-01 |
| YARS2 | 531,65 | 0,07 | 0,05 | 1,57 | 1,16E-01 | 3,18E-01 |
| FLT1 | 138,45 | -0,26 | 0,17 | -1,57 | 1,16E-01 | 3,18E-01 |
| RABGGTA | 95,97 | -0,17 | 0,11 | -1,57 | 1,16E-01 | 3,18E-01 |
| BHLHE40 | 836,29 | -0,10 | 0,06 | -1,57 | 1,16E-01 | 3,18E-01 |
| FAM13B | 2651,30 | 0,07 | 0,04 | 1,57 | 1,16E-01 | 3,18E-01 |
| TUBB6 | 171,45 | -0,17 | 0,11 | -1,57 | 1,16E-01 | 3,18E-01 |
| EFCAB7 | 233,66 | 0,13 | 0,08 | 1,57 | 1,16E-01 | 3,18E-01 |
| ALPK1 | 5773,76 | -0,12 | 0,08 | -1,57 | 1,16E-01 | 3,18E-01 |
| PEX6 | 748,00 | -0,09 | 0,06 | -1,57 | 1,16E-01 | 3,18E-01 |
| PAM | 1634,37 | -0,15 | 0,09 | -1,57 | 1,16E-01 | 3,18E-01 |
| INTS1 | 966,14 | -0,19 | 0,12 | -1,57 | 1,16E-01 | 3,18E-01 |
| LINGO3 | 48,28 | -0,15 | 0,10 | -1,57 | 1,16E-01 | 3,18E-01 |
| MAP1S | 254,02 | -0,15 | 0,09 | -1,57 | 1,17E-01 | 3,19E-01 |
| GUCY1B1 | 230,86 | -0,16 | 0,10 | -1,57 | 1,17E-01 | 3,19E-01 |
| RHOF | 572,20 | -0,14 | 0,09 | -1,57 | 1,17E-01 | 3,19E-01 |
| LSM11 | 328,48 | 0,06 | 0,04 | 1,57 | 1,17E-01 | 3,19E-01 |
| MDM1 | 912,82 | 0,09 | 0,06 | 1,57 | 1,17E-01 | 3,19E-01 |
| HSD17B4 | 2399,58 | 0,09 | 0,06 | 1,57 | 1,17E-01 | 3,19E-01 |
| MAGEH1 | 206,48 | 0,10 | 0,07 | 1,57 | 1,17E-01 | 3,20E-01 |
| RPL15 | 8534,25 | 0,12 | 0,07 | 1,57 | 1,17E-01 | 3,20E-01 |
| GAN | 945,31 | 0,05 | 0,03 | 1,57 | 1,17E-01 | 3,20E-01 |
| RWDD3 | 242,31 | 0,10 | 0,06 | 1,57 | 1,17E-01 | 3,20E-01 |
| PORCN | 48,36 | -0,18 | 0,12 | -1,57 | 1,17E-01 | 3,20E-01 |
| NKRF | 354,67 | 0,07 | 0,04 | 1,57 | 1,17E-01 | 3,20E-01 |
| DTNA | 69,05 | -0,30 | 0,19 | -1,57 | 1,17E-01 | 3,20E-01 |
| BTBD2 | 299,52 | -0,17 | 0,11 | -1,57 | 1,18E-01 | 3,20E-01 |
| DOCK4 | 1364,95 | -0,16 | 0,10 | -1,57 | 1,18E-01 | 3,20E-01 |
| TRAF7 | 432,30 | -0,14 | 0,09 | -1,56 | 1,18E-01 | 3,20E-01 |
| FHL3 | 386,16 | -0,15 | 0,10 | -1,56 | 1,18E-01 | 3,20E-01 |
| RPRD1A | 1819,89 | 0,08 | 0,05 | 1,56 | 1,18E-01 | 3,20E-01 |
| HNRNPD | 3768,88 | 0,06 | 0,04 | 1,56 | 1,18E-01 | 3,21E-01 |
| ANXA5 | 3054,78 | 0,11 | 0,07 | 1,56 | 1,18E-01 | 3,21E-01 |
| GPC2 | 108,52 | -0,15 | 0,09 | -1,56 | 1,18E-01 | 3,21E-01 |
| KISS1R | 28,74 | -0,22 | 0,14 | -1,56 | 1,18E-01 | 3,21E-01 |
| BSCL2 | 104,23 | -0,08 | 0,05 | -1,56 | 1,18E-01 | 3,21E-01 |
| UBE3B | 1769,03 | -0,06 | 0,04 | -1,56 | 1,18E-01 | 3,21E-01 |
| PSTPIP2 | 1454,22 | -0,11 | 0,07 | -1,56 | 1,18E-01 | 3,21E-01 |
| CES4A | 148,28 | -0,12 | 0,07 | -1,56 | 1,18E-01 | 3,21E-01 |
| FBXO33 | 1849,58 | 0,09 | 0,06 | 1,56 | 1,18E-01 | 3,21E-01 |
| STARD8 | 123,23 | -0,16 | 0,10 | -1,56 | 1,18E-01 | 3,21E-01 |
| ZNF257 | 225,37 | 0,16 | 0,10 | 1,56 | 1,18E-01 | 3,21E-01 |
| USP33 | 3633,65 | 0,08 | 0,05 | 1,56 | 1,18E-01 | 3,21E-01 |
| BTG2 | 1618,96 | -0,13 | 0,09 | -1,56 | 1,18E-01 | 3,21E-01 |
| POLR2J2 | 104,15 | -0,21 | 0,13 | -1,56 | 1,19E-01 | 3,21E-01 |

| | | | | | | |
|---|---|---|---|---|---|---|
| DBR1 | 603,49 | 0,08 | 0,05 | 1,56 | 1,19E-01 | 3,21E-01 |
| LRP10 | 3140,59 | -0,18 | 0,12 | -1,56 | 1,19E-01 | 3,22E-01 |
| ZNF75D | 1360,67 | 0,06 | 0,04 | 1,56 | 1,19E-01 | 3,22E-01 |
| FHIP2B | 286,48 | -0,17 | 0,11 | -1,56 | 1,19E-01 | 3,22E-01 |
| BRD7 | 1788,68 | 0,05 | 0,03 | 1,56 | 1,19E-01 | 3,22E-01 |
| LTA4H | 6714,07 | 0,11 | 0,07 | 1,56 | 1,19E-01 | 3,22E-01 |
| METTL2A | 340,95 | 0,06 | 0,04 | 1,56 | 1,19E-01 | 3,22E-01 |
| BMAL2 | 72,01 | -0,21 | 0,14 | -1,56 | 1,19E-01 | 3,22E-01 |
| SIRT6 | 184,74 | -0,09 | 0,06 | -1,56 | 1,19E-01 | 3,22E-01 |
| TEK | 63,16 | -0,32 | 0,20 | -1,56 | 1,19E-01 | 3,22E-01 |
| RAB36 | 63,42 | -0,22 | 0,14 | -1,56 | 1,19E-01 | 3,22E-01 |
| PLCB2 | 3705,70 | -0,15 | 0,10 | -1,56 | 1,19E-01 | 3,22E-01 |
| ZNF749 | 160,41 | 0,10 | 0,06 | 1,56 | 1,19E-01 | 3,22E-01 |
| NACA | 8712,05 | 0,11 | 0,07 | 1,56 | 1,19E-01 | 3,22E-01 |
| RHOG | 671,99 | -0,15 | 0,10 | -1,56 | 1,19E-01 | 3,22E-01 |
| TPST1 | 429,57 | -0,18 | 0,12 | -1,56 | 1,19E-01 | 3,23E-01 |
| NCK1 | 1072,95 | 0,09 | 0,06 | 1,56 | 1,19E-01 | 3,23E-01 |
| ARHGEF19 | 99,84 | -0,16 | 0,10 | -1,56 | 1,19E-01 | 3,23E-01 |
| PUF60 | 1028,85 | 0,05 | 0,03 | 1,56 | 1,19E-01 | 3,23E-01 |
| WDR7 | 2814,88 | 0,07 | 0,04 | 1,56 | 1,20E-01 | 3,23E-01 |
| UPF2 | 2730,86 | 0,08 | 0,05 | 1,56 | 1,20E-01 | 3,23E-01 |
| LGALS9 | 449,86 | -0,18 | 0,11 | -1,56 | 1,20E-01 | 3,23E-01 |
| ERH | 785,00 | 0,17 | 0,11 | 1,56 | 1,20E-01 | 3,23E-01 |
| UBXN8 | 95,18 | 0,12 | 0,08 | 1,56 | 1,20E-01 | 3,23E-01 |
| ME3 | 91,64 | -0,17 | 0,11 | -1,56 | 1,20E-01 | 3,23E-01 |
| ZNF280D | 934,23 | 0,13 | 0,08 | 1,56 | 1,20E-01 | 3,23E-01 |
| HACE1 | 722,28 | 0,08 | 0,05 | 1,56 | 1,20E-01 | 3,23E-01 |
| B3GNT8 | 463,64 | -0,13 | 0,08 | -1,55 | 1,20E-01 | 3,24E-01 |
| TBC1D4 | 1780,54 | 0,11 | 0,07 | 1,55 | 1,20E-01 | 3,24E-01 |
| AUH | 802,34 | 0,06 | 0,04 | 1,55 | 1,20E-01 | 3,24E-01 |
| C5orf15 | 854,18 | 0,09 | 0,06 | 1,55 | 1,20E-01 | 3,24E-01 |
| COL24A1 | 90,42 | -0,25 | 0,16 | -1,55 | 1,20E-01 | 3,24E-01 |
| IQSEC2 | 143,20 | -0,20 | 0,13 | -1,55 | 1,20E-01 | 3,24E-01 |
| PHF2 | 870,40 | -0,12 | 0,07 | -1,55 | 1,20E-01 | 3,24E-01 |
| IL20RB | 55,72 | -0,16 | 0,10 | -1,55 | 1,21E-01 | 3,24E-01 |
| ENSG000002 | 421,58 | 0,23 | 0,15 | 1,55 | 1,21E-01 | 3,24E-01 |
| TMEM39A | 1099,21 | 0,05 | 0,03 | 1,55 | 1,21E-01 | 3,24E-01 |
| RNF113A | 429,10 | 0,08 | 0,05 | 1,55 | 1,21E-01 | 3,24E-01 |
| NUDT8 | 25,22 | -0,19 | 0,12 | -1,55 | 1,21E-01 | 3,24E-01 |
| TUSC2 | 313,73 | -0,07 | 0,05 | -1,55 | 1,21E-01 | 3,24E-01 |
| SLC12A6 | 10002,54 | -0,09 | 0,06 | -1,55 | 1,21E-01 | 3,24E-01 |
| ADISSP | 158,11 | -0,15 | 0,10 | -1,55 | 1,21E-01 | 3,25E-01 |
| VPS9D1 | 452,95 | -0,15 | 0,09 | -1,55 | 1,21E-01 | 3,25E-01 |
| ENSG000002 | 230,99 | -0,11 | 0,07 | -1,55 | 1,21E-01 | 3,25E-01 |
| ERP29 | 740,14 | 0,08 | 0,05 | 1,55 | 1,21E-01 | 3,25E-01 |
| INO80 | 1462,45 | 0,05 | 0,03 | 1,55 | 1,21E-01 | 3,25E-01 |

| | | | | | | |
|---|---|---|---|---|---|---|
| PTGR2 | 89,88 | -0,14 | 0,09 | -1,55 | 1,21E-01 | 3,25E-01 |
| LPAR6 | 1205,57 | 0,24 | 0,15 | 1,55 | 1,21E-01 | 3,25E-01 |
| MLC1 | 183,77 | -0,16 | 0,10 | -1,55 | 1,21E-01 | 3,25E-01 |
| EHD3 | 298,13 | -0,15 | 0,10 | -1,55 | 1,21E-01 | 3,26E-01 |
| BCLAF3 | 1139,11 | 0,07 | 0,05 | 1,55 | 1,22E-01 | 3,26E-01 |
| ADH5 | 1213,60 | 0,10 | 0,06 | 1,55 | 1,22E-01 | 3,26E-01 |
| CDAN1 | 320,11 | -0,13 | 0,09 | -1,55 | 1,22E-01 | 3,26E-01 |
| PACS2 | 454,38 | -0,19 | 0,12 | -1,55 | 1,22E-01 | 3,26E-01 |
| RORC | 104,73 | -0,21 | 0,13 | -1,55 | 1,22E-01 | 3,26E-01 |
| TTC22 | 318,49 | -0,12 | 0,08 | -1,55 | 1,22E-01 | 3,26E-01 |
| CD1E | 34,86 | -0,25 | 0,16 | -1,55 | 1,22E-01 | 3,26E-01 |
| PDCL | 716,37 | 0,07 | 0,05 | 1,55 | 1,22E-01 | 3,26E-01 |
| GK5 | 1807,46 | 0,09 | 0,06 | 1,55 | 1,22E-01 | 3,26E-01 |
| VWF | 144,04 | -0,31 | 0,20 | -1,55 | 1,22E-01 | 3,26E-01 |
| PLA2G4C | 92,61 | -0,30 | 0,20 | -1,55 | 1,22E-01 | 3,26E-01 |
| TAX1BP1 | 4380,58 | 0,13 | 0,08 | 1,55 | 1,22E-01 | 3,26E-01 |
| TMEM208 | 401,09 | 0,07 | 0,05 | 1,55 | 1,22E-01 | 3,26E-01 |
| ASF1B | 63,73 | -0,20 | 0,13 | -1,55 | 1,22E-01 | 3,26E-01 |
| THOC7 | 309,71 | 0,10 | 0,07 | 1,55 | 1,22E-01 | 3,26E-01 |
| SLC25A16 | 895,65 | 0,04 | 0,02 | 1,55 | 1,22E-01 | 3,27E-01 |
| TNPO2 | 476,13 | -0,12 | 0,08 | -1,55 | 1,22E-01 | 3,27E-01 |
| STARD3 | 710,22 | -0,11 | 0,07 | -1,55 | 1,22E-01 | 3,27E-01 |
| CACFD1 | 111,92 | -0,13 | 0,08 | -1,55 | 1,22E-01 | 3,27E-01 |
| GPI | 654,67 | -0,13 | 0,09 | -1,54 | 1,22E-01 | 3,27E-01 |
| DUSP2 | 307,67 | 0,16 | 0,10 | 1,54 | 1,22E-01 | 3,27E-01 |
| SLC30A4 | 265,00 | 0,10 | 0,06 | 1,54 | 1,23E-01 | 3,27E-01 |
| MAST3 | 2414,57 | -0,17 | 0,11 | -1,54 | 1,22E-01 | 3,27E-01 |
| BCL2L1 | 13853,53 | -0,22 | 0,14 | -1,54 | 1,22E-01 | 3,27E-01 |
| DIS3L | 805,46 | 0,05 | 0,03 | 1,54 | 1,23E-01 | 3,27E-01 |
| NCAPG2 | 459,95 | -0,15 | 0,10 | -1,54 | 1,23E-01 | 3,27E-01 |
| NLRP3 | 1399,19 | 0,10 | 0,07 | 1,54 | 1,22E-01 | 3,27E-01 |
| DNLZ | 31,09 | -0,20 | 0,13 | -1,54 | 1,23E-01 | 3,27E-01 |
| SPDYA | 155,80 | 0,11 | 0,07 | 1,54 | 1,23E-01 | 3,27E-01 |
| ANXA2 | 5242,31 | 0,08 | 0,05 | 1,54 | 1,23E-01 | 3,27E-01 |
| COX7C | 1076,83 | 0,21 | 0,13 | 1,54 | 1,23E-01 | 3,27E-01 |
| NCOA7 | 1419,36 | 0,07 | 0,05 | 1,54 | 1,23E-01 | 3,27E-01 |
| DLG5 | 244,17 | -0,21 | 0,14 | -1,54 | 1,23E-01 | 3,28E-01 |
| CDKN2AIP | 1005,59 | 0,11 | 0,07 | 1,54 | 1,23E-01 | 3,28E-01 |
| REM2 | 254,72 | -0,14 | 0,09 | -1,54 | 1,23E-01 | 3,28E-01 |
| FBXO6 | 273,13 | -0,12 | 0,08 | -1,54 | 1,23E-01 | 3,28E-01 |
| SEMA3A | 38,57 | -0,26 | 0,17 | -1,54 | 1,23E-01 | 3,28E-01 |
| ITGB3BP | 367,82 | 0,14 | 0,09 | 1,54 | 1,23E-01 | 3,28E-01 |
| FCRL3 | 1512,45 | 0,15 | 0,10 | 1,54 | 1,23E-01 | 3,28E-01 |
| ABHD11 | 127,23 | -0,10 | 0,06 | -1,54 | 1,23E-01 | 3,28E-01 |
| ZNF584 | 136,93 | -0,07 | 0,05 | -1,54 | 1,23E-01 | 3,28E-01 |
| NADK | 5615,92 | -0,13 | 0,08 | -1,54 | 1,23E-01 | 3,28E-01 |

| | | | | | | |
|---|---|---|---|---|---|---|
| USP4 | 4160,95 | -0,08 | 0,05 | -1,54 | 1,23E-01 | 3,28E-01 |
| STAT3 | 6518,73 | -0,11 | 0,07 | -1,54 | 1,23E-01 | 3,28E-01 |
| COX16 | 67,34 | 0,08 | 0,05 | 1,54 | 1,23E-01 | 3,28E-01 |
| FBXO41 | 244,24 | -0,20 | 0,13 | -1,54 | 1,24E-01 | 3,28E-01 |
| REV3L | 2497,21 | 0,07 | 0,04 | 1,54 | 1,24E-01 | 3,28E-01 |
| SEMA4B | 1117,26 | -0,14 | 0,09 | -1,54 | 1,23E-01 | 3,28E-01 |
| LRRK1 | 995,55 | -0,13 | 0,08 | -1,54 | 1,24E-01 | 3,28E-01 |
| METRN | 77,70 | -0,16 | 0,10 | -1,54 | 1,24E-01 | 3,28E-01 |
| PRPF31 | 787,32 | 0,07 | 0,04 | 1,54 | 1,24E-01 | 3,28E-01 |
| MTG1 | 206,36 | -0,11 | 0,07 | -1,54 | 1,24E-01 | 3,28E-01 |
| CAMK2G | 2610,98 | -0,11 | 0,07 | -1,54 | 1,24E-01 | 3,28E-01 |
| ARRB2 | 4057,13 | -0,13 | 0,09 | -1,54 | 1,24E-01 | 3,28E-01 |
| ZNF446 | 127,59 | -0,12 | 0,08 | -1,54 | 1,24E-01 | 3,28E-01 |
| C8orf58 | 88,18 | -0,11 | 0,07 | -1,54 | 1,24E-01 | 3,28E-01 |
| IDS | 3829,88 | -0,10 | 0,06 | -1,54 | 1,24E-01 | 3,28E-01 |
| PTGER4 | 2042,13 | 0,07 | 0,04 | 1,54 | 1,24E-01 | 3,29E-01 |
| CEACAM3 | 949,92 | -0,15 | 0,10 | -1,54 | 1,24E-01 | 3,29E-01 |
| CLK4 | 3779,22 | 0,11 | 0,07 | 1,54 | 1,24E-01 | 3,29E-01 |
| ISG20 | 2628,54 | -0,09 | 0,06 | -1,54 | 1,24E-01 | 3,29E-01 |
| CDK2 | 206,58 | -0,06 | 0,04 | -1,54 | 1,24E-01 | 3,29E-01 |
| ATP5IF1 | 354,06 | 0,10 | 0,07 | 1,54 | 1,24E-01 | 3,29E-01 |
| NSD2 | 1482,85 | -0,07 | 0,05 | -1,54 | 1,24E-01 | 3,29E-01 |
| SIRPB2 | 2326,23 | -0,16 | 0,10 | -1,54 | 1,24E-01 | 3,29E-01 |
| CD200 | 157,00 | 0,20 | 0,13 | 1,54 | 1,25E-01 | 3,29E-01 |
| TAS2R43 | 88,70 | -0,33 | 0,21 | -1,54 | 1,25E-01 | 3,29E-01 |
| LRRC8D | 1171,09 | 0,06 | 0,04 | 1,54 | 1,25E-01 | 3,29E-01 |
| CROCC | 678,59 | -0,14 | 0,09 | -1,54 | 1,24E-01 | 3,29E-01 |
| SLC38A2 | 6629,21 | 0,12 | 0,08 | 1,54 | 1,24E-01 | 3,29E-01 |
| C17orf100 | 25,42 | -0,20 | 0,13 | -1,54 | 1,25E-01 | 3,29E-01 |
| INSIG1 | 1229,34 | 0,07 | 0,05 | 1,54 | 1,25E-01 | 3,29E-01 |
| MLLT3 | 1171,63 | 0,10 | 0,06 | 1,54 | 1,25E-01 | 3,29E-01 |
| MBTD1 | 2133,76 | 0,07 | 0,04 | 1,54 | 1,25E-01 | 3,29E-01 |
| GNAL | 92,73 | -0,22 | 0,14 | -1,54 | 1,24E-01 | 3,29E-01 |
| ELP5 | 260,16 | 0,05 | 0,04 | 1,54 | 1,25E-01 | 3,29E-01 |
| SHC1 | 664,04 | -0,10 | 0,07 | -1,54 | 1,25E-01 | 3,29E-01 |
| NSUN7 | 324,20 | -0,14 | 0,09 | -1,54 | 1,25E-01 | 3,29E-01 |
| FNTB | 196,87 | -0,07 | 0,05 | -1,53 | 1,25E-01 | 3,29E-01 |
| RBM15 | 2399,92 | 0,06 | 0,04 | 1,53 | 1,25E-01 | 3,30E-01 |
| ZNF717 | 319,06 | 0,08 | 0,05 | 1,53 | 1,25E-01 | 3,30E-01 |
| LYNX1 | 69,66 | -0,23 | 0,15 | -1,53 | 1,25E-01 | 3,30E-01 |
| ZYX | 1319,16 | -0,21 | 0,14 | -1,53 | 1,25E-01 | 3,30E-01 |
| TIPARP | 1774,90 | 0,08 | 0,05 | 1,53 | 1,25E-01 | 3,30E-01 |
| MTM1 | 1192,30 | 0,07 | 0,05 | 1,53 | 1,25E-01 | 3,30E-01 |
| MYBPC3 | 202,57 | -0,18 | 0,12 | -1,53 | 1,25E-01 | 3,30E-01 |
| SEH1L | 665,02 | 0,05 | 0,03 | 1,53 | 1,25E-01 | 3,30E-01 |
| DCHS1 | 128,51 | -0,26 | 0,17 | -1,53 | 1,25E-01 | 3,30E-01 |

| | | | | | | |
|---|---|---|---|---|---|---|
| DIXDC1 | 112,04 | -0,19 | 0,12 | -1,53 | 1,25E-01 | 3,30E-01 |
| FDXACB1 | 51,04 | 0,08 | 0,06 | 1,53 | 1,25E-01 | 3,30E-01 |
| MACROH2A2 | 61,52 | -0,11 | 0,07 | -1,53 | 1,26E-01 | 3,30E-01 |
| TMEM134 | 174,94 | -0,12 | 0,08 | -1,53 | 1,26E-01 | 3,30E-01 |
| STK36 | 237,22 | -0,10 | 0,06 | -1,53 | 1,26E-01 | 3,31E-01 |
| ZNF496 | 211,91 | -0,17 | 0,11 | -1,53 | 1,26E-01 | 3,31E-01 |
| CACNB1 | 152,21 | -0,15 | 0,10 | -1,53 | 1,26E-01 | 3,31E-01 |
| FITM1 | 26,75 | -0,13 | 0,08 | -1,53 | 1,26E-01 | 3,31E-01 |
| PPA2 | 1166,41 | 0,08 | 0,05 | 1,53 | 1,26E-01 | 3,31E-01 |
| RERE | 2545,05 | -0,11 | 0,07 | -1,53 | 1,26E-01 | 3,31E-01 |
| ZDHHC14 | 175,01 | -0,16 | 0,11 | -1,53 | 1,26E-01 | 3,31E-01 |
| PMAIP1 | 437,49 | 0,17 | 0,11 | 1,53 | 1,26E-01 | 3,31E-01 |
| SIGMAR1 | 320,65 | -0,06 | 0,04 | -1,53 | 1,26E-01 | 3,31E-01 |
| KLC2 | 171,70 | -0,15 | 0,10 | -1,53 | 1,26E-01 | 3,31E-01 |
| NKTR | 13227,33 | 0,08 | 0,05 | 1,53 | 1,26E-01 | 3,31E-01 |
| LIPT1 | 134,92 | 0,09 | 0,06 | 1,53 | 1,26E-01 | 3,31E-01 |
| MICAL1 | 1107,88 | -0,18 | 0,12 | -1,53 | 1,26E-01 | 3,31E-01 |
| STX17 | 1323,77 | 0,07 | 0,04 | 1,53 | 1,26E-01 | 3,31E-01 |
| CCNJ | 457,73 | 0,06 | 0,04 | 1,53 | 1,26E-01 | 3,31E-01 |
| LAMA2 | 120,82 | -0,20 | 0,13 | -1,53 | 1,26E-01 | 3,31E-01 |
| BTN3A2 | 4101,09 | -0,12 | 0,08 | -1,53 | 1,26E-01 | 3,31E-01 |
| POLN | 77,30 | -0,19 | 0,13 | -1,53 | 1,26E-01 | 3,31E-01 |
| NASP | 1595,61 | 0,06 | 0,04 | 1,53 | 1,26E-01 | 3,31E-01 |
| HNRNPH2 | 2173,63 | 0,13 | 0,08 | 1,53 | 1,26E-01 | 3,31E-01 |
| UCN | 33,28 | -0,14 | 0,09 | -1,53 | 1,26E-01 | 3,31E-01 |
| ITGA2B | 723,24 | -0,28 | 0,18 | -1,53 | 1,27E-01 | 3,31E-01 |
| NFKB2 | 1249,93 | -0,12 | 0,08 | -1,53 | 1,27E-01 | 3,31E-01 |
| PRPS1 | 724,96 | 0,08 | 0,05 | 1,53 | 1,27E-01 | 3,31E-01 |
| ANKRD50 | 819,03 | 0,11 | 0,07 | 1,53 | 1,27E-01 | 3,31E-01 |
| IMPA2 | 1046,47 | -0,11 | 0,07 | -1,53 | 1,27E-01 | 3,31E-01 |
| CEP20 | 482,14 | 0,11 | 0,07 | 1,53 | 1,27E-01 | 3,32E-01 |
| NDUFA12 | 618,12 | 0,10 | 0,07 | 1,53 | 1,27E-01 | 3,32E-01 |
| CLEC17A | 189,61 | 0,17 | 0,11 | 1,53 | 1,27E-01 | 3,32E-01 |
| BPTF | 5323,42 | 0,06 | 0,04 | 1,53 | 1,27E-01 | 3,32E-01 |
| ADAMTS4 | 59,64 | -0,23 | 0,15 | -1,53 | 1,27E-01 | 3,32E-01 |
| CENPN | 78,35 | -0,12 | 0,08 | -1,53 | 1,27E-01 | 3,32E-01 |
| ENSG000002 | 20,54 | -0,15 | 0,10 | -1,53 | 1,27E-01 | 3,32E-01 |
| UCP2 | 1487,67 | -0,18 | 0,12 | -1,52 | 1,27E-01 | 3,32E-01 |
| ZRANB2 | 4096,72 | 0,14 | 0,09 | 1,53 | 1,27E-01 | 3,32E-01 |
| ERRFI1 | 35,45 | -0,19 | 0,12 | -1,52 | 1,27E-01 | 3,32E-01 |
| TMEM200A | 57,36 | -0,21 | 0,14 | -1,52 | 1,27E-01 | 3,32E-01 |
| TXNRD2 | 718,07 | -0,11 | 0,07 | -1,52 | 1,27E-01 | 3,32E-01 |
| SLC7A6OS | 1251,29 | 0,04 | 0,02 | 1,53 | 1,27E-01 | 3,32E-01 |
| ZNF546 | 382,17 | 0,08 | 0,05 | 1,52 | 1,27E-01 | 3,32E-01 |
| PSMD5 | 882,74 | 0,05 | 0,03 | 1,53 | 1,27E-01 | 3,32E-01 |
| ADAM9 | 917,41 | 0,10 | 0,06 | 1,53 | 1,27E-01 | 3,32E-01 |

| | | | | | | |
|---|---|---|---|---|---|---|
| CTTN | 491,98 | -0,21 | 0,14 | -1,53 | 1,27E-01 | 3,32E-01 |
| TRABD | 637,87 | -0,15 | 0,10 | -1,52 | 1,27E-01 | 3,32E-01 |
| ZNF699 | 456,57 | 0,08 | 0,05 | 1,53 | 1,27E-01 | 3,32E-01 |
| RAPGEF5 | 69,91 | -0,30 | 0,20 | -1,52 | 1,27E-01 | 3,32E-01 |
| PLEKHM1 | 1274,99 | -0,15 | 0,10 | -1,52 | 1,27E-01 | 3,32E-01 |
| CDKN2A | 135,53 | -0,13 | 0,09 | -1,52 | 1,27E-01 | 3,32E-01 |
| SPTLC1 | 1910,29 | 0,06 | 0,04 | 1,52 | 1,27E-01 | 3,32E-01 |
| FBXO21 | 1115,02 | 0,06 | 0,04 | 1,52 | 1,28E-01 | 3,32E-01 |
| PIGM | 606,30 | 0,06 | 0,04 | 1,52 | 1,28E-01 | 3,32E-01 |
| MAP4K2 | 772,25 | -0,13 | 0,08 | -1,52 | 1,28E-01 | 3,32E-01 |
| TSHZ3 | 317,21 | -0,12 | 0,08 | -1,52 | 1,28E-01 | 3,32E-01 |
| UBE3C | 3061,51 | 0,04 | 0,03 | 1,52 | 1,28E-01 | 3,33E-01 |
| SLC12A9 | 3383,32 | -0,13 | 0,08 | -1,52 | 1,28E-01 | 3,33E-01 |
| GNPNAT1 | 292,54 | 0,08 | 0,05 | 1,52 | 1,28E-01 | 3,33E-01 |
| TNFSF4 | 325,05 | -0,12 | 0,08 | -1,52 | 1,28E-01 | 3,33E-01 |
| WDR46 | 645,37 | 0,06 | 0,04 | 1,52 | 1,28E-01 | 3,33E-01 |
| ITIH4 | 28,24 | -0,19 | 0,13 | -1,52 | 1,28E-01 | 3,33E-01 |
| ZFAND6 | 3163,71 | 0,09 | 0,06 | 1,52 | 1,28E-01 | 3,33E-01 |
| PRKCD | 1268,28 | -0,17 | 0,11 | -1,52 | 1,28E-01 | 3,33E-01 |
| LIMK2 | 4650,16 | -0,14 | 0,09 | -1,52 | 1,28E-01 | 3,33E-01 |
| FBLN7 | 151,18 | -0,15 | 0,10 | -1,52 | 1,28E-01 | 3,33E-01 |
| CCSER1 | 252,37 | 0,13 | 0,09 | 1,52 | 1,29E-01 | 3,33E-01 |
| CYTIP | 9468,44 | 0,08 | 0,05 | 1,52 | 1,29E-01 | 3,34E-01 |
| RNF126 | 483,91 | -0,11 | 0,07 | -1,52 | 1,29E-01 | 3,34E-01 |
| C12orf76 | 483,41 | -0,07 | 0,04 | -1,52 | 1,29E-01 | 3,34E-01 |
| FAM171B | 29,06 | -0,22 | 0,14 | -1,52 | 1,29E-01 | 3,34E-01 |
| OXLD1 | 762,93 | 0,08 | 0,05 | 1,52 | 1,29E-01 | 3,34E-01 |
| LAMTOR3 | 1042,14 | 0,13 | 0,09 | 1,52 | 1,29E-01 | 3,34E-01 |
| PRAG1 | 150,89 | -0,17 | 0,11 | -1,52 | 1,29E-01 | 3,34E-01 |
| EPHB4 | 231,03 | -0,19 | 0,13 | -1,52 | 1,29E-01 | 3,34E-01 |
| RUNDC1 | 527,18 | -0,08 | 0,05 | -1,52 | 1,29E-01 | 3,34E-01 |
| ENPP5 | 121,91 | 0,16 | 0,11 | 1,52 | 1,29E-01 | 3,34E-01 |
| STK39 | 879,87 | 0,09 | 0,06 | 1,52 | 1,29E-01 | 3,34E-01 |
| CLK1 | 7013,67 | 0,11 | 0,07 | 1,52 | 1,29E-01 | 3,34E-01 |
| PPAN | 82,98 | 0,08 | 0,05 | 1,52 | 1,29E-01 | 3,34E-01 |
| RABL2B | 232,92 | -0,13 | 0,09 | -1,52 | 1,29E-01 | 3,34E-01 |
| PRR12 | 254,55 | -0,29 | 0,19 | -1,52 | 1,29E-01 | 3,34E-01 |
| MTMR3 | 5962,28 | -0,11 | 0,07 | -1,52 | 1,29E-01 | 3,34E-01 |
| ALG6 | 1039,36 | 0,08 | 0,05 | 1,52 | 1,29E-01 | 3,34E-01 |
| PIGV | 469,00 | 0,05 | 0,03 | 1,52 | 1,29E-01 | 3,34E-01 |
| SCRIB | 325,62 | -0,18 | 0,12 | -1,52 | 1,29E-01 | 3,34E-01 |
| NEK7 | 5408,92 | 0,12 | 0,08 | 1,52 | 1,29E-01 | 3,34E-01 |
| C2CD5 | 2525,45 | 0,08 | 0,05 | 1,52 | 1,30E-01 | 3,35E-01 |
| CD24 | 615,20 | -0,21 | 0,14 | -1,52 | 1,30E-01 | 3,35E-01 |
| ANXA6 | 7119,50 | 0,08 | 0,05 | 1,52 | 1,30E-01 | 3,35E-01 |
| KIF21A | 539,32 | 0,13 | 0,09 | 1,52 | 1,30E-01 | 3,35E-01 |

| | | | | | | |
|---|---|---|---|---|---|---|
| ZNF213 | 224,15 | -0,13 | 0,08 | -1,52 | 1,30E-01 | 3,35E-01 |
| ELK1 | 253,97 | -0,08 | 0,05 | -1,52 | 1,30E-01 | 3,35E-01 |
| ATXN7L3 | 1794,27 | -0,07 | 0,05 | -1,52 | 1,30E-01 | 3,35E-01 |
| GPSM2 | 378,67 | -0,10 | 0,07 | -1,52 | 1,30E-01 | 3,35E-01 |
| CASP3 | 1109,42 | 0,14 | 0,09 | 1,52 | 1,30E-01 | 3,35E-01 |
| ZNF205 | 28,75 | -0,26 | 0,17 | -1,52 | 1,30E-01 | 3,35E-01 |
| MON1B | 1123,92 | -0,07 | 0,05 | -1,51 | 1,30E-01 | 3,35E-01 |
| DCUN1D4 | 858,76 | 0,09 | 0,06 | 1,51 | 1,30E-01 | 3,35E-01 |
| GSTO2 | 75,15 | -0,19 | 0,12 | -1,51 | 1,30E-01 | 3,35E-01 |
| GPSM1 | 109,14 | -0,20 | 0,13 | -1,51 | 1,30E-01 | 3,35E-01 |
| TRDMT1 | 1135,27 | 0,07 | 0,05 | 1,51 | 1,30E-01 | 3,35E-01 |
| PHLPP1 | 527,14 | -0,09 | 0,06 | -1,51 | 1,30E-01 | 3,35E-01 |
| TMCO6 | 407,35 | -0,08 | 0,05 | -1,51 | 1,30E-01 | 3,35E-01 |
| ATL3 | 2063,65 | 0,06 | 0,04 | 1,51 | 1,30E-01 | 3,35E-01 |
| ACVR1 | 431,30 | -0,07 | 0,05 | -1,51 | 1,30E-01 | 3,35E-01 |
| ADAR | 11790,86 | -0,12 | 0,08 | -1,51 | 1,30E-01 | 3,35E-01 |
| EDF1 | 2173,86 | 0,07 | 0,05 | 1,51 | 1,30E-01 | 3,35E-01 |
| LGALS3BP | 95,88 | -0,19 | 0,12 | -1,51 | 1,30E-01 | 3,35E-01 |
| FILIP1L | 42,61 | -0,32 | 0,21 | -1,51 | 1,30E-01 | 3,35E-01 |
| SGPL1 | 1461,63 | -0,07 | 0,04 | -1,51 | 1,31E-01 | 3,35E-01 |
| UEVLD | 706,16 | 0,07 | 0,05 | 1,51 | 1,30E-01 | 3,35E-01 |
| PLCH2 | 234,97 | -0,22 | 0,14 | -1,51 | 1,31E-01 | 3,36E-01 |
| PFDN5 | 2137,88 | 0,20 | 0,13 | 1,51 | 1,31E-01 | 3,36E-01 |
| RARA | 1383,80 | -0,15 | 0,10 | -1,51 | 1,31E-01 | 3,36E-01 |
| TNFRSF21 | 43,94 | -0,22 | 0,15 | -1,51 | 1,31E-01 | 3,36E-01 |
| LRWD1 | 377,15 | -0,14 | 0,09 | -1,51 | 1,31E-01 | 3,36E-01 |
| ZBTB17 | 588,49 | -0,06 | 0,04 | -1,51 | 1,31E-01 | 3,36E-01 |
| SLK | 4007,17 | 0,11 | 0,07 | 1,51 | 1,31E-01 | 3,36E-01 |
| CALM2 | 10139,59 | 0,12 | 0,08 | 1,51 | 1,31E-01 | 3,36E-01 |
| PGAP6 | 621,37 | -0,15 | 0,10 | -1,51 | 1,31E-01 | 3,36E-01 |
| MAGI3 | 166,51 | -0,13 | 0,08 | -1,51 | 1,31E-01 | 3,36E-01 |
| TMEM11 | 234,91 | -0,07 | 0,05 | -1,51 | 1,31E-01 | 3,36E-01 |
| TRAPPC2 | 1014,21 | 0,06 | 0,04 | 1,51 | 1,31E-01 | 3,36E-01 |
| MRPL58 | 169,99 | 0,07 | 0,05 | 1,51 | 1,31E-01 | 3,36E-01 |
| KLHDC8B | 231,31 | -0,15 | 0,10 | -1,51 | 1,31E-01 | 3,36E-01 |
| CHMP5 | 718,64 | 0,15 | 0,10 | 1,51 | 1,31E-01 | 3,36E-01 |
| NIPSNAP3A | 407,29 | 0,14 | 0,09 | 1,51 | 1,31E-01 | 3,36E-01 |
| UVRAG | 2132,60 | 0,06 | 0,04 | 1,51 | 1,31E-01 | 3,36E-01 |
| ADGRE5 | 8704,76 | -0,13 | 0,09 | -1,51 | 1,31E-01 | 3,36E-01 |
| TRMT1L | 744,64 | 0,10 | 0,06 | 1,51 | 1,31E-01 | 3,36E-01 |
| ZER1 | 1656,49 | -0,20 | 0,13 | -1,51 | 1,31E-01 | 3,36E-01 |
| ZNF600 | 703,33 | 0,14 | 0,09 | 1,51 | 1,31E-01 | 3,36E-01 |
| MAML3 | 634,24 | -0,12 | 0,08 | -1,51 | 1,31E-01 | 3,36E-01 |
| PATL1 | 982,54 | -0,09 | 0,06 | -1,51 | 1,32E-01 | 3,36E-01 |
| SNX11 | 1142,07 | -0,09 | 0,06 | -1,51 | 1,31E-01 | 3,36E-01 |
| PHF12 | 1914,52 | -0,12 | 0,08 | -1,51 | 1,31E-01 | 3,36E-01 |

| | | | | | | |
|---|---|---|---|---|---|---|
| FRYL | 7605,16 | 0,07 | 0,04 | 1,51 | 1,32E-01 | 3,36E-01 |
| SETD1A | 159,86 | -0,21 | 0,14 | -1,51 | 1,32E-01 | 3,36E-01 |
| POGLUT1 | 1013,20 | 0,06 | 0,04 | 1,51 | 1,32E-01 | 3,36E-01 |
| NDOR1 | 242,93 | -0,13 | 0,09 | -1,51 | 1,32E-01 | 3,36E-01 |
| RPP38 | 327,44 | 0,06 | 0,04 | 1,51 | 1,32E-01 | 3,36E-01 |
| RAB40C | 323,81 | -0,11 | 0,07 | -1,51 | 1,32E-01 | 3,36E-01 |
| EIF4EBP2 | 3948,01 | -0,08 | 0,06 | -1,51 | 1,32E-01 | 3,37E-01 |
| DOLPP1 | 159,06 | -0,06 | 0,04 | -1,51 | 1,32E-01 | 3,37E-01 |
| CEP192 | 3973,44 | 0,07 | 0,04 | 1,51 | 1,32E-01 | 3,37E-01 |
| SCAPER | 1522,55 | 0,08 | 0,06 | 1,51 | 1,32E-01 | 3,37E-01 |
| PTK2B | 4158,52 | -0,13 | 0,09 | -1,51 | 1,32E-01 | 3,37E-01 |
| MRPL20 | 654,47 | 0,06 | 0,04 | 1,51 | 1,32E-01 | 3,37E-01 |
| PEX7 | 198,04 | 0,07 | 0,05 | 1,51 | 1,32E-01 | 3,37E-01 |
| GRAMD1C | 592,99 | 0,14 | 0,09 | 1,51 | 1,32E-01 | 3,37E-01 |
| UBALD2 | 938,27 | -0,12 | 0,08 | -1,51 | 1,32E-01 | 3,37E-01 |
| IL10RB | 331,18 | -0,10 | 0,07 | -1,50 | 1,33E-01 | 3,38E-01 |
| TUBB2A | 840,57 | -0,46 | 0,31 | -1,50 | 1,33E-01 | 3,38E-01 |
| MAP3K15 | 39,22 | -0,27 | 0,18 | -1,50 | 1,33E-01 | 3,38E-01 |
| CSF2RB | 7335,74 | -0,13 | 0,08 | -1,50 | 1,33E-01 | 3,38E-01 |
| RAPGEFL1 | 123,18 | -0,21 | 0,14 | -1,50 | 1,33E-01 | 3,38E-01 |
| DUS3L | 388,35 | 0,07 | 0,04 | 1,50 | 1,33E-01 | 3,38E-01 |
| CATSPERG | 266,51 | -0,13 | 0,09 | -1,50 | 1,33E-01 | 3,38E-01 |
| SMIM13 | 318,10 | 0,10 | 0,06 | 1,50 | 1,33E-01 | 3,38E-01 |
| CEP72 | 219,25 | -0,12 | 0,08 | -1,50 | 1,33E-01 | 3,38E-01 |
| ITPR1 | 6186,78 | 0,08 | 0,05 | 1,50 | 1,33E-01 | 3,38E-01 |
| ZMYM3 | 243,30 | -0,13 | 0,09 | -1,50 | 1,33E-01 | 3,38E-01 |
| MTRF1 | 505,62 | 0,07 | 0,04 | 1,50 | 1,33E-01 | 3,38E-01 |
| REL | 2747,15 | 0,06 | 0,04 | 1,50 | 1,33E-01 | 3,39E-01 |
| CHFR | 2544,11 | -0,08 | 0,05 | -1,50 | 1,33E-01 | 3,39E-01 |
| ELP3 | 1150,25 | 0,06 | 0,04 | 1,50 | 1,33E-01 | 3,39E-01 |
| SNX16 | 461,37 | 0,15 | 0,10 | 1,50 | 1,33E-01 | 3,39E-01 |
| STX18 | 647,00 | 0,04 | 0,02 | 1,50 | 1,33E-01 | 3,39E-01 |
| NAPEPLD | 537,29 | 0,06 | 0,04 | 1,50 | 1,34E-01 | 3,39E-01 |
| GGPS1 | 304,61 | 0,08 | 0,05 | 1,50 | 1,34E-01 | 3,39E-01 |
| POGK | 1340,22 | 0,04 | 0,03 | 1,50 | 1,34E-01 | 3,39E-01 |
| UROS | 670,04 | -0,05 | 0,03 | -1,50 | 1,34E-01 | 3,39E-01 |
| NEK10 | 96,95 | -0,20 | 0,14 | -1,50 | 1,34E-01 | 3,39E-01 |
| NBN | 3189,54 | 0,12 | 0,08 | 1,50 | 1,34E-01 | 3,40E-01 |
| KLHL21 | 1784,75 | -0,12 | 0,08 | -1,50 | 1,34E-01 | 3,40E-01 |
| RFX3 | 1767,14 | 0,08 | 0,06 | 1,50 | 1,34E-01 | 3,40E-01 |
| METTL18 | 131,40 | 0,20 | 0,13 | 1,50 | 1,34E-01 | 3,40E-01 |
| RFC4 | 161,81 | 0,08 | 0,05 | 1,50 | 1,34E-01 | 3,40E-01 |
| TRIAP1 | 152,69 | 0,16 | 0,11 | 1,50 | 1,34E-01 | 3,40E-01 |
| DPCD | 187,99 | -0,17 | 0,11 | -1,50 | 1,34E-01 | 3,40E-01 |
| BCL6 | 5430,15 | -0,14 | 0,10 | -1,50 | 1,34E-01 | 3,41E-01 |
| ZNF195 | 1522,05 | 0,08 | 0,05 | 1,50 | 1,34E-01 | 3,41E-01 |

| | | | | | | |
|---|---|---|---|---|---|---|
| RABGAP1 | 1635,78 | 0,05 | 0,03 | 1,50 | 1,34E-01 | 3,41E-01 |
| NAV1 | 234,20 | -0,18 | 0,12 | -1,50 | 1,35E-01 | 3,41E-01 |
| ABCB4 | 279,10 | 0,18 | 0,12 | 1,50 | 1,35E-01 | 3,41E-01 |
| TTC32 | 701,66 | 0,11 | 0,07 | 1,50 | 1,35E-01 | 3,41E-01 |
| ZNF598 | 259,10 | -0,14 | 0,09 | -1,50 | 1,35E-01 | 3,41E-01 |
| ERI1 | 694,90 | 0,08 | 0,05 | 1,50 | 1,35E-01 | 3,41E-01 |
| PLPP6 | 346,04 | 0,09 | 0,06 | 1,50 | 1,35E-01 | 3,41E-01 |
| ZKSCAN4 | 383,37 | 0,05 | 0,04 | 1,50 | 1,35E-01 | 3,41E-01 |
| CFH | 280,18 | 0,21 | 0,14 | 1,50 | 1,35E-01 | 3,41E-01 |
| MOB2 | 1376,25 | -0,06 | 0,04 | -1,50 | 1,35E-01 | 3,41E-01 |
| RIMOC1 | 848,71 | 0,07 | 0,04 | 1,49 | 1,35E-01 | 3,41E-01 |
| FOCAD | 1383,66 | 0,08 | 0,05 | 1,49 | 1,35E-01 | 3,41E-01 |
| SLC46A3 | 718,18 | 0,06 | 0,04 | 1,49 | 1,35E-01 | 3,41E-01 |
| MFAP3L | 864,45 | -0,15 | 0,10 | -1,49 | 1,35E-01 | 3,41E-01 |
| GFOD2 | 882,11 | -0,10 | 0,07 | -1,49 | 1,35E-01 | 3,41E-01 |
| ARMT1 | 707,22 | 0,12 | 0,08 | 1,49 | 1,35E-01 | 3,41E-01 |
| AKT1S1 | 379,07 | -0,10 | 0,06 | -1,49 | 1,35E-01 | 3,41E-01 |
| EIF2AK1 | 3979,68 | -0,16 | 0,11 | -1,49 | 1,35E-01 | 3,41E-01 |
| SACM1L | 3754,60 | 0,09 | 0,06 | 1,49 | 1,35E-01 | 3,41E-01 |
| RPL27 | 3799,78 | 0,21 | 0,14 | 1,49 | 1,35E-01 | 3,41E-01 |
| FCAR | 1001,29 | -0,14 | 0,09 | -1,49 | 1,35E-01 | 3,41E-01 |
| AZI2 | 904,23 | 0,15 | 0,10 | 1,49 | 1,35E-01 | 3,42E-01 |
| GSTCD | 249,48 | 0,08 | 0,05 | 1,49 | 1,35E-01 | 3,42E-01 |
| CD2AP | 1467,41 | 0,11 | 0,07 | 1,49 | 1,35E-01 | 3,42E-01 |
| SAMD1 | 77,90 | -0,18 | 0,12 | -1,49 | 1,36E-01 | 3,42E-01 |
| SIRT3 | 263,55 | -0,08 | 0,05 | -1,49 | 1,36E-01 | 3,42E-01 |
| QRICH2 | 141,43 | -0,12 | 0,08 | -1,49 | 1,36E-01 | 3,42E-01 |
| NFKBIL1 | 89,26 | -0,10 | 0,07 | -1,49 | 1,36E-01 | 3,42E-01 |
| PIAS4 | 459,25 | -0,09 | 0,06 | -1,49 | 1,36E-01 | 3,42E-01 |
| TSPAN15 | 67,59 | -0,21 | 0,14 | -1,49 | 1,36E-01 | 3,42E-01 |
| SLCO3A1 | 2836,63 | -0,09 | 0,06 | -1,49 | 1,36E-01 | 3,42E-01 |
| CHD6 | 3414,34 | 0,07 | 0,04 | 1,49 | 1,36E-01 | 3,42E-01 |
| SESTD1 | 1114,13 | 0,10 | 0,07 | 1,49 | 1,36E-01 | 3,42E-01 |
| REXO1 | 324,24 | -0,20 | 0,14 | -1,49 | 1,36E-01 | 3,42E-01 |
| SMIM30 | 146,51 | 0,18 | 0,12 | 1,49 | 1,36E-01 | 3,42E-01 |
| PLEKHG7 | 48,95 | -0,23 | 0,16 | -1,49 | 1,36E-01 | 3,42E-01 |
| ZNF879 | 280,25 | 0,13 | 0,09 | 1,49 | 1,36E-01 | 3,42E-01 |
| TXNL1 | 950,50 | 0,07 | 0,04 | 1,49 | 1,36E-01 | 3,42E-01 |
| EBP | 305,22 | 0,07 | 0,05 | 1,49 | 1,36E-01 | 3,42E-01 |
| TAF1C | 211,41 | -0,19 | 0,12 | -1,49 | 1,36E-01 | 3,42E-01 |
| SHISA4 | 133,27 | -0,26 | 0,17 | -1,49 | 1,36E-01 | 3,42E-01 |
| SERINC1 | 7806,61 | 0,13 | 0,09 | 1,49 | 1,36E-01 | 3,42E-01 |
| SF3A2 | 338,14 | -0,11 | 0,08 | -1,49 | 1,36E-01 | 3,43E-01 |
| SLC9A1 | 196,15 | -0,21 | 0,14 | -1,49 | 1,37E-01 | 3,43E-01 |
| PTPN4 | 5591,24 | 0,12 | 0,08 | 1,49 | 1,36E-01 | 3,43E-01 |
| OGFRL1 | 17220,26 | 0,13 | 0,09 | 1,49 | 1,37E-01 | 3,43E-01 |

| | | | | | | |
|---|---|---|---|---|---|---|
| PLK1 | 80,27 | -0,16 | 0,11 | -1,49 | 1,37E-01 | 3,43E-01 |
| PTGFRN | 72,91 | -0,21 | 0,14 | -1,49 | 1,37E-01 | 3,43E-01 |
| GIMAP4 | 10295,81 | 0,11 | 0,08 | 1,49 | 1,37E-01 | 3,43E-01 |
| ARF5 | 502,43 | -0,08 | 0,05 | -1,49 | 1,37E-01 | 3,43E-01 |
| NLRC5 | 5432,55 | -0,14 | 0,10 | -1,49 | 1,37E-01 | 3,43E-01 |
| CHMP2B | 829,35 | 0,10 | 0,07 | 1,49 | 1,37E-01 | 3,43E-01 |
| ATP2A3 | 3669,40 | -0,13 | 0,09 | -1,49 | 1,37E-01 | 3,43E-01 |
| C9orf43 | 27,46 | -0,21 | 0,14 | -1,49 | 1,37E-01 | 3,43E-01 |
| BTF3L4 | 1101,33 | 0,09 | 0,06 | 1,49 | 1,37E-01 | 3,43E-01 |
| ENSG000002 | 50,15 | -0,23 | 0,16 | -1,49 | 1,37E-01 | 3,43E-01 |
| SV2A | 43,46 | -0,22 | 0,15 | -1,49 | 1,37E-01 | 3,43E-01 |
| JADE1 | 2460,46 | 0,05 | 0,03 | 1,49 | 1,37E-01 | 3,43E-01 |
| PPCS | 811,63 | 0,07 | 0,05 | 1,49 | 1,37E-01 | 3,43E-01 |
| ARHGEF3 | 2590,73 | 0,05 | 0,04 | 1,49 | 1,37E-01 | 3,43E-01 |
| THAP2 | 303,83 | 0,11 | 0,08 | 1,49 | 1,37E-01 | 3,43E-01 |
| RAP1A | 3565,35 | 0,08 | 0,06 | 1,49 | 1,37E-01 | 3,44E-01 |
| THBS4 | 71,20 | -0,20 | 0,13 | -1,48 | 1,38E-01 | 3,44E-01 |
| ZNF581 | 125,75 | -0,10 | 0,06 | -1,48 | 1,38E-01 | 3,44E-01 |
| STK24 | 3552,35 | -0,06 | 0,04 | -1,48 | 1,38E-01 | 3,44E-01 |
| ZNF563 | 123,84 | 0,11 | 0,07 | 1,48 | 1,38E-01 | 3,44E-01 |
| TLCD2 | 59,76 | -0,23 | 0,16 | -1,48 | 1,38E-01 | 3,45E-01 |
| RAB28 | 646,57 | 0,08 | 0,05 | 1,48 | 1,38E-01 | 3,45E-01 |
| GOLGA6L9 | 135,40 | 0,13 | 0,09 | 1,48 | 1,38E-01 | 3,45E-01 |
| DDOST | 2167,62 | 0,06 | 0,04 | 1,48 | 1,38E-01 | 3,45E-01 |
| FOXO1 | 2070,05 | 0,06 | 0,04 | 1,48 | 1,38E-01 | 3,45E-01 |
| AOC2 | 199,80 | -0,16 | 0,11 | -1,48 | 1,38E-01 | 3,45E-01 |
| PABIR2 | 820,61 | 0,08 | 0,05 | 1,48 | 1,38E-01 | 3,45E-01 |
| METTL26 | 131,45 | -0,11 | 0,07 | -1,48 | 1,38E-01 | 3,45E-01 |
| GPR183 | 1372,71 | 0,15 | 0,10 | 1,48 | 1,38E-01 | 3,45E-01 |
| PTCRA | 162,79 | -0,19 | 0,13 | -1,48 | 1,38E-01 | 3,45E-01 |
| INF2 | 353,48 | -0,21 | 0,14 | -1,48 | 1,38E-01 | 3,45E-01 |
| C2CD2L | 383,20 | -0,14 | 0,09 | -1,48 | 1,38E-01 | 3,45E-01 |
| SLC36A1 | 1381,16 | -0,10 | 0,07 | -1,48 | 1,38E-01 | 3,45E-01 |
| TMEM234 | 346,24 | -0,09 | 0,06 | -1,48 | 1,38E-01 | 3,45E-01 |
| PCDHGC3 | 30,86 | -0,22 | 0,15 | -1,48 | 1,39E-01 | 3,45E-01 |
| NCF1 | 3810,34 | -0,13 | 0,09 | -1,48 | 1,39E-01 | 3,45E-01 |
| VPS37B | 1023,91 | -0,11 | 0,07 | -1,48 | 1,39E-01 | 3,45E-01 |
| THG1L | 236,02 | 0,08 | 0,05 | 1,48 | 1,39E-01 | 3,45E-01 |
| PRXL2A | 114,46 | -0,12 | 0,08 | -1,48 | 1,39E-01 | 3,45E-01 |
| ARL13B | 248,44 | 0,07 | 0,05 | 1,48 | 1,39E-01 | 3,45E-01 |
| ROCK2 | 1909,40 | 0,08 | 0,05 | 1,48 | 1,39E-01 | 3,45E-01 |
| ZNF746 | 2206,06 | -0,12 | 0,08 | -1,48 | 1,39E-01 | 3,45E-01 |
| TATDN2 | 896,69 | -0,07 | 0,05 | -1,48 | 1,39E-01 | 3,45E-01 |
| RCE1 | 184,81 | -0,08 | 0,05 | -1,48 | 1,39E-01 | 3,45E-01 |
| KIF9 | 66,74 | -0,17 | 0,11 | -1,48 | 1,39E-01 | 3,45E-01 |
| POFUT2 | 407,61 | -0,12 | 0,08 | -1,48 | 1,39E-01 | 3,45E-01 |

| | | | | | | |
|---|---|---|---|---|---|---|
| PKN2 | 4755,82 | 0,11 | 0,07 | 1,48 | 1,39E-01 | 3,45E-01 |
| SPSB1 | 47,21 | -0,21 | 0,14 | -1,48 | 1,39E-01 | 3,45E-01 |
| TRIT1 | 570,29 | 0,06 | 0,04 | 1,48 | 1,39E-01 | 3,45E-01 |
| SIPA1L2 | 1747,91 | -0,14 | 0,10 | -1,48 | 1,39E-01 | 3,45E-01 |
| PTK2 | 432,13 | -0,10 | 0,07 | -1,48 | 1,39E-01 | 3,45E-01 |
| PTBP2 | 1529,95 | 0,10 | 0,07 | 1,48 | 1,39E-01 | 3,45E-01 |
| MMAA | 338,26 | 0,09 | 0,06 | 1,48 | 1,39E-01 | 3,45E-01 |
| XPO6 | 20084,18 | -0,10 | 0,07 | -1,48 | 1,39E-01 | 3,45E-01 |
| MARVELD1 | 49,31 | -0,24 | 0,16 | -1,48 | 1,39E-01 | 3,45E-01 |
| SELENOO | 673,53 | -0,09 | 0,06 | -1,48 | 1,39E-01 | 3,46E-01 |
| IFT88 | 518,10 | 0,09 | 0,06 | 1,48 | 1,39E-01 | 3,46E-01 |
| MBNL1 | 26427,74 | 0,08 | 0,05 | 1,48 | 1,40E-01 | 3,46E-01 |
| RORA | 8193,19 | 0,12 | 0,08 | 1,48 | 1,40E-01 | 3,46E-01 |
| FAAP20 | 266,83 | -0,12 | 0,08 | -1,48 | 1,40E-01 | 3,47E-01 |
| PRSS33 | 108,58 | -0,27 | 0,18 | -1,48 | 1,40E-01 | 3,47E-01 |
| CCT4 | 1699,97 | 0,08 | 0,05 | 1,48 | 1,40E-01 | 3,47E-01 |
| ATP5MJ | 427,77 | 0,13 | 0,09 | 1,48 | 1,40E-01 | 3,47E-01 |
| ZSCAN16 | 281,02 | 0,08 | 0,05 | 1,48 | 1,40E-01 | 3,47E-01 |
| SLX9 | 235,94 | -0,06 | 0,04 | -1,48 | 1,40E-01 | 3,47E-01 |
| ZNF69 | 155,87 | 0,08 | 0,06 | 1,48 | 1,40E-01 | 3,47E-01 |
| POLR1E | 385,56 | 0,06 | 0,04 | 1,48 | 1,40E-01 | 3,47E-01 |
| PITHD1 | 4467,43 | -0,19 | 0,13 | -1,48 | 1,40E-01 | 3,47E-01 |
| FADS3 | 88,52 | -0,17 | 0,11 | -1,47 | 1,40E-01 | 3,47E-01 |
| ATP13A2 | 231,58 | -0,18 | 0,12 | -1,48 | 1,40E-01 | 3,47E-01 |
| NREP | 234,94 | 0,09 | 0,06 | 1,48 | 1,40E-01 | 3,47E-01 |
| POLQ | 141,17 | -0,16 | 0,11 | -1,48 | 1,40E-01 | 3,47E-01 |
| PNKP | 968,14 | -0,11 | 0,07 | -1,48 | 1,40E-01 | 3,47E-01 |
| TMEM167B | 1806,36 | 0,10 | 0,07 | 1,48 | 1,40E-01 | 3,47E-01 |
| BACH2 | 1451,93 | 0,12 | 0,08 | 1,48 | 1,40E-01 | 3,47E-01 |
| PPM1B | 1794,16 | 0,12 | 0,08 | 1,47 | 1,40E-01 | 3,47E-01 |
| SF3B3 | 3311,04 | 0,05 | 0,03 | 1,47 | 1,40E-01 | 3,47E-01 |
| YIPF3 | 443,14 | -0,13 | 0,09 | -1,47 | 1,40E-01 | 3,47E-01 |
| LNPK | 1527,41 | 0,11 | 0,07 | 1,47 | 1,41E-01 | 3,47E-01 |
| ENSG000002 | 97,08 | -0,14 | 0,10 | -1,47 | 1,41E-01 | 3,47E-01 |
| ITPRIP | 3078,45 | -0,11 | 0,07 | -1,47 | 1,41E-01 | 3,48E-01 |
| ACTB | 26344,76 | -0,15 | 0,10 | -1,47 | 1,41E-01 | 3,48E-01 |
| DCLRE1A | 275,02 | 0,11 | 0,07 | 1,47 | 1,41E-01 | 3,48E-01 |
| WDR24 | 91,14 | -0,16 | 0,11 | -1,47 | 1,41E-01 | 3,48E-01 |
| CEP70 | 220,86 | 0,13 | 0,09 | 1,47 | 1,41E-01 | 3,48E-01 |
| COA4 | 473,75 | 0,05 | 0,04 | 1,47 | 1,41E-01 | 3,48E-01 |
| MFN1 | 2378,27 | 0,09 | 0,06 | 1,47 | 1,41E-01 | 3,48E-01 |
| GAL3ST4 | 33,91 | -0,25 | 0,17 | -1,47 | 1,41E-01 | 3,48E-01 |
| ASAP1 | 3370,94 | -0,10 | 0,07 | -1,47 | 1,41E-01 | 3,48E-01 |
| SAMD10 | 52,37 | -0,19 | 0,13 | -1,47 | 1,41E-01 | 3,48E-01 |
| TUBE1 | 605,62 | 0,11 | 0,07 | 1,47 | 1,41E-01 | 3,48E-01 |
| ZFP36 | 1406,13 | -0,12 | 0,08 | -1,47 | 1,41E-01 | 3,48E-01 |

| | | | | | | |
|---|---|---|---|---|---|---|
| CCS | 941,34 | -0,10 | 0,07 | -1,47 | 1,41E-01 | 3,48E-01 |
| PAQR3 | 450,02 | 0,09 | 0,06 | 1,47 | 1,42E-01 | 3,48E-01 |
| PDPK1 | 1656,39 | -0,10 | 0,07 | -1,47 | 1,42E-01 | 3,49E-01 |
| ATF1 | 669,64 | 0,07 | 0,05 | 1,47 | 1,42E-01 | 3,49E-01 |
| SNX18 | 5648,39 | 0,08 | 0,06 | 1,47 | 1,42E-01 | 3,49E-01 |
| ZNF790 | 238,56 | 0,11 | 0,07 | 1,47 | 1,42E-01 | 3,49E-01 |
| SOS1 | 2962,71 | 0,09 | 0,06 | 1,47 | 1,42E-01 | 3,49E-01 |
| TBCCD1 | 463,89 | 0,05 | 0,04 | 1,47 | 1,42E-01 | 3,49E-01 |
| RNPC3 | 1900,12 | 0,09 | 0,06 | 1,47 | 1,42E-01 | 3,49E-01 |
| SMAD2 | 8174,21 | 0,06 | 0,04 | 1,47 | 1,42E-01 | 3,49E-01 |
| CC2D1B | 963,37 | -0,09 | 0,06 | -1,47 | 1,42E-01 | 3,49E-01 |
| GNAZ | 409,40 | -0,19 | 0,13 | -1,47 | 1,42E-01 | 3,49E-01 |
| TRIM32 | 412,75 | 0,07 | 0,05 | 1,47 | 1,42E-01 | 3,49E-01 |
| UBR3 | 3569,51 | 0,08 | 0,05 | 1,47 | 1,42E-01 | 3,49E-01 |
| ABHD14A | 51,06 | -0,10 | 0,06 | -1,47 | 1,42E-01 | 3,49E-01 |
| LHFPL2 | 542,11 | -0,11 | 0,08 | -1,47 | 1,42E-01 | 3,49E-01 |
| ARID4A | 3524,47 | 0,10 | 0,07 | 1,47 | 1,42E-01 | 3,49E-01 |
| HIC1 | 143,80 | -0,18 | 0,12 | -1,47 | 1,42E-01 | 3,49E-01 |
| DEPDC5 | 743,63 | -0,10 | 0,06 | -1,47 | 1,42E-01 | 3,49E-01 |
| MYCT1 | 76,44 | -0,15 | 0,10 | -1,47 | 1,42E-01 | 3,49E-01 |
| SIT1 | 928,18 | 0,09 | 0,06 | 1,47 | 1,43E-01 | 3,50E-01 |
| SLC39A13 | 171,12 | -0,13 | 0,09 | -1,47 | 1,43E-01 | 3,50E-01 |
| HGS | 833,12 | -0,14 | 0,10 | -1,47 | 1,43E-01 | 3,50E-01 |
| LYPLA1 | 1784,84 | 0,09 | 0,06 | 1,47 | 1,43E-01 | 3,50E-01 |
| PFKFB4 | 2165,74 | -0,12 | 0,08 | -1,47 | 1,43E-01 | 3,50E-01 |
| TELO2 | 314,09 | -0,14 | 0,10 | -1,47 | 1,43E-01 | 3,50E-01 |
| CD96 | 2758,44 | 0,10 | 0,07 | 1,47 | 1,43E-01 | 3,50E-01 |
| C1GALT1C1 | 433,04 | 0,12 | 0,08 | 1,47 | 1,43E-01 | 3,50E-01 |
| RANBP9 | 1162,86 | 0,09 | 0,06 | 1,47 | 1,43E-01 | 3,50E-01 |
| ANXA11 | 4925,42 | -0,11 | 0,08 | -1,47 | 1,43E-01 | 3,50E-01 |
| BRF2 | 390,34 | 0,05 | 0,03 | 1,46 | 1,43E-01 | 3,50E-01 |
| SLC9A6 | 871,01 | 0,05 | 0,03 | 1,46 | 1,43E-01 | 3,50E-01 |
| CTSG | 45,55 | -0,31 | 0,21 | -1,46 | 1,43E-01 | 3,50E-01 |
| ADGRG6 | 49,57 | -0,25 | 0,17 | -1,46 | 1,43E-01 | 3,50E-01 |
| GSTM2 | 167,61 | -0,15 | 0,10 | -1,46 | 1,43E-01 | 3,50E-01 |
| CPAMD8 | 151,99 | -0,21 | 0,14 | -1,46 | 1,43E-01 | 3,50E-01 |
| UBE2F | 581,90 | -0,06 | 0,04 | -1,46 | 1,43E-01 | 3,50E-01 |
| VPS37A | 1187,69 | 0,07 | 0,05 | 1,46 | 1,43E-01 | 3,50E-01 |
| ZNF814 | 678,84 | 0,08 | 0,06 | 1,46 | 1,43E-01 | 3,50E-01 |
| CEP152 | 607,34 | 0,08 | 0,06 | 1,46 | 1,43E-01 | 3,50E-01 |
| PTTG1 | 94,51 | -0,11 | 0,07 | -1,46 | 1,43E-01 | 3,50E-01 |
| EIF4G2 | 7902,68 | 0,06 | 0,04 | 1,46 | 1,43E-01 | 3,50E-01 |
| WWC3 | 1939,23 | -0,11 | 0,08 | -1,46 | 1,43E-01 | 3,50E-01 |
| GEMIN8 | 266,23 | 0,06 | 0,04 | 1,46 | 1,43E-01 | 3,50E-01 |
| FDXR | 113,66 | -0,13 | 0,09 | -1,46 | 1,43E-01 | 3,50E-01 |
| CENPL | 199,00 | 0,07 | 0,05 | 1,46 | 1,44E-01 | 3,50E-01 |

| | | | | | | |
|---|---|---|---|---|---|---|
| DCAF1 | 1185,82 | 0,05 | 0,04 | 1,46 | 1,44E-01 | 3,50E-01 |
| XRN2 | 4147,62 | 0,07 | 0,05 | 1,46 | 1,44E-01 | 3,50E-01 |
| AGAP1 | 356,92 | -0,19 | 0,13 | -1,46 | 1,44E-01 | 3,50E-01 |
| KDM6B | 2200,08 | -0,20 | 0,14 | -1,46 | 1,44E-01 | 3,50E-01 |
| RFNG | 125,97 | -0,16 | 0,11 | -1,46 | 1,44E-01 | 3,51E-01 |
| CFAP53 | 35,50 | -0,19 | 0,13 | -1,46 | 1,44E-01 | 3,51E-01 |
| DENND1B | 2040,17 | 0,12 | 0,08 | 1,46 | 1,44E-01 | 3,51E-01 |
| CXCR5 | 280,44 | 0,14 | 0,10 | 1,46 | 1,44E-01 | 3,51E-01 |
| SRR | 60,99 | -0,12 | 0,08 | -1,46 | 1,44E-01 | 3,51E-01 |
| LIN54 | 1188,40 | 0,06 | 0,04 | 1,46 | 1,44E-01 | 3,51E-01 |
| KCTD15 | 319,13 | -0,10 | 0,07 | -1,46 | 1,44E-01 | 3,52E-01 |
| IL6ST | 5947,26 | 0,12 | 0,08 | 1,46 | 1,44E-01 | 3,52E-01 |
| ATP5PO | 421,63 | 0,10 | 0,07 | 1,46 | 1,44E-01 | 3,52E-01 |
| LCAT | 38,83 | -0,19 | 0,13 | -1,46 | 1,44E-01 | 3,52E-01 |
| METTL3 | 1376,72 | 0,05 | 0,03 | 1,46 | 1,44E-01 | 3,52E-01 |
| CEACAM8 | 133,11 | -0,28 | 0,20 | -1,46 | 1,45E-01 | 3,52E-01 |
| ATP5MK | 546,48 | 0,13 | 0,09 | 1,46 | 1,45E-01 | 3,52E-01 |
| MIB2 | 676,35 | -0,08 | 0,06 | -1,46 | 1,45E-01 | 3,52E-01 |
| TJAP1 | 273,38 | -0,12 | 0,08 | -1,46 | 1,45E-01 | 3,52E-01 |
| ZNF707 | 367,83 | -0,09 | 0,07 | -1,46 | 1,45E-01 | 3,52E-01 |
| ACADS | 306,63 | -0,07 | 0,05 | -1,46 | 1,45E-01 | 3,52E-01 |
| HNRNPL | 2887,77 | 0,04 | 0,03 | 1,46 | 1,45E-01 | 3,52E-01 |
| KLHL26 | 107,72 | -0,15 | 0,11 | -1,46 | 1,45E-01 | 3,52E-01 |
| BORCS8 | 301,21 | -0,15 | 0,10 | -1,46 | 1,45E-01 | 3,52E-01 |
| LRRN3 | 296,39 | 0,24 | 0,17 | 1,46 | 1,45E-01 | 3,52E-01 |
| ZFP30 | 521,17 | 0,13 | 0,09 | 1,46 | 1,45E-01 | 3,52E-01 |
| ADAM23 | 239,92 | -0,20 | 0,14 | -1,46 | 1,45E-01 | 3,52E-01 |
| PI4KA | 2083,32 | -0,12 | 0,08 | -1,46 | 1,45E-01 | 3,52E-01 |
| TMEM91 | 656,12 | -0,10 | 0,07 | -1,46 | 1,45E-01 | 3,52E-01 |
| KHDRBS2 | 122,67 | 0,17 | 0,11 | 1,46 | 1,45E-01 | 3,52E-01 |
| SLC6A8 | 1059,72 | -0,28 | 0,19 | -1,46 | 1,45E-01 | 3,52E-01 |
| TRAPPC9 | 1737,06 | -0,10 | 0,07 | -1,46 | 1,45E-01 | 3,52E-01 |
| ELOC | 457,08 | 0,07 | 0,05 | 1,46 | 1,45E-01 | 3,52E-01 |
| SNAPIN | 394,52 | 0,07 | 0,05 | 1,46 | 1,45E-01 | 3,52E-01 |
| YIPF1 | 600,19 | -0,06 | 0,04 | -1,46 | 1,45E-01 | 3,52E-01 |
| TAGLN2 | 11795,19 | -0,11 | 0,07 | -1,46 | 1,45E-01 | 3,52E-01 |
| MFHAS1 | 851,76 | -0,10 | 0,07 | -1,46 | 1,45E-01 | 3,52E-01 |
| CFAP58 | 134,08 | -0,14 | 0,10 | -1,46 | 1,46E-01 | 3,53E-01 |
| ZFP1 | 204,31 | 0,09 | 0,06 | 1,46 | 1,46E-01 | 3,53E-01 |
| TMEM222 | 318,16 | -0,09 | 0,06 | -1,46 | 1,46E-01 | 3,53E-01 |
| ENSG000002 | 96,17 | -0,19 | 0,13 | -1,45 | 1,46E-01 | 3,53E-01 |
| ELANE | 36,70 | -0,27 | 0,19 | -1,45 | 1,46E-01 | 3,53E-01 |
| REEP3 | 812,07 | 0,10 | 0,07 | 1,45 | 1,46E-01 | 3,53E-01 |
| SLC35F5 | 2797,97 | 0,08 | 0,06 | 1,45 | 1,46E-01 | 3,53E-01 |
| PHETA1 | 95,98 | -0,16 | 0,11 | -1,45 | 1,46E-01 | 3,53E-01 |
| FHOD1 | 316,79 | -0,15 | 0,10 | -1,45 | 1,46E-01 | 3,53E-01 |

| | | | | | | |
|---|---|---|---|---|---|---|
| DNAJC27 | 225,95 | 0,07 | 0,05 | 1,45 | 1,46E-01 | 3,53E-01 |
| WDR5 | 355,54 | -0,07 | 0,04 | -1,45 | 1,46E-01 | 3,53E-01 |
| ZDHHC17 | 3985,98 | 0,11 | 0,07 | 1,45 | 1,46E-01 | 3,53E-01 |
| KCNK13 | 37,85 | -0,18 | 0,13 | -1,45 | 1,46E-01 | 3,53E-01 |
| DAP3 | 1399,87 | 0,04 | 0,03 | 1,45 | 1,46E-01 | 3,53E-01 |
| COASY | 203,47 | -0,16 | 0,11 | -1,45 | 1,46E-01 | 3,53E-01 |
| NMNAT3 | 172,70 | -0,14 | 0,10 | -1,45 | 1,46E-01 | 3,53E-01 |
| DDX51 | 175,01 | -0,17 | 0,12 | -1,45 | 1,46E-01 | 3,53E-01 |
| ENSG000002 | 27,63 | -0,15 | 0,10 | -1,45 | 1,46E-01 | 3,53E-01 |
| TPMT | 375,62 | 0,11 | 0,07 | 1,45 | 1,46E-01 | 3,53E-01 |
| ABCA6 | 180,49 | -0,27 | 0,19 | -1,45 | 1,46E-01 | 3,53E-01 |
| IAH1 | 677,76 | 0,05 | 0,03 | 1,45 | 1,46E-01 | 3,53E-01 |
| WNT7A | 136,49 | -0,15 | 0,10 | -1,45 | 1,46E-01 | 3,53E-01 |
| TCEAL1 | 81,46 | 0,13 | 0,09 | 1,45 | 1,47E-01 | 3,53E-01 |
| ZNF512B | 156,76 | -0,21 | 0,14 | -1,45 | 1,46E-01 | 3,53E-01 |
| TP53RK | 971,66 | 0,08 | 0,06 | 1,45 | 1,46E-01 | 3,53E-01 |
| CUL7 | 253,27 | -0,12 | 0,08 | -1,45 | 1,47E-01 | 3,53E-01 |
| ZKSCAN1 | 2669,52 | 0,04 | 0,03 | 1,45 | 1,47E-01 | 3,53E-01 |
| ALKBH2 | 154,58 | 0,08 | 0,05 | 1,45 | 1,47E-01 | 3,53E-01 |
| PHLPP2 | 802,34 | -0,09 | 0,06 | -1,45 | 1,47E-01 | 3,54E-01 |
| LARGE1 | 220,78 | -0,20 | 0,14 | -1,45 | 1,47E-01 | 3,54E-01 |
| CDCA8 | 34,13 | -0,17 | 0,12 | -1,45 | 1,47E-01 | 3,54E-01 |
| RPL22 | 2582,00 | 0,12 | 0,08 | 1,45 | 1,47E-01 | 3,54E-01 |
| IGHMBP2 | 413,74 | -0,13 | 0,09 | -1,45 | 1,47E-01 | 3,54E-01 |
| LRRC25 | 1399,97 | -0,14 | 0,10 | -1,45 | 1,47E-01 | 3,54E-01 |
| YWHAG | 1375,93 | 0,05 | 0,03 | 1,45 | 1,47E-01 | 3,54E-01 |
| ZNF829 | 207,43 | 0,11 | 0,07 | 1,45 | 1,47E-01 | 3,54E-01 |
| EPB41L2 | 465,20 | 0,10 | 0,07 | 1,45 | 1,47E-01 | 3,54E-01 |
| ATG16L2 | 10067,08 | -0,15 | 0,10 | -1,45 | 1,47E-01 | 3,54E-01 |
| SNED1 | 353,78 | -0,17 | 0,12 | -1,45 | 1,47E-01 | 3,54E-01 |
| LRCH1 | 943,54 | 0,06 | 0,04 | 1,45 | 1,47E-01 | 3,54E-01 |
| GOLGA8B | 1386,63 | 0,14 | 0,09 | 1,45 | 1,47E-01 | 3,54E-01 |
| GPN3 | 262,85 | 0,10 | 0,07 | 1,45 | 1,47E-01 | 3,54E-01 |
| ERBIN | 11100,25 | 0,10 | 0,07 | 1,45 | 1,47E-01 | 3,54E-01 |
| RPS25 | 5981,39 | 0,16 | 0,11 | 1,45 | 1,47E-01 | 3,54E-01 |
| SNX27 | 3543,01 | -0,10 | 0,07 | -1,45 | 1,47E-01 | 3,54E-01 |
| ZCCHC24 | 106,09 | -0,14 | 0,10 | -1,45 | 1,47E-01 | 3,54E-01 |
| LIN37 | 171,22 | -0,06 | 0,04 | -1,45 | 1,47E-01 | 3,54E-01 |
| PRMT6 | 479,91 | 0,09 | 0,06 | 1,45 | 1,47E-01 | 3,54E-01 |
| WIPF2 | 1197,73 | -0,11 | 0,07 | -1,45 | 1,48E-01 | 3,54E-01 |
| KIF5C | 373,40 | -0,12 | 0,09 | -1,45 | 1,48E-01 | 3,54E-01 |
| MAGED1 | 147,85 | -0,12 | 0,08 | -1,45 | 1,48E-01 | 3,54E-01 |
| FOXK2 | 768,09 | -0,08 | 0,06 | -1,45 | 1,48E-01 | 3,54E-01 |
| PIGN | 1708,72 | 0,06 | 0,04 | 1,45 | 1,48E-01 | 3,55E-01 |
| CALU | 795,66 | 0,06 | 0,04 | 1,45 | 1,48E-01 | 3,55E-01 |
| CBLN3 | 43,36 | -0,19 | 0,13 | -1,45 | 1,48E-01 | 3,55E-01 |

| | | | | | | |
|---|---|---|---|---|---|---|
| SUCLG1 | 1084,75 | 0,06 | 0,04 | 1,45 | 1,48E-01 | 3,55E-01 |
| MAFF | 182,45 | -0,11 | 0,08 | -1,45 | 1,48E-01 | 3,55E-01 |
| ZNF148 | 4192,40 | 0,10 | 0,07 | 1,45 | 1,48E-01 | 3,55E-01 |
| DDX5 | 25731,86 | 0,07 | 0,05 | 1,45 | 1,48E-01 | 3,55E-01 |
| TMEM170B | 3293,26 | 0,14 | 0,10 | 1,45 | 1,48E-01 | 3,55E-01 |
| ZNF292 | 6804,21 | 0,12 | 0,08 | 1,45 | 1,48E-01 | 3,55E-01 |
| SENP1 | 944,40 | 0,05 | 0,04 | 1,45 | 1,48E-01 | 3,55E-01 |
| TAS2R41 | 49,68 | -0,20 | 0,14 | -1,45 | 1,48E-01 | 3,55E-01 |
| PCNT | 1850,37 | -0,08 | 0,06 | -1,45 | 1,48E-01 | 3,55E-01 |
| GLRX5 | 5252,96 | -0,19 | 0,13 | -1,44 | 1,49E-01 | 3,55E-01 |
| MAFG | 289,25 | -0,14 | 0,10 | -1,44 | 1,49E-01 | 3,55E-01 |
| POLI | 1150,45 | 0,09 | 0,06 | 1,44 | 1,49E-01 | 3,55E-01 |
| PTDSS1 | 1986,58 | 0,05 | 0,03 | 1,44 | 1,49E-01 | 3,56E-01 |
| ZNF544 | 480,12 | 0,06 | 0,04 | 1,44 | 1,49E-01 | 3,56E-01 |
| LATS2 | 636,17 | -0,08 | 0,05 | -1,44 | 1,49E-01 | 3,56E-01 |
| PPFIA4 | 96,93 | -0,26 | 0,18 | -1,44 | 1,49E-01 | 3,56E-01 |
| DPH1 | 188,23 | -0,12 | 0,09 | -1,44 | 1,49E-01 | 3,56E-01 |
| C18orf32 | 324,29 | -0,05 | 0,04 | -1,44 | 1,49E-01 | 3,56E-01 |
| ULK1 | 707,00 | -0,21 | 0,14 | -1,44 | 1,49E-01 | 3,56E-01 |
| PCMTD1 | 6030,79 | 0,12 | 0,08 | 1,44 | 1,49E-01 | 3,56E-01 |
| ZNF507 | 1603,34 | 0,08 | 0,05 | 1,44 | 1,49E-01 | 3,56E-01 |
| BLNK | 295,93 | 0,13 | 0,09 | 1,44 | 1,49E-01 | 3,56E-01 |
| KIAA0930 | 966,84 | -0,15 | 0,10 | -1,44 | 1,49E-01 | 3,57E-01 |
| PFN1 | 3502,99 | -0,08 | 0,06 | -1,44 | 1,49E-01 | 3,57E-01 |
| UQCRFS1 | 898,09 | 0,07 | 0,05 | 1,44 | 1,49E-01 | 3,57E-01 |
| SRCAP | 424,02 | -0,10 | 0,07 | -1,44 | 1,50E-01 | 3,57E-01 |
| ANKRD34A | 50,58 | -0,13 | 0,09 | -1,44 | 1,50E-01 | 3,57E-01 |
| C2 | 93,68 | -0,11 | 0,07 | -1,44 | 1,50E-01 | 3,57E-01 |
| RETREG2 | 2503,47 | -0,10 | 0,07 | -1,44 | 1,50E-01 | 3,57E-01 |
| MARCHF7 | 7954,75 | 0,09 | 0,07 | 1,44 | 1,50E-01 | 3,57E-01 |
| MATCAP1 | 84,68 | -0,14 | 0,10 | -1,44 | 1,50E-01 | 3,57E-01 |
| STRN3 | 1369,19 | 0,07 | 0,05 | 1,44 | 1,50E-01 | 3,57E-01 |
| PTPN18 | 1405,50 | -0,10 | 0,07 | -1,44 | 1,50E-01 | 3,58E-01 |
| SLC4A1AP | 851,93 | 0,05 | 0,03 | 1,44 | 1,50E-01 | 3,58E-01 |
| NIT2 | 656,54 | 0,06 | 0,04 | 1,44 | 1,50E-01 | 3,58E-01 |
| DNAJA4 | 1124,43 | -0,15 | 0,10 | -1,44 | 1,50E-01 | 3,58E-01 |
| TCF7L2 | 552,36 | -0,11 | 0,08 | -1,44 | 1,50E-01 | 3,58E-01 |
| ST3GAL2 | 2399,52 | -0,10 | 0,07 | -1,44 | 1,50E-01 | 3,58E-01 |
| ZNF85 | 229,69 | 0,09 | 0,07 | 1,44 | 1,50E-01 | 3,58E-01 |
| CCP110 | 900,73 | 0,08 | 0,06 | 1,44 | 1,51E-01 | 3,58E-01 |
| RPS13 | 6147,60 | 0,13 | 0,09 | 1,44 | 1,51E-01 | 3,58E-01 |
| UBE2H | 6379,62 | -0,13 | 0,09 | -1,44 | 1,50E-01 | 3,58E-01 |
| DENND6A | 2084,60 | 0,05 | 0,04 | 1,44 | 1,50E-01 | 3,58E-01 |
| NANP | 126,22 | 0,08 | 0,06 | 1,44 | 1,51E-01 | 3,58E-01 |
| PYCARD | 1575,88 | -0,09 | 0,06 | -1,44 | 1,50E-01 | 3,58E-01 |
| NLRX1 | 336,07 | -0,13 | 0,09 | -1,44 | 1,51E-01 | 3,58E-01 |

| | | | | | | |
|---|---|---|---|---|---|---|
| BOP1 | 101,12 | -0,15 | 0,11 | -1,44 | 1,51E-01 | 3,58E-01 |
| USP31 | 411,42 | -0,07 | 0,05 | -1,44 | 1,51E-01 | 3,58E-01 |
| MYL6B | 44,01 | -0,15 | 0,10 | -1,44 | 1,51E-01 | 3,58E-01 |
| POLDIP3 | 1759,83 | -0,06 | 0,04 | -1,44 | 1,51E-01 | 3,58E-01 |
| EEPD1 | 728,25 | -0,12 | 0,09 | -1,44 | 1,51E-01 | 3,58E-01 |
| PRDX4 | 227,40 | 0,11 | 0,08 | 1,44 | 1,51E-01 | 3,58E-01 |
| PPIF | 1007,95 | -0,08 | 0,06 | -1,44 | 1,51E-01 | 3,58E-01 |
| ZCCHC8 | 954,04 | 0,05 | 0,03 | 1,44 | 1,51E-01 | 3,58E-01 |
| CLSTN1 | 627,12 | -0,14 | 0,10 | -1,44 | 1,51E-01 | 3,58E-01 |
| ADM | 937,31 | -0,15 | 0,10 | -1,44 | 1,51E-01 | 3,59E-01 |
| RGCC | 997,25 | -0,11 | 0,08 | -1,43 | 1,51E-01 | 3,59E-01 |
| MKNK2 | 2618,58 | -0,15 | 0,10 | -1,43 | 1,51E-01 | 3,59E-01 |
| DXO | 101,98 | -0,08 | 0,06 | -1,43 | 1,51E-01 | 3,59E-01 |
| SSH1 | 1373,47 | -0,12 | 0,08 | -1,43 | 1,52E-01 | 3,59E-01 |
| MED23 | 3252,11 | 0,07 | 0,05 | 1,43 | 1,52E-01 | 3,59E-01 |
| PXN | 2441,07 | -0,20 | 0,14 | -1,43 | 1,52E-01 | 3,59E-01 |
| ENDOV | 215,86 | -0,11 | 0,07 | -1,43 | 1,52E-01 | 3,59E-01 |
| DENND5A | 6003,94 | -0,09 | 0,07 | -1,43 | 1,52E-01 | 3,60E-01 |
| TBCA | 408,87 | 0,08 | 0,05 | 1,43 | 1,52E-01 | 3,60E-01 |
| CBX4 | 478,73 | -0,09 | 0,06 | -1,43 | 1,52E-01 | 3,60E-01 |
| TBC1D9 | 1297,28 | 0,08 | 0,06 | 1,43 | 1,52E-01 | 3,60E-01 |
| SOX12 | 51,40 | -0,22 | 0,16 | -1,43 | 1,52E-01 | 3,60E-01 |
| PGS1 | 4004,06 | -0,10 | 0,07 | -1,43 | 1,52E-01 | 3,60E-01 |
| NOL7 | 774,44 | 0,07 | 0,05 | 1,43 | 1,52E-01 | 3,60E-01 |
| ZNF225 | 420,12 | 0,07 | 0,05 | 1,43 | 1,52E-01 | 3,60E-01 |
| TESK1 | 141,74 | -0,12 | 0,09 | -1,43 | 1,52E-01 | 3,60E-01 |
| ZSCAN21 | 148,77 | 0,05 | 0,03 | 1,43 | 1,52E-01 | 3,60E-01 |
| COL4A4 | 163,59 | -0,21 | 0,15 | -1,43 | 1,52E-01 | 3,60E-01 |
| FKBP1B | 77,39 | -0,17 | 0,12 | -1,43 | 1,52E-01 | 3,60E-01 |
| LPAR2 | 608,07 | -0,19 | 0,13 | -1,43 | 1,52E-01 | 3,60E-01 |
| PTPDC1 | 282,40 | 0,08 | 0,06 | 1,43 | 1,52E-01 | 3,60E-01 |
| SAMSN1 | 1240,44 | 0,10 | 0,07 | 1,43 | 1,53E-01 | 3,60E-01 |
| RAB8A | 1295,80 | -0,07 | 0,05 | -1,43 | 1,53E-01 | 3,60E-01 |
| DYNLT2B | 22,17 | -0,14 | 0,10 | -1,43 | 1,53E-01 | 3,60E-01 |
| SPNS1 | 29,17 | -0,27 | 0,19 | -1,43 | 1,53E-01 | 3,60E-01 |
| INTS3 | 1434,09 | -0,10 | 0,07 | -1,43 | 1,53E-01 | 3,60E-01 |
| PCK2 | 200,76 | -0,13 | 0,09 | -1,43 | 1,53E-01 | 3,60E-01 |
| CPNE3 | 2714,97 | 0,08 | 0,05 | 1,43 | 1,53E-01 | 3,61E-01 |
| HELZ2 | 2194,22 | -0,17 | 0,12 | -1,43 | 1,53E-01 | 3,61E-01 |
| INO80D | 4936,85 | 0,06 | 0,04 | 1,43 | 1,53E-01 | 3,61E-01 |
| BCL9 | 140,50 | -0,15 | 0,10 | -1,43 | 1,53E-01 | 3,61E-01 |
| THRA | 378,85 | -0,11 | 0,08 | -1,43 | 1,53E-01 | 3,61E-01 |
| KNOP1 | 539,33 | 0,06 | 0,04 | 1,43 | 1,53E-01 | 3,61E-01 |
| CEP78 | 2075,61 | 0,12 | 0,09 | 1,43 | 1,53E-01 | 3,61E-01 |
| DVL3 | 357,43 | -0,14 | 0,10 | -1,43 | 1,53E-01 | 3,61E-01 |
| RBM43 | 441,13 | 0,08 | 0,05 | 1,43 | 1,53E-01 | 3,61E-01 |

| | | | | | | |
|---|---|---|---|---|---|---|
| VPS4A | 503,41 | -0,05 | 0,03 | -1,43 | 1,53E-01 | 3,61E-01 |
| RNF141 | 3696,46 | 0,10 | 0,07 | 1,43 | 1,53E-01 | 3,61E-01 |
| ITGAM | 2572,66 | -0,14 | 0,10 | -1,43 | 1,53E-01 | 3,61E-01 |
| KRAS | 2649,80 | 0,08 | 0,05 | 1,43 | 1,53E-01 | 3,61E-01 |
| IL17RA | 6111,25 | -0,15 | 0,10 | -1,43 | 1,53E-01 | 3,61E-01 |
| CERT1 | 4376,84 | 0,06 | 0,04 | 1,43 | 1,53E-01 | 3,61E-01 |
| RSU1 | 1949,72 | -0,09 | 0,06 | -1,43 | 1,54E-01 | 3,61E-01 |
| TMEM144 | 395,30 | 0,14 | 0,10 | 1,43 | 1,54E-01 | 3,61E-01 |
| PDE3A | 93,11 | -0,21 | 0,15 | -1,43 | 1,54E-01 | 3,61E-01 |
| RBM18 | 715,41 | 0,07 | 0,05 | 1,43 | 1,54E-01 | 3,61E-01 |
| GXYLT1 | 995,90 | 0,08 | 0,06 | 1,43 | 1,54E-01 | 3,61E-01 |
| RPS14 | 24556,62 | 0,15 | 0,11 | 1,43 | 1,54E-01 | 3,61E-01 |
| BOD1L1 | 12437,33 | 0,08 | 0,06 | 1,43 | 1,54E-01 | 3,61E-01 |
| DENND1C | 461,57 | -0,16 | 0,11 | -1,43 | 1,54E-01 | 3,61E-01 |
| TFG | 1400,56 | 0,07 | 0,05 | 1,43 | 1,54E-01 | 3,61E-01 |
| SMYD2 | 1038,64 | 0,06 | 0,04 | 1,43 | 1,54E-01 | 3,61E-01 |
| CIRBP | 8422,61 | 0,06 | 0,04 | 1,43 | 1,54E-01 | 3,61E-01 |
| ATF7 | 450,80 | -0,15 | 0,10 | -1,43 | 1,54E-01 | 3,61E-01 |
| COQ9 | 544,52 | 0,03 | 0,02 | 1,43 | 1,54E-01 | 3,61E-01 |
| LSMEM1 | 304,85 | -0,11 | 0,07 | -1,43 | 1,54E-01 | 3,61E-01 |
| ZIK1 | 257,28 | 0,09 | 0,06 | 1,42 | 1,55E-01 | 3,62E-01 |
| MRPL9 | 386,07 | 0,06 | 0,04 | 1,42 | 1,54E-01 | 3,62E-01 |
| CCDC15 | 230,76 | 0,12 | 0,08 | 1,42 | 1,55E-01 | 3,62E-01 |
| GCOM1 | 86,19 | -0,23 | 0,16 | -1,42 | 1,55E-01 | 3,62E-01 |
| FBXW9 | 56,62 | -0,11 | 0,08 | -1,42 | 1,55E-01 | 3,62E-01 |
| SKIC2 | 865,15 | -0,10 | 0,07 | -1,42 | 1,54E-01 | 3,62E-01 |
| SLC38A9 | 1035,46 | 0,06 | 0,04 | 1,42 | 1,55E-01 | 3,62E-01 |
| PI4K2A | 284,50 | -0,07 | 0,05 | -1,42 | 1,54E-01 | 3,62E-01 |
| ABCC1 | 1224,41 | -0,13 | 0,09 | -1,42 | 1,55E-01 | 3,62E-01 |
| LRR1 | 356,76 | 0,09 | 0,06 | 1,42 | 1,55E-01 | 3,62E-01 |
| RUFY1 | 2321,23 | -0,08 | 0,06 | -1,42 | 1,54E-01 | 3,62E-01 |
| LDLR | 281,60 | -0,13 | 0,09 | -1,42 | 1,55E-01 | 3,62E-01 |
| WAS | 820,68 | -0,16 | 0,11 | -1,42 | 1,54E-01 | 3,62E-01 |
| SAXO4 | 33,33 | -0,24 | 0,17 | -1,42 | 1,54E-01 | 3,62E-01 |
| REDIC1 | 36,11 | -0,21 | 0,15 | -1,42 | 1,55E-01 | 3,62E-01 |
| ZNF775 | 62,69 | -0,11 | 0,08 | -1,42 | 1,55E-01 | 3,62E-01 |
| MTHFD1L | 135,84 | -0,17 | 0,12 | -1,42 | 1,55E-01 | 3,62E-01 |
| SLC35A1 | 201,04 | 0,15 | 0,11 | 1,42 | 1,54E-01 | 3,62E-01 |
| NCAPD2 | 1038,07 | -0,08 | 0,05 | -1,42 | 1,54E-01 | 3,62E-01 |
| EXOC3 | 1219,13 | -0,11 | 0,08 | -1,42 | 1,54E-01 | 3,62E-01 |
| ACTMAP | 41,90 | -0,19 | 0,13 | -1,42 | 1,55E-01 | 3,62E-01 |
| DENND1A | 1313,42 | -0,09 | 0,07 | -1,42 | 1,55E-01 | 3,62E-01 |
| CYB5R1 | 729,34 | -0,08 | 0,06 | -1,42 | 1,55E-01 | 3,62E-01 |
| TMEM209 | 348,64 | 0,07 | 0,05 | 1,42 | 1,55E-01 | 3,62E-01 |
| LILRA2 | 2028,03 | -0,11 | 0,08 | -1,42 | 1,55E-01 | 3,62E-01 |
| SPR | 33,26 | -0,18 | 0,12 | -1,42 | 1,55E-01 | 3,62E-01 |

| | | | | | | |
|---|---|---|---|---|---|---|
| ZNF562 | 799,73 | 0,05 | 0,04 | 1,42 | 1,55E-01 | 3,62E-01 |
| KDM5A | 12197,96 | 0,06 | 0,05 | 1,42 | 1,55E-01 | 3,62E-01 |
| BABAM1 | 717,85 | -0,11 | 0,08 | -1,42 | 1,55E-01 | 3,62E-01 |
| TACC3 | 1624,74 | -0,13 | 0,09 | -1,42 | 1,55E-01 | 3,62E-01 |
| SLC22A4 | 1218,71 | -0,12 | 0,08 | -1,42 | 1,55E-01 | 3,62E-01 |
| TCAIM | 670,90 | 0,07 | 0,05 | 1,42 | 1,55E-01 | 3,62E-01 |
| RAMACL | 55,35 | -0,15 | 0,11 | -1,42 | 1,55E-01 | 3,62E-01 |
| TMEM81 | 138,82 | -0,06 | 0,04 | -1,42 | 1,55E-01 | 3,62E-01 |
| NGLY1 | 2618,43 | 0,07 | 0,05 | 1,42 | 1,55E-01 | 3,62E-01 |
| PDK2 | 259,71 | -0,13 | 0,09 | -1,42 | 1,55E-01 | 3,62E-01 |
| ZC3H14 | 1298,41 | 0,05 | 0,03 | 1,42 | 1,55E-01 | 3,62E-01 |
| ATP6V1H | 1170,56 | 0,05 | 0,04 | 1,42 | 1,56E-01 | 3,63E-01 |
| ORAI1 | 453,52 | -0,09 | 0,06 | -1,42 | 1,56E-01 | 3,63E-01 |
| TRAPPC12 | 2275,52 | -0,08 | 0,05 | -1,42 | 1,56E-01 | 3,63E-01 |
| ERO1B | 1191,49 | 0,07 | 0,05 | 1,42 | 1,56E-01 | 3,63E-01 |
| PLD4 | 233,63 | -0,13 | 0,09 | -1,42 | 1,56E-01 | 3,63E-01 |
| PWWP2A | 1746,68 | 0,04 | 0,03 | 1,42 | 1,56E-01 | 3,63E-01 |
| CDC123 | 1734,52 | 0,07 | 0,05 | 1,42 | 1,56E-01 | 3,63E-01 |
| ZNF540 | 314,37 | 0,11 | 0,08 | 1,42 | 1,56E-01 | 3,63E-01 |
| TMIGD2 | 183,56 | 0,14 | 0,10 | 1,42 | 1,56E-01 | 3,63E-01 |
| MMP28 | 46,12 | -0,24 | 0,17 | -1,42 | 1,56E-01 | 3,63E-01 |
| PHF7 | 190,22 | -0,06 | 0,04 | -1,42 | 1,56E-01 | 3,63E-01 |
| TSPAN14 | 3822,78 | -0,09 | 0,07 | -1,42 | 1,56E-01 | 3,63E-01 |
| TENT4B | 1116,35 | 0,06 | 0,04 | 1,42 | 1,56E-01 | 3,63E-01 |
| SUCO | 2383,99 | 0,10 | 0,07 | 1,42 | 1,56E-01 | 3,63E-01 |
| FUT8 | 987,05 | 0,08 | 0,05 | 1,42 | 1,56E-01 | 3,63E-01 |
| NSUN2 | 2203,67 | 0,06 | 0,04 | 1,42 | 1,56E-01 | 3,63E-01 |
| GIT1 | 353,23 | -0,14 | 0,10 | -1,42 | 1,56E-01 | 3,63E-01 |
| TRIM9 | 76,36 | -0,22 | 0,16 | -1,42 | 1,56E-01 | 3,63E-01 |
| WDR74 | 465,14 | 0,05 | 0,03 | 1,42 | 1,56E-01 | 3,63E-01 |
| SETD2 | 4450,28 | 0,05 | 0,04 | 1,42 | 1,56E-01 | 3,63E-01 |
| SMPD4 | 407,76 | -0,10 | 0,07 | -1,42 | 1,56E-01 | 3,63E-01 |
| ITGAX | 6208,68 | -0,20 | 0,14 | -1,42 | 1,57E-01 | 3,63E-01 |
| PPP1R12A | 7987,01 | 0,10 | 0,07 | 1,42 | 1,57E-01 | 3,63E-01 |
| TNKS2 | 5112,18 | 0,06 | 0,04 | 1,42 | 1,57E-01 | 3,63E-01 |
| PLXNA2 | 410,01 | -0,14 | 0,10 | -1,42 | 1,57E-01 | 3,63E-01 |
| MARK2 | 1232,64 | -0,14 | 0,10 | -1,42 | 1,57E-01 | 3,64E-01 |
| RMC1 | 1321,43 | -0,05 | 0,03 | -1,42 | 1,57E-01 | 3,64E-01 |
| GLRX3 | 873,73 | 0,04 | 0,03 | 1,42 | 1,57E-01 | 3,64E-01 |
| RACK1 | 13962,55 | 0,08 | 0,05 | 1,41 | 1,57E-01 | 3,64E-01 |
| MFSD8 | 1421,11 | 0,06 | 0,04 | 1,42 | 1,57E-01 | 3,64E-01 |
| ST3GAL4 | 651,34 | -0,13 | 0,09 | -1,42 | 1,57E-01 | 3,64E-01 |
| LIG1 | 326,54 | -0,12 | 0,09 | -1,41 | 1,57E-01 | 3,64E-01 |
| FBF1 | 135,42 | -0,07 | 0,05 | -1,42 | 1,57E-01 | 3,64E-01 |
| ZBTB20 | 3653,60 | 0,07 | 0,05 | 1,41 | 1,57E-01 | 3,64E-01 |
| MAK | 642,60 | -0,13 | 0,09 | -1,41 | 1,57E-01 | 3,64E-01 |

| | | | | | | |
|---|---|---|---|---|---|---|
| ACBD4 | 124,92 | -0,13 | 0,09 | -1,41 | 1,57E-01 | 3,64E-01 |
| ZSCAN31 | 36,30 | -0,21 | 0,15 | -1,41 | 1,57E-01 | 3,64E-01 |
| DFFB | 429,46 | -0,05 | 0,04 | -1,41 | 1,57E-01 | 3,64E-01 |
| GPRIN3 | 4424,58 | 0,10 | 0,07 | 1,41 | 1,57E-01 | 3,64E-01 |
| DTNB | 361,33 | -0,09 | 0,06 | -1,41 | 1,57E-01 | 3,64E-01 |
| AMOTL1 | 111,66 | -0,20 | 0,14 | -1,41 | 1,57E-01 | 3,64E-01 |
| CD300LF | 1286,85 | -0,09 | 0,07 | -1,41 | 1,58E-01 | 3,64E-01 |
| COL9A3 | 220,12 | -0,23 | 0,16 | -1,41 | 1,58E-01 | 3,64E-01 |
| FAM169A | 627,57 | 0,12 | 0,08 | 1,41 | 1,57E-01 | 3,64E-01 |
| MTRR | 1205,78 | 0,06 | 0,04 | 1,41 | 1,57E-01 | 3,64E-01 |
| CZIB | 630,44 | 0,04 | 0,03 | 1,41 | 1,58E-01 | 3,64E-01 |
| TAF4 | 411,96 | -0,13 | 0,09 | -1,41 | 1,58E-01 | 3,64E-01 |
| MXI1 | 7142,62 | -0,17 | 0,12 | -1,41 | 1,58E-01 | 3,64E-01 |
| ZMYM5 | 818,92 | 0,08 | 0,05 | 1,41 | 1,58E-01 | 3,64E-01 |
| SLC50A1 | 373,20 | -0,06 | 0,04 | -1,41 | 1,58E-01 | 3,64E-01 |
| ARF3 | 1118,66 | -0,13 | 0,09 | -1,41 | 1,58E-01 | 3,65E-01 |
| CLEC1B | 114,80 | -0,19 | 0,13 | -1,41 | 1,58E-01 | 3,65E-01 |
| DR1 | 4151,42 | 0,08 | 0,05 | 1,41 | 1,58E-01 | 3,65E-01 |
| C19orf38 | 873,85 | -0,09 | 0,06 | -1,41 | 1,58E-01 | 3,65E-01 |
| EPS15 | 4537,42 | 0,06 | 0,04 | 1,41 | 1,58E-01 | 3,65E-01 |
| CDC42SE2 | 9590,76 | 0,08 | 0,06 | 1,41 | 1,58E-01 | 3,65E-01 |
| HACD3 | 604,85 | 0,06 | 0,04 | 1,41 | 1,58E-01 | 3,65E-01 |
| DYTN | 34,37 | -0,22 | 0,16 | -1,41 | 1,58E-01 | 3,65E-01 |
| ATAD2B | 4424,53 | 0,07 | 0,05 | 1,41 | 1,58E-01 | 3,65E-01 |
| PJA2 | 7306,89 | 0,09 | 0,06 | 1,41 | 1,59E-01 | 3,65E-01 |
| ZNF638 | 8184,07 | 0,09 | 0,06 | 1,41 | 1,59E-01 | 3,65E-01 |
| TMEM170A | 1593,14 | 0,05 | 0,03 | 1,41 | 1,59E-01 | 3,65E-01 |
| PSMD13 | 1729,42 | 0,05 | 0,04 | 1,41 | 1,59E-01 | 3,65E-01 |
| TMEM120A | 411,44 | -0,09 | 0,06 | -1,41 | 1,59E-01 | 3,65E-01 |
| RGL4 | 595,10 | -0,10 | 0,07 | -1,41 | 1,59E-01 | 3,65E-01 |
| IL1B | 942,27 | -0,11 | 0,08 | -1,41 | 1,59E-01 | 3,65E-01 |
| ANKS3 | 257,00 | -0,10 | 0,07 | -1,41 | 1,59E-01 | 3,65E-01 |
| MBD4 | 1773,91 | 0,08 | 0,06 | 1,41 | 1,59E-01 | 3,65E-01 |
| ANLN | 73,82 | -0,20 | 0,14 | -1,41 | 1,59E-01 | 3,65E-01 |
| FBLN5 | 105,79 | -0,17 | 0,12 | -1,41 | 1,59E-01 | 3,65E-01 |
| TUBB1 | 3012,37 | -0,18 | 0,13 | -1,41 | 1,59E-01 | 3,65E-01 |
| RSPRY1 | 1218,41 | 0,04 | 0,03 | 1,41 | 1,59E-01 | 3,65E-01 |
| STX16 | 2888,69 | -0,07 | 0,05 | -1,41 | 1,59E-01 | 3,65E-01 |
| MOB4 | 678,42 | 0,09 | 0,06 | 1,41 | 1,59E-01 | 3,65E-01 |
| MCM7 | 912,81 | -0,06 | 0,04 | -1,41 | 1,59E-01 | 3,65E-01 |
| BORCS6 | 470,10 | -0,06 | 0,04 | -1,41 | 1,59E-01 | 3,65E-01 |
| IGF2BP2 | 902,84 | -0,20 | 0,14 | -1,41 | 1,59E-01 | 3,65E-01 |
| ATE1 | 953,79 | 0,05 | 0,03 | 1,41 | 1,59E-01 | 3,66E-01 |
| SURF4 | 1910,34 | -0,05 | 0,03 | -1,41 | 1,59E-01 | 3,66E-01 |
| SELP | 364,45 | -0,14 | 0,10 | -1,41 | 1,59E-01 | 3,66E-01 |
| RALY | 2133,54 | -0,07 | 0,05 | -1,41 | 1,59E-01 | 3,66E-01 |

| | | | | | | |
|---|---|---|---|---|---|---|
| FAM216A | 233,89 | 0,08 | 0,05 | 1,41 | 1,59E-01 | 3,66E-01 |
| PCNX3 | 810,10 | -0,15 | 0,10 | -1,41 | 1,59E-01 | 3,66E-01 |
| IL1R2 | 2281,93 | -0,13 | 0,10 | -1,41 | 1,60E-01 | 3,66E-01 |
| TAF15 | 1945,64 | 0,05 | 0,03 | 1,41 | 1,60E-01 | 3,66E-01 |
| RPS6KB2 | 830,25 | -0,06 | 0,04 | -1,41 | 1,60E-01 | 3,66E-01 |
| SENP6 | 5359,23 | 0,09 | 0,06 | 1,41 | 1,60E-01 | 3,66E-01 |
| CALHM5 | 41,88 | -0,23 | 0,17 | -1,41 | 1,60E-01 | 3,66E-01 |
| DENND2C | 118,46 | -0,16 | 0,11 | -1,41 | 1,60E-01 | 3,66E-01 |
| SLC35B1 | 484,75 | -0,04 | 0,03 | -1,41 | 1,60E-01 | 3,66E-01 |
| NRBP2 | 122,43 | -0,15 | 0,11 | -1,41 | 1,60E-01 | 3,66E-01 |
| MALSU1 | 413,89 | 0,06 | 0,04 | 1,41 | 1,60E-01 | 3,66E-01 |
| MRPS7 | 441,52 | 0,07 | 0,05 | 1,40 | 1,60E-01 | 3,66E-01 |
| CLK3 | 3532,63 | -0,05 | 0,03 | -1,40 | 1,60E-01 | 3,66E-01 |
| TBC1D17 | 452,10 | -0,12 | 0,09 | -1,40 | 1,60E-01 | 3,66E-01 |
| ADAM10 | 8646,82 | 0,08 | 0,06 | 1,40 | 1,60E-01 | 3,66E-01 |
| WDTC1 | 2143,89 | -0,11 | 0,08 | -1,40 | 1,60E-01 | 3,66E-01 |
| ESCO2 | 51,01 | -0,22 | 0,16 | -1,40 | 1,60E-01 | 3,66E-01 |
| PPM1H | 173,20 | -0,14 | 0,10 | -1,40 | 1,60E-01 | 3,66E-01 |
| SPPL2B | 330,52 | -0,16 | 0,11 | -1,40 | 1,60E-01 | 3,66E-01 |
| TRIM36 | 78,16 | -0,17 | 0,12 | -1,40 | 1,60E-01 | 3,66E-01 |
| SPATS2 | 148,95 | -0,11 | 0,08 | -1,40 | 1,60E-01 | 3,66E-01 |
| GOLGA8H | 32,91 | -0,17 | 0,12 | -1,40 | 1,60E-01 | 3,66E-01 |
| ADAMTS5 | 122,31 | 0,16 | 0,11 | 1,40 | 1,60E-01 | 3,66E-01 |
| ABCA5 | 2209,61 | 0,09 | 0,07 | 1,40 | 1,61E-01 | 3,67E-01 |
| ATP5PB | 2055,79 | 0,11 | 0,08 | 1,40 | 1,61E-01 | 3,67E-01 |
| PCDHGB5 | 34,97 | -0,19 | 0,13 | -1,40 | 1,61E-01 | 3,67E-01 |
| GLIPR1L2 | 34,06 | -0,20 | 0,14 | -1,40 | 1,61E-01 | 3,67E-01 |
| IFT46 | 140,38 | -0,07 | 0,05 | -1,40 | 1,61E-01 | 3,67E-01 |
| TPP1 | 996,05 | -0,12 | 0,08 | -1,40 | 1,61E-01 | 3,68E-01 |
| IRS1 | 93,93 | -0,19 | 0,13 | -1,40 | 1,61E-01 | 3,68E-01 |
| CDK5R1 | 504,94 | -0,12 | 0,08 | -1,40 | 1,61E-01 | 3,68E-01 |
| HK2 | 1975,81 | -0,10 | 0,07 | -1,40 | 1,61E-01 | 3,68E-01 |
| EZR | 3507,96 | 0,08 | 0,06 | 1,40 | 1,61E-01 | 3,68E-01 |
| SNX3 | 4114,00 | -0,14 | 0,10 | -1,40 | 1,61E-01 | 3,68E-01 |
| SCLY | 204,35 | -0,11 | 0,08 | -1,40 | 1,61E-01 | 3,68E-01 |
| CSGALNACT: | 1265,45 | -0,10 | 0,07 | -1,40 | 1,61E-01 | 3,68E-01 |
| RNF7 | 778,08 | 0,07 | 0,05 | 1,40 | 1,62E-01 | 3,68E-01 |
| NEURL4 | 498,79 | -0,07 | 0,05 | -1,40 | 1,62E-01 | 3,68E-01 |
| RXRB | 934,45 | -0,08 | 0,06 | -1,40 | 1,62E-01 | 3,68E-01 |
| PLEKHG2 | 231,67 | -0,16 | 0,11 | -1,40 | 1,62E-01 | 3,68E-01 |
| COBLL1 | 621,47 | 0,13 | 0,09 | 1,40 | 1,62E-01 | 3,68E-01 |
| CFAP70 | 196,66 | -0,11 | 0,08 | -1,40 | 1,62E-01 | 3,68E-01 |
| MKLN1 | 7626,09 | 0,08 | 0,06 | 1,40 | 1,62E-01 | 3,68E-01 |
| STON2 | 552,84 | -0,14 | 0,10 | -1,40 | 1,62E-01 | 3,68E-01 |
| NOTCH2 | 11905,65 | -0,10 | 0,07 | -1,40 | 1,62E-01 | 3,69E-01 |
| GRK3 | 2891,15 | 0,08 | 0,06 | 1,40 | 1,62E-01 | 3,69E-01 |

| | | | | | | |
|---|---|---|---|---|---|---|
| LLCFC1 | 18,00 | -0,21 | 0,15 | -1,40 | 1,62E-01 | 3,69E-01 |
| GNPDA1 | 504,60 | 0,06 | 0,04 | 1,40 | 1,62E-01 | 3,69E-01 |
| CFLAR | 34436,32 | -0,07 | 0,05 | -1,40 | 1,62E-01 | 3,69E-01 |
| L3MBTL1 | 428,84 | -0,10 | 0,07 | -1,40 | 1,62E-01 | 3,69E-01 |
| CENPO | 162,28 | -0,09 | 0,07 | -1,40 | 1,62E-01 | 3,69E-01 |
| UGGT2 | 310,91 | 0,13 | 0,09 | 1,40 | 1,63E-01 | 3,69E-01 |
| TGFBR1 | 5302,79 | 0,10 | 0,07 | 1,40 | 1,63E-01 | 3,70E-01 |
| TMT1A | 2481,08 | 0,08 | 0,06 | 1,40 | 1,63E-01 | 3,70E-01 |
| BCS1L | 230,17 | -0,06 | 0,05 | -1,39 | 1,63E-01 | 3,70E-01 |
| TOGARAM2 | 315,39 | -0,16 | 0,11 | -1,39 | 1,63E-01 | 3,70E-01 |
| SRSF8 | 1561,70 | 0,07 | 0,05 | 1,39 | 1,63E-01 | 3,70E-01 |
| DHRS3 | 351,99 | 0,08 | 0,06 | 1,39 | 1,63E-01 | 3,70E-01 |
| CCDC102A | 192,19 | -0,10 | 0,07 | -1,40 | 1,63E-01 | 3,70E-01 |
| CDKN2D | 2377,71 | -0,11 | 0,08 | -1,39 | 1,63E-01 | 3,70E-01 |
| MTARC1 | 1193,87 | -0,13 | 0,09 | -1,39 | 1,63E-01 | 3,70E-01 |
| LMF1 | 318,59 | -0,13 | 0,09 | -1,39 | 1,63E-01 | 3,70E-01 |
| LHPP | 182,12 | -0,13 | 0,09 | -1,39 | 1,63E-01 | 3,70E-01 |
| WDR44 | 548,03 | 0,06 | 0,05 | 1,39 | 1,63E-01 | 3,70E-01 |
| SLC2A13 | 429,56 | 0,07 | 0,05 | 1,39 | 1,63E-01 | 3,70E-01 |
| TRAIP | 41,11 | -0,14 | 0,10 | -1,39 | 1,63E-01 | 3,70E-01 |
| RGS7 | 31,06 | -0,22 | 0,16 | -1,39 | 1,63E-01 | 3,71E-01 |
| BAG3 | 97,55 | -0,12 | 0,08 | -1,39 | 1,64E-01 | 3,71E-01 |
| PSMG1 | 265,91 | 0,07 | 0,05 | 1,39 | 1,64E-01 | 3,71E-01 |
| GNG2 | 2984,64 | 0,06 | 0,05 | 1,39 | 1,64E-01 | 3,71E-01 |
| TEX22 | 65,78 | -0,13 | 0,09 | -1,39 | 1,64E-01 | 3,71E-01 |
| PIK3R3 | 109,76 | -0,14 | 0,10 | -1,39 | 1,64E-01 | 3,71E-01 |
| TMEM43 | 2662,18 | -0,07 | 0,05 | -1,39 | 1,64E-01 | 3,71E-01 |
| CDC42SE1 | 14851,40 | -0,06 | 0,04 | -1,39 | 1,64E-01 | 3,71E-01 |
| ZNF616 | 282,55 | 0,09 | 0,07 | 1,39 | 1,64E-01 | 3,71E-01 |
| SHMT2 | 677,15 | 0,07 | 0,05 | 1,39 | 1,64E-01 | 3,71E-01 |
| MTR | 3323,19 | 0,06 | 0,04 | 1,39 | 1,64E-01 | 3,71E-01 |
| CHN1 | 81,58 | -0,17 | 0,12 | -1,39 | 1,64E-01 | 3,72E-01 |
| STIM2 | 1828,82 | 0,06 | 0,04 | 1,39 | 1,64E-01 | 3,72E-01 |
| USP21 | 280,08 | -0,08 | 0,05 | -1,39 | 1,64E-01 | 3,72E-01 |
| ZCCHC17 | 598,70 | 0,06 | 0,05 | 1,39 | 1,64E-01 | 3,72E-01 |
| TRAPPC6A | 351,78 | 0,06 | 0,05 | 1,39 | 1,64E-01 | 3,72E-01 |
| AMY2B | 666,85 | 0,07 | 0,05 | 1,39 | 1,64E-01 | 3,72E-01 |
| BTN3A3 | 2843,53 | -0,07 | 0,05 | -1,39 | 1,64E-01 | 3,72E-01 |
| ZKSCAN5 | 594,93 | 0,04 | 0,03 | 1,39 | 1,65E-01 | 3,72E-01 |
| H2BC21 | 2842,36 | -0,09 | 0,06 | -1,39 | 1,65E-01 | 3,72E-01 |
| TCEA2 | 155,93 | -0,12 | 0,08 | -1,39 | 1,65E-01 | 3,72E-01 |
| PIK3CD | 4807,82 | -0,13 | 0,09 | -1,39 | 1,65E-01 | 3,72E-01 |
| AGPAT3 | 1377,44 | -0,10 | 0,07 | -1,39 | 1,65E-01 | 3,73E-01 |
| FPR2 | 5571,45 | -0,12 | 0,09 | -1,39 | 1,65E-01 | 3,73E-01 |
| IVNS1ABP | 6788,62 | 0,08 | 0,06 | 1,39 | 1,65E-01 | 3,73E-01 |
| LDLRAD3 | 90,17 | -0,15 | 0,11 | -1,39 | 1,65E-01 | 3,73E-01 |

| | | | | | | |
|---|---|---|---|---|---|---|
| CCL4L2 | 100,44 | -0,21 | 0,15 | -1,39 | 1,66E-01 | 3,73E-01 |
| ADCY7 | 6913,82 | -0,07 | 0,05 | -1,39 | 1,66E-01 | 3,73E-01 |
| ARHGAP29 | 90,26 | -0,23 | 0,17 | -1,39 | 1,66E-01 | 3,74E-01 |
| PREB | 430,85 | -0,06 | 0,05 | -1,39 | 1,66E-01 | 3,74E-01 |
| TBX19 | 140,09 | -0,13 | 0,09 | -1,39 | 1,66E-01 | 3,74E-01 |
| NEMP2 | 730,56 | 0,08 | 0,06 | 1,39 | 1,66E-01 | 3,74E-01 |
| FCRL5 | 835,62 | 0,23 | 0,16 | 1,39 | 1,66E-01 | 3,74E-01 |
| HDAC3 | 630,35 | -0,06 | 0,04 | -1,38 | 1,66E-01 | 3,74E-01 |
| ARGLU1 | 8675,57 | 0,10 | 0,07 | 1,38 | 1,66E-01 | 3,74E-01 |
| NUAK2 | 1587,59 | -0,12 | 0,09 | -1,38 | 1,66E-01 | 3,75E-01 |
| MBOAT7 | 1789,97 | -0,16 | 0,12 | -1,38 | 1,66E-01 | 3,75E-01 |
| BAZ2B | 11550,83 | 0,09 | 0,07 | 1,38 | 1,66E-01 | 3,75E-01 |
| FCN1 | 20249,54 | 0,10 | 0,07 | 1,38 | 1,66E-01 | 3,75E-01 |
| NUMB | 4191,02 | -0,09 | 0,07 | -1,38 | 1,67E-01 | 3,75E-01 |
| CNOT10 | 1125,77 | 0,04 | 0,03 | 1,38 | 1,67E-01 | 3,75E-01 |
| SRRD | 940,51 | -0,13 | 0,09 | -1,38 | 1,67E-01 | 3,75E-01 |
| SF3B6 | 441,21 | 0,15 | 0,11 | 1,38 | 1,67E-01 | 3,75E-01 |
| CCNL1 | 9185,00 | 0,08 | 0,06 | 1,38 | 1,67E-01 | 3,75E-01 |
| TRAPPC8 | 4842,76 | 0,07 | 0,05 | 1,38 | 1,67E-01 | 3,76E-01 |
| TMEM241 | 121,50 | -0,13 | 0,10 | -1,38 | 1,67E-01 | 3,76E-01 |
| NIPA2 | 1295,24 | 0,05 | 0,04 | 1,38 | 1,67E-01 | 3,76E-01 |
| FANCL | 220,00 | 0,13 | 0,09 | 1,38 | 1,67E-01 | 3,76E-01 |
| FZD6 | 118,49 | 0,15 | 0,11 | 1,38 | 1,67E-01 | 3,76E-01 |
| CLDN23 | 39,42 | -0,19 | 0,14 | -1,38 | 1,67E-01 | 3,76E-01 |
| ZNF267 | 2743,25 | 0,12 | 0,08 | 1,38 | 1,67E-01 | 3,76E-01 |
| POLE | 874,93 | -0,09 | 0,06 | -1,38 | 1,67E-01 | 3,76E-01 |
| H3C11 | 484,92 | -0,10 | 0,07 | -1,38 | 1,67E-01 | 3,76E-01 |
| CTNNBL1 | 1162,02 | 0,06 | 0,04 | 1,38 | 1,67E-01 | 3,76E-01 |
| TCEA1 | 2204,49 | 0,09 | 0,06 | 1,38 | 1,67E-01 | 3,76E-01 |
| RNPEPL1 | 794,93 | -0,18 | 0,13 | -1,38 | 1,67E-01 | 3,76E-01 |
| MIER1 | 4721,28 | 0,11 | 0,08 | 1,38 | 1,68E-01 | 3,76E-01 |
| LRCH4 | 1033,35 | -0,09 | 0,07 | -1,38 | 1,68E-01 | 3,76E-01 |
| NEO1 | 261,15 | -0,13 | 0,09 | -1,38 | 1,68E-01 | 3,76E-01 |
| CERS5 | 888,73 | -0,05 | 0,03 | -1,38 | 1,68E-01 | 3,76E-01 |
| EIF2B2 | 526,59 | 0,05 | 0,03 | 1,38 | 1,68E-01 | 3,76E-01 |
| RNF10 | 10762,52 | -0,18 | 0,13 | -1,38 | 1,68E-01 | 3,76E-01 |
| TOR4A | 375,84 | -0,14 | 0,10 | -1,38 | 1,68E-01 | 3,76E-01 |
| MGAM2 | 522,07 | -0,16 | 0,11 | -1,38 | 1,68E-01 | 3,76E-01 |
| FN3KRP | 335,08 | -0,07 | 0,05 | -1,38 | 1,68E-01 | 3,76E-01 |
| ARSK | 208,26 | 0,09 | 0,06 | 1,38 | 1,68E-01 | 3,76E-01 |
| CAPZA2 | 4386,55 | 0,15 | 0,11 | 1,38 | 1,68E-01 | 3,76E-01 |
| QNG1 | 341,00 | 0,06 | 0,04 | 1,38 | 1,68E-01 | 3,76E-01 |
| RBL1 | 950,52 | 0,08 | 0,06 | 1,38 | 1,68E-01 | 3,76E-01 |
| ATP8B4 | 1693,74 | 0,11 | 0,08 | 1,38 | 1,68E-01 | 3,76E-01 |
| RICTOR | 13174,71 | 0,10 | 0,07 | 1,38 | 1,68E-01 | 3,77E-01 |
| SQSTM1 | 5159,65 | -0,07 | 0,05 | -1,38 | 1,68E-01 | 3,77E-01 |

| | | | | | | |
|---|---|---|---|---|---|---|
| ENSG000002 | 35,10 | -0,15 | 0,11 | -1,38 | 1,68E-01 | 3,77E-01 |
| NOL3 | 63,80 | -0,16 | 0,11 | -1,38 | 1,68E-01 | 3,77E-01 |
| PPP5C | 591,20 | -0,08 | 0,06 | -1,38 | 1,69E-01 | 3,77E-01 |
| PGLYRP1 | 99,62 | -0,16 | 0,12 | -1,38 | 1,69E-01 | 3,77E-01 |
| SLC36A4 | 1207,96 | 0,11 | 0,08 | 1,38 | 1,69E-01 | 3,77E-01 |
| SPTSSA | 271,94 | 0,10 | 0,08 | 1,38 | 1,69E-01 | 3,77E-01 |
| BMP6 | 196,93 | -0,14 | 0,10 | -1,38 | 1,69E-01 | 3,77E-01 |
| CXCL5 | 318,76 | -0,18 | 0,13 | -1,38 | 1,69E-01 | 3,77E-01 |
| LRRC8C | 2337,41 | 0,08 | 0,06 | 1,38 | 1,69E-01 | 3,78E-01 |
| RMND5B | 509,87 | -0,07 | 0,05 | -1,37 | 1,69E-01 | 3,78E-01 |
| PCMTD2 | 3183,57 | 0,06 | 0,04 | 1,37 | 1,69E-01 | 3,78E-01 |
| AMBRA1 | 956,68 | -0,08 | 0,06 | -1,37 | 1,69E-01 | 3,78E-01 |
| SNRPB2 | 611,65 | 0,10 | 0,07 | 1,37 | 1,69E-01 | 3,78E-01 |
| SPATA20 | 218,42 | -0,16 | 0,11 | -1,37 | 1,70E-01 | 3,78E-01 |
| SPRYD3 | 375,89 | -0,09 | 0,07 | -1,37 | 1,70E-01 | 3,79E-01 |
| ZNF419 | 350,97 | 0,06 | 0,04 | 1,37 | 1,70E-01 | 3,79E-01 |
| LYPLAL1 | 962,39 | 0,12 | 0,09 | 1,37 | 1,70E-01 | 3,79E-01 |
| DENND11 | 3048,54 | 0,07 | 0,05 | 1,37 | 1,70E-01 | 3,80E-01 |
| SRSF4 | 3923,51 | 0,05 | 0,04 | 1,37 | 1,70E-01 | 3,80E-01 |
| NOMO2 | 429,47 | -0,08 | 0,06 | -1,37 | 1,70E-01 | 3,80E-01 |
| CARMIL2 | 309,63 | -0,17 | 0,13 | -1,37 | 1,70E-01 | 3,80E-01 |
| DRICH1 | 48,65 | -0,13 | 0,10 | -1,37 | 1,71E-01 | 3,80E-01 |
| LMBR1 | 2436,21 | 0,05 | 0,04 | 1,37 | 1,71E-01 | 3,80E-01 |
| PROX2 | 25,37 | -0,22 | 0,16 | -1,37 | 1,71E-01 | 3,80E-01 |
| WASHC5 | 1597,35 | 0,05 | 0,03 | 1,37 | 1,71E-01 | 3,80E-01 |
| TRIM33 | 5171,87 | 0,07 | 0,05 | 1,37 | 1,71E-01 | 3,80E-01 |
| ENSG000002 | 259,00 | 0,08 | 0,06 | 1,37 | 1,71E-01 | 3,80E-01 |
| NCLN | 452,66 | -0,15 | 0,11 | -1,37 | 1,71E-01 | 3,80E-01 |
| PRR14 | 394,17 | -0,10 | 0,07 | -1,37 | 1,71E-01 | 3,81E-01 |
| FAM161A | 45,75 | -0,17 | 0,13 | -1,37 | 1,71E-01 | 3,81E-01 |
| COL13A1 | 86,07 | -0,28 | 0,21 | -1,37 | 1,71E-01 | 3,81E-01 |
| ZNF622 | 1047,56 | 0,08 | 0,06 | 1,37 | 1,71E-01 | 3,81E-01 |
| IKBKG | 306,65 | -0,10 | 0,07 | -1,37 | 1,71E-01 | 3,81E-01 |
| CRYZ | 292,63 | 0,15 | 0,11 | 1,37 | 1,71E-01 | 3,81E-01 |
| FCRL1 | 1976,36 | 0,17 | 0,12 | 1,37 | 1,71E-01 | 3,81E-01 |
| MECP2 | 2413,43 | -0,11 | 0,08 | -1,37 | 1,71E-01 | 3,81E-01 |
| TMC8 | 2612,36 | -0,14 | 0,10 | -1,37 | 1,71E-01 | 3,81E-01 |
| PCBP1 | 2222,51 | -0,14 | 0,10 | -1,37 | 1,71E-01 | 3,81E-01 |
| FGD3 | 3714,51 | -0,15 | 0,11 | -1,37 | 1,71E-01 | 3,81E-01 |
| PEDS1 | 240,22 | -0,10 | 0,08 | -1,37 | 1,71E-01 | 3,81E-01 |
| COG5 | 2382,51 | 0,08 | 0,06 | 1,37 | 1,72E-01 | 3,81E-01 |
| SLC1A4 | 162,33 | -0,11 | 0,08 | -1,37 | 1,72E-01 | 3,81E-01 |
| RHOU | 599,18 | 0,09 | 0,07 | 1,37 | 1,72E-01 | 3,82E-01 |
| NHEJ1 | 57,57 | -0,10 | 0,07 | -1,37 | 1,72E-01 | 3,82E-01 |
| PLPP5 | 500,19 | 0,11 | 0,08 | 1,37 | 1,72E-01 | 3,82E-01 |
| RBBP6 | 2651,74 | 0,05 | 0,04 | 1,37 | 1,72E-01 | 3,82E-01 |

| | | | | | | |
|---|---|---|---|---|---|---|
| POP4 | 594,11 | 0,04 | 0,03 | 1,37 | 1,72E-01 | 3,82E-01 |
| TMEM121B | 177,24 | -0,19 | 0,14 | -1,37 | 1,72E-01 | 3,82E-01 |
| UPRT | 584,30 | 0,07 | 0,05 | 1,37 | 1,72E-01 | 3,82E-01 |
| GDE1 | 1678,37 | -0,09 | 0,06 | -1,36 | 1,72E-01 | 3,82E-01 |
| NRG1 | 281,42 | 0,23 | 0,17 | 1,36 | 1,72E-01 | 3,82E-01 |
| AP3M2 | 872,18 | 0,07 | 0,05 | 1,36 | 1,73E-01 | 3,82E-01 |
| CSNK2B | 196,19 | -0,08 | 0,06 | -1,36 | 1,73E-01 | 3,82E-01 |
| HIBCH | 929,54 | 0,06 | 0,05 | 1,36 | 1,73E-01 | 3,82E-01 |
| FAM193A | 1020,54 | -0,11 | 0,08 | -1,36 | 1,73E-01 | 3,82E-01 |
| RAB2B | 4378,94 | -0,15 | 0,11 | -1,36 | 1,73E-01 | 3,83E-01 |
| RGS10 | 1744,10 | -0,14 | 0,10 | -1,36 | 1,73E-01 | 3,83E-01 |
| FRY | 8521,13 | -0,09 | 0,07 | -1,36 | 1,73E-01 | 3,83E-01 |
| KCTD2 | 603,19 | -0,06 | 0,05 | -1,36 | 1,73E-01 | 3,83E-01 |
| ACVR2A | 475,72 | 0,09 | 0,07 | 1,36 | 1,73E-01 | 3,83E-01 |
| MANF | 424,95 | 0,08 | 0,06 | 1,36 | 1,73E-01 | 3,83E-01 |
| WDFY1 | 2497,19 | 0,07 | 0,05 | 1,36 | 1,73E-01 | 3,83E-01 |
| MAP2K3 | 2970,94 | -0,17 | 0,13 | -1,36 | 1,73E-01 | 3,83E-01 |
| NCKIPSD | 132,97 | -0,12 | 0,09 | -1,36 | 1,73E-01 | 3,83E-01 |
| DNAJC14 | 756,70 | 0,04 | 0,03 | 1,36 | 1,73E-01 | 3,83E-01 |
| U2AF1L4 | 34,04 | -0,13 | 0,09 | -1,36 | 1,73E-01 | 3,83E-01 |
| SMIM19 | 205,42 | 0,08 | 0,06 | 1,36 | 1,73E-01 | 3,83E-01 |
| UBE4B | 1840,41 | -0,07 | 0,05 | -1,36 | 1,73E-01 | 3,83E-01 |
| TAS2R46 | 128,98 | 0,12 | 0,09 | 1,36 | 1,73E-01 | 3,83E-01 |
| POLR1D | 2141,63 | -0,09 | 0,07 | -1,36 | 1,74E-01 | 3,83E-01 |
| MTPAP | 678,23 | 0,04 | 0,03 | 1,36 | 1,74E-01 | 3,83E-01 |
| GNG11 | 563,13 | -0,20 | 0,14 | -1,36 | 1,74E-01 | 3,83E-01 |
| FCRLA | 315,39 | 0,16 | 0,12 | 1,36 | 1,74E-01 | 3,83E-01 |
| BBS9 | 837,79 | 0,07 | 0,05 | 1,36 | 1,74E-01 | 3,84E-01 |
| CIB1 | 1012,60 | -0,07 | 0,05 | -1,36 | 1,74E-01 | 3,84E-01 |
| ZFP3 | 264,74 | 0,08 | 0,06 | 1,36 | 1,74E-01 | 3,84E-01 |
| MRPS17 | 98,10 | 0,07 | 0,05 | 1,36 | 1,74E-01 | 3,84E-01 |
| LDHA | 2107,15 | 0,06 | 0,05 | 1,36 | 1,74E-01 | 3,84E-01 |
| THAP4 | 177,44 | -0,10 | 0,07 | -1,36 | 1,74E-01 | 3,84E-01 |
| IRF9 | 489,57 | -0,11 | 0,08 | -1,36 | 1,74E-01 | 3,84E-01 |
| POU6F1 | 186,19 | -0,13 | 0,10 | -1,36 | 1,74E-01 | 3,84E-01 |
| C3orf18 | 315,16 | 0,06 | 0,04 | 1,36 | 1,74E-01 | 3,85E-01 |
| MICOS10 | 477,74 | 0,04 | 0,03 | 1,36 | 1,75E-01 | 3,85E-01 |
| PLAUR | 1339,33 | -0,11 | 0,08 | -1,36 | 1,75E-01 | 3,85E-01 |
| IER5 | 2069,36 | 0,07 | 0,05 | 1,36 | 1,75E-01 | 3,85E-01 |
| SLC66A3 | 863,98 | 0,07 | 0,05 | 1,36 | 1,75E-01 | 3,85E-01 |
| TAP2 | 1769,07 | -0,09 | 0,07 | -1,36 | 1,75E-01 | 3,85E-01 |
| NDUFB9 | 968,40 | 0,06 | 0,04 | 1,36 | 1,75E-01 | 3,85E-01 |
| RBM4 | 1469,52 | 0,05 | 0,04 | 1,36 | 1,75E-01 | 3,85E-01 |
| GDI1 | 867,93 | -0,09 | 0,07 | -1,36 | 1,75E-01 | 3,85E-01 |
| MYH3 | 231,35 | -0,13 | 0,09 | -1,36 | 1,75E-01 | 3,85E-01 |
| NFIC | 328,57 | -0,19 | 0,14 | -1,36 | 1,75E-01 | 3,85E-01 |

| | | | | | | |
|---|---|---|---|---|---|---|
| RPL27A | 9589,79 | 0,12 | 0,09 | 1,36 | 1,75E-01 | 3,85E-01 |
| ZBTB37 | 7102,71 | 0,06 | 0,05 | 1,36 | 1,75E-01 | 3,85E-01 |
| PARP8 | 14046,10 | 0,09 | 0,07 | 1,36 | 1,75E-01 | 3,85E-01 |
| AFF2 | 244,75 | -0,11 | 0,08 | -1,36 | 1,75E-01 | 3,85E-01 |
| WDR41 | 1100,39 | 0,07 | 0,05 | 1,35 | 1,75E-01 | 3,86E-01 |
| TRRAP | 3405,39 | -0,10 | 0,07 | -1,35 | 1,76E-01 | 3,86E-01 |
| LRRC37A | 122,62 | -0,14 | 0,10 | -1,35 | 1,76E-01 | 3,86E-01 |
| CADM1 | 96,71 | -0,23 | 0,17 | -1,35 | 1,76E-01 | 3,86E-01 |
| S100A8 | 9120,54 | 0,22 | 0,16 | 1,35 | 1,76E-01 | 3,86E-01 |
| TAB3 | 1817,00 | 0,05 | 0,04 | 1,35 | 1,76E-01 | 3,86E-01 |
| RAB2A | 2711,82 | 0,08 | 0,06 | 1,35 | 1,76E-01 | 3,86E-01 |
| ZNF837 | 62,66 | -0,13 | 0,09 | -1,35 | 1,76E-01 | 3,86E-01 |
| TIMM29 | 663,83 | 0,06 | 0,04 | 1,35 | 1,76E-01 | 3,86E-01 |
| MARCHF9 | 219,68 | -0,11 | 0,08 | -1,35 | 1,76E-01 | 3,86E-01 |
| PLVAP | 171,19 | -0,22 | 0,16 | -1,35 | 1,76E-01 | 3,86E-01 |
| ST8SIA1 | 488,33 | -0,13 | 0,10 | -1,35 | 1,76E-01 | 3,86E-01 |
| PAWR | 310,91 | 0,13 | 0,10 | 1,35 | 1,76E-01 | 3,86E-01 |
| DNAL1 | 246,50 | 0,06 | 0,04 | 1,35 | 1,76E-01 | 3,86E-01 |
| DDX59 | 788,66 | 0,05 | 0,04 | 1,35 | 1,76E-01 | 3,86E-01 |
| EDEM3 | 4372,09 | 0,08 | 0,06 | 1,35 | 1,76E-01 | 3,86E-01 |
| SLC6A16 | 163,64 | -0,12 | 0,09 | -1,35 | 1,76E-01 | 3,86E-01 |
| CABP4 | 44,36 | -0,19 | 0,14 | -1,35 | 1,76E-01 | 3,86E-01 |
| NCSTN | 1078,29 | -0,08 | 0,06 | -1,35 | 1,76E-01 | 3,86E-01 |
| RNF187 | 524,19 | -0,17 | 0,12 | -1,35 | 1,76E-01 | 3,86E-01 |
| C14orf132 | 133,29 | -0,19 | 0,14 | -1,35 | 1,76E-01 | 3,86E-01 |
| SLC25A42 | 229,83 | -0,11 | 0,08 | -1,35 | 1,76E-01 | 3,86E-01 |
| CAPZA1 | 7765,94 | 0,12 | 0,09 | 1,35 | 1,76E-01 | 3,86E-01 |
| TNS1 | 4317,29 | -0,23 | 0,17 | -1,35 | 1,76E-01 | 3,86E-01 |
| LTBR | 548,72 | -0,11 | 0,08 | -1,35 | 1,77E-01 | 3,87E-01 |
| PDIA5 | 106,26 | -0,12 | 0,09 | -1,35 | 1,77E-01 | 3,87E-01 |
| GINS4 | 87,83 | -0,11 | 0,08 | -1,35 | 1,77E-01 | 3,87E-01 |
| AKT2 | 2692,89 | -0,07 | 0,05 | -1,35 | 1,77E-01 | 3,87E-01 |
| TAS2R10 | 30,15 | 0,13 | 0,10 | 1,35 | 1,77E-01 | 3,87E-01 |
| EEF1AKMT1 | 91,67 | 0,08 | 0,06 | 1,35 | 1,77E-01 | 3,87E-01 |
| MGME1 | 642,28 | 0,07 | 0,05 | 1,35 | 1,77E-01 | 3,87E-01 |
| XK | 2040,71 | -0,20 | 0,15 | -1,35 | 1,77E-01 | 3,87E-01 |
| AMACR | 134,16 | 0,09 | 0,07 | 1,35 | 1,77E-01 | 3,87E-01 |
| GRAMD1A | 1355,62 | -0,12 | 0,09 | -1,35 | 1,77E-01 | 3,87E-01 |
| HADHA | 3573,18 | 0,06 | 0,04 | 1,35 | 1,77E-01 | 3,87E-01 |
| SHOC2 | 4525,94 | 0,10 | 0,08 | 1,35 | 1,77E-01 | 3,88E-01 |
| BEX3 | 264,27 | -0,14 | 0,10 | -1,35 | 1,77E-01 | 3,88E-01 |
| FAM120AOS | 1776,44 | 0,05 | 0,03 | 1,35 | 1,77E-01 | 3,88E-01 |
| JDP2 | 951,09 | -0,09 | 0,06 | -1,35 | 1,78E-01 | 3,88E-01 |
| DERL3 | 38,85 | -0,22 | 0,16 | -1,35 | 1,78E-01 | 3,89E-01 |
| B4GALT5 | 3187,50 | -0,11 | 0,08 | -1,35 | 1,78E-01 | 3,89E-01 |
| LRGUK | 44,76 | -0,26 | 0,19 | -1,35 | 1,78E-01 | 3,89E-01 |

| | | | | | | |
|---|---|---|---|---|---|---|
| FRMPD3 | 85,27 | -0,20 | 0,15 | -1,35 | 1,78E-01 | 3,89E-01 |
| MARK3 | 5171,17 | -0,06 | 0,05 | -1,35 | 1,78E-01 | 3,89E-01 |
| WDR19 | 469,21 | 0,06 | 0,04 | 1,35 | 1,78E-01 | 3,89E-01 |
| CES1 | 273,18 | 0,21 | 0,16 | 1,35 | 1,78E-01 | 3,89E-01 |
| ABRAXAS2 | 272,45 | 0,06 | 0,04 | 1,35 | 1,78E-01 | 3,89E-01 |
| HNRNPF | 4537,75 | 0,06 | 0,05 | 1,35 | 1,78E-01 | 3,89E-01 |
| ZNF595 | 345,11 | 0,13 | 0,09 | 1,34 | 1,79E-01 | 3,90E-01 |
| GZMM | 1001,32 | 0,10 | 0,07 | 1,34 | 1,79E-01 | 3,90E-01 |
| KLHL33 | 34,40 | -0,26 | 0,19 | -1,34 | 1,79E-01 | 3,90E-01 |
| CAMK4 | 4132,38 | 0,11 | 0,08 | 1,34 | 1,79E-01 | 3,90E-01 |
| PPP3CB | 2020,27 | 0,07 | 0,05 | 1,34 | 1,79E-01 | 3,90E-01 |
| IL18RAP | 2180,59 | 0,14 | 0,10 | 1,34 | 1,79E-01 | 3,90E-01 |
| LY6G6F | 27,38 | -0,21 | 0,16 | -1,34 | 1,79E-01 | 3,90E-01 |
| MIS18A | 64,52 | -0,10 | 0,07 | -1,34 | 1,79E-01 | 3,90E-01 |
| VCL | 4641,73 | -0,10 | 0,07 | -1,34 | 1,79E-01 | 3,90E-01 |
| MBLAC1 | 64,67 | -0,09 | 0,07 | -1,34 | 1,79E-01 | 3,90E-01 |
| IPMK | 2961,93 | 0,13 | 0,09 | 1,34 | 1,79E-01 | 3,90E-01 |
| VTI1B | 2587,32 | -0,09 | 0,07 | -1,34 | 1,79E-01 | 3,90E-01 |
| RGS2 | 12107,90 | 0,13 | 0,10 | 1,34 | 1,79E-01 | 3,90E-01 |
| KIN | 1012,63 | 0,05 | 0,04 | 1,34 | 1,79E-01 | 3,90E-01 |
| ABT1 | 1388,19 | 0,06 | 0,04 | 1,34 | 1,79E-01 | 3,90E-01 |
| BBOF1 | 525,27 | -0,16 | 0,12 | -1,34 | 1,80E-01 | 3,91E-01 |
| PDE4D | 1438,84 | 0,09 | 0,06 | 1,34 | 1,80E-01 | 3,91E-01 |
| KCTD13 | 754,77 | -0,09 | 0,06 | -1,34 | 1,80E-01 | 3,91E-01 |
| TP53I11 | 379,86 | -0,17 | 0,13 | -1,34 | 1,80E-01 | 3,91E-01 |
| GALNT10 | 1931,88 | -0,07 | 0,06 | -1,34 | 1,80E-01 | 3,91E-01 |
| EIF4E3 | 5012,10 | -0,09 | 0,07 | -1,34 | 1,80E-01 | 3,91E-01 |
| GOLM2 | 3178,98 | 0,08 | 0,06 | 1,34 | 1,80E-01 | 3,91E-01 |
| TSC22D3 | 4908,79 | -0,10 | 0,08 | -1,34 | 1,80E-01 | 3,91E-01 |
| UBASH3B | 1177,98 | -0,07 | 0,05 | -1,34 | 1,80E-01 | 3,91E-01 |
| BST1 | 1642,92 | -0,09 | 0,07 | -1,34 | 1,80E-01 | 3,91E-01 |
| XPOT | 1560,30 | 0,06 | 0,05 | 1,34 | 1,80E-01 | 3,91E-01 |
| GRK5 | 908,22 | -0,06 | 0,04 | -1,34 | 1,80E-01 | 3,91E-01 |
| UTP4 | 305,94 | -0,06 | 0,05 | -1,34 | 1,80E-01 | 3,91E-01 |
| SNX30 | 2091,73 | 0,05 | 0,04 | 1,34 | 1,80E-01 | 3,91E-01 |
| SUMO2 | 2444,75 | 0,08 | 0,06 | 1,34 | 1,80E-01 | 3,91E-01 |
| AGAP2 | 956,35 | -0,12 | 0,09 | -1,34 | 1,80E-01 | 3,91E-01 |
| NRBP1 | 1120,70 | -0,07 | 0,05 | -1,34 | 1,81E-01 | 3,91E-01 |
| JMJD7 | 44,58 | -0,14 | 0,10 | -1,34 | 1,81E-01 | 3,91E-01 |
| COX7A2 | 985,61 | 0,14 | 0,10 | 1,34 | 1,80E-01 | 3,91E-01 |
| ARAP3 | 587,82 | -0,19 | 0,14 | -1,34 | 1,81E-01 | 3,91E-01 |
| BRCA2 | 275,01 | 0,13 | 0,10 | 1,34 | 1,81E-01 | 3,92E-01 |
| VRK3 | 1614,32 | -0,06 | 0,05 | -1,34 | 1,81E-01 | 3,92E-01 |
| LILRB3 | 2625,03 | -0,14 | 0,11 | -1,34 | 1,81E-01 | 3,92E-01 |
| SGPP1 | 879,48 | 0,09 | 0,07 | 1,34 | 1,81E-01 | 3,92E-01 |
| AXIN1 | 1044,44 | -0,09 | 0,07 | -1,34 | 1,81E-01 | 3,92E-01 |

| | | | | | | |
|---|---|---|---|---|---|---|
| SELENOK | 1800,01 | -0,14 | 0,11 | -1,34 | 1,81E-01 | 3,92E-01 |
| ZC3H12A | 523,66 | -0,07 | 0,05 | -1,34 | 1,81E-01 | 3,92E-01 |
| TRPM2 | 295,57 | -0,16 | 0,12 | -1,34 | 1,81E-01 | 3,92E-01 |
| ABHD17B | 636,17 | 0,06 | 0,04 | 1,34 | 1,81E-01 | 3,92E-01 |
| DCAF15 | 351,81 | -0,11 | 0,08 | -1,34 | 1,81E-01 | 3,92E-01 |
| YTHDF3 | 3775,42 | 0,07 | 0,05 | 1,34 | 1,81E-01 | 3,92E-01 |
| ABHD13 | 2109,74 | 0,13 | 0,10 | 1,34 | 1,81E-01 | 3,92E-01 |
| SPTA1 | 473,03 | -0,18 | 0,14 | -1,34 | 1,81E-01 | 3,92E-01 |
| SHTN1 | 553,07 | 0,11 | 0,08 | 1,34 | 1,81E-01 | 3,92E-01 |
| UPF1 | 3155,57 | -0,10 | 0,07 | -1,34 | 1,81E-01 | 3,92E-01 |
| PSMA5 | 1859,63 | 0,07 | 0,05 | 1,34 | 1,81E-01 | 3,92E-01 |
| ZDHHC20 | 4718,94 | 0,10 | 0,07 | 1,34 | 1,82E-01 | 3,92E-01 |
| SYCE1L | 42,75 | -0,13 | 0,09 | -1,34 | 1,82E-01 | 3,92E-01 |
| ATP11B | 8008,03 | 0,10 | 0,07 | 1,34 | 1,82E-01 | 3,92E-01 |
| IGF1R | 3967,58 | -0,11 | 0,08 | -1,34 | 1,82E-01 | 3,92E-01 |
| THBD | 782,85 | -0,12 | 0,09 | -1,34 | 1,82E-01 | 3,92E-01 |
| HSBP1 | 1160,38 | 0,10 | 0,08 | 1,34 | 1,82E-01 | 3,92E-01 |
| ARID2 | 2694,91 | 0,05 | 0,04 | 1,34 | 1,82E-01 | 3,92E-01 |
| NBEAL2 | 5363,33 | -0,19 | 0,14 | -1,33 | 1,82E-01 | 3,93E-01 |
| ALDOC | 60,13 | -0,12 | 0,09 | -1,33 | 1,82E-01 | 3,93E-01 |
| AFAP1 | 363,28 | -0,16 | 0,12 | -1,33 | 1,82E-01 | 3,93E-01 |
| ZNG1E | 1038,35 | 0,09 | 0,07 | 1,33 | 1,82E-01 | 3,93E-01 |
| BOLA1 | 159,38 | 0,06 | 0,05 | 1,33 | 1,82E-01 | 3,93E-01 |
| GATAD2A | 1398,12 | -0,13 | 0,10 | -1,33 | 1,82E-01 | 3,93E-01 |
| NDUFB1 | 767,17 | 0,08 | 0,06 | 1,33 | 1,82E-01 | 3,93E-01 |
| SLC25A53 | 550,37 | 0,07 | 0,05 | 1,33 | 1,82E-01 | 3,93E-01 |
| ARL4C | 4109,75 | 0,07 | 0,05 | 1,33 | 1,82E-01 | 3,93E-01 |
| SCPEP1 | 2238,45 | 0,08 | 0,06 | 1,33 | 1,82E-01 | 3,93E-01 |
| UBA3 | 3108,02 | 0,09 | 0,07 | 1,33 | 1,82E-01 | 3,93E-01 |
| MED12 | 822,27 | -0,16 | 0,12 | -1,33 | 1,82E-01 | 3,93E-01 |
| TMEM250 | 1154,54 | -0,06 | 0,04 | -1,33 | 1,82E-01 | 3,93E-01 |
| FAAP100 | 289,13 | -0,15 | 0,11 | -1,33 | 1,82E-01 | 3,93E-01 |
| GPR65 | 2322,16 | 0,12 | 0,09 | 1,33 | 1,82E-01 | 3,93E-01 |
| CLSTN3 | 556,93 | -0,13 | 0,10 | -1,33 | 1,82E-01 | 3,93E-01 |
| CENPT | 172,10 | -0,14 | 0,11 | -1,33 | 1,82E-01 | 3,93E-01 |
| ZNF668 | 114,39 | -0,10 | 0,08 | -1,33 | 1,82E-01 | 3,93E-01 |
| SAMD12 | 420,61 | 0,15 | 0,11 | 1,33 | 1,83E-01 | 3,93E-01 |
| CASTOR1 | 53,74 | -0,09 | 0,06 | -1,33 | 1,83E-01 | 3,93E-01 |
| LY75 | 973,09 | 0,07 | 0,05 | 1,33 | 1,83E-01 | 3,93E-01 |
| ADORA3 | 73,16 | -0,18 | 0,13 | -1,33 | 1,83E-01 | 3,93E-01 |
| CRTAM | 199,62 | 0,12 | 0,09 | 1,33 | 1,83E-01 | 3,93E-01 |
| HHEX | 689,38 | 0,07 | 0,06 | 1,33 | 1,83E-01 | 3,93E-01 |
| ZMPSTE24 | 1561,64 | 0,09 | 0,06 | 1,33 | 1,83E-01 | 3,93E-01 |
| PLXNA4 | 224,73 | -0,18 | 0,14 | -1,33 | 1,83E-01 | 3,94E-01 |
| NUDT4 | 2996,50 | -0,12 | 0,09 | -1,33 | 1,83E-01 | 3,94E-01 |
| HBP1 | 5608,76 | 0,08 | 0,06 | 1,33 | 1,84E-01 | 3,94E-01 |

| | | | | | | |
|---|---|---|---|---|---|---|
| FOXJ2 | 554,42 | -0,11 | 0,08 | -1,33 | 1,84E-01 | 3,94E-01 |
| SLC25A44 | 1532,38 | -0,09 | 0,07 | -1,33 | 1,84E-01 | 3,95E-01 |
| ZNF274 | 1596,34 | 0,05 | 0,04 | 1,33 | 1,84E-01 | 3,95E-01 |
| STX12 | 1374,58 | 0,05 | 0,04 | 1,33 | 1,84E-01 | 3,95E-01 |
| TNFRSF14 | 2121,13 | -0,09 | 0,07 | -1,33 | 1,84E-01 | 3,95E-01 |
| NFATC1 | 502,18 | -0,13 | 0,10 | -1,33 | 1,84E-01 | 3,95E-01 |
| ZNF232 | 200,03 | 0,06 | 0,05 | 1,33 | 1,84E-01 | 3,95E-01 |
| GALE | 148,21 | -0,07 | 0,05 | -1,33 | 1,84E-01 | 3,95E-01 |
| RNF115 | 1679,45 | 0,04 | 0,03 | 1,33 | 1,84E-01 | 3,95E-01 |
| GUCY1A1 | 272,64 | -0,13 | 0,10 | -1,33 | 1,84E-01 | 3,95E-01 |
| TRPV2 | 1144,98 | -0,07 | 0,06 | -1,33 | 1,84E-01 | 3,95E-01 |
| AOPEP | 645,73 | -0,08 | 0,06 | -1,33 | 1,84E-01 | 3,95E-01 |
| NADSYN1 | 2605,71 | -0,07 | 0,05 | -1,33 | 1,84E-01 | 3,95E-01 |
| TSC2 | 935,33 | -0,16 | 0,12 | -1,33 | 1,84E-01 | 3,95E-01 |
| RCBTB1 | 953,40 | 0,06 | 0,04 | 1,33 | 1,84E-01 | 3,95E-01 |
| ZSWIM8 | 1685,29 | -0,11 | 0,08 | -1,33 | 1,84E-01 | 3,95E-01 |
| MCM3 | 945,82 | 0,06 | 0,04 | 1,33 | 1,84E-01 | 3,95E-01 |
| LRRC70 | 44,23 | 0,14 | 0,11 | 1,33 | 1,84E-01 | 3,95E-01 |
| FAM91A1 | 4742,66 | 0,08 | 0,06 | 1,33 | 1,84E-01 | 3,95E-01 |
| CCDC62 | 58,36 | -0,12 | 0,09 | -1,33 | 1,85E-01 | 3,95E-01 |
| PHKB | 2978,46 | 0,05 | 0,04 | 1,33 | 1,85E-01 | 3,95E-01 |
| ABCD1 | 250,13 | -0,12 | 0,09 | -1,33 | 1,85E-01 | 3,95E-01 |
| SERPINH1 | 189,47 | -0,09 | 0,07 | -1,33 | 1,85E-01 | 3,96E-01 |
| STPG4 | 34,07 | -0,15 | 0,11 | -1,32 | 1,85E-01 | 3,96E-01 |
| MGAM | 9160,62 | -0,13 | 0,10 | -1,33 | 1,85E-01 | 3,96E-01 |
| ENSG000002 | 347,79 | -0,07 | 0,06 | -1,32 | 1,85E-01 | 3,96E-01 |
| C5AR1 | 1955,82 | -0,14 | 0,11 | -1,32 | 1,85E-01 | 3,96E-01 |
| VCF2 | 108,64 | -0,10 | 0,08 | -1,32 | 1,85E-01 | 3,96E-01 |
| NLE1 | 318,97 | -0,07 | 0,05 | -1,32 | 1,85E-01 | 3,96E-01 |
| TBCK | 2135,05 | 0,09 | 0,07 | 1,32 | 1,86E-01 | 3,96E-01 |
| F2R | 592,25 | -0,11 | 0,08 | -1,32 | 1,86E-01 | 3,96E-01 |
| STK38L | 2658,37 | 0,09 | 0,07 | 1,32 | 1,86E-01 | 3,96E-01 |
| ATP11A | 3903,19 | -0,10 | 0,08 | -1,32 | 1,86E-01 | 3,96E-01 |
| BAZ2A | 3068,67 | -0,12 | 0,09 | -1,32 | 1,86E-01 | 3,96E-01 |
| SMG9 | 304,68 | -0,11 | 0,08 | -1,32 | 1,86E-01 | 3,96E-01 |
| MAP7 | 132,96 | -0,12 | 0,09 | -1,32 | 1,86E-01 | 3,97E-01 |
| ZNF440 | 637,47 | 0,05 | 0,04 | 1,32 | 1,86E-01 | 3,97E-01 |
| RALGAPA2 | 5263,76 | -0,09 | 0,07 | -1,32 | 1,86E-01 | 3,97E-01 |
| MRPS5 | 1239,89 | 0,04 | 0,03 | 1,32 | 1,86E-01 | 3,97E-01 |
| MAPK13 | 1065,93 | -0,14 | 0,11 | -1,32 | 1,86E-01 | 3,97E-01 |
| IL31RA | 56,64 | -0,23 | 0,18 | -1,32 | 1,86E-01 | 3,97E-01 |
| ZNF623 | 983,64 | 0,04 | 0,03 | 1,32 | 1,86E-01 | 3,97E-01 |
| ENSG000002 | 39,91 | -0,08 | 0,06 | -1,32 | 1,86E-01 | 3,97E-01 |
| TMEM135 | 425,23 | 0,09 | 0,07 | 1,32 | 1,86E-01 | 3,97E-01 |
| NEDD9 | 5339,77 | -0,09 | 0,07 | -1,32 | 1,86E-01 | 3,97E-01 |
| ZBTB8OS | 448,65 | 0,05 | 0,04 | 1,32 | 1,86E-01 | 3,97E-01 |

| | | | | | | |
|---|---|---|---|---|---|---|
| ZNF576 | 173,34 | -0,05 | 0,04 | -1,32 | 1,87E-01 | 3,97E-01 |
| MYO1F | 14084,75 | -0,12 | 0,09 | -1,32 | 1,87E-01 | 3,97E-01 |
| ZNF473 | 292,11 | 0,05 | 0,04 | 1,32 | 1,87E-01 | 3,97E-01 |
| PSMC3 | 921,32 | 0,07 | 0,05 | 1,32 | 1,87E-01 | 3,98E-01 |
| FAM161B | 151,05 | -0,07 | 0,05 | -1,32 | 1,87E-01 | 3,98E-01 |
| WWP2 | 2412,07 | -0,10 | 0,08 | -1,32 | 1,87E-01 | 3,98E-01 |
| DGKZ | 2022,37 | -0,14 | 0,10 | -1,32 | 1,87E-01 | 3,98E-01 |
| TNRC18 | 1918,29 | -0,19 | 0,15 | -1,32 | 1,87E-01 | 3,98E-01 |
| ABI1 | 4996,03 | 0,08 | 0,06 | 1,32 | 1,87E-01 | 3,98E-01 |
| ACVR1B | 656,28 | -0,08 | 0,06 | -1,32 | 1,87E-01 | 3,98E-01 |
| DGKD | 2527,75 | -0,11 | 0,08 | -1,32 | 1,87E-01 | 3,98E-01 |
| PTPRC | 66718,73 | 0,08 | 0,06 | 1,32 | 1,87E-01 | 3,99E-01 |
| CDC34 | 6011,19 | -0,17 | 0,13 | -1,32 | 1,88E-01 | 3,99E-01 |
| NENF | 135,22 | -0,08 | 0,06 | -1,32 | 1,88E-01 | 3,99E-01 |
| APLF | 300,79 | 0,10 | 0,08 | 1,32 | 1,88E-01 | 3,99E-01 |
| ZFP2 | 34,64 | -0,18 | 0,13 | -1,32 | 1,88E-01 | 3,99E-01 |
| MAF1 | 3581,71 | -0,12 | 0,09 | -1,32 | 1,88E-01 | 3,99E-01 |
| HDAC7 | 1743,72 | -0,12 | 0,09 | -1,32 | 1,88E-01 | 3,99E-01 |
| ARB2A | 1361,39 | 0,08 | 0,06 | 1,32 | 1,88E-01 | 3,99E-01 |
| REXO4 | 617,13 | 0,05 | 0,04 | 1,32 | 1,88E-01 | 3,99E-01 |
| LAMP5 | 28,14 | -0,19 | 0,15 | -1,32 | 1,88E-01 | 3,99E-01 |
| ABHD14B | 197,04 | -0,10 | 0,08 | -1,32 | 1,88E-01 | 3,99E-01 |
| DPY30 | 319,50 | 0,10 | 0,07 | 1,32 | 1,88E-01 | 3,99E-01 |
| STXBP2 | 747,54 | -0,13 | 0,10 | -1,32 | 1,88E-01 | 3,99E-01 |
| ADCK5 | 156,01 | -0,09 | 0,07 | -1,32 | 1,88E-01 | 3,99E-01 |
| SUSD6 | 2120,11 | -0,13 | 0,10 | -1,32 | 1,88E-01 | 3,99E-01 |
| RAB11FIP3 | 391,24 | -0,11 | 0,09 | -1,32 | 1,88E-01 | 3,99E-01 |
| ITCH | 5279,76 | 0,06 | 0,05 | 1,32 | 1,88E-01 | 3,99E-01 |
| MICALL1 | 211,09 | -0,16 | 0,12 | -1,32 | 1,88E-01 | 3,99E-01 |
| SCAI | 707,74 | 0,07 | 0,05 | 1,32 | 1,88E-01 | 3,99E-01 |
| PLS1 | 57,77 | -0,17 | 0,13 | -1,32 | 1,88E-01 | 3,99E-01 |
| SPAST | 1934,45 | 0,07 | 0,06 | 1,32 | 1,88E-01 | 3,99E-01 |
| GTF2IRD2B | 443,21 | 0,06 | 0,05 | 1,31 | 1,89E-01 | 3,99E-01 |
| STK40 | 2136,65 | -0,12 | 0,09 | -1,32 | 1,89E-01 | 3,99E-01 |
| MCF2 | 57,93 | -0,13 | 0,10 | -1,32 | 1,89E-01 | 3,99E-01 |
| CLPTM1L | 1509,38 | -0,04 | 0,03 | -1,31 | 1,89E-01 | 3,99E-01 |
| FHIP1A | 77,74 | -0,17 | 0,13 | -1,31 | 1,89E-01 | 4,00E-01 |
| SPAG1 | 244,66 | 0,09 | 0,07 | 1,31 | 1,89E-01 | 4,00E-01 |
| ACYP2 | 351,81 | -0,05 | 0,04 | -1,31 | 1,89E-01 | 4,00E-01 |
| HINT3 | 915,01 | 0,08 | 0,06 | 1,31 | 1,89E-01 | 4,00E-01 |
| CDK17 | 3489,79 | 0,07 | 0,05 | 1,31 | 1,89E-01 | 4,00E-01 |
| FGFR2 | 136,13 | -0,23 | 0,18 | -1,31 | 1,89E-01 | 4,00E-01 |
| KCNMB3 | 99,68 | 0,09 | 0,07 | 1,31 | 1,89E-01 | 4,00E-01 |
| EIF5A2 | 243,89 | 0,11 | 0,08 | 1,31 | 1,89E-01 | 4,00E-01 |
| CYP2U1 | 331,12 | 0,07 | 0,05 | 1,31 | 1,89E-01 | 4,01E-01 |
| GRK2 | 4764,50 | -0,10 | 0,08 | -1,31 | 1,90E-01 | 4,01E-01 |

| | | | | | | |
|---|---|---|---|---|---|---|
| PRICKLE2 | 64,39 | -0,30 | 0,23 | -1,31 | 1,90E-01 | 4,01E-01 |
| MBOAT1 | 1107,28 | -0,06 | 0,05 | -1,31 | 1,90E-01 | 4,01E-01 |
| SRF | 602,38 | -0,07 | 0,05 | -1,31 | 1,90E-01 | 4,01E-01 |
| CD3D | 1082,43 | 0,11 | 0,08 | 1,31 | 1,90E-01 | 4,01E-01 |
| LRP12 | 144,40 | -0,11 | 0,09 | -1,31 | 1,90E-01 | 4,01E-01 |
| PARM1 | 51,46 | -0,17 | 0,13 | -1,31 | 1,90E-01 | 4,01E-01 |
| AP1M1 | 1218,79 | -0,08 | 0,06 | -1,31 | 1,90E-01 | 4,01E-01 |
| CSNK1D | 2288,55 | -0,12 | 0,09 | -1,31 | 1,90E-01 | 4,01E-01 |
| AGA | 510,78 | 0,07 | 0,06 | 1,31 | 1,90E-01 | 4,01E-01 |
| ENSG000002 | 19,35 | -0,14 | 0,11 | -1,31 | 1,90E-01 | 4,01E-01 |
| LCMT2 | 385,81 | 0,07 | 0,05 | 1,31 | 1,90E-01 | 4,01E-01 |
| WTAP | 4405,51 | 0,07 | 0,06 | 1,31 | 1,90E-01 | 4,01E-01 |
| CARD19 | 721,84 | -0,09 | 0,07 | -1,31 | 1,90E-01 | 4,01E-01 |
| ICOS | 588,54 | 0,10 | 0,08 | 1,31 | 1,90E-01 | 4,01E-01 |
| ATP5PF | 194,39 | 0,11 | 0,08 | 1,31 | 1,90E-01 | 4,01E-01 |
| CDK13 | 6735,56 | 0,04 | 0,03 | 1,31 | 1,90E-01 | 4,01E-01 |
| HSF1 | 357,95 | -0,12 | 0,09 | -1,31 | 1,90E-01 | 4,01E-01 |
| THADA | 2921,14 | 0,06 | 0,04 | 1,31 | 1,90E-01 | 4,01E-01 |
| SCARB1 | 169,66 | -0,15 | 0,12 | -1,31 | 1,91E-01 | 4,01E-01 |
| SPDL1 | 193,18 | 0,08 | 0,06 | 1,31 | 1,91E-01 | 4,01E-01 |
| SS18L2 | 261,34 | 0,13 | 0,10 | 1,31 | 1,91E-01 | 4,01E-01 |
| CYB5D1 | 42,18 | 0,09 | 0,07 | 1,31 | 1,91E-01 | 4,01E-01 |
| SLC30A7 | 2650,24 | 0,06 | 0,05 | 1,31 | 1,91E-01 | 4,01E-01 |
| MSL1 | 7959,88 | -0,08 | 0,06 | -1,31 | 1,91E-01 | 4,01E-01 |
| SCML1 | 321,37 | 0,12 | 0,09 | 1,31 | 1,91E-01 | 4,01E-01 |
| PTRH2 | 488,07 | 0,06 | 0,05 | 1,31 | 1,91E-01 | 4,01E-01 |
| C4orf46 | 163,49 | 0,08 | 0,06 | 1,31 | 1,91E-01 | 4,01E-01 |
| IGBP1 | 848,51 | 0,09 | 0,07 | 1,31 | 1,91E-01 | 4,01E-01 |
| ENKD1 | 150,86 | -0,07 | 0,06 | -1,31 | 1,91E-01 | 4,01E-01 |
| PPP1R13B | 226,72 | -0,13 | 0,10 | -1,31 | 1,91E-01 | 4,01E-01 |
| SLC49A3 | 292,73 | -0,11 | 0,09 | -1,31 | 1,91E-01 | 4,01E-01 |
| PHKA2 | 1283,50 | -0,08 | 0,06 | -1,31 | 1,91E-01 | 4,02E-01 |
| SAP130 | 615,31 | -0,06 | 0,05 | -1,31 | 1,91E-01 | 4,02E-01 |
| MAP3K9 | 172,89 | -0,16 | 0,12 | -1,31 | 1,91E-01 | 4,02E-01 |
| CNNM2 | 467,24 | -0,08 | 0,06 | -1,31 | 1,91E-01 | 4,02E-01 |
| BCL7C | 457,43 | 0,07 | 0,05 | 1,31 | 1,92E-01 | 4,03E-01 |
| PCYT2 | 308,10 | -0,07 | 0,05 | -1,31 | 1,92E-01 | 4,03E-01 |
| COCH | 90,00 | -0,23 | 0,18 | -1,31 | 1,92E-01 | 4,03E-01 |
| S1PR1 | 3199,16 | 0,08 | 0,06 | 1,31 | 1,92E-01 | 4,03E-01 |
| KCNE1 | 721,60 | -0,11 | 0,09 | -1,30 | 1,92E-01 | 4,03E-01 |
| VASP | 5248,24 | -0,10 | 0,07 | -1,30 | 1,92E-01 | 4,03E-01 |
| EMC10 | 1043,12 | -0,06 | 0,05 | -1,30 | 1,92E-01 | 4,03E-01 |
| LRRC7 | 66,45 | -0,32 | 0,24 | -1,30 | 1,92E-01 | 4,03E-01 |
| FANCG | 163,66 | -0,08 | 0,06 | -1,30 | 1,92E-01 | 4,03E-01 |
| PTGR3 | 2270,73 | 0,05 | 0,04 | 1,30 | 1,92E-01 | 4,03E-01 |
| TIMM50 | 349,28 | -0,06 | 0,04 | -1,30 | 1,92E-01 | 4,03E-01 |

| | | | | | | |
|---|---|---|---|---|---|---|
| TBCE | 230,73 | -0,08 | 0,06 | -1,30 | 1,92E-01 | 4,03E-01 |
| GLS2 | 36,73 | -0,15 | 0,12 | -1,30 | 1,92E-01 | 4,03E-01 |
| VCAN | 26474,10 | 0,10 | 0,08 | 1,30 | 1,92E-01 | 4,03E-01 |
| RPL18 | 11977,45 | 0,08 | 0,06 | 1,30 | 1,92E-01 | 4,03E-01 |
| NCOA6 | 1218,19 | -0,07 | 0,06 | -1,30 | 1,92E-01 | 4,03E-01 |
| ABCA13 | 508,36 | -0,27 | 0,20 | -1,30 | 1,92E-01 | 4,03E-01 |
| FBXL19 | 318,93 | -0,09 | 0,07 | -1,30 | 1,93E-01 | 4,03E-01 |
| ACP2 | 150,15 | -0,09 | 0,07 | -1,30 | 1,93E-01 | 4,03E-01 |
| XKR8 | 584,91 | -0,11 | 0,09 | -1,30 | 1,93E-01 | 4,03E-01 |
| GBE1 | 1009,61 | 0,08 | 0,06 | 1,30 | 1,93E-01 | 4,04E-01 |
| SELPLG | 4269,12 | -0,09 | 0,07 | -1,30 | 1,93E-01 | 4,04E-01 |
| ARHGAP1 | 596,19 | -0,13 | 0,10 | -1,30 | 1,93E-01 | 4,04E-01 |
| STAM | 935,38 | 0,06 | 0,05 | 1,30 | 1,93E-01 | 4,04E-01 |
| ZNF354B | 394,66 | 0,08 | 0,06 | 1,30 | 1,93E-01 | 4,04E-01 |
| NUDT19 | 311,64 | -0,04 | 0,03 | -1,30 | 1,93E-01 | 4,04E-01 |
| DTL | 60,37 | -0,16 | 0,12 | -1,30 | 1,93E-01 | 4,04E-01 |
| ACTR3B | 248,94 | -0,07 | 0,05 | -1,30 | 1,93E-01 | 4,04E-01 |
| AGAP4 | 314,17 | -0,07 | 0,06 | -1,30 | 1,93E-01 | 4,05E-01 |
| MCM8 | 474,31 | 0,06 | 0,04 | 1,30 | 1,94E-01 | 4,05E-01 |
| SLC39A8 | 579,24 | 0,09 | 0,07 | 1,30 | 1,94E-01 | 4,05E-01 |
| MGST3 | 866,75 | 0,05 | 0,04 | 1,30 | 1,94E-01 | 4,05E-01 |
| PFDN6 | 296,47 | 0,08 | 0,06 | 1,30 | 1,94E-01 | 4,05E-01 |
| TIRAP | 356,28 | -0,09 | 0,07 | -1,30 | 1,94E-01 | 4,05E-01 |
| VNN1 | 676,58 | 0,15 | 0,12 | 1,30 | 1,94E-01 | 4,05E-01 |
| PHB1 | 1032,94 | 0,07 | 0,06 | 1,30 | 1,94E-01 | 4,05E-01 |
| CNTNAP1 | 87,45 | -0,19 | 0,15 | -1,30 | 1,94E-01 | 4,05E-01 |
| ZNF672 | 726,12 | -0,05 | 0,04 | -1,30 | 1,94E-01 | 4,05E-01 |
| ZBTB7B | 2054,72 | -0,10 | 0,07 | -1,30 | 1,94E-01 | 4,05E-01 |
| SEC14L2 | 70,10 | -0,15 | 0,11 | -1,30 | 1,94E-01 | 4,05E-01 |
| SLC35F3 | 54,04 | -0,19 | 0,15 | -1,30 | 1,94E-01 | 4,06E-01 |
| F13A1 | 5382,93 | -0,15 | 0,12 | -1,30 | 1,95E-01 | 4,06E-01 |
| GPR89A | 358,48 | 0,06 | 0,05 | 1,30 | 1,95E-01 | 4,06E-01 |
| MAPKBP1 | 600,22 | -0,09 | 0,07 | -1,30 | 1,95E-01 | 4,06E-01 |
| EMC7 | 460,32 | 0,05 | 0,04 | 1,30 | 1,95E-01 | 4,06E-01 |
| HLA-DPA1 | 4884,59 | 0,08 | 0,06 | 1,30 | 1,95E-01 | 4,06E-01 |
| CLCN7 | 663,65 | -0,16 | 0,12 | -1,30 | 1,95E-01 | 4,06E-01 |
| ASB6 | 336,54 | -0,10 | 0,08 | -1,30 | 1,95E-01 | 4,06E-01 |
| SNTB2 | 1500,78 | -0,05 | 0,04 | -1,30 | 1,94E-01 | 4,06E-01 |
| SPIRE1 | 158,50 | -0,11 | 0,08 | -1,30 | 1,95E-01 | 4,06E-01 |
| ZNF891 | 968,04 | 0,06 | 0,04 | 1,30 | 1,95E-01 | 4,06E-01 |
| WIPI1 | 901,73 | -0,08 | 0,06 | -1,30 | 1,95E-01 | 4,06E-01 |
| GP1BA | 30,68 | -0,25 | 0,20 | -1,30 | 1,95E-01 | 4,06E-01 |
| MAML1 | 1554,48 | -0,10 | 0,08 | -1,30 | 1,95E-01 | 4,06E-01 |
| SLC35C1 | 167,46 | -0,12 | 0,10 | -1,30 | 1,95E-01 | 4,06E-01 |
| KATNIP | 839,93 | -0,09 | 0,07 | -1,30 | 1,95E-01 | 4,06E-01 |
| KIF21B | 2547,27 | -0,12 | 0,09 | -1,30 | 1,95E-01 | 4,06E-01 |

| | | | | | | |
|---|---|---|---|---|---|---|
| GUCD1 | 7382,62 | -0,14 | 0,11 | -1,30 | 1,95E-01 | 4,06E-01 |
| ARAF | 489,24 | -0,09 | 0,07 | -1,30 | 1,95E-01 | 4,06E-01 |
| PSMC5 | 507,78 | -0,06 | 0,04 | -1,30 | 1,95E-01 | 4,06E-01 |
| PI4KB | 1252,14 | -0,08 | 0,06 | -1,30 | 1,95E-01 | 4,06E-01 |
| MTMR6 | 2578,64 | 0,09 | 0,07 | 1,29 | 1,95E-01 | 4,06E-01 |
| MRPL11 | 287,13 | 0,08 | 0,06 | 1,30 | 1,95E-01 | 4,06E-01 |
| NAA10 | 250,97 | -0,07 | 0,05 | -1,29 | 1,95E-01 | 4,06E-01 |
| GLCE | 474,70 | 0,07 | 0,06 | 1,29 | 1,96E-01 | 4,06E-01 |
| GPR18 | 201,28 | 0,12 | 0,09 | 1,29 | 1,96E-01 | 4,06E-01 |
| PRCC | 413,77 | -0,06 | 0,05 | -1,29 | 1,96E-01 | 4,06E-01 |
| CDNF | 36,79 | -0,12 | 0,09 | -1,29 | 1,96E-01 | 4,07E-01 |
| HDAC6 | 941,30 | -0,06 | 0,05 | -1,29 | 1,96E-01 | 4,07E-01 |
| GLT1D1 | 2931,03 | -0,11 | 0,08 | -1,29 | 1,96E-01 | 4,07E-01 |
| STIM1 | 1828,85 | -0,09 | 0,07 | -1,29 | 1,96E-01 | 4,07E-01 |
| ZNF142 | 350,14 | -0,13 | 0,10 | -1,29 | 1,96E-01 | 4,07E-01 |
| YAF2 | 1931,52 | 0,06 | 0,05 | 1,29 | 1,96E-01 | 4,07E-01 |
| PSME3IP1 | 2980,06 | 0,04 | 0,03 | 1,29 | 1,96E-01 | 4,07E-01 |
| SRSF9 | 2411,13 | 0,04 | 0,03 | 1,29 | 1,96E-01 | 4,07E-01 |
| SLC24A1 | 158,31 | -0,12 | 0,09 | -1,29 | 1,96E-01 | 4,07E-01 |
| C1QBP | 912,16 | 0,06 | 0,05 | 1,29 | 1,96E-01 | 4,07E-01 |
| SMIM27 | 181,15 | -0,08 | 0,06 | -1,29 | 1,96E-01 | 4,07E-01 |
| COX4I1 | 3144,92 | 0,08 | 0,06 | 1,29 | 1,96E-01 | 4,07E-01 |
| CDK7 | 472,95 | 0,05 | 0,04 | 1,29 | 1,96E-01 | 4,07E-01 |
| ACOT2 | 135,73 | -0,07 | 0,05 | -1,29 | 1,96E-01 | 4,07E-01 |
| FKBP11 | 281,70 | -0,12 | 0,09 | -1,29 | 1,96E-01 | 4,07E-01 |
| CARS1 | 1678,77 | -0,05 | 0,04 | -1,29 | 1,97E-01 | 4,07E-01 |
| ZNF692 | 444,05 | -0,07 | 0,05 | -1,29 | 1,97E-01 | 4,08E-01 |
| MYC | 506,01 | 0,08 | 0,06 | 1,29 | 1,97E-01 | 4,08E-01 |
| NMI | 2310,03 | 0,09 | 0,07 | 1,29 | 1,97E-01 | 4,08E-01 |
| SNX13 | 5209,18 | 0,09 | 0,07 | 1,29 | 1,97E-01 | 4,08E-01 |
| KL | 45,80 | -0,17 | 0,13 | -1,29 | 1,97E-01 | 4,08E-01 |
| SPATA6L | 92,48 | -0,15 | 0,12 | -1,29 | 1,97E-01 | 4,08E-01 |
| MRPL17 | 418,61 | 0,08 | 0,06 | 1,29 | 1,97E-01 | 4,08E-01 |
| UBE2E1 | 1039,85 | 0,06 | 0,05 | 1,29 | 1,97E-01 | 4,08E-01 |
| CYB561D1 | 976,20 | -0,08 | 0,06 | -1,29 | 1,97E-01 | 4,08E-01 |
| SOAT1 | 2727,20 | 0,05 | 0,04 | 1,29 | 1,98E-01 | 4,09E-01 |
| REPS2 | 2832,49 | -0,11 | 0,09 | -1,29 | 1,98E-01 | 4,09E-01 |
| DOP1B | 902,03 | -0,10 | 0,07 | -1,29 | 1,98E-01 | 4,09E-01 |
| SIAH1 | 901,61 | 0,05 | 0,04 | 1,29 | 1,98E-01 | 4,09E-01 |
| STK11 | 3243,54 | -0,10 | 0,08 | -1,29 | 1,98E-01 | 4,09E-01 |
| FGFR1OP2 | 3267,64 | 0,11 | 0,08 | 1,29 | 1,98E-01 | 4,09E-01 |
| CHUK | 1572,44 | 0,09 | 0,07 | 1,29 | 1,98E-01 | 4,09E-01 |
| MBD6 | 410,83 | -0,19 | 0,15 | -1,29 | 1,98E-01 | 4,09E-01 |
| HMOX2 | 872,38 | -0,05 | 0,04 | -1,29 | 1,98E-01 | 4,09E-01 |
| ARL17A | 257,25 | 0,20 | 0,15 | 1,29 | 1,98E-01 | 4,09E-01 |
| ZBTB43 | 1030,38 | 0,04 | 0,03 | 1,29 | 1,98E-01 | 4,09E-01 |

| | | | | | | |
|---|---|---|---|---|---|---|
| UHMK1 | 8634,88 | 0,08 | 0,06 | 1,29 | 1,98E-01 | 4,09E-01 |
| ZNF519 | 440,84 | 0,08 | 0,06 | 1,29 | 1,98E-01 | 4,09E-01 |
| COX17 | 403,23 | 0,07 | 0,05 | 1,29 | 1,98E-01 | 4,10E-01 |
| IL15RA | 261,70 | -0,07 | 0,06 | -1,29 | 1,98E-01 | 4,10E-01 |
| CSTPP1 | 138,38 | -0,11 | 0,09 | -1,29 | 1,99E-01 | 4,10E-01 |
| CASP7 | 595,38 | 0,07 | 0,05 | 1,29 | 1,99E-01 | 4,10E-01 |
| PIP4K2A | 13712,89 | -0,12 | 0,09 | -1,28 | 1,99E-01 | 4,10E-01 |
| ENSG000002 | 44,29 | -0,10 | 0,08 | -1,28 | 1,99E-01 | 4,10E-01 |
| FCHSD1 | 721,33 | -0,10 | 0,08 | -1,28 | 1,99E-01 | 4,10E-01 |
| HBA2 | 328,56 | -0,29 | 0,23 | -1,28 | 1,99E-01 | 4,10E-01 |
| FABP5 | 41,96 | -0,16 | 0,12 | -1,28 | 1,99E-01 | 4,10E-01 |
| MARCO | 219,16 | 0,18 | 0,14 | 1,28 | 1,99E-01 | 4,10E-01 |
| MTA1 | 391,50 | -0,14 | 0,11 | -1,28 | 1,99E-01 | 4,10E-01 |
| LTK | 158,68 | -0,17 | 0,13 | -1,28 | 1,99E-01 | 4,10E-01 |
| NSMF | 388,81 | -0,11 | 0,08 | -1,28 | 1,99E-01 | 4,10E-01 |
| CCN3 | 292,88 | -0,13 | 0,10 | -1,28 | 1,99E-01 | 4,10E-01 |
| TMX2 | 614,60 | 0,04 | 0,03 | 1,28 | 1,99E-01 | 4,10E-01 |
| MARS1 | 1970,17 | -0,07 | 0,06 | -1,28 | 1,99E-01 | 4,11E-01 |
| SP110 | 4945,35 | -0,08 | 0,06 | -1,28 | 1,99E-01 | 4,11E-01 |
| CORO1C | 2076,50 | -0,08 | 0,06 | -1,28 | 2,00E-01 | 4,11E-01 |
| SEC16A | 1482,13 | -0,14 | 0,11 | -1,28 | 2,00E-01 | 4,11E-01 |
| ATG4B | 694,36 | -0,08 | 0,06 | -1,28 | 2,00E-01 | 4,11E-01 |
| TARDBP | 1471,52 | 0,05 | 0,04 | 1,28 | 2,00E-01 | 4,11E-01 |
| SEC31B | 301,70 | -0,15 | 0,12 | -1,28 | 2,00E-01 | 4,11E-01 |
| ATP5MC2 | 1423,03 | 0,05 | 0,04 | 1,28 | 2,00E-01 | 4,11E-01 |
| PPME1 | 380,98 | -0,05 | 0,04 | -1,28 | 2,00E-01 | 4,11E-01 |
| SLC35G2 | 31,61 | -0,25 | 0,20 | -1,28 | 2,00E-01 | 4,11E-01 |
| AP3S1 | 751,42 | 0,09 | 0,07 | 1,28 | 2,00E-01 | 4,11E-01 |
| ZNF443 | 138,79 | 0,12 | 0,09 | 1,28 | 2,00E-01 | 4,11E-01 |
| LACC1 | 157,52 | 0,12 | 0,09 | 1,28 | 2,00E-01 | 4,12E-01 |
| DHRS11 | 73,34 | -0,13 | 0,10 | -1,28 | 2,00E-01 | 4,12E-01 |
| MAP1LC3B | 1797,06 | -0,07 | 0,05 | -1,28 | 2,01E-01 | 4,12E-01 |
| CAB39 | 6824,87 | 0,06 | 0,05 | 1,28 | 2,01E-01 | 4,12E-01 |
| MCM4 | 253,18 | -0,07 | 0,05 | -1,28 | 2,01E-01 | 4,12E-01 |
| FSIP1 | 53,92 | -0,16 | 0,12 | -1,28 | 2,01E-01 | 4,12E-01 |
| SLC8B1 | 1294,56 | -0,08 | 0,06 | -1,28 | 2,01E-01 | 4,12E-01 |
| TNF | 86,86 | -0,09 | 0,07 | -1,28 | 2,01E-01 | 4,12E-01 |
| DDX28 | 330,75 | 0,05 | 0,04 | 1,28 | 2,01E-01 | 4,13E-01 |
| CMIP | 1314,53 | -0,15 | 0,12 | -1,28 | 2,01E-01 | 4,13E-01 |
| MEIS1 | 217,33 | -0,15 | 0,11 | -1,28 | 2,01E-01 | 4,13E-01 |
| ZWINT | 68,86 | -0,12 | 0,09 | -1,28 | 2,01E-01 | 4,13E-01 |
| LIPC | 61,78 | -0,18 | 0,14 | -1,28 | 2,01E-01 | 4,13E-01 |
| RIMKLB | 627,30 | 0,07 | 0,06 | 1,28 | 2,01E-01 | 4,13E-01 |
| SCYL1 | 832,87 | -0,08 | 0,06 | -1,28 | 2,01E-01 | 4,13E-01 |
| PSD4 | 4988,27 | -0,09 | 0,07 | -1,28 | 2,02E-01 | 4,13E-01 |
| XRCC3 | 163,47 | -0,12 | 0,10 | -1,28 | 2,02E-01 | 4,13E-01 |

| | | | | | | |
|---|---|---|---|---|---|---|
| VPS13A | 3866,23 | 0,09 | 0,07 | 1,28 | 2,02E-01 | 4,13E-01 |
| DDX3X | 21753,34 | 0,08 | 0,06 | 1,28 | 2,02E-01 | 4,14E-01 |
| LAS1L | 717,15 | 0,05 | 0,04 | 1,28 | 2,02E-01 | 4,14E-01 |
| DCTN2 | 584,60 | -0,07 | 0,05 | -1,28 | 2,02E-01 | 4,14E-01 |
| PDE8A | 1116,65 | 0,05 | 0,04 | 1,28 | 2,02E-01 | 4,14E-01 |
| CREBL2 | 1889,89 | 0,07 | 0,05 | 1,28 | 2,02E-01 | 4,14E-01 |
| TBC1D19 | 454,02 | 0,10 | 0,08 | 1,28 | 2,02E-01 | 4,14E-01 |
| RPAP1 | 276,64 | -0,10 | 0,08 | -1,28 | 2,02E-01 | 4,14E-01 |
| ATP5PD | 886,12 | 0,07 | 0,06 | 1,28 | 2,02E-01 | 4,14E-01 |
| ZSWIM9 | 141,68 | -0,11 | 0,09 | -1,28 | 2,02E-01 | 4,14E-01 |
| PTPN23 | 319,49 | -0,13 | 0,10 | -1,28 | 2,02E-01 | 4,14E-01 |
| UNKL | 1269,95 | -0,11 | 0,08 | -1,28 | 2,02E-01 | 4,14E-01 |
| EIF3L | 2482,23 | 0,06 | 0,04 | 1,27 | 2,02E-01 | 4,14E-01 |
| COX14 | 312,00 | 0,08 | 0,06 | 1,27 | 2,03E-01 | 4,14E-01 |
| PRKD2 | 1386,72 | -0,12 | 0,09 | -1,27 | 2,03E-01 | 4,14E-01 |
| GLIPR1 | 7782,20 | 0,10 | 0,08 | 1,27 | 2,03E-01 | 4,14E-01 |
| COQ5 | 310,21 | 0,08 | 0,06 | 1,27 | 2,03E-01 | 4,14E-01 |
| CDKN2AIPNL | 211,00 | 0,07 | 0,05 | 1,27 | 2,03E-01 | 4,14E-01 |
| ZZEF1 | 5449,64 | -0,08 | 0,06 | -1,27 | 2,03E-01 | 4,14E-01 |
| PROCA1 | 115,40 | -0,14 | 0,11 | -1,27 | 2,03E-01 | 4,14E-01 |
| IZUMO4 | 46,67 | -0,13 | 0,10 | -1,27 | 2,03E-01 | 4,14E-01 |
| METTL23 | 532,52 | 0,06 | 0,04 | 1,27 | 2,03E-01 | 4,15E-01 |
| MXD4 | 2005,06 | -0,07 | 0,05 | -1,27 | 2,03E-01 | 4,15E-01 |
| GCSAML | 86,71 | -0,19 | 0,15 | -1,27 | 2,03E-01 | 4,15E-01 |
| FEM1A | 1991,22 | -0,07 | 0,05 | -1,27 | 2,03E-01 | 4,15E-01 |
| RNF13 | 5477,14 | 0,09 | 0,07 | 1,27 | 2,04E-01 | 4,15E-01 |
| COMMD9 | 643,81 | 0,04 | 0,03 | 1,27 | 2,04E-01 | 4,15E-01 |
| RRM2B | 2769,54 | 0,11 | 0,09 | 1,27 | 2,04E-01 | 4,16E-01 |
| RNF19A | 4655,23 | 0,07 | 0,06 | 1,27 | 2,04E-01 | 4,16E-01 |
| PPP2R1B | 1299,16 | 0,05 | 0,04 | 1,27 | 2,04E-01 | 4,16E-01 |
| CREBRF | 7367,77 | 0,07 | 0,06 | 1,27 | 2,04E-01 | 4,16E-01 |
| RAP2C | 1425,78 | 0,11 | 0,09 | 1,27 | 2,04E-01 | 4,16E-01 |
| CAMLG | 578,99 | 0,08 | 0,06 | 1,27 | 2,04E-01 | 4,16E-01 |
| BCO2 | 75,11 | -0,12 | 0,10 | -1,27 | 2,04E-01 | 4,16E-01 |
| ZMYND15 | 35,22 | -0,24 | 0,19 | -1,27 | 2,04E-01 | 4,16E-01 |
| FBXL12 | 290,00 | -0,06 | 0,05 | -1,27 | 2,04E-01 | 4,16E-01 |
| MRTFA | 1471,64 | -0,09 | 0,07 | -1,27 | 2,04E-01 | 4,16E-01 |
| ZNF335 | 808,88 | -0,10 | 0,07 | -1,27 | 2,04E-01 | 4,16E-01 |
| COL8A2 | 43,36 | -0,19 | 0,15 | -1,27 | 2,05E-01 | 4,16E-01 |
| TOP3B | 761,65 | -0,06 | 0,05 | -1,27 | 2,05E-01 | 4,16E-01 |
| NOL12 | 175,81 | -0,10 | 0,08 | -1,27 | 2,05E-01 | 4,16E-01 |
| MED10 | 522,15 | 0,05 | 0,04 | 1,27 | 2,05E-01 | 4,17E-01 |
| PTPN9 | 686,63 | -0,05 | 0,04 | -1,27 | 2,05E-01 | 4,17E-01 |
| OTUD7A | 196,98 | -0,14 | 0,11 | -1,27 | 2,05E-01 | 4,17E-01 |
| RIC1 | 3283,94 | 0,05 | 0,04 | 1,27 | 2,05E-01 | 4,17E-01 |
| SUMO3 | 362,94 | -0,06 | 0,05 | -1,27 | 2,05E-01 | 4,17E-01 |

| | | | | | | |
|---|---|---|---|---|---|---|
| RBM42 | 157,39 | -0,11 | 0,08 | -1,27 | 2,05E-01 | 4,17E-01 |
| RAG1 | 52,72 | -0,18 | 0,14 | -1,27 | 2,05E-01 | 4,17E-01 |
| MRPL16 | 404,48 | 0,04 | 0,04 | 1,27 | 2,05E-01 | 4,17E-01 |
| ANKH | 1653,15 | -0,11 | 0,09 | -1,27 | 2,05E-01 | 4,17E-01 |
| ZNF280B | 65,97 | -0,13 | 0,10 | -1,27 | 2,05E-01 | 4,17E-01 |
| ZNF345 | 316,43 | 0,06 | 0,05 | 1,27 | 2,05E-01 | 4,17E-01 |
| HOOK3 | 3475,53 | 0,05 | 0,04 | 1,27 | 2,05E-01 | 4,17E-01 |
| TMA7B | 222,87 | -0,11 | 0,08 | -1,27 | 2,06E-01 | 4,18E-01 |
| IL23A | 137,93 | 0,09 | 0,07 | 1,27 | 2,06E-01 | 4,18E-01 |
| ZNF821 | 155,33 | -0,07 | 0,05 | -1,26 | 2,06E-01 | 4,18E-01 |
| HOXB4 | 76,85 | -0,11 | 0,09 | -1,26 | 2,06E-01 | 4,18E-01 |
| KLHL12 | 1286,65 | 0,05 | 0,04 | 1,26 | 2,06E-01 | 4,19E-01 |
| TECPR2 | 3474,64 | -0,11 | 0,09 | -1,26 | 2,06E-01 | 4,19E-01 |
| BANP | 2344,12 | -0,06 | 0,05 | -1,26 | 2,06E-01 | 4,19E-01 |
| AP1B1 | 557,80 | -0,12 | 0,09 | -1,26 | 2,06E-01 | 4,19E-01 |
| SMARCD2 | 2179,74 | 0,05 | 0,04 | 1,26 | 2,06E-01 | 4,19E-01 |
| SLC27A2 | 36,22 | -0,17 | 0,13 | -1,26 | 2,06E-01 | 4,19E-01 |
| MFF | 1481,16 | 0,08 | 0,06 | 1,26 | 2,06E-01 | 4,19E-01 |
| KDM1B | 1556,34 | -0,08 | 0,06 | -1,26 | 2,07E-01 | 4,19E-01 |
| CENPU | 79,30 | 0,12 | 0,10 | 1,26 | 2,07E-01 | 4,19E-01 |
| SMYD5 | 167,85 | -0,11 | 0,09 | -1,26 | 2,07E-01 | 4,19E-01 |
| ANKS1A | 1041,28 | -0,08 | 0,07 | -1,26 | 2,07E-01 | 4,19E-01 |
| NDUFC1 | 334,56 | 0,06 | 0,05 | 1,26 | 2,07E-01 | 4,19E-01 |
| EPC2 | 2240,02 | 0,06 | 0,05 | 1,26 | 2,07E-01 | 4,19E-01 |
| ZNF154 | 536,30 | 0,13 | 0,10 | 1,26 | 2,07E-01 | 4,19E-01 |
| SPICE1 | 285,59 | 0,06 | 0,05 | 1,26 | 2,07E-01 | 4,19E-01 |
| GALNS | 734,95 | -0,15 | 0,12 | -1,26 | 2,07E-01 | 4,19E-01 |
| ELP1 | 1756,50 | 0,05 | 0,04 | 1,26 | 2,07E-01 | 4,19E-01 |
| IQCN | 255,41 | -0,17 | 0,13 | -1,26 | 2,07E-01 | 4,19E-01 |
| MPO | 170,95 | -0,21 | 0,16 | -1,26 | 2,07E-01 | 4,19E-01 |
| PREX1 | 14510,59 | -0,10 | 0,08 | -1,26 | 2,07E-01 | 4,19E-01 |
| KIAA1328 | 521,31 | 0,06 | 0,05 | 1,26 | 2,07E-01 | 4,19E-01 |
| ZNF592 | 1799,09 | -0,10 | 0,08 | -1,26 | 2,07E-01 | 4,19E-01 |
| ERG28 | 163,86 | -0,05 | 0,04 | -1,26 | 2,07E-01 | 4,19E-01 |
| TTYH2 | 267,56 | -0,13 | 0,10 | -1,26 | 2,07E-01 | 4,19E-01 |
| TMEM126A | 135,88 | 0,13 | 0,10 | 1,26 | 2,08E-01 | 4,20E-01 |
| GTPBP3 | 153,54 | -0,10 | 0,08 | -1,26 | 2,08E-01 | 4,20E-01 |
| SATB1 | 3866,20 | 0,07 | 0,05 | 1,26 | 2,08E-01 | 4,20E-01 |
| RTN4 | 2273,49 | 0,06 | 0,05 | 1,26 | 2,08E-01 | 4,20E-01 |
| COG2 | 1185,33 | 0,04 | 0,03 | 1,26 | 2,08E-01 | 4,20E-01 |
| GCC1 | 1763,95 | 0,06 | 0,05 | 1,26 | 2,08E-01 | 4,20E-01 |
| TMEM175 | 269,17 | -0,12 | 0,10 | -1,26 | 2,08E-01 | 4,20E-01 |
| SNAPC4 | 269,35 | -0,13 | 0,10 | -1,26 | 2,08E-01 | 4,20E-01 |
| RDH16 | 50,25 | -0,15 | 0,12 | -1,26 | 2,08E-01 | 4,20E-01 |
| BACE1 | 75,75 | -0,17 | 0,13 | -1,26 | 2,08E-01 | 4,20E-01 |
| PRSS53 | 60,56 | -0,14 | 0,11 | -1,26 | 2,08E-01 | 4,21E-01 |

| | | | | | | |
|---|---|---|---|---|---|---|
| BOLA3 | 81,96 | -0,09 | 0,07 | -1,26 | 2,08E-01 | 4,21E-01 |
| GLIS3 | 85,58 | -0,24 | 0,19 | -1,26 | 2,08E-01 | 4,21E-01 |
| CTDNEP1 | 595,13 | -0,11 | 0,08 | -1,26 | 2,09E-01 | 4,21E-01 |
| ANO8 | 202,54 | -0,13 | 0,10 | -1,26 | 2,09E-01 | 4,21E-01 |
| EML4 | 4978,63 | 0,06 | 0,05 | 1,26 | 2,09E-01 | 4,21E-01 |
| BBS1 | 56,71 | -0,10 | 0,08 | -1,26 | 2,09E-01 | 4,21E-01 |
| SPATA2L | 136,72 | -0,10 | 0,08 | -1,26 | 2,09E-01 | 4,21E-01 |
| RTTN | 1748,00 | 0,06 | 0,05 | 1,26 | 2,09E-01 | 4,21E-01 |
| CD300LB | 789,46 | -0,10 | 0,08 | -1,26 | 2,09E-01 | 4,21E-01 |
| TYK2 | 5142,95 | -0,08 | 0,06 | -1,26 | 2,09E-01 | 4,21E-01 |
| ITGB3 | 1420,19 | -0,24 | 0,19 | -1,26 | 2,09E-01 | 4,21E-01 |
| YIF1B | 151,53 | -0,10 | 0,08 | -1,26 | 2,09E-01 | 4,21E-01 |
| MROH8 | 60,32 | -0,13 | 0,11 | -1,26 | 2,09E-01 | 4,21E-01 |
| DNTTIP1 | 744,97 | -0,07 | 0,06 | -1,26 | 2,09E-01 | 4,21E-01 |
| SLC5A3 | 4271,77 | 0,08 | 0,07 | 1,26 | 2,09E-01 | 4,22E-01 |
| CUEDC1 | 324,12 | -0,11 | 0,09 | -1,26 | 2,09E-01 | 4,22E-01 |
| HIPK1 | 7777,86 | -0,06 | 0,05 | -1,25 | 2,10E-01 | 4,22E-01 |
| SNRNP27 | 291,06 | 0,08 | 0,06 | 1,25 | 2,10E-01 | 4,22E-01 |
| TSSK6 | 90,64 | -0,07 | 0,06 | -1,25 | 2,10E-01 | 4,22E-01 |
| G6PD | 656,48 | -0,13 | 0,11 | -1,25 | 2,10E-01 | 4,22E-01 |
| EMD | 202,33 | -0,08 | 0,07 | -1,25 | 2,10E-01 | 4,22E-01 |
| TACO1 | 265,48 | 0,04 | 0,03 | 1,25 | 2,10E-01 | 4,22E-01 |
| THEM6 | 234,62 | -0,07 | 0,05 | -1,25 | 2,10E-01 | 4,22E-01 |
| GART | 1171,57 | 0,04 | 0,03 | 1,25 | 2,10E-01 | 4,22E-01 |
| SEC22C | 1263,07 | 0,04 | 0,03 | 1,25 | 2,10E-01 | 4,22E-01 |
| RAB8B | 6592,94 | 0,11 | 0,09 | 1,25 | 2,10E-01 | 4,22E-01 |
| C14orf93 | 295,39 | -0,04 | 0,03 | -1,25 | 2,10E-01 | 4,22E-01 |
| FUT4 | 652,54 | 0,06 | 0,05 | 1,25 | 2,10E-01 | 4,23E-01 |
| GNB1 | 3595,83 | -0,07 | 0,05 | -1,25 | 2,10E-01 | 4,23E-01 |
| TNFSF13B | 3097,03 | 0,11 | 0,09 | 1,25 | 2,11E-01 | 4,23E-01 |
| NPIPB2 | 42,29 | -0,12 | 0,10 | -1,25 | 2,11E-01 | 4,23E-01 |
| ME2 | 2828,45 | 0,05 | 0,04 | 1,25 | 2,11E-01 | 4,23E-01 |
| ARMC2 | 354,18 | -0,06 | 0,05 | -1,25 | 2,11E-01 | 4,23E-01 |
| POLA2 | 109,64 | -0,09 | 0,07 | -1,25 | 2,10E-01 | 4,23E-01 |
| ZC3HC1 | 266,47 | 0,04 | 0,03 | 1,25 | 2,10E-01 | 4,23E-01 |
| RPS6KA3 | 7753,70 | 0,06 | 0,05 | 1,25 | 2,11E-01 | 4,23E-01 |
| GGH | 52,82 | -0,14 | 0,11 | -1,25 | 2,11E-01 | 4,23E-01 |
| DDB2 | 349,69 | -0,10 | 0,08 | -1,25 | 2,11E-01 | 4,23E-01 |
| ITPKB | 3409,43 | -0,08 | 0,07 | -1,25 | 2,11E-01 | 4,23E-01 |
| PXYLP1 | 492,41 | -0,07 | 0,06 | -1,25 | 2,11E-01 | 4,23E-01 |
| KLHL14 | 181,80 | 0,16 | 0,13 | 1,25 | 2,11E-01 | 4,23E-01 |
| GRK6 | 3658,12 | -0,08 | 0,06 | -1,25 | 2,11E-01 | 4,23E-01 |
| DZANK1 | 155,77 | -0,11 | 0,09 | -1,25 | 2,11E-01 | 4,23E-01 |
| BACE2 | 363,15 | -0,12 | 0,10 | -1,25 | 2,11E-01 | 4,23E-01 |
| XKR3 | 51,81 | -0,29 | 0,23 | -1,25 | 2,11E-01 | 4,23E-01 |
| NOXRED1 | 93,93 | -0,07 | 0,05 | -1,25 | 2,11E-01 | 4,23E-01 |

| | | | | | | |
|---|---|---|---|---|---|---|
| MPHOSPH8 | 2742,50 | 0,04 | 0,03 | 1,25 | 2,11E-01 | 4,23E-01 |
| FAM210A | 315,22 | 0,06 | 0,05 | 1,25 | 2,11E-01 | 4,23E-01 |
| OFD1 | 1997,28 | 0,07 | 0,05 | 1,25 | 2,11E-01 | 4,23E-01 |
| SUPT20H | 3625,58 | 0,07 | 0,06 | 1,25 | 2,11E-01 | 4,23E-01 |
| ZNF708 | 1308,85 | 0,10 | 0,08 | 1,25 | 2,12E-01 | 4,23E-01 |
| PIGC | 1403,89 | 0,14 | 0,11 | 1,25 | 2,12E-01 | 4,24E-01 |
| USP15 | 21792,83 | 0,11 | 0,09 | 1,25 | 2,12E-01 | 4,24E-01 |
| NAA60 | 531,05 | -0,07 | 0,06 | -1,25 | 2,12E-01 | 4,24E-01 |
| PKD1 | 664,36 | -0,18 | 0,14 | -1,25 | 2,12E-01 | 4,24E-01 |
| S100A13 | 33,46 | -0,10 | 0,08 | -1,25 | 2,12E-01 | 4,24E-01 |
| ANAPC10 | 173,36 | 0,08 | 0,06 | 1,25 | 2,12E-01 | 4,24E-01 |
| CIMAP2 | 85,15 | -0,16 | 0,13 | -1,25 | 2,12E-01 | 4,24E-01 |
| BCORL1 | 233,08 | -0,14 | 0,11 | -1,25 | 2,12E-01 | 4,24E-01 |
| CERK | 3974,45 | -0,04 | 0,03 | -1,25 | 2,12E-01 | 4,24E-01 |
| CHI3L1 | 2119,86 | 0,18 | 0,15 | 1,25 | 2,12E-01 | 4,24E-01 |
| GSTA4 | 32,68 | -0,15 | 0,12 | -1,25 | 2,12E-01 | 4,25E-01 |
| CLASRP | 380,25 | -0,12 | 0,10 | -1,25 | 2,13E-01 | 4,25E-01 |
| PFKFB3 | 1873,29 | -0,12 | 0,09 | -1,25 | 2,13E-01 | 4,25E-01 |
| JAG1 | 246,54 | -0,10 | 0,08 | -1,25 | 2,13E-01 | 4,25E-01 |
| KDM5B | 2203,84 | -0,05 | 0,04 | -1,25 | 2,13E-01 | 4,25E-01 |
| FBXO34 | 1253,13 | 0,04 | 0,04 | 1,25 | 2,13E-01 | 4,25E-01 |
| LSM2 | 51,54 | -0,09 | 0,07 | -1,25 | 2,13E-01 | 4,25E-01 |
| GABARAPL2 | 2296,43 | -0,11 | 0,09 | -1,25 | 2,13E-01 | 4,25E-01 |
| ATP6V1C1 | 1481,21 | 0,07 | 0,05 | 1,25 | 2,13E-01 | 4,25E-01 |
| MRPL23 | 147,22 | -0,15 | 0,12 | -1,25 | 2,13E-01 | 4,25E-01 |
| MARCHF5 | 702,19 | 0,05 | 0,04 | 1,25 | 2,13E-01 | 4,25E-01 |
| ABR | 1750,38 | -0,13 | 0,10 | -1,25 | 2,13E-01 | 4,25E-01 |
| ABCB8 | 297,61 | -0,11 | 0,09 | -1,25 | 2,13E-01 | 4,25E-01 |
| SEMA4A | 1563,97 | -0,12 | 0,10 | -1,25 | 2,13E-01 | 4,25E-01 |
| EDA | 98,34 | -0,16 | 0,13 | -1,25 | 2,13E-01 | 4,25E-01 |
| CDKN3 | 30,96 | -0,17 | 0,14 | -1,24 | 2,13E-01 | 4,25E-01 |
| ZBP1 | 1629,37 | -0,10 | 0,08 | -1,24 | 2,13E-01 | 4,25E-01 |
| TPM2 | 64,30 | -0,16 | 0,13 | -1,24 | 2,13E-01 | 4,25E-01 |
| GPR135 | 138,96 | -0,10 | 0,08 | -1,24 | 2,14E-01 | 4,25E-01 |
| ENSG000002 | 66,43 | -0,17 | 0,14 | -1,24 | 2,13E-01 | 4,25E-01 |
| PIGH | 337,74 | 0,05 | 0,04 | 1,24 | 2,14E-01 | 4,25E-01 |
| ATRN | 1208,77 | 0,04 | 0,03 | 1,24 | 2,14E-01 | 4,25E-01 |
| ARL8A | 402,38 | -0,09 | 0,07 | -1,24 | 2,14E-01 | 4,26E-01 |
| CAPG | 968,87 | 0,08 | 0,07 | 1,24 | 2,14E-01 | 4,26E-01 |
| FAM111B | 73,29 | 0,18 | 0,14 | 1,24 | 2,14E-01 | 4,26E-01 |
| CTNNB1 | 6557,50 | 0,05 | 0,04 | 1,24 | 2,14E-01 | 4,26E-01 |
| MIA2 | 2216,38 | 0,06 | 0,05 | 1,24 | 2,14E-01 | 4,26E-01 |
| SPOPL | 6130,26 | 0,11 | 0,09 | 1,24 | 2,14E-01 | 4,26E-01 |
| CHST12 | 974,74 | -0,14 | 0,11 | -1,24 | 2,14E-01 | 4,26E-01 |
| WDR91 | 773,18 | -0,06 | 0,05 | -1,24 | 2,15E-01 | 4,26E-01 |
| GAA | 970,85 | -0,12 | 0,10 | -1,24 | 2,14E-01 | 4,26E-01 |

| | | | | | | |
|---|---|---|---|---|---|---|
| PDP2 | 286,87 | -0,06 | 0,05 | -1,24 | 2,14E-01 | 4,26E-01 |
| PGP | 439,46 | -0,05 | 0,04 | -1,24 | 2,15E-01 | 4,26E-01 |
| PRDX3 | 1513,93 | 0,06 | 0,05 | 1,24 | 2,15E-01 | 4,26E-01 |
| GAK | 2210,17 | -0,11 | 0,09 | -1,24 | 2,15E-01 | 4,26E-01 |
| EDC4 | 471,33 | -0,11 | 0,09 | -1,24 | 2,14E-01 | 4,26E-01 |
| C5orf22 | 701,03 | 0,05 | 0,04 | 1,24 | 2,15E-01 | 4,26E-01 |
| PTH2R | 57,65 | -0,15 | 0,12 | -1,24 | 2,15E-01 | 4,26E-01 |
| AP4S1 | 502,77 | 0,05 | 0,04 | 1,24 | 2,14E-01 | 4,26E-01 |
| ARHGAP27 | 3165,22 | -0,12 | 0,10 | -1,24 | 2,14E-01 | 4,26E-01 |
| MRPL24 | 332,03 | 0,06 | 0,05 | 1,24 | 2,14E-01 | 4,26E-01 |
| STAU2 | 1432,32 | 0,06 | 0,05 | 1,24 | 2,15E-01 | 4,26E-01 |
| RPL30 | 9525,47 | 0,15 | 0,12 | 1,24 | 2,15E-01 | 4,26E-01 |
| RAB29 | 1803,57 | 0,05 | 0,04 | 1,24 | 2,15E-01 | 4,27E-01 |
| SNX15 | 54,83 | -0,10 | 0,08 | -1,24 | 2,15E-01 | 4,27E-01 |
| VANGL1 | 202,61 | -0,10 | 0,08 | -1,24 | 2,15E-01 | 4,27E-01 |
| KRT18 | 57,44 | -0,14 | 0,11 | -1,24 | 2,15E-01 | 4,27E-01 |
| COPS7A | 575,33 | -0,04 | 0,04 | -1,24 | 2,15E-01 | 4,27E-01 |
| TLR4 | 13476,83 | 0,10 | 0,08 | 1,24 | 2,15E-01 | 4,27E-01 |
| B3GNT2 | 821,94 | 0,08 | 0,06 | 1,24 | 2,15E-01 | 4,27E-01 |
| RBM4B | 383,90 | 0,04 | 0,03 | 1,24 | 2,16E-01 | 4,27E-01 |
| CCDC89 | 53,75 | 0,14 | 0,11 | 1,24 | 2,16E-01 | 4,27E-01 |
| CDIP1 | 241,79 | -0,11 | 0,09 | -1,24 | 2,16E-01 | 4,27E-01 |
| CREBBP | 4195,64 | -0,10 | 0,08 | -1,24 | 2,16E-01 | 4,27E-01 |
| WDPCP | 673,86 | 0,06 | 0,05 | 1,24 | 2,16E-01 | 4,27E-01 |
| SLC37A4 | 186,23 | -0,07 | 0,06 | -1,24 | 2,16E-01 | 4,27E-01 |
| CHPT1 | 2316,86 | -0,11 | 0,09 | -1,24 | 2,16E-01 | 4,27E-01 |
| TCIRG1 | 3679,44 | -0,11 | 0,09 | -1,24 | 2,16E-01 | 4,27E-01 |
| LYZ | 76925,04 | 0,13 | 0,10 | 1,24 | 2,16E-01 | 4,28E-01 |
| RPS27L | 831,62 | 0,09 | 0,07 | 1,24 | 2,16E-01 | 4,28E-01 |
| RNF168 | 1449,99 | 0,05 | 0,04 | 1,24 | 2,16E-01 | 4,28E-01 |
| MCF2L2 | 299,85 | -0,14 | 0,11 | -1,24 | 2,16E-01 | 4,28E-01 |
| PIDD1 | 155,63 | -0,13 | 0,11 | -1,24 | 2,16E-01 | 4,28E-01 |
| RPP21 | 51,66 | -0,09 | 0,07 | -1,24 | 2,16E-01 | 4,28E-01 |
| RUVBL1 | 663,42 | 0,05 | 0,04 | 1,24 | 2,16E-01 | 4,28E-01 |
| RBFA | 191,88 | -0,06 | 0,05 | -1,24 | 2,17E-01 | 4,28E-01 |
| NR1H3 | 102,76 | -0,10 | 0,08 | -1,24 | 2,17E-01 | 4,28E-01 |
| KIT | 116,31 | -0,12 | 0,10 | -1,24 | 2,17E-01 | 4,28E-01 |
| COQ10B | 1394,53 | 0,07 | 0,06 | 1,24 | 2,17E-01 | 4,28E-01 |
| PIK3R5 | 3788,80 | -0,10 | 0,08 | -1,24 | 2,16E-01 | 4,28E-01 |
| GPR137B | 190,23 | -0,07 | 0,06 | -1,24 | 2,17E-01 | 4,28E-01 |
| BAZ1B | 2313,52 | 0,05 | 0,04 | 1,24 | 2,17E-01 | 4,28E-01 |
| IFIT1B | 1011,12 | -0,19 | 0,15 | -1,24 | 2,17E-01 | 4,28E-01 |
| BBS7 | 295,83 | 0,08 | 0,06 | 1,23 | 2,17E-01 | 4,28E-01 |
| PRXL2B | 156,69 | -0,09 | 0,07 | -1,24 | 2,17E-01 | 4,28E-01 |
| PFKFB2 | 981,22 | -0,09 | 0,07 | -1,24 | 2,17E-01 | 4,28E-01 |
| TNIP3 | 82,83 | 0,17 | 0,14 | 1,23 | 2,17E-01 | 4,28E-01 |

| | | | | | | |
|---|---|---|---|---|---|---|
| GNB2 | 3090,84 | -0,10 | 0,08 | -1,23 | 2,17E-01 | 4,28E-01 |
| OSTM1 | 2455,71 | 0,09 | 0,07 | 1,23 | 2,17E-01 | 4,29E-01 |
| LEPROTL1 | 2960,26 | 0,07 | 0,05 | 1,23 | 2,17E-01 | 4,29E-01 |
| SVIL | 3953,11 | -0,09 | 0,08 | -1,23 | 2,17E-01 | 4,29E-01 |
| AFF4 | 3840,52 | 0,04 | 0,04 | 1,23 | 2,17E-01 | 4,29E-01 |
| IWS1 | 1760,47 | 0,05 | 0,04 | 1,23 | 2,17E-01 | 4,29E-01 |
| CCDC180 | 100,38 | -0,15 | 0,13 | -1,23 | 2,17E-01 | 4,29E-01 |
| COQ8B | 625,40 | -0,06 | 0,05 | -1,23 | 2,18E-01 | 4,29E-01 |
| TMEM30A | 4706,79 | 0,09 | 0,07 | 1,23 | 2,18E-01 | 4,29E-01 |
| PARP15 | 2105,88 | 0,09 | 0,07 | 1,23 | 2,18E-01 | 4,29E-01 |
| PLCL2 | 4897,78 | 0,06 | 0,05 | 1,23 | 2,18E-01 | 4,29E-01 |
| CTC1 | 1927,75 | -0,09 | 0,07 | -1,23 | 2,18E-01 | 4,29E-01 |
| ZFP90 | 1100,41 | 0,06 | 0,05 | 1,23 | 2,18E-01 | 4,29E-01 |
| TMEM65 | 3334,82 | 0,07 | 0,05 | 1,23 | 2,18E-01 | 4,29E-01 |
| SLC35A5 | 1356,10 | 0,07 | 0,05 | 1,23 | 2,18E-01 | 4,29E-01 |
| PPA1 | 573,03 | 0,11 | 0,09 | 1,23 | 2,18E-01 | 4,29E-01 |
| GNB4 | 3459,39 | 0,09 | 0,07 | 1,23 | 2,18E-01 | 4,29E-01 |
| MRPL40 | 138,21 | 0,10 | 0,08 | 1,23 | 2,18E-01 | 4,29E-01 |
| CCDC92 | 534,85 | -0,06 | 0,05 | -1,23 | 2,18E-01 | 4,29E-01 |
| ACTN1 | 3562,51 | -0,12 | 0,10 | -1,23 | 2,18E-01 | 4,29E-01 |
| ATP6V1F | 155,11 | -0,07 | 0,06 | -1,23 | 2,18E-01 | 4,29E-01 |
| CCDC7 | 863,25 | 0,10 | 0,08 | 1,23 | 2,18E-01 | 4,29E-01 |
| SMAP2 | 9414,00 | -0,09 | 0,08 | -1,23 | 2,18E-01 | 4,29E-01 |
| HS3ST1 | 59,41 | -0,20 | 0,16 | -1,23 | 2,18E-01 | 4,29E-01 |
| MANBAL | 344,99 | -0,05 | 0,04 | -1,23 | 2,18E-01 | 4,29E-01 |
| IPPK | 509,77 | -0,04 | 0,03 | -1,23 | 2,18E-01 | 4,29E-01 |
| TRIOBP | 1626,33 | -0,09 | 0,07 | -1,23 | 2,18E-01 | 4,29E-01 |
| BICD1 | 421,04 | 0,07 | 0,06 | 1,23 | 2,18E-01 | 4,29E-01 |
| FLOT2 | 2403,11 | -0,12 | 0,10 | -1,23 | 2,18E-01 | 4,29E-01 |
| RNF31 | 701,05 | -0,08 | 0,06 | -1,23 | 2,18E-01 | 4,29E-01 |
| PHC2 | 2148,23 | -0,14 | 0,11 | -1,23 | 2,18E-01 | 4,29E-01 |
| SH2B2 | 252,89 | -0,13 | 0,11 | -1,23 | 2,19E-01 | 4,29E-01 |
| TMEM214 | 273,54 | -0,10 | 0,08 | -1,23 | 2,19E-01 | 4,29E-01 |
| DYNC2H1 | 465,03 | 0,09 | 0,08 | 1,23 | 2,19E-01 | 4,29E-01 |
| EIF3K | 2384,34 | 0,06 | 0,05 | 1,23 | 2,19E-01 | 4,29E-01 |
| FAM50B | 181,19 | -0,07 | 0,06 | -1,23 | 2,19E-01 | 4,29E-01 |
| TTBK2 | 1725,98 | 0,04 | 0,03 | 1,23 | 2,19E-01 | 4,29E-01 |
| KBTBD4 | 674,02 | 0,05 | 0,04 | 1,23 | 2,19E-01 | 4,29E-01 |
| ACO2 | 604,88 | -0,08 | 0,07 | -1,23 | 2,19E-01 | 4,29E-01 |
| KMT2B | 597,50 | -0,20 | 0,16 | -1,23 | 2,19E-01 | 4,30E-01 |
| ACCS | 562,11 | -0,16 | 0,13 | -1,23 | 2,19E-01 | 4,30E-01 |
| IFIT2 | 6364,41 | 0,17 | 0,14 | 1,23 | 2,19E-01 | 4,30E-01 |
| ABCC10 | 914,97 | -0,09 | 0,07 | -1,23 | 2,19E-01 | 4,30E-01 |
| ZSWIM1 | 137,00 | -0,09 | 0,07 | -1,23 | 2,19E-01 | 4,30E-01 |
| NDUFA4 | 822,08 | 0,15 | 0,12 | 1,23 | 2,19E-01 | 4,30E-01 |
| ZNF558 | 441,88 | 0,06 | 0,05 | 1,23 | 2,20E-01 | 4,30E-01 |

| | | | | | | |
|---|---|---|---|---|---|---|
| LONRF1 | 918,62 | 0,06 | 0,05 | 1,23 | 2,20E-01 | 4,30E-01 |
| ODF2L | 2384,36 | 0,09 | 0,08 | 1,23 | 2,20E-01 | 4,31E-01 |
| PPP1R26 | 217,25 | -0,10 | 0,08 | -1,23 | 2,20E-01 | 4,31E-01 |
| ZNF394 | 2405,63 | 0,04 | 0,04 | 1,23 | 2,20E-01 | 4,31E-01 |
| RRP7A | 1296,11 | -0,08 | 0,06 | -1,23 | 2,20E-01 | 4,31E-01 |
| EEF1D | 7009,43 | 0,08 | 0,06 | 1,23 | 2,20E-01 | 4,31E-01 |
| UBA7 | 1336,70 | -0,09 | 0,07 | -1,23 | 2,20E-01 | 4,31E-01 |
| PSMD2 | 2044,11 | -0,05 | 0,04 | -1,23 | 2,20E-01 | 4,31E-01 |
| KIF1C | 478,91 | -0,11 | 0,09 | -1,23 | 2,20E-01 | 4,31E-01 |
| BRI3 | 1163,37 | -0,07 | 0,06 | -1,23 | 2,20E-01 | 4,31E-01 |
| F5 | 1559,16 | -0,10 | 0,08 | -1,23 | 2,20E-01 | 4,31E-01 |
| NABP2 | 122,04 | -0,06 | 0,05 | -1,23 | 2,20E-01 | 4,31E-01 |
| UXT | 792,49 | 0,08 | 0,07 | 1,22 | 2,21E-01 | 4,31E-01 |
| ACBD6 | 1250,04 | 0,04 | 0,03 | 1,22 | 2,21E-01 | 4,32E-01 |
| JAK2 | 5736,96 | 0,10 | 0,08 | 1,22 | 2,21E-01 | 4,32E-01 |
| PPP1CB | 8683,31 | 0,12 | 0,10 | 1,22 | 2,21E-01 | 4,32E-01 |
| CHCHD6 | 149,41 | -0,09 | 0,07 | -1,22 | 2,21E-01 | 4,32E-01 |
| SVIP | 836,21 | 0,14 | 0,12 | 1,22 | 2,21E-01 | 4,32E-01 |
| UBE2O | 1184,04 | -0,16 | 0,13 | -1,22 | 2,21E-01 | 4,32E-01 |
| ZNHIT1 | 733,51 | -0,04 | 0,04 | -1,22 | 2,21E-01 | 4,32E-01 |
| TMPPE | 662,68 | -0,09 | 0,07 | -1,22 | 2,21E-01 | 4,32E-01 |
| GIT2 | 4785,57 | -0,06 | 0,05 | -1,22 | 2,21E-01 | 4,32E-01 |
| REELD1 | 47,31 | -0,16 | 0,13 | -1,22 | 2,21E-01 | 4,32E-01 |
| MED24 | 388,43 | -0,09 | 0,07 | -1,22 | 2,21E-01 | 4,32E-01 |
| BRD3 | 847,72 | -0,08 | 0,07 | -1,22 | 2,21E-01 | 4,32E-01 |
| PPP1R18 | 1453,70 | -0,15 | 0,12 | -1,22 | 2,21E-01 | 4,32E-01 |
| TUBA1B | 3227,32 | -0,05 | 0,04 | -1,22 | 2,21E-01 | 4,32E-01 |
| STPG2 | 22,87 | -0,16 | 0,13 | -1,22 | 2,21E-01 | 4,32E-01 |
| ZC3H4 | 912,99 | -0,08 | 0,06 | -1,22 | 2,21E-01 | 4,32E-01 |
| AKT1 | 3352,29 | -0,07 | 0,05 | -1,22 | 2,21E-01 | 4,32E-01 |
| YDJC | 268,68 | -0,06 | 0,05 | -1,22 | 2,21E-01 | 4,32E-01 |
| SNAPC1 | 134,07 | 0,10 | 0,08 | 1,22 | 2,22E-01 | 4,32E-01 |
| PET117 | 61,92 | 0,09 | 0,07 | 1,22 | 2,22E-01 | 4,32E-01 |
| NBAS | 3029,77 | 0,06 | 0,05 | 1,22 | 2,22E-01 | 4,32E-01 |
| PIKFYVE | 5242,41 | 0,05 | 0,04 | 1,22 | 2,22E-01 | 4,33E-01 |
| KCNJ15 | 6803,33 | -0,13 | 0,11 | -1,22 | 2,22E-01 | 4,33E-01 |
| MEGF6 | 1527,28 | -0,10 | 0,08 | -1,22 | 2,22E-01 | 4,33E-01 |
| HLA-DMA | 899,64 | 0,06 | 0,05 | 1,22 | 2,22E-01 | 4,33E-01 |
| FXR2 | 560,21 | -0,05 | 0,04 | -1,22 | 2,22E-01 | 4,33E-01 |
| HIPK2 | 1589,83 | -0,11 | 0,09 | -1,22 | 2,22E-01 | 4,33E-01 |
| CERS6 | 1230,98 | 0,06 | 0,05 | 1,22 | 2,23E-01 | 4,33E-01 |
| MON1A | 164,69 | -0,07 | 0,05 | -1,22 | 2,23E-01 | 4,33E-01 |
| UCHL3 | 98,57 | 0,09 | 0,07 | 1,22 | 2,23E-01 | 4,33E-01 |
| CSNK1G1 | 678,31 | -0,06 | 0,05 | -1,22 | 2,23E-01 | 4,33E-01 |
| UBR4 | 6958,49 | -0,09 | 0,07 | -1,22 | 2,23E-01 | 4,33E-01 |
| CBFA2T3 | 335,13 | -0,15 | 0,12 | -1,22 | 2,23E-01 | 4,34E-01 |

| | | | | | | |
|---|---|---|---|---|---|---|
| CLN5 | 740,96 | 0,06 | 0,05 | 1,22 | 2,23E-01 | 4,34E-01 |
| SLC19A1 | 1851,13 | -0,14 | 0,12 | -1,22 | 2,23E-01 | 4,34E-01 |
| HSPH1 | 1039,75 | 0,09 | 0,07 | 1,22 | 2,23E-01 | 4,34E-01 |
| HOOK1 | 289,19 | 0,12 | 0,10 | 1,22 | 2,23E-01 | 4,34E-01 |
| STX11 | 1884,62 | -0,09 | 0,08 | -1,22 | 2,23E-01 | 4,34E-01 |
| HS6ST1 | 227,12 | -0,09 | 0,07 | -1,22 | 2,23E-01 | 4,34E-01 |
| GALNT6 | 664,78 | -0,08 | 0,06 | -1,22 | 2,23E-01 | 4,34E-01 |
| UBOX5 | 193,86 | -0,10 | 0,08 | -1,22 | 2,23E-01 | 4,34E-01 |
| SLC35E3 | 898,21 | -0,08 | 0,07 | -1,22 | 2,23E-01 | 4,34E-01 |
| EFCAB13 | 351,88 | 0,12 | 0,10 | 1,22 | 2,23E-01 | 4,34E-01 |
| SERP1 | 3814,25 | 0,09 | 0,07 | 1,22 | 2,23E-01 | 4,34E-01 |
| ADGRG1 | 642,45 | -0,16 | 0,13 | -1,22 | 2,24E-01 | 4,34E-01 |
| SLC35E2B | 1016,00 | -0,12 | 0,10 | -1,22 | 2,24E-01 | 4,34E-01 |
| C11orf24 | 167,58 | -0,07 | 0,06 | -1,22 | 2,24E-01 | 4,34E-01 |
| IMMP1L | 364,52 | 0,08 | 0,07 | 1,22 | 2,24E-01 | 4,34E-01 |
| RFWD3 | 883,16 | -0,04 | 0,03 | -1,22 | 2,24E-01 | 4,34E-01 |
| BRPF3 | 672,68 | -0,07 | 0,06 | -1,22 | 2,24E-01 | 4,34E-01 |
| TRIM27 | 2676,45 | -0,05 | 0,04 | -1,22 | 2,24E-01 | 4,34E-01 |
| NOP16 | 130,41 | 0,05 | 0,04 | 1,22 | 2,24E-01 | 4,34E-01 |
| LEAP2 | 84,76 | -0,06 | 0,05 | -1,22 | 2,24E-01 | 4,34E-01 |
| MAU2 | 1593,01 | -0,11 | 0,09 | -1,22 | 2,24E-01 | 4,34E-01 |
| PARP16 | 208,09 | -0,08 | 0,07 | -1,22 | 2,24E-01 | 4,35E-01 |
| CAPN15 | 501,69 | -0,19 | 0,15 | -1,21 | 2,24E-01 | 4,35E-01 |
| NOB1 | 148,04 | -0,07 | 0,06 | -1,21 | 2,25E-01 | 4,35E-01 |
| SNTG2 | 69,63 | -0,22 | 0,18 | -1,21 | 2,24E-01 | 4,35E-01 |
| HLA-DRB1 | 4588,89 | 0,09 | 0,08 | 1,21 | 2,24E-01 | 4,35E-01 |
| SELENOW | 400,80 | -0,06 | 0,05 | -1,21 | 2,25E-01 | 4,35E-01 |
| MGAT4B | 853,88 | -0,09 | 0,07 | -1,21 | 2,25E-01 | 4,35E-01 |
| ZFYVE28 | 373,14 | -0,13 | 0,11 | -1,21 | 2,25E-01 | 4,36E-01 |
| ZNF207 | 11844,78 | 0,06 | 0,05 | 1,21 | 2,25E-01 | 4,36E-01 |
| CYP4F12 | 189,74 | -0,14 | 0,11 | -1,21 | 2,25E-01 | 4,36E-01 |
| MARK4 | 255,33 | -0,13 | 0,11 | -1,21 | 2,26E-01 | 4,37E-01 |
| SAMD3 | 1665,24 | 0,11 | 0,09 | 1,21 | 2,26E-01 | 4,37E-01 |
| KLRF1 | 977,42 | 0,16 | 0,13 | 1,21 | 2,26E-01 | 4,37E-01 |
| FBH1 | 1769,50 | -0,05 | 0,04 | -1,21 | 2,26E-01 | 4,37E-01 |
| ARSD | 428,45 | -0,07 | 0,06 | -1,21 | 2,26E-01 | 4,37E-01 |
| ZBTB2 | 394,97 | 0,05 | 0,04 | 1,21 | 2,26E-01 | 4,37E-01 |
| ABCA7 | 2218,33 | -0,15 | 0,12 | -1,21 | 2,26E-01 | 4,37E-01 |
| ZNF81 | 1074,51 | 0,05 | 0,04 | 1,21 | 2,26E-01 | 4,37E-01 |
| NUBPL | 302,53 | 0,06 | 0,05 | 1,21 | 2,26E-01 | 4,37E-01 |
| PTBP1 | 1971,41 | -0,07 | 0,06 | -1,21 | 2,26E-01 | 4,37E-01 |
| HAX1 | 913,70 | 0,06 | 0,05 | 1,21 | 2,26E-01 | 4,37E-01 |
| SH3GLB2 | 1152,31 | -0,08 | 0,06 | -1,21 | 2,26E-01 | 4,38E-01 |
| IRF1 | 16477,34 | -0,07 | 0,06 | -1,21 | 2,27E-01 | 4,38E-01 |
| TKFC | 188,47 | -0,10 | 0,08 | -1,21 | 2,27E-01 | 4,38E-01 |
| SMIM7 | 1069,45 | 0,04 | 0,03 | 1,21 | 2,27E-01 | 4,38E-01 |

| | | | | | | |
|---|---|---|---|---|---|---|
| LGR6 | 114,26 | -0,18 | 0,15 | -1,21 | 2,27E-01 | 4,38E-01 |
| GTF2B | 759,68 | 0,08 | 0,07 | 1,21 | 2,27E-01 | 4,38E-01 |
| YJU2B | 424,24 | -0,08 | 0,06 | -1,21 | 2,27E-01 | 4,38E-01 |
| ZHX2 | 555,97 | -0,08 | 0,07 | -1,21 | 2,27E-01 | 4,38E-01 |
| PTPRK | 210,60 | 0,13 | 0,11 | 1,21 | 2,27E-01 | 4,38E-01 |
| MCU | 777,01 | -0,05 | 0,04 | -1,21 | 2,27E-01 | 4,38E-01 |
| MTCH1 | 2128,72 | -0,05 | 0,04 | -1,21 | 2,27E-01 | 4,38E-01 |
| PAFAH1B3 | 121,38 | -0,06 | 0,05 | -1,21 | 2,27E-01 | 4,38E-01 |
| IFNGR1 | 5701,67 | 0,10 | 0,09 | 1,21 | 2,27E-01 | 4,38E-01 |
| PSPH | 75,27 | -0,11 | 0,09 | -1,21 | 2,27E-01 | 4,38E-01 |
| PELP1 | 225,02 | -0,11 | 0,09 | -1,21 | 2,27E-01 | 4,38E-01 |
| CUX1 | 1854,61 | -0,09 | 0,08 | -1,21 | 2,27E-01 | 4,38E-01 |
| PIP5K1B | 519,92 | -0,08 | 0,07 | -1,21 | 2,28E-01 | 4,39E-01 |
| PFDN4 | 137,61 | 0,13 | 0,11 | 1,21 | 2,28E-01 | 4,39E-01 |
| PPP1R14B | 192,35 | 0,07 | 0,06 | 1,21 | 2,28E-01 | 4,39E-01 |
| PGLS | 456,99 | -0,09 | 0,07 | -1,21 | 2,28E-01 | 4,39E-01 |
| DLG3 | 242,47 | -0,09 | 0,07 | -1,21 | 2,28E-01 | 4,39E-01 |
| TAS2R30 | 391,63 | 0,10 | 0,08 | 1,21 | 2,28E-01 | 4,39E-01 |
| STK17B | 19061,24 | 0,09 | 0,07 | 1,21 | 2,28E-01 | 4,39E-01 |
| SMAD7 | 554,13 | -0,08 | 0,06 | -1,21 | 2,28E-01 | 4,39E-01 |
| FBXO30 | 1597,37 | 0,10 | 0,08 | 1,20 | 2,28E-01 | 4,39E-01 |
| OBSCN | 1521,44 | -0,14 | 0,12 | -1,20 | 2,28E-01 | 4,39E-01 |
| IRAG2 | 5598,02 | 0,08 | 0,06 | 1,21 | 2,28E-01 | 4,39E-01 |
| PRDM11 | 233,34 | -0,13 | 0,11 | -1,20 | 2,28E-01 | 4,39E-01 |
| RNF146 | 2200,79 | 0,08 | 0,07 | 1,20 | 2,28E-01 | 4,39E-01 |
| ATP5F1C | 1086,26 | 0,10 | 0,08 | 1,20 | 2,28E-01 | 4,39E-01 |
| EDC3 | 437,46 | -0,07 | 0,05 | -1,20 | 2,28E-01 | 4,39E-01 |
| GTPBP1 | 2094,13 | -0,09 | 0,07 | -1,20 | 2,29E-01 | 4,39E-01 |
| NEK4 | 881,62 | 0,04 | 0,03 | 1,20 | 2,29E-01 | 4,39E-01 |
| MRPL54 | 402,45 | 0,08 | 0,07 | 1,20 | 2,29E-01 | 4,39E-01 |
| ITGAL | 5083,58 | -0,10 | 0,08 | -1,20 | 2,29E-01 | 4,40E-01 |
| CEBPB | 4158,71 | -0,09 | 0,07 | -1,20 | 2,29E-01 | 4,40E-01 |
| PLCXD2 | 959,80 | 0,08 | 0,06 | 1,20 | 2,29E-01 | 4,40E-01 |
| ZFAND5 | 5208,60 | 0,07 | 0,06 | 1,20 | 2,29E-01 | 4,40E-01 |
| PEF1 | 689,37 | -0,04 | 0,03 | -1,20 | 2,29E-01 | 4,40E-01 |
| SMIM12 | 1267,69 | 0,03 | 0,02 | 1,20 | 2,29E-01 | 4,40E-01 |
| PCGF6 | 246,28 | 0,07 | 0,06 | 1,20 | 2,29E-01 | 4,40E-01 |
| FOXO3 | 10261,00 | -0,12 | 0,10 | -1,20 | 2,29E-01 | 4,40E-01 |
| CBFA2T2 | 588,44 | -0,10 | 0,08 | -1,20 | 2,29E-01 | 4,40E-01 |
| NSUN5 | 496,68 | -0,05 | 0,04 | -1,20 | 2,29E-01 | 4,40E-01 |
| APOBEC3F | 151,48 | -0,11 | 0,09 | -1,20 | 2,29E-01 | 4,40E-01 |
| LAX1 | 707,78 | 0,07 | 0,06 | 1,20 | 2,29E-01 | 4,40E-01 |
| ZMAT4 | 42,11 | -0,26 | 0,22 | -1,20 | 2,29E-01 | 4,40E-01 |
| ZNF605 | 525,00 | 0,08 | 0,07 | 1,20 | 2,30E-01 | 4,41E-01 |
| BID | 2338,45 | -0,06 | 0,05 | -1,20 | 2,30E-01 | 4,41E-01 |
| EMC8 | 236,79 | -0,06 | 0,05 | -1,20 | 2,30E-01 | 4,41E-01 |

| | | | | | | |
|---|---|---|---|---|---|---|
| TRAF2 | 416,22 | -0,06 | 0,05 | -1,20 | 2,30E-01 | 4,41E-01 |
| CALML4 | 94,32 | 0,06 | 0,05 | 1,20 | 2,30E-01 | 4,41E-01 |
| CCAR2 | 652,31 | -0,10 | 0,08 | -1,20 | 2,30E-01 | 4,41E-01 |
| PLA2G4B | 53,64 | -0,13 | 0,11 | -1,20 | 2,30E-01 | 4,41E-01 |
| MAD2L1 | 120,98 | 0,11 | 0,09 | 1,20 | 2,30E-01 | 4,41E-01 |
| FBXL17 | 637,59 | 0,05 | 0,04 | 1,20 | 2,30E-01 | 4,41E-01 |
| PGGHG | 2183,14 | -0,15 | 0,13 | -1,20 | 2,30E-01 | 4,42E-01 |
| IFI16 | 5382,10 | 0,08 | 0,07 | 1,20 | 2,31E-01 | 4,42E-01 |
| MYO1B | 91,89 | -0,21 | 0,18 | -1,20 | 2,31E-01 | 4,42E-01 |
| ENSG000002 | 23,89 | 0,11 | 0,09 | 1,20 | 2,31E-01 | 4,42E-01 |
| CAMTA2 | 356,16 | -0,17 | 0,14 | -1,20 | 2,31E-01 | 4,42E-01 |
| PLD3 | 475,30 | -0,10 | 0,09 | -1,20 | 2,31E-01 | 4,42E-01 |
| ENSG000002 | 23,11 | -0,09 | 0,07 | -1,20 | 2,31E-01 | 4,42E-01 |
| UBL4A | 641,88 | -0,06 | 0,05 | -1,20 | 2,31E-01 | 4,42E-01 |
| PLEKHM3 | 1700,27 | -0,06 | 0,05 | -1,20 | 2,31E-01 | 4,42E-01 |
| TMEM64 | 630,28 | 0,08 | 0,07 | 1,20 | 2,31E-01 | 4,42E-01 |
| SKI | 1031,99 | -0,14 | 0,11 | -1,20 | 2,31E-01 | 4,42E-01 |
| ZNF444 | 224,78 | -0,13 | 0,11 | -1,20 | 2,31E-01 | 4,42E-01 |
| ENSG000002 | 25,25 | -0,15 | 0,12 | -1,20 | 2,31E-01 | 4,42E-01 |
| VPS4B | 4312,89 | 0,06 | 0,05 | 1,20 | 2,31E-01 | 4,42E-01 |
| SLFN11 | 1915,29 | 0,06 | 0,05 | 1,20 | 2,31E-01 | 4,42E-01 |
| FMNL1 | 3393,50 | -0,18 | 0,15 | -1,20 | 2,31E-01 | 4,42E-01 |
| COL6A2 | 356,93 | -0,16 | 0,13 | -1,20 | 2,31E-01 | 4,42E-01 |
| PLIN4 | 318,88 | -0,16 | 0,13 | -1,20 | 2,31E-01 | 4,42E-01 |
| H1-0 | 839,59 | -0,18 | 0,15 | -1,20 | 2,31E-01 | 4,42E-01 |
| MRPL46 | 333,94 | 0,06 | 0,05 | 1,20 | 2,31E-01 | 4,42E-01 |
| BRMS1 | 699,37 | 0,05 | 0,05 | 1,20 | 2,31E-01 | 4,42E-01 |
| SZT2 | 2101,44 | -0,10 | 0,08 | -1,20 | 2,31E-01 | 4,42E-01 |
| C12orf60 | 28,59 | -0,17 | 0,14 | -1,20 | 2,31E-01 | 4,42E-01 |
| CLC | 2103,60 | -0,20 | 0,16 | -1,20 | 2,31E-01 | 4,42E-01 |
| PHYH | 114,63 | -0,08 | 0,07 | -1,20 | 2,32E-01 | 4,42E-01 |
| HINFP | 435,96 | -0,07 | 0,06 | -1,20 | 2,32E-01 | 4,42E-01 |
| USP48 | 3344,40 | 0,05 | 0,04 | 1,20 | 2,32E-01 | 4,42E-01 |
| FZD2 | 77,36 | -0,10 | 0,08 | -1,20 | 2,32E-01 | 4,42E-01 |
| CSNK1G2 | 2492,35 | -0,07 | 0,06 | -1,20 | 2,32E-01 | 4,42E-01 |
| SH3RF3 | 201,98 | -0,14 | 0,12 | -1,20 | 2,32E-01 | 4,42E-01 |
| PSMC2 | 763,77 | 0,08 | 0,07 | 1,20 | 2,32E-01 | 4,42E-01 |
| SYS1 | 514,68 | -0,05 | 0,04 | -1,20 | 2,32E-01 | 4,42E-01 |
| PPIA | 5244,75 | 0,08 | 0,07 | 1,20 | 2,32E-01 | 4,42E-01 |
| BEST1 | 1410,00 | -0,11 | 0,09 | -1,19 | 2,32E-01 | 4,43E-01 |
| BCAS3 | 1831,77 | -0,05 | 0,04 | -1,19 | 2,32E-01 | 4,43E-01 |
| MTG2 | 580,94 | -0,09 | 0,07 | -1,19 | 2,32E-01 | 4,43E-01 |
| MYCBP2 | 13671,64 | 0,06 | 0,05 | 1,19 | 2,32E-01 | 4,43E-01 |
| LARP1 | 2148,14 | -0,08 | 0,07 | -1,19 | 2,33E-01 | 4,43E-01 |
| HRH2 | 2289,50 | -0,10 | 0,09 | -1,19 | 2,33E-01 | 4,43E-01 |
| CCNI2 | 84,36 | -0,09 | 0,08 | -1,19 | 2,33E-01 | 4,43E-01 |

| | | | | | | |
|---|---|---|---|---|---|---|
| MYO9B | 5122,44 | -0,11 | 0,09 | -1,19 | 2,33E-01 | 4,43E-01 |
| TMEM38A | 84,18 | -0,09 | 0,08 | -1,19 | 2,33E-01 | 4,43E-01 |
| LGALS3 | 1498,60 | -0,13 | 0,11 | -1,19 | 2,33E-01 | 4,43E-01 |
| HAL | 2432,15 | -0,09 | 0,08 | -1,19 | 2,33E-01 | 4,43E-01 |
| CLDND1 | 1927,43 | 0,08 | 0,07 | 1,19 | 2,33E-01 | 4,43E-01 |
| ZNF276 | 3804,43 | -0,05 | 0,04 | -1,19 | 2,33E-01 | 4,43E-01 |
| TIMM8B | 320,69 | 0,13 | 0,11 | 1,19 | 2,33E-01 | 4,43E-01 |
| EEF1AKMT4 | 22,95 | -0,09 | 0,08 | -1,19 | 2,33E-01 | 4,43E-01 |
| NDUFC2 | 374,57 | 0,07 | 0,06 | 1,19 | 2,33E-01 | 4,43E-01 |
| TTLL3 | 779,54 | -0,13 | 0,11 | -1,19 | 2,33E-01 | 4,43E-01 |
| ACTR5 | 420,41 | 0,04 | 0,04 | 1,19 | 2,33E-01 | 4,43E-01 |
| NSL1 | 1040,60 | 0,07 | 0,06 | 1,19 | 2,33E-01 | 4,43E-01 |
| GPR171 | 455,28 | 0,12 | 0,10 | 1,19 | 2,33E-01 | 4,43E-01 |
| GATD1 | 344,98 | -0,11 | 0,09 | -1,19 | 2,33E-01 | 4,43E-01 |
| CCZ1B | 581,40 | 0,07 | 0,06 | 1,19 | 2,34E-01 | 4,44E-01 |
| NRROS | 195,48 | -0,14 | 0,12 | -1,19 | 2,34E-01 | 4,44E-01 |
| ABHD16A | 275,17 | -0,05 | 0,04 | -1,19 | 2,34E-01 | 4,44E-01 |
| DCK | 2117,09 | 0,09 | 0,07 | 1,19 | 2,34E-01 | 4,44E-01 |
| CELF2 | 14484,19 | -0,05 | 0,04 | -1,19 | 2,34E-01 | 4,44E-01 |
| C18orf54 | 189,41 | 0,09 | 0,08 | 1,19 | 2,34E-01 | 4,44E-01 |
| FGL2 | 20062,13 | 0,11 | 0,10 | 1,19 | 2,34E-01 | 4,44E-01 |
| MCRS1 | 402,34 | -0,07 | 0,06 | -1,19 | 2,34E-01 | 4,44E-01 |
| TRIM58 | 14643,36 | -0,18 | 0,15 | -1,19 | 2,34E-01 | 4,44E-01 |
| FTSJ1 | 214,95 | -0,06 | 0,05 | -1,19 | 2,34E-01 | 4,44E-01 |
| PHACTR2 | 1666,29 | 0,05 | 0,05 | 1,19 | 2,34E-01 | 4,44E-01 |
| CXCR1 | 7617,81 | -0,10 | 0,08 | -1,19 | 2,34E-01 | 4,44E-01 |
| SIRPB1 | 1584,82 | -0,12 | 0,10 | -1,19 | 2,34E-01 | 4,44E-01 |
| EML3 | 678,76 | -0,12 | 0,10 | -1,19 | 2,34E-01 | 4,44E-01 |
| SLC25A33 | 129,48 | -0,06 | 0,05 | -1,19 | 2,34E-01 | 4,44E-01 |
| TMEM94 | 327,51 | -0,13 | 0,11 | -1,19 | 2,34E-01 | 4,44E-01 |
| ZSCAN22 | 141,49 | -0,05 | 0,05 | -1,19 | 2,34E-01 | 4,44E-01 |
| SLFN12L | 4030,90 | 0,08 | 0,07 | 1,19 | 2,34E-01 | 4,44E-01 |
| EIF2B1 | 943,55 | 0,03 | 0,03 | 1,19 | 2,35E-01 | 4,44E-01 |
| NECAP2 | 1887,99 | -0,04 | 0,03 | -1,19 | 2,35E-01 | 4,44E-01 |
| ING4 | 826,13 | 0,05 | 0,04 | 1,19 | 2,35E-01 | 4,44E-01 |
| KIZ | 800,55 | 0,04 | 0,03 | 1,19 | 2,35E-01 | 4,44E-01 |
| NAT1 | 252,91 | 0,08 | 0,07 | 1,19 | 2,35E-01 | 4,44E-01 |
| FKRP | 221,02 | -0,11 | 0,09 | -1,19 | 2,35E-01 | 4,44E-01 |
| SLC18A2 | 101,98 | -0,13 | 0,11 | -1,19 | 2,35E-01 | 4,44E-01 |
| FAM193B | 791,22 | -0,14 | 0,12 | -1,19 | 2,35E-01 | 4,44E-01 |
| NFKBID | 337,41 | -0,09 | 0,08 | -1,19 | 2,35E-01 | 4,44E-01 |
| INPP4B | 5042,85 | 0,10 | 0,08 | 1,19 | 2,35E-01 | 4,44E-01 |
| CMTM3 | 889,56 | -0,05 | 0,04 | -1,19 | 2,35E-01 | 4,44E-01 |
| SLC35F6 | 368,05 | -0,10 | 0,08 | -1,19 | 2,35E-01 | 4,44E-01 |
| TBC1D10B | 421,55 | -0,14 | 0,12 | -1,19 | 2,35E-01 | 4,44E-01 |
| MKKS | 458,91 | 0,07 | 0,06 | 1,19 | 2,35E-01 | 4,45E-01 |

| | | | | | | |
|---|---|---|---|---|---|---|
| UQCRH | 849,96 | 0,11 | 0,10 | 1,19 | 2,35E-01 | 4,45E-01 |
| NOD1 | 722,65 | -0,07 | 0,06 | -1,19 | 2,35E-01 | 4,45E-01 |
| CDA | 447,38 | -0,12 | 0,10 | -1,19 | 2,36E-01 | 4,45E-01 |
| ZCRB1 | 203,56 | 0,08 | 0,07 | 1,19 | 2,36E-01 | 4,45E-01 |
| DHX35 | 597,07 | 0,04 | 0,03 | 1,19 | 2,36E-01 | 4,45E-01 |
| OTUD3 | 751,39 | 0,05 | 0,04 | 1,19 | 2,36E-01 | 4,45E-01 |
| N4BP2L2 | 12129,50 | 0,06 | 0,05 | 1,19 | 2,36E-01 | 4,45E-01 |
| DENND4B | 2042,32 | -0,11 | 0,09 | -1,19 | 2,36E-01 | 4,45E-01 |
| ATAD3A | 153,27 | -0,09 | 0,08 | -1,19 | 2,36E-01 | 4,45E-01 |
| MAP4K5 | 1671,09 | 0,07 | 0,06 | 1,19 | 2,36E-01 | 4,45E-01 |
| PLA2G4A | 306,48 | 0,10 | 0,08 | 1,19 | 2,36E-01 | 4,45E-01 |
| SCARF1 | 1021,06 | -0,13 | 0,11 | -1,19 | 2,36E-01 | 4,45E-01 |
| PPM1N | 98,61 | -0,09 | 0,08 | -1,18 | 2,36E-01 | 4,46E-01 |
| JMJD1C | 12272,52 | 0,07 | 0,06 | 1,18 | 2,36E-01 | 4,46E-01 |
| DHX34 | 1805,95 | -0,10 | 0,08 | -1,18 | 2,36E-01 | 4,46E-01 |
| SAMD8 | 2611,28 | 0,07 | 0,06 | 1,18 | 2,36E-01 | 4,46E-01 |
| GCNT1 | 815,82 | 0,07 | 0,06 | 1,18 | 2,37E-01 | 4,46E-01 |
| TMEM185B | 518,85 | -0,05 | 0,05 | -1,18 | 2,37E-01 | 4,46E-01 |
| INVS | 885,80 | 0,05 | 0,04 | 1,18 | 2,37E-01 | 4,46E-01 |
| PERP | 55,78 | -0,14 | 0,12 | -1,18 | 2,37E-01 | 4,46E-01 |
| CD300A | 3083,96 | -0,07 | 0,06 | -1,18 | 2,37E-01 | 4,46E-01 |
| SLC9A8 | 1319,52 | -0,08 | 0,07 | -1,18 | 2,37E-01 | 4,46E-01 |
| LTF | 609,07 | -0,25 | 0,21 | -1,18 | 2,37E-01 | 4,46E-01 |
| TRANK1 | 11617,57 | -0,08 | 0,07 | -1,18 | 2,37E-01 | 4,46E-01 |
| TSEN54 | 313,76 | -0,09 | 0,08 | -1,18 | 2,37E-01 | 4,47E-01 |
| SVBP | 389,46 | -0,10 | 0,09 | -1,18 | 2,37E-01 | 4,47E-01 |
| NLN | 308,83 | 0,09 | 0,07 | 1,18 | 2,37E-01 | 4,47E-01 |
| EPS15L1 | 1084,76 | -0,11 | 0,10 | -1,18 | 2,37E-01 | 4,47E-01 |
| CKS2 | 62,60 | 0,14 | 0,12 | 1,18 | 2,37E-01 | 4,47E-01 |
| KNTC1 | 1026,75 | 0,06 | 0,05 | 1,18 | 2,38E-01 | 4,47E-01 |
| POLR3B | 467,69 | 0,05 | 0,04 | 1,18 | 2,38E-01 | 4,48E-01 |
| HENMT1 | 465,66 | 0,07 | 0,06 | 1,18 | 2,38E-01 | 4,48E-01 |
| ARHGEF2 | 4374,51 | -0,08 | 0,07 | -1,18 | 2,38E-01 | 4,48E-01 |
| KCNH8 | 118,08 | 0,15 | 0,13 | 1,18 | 2,38E-01 | 4,48E-01 |
| CYP1B1 | 893,83 | 0,11 | 0,10 | 1,18 | 2,38E-01 | 4,48E-01 |
| UBAP1L | 401,99 | -0,12 | 0,10 | -1,18 | 2,38E-01 | 4,48E-01 |
| ISCU | 3076,26 | -0,07 | 0,06 | -1,18 | 2,38E-01 | 4,48E-01 |
| RNF40 | 2299,01 | -0,07 | 0,06 | -1,18 | 2,38E-01 | 4,48E-01 |
| PTOV1 | 820,60 | -0,07 | 0,06 | -1,18 | 2,38E-01 | 4,48E-01 |
| SPART | 1470,36 | 0,07 | 0,06 | 1,18 | 2,38E-01 | 4,48E-01 |
| CCDC93 | 3324,13 | 0,04 | 0,04 | 1,18 | 2,38E-01 | 4,48E-01 |
| ENKUR | 123,71 | -0,13 | 0,11 | -1,18 | 2,39E-01 | 4,48E-01 |
| INTS5 | 215,58 | -0,09 | 0,08 | -1,18 | 2,39E-01 | 4,48E-01 |
| AMMECR1 | 624,38 | 0,06 | 0,05 | 1,18 | 2,39E-01 | 4,48E-01 |
| DEF8 | 3205,68 | -0,08 | 0,07 | -1,18 | 2,39E-01 | 4,48E-01 |
| ATP6V1G1 | 1782,87 | 0,11 | 0,09 | 1,18 | 2,39E-01 | 4,48E-01 |

| | | | | | | |
|---|---|---|---|---|---|---|
| PYROXD2 | 443,22 | -0,07 | 0,06 | -1,18 | 2,39E-01 | 4,49E-01 |
| SHKBP1 | 1980,14 | -0,12 | 0,10 | -1,18 | 2,39E-01 | 4,49E-01 |
| CIMAP1B | 528,51 | -0,13 | 0,11 | -1,18 | 2,39E-01 | 4,49E-01 |
| CNTNAP2 | 116,36 | -0,27 | 0,23 | -1,18 | 2,39E-01 | 4,49E-01 |
| GCAT | 148,95 | -0,19 | 0,16 | -1,18 | 2,39E-01 | 4,49E-01 |
| UBE2D2 | 1462,37 | 0,04 | 0,04 | 1,18 | 2,39E-01 | 4,49E-01 |
| MFAP1 | 884,13 | 0,07 | 0,06 | 1,18 | 2,39E-01 | 4,49E-01 |
| FAM200B | 945,72 | 0,10 | 0,09 | 1,18 | 2,39E-01 | 4,49E-01 |
| DENND2D | 3415,91 | 0,06 | 0,05 | 1,18 | 2,39E-01 | 4,49E-01 |
| SLC35D2 | 210,34 | -0,04 | 0,04 | -1,18 | 2,39E-01 | 4,49E-01 |
| LILRB4 | 123,22 | -0,13 | 0,11 | -1,18 | 2,40E-01 | 4,49E-01 |
| IPP | 387,02 | 0,04 | 0,03 | 1,18 | 2,40E-01 | 4,49E-01 |
| CRTC1 | 326,14 | -0,13 | 0,11 | -1,18 | 2,40E-01 | 4,49E-01 |
| POLR3K | 193,07 | -0,05 | 0,04 | -1,18 | 2,40E-01 | 4,49E-01 |
| DDTL | 29,52 | -0,24 | 0,21 | -1,18 | 2,40E-01 | 4,49E-01 |
| IFT81 | 79,47 | -0,11 | 0,10 | -1,18 | 2,40E-01 | 4,49E-01 |
| GOLGA3 | 2576,35 | -0,07 | 0,06 | -1,18 | 2,40E-01 | 4,49E-01 |
| CDYL2 | 356,69 | -0,07 | 0,06 | -1,17 | 2,40E-01 | 4,49E-01 |
| ILRUN | 1961,98 | -0,06 | 0,05 | -1,17 | 2,40E-01 | 4,50E-01 |
| DUS4L | 214,27 | 0,06 | 0,05 | 1,17 | 2,40E-01 | 4,50E-01 |
| ATF7IP | 7482,75 | 0,05 | 0,04 | 1,17 | 2,40E-01 | 4,50E-01 |
| SEC24A | 817,01 | 0,06 | 0,05 | 1,17 | 2,41E-01 | 4,50E-01 |
| TAFA1 | 83,35 | 0,12 | 0,10 | 1,17 | 2,41E-01 | 4,50E-01 |
| TIMM10 | 224,26 | 0,09 | 0,08 | 1,17 | 2,41E-01 | 4,50E-01 |
| PIK3IP1 | 4304,86 | 0,07 | 0,06 | 1,17 | 2,41E-01 | 4,50E-01 |
| PRKAA1 | 3607,47 | 0,06 | 0,05 | 1,17 | 2,41E-01 | 4,50E-01 |
| RNF19B | 1442,58 | -0,08 | 0,07 | -1,17 | 2,41E-01 | 4,50E-01 |
| SCML4 | 1710,54 | -0,07 | 0,06 | -1,17 | 2,41E-01 | 4,50E-01 |
| MIF4GD | 205,15 | -0,05 | 0,04 | -1,17 | 2,41E-01 | 4,50E-01 |
| FERMT3 | 1620,84 | -0,10 | 0,09 | -1,17 | 2,41E-01 | 4,50E-01 |
| WASF1 | 45,79 | -0,13 | 0,11 | -1,17 | 2,41E-01 | 4,50E-01 |
| R3HCC1 | 430,62 | 0,04 | 0,03 | 1,17 | 2,41E-01 | 4,50E-01 |
| FOXN2 | 3773,12 | 0,09 | 0,08 | 1,17 | 2,41E-01 | 4,50E-01 |
| NDUFAF1 | 382,98 | 0,06 | 0,05 | 1,17 | 2,41E-01 | 4,50E-01 |
| LTB4R | 1355,96 | -0,08 | 0,07 | -1,17 | 2,41E-01 | 4,50E-01 |
| KMT5B | 2851,40 | 0,05 | 0,04 | 1,17 | 2,41E-01 | 4,50E-01 |
| RPL21 | 8167,47 | 0,19 | 0,16 | 1,17 | 2,41E-01 | 4,50E-01 |
| APC | 3776,22 | 0,07 | 0,06 | 1,17 | 2,41E-01 | 4,50E-01 |
| KLF6 | 7733,18 | -0,05 | 0,05 | -1,17 | 2,41E-01 | 4,50E-01 |
| NT5C3A | 1754,23 | 0,09 | 0,08 | 1,17 | 2,41E-01 | 4,50E-01 |
| RBM47 | 4441,09 | -0,09 | 0,07 | -1,17 | 2,41E-01 | 4,50E-01 |
| SURF6 | 902,53 | 0,04 | 0,03 | 1,17 | 2,41E-01 | 4,50E-01 |
| NAA40 | 200,65 | -0,14 | 0,12 | -1,17 | 2,42E-01 | 4,50E-01 |
| NLRP2 | 139,93 | -0,18 | 0,15 | -1,17 | 2,42E-01 | 4,50E-01 |
| GET4 | 154,21 | -0,10 | 0,08 | -1,17 | 2,42E-01 | 4,50E-01 |
| IKZF1 | 11321,61 | -0,05 | 0,04 | -1,17 | 2,42E-01 | 4,50E-01 |

| | | | | | | |
|---|---|---|---|---|---|---|
| TTC39B | 1974,00 | 0,07 | 0,06 | 1,17 | 2,42E-01 | 4,51E-01 |
| NAPRT | 577,82 | -0,09 | 0,07 | -1,17 | 2,42E-01 | 4,51E-01 |
| UBE3D | 403,82 | 0,06 | 0,05 | 1,17 | 2,42E-01 | 4,51E-01 |
| MOSPD2 | 2262,97 | 0,09 | 0,07 | 1,17 | 2,42E-01 | 4,51E-01 |
| FEZ2 | 1759,10 | 0,06 | 0,05 | 1,17 | 2,42E-01 | 4,51E-01 |
| PLA2G6 | 445,62 | -0,09 | 0,08 | -1,17 | 2,42E-01 | 4,51E-01 |
| KLHDC1 | 298,92 | 0,07 | 0,06 | 1,17 | 2,42E-01 | 4,52E-01 |
| FBXW5 | 1710,49 | -0,06 | 0,05 | -1,17 | 2,42E-01 | 4,52E-01 |
| OSCAR | 334,69 | -0,11 | 0,10 | -1,17 | 2,43E-01 | 4,52E-01 |
| EML6 | 188,01 | -0,15 | 0,13 | -1,17 | 2,43E-01 | 4,52E-01 |
| CLPP | 243,85 | -0,05 | 0,05 | -1,17 | 2,43E-01 | 4,52E-01 |
| KCTD10 | 760,62 | -0,06 | 0,05 | -1,17 | 2,43E-01 | 4,52E-01 |
| ATF6B | 561,49 | -0,11 | 0,09 | -1,17 | 2,43E-01 | 4,52E-01 |
| HGF | 501,21 | 0,10 | 0,08 | 1,17 | 2,43E-01 | 4,52E-01 |
| POU5F1B | 46,76 | -0,14 | 0,12 | -1,17 | 2,43E-01 | 4,52E-01 |
| FBXO36 | 33,13 | -0,12 | 0,10 | -1,17 | 2,43E-01 | 4,52E-01 |
| GSS | 301,91 | -0,05 | 0,04 | -1,17 | 2,43E-01 | 4,52E-01 |
| ZNF583 | 224,55 | 0,11 | 0,10 | 1,17 | 2,43E-01 | 4,53E-01 |
| AGO3 | 3687,42 | 0,04 | 0,04 | 1,17 | 2,43E-01 | 4,53E-01 |
| SP140L | 1899,27 | 0,05 | 0,04 | 1,17 | 2,43E-01 | 4,53E-01 |
| GADD45B | 614,16 | -0,08 | 0,07 | -1,17 | 2,44E-01 | 4,53E-01 |
| UBXN10 | 126,73 | -0,13 | 0,11 | -1,17 | 2,44E-01 | 4,53E-01 |
| ANKRD52 | 582,09 | -0,10 | 0,09 | -1,17 | 2,44E-01 | 4,53E-01 |
| LGALSL | 570,66 | -0,11 | 0,10 | -1,17 | 2,44E-01 | 4,53E-01 |
| TIMP2 | 779,03 | -0,14 | 0,12 | -1,17 | 2,44E-01 | 4,53E-01 |
| ATP5MC1 | 300,96 | 0,05 | 0,04 | 1,17 | 2,44E-01 | 4,53E-01 |
| SCAF1 | 322,35 | -0,14 | 0,12 | -1,16 | 2,44E-01 | 4,53E-01 |
| SCAMP2 | 886,46 | -0,07 | 0,06 | -1,16 | 2,44E-01 | 4,53E-01 |
| MINPP1 | 292,96 | 0,09 | 0,08 | 1,16 | 2,44E-01 | 4,53E-01 |
| NUP88 | 737,35 | 0,06 | 0,05 | 1,16 | 2,44E-01 | 4,53E-01 |
| ORC6 | 53,84 | -0,12 | 0,10 | -1,16 | 2,44E-01 | 4,53E-01 |
| ZNF568 | 449,45 | 0,06 | 0,05 | 1,16 | 2,44E-01 | 4,53E-01 |
| PRH1 | 360,04 | 0,09 | 0,08 | 1,16 | 2,44E-01 | 4,53E-01 |
| CEP15 | 42,78 | -0,16 | 0,14 | -1,16 | 2,45E-01 | 4,54E-01 |
| MAT2B | 5077,51 | 0,07 | 0,06 | 1,16 | 2,45E-01 | 4,54E-01 |
| LRP1B | 48,51 | -0,25 | 0,22 | -1,16 | 2,45E-01 | 4,54E-01 |
| ARHGAP9 | 3136,38 | -0,11 | 0,09 | -1,16 | 2,45E-01 | 4,54E-01 |
| HEXIM1 | 1331,86 | -0,05 | 0,05 | -1,16 | 2,45E-01 | 4,54E-01 |
| ARID5A | 617,13 | -0,10 | 0,08 | -1,16 | 2,45E-01 | 4,54E-01 |
| AATK | 1015,87 | -0,15 | 0,13 | -1,16 | 2,45E-01 | 4,54E-01 |
| SOX5 | 73,17 | -0,20 | 0,17 | -1,16 | 2,45E-01 | 4,54E-01 |
| SPI1 | 1060,97 | -0,13 | 0,11 | -1,16 | 2,45E-01 | 4,54E-01 |
| MED18 | 612,97 | 0,05 | 0,05 | 1,16 | 2,45E-01 | 4,54E-01 |
| PPP2R3C | 1268,99 | 0,08 | 0,07 | 1,16 | 2,45E-01 | 4,54E-01 |
| RAB1B | 714,85 | -0,11 | 0,09 | -1,16 | 2,45E-01 | 4,54E-01 |
| GSK3A | 446,73 | -0,09 | 0,08 | -1,16 | 2,45E-01 | 4,54E-01 |

| | | | | | | |
|---|---|---|---|---|---|---|
| CPSF7 | 1035,85 | -0,09 | 0,07 | -1,16 | 2,45E-01 | 4,54E-01 |
| DIAPH1 | 3830,32 | -0,09 | 0,08 | -1,16 | 2,46E-01 | 4,55E-01 |
| CD37 | 1983,80 | -0,11 | 0,09 | -1,16 | 2,46E-01 | 4,55E-01 |
| MEN1 | 174,85 | -0,11 | 0,09 | -1,16 | 2,46E-01 | 4,55E-01 |
| MAN2A2 | 3801,63 | -0,07 | 0,06 | -1,16 | 2,46E-01 | 4,55E-01 |
| DHX37 | 303,22 | -0,10 | 0,09 | -1,16 | 2,46E-01 | 4,55E-01 |
| PLEKHM2 | 571,13 | -0,11 | 0,09 | -1,16 | 2,46E-01 | 4,55E-01 |
| JPT1 | 1540,51 | -0,07 | 0,06 | -1,16 | 2,46E-01 | 4,55E-01 |
| GSK3B | 2827,48 | -0,05 | 0,05 | -1,16 | 2,46E-01 | 4,55E-01 |
| BRAP | 1309,47 | 0,04 | 0,04 | 1,16 | 2,46E-01 | 4,55E-01 |
| PRPF40B | 188,14 | -0,11 | 0,10 | -1,16 | 2,46E-01 | 4,55E-01 |
| ANKS6 | 240,47 | -0,09 | 0,08 | -1,16 | 2,46E-01 | 4,55E-01 |
| PRPF3 | 1767,14 | 0,03 | 0,03 | 1,16 | 2,46E-01 | 4,55E-01 |
| FLYWCH2 | 163,69 | 0,05 | 0,05 | 1,16 | 2,46E-01 | 4,55E-01 |
| GMIP | 2275,12 | -0,14 | 0,12 | -1,16 | 2,46E-01 | 4,55E-01 |
| KMT2E | 6723,10 | 0,04 | 0,04 | 1,16 | 2,47E-01 | 4,55E-01 |
| ANKRD13A | 7820,38 | -0,06 | 0,05 | -1,16 | 2,47E-01 | 4,56E-01 |
| DNAJA2 | 2166,97 | 0,07 | 0,06 | 1,16 | 2,47E-01 | 4,56E-01 |
| CDK5RAP1 | 770,55 | 0,03 | 0,03 | 1,16 | 2,47E-01 | 4,56E-01 |
| R3HDM1 | 767,49 | 0,05 | 0,04 | 1,16 | 2,47E-01 | 4,56E-01 |
| TSC22D4 | 728,32 | -0,15 | 0,13 | -1,16 | 2,47E-01 | 4,56E-01 |
| NAP1L5 | 37,34 | -0,12 | 0,11 | -1,16 | 2,47E-01 | 4,56E-01 |
| TMEM41B | 1111,62 | 0,06 | 0,05 | 1,16 | 2,47E-01 | 4,56E-01 |
| PLPBP | 849,18 | 0,03 | 0,03 | 1,16 | 2,47E-01 | 4,56E-01 |
| ZNF20 | 74,78 | -0,07 | 0,06 | -1,16 | 2,47E-01 | 4,56E-01 |
| KLRC2 | 166,31 | -0,25 | 0,21 | -1,16 | 2,47E-01 | 4,56E-01 |
| MCM3AP | 2252,54 | -0,07 | 0,06 | -1,16 | 2,47E-01 | 4,56E-01 |
| CCNG1 | 1505,75 | 0,09 | 0,08 | 1,16 | 2,47E-01 | 4,56E-01 |
| MAEA | 1867,75 | -0,07 | 0,06 | -1,16 | 2,47E-01 | 4,56E-01 |
| ZNF486 | 725,64 | 0,08 | 0,07 | 1,16 | 2,47E-01 | 4,56E-01 |
| GGTA1 | 140,85 | -0,13 | 0,11 | -1,16 | 2,47E-01 | 4,56E-01 |
| TTC38 | 898,57 | -0,08 | 0,07 | -1,16 | 2,47E-01 | 4,56E-01 |
| SPTB | 1936,95 | -0,21 | 0,18 | -1,16 | 2,47E-01 | 4,56E-01 |
| SOCS3 | 979,89 | -0,12 | 0,10 | -1,16 | 2,48E-01 | 4,56E-01 |
| KDM2A | 5062,39 | -0,06 | 0,05 | -1,16 | 2,48E-01 | 4,56E-01 |
| OXNAD1 | 5207,38 | 0,09 | 0,08 | 1,16 | 2,48E-01 | 4,56E-01 |
| ATG13 | 1188,80 | -0,06 | 0,05 | -1,16 | 2,48E-01 | 4,56E-01 |
| MAP3K2 | 11571,37 | 0,08 | 0,07 | 1,16 | 2,48E-01 | 4,56E-01 |
| EPHB6 | 197,12 | -0,13 | 0,11 | -1,16 | 2,48E-01 | 4,56E-01 |
| ANO5 | 115,36 | 0,14 | 0,12 | 1,16 | 2,48E-01 | 4,56E-01 |
| ABCC3 | 451,43 | -0,15 | 0,13 | -1,16 | 2,48E-01 | 4,57E-01 |
| RASAL3 | 867,27 | -0,13 | 0,11 | -1,15 | 2,48E-01 | 4,57E-01 |
| TOPORS | 2461,24 | 0,10 | 0,08 | 1,15 | 2,48E-01 | 4,57E-01 |
| BTBD10 | 1725,00 | 0,07 | 0,06 | 1,15 | 2,48E-01 | 4,57E-01 |
| PHF24 | 31,62 | -0,24 | 0,21 | -1,15 | 2,48E-01 | 4,57E-01 |
| ALPL | 1192,86 | -0,16 | 0,14 | -1,15 | 2,48E-01 | 4,57E-01 |

| | | | | | | |
|---|---|---|---|---|---|---|
| C12orf57 | 860,71 | 0,10 | 0,09 | 1,15 | 2,48E-01 | 4,57E-01 |
| PCSK5 | 721,48 | 0,06 | 0,06 | 1,15 | 2,48E-01 | 4,57E-01 |
| INTS6L | 1832,00 | 0,06 | 0,05 | 1,15 | 2,49E-01 | 4,57E-01 |
| ALYREF | 626,95 | 0,05 | 0,04 | 1,15 | 2,48E-01 | 4,57E-01 |
| CIAO1 | 1759,18 | 0,02 | 0,02 | 1,15 | 2,49E-01 | 4,57E-01 |
| ZNF248 | 475,86 | 0,06 | 0,05 | 1,15 | 2,49E-01 | 4,57E-01 |
| BAIAP3 | 248,31 | -0,17 | 0,14 | -1,15 | 2,49E-01 | 4,57E-01 |
| CITED2 | 926,08 | -0,05 | 0,04 | -1,15 | 2,49E-01 | 4,57E-01 |
| LDHB | 727,57 | 0,10 | 0,08 | 1,15 | 2,49E-01 | 4,57E-01 |
| APTX | 517,09 | 0,03 | 0,03 | 1,15 | 2,49E-01 | 4,57E-01 |
| M6PR | 3860,63 | 0,05 | 0,05 | 1,15 | 2,49E-01 | 4,57E-01 |
| SGSH | 378,39 | -0,16 | 0,14 | -1,15 | 2,49E-01 | 4,57E-01 |
| SYMPK | 804,17 | -0,12 | 0,11 | -1,15 | 2,49E-01 | 4,57E-01 |
| GPR157 | 180,26 | -0,10 | 0,09 | -1,15 | 2,49E-01 | 4,57E-01 |
| PGAP1 | 446,69 | 0,09 | 0,08 | 1,15 | 2,49E-01 | 4,57E-01 |
| ZNF850 | 186,05 | 0,07 | 0,06 | 1,15 | 2,49E-01 | 4,57E-01 |
| ARV1 | 176,48 | 0,07 | 0,06 | 1,15 | 2,49E-01 | 4,57E-01 |
| PAPOLG | 1844,78 | 0,05 | 0,04 | 1,15 | 2,49E-01 | 4,57E-01 |
| RAB10 | 3946,46 | 0,06 | 0,05 | 1,15 | 2,49E-01 | 4,57E-01 |
| SYVN1 | 358,77 | -0,10 | 0,08 | -1,15 | 2,49E-01 | 4,57E-01 |
| ZNF559 | 590,40 | 0,07 | 0,06 | 1,15 | 2,49E-01 | 4,57E-01 |
| TENT5C | 28165,20 | -0,18 | 0,15 | -1,15 | 2,49E-01 | 4,57E-01 |
| ME1 | 49,84 | -0,14 | 0,12 | -1,15 | 2,49E-01 | 4,57E-01 |
| DPPA4 | 29,10 | -0,19 | 0,17 | -1,15 | 2,49E-01 | 4,57E-01 |
| PLCL1 | 600,52 | 0,08 | 0,07 | 1,15 | 2,49E-01 | 4,57E-01 |
| NPLOC4 | 2356,35 | -0,06 | 0,05 | -1,15 | 2,50E-01 | 4,57E-01 |
| TMEM131 | 2662,96 | 0,05 | 0,04 | 1,15 | 2,50E-01 | 4,57E-01 |
| LMBR1L | 1381,96 | -0,05 | 0,04 | -1,15 | 2,50E-01 | 4,57E-01 |
| CCDC28B | 120,95 | -0,07 | 0,06 | -1,15 | 2,50E-01 | 4,57E-01 |
| POLR2J3 | 426,50 | -0,13 | 0,11 | -1,15 | 2,50E-01 | 4,57E-01 |
| SF3A1 | 1247,18 | -0,08 | 0,07 | -1,15 | 2,50E-01 | 4,58E-01 |
| TMEM14C | 437,36 | -0,09 | 0,08 | -1,15 | 2,50E-01 | 4,58E-01 |
| HEXB | 1384,48 | 0,05 | 0,04 | 1,15 | 2,50E-01 | 4,58E-01 |
| CHKB-CPT1E | 43,43 | -0,10 | 0,09 | -1,15 | 2,50E-01 | 4,58E-01 |
| CRYGS | 178,12 | 0,06 | 0,05 | 1,15 | 2,50E-01 | 4,58E-01 |
| SLC4A2 | 398,42 | -0,10 | 0,09 | -1,15 | 2,50E-01 | 4,58E-01 |
| SMARCC2 | 1160,14 | -0,07 | 0,06 | -1,15 | 2,50E-01 | 4,58E-01 |
| ZNF143 | 1278,21 | 0,05 | 0,04 | 1,15 | 2,50E-01 | 4,58E-01 |
| PXMP4 | 385,52 | 0,03 | 0,03 | 1,15 | 2,51E-01 | 4,58E-01 |
| MRPL45 | 194,39 | 0,06 | 0,05 | 1,15 | 2,51E-01 | 4,58E-01 |
| BRI3BP | 1097,10 | -0,05 | 0,05 | -1,15 | 2,51E-01 | 4,59E-01 |
| ZNF16 | 181,25 | 0,05 | 0,05 | 1,15 | 2,51E-01 | 4,59E-01 |
| LMTK2 | 1906,65 | -0,08 | 0,07 | -1,15 | 2,51E-01 | 4,59E-01 |
| BIN2 | 5959,72 | -0,04 | 0,04 | -1,15 | 2,51E-01 | 4,59E-01 |
| PLIN5 | 309,98 | -0,13 | 0,11 | -1,15 | 2,51E-01 | 4,59E-01 |
| APH1A | 2089,50 | 0,05 | 0,04 | 1,15 | 2,51E-01 | 4,59E-01 |

| | | | | | | |
|---|---|---|---|---|---|---|
| BCKDK | 513,24 | -0,07 | 0,06 | -1,15 | 2,51E-01 | 4,59E-01 |
| LRRC45 | 308,23 | -0,05 | 0,05 | -1,15 | 2,51E-01 | 4,59E-01 |
| LAT | 67,93 | -0,17 | 0,14 | -1,15 | 2,52E-01 | 4,59E-01 |
| PRELID2 | 89,47 | -0,11 | 0,10 | -1,15 | 2,51E-01 | 4,59E-01 |
| ARID1A | 3096,89 | -0,09 | 0,08 | -1,15 | 2,51E-01 | 4,59E-01 |
| ZNF222 | 105,41 | 0,08 | 0,07 | 1,15 | 2,52E-01 | 4,59E-01 |
| NEXN | 116,34 | -0,11 | 0,10 | -1,15 | 2,51E-01 | 4,59E-01 |
| SPHK2 | 186,46 | -0,10 | 0,09 | -1,15 | 2,52E-01 | 4,59E-01 |
| TANC2 | 1339,40 | -0,07 | 0,06 | -1,15 | 2,52E-01 | 4,59E-01 |
| GIMAP5 | 2243,50 | 0,07 | 0,06 | 1,15 | 2,52E-01 | 4,59E-01 |
| YY2 | 51,08 | 0,07 | 0,06 | 1,15 | 2,52E-01 | 4,59E-01 |
| PSMC3IP | 61,46 | -0,07 | 0,06 | -1,15 | 2,52E-01 | 4,59E-01 |
| PLCG2 | 5258,35 | -0,07 | 0,06 | -1,15 | 2,52E-01 | 4,59E-01 |
| RPUSD2 | 179,94 | 0,05 | 0,05 | 1,15 | 2,52E-01 | 4,59E-01 |
| USP24 | 6734,41 | 0,05 | 0,04 | 1,15 | 2,52E-01 | 4,59E-01 |
| SPTSSB | 55,52 | -0,16 | 0,14 | -1,15 | 2,52E-01 | 4,59E-01 |
| PON2 | 237,57 | 0,06 | 0,05 | 1,14 | 2,52E-01 | 4,60E-01 |
| TMTC1 | 451,50 | 0,22 | 0,19 | 1,14 | 2,52E-01 | 4,60E-01 |
| ABCA10 | 163,39 | -0,11 | 0,09 | -1,14 | 2,53E-01 | 4,60E-01 |
| ZNF264 | 2744,10 | 0,03 | 0,03 | 1,14 | 2,53E-01 | 4,60E-01 |
| TBC1D13 | 252,71 | -0,10 | 0,09 | -1,14 | 2,53E-01 | 4,60E-01 |
| ADGRF3 | 81,36 | -0,12 | 0,10 | -1,14 | 2,53E-01 | 4,60E-01 |
| NDUFB3 | 228,55 | 0,15 | 0,13 | 1,14 | 2,53E-01 | 4,60E-01 |
| TAF5L | 630,58 | 0,03 | 0,03 | 1,14 | 2,53E-01 | 4,60E-01 |
| PLAGL2 | 587,07 | -0,08 | 0,07 | -1,14 | 2,53E-01 | 4,60E-01 |
| ZC3H3 | 402,77 | -0,11 | 0,09 | -1,14 | 2,53E-01 | 4,61E-01 |
| MCFD2 | 919,59 | 0,03 | 0,02 | 1,14 | 2,53E-01 | 4,61E-01 |
| P4HTM | 616,37 | -0,05 | 0,05 | -1,14 | 2,53E-01 | 4,61E-01 |
| MGRN1 | 1228,59 | -0,14 | 0,12 | -1,14 | 2,53E-01 | 4,61E-01 |
| RPA3 | 242,63 | 0,09 | 0,08 | 1,14 | 2,53E-01 | 4,61E-01 |
| DHRS4 | 312,68 | 0,05 | 0,05 | 1,14 | 2,53E-01 | 4,61E-01 |
| PTP4A1 | 71,93 | -0,07 | 0,07 | -1,14 | 2,54E-01 | 4,61E-01 |
| SGF29 | 253,02 | -0,05 | 0,04 | -1,14 | 2,54E-01 | 4,61E-01 |
| MRPL52 | 363,28 | 0,04 | 0,03 | 1,14 | 2,54E-01 | 4,62E-01 |
| PCP2 | 30,41 | -0,11 | 0,10 | -1,14 | 2,54E-01 | 4,62E-01 |
| PPARGC1A | 43,68 | -0,30 | 0,27 | -1,14 | 2,54E-01 | 4,62E-01 |
| CALHM2 | 405,13 | -0,08 | 0,07 | -1,14 | 2,54E-01 | 4,62E-01 |
| AXIN2 | 233,32 | -0,10 | 0,09 | -1,14 | 2,54E-01 | 4,62E-01 |
| SIGLEC9 | 1447,69 | -0,09 | 0,08 | -1,14 | 2,54E-01 | 4,62E-01 |
| GRAP2 | 2638,93 | -0,06 | 0,06 | -1,14 | 2,54E-01 | 4,62E-01 |
| PCTP | 1278,91 | -0,10 | 0,08 | -1,14 | 2,55E-01 | 4,62E-01 |
| ZC3H7A | 1679,03 | 0,04 | 0,03 | 1,14 | 2,55E-01 | 4,62E-01 |
| UBAC2 | 1717,14 | -0,04 | 0,03 | -1,14 | 2,55E-01 | 4,62E-01 |
| PUM2 | 7278,93 | 0,05 | 0,04 | 1,14 | 2,55E-01 | 4,62E-01 |
| RPLP0 | 12732,36 | 0,09 | 0,08 | 1,14 | 2,55E-01 | 4,62E-01 |
| RBBP8 | 294,12 | 0,10 | 0,09 | 1,14 | 2,55E-01 | 4,62E-01 |

| | | | | | | |
|---|---|---|---|---|---|---|
| CENPJ | 273,56 | 0,07 | 0,06 | 1,14 | 2,55E-01 | 4,63E-01 |
| FGF9 | 50,90 | -0,16 | 0,14 | -1,14 | 2,55E-01 | 4,63E-01 |
| RAP1B | 6878,80 | 0,06 | 0,05 | 1,14 | 2,55E-01 | 4,63E-01 |
| SCAP | 1183,62 | -0,10 | 0,09 | -1,14 | 2,55E-01 | 4,63E-01 |
| PAQR6 | 67,61 | -0,15 | 0,13 | -1,14 | 2,55E-01 | 4,63E-01 |
| ZSCAN23 | 66,20 | 0,15 | 0,14 | 1,14 | 2,55E-01 | 4,63E-01 |
| RASSF3 | 5596,57 | -0,06 | 0,06 | -1,14 | 2,56E-01 | 4,64E-01 |
| HLA-DQA1 | 1860,55 | 0,10 | 0,09 | 1,14 | 2,56E-01 | 4,64E-01 |
| WDR1 | 6815,81 | -0,06 | 0,05 | -1,14 | 2,56E-01 | 4,64E-01 |
| CIAO3 | 290,02 | -0,09 | 0,08 | -1,14 | 2,56E-01 | 4,64E-01 |
| ARSG | 1651,98 | -0,07 | 0,06 | -1,14 | 2,56E-01 | 4,64E-01 |
| OSGEP | 1003,52 | 0,04 | 0,03 | 1,14 | 2,56E-01 | 4,64E-01 |
| NUFIP2 | 9171,72 | 0,06 | 0,05 | 1,14 | 2,56E-01 | 4,64E-01 |
| IDH1 | 1189,46 | 0,07 | 0,06 | 1,14 | 2,56E-01 | 4,64E-01 |
| DDX55 | 600,95 | 0,04 | 0,03 | 1,13 | 2,57E-01 | 4,64E-01 |
| HAUS1 | 252,51 | 0,08 | 0,07 | 1,13 | 2,57E-01 | 4,64E-01 |
| TNK2 | 1941,85 | -0,13 | 0,11 | -1,13 | 2,57E-01 | 4,64E-01 |
| ATP2B4 | 3840,09 | -0,07 | 0,06 | -1,13 | 2,57E-01 | 4,64E-01 |
| GCNA | 239,08 | -0,05 | 0,04 | -1,13 | 2,57E-01 | 4,64E-01 |
| TECR | 241,78 | -0,09 | 0,08 | -1,13 | 2,57E-01 | 4,64E-01 |
| AGBL3 | 213,34 | 0,09 | 0,08 | 1,13 | 2,57E-01 | 4,64E-01 |
| PAK1 | 3121,42 | -0,07 | 0,06 | -1,13 | 2,57E-01 | 4,64E-01 |
| TLK2 | 2279,09 | 0,03 | 0,03 | 1,13 | 2,57E-01 | 4,65E-01 |
| NDE1 | 2466,91 | -0,09 | 0,08 | -1,13 | 2,57E-01 | 4,65E-01 |
| TMEM229B | 508,35 | -0,08 | 0,07 | -1,13 | 2,57E-01 | 4,65E-01 |
| KCNC4 | 255,47 | -0,16 | 0,15 | -1,13 | 2,57E-01 | 4,65E-01 |
| XNDC1N | 528,52 | 0,06 | 0,05 | 1,13 | 2,57E-01 | 4,65E-01 |
| KLRG1 | 1628,68 | 0,12 | 0,11 | 1,13 | 2,57E-01 | 4,65E-01 |
| DONSON | 155,16 | -0,04 | 0,04 | -1,13 | 2,57E-01 | 4,65E-01 |
| PLXNC1 | 16001,66 | -0,08 | 0,07 | -1,13 | 2,57E-01 | 4,65E-01 |
| HCFC1 | 653,48 | -0,13 | 0,11 | -1,13 | 2,57E-01 | 4,65E-01 |
| BRD4 | 1675,02 | -0,13 | 0,11 | -1,13 | 2,57E-01 | 4,65E-01 |
| BAG1 | 6757,30 | -0,13 | 0,12 | -1,13 | 2,57E-01 | 4,65E-01 |
| ZNF569 | 261,48 | 0,08 | 0,07 | 1,13 | 2,58E-01 | 4,65E-01 |
| EEFSEC | 185,68 | -0,07 | 0,07 | -1,13 | 2,58E-01 | 4,65E-01 |
| TAGAP | 4982,10 | -0,06 | 0,05 | -1,13 | 2,58E-01 | 4,65E-01 |
| SDHAF2 | 562,72 | -0,05 | 0,04 | -1,13 | 2,58E-01 | 4,65E-01 |
| VHL | 2848,16 | 0,03 | 0,03 | 1,13 | 2,58E-01 | 4,65E-01 |
| SMARCD1 | 592,50 | -0,05 | 0,04 | -1,13 | 2,58E-01 | 4,66E-01 |
| KCNQ5 | 203,09 | 0,09 | 0,08 | 1,13 | 2,58E-01 | 4,66E-01 |
| CDC42BPA | 383,74 | -0,13 | 0,12 | -1,13 | 2,58E-01 | 4,66E-01 |
| USP36 | 717,92 | -0,10 | 0,08 | -1,13 | 2,58E-01 | 4,66E-01 |
| BAZ1A | 5851,53 | 0,07 | 0,06 | 1,13 | 2,59E-01 | 4,66E-01 |
| LITAF | 12970,15 | -0,08 | 0,07 | -1,13 | 2,59E-01 | 4,66E-01 |
| HTRA4 | 60,26 | -0,10 | 0,09 | -1,13 | 2,59E-01 | 4,66E-01 |
| SERPINF2 | 46,92 | -0,14 | 0,12 | -1,13 | 2,59E-01 | 4,66E-01 |

| | | | | | | |
|---|---|---|---|---|---|---|
| CREB1 | 6243,20 | 0,07 | 0,06 | 1,13 | 2,59E-01 | 4,67E-01 |
| ORC2 | 1195,18 | 0,04 | 0,03 | 1,13 | 2,59E-01 | 4,67E-01 |
| BDH1 | 303,54 | -0,09 | 0,08 | -1,13 | 2,59E-01 | 4,67E-01 |
| LMO4 | 888,36 | 0,04 | 0,04 | 1,13 | 2,59E-01 | 4,67E-01 |
| CRISPLD2 | 2686,58 | -0,13 | 0,11 | -1,13 | 2,59E-01 | 4,67E-01 |
| THEMIS2 | 8882,26 | -0,08 | 0,07 | -1,13 | 2,59E-01 | 4,67E-01 |
| EEF2KMT | 121,51 | -0,06 | 0,05 | -1,13 | 2,59E-01 | 4,67E-01 |
| IFIH1 | 1582,68 | 0,10 | 0,09 | 1,13 | 2,60E-01 | 4,67E-01 |
| SH2D3C | 1466,29 | -0,10 | 0,09 | -1,13 | 2,60E-01 | 4,67E-01 |
| SRP14 | 1494,92 | 0,11 | 0,10 | 1,13 | 2,60E-01 | 4,67E-01 |
| TRIM59 | 402,75 | 0,07 | 0,07 | 1,13 | 2,60E-01 | 4,67E-01 |
| ACAP3 | 801,39 | -0,10 | 0,09 | -1,13 | 2,60E-01 | 4,67E-01 |
| NDC1 | 664,07 | 0,05 | 0,04 | 1,13 | 2,60E-01 | 4,67E-01 |
| TMEM69 | 353,21 | 0,05 | 0,04 | 1,13 | 2,60E-01 | 4,67E-01 |
| ZNF384 | 838,74 | -0,05 | 0,04 | -1,13 | 2,60E-01 | 4,68E-01 |
| RHNO1 | 270,07 | 0,05 | 0,05 | 1,13 | 2,60E-01 | 4,68E-01 |
| ACTR3 | 16418,44 | 0,07 | 0,06 | 1,13 | 2,60E-01 | 4,68E-01 |
| C1orf43 | 1320,26 | -0,04 | 0,04 | -1,13 | 2,60E-01 | 4,68E-01 |
| PTPMT1 | 176,53 | 0,04 | 0,04 | 1,12 | 2,61E-01 | 4,68E-01 |
| ENSG000002 | 45,22 | -0,05 | 0,05 | -1,12 | 2,61E-01 | 4,68E-01 |
| MTSS1 | 1765,89 | 0,07 | 0,06 | 1,12 | 2,61E-01 | 4,68E-01 |
| RGL1 | 132,89 | -0,09 | 0,08 | -1,12 | 2,61E-01 | 4,68E-01 |
| PPP3CC | 1638,90 | 0,06 | 0,06 | 1,12 | 2,61E-01 | 4,68E-01 |
| CASP9 | 408,60 | -0,06 | 0,05 | -1,12 | 2,61E-01 | 4,68E-01 |
| CLDN7 | 30,54 | -0,15 | 0,14 | -1,12 | 2,61E-01 | 4,69E-01 |
| ZMIZ2 | 705,09 | -0,12 | 0,11 | -1,12 | 2,61E-01 | 4,69E-01 |
| ITM2C | 196,18 | -0,10 | 0,09 | -1,12 | 2,61E-01 | 4,69E-01 |
| MMADHC | 1630,41 | 0,08 | 0,07 | 1,12 | 2,62E-01 | 4,69E-01 |
| NLK | 930,53 | 0,04 | 0,03 | 1,12 | 2,62E-01 | 4,70E-01 |
| DUSP14 | 71,91 | -0,09 | 0,08 | -1,12 | 2,62E-01 | 4,70E-01 |
| KDSR | 506,55 | -0,05 | 0,04 | -1,12 | 2,62E-01 | 4,70E-01 |
| RSBN1L | 3548,40 | 0,06 | 0,05 | 1,12 | 2,62E-01 | 4,70E-01 |
| ELK3 | 732,58 | 0,05 | 0,04 | 1,12 | 2,62E-01 | 4,70E-01 |
| GTF3C2 | 784,67 | -0,07 | 0,06 | -1,12 | 2,62E-01 | 4,70E-01 |
| ACSM3 | 367,53 | -0,11 | 0,09 | -1,12 | 2,62E-01 | 4,70E-01 |
| KCNAB2 | 2920,91 | -0,09 | 0,08 | -1,12 | 2,62E-01 | 4,70E-01 |
| SULT1B1 | 3893,42 | -0,10 | 0,09 | -1,12 | 2,62E-01 | 4,70E-01 |
| CLTA | 1053,84 | 0,04 | 0,04 | 1,12 | 2,62E-01 | 4,70E-01 |
| ZBED6 | 44,68 | -0,10 | 0,09 | -1,12 | 2,63E-01 | 4,70E-01 |
| ITPA | 561,61 | 0,04 | 0,03 | 1,12 | 2,63E-01 | 4,70E-01 |
| PYGO2 | 377,98 | -0,09 | 0,08 | -1,12 | 2,63E-01 | 4,70E-01 |
| ID3 | 124,08 | 0,11 | 0,10 | 1,12 | 2,63E-01 | 4,70E-01 |
| UBA1 | 1853,00 | -0,10 | 0,09 | -1,12 | 2,63E-01 | 4,70E-01 |
| MFSD11 | 756,83 | -0,03 | 0,03 | -1,12 | 2,63E-01 | 4,70E-01 |
| ZNF853 | 66,49 | -0,13 | 0,12 | -1,12 | 2,63E-01 | 4,70E-01 |
| PLAC8 | 3816,65 | 0,05 | 0,05 | 1,12 | 2,63E-01 | 4,70E-01 |

| | | | | | | |
|---|---|---|---|---|---|---|
| PYCR2 | 432,58 | -0,07 | 0,07 | -1,12 | 2,63E-01 | 4,71E-01 |
| TSPOAP1 | 452,98 | -0,16 | 0,14 | -1,12 | 2,63E-01 | 4,71E-01 |
| MKRN1 | 29826,95 | -0,13 | 0,12 | -1,12 | 2,63E-01 | 4,71E-01 |
| PEAK1 | 952,86 | 0,05 | 0,05 | 1,12 | 2,63E-01 | 4,71E-01 |
| LCOR | 6117,81 | 0,04 | 0,04 | 1,12 | 2,63E-01 | 4,71E-01 |
| OR2W3 | 481,83 | -0,19 | 0,17 | -1,12 | 2,63E-01 | 4,71E-01 |
| RPS19 | 10230,30 | 0,08 | 0,07 | 1,12 | 2,63E-01 | 4,71E-01 |
| NEURL2 | 37,95 | -0,09 | 0,08 | -1,12 | 2,63E-01 | 4,71E-01 |
| MVD | 370,86 | -0,06 | 0,05 | -1,12 | 2,64E-01 | 4,71E-01 |
| GREM2 | 36,23 | -0,18 | 0,16 | -1,12 | 2,64E-01 | 4,71E-01 |
| DGCR2 | 2723,81 | -0,09 | 0,08 | -1,12 | 2,64E-01 | 4,72E-01 |
| PPP6C | 1709,07 | 0,05 | 0,04 | 1,12 | 2,64E-01 | 4,72E-01 |
| SLC25A5 | 1526,42 | 0,05 | 0,05 | 1,12 | 2,64E-01 | 4,72E-01 |
| VEGFA | 393,71 | -0,09 | 0,08 | -1,12 | 2,64E-01 | 4,72E-01 |
| RASSF6 | 44,17 | -0,17 | 0,15 | -1,12 | 2,64E-01 | 4,72E-01 |
| CD28 | 2789,32 | 0,08 | 0,07 | 1,12 | 2,64E-01 | 4,72E-01 |
| SLC26A6 | 258,12 | -0,08 | 0,07 | -1,12 | 2,64E-01 | 4,72E-01 |
| ZNF763 | 165,04 | 0,06 | 0,06 | 1,12 | 2,64E-01 | 4,72E-01 |
| KIAA2013 | 1089,56 | -0,06 | 0,05 | -1,12 | 2,65E-01 | 4,72E-01 |
| PTRHD1 | 238,50 | 0,09 | 0,09 | 1,12 | 2,65E-01 | 4,72E-01 |
| CDC42 | 749,33 | -0,04 | 0,03 | -1,12 | 2,65E-01 | 4,72E-01 |
| SLC27A4 | 215,39 | -0,07 | 0,06 | -1,12 | 2,65E-01 | 4,72E-01 |
| PRDM10 | 702,29 | -0,03 | 0,03 | -1,12 | 2,65E-01 | 4,72E-01 |
| VAMP1 | 966,08 | -0,06 | 0,05 | -1,12 | 2,65E-01 | 4,72E-01 |
| TRPM6 | 1304,38 | -0,10 | 0,09 | -1,12 | 2,65E-01 | 4,72E-01 |
| FAM219A | 224,64 | -0,10 | 0,09 | -1,11 | 2,65E-01 | 4,72E-01 |
| TNFSF14 | 1994,37 | -0,07 | 0,06 | -1,11 | 2,65E-01 | 4,72E-01 |
| SLC52A2 | 190,47 | -0,06 | 0,06 | -1,11 | 2,65E-01 | 4,72E-01 |
| TMEM273 | 1205,58 | -0,09 | 0,08 | -1,11 | 2,65E-01 | 4,72E-01 |
| MFGE8 | 178,93 | -0,13 | 0,12 | -1,11 | 2,65E-01 | 4,72E-01 |
| GPR35 | 179,49 | -0,10 | 0,09 | -1,11 | 2,65E-01 | 4,73E-01 |
| CCDC159 | 267,30 | -0,07 | 0,06 | -1,11 | 2,65E-01 | 4,73E-01 |
| GAB2 | 2395,57 | -0,10 | 0,09 | -1,11 | 2,65E-01 | 4,73E-01 |
| HLA-DPB1 | 4602,28 | 0,07 | 0,06 | 1,11 | 2,65E-01 | 4,73E-01 |
| TBXAS1 | 6892,00 | -0,07 | 0,07 | -1,11 | 2,65E-01 | 4,73E-01 |
| JAKMIP2 | 455,59 | 0,12 | 0,11 | 1,11 | 2,65E-01 | 4,73E-01 |
| CD72 | 284,73 | 0,11 | 0,10 | 1,11 | 2,66E-01 | 4,73E-01 |
| DHRS13 | 818,48 | -0,08 | 0,07 | -1,11 | 2,66E-01 | 4,73E-01 |
| ASAH1 | 10084,36 | 0,08 | 0,07 | 1,11 | 2,66E-01 | 4,73E-01 |
| P2RX5 | 159,46 | -0,12 | 0,11 | -1,11 | 2,66E-01 | 4,73E-01 |
| FAM217B | 1190,02 | 0,05 | 0,05 | 1,11 | 2,66E-01 | 4,73E-01 |
| CAPZB | 1161,24 | -0,10 | 0,09 | -1,11 | 2,66E-01 | 4,73E-01 |
| MKRN2 | 708,73 | 0,03 | 0,02 | 1,11 | 2,66E-01 | 4,73E-01 |
| UFSP1 | 111,29 | 0,07 | 0,06 | 1,11 | 2,66E-01 | 4,73E-01 |
| FAM177A1 | 747,21 | -0,04 | 0,03 | -1,11 | 2,66E-01 | 4,74E-01 |
| GTF3C4 | 624,94 | 0,03 | 0,02 | 1,11 | 2,67E-01 | 4,74E-01 |

| | | | | | | |
|---|---|---|---|---|---|---|
| C2CD2 | 343,92 | -0,07 | 0,06 | -1,11 | 2,67E-01 | 4,74E-01 |
| CTSD | 975,87 | -0,08 | 0,07 | -1,11 | 2,67E-01 | 4,74E-01 |
| MIDN | 1295,71 | -0,11 | 0,10 | -1,11 | 2,67E-01 | 4,74E-01 |
| THBS3 | 477,94 | -0,07 | 0,06 | -1,11 | 2,67E-01 | 4,74E-01 |
| GHRL | 150,20 | -0,11 | 0,09 | -1,11 | 2,67E-01 | 4,75E-01 |
| RNF125 | 1663,33 | 0,05 | 0,04 | 1,11 | 2,67E-01 | 4,75E-01 |
| DRAM1 | 451,97 | -0,06 | 0,06 | -1,11 | 2,67E-01 | 4,75E-01 |
| PIGT | 951,82 | -0,05 | 0,05 | -1,11 | 2,67E-01 | 4,75E-01 |
| DNAJA1 | 2215,61 | 0,10 | 0,09 | 1,11 | 2,67E-01 | 4,75E-01 |
| SANBR | 343,71 | -0,05 | 0,04 | -1,11 | 2,67E-01 | 4,75E-01 |
| CACNA2D4 | 232,79 | -0,09 | 0,08 | -1,11 | 2,68E-01 | 4,75E-01 |
| AGTPBP1 | 6772,37 | 0,07 | 0,07 | 1,11 | 2,68E-01 | 4,75E-01 |
| PRR13 | 610,32 | -0,08 | 0,07 | -1,11 | 2,68E-01 | 4,75E-01 |
| NCOA4 | 38278,42 | -0,10 | 0,09 | -1,11 | 2,68E-01 | 4,75E-01 |
| NID1 | 147,30 | -0,14 | 0,13 | -1,11 | 2,68E-01 | 4,75E-01 |
| ZNF132 | 296,28 | 0,09 | 0,08 | 1,11 | 2,68E-01 | 4,75E-01 |
| SLC2A11 | 187,17 | -0,09 | 0,08 | -1,11 | 2,68E-01 | 4,75E-01 |
| PALS1 | 445,10 | 0,06 | 0,05 | 1,11 | 2,68E-01 | 4,75E-01 |
| PAIP2 | 2300,00 | 0,08 | 0,07 | 1,11 | 2,68E-01 | 4,75E-01 |
| RGS3 | 1413,62 | -0,07 | 0,06 | -1,11 | 2,68E-01 | 4,75E-01 |
| PLD2 | 199,87 | -0,11 | 0,10 | -1,11 | 2,68E-01 | 4,75E-01 |
| RPS4X | 10321,18 | 0,08 | 0,07 | 1,11 | 2,68E-01 | 4,75E-01 |
| DAPK1 | 1663,12 | -0,09 | 0,08 | -1,11 | 2,68E-01 | 4,75E-01 |
| AFF3 | 1047,49 | 0,12 | 0,11 | 1,11 | 2,68E-01 | 4,75E-01 |
| SLC10A7 | 512,44 | 0,05 | 0,05 | 1,11 | 2,68E-01 | 4,76E-01 |
| BLOC1S3 | 205,56 | -0,09 | 0,08 | -1,11 | 2,68E-01 | 4,76E-01 |
| GPBAR1 | 189,60 | -0,08 | 0,07 | -1,11 | 2,68E-01 | 4,76E-01 |
| CHSY1 | 2327,91 | 0,07 | 0,06 | 1,11 | 2,69E-01 | 4,76E-01 |
| ARMC8 | 2570,15 | 0,04 | 0,04 | 1,11 | 2,69E-01 | 4,76E-01 |
| H2BC17 | 198,30 | -0,10 | 0,09 | -1,11 | 2,69E-01 | 4,76E-01 |
| TMEM33 | 4335,44 | 0,06 | 0,06 | 1,11 | 2,69E-01 | 4,76E-01 |
| CHAF1A | 176,10 | -0,08 | 0,07 | -1,11 | 2,69E-01 | 4,76E-01 |
| MON2 | 6238,32 | 0,06 | 0,05 | 1,10 | 2,69E-01 | 4,77E-01 |
| UGDH | 319,28 | 0,06 | 0,05 | 1,10 | 2,69E-01 | 4,77E-01 |
| DNMT3A | 1600,78 | -0,07 | 0,06 | -1,10 | 2,69E-01 | 4,77E-01 |
| WDR76 | 306,95 | 0,06 | 0,06 | 1,10 | 2,69E-01 | 4,77E-01 |
| HNRNPLL | 1988,03 | 0,07 | 0,07 | 1,10 | 2,70E-01 | 4,77E-01 |
| NUDT1 | 286,96 | -0,07 | 0,06 | -1,10 | 2,70E-01 | 4,78E-01 |
| AKAP5 | 145,20 | 0,14 | 0,13 | 1,10 | 2,70E-01 | 4,78E-01 |
| SGSM2 | 813,14 | -0,10 | 0,09 | -1,10 | 2,70E-01 | 4,78E-01 |
| MESD | 1045,75 | 0,04 | 0,03 | 1,10 | 2,70E-01 | 4,78E-01 |
| FAM98A | 503,65 | 0,05 | 0,05 | 1,10 | 2,70E-01 | 4,78E-01 |
| RAVER1 | 219,72 | -0,12 | 0,11 | -1,10 | 2,70E-01 | 4,78E-01 |
| KXD1 | 724,81 | -0,06 | 0,05 | -1,10 | 2,70E-01 | 4,78E-01 |
| SCRN1 | 878,35 | 0,07 | 0,06 | 1,10 | 2,70E-01 | 4,78E-01 |
| NAB1 | 1926,08 | 0,07 | 0,06 | 1,10 | 2,71E-01 | 4,78E-01 |

| | | | | | | |
|---|---|---|---|---|---|---|
| UBXN2B | 2887,65 | -0,07 | 0,06 | -1,10 | 2,71E-01 | 4,78E-01 |
| CPNE5 | 200,99 | -0,11 | 0,10 | -1,10 | 2,71E-01 | 4,78E-01 |
| RFESD | 169,23 | 0,09 | 0,08 | 1,10 | 2,71E-01 | 4,78E-01 |
| IL7 | 102,89 | 0,12 | 0,11 | 1,10 | 2,71E-01 | 4,78E-01 |
| RAB18 | 3277,97 | 0,08 | 0,07 | 1,10 | 2,71E-01 | 4,78E-01 |
| SF3A3 | 1571,72 | 0,04 | 0,04 | 1,10 | 2,71E-01 | 4,79E-01 |
| LEPROT | 1283,37 | -0,06 | 0,05 | -1,10 | 2,72E-01 | 4,79E-01 |
| ALDH1B1 | 122,05 | -0,07 | 0,06 | -1,10 | 2,71E-01 | 4,79E-01 |
| HDGF | 5740,84 | -0,12 | 0,11 | -1,10 | 2,72E-01 | 4,79E-01 |
| BROX | 3092,92 | 0,06 | 0,06 | 1,10 | 2,71E-01 | 4,79E-01 |
| GNAI2 | 4220,08 | -0,09 | 0,08 | -1,10 | 2,71E-01 | 4,79E-01 |
| CAPS2 | 237,17 | 0,07 | 0,07 | 1,10 | 2,72E-01 | 4,79E-01 |
| SLC26A11 | 286,31 | -0,09 | 0,08 | -1,10 | 2,72E-01 | 4,79E-01 |
| RNF111 | 2292,40 | 0,05 | 0,04 | 1,10 | 2,71E-01 | 4,79E-01 |
| ERI2 | 185,56 | 0,07 | 0,07 | 1,10 | 2,71E-01 | 4,79E-01 |
| ACAP1 | 2576,54 | -0,08 | 0,07 | -1,10 | 2,72E-01 | 4,79E-01 |
| LYVE1 | 55,62 | -0,11 | 0,10 | -1,10 | 2,72E-01 | 4,79E-01 |
| TRNAU1AP | 633,30 | 0,03 | 0,03 | 1,10 | 2,72E-01 | 4,79E-01 |
| CDCA4 | 259,50 | -0,05 | 0,04 | -1,10 | 2,72E-01 | 4,79E-01 |
| NR2C2AP | 218,66 | 0,05 | 0,05 | 1,10 | 2,72E-01 | 4,79E-01 |
| VAT1 | 158,29 | -0,09 | 0,09 | -1,10 | 2,72E-01 | 4,79E-01 |
| ITPR3 | 965,82 | -0,11 | 0,10 | -1,10 | 2,72E-01 | 4,79E-01 |
| UQCC3 | 178,15 | 0,06 | 0,05 | 1,10 | 2,72E-01 | 4,79E-01 |
| ZNF287 | 234,40 | 0,07 | 0,06 | 1,10 | 2,72E-01 | 4,79E-01 |
| B4GALT7 | 285,08 | -0,06 | 0,05 | -1,10 | 2,72E-01 | 4,79E-01 |
| ATP6AP1 | 738,09 | -0,07 | 0,06 | -1,10 | 2,73E-01 | 4,80E-01 |
| SLC29A3 | 117,77 | -0,09 | 0,08 | -1,10 | 2,73E-01 | 4,80E-01 |
| SIPA1 | 2155,49 | -0,12 | 0,11 | -1,10 | 2,73E-01 | 4,80E-01 |
| ZDHHC15 | 25,42 | -0,16 | 0,15 | -1,10 | 2,73E-01 | 4,80E-01 |
| LMNA | 318,93 | -0,12 | 0,11 | -1,10 | 2,73E-01 | 4,80E-01 |
| GRIP1 | 73,69 | -0,18 | 0,16 | -1,10 | 2,73E-01 | 4,80E-01 |
| SYTL4 | 104,26 | -0,14 | 0,13 | -1,10 | 2,73E-01 | 4,80E-01 |
| PICK1 | 280,76 | -0,06 | 0,05 | -1,10 | 2,73E-01 | 4,80E-01 |
| AMOT | 116,28 | -0,08 | 0,07 | -1,10 | 2,73E-01 | 4,80E-01 |
| MFNG | 954,84 | -0,08 | 0,07 | -1,10 | 2,73E-01 | 4,80E-01 |
| OAF | 134,50 | -0,12 | 0,11 | -1,10 | 2,73E-01 | 4,80E-01 |
| QTRT1 | 214,77 | -0,06 | 0,06 | -1,10 | 2,73E-01 | 4,80E-01 |
| RAB43 | 376,67 | -0,06 | 0,06 | -1,10 | 2,73E-01 | 4,80E-01 |
| MKNK1 | 2419,43 | -0,07 | 0,06 | -1,09 | 2,74E-01 | 4,81E-01 |
| LRP8 | 333,52 | -0,07 | 0,06 | -1,09 | 2,74E-01 | 4,81E-01 |
| CLEC4C | 210,17 | 0,18 | 0,17 | 1,09 | 2,74E-01 | 4,81E-01 |
| TNFRSF13B | 78,31 | -0,16 | 0,14 | -1,09 | 2,74E-01 | 4,81E-01 |
| TANK | 5698,45 | 0,07 | 0,06 | 1,09 | 2,74E-01 | 4,81E-01 |
| EP400 | 1476,69 | -0,12 | 0,11 | -1,09 | 2,74E-01 | 4,82E-01 |
| DNAAF2 | 190,15 | 0,07 | 0,07 | 1,09 | 2,75E-01 | 4,82E-01 |
| BBIP1 | 1132,16 | 0,04 | 0,04 | 1,09 | 2,74E-01 | 4,82E-01 |

| | | | | | | |
|---|---|---|---|---|---|---|
| P2RX1 | 730,07 | -0,11 | 0,10 | -1,09 | 2,74E-01 | 4,82E-01 |
| FASTKD5 | 689,94 | 0,04 | 0,04 | 1,09 | 2,75E-01 | 4,82E-01 |
| SRGAP2 | 1050,60 | -0,08 | 0,07 | -1,09 | 2,75E-01 | 4,82E-01 |
| GCSH | 23,41 | -0,13 | 0,12 | -1,09 | 2,75E-01 | 4,82E-01 |
| PTPRJ | 9873,43 | -0,07 | 0,07 | -1,09 | 2,75E-01 | 4,82E-01 |
| USF2 | 807,77 | -0,10 | 0,09 | -1,09 | 2,75E-01 | 4,82E-01 |
| KYNU | 1736,15 | 0,08 | 0,07 | 1,09 | 2,75E-01 | 4,82E-01 |
| TGIF1 | 732,19 | 0,05 | 0,04 | 1,09 | 2,75E-01 | 4,82E-01 |
| CYB5RL | 143,83 | -0,10 | 0,09 | -1,09 | 2,75E-01 | 4,82E-01 |
| PTGDS | 88,50 | -0,14 | 0,13 | -1,09 | 2,75E-01 | 4,82E-01 |
| FAM234B | 208,11 | -0,06 | 0,05 | -1,09 | 2,75E-01 | 4,82E-01 |
| KLF5 | 98,78 | -0,09 | 0,08 | -1,09 | 2,75E-01 | 4,82E-01 |
| RAB11FIP4 | 3872,51 | -0,06 | 0,06 | -1,09 | 2,75E-01 | 4,82E-01 |
| HMGN5 | 34,09 | -0,11 | 0,10 | -1,09 | 2,75E-01 | 4,82E-01 |
| SREBF2 | 631,98 | -0,12 | 0,11 | -1,09 | 2,75E-01 | 4,82E-01 |
| SPRED1 | 358,96 | 0,11 | 0,10 | 1,09 | 2,75E-01 | 4,82E-01 |
| NME6 | 1012,50 | 0,03 | 0,03 | 1,09 | 2,75E-01 | 4,82E-01 |
| XPNPEP1 | 1630,81 | 0,03 | 0,03 | 1,09 | 2,75E-01 | 4,82E-01 |
| CHRNE | 76,22 | -0,14 | 0,13 | -1,09 | 2,76E-01 | 4,82E-01 |
| NOL4L | 1302,30 | -0,08 | 0,07 | -1,09 | 2,76E-01 | 4,82E-01 |
| ADTRP | 407,14 | 0,14 | 0,13 | 1,09 | 2,76E-01 | 4,82E-01 |
| ARG1 | 267,95 | -0,16 | 0,15 | -1,09 | 2,76E-01 | 4,83E-01 |
| SKA2 | 256,49 | 0,08 | 0,07 | 1,09 | 2,76E-01 | 4,83E-01 |
| RMDN3 | 492,13 | -0,04 | 0,04 | -1,09 | 2,76E-01 | 4,83E-01 |
| COA1 | 2388,67 | 0,06 | 0,05 | 1,09 | 2,77E-01 | 4,84E-01 |
| SPIB | 164,45 | 0,11 | 0,10 | 1,09 | 2,77E-01 | 4,84E-01 |
| PDLIM2 | 1084,67 | -0,09 | 0,09 | -1,09 | 2,77E-01 | 4,84E-01 |
| GRAP | 736,88 | 0,07 | 0,06 | 1,09 | 2,77E-01 | 4,84E-01 |
| MLLT10 | 1705,94 | 0,04 | 0,04 | 1,09 | 2,77E-01 | 4,84E-01 |
| ARAP1 | 7304,68 | -0,12 | 0,11 | -1,09 | 2,77E-01 | 4,84E-01 |
| UBE2W | 3211,43 | 0,09 | 0,08 | 1,09 | 2,77E-01 | 4,84E-01 |
| DNHD1 | 945,81 | -0,10 | 0,09 | -1,09 | 2,77E-01 | 4,84E-01 |
| DENND10 | 1636,75 | -0,05 | 0,04 | -1,09 | 2,77E-01 | 4,84E-01 |
| ABCA1 | 4184,03 | -0,11 | 0,10 | -1,09 | 2,77E-01 | 4,84E-01 |
| SYNJ2 | 561,66 | -0,08 | 0,08 | -1,09 | 2,77E-01 | 4,84E-01 |
| SIL1 | 550,79 | 0,04 | 0,03 | 1,09 | 2,78E-01 | 4,85E-01 |
| CTSH | 1457,81 | 0,06 | 0,06 | 1,09 | 2,78E-01 | 4,85E-01 |
| ZFYVE16 | 5252,69 | 0,08 | 0,07 | 1,09 | 2,78E-01 | 4,85E-01 |
| WDR47 | 738,87 | 0,04 | 0,04 | 1,09 | 2,78E-01 | 4,85E-01 |
| TTI2 | 433,15 | 0,03 | 0,03 | 1,09 | 2,78E-01 | 4,85E-01 |
| WDFY4 | 3779,11 | -0,09 | 0,09 | -1,09 | 2,78E-01 | 4,85E-01 |
| FLNB | 1319,08 | -0,08 | 0,08 | -1,09 | 2,78E-01 | 4,85E-01 |
| KCNMB1 | 127,59 | -0,11 | 0,10 | -1,09 | 2,78E-01 | 4,85E-01 |
| VPS33B | 99,31 | -0,05 | 0,04 | -1,08 | 2,78E-01 | 4,85E-01 |
| CNDP2 | 2106,81 | 0,05 | 0,04 | 1,08 | 2,78E-01 | 4,85E-01 |
| VCPIP1 | 6995,38 | 0,08 | 0,07 | 1,08 | 2,78E-01 | 4,85E-01 |

| | | | | | | |
|---|---|---|---|---|---|---|
| ZNF490 | 235,53 | 0,03 | 0,03 | 1,08 | 2,78E-01 | 4,85E-01 |
| TGOLN2 | 7753,38 | -0,06 | 0,06 | -1,08 | 2,78E-01 | 4,85E-01 |
| NUP58 | 3639,95 | 0,06 | 0,06 | 1,08 | 2,79E-01 | 4,85E-01 |
| AP5S1 | 223,45 | -0,06 | 0,06 | -1,08 | 2,79E-01 | 4,85E-01 |
| BAG2 | 263,55 | 0,08 | 0,08 | 1,08 | 2,79E-01 | 4,85E-01 |
| TGFB1 | 875,63 | -0,16 | 0,15 | -1,08 | 2,79E-01 | 4,85E-01 |
| INTS7 | 693,94 | 0,05 | 0,05 | 1,08 | 2,79E-01 | 4,85E-01 |
| USP44 | 150,58 | 0,12 | 0,11 | 1,08 | 2,79E-01 | 4,85E-01 |
| ERMN | 416,34 | 0,07 | 0,06 | 1,08 | 2,79E-01 | 4,85E-01 |
| ZNF641 | 3521,22 | 0,07 | 0,06 | 1,08 | 2,79E-01 | 4,85E-01 |
| PRKAR2B | 2143,50 | -0,13 | 0,12 | -1,08 | 2,79E-01 | 4,85E-01 |
| OVGP1 | 78,63 | -0,09 | 0,09 | -1,08 | 2,79E-01 | 4,86E-01 |
| WDR54 | 134,31 | -0,06 | 0,05 | -1,08 | 2,79E-01 | 4,86E-01 |
| TTC33 | 540,38 | 0,11 | 0,10 | 1,08 | 2,79E-01 | 4,86E-01 |
| PSMB5 | 424,64 | 0,05 | 0,04 | 1,08 | 2,79E-01 | 4,86E-01 |
| MMP25 | 3870,52 | -0,15 | 0,13 | -1,08 | 2,79E-01 | 4,86E-01 |
| NAAA | 2113,12 | -0,07 | 0,07 | -1,08 | 2,79E-01 | 4,86E-01 |
| SREBF1 | 409,01 | -0,13 | 0,12 | -1,08 | 2,79E-01 | 4,86E-01 |
| POLR1C | 369,27 | 0,04 | 0,04 | 1,08 | 2,79E-01 | 4,86E-01 |
| GBF1 | 1214,89 | -0,07 | 0,07 | -1,08 | 2,80E-01 | 4,86E-01 |
| COPS6 | 394,67 | -0,04 | 0,03 | -1,08 | 2,80E-01 | 4,86E-01 |
| LAPTM4A | 1365,46 | 0,06 | 0,05 | 1,08 | 2,80E-01 | 4,86E-01 |
| HYOU1 | 568,86 | -0,08 | 0,08 | -1,08 | 2,80E-01 | 4,86E-01 |
| NEURL1 | 188,18 | -0,12 | 0,11 | -1,08 | 2,80E-01 | 4,86E-01 |
| HRAS | 92,90 | -0,09 | 0,08 | -1,08 | 2,80E-01 | 4,86E-01 |
| TEX261 | 322,46 | -0,09 | 0,08 | -1,08 | 2,80E-01 | 4,86E-01 |
| CD47 | 9310,95 | 0,05 | 0,05 | 1,08 | 2,80E-01 | 4,87E-01 |
| UACA | 151,52 | -0,14 | 0,13 | -1,08 | 2,80E-01 | 4,87E-01 |
| CAP1 | 7222,58 | -0,05 | 0,05 | -1,08 | 2,80E-01 | 4,87E-01 |
| ATXN7L3B | 1785,78 | 0,04 | 0,03 | 1,08 | 2,81E-01 | 4,87E-01 |
| CFAP20 | 364,34 | 0,04 | 0,04 | 1,08 | 2,81E-01 | 4,87E-01 |
| ATP13A1 | 574,83 | -0,09 | 0,09 | -1,08 | 2,81E-01 | 4,87E-01 |
| ZNF577 | 451,22 | 0,05 | 0,05 | 1,08 | 2,81E-01 | 4,87E-01 |
| STRIP1 | 1054,48 | -0,06 | 0,05 | -1,08 | 2,81E-01 | 4,87E-01 |
| PLCD1 | 358,88 | -0,05 | 0,05 | -1,08 | 2,81E-01 | 4,87E-01 |
| ALS2CL | 327,91 | -0,11 | 0,11 | -1,08 | 2,81E-01 | 4,87E-01 |
| DPF2 | 917,01 | -0,06 | 0,06 | -1,08 | 2,81E-01 | 4,88E-01 |
| MCPH1 | 1067,07 | 0,05 | 0,05 | 1,08 | 2,81E-01 | 4,88E-01 |
| CTLA4 | 250,68 | 0,09 | 0,09 | 1,08 | 2,82E-01 | 4,88E-01 |
| PIPOX | 209,07 | -0,05 | 0,04 | -1,08 | 2,82E-01 | 4,88E-01 |
| CREB5 | 6073,96 | -0,10 | 0,09 | -1,08 | 2,82E-01 | 4,88E-01 |
| TRA2A | 3516,06 | 0,05 | 0,04 | 1,08 | 2,82E-01 | 4,88E-01 |
| CABIN1 | 1759,09 | -0,09 | 0,08 | -1,08 | 2,82E-01 | 4,88E-01 |
| MKS1 | 170,13 | -0,06 | 0,05 | -1,08 | 2,82E-01 | 4,88E-01 |
| TPRG1 | 310,70 | 0,13 | 0,12 | 1,08 | 2,82E-01 | 4,88E-01 |
| MMP17 | 64,69 | -0,22 | 0,20 | -1,08 | 2,82E-01 | 4,89E-01 |

| | | | | | | |
|---|---|---|---|---|---|---|
| TBC1D12 | 307,08 | 0,08 | 0,08 | 1,08 | 2,82E-01 | 4,89E-01 |
| RPA4 | 60,58 | 0,07 | 0,07 | 1,08 | 2,82E-01 | 4,89E-01 |
| P2RY13 | 1466,27 | -0,08 | 0,08 | -1,08 | 2,82E-01 | 4,89E-01 |
| PFAS | 163,03 | -0,12 | 0,11 | -1,08 | 2,82E-01 | 4,89E-01 |
| FAM111A | 3569,35 | 0,05 | 0,04 | 1,08 | 2,82E-01 | 4,89E-01 |
| TMEM127 | 2499,48 | -0,09 | 0,08 | -1,07 | 2,82E-01 | 4,89E-01 |
| CREG1 | 2634,68 | -0,10 | 0,09 | -1,07 | 2,83E-01 | 4,89E-01 |
| OPTN | 3406,71 | -0,09 | 0,09 | -1,07 | 2,83E-01 | 4,89E-01 |
| SNCA | 38538,61 | -0,15 | 0,14 | -1,07 | 2,83E-01 | 4,89E-01 |
| ZBTB47 | 326,05 | -0,07 | 0,07 | -1,07 | 2,83E-01 | 4,89E-01 |
| EXOC7 | 2195,53 | -0,06 | 0,05 | -1,07 | 2,83E-01 | 4,89E-01 |
| B3GLCT | 245,61 | 0,06 | 0,05 | 1,07 | 2,83E-01 | 4,89E-01 |
| BPI | 138,38 | -0,19 | 0,18 | -1,07 | 2,83E-01 | 4,89E-01 |
| TXK | 2739,05 | 0,08 | 0,08 | 1,07 | 2,83E-01 | 4,89E-01 |
| CAMP | 418,51 | -0,19 | 0,18 | -1,07 | 2,83E-01 | 4,89E-01 |
| ATP6V0E2 | 290,77 | -0,08 | 0,07 | -1,07 | 2,83E-01 | 4,89E-01 |
| GSTP1 | 1521,34 | 0,06 | 0,05 | 1,07 | 2,83E-01 | 4,90E-01 |
| USP22 | 1875,41 | -0,08 | 0,07 | -1,07 | 2,83E-01 | 4,90E-01 |
| PRORP | 451,04 | 0,05 | 0,05 | 1,07 | 2,83E-01 | 4,90E-01 |
| SETMAR | 121,87 | 0,06 | 0,05 | 1,07 | 2,84E-01 | 4,90E-01 |
| TADA2B | 1805,12 | -0,04 | 0,04 | -1,07 | 2,84E-01 | 4,90E-01 |
| GLCCI1 | 1130,08 | 0,05 | 0,05 | 1,07 | 2,84E-01 | 4,90E-01 |
| EHMT2 | 522,91 | -0,07 | 0,07 | -1,07 | 2,84E-01 | 4,90E-01 |
| GATAD2B | 1505,87 | -0,10 | 0,09 | -1,07 | 2,84E-01 | 4,90E-01 |
| PKNOX1 | 581,21 | -0,05 | 0,05 | -1,07 | 2,84E-01 | 4,90E-01 |
| MED8 | 731,08 | 0,03 | 0,03 | 1,07 | 2,84E-01 | 4,90E-01 |
| PLAG1 | 360,58 | 0,10 | 0,09 | 1,07 | 2,84E-01 | 4,91E-01 |
| ZNF835 | 92,62 | -0,12 | 0,11 | -1,07 | 2,85E-01 | 4,91E-01 |
| CTCF | 1136,73 | -0,05 | 0,04 | -1,07 | 2,85E-01 | 4,91E-01 |
| TMEM106C | 290,65 | -0,05 | 0,05 | -1,07 | 2,85E-01 | 4,91E-01 |
| BCL2L15 | 106,69 | -0,10 | 0,09 | -1,07 | 2,85E-01 | 4,91E-01 |
| IRF2 | 2576,87 | -0,05 | 0,05 | -1,07 | 2,85E-01 | 4,91E-01 |
| ZC3H10 | 671,03 | 0,03 | 0,03 | 1,07 | 2,85E-01 | 4,91E-01 |
| KLHL7 | 660,62 | 0,05 | 0,04 | 1,07 | 2,85E-01 | 4,91E-01 |
| NCOR2 | 1133,14 | -0,12 | 0,11 | -1,07 | 2,85E-01 | 4,91E-01 |
| GTF3C6 | 317,37 | 0,07 | 0,06 | 1,07 | 2,85E-01 | 4,91E-01 |
| CAMTA1 | 457,84 | -0,06 | 0,06 | -1,07 | 2,85E-01 | 4,91E-01 |
| FAM8A1 | 3727,16 | 0,08 | 0,08 | 1,07 | 2,85E-01 | 4,91E-01 |
| DOCK8 | 19855,17 | -0,06 | 0,06 | -1,07 | 2,85E-01 | 4,91E-01 |
| TMEM25 | 71,06 | -0,12 | 0,11 | -1,07 | 2,85E-01 | 4,91E-01 |
| FOSL2 | 4211,52 | -0,09 | 0,09 | -1,07 | 2,85E-01 | 4,92E-01 |
| RAB44 | 356,95 | -0,13 | 0,12 | -1,07 | 2,85E-01 | 4,92E-01 |
| H1-3 | 3463,61 | 0,08 | 0,08 | 1,07 | 2,86E-01 | 4,92E-01 |
| COMMD1 | 322,82 | 0,04 | 0,04 | 1,07 | 2,86E-01 | 4,92E-01 |
| IGLL5 | 84,90 | 0,17 | 0,16 | 1,07 | 2,86E-01 | 4,92E-01 |
| BPGM | 4060,18 | -0,15 | 0,14 | -1,07 | 2,86E-01 | 4,92E-01 |

| | | | | | | |
|---|---|---|---|---|---|---|
| PRXL2C | 310,11 | 0,05 | 0,05 | 1,07 | 2,86E-01 | 4,92E-01 |
| LTBP3 | 443,53 | -0,14 | 0,13 | -1,07 | 2,86E-01 | 4,92E-01 |
| BUB1B | 58,95 | -0,13 | 0,12 | -1,07 | 2,86E-01 | 4,92E-01 |
| NADK2 | 497,97 | 0,06 | 0,05 | 1,07 | 2,86E-01 | 4,92E-01 |
| UBE2D4 | 139,63 | -0,06 | 0,05 | -1,07 | 2,86E-01 | 4,92E-01 |
| GASK1B | 1334,74 | 0,10 | 0,09 | 1,07 | 2,86E-01 | 4,92E-01 |
| CDKN1B | 6483,53 | 0,06 | 0,05 | 1,07 | 2,87E-01 | 4,93E-01 |
| GGA3 | 832,96 | -0,08 | 0,08 | -1,07 | 2,87E-01 | 4,93E-01 |
| LARP1B | 445,85 | 0,07 | 0,06 | 1,07 | 2,87E-01 | 4,93E-01 |
| TNFRSF9 | 370,64 | -0,09 | 0,08 | -1,07 | 2,87E-01 | 4,93E-01 |
| CDRT4 | 76,29 | -0,14 | 0,13 | -1,06 | 2,87E-01 | 4,93E-01 |
| CYFIP2 | 7344,95 | -0,05 | 0,05 | -1,06 | 2,87E-01 | 4,93E-01 |
| LAMB2 | 56,18 | -0,19 | 0,17 | -1,06 | 2,87E-01 | 4,93E-01 |
| GDPGP1 | 74,91 | -0,09 | 0,08 | -1,06 | 2,87E-01 | 4,93E-01 |
| CALCOCO2 | 2083,31 | -0,06 | 0,05 | -1,06 | 2,87E-01 | 4,93E-01 |
| CASK | 1390,82 | 0,05 | 0,05 | 1,06 | 2,87E-01 | 4,93E-01 |
| SORL1 | 69068,00 | -0,08 | 0,08 | -1,06 | 2,87E-01 | 4,93E-01 |
| STK10 | 7074,18 | -0,09 | 0,08 | -1,06 | 2,87E-01 | 4,93E-01 |
| SUSD4 | 131,51 | -0,13 | 0,12 | -1,06 | 2,88E-01 | 4,94E-01 |
| UBE2R2 | 1704,25 | -0,06 | 0,06 | -1,06 | 2,88E-01 | 4,94E-01 |
| PPFIA1 | 1754,68 | -0,06 | 0,05 | -1,06 | 2,88E-01 | 4,94E-01 |
| VWA8 | 1473,40 | 0,05 | 0,05 | 1,06 | 2,88E-01 | 4,94E-01 |
| SIK3 | 2706,39 | -0,07 | 0,07 | -1,06 | 2,88E-01 | 4,94E-01 |
| CEP164 | 929,58 | -0,07 | 0,07 | -1,06 | 2,88E-01 | 4,94E-01 |
| OLR1 | 39,35 | -0,22 | 0,21 | -1,06 | 2,88E-01 | 4,94E-01 |
| PACC1 | 461,84 | 0,03 | 0,03 | 1,06 | 2,88E-01 | 4,94E-01 |
| TSPYL1 | 2487,45 | 0,05 | 0,04 | 1,06 | 2,88E-01 | 4,95E-01 |
| BMF | 538,46 | -0,08 | 0,07 | -1,06 | 2,89E-01 | 4,95E-01 |
| KYAT3 | 543,28 | 0,05 | 0,05 | 1,06 | 2,89E-01 | 4,95E-01 |
| KIR3DL2 | 32,54 | -0,19 | 0,18 | -1,06 | 2,89E-01 | 4,95E-01 |
| CENPK | 452,90 | 0,16 | 0,15 | 1,06 | 2,88E-01 | 4,95E-01 |
| MRPL51 | 425,94 | 0,09 | 0,08 | 1,06 | 2,89E-01 | 4,95E-01 |
| FSD1L | 331,10 | 0,09 | 0,09 | 1,06 | 2,89E-01 | 4,95E-01 |
| SORT1 | 1274,44 | -0,09 | 0,08 | -1,06 | 2,89E-01 | 4,95E-01 |
| ATP6V0A2 | 1575,99 | 0,03 | 0,03 | 1,06 | 2,89E-01 | 4,95E-01 |
| SULT1A1 | 1460,57 | 0,12 | 0,11 | 1,06 | 2,89E-01 | 4,95E-01 |
| ENSG000002 | 296,07 | -0,12 | 0,11 | -1,06 | 2,89E-01 | 4,95E-01 |
| RNF149 | 20022,97 | -0,06 | 0,06 | -1,06 | 2,89E-01 | 4,95E-01 |
| ATXN2L | 713,66 | -0,12 | 0,11 | -1,06 | 2,89E-01 | 4,95E-01 |
| SELENOT | 2672,32 | 0,08 | 0,08 | 1,06 | 2,90E-01 | 4,96E-01 |
| ETS1 | 11138,24 | 0,05 | 0,05 | 1,06 | 2,90E-01 | 4,96E-01 |
| RCN3 | 188,40 | -0,17 | 0,16 | -1,06 | 2,90E-01 | 4,96E-01 |
| CLIP2 | 230,95 | -0,12 | 0,12 | -1,06 | 2,90E-01 | 4,96E-01 |
| GDAP2 | 2523,36 | 0,05 | 0,05 | 1,06 | 2,90E-01 | 4,96E-01 |
| KPNA6 | 1803,32 | -0,05 | 0,05 | -1,06 | 2,90E-01 | 4,96E-01 |
| KANSL1L | 1100,95 | 0,06 | 0,06 | 1,06 | 2,90E-01 | 4,96E-01 |

| | | | | | | |
|---|---|---|---|---|---|---|
| ARFGAP1 | 665,92 | -0,10 | 0,09 | -1,06 | 2,90E-01 | 4,96E-01 |
| TMEM106A | 347,90 | -0,11 | 0,10 | -1,06 | 2,90E-01 | 4,96E-01 |
| AKIP1 | 187,17 | 0,05 | 0,04 | 1,06 | 2,90E-01 | 4,96E-01 |
| PNMA1 | 383,40 | -0,06 | 0,05 | -1,06 | 2,90E-01 | 4,96E-01 |
| DIP2C | 309,61 | -0,11 | 0,11 | -1,06 | 2,91E-01 | 4,97E-01 |
| C15orf39 | 1728,41 | -0,13 | 0,13 | -1,06 | 2,91E-01 | 4,97E-01 |
| ARHGEF6 | 4685,19 | 0,04 | 0,04 | 1,06 | 2,91E-01 | 4,97E-01 |
| SMG7 | 1939,36 | -0,05 | 0,05 | -1,06 | 2,91E-01 | 4,97E-01 |
| CGRRF1 | 375,17 | 0,08 | 0,08 | 1,06 | 2,91E-01 | 4,97E-01 |
| MTERF4 | 1900,09 | 0,04 | 0,04 | 1,06 | 2,91E-01 | 4,97E-01 |
| ADAMTS10 | 294,52 | -0,11 | 0,10 | -1,06 | 2,91E-01 | 4,98E-01 |
| MLF1 | 26,46 | -0,16 | 0,15 | -1,06 | 2,91E-01 | 4,98E-01 |
| H3C1 | 260,16 | 0,09 | 0,08 | 1,05 | 2,91E-01 | 4,98E-01 |
| H2BC13 | 193,30 | -0,10 | 0,10 | -1,05 | 2,92E-01 | 4,98E-01 |
| POLD1 | 373,99 | -0,06 | 0,05 | -1,05 | 2,92E-01 | 4,98E-01 |
| RTN3 | 3050,43 | -0,07 | 0,06 | -1,05 | 2,92E-01 | 4,98E-01 |
| KCTD7 | 1232,59 | 0,05 | 0,05 | 1,05 | 2,92E-01 | 4,98E-01 |
| TTC16 | 301,13 | -0,13 | 0,12 | -1,05 | 2,92E-01 | 4,98E-01 |
| CHD2 | 12970,05 | 0,04 | 0,04 | 1,05 | 2,92E-01 | 4,98E-01 |
| TBC1D3L | 42,03 | -0,21 | 0,20 | -1,05 | 2,92E-01 | 4,98E-01 |
| PROK2 | 2495,85 | -0,11 | 0,10 | -1,05 | 2,92E-01 | 4,98E-01 |
| CTNNAL1 | 241,45 | -0,15 | 0,15 | -1,05 | 2,92E-01 | 4,98E-01 |
| ZFC3H1 | 5579,02 | 0,06 | 0,05 | 1,05 | 2,92E-01 | 4,98E-01 |
| BCDIN3D | 316,21 | 0,06 | 0,05 | 1,05 | 2,92E-01 | 4,98E-01 |
| PYHIN1 | 1510,79 | 0,09 | 0,08 | 1,05 | 2,92E-01 | 4,98E-01 |
| TRMT2B | 1162,34 | 0,04 | 0,04 | 1,05 | 2,93E-01 | 4,99E-01 |
| KMT2D | 4397,21 | -0,13 | 0,12 | -1,05 | 2,93E-01 | 4,99E-01 |
| DPYSL2 | 2252,62 | 0,07 | 0,06 | 1,05 | 2,93E-01 | 4,99E-01 |
| CD40 | 321,84 | 0,08 | 0,07 | 1,05 | 2,93E-01 | 4,99E-01 |
| SLC17A9 | 134,80 | -0,12 | 0,11 | -1,05 | 2,93E-01 | 4,99E-01 |
| STRN | 3178,12 | 0,05 | 0,05 | 1,05 | 2,93E-01 | 4,99E-01 |
| TDRD7 | 1288,88 | 0,06 | 0,06 | 1,05 | 2,93E-01 | 4,99E-01 |
| SPEN | 5602,46 | 0,04 | 0,04 | 1,05 | 2,93E-01 | 4,99E-01 |
| MAT2A | 2305,63 | 0,04 | 0,04 | 1,05 | 2,93E-01 | 4,99E-01 |
| DERA | 561,52 | 0,04 | 0,04 | 1,05 | 2,93E-01 | 4,99E-01 |
| NAP1L2 | 166,08 | 0,11 | 0,10 | 1,05 | 2,93E-01 | 4,99E-01 |
| EVI5 | 1363,44 | 0,06 | 0,06 | 1,05 | 2,93E-01 | 4,99E-01 |
| H2BC18 | 2603,73 | -0,07 | 0,07 | -1,05 | 2,93E-01 | 4,99E-01 |
| STIMATE | 87,05 | -0,08 | 0,07 | -1,05 | 2,94E-01 | 4,99E-01 |
| LGSN | 29,70 | -0,25 | 0,24 | -1,05 | 2,94E-01 | 4,99E-01 |
| PHETA2 | 127,02 | -0,07 | 0,06 | -1,05 | 2,94E-01 | 4,99E-01 |
| CGGBP1 | 4755,24 | 0,08 | 0,07 | 1,05 | 2,94E-01 | 5,00E-01 |
| PKIA | 283,17 | 0,09 | 0,09 | 1,05 | 2,94E-01 | 5,00E-01 |
| CNST | 2315,92 | 0,05 | 0,04 | 1,05 | 2,94E-01 | 5,00E-01 |
| FAM72B | 25,51 | -0,11 | 0,11 | -1,05 | 2,94E-01 | 5,00E-01 |
| SLC40A1 | 3011,89 | -0,08 | 0,07 | -1,05 | 2,94E-01 | 5,00E-01 |

| | | | | | | |
|---|---|---|---|---|---|---|
| TTC9 | 478,26 | -0,08 | 0,08 | -1,05 | 2,94E-01 | 5,00E-01 |
| XXYLT1 | 241,64 | -0,07 | 0,06 | -1,05 | 2,94E-01 | 5,00E-01 |
| EPG5 | 4006,55 | -0,05 | 0,05 | -1,05 | 2,94E-01 | 5,00E-01 |
| TNFRSF1B | 8082,81 | -0,08 | 0,08 | -1,05 | 2,94E-01 | 5,00E-01 |
| CUL9 | 1041,83 | -0,08 | 0,08 | -1,05 | 2,95E-01 | 5,00E-01 |
| SURF2 | 159,86 | -0,04 | 0,04 | -1,05 | 2,95E-01 | 5,01E-01 |
| KDM4B | 2883,41 | -0,10 | 0,10 | -1,05 | 2,95E-01 | 5,01E-01 |
| OSGIN2 | 1678,31 | 0,09 | 0,09 | 1,05 | 2,95E-01 | 5,01E-01 |
| PCDH12 | 41,48 | -0,18 | 0,17 | -1,05 | 2,95E-01 | 5,01E-01 |
| RASSF4 | 1219,48 | 0,07 | 0,07 | 1,05 | 2,95E-01 | 5,01E-01 |
| FAM3B | 36,20 | -0,40 | 0,38 | -1,05 | 2,96E-01 | 5,01E-01 |
| C2orf88 | 828,37 | -0,11 | 0,11 | -1,05 | 2,96E-01 | 5,01E-01 |
| SMARCA4 | 969,46 | -0,10 | 0,10 | -1,05 | 2,96E-01 | 5,01E-01 |
| ERGIC1 | 5018,10 | -0,07 | 0,07 | -1,05 | 2,96E-01 | 5,01E-01 |
| CHEK2 | 194,68 | -0,06 | 0,06 | -1,05 | 2,96E-01 | 5,01E-01 |
| VEZF1 | 2238,99 | 0,05 | 0,05 | 1,05 | 2,96E-01 | 5,01E-01 |
| SH3BP2 | 4227,38 | -0,10 | 0,10 | -1,05 | 2,95E-01 | 5,01E-01 |
| COX10 | 342,66 | 0,03 | 0,03 | 1,05 | 2,96E-01 | 5,01E-01 |
| MVK | 189,29 | -0,08 | 0,08 | -1,05 | 2,96E-01 | 5,01E-01 |
| SEC31A | 2157,03 | 0,04 | 0,03 | 1,05 | 2,96E-01 | 5,01E-01 |
| TBL3 | 320,59 | -0,06 | 0,06 | -1,05 | 2,96E-01 | 5,01E-01 |
| EXOC8 | 2363,06 | 0,06 | 0,05 | 1,05 | 2,96E-01 | 5,01E-01 |
| TRAPPC4 | 904,13 | 0,04 | 0,04 | 1,04 | 2,96E-01 | 5,01E-01 |
| SIN3B | 621,13 | -0,13 | 0,12 | -1,04 | 2,96E-01 | 5,01E-01 |
| EXOC4 | 2681,38 | 0,04 | 0,04 | 1,05 | 2,96E-01 | 5,01E-01 |
| PPP6R2 | 1365,30 | -0,07 | 0,07 | -1,05 | 2,96E-01 | 5,01E-01 |
| TCEA3 | 141,32 | -0,11 | 0,10 | -1,04 | 2,96E-01 | 5,02E-01 |
| CEP68 | 667,11 | 0,06 | 0,05 | 1,04 | 2,97E-01 | 5,02E-01 |
| CALCOCO1 | 2969,70 | -0,05 | 0,05 | -1,04 | 2,97E-01 | 5,02E-01 |
| CSK | 2449,12 | -0,10 | 0,09 | -1,04 | 2,97E-01 | 5,03E-01 |
| MLLT1 | 322,14 | -0,14 | 0,13 | -1,04 | 2,97E-01 | 5,03E-01 |
| MIDEAS | 2156,23 | -0,07 | 0,07 | -1,04 | 2,97E-01 | 5,03E-01 |
| TEC | 421,45 | -0,07 | 0,07 | -1,04 | 2,97E-01 | 5,03E-01 |
| RAF1 | 7091,43 | -0,06 | 0,05 | -1,04 | 2,97E-01 | 5,03E-01 |
| EBPL | 217,56 | 0,06 | 0,05 | 1,04 | 2,97E-01 | 5,03E-01 |
| STK17A | 4059,93 | 0,06 | 0,06 | 1,04 | 2,97E-01 | 5,03E-01 |
| NOP2 | 57,03 | -0,07 | 0,07 | -1,04 | 2,97E-01 | 5,03E-01 |
| L1TD1 | 40,14 | -0,21 | 0,20 | -1,04 | 2,97E-01 | 5,03E-01 |
| TP53I3 | 129,35 | -0,08 | 0,08 | -1,04 | 2,98E-01 | 5,03E-01 |
| GPD2 | 1436,22 | 0,06 | 0,06 | 1,04 | 2,98E-01 | 5,03E-01 |
| CHD4 | 423,74 | -0,05 | 0,05 | -1,04 | 2,98E-01 | 5,03E-01 |
| SRRM2 | 5857,98 | -0,11 | 0,11 | -1,04 | 2,98E-01 | 5,03E-01 |
| SDHAF4 | 214,24 | 0,05 | 0,05 | 1,04 | 2,98E-01 | 5,03E-01 |
| RAP1GAP2 | 2562,89 | -0,10 | 0,10 | -1,04 | 2,98E-01 | 5,03E-01 |
| TPCN1 | 432,25 | -0,13 | 0,12 | -1,04 | 2,98E-01 | 5,04E-01 |
| ANXA3 | 912,13 | -0,13 | 0,12 | -1,04 | 2,98E-01 | 5,04E-01 |

| | | | | | | |
|---|---|---|---|---|---|---|
| CCR9 | 44,59 | 0,12 | 0,12 | 1,04 | 2,98E-01 | 5,04E-01 |
| STOML1 | 207,95 | -0,07 | 0,07 | -1,04 | 2,98E-01 | 5,04E-01 |
| NME7 | 228,89 | 0,07 | 0,07 | 1,04 | 2,98E-01 | 5,04E-01 |
| DPP4 | 1046,44 | 0,08 | 0,07 | 1,04 | 2,99E-01 | 5,04E-01 |
| KCMF1 | 2161,76 | -0,03 | 0,03 | -1,04 | 2,99E-01 | 5,04E-01 |
| TIAL1 | 1918,93 | 0,06 | 0,06 | 1,04 | 2,99E-01 | 5,04E-01 |
| IFIT1 | 1634,73 | 0,23 | 0,22 | 1,04 | 2,99E-01 | 5,04E-01 |
| PDZD8 | 3312,84 | 0,07 | 0,07 | 1,04 | 2,99E-01 | 5,04E-01 |
| NOTCH2NLA | 506,58 | 0,08 | 0,08 | 1,04 | 2,99E-01 | 5,04E-01 |
| P3H3 | 42,91 | -0,17 | 0,16 | -1,04 | 2,99E-01 | 5,04E-01 |
| TPCN2 | 836,39 | -0,10 | 0,10 | -1,04 | 2,99E-01 | 5,04E-01 |
| ENO1 | 7375,12 | 0,06 | 0,06 | 1,04 | 2,99E-01 | 5,04E-01 |
| TXNIP | 70242,80 | 0,05 | 0,05 | 1,04 | 2,99E-01 | 5,04E-01 |
| B3GALT2 | 70,96 | 0,16 | 0,15 | 1,04 | 2,99E-01 | 5,05E-01 |
| MAD2L1BP | 349,12 | -0,04 | 0,04 | -1,04 | 3,00E-01 | 5,05E-01 |
| CD52 | 4379,19 | 0,11 | 0,11 | 1,04 | 3,00E-01 | 5,05E-01 |
| MARCHF8 | 12168,71 | -0,13 | 0,13 | -1,04 | 3,00E-01 | 5,05E-01 |
| TBC1D2 | 391,28 | -0,08 | 0,08 | -1,04 | 3,00E-01 | 5,05E-01 |
| TEPSIN | 629,73 | -0,09 | 0,09 | -1,04 | 3,00E-01 | 5,05E-01 |
| TBC1D9B | 848,66 | -0,10 | 0,10 | -1,04 | 3,00E-01 | 5,05E-01 |
| VKORC1L1 | 620,87 | 0,03 | 0,03 | 1,04 | 3,00E-01 | 5,05E-01 |
| RARG | 245,22 | -0,08 | 0,08 | -1,04 | 3,00E-01 | 5,06E-01 |
| IL32 | 673,01 | -0,10 | 0,09 | -1,04 | 3,00E-01 | 5,06E-01 |
| TAF1D | 3933,14 | 0,10 | 0,10 | 1,04 | 3,00E-01 | 5,06E-01 |
| ACADVL | 2236,00 | -0,05 | 0,05 | -1,04 | 3,01E-01 | 5,06E-01 |
| SEC61B | 943,01 | 0,06 | 0,06 | 1,03 | 3,01E-01 | 5,06E-01 |
| ACSL3 | 1352,00 | 0,05 | 0,05 | 1,03 | 3,01E-01 | 5,06E-01 |
| MFSD10 | 435,60 | -0,07 | 0,07 | -1,03 | 3,01E-01 | 5,06E-01 |
| ATAD5 | 314,41 | 0,06 | 0,06 | 1,03 | 3,01E-01 | 5,06E-01 |
| RRP8 | 891,40 | 0,03 | 0,03 | 1,03 | 3,01E-01 | 5,06E-01 |
| IQCG | 315,72 | 0,05 | 0,05 | 1,03 | 3,01E-01 | 5,06E-01 |
| DRG1 | 666,53 | 0,04 | 0,04 | 1,03 | 3,01E-01 | 5,06E-01 |
| ADGRA2 | 76,03 | -0,18 | 0,17 | -1,03 | 3,01E-01 | 5,06E-01 |
| CIC | 581,34 | -0,15 | 0,14 | -1,03 | 3,01E-01 | 5,07E-01 |
| LPL | 37,24 | -0,22 | 0,21 | -1,03 | 3,01E-01 | 5,07E-01 |
| MIGA2 | 610,68 | -0,06 | 0,06 | -1,03 | 3,02E-01 | 5,07E-01 |
| ACAD10 | 735,35 | -0,06 | 0,05 | -1,03 | 3,02E-01 | 5,07E-01 |
| ZDHHC24 | 396,65 | -0,08 | 0,07 | -1,03 | 3,02E-01 | 5,07E-01 |
| NIN | 14509,81 | 0,05 | 0,05 | 1,03 | 3,02E-01 | 5,07E-01 |
| PRKAB1 | 887,95 | -0,03 | 0,03 | -1,03 | 3,02E-01 | 5,07E-01 |
| CHMP2A | 1202,27 | -0,07 | 0,06 | -1,03 | 3,02E-01 | 5,08E-01 |
| OPHN1 | 106,41 | -0,10 | 0,10 | -1,03 | 3,02E-01 | 5,08E-01 |
| SNRNP35 | 708,40 | -0,03 | 0,03 | -1,03 | 3,03E-01 | 5,08E-01 |
| PSMA7 | 1987,68 | 0,05 | 0,05 | 1,03 | 3,02E-01 | 5,08E-01 |
| DOCK11 | 9744,35 | 0,05 | 0,05 | 1,03 | 3,03E-01 | 5,08E-01 |
| GPR89B | 532,94 | 0,05 | 0,05 | 1,03 | 3,03E-01 | 5,08E-01 |

| | | | | | | |
|---|---|---|---|---|---|---|
| FBXW2 | 3959,10 | 0,04 | 0,04 | 1,03 | 3,03E-01 | 5,08E-01 |
| TMEM60 | 316,72 | 0,06 | 0,06 | 1,03 | 3,03E-01 | 5,08E-01 |
| CDKL5 | 847,78 | -0,06 | 0,05 | -1,03 | 3,03E-01 | 5,08E-01 |
| DGCR8 | 620,17 | -0,07 | 0,07 | -1,03 | 3,03E-01 | 5,08E-01 |
| C1GALT1 | 3009,53 | 0,07 | 0,07 | 1,03 | 3,03E-01 | 5,08E-01 |
| UBXN7 | 3782,26 | 0,04 | 0,04 | 1,03 | 3,03E-01 | 5,08E-01 |
| NAIF1 | 372,06 | -0,04 | 0,04 | -1,03 | 3,03E-01 | 5,08E-01 |
| DNAH1 | 2155,52 | -0,09 | 0,09 | -1,03 | 3,03E-01 | 5,08E-01 |
| TCN2 | 67,06 | -0,12 | 0,11 | -1,03 | 3,03E-01 | 5,08E-01 |
| TBL1X | 2656,14 | -0,08 | 0,08 | -1,03 | 3,03E-01 | 5,08E-01 |
| NPEPPS | 3510,30 | -0,04 | 0,04 | -1,03 | 3,03E-01 | 5,08E-01 |
| CNN3 | 33,89 | -0,15 | 0,15 | -1,03 | 3,03E-01 | 5,08E-01 |
| ST13 | 2002,85 | -0,07 | 0,07 | -1,03 | 3,04E-01 | 5,09E-01 |
| MS4A3 | 187,83 | -0,19 | 0,18 | -1,03 | 3,04E-01 | 5,09E-01 |
| VGLL4 | 596,65 | -0,05 | 0,05 | -1,03 | 3,04E-01 | 5,09E-01 |
| RALBP1 | 3134,64 | -0,07 | 0,07 | -1,03 | 3,04E-01 | 5,09E-01 |
| POLR2A | 1474,71 | -0,12 | 0,12 | -1,03 | 3,04E-01 | 5,09E-01 |
| KIF1B | 2419,49 | -0,07 | 0,07 | -1,03 | 3,04E-01 | 5,09E-01 |
| OLFM4 | 135,91 | -0,25 | 0,24 | -1,03 | 3,04E-01 | 5,09E-01 |
| EPPK1 | 260,22 | -0,13 | 0,13 | -1,03 | 3,04E-01 | 5,09E-01 |
| PRDX5 | 3890,89 | -0,11 | 0,10 | -1,03 | 3,04E-01 | 5,09E-01 |
| ATXN7L1 | 797,77 | -0,06 | 0,05 | -1,03 | 3,04E-01 | 5,09E-01 |
| FAM98C | 362,06 | -0,06 | 0,06 | -1,03 | 3,04E-01 | 5,09E-01 |
| ARFGAP3 | 1385,99 | 0,04 | 0,04 | 1,03 | 3,05E-01 | 5,09E-01 |
| PYCR3 | 71,87 | -0,10 | 0,10 | -1,03 | 3,05E-01 | 5,09E-01 |
| MINK1 | 636,31 | -0,13 | 0,13 | -1,03 | 3,05E-01 | 5,09E-01 |
| AKAP8 | 1229,92 | 0,03 | 0,02 | 1,03 | 3,05E-01 | 5,09E-01 |
| NDUFAF6 | 385,39 | 0,04 | 0,04 | 1,03 | 3,05E-01 | 5,09E-01 |
| PITPNM2 | 287,32 | -0,13 | 0,13 | -1,03 | 3,05E-01 | 5,10E-01 |
| TBC1D2B | 2995,81 | -0,05 | 0,04 | -1,03 | 3,05E-01 | 5,10E-01 |
| MORN3 | 134,63 | -0,08 | 0,08 | -1,03 | 3,05E-01 | 5,10E-01 |
| HNRNPC | 13360,12 | 0,05 | 0,05 | 1,02 | 3,05E-01 | 5,10E-01 |
| NOP9 | 1418,80 | 0,05 | 0,05 | 1,02 | 3,05E-01 | 5,10E-01 |
| NDST1 | 1539,93 | -0,11 | 0,11 | -1,02 | 3,05E-01 | 5,10E-01 |
| PRMT5 | 475,09 | -0,05 | 0,05 | -1,02 | 3,06E-01 | 5,10E-01 |
| SUFU | 242,97 | -0,10 | 0,10 | -1,02 | 3,06E-01 | 5,10E-01 |
| PRRC2A | 1117,49 | -0,14 | 0,14 | -1,02 | 3,06E-01 | 5,10E-01 |
| DNAL4 | 126,73 | 0,05 | 0,04 | 1,02 | 3,06E-01 | 5,10E-01 |
| TSNARE1 | 285,16 | -0,09 | 0,09 | -1,02 | 3,06E-01 | 5,11E-01 |
| PDCD7 | 639,26 | 0,04 | 0,04 | 1,02 | 3,06E-01 | 5,11E-01 |
| PAK2 | 4443,65 | -0,06 | 0,05 | -1,02 | 3,06E-01 | 5,11E-01 |
| GOT2 | 378,73 | -0,04 | 0,04 | -1,02 | 3,06E-01 | 5,11E-01 |
| GALK1 | 328,65 | -0,06 | 0,05 | -1,02 | 3,06E-01 | 5,11E-01 |
| FAM200C | 64,29 | 0,09 | 0,09 | 1,02 | 3,06E-01 | 5,11E-01 |
| VAMP2 | 377,11 | -0,10 | 0,10 | -1,02 | 3,06E-01 | 5,11E-01 |
| RELA | 598,38 | -0,08 | 0,08 | -1,02 | 3,06E-01 | 5,11E-01 |

| | | | | | | |
|---|---|---|---|---|---|---|
| ICA1L | 134,48 | -0,10 | 0,09 | -1,02 | 3,06E-01 | 5,11E-01 |
| FCRL2 | 831,67 | 0,14 | 0,13 | 1,02 | 3,06E-01 | 5,11E-01 |
| ESYT1 | 2420,69 | -0,07 | 0,07 | -1,02 | 3,06E-01 | 5,11E-01 |
| PITPNA | 1910,23 | -0,06 | 0,06 | -1,02 | 3,07E-01 | 5,11E-01 |
| CARD8 | 6312,19 | -0,05 | 0,05 | -1,02 | 3,07E-01 | 5,11E-01 |
| ZEB1 | 2307,69 | 0,05 | 0,05 | 1,02 | 3,07E-01 | 5,11E-01 |
| PSMG2 | 748,14 | 0,04 | 0,04 | 1,02 | 3,07E-01 | 5,11E-01 |
| FMN1 | 663,96 | -0,19 | 0,19 | -1,02 | 3,07E-01 | 5,11E-01 |
| ZNF493 | 2220,15 | 0,06 | 0,06 | 1,02 | 3,07E-01 | 5,11E-01 |
| PHF21A | 3755,84 | -0,06 | 0,05 | -1,02 | 3,07E-01 | 5,11E-01 |
| GIGYF2 | 4446,41 | 0,04 | 0,04 | 1,02 | 3,07E-01 | 5,11E-01 |
| GCDH | 452,35 | -0,04 | 0,04 | -1,02 | 3,07E-01 | 5,11E-01 |
| CHPF2 | 1189,24 | -0,08 | 0,07 | -1,02 | 3,07E-01 | 5,11E-01 |
| PLSCR2 | 49,25 | -0,12 | 0,12 | -1,02 | 3,07E-01 | 5,11E-01 |
| CCDC183 | 28,31 | -0,10 | 0,10 | -1,02 | 3,07E-01 | 5,12E-01 |
| KLF11 | 470,84 | 0,05 | 0,05 | 1,02 | 3,07E-01 | 5,12E-01 |
| MTMR12 | 1103,82 | -0,03 | 0,03 | -1,02 | 3,08E-01 | 5,12E-01 |
| UQCC6 | 185,33 | 0,06 | 0,06 | 1,02 | 3,08E-01 | 5,12E-01 |
| PTPN6 | 2774,44 | -0,10 | 0,10 | -1,02 | 3,08E-01 | 5,12E-01 |
| ACAD11 | 246,79 | 0,05 | 0,05 | 1,02 | 3,08E-01 | 5,12E-01 |
| MAD2L2 | 283,77 | -0,04 | 0,04 | -1,02 | 3,08E-01 | 5,12E-01 |
| PLIN3 | 1058,11 | -0,07 | 0,07 | -1,02 | 3,08E-01 | 5,12E-01 |
| GGA2 | 2346,43 | 0,05 | 0,05 | 1,02 | 3,08E-01 | 5,12E-01 |
| NEK8 | 82,36 | -0,11 | 0,11 | -1,02 | 3,08E-01 | 5,12E-01 |
| N4BP3 | 95,16 | -0,13 | 0,13 | -1,02 | 3,08E-01 | 5,12E-01 |
| JMJD4 | 561,28 | 0,05 | 0,05 | 1,02 | 3,09E-01 | 5,13E-01 |
| HS1BP3 | 583,99 | -0,08 | 0,08 | -1,02 | 3,09E-01 | 5,13E-01 |
| RABGAP1L | 9063,84 | 0,05 | 0,05 | 1,02 | 3,09E-01 | 5,13E-01 |
| FRAT2 | 7735,62 | -0,08 | 0,07 | -1,02 | 3,09E-01 | 5,13E-01 |
| SH2D2A | 148,37 | -0,14 | 0,14 | -1,02 | 3,09E-01 | 5,13E-01 |
| SNTB1 | 1081,79 | -0,04 | 0,04 | -1,02 | 3,09E-01 | 5,13E-01 |
| INO80C | 195,48 | -0,05 | 0,05 | -1,02 | 3,09E-01 | 5,13E-01 |
| TIGIT | 1261,46 | 0,08 | 0,08 | 1,02 | 3,09E-01 | 5,13E-01 |
| CORO7 | 555,34 | -0,09 | 0,08 | -1,02 | 3,09E-01 | 5,13E-01 |
| YY1 | 3361,84 | 0,03 | 0,03 | 1,02 | 3,09E-01 | 5,13E-01 |
| SAT1 | 9248,27 | 0,08 | 0,08 | 1,02 | 3,09E-01 | 5,13E-01 |
| ACP5 | 341,20 | -0,10 | 0,10 | -1,02 | 3,09E-01 | 5,13E-01 |
| RHOQ | 2530,36 | 0,05 | 0,05 | 1,02 | 3,09E-01 | 5,13E-01 |
| ZNF133 | 564,11 | 0,03 | 0,03 | 1,02 | 3,09E-01 | 5,13E-01 |
| CTSC | 6388,40 | 0,06 | 0,06 | 1,02 | 3,10E-01 | 5,14E-01 |
| TENT2 | 4351,20 | 0,05 | 0,05 | 1,02 | 3,10E-01 | 5,14E-01 |
| SEC23B | 1572,56 | 0,04 | 0,04 | 1,02 | 3,10E-01 | 5,14E-01 |
| WDR13 | 1225,10 | -0,05 | 0,05 | -1,02 | 3,10E-01 | 5,14E-01 |
| ATG3 | 2572,95 | 0,07 | 0,07 | 1,02 | 3,10E-01 | 5,14E-01 |
| NOP10 | 1067,23 | -0,07 | 0,07 | -1,01 | 3,10E-01 | 5,14E-01 |
| ADAM19 | 3560,56 | -0,07 | 0,07 | -1,01 | 3,10E-01 | 5,14E-01 |

| | | | | | | |
|---|---|---|---|---|---|---|
| TBC1D5 | 3352,44 | 0,04 | 0,04 | 1,01 | 3,10E-01 | 5,14E-01 |
| RCSD1 | 6895,78 | 0,05 | 0,05 | 1,01 | 3,10E-01 | 5,14E-01 |
| MAZ | 562,29 | -0,11 | 0,11 | -1,01 | 3,11E-01 | 5,14E-01 |
| PDSS1 | 151,04 | 0,05 | 0,05 | 1,01 | 3,11E-01 | 5,15E-01 |
| OCIAD2 | 228,85 | 0,07 | 0,06 | 1,01 | 3,11E-01 | 5,15E-01 |
| CSNK1A1 | 3045,13 | 0,04 | 0,04 | 1,01 | 3,11E-01 | 5,15E-01 |
| PHB2 | 1513,44 | 0,05 | 0,04 | 1,01 | 3,11E-01 | 5,15E-01 |
| NPIPB3 | 70,15 | -0,15 | 0,15 | -1,01 | 3,11E-01 | 5,15E-01 |
| CASP8 | 8806,08 | 0,05 | 0,05 | 1,01 | 3,11E-01 | 5,15E-01 |
| ALOX5 | 5660,21 | -0,07 | 0,07 | -1,01 | 3,11E-01 | 5,15E-01 |
| RLIG1 | 578,16 | 0,09 | 0,08 | 1,01 | 3,11E-01 | 5,15E-01 |
| PLXND1 | 594,12 | -0,13 | 0,13 | -1,01 | 3,11E-01 | 5,15E-01 |
| APEH | 1298,84 | -0,03 | 0,03 | -1,01 | 3,12E-01 | 5,16E-01 |
| QKI | 10201,95 | 0,07 | 0,07 | 1,01 | 3,12E-01 | 5,16E-01 |
| HLX | 494,87 | -0,09 | 0,09 | -1,01 | 3,12E-01 | 5,16E-01 |
| MADD | 1976,85 | -0,06 | 0,06 | -1,01 | 3,12E-01 | 5,16E-01 |
| CD22 | 625,60 | 0,12 | 0,11 | 1,01 | 3,12E-01 | 5,16E-01 |
| INPPL1 | 936,21 | -0,10 | 0,10 | -1,01 | 3,12E-01 | 5,16E-01 |
| RPS5 | 6618,98 | 0,08 | 0,08 | 1,01 | 3,12E-01 | 5,16E-01 |
| C2orf92 | 139,16 | -0,09 | 0,09 | -1,01 | 3,12E-01 | 5,16E-01 |
| TMCO4 | 456,71 | -0,06 | 0,06 | -1,01 | 3,12E-01 | 5,16E-01 |
| CS | 962,05 | -0,06 | 0,06 | -1,01 | 3,12E-01 | 5,16E-01 |
| VOPP1 | 2148,44 | -0,05 | 0,05 | -1,01 | 3,12E-01 | 5,16E-01 |
| TMEM140 | 4051,66 | -0,07 | 0,07 | -1,01 | 3,13E-01 | 5,16E-01 |
| DOK3 | 1842,25 | -0,10 | 0,10 | -1,01 | 3,13E-01 | 5,16E-01 |
| SUGP1 | 317,35 | -0,08 | 0,08 | -1,01 | 3,13E-01 | 5,16E-01 |
| PLGLB1 | 122,85 | 0,10 | 0,10 | 1,01 | 3,13E-01 | 5,16E-01 |
| CLHC1 | 238,38 | 0,10 | 0,09 | 1,01 | 3,13E-01 | 5,17E-01 |
| RPL26L1 | 66,09 | 0,12 | 0,12 | 1,01 | 3,13E-01 | 5,17E-01 |
| CHAC2 | 52,26 | 0,10 | 0,10 | 1,01 | 3,13E-01 | 5,17E-01 |
| LLGL2 | 489,68 | -0,10 | 0,09 | -1,01 | 3,13E-01 | 5,17E-01 |
| ACSL4 | 4286,26 | 0,08 | 0,08 | 1,01 | 3,13E-01 | 5,17E-01 |
| TSPO | 3705,76 | -0,07 | 0,07 | -1,01 | 3,13E-01 | 5,17E-01 |
| THRAP3 | 5561,72 | 0,05 | 0,05 | 1,01 | 3,14E-01 | 5,17E-01 |
| SOS2 | 4029,57 | 0,06 | 0,06 | 1,01 | 3,14E-01 | 5,17E-01 |
| AMPD3 | 1837,14 | -0,05 | 0,05 | -1,01 | 3,14E-01 | 5,17E-01 |
| P3H1 | 491,08 | -0,04 | 0,04 | -1,01 | 3,14E-01 | 5,17E-01 |
| UQCRQ | 448,64 | 0,09 | 0,09 | 1,01 | 3,14E-01 | 5,17E-01 |
| FAM184A | 123,52 | -0,11 | 0,11 | -1,01 | 3,14E-01 | 5,17E-01 |
| MRPS16 | 1355,30 | 0,04 | 0,04 | 1,01 | 3,14E-01 | 5,17E-01 |
| SCAF8 | 3026,04 | 0,03 | 0,03 | 1,01 | 3,14E-01 | 5,17E-01 |
| SLF1 | 1005,16 | 0,08 | 0,08 | 1,01 | 3,14E-01 | 5,17E-01 |
| PTAFR | 7040,29 | -0,08 | 0,08 | -1,01 | 3,14E-01 | 5,17E-01 |
| WSB1 | 7509,91 | 0,06 | 0,06 | 1,01 | 3,14E-01 | 5,17E-01 |
| RNF166 | 1436,84 | -0,11 | 0,11 | -1,01 | 3,14E-01 | 5,17E-01 |
| TEP1 | 3562,92 | -0,06 | 0,06 | -1,01 | 3,14E-01 | 5,18E-01 |

| | | | | | | |
|---|---|---|---|---|---|---|
| CMTM7 | 935,45 | -0,05 | 0,05 | -1,01 | 3,14E-01 | 5,18E-01 |
| PPP1R16B | 1363,30 | -0,06 | 0,06 | -1,01 | 3,14E-01 | 5,18E-01 |
| NPIPB12 | 104,41 | -0,11 | 0,11 | -1,01 | 3,14E-01 | 5,18E-01 |
| PLXDC1 | 163,76 | -0,13 | 0,12 | -1,01 | 3,14E-01 | 5,18E-01 |
| CXXC1 | 841,13 | -0,04 | 0,04 | -1,01 | 3,14E-01 | 5,18E-01 |
| HRH4 | 448,82 | -0,13 | 0,13 | -1,01 | 3,14E-01 | 5,18E-01 |
| ITPRIPL2 | 766,36 | 0,06 | 0,06 | 1,01 | 3,15E-01 | 5,18E-01 |
| TUBD1 | 342,06 | 0,04 | 0,04 | 1,01 | 3,15E-01 | 5,18E-01 |
| HMGA1 | 197,10 | -0,11 | 0,11 | -1,01 | 3,15E-01 | 5,18E-01 |
| PWWP2B | 279,81 | -0,06 | 0,06 | -1,01 | 3,15E-01 | 5,18E-01 |
| BORCS5 | 568,05 | -0,07 | 0,07 | -1,01 | 3,15E-01 | 5,18E-01 |
| MTX2 | 323,61 | 0,07 | 0,07 | 1,01 | 3,15E-01 | 5,18E-01 |
| ACP6 | 509,20 | -0,06 | 0,06 | -1,01 | 3,15E-01 | 5,18E-01 |
| MTNAP1 | 449,83 | 0,05 | 0,05 | 1,01 | 3,15E-01 | 5,18E-01 |
| SSR2 | 2587,50 | 0,05 | 0,05 | 1,00 | 3,15E-01 | 5,18E-01 |
| SRD5A3 | 155,41 | 0,08 | 0,08 | 1,00 | 3,15E-01 | 5,18E-01 |
| CCNL2 | 4005,72 | -0,03 | 0,03 | -1,00 | 3,15E-01 | 5,18E-01 |
| RAB22A | 1787,80 | 0,03 | 0,03 | 1,00 | 3,15E-01 | 5,18E-01 |
| COQ4 | 249,22 | -0,08 | 0,08 | -1,00 | 3,15E-01 | 5,18E-01 |
| RENBP | 458,93 | -0,07 | 0,07 | -1,00 | 3,15E-01 | 5,18E-01 |
| PARN | 1156,04 | 0,03 | 0,03 | 1,00 | 3,15E-01 | 5,18E-01 |
| KARS1 | 2056,32 | 0,04 | 0,04 | 1,00 | 3,16E-01 | 5,18E-01 |
| AARS1 | 741,90 | -0,05 | 0,05 | -1,00 | 3,15E-01 | 5,18E-01 |
| ZNF362 | 311,24 | -0,08 | 0,08 | -1,00 | 3,16E-01 | 5,18E-01 |
| C11orf65 | 31,58 | -0,14 | 0,14 | -1,00 | 3,16E-01 | 5,18E-01 |
| GPHN | 189,18 | -0,07 | 0,07 | -1,00 | 3,16E-01 | 5,19E-01 |
| SETD1B | 769,01 | -0,14 | 0,14 | -1,00 | 3,16E-01 | 5,19E-01 |
| TNFAIP3 | 1009,37 | -0,06 | 0,06 | -1,00 | 3,16E-01 | 5,19E-01 |
| ACOT13 | 276,81 | 0,05 | 0,05 | 1,00 | 3,16E-01 | 5,19E-01 |
| MYBBP1A | 345,00 | -0,09 | 0,09 | -1,00 | 3,16E-01 | 5,19E-01 |
| INTS11 | 791,38 | -0,06 | 0,06 | -1,00 | 3,16E-01 | 5,19E-01 |
| RNFT1 | 655,82 | 0,06 | 0,06 | 1,00 | 3,17E-01 | 5,19E-01 |
| C13orf46 | 272,38 | -0,13 | 0,13 | -1,00 | 3,17E-01 | 5,19E-01 |
| CTDSP1 | 1498,90 | -0,07 | 0,07 | -1,00 | 3,17E-01 | 5,19E-01 |
| VAV2 | 292,06 | -0,11 | 0,11 | -1,00 | 3,17E-01 | 5,20E-01 |
| TPD52L2 | 2868,83 | -0,06 | 0,06 | -1,00 | 3,17E-01 | 5,20E-01 |
| CLTC | 6572,37 | 0,04 | 0,04 | 1,00 | 3,17E-01 | 5,20E-01 |
| SLC25A1 | 239,43 | -0,06 | 0,06 | -1,00 | 3,18E-01 | 5,20E-01 |
| SMIM10L1 | 422,25 | 0,08 | 0,08 | 1,00 | 3,18E-01 | 5,20E-01 |
| VPS39 | 2003,80 | -0,06 | 0,06 | -1,00 | 3,18E-01 | 5,20E-01 |
| DVL2 | 179,05 | -0,09 | 0,09 | -1,00 | 3,18E-01 | 5,21E-01 |
| NDUFA6 | 400,70 | 0,09 | 0,09 | 1,00 | 3,18E-01 | 5,21E-01 |
| ELOVL1 | 838,67 | -0,04 | 0,04 | -1,00 | 3,18E-01 | 5,21E-01 |
| FAM110A | 368,20 | -0,08 | 0,08 | -1,00 | 3,18E-01 | 5,21E-01 |
| TMEM138 | 621,15 | -0,04 | 0,04 | -1,00 | 3,18E-01 | 5,21E-01 |
| VPS26C | 1980,57 | -0,02 | 0,02 | -1,00 | 3,18E-01 | 5,21E-01 |

| | | | | | | |
|---|---|---|---|---|---|---|
| EFNA4 | 109,10 | -0,05 | 0,05 | -1,00 | 3,18E-01 | 5,21E-01 |
| USP34 | 11300,99 | 0,05 | 0,05 | 1,00 | 3,18E-01 | 5,21E-01 |
| GATC | 604,07 | 0,04 | 0,04 | 1,00 | 3,18E-01 | 5,21E-01 |
| AKAP8L | 1359,89 | -0,05 | 0,05 | -1,00 | 3,19E-01 | 5,22E-01 |
| N4BP1 | 3290,35 | 0,05 | 0,05 | 1,00 | 3,19E-01 | 5,22E-01 |
| HIVEP3 | 364,29 | -0,12 | 0,12 | -1,00 | 3,19E-01 | 5,22E-01 |
| BRD2 | 3584,87 | -0,04 | 0,04 | -1,00 | 3,19E-01 | 5,22E-01 |
| PLOD1 | 585,94 | -0,07 | 0,08 | -1,00 | 3,19E-01 | 5,22E-01 |
| SUMO4 | 356,37 | 0,08 | 0,08 | 1,00 | 3,19E-01 | 5,22E-01 |
| XCL2 | 115,01 | 0,15 | 0,15 | 1,00 | 3,19E-01 | 5,22E-01 |
| KIAA0319L | 2420,69 | -0,06 | 0,06 | -1,00 | 3,19E-01 | 5,22E-01 |
| MOB1B | 1445,84 | 0,07 | 0,07 | 1,00 | 3,19E-01 | 5,22E-01 |
| BIRC2 | 4650,55 | 0,09 | 0,09 | 1,00 | 3,19E-01 | 5,22E-01 |
| RNF11 | 7128,18 | -0,12 | 0,12 | -1,00 | 3,19E-01 | 5,22E-01 |
| APH1B | 1181,70 | 0,04 | 0,04 | 1,00 | 3,19E-01 | 5,22E-01 |
| KCNA3 | 1091,66 | -0,06 | 0,06 | -1,00 | 3,19E-01 | 5,22E-01 |
| MAPKAP1 | 1953,24 | 0,03 | 0,03 | 1,00 | 3,20E-01 | 5,22E-01 |
| RSKR | 221,79 | -0,09 | 0,09 | -1,00 | 3,20E-01 | 5,22E-01 |
| EFCAB5 | 73,86 | -0,13 | 0,13 | -0,99 | 3,20E-01 | 5,22E-01 |
| FBXO42 | 1399,39 | -0,03 | 0,03 | -0,99 | 3,20E-01 | 5,22E-01 |
| APOBEC3D | 60,07 | -0,08 | 0,08 | -0,99 | 3,20E-01 | 5,22E-01 |
| C2orf76 | 154,02 | 0,06 | 0,06 | 0,99 | 3,20E-01 | 5,22E-01 |
| MICAL2 | 7402,25 | -0,10 | 0,10 | -0,99 | 3,20E-01 | 5,22E-01 |
| AKR1C3 | 177,78 | 0,12 | 0,12 | 0,99 | 3,20E-01 | 5,22E-01 |
| NUP98 | 6260,06 | -0,05 | 0,05 | -0,99 | 3,20E-01 | 5,23E-01 |
| SAAL1 | 305,87 | 0,05 | 0,05 | 0,99 | 3,20E-01 | 5,23E-01 |
| DDX47 | 175,95 | 0,04 | 0,04 | 0,99 | 3,20E-01 | 5,23E-01 |
| TSC22D2 | 1051,79 | 0,04 | 0,04 | 0,99 | 3,20E-01 | 5,23E-01 |
| COTL1 | 4841,61 | -0,09 | 0,09 | -0,99 | 3,20E-01 | 5,23E-01 |
| SBF1 | 1566,92 | -0,11 | 0,12 | -0,99 | 3,21E-01 | 5,23E-01 |
| LILRB2 | 1913,06 | -0,09 | 0,09 | -0,99 | 3,21E-01 | 5,23E-01 |
| NXF1 | 1686,08 | -0,05 | 0,05 | -0,99 | 3,21E-01 | 5,23E-01 |
| UBN2 | 2682,06 | 0,04 | 0,04 | 0,99 | 3,21E-01 | 5,24E-01 |
| RAPGEF6 | 1760,00 | 0,06 | 0,06 | 0,99 | 3,21E-01 | 5,24E-01 |
| ABCD4 | 587,47 | -0,06 | 0,06 | -0,99 | 3,21E-01 | 5,24E-01 |
| SLBP | 1071,89 | 0,05 | 0,05 | 0,99 | 3,22E-01 | 5,24E-01 |
| CREM | 393,10 | 0,04 | 0,04 | 0,99 | 3,21E-01 | 5,24E-01 |
| ORMDL3 | 3610,51 | -0,07 | 0,07 | -0,99 | 3,22E-01 | 5,24E-01 |
| TSGA10 | 403,50 | 0,08 | 0,08 | 0,99 | 3,22E-01 | 5,24E-01 |
| OR52K2 | 100,37 | -0,15 | 0,15 | -0,99 | 3,22E-01 | 5,24E-01 |
| UBQLN4 | 171,82 | -0,08 | 0,08 | -0,99 | 3,22E-01 | 5,24E-01 |
| SRRT | 492,31 | -0,09 | 0,09 | -0,99 | 3,22E-01 | 5,24E-01 |
| RAI1 | 345,76 | -0,11 | 0,11 | -0,99 | 3,22E-01 | 5,25E-01 |
| RBM14 | 1678,38 | -0,06 | 0,06 | -0,99 | 3,22E-01 | 5,25E-01 |
| PABIR3 | 314,15 | -0,05 | 0,05 | -0,99 | 3,22E-01 | 5,25E-01 |
| WDFY3 | 10233,18 | -0,08 | 0,08 | -0,99 | 3,23E-01 | 5,25E-01 |

| | | | | | | |
|---|---|---|---|---|---|---|
| BTBD8 | 303,40 | 0,07 | 0,07 | 0,99 | 3,23E-01 | 5,25E-01 |
| HPSE | 856,00 | -0,07 | 0,07 | -0,99 | 3,23E-01 | 5,25E-01 |
| PIP5K1A | 1212,23 | -0,03 | 0,03 | -0,99 | 3,23E-01 | 5,25E-01 |
| XRN1 | 7630,01 | 0,06 | 0,07 | 0,99 | 3,23E-01 | 5,25E-01 |
| TNRC6C | 1789,73 | -0,06 | 0,06 | -0,99 | 3,23E-01 | 5,26E-01 |
| CNPY4 | 388,93 | 0,04 | 0,04 | 0,99 | 3,23E-01 | 5,26E-01 |
| HNRNPK | 14756,09 | 0,05 | 0,05 | 0,99 | 3,24E-01 | 5,26E-01 |
| QSOX2 | 480,09 | -0,07 | 0,07 | -0,99 | 3,24E-01 | 5,26E-01 |
| PJVK | 47,43 | -0,10 | 0,10 | -0,99 | 3,24E-01 | 5,26E-01 |
| LATS1 | 1687,69 | 0,03 | 0,04 | 0,99 | 3,24E-01 | 5,26E-01 |
| GMEB1 | 702,82 | 0,02 | 0,02 | 0,99 | 3,24E-01 | 5,26E-01 |
| RABGEF1 | 241,80 | -0,03 | 0,03 | -0,99 | 3,24E-01 | 5,26E-01 |
| ZEB2 | 16471,46 | 0,05 | 0,05 | 0,99 | 3,24E-01 | 5,26E-01 |
| IFT172 | 370,03 | -0,06 | 0,07 | -0,99 | 3,24E-01 | 5,26E-01 |
| BLTP3A | 776,43 | -0,06 | 0,06 | -0,99 | 3,24E-01 | 5,27E-01 |
| PCYOX1 | 636,54 | 0,04 | 0,04 | 0,99 | 3,24E-01 | 5,27E-01 |
| DACH1 | 161,85 | -0,09 | 0,09 | -0,99 | 3,24E-01 | 5,27E-01 |
| RELT | 3456,17 | -0,07 | 0,08 | -0,99 | 3,25E-01 | 5,27E-01 |
| TUBB | 3090,17 | -0,04 | 0,04 | -0,99 | 3,25E-01 | 5,27E-01 |
| ZFP41 | 128,75 | -0,10 | 0,10 | -0,98 | 3,25E-01 | 5,27E-01 |
| ZBTB25 | 2407,21 | 0,05 | 0,05 | 0,98 | 3,25E-01 | 5,27E-01 |
| SLC25A4 | 118,77 | -0,07 | 0,08 | -0,98 | 3,25E-01 | 5,27E-01 |
| GLUL | 27647,70 | -0,08 | 0,08 | -0,98 | 3,25E-01 | 5,27E-01 |
| ANKRD55 | 161,60 | -0,11 | 0,11 | -0,98 | 3,25E-01 | 5,27E-01 |
| BMERB1 | 47,42 | -0,10 | 0,11 | -0,98 | 3,25E-01 | 5,27E-01 |
| GOT1 | 187,17 | -0,05 | 0,05 | -0,98 | 3,25E-01 | 5,28E-01 |
| DDX23 | 2915,90 | 0,05 | 0,05 | 0,98 | 3,25E-01 | 5,28E-01 |
| MVB12B | 221,78 | -0,11 | 0,11 | -0,98 | 3,26E-01 | 5,28E-01 |
| COMMD7 | 97,45 | -0,05 | 0,06 | -0,98 | 3,26E-01 | 5,28E-01 |
| FECH | 5685,39 | -0,13 | 0,13 | -0,98 | 3,26E-01 | 5,28E-01 |
| RHOT1 | 2226,82 | 0,06 | 0,06 | 0,98 | 3,26E-01 | 5,28E-01 |
| MTRES1 | 195,64 | 0,04 | 0,05 | 0,98 | 3,26E-01 | 5,28E-01 |
| VPS53 | 1765,54 | -0,06 | 0,06 | -0,98 | 3,26E-01 | 5,28E-01 |
| NFYA | 1761,23 | 0,03 | 0,03 | 0,98 | 3,26E-01 | 5,28E-01 |
| RIPOR1 | 779,04 | -0,08 | 0,08 | -0,98 | 3,27E-01 | 5,29E-01 |
| RPS9 | 13014,46 | 0,08 | 0,08 | 0,98 | 3,27E-01 | 5,29E-01 |
| UPK3A | 39,22 | -0,10 | 0,11 | -0,98 | 3,27E-01 | 5,29E-01 |
| NDUFS6 | 464,07 | 0,05 | 0,05 | 0,98 | 3,27E-01 | 5,29E-01 |
| VKORC1 | 156,18 | -0,04 | 0,05 | -0,98 | 3,27E-01 | 5,29E-01 |
| DTNBP1 | 439,84 | -0,05 | 0,05 | -0,98 | 3,27E-01 | 5,29E-01 |
| PHF8 | 1521,75 | -0,04 | 0,05 | -0,98 | 3,27E-01 | 5,29E-01 |
| PRKAR1A | 10304,71 | 0,07 | 0,07 | 0,98 | 3,27E-01 | 5,29E-01 |
| ZNF57 | 64,24 | 0,06 | 0,07 | 0,98 | 3,27E-01 | 5,29E-01 |
| LYN | 10045,20 | -0,06 | 0,07 | -0,98 | 3,27E-01 | 5,29E-01 |
| TOP3A | 1466,81 | -0,05 | 0,05 | -0,98 | 3,27E-01 | 5,29E-01 |
| ACD | 118,23 | -0,07 | 0,08 | -0,98 | 3,27E-01 | 5,29E-01 |

| | | | | | | |
|---|---|---|---|---|---|---|
| RPA2 | 1266,47 | 0,05 | 0,05 | 0,98 | 3,27E-01 | 5,29E-01 |
| ITFG2 | 1300,34 | 0,04 | 0,04 | 0,98 | 3,27E-01 | 5,30E-01 |
| IL7R | 18957,47 | 0,07 | 0,07 | 0,98 | 3,28E-01 | 5,30E-01 |
| AKNA | 7677,05 | -0,10 | 0,11 | -0,98 | 3,28E-01 | 5,31E-01 |
| CRLF3 | 4825,00 | 0,05 | 0,05 | 0,98 | 3,29E-01 | 5,31E-01 |
| TMEM115 | 313,91 | -0,05 | 0,05 | -0,98 | 3,29E-01 | 5,31E-01 |
| PPFIBP2 | 769,66 | 0,06 | 0,07 | 0,98 | 3,29E-01 | 5,31E-01 |
| DHX40 | 1392,45 | 0,06 | 0,06 | 0,98 | 3,29E-01 | 5,32E-01 |
| CA5B | 1252,07 | 0,04 | 0,04 | 0,98 | 3,29E-01 | 5,32E-01 |
| ITGB1BP1 | 1038,53 | 0,03 | 0,03 | 0,98 | 3,29E-01 | 5,32E-01 |
| ENSG000002 | 20,26 | 0,10 | 0,10 | 0,98 | 3,30E-01 | 5,32E-01 |
| DPH2 | 128,93 | -0,07 | 0,07 | -0,97 | 3,30E-01 | 5,32E-01 |
| MED13 | 6981,38 | 0,04 | 0,04 | 0,97 | 3,30E-01 | 5,32E-01 |
| ACSS1 | 1455,95 | -0,05 | 0,05 | -0,97 | 3,30E-01 | 5,32E-01 |
| GPR82 | 90,89 | 0,14 | 0,14 | 0,97 | 3,30E-01 | 5,32E-01 |
| ITGA5 | 3914,01 | -0,06 | 0,07 | -0,97 | 3,30E-01 | 5,32E-01 |
| PDE4B | 3516,39 | 0,05 | 0,05 | 0,97 | 3,30E-01 | 5,32E-01 |
| ECHS1 | 480,97 | 0,04 | 0,04 | 0,97 | 3,30E-01 | 5,32E-01 |
| KGD4 | 340,78 | 0,06 | 0,06 | 0,97 | 3,30E-01 | 5,32E-01 |
| GPX3 | 25,59 | -0,16 | 0,17 | -0,97 | 3,30E-01 | 5,32E-01 |
| AUP1 | 1274,41 | -0,03 | 0,03 | -0,97 | 3,30E-01 | 5,32E-01 |
| PPDPF | 229,47 | -0,15 | 0,15 | -0,97 | 3,30E-01 | 5,32E-01 |
| STXBP4 | 506,32 | 0,06 | 0,06 | 0,97 | 3,30E-01 | 5,32E-01 |
| RABIF | 430,03 | -0,03 | 0,03 | -0,97 | 3,30E-01 | 5,32E-01 |
| JOSD1 | 1136,44 | 0,03 | 0,03 | 0,97 | 3,30E-01 | 5,32E-01 |
| ZNF48 | 145,20 | -0,06 | 0,06 | -0,97 | 3,30E-01 | 5,32E-01 |
| MAML2 | 1445,84 | -0,04 | 0,04 | -0,97 | 3,30E-01 | 5,32E-01 |
| NBPF3 | 390,52 | 0,08 | 0,08 | 0,97 | 3,31E-01 | 5,32E-01 |
| AK9 | 488,20 | 0,06 | 0,06 | 0,97 | 3,31E-01 | 5,32E-01 |
| GABBR1 | 651,44 | -0,11 | 0,11 | -0,97 | 3,31E-01 | 5,32E-01 |
| SYCP2L | 45,51 | -0,24 | 0,24 | -0,97 | 3,31E-01 | 5,32E-01 |
| VPS26A | 814,43 | 0,06 | 0,06 | 0,97 | 3,30E-01 | 5,32E-01 |
| CYC1 | 726,19 | 0,04 | 0,04 | 0,97 | 3,31E-01 | 5,32E-01 |
| POR | 619,28 | -0,07 | 0,07 | -0,97 | 3,31E-01 | 5,32E-01 |
| GRIPAP1 | 982,80 | -0,07 | 0,07 | -0,97 | 3,31E-01 | 5,32E-01 |
| TEDC1 | 313,49 | -0,07 | 0,07 | -0,97 | 3,31E-01 | 5,32E-01 |
| HMCES | 853,24 | 0,04 | 0,04 | 0,97 | 3,31E-01 | 5,32E-01 |
| H2BC12 | 2190,79 | -0,06 | 0,06 | -0,97 | 3,31E-01 | 5,33E-01 |
| RWDD2B | 86,35 | 0,07 | 0,07 | 0,97 | 3,31E-01 | 5,33E-01 |
| TDRD9 | 244,89 | -0,13 | 0,14 | -0,97 | 3,31E-01 | 5,33E-01 |
| MAN1C1 | 379,86 | -0,11 | 0,11 | -0,97 | 3,31E-01 | 5,33E-01 |
| DPP7 | 739,42 | 0,06 | 0,06 | 0,97 | 3,32E-01 | 5,33E-01 |
| SNX33 | 175,79 | -0,09 | 0,09 | -0,97 | 3,32E-01 | 5,33E-01 |
| DNAJC1 | 661,32 | 0,05 | 0,05 | 0,97 | 3,31E-01 | 5,33E-01 |
| PPM1M | 1735,80 | -0,04 | 0,04 | -0,97 | 3,31E-01 | 5,33E-01 |
| DPEP2 | 3040,49 | -0,08 | 0,09 | -0,97 | 3,32E-01 | 5,34E-01 |

| | | | | | | |
|---|---|---|---|---|---|---|
| ECT2 | 290,73 | 0,08 | 0,09 | 0,97 | 3,32E-01 | 5,34E-01 |
| TRIM65 | 396,58 | -0,06 | 0,06 | -0,97 | 3,32E-01 | 5,34E-01 |
| LAPTM5 | 7655,26 | -0,07 | 0,07 | -0,97 | 3,32E-01 | 5,34E-01 |
| MAP4K1 | 447,41 | -0,10 | 0,11 | -0,97 | 3,32E-01 | 5,34E-01 |
| ZNF30 | 107,53 | 0,07 | 0,07 | 0,97 | 3,33E-01 | 5,34E-01 |
| MEF2D | 1330,53 | -0,08 | 0,08 | -0,97 | 3,33E-01 | 5,34E-01 |
| BTNL9 | 48,61 | -0,22 | 0,23 | -0,97 | 3,33E-01 | 5,34E-01 |
| DDX60 | 2548,74 | 0,11 | 0,12 | 0,97 | 3,33E-01 | 5,34E-01 |
| PISD | 2304,46 | -0,08 | 0,09 | -0,97 | 3,33E-01 | 5,34E-01 |
| TMEM9 | 250,26 | -0,06 | 0,06 | -0,97 | 3,33E-01 | 5,35E-01 |
| POM121 | 723,84 | -0,07 | 0,07 | -0,97 | 3,33E-01 | 5,35E-01 |
| YWHAB | 7696,90 | 0,04 | 0,04 | 0,97 | 3,33E-01 | 5,35E-01 |
| PTPRA | 1604,95 | -0,05 | 0,06 | -0,97 | 3,33E-01 | 5,35E-01 |
| MPC2 | 916,63 | -0,06 | 0,06 | -0,97 | 3,33E-01 | 5,35E-01 |
| GALNT2 | 822,64 | -0,05 | 0,05 | -0,97 | 3,33E-01 | 5,35E-01 |
| TTC21A | 185,62 | -0,10 | 0,10 | -0,97 | 3,33E-01 | 5,35E-01 |
| IRF7 | 440,65 | -0,10 | 0,11 | -0,97 | 3,34E-01 | 5,35E-01 |
| UTRN | 20708,64 | 0,05 | 0,06 | 0,97 | 3,34E-01 | 5,35E-01 |
| LDB1 | 1216,92 | -0,05 | 0,05 | -0,97 | 3,34E-01 | 5,36E-01 |
| CPSF4 | 397,60 | -0,03 | 0,04 | -0,97 | 3,34E-01 | 5,36E-01 |
| CLK2 | 899,53 | -0,04 | 0,04 | -0,97 | 3,34E-01 | 5,36E-01 |
| TMEM260 | 2139,51 | 0,05 | 0,05 | 0,97 | 3,34E-01 | 5,36E-01 |
| FBXO31 | 348,11 | -0,07 | 0,08 | -0,97 | 3,34E-01 | 5,36E-01 |
| URB2 | 471,33 | 0,04 | 0,04 | 0,97 | 3,34E-01 | 5,36E-01 |
| MSTO1 | 147,90 | -0,06 | 0,06 | -0,97 | 3,34E-01 | 5,36E-01 |
| CD6 | 1454,88 | -0,09 | 0,09 | -0,96 | 3,35E-01 | 5,36E-01 |
| CENPI | 92,73 | -0,09 | 0,09 | -0,96 | 3,35E-01 | 5,36E-01 |
| POLD2 | 129,12 | -0,08 | 0,08 | -0,96 | 3,35E-01 | 5,36E-01 |
| MUS81 | 632,39 | -0,04 | 0,04 | -0,96 | 3,35E-01 | 5,36E-01 |
| CGAS | 747,98 | -0,04 | 0,04 | -0,96 | 3,35E-01 | 5,36E-01 |
| SCYL2 | 2641,98 | 0,05 | 0,05 | 0,96 | 3,35E-01 | 5,36E-01 |
| SPICE1-CFAI | 24,15 | -0,09 | 0,10 | -0,96 | 3,35E-01 | 5,36E-01 |
| SEC14L1 | 9387,16 | -0,09 | 0,09 | -0,96 | 3,35E-01 | 5,36E-01 |
| COLQ | 232,13 | -0,09 | 0,09 | -0,96 | 3,35E-01 | 5,36E-01 |
| SEMA4F | 320,41 | -0,06 | 0,06 | -0,96 | 3,35E-01 | 5,36E-01 |
| RNF44 | 865,70 | -0,09 | 0,10 | -0,96 | 3,35E-01 | 5,36E-01 |
| USP6NL | 1101,74 | 0,05 | 0,05 | 0,96 | 3,35E-01 | 5,36E-01 |
| EIF3F | 2697,49 | 0,05 | 0,05 | 0,96 | 3,35E-01 | 5,36E-01 |
| PSMC1 | 1050,71 | 0,04 | 0,04 | 0,96 | 3,35E-01 | 5,36E-01 |
| ENSG000002 | 836,99 | 0,05 | 0,05 | 0,96 | 3,35E-01 | 5,36E-01 |
| SMCHD1 | 53641,84 | 0,07 | 0,08 | 0,96 | 3,35E-01 | 5,36E-01 |
| HTT | 3023,90 | -0,06 | 0,06 | -0,96 | 3,36E-01 | 5,37E-01 |
| PSPC1 | 3537,62 | 0,04 | 0,04 | 0,96 | 3,36E-01 | 5,37E-01 |
| CXCL16 | 510,54 | -0,09 | 0,10 | -0,96 | 3,36E-01 | 5,37E-01 |
| BCL2L2 | 264,69 | 0,05 | 0,05 | 0,96 | 3,36E-01 | 5,37E-01 |
| TIMM23 | 551,81 | 0,04 | 0,05 | 0,96 | 3,36E-01 | 5,37E-01 |

| | | | | | | |
|---|---|---|---|---|---|---|
| FBXO9 | 2848,90 | -0,07 | 0,07 | -0,96 | 3,36E-01 | 5,37E-01 |
| BTNL3 | 68,43 | -0,39 | 0,41 | -0,96 | 3,36E-01 | 5,37E-01 |
| UCKL1 | 271,68 | -0,07 | 0,07 | -0,96 | 3,36E-01 | 5,37E-01 |
| TAF6L | 236,50 | 0,04 | 0,04 | 0,96 | 3,36E-01 | 5,37E-01 |
| ECE1 | 2776,74 | -0,09 | 0,09 | -0,96 | 3,36E-01 | 5,37E-01 |
| INPP5D | 4486,82 | -0,08 | 0,08 | -0,96 | 3,36E-01 | 5,37E-01 |
| UBE2K | 1921,87 | 0,06 | 0,06 | 0,96 | 3,36E-01 | 5,37E-01 |
| YY1AP1 | 1903,94 | -0,05 | 0,05 | -0,96 | 3,36E-01 | 5,37E-01 |
| ALDH2 | 472,50 | 0,08 | 0,08 | 0,96 | 3,36E-01 | 5,37E-01 |
| SDC2 | 84,15 | -0,13 | 0,13 | -0,96 | 3,37E-01 | 5,37E-01 |
| HMMR | 34,74 | -0,16 | 0,17 | -0,96 | 3,37E-01 | 5,37E-01 |
| ZNF407 | 3186,06 | 0,05 | 0,05 | 0,96 | 3,37E-01 | 5,37E-01 |
| MAL | 317,67 | -0,08 | 0,08 | -0,96 | 3,37E-01 | 5,37E-01 |
| SFI1 | 2092,26 | -0,07 | 0,07 | -0,96 | 3,37E-01 | 5,37E-01 |
| DHRS12 | 456,39 | -0,08 | 0,08 | -0,96 | 3,37E-01 | 5,38E-01 |
| CENPF | 177,90 | -0,11 | 0,11 | -0,96 | 3,37E-01 | 5,38E-01 |
| PHTF1 | 1053,23 | -0,04 | 0,04 | -0,96 | 3,37E-01 | 5,38E-01 |
| RRH | 28,52 | -0,09 | 0,09 | -0,96 | 3,37E-01 | 5,38E-01 |
| NFYC | 1576,47 | -0,05 | 0,05 | -0,96 | 3,37E-01 | 5,38E-01 |
| PM20D2 | 603,71 | 0,05 | 0,05 | 0,96 | 3,38E-01 | 5,38E-01 |
| SAMD9L | 5772,99 | 0,11 | 0,12 | 0,96 | 3,38E-01 | 5,38E-01 |
| SYCP2 | 400,59 | 0,09 | 0,10 | 0,96 | 3,38E-01 | 5,39E-01 |
| FBRS | 1500,61 | -0,08 | 0,08 | -0,96 | 3,38E-01 | 5,39E-01 |
| UNC45A | 788,60 | -0,04 | 0,05 | -0,96 | 3,38E-01 | 5,39E-01 |
| ARL6IP5 | 4177,67 | 0,06 | 0,06 | 0,96 | 3,38E-01 | 5,39E-01 |
| GPR63 | 46,00 | -0,13 | 0,14 | -0,96 | 3,38E-01 | 5,39E-01 |
| AGFG1 | 4427,22 | 0,05 | 0,06 | 0,96 | 3,38E-01 | 5,39E-01 |
| AFF1 | 5252,48 | -0,05 | 0,05 | -0,96 | 3,38E-01 | 5,39E-01 |
| ATRAID | 893,15 | 0,04 | 0,04 | 0,96 | 3,38E-01 | 5,39E-01 |
| CYBA | 9119,23 | -0,05 | 0,06 | -0,96 | 3,38E-01 | 5,39E-01 |
| IMMP2L | 514,32 | 0,06 | 0,06 | 0,96 | 3,39E-01 | 5,39E-01 |
| JKAMP | 1313,94 | 0,06 | 0,06 | 0,96 | 3,39E-01 | 5,39E-01 |
| ARL11 | 1676,60 | -0,06 | 0,06 | -0,96 | 3,39E-01 | 5,39E-01 |
| GSDMB | 1218,24 | -0,07 | 0,07 | -0,96 | 3,39E-01 | 5,39E-01 |
| GATA2 | 243,54 | -0,19 | 0,20 | -0,96 | 3,39E-01 | 5,39E-01 |
| METRNL | 1358,23 | -0,06 | 0,06 | -0,96 | 3,39E-01 | 5,39E-01 |
| GDF7 | 152,53 | -0,17 | 0,18 | -0,96 | 3,39E-01 | 5,39E-01 |
| TSHZ1 | 521,48 | -0,05 | 0,06 | -0,96 | 3,39E-01 | 5,39E-01 |
| ZBTB7A | 642,98 | -0,10 | 0,10 | -0,96 | 3,39E-01 | 5,39E-01 |
| MAPK7 | 178,92 | -0,08 | 0,09 | -0,96 | 3,39E-01 | 5,39E-01 |
| TM2D1 | 1386,68 | 0,03 | 0,04 | 0,96 | 3,39E-01 | 5,39E-01 |
| MAP3K8 | 1591,87 | 0,04 | 0,04 | 0,96 | 3,39E-01 | 5,39E-01 |
| DNM2 | 4151,91 | -0,07 | 0,07 | -0,96 | 3,39E-01 | 5,39E-01 |
| TWF2 | 1617,84 | -0,05 | 0,05 | -0,96 | 3,39E-01 | 5,39E-01 |
| PSMB4 | 2366,48 | 0,05 | 0,05 | 0,96 | 3,40E-01 | 5,39E-01 |
| AHSA1 | 983,22 | 0,04 | 0,04 | 0,95 | 3,40E-01 | 5,40E-01 |

| | | | | | | |
|---|---|---|---|---|---|---|
| PIGW | 142,55 | 0,04 | 0,05 | 0,95 | 3,40E-01 | 5,40E-01 |
| MIEF1 | 558,03 | -0,05 | 0,05 | -0,95 | 3,40E-01 | 5,40E-01 |
| ENPP3 | 91,16 | -0,14 | 0,15 | -0,95 | 3,40E-01 | 5,40E-01 |
| PDCD11 | 1139,96 | 0,04 | 0,04 | 0,95 | 3,40E-01 | 5,40E-01 |
| GNA15 | 368,34 | -0,07 | 0,07 | -0,95 | 3,40E-01 | 5,40E-01 |
| NCKAP5L | 261,25 | -0,11 | 0,11 | -0,95 | 3,40E-01 | 5,40E-01 |
| CLIC5 | 184,07 | -0,09 | 0,09 | -0,95 | 3,40E-01 | 5,40E-01 |
| TOR3A | 675,78 | -0,03 | 0,04 | -0,95 | 3,40E-01 | 5,40E-01 |
| ZRANB3 | 209,60 | 0,06 | 0,06 | 0,95 | 3,40E-01 | 5,40E-01 |
| SNN | 3086,65 | -0,07 | 0,07 | -0,95 | 3,40E-01 | 5,40E-01 |
| B4GALT4 | 781,95 | 0,05 | 0,05 | 0,95 | 3,40E-01 | 5,40E-01 |
| ARHGEF12 | 3762,87 | -0,10 | 0,10 | -0,95 | 3,40E-01 | 5,40E-01 |
| STX16-NPEPL | 125,49 | -0,08 | 0,08 | -0,95 | 3,40E-01 | 5,40E-01 |
| YIPF4 | 5647,59 | 0,04 | 0,04 | 0,95 | 3,41E-01 | 5,41E-01 |
| CEP85 | 604,14 | -0,03 | 0,04 | -0,95 | 3,41E-01 | 5,41E-01 |
| XPO5 | 639,77 | -0,05 | 0,05 | -0,95 | 3,41E-01 | 5,41E-01 |
| PER2 | 259,46 | -0,08 | 0,08 | -0,95 | 3,41E-01 | 5,41E-01 |
| HERC6 | 930,73 | 0,08 | 0,08 | 0,95 | 3,41E-01 | 5,41E-01 |
| PDCD2L | 49,34 | 0,07 | 0,07 | 0,95 | 3,41E-01 | 5,41E-01 |
| SLC45A3 | 153,50 | -0,16 | 0,16 | -0,95 | 3,42E-01 | 5,41E-01 |
| NOL6 | 332,93 | -0,08 | 0,08 | -0,95 | 3,42E-01 | 5,41E-01 |
| KCNQ1 | 679,21 | -0,08 | 0,08 | -0,95 | 3,42E-01 | 5,41E-01 |
| HARS2 | 1024,92 | -0,03 | 0,03 | -0,95 | 3,42E-01 | 5,41E-01 |
| CNNM3 | 632,71 | -0,07 | 0,07 | -0,95 | 3,42E-01 | 5,41E-01 |
| HELZ | 6509,14 | 0,04 | 0,04 | 0,95 | 3,42E-01 | 5,42E-01 |
| ECHDC3 | 175,15 | -0,10 | 0,11 | -0,95 | 3,42E-01 | 5,42E-01 |
| YLPM1 | 3058,54 | 0,04 | 0,04 | 0,95 | 3,43E-01 | 5,42E-01 |
| ATG101 | 536,60 | 0,04 | 0,04 | 0,95 | 3,42E-01 | 5,42E-01 |
| ALDH8A1 | 70,44 | -0,10 | 0,11 | -0,95 | 3,43E-01 | 5,42E-01 |
| SAPCD2 | 48,53 | -0,18 | 0,19 | -0,95 | 3,43E-01 | 5,42E-01 |
| TBCD | 1401,49 | -0,07 | 0,07 | -0,95 | 3,43E-01 | 5,42E-01 |
| FEM1C | 1683,63 | 0,06 | 0,07 | 0,95 | 3,43E-01 | 5,42E-01 |
| FOXP1 | 7617,24 | 0,05 | 0,05 | 0,95 | 3,43E-01 | 5,43E-01 |
| UBL3 | 2111,57 | 0,05 | 0,05 | 0,95 | 3,43E-01 | 5,43E-01 |
| ZBTB45 | 263,22 | -0,04 | 0,05 | -0,95 | 3,43E-01 | 5,43E-01 |
| TMX4 | 6150,03 | 0,05 | 0,06 | 0,95 | 3,43E-01 | 5,43E-01 |
| RRAGD | 982,02 | -0,06 | 0,07 | -0,95 | 3,43E-01 | 5,43E-01 |
| ZNF316 | 1571,82 | -0,06 | 0,06 | -0,95 | 3,43E-01 | 5,43E-01 |
| PAPOLA | 9236,36 | 0,05 | 0,06 | 0,95 | 3,44E-01 | 5,43E-01 |
| SLAIN1 | 597,10 | 0,05 | 0,05 | 0,95 | 3,44E-01 | 5,43E-01 |
| QSER1 | 1185,77 | 0,04 | 0,05 | 0,95 | 3,44E-01 | 5,43E-01 |
| MED15 | 1849,46 | -0,08 | 0,09 | -0,95 | 3,44E-01 | 5,43E-01 |
| LSR | 118,14 | -0,09 | 0,10 | -0,95 | 3,44E-01 | 5,43E-01 |
| TTC9C | 559,68 | 0,03 | 0,03 | 0,95 | 3,44E-01 | 5,43E-01 |
| DSC2 | 1323,18 | -0,11 | 0,12 | -0,95 | 3,44E-01 | 5,43E-01 |
| TCP11L2 | 9869,42 | -0,06 | 0,06 | -0,95 | 3,44E-01 | 5,43E-01 |

| | | | | | | |
|---|---|---|---|---|---|---|
| COQ10A | 176,32 | -0,04 | 0,05 | -0,95 | 3,44E-01 | 5,43E-01 |
| MBOAT2 | 2647,24 | -0,07 | 0,08 | -0,95 | 3,44E-01 | 5,43E-01 |
| GSPT1 | 10494,02 | -0,11 | 0,11 | -0,95 | 3,44E-01 | 5,43E-01 |
| CNTLN | 413,16 | 0,07 | 0,07 | 0,95 | 3,44E-01 | 5,43E-01 |
| KDELR1 | 156,44 | -0,09 | 0,10 | -0,95 | 3,44E-01 | 5,43E-01 |
| YPEL5 | 3336,17 | 0,07 | 0,08 | 0,95 | 3,44E-01 | 5,43E-01 |
| SEC24C | 2006,81 | -0,05 | 0,05 | -0,95 | 3,44E-01 | 5,43E-01 |
| NKG7 | 599,24 | -0,12 | 0,12 | -0,95 | 3,45E-01 | 5,43E-01 |
| KIAA0753 | 758,89 | -0,04 | 0,04 | -0,95 | 3,44E-01 | 5,43E-01 |
| RFLNB | 1716,07 | -0,09 | 0,09 | -0,94 | 3,45E-01 | 5,44E-01 |
| HECW2 | 482,78 | -0,09 | 0,09 | -0,94 | 3,45E-01 | 5,44E-01 |
| VSIR | 10300,95 | -0,07 | 0,08 | -0,94 | 3,45E-01 | 5,44E-01 |
| MDH2 | 487,35 | -0,05 | 0,05 | -0,94 | 3,45E-01 | 5,44E-01 |
| CHST10 | 91,08 | -0,11 | 0,11 | -0,94 | 3,45E-01 | 5,44E-01 |
| ZFP82 | 248,69 | 0,08 | 0,08 | 0,94 | 3,45E-01 | 5,44E-01 |
| GPAA1 | 386,57 | -0,05 | 0,06 | -0,94 | 3,45E-01 | 5,44E-01 |
| MRTO4 | 339,81 | 0,05 | 0,05 | 0,94 | 3,45E-01 | 5,44E-01 |
| CEBPE | 138,72 | -0,11 | 0,11 | -0,94 | 3,45E-01 | 5,44E-01 |
| ZNF333 | 1321,57 | -0,04 | 0,05 | -0,94 | 3,45E-01 | 5,44E-01 |
| PDXK | 1787,09 | -0,07 | 0,07 | -0,94 | 3,45E-01 | 5,44E-01 |
| POLR1H | 420,57 | 0,03 | 0,04 | 0,94 | 3,45E-01 | 5,44E-01 |
| AASS | 150,00 | -0,09 | 0,09 | -0,94 | 3,45E-01 | 5,44E-01 |
| ZNF281 | 2174,92 | 0,07 | 0,07 | 0,94 | 3,45E-01 | 5,44E-01 |
| ANKEF1 | 202,15 | 0,07 | 0,07 | 0,94 | 3,46E-01 | 5,44E-01 |
| HDAC5 | 918,32 | -0,08 | 0,09 | -0,94 | 3,46E-01 | 5,44E-01 |
| FLNA | 19031,67 | -0,10 | 0,10 | -0,94 | 3,46E-01 | 5,44E-01 |
| PSMB7 | 1110,93 | 0,04 | 0,05 | 0,94 | 3,46E-01 | 5,44E-01 |
| LRG1 | 1212,15 | -0,09 | 0,09 | -0,94 | 3,46E-01 | 5,44E-01 |
| CTBP1 | 3814,74 | 0,04 | 0,04 | 0,94 | 3,46E-01 | 5,44E-01 |
| PPM1F | 4290,76 | -0,08 | 0,08 | -0,94 | 3,46E-01 | 5,45E-01 |
| CD244 | 1609,58 | 0,06 | 0,06 | 0,94 | 3,46E-01 | 5,45E-01 |
| PHAF1 | 337,54 | -0,06 | 0,07 | -0,94 | 3,47E-01 | 5,45E-01 |
| OSBPL5 | 716,92 | -0,10 | 0,10 | -0,94 | 3,47E-01 | 5,45E-01 |
| RIPOR2 | 25541,02 | -0,05 | 0,05 | -0,94 | 3,47E-01 | 5,45E-01 |
| RPS19BP1 | 661,81 | -0,04 | 0,04 | -0,94 | 3,47E-01 | 5,45E-01 |
| HSP90AB1 | 2138,72 | 0,05 | 0,05 | 0,94 | 3,47E-01 | 5,46E-01 |
| GBP3 | 1082,35 | 0,07 | 0,08 | 0,94 | 3,47E-01 | 5,46E-01 |
| EXOSC10 | 1061,23 | 0,03 | 0,03 | 0,94 | 3,47E-01 | 5,46E-01 |
| MARF1 | 5529,20 | -0,07 | 0,07 | -0,94 | 3,47E-01 | 5,46E-01 |
| CNKSR2 | 557,16 | 0,08 | 0,08 | 0,94 | 3,47E-01 | 5,46E-01 |
| CXCR4 | 5959,26 | 0,06 | 0,06 | 0,94 | 3,47E-01 | 5,46E-01 |
| ZDHHC11B | 53,41 | -0,18 | 0,19 | -0,94 | 3,48E-01 | 5,46E-01 |
| KDM4C | 4230,58 | 0,04 | 0,05 | 0,94 | 3,48E-01 | 5,47E-01 |
| REPS1 | 1828,18 | 0,04 | 0,04 | 0,94 | 3,48E-01 | 5,47E-01 |
| PNMA3 | 211,66 | 0,12 | 0,13 | 0,94 | 3,48E-01 | 5,47E-01 |
| ZC3HAV1 | 6679,85 | -0,04 | 0,04 | -0,94 | 3,48E-01 | 5,47E-01 |

| | | | | | | |
|---|---|---|---|---|---|---|
| SNRK | 7734,50 | 0,05 | 0,05 | 0,94 | 3,49E-01 | 5,48E-01 |
| RMI1 | 350,62 | 0,06 | 0,07 | 0,94 | 3,49E-01 | 5,48E-01 |
| POMP | 686,42 | 0,06 | 0,07 | 0,94 | 3,49E-01 | 5,48E-01 |
| FLAD1 | 626,87 | 0,03 | 0,03 | 0,94 | 3,49E-01 | 5,48E-01 |
| LIMA1 | 260,51 | 0,06 | 0,07 | 0,94 | 3,49E-01 | 5,48E-01 |
| AZIN2 | 142,71 | -0,08 | 0,09 | -0,94 | 3,50E-01 | 5,48E-01 |
| PPM1D | 760,61 | -0,03 | 0,03 | -0,94 | 3,49E-01 | 5,48E-01 |
| B9D2 | 347,76 | -0,05 | 0,06 | -0,93 | 3,50E-01 | 5,49E-01 |
| SIGLEC7 | 310,14 | -0,07 | 0,07 | -0,93 | 3,50E-01 | 5,49E-01 |
| CACNA2D3 | 187,59 | 0,08 | 0,09 | 0,93 | 3,50E-01 | 5,49E-01 |
| CHCHD1 | 277,68 | 0,06 | 0,07 | 0,93 | 3,50E-01 | 5,49E-01 |
| FANCC | 293,09 | -0,09 | 0,09 | -0,93 | 3,50E-01 | 5,49E-01 |
| SLC35C2 | 1618,83 | -0,04 | 0,04 | -0,93 | 3,51E-01 | 5,49E-01 |
| CSF3R | 30733,40 | -0,09 | 0,09 | -0,93 | 3,51E-01 | 5,49E-01 |
| UTY | 4481,92 | -0,23 | 0,24 | -0,93 | 3,51E-01 | 5,49E-01 |
| IFFO1 | 680,34 | -0,07 | 0,07 | -0,93 | 3,51E-01 | 5,50E-01 |
| TIMELESS | 210,93 | -0,06 | 0,07 | -0,93 | 3,51E-01 | 5,50E-01 |
| MARCKS | 2555,39 | 0,08 | 0,08 | 0,93 | 3,51E-01 | 5,50E-01 |
| FMNL2 | 181,91 | -0,09 | 0,10 | -0,93 | 3,51E-01 | 5,50E-01 |
| RAB4A | 796,05 | 0,04 | 0,04 | 0,93 | 3,51E-01 | 5,50E-01 |
| OXSR1 | 2437,24 | 0,04 | 0,04 | 0,93 | 3,51E-01 | 5,50E-01 |
| ALDH3B1 | 533,01 | -0,08 | 0,08 | -0,93 | 3,51E-01 | 5,50E-01 |
| TBPL1 | 914,64 | 0,06 | 0,06 | 0,93 | 3,52E-01 | 5,50E-01 |
| SH3BP5L | 2107,64 | -0,05 | 0,05 | -0,93 | 3,52E-01 | 5,50E-01 |
| SLC4A10 | 337,62 | 0,16 | 0,18 | 0,93 | 3,52E-01 | 5,50E-01 |
| ATOX1 | 544,61 | -0,04 | 0,04 | -0,93 | 3,52E-01 | 5,50E-01 |
| CALM1 | 8376,37 | 0,03 | 0,04 | 0,93 | 3,52E-01 | 5,51E-01 |
| ELMO2 | 1444,91 | -0,04 | 0,04 | -0,93 | 3,52E-01 | 5,51E-01 |
| NXT1 | 126,02 | 0,05 | 0,05 | 0,93 | 3,52E-01 | 5,51E-01 |
| SPATA2 | 197,81 | -0,07 | 0,07 | -0,93 | 3,52E-01 | 5,51E-01 |
| GRB2 | 8182,82 | -0,05 | 0,05 | -0,93 | 3,52E-01 | 5,51E-01 |
| USP28 | 1275,39 | 0,06 | 0,06 | 0,93 | 3,52E-01 | 5,51E-01 |
| UNC50 | 1006,28 | 0,04 | 0,04 | 0,93 | 3,52E-01 | 5,51E-01 |
| TCN1 | 136,21 | -0,14 | 0,16 | -0,93 | 3,52E-01 | 5,51E-01 |
| LY86 | 508,60 | 0,07 | 0,07 | 0,93 | 3,52E-01 | 5,51E-01 |
| KBTBD2 | 3581,11 | 0,05 | 0,05 | 0,93 | 3,52E-01 | 5,51E-01 |
| BTN2A1 | 1406,71 | -0,06 | 0,06 | -0,93 | 3,52E-01 | 5,51E-01 |
| GALNT12 | 178,77 | -0,06 | 0,06 | -0,93 | 3,52E-01 | 5,51E-01 |
| TES | 3194,06 | 0,05 | 0,05 | 0,93 | 3,52E-01 | 5,51E-01 |
| OSBPL7 | 270,26 | -0,08 | 0,09 | -0,93 | 3,53E-01 | 5,51E-01 |
| FBXW8 | 347,05 | -0,06 | 0,06 | -0,93 | 3,53E-01 | 5,51E-01 |
| DHX16 | 1652,90 | 0,03 | 0,04 | 0,93 | 3,53E-01 | 5,51E-01 |
| GLE1 | 1264,50 | 0,03 | 0,03 | 0,93 | 3,53E-01 | 5,51E-01 |
| TALDO1 | 10156,92 | -0,07 | 0,08 | -0,93 | 3,53E-01 | 5,51E-01 |
| AGFG2 | 352,86 | -0,07 | 0,08 | -0,93 | 3,53E-01 | 5,52E-01 |
| TFEB | 973,69 | -0,08 | 0,08 | -0,93 | 3,54E-01 | 5,52E-01 |

| | | | | | | |
|---|---|---|---|---|---|---|
| CCDC141 | 787,00 | 0,08 | 0,09 | 0,93 | 3,54E-01 | 5,52E-01 |
| TIPIN | 86,98 | 0,07 | 0,07 | 0,93 | 3,54E-01 | 5,53E-01 |
| USP11 | 303,43 | -0,06 | 0,07 | -0,93 | 3,54E-01 | 5,53E-01 |
| VSIG4 | 49,61 | 0,11 | 0,12 | 0,93 | 3,55E-01 | 5,53E-01 |
| OTUD5 | 1075,45 | -0,07 | 0,07 | -0,93 | 3,55E-01 | 5,53E-01 |
| PSMD10 | 646,19 | 0,08 | 0,08 | 0,93 | 3,55E-01 | 5,53E-01 |
| NDUFB2 | 655,41 | 0,06 | 0,06 | 0,93 | 3,55E-01 | 5,53E-01 |
| GORASP2 | 529,10 | 0,03 | 0,03 | 0,92 | 3,55E-01 | 5,54E-01 |
| PHF19 | 731,15 | -0,05 | 0,06 | -0,92 | 3,55E-01 | 5,54E-01 |
| CHMP1A | 668,65 | -0,06 | 0,07 | -0,92 | 3,55E-01 | 5,54E-01 |
| BTBD19 | 84,13 | -0,13 | 0,14 | -0,92 | 3,55E-01 | 5,54E-01 |
| ERAP2 | 3309,07 | 0,08 | 0,09 | 0,92 | 3,55E-01 | 5,54E-01 |
| IRGQ | 524,58 | -0,05 | 0,05 | -0,92 | 3,56E-01 | 5,54E-01 |
| KRBOX4 | 373,32 | 0,03 | 0,03 | 0,92 | 3,56E-01 | 5,55E-01 |
| AARS2 | 581,78 | 0,04 | 0,04 | 0,92 | 3,56E-01 | 5,55E-01 |
| CDC25B | 915,62 | -0,10 | 0,11 | -0,92 | 3,56E-01 | 5,55E-01 |
| PRSS21 | 27,72 | -0,18 | 0,19 | -0,92 | 3,56E-01 | 5,55E-01 |
| TIMM22 | 365,84 | 0,03 | 0,03 | 0,92 | 3,56E-01 | 5,55E-01 |
| U2AF2 | 1358,07 | -0,05 | 0,06 | -0,92 | 3,56E-01 | 5,55E-01 |
| ZNF66 | 582,32 | 0,08 | 0,09 | 0,92 | 3,56E-01 | 5,55E-01 |
| PER1 | 176,85 | -0,14 | 0,15 | -0,92 | 3,56E-01 | 5,55E-01 |
| TAS2R60 | 47,08 | -0,14 | 0,15 | -0,92 | 3,56E-01 | 5,55E-01 |
| CCDC122 | 165,07 | 0,10 | 0,11 | 0,92 | 3,57E-01 | 5,55E-01 |
| GLB1L | 51,29 | -0,11 | 0,12 | -0,92 | 3,57E-01 | 5,55E-01 |
| ANKMY1 | 1148,72 | -0,06 | 0,06 | -0,92 | 3,57E-01 | 5,55E-01 |
| LRRC37A3 | 301,99 | -0,11 | 0,12 | -0,92 | 3,57E-01 | 5,55E-01 |
| AGAP9 | 136,21 | -0,11 | 0,12 | -0,92 | 3,57E-01 | 5,55E-01 |
| TERF2 | 555,76 | -0,04 | 0,04 | -0,92 | 3,57E-01 | 5,55E-01 |
| YPEL1 | 409,41 | -0,06 | 0,07 | -0,92 | 3,57E-01 | 5,55E-01 |
| UBE2Z | 1278,13 | -0,05 | 0,06 | -0,92 | 3,57E-01 | 5,55E-01 |
| MMP8 | 175,57 | -0,21 | 0,23 | -0,92 | 3,57E-01 | 5,55E-01 |
| NPC1 | 1599,86 | -0,05 | 0,06 | -0,92 | 3,57E-01 | 5,56E-01 |
| MSS51 | 265,67 | -0,05 | 0,05 | -0,92 | 3,57E-01 | 5,56E-01 |
| PTPN13 | 356,00 | -0,08 | 0,09 | -0,92 | 3,58E-01 | 5,56E-01 |
| SMURF1 | 1155,90 | -0,04 | 0,04 | -0,92 | 3,58E-01 | 5,56E-01 |
| ST3GAL3 | 470,46 | -0,08 | 0,09 | -0,92 | 3,58E-01 | 5,56E-01 |
| NPRL3 | 2421,03 | -0,20 | 0,21 | -0,92 | 3,58E-01 | 5,57E-01 |
| RHBDF2 | 1044,29 | -0,09 | 0,09 | -0,92 | 3,58E-01 | 5,57E-01 |
| TSR3 | 623,65 | -0,04 | 0,04 | -0,92 | 3,58E-01 | 5,57E-01 |
| SFXN5 | 932,13 | -0,06 | 0,06 | -0,92 | 3,58E-01 | 5,57E-01 |
| SLC25A45 | 402,89 | -0,10 | 0,10 | -0,92 | 3,58E-01 | 5,57E-01 |
| MRPS21 | 586,31 | 0,06 | 0,06 | 0,92 | 3,58E-01 | 5,57E-01 |
| PILRA | 523,80 | -0,11 | 0,12 | -0,92 | 3,58E-01 | 5,57E-01 |
| CIR1 | 1413,84 | 0,05 | 0,06 | 0,92 | 3,58E-01 | 5,57E-01 |
| SEMA5A | 97,99 | -0,15 | 0,16 | -0,92 | 3,59E-01 | 5,57E-01 |
| PIM3 | 1110,58 | -0,06 | 0,07 | -0,92 | 3,59E-01 | 5,57E-01 |

| | | | | | | |
|---|---|---|---|---|---|---|
| LEPR | 433,83 | -0,10 | 0,11 | -0,92 | 3,59E-01 | 5,57E-01 |
| CYTH4 | 11103,96 | -0,05 | 0,06 | -0,92 | 3,59E-01 | 5,57E-01 |
| CCDC134 | 131,16 | -0,08 | 0,09 | -0,92 | 3,59E-01 | 5,57E-01 |
| PPM1L | 916,39 | -0,05 | 0,06 | -0,92 | 3,59E-01 | 5,57E-01 |
| OS9 | 2056,02 | -0,06 | 0,06 | -0,92 | 3,59E-01 | 5,57E-01 |
| FBXL6 | 154,43 | -0,08 | 0,08 | -0,92 | 3,59E-01 | 5,57E-01 |
| ADGRG3 | 1263,30 | -0,11 | 0,12 | -0,92 | 3,59E-01 | 5,57E-01 |
| VDAC3 | 1369,57 | 0,05 | 0,06 | 0,92 | 3,59E-01 | 5,57E-01 |
| DCAKD | 304,62 | -0,04 | 0,05 | -0,92 | 3,59E-01 | 5,57E-01 |
| B3GALNT2 | 342,46 | 0,06 | 0,06 | 0,92 | 3,59E-01 | 5,57E-01 |
| NUMA1 | 5770,40 | 0,05 | 0,05 | 0,92 | 3,59E-01 | 5,57E-01 |
| LETMD1 | 1097,71 | 0,03 | 0,03 | 0,92 | 3,60E-01 | 5,57E-01 |
| STRN4 | 1101,94 | -0,07 | 0,08 | -0,92 | 3,60E-01 | 5,58E-01 |
| IQGAP2 | 6667,44 | 0,05 | 0,05 | 0,92 | 3,60E-01 | 5,58E-01 |
| CCDC97 | 365,23 | -0,08 | 0,09 | -0,91 | 3,60E-01 | 5,58E-01 |
| ENSG000002 | 36,77 | -0,06 | 0,07 | -0,91 | 3,60E-01 | 5,58E-01 |
| CALHM6 | 269,19 | -0,12 | 0,13 | -0,91 | 3,60E-01 | 5,58E-01 |
| RANBP1 | 533,26 | 0,04 | 0,04 | 0,91 | 3,60E-01 | 5,58E-01 |
| IFNG | 40,05 | 0,16 | 0,17 | 0,91 | 3,60E-01 | 5,58E-01 |
| CYTH2 | 842,43 | -0,05 | 0,06 | -0,91 | 3,60E-01 | 5,58E-01 |
| RHEBL1 | 42,99 | -0,08 | 0,09 | -0,91 | 3,61E-01 | 5,58E-01 |
| CCL5 | 4369,85 | -0,08 | 0,09 | -0,91 | 3,61E-01 | 5,58E-01 |
| GHDC | 155,54 | -0,08 | 0,09 | -0,91 | 3,61E-01 | 5,58E-01 |
| CDK14 | 1104,96 | 0,07 | 0,08 | 0,91 | 3,61E-01 | 5,58E-01 |
| BLK | 591,62 | 0,10 | 0,11 | 0,91 | 3,61E-01 | 5,58E-01 |
| SENP8 | 34,60 | -0,12 | 0,13 | -0,91 | 3,61E-01 | 5,58E-01 |
| SGMS2 | 804,15 | 0,08 | 0,08 | 0,91 | 3,61E-01 | 5,58E-01 |
| RBM15B | 1396,55 | 0,02 | 0,03 | 0,91 | 3,61E-01 | 5,58E-01 |
| FSBP | 28,75 | 0,11 | 0,12 | 0,91 | 3,61E-01 | 5,58E-01 |
| BIN1 | 1566,20 | 0,06 | 0,06 | 0,91 | 3,61E-01 | 5,58E-01 |
| CCL4 | 433,95 | 0,08 | 0,09 | 0,91 | 3,61E-01 | 5,59E-01 |
| ATP1B3 | 476,68 | 0,04 | 0,04 | 0,91 | 3,61E-01 | 5,59E-01 |
| EEF1AKMT3 | 48,16 | -0,09 | 0,10 | -0,91 | 3,61E-01 | 5,59E-01 |
| GPS1 | 619,58 | -0,04 | 0,04 | -0,91 | 3,62E-01 | 5,59E-01 |
| RBM38 | 7422,90 | -0,13 | 0,14 | -0,91 | 3,62E-01 | 5,59E-01 |
| RILPL2 | 2040,84 | -0,05 | 0,05 | -0,91 | 3,62E-01 | 5,60E-01 |
| PPP1R15A | 909,53 | -0,07 | 0,08 | -0,91 | 3,62E-01 | 5,60E-01 |
| RIN2 | 527,16 | 0,06 | 0,07 | 0,91 | 3,63E-01 | 5,60E-01 |
| ACSL6 | 482,05 | -0,06 | 0,07 | -0,91 | 3,63E-01 | 5,60E-01 |
| TGFBRAP1 | 637,62 | -0,04 | 0,04 | -0,91 | 3,63E-01 | 5,60E-01 |
| PSMG4 | 1218,42 | 0,04 | 0,05 | 0,91 | 3,63E-01 | 5,60E-01 |
| GTF3C1 | 1325,97 | -0,05 | 0,06 | -0,91 | 3,62E-01 | 5,60E-01 |
| KDM3B | 3389,80 | -0,06 | 0,07 | -0,91 | 3,63E-01 | 5,60E-01 |
| SHF | 89,97 | -0,09 | 0,09 | -0,91 | 3,62E-01 | 5,60E-01 |
| NACC2 | 1549,93 | -0,08 | 0,08 | -0,91 | 3,62E-01 | 5,60E-01 |
| HLA-DRB5 | 1509,45 | 0,19 | 0,21 | 0,91 | 3,63E-01 | 5,60E-01 |

| | | | | | | |
|---|---|---|---|---|---|---|
| HERC4 | 6057,40 | 0,04 | 0,04 | 0,91 | 3,63E-01 | 5,60E-01 |
| CCDC30 | 377,12 | -0,06 | 0,06 | -0,91 | 3,63E-01 | 5,60E-01 |
| ZNF852 | 573,78 | 0,03 | 0,04 | 0,91 | 3,63E-01 | 5,60E-01 |
| ACIN1 | 4538,11 | -0,04 | 0,05 | -0,91 | 3,63E-01 | 5,60E-01 |
| ADPRS | 647,22 | 0,04 | 0,05 | 0,91 | 3,64E-01 | 5,61E-01 |
| AARSD1 | 69,67 | -0,07 | 0,08 | -0,91 | 3,64E-01 | 5,61E-01 |
| YKT6 | 1237,61 | -0,03 | 0,03 | -0,91 | 3,64E-01 | 5,61E-01 |
| IDH3B | 905,59 | 0,04 | 0,04 | 0,91 | 3,64E-01 | 5,61E-01 |
| ASCL2 | 288,54 | -0,07 | 0,08 | -0,91 | 3,64E-01 | 5,61E-01 |
| TMEM63A | 2983,70 | -0,09 | 0,10 | -0,91 | 3,64E-01 | 5,61E-01 |
| EPS8 | 136,81 | -0,10 | 0,11 | -0,91 | 3,64E-01 | 5,61E-01 |
| RTL6 | 367,65 | -0,07 | 0,07 | -0,91 | 3,64E-01 | 5,61E-01 |
| ACTN4 | 2376,18 | -0,08 | 0,09 | -0,91 | 3,65E-01 | 5,61E-01 |
| SH2D3A | 147,23 | -0,12 | 0,13 | -0,91 | 3,65E-01 | 5,61E-01 |
| APEX2 | 174,80 | -0,05 | 0,06 | -0,91 | 3,65E-01 | 5,61E-01 |
| AP5B1 | 2462,89 | -0,10 | 0,11 | -0,91 | 3,65E-01 | 5,62E-01 |
| SNX8 | 254,98 | -0,07 | 0,08 | -0,91 | 3,65E-01 | 5,62E-01 |
| DDB1 | 4024,31 | -0,05 | 0,06 | -0,91 | 3,65E-01 | 5,62E-01 |
| WDHD1 | 181,85 | 0,08 | 0,08 | 0,90 | 3,65E-01 | 5,62E-01 |
| FAM221A | 182,37 | 0,05 | 0,05 | 0,90 | 3,65E-01 | 5,62E-01 |
| TULP4 | 1264,77 | 0,03 | 0,04 | 0,90 | 3,66E-01 | 5,62E-01 |
| KLF8 | 186,95 | 0,08 | 0,09 | 0,90 | 3,66E-01 | 5,62E-01 |
| TMLHE | 847,71 | -0,04 | 0,05 | -0,91 | 3,65E-01 | 5,62E-01 |
| ZFY | 553,86 | 0,15 | 0,16 | 0,91 | 3,65E-01 | 5,62E-01 |
| GAS7 | 2404,21 | -0,08 | 0,09 | -0,90 | 3,66E-01 | 5,63E-01 |
| C16orf54 | 2813,09 | -0,06 | 0,07 | -0,90 | 3,66E-01 | 5,63E-01 |
| STAT2 | 3596,73 | -0,08 | 0,08 | -0,90 | 3,66E-01 | 5,63E-01 |
| ZNF76 | 546,47 | -0,06 | 0,06 | -0,90 | 3,66E-01 | 5,63E-01 |
| AFG2B | 688,66 | 0,04 | 0,05 | 0,90 | 3,66E-01 | 5,63E-01 |
| SPINT1 | 213,52 | -0,08 | 0,09 | -0,90 | 3,67E-01 | 5,63E-01 |
| CRIP2 | 111,09 | -0,13 | 0,14 | -0,90 | 3,67E-01 | 5,63E-01 |
| SLC27A1 | 147,32 | -0,11 | 0,13 | -0,90 | 3,67E-01 | 5,63E-01 |
| EFCAB14 | 3086,54 | 0,02 | 0,03 | 0,90 | 3,67E-01 | 5,63E-01 |
| GATB | 346,89 | -0,05 | 0,05 | -0,90 | 3,67E-01 | 5,64E-01 |
| TRAF3 | 1336,78 | -0,04 | 0,05 | -0,90 | 3,67E-01 | 5,64E-01 |
| MYCBP | 385,03 | 0,06 | 0,06 | 0,90 | 3,67E-01 | 5,64E-01 |
| C19orf53 | 307,78 | -0,05 | 0,05 | -0,90 | 3,67E-01 | 5,64E-01 |
| BMPR1A | 455,60 | 0,06 | 0,07 | 0,90 | 3,68E-01 | 5,64E-01 |
| ACSL1 | 24065,75 | -0,09 | 0,10 | -0,90 | 3,67E-01 | 5,64E-01 |
| KEAP1 | 502,48 | -0,04 | 0,04 | -0,90 | 3,67E-01 | 5,64E-01 |
| PAQR4 | 74,90 | -0,09 | 0,10 | -0,90 | 3,68E-01 | 5,64E-01 |
| AHCTF1 | 6987,58 | 0,05 | 0,06 | 0,90 | 3,68E-01 | 5,64E-01 |
| PLEKHG4 | 195,97 | -0,08 | 0,09 | -0,90 | 3,68E-01 | 5,64E-01 |
| SH3GLB1 | 4652,63 | 0,05 | 0,06 | 0,90 | 3,68E-01 | 5,64E-01 |
| ACAP2 | 12402,86 | 0,05 | 0,06 | 0,90 | 3,68E-01 | 5,64E-01 |
| BCL2L11 | 1671,73 | -0,05 | 0,05 | -0,90 | 3,68E-01 | 5,64E-01 |

| | | | | | | |
|---|---|---|---|---|---|---|
| RESF1 | 29300,92 | 0,07 | 0,08 | 0,90 | 3,68E-01 | 5,65E-01 |
| RPS3 | 9638,09 | 0,07 | 0,08 | 0,90 | 3,68E-01 | 5,65E-01 |
| BTG3 | 130,84 | 0,06 | 0,07 | 0,90 | 3,68E-01 | 5,65E-01 |
| ZNFX1 | 5659,10 | -0,05 | 0,06 | -0,90 | 3,69E-01 | 5,65E-01 |
| FAU | 4912,60 | 0,08 | 0,09 | 0,90 | 3,69E-01 | 5,65E-01 |
| RGS18 | 4671,13 | 0,12 | 0,13 | 0,90 | 3,69E-01 | 5,65E-01 |
| DNAJC5 | 2976,85 | -0,07 | 0,07 | -0,90 | 3,69E-01 | 5,65E-01 |
| SCO1 | 964,91 | 0,03 | 0,03 | 0,90 | 3,69E-01 | 5,65E-01 |
| EMC1 | 598,15 | -0,05 | 0,06 | -0,90 | 3,69E-01 | 5,66E-01 |
| FAM174B | 333,47 | -0,07 | 0,08 | -0,90 | 3,70E-01 | 5,66E-01 |
| SCD | 90,51 | -0,10 | 0,11 | -0,90 | 3,70E-01 | 5,66E-01 |
| PDZD4 | 474,81 | -0,09 | 0,10 | -0,90 | 3,70E-01 | 5,66E-01 |
| HUS1B | 77,35 | -0,07 | 0,08 | -0,90 | 3,69E-01 | 5,66E-01 |
| TUBG2 | 107,36 | -0,06 | 0,06 | -0,90 | 3,70E-01 | 5,66E-01 |
| EPB41 | 46588,13 | -0,10 | 0,11 | -0,90 | 3,70E-01 | 5,66E-01 |
| SIPA1L1 | 5127,50 | -0,06 | 0,07 | -0,90 | 3,70E-01 | 5,66E-01 |
| NUSAP1 | 35,82 | -0,08 | 0,09 | -0,90 | 3,70E-01 | 5,67E-01 |
| TMEM41A | 688,87 | 0,03 | 0,03 | 0,90 | 3,70E-01 | 5,67E-01 |
| VAV3 | 2543,55 | 0,04 | 0,05 | 0,90 | 3,70E-01 | 5,67E-01 |
| AVPR1A | 90,58 | -0,11 | 0,12 | -0,90 | 3,70E-01 | 5,67E-01 |
| RPL13 | 20208,45 | 0,06 | 0,07 | 0,90 | 3,70E-01 | 5,67E-01 |
| PKN1 | 1036,21 | -0,09 | 0,10 | -0,90 | 3,71E-01 | 5,67E-01 |
| ARL6IP6 | 1007,17 | 0,05 | 0,05 | 0,90 | 3,71E-01 | 5,67E-01 |
| KANK1 | 361,69 | -0,10 | 0,11 | -0,89 | 3,71E-01 | 5,68E-01 |
| PEX14 | 204,11 | -0,06 | 0,07 | -0,89 | 3,71E-01 | 5,68E-01 |
| CD8A | 1642,19 | 0,11 | 0,12 | 0,89 | 3,71E-01 | 5,68E-01 |
| TIGD6 | 137,54 | 0,06 | 0,06 | 0,89 | 3,71E-01 | 5,68E-01 |
| MUTYH | 296,40 | -0,04 | 0,04 | -0,89 | 3,72E-01 | 5,68E-01 |
| DLGAP5 | 60,10 | -0,14 | 0,15 | -0,89 | 3,72E-01 | 5,68E-01 |
| CDS2 | 4997,35 | -0,05 | 0,05 | -0,89 | 3,72E-01 | 5,69E-01 |
| PGAM1 | 1012,32 | -0,04 | 0,04 | -0,89 | 3,72E-01 | 5,69E-01 |
| DLST | 1572,02 | -0,03 | 0,03 | -0,89 | 3,72E-01 | 5,69E-01 |
| ITSN1 | 496,42 | -0,08 | 0,09 | -0,89 | 3,72E-01 | 5,69E-01 |
| HSH2D | 1492,18 | -0,08 | 0,10 | -0,89 | 3,72E-01 | 5,69E-01 |
| TRPC4AP | 2744,90 | -0,04 | 0,04 | -0,89 | 3,72E-01 | 5,69E-01 |
| ANKDD1A | 218,47 | -0,12 | 0,13 | -0,89 | 3,72E-01 | 5,69E-01 |
| F8 | 113,53 | -0,07 | 0,08 | -0,89 | 3,73E-01 | 5,69E-01 |
| CCNK | 1176,74 | 0,03 | 0,03 | 0,89 | 3,73E-01 | 5,69E-01 |
| PLIN2 | 767,38 | -0,05 | 0,05 | -0,89 | 3,73E-01 | 5,69E-01 |
| PDHA1 | 785,93 | 0,03 | 0,03 | 0,89 | 3,73E-01 | 5,69E-01 |
| PYGB | 1209,47 | -0,05 | 0,06 | -0,89 | 3,73E-01 | 5,69E-01 |
| ZNF710 | 721,94 | -0,08 | 0,09 | -0,89 | 3,73E-01 | 5,69E-01 |
| PNKD | 269,52 | -0,05 | 0,06 | -0,89 | 3,73E-01 | 5,70E-01 |
| ARNT | 1713,32 | -0,03 | 0,04 | -0,89 | 3,73E-01 | 5,70E-01 |
| CNOT6 | 1729,30 | 0,04 | 0,05 | 0,89 | 3,73E-01 | 5,70E-01 |
| IFNLR1 | 150,70 | -0,08 | 0,09 | -0,89 | 3,73E-01 | 5,70E-01 |

| | | | | | | |
|---|---|---|---|---|---|---|
| GARRE1 | 1017,60 | -0,04 | 0,05 | -0,89 | 3,74E-01 | 5,70E-01 |
| MTHFS | 290,25 | -0,05 | 0,06 | -0,89 | 3,74E-01 | 5,70E-01 |
| FITM2 | 232,40 | -0,06 | 0,06 | -0,89 | 3,74E-01 | 5,70E-01 |
| AMDHD1 | 54,18 | -0,09 | 0,10 | -0,89 | 3,74E-01 | 5,70E-01 |
| PXK | 1314,53 | -0,05 | 0,05 | -0,89 | 3,74E-01 | 5,70E-01 |
| ADD1 | 4293,22 | -0,06 | 0,07 | -0,89 | 3,74E-01 | 5,70E-01 |
| CAMK1D | 2270,61 | -0,07 | 0,08 | -0,89 | 3,74E-01 | 5,70E-01 |
| RAP2A | 1650,90 | 0,08 | 0,09 | 0,89 | 3,74E-01 | 5,70E-01 |
| TGIF2 | 368,95 | -0,05 | 0,06 | -0,89 | 3,74E-01 | 5,70E-01 |
| ZNF282 | 487,40 | -0,05 | 0,05 | -0,89 | 3,74E-01 | 5,70E-01 |
| MX2 | 5760,72 | -0,08 | 0,09 | -0,89 | 3,74E-01 | 5,70E-01 |
| WNT10B | 68,98 | -0,10 | 0,11 | -0,89 | 3,74E-01 | 5,70E-01 |
| CMTM6 | 11504,95 | 0,07 | 0,08 | 0,89 | 3,74E-01 | 5,70E-01 |
| MAP3K11 | 2700,52 | -0,07 | 0,07 | -0,89 | 3,75E-01 | 5,71E-01 |
| COMT | 628,18 | -0,04 | 0,05 | -0,89 | 3,75E-01 | 5,71E-01 |
| COPS9 | 336,32 | 0,06 | 0,07 | 0,89 | 3,75E-01 | 5,71E-01 |
| PIEZO1 | 1575,02 | -0,11 | 0,12 | -0,89 | 3,75E-01 | 5,72E-01 |
| PLGLB2 | 49,25 | 0,17 | 0,20 | 0,89 | 3,76E-01 | 5,72E-01 |
| MATR3 | 1765,82 | 0,04 | 0,05 | 0,89 | 3,76E-01 | 5,72E-01 |
| LSM1 | 437,74 | 0,07 | 0,08 | 0,89 | 3,76E-01 | 5,72E-01 |
| VCP | 4402,61 | 0,04 | 0,05 | 0,89 | 3,76E-01 | 5,72E-01 |
| ITPKC | 286,18 | -0,04 | 0,04 | -0,89 | 3,76E-01 | 5,72E-01 |
| C7orf25 | 128,54 | 0,06 | 0,07 | 0,89 | 3,76E-01 | 5,72E-01 |
| COPS3 | 1136,02 | 0,05 | 0,06 | 0,89 | 3,76E-01 | 5,72E-01 |
| FOXK1 | 1239,33 | -0,08 | 0,09 | -0,89 | 3,76E-01 | 5,72E-01 |
| CYB5R4 | 5930,86 | 0,06 | 0,07 | 0,89 | 3,76E-01 | 5,72E-01 |
| NINJ1 | 870,83 | -0,08 | 0,09 | -0,88 | 3,76E-01 | 5,72E-01 |
| COX5A | 148,24 | 0,05 | 0,05 | 0,88 | 3,76E-01 | 5,72E-01 |
| TOR1AIP1 | 3159,68 | 0,04 | 0,04 | 0,88 | 3,76E-01 | 5,72E-01 |
| LYPLA2 | 380,98 | -0,05 | 0,05 | -0,88 | 3,76E-01 | 5,72E-01 |
| DCBLD1 | 217,30 | 0,07 | 0,08 | 0,88 | 3,77E-01 | 5,72E-01 |
| NACC1 | 263,92 | -0,10 | 0,11 | -0,88 | 3,77E-01 | 5,72E-01 |
| LACTB | 1137,77 | 0,05 | 0,06 | 0,88 | 3,77E-01 | 5,72E-01 |
| PMPCA | 417,82 | -0,04 | 0,05 | -0,88 | 3,77E-01 | 5,72E-01 |
| PIK3C2B | 497,75 | -0,08 | 0,09 | -0,88 | 3,77E-01 | 5,72E-01 |
| BEX4 | 295,83 | 0,06 | 0,06 | 0,88 | 3,77E-01 | 5,72E-01 |
| RDH11 | 1067,40 | 0,04 | 0,04 | 0,88 | 3,77E-01 | 5,72E-01 |
| URM1 | 138,87 | -0,07 | 0,07 | -0,88 | 3,77E-01 | 5,72E-01 |
| RNF157 | 411,87 | -0,08 | 0,09 | -0,88 | 3,77E-01 | 5,72E-01 |
| NOM1 | 918,93 | 0,03 | 0,03 | 0,88 | 3,77E-01 | 5,72E-01 |
| RNPS1 | 1016,52 | -0,05 | 0,05 | -0,88 | 3,77E-01 | 5,72E-01 |
| LILRA1 | 1461,23 | -0,07 | 0,08 | -0,88 | 3,77E-01 | 5,72E-01 |
| PHF20L1 | 9225,52 | 0,07 | 0,08 | 0,88 | 3,77E-01 | 5,72E-01 |
| FMNL3 | 926,48 | -0,07 | 0,08 | -0,88 | 3,77E-01 | 5,72E-01 |
| SLC6A6 | 13846,74 | -0,06 | 0,07 | -0,88 | 3,77E-01 | 5,72E-01 |
| CD84 | 3286,48 | 0,04 | 0,04 | 0,88 | 3,77E-01 | 5,72E-01 |

| | | | | | | |
|---|---|---|---|---|---|---|
| RAB11FIP1 | 9392,27 | -0,06 | 0,07 | -0,88 | 3,77E-01 | 5,72E-01 |
| SWT1 | 851,55 | 0,09 | 0,11 | 0,88 | 3,78E-01 | 5,73E-01 |
| ARHGEF9 | 924,57 | 0,04 | 0,05 | 0,88 | 3,78E-01 | 5,74E-01 |
| KIF23 | 96,33 | -0,10 | 0,12 | -0,88 | 3,79E-01 | 5,74E-01 |
| SYCP3 | 51,92 | -0,09 | 0,10 | -0,88 | 3,79E-01 | 5,74E-01 |
| NSD3 | 7165,20 | 0,04 | 0,04 | 0,88 | 3,79E-01 | 5,74E-01 |
| CKAP4 | 836,84 | -0,07 | 0,08 | -0,88 | 3,79E-01 | 5,74E-01 |
| MLLT6 | 2719,55 | -0,09 | 0,10 | -0,88 | 3,79E-01 | 5,74E-01 |
| ATF7IP2 | 953,17 | 0,05 | 0,05 | 0,88 | 3,78E-01 | 5,74E-01 |
| ISG20L2 | 949,93 | -0,03 | 0,03 | -0,88 | 3,78E-01 | 5,74E-01 |
| SBNO1 | 3306,17 | 0,03 | 0,03 | 0,88 | 3,78E-01 | 5,74E-01 |
| TMEM9B | 1262,69 | 0,05 | 0,06 | 0,88 | 3,79E-01 | 5,74E-01 |
| ARHGEF1 | 4077,50 | -0,10 | 0,11 | -0,88 | 3,79E-01 | 5,74E-01 |
| MMP9 | 761,64 | -0,13 | 0,15 | -0,88 | 3,80E-01 | 5,75E-01 |
| RBKS | 94,87 | -0,06 | 0,07 | -0,88 | 3,80E-01 | 5,75E-01 |
| EHMT1 | 2617,92 | -0,06 | 0,07 | -0,88 | 3,80E-01 | 5,75E-01 |
| AIG1 | 382,89 | -0,04 | 0,05 | -0,88 | 3,80E-01 | 5,75E-01 |
| TAF8 | 1525,80 | -0,04 | 0,04 | -0,88 | 3,80E-01 | 5,75E-01 |
| RECQL5 | 504,72 | -0,05 | 0,05 | -0,88 | 3,80E-01 | 5,75E-01 |
| ENC1 | 856,83 | 0,11 | 0,12 | 0,88 | 3,80E-01 | 5,75E-01 |
| NCF2 | 21692,49 | -0,06 | 0,06 | -0,88 | 3,80E-01 | 5,75E-01 |
| PSKH1 | 338,46 | -0,06 | 0,07 | -0,88 | 3,80E-01 | 5,75E-01 |
| PUS10 | 481,19 | 0,05 | 0,06 | 0,88 | 3,80E-01 | 5,75E-01 |
| ERVK3-1 | 821,70 | 0,04 | 0,04 | 0,88 | 3,80E-01 | 5,75E-01 |
| FOXO4 | 3664,75 | -0,11 | 0,12 | -0,88 | 3,81E-01 | 5,76E-01 |
| CLEC5A | 161,26 | -0,10 | 0,11 | -0,88 | 3,81E-01 | 5,76E-01 |
| MSN | 5731,49 | -0,08 | 0,09 | -0,88 | 3,81E-01 | 5,76E-01 |
| ATP6V0B | 4282,34 | -0,06 | 0,06 | -0,88 | 3,81E-01 | 5,76E-01 |
| DAAM2 | 220,95 | -0,18 | 0,21 | -0,88 | 3,81E-01 | 5,76E-01 |
| KLRK1 | 421,14 | 0,08 | 0,10 | 0,88 | 3,81E-01 | 5,76E-01 |
| ZNF366 | 52,34 | -0,11 | 0,13 | -0,88 | 3,81E-01 | 5,76E-01 |
| ST6GALNAC2 | 472,03 | -0,09 | 0,10 | -0,88 | 3,81E-01 | 5,76E-01 |
| TTC31 | 366,24 | -0,06 | 0,06 | -0,88 | 3,81E-01 | 5,76E-01 |
| MTFR2 | 39,12 | -0,08 | 0,10 | -0,88 | 3,81E-01 | 5,76E-01 |
| NTMT1 | 215,35 | -0,04 | 0,05 | -0,88 | 3,81E-01 | 5,76E-01 |
| KLF7 | 2702,37 | -0,06 | 0,07 | -0,87 | 3,82E-01 | 5,76E-01 |
| PIGBOS1 | 231,07 | 0,04 | 0,05 | 0,87 | 3,82E-01 | 5,77E-01 |
| GEMIN7 | 240,41 | -0,04 | 0,05 | -0,87 | 3,82E-01 | 5,77E-01 |
| SMUG1 | 736,98 | 0,03 | 0,03 | 0,87 | 3,82E-01 | 5,77E-01 |
| SFXN3 | 604,28 | -0,04 | 0,05 | -0,87 | 3,82E-01 | 5,77E-01 |
| GPX4 | 1268,92 | -0,05 | 0,06 | -0,87 | 3,82E-01 | 5,77E-01 |
| CXXC5 | 460,78 | 0,06 | 0,07 | 0,87 | 3,83E-01 | 5,77E-01 |
| IMMT | 1089,20 | 0,03 | 0,03 | 0,87 | 3,83E-01 | 5,77E-01 |
| FDFT1 | 1956,68 | -0,03 | 0,04 | -0,87 | 3,83E-01 | 5,77E-01 |
| COG4 | 1360,07 | 0,02 | 0,03 | 0,87 | 3,83E-01 | 5,78E-01 |
| BTBD7 | 1434,63 | 0,03 | 0,03 | 0,87 | 3,83E-01 | 5,78E-01 |

| | | | | | | |
|---|---|---|---|---|---|---|
| TRAF6 | 1329,82 | 0,03 | 0,04 | 0,87 | 3,83E-01 | 5,78E-01 |
| PRKCSH | 945,88 | -0,07 | 0,08 | -0,87 | 3,83E-01 | 5,78E-01 |
| CD2 | 1176,93 | 0,07 | 0,08 | 0,87 | 3,83E-01 | 5,78E-01 |
| DPH3 | 474,86 | 0,07 | 0,08 | 0,87 | 3,83E-01 | 5,78E-01 |
| TNNI2 | 81,62 | -0,07 | 0,08 | -0,87 | 3,83E-01 | 5,78E-01 |
| MFSD14B | 2407,59 | -0,05 | 0,05 | -0,87 | 3,84E-01 | 5,78E-01 |
| RIN3 | 1290,26 | -0,10 | 0,12 | -0,87 | 3,84E-01 | 5,78E-01 |
| SMIM1 | 247,11 | -0,21 | 0,24 | -0,87 | 3,84E-01 | 5,78E-01 |
| PID1 | 350,29 | 0,07 | 0,08 | 0,87 | 3,84E-01 | 5,79E-01 |
| GID8 | 1560,11 | -0,04 | 0,05 | -0,87 | 3,84E-01 | 5,79E-01 |
| KIDINS220 | 6972,91 | 0,04 | 0,05 | 0,87 | 3,84E-01 | 5,79E-01 |
| ITPR2 | 7377,52 | 0,05 | 0,05 | 0,87 | 3,85E-01 | 5,79E-01 |
| ZNF778 | 662,50 | 0,02 | 0,03 | 0,87 | 3,85E-01 | 5,79E-01 |
| PDK3 | 1798,72 | -0,05 | 0,05 | -0,87 | 3,85E-01 | 5,79E-01 |
| LMF2 | 604,70 | -0,07 | 0,08 | -0,87 | 3,85E-01 | 5,80E-01 |
| TMEM199 | 604,23 | 0,02 | 0,02 | 0,87 | 3,85E-01 | 5,80E-01 |
| LRRFIP2 | 2396,01 | -0,04 | 0,04 | -0,87 | 3,85E-01 | 5,80E-01 |
| CCDC28A | 763,84 | 0,04 | 0,05 | 0,87 | 3,85E-01 | 5,80E-01 |
| S100P | 1719,93 | -0,15 | 0,17 | -0,87 | 3,86E-01 | 5,80E-01 |
| HOPX | 597,49 | 0,09 | 0,10 | 0,87 | 3,86E-01 | 5,80E-01 |
| RPL10 | 15734,04 | 0,05 | 0,06 | 0,87 | 3,86E-01 | 5,81E-01 |
| BLCAP | 1630,27 | -0,05 | 0,06 | -0,87 | 3,86E-01 | 5,81E-01 |
| SAP30BP | 1833,61 | 0,03 | 0,03 | 0,87 | 3,86E-01 | 5,81E-01 |
| RASGRP1 | 2936,75 | 0,05 | 0,05 | 0,87 | 3,86E-01 | 5,81E-01 |
| CCDC126 | 528,17 | 0,07 | 0,08 | 0,87 | 3,87E-01 | 5,81E-01 |
| HAAO | 49,84 | -0,10 | 0,11 | -0,87 | 3,86E-01 | 5,81E-01 |
| KHSRP | 637,54 | -0,07 | 0,09 | -0,87 | 3,87E-01 | 5,81E-01 |
| TBX21 | 659,77 | -0,09 | 0,10 | -0,87 | 3,87E-01 | 5,81E-01 |
| TXNDC15 | 944,65 | 0,03 | 0,03 | 0,87 | 3,87E-01 | 5,81E-01 |
| DIPK2A | 1158,43 | 0,05 | 0,06 | 0,87 | 3,87E-01 | 5,81E-01 |
| GPR162 | 142,68 | -0,11 | 0,12 | -0,87 | 3,87E-01 | 5,81E-01 |
| C19orf25 | 820,47 | -0,05 | 0,05 | -0,87 | 3,87E-01 | 5,81E-01 |
| MAP3K4 | 1734,07 | 0,04 | 0,05 | 0,87 | 3,87E-01 | 5,81E-01 |
| LENG8 | 2144,53 | -0,12 | 0,14 | -0,87 | 3,87E-01 | 5,81E-01 |
| PPP2R1A | 1571,33 | -0,04 | 0,05 | -0,87 | 3,87E-01 | 5,81E-01 |
| COPG2 | 399,81 | 0,04 | 0,04 | 0,87 | 3,87E-01 | 5,81E-01 |
| RFC5 | 287,59 | -0,02 | 0,03 | -0,86 | 3,87E-01 | 5,81E-01 |
| PANK4 | 838,86 | -0,04 | 0,04 | -0,86 | 3,87E-01 | 5,82E-01 |
| ZKSCAN2 | 186,65 | -0,06 | 0,07 | -0,86 | 3,87E-01 | 5,82E-01 |
| DNAAF11 | 382,93 | 0,10 | 0,12 | 0,86 | 3,87E-01 | 5,82E-01 |
| CEP97 | 939,93 | 0,05 | 0,06 | 0,86 | 3,88E-01 | 5,82E-01 |
| HIC2 | 384,03 | -0,04 | 0,05 | -0,86 | 3,88E-01 | 5,82E-01 |
| TMED4 | 1914,47 | 0,02 | 0,03 | 0,86 | 3,88E-01 | 5,82E-01 |
| SLC3A2 | 1185,16 | -0,04 | 0,04 | -0,86 | 3,88E-01 | 5,82E-01 |
| CFD | 3600,43 | -0,14 | 0,16 | -0,86 | 3,88E-01 | 5,83E-01 |
| STAC3 | 120,87 | -0,06 | 0,07 | -0,86 | 3,88E-01 | 5,83E-01 |

| | | | | | | |
|---|---|---|---|---|---|---|
| SEMA4C | 511,56 | -0,06 | 0,07 | -0,86 | 3,88E-01 | 5,83E-01 |
| GOSR2 | 2035,53 | 0,03 | 0,04 | 0,86 | 3,89E-01 | 5,83E-01 |
| PPP3CA | 3149,79 | 0,04 | 0,05 | 0,86 | 3,89E-01 | 5,83E-01 |
| CDYL | 1390,25 | -0,07 | 0,09 | -0,86 | 3,89E-01 | 5,83E-01 |
| RNF217 | 418,70 | 0,07 | 0,08 | 0,86 | 3,89E-01 | 5,83E-01 |
| PDGFB | 79,19 | -0,11 | 0,13 | -0,86 | 3,89E-01 | 5,83E-01 |
| TARS1 | 1055,27 | 0,05 | 0,05 | 0,86 | 3,89E-01 | 5,83E-01 |
| CCDC12 | 814,01 | 0,03 | 0,04 | 0,86 | 3,89E-01 | 5,84E-01 |
| IL4R | 5775,26 | 0,06 | 0,07 | 0,86 | 3,90E-01 | 5,84E-01 |
| PRKCA | 1455,72 | -0,06 | 0,07 | -0,86 | 3,90E-01 | 5,84E-01 |
| AMD1 | 4951,98 | 0,05 | 0,06 | 0,86 | 3,90E-01 | 5,84E-01 |
| MLF2 | 1206,12 | -0,05 | 0,06 | -0,86 | 3,90E-01 | 5,84E-01 |
| DUSP19 | 101,72 | -0,05 | 0,06 | -0,86 | 3,90E-01 | 5,84E-01 |
| LMNB2 | 392,95 | -0,05 | 0,06 | -0,86 | 3,90E-01 | 5,84E-01 |
| JMJD8 | 135,77 | -0,07 | 0,09 | -0,86 | 3,90E-01 | 5,84E-01 |
| DCP2 | 10954,36 | 0,06 | 0,07 | 0,86 | 3,90E-01 | 5,84E-01 |
| RREB1 | 2302,39 | -0,05 | 0,06 | -0,86 | 3,90E-01 | 5,84E-01 |
| KLF16 | 385,82 | -0,07 | 0,08 | -0,86 | 3,91E-01 | 5,85E-01 |
| RAB5A | 1756,07 | 0,04 | 0,05 | 0,86 | 3,91E-01 | 5,85E-01 |
| NDUFV1 | 992,61 | 0,03 | 0,04 | 0,86 | 3,91E-01 | 5,85E-01 |
| EMC4 | 886,56 | 0,04 | 0,04 | 0,86 | 3,91E-01 | 5,85E-01 |
| PCID2 | 1450,69 | 0,03 | 0,04 | 0,86 | 3,91E-01 | 5,86E-01 |
| CUEDC2 | 277,92 | 0,03 | 0,04 | 0,86 | 3,91E-01 | 5,86E-01 |
| ENSG000002 | 45,89 | -0,05 | 0,06 | -0,86 | 3,92E-01 | 5,86E-01 |
| FCGR1BP | 662,90 | -0,10 | 0,12 | -0,86 | 3,92E-01 | 5,86E-01 |
| ZNF174 | 235,53 | -0,02 | 0,03 | -0,86 | 3,92E-01 | 5,86E-01 |
| WDR4 | 274,22 | -0,06 | 0,07 | -0,86 | 3,92E-01 | 5,86E-01 |
| LETM2 | 214,18 | -0,06 | 0,07 | -0,86 | 3,92E-01 | 5,86E-01 |
| HIPK3 | 10618,41 | 0,05 | 0,06 | 0,86 | 3,92E-01 | 5,86E-01 |
| TACSTD2 | 46,55 | 0,19 | 0,23 | 0,86 | 3,92E-01 | 5,86E-01 |
| ARPC1B | 1095,09 | -0,04 | 0,05 | -0,86 | 3,92E-01 | 5,86E-01 |
| RAPGEF1 | 2284,94 | -0,09 | 0,10 | -0,86 | 3,92E-01 | 5,86E-01 |
| RAPH1 | 235,91 | 0,07 | 0,09 | 0,86 | 3,92E-01 | 5,86E-01 |
| SUV39H2 | 99,56 | 0,06 | 0,08 | 0,86 | 3,92E-01 | 5,86E-01 |
| MFSD3 | 107,42 | -0,05 | 0,06 | -0,86 | 3,92E-01 | 5,86E-01 |
| ZFHX3 | 425,66 | -0,10 | 0,11 | -0,86 | 3,92E-01 | 5,86E-01 |
| TRIM14 | 1026,97 | -0,07 | 0,08 | -0,86 | 3,92E-01 | 5,86E-01 |
| ST7L | 458,34 | 0,03 | 0,04 | 0,86 | 3,92E-01 | 5,86E-01 |
| EBNA1BP2 | 359,53 | 0,04 | 0,05 | 0,85 | 3,93E-01 | 5,86E-01 |
| H2AC20 | 5990,19 | 0,07 | 0,08 | 0,85 | 3,93E-01 | 5,87E-01 |
| PES1 | 766,32 | 0,03 | 0,04 | 0,85 | 3,94E-01 | 5,87E-01 |
| TEX30 | 148,88 | 0,07 | 0,08 | 0,85 | 3,93E-01 | 5,87E-01 |
| TMEM204 | 648,86 | 0,07 | 0,08 | 0,85 | 3,93E-01 | 5,87E-01 |
| AP4B1 | 1196,20 | 0,03 | 0,03 | 0,85 | 3,93E-01 | 5,87E-01 |
| KIAA0040 | 3583,45 | -0,04 | 0,05 | -0,85 | 3,94E-01 | 5,87E-01 |
| GPR108 | 676,52 | -0,04 | 0,05 | -0,85 | 3,94E-01 | 5,88E-01 |

| | | | | | | |
|---|---|---|---|---|---|---|
| TMEM205 | 404,64 | 0,04 | 0,05 | 0,85 | 3,94E-01 | 5,88E-01 |
| SSH2 | 16053,48 | -0,06 | 0,07 | -0,85 | 3,94E-01 | 5,88E-01 |
| SERTAD3 | 266,38 | -0,04 | 0,05 | -0,85 | 3,95E-01 | 5,89E-01 |
| TRIM41 | 989,81 | -0,03 | 0,04 | -0,85 | 3,95E-01 | 5,89E-01 |
| ALG1 | 396,11 | -0,03 | 0,03 | -0,85 | 3,96E-01 | 5,90E-01 |
| SDF4 | 848,76 | -0,06 | 0,06 | -0,85 | 3,96E-01 | 5,90E-01 |
| CDC42BPB | 396,30 | -0,07 | 0,08 | -0,85 | 3,96E-01 | 5,90E-01 |
| TBC1D1 | 4360,17 | -0,05 | 0,06 | -0,85 | 3,96E-01 | 5,90E-01 |
| FICD | 255,68 | 0,03 | 0,03 | 0,85 | 3,96E-01 | 5,90E-01 |
| ZFP28 | 202,71 | 0,07 | 0,08 | 0,85 | 3,96E-01 | 5,90E-01 |
| PGRMC1 | 1021,93 | -0,08 | 0,09 | -0,85 | 3,96E-01 | 5,90E-01 |
| ARHGEF5 | 58,66 | -0,12 | 0,14 | -0,85 | 3,96E-01 | 5,90E-01 |
| TUBGCP3 | 2180,81 | -0,04 | 0,04 | -0,85 | 3,96E-01 | 5,90E-01 |
| ADGRD1 | 190,24 | -0,13 | 0,15 | -0,85 | 3,97E-01 | 5,91E-01 |
| E2F2 | 1287,64 | -0,12 | 0,15 | -0,85 | 3,97E-01 | 5,91E-01 |
| FAM131A | 525,84 | -0,05 | 0,06 | -0,85 | 3,97E-01 | 5,91E-01 |
| LMO2 | 1949,00 | 0,05 | 0,06 | 0,85 | 3,97E-01 | 5,91E-01 |
| METTL13 | 726,14 | 0,03 | 0,03 | 0,85 | 3,97E-01 | 5,91E-01 |
| WLS | 1703,17 | 0,11 | 0,13 | 0,85 | 3,97E-01 | 5,91E-01 |
| SFXN2 | 255,19 | -0,07 | 0,08 | -0,85 | 3,97E-01 | 5,91E-01 |
| PIP4K2C | 588,33 | -0,03 | 0,04 | -0,85 | 3,97E-01 | 5,91E-01 |
| TNFRSF10D | 316,76 | -0,04 | 0,05 | -0,85 | 3,97E-01 | 5,91E-01 |
| ORAI2 | 3339,55 | -0,07 | 0,08 | -0,85 | 3,97E-01 | 5,91E-01 |
| NFATC3 | 3350,76 | -0,03 | 0,04 | -0,85 | 3,97E-01 | 5,91E-01 |
| KLF4 | 481,69 | 0,06 | 0,07 | 0,85 | 3,97E-01 | 5,91E-01 |
| ACAT2 | 380,15 | 0,04 | 0,05 | 0,85 | 3,97E-01 | 5,91E-01 |
| TFAP4 | 176,68 | -0,08 | 0,10 | -0,85 | 3,98E-01 | 5,91E-01 |
| CAT | 6430,70 | -0,06 | 0,07 | -0,84 | 3,98E-01 | 5,92E-01 |
| TRIM28 | 916,45 | -0,07 | 0,08 | -0,84 | 3,98E-01 | 5,92E-01 |
| STX5 | 741,13 | -0,03 | 0,04 | -0,84 | 3,98E-01 | 5,92E-01 |
| BMP2K | 4407,56 | 0,05 | 0,06 | 0,84 | 3,98E-01 | 5,92E-01 |
| CSTF1 | 679,46 | 0,02 | 0,03 | 0,84 | 3,98E-01 | 5,92E-01 |
| INAFM1 | 127,82 | -0,06 | 0,07 | -0,84 | 3,99E-01 | 5,92E-01 |
| STEAP4 | 10863,20 | -0,08 | 0,09 | -0,84 | 3,99E-01 | 5,92E-01 |
| IL6R | 10077,12 | -0,06 | 0,07 | -0,84 | 3,99E-01 | 5,92E-01 |
| HSPA1B | 303,06 | -0,06 | 0,07 | -0,84 | 3,99E-01 | 5,92E-01 |
| OGFOD2 | 64,96 | -0,04 | 0,05 | -0,84 | 3,99E-01 | 5,92E-01 |
| KHDC4 | 1516,48 | 0,03 | 0,03 | 0,84 | 3,99E-01 | 5,92E-01 |
| RAD51C | 271,98 | 0,05 | 0,06 | 0,84 | 3,99E-01 | 5,93E-01 |
| CHI3L2 | 233,82 | -0,07 | 0,09 | -0,84 | 3,99E-01 | 5,93E-01 |
| DMXL2 | 10352,72 | 0,06 | 0,07 | 0,84 | 3,99E-01 | 5,93E-01 |
| MACF1 | 26844,96 | 0,04 | 0,05 | 0,84 | 4,00E-01 | 5,93E-01 |
| KAT2A | 192,67 | -0,08 | 0,10 | -0,84 | 4,00E-01 | 5,93E-01 |
| BLMH | 485,49 | 0,03 | 0,04 | 0,84 | 4,00E-01 | 5,93E-01 |
| FAM162A | 265,75 | 0,04 | 0,05 | 0,84 | 4,00E-01 | 5,93E-01 |
| SDHC | 1617,37 | 0,03 | 0,03 | 0,84 | 4,00E-01 | 5,93E-01 |

| | | | | | | |
|---|---|---|---|---|---|---|
| SIPA1L3 | 582,37 | -0,09 | 0,11 | -0,84 | 4,00E-01 | 5,94E-01 |
| ITPK1 | 1876,40 | -0,08 | 0,09 | -0,84 | 4,00E-01 | 5,94E-01 |
| GSTZ1 | 309,25 | 0,04 | 0,05 | 0,84 | 4,00E-01 | 5,94E-01 |
| FCHO2 | 1828,52 | -0,06 | 0,07 | -0,84 | 4,01E-01 | 5,94E-01 |
| BUB1 | 109,14 | -0,09 | 0,11 | -0,84 | 4,01E-01 | 5,94E-01 |
| PHEX | 85,51 | 0,10 | 0,12 | 0,84 | 4,01E-01 | 5,94E-01 |
| RBM5 | 9112,91 | 0,03 | 0,03 | 0,84 | 4,01E-01 | 5,94E-01 |
| RPRD2 | 982,19 | 0,03 | 0,03 | 0,84 | 4,01E-01 | 5,94E-01 |
| RAB39A | 122,97 | 0,07 | 0,08 | 0,84 | 4,01E-01 | 5,94E-01 |
| PPP4R4 | 71,97 | -0,14 | 0,17 | -0,84 | 4,01E-01 | 5,94E-01 |
| RCC2 | 1874,71 | 0,04 | 0,05 | 0,84 | 4,01E-01 | 5,94E-01 |
| METTL4 | 784,10 | 0,05 | 0,06 | 0,84 | 4,01E-01 | 5,94E-01 |
| TMEM154 | 18729,12 | -0,05 | 0,06 | -0,84 | 4,01E-01 | 5,95E-01 |
| UBE2F-SCLY | 19,88 | -0,07 | 0,08 | -0,84 | 4,02E-01 | 5,95E-01 |
| NEK11 | 155,84 | -0,07 | 0,08 | -0,84 | 4,02E-01 | 5,95E-01 |
| CDK4 | 174,98 | -0,04 | 0,05 | -0,84 | 4,02E-01 | 5,95E-01 |
| ZNF578 | 59,61 | -0,09 | 0,11 | -0,84 | 4,02E-01 | 5,95E-01 |
| LYSMD4 | 465,49 | -0,04 | 0,05 | -0,84 | 4,02E-01 | 5,95E-01 |
| MNT | 707,83 | -0,08 | 0,10 | -0,84 | 4,02E-01 | 5,95E-01 |
| MITF | 152,11 | -0,07 | 0,09 | -0,84 | 4,02E-01 | 5,95E-01 |
| DCUN1D2 | 720,08 | 0,02 | 0,03 | 0,84 | 4,02E-01 | 5,95E-01 |
| CLYBL | 225,66 | -0,06 | 0,07 | -0,84 | 4,02E-01 | 5,95E-01 |
| NCOA1 | 5691,87 | -0,04 | 0,05 | -0,84 | 4,03E-01 | 5,96E-01 |
| SLC38A7 | 141,28 | -0,08 | 0,09 | -0,84 | 4,03E-01 | 5,96E-01 |
| LRMDA | 234,81 | -0,11 | 0,13 | -0,84 | 4,03E-01 | 5,96E-01 |
| DCAF7 | 1645,21 | -0,05 | 0,07 | -0,84 | 4,03E-01 | 5,96E-01 |
| COL5A3 | 115,39 | -0,22 | 0,26 | -0,84 | 4,03E-01 | 5,96E-01 |
| TOM1L2 | 672,05 | -0,06 | 0,07 | -0,84 | 4,03E-01 | 5,96E-01 |
| IFIT3 | 3948,96 | 0,14 | 0,17 | 0,84 | 4,03E-01 | 5,96E-01 |
| ZNF554 | 173,14 | -0,05 | 0,05 | -0,84 | 4,03E-01 | 5,96E-01 |
| ZNF517 | 207,61 | -0,05 | 0,06 | -0,84 | 4,03E-01 | 5,96E-01 |
| RIGI | 4315,20 | 0,08 | 0,09 | 0,84 | 4,04E-01 | 5,96E-01 |
| ATL1 | 122,90 | -0,07 | 0,08 | -0,84 | 4,04E-01 | 5,96E-01 |
| ZNF467 | 521,71 | -0,09 | 0,11 | -0,84 | 4,04E-01 | 5,96E-01 |
| FBXL5 | 14788,61 | 0,07 | 0,08 | 0,84 | 4,04E-01 | 5,96E-01 |
| PLD1 | 701,97 | -0,06 | 0,07 | -0,83 | 4,04E-01 | 5,97E-01 |
| ENSG000002 | 23,22 | -0,08 | 0,09 | -0,83 | 4,04E-01 | 5,97E-01 |
| IRAK3 | 6475,55 | -0,07 | 0,08 | -0,83 | 4,04E-01 | 5,97E-01 |
| CCDC146 | 478,52 | -0,06 | 0,07 | -0,83 | 4,04E-01 | 5,97E-01 |
| EDEM1 | 3749,98 | -0,04 | 0,04 | -0,83 | 4,05E-01 | 5,97E-01 |
| POLDIP2 | 828,44 | -0,03 | 0,04 | -0,83 | 4,05E-01 | 5,97E-01 |
| HECTD2 | 148,65 | 0,10 | 0,12 | 0,83 | 4,05E-01 | 5,98E-01 |
| SNAP29 | 732,19 | -0,03 | 0,04 | -0,83 | 4,05E-01 | 5,98E-01 |
| ZNF630 | 143,95 | 0,05 | 0,06 | 0,83 | 4,05E-01 | 5,98E-01 |
| DCTN1 | 1512,40 | -0,05 | 0,06 | -0,83 | 4,06E-01 | 5,99E-01 |
| ZNF526 | 164,58 | -0,08 | 0,09 | -0,83 | 4,06E-01 | 5,99E-01 |

| | | | | | | |
|---|---|---|---|---|---|---|
| HIF1A | 4150,81 | 0,05 | 0,06 | 0,83 | 4,06E-01 | 5,99E-01 |
| PACS1 | 2677,64 | -0,07 | 0,09 | -0,83 | 4,06E-01 | 5,99E-01 |
| AFG1L | 207,53 | 0,04 | 0,05 | 0,83 | 4,06E-01 | 5,99E-01 |
| JUNB | 3989,09 | -0,06 | 0,07 | -0,83 | 4,07E-01 | 5,99E-01 |
| MYO1D | 290,28 | -0,06 | 0,08 | -0,83 | 4,07E-01 | 5,99E-01 |
| OGG1 | 576,32 | -0,05 | 0,07 | -0,83 | 4,07E-01 | 5,99E-01 |
| CLDN5 | 202,54 | -0,11 | 0,13 | -0,83 | 4,07E-01 | 5,99E-01 |
| GEMIN5 | 555,64 | 0,03 | 0,03 | 0,83 | 4,07E-01 | 5,99E-01 |
| SNRNP25 | 151,81 | 0,05 | 0,06 | 0,83 | 4,07E-01 | 5,99E-01 |
| GMEB2 | 413,30 | -0,07 | 0,08 | -0,83 | 4,07E-01 | 5,99E-01 |
| SIGLEC11 | 26,89 | -0,17 | 0,20 | -0,83 | 4,07E-01 | 5,99E-01 |
| AHR | 2081,06 | 0,06 | 0,07 | 0,83 | 4,07E-01 | 6,00E-01 |
| TOE1 | 249,35 | 0,03 | 0,03 | 0,83 | 4,07E-01 | 6,00E-01 |
| TFDP1 | 1768,32 | -0,08 | 0,10 | -0,83 | 4,07E-01 | 6,00E-01 |
| MRPL18 | 475,52 | 0,04 | 0,05 | 0,83 | 4,07E-01 | 6,00E-01 |
| FRMD3 | 301,48 | -0,08 | 0,09 | -0,83 | 4,07E-01 | 6,00E-01 |
| AHI1 | 348,54 | 0,08 | 0,09 | 0,83 | 4,08E-01 | 6,01E-01 |
| FOXN3 | 4440,07 | -0,04 | 0,05 | -0,83 | 4,08E-01 | 6,01E-01 |
| INTS8 | 2515,26 | 0,03 | 0,04 | 0,83 | 4,08E-01 | 6,01E-01 |
| GPR107 | 1764,18 | -0,05 | 0,06 | -0,83 | 4,08E-01 | 6,01E-01 |
| ZFP57 | 37,13 | -0,40 | 0,48 | -0,83 | 4,08E-01 | 6,01E-01 |
| MAST4 | 586,58 | -0,05 | 0,06 | -0,83 | 4,08E-01 | 6,01E-01 |
| DOCK5 | 14326,23 | -0,06 | 0,08 | -0,83 | 4,08E-01 | 6,01E-01 |
| KCTD9 | 966,02 | 0,05 | 0,06 | 0,83 | 4,09E-01 | 6,01E-01 |
| POU2F2 | 754,16 | -0,09 | 0,11 | -0,83 | 4,09E-01 | 6,02E-01 |
| MFSD2A | 108,26 | -0,07 | 0,09 | -0,83 | 4,09E-01 | 6,02E-01 |
| KLHDC2 | 1540,22 | 0,05 | 0,06 | 0,83 | 4,09E-01 | 6,02E-01 |
| MTPN | 9376,66 | -0,05 | 0,06 | -0,83 | 4,09E-01 | 6,02E-01 |
| ACOX3 | 493,75 | -0,03 | 0,04 | -0,83 | 4,09E-01 | 6,02E-01 |
| ENSG000001 | 28,64 | -0,07 | 0,09 | -0,83 | 4,09E-01 | 6,02E-01 |
| ATG16L1 | 1755,85 | 0,02 | 0,03 | 0,83 | 4,09E-01 | 6,02E-01 |
| GIMAP1 | 2399,95 | 0,03 | 0,04 | 0,82 | 4,09E-01 | 6,02E-01 |
| JMJD7-PLA2( | 311,60 | -0,08 | 0,10 | -0,82 | 4,09E-01 | 6,02E-01 |
| R3HDM2 | 758,58 | -0,06 | 0,07 | -0,82 | 4,10E-01 | 6,02E-01 |
| PTCH1 | 549,51 | -0,09 | 0,11 | -0,82 | 4,10E-01 | 6,02E-01 |
| TOMM40L | 280,52 | -0,03 | 0,04 | -0,82 | 4,10E-01 | 6,02E-01 |
| MFSD9 | 564,89 | -0,06 | 0,07 | -0,82 | 4,10E-01 | 6,02E-01 |
| MCMBP | 2949,74 | 0,04 | 0,05 | 0,82 | 4,10E-01 | 6,02E-01 |
| TNFAIP2 | 7673,71 | -0,08 | 0,10 | -0,82 | 4,10E-01 | 6,02E-01 |
| SRSF2 | 2878,18 | 0,03 | 0,04 | 0,82 | 4,10E-01 | 6,02E-01 |
| CBR1 | 390,35 | 0,05 | 0,06 | 0,82 | 4,10E-01 | 6,02E-01 |
| TRIB1 | 726,92 | -0,05 | 0,06 | -0,82 | 4,10E-01 | 6,02E-01 |
| SGMS1 | 1807,38 | 0,03 | 0,04 | 0,82 | 4,10E-01 | 6,02E-01 |
| SH2B1 | 482,94 | -0,09 | 0,11 | -0,82 | 4,10E-01 | 6,02E-01 |
| PDE3B | 6405,50 | 0,05 | 0,06 | 0,82 | 4,10E-01 | 6,02E-01 |
| NUDT5 | 1617,76 | 0,03 | 0,04 | 0,82 | 4,11E-01 | 6,02E-01 |

| | | | | | | |
|---|---|---|---|---|---|---|
| NHERF1 | 2050,44 | -0,06 | 0,07 | -0,82 | 4,11E-01 | 6,02E-01 |
| LRATD2 | 526,05 | 0,05 | 0,06 | 0,82 | 4,11E-01 | 6,02E-01 |
| UBE2D1 | 2635,59 | 0,07 | 0,09 | 0,82 | 4,11E-01 | 6,02E-01 |
| DDX39B | 1559,95 | -0,09 | 0,10 | -0,82 | 4,11E-01 | 6,02E-01 |
| TNFSF15 | 54,63 | -0,12 | 0,14 | -0,82 | 4,11E-01 | 6,02E-01 |
| CPED1 | 343,95 | 0,07 | 0,08 | 0,82 | 4,11E-01 | 6,03E-01 |
| NIT1 | 572,18 | -0,03 | 0,03 | -0,82 | 4,11E-01 | 6,03E-01 |
| DYNC1LI1 | 2212,39 | 0,05 | 0,06 | 0,82 | 4,11E-01 | 6,03E-01 |
| CRADD | 424,31 | -0,03 | 0,04 | -0,82 | 4,11E-01 | 6,03E-01 |
| TET3 | 2712,17 | -0,06 | 0,07 | -0,82 | 4,11E-01 | 6,03E-01 |
| DPY19L3 | 1296,70 | 0,04 | 0,05 | 0,82 | 4,12E-01 | 6,03E-01 |
| VWA5A | 237,58 | -0,04 | 0,05 | -0,82 | 4,12E-01 | 6,03E-01 |
| CD160 | 257,01 | 0,10 | 0,12 | 0,82 | 4,12E-01 | 6,03E-01 |
| BTNL8 | 638,50 | -0,07 | 0,09 | -0,82 | 4,12E-01 | 6,03E-01 |
| ETFBKMT | 100,11 | -0,05 | 0,06 | -0,82 | 4,12E-01 | 6,03E-01 |
| ZNF532 | 418,35 | 0,06 | 0,08 | 0,82 | 4,12E-01 | 6,03E-01 |
| RWDD2A | 91,47 | 0,06 | 0,07 | 0,82 | 4,12E-01 | 6,03E-01 |
| ANKFY1 | 2635,04 | -0,04 | 0,05 | -0,82 | 4,12E-01 | 6,03E-01 |
| ZNF318 | 1330,18 | 0,04 | 0,05 | 0,82 | 4,12E-01 | 6,03E-01 |
| EMC9 | 88,85 | -0,04 | 0,05 | -0,82 | 4,12E-01 | 6,03E-01 |
| METTL16 | 567,83 | -0,05 | 0,06 | -0,82 | 4,12E-01 | 6,03E-01 |
| PHRF1 | 971,18 | -0,07 | 0,08 | -0,82 | 4,12E-01 | 6,03E-01 |
| CPOX | 822,39 | 0,03 | 0,04 | 0,82 | 4,12E-01 | 6,03E-01 |
| TP53BP2 | 1287,08 | 0,02 | 0,03 | 0,82 | 4,12E-01 | 6,03E-01 |
| FCGR3B | 37037,79 | -0,09 | 0,10 | -0,82 | 4,12E-01 | 6,03E-01 |
| ZNF18 | 834,85 | 0,03 | 0,03 | 0,82 | 4,13E-01 | 6,04E-01 |
| PACSIN2 | 5183,91 | -0,06 | 0,07 | -0,82 | 4,13E-01 | 6,04E-01 |
| SH3BP1 | 247,68 | -0,07 | 0,09 | -0,82 | 4,13E-01 | 6,04E-01 |
| XIAP | 3819,64 | 0,03 | 0,04 | 0,82 | 4,13E-01 | 6,04E-01 |
| FCHO1 | 1096,75 | -0,06 | 0,07 | -0,82 | 4,13E-01 | 6,04E-01 |
| SP1 | 6390,55 | -0,04 | 0,05 | -0,82 | 4,14E-01 | 6,05E-01 |
| RPN2 | 1843,22 | 0,03 | 0,03 | 0,82 | 4,14E-01 | 6,05E-01 |
| PARP1 | 2246,99 | 0,04 | 0,05 | 0,82 | 4,14E-01 | 6,05E-01 |
| ING2 | 117,76 | 0,05 | 0,06 | 0,82 | 4,14E-01 | 6,05E-01 |
| TIAM1 | 2121,59 | 0,04 | 0,05 | 0,82 | 4,14E-01 | 6,05E-01 |
| CCDC115 | 835,21 | 0,03 | 0,03 | 0,82 | 4,14E-01 | 6,05E-01 |
| COQ2 | 528,14 | 0,04 | 0,05 | 0,82 | 4,14E-01 | 6,05E-01 |
| LRRC63 | 31,63 | -0,08 | 0,10 | -0,82 | 4,14E-01 | 6,05E-01 |
| SOWAHD | 269,37 | -0,06 | 0,07 | -0,82 | 4,14E-01 | 6,05E-01 |
| YES1 | 336,72 | 0,06 | 0,07 | 0,82 | 4,14E-01 | 6,05E-01 |
| TAGLN | 431,96 | 0,09 | 0,11 | 0,82 | 4,14E-01 | 6,05E-01 |
| TRMU | 895,81 | -0,03 | 0,04 | -0,82 | 4,14E-01 | 6,05E-01 |
| SHPK | 32,70 | -0,06 | 0,08 | -0,82 | 4,14E-01 | 6,05E-01 |
| APOBEC3A | 4626,71 | -0,07 | 0,09 | -0,82 | 4,15E-01 | 6,05E-01 |
| PHYKPL | 3139,17 | -0,05 | 0,07 | -0,82 | 4,15E-01 | 6,05E-01 |
| TMEM183A | 2067,63 | -0,05 | 0,06 | -0,82 | 4,15E-01 | 6,05E-01 |

| | | | | | | |
|---|---|---|---|---|---|---|
| ST8SIA4 | 8742,63 | 0,06 | 0,07 | 0,82 | 4,15E-01 | 6,05E-01 |
| ZBTB3 | 173,09 | -0,05 | 0,06 | -0,82 | 4,15E-01 | 6,05E-01 |
| RCC1 | 261,47 | -0,04 | 0,05 | -0,82 | 4,15E-01 | 6,05E-01 |
| DHRS4L2 | 172,80 | -0,04 | 0,05 | -0,81 | 4,15E-01 | 6,05E-01 |
| ARID3A | 1359,02 | -0,07 | 0,08 | -0,81 | 4,15E-01 | 6,05E-01 |
| NFAT5 | 5113,80 | -0,05 | 0,06 | -0,81 | 4,16E-01 | 6,06E-01 |
| JADE3 | 81,16 | -0,07 | 0,09 | -0,81 | 4,16E-01 | 6,06E-01 |
| RIPK1 | 1324,41 | -0,03 | 0,04 | -0,81 | 4,16E-01 | 6,06E-01 |
| PSME3 | 1445,88 | -0,05 | 0,06 | -0,81 | 4,16E-01 | 6,06E-01 |
| EXOSC5 | 186,91 | 0,03 | 0,04 | 0,81 | 4,16E-01 | 6,06E-01 |
| MED17 | 1392,26 | 0,02 | 0,03 | 0,81 | 4,16E-01 | 6,06E-01 |
| LGALS8 | 2988,63 | 0,03 | 0,04 | 0,81 | 4,16E-01 | 6,06E-01 |
| HVCN1 | 1142,30 | 0,05 | 0,06 | 0,81 | 4,16E-01 | 6,06E-01 |
| AHCY | 171,73 | -0,06 | 0,08 | -0,81 | 4,17E-01 | 6,07E-01 |
| PIH1D1 | 730,30 | 0,03 | 0,04 | 0,81 | 4,17E-01 | 6,07E-01 |
| HERC3 | 5605,50 | -0,04 | 0,05 | -0,81 | 4,17E-01 | 6,07E-01 |
| SLC15A4 | 3517,83 | -0,04 | 0,05 | -0,81 | 4,17E-01 | 6,07E-01 |
| INKA2 | 341,39 | -0,06 | 0,08 | -0,81 | 4,17E-01 | 6,07E-01 |
| DGKK | 99,16 | -0,14 | 0,17 | -0,81 | 4,17E-01 | 6,07E-01 |
| TSPAN32 | 1122,38 | -0,07 | 0,08 | -0,81 | 4,17E-01 | 6,07E-01 |
| C19orf12 | 761,92 | 0,03 | 0,04 | 0,81 | 4,17E-01 | 6,07E-01 |
| CTNNBIP1 | 182,76 | -0,07 | 0,08 | -0,81 | 4,17E-01 | 6,08E-01 |
| PHLDB3 | 174,22 | -0,07 | 0,08 | -0,81 | 4,18E-01 | 6,08E-01 |
| TXNDC16 | 787,71 | 0,06 | 0,07 | 0,81 | 4,18E-01 | 6,08E-01 |
| OSBPL8 | 13807,45 | 0,08 | 0,10 | 0,81 | 4,18E-01 | 6,08E-01 |
| ZG16B | 27,52 | -0,11 | 0,14 | -0,81 | 4,18E-01 | 6,08E-01 |
| ASL | 289,01 | -0,04 | 0,05 | -0,81 | 4,18E-01 | 6,08E-01 |
| TRAFD1 | 2151,34 | -0,04 | 0,06 | -0,81 | 4,18E-01 | 6,08E-01 |
| CHST15 | 9242,52 | -0,06 | 0,08 | -0,81 | 4,18E-01 | 6,08E-01 |
| BICDL1 | 317,04 | -0,07 | 0,09 | -0,81 | 4,18E-01 | 6,08E-01 |
| NRBF2 | 1204,78 | 0,06 | 0,07 | 0,81 | 4,18E-01 | 6,08E-01 |
| SBF2 | 3730,59 | -0,05 | 0,06 | -0,81 | 4,18E-01 | 6,08E-01 |
| ROGDI | 434,97 | -0,07 | 0,09 | -0,81 | 4,18E-01 | 6,08E-01 |
| STN1 | 1142,36 | 0,03 | 0,04 | 0,81 | 4,19E-01 | 6,09E-01 |
| ZNF470 | 201,65 | 0,07 | 0,08 | 0,81 | 4,19E-01 | 6,09E-01 |
| BACH1 | 14066,03 | 0,05 | 0,07 | 0,81 | 4,19E-01 | 6,09E-01 |
| SRD5A1 | 550,31 | -0,04 | 0,05 | -0,81 | 4,19E-01 | 6,09E-01 |
| GPCPD1 | 10952,47 | 0,06 | 0,07 | 0,81 | 4,19E-01 | 6,09E-01 |
| MTMR11 | 339,03 | 0,06 | 0,07 | 0,81 | 4,19E-01 | 6,09E-01 |
| ADIPOR2 | 1459,20 | -0,03 | 0,03 | -0,81 | 4,19E-01 | 6,09E-01 |
| ATP6V1B2 | 14221,97 | -0,05 | 0,06 | -0,81 | 4,19E-01 | 6,09E-01 |
| ZFP69 | 172,36 | 0,04 | 0,04 | 0,81 | 4,20E-01 | 6,09E-01 |
| EME2 | 610,21 | -0,06 | 0,08 | -0,81 | 4,20E-01 | 6,09E-01 |
| SND1 | 2653,30 | 0,03 | 0,04 | 0,81 | 4,20E-01 | 6,09E-01 |
| ANXA7 | 2406,25 | 0,04 | 0,04 | 0,81 | 4,20E-01 | 6,09E-01 |
| UNG | 262,63 | 0,04 | 0,05 | 0,81 | 4,20E-01 | 6,09E-01 |

| | | | | | | |
|---|---|---|---|---|---|---|
| TAOK3 | 3767,13 | 0,03 | 0,03 | 0,81 | 4,20E-01 | 6,09E-01 |
| EZH1 | 2325,66 | -0,03 | 0,04 | -0,81 | 4,20E-01 | 6,10E-01 |
| ADRM1 | 400,58 | -0,04 | 0,06 | -0,81 | 4,20E-01 | 6,10E-01 |
| STBD1 | 43,60 | -0,08 | 0,10 | -0,81 | 4,20E-01 | 6,10E-01 |
| ZNF223 | 56,16 | 0,05 | 0,07 | 0,81 | 4,21E-01 | 6,10E-01 |
| SEPTIN11 | 1069,95 | -0,04 | 0,05 | -0,81 | 4,21E-01 | 6,10E-01 |
| ARHGAP15 | 7665,02 | 0,06 | 0,07 | 0,80 | 4,21E-01 | 6,11E-01 |
| MTA2 | 2107,03 | 0,03 | 0,04 | 0,80 | 4,21E-01 | 6,11E-01 |
| COQ3 | 69,31 | 0,05 | 0,06 | 0,80 | 4,21E-01 | 6,11E-01 |
| MORC4 | 202,86 | 0,05 | 0,06 | 0,80 | 4,21E-01 | 6,11E-01 |
| ZNF483 | 317,90 | 0,05 | 0,07 | 0,80 | 4,21E-01 | 6,11E-01 |
| ENSG000002 | 181,57 | -0,09 | 0,11 | -0,80 | 4,21E-01 | 6,11E-01 |
| SPATA6 | 346,36 | -0,06 | 0,07 | -0,80 | 4,22E-01 | 6,11E-01 |
| YWHAH | 406,60 | -0,05 | 0,07 | -0,80 | 4,22E-01 | 6,11E-01 |
| RELL1 | 1473,31 | -0,05 | 0,07 | -0,80 | 4,22E-01 | 6,11E-01 |
| C5orf63 | 405,21 | 0,05 | 0,06 | 0,80 | 4,22E-01 | 6,11E-01 |
| TLN1 | 14193,89 | -0,09 | 0,11 | -0,80 | 4,22E-01 | 6,11E-01 |
| KDM5D | 2504,13 | -0,21 | 0,27 | -0,80 | 4,22E-01 | 6,11E-01 |
| XYLT2 | 409,00 | -0,05 | 0,06 | -0,80 | 4,22E-01 | 6,11E-01 |
| ZNF445 | 2603,32 | -0,06 | 0,07 | -0,80 | 4,22E-01 | 6,11E-01 |
| TMEM129 | 323,81 | -0,06 | 0,07 | -0,80 | 4,22E-01 | 6,12E-01 |
| DUSP5 | 351,81 | 0,05 | 0,07 | 0,80 | 4,23E-01 | 6,12E-01 |
| CHRFAM7A | 24,76 | -0,12 | 0,15 | -0,80 | 4,23E-01 | 6,12E-01 |
| TLNRD1 | 531,00 | -0,05 | 0,06 | -0,80 | 4,23E-01 | 6,12E-01 |
| GNL1 | 863,35 | 0,03 | 0,03 | 0,80 | 4,23E-01 | 6,12E-01 |
| PRAM1 | 537,27 | -0,10 | 0,13 | -0,80 | 4,23E-01 | 6,12E-01 |
| PARVB | 234,13 | -0,09 | 0,11 | -0,80 | 4,23E-01 | 6,13E-01 |
| SUPT3H | 289,59 | 0,05 | 0,07 | 0,80 | 4,24E-01 | 6,13E-01 |
| DYNC2LI1 | 100,50 | 0,07 | 0,09 | 0,80 | 4,24E-01 | 6,13E-01 |
| ST6GALNAC6 | 275,96 | -0,06 | 0,08 | -0,80 | 4,24E-01 | 6,13E-01 |
| ATF5 | 82,37 | -0,08 | 0,10 | -0,80 | 4,24E-01 | 6,13E-01 |
| ARPC5L | 607,72 | 0,03 | 0,04 | 0,80 | 4,24E-01 | 6,13E-01 |
| SERPINB1 | 3799,95 | 0,05 | 0,06 | 0,80 | 4,24E-01 | 6,13E-01 |
| PRKG2 | 73,97 | -0,13 | 0,17 | -0,80 | 4,24E-01 | 6,13E-01 |
| P2RY12 | 218,82 | -0,09 | 0,11 | -0,80 | 4,24E-01 | 6,13E-01 |
| HOXB2 | 123,98 | -0,08 | 0,11 | -0,80 | 4,25E-01 | 6,14E-01 |
| ATXN2 | 1170,25 | -0,05 | 0,06 | -0,80 | 4,25E-01 | 6,14E-01 |
| C22orf39 | 760,85 | 0,02 | 0,03 | 0,80 | 4,26E-01 | 6,15E-01 |
| ANAPC15 | 133,53 | 0,04 | 0,05 | 0,80 | 4,26E-01 | 6,15E-01 |
| TMEM104 | 319,91 | -0,07 | 0,09 | -0,80 | 4,25E-01 | 6,15E-01 |
| AKAP10 | 2566,05 | 0,03 | 0,03 | 0,80 | 4,25E-01 | 6,15E-01 |
| ARHGAP22 | 161,42 | -0,10 | 0,13 | -0,80 | 4,25E-01 | 6,15E-01 |
| RANGAP1 | 676,12 | -0,05 | 0,07 | -0,80 | 4,26E-01 | 6,15E-01 |
| SHISAL2A | 279,27 | -0,08 | 0,10 | -0,80 | 4,26E-01 | 6,15E-01 |
| FAM120B | 2118,27 | -0,04 | 0,05 | -0,80 | 4,26E-01 | 6,15E-01 |
| RAB35 | 1047,97 | -0,04 | 0,05 | -0,80 | 4,26E-01 | 6,15E-01 |

| | | | | | | |
|---|---|---|---|---|---|---|
| CRTC2 | 511,25 | -0,06 | 0,08 | -0,80 | 4,26E-01 | 6,15E-01 |
| DNAJB12 | 1316,40 | -0,04 | 0,05 | -0,80 | 4,26E-01 | 6,15E-01 |
| LCMT1 | 241,11 | -0,04 | 0,05 | -0,80 | 4,26E-01 | 6,15E-01 |
| CNTRL | 3567,22 | 0,04 | 0,05 | 0,80 | 4,26E-01 | 6,15E-01 |
| MAP3K21 | 109,58 | -0,08 | 0,10 | -0,80 | 4,26E-01 | 6,15E-01 |
| AMIGO2 | 244,45 | 0,05 | 0,06 | 0,80 | 4,27E-01 | 6,15E-01 |
| TLE3 | 5089,14 | -0,07 | 0,09 | -0,79 | 4,27E-01 | 6,15E-01 |
| VENTX | 95,98 | -0,10 | 0,13 | -0,79 | 4,27E-01 | 6,15E-01 |
| IKZF3 | 4793,11 | 0,05 | 0,07 | 0,79 | 4,27E-01 | 6,15E-01 |
| ATP6V0A1 | 1532,42 | -0,04 | 0,05 | -0,79 | 4,27E-01 | 6,15E-01 |
| OSBPL11 | 2243,81 | 0,04 | 0,05 | 0,79 | 4,27E-01 | 6,15E-01 |
| TENM1 | 526,82 | 0,08 | 0,10 | 0,79 | 4,27E-01 | 6,15E-01 |
| GYPE | 75,11 | -0,09 | 0,11 | -0,79 | 4,27E-01 | 6,15E-01 |
| PLXNB2 | 927,92 | -0,11 | 0,14 | -0,79 | 4,27E-01 | 6,16E-01 |
| COPE | 1017,41 | -0,04 | 0,05 | -0,79 | 4,27E-01 | 6,16E-01 |
| PPP1R3D | 1120,38 | -0,04 | 0,05 | -0,79 | 4,28E-01 | 6,16E-01 |
| NAALADL1 | 156,20 | -0,05 | 0,06 | -0,79 | 4,28E-01 | 6,16E-01 |
| CNEP1R1 | 1373,40 | 0,05 | 0,06 | 0,79 | 4,28E-01 | 6,16E-01 |
| IRF5 | 595,81 | -0,05 | 0,07 | -0,79 | 4,28E-01 | 6,16E-01 |
| TYW1 | 591,15 | 0,03 | 0,04 | 0,79 | 4,28E-01 | 6,16E-01 |
| GPR34 | 148,26 | 0,11 | 0,14 | 0,79 | 4,28E-01 | 6,16E-01 |
| CCHCR1 | 257,61 | -0,04 | 0,05 | -0,79 | 4,28E-01 | 6,16E-01 |
| CCNY | 3546,25 | 0,03 | 0,04 | 0,79 | 4,28E-01 | 6,16E-01 |
| HP | 99,10 | -0,14 | 0,17 | -0,79 | 4,28E-01 | 6,16E-01 |
| CYB5D2 | 177,64 | -0,03 | 0,04 | -0,79 | 4,28E-01 | 6,16E-01 |
| ZNF71 | 283,98 | -0,04 | 0,05 | -0,79 | 4,28E-01 | 6,16E-01 |
| SYNE1 | 18887,88 | -0,05 | 0,07 | -0,79 | 4,28E-01 | 6,16E-01 |
| HM13 | 1200,18 | -0,07 | 0,09 | -0,79 | 4,29E-01 | 6,17E-01 |
| ENSG000002 | 142,43 | -0,08 | 0,10 | -0,79 | 4,29E-01 | 6,17E-01 |
| ADAM22 | 348,10 | 0,06 | 0,08 | 0,79 | 4,29E-01 | 6,17E-01 |
| PCYT1A | 2770,52 | -0,03 | 0,04 | -0,79 | 4,29E-01 | 6,17E-01 |
| ZNF652 | 4189,37 | -0,04 | 0,05 | -0,79 | 4,29E-01 | 6,17E-01 |
| GPRASP1 | 1160,42 | 0,05 | 0,07 | 0,79 | 4,29E-01 | 6,17E-01 |
| POMC | 68,44 | -0,09 | 0,12 | -0,79 | 4,30E-01 | 6,18E-01 |
| EIF4A3 | 1066,25 | 0,04 | 0,05 | 0,79 | 4,30E-01 | 6,18E-01 |
| MICB | 764,23 | -0,04 | 0,05 | -0,79 | 4,30E-01 | 6,18E-01 |
| RHEX | 40,13 | -0,14 | 0,18 | -0,79 | 4,30E-01 | 6,18E-01 |
| C2orf42 | 351,11 | -0,02 | 0,03 | -0,79 | 4,30E-01 | 6,18E-01 |
| ZBED10P | 185,36 | -0,06 | 0,08 | -0,79 | 4,30E-01 | 6,18E-01 |
| CTSF | 118,44 | -0,06 | 0,08 | -0,79 | 4,30E-01 | 6,18E-01 |
| CCNB1 | 96,22 | -0,07 | 0,09 | -0,79 | 4,30E-01 | 6,18E-01 |
| SUMF1 | 1363,12 | 0,03 | 0,03 | 0,79 | 4,30E-01 | 6,18E-01 |
| CD320 | 86,80 | -0,08 | 0,11 | -0,79 | 4,30E-01 | 6,18E-01 |
| SF1 | 3784,79 | -0,05 | 0,06 | -0,79 | 4,31E-01 | 6,18E-01 |
| MAF | 867,08 | 0,06 | 0,07 | 0,79 | 4,31E-01 | 6,19E-01 |
| TNFRSF17 | 34,29 | 0,13 | 0,16 | 0,79 | 4,31E-01 | 6,19E-01 |

| | | | | | | |
|---|---|---|---|---|---|---|
| CNBP | 8150,82 | 0,04 | 0,06 | 0,79 | 4,31E-01 | 6,19E-01 |
| AAMDC | 88,68 | -0,04 | 0,06 | -0,79 | 4,31E-01 | 6,19E-01 |
| ADGRE3 | 2443,03 | -0,06 | 0,07 | -0,79 | 4,31E-01 | 6,19E-01 |
| ITGA6 | 2592,62 | 0,05 | 0,06 | 0,79 | 4,32E-01 | 6,19E-01 |
| SPRY1 | 53,74 | -0,10 | 0,13 | -0,79 | 4,32E-01 | 6,19E-01 |
| IVD | 852,76 | -0,05 | 0,06 | -0,79 | 4,32E-01 | 6,19E-01 |
| IBA57 | 595,89 | -0,05 | 0,07 | -0,79 | 4,32E-01 | 6,20E-01 |
| C11orf71 | 305,29 | 0,05 | 0,07 | 0,79 | 4,32E-01 | 6,20E-01 |
| NEU1 | 792,89 | -0,04 | 0,05 | -0,79 | 4,32E-01 | 6,20E-01 |
| ELF2 | 4876,79 | 0,04 | 0,05 | 0,79 | 4,32E-01 | 6,20E-01 |
| LAMTOR1 | 1807,36 | -0,04 | 0,05 | -0,79 | 4,32E-01 | 6,20E-01 |
| TEF | 206,50 | -0,05 | 0,06 | -0,78 | 4,33E-01 | 6,20E-01 |
| COX19 | 1189,44 | 0,03 | 0,03 | 0,78 | 4,33E-01 | 6,20E-01 |
| FAHD2B | 69,20 | -0,06 | 0,07 | -0,78 | 4,33E-01 | 6,21E-01 |
| RAC1 | 2783,98 | -0,04 | 0,05 | -0,78 | 4,33E-01 | 6,21E-01 |
| STX4 | 931,95 | -0,04 | 0,05 | -0,78 | 4,33E-01 | 6,21E-01 |
| THYN1 | 382,94 | 0,03 | 0,04 | 0,78 | 4,33E-01 | 6,21E-01 |
| RCAN3 | 2773,67 | 0,06 | 0,07 | 0,78 | 4,33E-01 | 6,21E-01 |
| POLR3G | 44,36 | -0,09 | 0,11 | -0,78 | 4,34E-01 | 6,21E-01 |
| SARM1 | 537,79 | -0,07 | 0,09 | -0,78 | 4,34E-01 | 6,22E-01 |
| ELMOD3 | 796,49 | -0,02 | 0,03 | -0,78 | 4,34E-01 | 6,22E-01 |
| BCL7A | 121,21 | -0,08 | 0,11 | -0,78 | 4,34E-01 | 6,22E-01 |
| C12orf43 | 259,05 | -0,03 | 0,04 | -0,78 | 4,35E-01 | 6,22E-01 |
| BIN3 | 1265,56 | -0,04 | 0,05 | -0,78 | 4,35E-01 | 6,22E-01 |
| CDK10 | 511,62 | -0,06 | 0,08 | -0,78 | 4,35E-01 | 6,23E-01 |
| UBN1 | 3942,31 | -0,07 | 0,09 | -0,78 | 4,35E-01 | 6,23E-01 |
| RXRA | 4909,69 | -0,07 | 0,09 | -0,78 | 4,35E-01 | 6,23E-01 |
| PRPF8 | 4206,42 | -0,05 | 0,07 | -0,78 | 4,35E-01 | 6,23E-01 |
| RAD18 | 472,15 | 0,04 | 0,05 | 0,78 | 4,35E-01 | 6,23E-01 |
| HLA-E | 62242,19 | -0,04 | 0,05 | -0,78 | 4,36E-01 | 6,23E-01 |
| MYOCOS | 27,69 | -0,09 | 0,11 | -0,78 | 4,36E-01 | 6,23E-01 |
| GYG1 | 753,04 | 0,04 | 0,05 | 0,78 | 4,36E-01 | 6,23E-01 |
| HLA-DOB | 213,76 | 0,09 | 0,11 | 0,78 | 4,36E-01 | 6,23E-01 |
| ZXDB | 263,33 | 0,04 | 0,05 | 0,78 | 4,36E-01 | 6,23E-01 |
| SPCS1 | 359,81 | 0,03 | 0,04 | 0,78 | 4,36E-01 | 6,23E-01 |
| TMEM165 | 3021,04 | -0,03 | 0,04 | -0,78 | 4,36E-01 | 6,23E-01 |
| ST14 | 253,48 | -0,08 | 0,10 | -0,78 | 4,36E-01 | 6,24E-01 |
| ZMIZ1 | 1463,39 | -0,07 | 0,09 | -0,78 | 4,37E-01 | 6,24E-01 |
| GSTM4 | 197,51 | -0,09 | 0,11 | -0,78 | 4,37E-01 | 6,24E-01 |
| NOXA1 | 123,04 | -0,07 | 0,09 | -0,78 | 4,37E-01 | 6,24E-01 |
| SCMH1 | 388,24 | -0,04 | 0,05 | -0,78 | 4,37E-01 | 6,24E-01 |
| NDRG1 | 1840,18 | -0,04 | 0,05 | -0,78 | 4,37E-01 | 6,24E-01 |
| ZBTB34 | 3428,99 | -0,04 | 0,06 | -0,78 | 4,37E-01 | 6,24E-01 |
| ZNF688 | 346,52 | -0,03 | 0,04 | -0,78 | 4,37E-01 | 6,24E-01 |
| CDK5RAP3 | 1102,16 | -0,04 | 0,05 | -0,78 | 4,37E-01 | 6,24E-01 |
| EIF2B5 | 1169,55 | 0,02 | 0,03 | 0,78 | 4,37E-01 | 6,24E-01 |

| | | | | | | |
|---|---|---|---|---|---|---|
| IER5L | 188,17 | -0,06 | 0,08 | -0,78 | 4,37E-01 | 6,25E-01 |
| DNAJC3 | 3547,59 | 0,04 | 0,06 | 0,78 | 4,38E-01 | 6,25E-01 |
| DNASE1 | 2018,03 | -0,03 | 0,04 | -0,78 | 4,38E-01 | 6,25E-01 |
| TNRC6B | 10418,41 | -0,04 | 0,05 | -0,78 | 4,38E-01 | 6,25E-01 |
| FAM118A | 1298,22 | 0,09 | 0,12 | 0,78 | 4,38E-01 | 6,25E-01 |
| FAS | 2260,74 | 0,06 | 0,08 | 0,78 | 4,38E-01 | 6,25E-01 |
| MBNL3 | 19715,85 | -0,10 | 0,13 | -0,78 | 4,38E-01 | 6,25E-01 |
| MAP7D3 | 301,34 | 0,03 | 0,04 | 0,77 | 4,38E-01 | 6,25E-01 |
| ENSG000002 | 51,20 | -0,06 | 0,08 | -0,77 | 4,38E-01 | 6,25E-01 |
| ARK2C | 154,86 | -0,12 | 0,15 | -0,77 | 4,38E-01 | 6,25E-01 |
| ILVBL | 269,56 | -0,04 | 0,06 | -0,77 | 4,39E-01 | 6,26E-01 |
| KLC1 | 3046,89 | -0,05 | 0,06 | -0,77 | 4,39E-01 | 6,26E-01 |
| AGPAT2 | 493,20 | -0,04 | 0,06 | -0,77 | 4,39E-01 | 6,26E-01 |
| H2AZ2 | 2601,72 | 0,03 | 0,03 | 0,77 | 4,39E-01 | 6,26E-01 |
| MYO15B | 2099,77 | -0,08 | 0,11 | -0,77 | 4,39E-01 | 6,26E-01 |
| RDH13 | 255,53 | -0,05 | 0,07 | -0,77 | 4,40E-01 | 6,26E-01 |
| RIC3 | 681,67 | 0,06 | 0,08 | 0,77 | 4,40E-01 | 6,26E-01 |
| ZNF726 | 96,08 | 0,06 | 0,08 | 0,77 | 4,39E-01 | 6,26E-01 |
| GALNT11 | 958,28 | 0,03 | 0,03 | 0,77 | 4,40E-01 | 6,26E-01 |
| AKTIP | 1479,31 | 0,04 | 0,05 | 0,77 | 4,39E-01 | 6,26E-01 |
| CD82 | 1748,73 | -0,04 | 0,06 | -0,77 | 4,39E-01 | 6,26E-01 |
| NLGN4Y | 32,80 | -0,11 | 0,14 | -0,77 | 4,39E-01 | 6,26E-01 |
| DOCK7 | 568,06 | 0,05 | 0,07 | 0,77 | 4,39E-01 | 6,26E-01 |
| LASP1 | 3501,38 | -0,07 | 0,08 | -0,77 | 4,40E-01 | 6,26E-01 |
| MLXIP | 1251,46 | -0,09 | 0,12 | -0,77 | 4,40E-01 | 6,27E-01 |
| PARVG | 3714,12 | -0,04 | 0,05 | -0,77 | 4,40E-01 | 6,27E-01 |
| ZNF687 | 681,31 | -0,06 | 0,08 | -0,77 | 4,41E-01 | 6,27E-01 |
| TAB1 | 347,73 | -0,06 | 0,08 | -0,77 | 4,41E-01 | 6,28E-01 |
| PPP2CB | 611,06 | 0,03 | 0,04 | 0,77 | 4,41E-01 | 6,28E-01 |
| KIAA0825 | 1306,42 | -0,06 | 0,08 | -0,77 | 4,41E-01 | 6,28E-01 |
| AMMECR1L | 688,02 | -0,02 | 0,03 | -0,77 | 4,41E-01 | 6,28E-01 |
| MAPRE1 | 3272,44 | 0,04 | 0,05 | 0,77 | 4,41E-01 | 6,28E-01 |
| SNX10 | 3004,56 | 0,06 | 0,07 | 0,77 | 4,41E-01 | 6,28E-01 |
| NRM | 62,61 | -0,05 | 0,07 | -0,77 | 4,42E-01 | 6,28E-01 |
| LIPN | 470,57 | 0,09 | 0,12 | 0,77 | 4,42E-01 | 6,28E-01 |
| CNR1 | 80,31 | -0,11 | 0,14 | -0,77 | 4,42E-01 | 6,28E-01 |
| IL18R1 | 1405,47 | -0,06 | 0,08 | -0,77 | 4,42E-01 | 6,28E-01 |
| EXT1 | 778,08 | -0,04 | 0,05 | -0,77 | 4,42E-01 | 6,28E-01 |
| RCOR3 | 2450,41 | 0,03 | 0,04 | 0,77 | 4,42E-01 | 6,29E-01 |
| STING1 | 504,35 | -0,06 | 0,08 | -0,77 | 4,42E-01 | 6,29E-01 |
| TRIM11 | 905,25 | -0,05 | 0,07 | -0,77 | 4,43E-01 | 6,29E-01 |
| YBEY | 219,49 | 0,05 | 0,06 | 0,77 | 4,43E-01 | 6,29E-01 |
| RPA1 | 1055,27 | -0,04 | 0,05 | -0,77 | 4,43E-01 | 6,29E-01 |
| YWHAZ | 15848,40 | 0,03 | 0,04 | 0,77 | 4,43E-01 | 6,29E-01 |
| KDM5C | 3113,56 | -0,04 | 0,05 | -0,77 | 4,43E-01 | 6,29E-01 |
| SDCCAG8 | 1096,43 | 0,03 | 0,04 | 0,77 | 4,43E-01 | 6,29E-01 |

| | | | | | | |
|---|---|---|---|---|---|---|
| BTN3A1 | 6892,73 | -0,05 | 0,06 | -0,77 | 4,43E-01 | 6,29E-01 |
| SERPINB6 | 1507,17 | -0,04 | 0,06 | -0,77 | 4,43E-01 | 6,29E-01 |
| NPIPA1 | 53,23 | -0,09 | 0,12 | -0,77 | 4,43E-01 | 6,29E-01 |
| ACAA2 | 582,19 | 0,04 | 0,06 | 0,77 | 4,43E-01 | 6,29E-01 |
| PRF1 | 1787,15 | -0,08 | 0,11 | -0,77 | 4,43E-01 | 6,29E-01 |
| TCEAL4 | 147,28 | 0,06 | 0,07 | 0,77 | 4,44E-01 | 6,30E-01 |
| TCTA | 564,41 | -0,03 | 0,04 | -0,77 | 4,44E-01 | 6,30E-01 |
| RCC1L | 526,50 | 0,03 | 0,04 | 0,77 | 4,44E-01 | 6,30E-01 |
| FAM185A | 306,79 | 0,04 | 0,05 | 0,77 | 4,43E-01 | 6,30E-01 |
| ANXA2R | 555,77 | 0,05 | 0,06 | 0,77 | 4,44E-01 | 6,30E-01 |
| ALKBH5 | 2065,78 | -0,03 | 0,04 | -0,77 | 4,44E-01 | 6,30E-01 |
| TMED3 | 879,30 | -0,03 | 0,04 | -0,76 | 4,44E-01 | 6,30E-01 |
| ZMYND8 | 2073,81 | -0,03 | 0,04 | -0,76 | 4,44E-01 | 6,30E-01 |
| KIF13A | 3311,92 | -0,05 | 0,07 | -0,76 | 4,44E-01 | 6,30E-01 |
| APOL3 | 4008,13 | 0,04 | 0,05 | 0,76 | 4,44E-01 | 6,30E-01 |
| JAML | 23753,63 | -0,05 | 0,06 | -0,76 | 4,45E-01 | 6,30E-01 |
| NAB2 | 219,31 | 0,05 | 0,06 | 0,76 | 4,45E-01 | 6,31E-01 |
| RUNX3 | 1113,43 | -0,07 | 0,09 | -0,76 | 4,45E-01 | 6,31E-01 |
| YOD1 | 12527,74 | -0,10 | 0,13 | -0,76 | 4,45E-01 | 6,31E-01 |
| MPZL1 | 2746,22 | -0,07 | 0,09 | -0,76 | 4,45E-01 | 6,31E-01 |
| HSPBAP1 | 1706,85 | 0,04 | 0,05 | 0,76 | 4,45E-01 | 6,31E-01 |
| C11orf54 | 693,65 | 0,03 | 0,04 | 0,76 | 4,45E-01 | 6,31E-01 |
| CRYL1 | 339,50 | -0,03 | 0,04 | -0,76 | 4,45E-01 | 6,31E-01 |
| MED30 | 480,28 | 0,05 | 0,06 | 0,76 | 4,46E-01 | 6,31E-01 |
| GCSAM | 389,18 | 0,06 | 0,07 | 0,76 | 4,46E-01 | 6,32E-01 |
| ERGIC3 | 755,34 | -0,04 | 0,05 | -0,76 | 4,46E-01 | 6,32E-01 |
| NIPA1 | 594,18 | 0,04 | 0,05 | 0,76 | 4,46E-01 | 6,32E-01 |
| DET1 | 206,88 | 0,04 | 0,06 | 0,76 | 4,46E-01 | 6,32E-01 |
| UBE2T | 36,07 | -0,07 | 0,09 | -0,76 | 4,46E-01 | 6,32E-01 |
| ARL2BP | 996,33 | 0,03 | 0,04 | 0,76 | 4,47E-01 | 6,32E-01 |
| SETD4 | 667,56 | 0,03 | 0,03 | 0,76 | 4,47E-01 | 6,32E-01 |
| FLII | 3387,47 | -0,05 | 0,07 | -0,76 | 4,47E-01 | 6,32E-01 |
| BEX5 | 30,16 | -0,08 | 0,11 | -0,76 | 4,47E-01 | 6,32E-01 |
| RBM23 | 3824,76 | -0,04 | 0,05 | -0,76 | 4,47E-01 | 6,32E-01 |
| AKR7A2 | 369,04 | 0,03 | 0,04 | 0,76 | 4,47E-01 | 6,32E-01 |
| STT3A | 1345,22 | 0,02 | 0,03 | 0,76 | 4,47E-01 | 6,32E-01 |
| GDF11 | 403,30 | -0,04 | 0,06 | -0,76 | 4,47E-01 | 6,32E-01 |
| NR3C2 | 902,90 | 0,05 | 0,06 | 0,76 | 4,47E-01 | 6,32E-01 |
| FAM120A | 3619,33 | -0,03 | 0,04 | -0,76 | 4,47E-01 | 6,33E-01 |
| ZFAT | 926,96 | -0,03 | 0,04 | -0,76 | 4,48E-01 | 6,33E-01 |
| ABCF3 | 804,31 | -0,03 | 0,04 | -0,76 | 4,48E-01 | 6,33E-01 |
| SDF2L1 | 233,82 | -0,04 | 0,06 | -0,76 | 4,48E-01 | 6,33E-01 |
| TMEM107 | 102,33 | -0,04 | 0,05 | -0,76 | 4,48E-01 | 6,33E-01 |
| CHMP4B | 775,34 | -0,05 | 0,06 | -0,76 | 4,48E-01 | 6,33E-01 |
| PPCDC | 935,62 | -0,07 | 0,09 | -0,76 | 4,48E-01 | 6,33E-01 |
| FMO4 | 135,88 | 0,04 | 0,05 | 0,76 | 4,48E-01 | 6,34E-01 |

| | | | | | | |
|---|---|---|---|---|---|---|
| DLEU7 | 49,47 | -0,08 | 0,11 | -0,76 | 4,48E-01 | 6,34E-01 |
| ZNF280C | 217,25 | 0,03 | 0,05 | 0,76 | 4,49E-01 | 6,34E-01 |
| NFATC2 | 1401,28 | -0,05 | 0,06 | -0,76 | 4,49E-01 | 6,34E-01 |
| AGAP6 | 444,10 | 0,05 | 0,06 | 0,76 | 4,49E-01 | 6,34E-01 |
| STEAP1B | 118,15 | -0,10 | 0,14 | -0,76 | 4,49E-01 | 6,34E-01 |
| MS4A7 | 2580,39 | 0,06 | 0,08 | 0,76 | 4,49E-01 | 6,34E-01 |
| RFTN1 | 1202,88 | 0,03 | 0,04 | 0,76 | 4,49E-01 | 6,34E-01 |
| NR2C2 | 3315,07 | -0,04 | 0,05 | -0,76 | 4,49E-01 | 6,34E-01 |
| TRAPPC3L | 41,84 | -0,11 | 0,14 | -0,76 | 4,49E-01 | 6,34E-01 |
| SAE1 | 787,51 | 0,03 | 0,04 | 0,76 | 4,50E-01 | 6,35E-01 |
| SSTR3 | 165,54 | -0,08 | 0,11 | -0,76 | 4,50E-01 | 6,35E-01 |
| ARL16 | 370,25 | 0,05 | 0,06 | 0,76 | 4,50E-01 | 6,35E-01 |
| PDGFD | 306,74 | 0,08 | 0,11 | 0,76 | 4,50E-01 | 6,35E-01 |
| CISD2 | 1272,82 | -0,08 | 0,11 | -0,75 | 4,50E-01 | 6,35E-01 |
| TNKS | 2431,70 | 0,03 | 0,04 | 0,75 | 4,50E-01 | 6,35E-01 |
| ESRRA | 1472,52 | -0,04 | 0,05 | -0,75 | 4,51E-01 | 6,36E-01 |
| DNASE1L1 | 750,31 | -0,04 | 0,05 | -0,75 | 4,51E-01 | 6,36E-01 |
| SEC14L5 | 86,07 | -0,15 | 0,20 | -0,75 | 4,51E-01 | 6,36E-01 |
| ARMC6 | 303,70 | -0,03 | 0,04 | -0,75 | 4,51E-01 | 6,36E-01 |
| GANAB | 1666,56 | -0,05 | 0,07 | -0,75 | 4,51E-01 | 6,36E-01 |
| RRS1 | 245,55 | 0,03 | 0,04 | 0,75 | 4,51E-01 | 6,36E-01 |
| ATG2B | 3350,42 | 0,03 | 0,04 | 0,75 | 4,51E-01 | 6,36E-01 |
| KLRC1 | 358,13 | 0,11 | 0,15 | 0,75 | 4,52E-01 | 6,36E-01 |
| CCSAP | 1600,35 | 0,05 | 0,07 | 0,75 | 4,52E-01 | 6,36E-01 |
| ZNF8 | 364,56 | -0,05 | 0,06 | -0,75 | 4,52E-01 | 6,36E-01 |
| MRC1 | 66,54 | -0,13 | 0,18 | -0,75 | 4,51E-01 | 6,36E-01 |
| DAZAP1 | 1704,83 | -0,05 | 0,06 | -0,75 | 4,52E-01 | 6,36E-01 |
| BCAP31 | 1111,16 | -0,03 | 0,05 | -0,75 | 4,52E-01 | 6,36E-01 |
| PRMT1 | 442,98 | 0,03 | 0,04 | 0,75 | 4,52E-01 | 6,36E-01 |
| PFDN1 | 164,36 | 0,03 | 0,04 | 0,75 | 4,52E-01 | 6,36E-01 |
| DCP1A | 894,62 | 0,02 | 0,03 | 0,75 | 4,52E-01 | 6,36E-01 |
| RUSC2 | 121,79 | -0,09 | 0,12 | -0,75 | 4,52E-01 | 6,37E-01 |
| FBXW4 | 1486,20 | -0,03 | 0,04 | -0,75 | 4,52E-01 | 6,37E-01 |
| SLC43A2 | 3057,06 | -0,07 | 0,09 | -0,75 | 4,52E-01 | 6,37E-01 |
| KANSL3 | 1400,96 | -0,05 | 0,06 | -0,75 | 4,53E-01 | 6,37E-01 |
| EPHA1 | 193,66 | -0,07 | 0,10 | -0,75 | 4,53E-01 | 6,37E-01 |
| ZMAT3 | 1466,17 | 0,04 | 0,05 | 0,75 | 4,53E-01 | 6,38E-01 |
| SLC45A4 | 3497,38 | -0,07 | 0,09 | -0,75 | 4,54E-01 | 6,38E-01 |
| KMT2C | 12246,48 | -0,04 | 0,05 | -0,75 | 4,54E-01 | 6,38E-01 |
| BTBD3 | 194,71 | 0,04 | 0,06 | 0,75 | 4,54E-01 | 6,38E-01 |
| PSMC4 | 925,88 | 0,03 | 0,04 | 0,75 | 4,54E-01 | 6,38E-01 |
| CRIP1 | 264,63 | -0,05 | 0,06 | -0,75 | 4,54E-01 | 6,38E-01 |
| GCLC | 1319,91 | -0,05 | 0,07 | -0,75 | 4,54E-01 | 6,38E-01 |
| RALGDS | 498,81 | -0,06 | 0,09 | -0,75 | 4,54E-01 | 6,38E-01 |
| TRAF3IP3 | 5265,09 | 0,03 | 0,04 | 0,75 | 4,54E-01 | 6,38E-01 |
| EMSY | 2122,37 | 0,03 | 0,04 | 0,75 | 4,54E-01 | 6,38E-01 |

| | | | | | | |
|---|---|---|---|---|---|---|
| TTC1 | 979,75 | 0,03 | 0,05 | 0,75 | 4,54E-01 | 6,38E-01 |
| LBR | 8412,64 | 0,05 | 0,06 | 0,75 | 4,54E-01 | 6,39E-01 |
| SLC22A18 | 353,16 | -0,07 | 0,09 | -0,75 | 4,54E-01 | 6,39E-01 |
| RANBP17 | 94,39 | -0,13 | 0,17 | -0,75 | 4,55E-01 | 6,39E-01 |
| IFNGR2 | 4017,30 | -0,05 | 0,06 | -0,75 | 4,55E-01 | 6,39E-01 |
| BRICD5 | 140,37 | -0,05 | 0,07 | -0,75 | 4,55E-01 | 6,39E-01 |
| COMMD5 | 544,08 | 0,02 | 0,03 | 0,75 | 4,55E-01 | 6,39E-01 |
| POFUT1 | 493,70 | -0,04 | 0,05 | -0,75 | 4,55E-01 | 6,39E-01 |
| PAQR8 | 614,75 | 0,04 | 0,05 | 0,75 | 4,55E-01 | 6,39E-01 |
| SNU13 | 1069,50 | 0,02 | 0,03 | 0,75 | 4,55E-01 | 6,39E-01 |
| C14orf119 | 915,97 | 0,03 | 0,04 | 0,75 | 4,55E-01 | 6,39E-01 |
| PPP1R9B | 2513,46 | -0,07 | 0,09 | -0,75 | 4,55E-01 | 6,39E-01 |
| FADS1 | 350,77 | -0,06 | 0,07 | -0,75 | 4,55E-01 | 6,39E-01 |
| TAF1 | 3344,51 | 0,03 | 0,04 | 0,75 | 4,55E-01 | 6,39E-01 |
| PDE5A | 917,81 | -0,08 | 0,11 | -0,75 | 4,56E-01 | 6,39E-01 |
| HERPUD1 | 1897,35 | 0,02 | 0,03 | 0,75 | 4,56E-01 | 6,39E-01 |
| FAM229B | 33,00 | -0,10 | 0,14 | -0,75 | 4,56E-01 | 6,39E-01 |
| WDR27 | 864,68 | -0,05 | 0,07 | -0,75 | 4,56E-01 | 6,39E-01 |
| PTX3 | 50,46 | -0,08 | 0,11 | -0,75 | 4,56E-01 | 6,39E-01 |
| CAPS | 377,28 | -0,05 | 0,07 | -0,75 | 4,56E-01 | 6,39E-01 |
| EFHC1 | 418,06 | -0,05 | 0,07 | -0,75 | 4,56E-01 | 6,39E-01 |
| FAM78A | 1742,76 | -0,04 | 0,06 | -0,75 | 4,56E-01 | 6,39E-01 |
| MAN1B1 | 1309,36 | 0,03 | 0,04 | 0,75 | 4,56E-01 | 6,39E-01 |
| SLC16A3 | 7953,69 | -0,06 | 0,07 | -0,75 | 4,56E-01 | 6,40E-01 |
| TGFBI | 1810,58 | 0,05 | 0,07 | 0,74 | 4,56E-01 | 6,40E-01 |
| RCOR1 | 2121,12 | -0,05 | 0,06 | -0,74 | 4,57E-01 | 6,40E-01 |
| ZNF629 | 156,24 | -0,05 | 0,07 | -0,74 | 4,57E-01 | 6,40E-01 |
| ZNF317 | 1249,15 | 0,02 | 0,03 | 0,74 | 4,57E-01 | 6,40E-01 |
| BAK1 | 622,30 | -0,04 | 0,05 | -0,74 | 4,57E-01 | 6,40E-01 |
| ENSG000002 | 51,75 | -0,05 | 0,07 | -0,74 | 4,57E-01 | 6,40E-01 |
| TBC1D14 | 4964,49 | -0,05 | 0,06 | -0,74 | 4,57E-01 | 6,40E-01 |
| POLH | 536,90 | 0,03 | 0,04 | 0,74 | 4,57E-01 | 6,40E-01 |
| LRRFIP1 | 13393,41 | -0,03 | 0,04 | -0,74 | 4,57E-01 | 6,40E-01 |
| ARHGAP24 | 1033,71 | 0,06 | 0,08 | 0,74 | 4,57E-01 | 6,40E-01 |
| EXPH5 | 145,19 | -0,07 | 0,09 | -0,74 | 4,57E-01 | 6,40E-01 |
| NLRC3 | 2966,87 | -0,05 | 0,06 | -0,74 | 4,57E-01 | 6,40E-01 |
| ATIC | 419,70 | 0,04 | 0,05 | 0,74 | 4,57E-01 | 6,40E-01 |
| WNT16 | 51,61 | 0,13 | 0,17 | 0,74 | 4,58E-01 | 6,40E-01 |
| SPOP | 2633,29 | 0,03 | 0,04 | 0,74 | 4,58E-01 | 6,40E-01 |
| ARMCX1 | 50,78 | -0,06 | 0,08 | -0,74 | 4,58E-01 | 6,41E-01 |
| RTKN2 | 455,71 | 0,08 | 0,11 | 0,74 | 4,58E-01 | 6,41E-01 |
| ITGB1 | 7309,50 | 0,05 | 0,07 | 0,74 | 4,58E-01 | 6,41E-01 |
| C11orf21 | 3071,22 | -0,04 | 0,06 | -0,74 | 4,59E-01 | 6,42E-01 |
| OSM | 146,45 | -0,07 | 0,10 | -0,74 | 4,59E-01 | 6,42E-01 |
| NPDC1 | 101,25 | -0,11 | 0,14 | -0,74 | 4,59E-01 | 6,42E-01 |
| AIM2 | 447,02 | 0,07 | 0,10 | 0,74 | 4,59E-01 | 6,42E-01 |

| | | | | | | |
|---|---|---|---|---|---|---|
| SMCR8 | 2129,82 | -0,04 | 0,05 | -0,74 | 4,59E-01 | 6,42E-01 |
| TMOD2 | 2536,77 | 0,05 | 0,06 | 0,74 | 4,59E-01 | 6,42E-01 |
| RHOH | 2940,69 | 0,04 | 0,05 | 0,74 | 4,59E-01 | 6,42E-01 |
| UXS1 | 1339,86 | 0,03 | 0,04 | 0,74 | 4,59E-01 | 6,42E-01 |
| MAP2K4 | 2560,81 | 0,04 | 0,06 | 0,74 | 4,59E-01 | 6,42E-01 |
| IL1RL1 | 475,65 | -0,11 | 0,15 | -0,74 | 4,60E-01 | 6,42E-01 |
| CEP41 | 225,23 | -0,05 | 0,07 | -0,74 | 4,60E-01 | 6,42E-01 |
| DAAM1 | 500,85 | 0,03 | 0,05 | 0,74 | 4,60E-01 | 6,43E-01 |
| CLCF1 | 54,39 | -0,07 | 0,10 | -0,74 | 4,60E-01 | 6,43E-01 |
| GALNT7 | 1426,08 | 0,04 | 0,05 | 0,74 | 4,60E-01 | 6,43E-01 |
| ABL2 | 1143,80 | -0,03 | 0,04 | -0,74 | 4,60E-01 | 6,43E-01 |
| PAM16 | 200,53 | -0,08 | 0,11 | -0,74 | 4,60E-01 | 6,43E-01 |
| CHMP1B | 3597,93 | 0,05 | 0,07 | 0,74 | 4,61E-01 | 6,43E-01 |
| FLVCR1 | 984,46 | -0,04 | 0,06 | -0,74 | 4,61E-01 | 6,43E-01 |
| TYMP | 5388,48 | -0,07 | 0,09 | -0,74 | 4,61E-01 | 6,43E-01 |
| PLEKHG3 | 5770,96 | -0,06 | 0,08 | -0,74 | 4,61E-01 | 6,43E-01 |
| FUS | 5178,83 | -0,05 | 0,06 | -0,74 | 4,61E-01 | 6,43E-01 |
| DDI2 | 2199,54 | 0,02 | 0,03 | 0,74 | 4,61E-01 | 6,43E-01 |
| TAPBPL | 1014,81 | -0,04 | 0,05 | -0,74 | 4,61E-01 | 6,43E-01 |
| ATG2A | 910,72 | -0,10 | 0,13 | -0,74 | 4,61E-01 | 6,43E-01 |
| CCNQ | 317,33 | 0,03 | 0,04 | 0,74 | 4,61E-01 | 6,43E-01 |
| UNC93B1 | 1817,22 | -0,05 | 0,06 | -0,74 | 4,62E-01 | 6,44E-01 |
| TNFSF8 | 1046,77 | 0,04 | 0,06 | 0,74 | 4,62E-01 | 6,44E-01 |
| ABLIM1 | 3962,31 | 0,04 | 0,06 | 0,74 | 4,62E-01 | 6,44E-01 |
| GNE | 514,79 | 0,02 | 0,03 | 0,74 | 4,62E-01 | 6,44E-01 |
| CD44 | 9520,55 | 0,03 | 0,04 | 0,74 | 4,61E-01 | 6,44E-01 |
| SLC41A2 | 89,35 | 0,07 | 0,09 | 0,74 | 4,62E-01 | 6,44E-01 |
| PCDHGA9 | 33,38 | -0,10 | 0,13 | -0,74 | 4,62E-01 | 6,44E-01 |
| ZNF792 | 737,12 | -0,03 | 0,04 | -0,74 | 4,62E-01 | 6,44E-01 |
| UBE2I | 1629,49 | -0,03 | 0,04 | -0,74 | 4,62E-01 | 6,44E-01 |
| C9orf153 | 40,17 | -0,07 | 0,10 | -0,74 | 4,62E-01 | 6,44E-01 |
| FAM32A | 1404,89 | 0,04 | 0,05 | 0,73 | 4,62E-01 | 6,44E-01 |
| TAOK2 | 1025,93 | -0,04 | 0,06 | -0,73 | 4,62E-01 | 6,44E-01 |
| CCDC136 | 260,77 | -0,05 | 0,07 | -0,73 | 4,63E-01 | 6,44E-01 |
| SUCNR1 | 54,11 | -0,09 | 0,13 | -0,73 | 4,63E-01 | 6,45E-01 |
| THSD7A | 57,12 | -0,16 | 0,22 | -0,73 | 4,63E-01 | 6,45E-01 |
| PUDP | 241,02 | 0,02 | 0,03 | 0,73 | 4,63E-01 | 6,45E-01 |
| H2AC15 | 317,44 | 0,06 | 0,08 | 0,73 | 4,63E-01 | 6,45E-01 |
| ZNF415 | 42,20 | -0,11 | 0,16 | -0,73 | 4,64E-01 | 6,45E-01 |
| HUWE1 | 4675,04 | -0,05 | 0,07 | -0,73 | 4,64E-01 | 6,45E-01 |
| GPR75 | 96,55 | -0,05 | 0,07 | -0,73 | 4,64E-01 | 6,46E-01 |
| BRAF | 3927,89 | 0,03 | 0,04 | 0,73 | 4,64E-01 | 6,46E-01 |
| DENND4A | 6612,14 | 0,04 | 0,06 | 0,73 | 4,64E-01 | 6,46E-01 |
| KIR3DL1 | 42,15 | -0,13 | 0,18 | -0,73 | 4,65E-01 | 6,46E-01 |
| CARS2 | 2742,15 | -0,03 | 0,05 | -0,73 | 4,65E-01 | 6,46E-01 |
| IGFLR1 | 184,41 | 0,04 | 0,06 | 0,73 | 4,65E-01 | 6,47E-01 |

| | | | | | | |
|---|---|---|---|---|---|---|
| SGCE | 104,73 | -0,13 | 0,18 | -0,73 | 4,65E-01 | 6,47E-01 |
| SLC35D1 | 1244,11 | 0,03 | 0,04 | 0,73 | 4,65E-01 | 6,47E-01 |
| TTLL4 | 457,72 | -0,06 | 0,08 | -0,73 | 4,65E-01 | 6,47E-01 |
| SECISBP2 | 4799,23 | 0,03 | 0,04 | 0,73 | 4,65E-01 | 6,47E-01 |
| HIRA | 294,69 | -0,03 | 0,05 | -0,73 | 4,66E-01 | 6,47E-01 |
| RASSF2 | 20691,81 | -0,05 | 0,07 | -0,73 | 4,65E-01 | 6,47E-01 |
| AREL1 | 4166,66 | -0,04 | 0,05 | -0,73 | 4,65E-01 | 6,47E-01 |
| SS18L1 | 477,38 | -0,03 | 0,04 | -0,73 | 4,65E-01 | 6,47E-01 |
| CAMSAP2 | 211,97 | 0,06 | 0,08 | 0,73 | 4,65E-01 | 6,47E-01 |
| MEI1 | 396,99 | -0,04 | 0,05 | -0,73 | 4,65E-01 | 6,47E-01 |
| RGMB | 195,95 | -0,07 | 0,10 | -0,73 | 4,65E-01 | 6,47E-01 |
| LZTFL1 | 411,55 | 0,04 | 0,06 | 0,73 | 4,66E-01 | 6,47E-01 |
| EOLA1 | 347,22 | 0,02 | 0,03 | 0,73 | 4,66E-01 | 6,47E-01 |
| PDIA3 | 1217,02 | 0,03 | 0,04 | 0,73 | 4,66E-01 | 6,47E-01 |
| SRPRA | 1569,18 | -0,04 | 0,05 | -0,73 | 4,66E-01 | 6,47E-01 |
| FAHD2A | 301,20 | -0,04 | 0,05 | -0,73 | 4,66E-01 | 6,47E-01 |
| CSGALNACT: | 2271,40 | 0,04 | 0,05 | 0,73 | 4,66E-01 | 6,47E-01 |
| HMGB2 | 1816,08 | 0,08 | 0,11 | 0,73 | 4,66E-01 | 6,47E-01 |
| CMTM2 | 1613,29 | -0,08 | 0,11 | -0,73 | 4,67E-01 | 6,48E-01 |
| ALOX5AP | 3107,70 | -0,06 | 0,09 | -0,73 | 4,67E-01 | 6,48E-01 |
| DGAT2 | 3257,90 | -0,06 | 0,08 | -0,73 | 4,67E-01 | 6,48E-01 |
| SMG6 | 1033,37 | -0,05 | 0,07 | -0,73 | 4,67E-01 | 6,48E-01 |
| MORN1 | 286,77 | -0,05 | 0,07 | -0,73 | 4,67E-01 | 6,48E-01 |
| ALDH5A1 | 745,14 | -0,07 | 0,09 | -0,73 | 4,67E-01 | 6,49E-01 |
| IDH2 | 880,53 | -0,03 | 0,04 | -0,73 | 4,68E-01 | 6,49E-01 |
| ANKRD11 | 4313,31 | -0,06 | 0,08 | -0,73 | 4,68E-01 | 6,49E-01 |
| OSER1 | 1562,09 | -0,05 | 0,06 | -0,73 | 4,68E-01 | 6,49E-01 |
| CCDC149 | 306,82 | -0,07 | 0,09 | -0,73 | 4,68E-01 | 6,49E-01 |
| CELF1 | 6123,22 | -0,03 | 0,04 | -0,73 | 4,68E-01 | 6,49E-01 |
| ANTKMT | 322,45 | 0,04 | 0,05 | 0,73 | 4,68E-01 | 6,49E-01 |
| BIRC6 | 14965,22 | 0,03 | 0,05 | 0,73 | 4,68E-01 | 6,49E-01 |
| USP32 | 7683,50 | -0,04 | 0,06 | -0,73 | 4,68E-01 | 6,49E-01 |
| TOR1AIP2 | 3743,15 | 0,02 | 0,03 | 0,73 | 4,68E-01 | 6,49E-01 |
| DNAJB5 | 82,13 | -0,10 | 0,13 | -0,73 | 4,68E-01 | 6,49E-01 |
| B2M | 111430,26 | 0,08 | 0,11 | 0,72 | 4,68E-01 | 6,49E-01 |
| ZNF266 | 1928,33 | 0,04 | 0,05 | 0,72 | 4,68E-01 | 6,49E-01 |
| FAM168A | 1162,37 | -0,04 | 0,05 | -0,73 | 4,68E-01 | 6,49E-01 |
| PATJ | 2486,42 | 0,04 | 0,06 | 0,72 | 4,69E-01 | 6,49E-01 |
| BRD1 | 1385,27 | -0,04 | 0,06 | -0,72 | 4,69E-01 | 6,49E-01 |
| C1orf216 | 240,17 | 0,04 | 0,05 | 0,72 | 4,69E-01 | 6,49E-01 |
| DPYD | 10330,06 | 0,05 | 0,07 | 0,72 | 4,69E-01 | 6,50E-01 |
| KAT8 | 1632,90 | 0,03 | 0,04 | 0,72 | 4,70E-01 | 6,50E-01 |
| SHMT1 | 228,71 | -0,05 | 0,07 | -0,72 | 4,70E-01 | 6,50E-01 |
| NLRP1 | 5497,78 | -0,08 | 0,11 | -0,72 | 4,70E-01 | 6,50E-01 |
| RHOT2 | 1087,84 | -0,04 | 0,05 | -0,72 | 4,70E-01 | 6,50E-01 |
| SAMD4B | 716,07 | -0,08 | 0,10 | -0,72 | 4,70E-01 | 6,50E-01 |

| | | | | | | |
|---|---|---|---|---|---|---|
| ZMAT2 | 2715,65 | -0,07 | 0,10 | -0,72 | 4,70E-01 | 6,50E-01 |
| TMTC4 | 529,38 | -0,03 | 0,05 | -0,72 | 4,70E-01 | 6,50E-01 |
| PLEKHG1 | 462,71 | 0,07 | 0,10 | 0,72 | 4,70E-01 | 6,50E-01 |
| NELFE | 438,68 | -0,03 | 0,04 | -0,72 | 4,70E-01 | 6,50E-01 |
| LRRC69 | 205,03 | 0,04 | 0,05 | 0,72 | 4,70E-01 | 6,50E-01 |
| ACOT4 | 36,45 | -0,08 | 0,11 | -0,72 | 4,70E-01 | 6,50E-01 |
| ZNF582 | 124,79 | 0,05 | 0,07 | 0,72 | 4,70E-01 | 6,50E-01 |
| CD177 | 70,65 | -0,17 | 0,24 | -0,72 | 4,70E-01 | 6,50E-01 |
| GCN1 | 2059,34 | -0,04 | 0,06 | -0,72 | 4,70E-01 | 6,50E-01 |
| NCR1 | 301,95 | -0,07 | 0,09 | -0,72 | 4,71E-01 | 6,50E-01 |
| LTO1 | 922,72 | -0,03 | 0,04 | -0,72 | 4,71E-01 | 6,51E-01 |
| ACTR10 | 1151,33 | 0,04 | 0,05 | 0,72 | 4,71E-01 | 6,51E-01 |
| SLC25A43 | 167,79 | -0,03 | 0,05 | -0,72 | 4,71E-01 | 6,51E-01 |
| URGCP | 637,05 | -0,03 | 0,04 | -0,72 | 4,71E-01 | 6,51E-01 |
| FUT10 | 259,83 | -0,08 | 0,11 | -0,72 | 4,71E-01 | 6,51E-01 |
| ZBTB16 | 414,64 | -0,08 | 0,11 | -0,72 | 4,71E-01 | 6,51E-01 |
| FCMR | 1251,09 | 0,06 | 0,08 | 0,72 | 4,71E-01 | 6,51E-01 |
| NUB1 | 2038,51 | -0,03 | 0,04 | -0,72 | 4,71E-01 | 6,51E-01 |
| EEIG1 | 706,97 | -0,09 | 0,13 | -0,72 | 4,71E-01 | 6,51E-01 |
| PRKAR2A | 1404,99 | 0,02 | 0,02 | 0,72 | 4,71E-01 | 6,51E-01 |
| HDAC1 | 2889,85 | 0,03 | 0,04 | 0,72 | 4,71E-01 | 6,51E-01 |
| OPA3 | 258,33 | -0,05 | 0,07 | -0,72 | 4,72E-01 | 6,51E-01 |
| CBX6 | 1109,19 | -0,05 | 0,07 | -0,72 | 4,72E-01 | 6,51E-01 |
| DHX30 | 926,67 | -0,04 | 0,05 | -0,72 | 4,72E-01 | 6,51E-01 |
| GUSB | 495,00 | -0,04 | 0,05 | -0,72 | 4,72E-01 | 6,51E-01 |
| HOMER3 | 218,25 | 0,05 | 0,07 | 0,72 | 4,72E-01 | 6,51E-01 |
| FARSA | 289,41 | -0,04 | 0,05 | -0,72 | 4,72E-01 | 6,52E-01 |
| SAP18 | 2235,37 | 0,04 | 0,06 | 0,72 | 4,73E-01 | 6,52E-01 |
| DBP | 407,15 | -0,04 | 0,06 | -0,72 | 4,73E-01 | 6,52E-01 |
| SPRY2 | 63,26 | 0,07 | 0,09 | 0,72 | 4,73E-01 | 6,52E-01 |
| PSAP | 13506,12 | -0,06 | 0,08 | -0,72 | 4,73E-01 | 6,52E-01 |
| RPS10 | 112,30 | 0,03 | 0,04 | 0,72 | 4,73E-01 | 6,52E-01 |
| TMEM272 | 956,10 | -0,06 | 0,09 | -0,72 | 4,73E-01 | 6,52E-01 |
| SLC4A4 | 225,51 | 0,07 | 0,10 | 0,72 | 4,73E-01 | 6,53E-01 |
| VPS37C | 305,27 | -0,06 | 0,09 | -0,72 | 4,73E-01 | 6,53E-01 |
| CCR4 | 1225,79 | 0,05 | 0,07 | 0,72 | 4,74E-01 | 6,53E-01 |
| MZF1 | 625,18 | -0,04 | 0,06 | -0,72 | 4,74E-01 | 6,53E-01 |
| SIRPG | 159,31 | -0,06 | 0,09 | -0,72 | 4,74E-01 | 6,53E-01 |
| CCNJL | 1265,24 | -0,07 | 0,10 | -0,72 | 4,74E-01 | 6,53E-01 |
| XYLT1 | 1595,72 | -0,05 | 0,06 | -0,72 | 4,74E-01 | 6,53E-01 |
| CKLF | 862,47 | -0,04 | 0,06 | -0,72 | 4,74E-01 | 6,53E-01 |
| NELFCD | 857,02 | 0,03 | 0,04 | 0,71 | 4,75E-01 | 6,54E-01 |
| MSRA | 411,25 | -0,06 | 0,08 | -0,71 | 4,75E-01 | 6,54E-01 |
| BMAL1 | 2497,70 | -0,03 | 0,04 | -0,71 | 4,75E-01 | 6,54E-01 |
| USP46 | 482,56 | 0,04 | 0,06 | 0,71 | 4,75E-01 | 6,54E-01 |
| CYTH1 | 9642,83 | -0,03 | 0,05 | -0,71 | 4,75E-01 | 6,54E-01 |

| | | | | | | |
|---|---|---|---|---|---|---|
| ERCC1 | 582,14 | -0,03 | 0,04 | -0,71 | 4,75E-01 | 6,54E-01 |
| MAP3K14 | 602,45 | -0,06 | 0,09 | -0,71 | 4,75E-01 | 6,54E-01 |
| IL13RA1 | 2881,10 | -0,05 | 0,07 | -0,71 | 4,75E-01 | 6,54E-01 |
| XBP1 | 2428,14 | 0,04 | 0,05 | 0,71 | 4,76E-01 | 6,54E-01 |
| DCUN1D3 | 345,63 | -0,03 | 0,05 | -0,71 | 4,76E-01 | 6,54E-01 |
| TMEM268 | 425,36 | -0,06 | 0,08 | -0,71 | 4,76E-01 | 6,55E-01 |
| TMC6 | 3401,16 | -0,06 | 0,08 | -0,71 | 4,76E-01 | 6,55E-01 |
| MTMR2 | 447,82 | 0,03 | 0,05 | 0,71 | 4,76E-01 | 6,55E-01 |
| MCRIP1 | 370,01 | -0,04 | 0,05 | -0,71 | 4,76E-01 | 6,55E-01 |
| ASAH2B | 103,69 | 0,07 | 0,10 | 0,71 | 4,76E-01 | 6,55E-01 |
| CRY1 | 359,22 | 0,03 | 0,04 | 0,71 | 4,76E-01 | 6,55E-01 |
| HSD17B12 | 617,22 | 0,03 | 0,05 | 0,71 | 4,76E-01 | 6,55E-01 |
| DAB2 | 446,37 | -0,05 | 0,07 | -0,71 | 4,76E-01 | 6,55E-01 |
| RAD9B | 35,72 | -0,07 | 0,09 | -0,71 | 4,77E-01 | 6,55E-01 |
| TM6SF1 | 1284,80 | -0,05 | 0,07 | -0,71 | 4,77E-01 | 6,55E-01 |
| ANAPC7 | 900,74 | 0,02 | 0,03 | 0,71 | 4,77E-01 | 6,55E-01 |
| UVSSA | 1945,06 | -0,06 | 0,08 | -0,71 | 4,77E-01 | 6,55E-01 |
| MFSD5 | 430,30 | -0,03 | 0,04 | -0,71 | 4,77E-01 | 6,56E-01 |
| CLPB | 763,61 | -0,04 | 0,05 | -0,71 | 4,77E-01 | 6,56E-01 |
| AP3D1 | 1302,43 | -0,06 | 0,08 | -0,71 | 4,78E-01 | 6,56E-01 |
| RBM19 | 1281,36 | 0,04 | 0,05 | 0,71 | 4,78E-01 | 6,56E-01 |
| PLEKHO1 | 3227,85 | 0,03 | 0,05 | 0,71 | 4,78E-01 | 6,56E-01 |
| CPEB4 | 5383,95 | -0,04 | 0,06 | -0,71 | 4,78E-01 | 6,56E-01 |
| TNFRSF10B | 2331,37 | -0,03 | 0,05 | -0,71 | 4,78E-01 | 6,56E-01 |
| FGGY | 442,99 | -0,04 | 0,06 | -0,71 | 4,78E-01 | 6,56E-01 |
| HSPA1L | 211,45 | -0,04 | 0,05 | -0,71 | 4,78E-01 | 6,56E-01 |
| IL18 | 171,14 | 0,05 | 0,08 | 0,71 | 4,78E-01 | 6,56E-01 |
| SLC39A3 | 198,30 | -0,04 | 0,06 | -0,71 | 4,78E-01 | 6,56E-01 |
| PAN2 | 839,27 | -0,04 | 0,05 | -0,71 | 4,78E-01 | 6,56E-01 |
| NPM3 | 134,63 | 0,04 | 0,05 | 0,71 | 4,79E-01 | 6,57E-01 |
| INTS6 | 5793,72 | 0,03 | 0,05 | 0,71 | 4,79E-01 | 6,57E-01 |
| SLC16A6 | 1148,93 | 0,04 | 0,06 | 0,71 | 4,79E-01 | 6,57E-01 |
| EFTUD2 | 1261,30 | 0,03 | 0,04 | 0,71 | 4,79E-01 | 6,57E-01 |
| GATAD1 | 1132,99 | 0,02 | 0,03 | 0,71 | 4,79E-01 | 6,57E-01 |
| TMEM185A | 363,25 | -0,03 | 0,04 | -0,71 | 4,79E-01 | 6,57E-01 |
| SLC17A5 | 549,86 | -0,02 | 0,03 | -0,71 | 4,79E-01 | 6,57E-01 |
| ATPAF2 | 384,48 | -0,03 | 0,05 | -0,71 | 4,79E-01 | 6,57E-01 |
| DCPS | 314,58 | -0,03 | 0,04 | -0,71 | 4,79E-01 | 6,57E-01 |
| QPCT | 2196,98 | -0,07 | 0,09 | -0,71 | 4,79E-01 | 6,57E-01 |
| RNF182 | 578,40 | -0,22 | 0,31 | -0,71 | 4,80E-01 | 6,57E-01 |
| ATG4A | 314,34 | 0,03 | 0,05 | 0,71 | 4,80E-01 | 6,57E-01 |
| FXYD5 | 635,93 | -0,04 | 0,06 | -0,71 | 4,80E-01 | 6,57E-01 |
| NEIL1 | 217,24 | -0,06 | 0,09 | -0,71 | 4,80E-01 | 6,57E-01 |
| ADAMTS1 | 218,31 | 0,08 | 0,12 | 0,71 | 4,80E-01 | 6,58E-01 |
| GATA3 | 176,37 | -0,06 | 0,09 | -0,71 | 4,80E-01 | 6,58E-01 |
| XAB2 | 633,86 | -0,04 | 0,05 | -0,71 | 4,80E-01 | 6,58E-01 |

| | | | | | | |
|---|---|---|---|---|---|---|
| NOG | 202,20 | 0,10 | 0,14 | 0,71 | 4,80E-01 | 6,58E-01 |
| TAFAZZIN | 613,52 | -0,04 | 0,06 | -0,71 | 4,81E-01 | 6,58E-01 |
| EID3 | 230,53 | 0,04 | 0,05 | 0,70 | 4,81E-01 | 6,59E-01 |
| IL21R | 657,96 | -0,05 | 0,07 | -0,70 | 4,81E-01 | 6,59E-01 |
| KLHL23 | 41,17 | -0,08 | 0,11 | -0,70 | 4,82E-01 | 6,59E-01 |
| PPIAL4H | 80,95 | 0,10 | 0,14 | 0,70 | 4,82E-01 | 6,59E-01 |
| PAG1 | 7991,99 | 0,04 | 0,05 | 0,70 | 4,82E-01 | 6,60E-01 |
| PIN1 | 884,52 | -0,02 | 0,03 | -0,70 | 4,82E-01 | 6,60E-01 |
| KAT6B | 2237,75 | 0,03 | 0,04 | 0,70 | 4,82E-01 | 6,60E-01 |
| USP51 | 47,82 | 0,07 | 0,09 | 0,70 | 4,82E-01 | 6,60E-01 |
| CLEC9A | 73,90 | -0,09 | 0,12 | -0,70 | 4,82E-01 | 6,60E-01 |
| IK | 2879,85 | 0,04 | 0,05 | 0,70 | 4,82E-01 | 6,60E-01 |
| NUDT22 | 438,99 | -0,03 | 0,05 | -0,70 | 4,83E-01 | 6,60E-01 |
| PAAF1 | 365,59 | 0,03 | 0,04 | 0,70 | 4,83E-01 | 6,60E-01 |
| GRN | 1681,21 | -0,07 | 0,10 | -0,70 | 4,83E-01 | 6,61E-01 |
| PPIB | 2438,42 | 0,04 | 0,05 | 0,70 | 4,83E-01 | 6,61E-01 |
| UMAD1 | 647,94 | 0,03 | 0,05 | 0,70 | 4,83E-01 | 6,61E-01 |
| SLC39A1 | 112,45 | 0,04 | 0,06 | 0,70 | 4,83E-01 | 6,61E-01 |
| PTGER2 | 784,53 | 0,04 | 0,05 | 0,70 | 4,83E-01 | 6,61E-01 |
| SAMD4A | 295,76 | -0,07 | 0,11 | -0,70 | 4,84E-01 | 6,61E-01 |
| SMIM20 | 163,85 | 0,05 | 0,08 | 0,70 | 4,84E-01 | 6,61E-01 |
| METTL17 | 623,26 | 0,02 | 0,03 | 0,70 | 4,84E-01 | 6,61E-01 |
| MYO6 | 348,20 | 0,08 | 0,12 | 0,70 | 4,84E-01 | 6,61E-01 |
| RUNX2 | 1325,67 | -0,04 | 0,06 | -0,70 | 4,84E-01 | 6,61E-01 |
| ATP1B1 | 96,77 | -0,08 | 0,11 | -0,70 | 4,84E-01 | 6,62E-01 |
| LBH | 5456,18 | 0,04 | 0,06 | 0,70 | 4,84E-01 | 6,62E-01 |
| S100A6 | 2612,29 | -0,04 | 0,06 | -0,70 | 4,85E-01 | 6,62E-01 |
| PECAM1 | 9542,99 | -0,04 | 0,06 | -0,70 | 4,85E-01 | 6,62E-01 |
| TINF2 | 1885,96 | 0,02 | 0,03 | 0,70 | 4,85E-01 | 6,62E-01 |
| PEX26 | 1304,10 | -0,04 | 0,06 | -0,70 | 4,85E-01 | 6,62E-01 |
| COL4A3 | 195,09 | -0,10 | 0,14 | -0,70 | 4,85E-01 | 6,62E-01 |
| TNFRSF18 | 96,87 | 0,07 | 0,10 | 0,70 | 4,86E-01 | 6,62E-01 |
| MAPK9 | 978,60 | 0,02 | 0,03 | 0,70 | 4,86E-01 | 6,62E-01 |
| TMEM80 | 361,16 | -0,04 | 0,06 | -0,70 | 4,85E-01 | 6,62E-01 |
| CRELD2 | 402,39 | -0,03 | 0,04 | -0,70 | 4,85E-01 | 6,62E-01 |
| IFT70A | 104,82 | 0,06 | 0,09 | 0,70 | 4,86E-01 | 6,62E-01 |
| SLC25A29 | 601,26 | -0,07 | 0,11 | -0,70 | 4,85E-01 | 6,62E-01 |
| DPP3 | 236,67 | -0,04 | 0,06 | -0,70 | 4,85E-01 | 6,62E-01 |
| GSR | 2415,48 | -0,03 | 0,04 | -0,70 | 4,86E-01 | 6,62E-01 |
| NIBAN2 | 577,61 | 0,05 | 0,07 | 0,70 | 4,86E-01 | 6,62E-01 |
| ENPP2 | 103,75 | -0,09 | 0,13 | -0,70 | 4,85E-01 | 6,62E-01 |
| CIAPIN1 | 476,64 | 0,02 | 0,02 | 0,70 | 4,85E-01 | 6,62E-01 |
| MYH9 | 15858,83 | -0,07 | 0,10 | -0,70 | 4,86E-01 | 6,62E-01 |
| ZFTRAF1 | 947,81 | -0,06 | 0,08 | -0,70 | 4,86E-01 | 6,63E-01 |
| BAX | 784,11 | -0,03 | 0,04 | -0,70 | 4,86E-01 | 6,63E-01 |
| GIPC1 | 181,56 | -0,05 | 0,07 | -0,70 | 4,86E-01 | 6,63E-01 |

| | | | | | | |
|---|---|---|---|---|---|---|
| NUP210 | 3230,34 | -0,05 | 0,08 | -0,70 | 4,86E-01 | 6,63E-01 |
| NAPB | 942,30 | 0,03 | 0,05 | 0,70 | 4,86E-01 | 6,63E-01 |
| PREP | 783,55 | 0,03 | 0,04 | 0,70 | 4,87E-01 | 6,63E-01 |
| ZNF112 | 28,37 | -0,12 | 0,17 | -0,69 | 4,87E-01 | 6,64E-01 |
| JAK1 | 13092,72 | 0,03 | 0,05 | 0,69 | 4,87E-01 | 6,64E-01 |
| GPRASP3 | 93,40 | -0,05 | 0,06 | -0,69 | 4,87E-01 | 6,64E-01 |
| LAP3 | 1994,52 | 0,08 | 0,12 | 0,69 | 4,87E-01 | 6,64E-01 |
| SNRPB | 635,35 | 0,03 | 0,04 | 0,69 | 4,88E-01 | 6,64E-01 |
| PDZD11 | 98,40 | -0,03 | 0,04 | -0,69 | 4,88E-01 | 6,64E-01 |
| CCDC142 | 1100,99 | 0,03 | 0,04 | 0,69 | 4,88E-01 | 6,64E-01 |
| PLEKHJ1 | 748,97 | -0,03 | 0,04 | -0,69 | 4,88E-01 | 6,64E-01 |
| DTX4 | 267,39 | -0,06 | 0,08 | -0,69 | 4,88E-01 | 6,64E-01 |
| TRIB2 | 1913,36 | 0,04 | 0,06 | 0,69 | 4,88E-01 | 6,64E-01 |
| TENT4A | 851,48 | -0,04 | 0,05 | -0,69 | 4,88E-01 | 6,64E-01 |
| TCOF1 | 645,68 | -0,05 | 0,07 | -0,69 | 4,88E-01 | 6,64E-01 |
| PRKACA | 543,70 | -0,08 | 0,11 | -0,69 | 4,88E-01 | 6,64E-01 |
| DDIT4 | 1108,57 | 0,06 | 0,09 | 0,69 | 4,88E-01 | 6,64E-01 |
| IL10RA | 7897,77 | 0,03 | 0,04 | 0,69 | 4,88E-01 | 6,64E-01 |
| PARP2 | 118,18 | 0,04 | 0,06 | 0,69 | 4,88E-01 | 6,64E-01 |
| HYLS1 | 152,82 | 0,04 | 0,06 | 0,69 | 4,89E-01 | 6,65E-01 |
| C1orf174 | 562,62 | -0,02 | 0,03 | -0,69 | 4,89E-01 | 6,65E-01 |
| DAPP1 | 4415,23 | 0,05 | 0,07 | 0,69 | 4,89E-01 | 6,65E-01 |
| CPVL | 2733,69 | 0,05 | 0,07 | 0,69 | 4,89E-01 | 6,65E-01 |
| YEATS2 | 1624,01 | -0,02 | 0,03 | -0,69 | 4,89E-01 | 6,65E-01 |
| CST7 | 383,62 | -0,07 | 0,10 | -0,69 | 4,89E-01 | 6,65E-01 |
| TMEM177 | 137,89 | -0,04 | 0,06 | -0,69 | 4,90E-01 | 6,65E-01 |
| FZD1 | 227,58 | -0,05 | 0,08 | -0,69 | 4,89E-01 | 6,65E-01 |
| STK4 | 19984,70 | 0,03 | 0,04 | 0,69 | 4,89E-01 | 6,65E-01 |
| MSI2 | 2486,06 | -0,03 | 0,05 | -0,69 | 4,90E-01 | 6,65E-01 |
| PHACTR4 | 453,13 | -0,04 | 0,06 | -0,69 | 4,90E-01 | 6,65E-01 |
| TANGO6 | 1111,26 | 0,02 | 0,03 | 0,69 | 4,90E-01 | 6,65E-01 |
| CLASP1 | 3632,37 | -0,03 | 0,04 | -0,69 | 4,90E-01 | 6,65E-01 |
| ARRB1 | 1533,07 | -0,06 | 0,09 | -0,69 | 4,90E-01 | 6,65E-01 |
| MAP4K4 | 6263,95 | -0,04 | 0,05 | -0,69 | 4,90E-01 | 6,65E-01 |
| ADNP2 | 564,52 | -0,02 | 0,03 | -0,69 | 4,90E-01 | 6,65E-01 |
| HIGD1A | 406,94 | 0,07 | 0,11 | 0,69 | 4,90E-01 | 6,65E-01 |
| PTGES3 | 1714,01 | 0,05 | 0,08 | 0,69 | 4,90E-01 | 6,66E-01 |
| ANKZF1 | 2045,30 | -0,03 | 0,04 | -0,69 | 4,90E-01 | 6,66E-01 |
| SULT1A2 | 36,50 | -0,11 | 0,16 | -0,69 | 4,91E-01 | 6,66E-01 |
| QSOX1 | 674,76 | -0,06 | 0,08 | -0,69 | 4,91E-01 | 6,66E-01 |
| ASH1L | 6132,75 | 0,02 | 0,03 | 0,69 | 4,91E-01 | 6,66E-01 |
| MTHFR | 896,30 | -0,06 | 0,09 | -0,69 | 4,91E-01 | 6,66E-01 |
| BCL9L | 838,47 | -0,09 | 0,13 | -0,69 | 4,91E-01 | 6,66E-01 |
| BAD | 347,16 | -0,04 | 0,05 | -0,69 | 4,91E-01 | 6,66E-01 |
| ZNF607 | 194,17 | 0,04 | 0,06 | 0,69 | 4,91E-01 | 6,66E-01 |
| KCNJ2 | 4527,65 | -0,06 | 0,09 | -0,69 | 4,91E-01 | 6,66E-01 |

| | | | | | | |
|---|---|---|---|---|---|---|
| UNK | 532,43 | -0,04 | 0,06 | -0,69 | 4,92E-01 | 6,67E-01 |
| SLC19A2 | 220,76 | 0,04 | 0,05 | 0,69 | 4,92E-01 | 6,67E-01 |
| SLC25A13 | 636,60 | 0,03 | 0,04 | 0,69 | 4,92E-01 | 6,67E-01 |
| TIAM2 | 558,15 | -0,05 | 0,07 | -0,69 | 4,92E-01 | 6,67E-01 |
| EXOC6 | 1533,87 | 0,05 | 0,07 | 0,69 | 4,92E-01 | 6,67E-01 |
| NOC2L | 394,42 | -0,06 | 0,08 | -0,69 | 4,92E-01 | 6,67E-01 |
| CHKA | 193,35 | -0,05 | 0,07 | -0,69 | 4,92E-01 | 6,67E-01 |
| CRY2 | 833,88 | -0,03 | 0,04 | -0,69 | 4,92E-01 | 6,67E-01 |
| AAMP | 876,85 | 0,03 | 0,05 | 0,69 | 4,93E-01 | 6,67E-01 |
| CPSF1 | 653,88 | -0,07 | 0,10 | -0,69 | 4,93E-01 | 6,67E-01 |
| RPS11 | 19335,89 | 0,06 | 0,09 | 0,69 | 4,93E-01 | 6,67E-01 |
| PARK7 | 1710,88 | 0,04 | 0,06 | 0,69 | 4,93E-01 | 6,67E-01 |
| NCOA3 | 5026,90 | 0,03 | 0,04 | 0,69 | 4,93E-01 | 6,67E-01 |
| BCL10 | 1201,97 | 0,03 | 0,04 | 0,69 | 4,93E-01 | 6,67E-01 |
| ARHGAP42 | 129,30 | 0,07 | 0,10 | 0,69 | 4,93E-01 | 6,67E-01 |
| LRRC59 | 1159,47 | 0,03 | 0,04 | 0,69 | 4,93E-01 | 6,68E-01 |
| PSME2 | 470,22 | -0,04 | 0,06 | -0,69 | 4,93E-01 | 6,68E-01 |
| IFT27 | 353,41 | -0,03 | 0,05 | -0,68 | 4,93E-01 | 6,68E-01 |
| FAM219B | 1064,86 | -0,02 | 0,03 | -0,68 | 4,93E-01 | 6,68E-01 |
| PCGF5 | 4851,33 | 0,05 | 0,07 | 0,68 | 4,94E-01 | 6,68E-01 |
| MDC1 | 410,26 | -0,04 | 0,06 | -0,68 | 4,94E-01 | 6,68E-01 |
| NDUFS8 | 259,27 | -0,04 | 0,06 | -0,68 | 4,94E-01 | 6,68E-01 |
| BRD9 | 1618,39 | -0,03 | 0,04 | -0,68 | 4,94E-01 | 6,68E-01 |
| SERINC5 | 3078,62 | -0,05 | 0,07 | -0,68 | 4,94E-01 | 6,68E-01 |
| SCLT1 | 4809,18 | 0,05 | 0,07 | 0,68 | 4,94E-01 | 6,68E-01 |
| SFMBT2 | 1487,13 | -0,03 | 0,04 | -0,68 | 4,94E-01 | 6,68E-01 |
| KLHL5 | 1315,04 | 0,04 | 0,05 | 0,68 | 4,94E-01 | 6,68E-01 |
| MTHFD2 | 842,18 | 0,05 | 0,07 | 0,68 | 4,94E-01 | 6,68E-01 |
| CDH26 | 242,40 | -0,05 | 0,07 | -0,68 | 4,94E-01 | 6,68E-01 |
| SPTAN1 | 3721,14 | -0,05 | 0,07 | -0,68 | 4,94E-01 | 6,68E-01 |
| B3GALT6 | 499,95 | 0,02 | 0,04 | 0,68 | 4,95E-01 | 6,69E-01 |
| LILRA4 | 57,70 | 0,09 | 0,13 | 0,68 | 4,95E-01 | 6,69E-01 |
| RASGEF1A | 225,60 | -0,05 | 0,08 | -0,68 | 4,95E-01 | 6,69E-01 |
| MRPL41 | 445,18 | 0,04 | 0,05 | 0,68 | 4,95E-01 | 6,69E-01 |
| GNB5 | 1172,96 | 0,03 | 0,04 | 0,68 | 4,95E-01 | 6,69E-01 |
| MTCP1 | 52,42 | 0,04 | 0,06 | 0,68 | 4,95E-01 | 6,69E-01 |
| PATL2 | 1225,96 | -0,05 | 0,08 | -0,68 | 4,96E-01 | 6,69E-01 |
| TUBGCP6 | 1699,24 | -0,05 | 0,07 | -0,68 | 4,96E-01 | 6,70E-01 |
| RPF2 | 217,79 | 0,04 | 0,06 | 0,68 | 4,96E-01 | 6,70E-01 |
| IGF2R | 30885,08 | -0,06 | 0,09 | -0,68 | 4,96E-01 | 6,70E-01 |
| UST | 296,76 | 0,05 | 0,07 | 0,68 | 4,96E-01 | 6,70E-01 |
| DNAJC18 | 127,45 | -0,04 | 0,06 | -0,68 | 4,96E-01 | 6,70E-01 |
| MGMT | 405,35 | 0,03 | 0,05 | 0,68 | 4,96E-01 | 6,70E-01 |
| LOXL3 | 207,93 | -0,06 | 0,08 | -0,68 | 4,96E-01 | 6,70E-01 |
| IFT20 | 309,61 | 0,03 | 0,05 | 0,68 | 4,96E-01 | 6,70E-01 |
| PVR | 311,84 | 0,05 | 0,07 | 0,68 | 4,96E-01 | 6,70E-01 |

| | | | | | | |
|---|---|---|---|---|---|---|
| IKBKB | 2851,75 | -0,04 | 0,06 | -0,68 | 4,97E-01 | 6,70E-01 |
| SPON2 | 642,84 | -0,07 | 0,11 | -0,68 | 4,97E-01 | 6,70E-01 |
| NUF2 | 54,29 | -0,09 | 0,13 | -0,68 | 4,97E-01 | 6,70E-01 |
| SKAP2 | 2991,49 | 0,05 | 0,08 | 0,68 | 4,97E-01 | 6,70E-01 |
| VAV1 | 4479,70 | -0,04 | 0,05 | -0,68 | 4,97E-01 | 6,70E-01 |
| SRA1 | 566,87 | -0,03 | 0,04 | -0,68 | 4,97E-01 | 6,70E-01 |
| ETFA | 1640,26 | 0,04 | 0,05 | 0,68 | 4,97E-01 | 6,70E-01 |
| JUN | 290,78 | 0,05 | 0,08 | 0,68 | 4,97E-01 | 6,70E-01 |
| BCL6-AS1 | 101,71 | 0,09 | 0,13 | 0,68 | 4,97E-01 | 6,70E-01 |
| TACC1 | 7475,26 | -0,03 | 0,04 | -0,68 | 4,98E-01 | 6,71E-01 |
| SLC11A1 | 6456,25 | -0,07 | 0,10 | -0,68 | 4,98E-01 | 6,71E-01 |
| ZNF500 | 244,15 | -0,07 | 0,10 | -0,68 | 4,98E-01 | 6,71E-01 |
| PIP4K2B | 1563,51 | 0,03 | 0,04 | 0,68 | 4,98E-01 | 6,71E-01 |
| TTLL1 | 106,06 | -0,04 | 0,06 | -0,68 | 4,98E-01 | 6,71E-01 |
| SETDB1 | 835,32 | -0,04 | 0,06 | -0,68 | 4,98E-01 | 6,71E-01 |
| FAM24B | 128,63 | 0,04 | 0,06 | 0,68 | 4,98E-01 | 6,71E-01 |
| APMAP | 4316,80 | -0,05 | 0,07 | -0,68 | 4,99E-01 | 6,71E-01 |
| TAF10 | 864,44 | -0,02 | 0,04 | -0,68 | 4,99E-01 | 6,71E-01 |
| KLHDC4 | 1319,76 | -0,05 | 0,08 | -0,68 | 4,99E-01 | 6,72E-01 |
| FAR2 | 1445,25 | -0,05 | 0,07 | -0,68 | 4,99E-01 | 6,72E-01 |
| TIMM23B-AG | 65,85 | 0,06 | 0,09 | 0,68 | 4,99E-01 | 6,72E-01 |
| TTC4 | 77,44 | 0,03 | 0,05 | 0,68 | 4,99E-01 | 6,72E-01 |
| MYL12B | 3800,23 | 0,05 | 0,08 | 0,68 | 4,99E-01 | 6,72E-01 |
| MAP7D1 | 972,67 | -0,07 | 0,10 | -0,68 | 4,99E-01 | 6,72E-01 |
| RNLS | 90,76 | 0,04 | 0,06 | 0,68 | 5,00E-01 | 6,72E-01 |
| SLC25A20 | 907,54 | -0,04 | 0,07 | -0,68 | 5,00E-01 | 6,72E-01 |
| SPATA24 | 29,32 | -0,05 | 0,07 | -0,68 | 5,00E-01 | 6,72E-01 |
| EXOC6B | 674,13 | 0,03 | 0,04 | 0,68 | 5,00E-01 | 6,72E-01 |
| SMAP1 | 807,77 | 0,02 | 0,03 | 0,68 | 4,99E-01 | 6,72E-01 |
| KLHL18 | 1212,89 | -0,03 | 0,05 | -0,68 | 4,99E-01 | 6,72E-01 |
| NAT9 | 447,43 | 0,03 | 0,04 | 0,68 | 4,99E-01 | 6,72E-01 |
| SCML2 | 44,16 | -0,06 | 0,09 | -0,68 | 5,00E-01 | 6,72E-01 |
| CENPP | 146,35 | -0,07 | 0,10 | -0,68 | 4,99E-01 | 6,72E-01 |
| MYDGF | 591,06 | 0,04 | 0,06 | 0,67 | 5,00E-01 | 6,72E-01 |
| D2HGDH | 432,83 | -0,06 | 0,09 | -0,67 | 5,00E-01 | 6,72E-01 |
| SP140 | 2665,63 | 0,04 | 0,06 | 0,67 | 5,00E-01 | 6,72E-01 |
| TOM1 | 963,05 | -0,05 | 0,07 | -0,67 | 5,00E-01 | 6,72E-01 |
| CAPN3 | 152,24 | -0,06 | 0,10 | -0,67 | 5,00E-01 | 6,72E-01 |
| SPSB2 | 385,87 | 0,04 | 0,06 | 0,67 | 5,00E-01 | 6,72E-01 |
| FAT4 | 122,01 | -0,10 | 0,14 | -0,67 | 5,00E-01 | 6,72E-01 |
| SULF2 | 3645,53 | 0,05 | 0,08 | 0,67 | 5,00E-01 | 6,72E-01 |
| ZNHIT2 | 113,26 | -0,03 | 0,05 | -0,67 | 5,01E-01 | 6,72E-01 |
| CHTF8 | 1059,13 | -0,03 | 0,04 | -0,67 | 5,01E-01 | 6,72E-01 |
| ISM1 | 56,41 | -0,10 | 0,15 | -0,67 | 5,01E-01 | 6,72E-01 |
| ZNF747 | 188,76 | -0,03 | 0,04 | -0,67 | 5,01E-01 | 6,72E-01 |
| ZNF487 | 396,52 | -0,03 | 0,05 | -0,67 | 5,01E-01 | 6,72E-01 |

| | | | | | | |
|---|---|---|---|---|---|---|
| ATG10 | 277,39 | -0,04 | 0,06 | -0,67 | 5,01E-01 | 6,72E-01 |
| USP7 | 4585,78 | 0,02 | 0,03 | 0,67 | 5,00E-01 | 6,72E-01 |
| TAOK1 | 7874,74 | 0,03 | 0,04 | 0,67 | 5,01E-01 | 6,72E-01 |
| ZC3H18 | 658,59 | -0,05 | 0,08 | -0,67 | 5,01E-01 | 6,72E-01 |
| SERINC3 | 3983,97 | -0,03 | 0,04 | -0,67 | 5,01E-01 | 6,72E-01 |
| FGD4 | 2950,13 | 0,05 | 0,07 | 0,67 | 5,01E-01 | 6,72E-01 |
| ALMS1 | 2346,57 | 0,03 | 0,05 | 0,67 | 5,01E-01 | 6,72E-01 |
| CASP4 | 6720,64 | 0,05 | 0,07 | 0,67 | 5,01E-01 | 6,72E-01 |
| MAP3K3 | 3027,56 | -0,05 | 0,08 | -0,67 | 5,01E-01 | 6,72E-01 |
| LRRC14 | 389,34 | -0,06 | 0,09 | -0,67 | 5,01E-01 | 6,72E-01 |
| MCRIP2 | 197,38 | -0,05 | 0,08 | -0,67 | 5,02E-01 | 6,72E-01 |
| TMEM59 | 3669,06 | 0,04 | 0,06 | 0,67 | 5,02E-01 | 6,73E-01 |
| RETSAT | 443,10 | -0,03 | 0,04 | -0,67 | 5,02E-01 | 6,73E-01 |
| TTL | 760,52 | -0,03 | 0,04 | -0,67 | 5,02E-01 | 6,73E-01 |
| ANKRD16 | 131,84 | 0,04 | 0,06 | 0,67 | 5,03E-01 | 6,74E-01 |
| MTHFSD | 573,79 | -0,03 | 0,04 | -0,67 | 5,03E-01 | 6,74E-01 |
| ENSG000002 | 244,70 | -0,08 | 0,12 | -0,67 | 5,03E-01 | 6,74E-01 |
| ITM2A | 895,31 | 0,07 | 0,11 | 0,67 | 5,03E-01 | 6,74E-01 |
| TICAM2 | 129,53 | -0,04 | 0,06 | -0,67 | 5,03E-01 | 6,74E-01 |
| NUP62CL | 39,56 | -0,07 | 0,11 | -0,67 | 5,04E-01 | 6,75E-01 |
| FBXW11 | 2100,20 | 0,02 | 0,03 | 0,67 | 5,04E-01 | 6,75E-01 |
| SPRYD4 | 343,92 | -0,04 | 0,06 | -0,67 | 5,05E-01 | 6,76E-01 |
| AAAS | 152,02 | -0,05 | 0,07 | -0,67 | 5,05E-01 | 6,76E-01 |
| UBP1 | 1882,29 | 0,02 | 0,04 | 0,67 | 5,05E-01 | 6,76E-01 |
| GMCL1 | 1788,10 | 0,04 | 0,05 | 0,67 | 5,05E-01 | 6,76E-01 |
| ANGEL1 | 565,95 | 0,03 | 0,04 | 0,67 | 5,05E-01 | 6,76E-01 |
| SPTY2D1 | 1020,37 | 0,02 | 0,03 | 0,67 | 5,05E-01 | 6,76E-01 |
| RALGAPB | 4952,00 | 0,02 | 0,04 | 0,67 | 5,05E-01 | 6,76E-01 |
| CNPY2 | 249,17 | 0,03 | 0,04 | 0,67 | 5,05E-01 | 6,76E-01 |
| AQP9 | 13279,41 | -0,05 | 0,08 | -0,67 | 5,06E-01 | 6,76E-01 |
| FAM149B1 | 504,58 | 0,02 | 0,03 | 0,67 | 5,06E-01 | 6,76E-01 |
| VARS1 | 301,43 | -0,06 | 0,09 | -0,67 | 5,05E-01 | 6,76E-01 |
| ITGAE | 384,25 | -0,03 | 0,04 | -0,67 | 5,05E-01 | 6,76E-01 |
| ENGASE | 1147,07 | -0,04 | 0,06 | -0,67 | 5,06E-01 | 6,76E-01 |
| SEPTIN9 | 3341,98 | -0,06 | 0,09 | -0,67 | 5,05E-01 | 6,76E-01 |
| NDEL1 | 6185,59 | -0,04 | 0,07 | -0,67 | 5,06E-01 | 6,76E-01 |
| PCGF1 | 144,15 | -0,03 | 0,04 | -0,67 | 5,06E-01 | 6,76E-01 |
| KPNB1 | 5569,24 | -0,03 | 0,05 | -0,67 | 5,06E-01 | 6,76E-01 |
| GDPD1 | 105,07 | 0,04 | 0,06 | 0,66 | 5,06E-01 | 6,77E-01 |
| KDM7A | 8707,22 | 0,04 | 0,06 | 0,66 | 5,06E-01 | 6,77E-01 |
| ALG8 | 401,64 | 0,02 | 0,03 | 0,66 | 5,06E-01 | 6,77E-01 |
| GORASP1 | 688,93 | -0,03 | 0,04 | -0,66 | 5,06E-01 | 6,77E-01 |
| MRPL2 | 120,63 | -0,04 | 0,06 | -0,66 | 5,06E-01 | 6,77E-01 |
| SRPK1 | 2572,69 | -0,04 | 0,07 | -0,66 | 5,07E-01 | 6,77E-01 |
| GLG1 | 4963,10 | -0,03 | 0,05 | -0,66 | 5,07E-01 | 6,77E-01 |
| RPP40 | 86,33 | 0,05 | 0,08 | 0,66 | 5,07E-01 | 6,77E-01 |

| | | | | | | |
|---|---|---|---|---|---|---|
| MRPS6 | 341,40 | 0,04 | 0,05 | 0,66 | 5,07E-01 | 6,77E-01 |
| MED22 | 1262,91 | -0,03 | 0,04 | -0,66 | 5,07E-01 | 6,77E-01 |
| RPL18A | 9454,33 | 0,04 | 0,07 | 0,66 | 5,07E-01 | 6,77E-01 |
| SNX24 | 168,32 | 0,04 | 0,05 | 0,66 | 5,07E-01 | 6,77E-01 |
| BRD10 | 3395,04 | 0,02 | 0,04 | 0,66 | 5,07E-01 | 6,77E-01 |
| UQCRC1 | 1346,09 | 0,03 | 0,04 | 0,66 | 5,07E-01 | 6,77E-01 |
| SELL | 18687,68 | -0,05 | 0,07 | -0,66 | 5,07E-01 | 6,77E-01 |
| FBXO32 | 421,00 | -0,04 | 0,06 | -0,66 | 5,08E-01 | 6,78E-01 |
| CDC42EP3 | 3673,67 | 0,04 | 0,06 | 0,66 | 5,08E-01 | 6,78E-01 |
| KLRD1 | 5048,52 | 0,07 | 0,10 | 0,66 | 5,08E-01 | 6,78E-01 |
| CDK9 | 1309,90 | -0,02 | 0,04 | -0,66 | 5,08E-01 | 6,78E-01 |
| EHBP1L1 | 7803,69 | -0,05 | 0,08 | -0,66 | 5,08E-01 | 6,78E-01 |
| DAGLB | 592,88 | -0,04 | 0,06 | -0,66 | 5,08E-01 | 6,78E-01 |
| SGO1 | 34,48 | -0,08 | 0,13 | -0,66 | 5,08E-01 | 6,78E-01 |
| IRAK1 | 760,09 | -0,05 | 0,08 | -0,66 | 5,09E-01 | 6,78E-01 |
| FYN | 7023,02 | 0,04 | 0,05 | 0,66 | 5,09E-01 | 6,78E-01 |
| NUDT13 | 85,63 | 0,04 | 0,07 | 0,66 | 5,09E-01 | 6,78E-01 |
| TSC1 | 1935,11 | -0,02 | 0,03 | -0,66 | 5,09E-01 | 6,78E-01 |
| PRDM4 | 921,95 | 0,02 | 0,03 | 0,66 | 5,09E-01 | 6,78E-01 |
| TOB1 | 595,29 | 0,03 | 0,04 | 0,66 | 5,09E-01 | 6,78E-01 |
| ARID1B | 6511,62 | 0,03 | 0,04 | 0,66 | 5,09E-01 | 6,78E-01 |
| SHLD1 | 284,26 | -0,04 | 0,06 | -0,66 | 5,09E-01 | 6,78E-01 |
| PLA2G7 | 245,24 | 0,07 | 0,10 | 0,66 | 5,09E-01 | 6,78E-01 |
| GFOD1 | 1273,96 | -0,05 | 0,07 | -0,66 | 5,09E-01 | 6,78E-01 |
| NIPSNAP3B | 89,92 | 0,06 | 0,09 | 0,66 | 5,09E-01 | 6,78E-01 |
| HIP1 | 1285,15 | -0,08 | 0,13 | -0,66 | 5,09E-01 | 6,78E-01 |
| NSRP1 | 809,90 | 0,02 | 0,03 | 0,66 | 5,09E-01 | 6,78E-01 |
| CEP76 | 244,42 | 0,03 | 0,05 | 0,66 | 5,09E-01 | 6,78E-01 |
| IRF3 | 1123,56 | -0,03 | 0,05 | -0,66 | 5,10E-01 | 6,79E-01 |
| CISH | 518,98 | -0,07 | 0,11 | -0,66 | 5,10E-01 | 6,79E-01 |
| C6orf163 | 343,41 | -0,04 | 0,06 | -0,66 | 5,10E-01 | 6,79E-01 |
| OSBPL2 | 3530,39 | -0,04 | 0,06 | -0,66 | 5,10E-01 | 6,79E-01 |
| LST1 | 5086,12 | -0,04 | 0,07 | -0,66 | 5,10E-01 | 6,79E-01 |
| FRAT1 | 2331,64 | -0,05 | 0,07 | -0,66 | 5,10E-01 | 6,79E-01 |
| HMGXB4 | 449,09 | 0,02 | 0,03 | 0,66 | 5,10E-01 | 6,79E-01 |
| FYB1 | 16058,53 | 0,04 | 0,06 | 0,66 | 5,10E-01 | 6,79E-01 |
| MC1R | 212,72 | -0,05 | 0,08 | -0,66 | 5,10E-01 | 6,79E-01 |
| ATF7-NPFF | 29,33 | -0,06 | 0,09 | -0,66 | 5,10E-01 | 6,79E-01 |
| AP1S3 | 123,09 | 0,06 | 0,09 | 0,66 | 5,10E-01 | 6,79E-01 |
| BLTP3B | 2985,10 | 0,04 | 0,06 | 0,66 | 5,11E-01 | 6,79E-01 |
| HIRIP3 | 209,84 | 0,03 | 0,05 | 0,66 | 5,11E-01 | 6,79E-01 |
| ARHGAP35 | 1489,42 | -0,03 | 0,05 | -0,66 | 5,11E-01 | 6,79E-01 |
| NPHP3 | 528,94 | 0,03 | 0,05 | 0,66 | 5,11E-01 | 6,79E-01 |
| SLC16A5 | 328,52 | -0,06 | 0,10 | -0,66 | 5,11E-01 | 6,79E-01 |
| IPO9 | 1640,85 | 0,02 | 0,04 | 0,66 | 5,11E-01 | 6,80E-01 |
| HTATIP2 | 1119,11 | -0,04 | 0,06 | -0,66 | 5,11E-01 | 6,80E-01 |

| | | | | | | |
|---|---|---|---|---|---|---|
| SEPTIN2 | 5556,33 | 0,03 | 0,04 | 0,66 | 5,12E-01 | 6,80E-01 |
| CFAP45 | 219,41 | -0,07 | 0,10 | -0,66 | 5,11E-01 | 6,80E-01 |
| PIGO | 397,54 | -0,04 | 0,06 | -0,66 | 5,12E-01 | 6,80E-01 |
| ENSG000002 | 33,05 | -0,07 | 0,11 | -0,66 | 5,12E-01 | 6,80E-01 |
| BTRC | 489,50 | -0,03 | 0,04 | -0,66 | 5,12E-01 | 6,80E-01 |
| ELOVL5 | 6308,66 | 0,03 | 0,05 | 0,66 | 5,12E-01 | 6,80E-01 |
| VPS13D | 5604,76 | -0,04 | 0,06 | -0,66 | 5,12E-01 | 6,80E-01 |
| CLEC10A | 184,97 | 0,06 | 0,09 | 0,66 | 5,12E-01 | 6,80E-01 |
| CLEC18A | 61,81 | -0,12 | 0,18 | -0,66 | 5,12E-01 | 6,80E-01 |
| EIF1AY | 874,91 | -0,16 | 0,24 | -0,65 | 5,13E-01 | 6,80E-01 |
| LPGAT1 | 7378,03 | 0,04 | 0,06 | 0,65 | 5,13E-01 | 6,80E-01 |
| GFI1 | 319,32 | -0,06 | 0,09 | -0,65 | 5,13E-01 | 6,80E-01 |
| RABL6 | 506,25 | -0,05 | 0,08 | -0,65 | 5,12E-01 | 6,80E-01 |
| SGCB | 150,52 | 0,05 | 0,07 | 0,65 | 5,13E-01 | 6,80E-01 |
| ZNF804A | 91,31 | 0,05 | 0,08 | 0,65 | 5,13E-01 | 6,80E-01 |
| NDRG2 | 154,49 | -0,06 | 0,09 | -0,65 | 5,13E-01 | 6,80E-01 |
| ULK4 | 1010,42 | 0,03 | 0,05 | 0,65 | 5,13E-01 | 6,80E-01 |
| TKT | 9040,00 | -0,04 | 0,07 | -0,65 | 5,13E-01 | 6,80E-01 |
| RITA1 | 212,07 | -0,02 | 0,04 | -0,65 | 5,13E-01 | 6,80E-01 |
| PGAP3 | 181,19 | -0,05 | 0,07 | -0,65 | 5,13E-01 | 6,80E-01 |
| NEBL | 229,29 | -0,22 | 0,33 | -0,65 | 5,13E-01 | 6,80E-01 |
| NCK2 | 2090,11 | -0,03 | 0,04 | -0,65 | 5,13E-01 | 6,80E-01 |
| SUPT5H | 1371,54 | -0,05 | 0,07 | -0,65 | 5,13E-01 | 6,80E-01 |
| APAF1 | 7918,09 | -0,04 | 0,06 | -0,65 | 5,13E-01 | 6,80E-01 |
| FBXL4 | 2289,12 | 0,03 | 0,05 | 0,65 | 5,13E-01 | 6,80E-01 |
| RP9 | 145,08 | 0,03 | 0,05 | 0,65 | 5,13E-01 | 6,81E-01 |
| HSPBP1 | 99,86 | -0,05 | 0,08 | -0,65 | 5,14E-01 | 6,81E-01 |
| FTSJ3 | 972,08 | 0,03 | 0,04 | 0,65 | 5,14E-01 | 6,81E-01 |
| MFSD4B | 1195,64 | 0,02 | 0,04 | 0,65 | 5,14E-01 | 6,81E-01 |
| RAD23B | 3638,37 | 0,03 | 0,05 | 0,65 | 5,14E-01 | 6,81E-01 |
| TMEM259 | 1239,52 | -0,06 | 0,09 | -0,65 | 5,15E-01 | 6,81E-01 |
| HSD17B7 | 519,76 | 0,02 | 0,03 | 0,65 | 5,14E-01 | 6,81E-01 |
| SDHB | 1144,83 | 0,03 | 0,05 | 0,65 | 5,15E-01 | 6,81E-01 |
| RBM6 | 2902,07 | 0,03 | 0,05 | 0,65 | 5,14E-01 | 6,81E-01 |
| SIDT2 | 582,10 | -0,08 | 0,12 | -0,65 | 5,15E-01 | 6,81E-01 |
| NAXD | 729,24 | 0,03 | 0,04 | 0,65 | 5,14E-01 | 6,81E-01 |
| ACTR3C | 476,65 | -0,03 | 0,05 | -0,65 | 5,15E-01 | 6,81E-01 |
| YTHDF1 | 784,84 | -0,02 | 0,03 | -0,65 | 5,14E-01 | 6,81E-01 |
| ADAMTS6 | 138,33 | 0,07 | 0,11 | 0,65 | 5,15E-01 | 6,81E-01 |
| ITGB2 | 18988,21 | -0,05 | 0,07 | -0,65 | 5,14E-01 | 6,81E-01 |
| NPTN | 1690,92 | -0,03 | 0,04 | -0,65 | 5,14E-01 | 6,81E-01 |
| KRIT1 | 893,42 | 0,04 | 0,06 | 0,65 | 5,14E-01 | 6,81E-01 |
| PPID | 353,46 | 0,03 | 0,04 | 0,65 | 5,15E-01 | 6,81E-01 |
| TRUB2 | 394,38 | 0,02 | 0,04 | 0,65 | 5,15E-01 | 6,81E-01 |
| DUSP10 | 291,45 | 0,03 | 0,04 | 0,65 | 5,15E-01 | 6,82E-01 |
| PGD | 6106,70 | -0,05 | 0,08 | -0,65 | 5,15E-01 | 6,82E-01 |

| | | | | | | |
|---|---|---|---|---|---|---|
| EHD4 | 672,86 | 0,03 | 0,04 | 0,65 | 5,16E-01 | 6,82E-01 |
| PICALM | 19412,40 | 0,04 | 0,06 | 0,65 | 5,15E-01 | 6,82E-01 |
| LRP2BP | 58,95 | -0,08 | 0,12 | -0,65 | 5,16E-01 | 6,82E-01 |
| CSDE1 | 14487,06 | 0,04 | 0,06 | 0,65 | 5,16E-01 | 6,82E-01 |
| ZMYND19 | 207,22 | -0,03 | 0,04 | -0,65 | 5,16E-01 | 6,82E-01 |
| EDEM2 | 1291,29 | -0,03 | 0,04 | -0,65 | 5,16E-01 | 6,82E-01 |
| DUSP6 | 2513,06 | 0,04 | 0,07 | 0,65 | 5,16E-01 | 6,82E-01 |
| DDX17 | 48058,84 | 0,02 | 0,04 | 0,65 | 5,17E-01 | 6,83E-01 |
| SLC37A3 | 1151,15 | -0,04 | 0,06 | -0,65 | 5,17E-01 | 6,83E-01 |
| CCDC167 | 106,36 | -0,06 | 0,09 | -0,65 | 5,17E-01 | 6,83E-01 |
| SOCS2 | 301,58 | -0,04 | 0,06 | -0,65 | 5,17E-01 | 6,83E-01 |
| NECTIN2 | 90,86 | -0,16 | 0,24 | -0,65 | 5,17E-01 | 6,83E-01 |
| GRHPR | 830,87 | 0,03 | 0,05 | 0,65 | 5,17E-01 | 6,83E-01 |
| C7orf50 | 428,13 | -0,04 | 0,06 | -0,65 | 5,17E-01 | 6,83E-01 |
| RACGAP1 | 287,23 | -0,03 | 0,04 | -0,65 | 5,17E-01 | 6,83E-01 |
| KATNBL1 | 3011,60 | -0,04 | 0,07 | -0,65 | 5,17E-01 | 6,83E-01 |
| NIBAN1 | 20692,76 | -0,06 | 0,09 | -0,65 | 5,17E-01 | 6,83E-01 |
| MGAT5 | 2562,97 | 0,03 | 0,04 | 0,65 | 5,18E-01 | 6,83E-01 |
| RBMS1 | 4266,41 | 0,03 | 0,05 | 0,65 | 5,18E-01 | 6,83E-01 |
| CCDC163 | 39,27 | -0,11 | 0,17 | -0,65 | 5,18E-01 | 6,83E-01 |
| PPP2R5A | 3944,13 | 0,04 | 0,06 | 0,65 | 5,18E-01 | 6,83E-01 |
| ENSG000002 | 83,07 | -0,06 | 0,09 | -0,65 | 5,18E-01 | 6,83E-01 |
| TMEM232 | 49,15 | -0,09 | 0,14 | -0,65 | 5,18E-01 | 6,83E-01 |
| SEC24D | 1054,53 | -0,02 | 0,04 | -0,65 | 5,18E-01 | 6,83E-01 |
| PRMT7 | 809,58 | -0,03 | 0,05 | -0,65 | 5,19E-01 | 6,84E-01 |
| DSTN | 525,79 | 0,03 | 0,05 | 0,65 | 5,19E-01 | 6,84E-01 |
| G6PC3 | 131,44 | -0,05 | 0,08 | -0,65 | 5,19E-01 | 6,84E-01 |
| ZSCAN25 | 1109,03 | -0,02 | 0,04 | -0,65 | 5,19E-01 | 6,84E-01 |
| CDK1 | 43,92 | -0,09 | 0,14 | -0,65 | 5,19E-01 | 6,84E-01 |
| DYM | 1866,88 | 0,02 | 0,03 | 0,65 | 5,19E-01 | 6,84E-01 |
| MEPCE | 748,63 | -0,03 | 0,05 | -0,64 | 5,19E-01 | 6,84E-01 |
| MFSD1 | 4108,86 | 0,04 | 0,07 | 0,64 | 5,19E-01 | 6,84E-01 |
| PEX16 | 863,78 | 0,02 | 0,04 | 0,64 | 5,19E-01 | 6,84E-01 |
| S1PR4 | 2400,84 | -0,04 | 0,06 | -0,64 | 5,19E-01 | 6,84E-01 |
| H3-3A | 10132,26 | -0,04 | 0,06 | -0,64 | 5,19E-01 | 6,84E-01 |
| SPATA1 | 62,18 | -0,07 | 0,10 | -0,64 | 5,19E-01 | 6,84E-01 |
| C1orf74 | 100,76 | 0,04 | 0,06 | 0,64 | 5,20E-01 | 6,85E-01 |
| TNFRSF10C | 4931,93 | -0,06 | 0,09 | -0,64 | 5,20E-01 | 6,85E-01 |
| IRAK4 | 3088,34 | 0,03 | 0,04 | 0,64 | 5,20E-01 | 6,85E-01 |
| MPI | 406,67 | -0,04 | 0,06 | -0,64 | 5,20E-01 | 6,85E-01 |
| ARIH1 | 4038,89 | 0,02 | 0,03 | 0,64 | 5,20E-01 | 6,85E-01 |
| POU5F2 | 1084,32 | -0,05 | 0,08 | -0,64 | 5,21E-01 | 6,86E-01 |
| UBQLN1 | 4605,93 | 0,03 | 0,05 | 0,64 | 5,21E-01 | 6,86E-01 |
| CDKL1 | 506,71 | -0,04 | 0,07 | -0,64 | 5,22E-01 | 6,87E-01 |
| WDR83 | 186,25 | -0,02 | 0,04 | -0,64 | 5,22E-01 | 6,87E-01 |
| TMEM254 | 186,78 | -0,02 | 0,04 | -0,64 | 5,22E-01 | 6,87E-01 |

| | | | | | | |
|---|---|---|---|---|---|---|
| POMK | 506,55 | -0,02 | 0,03 | -0,64 | 5,22E-01 | 6,87E-01 |
| MNDA | 26899,11 | 0,06 | 0,09 | 0,64 | 5,22E-01 | 6,87E-01 |
| WDR49 | 242,56 | 0,06 | 0,09 | 0,64 | 5,22E-01 | 6,87E-01 |
| CCR7 | 2008,76 | 0,06 | 0,10 | 0,64 | 5,22E-01 | 6,87E-01 |
| TEKTIP1 | 39,20 | -0,10 | 0,16 | -0,64 | 5,22E-01 | 6,87E-01 |
| OAT | 942,98 | 0,04 | 0,07 | 0,64 | 5,23E-01 | 6,88E-01 |
| ZNF135 | 83,63 | -0,08 | 0,13 | -0,64 | 5,23E-01 | 6,88E-01 |
| FBXO38 | 4521,34 | 0,03 | 0,05 | 0,64 | 5,23E-01 | 6,88E-01 |
| ABCC5 | 2646,39 | 0,04 | 0,07 | 0,64 | 5,23E-01 | 6,88E-01 |
| TRIR | 1123,21 | -0,03 | 0,05 | -0,64 | 5,23E-01 | 6,88E-01 |
| CDC26 | 316,89 | 0,04 | 0,06 | 0,64 | 5,23E-01 | 6,88E-01 |
| PRR5L | 2390,79 | -0,05 | 0,08 | -0,64 | 5,24E-01 | 6,89E-01 |
| USP9Y | 2008,77 | -0,15 | 0,24 | -0,64 | 5,24E-01 | 6,89E-01 |
| ELOVL4 | 55,16 | -0,09 | 0,14 | -0,64 | 5,24E-01 | 6,89E-01 |
| FKBPL | 57,69 | -0,04 | 0,07 | -0,64 | 5,24E-01 | 6,89E-01 |
| CAVIN2 | 2138,30 | -0,07 | 0,11 | -0,64 | 5,25E-01 | 6,89E-01 |
| CD70 | 27,53 | -0,08 | 0,13 | -0,64 | 5,25E-01 | 6,90E-01 |
| SNX9 | 601,48 | 0,03 | 0,05 | 0,64 | 5,25E-01 | 6,90E-01 |
| NUP188 | 650,19 | -0,04 | 0,06 | -0,64 | 5,25E-01 | 6,90E-01 |
| CDK19 | 2228,13 | -0,03 | 0,05 | -0,64 | 5,25E-01 | 6,90E-01 |
| UCK2 | 368,91 | 0,03 | 0,05 | 0,64 | 5,25E-01 | 6,90E-01 |
| PSTPIP1 | 3875,61 | -0,04 | 0,06 | -0,63 | 5,26E-01 | 6,90E-01 |
| BTBD9 | 1252,28 | -0,03 | 0,04 | -0,63 | 5,26E-01 | 6,90E-01 |
| NAXE | 246,92 | 0,02 | 0,04 | 0,63 | 5,26E-01 | 6,90E-01 |
| SLAMF1 | 614,85 | 0,04 | 0,06 | 0,63 | 5,26E-01 | 6,90E-01 |
| TCFL5 | 297,32 | 0,03 | 0,04 | 0,63 | 5,26E-01 | 6,90E-01 |
| BICD2 | 2932,57 | -0,03 | 0,05 | -0,63 | 5,26E-01 | 6,90E-01 |
| RAB1A | 2638,72 | 0,04 | 0,06 | 0,63 | 5,26E-01 | 6,90E-01 |
| ANAPC2 | 659,43 | -0,05 | 0,08 | -0,63 | 5,26E-01 | 6,90E-01 |
| ASGR2 | 112,42 | -0,08 | 0,12 | -0,63 | 5,26E-01 | 6,90E-01 |
| MZT2B | 1188,77 | -0,05 | 0,08 | -0,63 | 5,26E-01 | 6,90E-01 |
| KRT23 | 2031,01 | -0,07 | 0,11 | -0,63 | 5,26E-01 | 6,90E-01 |
| PRKCE | 797,72 | -0,05 | 0,08 | -0,63 | 5,26E-01 | 6,90E-01 |
| IFRD1 | 5326,37 | 0,05 | 0,08 | 0,63 | 5,26E-01 | 6,90E-01 |
| ATP6V1E2 | 192,17 | -0,03 | 0,05 | -0,63 | 5,27E-01 | 6,91E-01 |
| DERL2 | 1154,19 | 0,02 | 0,04 | 0,63 | 5,27E-01 | 6,91E-01 |
| HILPDA | 67,69 | -0,04 | 0,06 | -0,63 | 5,27E-01 | 6,91E-01 |
| DOK1 | 996,32 | -0,03 | 0,05 | -0,63 | 5,27E-01 | 6,91E-01 |
| CSNK1E | 300,83 | -0,04 | 0,07 | -0,63 | 5,28E-01 | 6,92E-01 |
| CALR | 1544,04 | -0,04 | 0,06 | -0,63 | 5,28E-01 | 6,92E-01 |
| CDC37 | 2133,06 | 0,03 | 0,04 | 0,63 | 5,28E-01 | 6,92E-01 |
| ETS2 | 2110,79 | -0,04 | 0,06 | -0,63 | 5,28E-01 | 6,92E-01 |
| TTLL5 | 648,69 | -0,04 | 0,06 | -0,63 | 5,29E-01 | 6,92E-01 |
| L3MBTL4 | 278,63 | -0,05 | 0,08 | -0,63 | 5,29E-01 | 6,92E-01 |
| B4GALT6 | 112,28 | 0,06 | 0,09 | 0,63 | 5,29E-01 | 6,92E-01 |
| TEX9 | 103,03 | -0,08 | 0,13 | -0,63 | 5,29E-01 | 6,92E-01 |

| | | | | | | |
|---|---|---|---|---|---|---|
| TADA2A | 584,13 | 0,02 | 0,03 | 0,63 | 5,29E-01 | 6,92E-01 |
| DAPK3 | 723,47 | -0,03 | 0,05 | -0,63 | 5,29E-01 | 6,93E-01 |
| SSBP4 | 398,23 | -0,06 | 0,09 | -0,63 | 5,29E-01 | 6,93E-01 |
| B4GAT1 | 71,11 | -0,04 | 0,07 | -0,63 | 5,29E-01 | 6,93E-01 |
| ZSWIM5 | 133,47 | -0,06 | 0,09 | -0,63 | 5,30E-01 | 6,93E-01 |
| ADCK1 | 154,99 | -0,06 | 0,10 | -0,63 | 5,30E-01 | 6,93E-01 |
| ZNF251 | 425,54 | 0,02 | 0,04 | 0,63 | 5,29E-01 | 6,93E-01 |
| BTK | 2004,07 | 0,03 | 0,05 | 0,63 | 5,29E-01 | 6,93E-01 |
| FCGRT | 3961,25 | -0,05 | 0,07 | -0,63 | 5,30E-01 | 6,93E-01 |
| KIF16B | 647,90 | 0,04 | 0,07 | 0,63 | 5,30E-01 | 6,93E-01 |
| SLC25A14 | 247,36 | 0,02 | 0,04 | 0,63 | 5,29E-01 | 6,93E-01 |
| C15orf40 | 1178,67 | 0,02 | 0,04 | 0,63 | 5,30E-01 | 6,93E-01 |
| SPATS2L | 191,53 | -0,08 | 0,13 | -0,63 | 5,29E-01 | 6,93E-01 |
| EIF4H | 4891,64 | 0,02 | 0,04 | 0,63 | 5,30E-01 | 6,93E-01 |
| CCT6B | 79,96 | -0,06 | 0,09 | -0,63 | 5,30E-01 | 6,93E-01 |
| TMCC1 | 2759,64 | -0,04 | 0,06 | -0,63 | 5,30E-01 | 6,93E-01 |
| SH3BGRL | 4011,78 | 0,04 | 0,06 | 0,63 | 5,30E-01 | 6,93E-01 |
| HELLS | 470,56 | 0,04 | 0,06 | 0,63 | 5,29E-01 | 6,93E-01 |
| TRAK2 | 5143,91 | -0,05 | 0,08 | -0,63 | 5,30E-01 | 6,93E-01 |
| CLNK | 68,19 | -0,11 | 0,17 | -0,63 | 5,30E-01 | 6,93E-01 |
| NDUFS3 | 372,96 | 0,03 | 0,05 | 0,63 | 5,30E-01 | 6,93E-01 |
| ZNF585B | 318,39 | 0,03 | 0,04 | 0,63 | 5,30E-01 | 6,93E-01 |
| TBC1D10C | 3498,08 | -0,03 | 0,05 | -0,63 | 5,29E-01 | 6,93E-01 |
| PARD6B | 206,54 | 0,03 | 0,05 | 0,63 | 5,30E-01 | 6,93E-01 |
| ATF6 | 4584,09 | 0,03 | 0,05 | 0,63 | 5,30E-01 | 6,93E-01 |
| CD53 | 12380,42 | -0,02 | 0,04 | -0,63 | 5,31E-01 | 6,94E-01 |
| SPECC1 | 4814,33 | -0,06 | 0,10 | -0,63 | 5,31E-01 | 6,94E-01 |
| CSRNP1 | 868,65 | -0,04 | 0,07 | -0,62 | 5,32E-01 | 6,95E-01 |
| NAGK | 4497,97 | 0,04 | 0,06 | 0,62 | 5,33E-01 | 6,95E-01 |
| GNRH1 | 112,34 | 0,03 | 0,05 | 0,62 | 5,33E-01 | 6,95E-01 |
| GPAT4 | 1917,72 | 0,02 | 0,03 | 0,62 | 5,33E-01 | 6,95E-01 |
| TMEM181 | 1960,89 | 0,03 | 0,05 | 0,62 | 5,33E-01 | 6,95E-01 |
| TXLNB | 73,10 | -0,05 | 0,09 | -0,62 | 5,33E-01 | 6,95E-01 |
| SPIN3 | 574,10 | 0,03 | 0,05 | 0,62 | 5,33E-01 | 6,95E-01 |
| ANKRD44 | 22030,77 | 0,03 | 0,04 | 0,62 | 5,32E-01 | 6,95E-01 |
| SQLE | 216,15 | 0,04 | 0,06 | 0,62 | 5,33E-01 | 6,95E-01 |
| DCTD | 565,27 | 0,02 | 0,03 | 0,62 | 5,33E-01 | 6,95E-01 |
| ASPH | 2776,91 | -0,05 | 0,08 | -0,62 | 5,34E-01 | 6,97E-01 |
| COPRS | 87,56 | 0,04 | 0,06 | 0,62 | 5,34E-01 | 6,97E-01 |
| SCART1 | 371,64 | -0,07 | 0,11 | -0,62 | 5,35E-01 | 6,97E-01 |
| CRAMP1 | 333,36 | -0,05 | 0,09 | -0,62 | 5,35E-01 | 6,97E-01 |
| FASLG | 198,94 | 0,06 | 0,09 | 0,62 | 5,35E-01 | 6,97E-01 |
| EXOSC4 | 171,82 | -0,03 | 0,05 | -0,62 | 5,35E-01 | 6,98E-01 |
| CAMK1 | 564,02 | -0,05 | 0,08 | -0,62 | 5,35E-01 | 6,98E-01 |
| GNG7 | 169,39 | -0,06 | 0,10 | -0,62 | 5,35E-01 | 6,98E-01 |
| RINL | 906,85 | -0,03 | 0,04 | -0,62 | 5,35E-01 | 6,98E-01 |

| | | | | | | |
|---|---|---|---|---|---|---|
| FES | 2412,65 | -0,04 | 0,07 | -0,62 | 5,35E-01 | 6,98E-01 |
| RAB38 | 29,85 | -0,07 | 0,11 | -0,62 | 5,35E-01 | 6,98E-01 |
| H3-3B | 26574,25 | 0,04 | 0,07 | 0,62 | 5,36E-01 | 6,98E-01 |
| NETO1 | 54,99 | -0,10 | 0,16 | -0,62 | 5,36E-01 | 6,98E-01 |
| DDX3Y | 2323,32 | -0,15 | 0,24 | -0,62 | 5,36E-01 | 6,98E-01 |
| ATXN1L | 1430,46 | -0,03 | 0,05 | -0,62 | 5,36E-01 | 6,98E-01 |
| PABPC4 | 1317,56 | -0,03 | 0,04 | -0,62 | 5,36E-01 | 6,98E-01 |
| PSMG3 | 275,91 | -0,03 | 0,04 | -0,62 | 5,36E-01 | 6,98E-01 |
| CD4 | 1232,49 | -0,05 | 0,08 | -0,62 | 5,36E-01 | 6,98E-01 |
| ARPC3 | 4740,53 | 0,04 | 0,07 | 0,62 | 5,36E-01 | 6,98E-01 |
| TBC1D10A | 476,77 | -0,05 | 0,08 | -0,62 | 5,36E-01 | 6,98E-01 |
| TWNK | 254,30 | 0,03 | 0,05 | 0,62 | 5,36E-01 | 6,98E-01 |
| GLTP | 970,71 | 0,02 | 0,04 | 0,62 | 5,36E-01 | 6,98E-01 |
| ERAP1 | 3372,02 | 0,02 | 0,04 | 0,62 | 5,36E-01 | 6,98E-01 |
| LPCAT1 | 2166,76 | -0,04 | 0,07 | -0,62 | 5,37E-01 | 6,98E-01 |
| COL9A2 | 187,96 | -0,08 | 0,13 | -0,62 | 5,37E-01 | 6,98E-01 |
| UPF3A | 985,27 | 0,03 | 0,04 | 0,62 | 5,37E-01 | 6,98E-01 |
| CD247 | 3990,88 | 0,04 | 0,06 | 0,62 | 5,37E-01 | 6,98E-01 |
| TMEM117 | 149,81 | 0,04 | 0,07 | 0,62 | 5,37E-01 | 6,98E-01 |
| DNAJA3 | 828,92 | 0,02 | 0,04 | 0,62 | 5,37E-01 | 6,98E-01 |
| ZNF862 | 1046,30 | -0,03 | 0,05 | -0,62 | 5,37E-01 | 6,98E-01 |
| ENSG000002 | 157,36 | -0,07 | 0,12 | -0,62 | 5,37E-01 | 6,98E-01 |
| ASXL1 | 2293,39 | -0,03 | 0,05 | -0,62 | 5,37E-01 | 6,99E-01 |
| SMARCA2 | 7407,94 | 0,02 | 0,04 | 0,62 | 5,38E-01 | 6,99E-01 |
| GTF3C5 | 700,73 | -0,02 | 0,03 | -0,62 | 5,38E-01 | 6,99E-01 |
| STRADA | 809,26 | -0,03 | 0,04 | -0,62 | 5,38E-01 | 6,99E-01 |
| NECAP1 | 1671,80 | 0,02 | 0,03 | 0,62 | 5,38E-01 | 6,99E-01 |
| GIGYF1 | 1034,82 | -0,07 | 0,12 | -0,62 | 5,38E-01 | 6,99E-01 |
| CD1A | 41,56 | -0,08 | 0,13 | -0,62 | 5,38E-01 | 6,99E-01 |
| MED31 | 451,89 | 0,04 | 0,06 | 0,61 | 5,39E-01 | 7,00E-01 |
| NBPF1 | 549,93 | -0,05 | 0,08 | -0,61 | 5,39E-01 | 7,00E-01 |
| RC3H2 | 4194,33 | 0,02 | 0,04 | 0,61 | 5,39E-01 | 7,00E-01 |
| NFAM1 | 2794,14 | -0,09 | 0,14 | -0,61 | 5,39E-01 | 7,00E-01 |
| RBL2 | 11598,14 | 0,03 | 0,04 | 0,61 | 5,39E-01 | 7,00E-01 |
| FBXL15 | 366,07 | -0,03 | 0,04 | -0,61 | 5,39E-01 | 7,00E-01 |
| VIPR1 | 432,75 | -0,06 | 0,09 | -0,61 | 5,39E-01 | 7,00E-01 |
| BEX2 | 86,43 | 0,06 | 0,10 | 0,61 | 5,40E-01 | 7,00E-01 |
| GADD45GIP1 | 206,89 | -0,04 | 0,06 | -0,61 | 5,40E-01 | 7,00E-01 |
| FCGR1A | 535,59 | -0,09 | 0,15 | -0,61 | 5,40E-01 | 7,01E-01 |
| BET1L | 480,55 | -0,05 | 0,08 | -0,61 | 5,40E-01 | 7,01E-01 |
| CD55 | 5761,88 | 0,04 | 0,07 | 0,61 | 5,40E-01 | 7,01E-01 |
| KCNMB4 | 59,95 | -0,07 | 0,11 | -0,61 | 5,40E-01 | 7,01E-01 |
| ZNF677 | 169,98 | -0,06 | 0,09 | -0,61 | 5,41E-01 | 7,02E-01 |
| RAB11A | 1859,13 | 0,04 | 0,06 | 0,61 | 5,41E-01 | 7,02E-01 |
| NT5C | 491,00 | 0,03 | 0,05 | 0,61 | 5,41E-01 | 7,02E-01 |
| ADARB1 | 852,24 | -0,05 | 0,08 | -0,61 | 5,41E-01 | 7,02E-01 |

| | | | | | | |
|---|---|---|---|---|---|---|
| ATP6V0D1 | 3413,70 | -0,04 | 0,07 | -0,61 | 5,41E-01 | 7,02E-01 |
| SH3KBP1 | 3017,90 | -0,03 | 0,05 | -0,61 | 5,42E-01 | 7,02E-01 |
| GPAT3 | 1251,09 | -0,05 | 0,08 | -0,61 | 5,42E-01 | 7,03E-01 |
| CEP19 | 720,46 | -0,05 | 0,08 | -0,61 | 5,43E-01 | 7,04E-01 |
| CXorf38 | 1818,47 | -0,02 | 0,03 | -0,61 | 5,43E-01 | 7,04E-01 |
| CRIP3 | 32,00 | -0,08 | 0,14 | -0,61 | 5,44E-01 | 7,04E-01 |
| NOP53 | 5876,42 | 0,04 | 0,07 | 0,61 | 5,43E-01 | 7,04E-01 |
| ABHD17A | 3583,94 | -0,03 | 0,05 | -0,61 | 5,43E-01 | 7,04E-01 |
| PAOX | 176,45 | -0,04 | 0,06 | -0,61 | 5,44E-01 | 7,04E-01 |
| SNRPN | 297,81 | -0,03 | 0,05 | -0,61 | 5,43E-01 | 7,04E-01 |
| CYB5A | 383,49 | -0,03 | 0,05 | -0,61 | 5,44E-01 | 7,04E-01 |
| PCDHGB7 | 58,48 | -0,08 | 0,13 | -0,61 | 5,44E-01 | 7,04E-01 |
| TRIM22 | 9966,62 | 0,05 | 0,08 | 0,61 | 5,44E-01 | 7,05E-01 |
| LDAF1 | 333,40 | -0,03 | 0,04 | -0,61 | 5,44E-01 | 7,05E-01 |
| EPHX2 | 432,17 | -0,06 | 0,09 | -0,61 | 5,44E-01 | 7,05E-01 |
| DSE | 2005,13 | -0,04 | 0,06 | -0,61 | 5,45E-01 | 7,05E-01 |
| ZDHHC2 | 3053,02 | 0,05 | 0,08 | 0,61 | 5,45E-01 | 7,05E-01 |
| LZTS2 | 289,07 | -0,04 | 0,07 | -0,60 | 5,46E-01 | 7,06E-01 |
| CDK5RAP2 | 2283,92 | 0,03 | 0,05 | 0,60 | 5,46E-01 | 7,06E-01 |
| ATP6V0E1 | 3344,55 | -0,04 | 0,07 | -0,60 | 5,46E-01 | 7,06E-01 |
| NBPF20 | 98,98 | -0,05 | 0,08 | -0,60 | 5,46E-01 | 7,06E-01 |
| TRPC1 | 221,75 | 0,05 | 0,09 | 0,60 | 5,46E-01 | 7,06E-01 |
| KIF3B | 940,07 | 0,02 | 0,04 | 0,60 | 5,46E-01 | 7,06E-01 |
| LCK | 3963,40 | 0,03 | 0,05 | 0,60 | 5,46E-01 | 7,07E-01 |
| E4F1 | 828,27 | -0,02 | 0,04 | -0,60 | 5,46E-01 | 7,07E-01 |
| NRDE2 | 1777,50 | -0,02 | 0,04 | -0,60 | 5,46E-01 | 7,07E-01 |
| BCOR | 625,82 | -0,04 | 0,06 | -0,60 | 5,47E-01 | 7,07E-01 |
| LTB4R2 | 319,07 | -0,03 | 0,06 | -0,60 | 5,47E-01 | 7,07E-01 |
| PCED1A | 569,10 | -0,03 | 0,05 | -0,60 | 5,47E-01 | 7,07E-01 |
| ZNF93 | 198,30 | 0,03 | 0,05 | 0,60 | 5,47E-01 | 7,07E-01 |
| DHRS7 | 2464,29 | 0,04 | 0,06 | 0,60 | 5,47E-01 | 7,08E-01 |
| PUM1 | 2941,54 | 0,02 | 0,03 | 0,60 | 5,47E-01 | 7,08E-01 |
| CREB3L2 | 846,70 | -0,04 | 0,06 | -0,60 | 5,48E-01 | 7,08E-01 |
| ENSG000002 | 136,26 | -0,08 | 0,13 | -0,60 | 5,48E-01 | 7,08E-01 |
| ZWILCH | 209,16 | 0,04 | 0,07 | 0,60 | 5,48E-01 | 7,08E-01 |
| DIP2B | 6490,71 | -0,03 | 0,05 | -0,60 | 5,48E-01 | 7,08E-01 |
| MTMR4 | 1912,10 | -0,02 | 0,04 | -0,60 | 5,48E-01 | 7,08E-01 |
| DHRS9 | 935,91 | 0,06 | 0,11 | 0,60 | 5,48E-01 | 7,08E-01 |
| TMEM42 | 391,57 | 0,03 | 0,05 | 0,60 | 5,48E-01 | 7,08E-01 |
| GALK2 | 1140,10 | 0,02 | 0,04 | 0,60 | 5,49E-01 | 7,08E-01 |
| HSD17B13 | 53,45 | -0,05 | 0,08 | -0,60 | 5,48E-01 | 7,08E-01 |
| GAS2L3 | 92,48 | -0,07 | 0,11 | -0,60 | 5,48E-01 | 7,08E-01 |
| SLFN13 | 1398,19 | -0,04 | 0,06 | -0,60 | 5,48E-01 | 7,08E-01 |
| NOPCHAP1 | 889,35 | 0,04 | 0,06 | 0,60 | 5,48E-01 | 7,08E-01 |
| HDLBP | 4555,02 | -0,04 | 0,06 | -0,60 | 5,48E-01 | 7,08E-01 |
| SF3B4 | 178,82 | -0,04 | 0,06 | -0,60 | 5,49E-01 | 7,08E-01 |

| | | | | | | |
|---|---|---|---|---|---|---|
| MXRA7 | 249,39 | -0,09 | 0,15 | -0,60 | 5,49E-01 | 7,08E-01 |
| STARD9 | 720,07 | -0,05 | 0,08 | -0,60 | 5,49E-01 | 7,08E-01 |
| RGP1 | 805,49 | -0,04 | 0,06 | -0,60 | 5,49E-01 | 7,08E-01 |
| DNPEP | 604,13 | -0,02 | 0,04 | -0,60 | 5,49E-01 | 7,08E-01 |
| CCNYL1 | 465,01 | 0,03 | 0,04 | 0,60 | 5,49E-01 | 7,09E-01 |
| CLP1 | 385,23 | 0,03 | 0,04 | 0,60 | 5,49E-01 | 7,09E-01 |
| MTMR1 | 1897,04 | 0,02 | 0,04 | 0,60 | 5,49E-01 | 7,09E-01 |
| CCDC144A | 724,51 | 0,17 | 0,28 | 0,60 | 5,49E-01 | 7,09E-01 |
| SIK2 | 655,75 | -0,02 | 0,04 | -0,60 | 5,50E-01 | 7,09E-01 |
| TTYH3 | 455,50 | -0,08 | 0,14 | -0,60 | 5,50E-01 | 7,09E-01 |
| ELMO1 | 4770,59 | -0,03 | 0,06 | -0,60 | 5,50E-01 | 7,09E-01 |
| VNN2 | 16638,55 | -0,06 | 0,10 | -0,60 | 5,50E-01 | 7,09E-01 |
| NUDT16L1 | 462,47 | -0,03 | 0,04 | -0,60 | 5,50E-01 | 7,09E-01 |
| RALB | 3708,76 | -0,04 | 0,07 | -0,60 | 5,50E-01 | 7,09E-01 |
| ANO10 | 461,15 | -0,03 | 0,05 | -0,60 | 5,50E-01 | 7,09E-01 |
| LYSMD1 | 42,22 | -0,04 | 0,07 | -0,60 | 5,51E-01 | 7,09E-01 |
| KIFC2 | 333,62 | -0,05 | 0,08 | -0,60 | 5,51E-01 | 7,10E-01 |
| HLA-DQB1 | 2910,95 | -0,10 | 0,16 | -0,60 | 5,51E-01 | 7,10E-01 |
| CNTROB | 275,10 | -0,04 | 0,06 | -0,60 | 5,51E-01 | 7,10E-01 |
| TNFRSF12A | 220,03 | -0,05 | 0,08 | -0,60 | 5,51E-01 | 7,10E-01 |
| LRCH3 | 2200,81 | -0,02 | 0,04 | -0,60 | 5,51E-01 | 7,10E-01 |
| CD58 | 1388,41 | 0,03 | 0,06 | 0,60 | 5,52E-01 | 7,10E-01 |
| TUT7 | 11723,28 | 0,04 | 0,06 | 0,60 | 5,52E-01 | 7,10E-01 |
| TMEM102 | 283,69 | -0,02 | 0,04 | -0,60 | 5,52E-01 | 7,10E-01 |
| TBP | 395,71 | 0,02 | 0,03 | 0,60 | 5,52E-01 | 7,10E-01 |
| PTPA | 781,72 | -0,03 | 0,05 | -0,59 | 5,52E-01 | 7,11E-01 |
| PPRC1 | 220,52 | -0,06 | 0,10 | -0,59 | 5,52E-01 | 7,11E-01 |
| NAMPT | 56536,38 | 0,06 | 0,10 | 0,59 | 5,52E-01 | 7,11E-01 |
| HERC5 | 1331,96 | 0,10 | 0,17 | 0,59 | 5,53E-01 | 7,11E-01 |
| ACO1 | 369,60 | 0,03 | 0,05 | 0,59 | 5,53E-01 | 7,11E-01 |
| TGFBR2 | 9906,92 | -0,02 | 0,04 | -0,59 | 5,53E-01 | 7,11E-01 |
| KAT2B | 6977,83 | -0,05 | 0,08 | -0,59 | 5,53E-01 | 7,11E-01 |
| BCL11B | 4141,68 | 0,03 | 0,06 | 0,59 | 5,53E-01 | 7,11E-01 |
| POLE4 | 313,88 | 0,03 | 0,05 | 0,59 | 5,54E-01 | 7,12E-01 |
| AIMP2 | 161,96 | -0,02 | 0,04 | -0,59 | 5,54E-01 | 7,12E-01 |
| YJU2 | 179,91 | 0,03 | 0,05 | 0,59 | 5,54E-01 | 7,12E-01 |
| TSPAN31 | 496,98 | 0,02 | 0,03 | 0,59 | 5,54E-01 | 7,12E-01 |
| USP19 | 2218,76 | -0,03 | 0,05 | -0,59 | 5,54E-01 | 7,12E-01 |
| MSL3 | 2283,26 | 0,02 | 0,04 | 0,59 | 5,54E-01 | 7,13E-01 |
| FAM72A | 31,91 | -0,06 | 0,11 | -0,59 | 5,55E-01 | 7,13E-01 |
| FXYD2 | 38,73 | -0,10 | 0,17 | -0,59 | 5,55E-01 | 7,13E-01 |
| GNA13 | 5807,20 | 0,03 | 0,06 | 0,59 | 5,55E-01 | 7,13E-01 |
| CD7 | 144,99 | -0,07 | 0,12 | -0,59 | 5,55E-01 | 7,13E-01 |
| XYLB | 114,67 | -0,06 | 0,10 | -0,59 | 5,56E-01 | 7,14E-01 |
| SON | 13428,17 | 0,02 | 0,03 | 0,59 | 5,56E-01 | 7,14E-01 |
| MTA3 | 344,28 | -0,03 | 0,06 | -0,59 | 5,56E-01 | 7,14E-01 |

| | | | | | | |
|---|---|---|---|---|---|---|
| PER3 | 226,85 | -0,04 | 0,08 | -0,59 | 5,56E-01 | 7,14E-01 |
| MAPK1 | 11348,70 | -0,03 | 0,05 | -0,59 | 5,56E-01 | 7,14E-01 |
| NDFIP1 | 2867,36 | 0,02 | 0,04 | 0,59 | 5,56E-01 | 7,14E-01 |
| PSMD4 | 2873,76 | -0,03 | 0,06 | -0,59 | 5,56E-01 | 7,14E-01 |
| SH3PXD2A | 313,00 | -0,07 | 0,11 | -0,59 | 5,56E-01 | 7,14E-01 |
| PLK3 | 390,87 | -0,05 | 0,09 | -0,59 | 5,56E-01 | 7,14E-01 |
| PPIL2 | 913,42 | -0,04 | 0,06 | -0,59 | 5,57E-01 | 7,15E-01 |
| CDKL3 | 39,66 | -0,05 | 0,08 | -0,59 | 5,57E-01 | 7,15E-01 |
| SPATA13 | 3592,86 | 0,02 | 0,04 | 0,59 | 5,57E-01 | 7,15E-01 |
| CFAP68 | 83,14 | -0,05 | 0,08 | -0,59 | 5,57E-01 | 7,15E-01 |
| TUBGCP4 | 556,76 | -0,03 | 0,06 | -0,59 | 5,57E-01 | 7,15E-01 |
| ADSL | 376,70 | -0,02 | 0,04 | -0,59 | 5,57E-01 | 7,15E-01 |
| MUL1 | 489,62 | 0,02 | 0,04 | 0,59 | 5,58E-01 | 7,15E-01 |
| FUOM | 233,07 | 0,03 | 0,05 | 0,59 | 5,58E-01 | 7,16E-01 |
| B3GNTL1 | 801,29 | -0,05 | 0,09 | -0,59 | 5,58E-01 | 7,16E-01 |
| SLC46A1 | 157,00 | -0,05 | 0,09 | -0,59 | 5,58E-01 | 7,16E-01 |
| ENSG000002 | 113,97 | 0,03 | 0,06 | 0,59 | 5,58E-01 | 7,16E-01 |
| MOB1A | 8876,25 | 0,03 | 0,05 | 0,59 | 5,58E-01 | 7,16E-01 |
| SYNJ1 | 1936,14 | 0,03 | 0,05 | 0,59 | 5,58E-01 | 7,16E-01 |
| PPP1R12C | 995,46 | -0,06 | 0,10 | -0,59 | 5,58E-01 | 7,16E-01 |
| MAGED2 | 1061,93 | 0,03 | 0,05 | 0,58 | 5,59E-01 | 7,16E-01 |
| LIN9 | 132,66 | 0,05 | 0,09 | 0,58 | 5,59E-01 | 7,16E-01 |
| LYRM1 | 605,65 | -0,02 | 0,04 | -0,58 | 5,59E-01 | 7,16E-01 |
| LRRC47 | 912,80 | 0,02 | 0,03 | 0,58 | 5,59E-01 | 7,16E-01 |
| FNIP1 | 4880,04 | 0,04 | 0,06 | 0,58 | 5,59E-01 | 7,16E-01 |
| FMC1 | 50,59 | -0,03 | 0,06 | -0,58 | 5,59E-01 | 7,16E-01 |
| ZNF320 | 109,95 | -0,04 | 0,07 | -0,58 | 5,59E-01 | 7,16E-01 |
| ENSG000002 | 57,77 | -0,08 | 0,14 | -0,58 | 5,59E-01 | 7,16E-01 |
| EME1 | 25,94 | -0,08 | 0,14 | -0,58 | 5,60E-01 | 7,17E-01 |
| CCDC9 | 478,75 | -0,04 | 0,07 | -0,58 | 5,60E-01 | 7,17E-01 |
| RCBTB2 | 2845,83 | 0,04 | 0,06 | 0,58 | 5,60E-01 | 7,17E-01 |
| RCAN1 | 313,78 | 0,02 | 0,04 | 0,58 | 5,60E-01 | 7,17E-01 |
| ATXN7 | 941,32 | -0,03 | 0,05 | -0,58 | 5,60E-01 | 7,17E-01 |
| MRPS12 | 240,55 | 0,03 | 0,05 | 0,58 | 5,60E-01 | 7,17E-01 |
| PIAS1 | 5621,45 | 0,03 | 0,05 | 0,58 | 5,60E-01 | 7,17E-01 |
| SUSD1 | 1237,60 | -0,03 | 0,05 | -0,58 | 5,61E-01 | 7,18E-01 |
| ABHD3 | 2898,29 | -0,03 | 0,06 | -0,58 | 5,61E-01 | 7,18E-01 |
| ENTPD4 | 3968,63 | 0,02 | 0,03 | 0,58 | 5,61E-01 | 7,18E-01 |
| PDRG1 | 296,41 | 0,02 | 0,04 | 0,58 | 5,62E-01 | 7,18E-01 |
| TRPC6 | 77,42 | -0,08 | 0,15 | -0,58 | 5,62E-01 | 7,19E-01 |
| MAPK14 | 10856,20 | -0,04 | 0,07 | -0,58 | 5,62E-01 | 7,19E-01 |
| GZMB | 1107,44 | 0,06 | 0,10 | 0,58 | 5,62E-01 | 7,19E-01 |
| ABHD17C | 53,85 | -0,07 | 0,12 | -0,58 | 5,62E-01 | 7,19E-01 |
| CCDC171 | 150,95 | -0,06 | 0,11 | -0,58 | 5,62E-01 | 7,19E-01 |
| CIITA | 2685,20 | -0,07 | 0,12 | -0,58 | 5,62E-01 | 7,19E-01 |
| AGO4 | 8823,93 | -0,03 | 0,06 | -0,58 | 5,62E-01 | 7,19E-01 |

| | | | | | | |
|---|---|---|---|---|---|---|
| EHHADH | 83,56 | -0,05 | 0,09 | -0,58 | 5,63E-01 | 7,19E-01 |
| GPA33 | 189,08 | -0,05 | 0,08 | -0,58 | 5,63E-01 | 7,19E-01 |
| HNRNPM | 2441,58 | 0,02 | 0,04 | 0,58 | 5,63E-01 | 7,19E-01 |
| ZNF610 | 35,60 | -0,07 | 0,12 | -0,58 | 5,63E-01 | 7,19E-01 |
| VMAC | 319,54 | -0,04 | 0,07 | -0,58 | 5,63E-01 | 7,20E-01 |
| MAP2K5 | 652,19 | 0,03 | 0,06 | 0,58 | 5,64E-01 | 7,20E-01 |
| ACYP1 | 218,72 | -0,03 | 0,06 | -0,58 | 5,64E-01 | 7,20E-01 |
| HSCB | 148,11 | 0,03 | 0,05 | 0,58 | 5,64E-01 | 7,20E-01 |
| MVP | 5779,85 | -0,04 | 0,06 | -0,58 | 5,63E-01 | 7,20E-01 |
| SERPINI1 | 167,31 | 0,05 | 0,08 | 0,58 | 5,64E-01 | 7,20E-01 |
| P2RY11 | 151,72 | 0,04 | 0,07 | 0,58 | 5,64E-01 | 7,20E-01 |
| TRIM68 | 306,37 | 0,02 | 0,03 | 0,58 | 5,64E-01 | 7,20E-01 |
| STYK1 | 95,89 | -0,08 | 0,13 | -0,58 | 5,64E-01 | 7,20E-01 |
| DUSP1 | 9238,91 | -0,05 | 0,09 | -0,58 | 5,64E-01 | 7,20E-01 |
| DDX6 | 17503,88 | 0,03 | 0,05 | 0,58 | 5,64E-01 | 7,20E-01 |
| RETREG1 | 575,17 | 0,04 | 0,08 | 0,58 | 5,64E-01 | 7,20E-01 |
| INTS4 | 1348,46 | 0,02 | 0,03 | 0,58 | 5,64E-01 | 7,20E-01 |
| FGFBP3 | 115,77 | 0,04 | 0,06 | 0,58 | 5,64E-01 | 7,20E-01 |
| INTS15 | 679,75 | -0,02 | 0,04 | -0,58 | 5,65E-01 | 7,21E-01 |
| CEACAM21 | 433,42 | -0,02 | 0,04 | -0,58 | 5,65E-01 | 7,21E-01 |
| NUP93 | 964,01 | 0,02 | 0,04 | 0,58 | 5,65E-01 | 7,21E-01 |
| BST2 | 1557,69 | 0,04 | 0,07 | 0,58 | 5,65E-01 | 7,21E-01 |
| VPS51 | 2764,40 | 0,02 | 0,04 | 0,58 | 5,65E-01 | 7,21E-01 |
| LPAR1 | 311,61 | 0,06 | 0,10 | 0,58 | 5,65E-01 | 7,21E-01 |
| CLTB | 1104,97 | -0,04 | 0,07 | -0,58 | 5,65E-01 | 7,21E-01 |
| RGS17 | 33,37 | 0,11 | 0,20 | 0,57 | 5,66E-01 | 7,21E-01 |
| WDR81 | 989,94 | -0,04 | 0,07 | -0,57 | 5,66E-01 | 7,21E-01 |
| STXBP5 | 5960,34 | 0,04 | 0,07 | 0,57 | 5,66E-01 | 7,21E-01 |
| ENSG000002 | 62,23 | -0,04 | 0,07 | -0,57 | 5,66E-01 | 7,21E-01 |
| ASGR1 | 231,61 | -0,04 | 0,07 | -0,57 | 5,66E-01 | 7,21E-01 |
| SOD1 | 1229,92 | 0,04 | 0,07 | 0,57 | 5,66E-01 | 7,21E-01 |
| CA13 | 221,51 | -0,03 | 0,05 | -0,57 | 5,66E-01 | 7,22E-01 |
| OXA1L | 1590,79 | -0,02 | 0,03 | -0,57 | 5,67E-01 | 7,22E-01 |
| PARP6 | 814,82 | -0,03 | 0,05 | -0,57 | 5,67E-01 | 7,23E-01 |
| CSNK2A2 | 1023,58 | 0,02 | 0,04 | 0,57 | 5,67E-01 | 7,23E-01 |
| KIF18A | 163,93 | -0,06 | 0,11 | -0,57 | 5,67E-01 | 7,23E-01 |
| C5AR2 | 4326,26 | -0,04 | 0,08 | -0,57 | 5,68E-01 | 7,23E-01 |
| NT5C2 | 6249,76 | 0,03 | 0,06 | 0,57 | 5,68E-01 | 7,23E-01 |
| SMPD2 | 219,75 | -0,03 | 0,06 | -0,57 | 5,68E-01 | 7,23E-01 |
| HECA | 5857,49 | 0,02 | 0,04 | 0,57 | 5,68E-01 | 7,23E-01 |
| STAMBP | 1259,16 | 0,02 | 0,03 | 0,57 | 5,68E-01 | 7,23E-01 |
| SPEF2 | 485,67 | 0,05 | 0,08 | 0,57 | 5,68E-01 | 7,23E-01 |
| SMIM35 | 47,98 | -0,08 | 0,15 | -0,57 | 5,68E-01 | 7,23E-01 |
| LIMD2 | 6293,71 | -0,03 | 0,05 | -0,57 | 5,68E-01 | 7,23E-01 |
| USP40 | 564,71 | -0,03 | 0,05 | -0,57 | 5,68E-01 | 7,23E-01 |
| MICOS13 | 262,94 | 0,03 | 0,06 | 0,57 | 5,68E-01 | 7,23E-01 |

| | | | | | | |
|---|---|---|---|---|---|---|
| PTGDR | 1113,77 | 0,06 | 0,10 | 0,57 | 5,68E-01 | 7,23E-01 |
| HS3ST3B1 | 554,18 | -0,03 | 0,05 | -0,57 | 5,68E-01 | 7,23E-01 |
| KLHDC10 | 1498,82 | 0,02 | 0,03 | 0,57 | 5,69E-01 | 7,23E-01 |
| IKZF2 | 2783,09 | 0,05 | 0,09 | 0,57 | 5,69E-01 | 7,24E-01 |
| LMLN | 163,42 | -0,04 | 0,07 | -0,57 | 5,69E-01 | 7,24E-01 |
| LMO7 | 481,02 | 0,04 | 0,06 | 0,57 | 5,69E-01 | 7,24E-01 |
| HGH1 | 142,91 | -0,04 | 0,07 | -0,57 | 5,70E-01 | 7,24E-01 |
| MTOR | 1849,20 | -0,03 | 0,05 | -0,57 | 5,70E-01 | 7,24E-01 |
| MFSD14A | 264,52 | 0,03 | 0,06 | 0,57 | 5,70E-01 | 7,24E-01 |
| TAF4B | 339,26 | 0,03 | 0,06 | 0,57 | 5,70E-01 | 7,24E-01 |
| PHACTR1 | 415,32 | -0,05 | 0,09 | -0,57 | 5,70E-01 | 7,25E-01 |
| CARMIL1 | 422,17 | -0,05 | 0,09 | -0,57 | 5,70E-01 | 7,25E-01 |
| GPS2 | 132,73 | -0,04 | 0,08 | -0,57 | 5,70E-01 | 7,25E-01 |
| CASS4 | 1476,82 | -0,04 | 0,07 | -0,57 | 5,71E-01 | 7,25E-01 |
| CYB561A3 | 1046,79 | -0,03 | 0,06 | -0,57 | 5,71E-01 | 7,26E-01 |
| SPAG8 | 55,55 | -0,04 | 0,08 | -0,57 | 5,71E-01 | 7,26E-01 |
| REX1BD | 681,53 | -0,03 | 0,05 | -0,57 | 5,71E-01 | 7,26E-01 |
| PDE6D | 504,25 | 0,02 | 0,03 | 0,57 | 5,72E-01 | 7,26E-01 |
| RAB5IF | 1180,92 | -0,04 | 0,07 | -0,56 | 5,72E-01 | 7,27E-01 |
| ZNF418 | 218,48 | 0,05 | 0,08 | 0,56 | 5,72E-01 | 7,27E-01 |
| ENSG000002 | 107,14 | -0,06 | 0,10 | -0,56 | 5,72E-01 | 7,27E-01 |
| NPL | 4070,60 | -0,04 | 0,07 | -0,56 | 5,73E-01 | 7,27E-01 |
| RNF34 | 2112,90 | 0,02 | 0,03 | 0,56 | 5,73E-01 | 7,27E-01 |
| XRCC1 | 222,68 | -0,04 | 0,08 | -0,56 | 5,72E-01 | 7,27E-01 |
| SIGIRR | 778,59 | -0,05 | 0,09 | -0,56 | 5,73E-01 | 7,27E-01 |
| RAD51B | 683,59 | 0,03 | 0,05 | 0,56 | 5,73E-01 | 7,27E-01 |
| CKS1B | 96,11 | 0,05 | 0,09 | 0,56 | 5,73E-01 | 7,27E-01 |
| IPCEF1 | 3066,29 | -0,02 | 0,04 | -0,56 | 5,73E-01 | 7,27E-01 |
| NEB | 313,70 | -0,07 | 0,13 | -0,56 | 5,73E-01 | 7,27E-01 |
| TXNDC17 | 341,54 | 0,04 | 0,07 | 0,56 | 5,73E-01 | 7,27E-01 |
| GPR19 | 74,03 | -0,04 | 0,07 | -0,56 | 5,73E-01 | 7,27E-01 |
| SYNE3 | 4474,27 | 0,03 | 0,06 | 0,56 | 5,73E-01 | 7,27E-01 |
| SAP30 | 217,04 | 0,04 | 0,07 | 0,56 | 5,73E-01 | 7,27E-01 |
| ABHD15 | 577,51 | -0,02 | 0,04 | -0,56 | 5,73E-01 | 7,27E-01 |
| SPG7 | 3648,37 | -0,03 | 0,06 | -0,56 | 5,73E-01 | 7,27E-01 |
| UBAP2L | 1622,56 | -0,02 | 0,04 | -0,56 | 5,74E-01 | 7,27E-01 |
| ZNF596 | 97,78 | 0,06 | 0,10 | 0,56 | 5,74E-01 | 7,27E-01 |
| STX8 | 457,78 | 0,02 | 0,04 | 0,56 | 5,74E-01 | 7,27E-01 |
| IMP4 | 735,05 | 0,02 | 0,03 | 0,56 | 5,74E-01 | 7,28E-01 |
| PAF1 | 341,10 | -0,03 | 0,05 | -0,56 | 5,74E-01 | 7,28E-01 |
| MRPL34 | 281,60 | -0,03 | 0,05 | -0,56 | 5,74E-01 | 7,28E-01 |
| SELENOH | 880,41 | 0,03 | 0,06 | 0,56 | 5,74E-01 | 7,28E-01 |
| NFKBIZ | 3674,91 | 0,03 | 0,05 | 0,56 | 5,75E-01 | 7,28E-01 |
| ZFP36L2 | 3776,23 | 0,03 | 0,06 | 0,56 | 5,75E-01 | 7,28E-01 |
| ARHGEF18 | 254,68 | -0,05 | 0,09 | -0,56 | 5,75E-01 | 7,28E-01 |
| HNRNPAB | 1195,24 | 0,02 | 0,04 | 0,56 | 5,75E-01 | 7,28E-01 |

| | | | | | | |
|---|---|---|---|---|---|---|
| MRPL44 | 736,32 | 0,03 | 0,05 | 0,56 | 5,75E-01 | 7,28E-01 |
| C9orf40 | 268,71 | -0,05 | 0,08 | -0,56 | 5,75E-01 | 7,28E-01 |
| MTFR1 | 183,09 | 0,03 | 0,05 | 0,56 | 5,75E-01 | 7,28E-01 |
| NDUFB6 | 571,14 | 0,04 | 0,07 | 0,56 | 5,75E-01 | 7,28E-01 |
| AHCYL1 | 1529,65 | 0,01 | 0,03 | 0,56 | 5,75E-01 | 7,28E-01 |
| DUS1L | 777,57 | -0,03 | 0,05 | -0,56 | 5,75E-01 | 7,28E-01 |
| SLC24A4 | 1315,51 | 0,06 | 0,10 | 0,56 | 5,76E-01 | 7,28E-01 |
| FBXL14 | 406,79 | -0,02 | 0,04 | -0,56 | 5,76E-01 | 7,28E-01 |
| SLC35B2 | 424,23 | 0,02 | 0,04 | 0,56 | 5,76E-01 | 7,28E-01 |
| PCNX1 | 14420,17 | 0,03 | 0,05 | 0,56 | 5,76E-01 | 7,28E-01 |
| TTC23 | 52,37 | -0,08 | 0,15 | -0,56 | 5,76E-01 | 7,29E-01 |
| FUZ | 293,66 | -0,03 | 0,06 | -0,56 | 5,76E-01 | 7,29E-01 |
| INPP4A | 7088,34 | -0,03 | 0,05 | -0,56 | 5,76E-01 | 7,29E-01 |
| PSMB9 | 5135,02 | -0,04 | 0,07 | -0,56 | 5,76E-01 | 7,29E-01 |
| RUSF1 | 525,75 | -0,03 | 0,06 | -0,56 | 5,77E-01 | 7,29E-01 |
| SLC38A10 | 1925,59 | -0,05 | 0,09 | -0,56 | 5,77E-01 | 7,29E-01 |
| CLDND2 | 174,58 | -0,06 | 0,11 | -0,56 | 5,77E-01 | 7,29E-01 |
| CHRNB1 | 209,40 | 0,03 | 0,06 | 0,56 | 5,77E-01 | 7,29E-01 |
| ACVR1C | 205,24 | 0,06 | 0,10 | 0,56 | 5,77E-01 | 7,29E-01 |
| CRCP | 799,86 | -0,02 | 0,04 | -0,56 | 5,77E-01 | 7,29E-01 |
| ACER3 | 1083,36 | 0,03 | 0,05 | 0,56 | 5,77E-01 | 7,29E-01 |
| CRK | 2283,59 | -0,02 | 0,04 | -0,56 | 5,77E-01 | 7,29E-01 |
| ZNF589 | 985,72 | 0,03 | 0,05 | 0,56 | 5,77E-01 | 7,30E-01 |
| LSM14A | 3890,78 | 0,02 | 0,04 | 0,56 | 5,77E-01 | 7,30E-01 |
| ANKRD39 | 145,17 | -0,02 | 0,04 | -0,56 | 5,78E-01 | 7,30E-01 |
| USP53 | 1408,44 | 0,05 | 0,10 | 0,56 | 5,78E-01 | 7,30E-01 |
| MRPL43 | 438,05 | -0,02 | 0,03 | -0,56 | 5,78E-01 | 7,30E-01 |
| PAX5 | 536,76 | 0,07 | 0,13 | 0,56 | 5,78E-01 | 7,30E-01 |
| ANP32A | 5258,92 | 0,03 | 0,05 | 0,56 | 5,78E-01 | 7,30E-01 |
| LIMS1 | 4261,58 | -0,04 | 0,07 | -0,56 | 5,78E-01 | 7,30E-01 |
| RHOBTB2 | 226,86 | -0,05 | 0,09 | -0,56 | 5,78E-01 | 7,30E-01 |
| ZNF169 | 632,18 | 0,02 | 0,04 | 0,56 | 5,78E-01 | 7,30E-01 |
| VAC14 | 636,64 | -0,04 | 0,08 | -0,56 | 5,78E-01 | 7,30E-01 |
| DCUN1D1 | 4564,55 | 0,05 | 0,09 | 0,56 | 5,78E-01 | 7,30E-01 |
| MAPK8IP3 | 2216,39 | -0,05 | 0,08 | -0,56 | 5,78E-01 | 7,30E-01 |
| LRRC57 | 666,27 | -0,02 | 0,04 | -0,56 | 5,79E-01 | 7,30E-01 |
| PILRB | 471,78 | 0,06 | 0,11 | 0,56 | 5,79E-01 | 7,30E-01 |
| ST20 | 765,14 | -0,04 | 0,08 | -0,55 | 5,79E-01 | 7,30E-01 |
| RABEPK | 348,58 | -0,02 | 0,04 | -0,55 | 5,79E-01 | 7,30E-01 |
| SLC2A6 | 655,66 | -0,04 | 0,07 | -0,55 | 5,79E-01 | 7,30E-01 |
| ZNF100 | 797,13 | -0,06 | 0,10 | -0,55 | 5,79E-01 | 7,30E-01 |
| NDUFB7 | 542,46 | 0,03 | 0,06 | 0,55 | 5,79E-01 | 7,31E-01 |
| MLLT11 | 98,30 | -0,03 | 0,06 | -0,55 | 5,79E-01 | 7,31E-01 |
| ARHGAP26 | 13369,18 | -0,04 | 0,07 | -0,55 | 5,80E-01 | 7,31E-01 |
| SNX19 | 3169,82 | 0,02 | 0,04 | 0,55 | 5,80E-01 | 7,31E-01 |
| ZNF599 | 159,80 | -0,03 | 0,06 | -0,55 | 5,80E-01 | 7,31E-01 |

| | | | | | | |
|---|---|---|---|---|---|---|
| SMDT1 | 202,29 | -0,03 | 0,06 | -0,55 | 5,80E-01 | 7,31E-01 |
| CA8 | 99,07 | -0,09 | 0,16 | -0,55 | 5,80E-01 | 7,31E-01 |
| CNPY3 | 1371,75 | -0,04 | 0,06 | -0,55 | 5,80E-01 | 7,31E-01 |
| KLRC3 | 126,54 | -0,09 | 0,16 | -0,55 | 5,80E-01 | 7,31E-01 |
| NDUFB4 | 803,63 | 0,03 | 0,05 | 0,55 | 5,80E-01 | 7,31E-01 |
| MAGEF1 | 205,48 | 0,03 | 0,06 | 0,55 | 5,81E-01 | 7,31E-01 |
| POP1 | 266,79 | 0,02 | 0,04 | 0,55 | 5,81E-01 | 7,31E-01 |
| C4orf3 | 2958,36 | 0,04 | 0,07 | 0,55 | 5,81E-01 | 7,32E-01 |
| ZC4H2 | 116,88 | -0,04 | 0,07 | -0,55 | 5,81E-01 | 7,32E-01 |
| PCCA | 549,69 | 0,03 | 0,05 | 0,55 | 5,82E-01 | 7,32E-01 |
| SERPINI2 | 57,70 | 0,07 | 0,12 | 0,55 | 5,82E-01 | 7,32E-01 |
| SLC35F2 | 149,76 | -0,04 | 0,08 | -0,55 | 5,82E-01 | 7,32E-01 |
| TM9SF4 | 807,21 | -0,04 | 0,07 | -0,55 | 5,82E-01 | 7,32E-01 |
| NMRAL1 | 594,80 | -0,03 | 0,05 | -0,55 | 5,82E-01 | 7,32E-01 |
| C12orf75 | 442,80 | -0,05 | 0,08 | -0,55 | 5,82E-01 | 7,32E-01 |
| CCDC85B | 501,78 | -0,03 | 0,06 | -0,55 | 5,82E-01 | 7,33E-01 |
| CATSPERE | 69,48 | -0,05 | 0,10 | -0,55 | 5,82E-01 | 7,33E-01 |
| LDLRAP1 | 1296,15 | -0,05 | 0,09 | -0,55 | 5,82E-01 | 7,33E-01 |
| MLEC | 1912,29 | -0,02 | 0,04 | -0,55 | 5,83E-01 | 7,33E-01 |
| SSX2IP | 547,16 | 0,04 | 0,08 | 0,55 | 5,83E-01 | 7,33E-01 |
| ZFP69B | 94,16 | -0,02 | 0,04 | -0,55 | 5,83E-01 | 7,33E-01 |
| RNF121 | 453,97 | -0,02 | 0,04 | -0,55 | 5,83E-01 | 7,33E-01 |
| HP1BP3 | 6228,07 | 0,02 | 0,03 | 0,55 | 5,83E-01 | 7,33E-01 |
| RPL36AL | 2046,82 | 0,05 | 0,09 | 0,55 | 5,83E-01 | 7,33E-01 |
| TP53 | 485,11 | -0,04 | 0,07 | -0,55 | 5,84E-01 | 7,34E-01 |
| GPKOW | 418,87 | -0,02 | 0,04 | -0,55 | 5,84E-01 | 7,34E-01 |
| NPC2 | 1677,32 | 0,03 | 0,06 | 0,55 | 5,84E-01 | 7,34E-01 |
| RUVBL2 | 465,90 | 0,02 | 0,04 | 0,55 | 5,84E-01 | 7,34E-01 |
| TIFAB | 47,11 | -0,06 | 0,11 | -0,55 | 5,85E-01 | 7,35E-01 |
| PTCD2 | 427,18 | 0,02 | 0,05 | 0,55 | 5,85E-01 | 7,35E-01 |
| HSD17B10 | 478,87 | 0,03 | 0,05 | 0,55 | 5,85E-01 | 7,35E-01 |
| CCNG2 | 2386,01 | 0,03 | 0,06 | 0,55 | 5,85E-01 | 7,35E-01 |
| TRIQK | 677,48 | 0,05 | 0,09 | 0,55 | 5,85E-01 | 7,35E-01 |
| SLC22A5 | 292,02 | 0,04 | 0,08 | 0,54 | 5,86E-01 | 7,36E-01 |
| MICA | 487,03 | -0,02 | 0,04 | -0,54 | 5,86E-01 | 7,36E-01 |
| ATP8B2 | 3922,73 | -0,03 | 0,05 | -0,54 | 5,86E-01 | 7,36E-01 |
| RIC8A | 1606,05 | -0,02 | 0,04 | -0,54 | 5,87E-01 | 7,37E-01 |
| LENG9 | 104,27 | -0,03 | 0,06 | -0,54 | 5,87E-01 | 7,37E-01 |
| ITPRID2 | 3441,97 | 0,03 | 0,06 | 0,54 | 5,87E-01 | 7,37E-01 |
| ATRIP | 221,33 | -0,02 | 0,04 | -0,54 | 5,87E-01 | 7,37E-01 |
| IL2RB | 633,14 | -0,06 | 0,12 | -0,54 | 5,87E-01 | 7,37E-01 |
| PPT1 | 4142,35 | 0,04 | 0,07 | 0,54 | 5,87E-01 | 7,37E-01 |
| MMP14 | 54,46 | -0,08 | 0,14 | -0,54 | 5,88E-01 | 7,37E-01 |
| HDGFL2 | 583,32 | -0,03 | 0,06 | -0,54 | 5,88E-01 | 7,37E-01 |
| MICAL3 | 902,87 | -0,04 | 0,08 | -0,54 | 5,87E-01 | 7,37E-01 |
| SPAG7 | 242,78 | 0,03 | 0,06 | 0,54 | 5,88E-01 | 7,37E-01 |

| | | | | | | |
|---|---|---|---|---|---|---|
| SGCD | 67,94 | -0,12 | 0,21 | -0,54 | 5,88E-01 | 7,37E-01 |
| NTHL1 | 153,13 | -0,03 | 0,05 | -0,54 | 5,88E-01 | 7,37E-01 |
| FCGR3A | 4925,02 | -0,05 | 0,08 | -0,54 | 5,88E-01 | 7,37E-01 |
| AP1G1 | 4173,56 | 0,02 | 0,03 | 0,54 | 5,88E-01 | 7,38E-01 |
| SIRT5 | 546,38 | -0,02 | 0,03 | -0,54 | 5,88E-01 | 7,38E-01 |
| SDHAF1 | 285,76 | 0,03 | 0,05 | 0,54 | 5,88E-01 | 7,38E-01 |
| TMEM150B | 80,52 | -0,06 | 0,12 | -0,54 | 5,89E-01 | 7,38E-01 |
| RPRD1B | 670,37 | -0,03 | 0,05 | -0,54 | 5,89E-01 | 7,38E-01 |
| ILF3 | 3670,45 | 0,02 | 0,04 | 0,54 | 5,89E-01 | 7,38E-01 |
| NUDT6 | 119,02 | 0,04 | 0,08 | 0,54 | 5,89E-01 | 7,38E-01 |
| KAT5 | 848,17 | 0,02 | 0,03 | 0,54 | 5,89E-01 | 7,38E-01 |
| PTGES | 29,38 | 0,10 | 0,18 | 0,54 | 5,89E-01 | 7,38E-01 |
| TFDP2 | 1388,28 | -0,02 | 0,04 | -0,54 | 5,89E-01 | 7,38E-01 |
| SLC20A2 | 745,16 | 0,02 | 0,04 | 0,54 | 5,89E-01 | 7,38E-01 |
| LY6G5B | 72,61 | -0,06 | 0,12 | -0,54 | 5,89E-01 | 7,38E-01 |
| SERAC1 | 420,27 | 0,03 | 0,05 | 0,54 | 5,89E-01 | 7,38E-01 |
| IFI44 | 1309,81 | 0,10 | 0,19 | 0,54 | 5,89E-01 | 7,38E-01 |
| PPP1R15B | 4822,29 | 0,02 | 0,04 | 0,54 | 5,90E-01 | 7,38E-01 |
| PRSS57 | 32,85 | -0,08 | 0,14 | -0,54 | 5,90E-01 | 7,39E-01 |
| TRMT1 | 517,34 | 0,02 | 0,04 | 0,54 | 5,90E-01 | 7,39E-01 |
| BRWD3 | 4276,20 | 0,03 | 0,05 | 0,54 | 5,90E-01 | 7,39E-01 |
| APBB3 | 399,11 | -0,03 | 0,06 | -0,54 | 5,90E-01 | 7,39E-01 |
| DST | 1020,78 | 0,04 | 0,08 | 0,54 | 5,90E-01 | 7,39E-01 |
| BBS12 | 106,34 | 0,04 | 0,08 | 0,54 | 5,90E-01 | 7,39E-01 |
| SAT2 | 155,95 | -0,03 | 0,06 | -0,54 | 5,91E-01 | 7,39E-01 |
| FASTK | 590,80 | -0,03 | 0,05 | -0,54 | 5,91E-01 | 7,39E-01 |
| ARHGEF40 | 1726,34 | -0,05 | 0,10 | -0,54 | 5,91E-01 | 7,39E-01 |
| ZNF429 | 588,91 | -0,04 | 0,08 | -0,54 | 5,91E-01 | 7,39E-01 |
| SPON1 | 77,24 | -0,08 | 0,14 | -0,54 | 5,91E-01 | 7,39E-01 |
| UBE2J1 | 2516,73 | 0,04 | 0,07 | 0,54 | 5,91E-01 | 7,39E-01 |
| ZNF404 | 43,38 | 0,06 | 0,12 | 0,54 | 5,91E-01 | 7,39E-01 |
| PTPN1 | 1877,77 | -0,02 | 0,04 | -0,54 | 5,91E-01 | 7,39E-01 |
| LYRM9 | 226,87 | 0,02 | 0,05 | 0,54 | 5,92E-01 | 7,40E-01 |
| PAPSS1 | 1573,48 | -0,04 | 0,08 | -0,54 | 5,92E-01 | 7,40E-01 |
| SPTLC3 | 55,19 | -0,09 | 0,17 | -0,54 | 5,92E-01 | 7,40E-01 |
| ACSF3 | 1557,81 | 0,03 | 0,06 | 0,54 | 5,92E-01 | 7,40E-01 |
| DICER1 | 10531,63 | 0,03 | 0,05 | 0,54 | 5,92E-01 | 7,40E-01 |
| TUBGCP2 | 1482,08 | -0,02 | 0,05 | -0,54 | 5,92E-01 | 7,40E-01 |
| HEATR6 | 535,19 | 0,02 | 0,04 | 0,54 | 5,92E-01 | 7,40E-01 |
| CDR2 | 723,71 | 0,03 | 0,05 | 0,54 | 5,92E-01 | 7,40E-01 |
| SEPTIN6 | 2371,38 | 0,03 | 0,05 | 0,54 | 5,92E-01 | 7,40E-01 |
| ZNF367 | 193,84 | 0,03 | 0,06 | 0,54 | 5,92E-01 | 7,40E-01 |
| DBF4B | 128,05 | -0,05 | 0,09 | -0,54 | 5,92E-01 | 7,40E-01 |
| ERF | 334,90 | -0,05 | 0,09 | -0,54 | 5,92E-01 | 7,40E-01 |
| BNIP1 | 105,89 | -0,03 | 0,06 | -0,54 | 5,93E-01 | 7,40E-01 |
| IDO1 | 208,08 | -0,08 | 0,16 | -0,53 | 5,93E-01 | 7,40E-01 |

| | | | | | | |
|---|---|---|---|---|---|---|
| TRMT2A | 539,09 | -0,03 | 0,05 | -0,53 | 5,93E-01 | 7,40E-01 |
| HIVEP1 | 1982,93 | 0,02 | 0,04 | 0,53 | 5,93E-01 | 7,40E-01 |
| NXPE3 | 1500,10 | -0,02 | 0,04 | -0,53 | 5,93E-01 | 7,40E-01 |
| SCAMP3 | 299,63 | -0,02 | 0,04 | -0,53 | 5,93E-01 | 7,40E-01 |
| CPEB2 | 1967,17 | 0,04 | 0,08 | 0,53 | 5,93E-01 | 7,40E-01 |
| RASA3 | 3492,56 | -0,04 | 0,07 | -0,53 | 5,93E-01 | 7,40E-01 |
| PRPSAP2 | 1014,65 | 0,02 | 0,03 | 0,53 | 5,93E-01 | 7,40E-01 |
| TRIM66 | 891,26 | -0,04 | 0,08 | -0,53 | 5,93E-01 | 7,40E-01 |
| TMED1 | 377,46 | -0,03 | 0,06 | -0,53 | 5,94E-01 | 7,41E-01 |
| ANAPC5 | 3511,75 | 0,01 | 0,03 | 0,53 | 5,94E-01 | 7,41E-01 |
| USP5 | 787,28 | 0,02 | 0,04 | 0,53 | 5,94E-01 | 7,41E-01 |
| OGFOD3 | 355,53 | -0,03 | 0,05 | -0,53 | 5,94E-01 | 7,41E-01 |
| CCDC86 | 95,61 | -0,04 | 0,07 | -0,53 | 5,94E-01 | 7,41E-01 |
| SETD9 | 110,12 | 0,05 | 0,09 | 0,53 | 5,94E-01 | 7,41E-01 |
| TMOD3 | 4256,37 | -0,02 | 0,03 | -0,53 | 5,94E-01 | 7,41E-01 |
| IL11RA | 409,06 | -0,05 | 0,09 | -0,53 | 5,94E-01 | 7,41E-01 |
| C2orf68 | 1787,11 | -0,02 | 0,03 | -0,53 | 5,94E-01 | 7,41E-01 |
| GTF2H2C | 394,60 | 0,03 | 0,05 | 0,53 | 5,94E-01 | 7,41E-01 |
| RAD54L2 | 1449,47 | -0,02 | 0,04 | -0,53 | 5,95E-01 | 7,41E-01 |
| ZHX3 | 282,99 | -0,04 | 0,08 | -0,53 | 5,95E-01 | 7,42E-01 |
| ZNF230 | 740,25 | 0,04 | 0,07 | 0,53 | 5,95E-01 | 7,42E-01 |
| ADAP2 | 233,13 | -0,04 | 0,08 | -0,53 | 5,95E-01 | 7,42E-01 |
| HUS1 | 874,08 | 0,02 | 0,04 | 0,53 | 5,96E-01 | 7,42E-01 |
| KLRC4-KLRK: | 233,61 | 0,07 | 0,14 | 0,53 | 5,96E-01 | 7,42E-01 |
| PARP4 | 4874,75 | 0,02 | 0,04 | 0,53 | 5,96E-01 | 7,43E-01 |
| H2AC8 | 1509,45 | 0,04 | 0,08 | 0,53 | 5,96E-01 | 7,43E-01 |
| MRPS26 | 346,98 | 0,02 | 0,04 | 0,53 | 5,97E-01 | 7,43E-01 |
| GNS | 5367,24 | -0,03 | 0,06 | -0,53 | 5,97E-01 | 7,43E-01 |
| HSF5 | 103,40 | 0,04 | 0,08 | 0,53 | 5,97E-01 | 7,43E-01 |
| BRIP1 | 243,31 | 0,05 | 0,09 | 0,53 | 5,97E-01 | 7,43E-01 |
| PFKL | 1955,65 | 0,03 | 0,06 | 0,53 | 5,97E-01 | 7,43E-01 |
| TAMM41 | 423,73 | -0,04 | 0,07 | -0,53 | 5,97E-01 | 7,43E-01 |
| ADAM15 | 443,17 | -0,04 | 0,08 | -0,53 | 5,97E-01 | 7,43E-01 |
| BTN2A2 | 492,58 | 0,03 | 0,06 | 0,53 | 5,97E-01 | 7,43E-01 |
| RBM8A | 2492,33 | 0,02 | 0,04 | 0,53 | 5,98E-01 | 7,44E-01 |
| SLC22A16 | 107,39 | -0,06 | 0,12 | -0,53 | 5,98E-01 | 7,44E-01 |
| ZNF727 | 25,08 | 0,10 | 0,18 | 0,53 | 5,98E-01 | 7,44E-01 |
| DDX19B | 215,27 | -0,02 | 0,04 | -0,53 | 5,98E-01 | 7,44E-01 |
| ZNF514 | 887,09 | 0,03 | 0,06 | 0,53 | 5,98E-01 | 7,44E-01 |
| SLC25A19 | 178,44 | -0,03 | 0,05 | -0,53 | 5,99E-01 | 7,44E-01 |
| SASH1 | 249,41 | -0,06 | 0,12 | -0,53 | 5,99E-01 | 7,44E-01 |
| NPIPB4 | 232,40 | -0,07 | 0,12 | -0,53 | 5,99E-01 | 7,44E-01 |
| STK25 | 880,89 | -0,03 | 0,05 | -0,53 | 5,99E-01 | 7,45E-01 |
| HERC2 | 2099,09 | -0,04 | 0,07 | -0,53 | 5,99E-01 | 7,45E-01 |
| RPP25L | 347,41 | 0,02 | 0,04 | 0,52 | 6,00E-01 | 7,45E-01 |
| LRP6 | 200,34 | -0,06 | 0,11 | -0,52 | 6,00E-01 | 7,45E-01 |

| | | | | | | |
|---|---|---|---|---|---|---|
| IFT52 | 365,66 | -0,03 | 0,05 | -0,52 | 6,00E-01 | 7,45E-01 |
| CDIN1 | 308,02 | 0,03 | 0,06 | 0,52 | 6,00E-01 | 7,45E-01 |
| MSL2 | 4136,69 | 0,02 | 0,03 | 0,52 | 6,00E-01 | 7,45E-01 |
| RYBP | 3035,26 | -0,03 | 0,06 | -0,52 | 6,00E-01 | 7,45E-01 |
| RRP1B | 648,25 | -0,03 | 0,06 | -0,52 | 6,00E-01 | 7,45E-01 |
| HAPLN3 | 585,34 | -0,04 | 0,07 | -0,52 | 6,00E-01 | 7,45E-01 |
| TNFRSF4 | 112,14 | -0,04 | 0,09 | -0,52 | 6,00E-01 | 7,45E-01 |
| TSEN2 | 420,86 | 0,02 | 0,04 | 0,52 | 6,00E-01 | 7,45E-01 |
| HYCC2 | 8088,40 | -0,04 | 0,07 | -0,52 | 6,00E-01 | 7,45E-01 |
| UQCC5 | 146,09 | 0,03 | 0,05 | 0,52 | 6,00E-01 | 7,45E-01 |
| MYL12A | 7771,42 | 0,03 | 0,06 | 0,52 | 6,00E-01 | 7,45E-01 |
| RAB33A | 71,09 | -0,03 | 0,06 | -0,52 | 6,01E-01 | 7,46E-01 |
| TRIM8 | 1761,63 | -0,04 | 0,08 | -0,52 | 6,01E-01 | 7,46E-01 |
| LRP1 | 2570,68 | -0,07 | 0,13 | -0,52 | 6,01E-01 | 7,46E-01 |
| TOR1B | 1038,00 | 0,03 | 0,06 | 0,52 | 6,01E-01 | 7,46E-01 |
| CAPNS1 | 472,02 | -0,04 | 0,07 | -0,52 | 6,01E-01 | 7,46E-01 |
| MCM2 | 228,62 | -0,04 | 0,07 | -0,52 | 6,01E-01 | 7,46E-01 |
| NUFIP1 | 91,50 | 0,04 | 0,07 | 0,52 | 6,01E-01 | 7,46E-01 |
| RAD51AP1 | 51,46 | 0,06 | 0,12 | 0,52 | 6,02E-01 | 7,46E-01 |
| ACP3 | 1123,49 | -0,04 | 0,08 | -0,52 | 6,02E-01 | 7,46E-01 |
| BCR | 1535,40 | -0,04 | 0,08 | -0,52 | 6,02E-01 | 7,46E-01 |
| MS4A2 | 155,50 | -0,10 | 0,20 | -0,52 | 6,02E-01 | 7,47E-01 |
| PPTC7 | 2741,35 | 0,02 | 0,04 | 0,52 | 6,02E-01 | 7,47E-01 |
| EBF1 | 290,49 | 0,06 | 0,11 | 0,52 | 6,03E-01 | 7,47E-01 |
| HK3 | 5206,00 | -0,04 | 0,07 | -0,52 | 6,03E-01 | 7,48E-01 |
| PDCD6 | 1991,79 | -0,02 | 0,03 | -0,52 | 6,03E-01 | 7,48E-01 |
| PSMD11 | 950,42 | -0,01 | 0,03 | -0,52 | 6,04E-01 | 7,48E-01 |
| CEACAM4 | 1025,19 | -0,05 | 0,09 | -0,52 | 6,04E-01 | 7,48E-01 |
| S100B | 61,87 | -0,12 | 0,24 | -0,52 | 6,04E-01 | 7,48E-01 |
| DHRS7B | 665,00 | -0,02 | 0,04 | -0,52 | 6,04E-01 | 7,48E-01 |
| ELAPOR2 | 358,85 | 0,06 | 0,12 | 0,52 | 6,04E-01 | 7,49E-01 |
| HPCAL4 | 136,75 | 0,06 | 0,11 | 0,52 | 6,04E-01 | 7,49E-01 |
| MED9 | 247,95 | -0,02 | 0,05 | -0,52 | 6,04E-01 | 7,49E-01 |
| ZSWIM6 | 2157,55 | 0,02 | 0,04 | 0,52 | 6,05E-01 | 7,49E-01 |
| SLC4A5 | 88,02 | -0,04 | 0,07 | -0,52 | 6,05E-01 | 7,49E-01 |
| IP6K2 | 1146,91 | -0,02 | 0,03 | -0,52 | 6,04E-01 | 7,49E-01 |
| SYTL3 | 1331,74 | -0,03 | 0,06 | -0,52 | 6,05E-01 | 7,49E-01 |
| SCRN2 | 229,07 | 0,03 | 0,06 | 0,52 | 6,05E-01 | 7,49E-01 |
| KDM8 | 198,42 | -0,03 | 0,06 | -0,52 | 6,05E-01 | 7,49E-01 |
| RAB3IP | 844,99 | 0,03 | 0,05 | 0,52 | 6,05E-01 | 7,49E-01 |
| ARHGAP25 | 9625,63 | -0,03 | 0,05 | -0,52 | 6,05E-01 | 7,49E-01 |
| RASSF7 | 290,88 | -0,03 | 0,05 | -0,52 | 6,05E-01 | 7,49E-01 |
| HDHD5 | 502,18 | 0,02 | 0,04 | 0,52 | 6,05E-01 | 7,49E-01 |
| HMGXB3 | 1109,70 | -0,02 | 0,04 | -0,52 | 6,05E-01 | 7,49E-01 |
| NBEA | 358,31 | 0,04 | 0,08 | 0,52 | 6,06E-01 | 7,49E-01 |
| MPP7 | 1801,80 | 0,03 | 0,06 | 0,52 | 6,06E-01 | 7,49E-01 |

| | | | | | | |
|---|---|---|---|---|---|---|
| STMN1 | 332,71 | 0,04 | 0,07 | 0,52 | 6,06E-01 | 7,49E-01 |
| NCAPD3 | 877,74 | -0,02 | 0,04 | -0,52 | 6,06E-01 | 7,49E-01 |
| DNAJC17 | 499,32 | -0,02 | 0,04 | -0,52 | 6,06E-01 | 7,49E-01 |
| MYL6 | 9536,98 | -0,06 | 0,11 | -0,52 | 6,06E-01 | 7,49E-01 |
| ZNF713 | 53,63 | -0,05 | 0,10 | -0,52 | 6,06E-01 | 7,49E-01 |
| C1QA | 106,85 | 0,07 | 0,13 | 0,52 | 6,06E-01 | 7,49E-01 |
| ANKDD1B | 74,10 | -0,05 | 0,09 | -0,52 | 6,06E-01 | 7,49E-01 |
| VPS11 | 899,14 | -0,02 | 0,03 | -0,52 | 6,06E-01 | 7,49E-01 |
| ENSG000002 | 32,20 | -0,05 | 0,10 | -0,52 | 6,06E-01 | 7,49E-01 |
| ZC3H7B | 798,40 | -0,04 | 0,08 | -0,52 | 6,06E-01 | 7,49E-01 |
| TOX4 | 2111,62 | -0,02 | 0,03 | -0,52 | 6,06E-01 | 7,49E-01 |
| ARRDC2 | 1558,56 | -0,03 | 0,05 | -0,52 | 6,06E-01 | 7,49E-01 |
| MAPDA | 181,96 | 0,03 | 0,06 | 0,52 | 6,06E-01 | 7,49E-01 |
| SRGAP2C | 557,44 | -0,03 | 0,07 | -0,52 | 6,06E-01 | 7,49E-01 |
| H1-5 | 769,86 | -0,05 | 0,10 | -0,51 | 6,07E-01 | 7,49E-01 |
| MYEF2 | 502,86 | 0,04 | 0,09 | 0,51 | 6,07E-01 | 7,49E-01 |
| FPGS | 509,02 | -0,03 | 0,06 | -0,51 | 6,07E-01 | 7,50E-01 |
| RPS2 | 17308,51 | 0,03 | 0,06 | 0,51 | 6,07E-01 | 7,50E-01 |
| ARHGAP32 | 348,85 | 0,03 | 0,06 | 0,51 | 6,07E-01 | 7,50E-01 |
| ZNF785 | 228,61 | -0,04 | 0,08 | -0,51 | 6,07E-01 | 7,50E-01 |
| IKBKE | 1277,04 | 0,02 | 0,05 | 0,51 | 6,07E-01 | 7,50E-01 |
| DHX8 | 2682,72 | 0,02 | 0,04 | 0,51 | 6,08E-01 | 7,50E-01 |
| H1-2 | 6074,96 | 0,04 | 0,07 | 0,51 | 6,08E-01 | 7,50E-01 |
| DOCK9 | 2098,46 | -0,03 | 0,06 | -0,51 | 6,08E-01 | 7,50E-01 |
| MANBA | 3980,61 | -0,03 | 0,05 | -0,51 | 6,08E-01 | 7,50E-01 |
| EIF4G1 | 2133,80 | -0,04 | 0,07 | -0,51 | 6,08E-01 | 7,50E-01 |
| ZNF275 | 544,79 | -0,03 | 0,06 | -0,51 | 6,08E-01 | 7,50E-01 |
| PEPD | 664,49 | -0,02 | 0,04 | -0,51 | 6,08E-01 | 7,50E-01 |
| IGFBP7 | 322,86 | 0,04 | 0,08 | 0,51 | 6,08E-01 | 7,50E-01 |
| OTUD7B | 578,53 | -0,02 | 0,05 | -0,51 | 6,08E-01 | 7,50E-01 |
| CMTR1 | 2147,49 | 0,02 | 0,05 | 0,51 | 6,08E-01 | 7,50E-01 |
| RLN1 | 50,99 | 0,05 | 0,10 | 0,51 | 6,08E-01 | 7,50E-01 |
| UAP1L1 | 147,97 | -0,06 | 0,12 | -0,51 | 6,09E-01 | 7,51E-01 |
| MCTP2 | 8924,58 | 0,03 | 0,07 | 0,51 | 6,09E-01 | 7,51E-01 |
| RNH1 | 1358,91 | 0,03 | 0,05 | 0,51 | 6,09E-01 | 7,51E-01 |
| AKAP13 | 20230,68 | -0,03 | 0,05 | -0,51 | 6,09E-01 | 7,51E-01 |
| SLCO4C1 | 1050,81 | -0,04 | 0,08 | -0,51 | 6,09E-01 | 7,51E-01 |
| POPDC2 | 83,48 | -0,03 | 0,07 | -0,51 | 6,09E-01 | 7,51E-01 |
| CLN6 | 321,08 | -0,04 | 0,08 | -0,51 | 6,09E-01 | 7,51E-01 |
| TMEM50A | 3423,06 | -0,03 | 0,06 | -0,51 | 6,10E-01 | 7,51E-01 |
| ATXN1 | 5161,07 | -0,03 | 0,06 | -0,51 | 6,10E-01 | 7,51E-01 |
| PPP1R9A | 121,05 | -0,07 | 0,14 | -0,51 | 6,10E-01 | 7,52E-01 |
| RAD50 | 147,05 | -0,02 | 0,04 | -0,51 | 6,10E-01 | 7,52E-01 |
| NME3 | 561,26 | 0,03 | 0,05 | 0,51 | 6,10E-01 | 7,52E-01 |
| DERL1 | 2187,43 | 0,02 | 0,03 | 0,51 | 6,11E-01 | 7,52E-01 |
| IGSF8 | 249,93 | -0,05 | 0,10 | -0,51 | 6,11E-01 | 7,52E-01 |

| | | | | | | |
|---|---|---|---|---|---|---|
| MRNIP | 485,46 | -0,04 | 0,08 | -0,51 | 6,11E-01 | 7,52E-01 |
| DHX33 | 364,43 | -0,02 | 0,03 | -0,51 | 6,11E-01 | 7,52E-01 |
| PLSCR1 | 3115,36 | 0,06 | 0,11 | 0,51 | 6,11E-01 | 7,52E-01 |
| ATP6V1D | 930,56 | 0,03 | 0,06 | 0,51 | 6,11E-01 | 7,53E-01 |
| MYLIP | 2652,29 | -0,03 | 0,06 | -0,51 | 6,11E-01 | 7,53E-01 |
| DYRK1A | 10740,75 | 0,02 | 0,04 | 0,51 | 6,11E-01 | 7,53E-01 |
| FGD6 | 360,27 | 0,04 | 0,07 | 0,51 | 6,12E-01 | 7,53E-01 |
| GLMP | 113,91 | -0,05 | 0,09 | -0,51 | 6,12E-01 | 7,53E-01 |
| PDCL3 | 168,52 | 0,03 | 0,06 | 0,51 | 6,12E-01 | 7,53E-01 |
| APOL6 | 9645,51 | 0,04 | 0,08 | 0,51 | 6,12E-01 | 7,53E-01 |
| GABPB2 | 1139,98 | -0,02 | 0,04 | -0,51 | 6,12E-01 | 7,53E-01 |
| CLIC4 | 979,19 | 0,04 | 0,07 | 0,51 | 6,12E-01 | 7,53E-01 |
| PTPN7 | 561,08 | -0,04 | 0,08 | -0,51 | 6,12E-01 | 7,53E-01 |
| GNAI3 | 7932,59 | 0,02 | 0,05 | 0,51 | 6,12E-01 | 7,53E-01 |
| ABCF1 | 786,69 | -0,03 | 0,05 | -0,51 | 6,13E-01 | 7,54E-01 |
| H2AC13 | 853,12 | -0,05 | 0,09 | -0,51 | 6,13E-01 | 7,54E-01 |
| ESS2 | 540,97 | -0,02 | 0,04 | -0,51 | 6,13E-01 | 7,54E-01 |
| SETD6 | 318,64 | -0,02 | 0,04 | -0,51 | 6,13E-01 | 7,54E-01 |
| CHMP4A | 159,79 | 0,03 | 0,05 | 0,51 | 6,13E-01 | 7,54E-01 |
| EPB41L3 | 1045,10 | 0,04 | 0,08 | 0,51 | 6,13E-01 | 7,54E-01 |
| KCNK6 | 589,80 | -0,03 | 0,05 | -0,51 | 6,13E-01 | 7,54E-01 |
| LY9 | 900,99 | -0,04 | 0,07 | -0,51 | 6,13E-01 | 7,54E-01 |
| TNFRSF25 | 1085,76 | -0,04 | 0,07 | -0,51 | 6,13E-01 | 7,54E-01 |
| EI24 | 584,82 | 0,01 | 0,03 | 0,50 | 6,14E-01 | 7,54E-01 |
| PPP1R3B | 4167,97 | -0,04 | 0,07 | -0,50 | 6,14E-01 | 7,54E-01 |
| GOLGA1 | 736,29 | -0,02 | 0,04 | -0,50 | 6,14E-01 | 7,54E-01 |
| NLRP12 | 4670,54 | -0,04 | 0,07 | -0,50 | 6,14E-01 | 7,54E-01 |
| RGPD2 | 203,51 | 0,11 | 0,22 | 0,50 | 6,14E-01 | 7,54E-01 |
| ELAC2 | 1224,00 | 0,01 | 0,03 | 0,50 | 6,14E-01 | 7,54E-01 |
| NDUFA8 | 235,47 | 0,03 | 0,05 | 0,50 | 6,14E-01 | 7,55E-01 |
| VMO1 | 28,45 | 0,08 | 0,16 | 0,50 | 6,15E-01 | 7,55E-01 |
| LEMD2 | 2362,62 | 0,01 | 0,03 | 0,50 | 6,15E-01 | 7,55E-01 |
| HSDL2 | 1806,91 | 0,04 | 0,07 | 0,50 | 6,15E-01 | 7,55E-01 |
| SPTLC2 | 4565,67 | 0,02 | 0,05 | 0,50 | 6,15E-01 | 7,55E-01 |
| SEC24B | 1709,04 | 0,02 | 0,03 | 0,50 | 6,15E-01 | 7,55E-01 |
| ZNF23 | 175,42 | -0,03 | 0,05 | -0,50 | 6,15E-01 | 7,55E-01 |
| FUT11 | 1065,71 | 0,02 | 0,05 | 0,50 | 6,15E-01 | 7,55E-01 |
| CCDC170 | 258,51 | -0,04 | 0,08 | -0,50 | 6,15E-01 | 7,55E-01 |
| PEX10 | 202,38 | -0,02 | 0,05 | -0,50 | 6,16E-01 | 7,55E-01 |
| THOC6 | 322,40 | 0,02 | 0,04 | 0,50 | 6,16E-01 | 7,55E-01 |
| SLC16A10 | 301,65 | 0,05 | 0,10 | 0,50 | 6,16E-01 | 7,55E-01 |
| SEL1L | 5312,61 | 0,02 | 0,05 | 0,50 | 6,16E-01 | 7,55E-01 |
| SLC39A11 | 632,19 | -0,03 | 0,06 | -0,50 | 6,16E-01 | 7,55E-01 |
| EIF4EBP1 | 73,30 | -0,04 | 0,08 | -0,50 | 6,16E-01 | 7,55E-01 |
| WWC2 | 319,62 | -0,03 | 0,06 | -0,50 | 6,16E-01 | 7,55E-01 |
| NMUR1 | 203,42 | -0,06 | 0,11 | -0,50 | 6,16E-01 | 7,55E-01 |

| | | | | | | |
|---|---|---|---|---|---|---|
| DEPDC7 | 34,18 | -0,07 | 0,13 | -0,50 | 6,16E-01 | 7,55E-01 |
| ZNF438 | 921,67 | -0,03 | 0,07 | -0,50 | 6,17E-01 | 7,55E-01 |
| TPPP3 | 64,63 | -0,06 | 0,12 | -0,50 | 6,17E-01 | 7,55E-01 |
| EPHB1 | 3345,68 | -0,04 | 0,08 | -0,50 | 6,16E-01 | 7,55E-01 |
| ERO1A | 3489,94 | 0,03 | 0,06 | 0,50 | 6,16E-01 | 7,55E-01 |
| GTF2IRD2 | 175,97 | -0,04 | 0,08 | -0,50 | 6,17E-01 | 7,55E-01 |
| ST3GAL6 | 1363,56 | 0,04 | 0,07 | 0,50 | 6,17E-01 | 7,55E-01 |
| SLC39A7 | 870,82 | 0,02 | 0,05 | 0,50 | 6,17E-01 | 7,56E-01 |
| NCBP2AS2 | 252,10 | 0,03 | 0,05 | 0,50 | 6,17E-01 | 7,56E-01 |
| DNAJC6 | 499,77 | -0,07 | 0,13 | -0,50 | 6,17E-01 | 7,56E-01 |
| CLEC4F | 72,36 | -0,12 | 0,24 | -0,50 | 6,17E-01 | 7,56E-01 |
| IL1RAP | 4229,95 | -0,04 | 0,08 | -0,50 | 6,18E-01 | 7,56E-01 |
| TBC1D22A | 2401,87 | -0,03 | 0,05 | -0,50 | 6,17E-01 | 7,56E-01 |
| MACROH2A1 | 10323,17 | 0,02 | 0,05 | 0,50 | 6,18E-01 | 7,56E-01 |
| DSP | 96,39 | -0,13 | 0,26 | -0,50 | 6,18E-01 | 7,56E-01 |
| SLAMF6 | 1067,78 | -0,03 | 0,06 | -0,50 | 6,18E-01 | 7,57E-01 |
| NEDD4 | 308,77 | 0,04 | 0,09 | 0,50 | 6,19E-01 | 7,57E-01 |
| HLA-C | 46777,21 | -0,04 | 0,08 | -0,50 | 6,19E-01 | 7,57E-01 |
| TARBP2 | 172,74 | -0,03 | 0,05 | -0,50 | 6,19E-01 | 7,57E-01 |
| FAM156A | 43,57 | 0,03 | 0,06 | 0,50 | 6,19E-01 | 7,57E-01 |
| NUP210L | 98,23 | 0,11 | 0,21 | 0,50 | 6,19E-01 | 7,57E-01 |
| MTLN | 124,03 | 0,03 | 0,07 | 0,50 | 6,19E-01 | 7,57E-01 |
| ARHGDIB | 8627,32 | -0,02 | 0,04 | -0,50 | 6,19E-01 | 7,57E-01 |
| GLI4 | 161,02 | -0,05 | 0,11 | -0,50 | 6,20E-01 | 7,58E-01 |
| GPR52 | 130,58 | -0,05 | 0,09 | -0,50 | 6,20E-01 | 7,58E-01 |
| TMED9 | 1279,68 | 0,02 | 0,04 | 0,50 | 6,20E-01 | 7,58E-01 |
| FAAH | 191,58 | -0,05 | 0,09 | -0,50 | 6,20E-01 | 7,58E-01 |
| GNA12 | 623,62 | -0,06 | 0,12 | -0,50 | 6,21E-01 | 7,58E-01 |
| TAF12 | 648,91 | -0,02 | 0,04 | -0,50 | 6,20E-01 | 7,58E-01 |
| ABCC4 | 1403,38 | -0,04 | 0,09 | -0,50 | 6,20E-01 | 7,58E-01 |
| GK | 5172,30 | -0,04 | 0,08 | -0,50 | 6,20E-01 | 7,58E-01 |
| C15orf61 | 178,13 | 0,04 | 0,07 | 0,50 | 6,20E-01 | 7,58E-01 |
| FANCI | 381,00 | -0,02 | 0,05 | -0,50 | 6,20E-01 | 7,58E-01 |
| PRKCQ | 1449,28 | 0,03 | 0,05 | 0,50 | 6,20E-01 | 7,58E-01 |
| DGKA | 3506,45 | -0,03 | 0,07 | -0,49 | 6,21E-01 | 7,59E-01 |
| NCAPG | 84,62 | -0,07 | 0,14 | -0,49 | 6,21E-01 | 7,59E-01 |
| EZH2 | 321,36 | -0,03 | 0,06 | -0,49 | 6,21E-01 | 7,59E-01 |
| ZCWPW2 | 40,81 | 0,04 | 0,08 | 0,49 | 6,21E-01 | 7,59E-01 |
| CCM2 | 4021,57 | -0,02 | 0,04 | -0,49 | 6,22E-01 | 7,59E-01 |
| TXNRD1 | 4153,50 | 0,03 | 0,06 | 0,49 | 6,21E-01 | 7,59E-01 |
| ACKR3 | 129,58 | -0,03 | 0,06 | -0,49 | 6,22E-01 | 7,59E-01 |
| HCK | 11212,09 | -0,04 | 0,07 | -0,49 | 6,22E-01 | 7,59E-01 |
| DNMT1 | 1713,63 | -0,03 | 0,05 | -0,49 | 6,22E-01 | 7,59E-01 |
| FLVCR2 | 349,21 | -0,03 | 0,07 | -0,49 | 6,22E-01 | 7,59E-01 |
| CX3CR1 | 2399,55 | -0,04 | 0,08 | -0,49 | 6,22E-01 | 7,59E-01 |
| ZNF547 | 95,90 | 0,04 | 0,08 | 0,49 | 6,23E-01 | 7,60E-01 |

| | | | | | | |
|---|---|---|---|---|---|---|
| NUGGC | 275,09 | -0,05 | 0,10 | -0,49 | 6,23E-01 | 7,60E-01 |
| ZKSCAN3 | 403,30 | -0,02 | 0,03 | -0,49 | 6,23E-01 | 7,60E-01 |
| RAB27A | 2978,88 | -0,02 | 0,05 | -0,49 | 6,23E-01 | 7,60E-01 |
| HEG1 | 725,78 | -0,03 | 0,06 | -0,49 | 6,23E-01 | 7,60E-01 |
| CFAP410 | 317,12 | -0,03 | 0,07 | -0,49 | 6,23E-01 | 7,60E-01 |
| CD81 | 760,04 | -0,04 | 0,08 | -0,49 | 6,23E-01 | 7,60E-01 |
| BRPF1 | 806,04 | -0,02 | 0,04 | -0,49 | 6,23E-01 | 7,60E-01 |
| TRAP1 | 592,53 | 0,03 | 0,05 | 0,49 | 6,24E-01 | 7,60E-01 |
| TTF2 | 639,78 | 0,02 | 0,05 | 0,49 | 6,24E-01 | 7,60E-01 |
| TRAPPC14 | 992,86 | -0,03 | 0,07 | -0,49 | 6,24E-01 | 7,60E-01 |
| DDX56 | 872,56 | -0,02 | 0,04 | -0,49 | 6,24E-01 | 7,61E-01 |
| URB1 | 960,87 | -0,03 | 0,06 | -0,49 | 6,24E-01 | 7,61E-01 |
| MACROD2 | 117,75 | 0,08 | 0,17 | 0,49 | 6,25E-01 | 7,61E-01 |
| VAPA | 4347,02 | 0,03 | 0,05 | 0,49 | 6,25E-01 | 7,61E-01 |
| ZNF621 | 790,07 | 0,02 | 0,04 | 0,49 | 6,25E-01 | 7,61E-01 |
| BAHD1 | 529,55 | -0,03 | 0,06 | -0,49 | 6,25E-01 | 7,61E-01 |
| CREB3 | 264,40 | 0,02 | 0,03 | 0,49 | 6,25E-01 | 7,61E-01 |
| GVQW3 | 217,54 | 0,03 | 0,06 | 0,49 | 6,25E-01 | 7,62E-01 |
| SH2D1B | 444,73 | 0,05 | 0,11 | 0,49 | 6,26E-01 | 7,62E-01 |
| SFT2D2 | 4783,15 | -0,03 | 0,06 | -0,49 | 6,26E-01 | 7,62E-01 |
| DAPK2 | 2194,37 | -0,04 | 0,09 | -0,49 | 6,26E-01 | 7,62E-01 |
| NAGPA | 379,28 | -0,03 | 0,05 | -0,49 | 6,26E-01 | 7,62E-01 |
| ACOT8 | 651,42 | -0,02 | 0,03 | -0,49 | 6,26E-01 | 7,62E-01 |
| HMGCR | 1968,87 | 0,03 | 0,06 | 0,49 | 6,27E-01 | 7,63E-01 |
| CTNNA1 | 2378,46 | 0,03 | 0,07 | 0,49 | 6,27E-01 | 7,63E-01 |
| MRPL14 | 305,47 | 0,02 | 0,05 | 0,49 | 6,27E-01 | 7,63E-01 |
| MEGF9 | 9725,98 | -0,04 | 0,07 | -0,49 | 6,27E-01 | 7,63E-01 |
| KIAA0232 | 5419,12 | 0,03 | 0,06 | 0,49 | 6,27E-01 | 7,63E-01 |
| ZNF565 | 117,69 | -0,03 | 0,06 | -0,49 | 6,27E-01 | 7,63E-01 |
| ARPC4 | 4835,87 | -0,03 | 0,05 | -0,49 | 6,28E-01 | 7,64E-01 |
| ZNF7 | 560,19 | 0,02 | 0,03 | 0,49 | 6,28E-01 | 7,64E-01 |
| WWOX | 445,07 | -0,03 | 0,06 | -0,49 | 6,28E-01 | 7,64E-01 |
| SMPDL3A | 184,86 | 0,04 | 0,08 | 0,48 | 6,28E-01 | 7,64E-01 |
| ICA1 | 127,26 | -0,04 | 0,08 | -0,48 | 6,28E-01 | 7,64E-01 |
| ST3GAL1 | 4562,76 | -0,03 | 0,06 | -0,48 | 6,28E-01 | 7,64E-01 |
| HSD17B11 | 5193,97 | 0,03 | 0,06 | 0,48 | 6,28E-01 | 7,64E-01 |
| CYRIA | 3507,18 | -0,03 | 0,05 | -0,48 | 6,29E-01 | 7,64E-01 |
| NANS | 510,29 | -0,02 | 0,04 | -0,48 | 6,29E-01 | 7,64E-01 |
| GYS1 | 349,10 | -0,04 | 0,07 | -0,48 | 6,29E-01 | 7,64E-01 |
| BCKDHA | 31,12 | -0,03 | 0,07 | -0,48 | 6,29E-01 | 7,64E-01 |
| NUDT3 | 2149,43 | 0,02 | 0,04 | 0,48 | 6,29E-01 | 7,64E-01 |
| MEF2A | 2109,43 | 0,02 | 0,05 | 0,48 | 6,29E-01 | 7,64E-01 |
| VAPB | 1572,15 | 0,02 | 0,04 | 0,48 | 6,29E-01 | 7,64E-01 |
| SCAF4 | 778,44 | 0,02 | 0,04 | 0,48 | 6,29E-01 | 7,65E-01 |
| TRIM39 | 519,51 | 0,01 | 0,03 | 0,48 | 6,29E-01 | 7,65E-01 |
| FNBP1 | 3531,66 | -0,02 | 0,05 | -0,48 | 6,30E-01 | 7,65E-01 |

| | | | | | | |
|---|---|---|---|---|---|---|
| NCOR1 | 5844,68 | -0,02 | 0,04 | -0,48 | 6,30E-01 | 7,65E-01 |
| NETO2 | 148,41 | -0,04 | 0,07 | -0,48 | 6,30E-01 | 7,65E-01 |
| TDP2 | 2581,11 | 0,03 | 0,06 | 0,48 | 6,30E-01 | 7,65E-01 |
| CKAP2 | 421,75 | 0,03 | 0,06 | 0,48 | 6,30E-01 | 7,65E-01 |
| NCEH1 | 266,54 | 0,02 | 0,04 | 0,48 | 6,30E-01 | 7,65E-01 |
| TTN | 6471,84 | 0,04 | 0,09 | 0,48 | 6,30E-01 | 7,65E-01 |
| APOL1 | 903,88 | -0,04 | 0,09 | -0,48 | 6,31E-01 | 7,66E-01 |
| GLIPR2 | 4713,49 | -0,03 | 0,06 | -0,48 | 6,31E-01 | 7,66E-01 |
| GLYR1 | 2057,25 | -0,02 | 0,04 | -0,48 | 6,31E-01 | 7,66E-01 |
| ERC1 | 1512,67 | 0,03 | 0,06 | 0,48 | 6,31E-01 | 7,66E-01 |
| ZNF740 | 2255,88 | 0,02 | 0,03 | 0,48 | 6,31E-01 | 7,66E-01 |
| ABHD2 | 8098,34 | -0,04 | 0,08 | -0,48 | 6,31E-01 | 7,66E-01 |
| ENSG000002 | 237,46 | 0,13 | 0,27 | 0,48 | 6,31E-01 | 7,66E-01 |
| GOLPH3 | 2281,63 | 0,02 | 0,04 | 0,48 | 6,31E-01 | 7,66E-01 |
| TP53I13 | 595,34 | -0,03 | 0,06 | -0,48 | 6,31E-01 | 7,66E-01 |
| UBC | 11209,34 | 0,03 | 0,06 | 0,48 | 6,31E-01 | 7,66E-01 |
| ARHGAP31 | 634,03 | 0,03 | 0,06 | 0,48 | 6,32E-01 | 7,66E-01 |
| MYZAP | 27,12 | -0,08 | 0,16 | -0,48 | 6,32E-01 | 7,66E-01 |
| RUNX1 | 2879,26 | -0,02 | 0,05 | -0,48 | 6,32E-01 | 7,66E-01 |
| SPPL2A | 2297,20 | 0,02 | 0,04 | 0,48 | 6,32E-01 | 7,66E-01 |
| DHX32 | 339,18 | 0,02 | 0,05 | 0,48 | 6,32E-01 | 7,66E-01 |
| KMT2A | 3951,46 | -0,03 | 0,06 | -0,48 | 6,32E-01 | 7,66E-01 |
| FNBP1L | 278,15 | -0,04 | 0,08 | -0,48 | 6,32E-01 | 7,66E-01 |
| CLEC4E | 2409,05 | -0,05 | 0,10 | -0,48 | 6,32E-01 | 7,66E-01 |
| INTS9 | 441,65 | -0,02 | 0,03 | -0,48 | 6,32E-01 | 7,66E-01 |
| RAB33B | 1046,22 | 0,04 | 0,07 | 0,48 | 6,33E-01 | 7,67E-01 |
| ZNF619 | 378,27 | -0,02 | 0,04 | -0,48 | 6,33E-01 | 7,67E-01 |
| EIF1B | 943,91 | -0,03 | 0,07 | -0,48 | 6,33E-01 | 7,67E-01 |
| APOBEC3H | 38,23 | -0,05 | 0,11 | -0,48 | 6,33E-01 | 7,67E-01 |
| CAPN10 | 1030,66 | -0,02 | 0,05 | -0,48 | 6,33E-01 | 7,67E-01 |
| KIF24 | 57,62 | -0,05 | 0,10 | -0,48 | 6,33E-01 | 7,67E-01 |
| DACT1 | 123,90 | 0,07 | 0,15 | 0,48 | 6,33E-01 | 7,67E-01 |
| TLE5 | 1197,83 | -0,04 | 0,08 | -0,48 | 6,33E-01 | 7,67E-01 |
| C9orf78 | 7099,04 | -0,06 | 0,13 | -0,48 | 6,34E-01 | 7,68E-01 |
| UBR5 | 7178,17 | 0,02 | 0,04 | 0,48 | 6,34E-01 | 7,68E-01 |
| TNS3 | 742,88 | -0,04 | 0,09 | -0,48 | 6,34E-01 | 7,68E-01 |
| HKDC1 | 174,25 | 0,05 | 0,11 | 0,48 | 6,35E-01 | 7,68E-01 |
| TWSG1 | 309,98 | 0,03 | 0,06 | 0,48 | 6,35E-01 | 7,68E-01 |
| RPL39L | 35,66 | -0,05 | 0,10 | -0,48 | 6,35E-01 | 7,68E-01 |
| NUDT18 | 261,70 | -0,03 | 0,05 | -0,48 | 6,34E-01 | 7,68E-01 |
| LRRC27 | 213,31 | -0,03 | 0,06 | -0,48 | 6,35E-01 | 7,68E-01 |
| SCFD2 | 747,43 | 0,02 | 0,04 | 0,48 | 6,35E-01 | 7,68E-01 |
| LASP1NB | 171,41 | -0,03 | 0,06 | -0,47 | 6,35E-01 | 7,68E-01 |
| POMT1 | 503,97 | -0,02 | 0,05 | -0,47 | 6,35E-01 | 7,68E-01 |
| ZNF74 | 234,51 | -0,03 | 0,06 | -0,47 | 6,35E-01 | 7,68E-01 |
| RP2 | 3121,79 | 0,04 | 0,08 | 0,47 | 6,35E-01 | 7,69E-01 |

| | | | | | | |
|---|---|---|---|---|---|---|
| ZNF609 | 2043,28 | -0,02 | 0,05 | -0,47 | 6,36E-01 | 7,69E-01 |
| ESYT2 | 4446,39 | 0,02 | 0,04 | 0,47 | 6,36E-01 | 7,69E-01 |
| PRKN | 173,00 | -0,04 | 0,08 | -0,47 | 6,36E-01 | 7,69E-01 |
| FAHD1 | 470,70 | -0,03 | 0,07 | -0,47 | 6,36E-01 | 7,69E-01 |
| TMEM131L | 4581,52 | -0,02 | 0,04 | -0,47 | 6,36E-01 | 7,69E-01 |
| PPARGC1B | 459,77 | -0,04 | 0,08 | -0,47 | 6,36E-01 | 7,69E-01 |
| REXO2 | 708,51 | -0,03 | 0,06 | -0,47 | 6,36E-01 | 7,69E-01 |
| ATG7 | 2841,45 | -0,03 | 0,06 | -0,47 | 6,36E-01 | 7,69E-01 |
| CTPS2 | 287,20 | 0,02 | 0,04 | 0,47 | 6,36E-01 | 7,69E-01 |
| ENDOD1 | 1337,09 | -0,03 | 0,07 | -0,47 | 6,37E-01 | 7,69E-01 |
| KCTD11 | 121,98 | -0,03 | 0,06 | -0,47 | 6,37E-01 | 7,69E-01 |
| EIF4ENIF1 | 673,00 | -0,01 | 0,03 | -0,47 | 6,37E-01 | 7,69E-01 |
| TCEANC2 | 1010,68 | -0,01 | 0,03 | -0,47 | 6,37E-01 | 7,69E-01 |
| NRDC | 5643,18 | -0,02 | 0,05 | -0,47 | 6,37E-01 | 7,69E-01 |
| KPNA4 | 4100,87 | 0,02 | 0,04 | 0,47 | 6,37E-01 | 7,69E-01 |
| ERN1 | 3056,74 | -0,03 | 0,07 | -0,47 | 6,37E-01 | 7,69E-01 |
| HAUS4 | 722,36 | -0,04 | 0,08 | -0,47 | 6,38E-01 | 7,70E-01 |
| AIF1 | 1412,80 | 0,05 | 0,10 | 0,47 | 6,38E-01 | 7,70E-01 |
| ATP5F1B | 5993,80 | -0,02 | 0,05 | -0,47 | 6,38E-01 | 7,70E-01 |
| SAYSD1 | 410,22 | 0,02 | 0,05 | 0,47 | 6,38E-01 | 7,70E-01 |
| MAN2C1 | 1694,60 | -0,04 | 0,08 | -0,47 | 6,38E-01 | 7,70E-01 |
| NDC80 | 139,25 | 0,03 | 0,07 | 0,47 | 6,38E-01 | 7,70E-01 |
| RPS4Y1 | 2151,77 | -0,12 | 0,25 | -0,47 | 6,38E-01 | 7,70E-01 |
| NEIL2 | 316,07 | -0,03 | 0,07 | -0,47 | 6,39E-01 | 7,70E-01 |
| CCDC117 | 1081,41 | 0,02 | 0,04 | 0,47 | 6,39E-01 | 7,70E-01 |
| TAS2R3 | 130,69 | -0,04 | 0,09 | -0,47 | 6,39E-01 | 7,70E-01 |
| SGK1 | 2750,14 | -0,03 | 0,07 | -0,47 | 6,39E-01 | 7,70E-01 |
| QARS1 | 1893,17 | 0,01 | 0,03 | 0,47 | 6,39E-01 | 7,70E-01 |
| FAM228B | 395,45 | 0,02 | 0,03 | 0,47 | 6,39E-01 | 7,70E-01 |
| PGM2L1 | 632,44 | 0,03 | 0,07 | 0,47 | 6,39E-01 | 7,70E-01 |
| KATNAL1 | 489,09 | 0,03 | 0,06 | 0,47 | 6,39E-01 | 7,71E-01 |
| SLC27A5 | 145,23 | -0,03 | 0,07 | -0,47 | 6,39E-01 | 7,71E-01 |
| CD46 | 13826,11 | 0,03 | 0,06 | 0,47 | 6,39E-01 | 7,71E-01 |
| CAMKMT | 593,10 | 0,02 | 0,05 | 0,47 | 6,40E-01 | 7,71E-01 |
| ELL2 | 582,22 | -0,03 | 0,05 | -0,47 | 6,40E-01 | 7,71E-01 |
| IRF2BP1 | 419,14 | -0,02 | 0,04 | -0,47 | 6,40E-01 | 7,71E-01 |
| SLC25A32 | 319,57 | 0,03 | 0,06 | 0,47 | 6,40E-01 | 7,71E-01 |
| CATSPERB | 244,06 | -0,04 | 0,08 | -0,47 | 6,40E-01 | 7,71E-01 |
| LSP1 | 8802,90 | 0,04 | 0,08 | 0,47 | 6,40E-01 | 7,71E-01 |
| H2BC6 | 624,01 | -0,03 | 0,07 | -0,47 | 6,40E-01 | 7,71E-01 |
| ATP6V1E1 | 1621,57 | -0,03 | 0,05 | -0,47 | 6,41E-01 | 7,72E-01 |
| ZNF839 | 465,81 | -0,02 | 0,05 | -0,47 | 6,41E-01 | 7,72E-01 |
| BBS2 | 974,80 | 0,02 | 0,05 | 0,47 | 6,41E-01 | 7,72E-01 |
| RBPJ | 5054,64 | 0,03 | 0,06 | 0,47 | 6,41E-01 | 7,72E-01 |
| TMEM116 | 730,63 | 0,02 | 0,04 | 0,47 | 6,41E-01 | 7,72E-01 |
| SUPT6H | 2261,85 | -0,03 | 0,07 | -0,47 | 6,41E-01 | 7,72E-01 |

| | | | | | | |
|---|---|---|---|---|---|---|
| BTF3 | 3279,13 | -0,04 | 0,08 | -0,47 | 6,41E-01 | 7,72E-01 |
| TMSB15B | 44,31 | -0,03 | 0,07 | -0,47 | 6,42E-01 | 7,72E-01 |
| RAB27B | 1431,63 | -0,05 | 0,10 | -0,47 | 6,42E-01 | 7,73E-01 |
| ZNF823 | 101,33 | 0,03 | 0,08 | 0,46 | 6,43E-01 | 7,73E-01 |
| ISY1 | 474,70 | 0,02 | 0,03 | 0,46 | 6,43E-01 | 7,73E-01 |
| ZNF831 | 1815,86 | -0,03 | 0,07 | -0,46 | 6,43E-01 | 7,73E-01 |
| POLB | 950,00 | -0,02 | 0,05 | -0,46 | 6,43E-01 | 7,73E-01 |
| TMEM220 | 72,71 | -0,05 | 0,11 | -0,46 | 6,43E-01 | 7,74E-01 |
| TMEM176B | 190,37 | 0,12 | 0,25 | 0,46 | 6,43E-01 | 7,74E-01 |
| MAVS | 2164,51 | -0,04 | 0,08 | -0,46 | 6,44E-01 | 7,74E-01 |
| SIGLEC6 | 49,29 | -0,06 | 0,13 | -0,46 | 6,44E-01 | 7,74E-01 |
| CHMP3 | 2085,09 | -0,02 | 0,04 | -0,46 | 6,44E-01 | 7,74E-01 |
| MACIR | 210,06 | -0,03 | 0,06 | -0,46 | 6,44E-01 | 7,74E-01 |
| SLC44A5 | 59,78 | -0,15 | 0,32 | -0,46 | 6,44E-01 | 7,74E-01 |
| RXYLT1 | 287,10 | 0,02 | 0,05 | 0,46 | 6,44E-01 | 7,74E-01 |
| SWI5 | 95,26 | 0,02 | 0,04 | 0,46 | 6,44E-01 | 7,74E-01 |
| TMEM86A | 90,66 | -0,04 | 0,08 | -0,46 | 6,45E-01 | 7,75E-01 |
| XPNPEP3 | 492,49 | -0,02 | 0,05 | -0,46 | 6,45E-01 | 7,75E-01 |
| RNF14 | 2431,27 | -0,04 | 0,08 | -0,46 | 6,45E-01 | 7,75E-01 |
| ZNF586 | 1665,92 | 0,02 | 0,04 | 0,46 | 6,45E-01 | 7,75E-01 |
| AK3 | 673,98 | 0,02 | 0,05 | 0,46 | 6,45E-01 | 7,75E-01 |
| SLC37A2 | 723,25 | 0,03 | 0,07 | 0,46 | 6,45E-01 | 7,75E-01 |
| CEP43 | 904,61 | -0,02 | 0,05 | -0,46 | 6,45E-01 | 7,75E-01 |
| SDHAF3 | 138,25 | 0,03 | 0,06 | 0,46 | 6,45E-01 | 7,75E-01 |
| CRKL | 3742,69 | -0,02 | 0,04 | -0,46 | 6,45E-01 | 7,75E-01 |
| SH3RF1 | 136,59 | -0,04 | 0,09 | -0,46 | 6,46E-01 | 7,75E-01 |
| APBB1IP | 3806,78 | -0,03 | 0,06 | -0,46 | 6,46E-01 | 7,75E-01 |
| PPP3R1 | 3898,20 | -0,04 | 0,08 | -0,46 | 6,46E-01 | 7,76E-01 |
| CLEC12B | 294,63 | -0,06 | 0,14 | -0,46 | 6,46E-01 | 7,76E-01 |
| JAKMIP1 | 46,09 | -0,06 | 0,13 | -0,46 | 6,47E-01 | 7,76E-01 |
| CDHR3 | 419,89 | -0,04 | 0,08 | -0,46 | 6,47E-01 | 7,76E-01 |
| B3GALNT1 | 47,10 | -0,06 | 0,13 | -0,46 | 6,47E-01 | 7,76E-01 |
| POLR1G | 45,87 | -0,04 | 0,09 | -0,46 | 6,47E-01 | 7,76E-01 |
| CLEC6A | 90,75 | -0,06 | 0,12 | -0,46 | 6,47E-01 | 7,77E-01 |
| AOAH | 7019,78 | 0,03 | 0,06 | 0,46 | 6,47E-01 | 7,77E-01 |
| H1-4 | 5269,26 | 0,03 | 0,07 | 0,46 | 6,47E-01 | 7,77E-01 |
| SMARCAL1 | 476,44 | -0,02 | 0,04 | -0,46 | 6,47E-01 | 7,77E-01 |
| CFDP1 | 464,14 | 0,01 | 0,03 | 0,46 | 6,48E-01 | 7,77E-01 |
| ENSG000002 | 391,08 | 0,01 | 0,03 | 0,46 | 6,48E-01 | 7,77E-01 |
| CETN2 | 231,31 | -0,03 | 0,07 | -0,46 | 6,49E-01 | 7,78E-01 |
| CAMKK1 | 376,68 | -0,04 | 0,10 | -0,46 | 6,49E-01 | 7,78E-01 |
| NBDY | 114,66 | -0,02 | 0,05 | -0,46 | 6,49E-01 | 7,78E-01 |
| DDX19A | 307,50 | -0,02 | 0,04 | -0,46 | 6,49E-01 | 7,78E-01 |
| COLGALT2 | 149,20 | -0,07 | 0,14 | -0,46 | 6,49E-01 | 7,78E-01 |
| MACO1 | 718,94 | -0,02 | 0,03 | -0,46 | 6,49E-01 | 7,78E-01 |
| MEA1 | 408,71 | 0,02 | 0,04 | 0,46 | 6,49E-01 | 7,78E-01 |

| | | | | | | |
|---|---|---|---|---|---|---|
| SPOUT1 | 578,54 | 0,02 | 0,04 | 0,46 | 6,49E-01 | 7,78E-01 |
| ENSG000002 | 107,53 | 0,04 | 0,09 | 0,45 | 6,49E-01 | 7,78E-01 |
| EXTL3 | 2372,45 | -0,03 | 0,07 | -0,45 | 6,50E-01 | 7,78E-01 |
| CRELD1 | 329,98 | -0,02 | 0,05 | -0,45 | 6,50E-01 | 7,78E-01 |
| MATK | 700,79 | 0,04 | 0,09 | 0,45 | 6,50E-01 | 7,79E-01 |
| UBE2Q1 | 1169,24 | 0,01 | 0,03 | 0,45 | 6,50E-01 | 7,79E-01 |
| TRIP4 | 481,58 | -0,01 | 0,03 | -0,45 | 6,50E-01 | 7,79E-01 |
| SEC11A | 1068,82 | 0,01 | 0,03 | 0,45 | 6,50E-01 | 7,79E-01 |
| SLMAP | 2839,86 | 0,02 | 0,04 | 0,45 | 6,51E-01 | 7,79E-01 |
| CTSZ | 4522,36 | 0,03 | 0,06 | 0,45 | 6,51E-01 | 7,79E-01 |
| KDM4A | 1074,74 | 0,02 | 0,04 | 0,45 | 6,51E-01 | 7,79E-01 |
| ENSA | 1391,42 | -0,02 | 0,04 | -0,45 | 6,51E-01 | 7,79E-01 |
| COA7 | 185,84 | -0,03 | 0,06 | -0,45 | 6,51E-01 | 7,79E-01 |
| LTB | 4240,32 | 0,02 | 0,05 | 0,45 | 6,51E-01 | 7,79E-01 |
| SLC7A11 | 126,88 | 0,04 | 0,10 | 0,45 | 6,51E-01 | 7,79E-01 |
| MARS2 | 432,71 | 0,02 | 0,04 | 0,45 | 6,52E-01 | 7,80E-01 |
| VTI1A | 2463,86 | 0,02 | 0,04 | 0,45 | 6,52E-01 | 7,80E-01 |
| RECK | 333,51 | 0,02 | 0,04 | 0,45 | 6,52E-01 | 7,80E-01 |
| LUZP1 | 816,62 | 0,02 | 0,04 | 0,45 | 6,52E-01 | 7,80E-01 |
| TEX264 | 1055,18 | 0,01 | 0,03 | 0,45 | 6,52E-01 | 7,80E-01 |
| OSBPL1A | 379,43 | -0,03 | 0,07 | -0,45 | 6,52E-01 | 7,80E-01 |
| C1orf50 | 290,06 | -0,02 | 0,05 | -0,45 | 6,53E-01 | 7,81E-01 |
| KDM3A | 4220,67 | 0,02 | 0,04 | 0,45 | 6,53E-01 | 7,81E-01 |
| PCBD1 | 125,61 | -0,03 | 0,06 | -0,45 | 6,53E-01 | 7,81E-01 |
| SERTAD2 | 920,04 | -0,02 | 0,05 | -0,45 | 6,54E-01 | 7,81E-01 |
| ZNF718 | 619,82 | 0,03 | 0,07 | 0,45 | 6,54E-01 | 7,82E-01 |
| ZNF324 | 560,66 | -0,02 | 0,04 | -0,45 | 6,54E-01 | 7,82E-01 |
| LRIG1 | 1089,32 | -0,02 | 0,05 | -0,45 | 6,55E-01 | 7,83E-01 |
| GAB3 | 1605,08 | -0,02 | 0,05 | -0,45 | 6,56E-01 | 7,84E-01 |
| TPM3 | 12248,98 | 0,02 | 0,05 | 0,45 | 6,56E-01 | 7,84E-01 |
| AMPD2 | 2721,65 | -0,04 | 0,09 | -0,44 | 6,57E-01 | 7,85E-01 |
| KIAA0513 | 2975,55 | -0,04 | 0,09 | -0,44 | 6,57E-01 | 7,85E-01 |
| CD226 | 4495,98 | 0,03 | 0,07 | 0,44 | 6,57E-01 | 7,85E-01 |
| EIF6 | 787,82 | 0,02 | 0,05 | 0,44 | 6,57E-01 | 7,85E-01 |
| C8orf76 | 184,47 | -0,01 | 0,03 | -0,44 | 6,57E-01 | 7,85E-01 |
| GTSF1 | 55,68 | 0,05 | 0,11 | 0,44 | 6,57E-01 | 7,85E-01 |
| GTF2H5 | 226,04 | 0,03 | 0,07 | 0,44 | 6,58E-01 | 7,85E-01 |
| CCDC88C | 4646,18 | -0,03 | 0,07 | -0,44 | 6,58E-01 | 7,86E-01 |
| STIP1 | 1262,57 | 0,02 | 0,05 | 0,44 | 6,58E-01 | 7,86E-01 |
| ZAP70 | 3809,91 | -0,03 | 0,07 | -0,44 | 6,58E-01 | 7,86E-01 |
| WEE1 | 305,51 | -0,03 | 0,07 | -0,44 | 6,58E-01 | 7,86E-01 |
| CC2D2B | 196,57 | -0,04 | 0,09 | -0,44 | 6,58E-01 | 7,86E-01 |
| DSEL | 161,40 | -0,05 | 0,12 | -0,44 | 6,58E-01 | 7,86E-01 |
| LINC02218 | 285,63 | -0,04 | 0,09 | -0,44 | 6,58E-01 | 7,86E-01 |
| SSRP1 | 708,12 | 0,02 | 0,04 | 0,44 | 6,59E-01 | 7,86E-01 |
| TNFSF10 | 4962,56 | 0,03 | 0,08 | 0,44 | 6,59E-01 | 7,86E-01 |

| | | | | | | |
|---|---|---|---|---|---|---|
| SKA3 | 108,29 | -0,06 | 0,14 | -0,44 | 6,59E-01 | 7,86E-01 |
| PEX19 | 475,20 | -0,01 | 0,03 | -0,44 | 6,59E-01 | 7,86E-01 |
| LILRA5 | 2915,38 | -0,04 | 0,09 | -0,44 | 6,59E-01 | 7,86E-01 |
| PTPRE | 8611,55 | -0,02 | 0,05 | -0,44 | 6,59E-01 | 7,86E-01 |
| ACP1 | 1817,10 | -0,03 | 0,06 | -0,44 | 6,59E-01 | 7,86E-01 |
| RNF216 | 2166,38 | -0,02 | 0,04 | -0,44 | 6,59E-01 | 7,86E-01 |
| RC3H1 | 8348,52 | -0,02 | 0,04 | -0,44 | 6,60E-01 | 7,87E-01 |
| RPN1 | 4185,32 | 0,02 | 0,04 | 0,44 | 6,60E-01 | 7,87E-01 |
| NDRG3 | 676,47 | -0,02 | 0,04 | -0,44 | 6,60E-01 | 7,87E-01 |
| SF3B2 | 1370,93 | -0,03 | 0,06 | -0,44 | 6,61E-01 | 7,88E-01 |
| TBCC | 1528,92 | 0,02 | 0,05 | 0,44 | 6,61E-01 | 7,88E-01 |
| NCALD | 723,97 | 0,04 | 0,08 | 0,44 | 6,61E-01 | 7,88E-01 |
| IL16 | 12898,04 | -0,02 | 0,05 | -0,44 | 6,61E-01 | 7,88E-01 |
| FIRRM | 454,00 | 0,02 | 0,04 | 0,44 | 6,61E-01 | 7,88E-01 |
| CLIP1 | 2682,33 | -0,02 | 0,04 | -0,44 | 6,61E-01 | 7,88E-01 |
| HCLS1 | 17353,32 | -0,03 | 0,06 | -0,44 | 6,61E-01 | 7,88E-01 |
| SGO2 | 112,23 | 0,05 | 0,12 | 0,44 | 6,62E-01 | 7,88E-01 |
| ARK2N | 3234,51 | 0,02 | 0,04 | 0,44 | 6,62E-01 | 7,89E-01 |
| SMAD3 | 1784,32 | -0,03 | 0,07 | -0,44 | 6,62E-01 | 7,89E-01 |
| PARS2 | 52,98 | -0,03 | 0,06 | -0,44 | 6,62E-01 | 7,89E-01 |
| UBE2B | 2458,59 | -0,03 | 0,06 | -0,44 | 6,63E-01 | 7,89E-01 |
| ZNF410 | 389,73 | -0,01 | 0,03 | -0,44 | 6,63E-01 | 7,89E-01 |
| NELL2 | 1896,84 | 0,04 | 0,09 | 0,44 | 6,63E-01 | 7,89E-01 |
| UBXN2A | 475,98 | 0,02 | 0,04 | 0,44 | 6,63E-01 | 7,89E-01 |
| GON4L | 2062,07 | 0,02 | 0,05 | 0,44 | 6,63E-01 | 7,89E-01 |
| PGM1 | 752,62 | 0,02 | 0,04 | 0,44 | 6,63E-01 | 7,89E-01 |
| BLVRA | 451,18 | 0,03 | 0,07 | 0,44 | 6,63E-01 | 7,89E-01 |
| TRAPPC10 | 3031,85 | -0,02 | 0,04 | -0,44 | 6,63E-01 | 7,89E-01 |
| TBRG4 | 1052,85 | 0,02 | 0,04 | 0,44 | 6,64E-01 | 7,90E-01 |
| LIG3 | 895,57 | -0,02 | 0,04 | -0,43 | 6,64E-01 | 7,90E-01 |
| FKBP2 | 457,95 | 0,02 | 0,04 | 0,43 | 6,64E-01 | 7,90E-01 |
| LIPT2 | 82,97 | 0,03 | 0,07 | 0,43 | 6,64E-01 | 7,90E-01 |
| MX1 | 3054,26 | 0,07 | 0,16 | 0,43 | 6,64E-01 | 7,90E-01 |
| SDHA | 2107,22 | -0,01 | 0,03 | -0,43 | 6,64E-01 | 7,90E-01 |
| H2BK1 | 22,32 | -0,04 | 0,08 | -0,43 | 6,64E-01 | 7,90E-01 |
| ARL10 | 527,63 | 0,03 | 0,07 | 0,43 | 6,64E-01 | 7,90E-01 |
| RRP9 | 215,66 | -0,02 | 0,05 | -0,43 | 6,64E-01 | 7,90E-01 |
| DUS2 | 685,36 | 0,02 | 0,05 | 0,43 | 6,65E-01 | 7,90E-01 |
| VCPKMT | 950,97 | 0,02 | 0,05 | 0,43 | 6,65E-01 | 7,90E-01 |
| LSM14B | 876,41 | -0,02 | 0,05 | -0,43 | 6,65E-01 | 7,90E-01 |
| KDM2B | 800,35 | -0,03 | 0,07 | -0,43 | 6,65E-01 | 7,90E-01 |
| PBXIP1 | 4623,36 | 0,03 | 0,06 | 0,43 | 6,65E-01 | 7,90E-01 |
| ARHGAP30 | 13504,27 | 0,02 | 0,05 | 0,43 | 6,65E-01 | 7,91E-01 |
| CD33 | 496,57 | 0,03 | 0,07 | 0,43 | 6,65E-01 | 7,91E-01 |
| MRPS15 | 300,57 | 0,02 | 0,04 | 0,43 | 6,65E-01 | 7,91E-01 |
| ICAM2 | 1065,77 | -0,02 | 0,04 | -0,43 | 6,65E-01 | 7,91E-01 |

| | | | | | | |
|---|---|---|---|---|---|---|
| MOGS | 530,02 | -0,03 | 0,07 | -0,43 | 6,65E-01 | 7,91E-01 |
| HORMAD1 | 240,90 | 0,04 | 0,10 | 0,43 | 6,66E-01 | 7,91E-01 |
| GOLGA8R | 54,94 | 0,05 | 0,11 | 0,43 | 6,66E-01 | 7,91E-01 |
| EIF2AK2 | 2810,66 | -0,04 | 0,10 | -0,43 | 6,66E-01 | 7,91E-01 |
| TAMALIN | 102,86 | -0,04 | 0,10 | -0,43 | 6,66E-01 | 7,91E-01 |
| WASHC1 | 151,03 | -0,05 | 0,12 | -0,43 | 6,66E-01 | 7,91E-01 |
| ANGPT1 | 320,66 | 0,05 | 0,12 | 0,43 | 6,66E-01 | 7,91E-01 |
| KLHL22 | 975,41 | -0,02 | 0,05 | -0,43 | 6,66E-01 | 7,91E-01 |
| SNX17 | 2199,48 | -0,01 | 0,03 | -0,43 | 6,66E-01 | 7,91E-01 |
| SNRNP70 | 1770,83 | -0,04 | 0,09 | -0,43 | 6,67E-01 | 7,91E-01 |
| SUGP2 | 2073,28 | -0,02 | 0,05 | -0,43 | 6,67E-01 | 7,91E-01 |
| CLEC4A | 288,08 | 0,04 | 0,09 | 0,43 | 6,67E-01 | 7,91E-01 |
| GPR155 | 2976,23 | 0,02 | 0,05 | 0,43 | 6,67E-01 | 7,91E-01 |
| BTD | 497,14 | -0,02 | 0,04 | -0,43 | 6,67E-01 | 7,92E-01 |
| PRPF6 | 1712,98 | 0,02 | 0,05 | 0,43 | 6,67E-01 | 7,92E-01 |
| ARPIN | 277,29 | 0,06 | 0,14 | 0,43 | 6,68E-01 | 7,92E-01 |
| SLC7A6 | 2053,16 | -0,03 | 0,06 | -0,43 | 6,68E-01 | 7,92E-01 |
| CAVIN3 | 27,59 | 0,08 | 0,19 | 0,43 | 6,68E-01 | 7,92E-01 |
| AGO1 | 2524,23 | -0,03 | 0,06 | -0,43 | 6,68E-01 | 7,92E-01 |
| NFIL3 | 665,77 | 0,03 | 0,07 | 0,43 | 6,68E-01 | 7,92E-01 |
| KIR2DL1 | 21,69 | -0,09 | 0,20 | -0,43 | 6,68E-01 | 7,92E-01 |
| LAMC1 | 183,80 | -0,05 | 0,11 | -0,43 | 6,68E-01 | 7,92E-01 |
| POLR3A | 813,68 | -0,02 | 0,04 | -0,43 | 6,68E-01 | 7,92E-01 |
| CMSS1 | 268,06 | 0,03 | 0,06 | 0,43 | 6,69E-01 | 7,93E-01 |
| MCL1 | 53749,40 | -0,02 | 0,06 | -0,43 | 6,69E-01 | 7,93E-01 |
| ASRGL1 | 257,64 | -0,03 | 0,07 | -0,43 | 6,69E-01 | 7,93E-01 |
| SYAP1 | 1196,70 | 0,02 | 0,04 | 0,43 | 6,69E-01 | 7,93E-01 |
| CCR1 | 1491,61 | -0,04 | 0,09 | -0,43 | 6,70E-01 | 7,94E-01 |
| SMG1 | 11746,64 | -0,02 | 0,05 | -0,43 | 6,70E-01 | 7,94E-01 |
| IL27RA | 1025,25 | 0,02 | 0,04 | 0,43 | 6,70E-01 | 7,94E-01 |
| PCED1B | 612,88 | 0,03 | 0,07 | 0,43 | 6,70E-01 | 7,94E-01 |
| C1orf21 | 738,63 | 0,04 | 0,10 | 0,43 | 6,71E-01 | 7,94E-01 |
| RPIA | 2133,56 | -0,05 | 0,11 | -0,43 | 6,71E-01 | 7,94E-01 |
| TMEM67 | 177,51 | -0,03 | 0,07 | -0,43 | 6,71E-01 | 7,94E-01 |
| HARS1 | 981,60 | -0,01 | 0,03 | -0,43 | 6,71E-01 | 7,94E-01 |
| TSTD1 | 562,06 | 0,02 | 0,05 | 0,42 | 6,71E-01 | 7,94E-01 |
| GAPDH | 10575,96 | -0,02 | 0,05 | -0,42 | 6,72E-01 | 7,95E-01 |
| SYNRG | 4474,34 | -0,02 | 0,04 | -0,42 | 6,72E-01 | 7,95E-01 |
| OGT | 19189,51 | 0,02 | 0,04 | 0,42 | 6,72E-01 | 7,95E-01 |
| RNF144B | 4050,59 | 0,03 | 0,07 | 0,42 | 6,72E-01 | 7,95E-01 |
| CASP5 | 399,42 | -0,05 | 0,11 | -0,42 | 6,72E-01 | 7,95E-01 |
| FAAH2 | 247,56 | 0,04 | 0,09 | 0,42 | 6,72E-01 | 7,95E-01 |
| CCL28 | 174,01 | 0,02 | 0,06 | 0,42 | 6,72E-01 | 7,95E-01 |
| PARP14 | 14464,74 | 0,04 | 0,09 | 0,42 | 6,73E-01 | 7,96E-01 |
| DPH7 | 533,60 | -0,02 | 0,04 | -0,42 | 6,73E-01 | 7,96E-01 |
| SH3TC1 | 1480,71 | 0,03 | 0,06 | 0,42 | 6,73E-01 | 7,96E-01 |

| | | | | | | |
|---|---|---|---|---|---|---|
| PDSS2 | 225,21 | 0,01 | 0,03 | 0,42 | 6,73E-01 | 7,96E-01 |
| LMNB1 | 1698,42 | 0,03 | 0,08 | 0,42 | 6,73E-01 | 7,96E-01 |
| PTBP3 | 9685,53 | 0,02 | 0,05 | 0,42 | 6,73E-01 | 7,96E-01 |
| NSDHL | 130,76 | 0,02 | 0,04 | 0,42 | 6,73E-01 | 7,96E-01 |
| BMX | 203,29 | -0,05 | 0,12 | -0,42 | 6,73E-01 | 7,96E-01 |
| AAK1 | 5351,39 | -0,03 | 0,06 | -0,42 | 6,74E-01 | 7,96E-01 |
| MID2 | 150,01 | -0,03 | 0,06 | -0,42 | 6,74E-01 | 7,96E-01 |
| ADRB2 | 1299,93 | 0,03 | 0,07 | 0,42 | 6,74E-01 | 7,96E-01 |
| CCPG1 | 2257,81 | 0,03 | 0,06 | 0,42 | 6,74E-01 | 7,96E-01 |
| POLR3D | 409,88 | -0,02 | 0,05 | -0,42 | 6,74E-01 | 7,96E-01 |
| VPS35L | 765,46 | -0,02 | 0,04 | -0,42 | 6,74E-01 | 7,96E-01 |
| MSR1 | 176,23 | 0,06 | 0,15 | 0,42 | 6,74E-01 | 7,96E-01 |
| PITPNB | 828,44 | 0,02 | 0,04 | 0,42 | 6,74E-01 | 7,96E-01 |
| HDGFL3 | 256,37 | 0,05 | 0,11 | 0,42 | 6,74E-01 | 7,97E-01 |
| CORO1A | 11966,41 | -0,03 | 0,06 | -0,42 | 6,75E-01 | 7,97E-01 |
| RBM14-RBM | 260,74 | 0,02 | 0,05 | 0,42 | 6,75E-01 | 7,97E-01 |
| ADAM20 | 112,13 | 0,04 | 0,09 | 0,42 | 6,75E-01 | 7,97E-01 |
| PPP1R21 | 2109,23 | -0,02 | 0,04 | -0,42 | 6,75E-01 | 7,97E-01 |
| GOLGA2 | 992,96 | -0,02 | 0,06 | -0,42 | 6,75E-01 | 7,97E-01 |
| ECHDC2 | 888,23 | -0,03 | 0,08 | -0,42 | 6,75E-01 | 7,97E-01 |
| GPR141 | 932,19 | -0,04 | 0,10 | -0,42 | 6,76E-01 | 7,97E-01 |
| SYNGR2 | 1217,00 | -0,02 | 0,05 | -0,42 | 6,76E-01 | 7,98E-01 |
| TNFSF13 | 477,69 | -0,03 | 0,07 | -0,42 | 6,76E-01 | 7,98E-01 |
| RPGRIP1 | 276,43 | -0,03 | 0,08 | -0,42 | 6,76E-01 | 7,98E-01 |
| STMN3 | 663,23 | -0,03 | 0,06 | -0,42 | 6,76E-01 | 7,98E-01 |
| MPLKIP | 628,54 | 0,02 | 0,04 | 0,42 | 6,76E-01 | 7,98E-01 |
| MMS19 | 1106,11 | -0,02 | 0,04 | -0,42 | 6,76E-01 | 7,98E-01 |
| BBC3 | 475,28 | -0,03 | 0,07 | -0,42 | 6,76E-01 | 7,98E-01 |
| ARL6IP1 | 2935,20 | 0,03 | 0,08 | 0,42 | 6,76E-01 | 7,98E-01 |
| SYNM | 291,17 | 0,06 | 0,16 | 0,42 | 6,76E-01 | 7,98E-01 |
| POLM | 609,38 | -0,03 | 0,07 | -0,42 | 6,77E-01 | 7,98E-01 |
| TLR6 | 5052,25 | -0,03 | 0,07 | -0,42 | 6,77E-01 | 7,98E-01 |
| RIPK2 | 599,80 | 0,02 | 0,06 | 0,42 | 6,77E-01 | 7,98E-01 |
| CFAP298 | 253,15 | 0,02 | 0,04 | 0,42 | 6,77E-01 | 7,98E-01 |
| FAIM | 87,30 | -0,03 | 0,08 | -0,42 | 6,77E-01 | 7,98E-01 |
| CMKLR1 | 523,56 | -0,04 | 0,09 | -0,42 | 6,77E-01 | 7,98E-01 |
| SDR42E1 | 346,10 | 0,03 | 0,07 | 0,42 | 6,77E-01 | 7,98E-01 |
| RSPH3 | 545,62 | -0,02 | 0,05 | -0,42 | 6,77E-01 | 7,98E-01 |
| SLC8A1 | 3838,94 | -0,03 | 0,07 | -0,42 | 6,77E-01 | 7,98E-01 |
| PLAAT3 | 151,95 | 0,04 | 0,09 | 0,42 | 6,77E-01 | 7,98E-01 |
| ARHGAP4 | 3252,66 | -0,03 | 0,06 | -0,42 | 6,77E-01 | 7,98E-01 |
| SOCS7 | 653,37 | -0,02 | 0,04 | -0,42 | 6,77E-01 | 7,98E-01 |
| MYO5A | 4929,66 | 0,02 | 0,04 | 0,42 | 6,78E-01 | 7,98E-01 |
| UPB1 | 99,21 | -0,05 | 0,12 | -0,42 | 6,78E-01 | 7,98E-01 |
| PCDHGB6 | 67,08 | -0,05 | 0,13 | -0,42 | 6,78E-01 | 7,98E-01 |
| BRF1 | 958,90 | -0,03 | 0,06 | -0,42 | 6,78E-01 | 7,98E-01 |

| | | | | | | |
|---|---|---|---|---|---|---|
| FAM174A | 753,38 | 0,03 | 0,07 | 0,41 | 6,78E-01 | 7,99E-01 |
| GTF2F1 | 883,73 | 0,02 | 0,04 | 0,41 | 6,78E-01 | 7,99E-01 |
| FZD3 | 245,08 | 0,03 | 0,08 | 0,41 | 6,78E-01 | 7,99E-01 |
| POGLUT3 | 218,28 | -0,03 | 0,06 | -0,41 | 6,79E-01 | 7,99E-01 |
| PRICKLE1 | 186,43 | -0,04 | 0,10 | -0,41 | 6,79E-01 | 7,99E-01 |
| CNOT1 | 7217,46 | -0,02 | 0,04 | -0,41 | 6,80E-01 | 8,00E-01 |
| CLIC2 | 255,51 | -0,05 | 0,11 | -0,41 | 6,80E-01 | 8,00E-01 |
| ERLIN1 | 770,84 | -0,02 | 0,05 | -0,41 | 6,80E-01 | 8,00E-01 |
| ZFYVE26 | 1868,84 | -0,02 | 0,05 | -0,41 | 6,81E-01 | 8,01E-01 |
| C5orf34 | 75,97 | 0,03 | 0,08 | 0,41 | 6,81E-01 | 8,01E-01 |
| P2RX7 | 678,26 | -0,02 | 0,06 | -0,41 | 6,81E-01 | 8,01E-01 |
| RSAD2 | 1848,60 | 0,09 | 0,23 | 0,41 | 6,81E-01 | 8,01E-01 |
| NSUN3 | 1926,21 | 0,03 | 0,07 | 0,41 | 6,81E-01 | 8,01E-01 |
| TMEM97 | 67,12 | -0,03 | 0,08 | -0,41 | 6,81E-01 | 8,01E-01 |
| CPEB3 | 395,36 | 0,02 | 0,04 | 0,41 | 6,82E-01 | 8,01E-01 |
| MTHFD1 | 586,17 | 0,02 | 0,04 | 0,41 | 6,82E-01 | 8,01E-01 |
| TRIM34 | 40,07 | 0,03 | 0,07 | 0,41 | 6,82E-01 | 8,02E-01 |
| RABL2A | 225,37 | -0,03 | 0,08 | -0,41 | 6,82E-01 | 8,02E-01 |
| BNIP3 | 411,25 | 0,02 | 0,05 | 0,41 | 6,83E-01 | 8,02E-01 |
| B3GAT3 | 335,95 | -0,02 | 0,05 | -0,41 | 6,83E-01 | 8,02E-01 |
| KPNA1 | 2641,10 | 0,01 | 0,04 | 0,41 | 6,83E-01 | 8,03E-01 |
| MAGEE1 | 79,14 | 0,03 | 0,07 | 0,41 | 6,83E-01 | 8,03E-01 |
| TFRC | 2907,85 | -0,04 | 0,09 | -0,41 | 6,83E-01 | 8,03E-01 |
| KCNE3 | 4909,37 | -0,03 | 0,07 | -0,41 | 6,83E-01 | 8,03E-01 |
| TRIM24 | 1232,17 | -0,01 | 0,03 | -0,41 | 6,84E-01 | 8,03E-01 |
| SLC9B1 | 94,99 | -0,03 | 0,07 | -0,41 | 6,84E-01 | 8,04E-01 |
| SLX4IP | 486,89 | -0,02 | 0,04 | -0,41 | 6,84E-01 | 8,04E-01 |
| CAMSAP1 | 831,89 | -0,01 | 0,03 | -0,41 | 6,84E-01 | 8,04E-01 |
| BATF | 183,46 | -0,02 | 0,06 | -0,41 | 6,84E-01 | 8,04E-01 |
| TRIM38 | 5285,18 | 0,02 | 0,04 | 0,41 | 6,85E-01 | 8,04E-01 |
| ARMCX2 | 84,31 | 0,03 | 0,08 | 0,41 | 6,85E-01 | 8,04E-01 |
| IER2 | 1854,30 | -0,02 | 0,05 | -0,41 | 6,85E-01 | 8,04E-01 |
| ENSG000002 | 83,27 | -0,04 | 0,09 | -0,41 | 6,85E-01 | 8,04E-01 |
| EWSR1 | 6376,65 | 0,01 | 0,03 | 0,41 | 6,85E-01 | 8,04E-01 |
| NF2 | 818,24 | -0,02 | 0,05 | -0,41 | 6,85E-01 | 8,04E-01 |
| SLC25A25 | 555,39 | -0,02 | 0,04 | -0,41 | 6,85E-01 | 8,04E-01 |
| RASGRP2 | 5431,42 | -0,02 | 0,05 | -0,40 | 6,86E-01 | 8,05E-01 |
| ASCC1 | 242,19 | -0,02 | 0,04 | -0,40 | 6,86E-01 | 8,05E-01 |
| BRMS1L | 105,46 | 0,03 | 0,08 | 0,40 | 6,86E-01 | 8,05E-01 |
| CR2 | 163,76 | 0,05 | 0,12 | 0,40 | 6,86E-01 | 8,05E-01 |
| ADAM17 | 2798,42 | 0,02 | 0,04 | 0,40 | 6,86E-01 | 8,05E-01 |
| ZC3H12B | 62,31 | -0,04 | 0,10 | -0,40 | 6,86E-01 | 8,05E-01 |
| CORO1B | 941,61 | -0,02 | 0,04 | -0,40 | 6,86E-01 | 8,05E-01 |
| H6PD | 1048,83 | -0,04 | 0,10 | -0,40 | 6,86E-01 | 8,05E-01 |
| LAMP1 | 2441,08 | 0,02 | 0,06 | 0,40 | 6,86E-01 | 8,05E-01 |
| CASP1 | 4482,69 | 0,03 | 0,07 | 0,40 | 6,86E-01 | 8,05E-01 |

| | | | | | | |
|---|---|---|---|---|---|---|
| KLF2 | 3311,17 | -0,02 | 0,06 | -0,40 | 6,86E-01 | 8,05E-01 |
| GSTM3 | 91,36 | 0,08 | 0,19 | 0,40 | 6,87E-01 | 8,05E-01 |
| PLAAT4 | 768,80 | 0,02 | 0,06 | 0,40 | 6,87E-01 | 8,05E-01 |
| NAIP | 1415,07 | -0,04 | 0,10 | -0,40 | 6,87E-01 | 8,06E-01 |
| ARL2 | 158,39 | 0,02 | 0,04 | 0,40 | 6,87E-01 | 8,06E-01 |
| RNASEL | 2663,68 | 0,02 | 0,05 | 0,40 | 6,87E-01 | 8,06E-01 |
| THEM4 | 470,33 | -0,03 | 0,07 | -0,40 | 6,88E-01 | 8,06E-01 |
| SARS2 | 58,16 | -0,03 | 0,07 | -0,40 | 6,88E-01 | 8,06E-01 |
| JADE2 | 2371,70 | -0,03 | 0,06 | -0,40 | 6,88E-01 | 8,06E-01 |
| ZFP36L1 | 2577,60 | 0,03 | 0,06 | 0,40 | 6,88E-01 | 8,06E-01 |
| EMP1 | 85,56 | -0,06 | 0,15 | -0,40 | 6,89E-01 | 8,07E-01 |
| ZSCAN2 | 273,68 | -0,01 | 0,04 | -0,40 | 6,89E-01 | 8,07E-01 |
| ZC3H12D | 836,22 | 0,03 | 0,07 | 0,40 | 6,89E-01 | 8,07E-01 |
| MTMR14 | 2718,50 | -0,02 | 0,04 | -0,40 | 6,89E-01 | 8,07E-01 |
| ZFYVE27 | 1856,54 | -0,01 | 0,03 | -0,40 | 6,89E-01 | 8,07E-01 |
| NKAPL | 43,29 | -0,04 | 0,10 | -0,40 | 6,89E-01 | 8,07E-01 |
| TVP23C | 494,31 | 0,02 | 0,06 | 0,40 | 6,89E-01 | 8,07E-01 |
| DHFR | 426,79 | -0,02 | 0,04 | -0,40 | 6,89E-01 | 8,07E-01 |
| SPRYD7 | 145,07 | 0,02 | 0,04 | 0,40 | 6,90E-01 | 8,07E-01 |
| KLF13 | 6228,42 | -0,02 | 0,06 | -0,40 | 6,90E-01 | 8,07E-01 |
| ZNF433 | 35,00 | -0,05 | 0,12 | -0,40 | 6,90E-01 | 8,07E-01 |
| RHOC | 444,35 | 0,02 | 0,06 | 0,40 | 6,90E-01 | 8,07E-01 |
| APRT | 496,54 | -0,02 | 0,05 | -0,40 | 6,90E-01 | 8,08E-01 |
| DHPS | 139,11 | -0,03 | 0,07 | -0,40 | 6,91E-01 | 8,08E-01 |
| KRT73 | 169,98 | 0,07 | 0,18 | 0,40 | 6,91E-01 | 8,08E-01 |
| SNX29 | 2146,69 | 0,02 | 0,05 | 0,40 | 6,91E-01 | 8,08E-01 |
| LONP1 | 855,24 | -0,02 | 0,04 | -0,40 | 6,91E-01 | 8,08E-01 |
| CYP4F3 | 5502,62 | 0,04 | 0,09 | 0,40 | 6,91E-01 | 8,08E-01 |
| CCDC34 | 48,41 | -0,05 | 0,12 | -0,40 | 6,91E-01 | 8,08E-01 |
| TUT1 | 94,08 | -0,03 | 0,08 | -0,40 | 6,91E-01 | 8,08E-01 |
| CD3G | 1603,36 | 0,03 | 0,06 | 0,40 | 6,91E-01 | 8,08E-01 |
| PLAGL1 | 755,21 | -0,03 | 0,06 | -0,40 | 6,92E-01 | 8,08E-01 |
| SMN2 | 100,85 | 0,05 | 0,12 | 0,40 | 6,92E-01 | 8,08E-01 |
| TIMM44 | 304,33 | 0,02 | 0,05 | 0,40 | 6,92E-01 | 8,08E-01 |
| USP39 | 1004,46 | -0,01 | 0,02 | -0,40 | 6,92E-01 | 8,08E-01 |
| ARHGAP10 | 481,97 | -0,02 | 0,06 | -0,40 | 6,92E-01 | 8,08E-01 |
| CRACR2A | 1298,18 | -0,02 | 0,06 | -0,40 | 6,92E-01 | 8,08E-01 |
| TLR1 | 10291,51 | 0,03 | 0,08 | 0,40 | 6,92E-01 | 8,08E-01 |
| COQ8A | 1288,39 | 0,02 | 0,06 | 0,40 | 6,92E-01 | 8,08E-01 |
| KIF13B | 1543,86 | -0,02 | 0,05 | -0,40 | 6,92E-01 | 8,08E-01 |
| MECR | 164,38 | -0,02 | 0,06 | -0,40 | 6,92E-01 | 8,08E-01 |
| PDGFC | 185,47 | 0,03 | 0,08 | 0,40 | 6,92E-01 | 8,08E-01 |
| TNPO3 | 2806,99 | 0,02 | 0,04 | 0,40 | 6,92E-01 | 8,08E-01 |
| DMAC2L | 1190,99 | 0,02 | 0,04 | 0,40 | 6,93E-01 | 8,09E-01 |
| NRF1 | 987,99 | -0,01 | 0,03 | -0,40 | 6,93E-01 | 8,09E-01 |
| RIDA | 82,23 | 0,03 | 0,08 | 0,40 | 6,93E-01 | 8,09E-01 |

| | | | | | | |
|---|---|---|---|---|---|---|
| SLC15A3 | 1270,90 | 0,03 | 0,06 | 0,39 | 6,93E-01 | 8,09E-01 |
| FGR | 9526,48 | -0,02 | 0,06 | -0,39 | 6,94E-01 | 8,10E-01 |
| DDAH2 | 665,18 | -0,02 | 0,05 | -0,39 | 6,94E-01 | 8,10E-01 |
| SNF8 | 846,86 | -0,01 | 0,03 | -0,39 | 6,94E-01 | 8,10E-01 |
| AP1S1 | 178,61 | -0,02 | 0,06 | -0,39 | 6,94E-01 | 8,10E-01 |
| GPR27 | 505,78 | -0,03 | 0,08 | -0,39 | 6,94E-01 | 8,10E-01 |
| TRIM35 | 848,19 | 0,01 | 0,04 | 0,39 | 6,94E-01 | 8,10E-01 |
| UBE2E3 | 479,79 | -0,02 | 0,06 | -0,39 | 6,94E-01 | 8,10E-01 |
| HSDL1 | 617,31 | 0,02 | 0,04 | 0,39 | 6,95E-01 | 8,10E-01 |
| LPAR5 | 199,61 | 0,03 | 0,07 | 0,39 | 6,95E-01 | 8,10E-01 |
| CXCR6 | 293,00 | 0,05 | 0,12 | 0,39 | 6,95E-01 | 8,10E-01 |
| EIPR1 | 660,75 | -0,01 | 0,04 | -0,39 | 6,95E-01 | 8,10E-01 |
| SLAIN2 | 1694,34 | 0,01 | 0,04 | 0,39 | 6,95E-01 | 8,11E-01 |
| MTMR8 | 223,64 | 0,02 | 0,05 | 0,39 | 6,95E-01 | 8,11E-01 |
| ZNG1C | 560,93 | 0,02 | 0,05 | 0,39 | 6,96E-01 | 8,11E-01 |
| CRIM1 | 302,87 | -0,03 | 0,07 | -0,39 | 6,96E-01 | 8,11E-01 |
| SORCS3 | 139,83 | -0,06 | 0,16 | -0,39 | 6,96E-01 | 8,11E-01 |
| PTGS2 | 1249,29 | 0,04 | 0,09 | 0,39 | 6,96E-01 | 8,11E-01 |
| JPT2 | 513,95 | 0,01 | 0,03 | 0,39 | 6,96E-01 | 8,11E-01 |
| VASH1 | 333,88 | -0,04 | 0,11 | -0,39 | 6,97E-01 | 8,12E-01 |
| STIMATE-MUS | 31,48 | -0,04 | 0,11 | -0,39 | 6,97E-01 | 8,12E-01 |
| CARD11 | 1108,64 | -0,03 | 0,09 | -0,39 | 6,97E-01 | 8,12E-01 |
| DDX41 | 899,82 | 0,02 | 0,04 | 0,39 | 6,97E-01 | 8,12E-01 |
| TMEM147 | 274,93 | 0,02 | 0,04 | 0,39 | 6,97E-01 | 8,12E-01 |
| HIF1AN | 2670,28 | -0,02 | 0,05 | -0,39 | 6,98E-01 | 8,12E-01 |
| NMT1 | 1446,06 | -0,01 | 0,03 | -0,39 | 6,98E-01 | 8,12E-01 |
| IRF4 | 639,69 | 0,03 | 0,06 | 0,39 | 6,98E-01 | 8,13E-01 |
| PSMB2 | 603,81 | -0,01 | 0,03 | -0,39 | 6,98E-01 | 8,13E-01 |
| IFI6 | 2166,01 | 0,07 | 0,17 | 0,39 | 6,99E-01 | 8,13E-01 |
| NOP14 | 763,09 | 0,02 | 0,04 | 0,39 | 6,99E-01 | 8,14E-01 |
| PLEK | 4122,01 | -0,02 | 0,06 | -0,39 | 6,99E-01 | 8,14E-01 |
| THTPA | 155,31 | -0,02 | 0,05 | -0,39 | 6,99E-01 | 8,14E-01 |
| PLEC | 9566,13 | -0,03 | 0,09 | -0,39 | 7,00E-01 | 8,14E-01 |
| TRAPPC5 | 550,58 | -0,02 | 0,05 | -0,39 | 7,00E-01 | 8,14E-01 |
| NCBP3 | 1931,57 | -0,02 | 0,04 | -0,39 | 7,00E-01 | 8,14E-01 |
| PYGL | 7330,44 | -0,03 | 0,08 | -0,39 | 7,00E-01 | 8,14E-01 |
| AGPAT4 | 848,41 | -0,02 | 0,06 | -0,39 | 7,00E-01 | 8,14E-01 |
| FGFBP2 | 759,55 | 0,05 | 0,12 | 0,39 | 7,00E-01 | 8,14E-01 |
| GPBP1L1 | 3252,89 | -0,01 | 0,03 | -0,38 | 7,00E-01 | 8,15E-01 |
| OCRL | 239,26 | -0,02 | 0,05 | -0,38 | 7,01E-01 | 8,15E-01 |
| TCHP | 1111,84 | -0,01 | 0,03 | -0,38 | 7,01E-01 | 8,15E-01 |
| TFIP11 | 1655,48 | 0,02 | 0,05 | 0,38 | 7,01E-01 | 8,15E-01 |
| TSSK4 | 24,06 | -0,04 | 0,11 | -0,38 | 7,01E-01 | 8,15E-01 |
| HDC | 276,40 | -0,08 | 0,21 | -0,38 | 7,01E-01 | 8,15E-01 |
| ATF4 | 2489,89 | -0,02 | 0,04 | -0,38 | 7,01E-01 | 8,15E-01 |
| KIF22 | 652,33 | 0,02 | 0,04 | 0,38 | 7,01E-01 | 8,15E-01 |

| | | | | | | |
|---|---|---|---|---|---|---|
| GOLGA8O | 32,78 | 0,06 | 0,15 | 0,38 | 7,02E-01 | 8,15E-01 |
| THAP11 | 552,63 | -0,01 | 0,04 | -0,38 | 7,02E-01 | 8,16E-01 |
| CPQ | 3606,62 | -0,03 | 0,07 | -0,38 | 7,02E-01 | 8,16E-01 |
| TCP11L1 | 547,19 | -0,02 | 0,05 | -0,38 | 7,02E-01 | 8,16E-01 |
| RIT1 | 2565,34 | 0,03 | 0,07 | 0,38 | 7,02E-01 | 8,16E-01 |
| FBP1 | 233,95 | -0,03 | 0,07 | -0,38 | 7,02E-01 | 8,16E-01 |
| CD14 | 4218,28 | 0,03 | 0,08 | 0,38 | 7,03E-01 | 8,16E-01 |
| CCDC121 | 71,92 | 0,03 | 0,09 | 0,38 | 7,03E-01 | 8,16E-01 |
| AFMID | 104,41 | -0,03 | 0,07 | -0,38 | 7,03E-01 | 8,16E-01 |
| FAM168B | 1953,91 | 0,01 | 0,04 | 0,38 | 7,04E-01 | 8,17E-01 |
| ZDHHC16 | 491,44 | -0,02 | 0,04 | -0,38 | 7,04E-01 | 8,17E-01 |
| CREB3L4 | 104,83 | 0,02 | 0,05 | 0,38 | 7,04E-01 | 8,17E-01 |
| C8orf82 | 214,15 | -0,02 | 0,06 | -0,38 | 7,04E-01 | 8,18E-01 |
| DYNC1H1 | 7178,46 | -0,02 | 0,06 | -0,38 | 7,04E-01 | 8,18E-01 |
| NEK9 | 2672,95 | 0,01 | 0,03 | 0,38 | 7,04E-01 | 8,18E-01 |
| STAT6 | 7578,13 | -0,03 | 0,07 | -0,38 | 7,05E-01 | 8,18E-01 |
| CASP10 | 2134,96 | 0,02 | 0,04 | 0,38 | 7,05E-01 | 8,18E-01 |
| ENTPD6 | 635,80 | -0,02 | 0,06 | -0,38 | 7,05E-01 | 8,18E-01 |
| CDK8 | 429,36 | 0,02 | 0,05 | 0,38 | 7,05E-01 | 8,18E-01 |
| PLBD1 | 3967,75 | -0,03 | 0,08 | -0,38 | 7,05E-01 | 8,18E-01 |
| PROCR | 33,63 | -0,04 | 0,11 | -0,38 | 7,05E-01 | 8,18E-01 |
| DDX49 | 559,96 | -0,01 | 0,04 | -0,38 | 7,05E-01 | 8,18E-01 |
| PTTG2 | 157,79 | -0,04 | 0,09 | -0,38 | 7,06E-01 | 8,19E-01 |
| CDC14A | 2493,11 | 0,02 | 0,05 | 0,38 | 7,07E-01 | 8,19E-01 |
| OSBPL10 | 580,70 | 0,05 | 0,13 | 0,38 | 7,07E-01 | 8,19E-01 |
| C2orf74 | 58,18 | 0,02 | 0,07 | 0,38 | 7,07E-01 | 8,19E-01 |
| H2AC14 | 256,82 | -0,05 | 0,13 | -0,38 | 7,07E-01 | 8,20E-01 |
| EPB41L5 | 316,40 | -0,02 | 0,06 | -0,38 | 7,07E-01 | 8,20E-01 |
| CENPE | 139,70 | 0,05 | 0,13 | 0,38 | 7,07E-01 | 8,20E-01 |
| CYP27A1 | 678,67 | 0,05 | 0,12 | 0,38 | 7,07E-01 | 8,20E-01 |
| CNTNAP3C | 441,86 | 0,06 | 0,16 | 0,37 | 7,08E-01 | 8,20E-01 |
| MDM4 | 10801,72 | 0,01 | 0,04 | 0,37 | 7,08E-01 | 8,20E-01 |
| BCAS4 | 419,44 | 0,03 | 0,09 | 0,37 | 7,08E-01 | 8,20E-01 |
| CEP128 | 660,08 | 0,02 | 0,07 | 0,37 | 7,09E-01 | 8,21E-01 |
| DNASE2 | 717,63 | -0,02 | 0,06 | -0,37 | 7,09E-01 | 8,21E-01 |
| SFT2D1 | 1356,67 | 0,02 | 0,05 | 0,37 | 7,09E-01 | 8,21E-01 |
| DIABLO | 318,76 | -0,01 | 0,04 | -0,37 | 7,09E-01 | 8,22E-01 |
| PRELID1 | 2059,48 | 0,02 | 0,05 | 0,37 | 7,09E-01 | 8,22E-01 |
| H2AC6 | 11176,06 | -0,03 | 0,08 | -0,37 | 7,09E-01 | 8,22E-01 |
| HEMGN | 2976,00 | -0,05 | 0,13 | -0,37 | 7,10E-01 | 8,22E-01 |
| GOLGA8N | 252,23 | -0,03 | 0,08 | -0,37 | 7,10E-01 | 8,22E-01 |
| SEPTIN1 | 1400,04 | 0,02 | 0,05 | 0,37 | 7,10E-01 | 8,22E-01 |
| SENP2 | 1275,68 | 0,01 | 0,03 | 0,37 | 7,10E-01 | 8,22E-01 |
| JAZF1 | 1962,01 | -0,03 | 0,09 | -0,37 | 7,10E-01 | 8,22E-01 |
| TTC39C | 1963,43 | 0,02 | 0,05 | 0,37 | 7,10E-01 | 8,22E-01 |
| NOMO3 | 229,83 | -0,04 | 0,11 | -0,37 | 7,10E-01 | 8,22E-01 |

| | | | | | | |
|---|---|---|---|---|---|---|
| DEF6 | 3604,78 | -0,02 | 0,04 | -0,37 | 7,10E-01 | 8,22E-01 |
| ZNF787 | 877,85 | -0,02 | 0,05 | -0,37 | 7,10E-01 | 8,22E-01 |
| KLHL32 | 78,00 | -0,03 | 0,08 | -0,37 | 7,10E-01 | 8,22E-01 |
| RHOBTB3 | 490,25 | -0,03 | 0,09 | -0,37 | 7,11E-01 | 8,22E-01 |
| TCF20 | 2940,26 | -0,02 | 0,05 | -0,37 | 7,11E-01 | 8,22E-01 |
| REPIN1 | 831,76 | -0,02 | 0,06 | -0,37 | 7,11E-01 | 8,22E-01 |
| ARFGAP2 | 2367,98 | -0,01 | 0,04 | -0,37 | 7,11E-01 | 8,22E-01 |
| TRAK1 | 1242,31 | -0,02 | 0,07 | -0,37 | 7,11E-01 | 8,22E-01 |
| DUSP22 | 2214,34 | -0,01 | 0,04 | -0,37 | 7,11E-01 | 8,22E-01 |
| RNF169 | 5124,73 | 0,02 | 0,04 | 0,37 | 7,11E-01 | 8,22E-01 |
| EAF1 | 1240,88 | 0,01 | 0,03 | 0,37 | 7,11E-01 | 8,22E-01 |
| CFAP44 | 348,27 | -0,02 | 0,06 | -0,37 | 7,12E-01 | 8,22E-01 |
| ARIH2 | 2905,70 | 0,01 | 0,03 | 0,37 | 7,12E-01 | 8,22E-01 |
| SETD5 | 3551,38 | -0,02 | 0,05 | -0,37 | 7,12E-01 | 8,23E-01 |
| NKIRAS1 | 102,53 | -0,02 | 0,06 | -0,37 | 7,12E-01 | 8,23E-01 |
| ALDH16A1 | 711,61 | 0,02 | 0,06 | 0,37 | 7,12E-01 | 8,23E-01 |
| DUSP3 | 879,65 | -0,02 | 0,05 | -0,37 | 7,13E-01 | 8,23E-01 |
| NEXMIF | 83,67 | -0,04 | 0,11 | -0,37 | 7,13E-01 | 8,23E-01 |
| C8orf88 | 72,40 | -0,04 | 0,10 | -0,37 | 7,13E-01 | 8,23E-01 |
| NAP1L4 | 5284,96 | 0,01 | 0,04 | 0,37 | 7,13E-01 | 8,23E-01 |
| EFHC2 | 193,26 | 0,03 | 0,07 | 0,37 | 7,13E-01 | 8,23E-01 |
| GET1 | 423,19 | -0,02 | 0,05 | -0,37 | 7,13E-01 | 8,23E-01 |
| HSBP1L1 | 134,90 | -0,02 | 0,07 | -0,37 | 7,13E-01 | 8,23E-01 |
| RBBP5 | 1640,28 | 0,01 | 0,04 | 0,37 | 7,13E-01 | 8,24E-01 |
| TYSND1 | 642,32 | -0,02 | 0,05 | -0,37 | 7,14E-01 | 8,24E-01 |
| BLTP1 | 13885,46 | 0,02 | 0,06 | 0,37 | 7,14E-01 | 8,24E-01 |
| DNA2 | 303,99 | 0,03 | 0,08 | 0,37 | 7,14E-01 | 8,24E-01 |
| RNF145 | 3422,99 | 0,02 | 0,05 | 0,37 | 7,14E-01 | 8,24E-01 |
| ADA2 | 3231,13 | 0,02 | 0,07 | 0,37 | 7,14E-01 | 8,24E-01 |
| IMPG2 | 198,90 | -0,02 | 0,07 | -0,37 | 7,14E-01 | 8,24E-01 |
| PARL | 178,02 | 0,01 | 0,04 | 0,37 | 7,14E-01 | 8,24E-01 |
| ING5 | 1110,50 | -0,02 | 0,04 | -0,37 | 7,14E-01 | 8,24E-01 |
| RBMS2 | 230,72 | -0,04 | 0,11 | -0,37 | 7,15E-01 | 8,24E-01 |
| ORMDL2 | 656,39 | 0,01 | 0,03 | 0,37 | 7,15E-01 | 8,24E-01 |
| SERPINB9 | 2429,82 | -0,02 | 0,05 | -0,37 | 7,15E-01 | 8,24E-01 |
| L3MBTL2 | 467,65 | -0,02 | 0,05 | -0,37 | 7,15E-01 | 8,24E-01 |
| TTC12 | 849,38 | 0,02 | 0,06 | 0,37 | 7,15E-01 | 8,24E-01 |
| TRIO | 1805,74 | 0,02 | 0,07 | 0,37 | 7,15E-01 | 8,24E-01 |
| PARPBP | 146,67 | -0,03 | 0,08 | -0,37 | 7,15E-01 | 8,24E-01 |
| TP53INP1 | 5741,62 | 0,02 | 0,06 | 0,36 | 7,15E-01 | 8,25E-01 |
| CRTC3 | 1534,99 | -0,02 | 0,05 | -0,36 | 7,16E-01 | 8,25E-01 |
| UBE2D3 | 15665,09 | 0,02 | 0,06 | 0,36 | 7,16E-01 | 8,25E-01 |
| MORC2 | 1091,60 | -0,01 | 0,04 | -0,36 | 7,16E-01 | 8,25E-01 |
| CD59 | 1464,47 | 0,02 | 0,06 | 0,36 | 7,16E-01 | 8,26E-01 |
| PPFIBP1 | 163,32 | -0,04 | 0,10 | -0,36 | 7,17E-01 | 8,26E-01 |
| TTC24 | 55,98 | -0,05 | 0,13 | -0,36 | 7,17E-01 | 8,26E-01 |

| | | | | | | |
|---|---|---|---|---|---|---|
| TARS2 | 380,36 | 0,01 | 0,04 | 0,36 | 7,17E-01 | 8,26E-01 |
| TAS2R5 | 58,45 | -0,03 | 0,09 | -0,36 | 7,17E-01 | 8,26E-01 |
| STK38 | 5835,69 | 0,01 | 0,04 | 0,36 | 7,17E-01 | 8,26E-01 |
| CLEC7A | 10954,79 | 0,03 | 0,07 | 0,36 | 7,17E-01 | 8,26E-01 |
| RNF38 | 1849,62 | -0,01 | 0,04 | -0,36 | 7,18E-01 | 8,26E-01 |
| TSPYL2 | 351,35 | -0,02 | 0,06 | -0,36 | 7,18E-01 | 8,27E-01 |
| TIMD4 | 34,99 | -0,04 | 0,11 | -0,36 | 7,18E-01 | 8,27E-01 |
| NME8 | 338,47 | 0,03 | 0,09 | 0,36 | 7,18E-01 | 8,27E-01 |
| RSRP1 | 11826,57 | -0,02 | 0,06 | -0,36 | 7,18E-01 | 8,27E-01 |
| C6orf52 | 28,22 | 0,04 | 0,11 | 0,36 | 7,19E-01 | 8,27E-01 |
| TM9SF1 | 125,27 | -0,01 | 0,04 | -0,36 | 7,19E-01 | 8,28E-01 |
| AHCYL2 | 695,68 | -0,01 | 0,04 | -0,36 | 7,19E-01 | 8,28E-01 |
| SLC25A2 | 27,37 | -0,04 | 0,10 | -0,36 | 7,19E-01 | 8,28E-01 |
| RDH10 | 182,26 | -0,02 | 0,05 | -0,36 | 7,19E-01 | 8,28E-01 |
| PEX5 | 512,84 | 0,01 | 0,04 | 0,36 | 7,19E-01 | 8,28E-01 |
| CLUAP1 | 349,10 | 0,02 | 0,06 | 0,36 | 7,20E-01 | 8,28E-01 |
| IRGM | 40,63 | -0,03 | 0,09 | -0,36 | 7,20E-01 | 8,28E-01 |
| CCDC107 | 360,57 | -0,02 | 0,05 | -0,36 | 7,20E-01 | 8,28E-01 |
| PSEN1 | 4961,94 | -0,02 | 0,05 | -0,36 | 7,20E-01 | 8,28E-01 |
| YIPF6 | 1933,59 | -0,02 | 0,06 | -0,36 | 7,20E-01 | 8,28E-01 |
| ZNRD2 | 67,72 | -0,02 | 0,05 | -0,36 | 7,20E-01 | 8,28E-01 |
| ZBTB4 | 944,94 | -0,03 | 0,07 | -0,36 | 7,20E-01 | 8,28E-01 |
| NUDT16 | 1620,47 | 0,02 | 0,05 | 0,36 | 7,21E-01 | 8,28E-01 |
| WIPF1 | 11590,17 | -0,02 | 0,04 | -0,36 | 7,21E-01 | 8,29E-01 |
| CSRP1 | 809,77 | 0,01 | 0,03 | 0,36 | 7,21E-01 | 8,29E-01 |
| TPST2 | 2054,91 | -0,02 | 0,05 | -0,36 | 7,21E-01 | 8,29E-01 |
| MID1IP1 | 2581,21 | 0,02 | 0,06 | 0,36 | 7,21E-01 | 8,29E-01 |
| ZNF428 | 405,47 | -0,02 | 0,05 | -0,36 | 7,21E-01 | 8,29E-01 |
| ZFP37 | 90,10 | 0,04 | 0,11 | 0,36 | 7,21E-01 | 8,29E-01 |
| CATSPER2 | 286,56 | 0,03 | 0,09 | 0,36 | 7,22E-01 | 8,29E-01 |
| MPG | 514,12 | 0,01 | 0,04 | 0,36 | 7,22E-01 | 8,29E-01 |
| ARMH3 | 1004,89 | -0,01 | 0,04 | -0,35 | 7,24E-01 | 8,31E-01 |
| CPD | 7617,85 | -0,02 | 0,07 | -0,35 | 7,24E-01 | 8,32E-01 |
| RAD54B | 70,27 | 0,03 | 0,09 | 0,35 | 7,24E-01 | 8,32E-01 |
| MED27 | 280,50 | -0,01 | 0,04 | -0,35 | 7,25E-01 | 8,33E-01 |
| GARS1 | 734,81 | 0,01 | 0,04 | 0,35 | 7,25E-01 | 8,33E-01 |
| USP18 | 96,10 | -0,05 | 0,15 | -0,35 | 7,25E-01 | 8,33E-01 |
| NFE2L2 | 2276,62 | 0,02 | 0,05 | 0,35 | 7,25E-01 | 8,33E-01 |
| EGLN1 | 5536,04 | -0,02 | 0,07 | -0,35 | 7,25E-01 | 8,33E-01 |
| COX7A2L | 1477,07 | 0,02 | 0,06 | 0,35 | 7,26E-01 | 8,33E-01 |
| CENATAC | 1082,76 | 0,02 | 0,04 | 0,35 | 7,26E-01 | 8,33E-01 |
| ANKHD1 | 929,22 | 0,02 | 0,05 | 0,35 | 7,26E-01 | 8,33E-01 |
| MEFV | 6376,26 | -0,03 | 0,08 | -0,35 | 7,26E-01 | 8,34E-01 |
| SLC16A4 | 37,88 | -0,04 | 0,13 | -0,35 | 7,27E-01 | 8,34E-01 |
| GMFG | 4563,30 | 0,03 | 0,08 | 0,35 | 7,27E-01 | 8,35E-01 |
| ENSG000002 | 26,57 | 0,02 | 0,07 | 0,35 | 7,28E-01 | 8,35E-01 |

| | | | | | | |
|---|---|---|---|---|---|---|
| MBD1 | 1487,47 | -0,01 | 0,03 | -0,35 | 7,28E-01 | 8,35E-01 |
| HGSNAT | 1137,61 | 0,02 | 0,05 | 0,35 | 7,28E-01 | 8,35E-01 |
| GAB1 | 1700,71 | 0,02 | 0,06 | 0,35 | 7,29E-01 | 8,36E-01 |
| ZNF80 | 53,72 | 0,05 | 0,15 | 0,35 | 7,29E-01 | 8,36E-01 |
| CDK5 | 147,51 | -0,02 | 0,05 | -0,35 | 7,29E-01 | 8,36E-01 |
| ANKRD23 | 91,56 | -0,02 | 0,05 | -0,35 | 7,29E-01 | 8,36E-01 |
| SLC11A2 | 1265,46 | -0,01 | 0,03 | -0,35 | 7,30E-01 | 8,36E-01 |
| DSTYK | 1593,08 | 0,01 | 0,03 | 0,35 | 7,30E-01 | 8,36E-01 |
| ZNF208 | 88,45 | 0,05 | 0,14 | 0,35 | 7,30E-01 | 8,36E-01 |
| CACUL1 | 5839,54 | 0,02 | 0,05 | 0,35 | 7,30E-01 | 8,37E-01 |
| CCDC51 | 135,74 | 0,02 | 0,05 | 0,35 | 7,30E-01 | 8,37E-01 |
| SNX22 | 511,62 | -0,03 | 0,08 | -0,34 | 7,30E-01 | 8,37E-01 |
| CAPN2 | 3713,89 | 0,01 | 0,04 | 0,34 | 7,30E-01 | 8,37E-01 |
| MPPE1 | 3942,98 | -0,02 | 0,06 | -0,34 | 7,30E-01 | 8,37E-01 |
| FAM53B | 2332,78 | -0,02 | 0,06 | -0,34 | 7,30E-01 | 8,37E-01 |
| MED13L | 13889,52 | -0,01 | 0,04 | -0,34 | 7,30E-01 | 8,37E-01 |
| DAZAP2 | 8596,45 | -0,02 | 0,05 | -0,34 | 7,30E-01 | 8,37E-01 |
| ANO9 | 468,88 | -0,03 | 0,09 | -0,34 | 7,31E-01 | 8,37E-01 |
| CAPNS2 | 188,48 | -0,03 | 0,09 | -0,34 | 7,31E-01 | 8,37E-01 |
| ANKRD18A | 129,42 | -0,03 | 0,10 | -0,34 | 7,31E-01 | 8,37E-01 |
| ZNF620 | 112,86 | -0,02 | 0,06 | -0,34 | 7,31E-01 | 8,37E-01 |
| PARP9 | 4655,89 | -0,03 | 0,09 | -0,34 | 7,31E-01 | 8,37E-01 |
| MMRN1 | 360,76 | -0,05 | 0,14 | -0,34 | 7,32E-01 | 8,37E-01 |
| GATM | 106,99 | 0,04 | 0,12 | 0,34 | 7,32E-01 | 8,37E-01 |
| NAT10 | 1044,34 | 0,02 | 0,04 | 0,34 | 7,32E-01 | 8,37E-01 |
| YARS1 | 925,73 | -0,01 | 0,04 | -0,34 | 7,32E-01 | 8,37E-01 |
| ZNF79 | 464,08 | 0,01 | 0,03 | 0,34 | 7,32E-01 | 8,37E-01 |
| AEN | 848,63 | -0,02 | 0,05 | -0,34 | 7,32E-01 | 8,38E-01 |
| OR52K1 | 327,67 | -0,04 | 0,11 | -0,34 | 7,33E-01 | 8,38E-01 |
| CD79A | 180,58 | 0,05 | 0,14 | 0,34 | 7,33E-01 | 8,38E-01 |
| STX10 | 1707,96 | -0,02 | 0,06 | -0,34 | 7,33E-01 | 8,38E-01 |
| C1QTNF6 | 404,79 | 0,02 | 0,05 | 0,34 | 7,32E-01 | 8,38E-01 |
| ERP27 | 469,57 | -0,02 | 0,06 | -0,34 | 7,33E-01 | 8,38E-01 |
| GALNT3 | 1216,20 | 0,02 | 0,07 | 0,34 | 7,33E-01 | 8,38E-01 |
| SEC61A1 | 2811,95 | -0,01 | 0,03 | -0,34 | 7,33E-01 | 8,38E-01 |
| CWC25 | 1043,97 | 0,01 | 0,04 | 0,34 | 7,33E-01 | 8,38E-01 |
| RBM33 | 6839,75 | 0,01 | 0,04 | 0,34 | 7,34E-01 | 8,39E-01 |
| CTSW | 375,11 | 0,04 | 0,12 | 0,34 | 7,34E-01 | 8,39E-01 |
| EXD2 | 287,11 | 0,02 | 0,05 | 0,34 | 7,34E-01 | 8,39E-01 |
| STYXL1 | 409,07 | 0,01 | 0,04 | 0,34 | 7,34E-01 | 8,39E-01 |
| ODC1 | 5304,41 | -0,04 | 0,10 | -0,34 | 7,34E-01 | 8,39E-01 |
| RGPD1 | 32,19 | -0,06 | 0,17 | -0,34 | 7,35E-01 | 8,40E-01 |
| EML2 | 680,79 | -0,01 | 0,04 | -0,34 | 7,35E-01 | 8,40E-01 |
| MIPEP | 272,67 | 0,02 | 0,05 | 0,34 | 7,35E-01 | 8,40E-01 |
| SERPINB2 | 59,25 | 0,04 | 0,11 | 0,34 | 7,35E-01 | 8,40E-01 |
| HEATR5A | 966,41 | -0,02 | 0,05 | -0,34 | 7,36E-01 | 8,40E-01 |

| | | | | | | |
|---|---|---|---|---|---|---|
| DGKG | 672,77 | -0,03 | 0,08 | -0,34 | 7,36E-01 | 8,41E-01 |
| DOLK | 128,90 | 0,01 | 0,04 | 0,34 | 7,36E-01 | 8,41E-01 |
| UQCR10 | 523,44 | 0,02 | 0,07 | 0,34 | 7,36E-01 | 8,41E-01 |
| RNF227 | 112,62 | -0,01 | 0,04 | -0,34 | 7,36E-01 | 8,41E-01 |
| ANKRD42 | 386,46 | 0,01 | 0,03 | 0,34 | 7,37E-01 | 8,41E-01 |
| CYP7B1 | 32,85 | -0,05 | 0,16 | -0,34 | 7,37E-01 | 8,42E-01 |
| ZXDC | 2583,96 | -0,02 | 0,05 | -0,34 | 7,37E-01 | 8,42E-01 |
| LAMTOR4 | 1972,70 | 0,02 | 0,06 | 0,34 | 7,38E-01 | 8,42E-01 |
| MRPS24 | 31,47 | 0,03 | 0,08 | 0,33 | 7,38E-01 | 8,42E-01 |
| MBD5 | 3501,29 | 0,02 | 0,05 | 0,33 | 7,38E-01 | 8,42E-01 |
| MDN1 | 3610,06 | -0,02 | 0,06 | -0,34 | 7,37E-01 | 8,42E-01 |
| AKAP6 | 226,50 | 0,04 | 0,12 | 0,34 | 7,37E-01 | 8,42E-01 |
| PHLDA1 | 155,12 | 0,03 | 0,08 | 0,33 | 7,38E-01 | 8,42E-01 |
| S1PR5 | 1038,14 | 0,03 | 0,10 | 0,34 | 7,38E-01 | 8,42E-01 |
| GOLIM4 | 460,79 | 0,02 | 0,05 | 0,33 | 7,38E-01 | 8,42E-01 |
| MED20 | 396,25 | 0,01 | 0,03 | 0,33 | 7,38E-01 | 8,42E-01 |
| FBN2 | 527,10 | -0,04 | 0,10 | -0,33 | 7,38E-01 | 8,42E-01 |
| DGAT1 | 819,88 | -0,02 | 0,06 | -0,33 | 7,38E-01 | 8,42E-01 |
| SPAG9 | 8653,12 | 0,02 | 0,05 | 0,33 | 7,39E-01 | 8,42E-01 |
| PDPR | 1130,43 | 0,03 | 0,10 | 0,33 | 7,39E-01 | 8,42E-01 |
| ZCCHC18 | 111,45 | 0,03 | 0,09 | 0,33 | 7,39E-01 | 8,42E-01 |
| DEPTOR | 45,23 | -0,04 | 0,12 | -0,33 | 7,39E-01 | 8,43E-01 |
| ALG3 | 175,85 | -0,01 | 0,04 | -0,33 | 7,39E-01 | 8,43E-01 |
| LETM1 | 778,02 | -0,02 | 0,05 | -0,33 | 7,39E-01 | 8,43E-01 |
| MZT2A | 620,60 | 0,02 | 0,05 | 0,33 | 7,39E-01 | 8,43E-01 |
| XRCC2 | 66,73 | -0,04 | 0,11 | -0,33 | 7,40E-01 | 8,43E-01 |
| PRKDC | 6622,70 | 0,02 | 0,05 | 0,33 | 7,40E-01 | 8,43E-01 |
| DGKQ | 627,22 | -0,03 | 0,09 | -0,33 | 7,40E-01 | 8,43E-01 |
| CCDC88B | 5773,77 | -0,03 | 0,08 | -0,33 | 7,40E-01 | 8,43E-01 |
| TLR3 | 82,56 | -0,04 | 0,11 | -0,33 | 7,40E-01 | 8,43E-01 |
| NUP50 | 5167,28 | 0,01 | 0,04 | 0,33 | 7,40E-01 | 8,43E-01 |
| SQOR | 1188,59 | -0,02 | 0,06 | -0,33 | 7,41E-01 | 8,44E-01 |
| NT5DC3 | 281,39 | 0,03 | 0,09 | 0,33 | 7,41E-01 | 8,44E-01 |
| IQGAP1 | 21548,21 | 0,02 | 0,06 | 0,33 | 7,42E-01 | 8,44E-01 |
| TCAF2 | 183,88 | -0,02 | 0,07 | -0,33 | 7,42E-01 | 8,44E-01 |
| HMOX1 | 301,58 | -0,03 | 0,08 | -0,33 | 7,42E-01 | 8,45E-01 |
| WDR37 | 2533,18 | -0,01 | 0,04 | -0,33 | 7,42E-01 | 8,45E-01 |
| PAN3 | 5122,22 | 0,02 | 0,05 | 0,33 | 7,42E-01 | 8,45E-01 |
| CCR6 | 138,13 | 0,03 | 0,09 | 0,33 | 7,42E-01 | 8,45E-01 |
| PIGB | 2108,76 | 0,02 | 0,07 | 0,33 | 7,43E-01 | 8,45E-01 |
| EIF1 | 3117,56 | -0,02 | 0,07 | -0,33 | 7,43E-01 | 8,45E-01 |
| CLEC4D | 654,57 | 0,04 | 0,11 | 0,33 | 7,43E-01 | 8,46E-01 |
| TP53INP2 | 155,47 | -0,04 | 0,12 | -0,33 | 7,43E-01 | 8,46E-01 |
| ILKAP | 690,92 | 0,02 | 0,05 | 0,33 | 7,43E-01 | 8,46E-01 |
| MEMO1 | 621,74 | 0,02 | 0,05 | 0,33 | 7,44E-01 | 8,46E-01 |
| ARL3 | 158,49 | -0,01 | 0,04 | -0,33 | 7,44E-01 | 8,46E-01 |

| | | | | | | |
|---|---|---|---|---|---|---|
| RHBDD3 | 183,55 | -0,02 | 0,05 | -0,33 | 7,44E-01 | 8,46E-01 |
| BASP1 | 5722,84 | -0,03 | 0,10 | -0,33 | 7,43E-01 | 8,46E-01 |
| TXN | 689,83 | -0,04 | 0,11 | -0,33 | 7,43E-01 | 8,46E-01 |
| ELOA | 598,20 | -0,01 | 0,04 | -0,33 | 7,44E-01 | 8,46E-01 |
| MAPKAPK3 | 2825,27 | -0,02 | 0,05 | -0,33 | 7,44E-01 | 8,46E-01 |
| MRTFB | 615,98 | 0,02 | 0,06 | 0,33 | 7,44E-01 | 8,46E-01 |
| NFKBIA | 2481,58 | -0,02 | 0,05 | -0,33 | 7,44E-01 | 8,46E-01 |
| OGA | 16088,22 | 0,01 | 0,04 | 0,33 | 7,44E-01 | 8,46E-01 |
| TXNDC12 | 411,25 | 0,02 | 0,05 | 0,33 | 7,44E-01 | 8,46E-01 |
| OSTF1 | 5626,28 | 0,02 | 0,06 | 0,33 | 7,45E-01 | 8,46E-01 |
| SUSD3 | 430,42 | -0,02 | 0,05 | -0,33 | 7,45E-01 | 8,46E-01 |
| WBP2NL | 124,21 | -0,02 | 0,07 | -0,33 | 7,45E-01 | 8,47E-01 |
| HMGB3 | 48,42 | -0,04 | 0,11 | -0,32 | 7,45E-01 | 8,47E-01 |
| GOLPH3L | 443,80 | -0,01 | 0,03 | -0,32 | 7,45E-01 | 8,47E-01 |
| COPS7B | 577,07 | -0,02 | 0,05 | -0,32 | 7,46E-01 | 8,47E-01 |
| ZCCHC2 | 2474,27 | -0,03 | 0,08 | -0,32 | 7,46E-01 | 8,48E-01 |
| XPR1 | 1502,67 | -0,01 | 0,03 | -0,32 | 7,46E-01 | 8,48E-01 |
| LPCAT2 | 3943,73 | -0,03 | 0,08 | -0,32 | 7,46E-01 | 8,48E-01 |
| SLA | 13172,08 | -0,02 | 0,06 | -0,32 | 7,46E-01 | 8,48E-01 |
| ENSG000002 | 57,93 | -0,03 | 0,08 | -0,32 | 7,47E-01 | 8,48E-01 |
| NOL9 | 890,81 | 0,02 | 0,05 | 0,32 | 7,47E-01 | 8,48E-01 |
| ZBTB39 | 378,71 | -0,01 | 0,04 | -0,32 | 7,47E-01 | 8,48E-01 |
| TRAPPC2B | 59,05 | 0,03 | 0,09 | 0,32 | 7,47E-01 | 8,48E-01 |
| PRDM1 | 1213,78 | 0,02 | 0,05 | 0,32 | 7,47E-01 | 8,48E-01 |
| MARCHF3 | 293,32 | -0,02 | 0,08 | -0,32 | 7,47E-01 | 8,48E-01 |
| HNRNPUL2 | 630,32 | 0,01 | 0,03 | 0,32 | 7,47E-01 | 8,48E-01 |
| FKBP15 | 3815,36 | -0,01 | 0,04 | -0,32 | 7,48E-01 | 8,49E-01 |
| H2BC5 | 1979,89 | -0,02 | 0,07 | -0,32 | 7,48E-01 | 8,49E-01 |
| ARF4 | 1123,06 | -0,01 | 0,04 | -0,32 | 7,49E-01 | 8,49E-01 |
| FOXJ3 | 2044,33 | 0,01 | 0,04 | 0,32 | 7,49E-01 | 8,49E-01 |
| NDUFAF8 | 225,49 | 0,02 | 0,07 | 0,32 | 7,48E-01 | 8,49E-01 |
| TOP1 | 2243,35 | -0,01 | 0,05 | -0,32 | 7,49E-01 | 8,49E-01 |
| TERF2IP | 2900,70 | -0,02 | 0,06 | -0,32 | 7,49E-01 | 8,49E-01 |
| MAP2K6 | 2211,05 | 0,02 | 0,05 | 0,32 | 7,49E-01 | 8,49E-01 |
| RNF4 | 2136,72 | 0,01 | 0,03 | 0,32 | 7,49E-01 | 8,49E-01 |
| LIPA | 1572,31 | 0,02 | 0,07 | 0,32 | 7,49E-01 | 8,49E-01 |
| IL23R | 48,88 | -0,06 | 0,17 | -0,32 | 7,49E-01 | 8,49E-01 |
| ZCCHC3 | 1041,66 | 0,01 | 0,03 | 0,32 | 7,49E-01 | 8,50E-01 |
| GCHFR | 181,08 | -0,02 | 0,06 | -0,32 | 7,50E-01 | 8,50E-01 |
| GLRX | 2579,67 | -0,02 | 0,06 | -0,32 | 7,50E-01 | 8,50E-01 |
| PAFAH1B1 | 2497,37 | -0,01 | 0,04 | -0,32 | 7,50E-01 | 8,50E-01 |
| RIOK3 | 11395,67 | -0,04 | 0,11 | -0,32 | 7,50E-01 | 8,50E-01 |
| AK5 | 397,56 | 0,03 | 0,10 | 0,32 | 7,50E-01 | 8,50E-01 |
| CHTOP | 2957,33 | 0,01 | 0,03 | 0,32 | 7,50E-01 | 8,50E-01 |
| ULK2 | 826,91 | 0,01 | 0,04 | 0,32 | 7,51E-01 | 8,51E-01 |
| INPP1 | 293,58 | 0,02 | 0,05 | 0,32 | 7,51E-01 | 8,51E-01 |

| | | | | | | |
|---|---|---|---|---|---|---|
| ZNF875 | 1001,80 | -0,02 | 0,05 | -0,32 | 7,51E-01 | 8,51E-01 |
| SLC15A2 | 648,71 | 0,02 | 0,08 | 0,32 | 7,52E-01 | 8,51E-01 |
| HINT2 | 47,14 | 0,02 | 0,08 | 0,32 | 7,51E-01 | 8,51E-01 |
| ZNF284 | 84,13 | 0,01 | 0,05 | 0,32 | 7,51E-01 | 8,51E-01 |
| APLP2 | 6203,64 | 0,02 | 0,06 | 0,32 | 7,52E-01 | 8,51E-01 |
| SENP3 | 294,02 | -0,01 | 0,03 | -0,32 | 7,52E-01 | 8,51E-01 |
| UFC1 | 615,60 | 0,02 | 0,06 | 0,32 | 7,52E-01 | 8,51E-01 |
| HIP1R | 901,14 | -0,02 | 0,07 | -0,32 | 7,52E-01 | 8,51E-01 |
| SFSWAP | 2460,31 | 0,01 | 0,03 | 0,32 | 7,52E-01 | 8,52E-01 |
| BRCA1 | 477,42 | 0,02 | 0,06 | 0,32 | 7,53E-01 | 8,52E-01 |
| BLTP2 | 4559,96 | -0,01 | 0,04 | -0,32 | 7,53E-01 | 8,52E-01 |
| STMP1 | 1657,56 | -0,03 | 0,09 | -0,31 | 7,53E-01 | 8,52E-01 |
| DGKH | 1244,77 | -0,02 | 0,06 | -0,31 | 7,53E-01 | 8,52E-01 |
| HAVCR1 | 77,08 | -0,03 | 0,09 | -0,31 | 7,54E-01 | 8,52E-01 |
| ZNF324B | 174,50 | -0,02 | 0,05 | -0,31 | 7,54E-01 | 8,52E-01 |
| NUDT12 | 74,52 | 0,04 | 0,14 | 0,31 | 7,54E-01 | 8,52E-01 |
| ZNF331 | 507,22 | 0,02 | 0,05 | 0,31 | 7,54E-01 | 8,52E-01 |
| DRC12 | 98,14 | 0,03 | 0,09 | 0,31 | 7,54E-01 | 8,52E-01 |
| VSIG1 | 283,05 | 0,02 | 0,07 | 0,31 | 7,54E-01 | 8,52E-01 |
| FAF2 | 1185,34 | 0,01 | 0,03 | 0,31 | 7,54E-01 | 8,52E-01 |
| POLR2H | 401,65 | -0,01 | 0,03 | -0,31 | 7,54E-01 | 8,52E-01 |
| MR1 | 1935,99 | 0,01 | 0,03 | 0,31 | 7,54E-01 | 8,52E-01 |
| TRADD | 1542,62 | -0,01 | 0,04 | -0,31 | 7,54E-01 | 8,52E-01 |
| ANKRD31 | 62,00 | -0,04 | 0,13 | -0,31 | 7,54E-01 | 8,53E-01 |
| GBP1 | 2882,27 | -0,04 | 0,12 | -0,31 | 7,55E-01 | 8,53E-01 |
| UBQLN2 | 815,37 | 0,02 | 0,05 | 0,31 | 7,55E-01 | 8,53E-01 |
| SAXO2 | 55,15 | -0,06 | 0,20 | -0,31 | 7,55E-01 | 8,53E-01 |
| C1orf115 | 71,21 | -0,03 | 0,10 | -0,31 | 7,55E-01 | 8,53E-01 |
| GRAMD2B | 149,26 | 0,02 | 0,06 | 0,31 | 7,55E-01 | 8,53E-01 |
| PARP12 | 2332,89 | -0,02 | 0,07 | -0,31 | 7,55E-01 | 8,53E-01 |
| PELO | 330,74 | -0,02 | 0,06 | -0,31 | 7,55E-01 | 8,53E-01 |
| CKAP5 | 1179,55 | 0,01 | 0,04 | 0,31 | 7,56E-01 | 8,53E-01 |
| PRMT2 | 5064,66 | 0,01 | 0,04 | 0,31 | 7,55E-01 | 8,53E-01 |
| HMGCL | 604,25 | 0,01 | 0,03 | 0,31 | 7,56E-01 | 8,53E-01 |
| MS4A14 | 375,86 | 0,03 | 0,09 | 0,31 | 7,56E-01 | 8,54E-01 |
| ZNF491 | 99,56 | 0,02 | 0,07 | 0,31 | 7,56E-01 | 8,54E-01 |
| CHCHD2 | 2224,96 | 0,02 | 0,07 | 0,31 | 7,56E-01 | 8,54E-01 |
| TLE1 | 319,67 | -0,02 | 0,08 | -0,31 | 7,56E-01 | 8,54E-01 |
| PNPO | 282,62 | -0,01 | 0,04 | -0,31 | 7,56E-01 | 8,54E-01 |
| POLR1A | 921,44 | -0,02 | 0,06 | -0,31 | 7,56E-01 | 8,54E-01 |
| ECI1 | 341,18 | 0,01 | 0,04 | 0,31 | 7,57E-01 | 8,54E-01 |
| MED19 | 111,26 | -0,01 | 0,04 | -0,31 | 7,57E-01 | 8,54E-01 |
| ATP7A | 2402,77 | 0,02 | 0,05 | 0,31 | 7,57E-01 | 8,54E-01 |
| TRIP12 | 11729,89 | 0,01 | 0,04 | 0,31 | 7,57E-01 | 8,54E-01 |
| VPS16 | 1161,75 | 0,01 | 0,03 | 0,31 | 7,58E-01 | 8,56E-01 |
| SMC1A | 2061,95 | 0,01 | 0,04 | 0,31 | 7,59E-01 | 8,56E-01 |

| | | | | | | |
|---|---|---|---|---|---|---|
| AKAP12 | 219,59 | -0,06 | 0,19 | -0,31 | 7,59E-01 | 8,56E-01 |
| ICMT | 785,97 | -0,01 | 0,03 | -0,31 | 7,59E-01 | 8,56E-01 |
| DTX3L | 3815,85 | -0,02 | 0,07 | -0,31 | 7,59E-01 | 8,56E-01 |
| DVL1 | 536,46 | 0,01 | 0,04 | 0,31 | 7,59E-01 | 8,56E-01 |
| RAPGEF2 | 4598,44 | -0,02 | 0,06 | -0,31 | 7,59E-01 | 8,56E-01 |
| SCN3A | 185,98 | 0,04 | 0,15 | 0,31 | 7,60E-01 | 8,56E-01 |
| FREY1 | 23,91 | -0,02 | 0,07 | -0,31 | 7,60E-01 | 8,56E-01 |
| GPR68 | 257,40 | -0,02 | 0,08 | -0,31 | 7,60E-01 | 8,57E-01 |
| STS | 240,09 | -0,02 | 0,05 | -0,31 | 7,60E-01 | 8,57E-01 |
| HDAC8 | 844,68 | 0,02 | 0,06 | 0,30 | 7,60E-01 | 8,57E-01 |
| FAM227B | 142,41 | -0,03 | 0,09 | -0,31 | 7,60E-01 | 8,57E-01 |
| SPG21 | 2241,95 | -0,01 | 0,04 | -0,30 | 7,61E-01 | 8,57E-01 |
| BABAM2 | 573,88 | 0,01 | 0,05 | 0,30 | 7,61E-01 | 8,57E-01 |
| RNF41 | 2074,13 | -0,01 | 0,04 | -0,30 | 7,62E-01 | 8,58E-01 |
| SUN2 | 8440,80 | 0,02 | 0,05 | 0,30 | 7,62E-01 | 8,58E-01 |
| NKAP | 464,34 | 0,01 | 0,03 | 0,30 | 7,63E-01 | 8,59E-01 |
| THOC3 | 688,69 | 0,02 | 0,05 | 0,30 | 7,63E-01 | 8,59E-01 |
| ZNF343 | 260,62 | 0,01 | 0,04 | 0,30 | 7,63E-01 | 8,59E-01 |
| LRRIQ3 | 45,64 | -0,05 | 0,17 | -0,30 | 7,63E-01 | 8,59E-01 |
| GPATCH3 | 250,07 | -0,01 | 0,04 | -0,30 | 7,63E-01 | 8,59E-01 |
| PGPEP1 | 772,22 | -0,02 | 0,06 | -0,30 | 7,62E-01 | 8,59E-01 |
| ZCCHC14 | 540,86 | 0,01 | 0,04 | 0,30 | 7,63E-01 | 8,59E-01 |
| ING1 | 895,03 | 0,01 | 0,03 | 0,30 | 7,62E-01 | 8,59E-01 |
| GPANK1 | 643,22 | -0,01 | 0,04 | -0,30 | 7,63E-01 | 8,59E-01 |
| ZNF471 | 71,85 | -0,04 | 0,14 | -0,30 | 7,63E-01 | 8,59E-01 |
| SLC25A28 | 606,82 | 0,01 | 0,03 | 0,30 | 7,63E-01 | 8,59E-01 |
| TMCC3 | 2675,98 | -0,02 | 0,07 | -0,30 | 7,63E-01 | 8,59E-01 |
| TBC1D8 | 841,86 | 0,02 | 0,07 | 0,30 | 7,64E-01 | 8,59E-01 |
| H2BC8 | 688,13 | 0,02 | 0,08 | 0,30 | 7,64E-01 | 8,59E-01 |
| RAB6A | 3000,67 | -0,02 | 0,06 | -0,30 | 7,64E-01 | 8,60E-01 |
| RPL12 | 3573,66 | 0,02 | 0,07 | 0,30 | 7,64E-01 | 8,60E-01 |
| CRIPT | 687,22 | 0,01 | 0,05 | 0,30 | 7,64E-01 | 8,60E-01 |
| VPS52 | 888,64 | 0,01 | 0,04 | 0,30 | 7,65E-01 | 8,60E-01 |
| MILR1 | 401,44 | -0,02 | 0,08 | -0,30 | 7,65E-01 | 8,60E-01 |
| APOO | 69,69 | -0,02 | 0,05 | -0,30 | 7,66E-01 | 8,61E-01 |
| GPR15 | 201,12 | 0,05 | 0,16 | 0,30 | 7,66E-01 | 8,61E-01 |
| CYP4V2 | 1213,23 | 0,02 | 0,06 | 0,30 | 7,66E-01 | 8,61E-01 |
| IST1 | 4302,40 | 0,01 | 0,03 | 0,30 | 7,66E-01 | 8,61E-01 |
| POLE2 | 146,23 | -0,04 | 0,13 | -0,30 | 7,66E-01 | 8,61E-01 |
| TIGD5 | 252,47 | -0,02 | 0,06 | -0,30 | 7,67E-01 | 8,62E-01 |
| MTCL2 | 303,06 | -0,04 | 0,12 | -0,30 | 7,67E-01 | 8,62E-01 |
| RNASEH2A | 23,93 | 0,03 | 0,09 | 0,30 | 7,67E-01 | 8,62E-01 |
| PRLR | 136,81 | -0,03 | 0,11 | -0,30 | 7,68E-01 | 8,62E-01 |
| CBR3 | 56,94 | 0,03 | 0,11 | 0,30 | 7,68E-01 | 8,62E-01 |
| RAB20 | 344,52 | 0,03 | 0,10 | 0,30 | 7,68E-01 | 8,62E-01 |
| PIGL | 575,70 | 0,01 | 0,05 | 0,30 | 7,68E-01 | 8,63E-01 |

| | | | | | | |
|---|---|---|---|---|---|---|
| TMEM245 | 2164,59 | 0,01 | 0,05 | 0,29 | 7,68E-01 | 8,63E-01 |
| BBS4 | 371,66 | -0,01 | 0,04 | -0,29 | 7,68E-01 | 8,63E-01 |
| MTMR10 | 4515,80 | 0,01 | 0,05 | 0,29 | 7,68E-01 | 8,63E-01 |
| USP10 | 3107,47 | -0,02 | 0,06 | -0,29 | 7,68E-01 | 8,63E-01 |
| ZNF398 | 1104,16 | -0,01 | 0,03 | -0,29 | 7,69E-01 | 8,63E-01 |
| CD101 | 612,29 | -0,02 | 0,08 | -0,29 | 7,69E-01 | 8,63E-01 |
| LPP | 5802,94 | 0,01 | 0,05 | 0,29 | 7,69E-01 | 8,63E-01 |
| EFNB1 | 122,10 | -0,02 | 0,06 | -0,29 | 7,69E-01 | 8,63E-01 |
| PABPC1 | 8493,25 | 0,01 | 0,05 | 0,29 | 7,69E-01 | 8,64E-01 |
| METTL25B | 316,28 | -0,01 | 0,05 | -0,29 | 7,70E-01 | 8,64E-01 |
| CYBRD1 | 1276,29 | -0,02 | 0,08 | -0,29 | 7,70E-01 | 8,64E-01 |
| ENTR1 | 509,16 | 0,01 | 0,04 | 0,29 | 7,70E-01 | 8,64E-01 |
| LMAN2 | 2487,80 | -0,01 | 0,04 | -0,29 | 7,71E-01 | 8,64E-01 |
| DNAJB6 | 3849,98 | -0,02 | 0,05 | -0,29 | 7,70E-01 | 8,64E-01 |
| MREG | 202,80 | -0,01 | 0,05 | -0,29 | 7,70E-01 | 8,64E-01 |
| PABIR1 | 704,07 | -0,01 | 0,03 | -0,29 | 7,71E-01 | 8,64E-01 |
| ACACA | 826,02 | -0,02 | 0,06 | -0,29 | 7,71E-01 | 8,65E-01 |
| QRICH1 | 1549,78 | -0,01 | 0,04 | -0,29 | 7,71E-01 | 8,65E-01 |
| PHF11 | 1614,83 | 0,01 | 0,05 | 0,29 | 7,71E-01 | 8,65E-01 |
| LILRB1 | 670,34 | -0,03 | 0,09 | -0,29 | 7,71E-01 | 8,65E-01 |
| MAN2B1 | 1957,63 | -0,02 | 0,06 | -0,29 | 7,71E-01 | 8,65E-01 |
| RAD51D | 624,22 | -0,01 | 0,05 | -0,29 | 7,72E-01 | 8,65E-01 |
| BIVM | 182,10 | 0,02 | 0,06 | 0,29 | 7,72E-01 | 8,65E-01 |
| STARD3NL | 770,92 | 0,02 | 0,05 | 0,29 | 7,72E-01 | 8,65E-01 |
| ENSG000001 | 438,23 | -0,02 | 0,06 | -0,29 | 7,72E-01 | 8,66E-01 |
| ADI1 | 965,35 | 0,01 | 0,04 | 0,29 | 7,72E-01 | 8,66E-01 |
| KIF15 | 117,50 | -0,04 | 0,14 | -0,29 | 7,73E-01 | 8,66E-01 |
| TRAF3IP1 | 258,73 | -0,02 | 0,06 | -0,29 | 7,73E-01 | 8,66E-01 |
| AVPI1 | 32,94 | -0,03 | 0,09 | -0,29 | 7,73E-01 | 8,66E-01 |
| SLC43A3 | 268,63 | -0,02 | 0,06 | -0,29 | 7,73E-01 | 8,66E-01 |
| WASHC2C | 1650,33 | -0,01 | 0,05 | -0,29 | 7,73E-01 | 8,66E-01 |
| TSHZ2 | 474,60 | -0,03 | 0,11 | -0,29 | 7,73E-01 | 8,66E-01 |
| ABTB3 | 562,26 | -0,02 | 0,07 | -0,29 | 7,73E-01 | 8,66E-01 |
| SLC25A17 | 385,71 | 0,01 | 0,03 | 0,29 | 7,74E-01 | 8,66E-01 |
| TET2 | 11847,75 | -0,02 | 0,06 | -0,29 | 7,74E-01 | 8,66E-01 |
| CLTCL1 | 350,52 | -0,03 | 0,09 | -0,29 | 7,74E-01 | 8,66E-01 |
| CD300E | 1217,52 | -0,02 | 0,09 | -0,29 | 7,74E-01 | 8,66E-01 |
| CINP | 549,30 | -0,01 | 0,03 | -0,29 | 7,74E-01 | 8,66E-01 |
| DISC1 | 1416,29 | -0,02 | 0,07 | -0,29 | 7,74E-01 | 8,67E-01 |
| POLG | 3206,27 | -0,01 | 0,04 | -0,29 | 7,75E-01 | 8,67E-01 |
| HHAT | 220,12 | -0,02 | 0,08 | -0,29 | 7,75E-01 | 8,67E-01 |
| NKX3-1 | 211,80 | 0,07 | 0,25 | 0,29 | 7,76E-01 | 8,68E-01 |
| PPIH | 348,06 | 0,01 | 0,04 | 0,28 | 7,76E-01 | 8,68E-01 |
| COP1 | 6976,61 | 0,02 | 0,06 | 0,28 | 7,76E-01 | 8,68E-01 |
| CDH2 | 61,32 | 0,05 | 0,18 | 0,28 | 7,76E-01 | 8,68E-01 |
| CTDP1 | 2241,71 | -0,01 | 0,05 | -0,28 | 7,76E-01 | 8,68E-01 |

| | | | | | | |
|---|---|---|---|---|---|---|
| SLC22A15 | 2192,51 | -0,02 | 0,07 | -0,28 | 7,76E-01 | 8,68E-01 |
| LCP2 | 9324,06 | -0,01 | 0,04 | -0,28 | 7,76E-01 | 8,68E-01 |
| ZNF221 | 82,45 | 0,02 | 0,09 | 0,28 | 7,77E-01 | 8,68E-01 |
| SPMIP4 | 75,64 | -0,02 | 0,08 | -0,28 | 7,77E-01 | 8,68E-01 |
| ZNF382 | 259,06 | 0,02 | 0,06 | 0,28 | 7,77E-01 | 8,68E-01 |
| MED14 | 1466,70 | -0,01 | 0,04 | -0,28 | 7,77E-01 | 8,69E-01 |
| SAFB2 | 2460,68 | -0,01 | 0,03 | -0,28 | 7,78E-01 | 8,69E-01 |
| DUSP7 | 624,65 | -0,01 | 0,05 | -0,28 | 7,78E-01 | 8,70E-01 |
| CDC27 | 2845,52 | 0,02 | 0,06 | 0,28 | 7,78E-01 | 8,70E-01 |
| MMAB | 230,95 | 0,01 | 0,05 | 0,28 | 7,78E-01 | 8,70E-01 |
| EFCAB11 | 118,75 | -0,02 | 0,07 | -0,28 | 7,78E-01 | 8,70E-01 |
| IL2RA | 332,36 | -0,02 | 0,07 | -0,28 | 7,78E-01 | 8,70E-01 |
| HMGN2 | 3811,70 | 0,01 | 0,04 | 0,28 | 7,78E-01 | 8,70E-01 |
| TKTL1 | 104,76 | 0,03 | 0,11 | 0,28 | 7,79E-01 | 8,70E-01 |
| TRAPPC2L | 358,57 | -0,01 | 0,05 | -0,28 | 7,79E-01 | 8,70E-01 |
| CSF1R | 1301,35 | -0,03 | 0,10 | -0,28 | 7,80E-01 | 8,71E-01 |
| AAR2 | 245,25 | -0,01 | 0,04 | -0,28 | 7,80E-01 | 8,71E-01 |
| VPS13B | 9869,14 | 0,01 | 0,05 | 0,28 | 7,80E-01 | 8,71E-01 |
| DISP1 | 131,03 | 0,02 | 0,06 | 0,28 | 7,80E-01 | 8,71E-01 |
| SLC20A1 | 3069,93 | -0,01 | 0,05 | -0,28 | 7,80E-01 | 8,71E-01 |
| DYNLL1 | 686,82 | -0,02 | 0,06 | -0,28 | 7,80E-01 | 8,71E-01 |
| DEXI | 532,27 | 0,01 | 0,04 | 0,28 | 7,80E-01 | 8,71E-01 |
| PPP1R11 | 329,72 | -0,01 | 0,05 | -0,28 | 7,81E-01 | 8,71E-01 |
| PANK2 | 2984,72 | -0,01 | 0,04 | -0,28 | 7,81E-01 | 8,71E-01 |
| DCP1B | 372,41 | 0,01 | 0,05 | 0,28 | 7,81E-01 | 8,71E-01 |
| FAM124B | 83,83 | 0,02 | 0,08 | 0,28 | 7,81E-01 | 8,72E-01 |
| LAMP2 | 8462,94 | 0,02 | 0,07 | 0,28 | 7,81E-01 | 8,72E-01 |
| XRRA1 | 725,48 | -0,04 | 0,13 | -0,28 | 7,81E-01 | 8,72E-01 |
| DDX54 | 558,14 | -0,02 | 0,06 | -0,28 | 7,81E-01 | 8,72E-01 |
| PDIA4 | 1041,67 | 0,01 | 0,05 | 0,28 | 7,82E-01 | 8,72E-01 |
| LENG1 | 488,60 | 0,01 | 0,04 | 0,28 | 7,83E-01 | 8,72E-01 |
| AMN1 | 570,57 | 0,01 | 0,05 | 0,28 | 7,82E-01 | 8,72E-01 |
| PBX2 | 3360,34 | -0,02 | 0,07 | -0,28 | 7,83E-01 | 8,72E-01 |
| MED11 | 431,43 | 0,01 | 0,05 | 0,28 | 7,82E-01 | 8,72E-01 |
| RTF1 | 1118,55 | 0,01 | 0,03 | 0,28 | 7,82E-01 | 8,72E-01 |
| EMC6 | 184,37 | -0,02 | 0,06 | -0,28 | 7,82E-01 | 8,72E-01 |
| ITPRIPL1 | 218,07 | -0,02 | 0,08 | -0,28 | 7,82E-01 | 8,72E-01 |
| ARRDC4 | 496,76 | 0,02 | 0,08 | 0,28 | 7,82E-01 | 8,72E-01 |
| TXNL4B | 574,55 | 0,01 | 0,04 | 0,28 | 7,82E-01 | 8,72E-01 |
| GCA | 15652,89 | 0,03 | 0,10 | 0,28 | 7,83E-01 | 8,73E-01 |
| RETREG3 | 2469,66 | -0,01 | 0,03 | -0,28 | 7,83E-01 | 8,73E-01 |
| CDK11A | 192,11 | -0,02 | 0,07 | -0,28 | 7,83E-01 | 8,73E-01 |
| FXN | 314,03 | 0,02 | 0,06 | 0,28 | 7,83E-01 | 8,73E-01 |
| MAPKAPK5 | 1449,54 | 0,01 | 0,03 | 0,27 | 7,83E-01 | 8,73E-01 |
| AMER1 | 256,85 | -0,01 | 0,05 | -0,27 | 7,84E-01 | 8,73E-01 |
| SMYD4 | 725,24 | 0,01 | 0,04 | 0,27 | 7,84E-01 | 8,73E-01 |

| | | | | | | |
|---|---|---|---|---|---|---|
| WASHC2A | 1137,06 | -0,01 | 0,04 | -0,27 | 7,84E-01 | 8,73E-01 |
| ZDHHC7 | 3605,35 | 0,01 | 0,05 | 0,27 | 7,84E-01 | 8,73E-01 |
| ACSL5 | 1636,00 | 0,01 | 0,04 | 0,27 | 7,84E-01 | 8,73E-01 |
| THBS1 | 1091,67 | 0,04 | 0,14 | 0,27 | 7,84E-01 | 8,73E-01 |
| NBR1 | 4726,13 | 0,01 | 0,04 | 0,27 | 7,85E-01 | 8,74E-01 |
| TRMT10B | 339,97 | 0,01 | 0,03 | 0,27 | 7,85E-01 | 8,74E-01 |
| TMEM14B | 778,63 | -0,01 | 0,05 | -0,27 | 7,86E-01 | 8,75E-01 |
| SARS1 | 2192,03 | 0,01 | 0,04 | 0,27 | 7,86E-01 | 8,75E-01 |
| PUS7 | 158,55 | -0,01 | 0,05 | -0,27 | 7,86E-01 | 8,75E-01 |
| SEC61A2 | 467,99 | -0,01 | 0,05 | -0,27 | 7,86E-01 | 8,75E-01 |
| TMEM258 | 652,40 | -0,02 | 0,06 | -0,27 | 7,86E-01 | 8,75E-01 |
| ZNF442 | 89,56 | -0,03 | 0,10 | -0,27 | 7,87E-01 | 8,76E-01 |
| LFNG | 3137,58 | -0,01 | 0,06 | -0,27 | 7,87E-01 | 8,76E-01 |
| NHLRC1 | 38,60 | 0,02 | 0,08 | 0,27 | 7,87E-01 | 8,76E-01 |
| EPSTI1 | 1685,60 | -0,04 | 0,15 | -0,27 | 7,87E-01 | 8,76E-01 |
| TMEM186 | 178,84 | -0,01 | 0,03 | -0,27 | 7,87E-01 | 8,76E-01 |
| TSPYL5 | 103,06 | -0,02 | 0,09 | -0,27 | 7,87E-01 | 8,76E-01 |
| ENSG000002 | 122,40 | 0,08 | 0,31 | 0,27 | 7,87E-01 | 8,76E-01 |
| CCDC127 | 488,54 | -0,01 | 0,04 | -0,27 | 7,88E-01 | 8,76E-01 |
| ZNF646 | 1519,78 | -0,02 | 0,07 | -0,27 | 7,88E-01 | 8,76E-01 |
| FTCDNL1 | 107,34 | -0,02 | 0,07 | -0,27 | 7,88E-01 | 8,76E-01 |
| ANG | 26,16 | 0,03 | 0,10 | 0,27 | 7,88E-01 | 8,76E-01 |
| ASH2L | 943,53 | -0,01 | 0,04 | -0,27 | 7,88E-01 | 8,76E-01 |
| DHX38 | 2044,92 | 0,01 | 0,04 | 0,27 | 7,88E-01 | 8,76E-01 |
| LEKR1 | 56,11 | -0,04 | 0,14 | -0,27 | 7,88E-01 | 8,76E-01 |
| RER1 | 2703,00 | -0,01 | 0,03 | -0,27 | 7,88E-01 | 8,76E-01 |
| FPGT-TNNI3K | 43,19 | -0,03 | 0,12 | -0,27 | 7,88E-01 | 8,76E-01 |
| FARS2 | 657,23 | -0,01 | 0,04 | -0,27 | 7,88E-01 | 8,76E-01 |
| NFE2L1 | 1571,31 | -0,01 | 0,04 | -0,27 | 7,88E-01 | 8,76E-01 |
| ARHGDIA | 609,37 | -0,03 | 0,10 | -0,27 | 7,89E-01 | 8,76E-01 |
| CPPED1 | 7160,69 | -0,02 | 0,07 | -0,27 | 7,89E-01 | 8,76E-01 |
| APP | 1623,83 | 0,02 | 0,06 | 0,27 | 7,89E-01 | 8,76E-01 |
| DCAF4 | 190,30 | -0,02 | 0,08 | -0,27 | 7,89E-01 | 8,76E-01 |
| ZNF608 | 156,48 | -0,03 | 0,11 | -0,27 | 7,89E-01 | 8,76E-01 |
| NEFL | 82,81 | -0,04 | 0,14 | -0,27 | 7,89E-01 | 8,76E-01 |
| NTAQ1 | 189,55 | -0,02 | 0,06 | -0,27 | 7,90E-01 | 8,77E-01 |
| FOS | 1190,49 | 0,03 | 0,10 | 0,27 | 7,90E-01 | 8,77E-01 |
| C6orf89 | 2284,43 | 0,01 | 0,04 | 0,27 | 7,90E-01 | 8,77E-01 |
| EXOG | 537,71 | 0,01 | 0,05 | 0,27 | 7,90E-01 | 8,77E-01 |
| KLF3 | 6264,39 | 0,01 | 0,04 | 0,27 | 7,90E-01 | 8,77E-01 |
| TCEANC | 592,36 | -0,01 | 0,05 | -0,27 | 7,90E-01 | 8,77E-01 |
| HDDC3 | 322,32 | -0,01 | 0,04 | -0,27 | 7,90E-01 | 8,77E-01 |
| RNF114 | 2263,10 | -0,01 | 0,04 | -0,27 | 7,90E-01 | 8,77E-01 |
| PDK4 | 884,12 | 0,03 | 0,13 | 0,27 | 7,91E-01 | 8,77E-01 |
| ZNF236 | 1409,27 | -0,01 | 0,04 | -0,27 | 7,91E-01 | 8,77E-01 |
| TCEAL3 | 100,87 | -0,02 | 0,07 | -0,26 | 7,91E-01 | 8,78E-01 |

| | | | | | | |
|---|---|---|---|---|---|---|
| ALDH6A1 | 377,42 | -0,01 | 0,05 | -0,26 | 7,92E-01 | 8,78E-01 |
| LRRC8B | 1181,86 | 0,02 | 0,06 | 0,26 | 7,92E-01 | 8,78E-01 |
| IRF2BPL | 1181,46 | 0,02 | 0,07 | 0,26 | 7,92E-01 | 8,78E-01 |
| KRI1 | 603,87 | 0,02 | 0,06 | 0,26 | 7,92E-01 | 8,78E-01 |
| GPR84 | 29,54 | -0,03 | 0,13 | -0,26 | 7,92E-01 | 8,79E-01 |
| ACTR1B | 1017,79 | 0,01 | 0,03 | 0,26 | 7,92E-01 | 8,79E-01 |
| CNR2 | 334,55 | -0,03 | 0,10 | -0,26 | 7,92E-01 | 8,79E-01 |
| PLGRKT | 277,19 | 0,01 | 0,05 | 0,26 | 7,93E-01 | 8,79E-01 |
| ZYG11B | 2249,60 | 0,01 | 0,04 | 0,26 | 7,93E-01 | 8,79E-01 |
| DRG2 | 614,13 | -0,02 | 0,06 | -0,26 | 7,93E-01 | 8,79E-01 |
| TLR2 | 11927,44 | -0,02 | 0,08 | -0,26 | 7,93E-01 | 8,79E-01 |
| CTBP2 | 2107,08 | -0,01 | 0,05 | -0,26 | 7,93E-01 | 8,79E-01 |
| HEBP1 | 196,99 | -0,03 | 0,12 | -0,26 | 7,94E-01 | 8,79E-01 |
| ZFAND2A | 328,42 | 0,01 | 0,04 | 0,26 | 7,94E-01 | 8,79E-01 |
| NPIPB11 | 36,94 | -0,04 | 0,14 | -0,26 | 7,94E-01 | 8,79E-01 |
| DDX31 | 463,15 | 0,01 | 0,05 | 0,26 | 7,94E-01 | 8,80E-01 |
| P2RY14 | 455,30 | -0,03 | 0,12 | -0,26 | 7,94E-01 | 8,80E-01 |
| TNIK | 2189,78 | 0,01 | 0,05 | 0,26 | 7,94E-01 | 8,80E-01 |
| TECPR1 | 1858,41 | -0,02 | 0,06 | -0,26 | 7,94E-01 | 8,80E-01 |
| GZMH | 1198,19 | -0,04 | 0,16 | -0,26 | 7,94E-01 | 8,80E-01 |
| OAS2 | 2052,12 | -0,03 | 0,10 | -0,26 | 7,94E-01 | 8,80E-01 |
| EARS2 | 376,30 | 0,01 | 0,04 | 0,26 | 7,94E-01 | 8,80E-01 |
| CACTIN | 439,14 | -0,02 | 0,08 | -0,26 | 7,95E-01 | 8,80E-01 |
| POLR2G | 885,77 | 0,01 | 0,04 | 0,26 | 7,95E-01 | 8,80E-01 |
| TCF7 | 2706,26 | -0,03 | 0,11 | -0,26 | 7,95E-01 | 8,80E-01 |
| THAP7 | 405,56 | 0,01 | 0,04 | 0,26 | 7,95E-01 | 8,80E-01 |
| PAFAH2 | 735,05 | -0,01 | 0,04 | -0,26 | 7,95E-01 | 8,80E-01 |
| ZNF783 | 942,00 | 0,01 | 0,04 | 0,26 | 7,95E-01 | 8,80E-01 |
| GRAMD4 | 1075,01 | -0,01 | 0,06 | -0,26 | 7,95E-01 | 8,80E-01 |
| CEMIP2 | 5740,42 | 0,01 | 0,04 | 0,26 | 7,95E-01 | 8,80E-01 |
| ARHGAP21 | 918,60 | -0,01 | 0,05 | -0,26 | 7,95E-01 | 8,80E-01 |
| GGA1 | 663,68 | -0,02 | 0,08 | -0,26 | 7,96E-01 | 8,80E-01 |
| PYM1 | 408,17 | -0,01 | 0,03 | -0,26 | 7,96E-01 | 8,81E-01 |
| ELAPOR1 | 1201,56 | 0,04 | 0,16 | 0,26 | 7,96E-01 | 8,81E-01 |
| SUPT4H1 | 120,88 | 0,02 | 0,06 | 0,26 | 7,96E-01 | 8,81E-01 |
| ENSG000002 | 24,55 | -0,04 | 0,15 | -0,26 | 7,97E-01 | 8,81E-01 |
| MCM6 | 460,19 | 0,01 | 0,05 | 0,26 | 7,97E-01 | 8,81E-01 |
| UBR2 | 13349,26 | -0,01 | 0,05 | -0,26 | 7,97E-01 | 8,81E-01 |
| AURKAIP1 | 1158,88 | -0,01 | 0,05 | -0,26 | 7,97E-01 | 8,81E-01 |
| PPM1A | 9162,77 | -0,02 | 0,09 | -0,26 | 7,97E-01 | 8,81E-01 |
| H2BC9 | 574,81 | -0,02 | 0,07 | -0,26 | 7,97E-01 | 8,81E-01 |
| EREG | 62,61 | -0,04 | 0,17 | -0,26 | 7,97E-01 | 8,81E-01 |
| ACOX1 | 5306,27 | 0,02 | 0,07 | 0,26 | 7,98E-01 | 8,81E-01 |
| MEAK7 | 412,44 | -0,01 | 0,06 | -0,26 | 7,98E-01 | 8,81E-01 |
| KCTD5 | 839,46 | -0,01 | 0,03 | -0,26 | 7,99E-01 | 8,82E-01 |
| BTBD6 | 407,69 | -0,01 | 0,04 | -0,25 | 7,99E-01 | 8,83E-01 |

| | | | | | | |
|---|---|---|---|---|---|---|
| RNF181 | 570,08 | -0,01 | 0,05 | -0,25 | 8,00E-01 | 8,83E-01 |
| ZNF185 | 1138,30 | 0,02 | 0,09 | 0,25 | 8,00E-01 | 8,84E-01 |
| CXCL1 | 97,15 | 0,02 | 0,08 | 0,25 | 8,00E-01 | 8,84E-01 |
| STK16 | 619,51 | -0,01 | 0,05 | -0,25 | 8,00E-01 | 8,84E-01 |
| FLT3 | 212,75 | 0,03 | 0,10 | 0,25 | 8,00E-01 | 8,84E-01 |
| CARHSP1 | 950,76 | -0,01 | 0,04 | -0,25 | 8,00E-01 | 8,84E-01 |
| PSMB6 | 635,10 | 0,01 | 0,05 | 0,25 | 8,00E-01 | 8,84E-01 |
| ARHGEF10L | 623,85 | -0,02 | 0,08 | -0,25 | 8,00E-01 | 8,84E-01 |
| KIF27 | 1102,66 | 0,02 | 0,08 | 0,25 | 8,01E-01 | 8,84E-01 |
| SEC62 | 4233,19 | 0,02 | 0,09 | 0,25 | 8,01E-01 | 8,84E-01 |
| PCLAF | 27,84 | -0,03 | 0,13 | -0,25 | 8,01E-01 | 8,84E-01 |
| TMEM237 | 104,74 | -0,02 | 0,09 | -0,25 | 8,01E-01 | 8,84E-01 |
| BNC2 | 207,16 | 0,04 | 0,14 | 0,25 | 8,01E-01 | 8,84E-01 |
| NDUFV3 | 1086,38 | 0,01 | 0,03 | 0,25 | 8,01E-01 | 8,84E-01 |
| ADPRH | 250,21 | 0,02 | 0,06 | 0,25 | 8,01E-01 | 8,84E-01 |
| ABCB10 | 1472,31 | -0,02 | 0,07 | -0,25 | 8,02E-01 | 8,84E-01 |
| SLC7A7 | 2382,20 | 0,01 | 0,06 | 0,25 | 8,02E-01 | 8,84E-01 |
| TAB2 | 3979,25 | 0,01 | 0,04 | 0,25 | 8,02E-01 | 8,84E-01 |
| CCDC69 | 2153,58 | -0,02 | 0,07 | -0,25 | 8,02E-01 | 8,84E-01 |
| ANKRD40 | 507,30 | -0,01 | 0,04 | -0,25 | 8,02E-01 | 8,84E-01 |
| MCM5 | 1083,12 | 0,01 | 0,05 | 0,25 | 8,02E-01 | 8,84E-01 |
| PCDHGA10 | 74,30 | -0,03 | 0,13 | -0,25 | 8,02E-01 | 8,84E-01 |
| ANO6 | 2821,30 | -0,01 | 0,05 | -0,25 | 8,03E-01 | 8,85E-01 |
| ID2 | 1089,91 | 0,01 | 0,06 | 0,25 | 8,03E-01 | 8,85E-01 |
| HYPK | 323,20 | -0,01 | 0,05 | -0,25 | 8,03E-01 | 8,85E-01 |
| ADCK2 | 291,45 | -0,01 | 0,05 | -0,25 | 8,03E-01 | 8,85E-01 |
| MGAT1 | 4506,51 | -0,01 | 0,05 | -0,25 | 8,03E-01 | 8,85E-01 |
| TM9SF2 | 3162,57 | 0,01 | 0,05 | 0,25 | 8,03E-01 | 8,85E-01 |
| MYOM2 | 392,99 | 0,06 | 0,25 | 0,25 | 8,04E-01 | 8,85E-01 |
| TIMM13 | 413,18 | 0,01 | 0,05 | 0,25 | 8,03E-01 | 8,85E-01 |
| HELB | 4313,67 | -0,01 | 0,05 | -0,25 | 8,03E-01 | 8,85E-01 |
| LY96 | 539,26 | 0,02 | 0,10 | 0,25 | 8,04E-01 | 8,86E-01 |
| H2AC11 | 1432,83 | 0,02 | 0,09 | 0,25 | 8,04E-01 | 8,86E-01 |
| GMPR2 | 1851,95 | 0,01 | 0,05 | 0,25 | 8,04E-01 | 8,86E-01 |
| KCTD20 | 3572,97 | 0,01 | 0,04 | 0,25 | 8,04E-01 | 8,86E-01 |
| CLCN4 | 403,66 | 0,03 | 0,11 | 0,25 | 8,05E-01 | 8,86E-01 |
| KBTBD7 | 1264,67 | 0,02 | 0,08 | 0,25 | 8,05E-01 | 8,86E-01 |
| SYT17 | 99,95 | -0,03 | 0,14 | -0,25 | 8,05E-01 | 8,86E-01 |
| DIDO1 | 4207,51 | -0,01 | 0,03 | -0,25 | 8,05E-01 | 8,86E-01 |
| ZNF492 | 31,13 | 0,03 | 0,14 | 0,25 | 8,05E-01 | 8,86E-01 |
| ACAD8 | 695,62 | 0,01 | 0,03 | 0,25 | 8,05E-01 | 8,86E-01 |
| ODF2 | 685,24 | -0,01 | 0,04 | -0,25 | 8,05E-01 | 8,86E-01 |
| SLC27A3 | 584,90 | -0,01 | 0,06 | -0,25 | 8,06E-01 | 8,87E-01 |
| ZNF365 | 79,18 | 0,03 | 0,12 | 0,25 | 8,06E-01 | 8,87E-01 |
| TNNT1 | 79,57 | -0,05 | 0,19 | -0,25 | 8,06E-01 | 8,87E-01 |
| EXT2 | 1048,19 | 0,01 | 0,04 | 0,25 | 8,06E-01 | 8,87E-01 |

| | | | | | | |
|---|---|---|---|---|---|---|
| CRISP3 | 109,45 | -0,05 | 0,20 | -0,24 | 8,07E-01 | 8,87E-01 |
| RRP12 | 832,49 | -0,02 | 0,10 | -0,24 | 8,07E-01 | 8,87E-01 |
| C16orf86 | 86,27 | -0,01 | 0,06 | -0,24 | 8,07E-01 | 8,87E-01 |
| SPECC1L | 321,58 | -0,01 | 0,06 | -0,24 | 8,07E-01 | 8,87E-01 |
| TET1 | 169,17 | -0,02 | 0,07 | -0,24 | 8,07E-01 | 8,87E-01 |
| CDKN2B | 145,40 | 0,02 | 0,09 | 0,24 | 8,07E-01 | 8,87E-01 |
| NUDCD3 | 2408,83 | 0,01 | 0,04 | 0,24 | 8,07E-01 | 8,88E-01 |
| ZNF892 | 132,86 | -0,01 | 0,05 | -0,24 | 8,07E-01 | 8,88E-01 |
| FGD2 | 1416,03 | -0,02 | 0,10 | -0,24 | 8,07E-01 | 8,88E-01 |
| MAN1A1 | 3673,07 | -0,01 | 0,06 | -0,24 | 8,08E-01 | 8,88E-01 |
| IL18BP | 240,48 | -0,02 | 0,10 | -0,24 | 8,08E-01 | 8,88E-01 |
| LAMTOR2 | 561,36 | 0,01 | 0,05 | 0,24 | 8,08E-01 | 8,88E-01 |
| DENND5B | 277,95 | -0,02 | 0,10 | -0,24 | 8,08E-01 | 8,88E-01 |
| SNUPN | 242,15 | 0,01 | 0,03 | 0,24 | 8,08E-01 | 8,88E-01 |
| SMCO4 | 262,58 | -0,02 | 0,08 | -0,24 | 8,08E-01 | 8,88E-01 |
| OSBPL3 | 1281,89 | -0,01 | 0,05 | -0,24 | 8,08E-01 | 8,88E-01 |
| LYST | 37166,56 | 0,02 | 0,07 | 0,24 | 8,08E-01 | 8,88E-01 |
| PGK1 | 8746,36 | 0,01 | 0,05 | 0,24 | 8,08E-01 | 8,88E-01 |
| SP100 | 8850,16 | 0,01 | 0,05 | 0,24 | 8,09E-01 | 8,88E-01 |
| MYO1G | 6079,96 | -0,01 | 0,05 | -0,24 | 8,08E-01 | 8,88E-01 |
| ELP6 | 388,38 | 0,01 | 0,03 | 0,24 | 8,09E-01 | 8,88E-01 |
| GALM | 432,19 | 0,01 | 0,06 | 0,24 | 8,09E-01 | 8,88E-01 |
| ARL4A | 978,79 | 0,03 | 0,11 | 0,24 | 8,09E-01 | 8,88E-01 |
| ITK | 6158,09 | 0,01 | 0,06 | 0,24 | 8,09E-01 | 8,88E-01 |
| PPP1R35 | 332,06 | -0,01 | 0,06 | -0,24 | 8,09E-01 | 8,88E-01 |
| APOBEC3G | 1136,25 | 0,01 | 0,06 | 0,24 | 8,09E-01 | 8,88E-01 |
| PRKCH | 6314,32 | 0,01 | 0,06 | 0,24 | 8,09E-01 | 8,88E-01 |
| USP3 | 6145,58 | -0,01 | 0,05 | -0,24 | 8,09E-01 | 8,88E-01 |
| PCSK7 | 4541,68 | -0,01 | 0,06 | -0,24 | 8,10E-01 | 8,88E-01 |
| DPY19L1 | 791,41 | 0,01 | 0,06 | 0,24 | 8,09E-01 | 8,88E-01 |
| TXNL4A | 1214,38 | 0,01 | 0,03 | 0,24 | 8,10E-01 | 8,88E-01 |
| ATP2A2 | 2539,65 | -0,01 | 0,04 | -0,24 | 8,10E-01 | 8,88E-01 |
| HBEGF | 39,99 | -0,03 | 0,14 | -0,24 | 8,09E-01 | 8,88E-01 |
| TMEM109 | 321,79 | -0,01 | 0,06 | -0,24 | 8,09E-01 | 8,88E-01 |
| RTL10 | 441,46 | -0,01 | 0,05 | -0,24 | 8,10E-01 | 8,88E-01 |
| NOP56 | 1300,38 | 0,01 | 0,04 | 0,24 | 8,10E-01 | 8,88E-01 |
| HPS6 | 442,59 | 0,01 | 0,05 | 0,24 | 8,10E-01 | 8,88E-01 |
| KIF11 | 120,70 | -0,02 | 0,09 | -0,24 | 8,10E-01 | 8,88E-01 |
| MTFMT | 487,58 | -0,01 | 0,03 | -0,24 | 8,10E-01 | 8,89E-01 |
| TMBIM4 | 1514,60 | -0,01 | 0,05 | -0,24 | 8,10E-01 | 8,89E-01 |
| ZNF697 | 305,68 | -0,02 | 0,07 | -0,24 | 8,11E-01 | 8,89E-01 |
| ARPC5 | 15975,10 | 0,02 | 0,07 | 0,24 | 8,11E-01 | 8,89E-01 |
| TRABD2A | 2405,79 | 0,02 | 0,08 | 0,24 | 8,11E-01 | 8,89E-01 |
| LPXN | 1345,29 | 0,01 | 0,03 | 0,24 | 8,11E-01 | 8,89E-01 |
| ACLY | 2147,01 | 0,01 | 0,05 | 0,24 | 8,12E-01 | 8,90E-01 |
| VMP1 | 15762,32 | 0,02 | 0,06 | 0,24 | 8,12E-01 | 8,90E-01 |

| | | | | | | |
|---|---|---|---|---|---|---|
| SPATA7 | 141,28 | -0,02 | 0,07 | -0,24 | 8,12E-01 | 8,90E-01 |
| TAS2R4 | 202,54 | -0,02 | 0,08 | -0,24 | 8,12E-01 | 8,90E-01 |
| PHPT1 | 257,05 | -0,01 | 0,05 | -0,24 | 8,13E-01 | 8,91E-01 |
| PPARD | 455,38 | -0,02 | 0,07 | -0,24 | 8,13E-01 | 8,91E-01 |
| YPEL2 | 2065,06 | 0,01 | 0,04 | 0,24 | 8,13E-01 | 8,91E-01 |
| NECTIN3 | 113,54 | -0,03 | 0,11 | -0,24 | 8,13E-01 | 8,91E-01 |
| KLHDC3 | 1156,49 | -0,01 | 0,03 | -0,24 | 8,13E-01 | 8,91E-01 |
| AMIGO1 | 313,30 | -0,02 | 0,08 | -0,24 | 8,14E-01 | 8,91E-01 |
| ELAVL1 | 670,71 | -0,01 | 0,05 | -0,24 | 8,14E-01 | 8,91E-01 |
| PLXDC2 | 4974,24 | 0,01 | 0,06 | 0,23 | 8,14E-01 | 8,91E-01 |
| ZNF512 | 689,20 | 0,01 | 0,04 | 0,24 | 8,14E-01 | 8,91E-01 |
| SPRING1 | 1165,57 | -0,01 | 0,03 | -0,23 | 8,15E-01 | 8,92E-01 |
| DOCK2 | 14613,24 | -0,01 | 0,06 | -0,23 | 8,15E-01 | 8,92E-01 |
| ARMCX6 | 298,11 | 0,01 | 0,05 | 0,23 | 8,15E-01 | 8,92E-01 |
| SUN1 | 1537,56 | 0,01 | 0,04 | 0,23 | 8,15E-01 | 8,92E-01 |
| ADAP1 | 673,09 | 0,02 | 0,07 | 0,23 | 8,16E-01 | 8,93E-01 |
| NCR3LG1 | 146,90 | 0,02 | 0,08 | 0,23 | 8,16E-01 | 8,93E-01 |
| LBHD1 | 223,75 | -0,01 | 0,06 | -0,23 | 8,16E-01 | 8,93E-01 |
| PHF20 | 2759,08 | -0,01 | 0,03 | -0,23 | 8,16E-01 | 8,93E-01 |
| TMEM223 | 234,43 | 0,01 | 0,05 | 0,23 | 8,17E-01 | 8,93E-01 |
| YIF1A | 333,54 | 0,01 | 0,04 | 0,23 | 8,17E-01 | 8,93E-01 |
| DCLRE1B | 144,69 | 0,01 | 0,04 | 0,23 | 8,17E-01 | 8,93E-01 |
| TMSB4Y | 73,99 | 0,02 | 0,10 | 0,23 | 8,18E-01 | 8,94E-01 |
| GPR160 | 700,32 | -0,02 | 0,08 | -0,23 | 8,18E-01 | 8,94E-01 |
| USF1 | 3311,90 | 0,01 | 0,05 | 0,23 | 8,18E-01 | 8,94E-01 |
| WAC | 6724,16 | 0,01 | 0,04 | 0,23 | 8,18E-01 | 8,94E-01 |
| PIWIL4 | 161,93 | -0,02 | 0,08 | -0,23 | 8,18E-01 | 8,95E-01 |
| SMYD3 | 613,78 | -0,02 | 0,07 | -0,23 | 8,18E-01 | 8,95E-01 |
| RAE1 | 721,63 | -0,01 | 0,03 | -0,23 | 8,19E-01 | 8,95E-01 |
| FRMD4B | 684,82 | 0,01 | 0,05 | 0,23 | 8,19E-01 | 8,95E-01 |
| CBX7 | 1771,47 | -0,02 | 0,08 | -0,23 | 8,19E-01 | 8,95E-01 |
| LAGE3 | 260,14 | 0,01 | 0,05 | 0,23 | 8,19E-01 | 8,95E-01 |
| CST3 | 1651,03 | 0,02 | 0,08 | 0,23 | 8,19E-01 | 8,95E-01 |
| RNASEK | 126,45 | -0,02 | 0,07 | -0,23 | 8,19E-01 | 8,95E-01 |
| GBP5 | 11049,23 | -0,03 | 0,11 | -0,23 | 8,19E-01 | 8,95E-01 |
| TOX | 214,28 | -0,02 | 0,08 | -0,23 | 8,20E-01 | 8,95E-01 |
| VPS25 | 467,55 | -0,01 | 0,04 | -0,23 | 8,20E-01 | 8,95E-01 |
| ELOVL6 | 202,13 | -0,02 | 0,08 | -0,23 | 8,20E-01 | 8,96E-01 |
| CUTA | 324,97 | -0,01 | 0,04 | -0,23 | 8,21E-01 | 8,96E-01 |
| DCBLD2 | 106,13 | -0,03 | 0,12 | -0,23 | 8,21E-01 | 8,96E-01 |
| SERPINF1 | 74,22 | 0,03 | 0,11 | 0,23 | 8,21E-01 | 8,96E-01 |
| VILL | 637,04 | -0,01 | 0,06 | -0,23 | 8,21E-01 | 8,97E-01 |
| PECR | 501,90 | -0,01 | 0,03 | -0,23 | 8,21E-01 | 8,97E-01 |
| POU2AF1 | 354,12 | -0,03 | 0,11 | -0,23 | 8,21E-01 | 8,97E-01 |
| TCTN1 | 368,31 | 0,02 | 0,07 | 0,23 | 8,22E-01 | 8,97E-01 |
| VAMP3 | 1801,63 | -0,01 | 0,05 | -0,23 | 8,22E-01 | 8,97E-01 |

| | | | | | | |
|---|---|---|---|---|---|---|
| ZNF217 | 8579,65 | -0,01 | 0,05 | -0,23 | 8,22E-01 | 8,97E-01 |
| SDF2 | 1133,26 | -0,01 | 0,05 | -0,23 | 8,22E-01 | 8,97E-01 |
| SFXN1 | 1092,15 | -0,01 | 0,04 | -0,22 | 8,22E-01 | 8,97E-01 |
| SFMBT1 | 626,23 | 0,01 | 0,06 | 0,22 | 8,22E-01 | 8,97E-01 |
| FADD | 1216,86 | -0,01 | 0,05 | -0,22 | 8,22E-01 | 8,97E-01 |
| SOX4 | 240,29 | 0,02 | 0,07 | 0,22 | 8,22E-01 | 8,97E-01 |
| RPL8 | 10450,38 | 0,01 | 0,07 | 0,22 | 8,22E-01 | 8,97E-01 |
| ALDH18A1 | 442,56 | -0,01 | 0,05 | -0,22 | 8,23E-01 | 8,97E-01 |
| MYB | 257,81 | -0,02 | 0,08 | -0,22 | 8,23E-01 | 8,97E-01 |
| CEP295NL | 403,01 | -0,03 | 0,14 | -0,22 | 8,23E-01 | 8,97E-01 |
| TBC1D8B | 89,01 | -0,02 | 0,09 | -0,22 | 8,23E-01 | 8,97E-01 |
| PKP4 | 613,96 | -0,01 | 0,06 | -0,22 | 8,23E-01 | 8,97E-01 |
| DGUOK | 472,20 | -0,01 | 0,04 | -0,22 | 8,24E-01 | 8,98E-01 |
| PEA15 | 530,09 | -0,01 | 0,05 | -0,22 | 8,24E-01 | 8,98E-01 |
| SYNE2 | 22237,38 | 0,01 | 0,06 | 0,22 | 8,24E-01 | 8,98E-01 |
| PHF23 | 946,59 | -0,01 | 0,04 | -0,22 | 8,24E-01 | 8,98E-01 |
| ATP23 | 148,45 | 0,02 | 0,08 | 0,22 | 8,24E-01 | 8,98E-01 |
| H3C6 | 160,43 | 0,02 | 0,09 | 0,22 | 8,25E-01 | 8,98E-01 |
| UBE4A | 3664,37 | -0,01 | 0,03 | -0,22 | 8,25E-01 | 8,99E-01 |
| PSMB10 | 1533,37 | -0,01 | 0,06 | -0,22 | 8,25E-01 | 8,99E-01 |
| KCTD17 | 150,79 | -0,01 | 0,06 | -0,22 | 8,25E-01 | 8,99E-01 |
| SMARCC1 | 1900,27 | 0,01 | 0,04 | 0,22 | 8,25E-01 | 8,99E-01 |
| HSPA5 | 2361,64 | -0,01 | 0,05 | -0,22 | 8,25E-01 | 8,99E-01 |
| WBP1 | 325,74 | 0,01 | 0,04 | 0,22 | 8,25E-01 | 8,99E-01 |
| TRIM21 | 1735,82 | -0,01 | 0,06 | -0,22 | 8,25E-01 | 8,99E-01 |
| ETHE1 | 195,09 | -0,01 | 0,04 | -0,22 | 8,26E-01 | 8,99E-01 |
| VIM | 18545,21 | 0,01 | 0,06 | 0,22 | 8,26E-01 | 8,99E-01 |
| DUSP28 | 286,56 | 0,01 | 0,04 | 0,22 | 8,26E-01 | 8,99E-01 |
| PLEKHF1 | 492,24 | -0,02 | 0,07 | -0,22 | 8,26E-01 | 8,99E-01 |
| DELE1 | 1112,37 | -0,01 | 0,06 | -0,22 | 8,26E-01 | 8,99E-01 |
| PKIG | 178,06 | 0,02 | 0,10 | 0,22 | 8,27E-01 | 9,00E-01 |
| FYCO1 | 1322,69 | -0,01 | 0,06 | -0,22 | 8,27E-01 | 9,00E-01 |
| TNRC6A | 2935,06 | 0,01 | 0,05 | 0,22 | 8,27E-01 | 9,00E-01 |
| NMRK1 | 933,74 | 0,01 | 0,07 | 0,22 | 8,27E-01 | 9,00E-01 |
| NBPF9 | 354,35 | -0,01 | 0,07 | -0,22 | 8,27E-01 | 9,00E-01 |
| CDH23 | 1983,19 | 0,01 | 0,07 | 0,22 | 8,27E-01 | 9,00E-01 |
| CHMP7 | 3025,55 | 0,01 | 0,05 | 0,22 | 8,27E-01 | 9,00E-01 |
| PTPRM | 415,41 | 0,03 | 0,12 | 0,22 | 8,28E-01 | 9,00E-01 |
| CNOT9 | 1301,46 | 0,01 | 0,03 | 0,22 | 8,28E-01 | 9,00E-01 |
| METTL1 | 45,55 | -0,02 | 0,08 | -0,22 | 8,28E-01 | 9,00E-01 |
| RBM22 | 848,74 | -0,01 | 0,03 | -0,22 | 8,28E-01 | 9,00E-01 |
| PLEKHA5 | 271,70 | 0,02 | 0,11 | 0,22 | 8,28E-01 | 9,01E-01 |
| PI16 | 119,99 | -0,02 | 0,09 | -0,22 | 8,29E-01 | 9,01E-01 |
| CLCN3 | 2570,29 | 0,01 | 0,04 | 0,22 | 8,29E-01 | 9,01E-01 |
| TMTC2 | 900,15 | 0,01 | 0,05 | 0,22 | 8,29E-01 | 9,01E-01 |
| ST6GAL1 | 3627,21 | 0,01 | 0,06 | 0,22 | 8,29E-01 | 9,01E-01 |

| | | | | | | |
|---|---|---|---|---|---|---|
| SMIM8 | 737,59 | 0,01 | 0,05 | 0,22 | 8,29E-01 | 9,01E-01 |
| GNPTG | 805,63 | -0,01 | 0,04 | -0,22 | 8,29E-01 | 9,01E-01 |
| CPLANE2 | 60,26 | -0,02 | 0,07 | -0,22 | 8,29E-01 | 9,01E-01 |
| ANKRD17 | 3632,13 | -0,01 | 0,04 | -0,22 | 8,30E-01 | 9,01E-01 |
| CD38 | 663,60 | -0,02 | 0,08 | -0,21 | 8,30E-01 | 9,02E-01 |
| ARL17B | 108,50 | 0,03 | 0,14 | 0,21 | 8,30E-01 | 9,02E-01 |
| OGFR | 3025,01 | -0,01 | 0,06 | -0,21 | 8,30E-01 | 9,02E-01 |
| ALG14 | 106,50 | -0,02 | 0,07 | -0,21 | 8,30E-01 | 9,02E-01 |
| POLRMT | 524,27 | -0,01 | 0,06 | -0,21 | 8,31E-01 | 9,02E-01 |
| PRR4 | 25,28 | -0,02 | 0,11 | -0,21 | 8,31E-01 | 9,02E-01 |
| SWSAP1 | 97,97 | -0,01 | 0,06 | -0,21 | 8,31E-01 | 9,02E-01 |
| DHODH | 246,20 | -0,01 | 0,06 | -0,21 | 8,31E-01 | 9,03E-01 |
| ITGB8 | 162,26 | -0,04 | 0,20 | -0,21 | 8,31E-01 | 9,03E-01 |
| PCDHGA11 | 65,36 | -0,03 | 0,13 | -0,21 | 8,32E-01 | 9,03E-01 |
| BUD13 | 465,23 | 0,01 | 0,03 | 0,21 | 8,32E-01 | 9,03E-01 |
| LINGO2 | 49,81 | -0,03 | 0,16 | -0,21 | 8,33E-01 | 9,03E-01 |
| DHDDS | 434,36 | 0,01 | 0,03 | 0,21 | 8,32E-01 | 9,03E-01 |
| CFP | 4050,99 | 0,01 | 0,07 | 0,21 | 8,33E-01 | 9,03E-01 |
| DCAF6 | 1895,09 | -0,01 | 0,05 | -0,21 | 8,33E-01 | 9,03E-01 |
| TOMM34 | 217,27 | -0,01 | 0,05 | -0,21 | 8,33E-01 | 9,03E-01 |
| RBM28 | 1034,57 | -0,01 | 0,06 | -0,21 | 8,33E-01 | 9,03E-01 |
| MDGA1 | 252,82 | -0,05 | 0,26 | -0,21 | 8,33E-01 | 9,03E-01 |
| PPP2R5C | 8285,10 | 0,01 | 0,04 | 0,21 | 8,33E-01 | 9,03E-01 |
| CCDC22 | 384,31 | -0,01 | 0,04 | -0,21 | 8,33E-01 | 9,03E-01 |
| UQCC1 | 685,16 | -0,01 | 0,04 | -0,21 | 8,32E-01 | 9,03E-01 |
| BAP1 | 1250,17 | 0,01 | 0,04 | 0,21 | 8,33E-01 | 9,03E-01 |
| ASNS | 66,79 | -0,02 | 0,08 | -0,21 | 8,33E-01 | 9,03E-01 |
| LXN | 175,30 | -0,02 | 0,08 | -0,21 | 8,33E-01 | 9,03E-01 |
| CES2 | 489,80 | -0,01 | 0,05 | -0,21 | 8,33E-01 | 9,03E-01 |
| NAGLU | 246,98 | 0,01 | 0,06 | 0,21 | 8,32E-01 | 9,03E-01 |
| BUD23 | 471,58 | 0,01 | 0,04 | 0,21 | 8,32E-01 | 9,03E-01 |
| ZRANB1 | 2403,35 | -0,01 | 0,06 | -0,21 | 8,32E-01 | 9,03E-01 |
| ARHGAP18 | 844,12 | -0,01 | 0,07 | -0,21 | 8,33E-01 | 9,03E-01 |
| NISCH | 1611,13 | -0,01 | 0,06 | -0,21 | 8,32E-01 | 9,03E-01 |
| FAM151B | 204,77 | -0,01 | 0,07 | -0,21 | 8,33E-01 | 9,03E-01 |
| RTN1 | 555,47 | -0,02 | 0,08 | -0,21 | 8,34E-01 | 9,03E-01 |
| RUBCNL | 2139,54 | 0,02 | 0,09 | 0,21 | 8,34E-01 | 9,03E-01 |
| BATF3 | 73,39 | -0,02 | 0,09 | -0,21 | 8,34E-01 | 9,03E-01 |
| HAUS5 | 152,93 | -0,02 | 0,08 | -0,21 | 8,34E-01 | 9,03E-01 |
| RTF2 | 3929,77 | 0,01 | 0,06 | 0,21 | 8,34E-01 | 9,03E-01 |
| SAFB | 2557,10 | -0,01 | 0,04 | -0,21 | 8,34E-01 | 9,04E-01 |
| ARPC1A | 264,12 | -0,01 | 0,04 | -0,21 | 8,35E-01 | 9,04E-01 |
| COX5B | 1767,06 | 0,01 | 0,06 | 0,21 | 8,35E-01 | 9,04E-01 |
| MS4A4A | 157,29 | 0,02 | 0,10 | 0,21 | 8,35E-01 | 9,04E-01 |
| SSR4 | 834,19 | 0,01 | 0,06 | 0,21 | 8,35E-01 | 9,04E-01 |
| ZDHHC13 | 947,37 | 0,01 | 0,04 | 0,21 | 8,35E-01 | 9,04E-01 |

| | | | | | | |
|---|---|---|---|---|---|---|
| NAA38 | 513,81 | 0,01 | 0,06 | 0,21 | 8,36E-01 | 9,05E-01 |
| TFPT | 234,47 | 0,01 | 0,04 | 0,21 | 8,36E-01 | 9,05E-01 |
| NAGA | 969,86 | 0,01 | 0,05 | 0,21 | 8,36E-01 | 9,05E-01 |
| INAFM2 | 1195,93 | 0,01 | 0,06 | 0,21 | 8,36E-01 | 9,05E-01 |
| ZNF689 | 329,08 | 0,01 | 0,03 | 0,21 | 8,36E-01 | 9,05E-01 |
| SLC9B2 | 277,27 | 0,01 | 0,05 | 0,21 | 8,37E-01 | 9,06E-01 |
| ERCC3 | 1126,49 | -0,01 | 0,03 | -0,21 | 8,37E-01 | 9,06E-01 |
| ACSS3 | 721,72 | -0,02 | 0,09 | -0,21 | 8,37E-01 | 9,06E-01 |
| CIDEB | 608,39 | 0,01 | 0,06 | 0,21 | 8,37E-01 | 9,06E-01 |
| DEGS1 | 1467,19 | -0,01 | 0,04 | -0,20 | 8,38E-01 | 9,06E-01 |
| GPR55 | 121,13 | -0,02 | 0,10 | -0,20 | 8,38E-01 | 9,06E-01 |
| IKBIP | 1219,48 | 0,02 | 0,08 | 0,20 | 8,38E-01 | 9,06E-01 |
| HEMK1 | 1189,90 | 0,01 | 0,06 | 0,20 | 8,38E-01 | 9,06E-01 |
| LUC7L | 1366,00 | -0,01 | 0,05 | -0,20 | 8,38E-01 | 9,06E-01 |
| IFTAP | 35,12 | 0,02 | 0,12 | 0,20 | 8,38E-01 | 9,06E-01 |
| SLC38A11 | 161,40 | -0,03 | 0,15 | -0,20 | 8,38E-01 | 9,06E-01 |
| DNAJB1 | 1348,86 | 0,01 | 0,04 | 0,20 | 8,38E-01 | 9,06E-01 |
| ARFIP2 | 432,07 | 0,01 | 0,03 | 0,20 | 8,38E-01 | 9,06E-01 |
| PPM1G | 1228,28 | 0,01 | 0,05 | 0,20 | 8,39E-01 | 9,06E-01 |
| ARFRP1 | 1378,15 | 0,01 | 0,05 | 0,20 | 8,39E-01 | 9,07E-01 |
| FLYWCH1 | 582,85 | 0,02 | 0,09 | 0,20 | 8,40E-01 | 9,07E-01 |
| GALT | 827,38 | -0,01 | 0,05 | -0,20 | 8,40E-01 | 9,07E-01 |
| ELL | 1432,00 | -0,02 | 0,08 | -0,20 | 8,40E-01 | 9,07E-01 |
| CPT2 | 632,99 | 0,01 | 0,03 | 0,20 | 8,40E-01 | 9,07E-01 |
| STAT1 | 12095,73 | 0,02 | 0,09 | 0,20 | 8,40E-01 | 9,07E-01 |
| CLDN12 | 72,05 | 0,02 | 0,08 | 0,20 | 8,40E-01 | 9,08E-01 |
| ENDOG | 116,54 | -0,01 | 0,06 | -0,20 | 8,41E-01 | 9,08E-01 |
| ARL6IP4 | 52,95 | 0,01 | 0,06 | 0,20 | 8,41E-01 | 9,08E-01 |
| LRIG2 | 1277,34 | 0,01 | 0,04 | 0,20 | 8,41E-01 | 9,08E-01 |
| MAFK | 1225,44 | 0,01 | 0,05 | 0,20 | 8,41E-01 | 9,08E-01 |
| RASGRF2 | 1213,56 | -0,01 | 0,07 | -0,20 | 8,41E-01 | 9,08E-01 |
| PIGP | 242,88 | -0,01 | 0,06 | -0,20 | 8,41E-01 | 9,08E-01 |
| S100A10 | 751,03 | -0,01 | 0,05 | -0,20 | 8,41E-01 | 9,08E-01 |
| NBPF15 | 1240,34 | -0,01 | 0,06 | -0,20 | 8,42E-01 | 9,08E-01 |
| SLC22A1 | 304,92 | -0,02 | 0,11 | -0,20 | 8,42E-01 | 9,08E-01 |
| TCTN3 | 228,50 | 0,01 | 0,05 | 0,20 | 8,42E-01 | 9,08E-01 |
| ATP6V1A | 4965,86 | 0,01 | 0,06 | 0,20 | 8,42E-01 | 9,08E-01 |
| APBB1 | 489,10 | 0,01 | 0,07 | 0,20 | 8,42E-01 | 9,08E-01 |
| TNFAIP6 | 620,12 | -0,03 | 0,15 | -0,20 | 8,42E-01 | 9,08E-01 |
| RPUSD3 | 480,17 | -0,01 | 0,04 | -0,20 | 8,42E-01 | 9,08E-01 |
| EIF2AK4 | 907,35 | -0,01 | 0,04 | -0,20 | 8,42E-01 | 9,08E-01 |
| NCAPH2 | 580,60 | 0,01 | 0,04 | 0,20 | 8,42E-01 | 9,08E-01 |
| RPS6KA5 | 5370,79 | 0,01 | 0,05 | 0,20 | 8,43E-01 | 9,09E-01 |
| EIF2B4 | 392,40 | -0,01 | 0,03 | -0,20 | 8,43E-01 | 9,09E-01 |
| EDAR | 162,44 | -0,02 | 0,12 | -0,20 | 8,43E-01 | 9,09E-01 |
| HMBOX1 | 1851,47 | -0,01 | 0,05 | -0,20 | 8,43E-01 | 9,09E-01 |

| | | | | | | |
|---|---|---|---|---|---|---|
| CCZ1 | 540,34 | -0,01 | 0,04 | -0,20 | 8,43E-01 | 9,09E-01 |
| BPHL | 181,72 | 0,01 | 0,06 | 0,20 | 8,43E-01 | 9,09E-01 |
| TDRKH | 207,66 | -0,01 | 0,06 | -0,20 | 8,43E-01 | 9,09E-01 |
| ZNF552 | 733,08 | 0,01 | 0,05 | 0,20 | 8,44E-01 | 9,09E-01 |
| ZNF513 | 316,97 | -0,01 | 0,03 | -0,20 | 8,44E-01 | 9,09E-01 |
| CATSPER1 | 98,16 | 0,01 | 0,07 | 0,20 | 8,44E-01 | 9,09E-01 |
| ZNF786 | 343,71 | 0,01 | 0,04 | 0,20 | 8,44E-01 | 9,09E-01 |
| SYT11 | 660,04 | 0,01 | 0,06 | 0,20 | 8,44E-01 | 9,09E-01 |
| RNF130 | 5724,41 | -0,01 | 0,06 | -0,20 | 8,44E-01 | 9,09E-01 |
| TBCEL | 1289,79 | 0,02 | 0,08 | 0,20 | 8,44E-01 | 9,09E-01 |
| LRRC28 | 540,96 | -0,01 | 0,05 | -0,20 | 8,45E-01 | 9,10E-01 |
| PASK | 1131,80 | -0,01 | 0,07 | -0,20 | 8,45E-01 | 9,10E-01 |
| TOP6BL | 254,46 | -0,01 | 0,07 | -0,20 | 8,45E-01 | 9,10E-01 |
| FCRL6 | 1038,95 | 0,02 | 0,12 | 0,20 | 8,45E-01 | 9,10E-01 |
| PRDM2 | 3392,17 | 0,01 | 0,04 | 0,19 | 8,46E-01 | 9,11E-01 |
| MPZL3 | 4456,21 | -0,01 | 0,08 | -0,19 | 8,46E-01 | 9,11E-01 |
| PABPN1 | 1252,26 | 0,01 | 0,04 | 0,19 | 8,46E-01 | 9,11E-01 |
| PIGG | 1954,49 | -0,01 | 0,04 | -0,19 | 8,46E-01 | 9,11E-01 |
| DTD1 | 363,20 | 0,01 | 0,04 | 0,19 | 8,47E-01 | 9,11E-01 |
| SNAP47 | 474,49 | 0,01 | 0,03 | 0,19 | 8,47E-01 | 9,11E-01 |
| TEX2 | 577,78 | -0,01 | 0,04 | -0,19 | 8,47E-01 | 9,11E-01 |
| POU2F1 | 2454,44 | 0,01 | 0,05 | 0,19 | 8,47E-01 | 9,11E-01 |
| HLA-DQB2 | 65,77 | 0,05 | 0,27 | 0,19 | 8,47E-01 | 9,11E-01 |
| NBPF11 | 703,80 | 0,01 | 0,05 | 0,19 | 8,47E-01 | 9,12E-01 |
| NEU3 | 650,05 | 0,01 | 0,05 | 0,19 | 8,47E-01 | 9,12E-01 |
| TK2 | 926,07 | -0,01 | 0,06 | -0,19 | 8,48E-01 | 9,12E-01 |
| ACAT1 | 320,06 | 0,01 | 0,07 | 0,19 | 8,48E-01 | 9,13E-01 |
| PSME1 | 4614,31 | 0,01 | 0,06 | 0,19 | 8,48E-01 | 9,13E-01 |
| DCAF5 | 1738,50 | -0,01 | 0,03 | -0,19 | 8,49E-01 | 9,13E-01 |
| APPL2 | 1646,07 | -0,01 | 0,04 | -0,19 | 8,49E-01 | 9,13E-01 |
| NDUFB10 | 543,48 | 0,01 | 0,05 | 0,19 | 8,49E-01 | 9,13E-01 |
| MRM1 | 162,72 | -0,01 | 0,06 | -0,19 | 8,50E-01 | 9,13E-01 |
| EIF4A1 | 566,36 | -0,01 | 0,05 | -0,19 | 8,49E-01 | 9,13E-01 |
| POLR3H | 418,30 | 0,01 | 0,07 | 0,19 | 8,50E-01 | 9,13E-01 |
| CD99L2 | 488,17 | -0,01 | 0,04 | -0,19 | 8,49E-01 | 9,13E-01 |
| SOWAHC | 113,84 | 0,02 | 0,09 | 0,19 | 8,49E-01 | 9,13E-01 |
| PIK3CB | 1974,91 | -0,01 | 0,05 | -0,19 | 8,50E-01 | 9,13E-01 |
| FAM241A | 429,08 | 0,01 | 0,05 | 0,19 | 8,50E-01 | 9,13E-01 |
| ADGRA3 | 132,20 | -0,02 | 0,12 | -0,19 | 8,50E-01 | 9,14E-01 |
| TUFM | 1420,29 | -0,01 | 0,05 | -0,19 | 8,50E-01 | 9,14E-01 |
| SNX12 | 609,17 | -0,01 | 0,03 | -0,19 | 8,50E-01 | 9,14E-01 |
| TXLNA | 1026,04 | -0,01 | 0,05 | -0,19 | 8,51E-01 | 9,14E-01 |
| DDX39A | 760,88 | -0,01 | 0,04 | -0,19 | 8,51E-01 | 9,14E-01 |
| MPZ | 229,48 | -0,01 | 0,06 | -0,19 | 8,51E-01 | 9,14E-01 |
| TCF25 | 2395,11 | -0,01 | 0,05 | -0,19 | 8,51E-01 | 9,14E-01 |
| ARFIP1 | 1881,23 | 0,01 | 0,07 | 0,19 | 8,51E-01 | 9,14E-01 |

| | | | | | | |
|---|---|---|---|---|---|---|
| SLC25A51 | 644,54 | 0,01 | 0,03 | 0,19 | 8,51E-01 | 9,14E-01 |
| NSUN4 | 439,14 | -0,01 | 0,05 | -0,19 | 8,52E-01 | 9,15E-01 |
| MAD1L1 | 1377,34 | 0,01 | 0,06 | 0,19 | 8,52E-01 | 9,15E-01 |
| MPND | 221,58 | 0,01 | 0,05 | 0,19 | 8,53E-01 | 9,15E-01 |
| SKAP1 | 800,51 | 0,01 | 0,07 | 0,19 | 8,53E-01 | 9,15E-01 |
| EEF2 | 24434,68 | -0,01 | 0,05 | -0,19 | 8,53E-01 | 9,16E-01 |
| TUBA1A | 4014,28 | 0,01 | 0,07 | 0,19 | 8,53E-01 | 9,16E-01 |
| WDFY2 | 4397,09 | -0,01 | 0,05 | -0,18 | 8,54E-01 | 9,16E-01 |
| PHLDB2 | 366,51 | 0,02 | 0,08 | 0,18 | 8,54E-01 | 9,17E-01 |
| SPN | 2611,58 | 0,01 | 0,08 | 0,18 | 8,55E-01 | 9,17E-01 |
| ST8SIA6 | 147,29 | -0,01 | 0,08 | -0,18 | 8,55E-01 | 9,17E-01 |
| NDUFS2 | 1222,35 | 0,01 | 0,03 | 0,18 | 8,55E-01 | 9,17E-01 |
| POMGNT1 | 414,91 | 0,01 | 0,05 | 0,18 | 8,55E-01 | 9,17E-01 |
| PARP10 | 1914,11 | -0,01 | 0,07 | -0,18 | 8,55E-01 | 9,17E-01 |
| NQO1 | 41,03 | -0,02 | 0,09 | -0,18 | 8,55E-01 | 9,17E-01 |
| ZNF391 | 42,94 | 0,02 | 0,12 | 0,18 | 8,55E-01 | 9,17E-01 |
| NGRN | 752,46 | -0,01 | 0,03 | -0,18 | 8,55E-01 | 9,17E-01 |
| NBPF12 | 1198,80 | -0,01 | 0,06 | -0,18 | 8,55E-01 | 9,17E-01 |
| POLR2C | 812,07 | -0,01 | 0,03 | -0,18 | 8,56E-01 | 9,17E-01 |
| RHBDD1 | 1603,01 | 0,01 | 0,06 | 0,18 | 8,55E-01 | 9,17E-01 |
| PIK3AP1 | 6542,17 | 0,01 | 0,06 | 0,18 | 8,56E-01 | 9,17E-01 |
| NIPSNAP1 | 194,81 | -0,01 | 0,06 | -0,18 | 8,56E-01 | 9,17E-01 |
| SIRT4 | 36,98 | 0,01 | 0,07 | 0,18 | 8,56E-01 | 9,17E-01 |
| PFKP | 1030,76 | -0,01 | 0,06 | -0,18 | 8,56E-01 | 9,17E-01 |
| UIMC1 | 2582,19 | 0,01 | 0,04 | 0,18 | 8,56E-01 | 9,17E-01 |
| TRIM69 | 1614,30 | 0,01 | 0,06 | 0,18 | 8,56E-01 | 9,17E-01 |
| ZFYVE9 | 251,98 | 0,01 | 0,07 | 0,18 | 8,56E-01 | 9,17E-01 |
| STK3 | 606,98 | 0,01 | 0,06 | 0,18 | 8,56E-01 | 9,17E-01 |
| DRAP1 | 349,66 | -0,01 | 0,07 | -0,18 | 8,56E-01 | 9,17E-01 |
| GCLM | 683,84 | 0,01 | 0,06 | 0,18 | 8,56E-01 | 9,17E-01 |
| GOLGA8A | 1205,82 | 0,01 | 0,08 | 0,18 | 8,57E-01 | 9,17E-01 |
| MPZL2 | 358,11 | 0,02 | 0,09 | 0,18 | 8,57E-01 | 9,17E-01 |
| GSTM1 | 76,56 | -0,11 | 0,60 | -0,18 | 8,57E-01 | 9,17E-01 |
| L2HGDH | 217,43 | 0,01 | 0,05 | 0,18 | 8,57E-01 | 9,17E-01 |
| ZNF793 | 281,21 | 0,01 | 0,08 | 0,18 | 8,57E-01 | 9,17E-01 |
| CLN8 | 726,85 | 0,01 | 0,05 | 0,18 | 8,57E-01 | 9,18E-01 |
| EIF3B | 1835,78 | -0,01 | 0,06 | -0,18 | 8,58E-01 | 9,18E-01 |
| TRIM2 | 188,43 | 0,02 | 0,09 | 0,18 | 8,58E-01 | 9,19E-01 |
| RBM44 | 188,25 | -0,01 | 0,07 | -0,18 | 8,58E-01 | 9,19E-01 |
| PCYOX1L | 480,23 | 0,01 | 0,05 | 0,18 | 8,58E-01 | 9,19E-01 |
| PMVK | 233,59 | -0,01 | 0,04 | -0,18 | 8,58E-01 | 9,19E-01 |
| C12orf42 | 237,10 | 0,01 | 0,08 | 0,18 | 8,58E-01 | 9,19E-01 |
| IGFBP3 | 164,21 | -0,02 | 0,11 | -0,18 | 8,59E-01 | 9,19E-01 |
| NELFB | 909,39 | 0,01 | 0,04 | 0,18 | 8,59E-01 | 9,19E-01 |
| POC1B | 1241,34 | 0,01 | 0,06 | 0,18 | 8,59E-01 | 9,19E-01 |
| DDO | 41,19 | 0,02 | 0,12 | 0,18 | 8,59E-01 | 9,19E-01 |

| | | | | | | |
|---|---|---|---|---|---|---|
| FLI1 | 6759,60 | 0,01 | 0,04 | 0,18 | 8,59E-01 | 9,19E-01 |
| TCAF1 | 241,63 | -0,01 | 0,05 | -0,18 | 8,59E-01 | 9,19E-01 |
| TSPAN4 | 181,87 | 0,02 | 0,09 | 0,18 | 8,60E-01 | 9,20E-01 |
| LIME1 | 126,63 | -0,01 | 0,06 | -0,18 | 8,60E-01 | 9,20E-01 |
| UBTF | 1902,47 | 0,01 | 0,04 | 0,18 | 8,60E-01 | 9,20E-01 |
| METAP1D | 143,15 | -0,01 | 0,07 | -0,18 | 8,60E-01 | 9,20E-01 |
| C1orf35 | 359,57 | -0,01 | 0,04 | -0,18 | 8,60E-01 | 9,20E-01 |
| TTC7A | 1766,92 | 0,01 | 0,06 | 0,18 | 8,61E-01 | 9,20E-01 |
| MRPL28 | 580,03 | -0,01 | 0,05 | -0,18 | 8,61E-01 | 9,20E-01 |
| PLCG1 | 2775,13 | -0,01 | 0,06 | -0,17 | 8,61E-01 | 9,21E-01 |
| SUDS3 | 2708,22 | -0,01 | 0,04 | -0,17 | 8,62E-01 | 9,21E-01 |
| GRWD1 | 306,46 | 0,01 | 0,05 | 0,17 | 8,61E-01 | 9,21E-01 |
| PSRC1 | 114,49 | -0,01 | 0,07 | -0,17 | 8,62E-01 | 9,21E-01 |
| CD74 | 4487,88 | 0,02 | 0,11 | 0,17 | 8,63E-01 | 9,21E-01 |
| KLHL2 | 2932,33 | -0,01 | 0,07 | -0,17 | 8,62E-01 | 9,21E-01 |
| KDM6A | 2840,86 | 0,01 | 0,04 | 0,17 | 8,63E-01 | 9,21E-01 |
| GTDC1 | 1398,22 | 0,01 | 0,05 | 0,17 | 8,63E-01 | 9,21E-01 |
| SLC31A1 | 881,85 | 0,01 | 0,06 | 0,17 | 8,63E-01 | 9,21E-01 |
| LDLRAD4 | 1673,16 | 0,01 | 0,05 | 0,17 | 8,63E-01 | 9,22E-01 |
| DDIAS | 158,62 | -0,01 | 0,07 | -0,17 | 8,63E-01 | 9,22E-01 |
| MRPL49 | 665,36 | 0,00 | 0,03 | 0,17 | 8,63E-01 | 9,22E-01 |
| MAP4 | 1151,55 | -0,01 | 0,07 | -0,17 | 8,64E-01 | 9,22E-01 |
| ARHGAP45 | 9342,88 | -0,01 | 0,07 | -0,17 | 8,64E-01 | 9,22E-01 |
| DOK2 | 1343,27 | -0,01 | 0,05 | -0,17 | 8,64E-01 | 9,22E-01 |
| DYNC2I1 | 486,95 | 0,01 | 0,05 | 0,17 | 8,64E-01 | 9,22E-01 |
| CNP | 417,48 | -0,01 | 0,06 | -0,17 | 8,64E-01 | 9,22E-01 |
| ARRDC5 | 185,72 | -0,02 | 0,10 | -0,17 | 8,64E-01 | 9,22E-01 |
| VPS33A | 696,08 | 0,01 | 0,03 | 0,17 | 8,64E-01 | 9,22E-01 |
| VNN3P | 3046,10 | 0,02 | 0,09 | 0,17 | 8,64E-01 | 9,22E-01 |
| CCDC102B | 124,87 | -0,01 | 0,07 | -0,17 | 8,64E-01 | 9,22E-01 |
| SPATA25 | 25,36 | -0,01 | 0,09 | -0,17 | 8,65E-01 | 9,22E-01 |
| RBP7 | 149,48 | -0,02 | 0,09 | -0,17 | 8,65E-01 | 9,22E-01 |
| LRRC37A2 | 499,58 | 0,01 | 0,08 | 0,17 | 8,65E-01 | 9,23E-01 |
| CPA3 | 126,51 | -0,03 | 0,21 | -0,17 | 8,65E-01 | 9,23E-01 |
| NUP62 | 627,80 | -0,01 | 0,07 | -0,17 | 8,65E-01 | 9,23E-01 |
| AKIRIN1 | 2571,16 | 0,01 | 0,04 | 0,17 | 8,65E-01 | 9,23E-01 |
| MRPS34 | 429,59 | 0,01 | 0,04 | 0,17 | 8,65E-01 | 9,23E-01 |
| IGSF6 | 7313,93 | 0,01 | 0,07 | 0,17 | 8,66E-01 | 9,23E-01 |
| FAM120C | 300,87 | -0,01 | 0,04 | -0,17 | 8,66E-01 | 9,23E-01 |
| SSU72 | 2101,08 | -0,01 | 0,04 | -0,17 | 8,66E-01 | 9,23E-01 |
| DMAP1 | 877,58 | -0,01 | 0,04 | -0,17 | 8,66E-01 | 9,23E-01 |
| ZCWPW1 | 189,44 | -0,01 | 0,04 | -0,17 | 8,66E-01 | 9,23E-01 |
| MFN2 | 3156,06 | 0,01 | 0,07 | 0,17 | 8,66E-01 | 9,23E-01 |
| RFC2 | 422,20 | -0,01 | 0,04 | -0,17 | 8,67E-01 | 9,24E-01 |
| PITPNC1 | 1694,86 | -0,01 | 0,03 | -0,17 | 8,67E-01 | 9,24E-01 |
| CLCN6 | 555,44 | -0,01 | 0,07 | -0,17 | 8,67E-01 | 9,24E-01 |

| | | | | | | |
|---|---|---|---|---|---|---|
| ISOC1 | 350,04 | 0,01 | 0,06 | 0,17 | 8,67E-01 | 9,24E-01 |
| FAAP24 | 86,20 | -0,01 | 0,08 | -0,17 | 8,67E-01 | 9,24E-01 |
| PDK1 | 3625,30 | 0,01 | 0,05 | 0,17 | 8,68E-01 | 9,24E-01 |
| ZBTB48 | 672,26 | -0,01 | 0,05 | -0,17 | 8,68E-01 | 9,24E-01 |
| CD9 | 686,43 | 0,02 | 0,12 | 0,17 | 8,68E-01 | 9,24E-01 |
| STK32C | 350,74 | -0,01 | 0,05 | -0,17 | 8,68E-01 | 9,24E-01 |
| ANKRD34B | 72,42 | -0,03 | 0,17 | -0,17 | 8,68E-01 | 9,24E-01 |
| TMEM248 | 2388,83 | 0,00 | 0,03 | -0,17 | 8,68E-01 | 9,24E-01 |
| SRPK2 | 4636,67 | -0,01 | 0,05 | -0,17 | 8,68E-01 | 9,24E-01 |
| STAT5A | 2713,42 | -0,01 | 0,06 | -0,17 | 8,68E-01 | 9,24E-01 |
| ZNF827 | 294,29 | -0,01 | 0,08 | -0,17 | 8,69E-01 | 9,24E-01 |
| ZNF524 | 306,82 | -0,01 | 0,05 | -0,17 | 8,69E-01 | 9,25E-01 |
| LEF1 | 3193,85 | 0,01 | 0,08 | 0,17 | 8,69E-01 | 9,25E-01 |
| DROSHA | 923,18 | 0,01 | 0,05 | 0,16 | 8,70E-01 | 9,25E-01 |
| ENSG000002 | 37,70 | 0,01 | 0,07 | 0,16 | 8,70E-01 | 9,26E-01 |
| FADS2 | 307,68 | -0,03 | 0,17 | -0,16 | 8,71E-01 | 9,27E-01 |
| UBASH3A | 980,32 | -0,01 | 0,07 | -0,16 | 8,72E-01 | 9,27E-01 |
| FBXO48 | 247,53 | -0,01 | 0,04 | -0,16 | 8,72E-01 | 9,27E-01 |
| RHOA | 8163,17 | 0,01 | 0,05 | 0,16 | 8,72E-01 | 9,27E-01 |
| FCHSD2 | 1391,47 | 0,01 | 0,05 | 0,16 | 8,72E-01 | 9,27E-01 |
| NFKB1 | 3217,16 | 0,01 | 0,04 | 0,16 | 8,72E-01 | 9,27E-01 |
| PAFAH1B2 | 2411,73 | 0,01 | 0,04 | 0,16 | 8,72E-01 | 9,27E-01 |
| PLEKHB2 | 3810,05 | -0,01 | 0,04 | -0,16 | 8,72E-01 | 9,27E-01 |
| CLCN5 | 458,28 | -0,01 | 0,06 | -0,16 | 8,72E-01 | 9,27E-01 |
| ZNF587 | 1893,57 | -0,01 | 0,05 | -0,16 | 8,73E-01 | 9,28E-01 |
| CMTM4 | 276,00 | -0,01 | 0,06 | -0,16 | 8,73E-01 | 9,28E-01 |
| PPP1R10 | 2032,38 | 0,01 | 0,04 | 0,16 | 8,73E-01 | 9,28E-01 |
| NLRC4 | 1513,18 | 0,01 | 0,07 | 0,16 | 8,73E-01 | 9,28E-01 |
| MAP3K7CL | 957,75 | 0,01 | 0,08 | 0,16 | 8,73E-01 | 9,28E-01 |
| ZNF200 | 1137,69 | 0,01 | 0,04 | 0,16 | 8,73E-01 | 9,28E-01 |
| DECR1 | 3869,73 | 0,01 | 0,05 | 0,16 | 8,73E-01 | 9,28E-01 |
| ABHD5 | 6056,38 | 0,01 | 0,08 | 0,16 | 8,73E-01 | 9,28E-01 |
| LCLAT1 | 730,33 | -0,01 | 0,06 | -0,16 | 8,74E-01 | 9,28E-01 |
| MYCL | 421,62 | 0,01 | 0,06 | 0,16 | 8,74E-01 | 9,28E-01 |
| NCAM1 | 469,73 | -0,02 | 0,11 | -0,16 | 8,74E-01 | 9,29E-01 |
| NUDT7 | 69,61 | 0,01 | 0,08 | 0,16 | 8,74E-01 | 9,29E-01 |
| SMARCB1 | 503,58 | -0,01 | 0,09 | -0,16 | 8,75E-01 | 9,29E-01 |
| DYNLT1 | 1830,50 | -0,01 | 0,06 | -0,16 | 8,75E-01 | 9,29E-01 |
| CD80 | 55,28 | 0,02 | 0,10 | 0,16 | 8,75E-01 | 9,29E-01 |
| PALLD | 188,10 | -0,01 | 0,09 | -0,16 | 8,75E-01 | 9,29E-01 |
| MKRN3 | 86,44 | -0,03 | 0,16 | -0,16 | 8,75E-01 | 9,29E-01 |
| TMEM71 | 4697,24 | 0,01 | 0,06 | 0,16 | 8,75E-01 | 9,29E-01 |
| PGM2 | 2037,35 | -0,01 | 0,06 | -0,16 | 8,75E-01 | 9,29E-01 |
| INSR | 532,90 | -0,01 | 0,07 | -0,16 | 8,76E-01 | 9,29E-01 |
| AVL9 | 4252,81 | 0,01 | 0,03 | 0,16 | 8,76E-01 | 9,29E-01 |
| JAGN1 | 335,51 | 0,01 | 0,04 | 0,16 | 8,76E-01 | 9,29E-01 |

| | | | | | | |
|---|---|---|---|---|---|---|
| PIP4P2 | 1686,61 | -0,01 | 0,08 | -0,16 | 8,76E-01 | 9,29E-01 |
| MRPL48 | 211,40 | 0,01 | 0,06 | 0,16 | 8,76E-01 | 9,29E-01 |
| CDK11B | 669,45 | -0,01 | 0,04 | -0,16 | 8,75E-01 | 9,29E-01 |
| CMPK2 | 782,45 | -0,02 | 0,16 | -0,16 | 8,76E-01 | 9,29E-01 |
| SELENOM | 112,92 | -0,01 | 0,08 | -0,16 | 8,76E-01 | 9,29E-01 |
| SPG11 | 8275,18 | 0,01 | 0,04 | 0,16 | 8,76E-01 | 9,29E-01 |
| XKR9 | 39,60 | 0,02 | 0,15 | 0,15 | 8,77E-01 | 9,30E-01 |
| PKM | 2670,41 | -0,01 | 0,08 | -0,15 | 8,77E-01 | 9,30E-01 |
| UBE2G1 | 1222,37 | 0,00 | 0,03 | 0,15 | 8,77E-01 | 9,30E-01 |
| MRPL21 | 213,74 | -0,01 | 0,05 | -0,15 | 8,77E-01 | 9,30E-01 |
| RTP4 | 373,47 | -0,02 | 0,10 | -0,15 | 8,77E-01 | 9,30E-01 |
| AATF | 1496,26 | 0,01 | 0,04 | 0,15 | 8,78E-01 | 9,31E-01 |
| CACNG6 | 50,09 | -0,03 | 0,17 | -0,15 | 8,78E-01 | 9,31E-01 |
| ERI3 | 483,23 | 0,01 | 0,05 | 0,15 | 8,78E-01 | 9,31E-01 |
| UBXN1 | 1852,48 | 0,01 | 0,05 | 0,15 | 8,78E-01 | 9,31E-01 |
| MGST2 | 194,38 | -0,01 | 0,06 | -0,15 | 8,78E-01 | 9,31E-01 |
| IL15 | 622,88 | 0,01 | 0,07 | 0,15 | 8,78E-01 | 9,31E-01 |
| LPIN1 | 1900,27 | 0,01 | 0,04 | 0,15 | 8,78E-01 | 9,31E-01 |
| GLB1 | 1625,38 | 0,01 | 0,04 | 0,15 | 8,78E-01 | 9,31E-01 |
| MERTK | 178,30 | 0,02 | 0,12 | 0,15 | 8,79E-01 | 9,31E-01 |
| MPDU1 | 399,12 | -0,01 | 0,04 | -0,15 | 8,78E-01 | 9,31E-01 |
| INPP5B | 1609,03 | 0,01 | 0,04 | 0,15 | 8,78E-01 | 9,31E-01 |
| KLRC4 | 205,82 | -0,02 | 0,13 | -0,15 | 8,79E-01 | 9,31E-01 |
| DCAF8 | 2526,20 | 0,01 | 0,03 | 0,15 | 8,79E-01 | 9,31E-01 |
| NDUFA2 | 726,40 | -0,01 | 0,07 | -0,15 | 8,79E-01 | 9,31E-01 |
| CNIH4 | 1173,52 | -0,01 | 0,05 | -0,15 | 8,79E-01 | 9,31E-01 |
| ZBTB18 | 2448,55 | -0,01 | 0,05 | -0,15 | 8,79E-01 | 9,31E-01 |
| FAM13A | 1006,79 | -0,01 | 0,08 | -0,15 | 8,79E-01 | 9,31E-01 |
| LONRF3 | 380,98 | 0,01 | 0,05 | 0,15 | 8,79E-01 | 9,31E-01 |
| TMEM182 | 147,86 | 0,01 | 0,08 | 0,15 | 8,79E-01 | 9,31E-01 |
| TM2D3 | 1542,66 | 0,00 | 0,03 | 0,15 | 8,80E-01 | 9,31E-01 |
| CCDC32 | 677,85 | 0,01 | 0,05 | 0,15 | 8,80E-01 | 9,31E-01 |
| RNASEH2C | 777,62 | 0,01 | 0,04 | 0,15 | 8,80E-01 | 9,31E-01 |
| TSG101 | 1458,03 | 0,01 | 0,04 | 0,15 | 8,80E-01 | 9,31E-01 |
| IRS2 | 1414,77 | -0,01 | 0,08 | -0,15 | 8,80E-01 | 9,31E-01 |
| ANAPC11 | 453,14 | 0,01 | 0,05 | 0,15 | 8,80E-01 | 9,31E-01 |
| S100Z | 297,35 | -0,01 | 0,07 | -0,15 | 8,80E-01 | 9,31E-01 |
| ZNF300 | 130,57 | 0,02 | 0,11 | 0,15 | 8,80E-01 | 9,31E-01 |
| CCDC137 | 252,39 | 0,01 | 0,05 | 0,15 | 8,80E-01 | 9,31E-01 |
| GYPA | 349,81 | 0,02 | 0,15 | 0,15 | 8,81E-01 | 9,32E-01 |
| ASXL2 | 2984,83 | 0,00 | 0,03 | 0,15 | 8,81E-01 | 9,32E-01 |
| SLC2A3 | 8633,37 | -0,01 | 0,08 | -0,15 | 8,81E-01 | 9,32E-01 |
| TNFAIP8L2 | 1513,89 | 0,01 | 0,05 | 0,15 | 8,82E-01 | 9,32E-01 |
| CCRL2 | 143,25 | -0,01 | 0,06 | -0,15 | 8,82E-01 | 9,32E-01 |
| MYOF | 778,03 | 0,01 | 0,10 | 0,15 | 8,82E-01 | 9,32E-01 |
| TFCP2 | 689,79 | 0,01 | 0,04 | 0,15 | 8,82E-01 | 9,32E-01 |

| | | | | | | |
|---|---|---|---|---|---|---|
| HAPSTR1 | 9174,66 | -0,01 | 0,04 | -0,15 | 8,83E-01 | 9,33E-01 |
| AKAP1 | 535,63 | -0,01 | 0,05 | -0,15 | 8,83E-01 | 9,33E-01 |
| SRBD1 | 1423,16 | 0,01 | 0,06 | 0,15 | 8,83E-01 | 9,33E-01 |
| CBX5 | 1698,55 | -0,01 | 0,06 | -0,15 | 8,83E-01 | 9,33E-01 |
| PAXIP1 | 688,45 | 0,01 | 0,04 | 0,15 | 8,83E-01 | 9,33E-01 |
| TAF3 | 360,91 | -0,01 | 0,04 | -0,15 | 8,83E-01 | 9,33E-01 |
| ARHGAP11A | 190,14 | 0,01 | 0,09 | 0,15 | 8,84E-01 | 9,34E-01 |
| CHST2 | 1179,90 | 0,01 | 0,07 | 0,15 | 8,84E-01 | 9,34E-01 |
| PIP4P1 | 909,34 | 0,00 | 0,03 | 0,15 | 8,84E-01 | 9,34E-01 |
| RAB9B | 40,01 | -0,01 | 0,09 | -0,14 | 8,85E-01 | 9,35E-01 |
| RNF220 | 1186,30 | 0,00 | 0,03 | -0,14 | 8,86E-01 | 9,35E-01 |
| KIAA1958 | 254,75 | -0,01 | 0,07 | -0,14 | 8,86E-01 | 9,35E-01 |
| ANKLE2 | 2791,79 | 0,00 | 0,03 | -0,14 | 8,86E-01 | 9,35E-01 |
| OR2L2 | 26,09 | -0,02 | 0,17 | -0,14 | 8,86E-01 | 9,35E-01 |
| ZC3H12C | 207,24 | 0,01 | 0,08 | 0,14 | 8,86E-01 | 9,35E-01 |
| TRIM56 | 4762,53 | 0,01 | 0,05 | 0,14 | 8,86E-01 | 9,35E-01 |
| FCER1A | 442,10 | 0,02 | 0,15 | 0,14 | 8,86E-01 | 9,36E-01 |
| ZNF660 | 67,11 | -0,01 | 0,10 | -0,14 | 8,87E-01 | 9,36E-01 |
| TMEM176A | 149,96 | 0,04 | 0,26 | 0,14 | 8,87E-01 | 9,36E-01 |
| MLKL | 2321,18 | -0,01 | 0,06 | -0,14 | 8,87E-01 | 9,36E-01 |
| RSAD1 | 557,14 | 0,00 | 0,03 | 0,14 | 8,87E-01 | 9,36E-01 |
| GM2A | 1350,01 | 0,01 | 0,08 | 0,14 | 8,87E-01 | 9,36E-01 |
| CD40LG | 316,63 | 0,01 | 0,07 | 0,14 | 8,88E-01 | 9,36E-01 |
| MZB1 | 129,10 | -0,02 | 0,13 | -0,14 | 8,88E-01 | 9,36E-01 |
| SLAMF7 | 1060,22 | 0,01 | 0,08 | 0,14 | 8,88E-01 | 9,36E-01 |
| PIGX | 1395,10 | -0,01 | 0,06 | -0,14 | 8,88E-01 | 9,36E-01 |
| CD27 | 302,91 | -0,01 | 0,08 | -0,14 | 8,88E-01 | 9,36E-01 |
| PLA2G12A | 591,45 | -0,01 | 0,06 | -0,14 | 8,88E-01 | 9,36E-01 |
| TBC1D31 | 1086,03 | -0,01 | 0,06 | -0,14 | 8,88E-01 | 9,36E-01 |
| OASL | 659,37 | -0,02 | 0,14 | -0,14 | 8,88E-01 | 9,36E-01 |
| CNTNAP3 | 2274,08 | -0,02 | 0,13 | -0,14 | 8,88E-01 | 9,36E-01 |
| CTNND1 | 743,98 | 0,01 | 0,07 | 0,14 | 8,88E-01 | 9,36E-01 |
| IFNAR2 | 2251,20 | -0,01 | 0,05 | -0,14 | 8,88E-01 | 9,36E-01 |
| H2BC4 | 3952,42 | -0,01 | 0,08 | -0,14 | 8,89E-01 | 9,37E-01 |
| GPX7 | 139,93 | 0,01 | 0,06 | 0,14 | 8,89E-01 | 9,37E-01 |
| ZC3H11A | 111,56 | 0,01 | 0,05 | 0,14 | 8,89E-01 | 9,37E-01 |
| ZNF212 | 377,74 | -0,01 | 0,04 | -0,14 | 8,89E-01 | 9,37E-01 |
| CENPW | 25,61 | -0,01 | 0,10 | -0,14 | 8,89E-01 | 9,37E-01 |
| TMCO3 | 1869,50 | 0,01 | 0,05 | 0,14 | 8,89E-01 | 9,37E-01 |
| ADHFE1 | 144,70 | -0,01 | 0,06 | -0,14 | 8,90E-01 | 9,37E-01 |
| XKR6 | 315,16 | 0,01 | 0,07 | 0,14 | 8,90E-01 | 9,38E-01 |
| INO80E | 1075,24 | 0,01 | 0,05 | 0,14 | 8,91E-01 | 9,39E-01 |
| BRD8 | 922,57 | 0,01 | 0,04 | 0,14 | 8,91E-01 | 9,39E-01 |
| PPP1R3E | 661,23 | -0,01 | 0,05 | -0,14 | 8,92E-01 | 9,39E-01 |
| NCOA5 | 653,82 | 0,00 | 0,04 | -0,14 | 8,92E-01 | 9,39E-01 |
| SPINT2 | 1016,05 | -0,01 | 0,05 | -0,14 | 8,92E-01 | 9,39E-01 |

| | | | | | | |
|---|---|---|---|---|---|---|
| GOLGA7 | 1399,69 | 0,01 | 0,05 | 0,14 | 8,92E-01 | 9,39E-01 |
| CHST7 | 557,41 | -0,01 | 0,07 | -0,14 | 8,92E-01 | 9,39E-01 |
| C1orf162 | 1218,85 | -0,01 | 0,04 | -0,14 | 8,92E-01 | 9,39E-01 |
| ATAD3B | 694,06 | -0,01 | 0,06 | -0,14 | 8,92E-01 | 9,39E-01 |
| AUTS2 | 588,00 | -0,01 | 0,10 | -0,14 | 8,93E-01 | 9,39E-01 |
| RASGRP3 | 421,45 | 0,01 | 0,09 | 0,14 | 8,93E-01 | 9,39E-01 |
| ACVR2B | 651,40 | 0,01 | 0,07 | 0,13 | 8,93E-01 | 9,40E-01 |
| SLC2A9 | 301,18 | 0,01 | 0,08 | 0,13 | 8,93E-01 | 9,40E-01 |
| RBSN | 1336,87 | 0,00 | 0,03 | -0,13 | 8,93E-01 | 9,40E-01 |
| DNPH1 | 184,21 | -0,01 | 0,07 | -0,13 | 8,93E-01 | 9,40E-01 |
| PEX11A | 75,21 | -0,01 | 0,08 | -0,13 | 8,93E-01 | 9,40E-01 |
| VPS8 | 7864,62 | 0,01 | 0,06 | 0,13 | 8,93E-01 | 9,40E-01 |
| LYSMD2 | 461,82 | 0,01 | 0,05 | 0,13 | 8,94E-01 | 9,40E-01 |
| CTTNBP2NL | 120,72 | -0,01 | 0,08 | -0,13 | 8,94E-01 | 9,41E-01 |
| FCGR2B | 575,43 | -0,01 | 0,10 | -0,13 | 8,95E-01 | 9,41E-01 |
| DSC1 | 119,88 | -0,02 | 0,15 | -0,13 | 8,95E-01 | 9,41E-01 |
| ZSCAN20 | 157,42 | -0,01 | 0,07 | -0,13 | 8,95E-01 | 9,41E-01 |
| CCDC71L | 824,47 | -0,01 | 0,06 | -0,13 | 8,95E-01 | 9,41E-01 |
| PLPP1 | 125,20 | -0,01 | 0,06 | -0,13 | 8,96E-01 | 9,41E-01 |
| KDELR2 | 771,29 | 0,00 | 0,03 | -0,13 | 8,96E-01 | 9,41E-01 |
| CARD6 | 429,12 | -0,01 | 0,06 | -0,13 | 8,96E-01 | 9,42E-01 |
| DDIT3 | 359,72 | -0,01 | 0,07 | -0,13 | 8,96E-01 | 9,42E-01 |
| USP12 | 3324,38 | -0,01 | 0,10 | -0,13 | 8,97E-01 | 9,42E-01 |
| TRAPPC3 | 1039,88 | 0,00 | 0,03 | 0,13 | 8,97E-01 | 9,42E-01 |
| TMEM230 | 1193,02 | 0,01 | 0,04 | 0,13 | 8,97E-01 | 9,42E-01 |
| TNIP2 | 1012,40 | 0,00 | 0,03 | 0,13 | 8,97E-01 | 9,42E-01 |
| MOSPD1 | 384,84 | -0,01 | 0,09 | -0,13 | 8,97E-01 | 9,42E-01 |
| GSDMD | 2497,43 | -0,01 | 0,05 | -0,13 | 8,97E-01 | 9,42E-01 |
| LRRC41 | 652,27 | -0,01 | 0,04 | -0,13 | 8,97E-01 | 9,42E-01 |
| PPP2R2B | 387,39 | -0,01 | 0,10 | -0,13 | 8,97E-01 | 9,42E-01 |
| OMG | 40,55 | -0,01 | 0,11 | -0,13 | 8,98E-01 | 9,43E-01 |
| REEP5 | 1594,58 | 0,01 | 0,06 | 0,13 | 8,98E-01 | 9,43E-01 |
| NF1 | 3040,86 | 0,01 | 0,04 | 0,13 | 8,98E-01 | 9,43E-01 |
| SPIN2B | 86,94 | -0,01 | 0,05 | -0,13 | 8,98E-01 | 9,43E-01 |
| PSMD8 | 578,51 | 0,00 | 0,03 | 0,13 | 8,99E-01 | 9,43E-01 |
| EPM2A | 430,70 | -0,01 | 0,06 | -0,13 | 8,99E-01 | 9,43E-01 |
| ZNF674 | 486,24 | -0,01 | 0,04 | -0,13 | 8,99E-01 | 9,44E-01 |
| GEMIN4 | 425,65 | 0,01 | 0,04 | 0,13 | 8,99E-01 | 9,44E-01 |
| DUSP16 | 678,45 | -0,01 | 0,05 | -0,13 | 8,99E-01 | 9,44E-01 |
| THNSL2 | 66,80 | 0,03 | 0,20 | 0,13 | 8,99E-01 | 9,44E-01 |
| PCMT1 | 1396,42 | 0,01 | 0,07 | 0,13 | 9,00E-01 | 9,44E-01 |
| WDR73 | 950,24 | 0,00 | 0,03 | -0,13 | 9,00E-01 | 9,44E-01 |
| ENSG000002 | 62,03 | 0,01 | 0,07 | 0,13 | 9,00E-01 | 9,44E-01 |
| STX2 | 551,88 | 0,01 | 0,07 | 0,13 | 9,00E-01 | 9,44E-01 |
| NONO | 2085,04 | 0,00 | 0,04 | -0,13 | 9,00E-01 | 9,44E-01 |
| KLHL3 | 600,23 | 0,01 | 0,07 | 0,13 | 9,00E-01 | 9,44E-01 |

| | | | | | | |
|---|---|---|---|---|---|---|
| AIFM1 | 362,42 | 0,00 | 0,04 | 0,12 | 9,01E-01 | 9,44E-01 |
| IFT43 | 161,88 | -0,01 | 0,05 | -0,13 | 9,00E-01 | 9,44E-01 |
| RFT1 | 628,00 | 0,00 | 0,03 | 0,13 | 9,00E-01 | 9,44E-01 |
| RPS6KA4 | 626,97 | -0,01 | 0,06 | -0,12 | 9,01E-01 | 9,44E-01 |
| TMUB1 | 234,47 | -0,01 | 0,05 | -0,13 | 9,01E-01 | 9,44E-01 |
| WARS1 | 3627,37 | -0,01 | 0,09 | -0,13 | 9,00E-01 | 9,44E-01 |
| SLC23A2 | 1709,24 | 0,01 | 0,05 | 0,12 | 9,01E-01 | 9,44E-01 |
| HAUS7 | 60,97 | -0,01 | 0,09 | -0,12 | 9,01E-01 | 9,44E-01 |
| NPRL2 | 645,50 | 0,00 | 0,04 | -0,12 | 9,01E-01 | 9,44E-01 |
| LARP4B | 3927,42 | -0,01 | 0,04 | -0,12 | 9,01E-01 | 9,44E-01 |
| UGGT1 | 4676,42 | 0,00 | 0,03 | 0,12 | 9,01E-01 | 9,44E-01 |
| GFER | 190,12 | 0,01 | 0,05 | 0,12 | 9,01E-01 | 9,44E-01 |
| SDE2 | 1839,39 | -0,01 | 0,05 | -0,12 | 9,02E-01 | 9,45E-01 |
| TGFBR3 | 1364,53 | 0,01 | 0,09 | 0,12 | 9,02E-01 | 9,45E-01 |
| MTFP1 | 32,81 | -0,01 | 0,07 | -0,12 | 9,02E-01 | 9,45E-01 |
| DYNLL2 | 3060,58 | 0,01 | 0,04 | 0,12 | 9,02E-01 | 9,45E-01 |
| PRDX1 | 1755,54 | 0,01 | 0,07 | 0,12 | 9,02E-01 | 9,45E-01 |
| DGCR6L | 361,86 | -0,01 | 0,04 | -0,12 | 9,02E-01 | 9,45E-01 |
| SAMM50 | 652,53 | 0,00 | 0,03 | 0,12 | 9,02E-01 | 9,45E-01 |
| TMED8 | 1792,28 | 0,01 | 0,05 | 0,12 | 9,03E-01 | 9,45E-01 |
| LIN7A | 2218,84 | 0,01 | 0,10 | 0,12 | 9,03E-01 | 9,45E-01 |
| RABEP2 | 537,58 | -0,01 | 0,06 | -0,12 | 9,03E-01 | 9,45E-01 |
| ENSG00000 | 269,12 | -0,01 | 0,09 | -0,12 | 9,03E-01 | 9,45E-01 |
| PIGU | 246,14 | -0,01 | 0,04 | -0,12 | 9,03E-01 | 9,45E-01 |
| HNRNPUL1 | 2520,58 | -0,01 | 0,07 | -0,12 | 9,03E-01 | 9,45E-01 |
| METTL9 | 3775,80 | -0,01 | 0,07 | -0,12 | 9,03E-01 | 9,45E-01 |
| LAT2 | 3376,37 | -0,01 | 0,06 | -0,12 | 9,04E-01 | 9,45E-01 |
| DDX43 | 54,89 | 0,02 | 0,17 | 0,12 | 9,04E-01 | 9,45E-01 |
| NBPF26 | 1810,22 | 0,01 | 0,12 | 0,12 | 9,04E-01 | 9,46E-01 |
| HEXA | 1517,78 | 0,01 | 0,04 | 0,12 | 9,04E-01 | 9,46E-01 |
| SLC31A2 | 2693,05 | -0,01 | 0,08 | -0,12 | 9,05E-01 | 9,46E-01 |
| CACNB4 | 625,64 | -0,01 | 0,12 | -0,12 | 9,05E-01 | 9,46E-01 |
| SERGEF | 567,67 | 0,01 | 0,05 | 0,12 | 9,05E-01 | 9,47E-01 |
| MARCHF6 | 6563,42 | -0,01 | 0,04 | -0,12 | 9,05E-01 | 9,47E-01 |
| ITM2B | 25555,12 | -0,01 | 0,06 | -0,12 | 9,05E-01 | 9,47E-01 |
| SART1 | 1380,79 | -0,01 | 0,06 | -0,12 | 9,05E-01 | 9,47E-01 |
| NBPF19 | 3327,35 | -0,01 | 0,08 | -0,12 | 9,06E-01 | 9,47E-01 |
| TNFRSF8 | 278,61 | -0,01 | 0,07 | -0,12 | 9,06E-01 | 9,47E-01 |
| SIMC1 | 276,16 | -0,01 | 0,05 | -0,12 | 9,06E-01 | 9,47E-01 |
| FOLR2 | 48,92 | 0,01 | 0,10 | 0,12 | 9,06E-01 | 9,47E-01 |
| ARPC2 | 9470,78 | -0,01 | 0,04 | -0,12 | 9,07E-01 | 9,48E-01 |
| LRRC23 | 92,93 | 0,01 | 0,07 | 0,12 | 9,08E-01 | 9,48E-01 |
| ALKBH7 | 599,96 | 0,01 | 0,06 | 0,12 | 9,08E-01 | 9,48E-01 |
| RAB34 | 188,18 | 0,01 | 0,07 | 0,12 | 9,08E-01 | 9,48E-01 |
| MOB3B | 269,87 | 0,01 | 0,06 | 0,12 | 9,08E-01 | 9,49E-01 |
| NFXL1 | 989,97 | -0,01 | 0,11 | -0,12 | 9,08E-01 | 9,49E-01 |

| | | | | | | |
|---|---|---|---|---|---|---|
| MRI1 | 625,04 | -0,01 | 0,06 | -0,12 | 9,08E-01 | 9,49E-01 |
| SDCBP | 12041,17 | 0,01 | 0,07 | 0,12 | 9,08E-01 | 9,49E-01 |
| GPM6A | 92,70 | -0,02 | 0,21 | -0,11 | 9,08E-01 | 9,49E-01 |
| TRAF1 | 1493,21 | -0,01 | 0,07 | -0,11 | 9,09E-01 | 9,49E-01 |
| CYB561D2 | 394,78 | 0,00 | 0,03 | -0,11 | 9,09E-01 | 9,49E-01 |
| DCTPP1 | 194,29 | -0,01 | 0,05 | -0,11 | 9,09E-01 | 9,49E-01 |
| DDRGK1 | 702,03 | 0,00 | 0,03 | -0,11 | 9,09E-01 | 9,49E-01 |
| HPS4 | 920,53 | 0,00 | 0,04 | -0,11 | 9,09E-01 | 9,49E-01 |
| KLHL8 | 1443,45 | 0,01 | 0,06 | 0,11 | 9,10E-01 | 9,49E-01 |
| BRAT1 | 790,73 | -0,01 | 0,05 | -0,11 | 9,10E-01 | 9,49E-01 |
| EIF1AD | 672,62 | 0,00 | 0,03 | 0,11 | 9,10E-01 | 9,50E-01 |
| SUV39H1 | 135,81 | 0,00 | 0,04 | -0,11 | 9,10E-01 | 9,50E-01 |
| MAP2K1 | 1528,06 | 0,00 | 0,04 | -0,11 | 9,10E-01 | 9,50E-01 |
| JRK | 1411,01 | -0,01 | 0,05 | -0,11 | 9,10E-01 | 9,50E-01 |
| UBR7 | 475,73 | 0,00 | 0,03 | 0,11 | 9,10E-01 | 9,50E-01 |
| USP49 | 445,73 | -0,01 | 0,05 | -0,11 | 9,10E-01 | 9,50E-01 |
| FTO | 1458,62 | 0,01 | 0,06 | 0,11 | 9,11E-01 | 9,50E-01 |
| RAD52 | 325,22 | -0,01 | 0,05 | -0,11 | 9,11E-01 | 9,50E-01 |
| TSPAN2 | 1135,00 | 0,01 | 0,08 | 0,11 | 9,11E-01 | 9,50E-01 |
| EOGT | 273,32 | 0,01 | 0,05 | 0,11 | 9,11E-01 | 9,50E-01 |
| SERTAD1 | 364,92 | 0,01 | 0,05 | 0,11 | 9,11E-01 | 9,51E-01 |
| XCL1 | 53,02 | -0,01 | 0,14 | -0,11 | 9,12E-01 | 9,51E-01 |
| CDC7 | 277,94 | -0,01 | 0,07 | -0,11 | 9,12E-01 | 9,51E-01 |
| BEND4 | 144,69 | 0,01 | 0,12 | 0,11 | 9,12E-01 | 9,51E-01 |
| EGLN2 | 1505,77 | 0,01 | 0,06 | 0,11 | 9,12E-01 | 9,51E-01 |
| NCR3 | 121,72 | 0,01 | 0,08 | 0,11 | 9,12E-01 | 9,51E-01 |
| PPP1R12B | 5291,98 | -0,01 | 0,08 | -0,11 | 9,12E-01 | 9,51E-01 |
| ZNF781 | 75,61 | 0,01 | 0,10 | 0,11 | 9,12E-01 | 9,51E-01 |
| PRDM5 | 520,35 | -0,01 | 0,12 | -0,11 | 9,14E-01 | 9,52E-01 |
| PDXDC1 | 2062,21 | 0,00 | 0,04 | -0,11 | 9,14E-01 | 9,52E-01 |
| BCAT1 | 428,15 | -0,01 | 0,11 | -0,11 | 9,14E-01 | 9,52E-01 |
| ASB13 | 258,32 | 0,01 | 0,05 | 0,11 | 9,14E-01 | 9,52E-01 |
| ADORA2A | 165,79 | 0,01 | 0,06 | 0,11 | 9,15E-01 | 9,53E-01 |
| ENSG000002 | 41,01 | -0,01 | 0,07 | -0,11 | 9,15E-01 | 9,53E-01 |
| ACAD9 | 976,98 | 0,00 | 0,04 | 0,11 | 9,15E-01 | 9,53E-01 |
| IFFO2 | 961,57 | -0,01 | 0,05 | -0,11 | 9,16E-01 | 9,53E-01 |
| LRRC39 | 27,06 | 0,01 | 0,11 | 0,11 | 9,16E-01 | 9,53E-01 |
| ZNF784 | 136,84 | 0,01 | 0,05 | 0,11 | 9,16E-01 | 9,53E-01 |
| DAD1 | 1264,28 | 0,01 | 0,06 | 0,11 | 9,16E-01 | 9,54E-01 |
| C6orf62 | 11400,18 | 0,00 | 0,05 | 0,11 | 9,16E-01 | 9,54E-01 |
| ACOT9 | 1365,57 | -0,01 | 0,05 | -0,11 | 9,16E-01 | 9,54E-01 |
| MCOLN2 | 522,87 | 0,01 | 0,09 | 0,11 | 9,16E-01 | 9,54E-01 |
| SLC25A11 | 695,80 | 0,00 | 0,04 | -0,11 | 9,16E-01 | 9,54E-01 |
| TLR8 | 5154,62 | 0,01 | 0,08 | 0,10 | 9,16E-01 | 9,54E-01 |
| YWHAE | 788,72 | 0,00 | 0,04 | -0,10 | 9,16E-01 | 9,54E-01 |
| GTF2I | 904,71 | 0,00 | 0,04 | -0,11 | 9,16E-01 | 9,54E-01 |

| | | | | | | |
|---|---|---|---|---|---|---|
| HACD4 | 3728,86 | -0,01 | 0,06 | -0,10 | 9,17E-01 | 9,54E-01 |
| ZNF665 | 143,30 | 0,01 | 0,09 | 0,10 | 9,17E-01 | 9,54E-01 |
| LRRC34 | 42,03 | 0,01 | 0,12 | 0,10 | 9,17E-01 | 9,54E-01 |
| CBY1 | 224,15 | 0,00 | 0,04 | -0,10 | 9,18E-01 | 9,55E-01 |
| ITGB7 | 1821,69 | -0,01 | 0,06 | -0,10 | 9,18E-01 | 9,55E-01 |
| SLITRK5 | 49,38 | -0,02 | 0,17 | -0,10 | 9,18E-01 | 9,55E-01 |
| RRP1 | 311,69 | 0,00 | 0,05 | -0,10 | 9,18E-01 | 9,55E-01 |
| DNAJC16 | 723,35 | 0,00 | 0,03 | -0,10 | 9,18E-01 | 9,55E-01 |
| TREM1 | 5642,70 | -0,01 | 0,08 | -0,10 | 9,18E-01 | 9,55E-01 |
| TXNDC11 | 789,47 | 0,00 | 0,03 | 0,10 | 9,19E-01 | 9,55E-01 |
| GOLM1 | 202,52 | -0,01 | 0,09 | -0,10 | 9,19E-01 | 9,55E-01 |
| ASB3 | 342,51 | 0,00 | 0,03 | 0,10 | 9,19E-01 | 9,55E-01 |
| H2AC4 | 183,48 | 0,01 | 0,12 | 0,10 | 9,19E-01 | 9,55E-01 |
| RCN1 | 150,98 | -0,01 | 0,07 | -0,10 | 9,19E-01 | 9,56E-01 |
| AURKA | 129,84 | -0,01 | 0,10 | -0,10 | 9,20E-01 | 9,56E-01 |
| FHIT | 539,67 | 0,01 | 0,11 | 0,10 | 9,20E-01 | 9,56E-01 |
| MME | 13212,09 | 0,01 | 0,10 | 0,10 | 9,20E-01 | 9,56E-01 |
| WDR45B | 1255,57 | 0,00 | 0,03 | 0,10 | 9,20E-01 | 9,56E-01 |
| ZSCAN18 | 596,19 | -0,01 | 0,07 | -0,10 | 9,20E-01 | 9,56E-01 |
| AIP | 372,83 | -0,01 | 0,06 | -0,10 | 9,20E-01 | 9,56E-01 |
| NHLRC4 | 86,81 | -0,01 | 0,07 | -0,10 | 9,21E-01 | 9,56E-01 |
| ZBTB40 | 2015,43 | 0,00 | 0,05 | -0,10 | 9,21E-01 | 9,56E-01 |
| KANSL1 | 6803,43 | 0,01 | 0,07 | 0,10 | 9,21E-01 | 9,57E-01 |
| FOXRED1 | 279,99 | 0,01 | 0,06 | 0,10 | 9,22E-01 | 9,57E-01 |
| XAF1 | 2818,42 | -0,01 | 0,11 | -0,10 | 9,21E-01 | 9,57E-01 |
| ALKBH4 | 178,31 | 0,00 | 0,04 | 0,10 | 9,22E-01 | 9,57E-01 |
| CD2BP2 | 1447,48 | 0,00 | 0,04 | 0,10 | 9,22E-01 | 9,57E-01 |
| CYSLTR2 | 704,86 | 0,01 | 0,09 | 0,10 | 9,22E-01 | 9,57E-01 |
| CASP6 | 295,85 | 0,00 | 0,05 | -0,10 | 9,22E-01 | 9,57E-01 |
| ZNF395 | 303,82 | -0,01 | 0,09 | -0,10 | 9,22E-01 | 9,57E-01 |
| COMMD4 | 268,61 | 0,00 | 0,05 | -0,10 | 9,22E-01 | 9,57E-01 |
| NFATC2IP | 1499,21 | 0,00 | 0,04 | -0,10 | 9,22E-01 | 9,57E-01 |
| FRRS1 | 267,00 | -0,01 | 0,10 | -0,10 | 9,22E-01 | 9,57E-01 |
| EML5 | 382,73 | 0,01 | 0,07 | 0,10 | 9,23E-01 | 9,57E-01 |
| PCBD2 | 450,01 | 0,00 | 0,04 | -0,10 | 9,23E-01 | 9,57E-01 |
| IRAG1 | 1567,48 | -0,01 | 0,08 | -0,10 | 9,23E-01 | 9,57E-01 |
| CHD3 | 4862,93 | -0,01 | 0,06 | -0,10 | 9,23E-01 | 9,57E-01 |
| KLHL6 | 2486,02 | 0,00 | 0,04 | -0,10 | 9,23E-01 | 9,57E-01 |
| HLA-B | 69661,96 | -0,01 | 0,06 | -0,10 | 9,23E-01 | 9,58E-01 |
| ADGRB3 | 46,84 | 0,02 | 0,19 | 0,10 | 9,24E-01 | 9,58E-01 |
| FNDC3B | 5909,90 | -0,01 | 0,06 | -0,10 | 9,24E-01 | 9,58E-01 |
| WDR26 | 9853,63 | 0,01 | 0,06 | 0,10 | 9,24E-01 | 9,58E-01 |
| KRTCAP2 | 160,68 | 0,00 | 0,05 | 0,09 | 9,24E-01 | 9,59E-01 |
| CCDC71 | 431,92 | 0,00 | 0,04 | -0,09 | 9,25E-01 | 9,59E-01 |
| HTD2 | 21,09 | -0,01 | 0,08 | -0,09 | 9,25E-01 | 9,59E-01 |
| PRR14L | 3924,34 | 0,00 | 0,03 | -0,09 | 9,25E-01 | 9,59E-01 |

| | | | | | | |
|---|---|---|---|---|---|---|
| CD93 | 8960,62 | -0,01 | 0,09 | -0,09 | 9,25E-01 | 9,59E-01 |
| KPNA2 | 100,30 | 0,01 | 0,07 | 0,09 | 9,25E-01 | 9,59E-01 |
| DDX60L | 9937,76 | 0,01 | 0,09 | 0,09 | 9,25E-01 | 9,59E-01 |
| TMBIM6 | 11718,12 | 0,00 | 0,05 | -0,09 | 9,26E-01 | 9,59E-01 |
| PELI3 | 209,90 | -0,01 | 0,06 | -0,09 | 9,26E-01 | 9,59E-01 |
| WDR82 | 5728,34 | 0,00 | 0,04 | 0,09 | 9,26E-01 | 9,59E-01 |
| RRAGA | 525,92 | 0,00 | 0,04 | -0,09 | 9,26E-01 | 9,59E-01 |
| PCCB | 502,28 | 0,00 | 0,03 | 0,09 | 9,26E-01 | 9,59E-01 |
| ZBED4 | 995,32 | 0,00 | 0,04 | 0,09 | 9,26E-01 | 9,59E-01 |
| NSD1 | 6169,00 | 0,00 | 0,04 | 0,09 | 9,26E-01 | 9,59E-01 |
| NEIL3 | 307,54 | -0,01 | 0,15 | -0,09 | 9,27E-01 | 9,60E-01 |
| PSME4 | 4291,37 | 0,00 | 0,05 | -0,09 | 9,27E-01 | 9,60E-01 |
| MTRFR | 547,33 | 0,00 | 0,04 | 0,09 | 9,27E-01 | 9,60E-01 |
| NABP1 | 19501,97 | -0,01 | 0,07 | -0,09 | 9,27E-01 | 9,60E-01 |
| MLYCD | 970,22 | 0,01 | 0,06 | 0,09 | 9,27E-01 | 9,60E-01 |
| PATZ1 | 584,04 | 0,00 | 0,05 | 0,09 | 9,28E-01 | 9,61E-01 |
| ALG9 | 203,89 | 0,00 | 0,05 | 0,09 | 9,28E-01 | 9,61E-01 |
| GTF2H2 | 315,48 | -0,01 | 0,13 | -0,09 | 9,28E-01 | 9,61E-01 |
| ASPM | 192,71 | -0,01 | 0,11 | -0,09 | 9,28E-01 | 9,61E-01 |
| GNGT2 | 119,97 | -0,01 | 0,06 | -0,09 | 9,29E-01 | 9,61E-01 |
| CLEC12A | 4576,97 | -0,01 | 0,15 | -0,09 | 9,29E-01 | 9,61E-01 |
| FAM50A | 418,57 | 0,00 | 0,04 | 0,09 | 9,29E-01 | 9,61E-01 |
| TIGAR | 147,52 | 0,00 | 0,04 | 0,09 | 9,29E-01 | 9,61E-01 |
| ENSG000002 | 66,89 | -0,01 | 0,14 | -0,09 | 9,29E-01 | 9,61E-01 |
| CHD8 | 3699,32 | 0,00 | 0,04 | 0,09 | 9,29E-01 | 9,61E-01 |
| MOB3C | 1430,42 | 0,00 | 0,03 | 0,09 | 9,29E-01 | 9,61E-01 |
| ARHGAP19 | 1277,68 | 0,00 | 0,06 | -0,09 | 9,29E-01 | 9,61E-01 |
| HLA-DQA2 | 178,70 | 0,03 | 0,37 | 0,09 | 9,29E-01 | 9,61E-01 |
| SNAP23 | 5573,85 | -0,01 | 0,06 | -0,09 | 9,29E-01 | 9,61E-01 |
| TPI1 | 3127,70 | 0,00 | 0,04 | -0,09 | 9,29E-01 | 9,61E-01 |
| SYK | 8921,78 | 0,01 | 0,06 | 0,09 | 9,30E-01 | 9,61E-01 |
| ADAT1 | 1425,78 | 0,00 | 0,04 | 0,09 | 9,30E-01 | 9,61E-01 |
| SDR39U1 | 402,29 | 0,00 | 0,04 | -0,09 | 9,30E-01 | 9,62E-01 |
| SEC13 | 1154,09 | 0,00 | 0,03 | 0,09 | 9,30E-01 | 9,62E-01 |
| TBCB | 928,57 | 0,00 | 0,04 | -0,09 | 9,31E-01 | 9,62E-01 |
| TRIM26 | 801,13 | 0,00 | 0,05 | -0,09 | 9,32E-01 | 9,63E-01 |
| PSMB8 | 3250,64 | 0,00 | 0,05 | -0,09 | 9,31E-01 | 9,63E-01 |
| MPV17 | 724,19 | 0,00 | 0,03 | -0,09 | 9,31E-01 | 9,63E-01 |
| TLE4 | 4451,26 | 0,00 | 0,04 | -0,09 | 9,32E-01 | 9,63E-01 |
| ST6GALNAC3 | 239,90 | 0,01 | 0,07 | 0,08 | 9,32E-01 | 9,63E-01 |
| MRPL36 | 153,08 | 0,01 | 0,07 | 0,08 | 9,32E-01 | 9,63E-01 |
| CXXC4 | 28,07 | -0,01 | 0,17 | -0,08 | 9,33E-01 | 9,63E-01 |
| POLR3C | 429,32 | 0,00 | 0,03 | 0,08 | 9,33E-01 | 9,63E-01 |
| KIF14 | 123,05 | -0,01 | 0,13 | -0,08 | 9,33E-01 | 9,64E-01 |
| RRAGC | 441,44 | 0,00 | 0,04 | -0,08 | 9,33E-01 | 9,64E-01 |
| MSANTD3 | 201,01 | 0,00 | 0,05 | 0,08 | 9,34E-01 | 9,64E-01 |

| | | | | | | |
|---|---|---|---|---|---|---|
| RAB23 | 70,63 | -0,01 | 0,11 | -0,08 | 9,34E-01 | 9,64E-01 |
| PANK1 | 108,11 | -0,01 | 0,10 | -0,08 | 9,34E-01 | 9,64E-01 |
| JARID2 | 3660,89 | -0,01 | 0,06 | -0,08 | 9,33E-01 | 9,64E-01 |
| MDM2 | 6346,25 | 0,00 | 0,05 | 0,08 | 9,34E-01 | 9,64E-01 |
| ZNF585A | 320,46 | 0,00 | 0,04 | 0,08 | 9,35E-01 | 9,65E-01 |
| COQ6 | 496,20 | 0,00 | 0,03 | 0,08 | 9,35E-01 | 9,65E-01 |
| C17orf67 | 226,03 | 0,00 | 0,05 | -0,08 | 9,35E-01 | 9,65E-01 |
| RPGRIP1L | 194,06 | 0,01 | 0,09 | 0,08 | 9,35E-01 | 9,65E-01 |
| DYRK4 | 295,48 | 0,00 | 0,06 | 0,08 | 9,35E-01 | 9,65E-01 |
| ZNF285 | 75,71 | 0,01 | 0,12 | 0,08 | 9,36E-01 | 9,65E-01 |
| MCTP1 | 3061,77 | -0,01 | 0,08 | -0,08 | 9,36E-01 | 9,65E-01 |
| ZNF768 | 177,61 | 0,00 | 0,05 | -0,08 | 9,36E-01 | 9,66E-01 |
| CARD9 | 729,37 | 0,01 | 0,07 | 0,08 | 9,37E-01 | 9,66E-01 |
| CRPPA | 123,60 | -0,01 | 0,10 | -0,08 | 9,37E-01 | 9,66E-01 |
| LYRM4 | 330,31 | 0,00 | 0,06 | 0,08 | 9,37E-01 | 9,66E-01 |
| ZNG1F | 529,03 | 0,01 | 0,07 | 0,08 | 9,37E-01 | 9,66E-01 |
| ZGPAT | 568,87 | 0,00 | 0,04 | -0,08 | 9,37E-01 | 9,66E-01 |
| SMG5 | 1146,03 | -0,01 | 0,08 | -0,08 | 9,37E-01 | 9,66E-01 |
| CCDC125 | 1435,17 | 0,00 | 0,06 | -0,08 | 9,37E-01 | 9,66E-01 |
| CDKN1C | 439,79 | 0,01 | 0,12 | 0,08 | 9,37E-01 | 9,66E-01 |
| G0S2 | 47,15 | -0,01 | 0,18 | -0,08 | 9,37E-01 | 9,66E-01 |
| CD3E | 651,65 | -0,01 | 0,08 | -0,08 | 9,37E-01 | 9,66E-01 |
| SNRNP200 | 4626,49 | 0,00 | 0,04 | -0,08 | 9,37E-01 | 9,66E-01 |
| U2AF1 | 369,17 | 0,02 | 0,20 | 0,08 | 9,37E-01 | 9,66E-01 |
| PGBD1 | 56,82 | 0,01 | 0,08 | 0,08 | 9,37E-01 | 9,66E-01 |
| KLHL36 | 2766,25 | 0,00 | 0,04 | 0,08 | 9,38E-01 | 9,66E-01 |
| GHITM | 4686,66 | -0,01 | 0,07 | -0,08 | 9,38E-01 | 9,66E-01 |
| SESN3 | 11741,69 | 0,01 | 0,09 | 0,08 | 9,38E-01 | 9,66E-01 |
| SLC49A4 | 1291,01 | 0,00 | 0,06 | 0,08 | 9,38E-01 | 9,66E-01 |
| TESPA1 | 1528,01 | -0,01 | 0,06 | -0,08 | 9,38E-01 | 9,66E-01 |
| PPP4R1 | 6642,41 | 0,00 | 0,06 | 0,08 | 9,39E-01 | 9,67E-01 |
| EIF4G3 | 1991,84 | 0,00 | 0,05 | 0,08 | 9,39E-01 | 9,67E-01 |
| GBP4 | 2518,73 | -0,01 | 0,09 | -0,08 | 9,39E-01 | 9,67E-01 |
| XPC | 2721,92 | 0,00 | 0,05 | 0,08 | 9,39E-01 | 9,67E-01 |
| TMSB4X | 70700,76 | 0,01 | 0,08 | 0,08 | 9,39E-01 | 9,67E-01 |
| ARCN1 | 2170,82 | 0,00 | 0,04 | 0,08 | 9,39E-01 | 9,67E-01 |
| ZNF385A | 211,18 | 0,01 | 0,13 | 0,08 | 9,39E-01 | 9,67E-01 |
| NUP153 | 3963,95 | 0,00 | 0,04 | -0,08 | 9,40E-01 | 9,67E-01 |
| ZNF106 | 8180,07 | 0,00 | 0,05 | 0,08 | 9,40E-01 | 9,67E-01 |
| WDR6 | 1325,20 | 0,00 | 0,06 | -0,08 | 9,40E-01 | 9,67E-01 |
| ENSG000002 | 37,95 | 0,00 | 0,06 | -0,07 | 9,40E-01 | 9,68E-01 |
| LGR4 | 83,20 | -0,01 | 0,12 | -0,07 | 9,41E-01 | 9,68E-01 |
| UBE2V1 | 695,85 | 0,00 | 0,05 | -0,07 | 9,41E-01 | 9,68E-01 |
| PTPN12 | 2646,86 | 0,00 | 0,05 | -0,07 | 9,41E-01 | 9,68E-01 |
| CEP63 | 2807,29 | 0,00 | 0,06 | -0,07 | 9,41E-01 | 9,68E-01 |
| RNF135 | 672,78 | 0,00 | 0,05 | -0,07 | 9,41E-01 | 9,68E-01 |

| | | | | | | |
|---|---|---|---|---|---|---|
| FUT7 | 1083,75 | -0,01 | 0,08 | -0,07 | 9,41E-01 | 9,68E-01 |
| MGST1 | 169,55 | -0,01 | 0,08 | -0,07 | 9,42E-01 | 9,69E-01 |
| SRP19 | 593,34 | 0,00 | 0,03 | 0,07 | 9,42E-01 | 9,69E-01 |
| MED26 | 193,96 | 0,00 | 0,06 | -0,07 | 9,42E-01 | 9,69E-01 |
| BNIP3L | 40044,02 | 0,01 | 0,12 | 0,07 | 9,42E-01 | 9,69E-01 |
| POLR3E | 1113,55 | 0,00 | 0,04 | 0,07 | 9,42E-01 | 9,69E-01 |
| GAPVD1 | 3862,47 | 0,00 | 0,04 | -0,07 | 9,42E-01 | 9,69E-01 |
| TRMT44 | 560,41 | 0,00 | 0,04 | -0,07 | 9,43E-01 | 9,69E-01 |
| MIEF2 | 348,73 | 0,00 | 0,04 | 0,07 | 9,43E-01 | 9,70E-01 |
| ANKHD1-EIF | 261,27 | 0,00 | 0,06 | 0,07 | 9,43E-01 | 9,70E-01 |
| EVL | 7349,05 | 0,00 | 0,07 | -0,07 | 9,44E-01 | 9,70E-01 |
| MB21D2 | 121,52 | -0,01 | 0,08 | -0,07 | 9,44E-01 | 9,70E-01 |
| MPV17L2 | 62,01 | 0,00 | 0,06 | -0,07 | 9,44E-01 | 9,70E-01 |
| STK19 | 315,88 | 0,00 | 0,04 | 0,07 | 9,44E-01 | 9,70E-01 |
| PINX1 | 150,11 | 0,00 | 0,04 | -0,07 | 9,44E-01 | 9,70E-01 |
| PGAM5 | 349,10 | 0,00 | 0,04 | -0,07 | 9,44E-01 | 9,70E-01 |
| TNFAIP8L1 | 294,78 | 0,00 | 0,06 | -0,07 | 9,45E-01 | 9,70E-01 |
| SERF1B | 42,26 | -0,01 | 0,08 | -0,07 | 9,45E-01 | 9,70E-01 |
| CDK12 | 3497,89 | 0,00 | 0,03 | 0,07 | 9,45E-01 | 9,71E-01 |
| SIN3A | 1972,96 | 0,00 | 0,04 | 0,07 | 9,45E-01 | 9,71E-01 |
| POGZ | 1931,39 | 0,00 | 0,04 | -0,07 | 9,45E-01 | 9,71E-01 |
| CHN2 | 445,29 | -0,01 | 0,08 | -0,07 | 9,46E-01 | 9,71E-01 |
| KHNYN | 4924,42 | 0,00 | 0,04 | -0,07 | 9,45E-01 | 9,71E-01 |
| NCKAP1L | 7965,82 | 0,00 | 0,04 | -0,07 | 9,45E-01 | 9,71E-01 |
| NFKBIE | 454,44 | 0,00 | 0,05 | 0,07 | 9,46E-01 | 9,71E-01 |
| WHAMM | 932,32 | 0,00 | 0,04 | -0,07 | 9,46E-01 | 9,71E-01 |
| CTPS1 | 675,62 | 0,00 | 0,05 | 0,07 | 9,46E-01 | 9,71E-01 |
| ANKUB1 | 116,42 | -0,01 | 0,10 | -0,07 | 9,46E-01 | 9,71E-01 |
| SIDT1 | 1585,38 | 0,00 | 0,05 | 0,07 | 9,46E-01 | 9,71E-01 |
| ZNF662 | 74,05 | 0,01 | 0,11 | 0,07 | 9,46E-01 | 9,71E-01 |
| KAT6A | 10264,41 | 0,00 | 0,04 | 0,07 | 9,46E-01 | 9,71E-01 |
| RAB32 | 881,99 | 0,00 | 0,07 | 0,07 | 9,47E-01 | 9,71E-01 |
| CCDC57 | 980,40 | -0,01 | 0,08 | -0,07 | 9,47E-01 | 9,71E-01 |
| UBIAD1 | 497,74 | 0,00 | 0,04 | 0,07 | 9,47E-01 | 9,71E-01 |
| MAP3K5 | 5930,03 | 0,00 | 0,05 | -0,07 | 9,47E-01 | 9,71E-01 |
| ZNF705EP | 23,17 | -0,01 | 0,10 | -0,07 | 9,47E-01 | 9,72E-01 |
| WDR77 | 414,26 | 0,00 | 0,04 | -0,07 | 9,48E-01 | 9,72E-01 |
| COG8 | 909,62 | 0,00 | 0,03 | 0,07 | 9,48E-01 | 9,72E-01 |
| PRADC1 | 126,75 | 0,00 | 0,05 | 0,06 | 9,48E-01 | 9,72E-01 |
| GALC | 1910,21 | 0,00 | 0,05 | 0,06 | 9,49E-01 | 9,73E-01 |
| STX6 | 2352,32 | 0,00 | 0,04 | 0,06 | 9,49E-01 | 9,73E-01 |
| ENTPD1 | 5302,21 | 0,00 | 0,07 | 0,06 | 9,49E-01 | 9,73E-01 |
| SLC25A15 | 167,19 | 0,00 | 0,04 | 0,06 | 9,49E-01 | 9,73E-01 |
| TSPYL4 | 450,37 | 0,00 | 0,03 | -0,06 | 9,49E-01 | 9,73E-01 |
| LIAS | 480,74 | 0,00 | 0,03 | -0,06 | 9,49E-01 | 9,73E-01 |
| PPP2R2D | 1858,99 | 0,00 | 0,03 | 0,06 | 9,50E-01 | 9,73E-01 |

| | | | | | | |
|---|---|---|---|---|---|---|
| ANKRD28 | 1870,76 | 0,00 | 0,06 | -0,06 | 9,50E-01 | 9,73E-01 |
| ZNF2 | 218,47 | 0,00 | 0,03 | -0,06 | 9,50E-01 | 9,73E-01 |
| SAMHD1 | 8776,01 | 0,00 | 0,05 | 0,06 | 9,50E-01 | 9,73E-01 |
| STAB1 | 1466,80 | -0,01 | 0,11 | -0,06 | 9,50E-01 | 9,73E-01 |
| RNF25 | 244,30 | 0,00 | 0,03 | -0,06 | 9,51E-01 | 9,74E-01 |
| CEBPA | 1231,74 | 0,00 | 0,06 | -0,06 | 9,51E-01 | 9,74E-01 |
| AIDA | 1435,02 | 0,00 | 0,07 | 0,06 | 9,51E-01 | 9,74E-01 |
| ANAPC13 | 654,94 | 0,00 | 0,07 | 0,06 | 9,51E-01 | 9,74E-01 |
| GCH1 | 1704,26 | 0,00 | 0,07 | -0,06 | 9,52E-01 | 9,74E-01 |
| RHOB | 2726,41 | 0,00 | 0,07 | 0,06 | 9,52E-01 | 9,74E-01 |
| BYSL | 104,82 | 0,00 | 0,05 | -0,06 | 9,52E-01 | 9,75E-01 |
| IGFBP4 | 64,17 | -0,01 | 0,14 | -0,06 | 9,52E-01 | 9,75E-01 |
| NCOA2 | 7514,85 | 0,00 | 0,04 | -0,06 | 9,53E-01 | 9,75E-01 |
| FUCA1 | 588,58 | 0,00 | 0,06 | -0,06 | 9,52E-01 | 9,75E-01 |
| RAB31 | 5053,22 | 0,00 | 0,07 | -0,06 | 9,52E-01 | 9,75E-01 |
| PNOC | 35,57 | -0,01 | 0,12 | -0,06 | 9,52E-01 | 9,75E-01 |
| ISCA1 | 2819,35 | -0,01 | 0,10 | -0,06 | 9,53E-01 | 9,75E-01 |
| KNL1 | 258,01 | 0,01 | 0,11 | 0,06 | 9,53E-01 | 9,75E-01 |
| MED28 | 4959,55 | 0,00 | 0,03 | 0,06 | 9,53E-01 | 9,75E-01 |
| TYW1B | 283,59 | 0,00 | 0,05 | -0,06 | 9,53E-01 | 9,75E-01 |
| PTEN | 23055,17 | 0,00 | 0,08 | 0,06 | 9,53E-01 | 9,75E-01 |
| HABP4 | 431,50 | 0,00 | 0,05 | -0,06 | 9,53E-01 | 9,75E-01 |
| ANXA4 | 807,00 | 0,00 | 0,04 | -0,06 | 9,54E-01 | 9,75E-01 |
| TM7SF3 | 1045,04 | 0,00 | 0,03 | -0,06 | 9,54E-01 | 9,75E-01 |
| NDFIP2 | 164,98 | 0,01 | 0,09 | 0,06 | 9,54E-01 | 9,75E-01 |
| SKP1 | 3102,79 | 0,00 | 0,05 | 0,06 | 9,54E-01 | 9,75E-01 |
| MAP3K20 | 911,03 | 0,00 | 0,06 | -0,06 | 9,54E-01 | 9,76E-01 |
| SLC16A1 | 336,09 | 0,00 | 0,05 | -0,06 | 9,54E-01 | 9,76E-01 |
| COLGALT1 | 1489,95 | 0,00 | 0,05 | 0,06 | 9,55E-01 | 9,76E-01 |
| SETBP1 | 581,39 | 0,00 | 0,08 | 0,06 | 9,55E-01 | 9,76E-01 |
| FPR3 | 55,35 | 0,01 | 0,12 | 0,06 | 9,55E-01 | 9,76E-01 |
| TCF19 | 115,40 | 0,00 | 0,08 | 0,06 | 9,55E-01 | 9,76E-01 |
| MAFB | 543,38 | 0,00 | 0,08 | -0,06 | 9,55E-01 | 9,76E-01 |
| CUL4A | 3073,06 | 0,00 | 0,04 | 0,06 | 9,55E-01 | 9,76E-01 |
| DHX58 | 584,49 | 0,00 | 0,08 | 0,06 | 9,56E-01 | 9,77E-01 |
| TRMT61A | 219,27 | 0,00 | 0,05 | -0,06 | 9,56E-01 | 9,77E-01 |
| HSPA1A | 1302,15 | 0,00 | 0,09 | 0,05 | 9,57E-01 | 9,77E-01 |
| MACC1 | 93,11 | 0,01 | 0,11 | 0,05 | 9,57E-01 | 9,77E-01 |
| RASSF1 | 3015,19 | 0,00 | 0,05 | 0,05 | 9,56E-01 | 9,77E-01 |
| P4HB | 2193,31 | 0,00 | 0,05 | -0,05 | 9,57E-01 | 9,77E-01 |
| HADHB | 2239,31 | 0,00 | 0,04 | 0,05 | 9,57E-01 | 9,77E-01 |
| CMAS | 537,34 | 0,00 | 0,08 | -0,05 | 9,57E-01 | 9,77E-01 |
| MAPRE2 | 1641,88 | 0,00 | 0,04 | 0,05 | 9,57E-01 | 9,77E-01 |
| PLK4 | 86,53 | 0,00 | 0,09 | 0,05 | 9,57E-01 | 9,77E-01 |
| WRNIP1 | 1311,31 | 0,00 | 0,03 | 0,05 | 9,58E-01 | 9,77E-01 |
| ERMP1 | 1157,16 | 0,00 | 0,05 | 0,05 | 9,58E-01 | 9,77E-01 |

| | | | | | | |
|---|---|---|---|---|---|---|
| CDC16 | 1376,57 | 0,00 | 0,05 | -0,05 | 9,58E-01 | 9,77E-01 |
| ERV3-1 | 2847,70 | 0,00 | 0,07 | -0,05 | 9,58E-01 | 9,78E-01 |
| COPG1 | 2188,22 | 0,00 | 0,04 | 0,05 | 9,58E-01 | 9,78E-01 |
| PPP4C | 1971,47 | 0,00 | 0,06 | 0,05 | 9,58E-01 | 9,78E-01 |
| NMNAT1 | 436,31 | 0,00 | 0,05 | 0,05 | 9,58E-01 | 9,78E-01 |
| FAM20B | 1409,60 | 0,00 | 0,05 | -0,05 | 9,59E-01 | 9,78E-01 |
| S100PBP | 1963,79 | 0,00 | 0,04 | -0,05 | 9,59E-01 | 9,78E-01 |
| AKIRIN2 | 998,23 | 0,00 | 0,05 | -0,05 | 9,59E-01 | 9,78E-01 |
| RPGR | 1311,17 | 0,00 | 0,06 | 0,05 | 9,59E-01 | 9,78E-01 |
| PRKRIP1 | 757,85 | 0,00 | 0,03 | 0,05 | 9,59E-01 | 9,78E-01 |
| TP53BP1 | 1480,67 | 0,00 | 0,04 | -0,05 | 9,59E-01 | 9,79E-01 |
| HLCS | 491,32 | 0,00 | 0,06 | 0,05 | 9,60E-01 | 9,79E-01 |
| TNFRSF13C | 173,57 | -0,01 | 0,15 | -0,05 | 9,60E-01 | 9,79E-01 |
| XKRX | 66,15 | -0,01 | 0,13 | -0,05 | 9,60E-01 | 9,79E-01 |
| TSSC4 | 1003,29 | 0,00 | 0,05 | -0,05 | 9,60E-01 | 9,79E-01 |
| GMNN | 93,39 | 0,00 | 0,07 | 0,05 | 9,60E-01 | 9,79E-01 |
| CUBN | 824,74 | 0,00 | 0,08 | 0,05 | 9,60E-01 | 9,79E-01 |
| BUD31 | 774,59 | 0,00 | 0,05 | -0,05 | 9,61E-01 | 9,79E-01 |
| HTR7 | 39,75 | -0,01 | 0,12 | -0,05 | 9,61E-01 | 9,79E-01 |
| SRGAP2B | 387,91 | 0,00 | 0,05 | -0,05 | 9,61E-01 | 9,79E-01 |
| WDR17 | 86,92 | 0,00 | 0,10 | 0,05 | 9,61E-01 | 9,79E-01 |
| PDLIM5 | 1617,44 | 0,00 | 0,04 | -0,05 | 9,61E-01 | 9,79E-01 |
| SFXN4 | 197,97 | 0,00 | 0,05 | -0,05 | 9,61E-01 | 9,79E-01 |
| ZFAND4 | 1038,49 | 0,00 | 0,05 | -0,05 | 9,61E-01 | 9,79E-01 |
| PIK3CG | 3871,19 | 0,00 | 0,04 | -0,05 | 9,61E-01 | 9,79E-01 |
| CD302 | 2347,96 | 0,00 | 0,08 | 0,05 | 9,62E-01 | 9,79E-01 |
| KLKB1 | 67,43 | -0,01 | 0,11 | -0,05 | 9,62E-01 | 9,80E-01 |
| ZBTB49 | 439,17 | 0,00 | 0,03 | 0,05 | 9,62E-01 | 9,80E-01 |
| SYDE2 | 139,34 | 0,00 | 0,09 | -0,05 | 9,62E-01 | 9,80E-01 |
| DCAF10 | 1813,42 | 0,00 | 0,04 | -0,05 | 9,62E-01 | 9,80E-01 |
| CCND2 | 3180,94 | 0,00 | 0,05 | 0,05 | 9,62E-01 | 9,80E-01 |
| PPP1R3F | 460,30 | 0,00 | 0,05 | -0,05 | 9,62E-01 | 9,80E-01 |
| PTGES2 | 393,73 | 0,00 | 0,06 | 0,05 | 9,63E-01 | 9,80E-01 |
| TOP2A | 132,72 | -0,01 | 0,11 | -0,05 | 9,63E-01 | 9,80E-01 |
| GMPPB | 539,82 | 0,00 | 0,04 | 0,05 | 9,63E-01 | 9,80E-01 |
| USP13 | 324,83 | 0,00 | 0,05 | 0,05 | 9,63E-01 | 9,80E-01 |
| NOSIP | 1936,01 | 0,00 | 0,05 | 0,05 | 9,63E-01 | 9,80E-01 |
| CEP250 | 1792,65 | 0,00 | 0,05 | -0,05 | 9,64E-01 | 9,81E-01 |
| USP9X | 7101,44 | 0,00 | 0,04 | 0,04 | 9,64E-01 | 9,81E-01 |
| CTSL | 286,63 | 0,00 | 0,10 | -0,04 | 9,64E-01 | 9,81E-01 |
| H2AZ1 | 2913,41 | 0,00 | 0,07 | 0,04 | 9,64E-01 | 9,81E-01 |
| MTX1 | 1073,55 | 0,00 | 0,07 | -0,04 | 9,64E-01 | 9,81E-01 |
| MPRIP | 4484,08 | 0,00 | 0,07 | 0,04 | 9,64E-01 | 9,81E-01 |
| DCTN4 | 2018,64 | 0,00 | 0,04 | 0,04 | 9,64E-01 | 9,81E-01 |
| DIS3L2 | 904,32 | 0,00 | 0,05 | -0,04 | 9,64E-01 | 9,81E-01 |
| PNPLA4 | 92,79 | 0,00 | 0,05 | -0,04 | 9,64E-01 | 9,81E-01 |

| | | | | | | |
|---|---|---|---|---|---|---|
| TDRD15 | 33,82 | -0,01 | 0,21 | -0,04 | 9,65E-01 | 9,81E-01 |
| ANK3 | 2154,89 | 0,00 | 0,08 | 0,04 | 9,65E-01 | 9,81E-01 |
| CIAO2B | 537,71 | 0,00 | 0,05 | -0,04 | 9,65E-01 | 9,81E-01 |
| CD63 | 2936,81 | 0,00 | 0,07 | -0,04 | 9,65E-01 | 9,81E-01 |
| VSTM1 | 232,46 | -0,01 | 0,12 | -0,04 | 9,65E-01 | 9,81E-01 |
| BECN1 | 2781,37 | 0,00 | 0,04 | -0,04 | 9,66E-01 | 9,82E-01 |
| AKR1B1 | 389,11 | 0,00 | 0,04 | 0,04 | 9,66E-01 | 9,82E-01 |
| ARHGAP17 | 1172,71 | 0,00 | 0,08 | -0,04 | 9,66E-01 | 9,82E-01 |
| TARS3 | 842,49 | 0,00 | 0,05 | -0,04 | 9,66E-01 | 9,82E-01 |
| HEBP2 | 3601,12 | 0,00 | 0,05 | 0,04 | 9,66E-01 | 9,82E-01 |
| ATP10A | 1060,52 | 0,00 | 0,07 | -0,04 | 9,66E-01 | 9,82E-01 |
| SEPTIN10 | 111,02 | 0,00 | 0,10 | 0,04 | 9,67E-01 | 9,82E-01 |
| ARMH2 | 54,45 | 0,00 | 0,07 | -0,04 | 9,67E-01 | 9,82E-01 |
| ZNF396 | 145,74 | 0,00 | 0,07 | 0,04 | 9,67E-01 | 9,82E-01 |
| PELI1 | 8634,31 | 0,00 | 0,07 | 0,04 | 9,67E-01 | 9,82E-01 |
| CUZD1 | 134,42 | 0,00 | 0,07 | 0,04 | 9,67E-01 | 9,82E-01 |
| FHIP2A | 2359,52 | 0,00 | 0,06 | 0,04 | 9,67E-01 | 9,82E-01 |
| CYP51A1 | 85,37 | 0,00 | 0,06 | -0,04 | 9,67E-01 | 9,82E-01 |
| GOSR1 | 1217,77 | 0,00 | 0,03 | -0,04 | 9,67E-01 | 9,82E-01 |
| CTBS | 3687,38 | 0,00 | 0,08 | 0,04 | 9,67E-01 | 9,82E-01 |
| SRP68 | 633,07 | 0,00 | 0,05 | 0,04 | 9,67E-01 | 9,82E-01 |
| COG1 | 648,65 | 0,00 | 0,04 | 0,04 | 9,67E-01 | 9,82E-01 |
| DNMBP | 386,82 | 0,00 | 0,08 | -0,04 | 9,68E-01 | 9,82E-01 |
| BFAR | 644,87 | 0,00 | 0,03 | 0,04 | 9,68E-01 | 9,82E-01 |
| HPGD | 158,79 | 0,00 | 0,10 | 0,04 | 9,68E-01 | 9,82E-01 |
| TRIM5 | 932,28 | 0,00 | 0,06 | 0,04 | 9,68E-01 | 9,82E-01 |
| ABI2 | 871,83 | 0,00 | 0,05 | 0,04 | 9,68E-01 | 9,83E-01 |
| NBPF10 | 422,64 | 0,00 | 0,06 | 0,04 | 9,69E-01 | 9,83E-01 |
| PMF1 | 271,69 | 0,00 | 0,05 | 0,04 | 9,69E-01 | 9,83E-01 |
| PAQR7 | 214,19 | 0,00 | 0,06 | -0,04 | 9,69E-01 | 9,83E-01 |
| IFI35 | 796,63 | 0,00 | 0,09 | -0,04 | 9,69E-01 | 9,83E-01 |
| DBNL | 10289,28 | 0,00 | 0,05 | 0,04 | 9,69E-01 | 9,83E-01 |
| MICU1 | 1006,16 | 0,00 | 0,04 | -0,04 | 9,69E-01 | 9,83E-01 |
| CLIC1 | 8175,26 | 0,00 | 0,07 | 0,04 | 9,69E-01 | 9,83E-01 |
| VIPAS39 | 862,47 | 0,00 | 0,03 | 0,04 | 9,70E-01 | 9,83E-01 |
| LIMD1 | 1595,30 | 0,00 | 0,06 | 0,04 | 9,70E-01 | 9,83E-01 |
| POLD4 | 851,99 | 0,00 | 0,05 | 0,04 | 9,70E-01 | 9,83E-01 |
| NR4A2 | 94,87 | 0,00 | 0,08 | 0,04 | 9,70E-01 | 9,83E-01 |
| C1QTNF3 | 117,01 | 0,00 | 0,07 | -0,04 | 9,70E-01 | 9,83E-01 |
| BEND5 | 87,87 | 0,00 | 0,10 | 0,04 | 9,70E-01 | 9,83E-01 |
| PRCP | 2503,16 | 0,00 | 0,05 | 0,04 | 9,70E-01 | 9,83E-01 |
| EPHA4 | 718,00 | 0,00 | 0,08 | 0,04 | 9,70E-01 | 9,84E-01 |
| USF3 | 2644,64 | 0,00 | 0,04 | 0,04 | 9,71E-01 | 9,84E-01 |
| ZNF19 | 124,65 | 0,00 | 0,09 | 0,04 | 9,71E-01 | 9,84E-01 |
| PRSS23 | 471,67 | 0,00 | 0,12 | 0,04 | 9,71E-01 | 9,84E-01 |
| PDE7B | 90,39 | 0,00 | 0,10 | -0,04 | 9,71E-01 | 9,84E-01 |

| | | | | | | |
|---|---|---|---|---|---|---|
| CAB39L | 176,35 | 0,00 | 0,05 | -0,04 | 9,71E-01 | 9,84E-01 |
| H1-10 | 562,84 | 0,00 | 0,09 | 0,04 | 9,71E-01 | 9,84E-01 |
| NRCAM | 88,99 | 0,01 | 0,20 | 0,04 | 9,71E-01 | 9,84E-01 |
| GUCA1B | 63,58 | 0,00 | 0,08 | -0,03 | 9,72E-01 | 9,85E-01 |
| PELI2 | 3543,70 | 0,00 | 0,06 | -0,03 | 9,72E-01 | 9,85E-01 |
| NHS | 377,18 | 0,00 | 0,07 | -0,03 | 9,72E-01 | 9,85E-01 |
| AMDHD2 | 738,55 | 0,00 | 0,06 | -0,03 | 9,73E-01 | 9,85E-01 |
| PRKCB | 10389,50 | 0,00 | 0,05 | 0,03 | 9,73E-01 | 9,85E-01 |
| ATP1A1 | 3254,17 | 0,00 | 0,04 | -0,03 | 9,73E-01 | 9,85E-01 |
| HFE | 209,16 | 0,00 | 0,05 | -0,03 | 9,73E-01 | 9,85E-01 |
| KAT7 | 2866,64 | 0,00 | 0,04 | 0,03 | 9,74E-01 | 9,85E-01 |
| LRRK2 | 41725,55 | 0,00 | 0,09 | -0,03 | 9,73E-01 | 9,85E-01 |
| S1PR3 | 454,76 | 0,00 | 0,08 | -0,03 | 9,73E-01 | 9,85E-01 |
| PRRC2C | 8966,38 | 0,00 | 0,04 | 0,03 | 9,74E-01 | 9,85E-01 |
| SARNP | 187,69 | 0,00 | 0,04 | -0,03 | 9,74E-01 | 9,85E-01 |
| CHD7 | 2868,42 | 0,00 | 0,05 | -0,03 | 9,73E-01 | 9,85E-01 |
| NIPAL3 | 2105,49 | 0,00 | 0,05 | -0,03 | 9,73E-01 | 9,85E-01 |
| ABI3 | 441,47 | 0,00 | 0,07 | -0,03 | 9,74E-01 | 9,85E-01 |
| PBX3 | 695,08 | 0,00 | 0,04 | 0,03 | 9,73E-01 | 9,85E-01 |
| FDPS | 1126,25 | 0,00 | 0,03 | -0,03 | 9,74E-01 | 9,85E-01 |
| BRK1 | 1782,91 | 0,00 | 0,07 | -0,03 | 9,74E-01 | 9,85E-01 |
| SF3B5 | 790,76 | 0,00 | 0,06 | 0,03 | 9,74E-01 | 9,86E-01 |
| UQCC2 | 311,21 | 0,00 | 0,04 | 0,03 | 9,75E-01 | 9,86E-01 |
| PSTK | 128,27 | 0,00 | 0,07 | 0,03 | 9,75E-01 | 9,86E-01 |
| TPM4 | 5191,69 | 0,00 | 0,05 | -0,03 | 9,75E-01 | 9,86E-01 |
| ANKRD36B | 1262,52 | 0,00 | 0,09 | 0,03 | 9,75E-01 | 9,86E-01 |
| DUSP23 | 228,50 | 0,00 | 0,05 | 0,03 | 9,76E-01 | 9,86E-01 |
| P2RX4 | 631,81 | 0,00 | 0,04 | 0,03 | 9,76E-01 | 9,86E-01 |
| PSMD3 | 771,31 | 0,00 | 0,04 | -0,03 | 9,76E-01 | 9,86E-01 |
| ZBTB22 | 357,10 | 0,00 | 0,05 | 0,03 | 9,75E-01 | 9,86E-01 |
| SNX1 | 3845,37 | 0,00 | 0,03 | -0,03 | 9,75E-01 | 9,86E-01 |
| GLA | 459,26 | 0,00 | 0,06 | 0,03 | 9,76E-01 | 9,86E-01 |
| CXCL6 | 29,63 | 0,00 | 0,12 | -0,03 | 9,77E-01 | 9,87E-01 |
| IFNAR1 | 4796,98 | 0,00 | 0,05 | -0,03 | 9,77E-01 | 9,87E-01 |
| UBXN11 | 688,76 | 0,00 | 0,10 | 0,03 | 9,77E-01 | 9,87E-01 |
| ARL15 | 1032,70 | 0,00 | 0,06 | 0,03 | 9,77E-01 | 9,87E-01 |
| GPATCH1 | 358,37 | 0,00 | 0,03 | 0,03 | 9,77E-01 | 9,87E-01 |
| IFT56 | 161,94 | 0,00 | 0,10 | -0,03 | 9,77E-01 | 9,87E-01 |
| TTC8 | 106,19 | 0,00 | 0,08 | 0,03 | 9,77E-01 | 9,87E-01 |
| UTS2 | 39,84 | -0,01 | 0,19 | -0,03 | 9,77E-01 | 9,87E-01 |
| CD19 | 169,33 | 0,00 | 0,12 | -0,03 | 9,77E-01 | 9,87E-01 |
| MSRB2 | 329,88 | 0,00 | 0,07 | 0,03 | 9,78E-01 | 9,87E-01 |
| NUBP2 | 775,45 | 0,00 | 0,04 | 0,03 | 9,78E-01 | 9,88E-01 |
| GPR132 | 1194,47 | 0,00 | 0,05 | -0,03 | 9,78E-01 | 9,88E-01 |
| LCP1 | 40886,78 | 0,00 | 0,06 | 0,03 | 9,79E-01 | 9,88E-01 |
| HLA-F | 5810,31 | 0,00 | 0,05 | 0,03 | 9,79E-01 | 9,88E-01 |

| | | | | | | |
|---|---|---|---|---|---|---|
| NUDT14 | 87,35 | 0,00 | 0,07 | 0,03 | 9,79E-01 | 9,88E-01 |
| TRAF4 | 169,65 | 0,00 | 0,07 | -0,03 | 9,79E-01 | 9,88E-01 |
| TOMM40 | 284,18 | 0,00 | 0,06 | 0,03 | 9,79E-01 | 9,88E-01 |
| SIVA1 | 690,36 | 0,00 | 0,04 | -0,03 | 9,80E-01 | 9,88E-01 |
| APBA2 | 856,33 | 0,00 | 0,09 | -0,03 | 9,80E-01 | 9,88E-01 |
| RBM34 | 90,54 | 0,00 | 0,05 | -0,03 | 9,79E-01 | 9,88E-01 |
| TRIM16 | 198,38 | 0,00 | 0,06 | 0,03 | 9,80E-01 | 9,88E-01 |
| JHY | 91,39 | 0,00 | 0,09 | -0,03 | 9,79E-01 | 9,88E-01 |
| ARHGEF7 | 2053,37 | 0,00 | 0,04 | -0,03 | 9,80E-01 | 9,88E-01 |
| MRPL55 | 257,00 | 0,00 | 0,04 | 0,02 | 9,80E-01 | 9,89E-01 |
| AIRIM | 302,42 | 0,00 | 0,03 | 0,02 | 9,80E-01 | 9,89E-01 |
| RMND5A | 7299,93 | 0,00 | 0,06 | -0,02 | 9,81E-01 | 9,89E-01 |
| F2RL1 | 835,17 | 0,00 | 0,10 | 0,02 | 9,81E-01 | 9,89E-01 |
| NUDC | 608,48 | 0,00 | 0,04 | -0,02 | 9,80E-01 | 9,89E-01 |
| JMJD6 | 935,16 | 0,00 | 0,04 | -0,02 | 9,81E-01 | 9,89E-01 |
| RBX1 | 1196,72 | 0,00 | 0,08 | 0,02 | 9,81E-01 | 9,89E-01 |
| PSMB3 | 2612,37 | 0,00 | 0,06 | 0,02 | 9,81E-01 | 9,89E-01 |
| DNAAF9 | 677,53 | 0,00 | 0,06 | -0,02 | 9,81E-01 | 9,89E-01 |
| H2AC12 | 330,97 | 0,00 | 0,11 | 0,02 | 9,81E-01 | 9,89E-01 |
| POLD3 | 721,34 | 0,00 | 0,04 | -0,02 | 9,81E-01 | 9,89E-01 |
| SLC46A2 | 256,59 | 0,00 | 0,07 | 0,02 | 9,81E-01 | 9,89E-01 |
| HERC1 | 14078,55 | 0,00 | 0,04 | -0,02 | 9,81E-01 | 9,89E-01 |
| PRKAG2 | 995,39 | 0,00 | 0,04 | -0,02 | 9,81E-01 | 9,89E-01 |
| NUDT2 | 206,87 | 0,00 | 0,05 | 0,02 | 9,82E-01 | 9,89E-01 |
| NME1 | 104,26 | 0,00 | 0,08 | -0,02 | 9,82E-01 | 9,89E-01 |
| C3AR1 | 313,78 | 0,00 | 0,09 | 0,02 | 9,82E-01 | 9,89E-01 |
| GSTO1 | 1250,60 | 0,00 | 0,07 | -0,02 | 9,82E-01 | 9,89E-01 |
| CYRIB | 8305,39 | 0,00 | 0,05 | 0,02 | 9,82E-01 | 9,89E-01 |
| LMAN2L | 303,88 | 0,00 | 0,03 | -0,02 | 9,82E-01 | 9,89E-01 |
| CFAP418 | 72,48 | 0,00 | 0,07 | -0,02 | 9,82E-01 | 9,89E-01 |
| SPOCK2 | 2959,99 | 0,00 | 0,08 | 0,02 | 9,82E-01 | 9,89E-01 |
| B3GALT4 | 780,48 | 0,00 | 0,04 | -0,02 | 9,82E-01 | 9,89E-01 |
| AGMAT | 156,23 | 0,00 | 0,07 | 0,02 | 9,83E-01 | 9,89E-01 |
| GBP2 | 9477,64 | 0,00 | 0,07 | 0,02 | 9,83E-01 | 9,89E-01 |
| CAPRIN2 | 1223,40 | 0,00 | 0,05 | -0,02 | 9,83E-01 | 9,89E-01 |
| SPTBN1 | 4654,89 | 0,00 | 0,06 | -0,02 | 9,83E-01 | 9,89E-01 |
| DNAJC11 | 608,35 | 0,00 | 0,05 | -0,02 | 9,83E-01 | 9,89E-01 |
| RBM10 | 1158,12 | 0,00 | 0,05 | -0,02 | 9,83E-01 | 9,89E-01 |
| GGACT | 195,68 | 0,00 | 0,05 | -0,02 | 9,84E-01 | 9,90E-01 |
| CYFIP1 | 966,76 | 0,00 | 0,07 | -0,02 | 9,84E-01 | 9,90E-01 |
| CYTH3 | 309,86 | 0,00 | 0,07 | 0,02 | 9,84E-01 | 9,90E-01 |
| PITRM1 | 975,96 | 0,00 | 0,04 | 0,02 | 9,84E-01 | 9,90E-01 |
| WDR20 | 1799,45 | 0,00 | 0,04 | 0,02 | 9,84E-01 | 9,90E-01 |
| P2RY1 | 208,43 | 0,00 | 0,09 | -0,02 | 9,84E-01 | 9,90E-01 |
| ENSG000002 | 125,60 | 0,00 | 0,05 | -0,02 | 9,85E-01 | 9,90E-01 |
| EMILIN2 | 2059,00 | 0,00 | 0,06 | -0,02 | 9,85E-01 | 9,90E-01 |

| | | | | | | |
|---|---|---|---|---|---|---|
| MISFA | 40,20 | 0,00 | 0,09 | -0,02 | 9,85E-01 | 9,90E-01 |
| CHIC2 | 2091,81 | 0,00 | 0,06 | 0,02 | 9,85E-01 | 9,90E-01 |
| TPGS1 | 560,57 | 0,00 | 0,04 | 0,02 | 9,85E-01 | 9,90E-01 |
| IFI30 | 245,93 | 0,00 | 0,07 | 0,02 | 9,86E-01 | 9,91E-01 |
| GNL3L | 1330,46 | 0,00 | 0,06 | -0,02 | 9,86E-01 | 9,91E-01 |
| ENSG000001 | 44684,85 | 0,00 | 0,08 | 0,02 | 9,86E-01 | 9,91E-01 |
| HOMER1 | 102,85 | 0,00 | 0,09 | 0,02 | 9,86E-01 | 9,91E-01 |
| CAPN12 | 149,71 | 0,00 | 0,11 | -0,02 | 9,86E-01 | 9,91E-01 |
| ZNHIT3 | 588,21 | 0,00 | 0,05 | -0,02 | 9,86E-01 | 9,91E-01 |
| ANTXR2 | 7005,86 | 0,00 | 0,05 | 0,02 | 9,86E-01 | 9,91E-01 |
| LAIR1 | 1658,18 | 0,00 | 0,04 | 0,02 | 9,86E-01 | 9,91E-01 |
| PPP1R7 | 827,60 | 0,00 | 0,03 | 0,02 | 9,86E-01 | 9,91E-01 |
| HADH | 466,11 | 0,00 | 0,05 | 0,02 | 9,87E-01 | 9,91E-01 |
| C17orf107 | 33,53 | 0,00 | 0,17 | 0,02 | 9,87E-01 | 9,91E-01 |
| LNX2 | 550,33 | 0,00 | 0,04 | 0,02 | 9,87E-01 | 9,91E-01 |
| TPH1 | 72,76 | 0,00 | 0,11 | 0,02 | 9,87E-01 | 9,91E-01 |
| KRT72 | 114,94 | 0,00 | 0,19 | -0,02 | 9,87E-01 | 9,91E-01 |
| PLEKHA2 | 3587,42 | 0,00 | 0,04 | 0,02 | 9,88E-01 | 9,92E-01 |
| CMC2 | 595,59 | 0,00 | 0,04 | 0,01 | 9,89E-01 | 9,93E-01 |
| PUS1 | 442,57 | 0,00 | 0,05 | -0,01 | 9,90E-01 | 9,94E-01 |
| ZSCAN5A | 276,98 | 0,00 | 0,06 | -0,01 | 9,90E-01 | 9,94E-01 |
| GLYCTK | 395,64 | 0,00 | 0,07 | -0,01 | 9,90E-01 | 9,94E-01 |
| TBC1D7 | 504,69 | 0,00 | 0,05 | -0,01 | 9,90E-01 | 9,94E-01 |
| OAS1 | 1767,61 | 0,00 | 0,12 | 0,01 | 9,90E-01 | 9,94E-01 |
| B3GNT5 | 583,46 | 0,00 | 0,08 | -0,01 | 9,91E-01 | 9,95E-01 |
| FIBP | 352,57 | 0,00 | 0,03 | -0,01 | 9,92E-01 | 9,95E-01 |
| ENTPD7 | 323,15 | 0,00 | 0,04 | 0,01 | 9,92E-01 | 9,95E-01 |
| NEK3 | 197,04 | 0,00 | 0,05 | -0,01 | 9,92E-01 | 9,95E-01 |
| HERPUD2 | 2044,47 | 0,00 | 0,03 | 0,01 | 9,92E-01 | 9,95E-01 |
| OGFOD1 | 442,85 | 0,00 | 0,03 | 0,01 | 9,92E-01 | 9,95E-01 |
| ASB8 | 1298,55 | 0,00 | 0,03 | 0,01 | 9,92E-01 | 9,95E-01 |
| NET1 | 280,62 | 0,00 | 0,08 | 0,01 | 9,93E-01 | 9,96E-01 |
| COPA | 4570,67 | 0,00 | 0,04 | 0,01 | 9,93E-01 | 9,96E-01 |
| ALDH9A1 | 1286,07 | 0,00 | 0,04 | 0,01 | 9,93E-01 | 9,96E-01 |
| PCNX2 | 1636,52 | 0,00 | 0,06 | -0,01 | 9,93E-01 | 9,96E-01 |
| ALS2 | 978,08 | 0,00 | 0,04 | -0,01 | 9,93E-01 | 9,96E-01 |
| GPN2 | 691,42 | 0,00 | 0,03 | -0,01 | 9,94E-01 | 9,96E-01 |
| CD300C | 285,88 | 0,00 | 0,08 | 0,01 | 9,94E-01 | 9,96E-01 |
| RRBP1 | 1540,09 | 0,00 | 0,06 | 0,01 | 9,95E-01 | 9,97E-01 |
| CCL3 | 78,70 | 0,00 | 0,11 | 0,01 | 9,95E-01 | 9,97E-01 |
| PPIP5K1 | 149,49 | 0,00 | 0,06 | -0,01 | 9,95E-01 | 9,97E-01 |
| AQP3 | 303,12 | 0,00 | 0,08 | -0,01 | 9,95E-01 | 9,97E-01 |
| GPRASP2 | 63,08 | 0,00 | 0,07 | -0,01 | 9,95E-01 | 9,97E-01 |
| KCNN4 | 283,94 | 0,00 | 0,09 | 0,01 | 9,96E-01 | 9,98E-01 |
| ST3GAL5 | 1580,94 | 0,00 | 0,05 | -0,01 | 9,96E-01 | 9,98E-01 |
| TREML2 | 1485,45 | 0,00 | 0,07 | 0,00 | 9,96E-01 | 9,98E-01 |

| | | | | | | |
|---|---|---|---|---|---|---|
| TAFA2 | 1019,81 | 0,00 | 0,08 | 0,00 | 9,96E-01 | 9,98E-01 |
| DPY19L2 | 88,27 | 0,00 | 0,14 | 0,00 | 9,96E-01 | 9,98E-01 |
| FARP2 | 733,31 | 0,00 | 0,05 | 0,00 | 9,96E-01 | 9,98E-01 |
| MBD3 | 696,47 | 0,00 | 0,04 | 0,00 | 9,96E-01 | 9,98E-01 |
| SRGN | 29170,26 | 0,00 | 0,11 | 0,00 | 9,97E-01 | 9,98E-01 |
| ASB7 | 1043,17 | 0,00 | 0,04 | 0,00 | 9,97E-01 | 9,98E-01 |
| EIF2D | 799,76 | 0,00 | 0,03 | 0,00 | 9,97E-01 | 9,98E-01 |
| PNRC1 | 5074,19 | 0,00 | 0,06 | 0,00 | 9,97E-01 | 9,98E-01 |
| LRRC1 | 133,23 | 0,00 | 0,08 | 0,00 | 9,97E-01 | 9,98E-01 |
| MED29 | 668,48 | 0,00 | 0,04 | 0,00 | 9,97E-01 | 9,98E-01 |
| ICAM3 | 7765,95 | 0,00 | 0,07 | 0,00 | 9,98E-01 | 9,98E-01 |
| VPS18 | 1007,86 | 0,00 | 0,04 | 0,00 | 9,98E-01 | 9,99E-01 |
| GABARAPL1 | 2838,86 | 0,00 | 0,07 | 0,00 | 9,98E-01 | 9,99E-01 |
| ASB1 | 1121,35 | 0,00 | 0,03 | 0,00 | 9,98E-01 | 9,99E-01 |
| GNAQ | 5669,55 | 0,00 | 0,06 | 0,00 | 9,99E-01 | 9,99E-01 |
| C16orf74 | 138,40 | 0,00 | 0,09 | 0,00 | 9,99E-01 | 9,99E-01 |
| CLMN | 852,79 | 0,00 | 0,07 | 0,00 | 9,99E-01 | 1,00E+00 |
| CCNE2 | 56,06 | 0,00 | 0,11 | 0,00 | 9,99E-01 | 1,00E+00 |
| MBTPS1 | 2947,29 | 0,00 | 0,03 | 0,00 | 1,00E+00 | 1,00E+00 |
| LNP1 | 50,25 | 0,00 | 0,09 | 0,00 | 1,00E+00 | 1,00E+00 |
| CPM | 175,78 | 0,00 | 0,08 | 0,00 | 1,00E+00 | 1,00E+00 |
| APOL2 | 3881,55 | 0,00 | 0,07 | 0,00 | 1,00E+00 | 1,00E+00 |
