## Supplementary material for "Longitudinal Assessment of DNA Repair Signature Trajectory in Prodromal versus Established Parkinson’s Disease": Table S4

| | baseMean | log2FoldCha | lfcSE | stat | pvalue |
|---|---|---|---|---|---|
| SUPT16H | 2028,64 | -0,18 | 0,03 | -5,54 | 2,97E-08 |
| HBB | 1278,97 | 1,14 | 0,21 | 5,43 | 5,76E-08 |
| LMTK3 | 66,97 | 0,86 | 0,16 | 5,47 | 4,47E-08 |
| RABEP1 | 2290,74 | -0,19 | 0,03 | -5,42 | 5,81E-08 |
| RTN2 | 97,75 | 0,45 | 0,08 | 5,41 | 6,28E-08 |
| MST1 | 57,85 | 0,47 | 0,09 | 5,46 | 4,86E-08 |
| HYAL3 | 45,01 | 0,42 | 0,08 | 5,43 | 5,63E-08 |
| CTR9 | 1686,99 | -0,16 | 0,03 | -5,35 | 8,93E-08 |
| CACNA1A | 119,18 | 1,59 | 0,30 | 5,34 | 9,46E-08 |
| TTF1 | 770,12 | -0,19 | 0,04 | -5,37 | 7,92E-08 |
| NRXN2 | 54,11 | 1,22 | 0,23 | 5,31 | 1,11E-07 |
| NR2F6 | 44,89 | 0,53 | 0,10 | 5,23 | 1,68E-07 |
| HSP90B1 | 2256,37 | -0,21 | 0,04 | -5,24 | 1,57E-07 |
| COL11A2 | 51,30 | 1,10 | 0,21 | 5,19 | 2,09E-07 |
| HSD17B1 | 32,14 | 0,61 | 0,12 | 5,18 | 2,24E-07 |
| ZNF136 | 952,87 | -0,17 | 0,03 | -5,18 | 2,19E-07 |
| FBXO24 | 39,69 | 0,54 | 0,11 | 5,08 | 3,71E-07 |
| DENND6B | 48,76 | 0,68 | 0,13 | 5,09 | 3,57E-07 |
| SMC3 | 1566,23 | -0,32 | 0,06 | -5,08 | 3,77E-07 |
| B4GALT3 | 382,17 | 0,35 | 0,07 | 5,14 | 2,77E-07 |
| FAM83H | 56,24 | 0,69 | 0,14 | 5,10 | 3,43E-07 |
| CYP4F22 | 25,00 | 0,89 | 0,17 | 5,12 | 3,14E-07 |
| HID1 | 34,71 | 0,92 | 0,18 | 5,09 | 3,59E-07 |
| ITGA7 | 51,22 | 0,89 | 0,17 | 5,10 | 3,48E-07 |
| RYR1 | 114,76 | 1,03 | 0,20 | 5,07 | 3,96E-07 |
| DLGAP1 | 48,79 | 1,13 | 0,22 | 5,05 | 4,44E-07 |
| ATP8B3 | 75,25 | 0,60 | 0,12 | 5,05 | 4,47E-07 |
| C17orf99 | 22,83 | 0,71 | 0,14 | 5,06 | 4,20E-07 |
| SMTN | 65,36 | 0,91 | 0,18 | 5,03 | 4,90E-07 |
| KNDC1 | 70,49 | 1,00 | 0,20 | 5,04 | 4,74E-07 |
| ENSG00000258472 | 24,89 | 0,75 | 0,15 | 5,02 | 5,08E-07 |
| WDR62 | 57,37 | 0,78 | 0,16 | 5,01 | 5,32E-07 |
| GOLGB1 | 4703,26 | -0,21 | 0,04 | -5,00 | 5,85E-07 |
| CLCN2 | 41,11 | 0,82 | 0,17 | 4,98 | 6,39E-07 |
| SLC7A5 | 199,50 | 0,59 | 0,12 | 4,98 | 6,50E-07 |
| GPRC5C | 36,12 | 1,05 | 0,21 | 4,98 | 6,40E-07 |
| WDR90 | 115,25 | 0,67 | 0,13 | 4,95 | 7,31E-07 |
| GOLGA4 | 3785,45 | -0,27 | 0,05 | -4,95 | 7,27E-07 |
| DECR2 | 41,89 | 0,55 | 0,11 | 4,95 | 7,48E-07 |
| CDC5L | 2154,20 | -0,25 | 0,05 | -4,92 | 8,61E-07 |
| ZNF703 | 52,50 | 0,71 | 0,14 | 4,93 | 8,07E-07 |
| PIP5KL1 | 36,49 | 0,59 | 0,12 | 4,93 | 8,08E-07 |
| SEMA6B | 39,53 | 0,85 | 0,17 | 4,92 | 8,60E-07 |
| PLXNB1 | 49,78 | 0,95 | 0,19 | 4,92 | 8,63E-07 |
| SEMA3G | 37,00 | 0,81 | 0,17 | 4,91 | 8,92E-07 |

| | | | | | |
|---|---|---|---|---|---|
| PLCD3 | 33,51 | 0,96 | 0,20 | 4,90 | 9,62E-07 |
| ZSCAN26 | 429,54 | -0,18 | 0,04 | -4,86 | 1,19E-06 |
| PDZD7 | 39,79 | 0,99 | 0,20 | 4,84 | 1,28E-06 |
| BAHCC1 | 86,33 | 0,94 | 0,19 | 4,85 | 1,26E-06 |
| CDC42BPG | 89,50 | 0,55 | 0,11 | 4,85 | 1,26E-06 |
| LIMS2 | 62,93 | 0,71 | 0,15 | 4,84 | 1,33E-06 |
| DNAH17 | 63,20 | 1,24 | 0,26 | 4,83 | 1,37E-06 |
| GLB1L3 | 44,25 | 1,04 | 0,22 | 4,82 | 1,47E-06 |
| ACHE | 35,60 | 0,83 | 0,17 | 4,80 | 1,55E-06 |
| CAND2 | 37,42 | 0,79 | 0,16 | 4,81 | 1,53E-06 |
| NHSL3 | 39,04 | 0,86 | 0,18 | 4,80 | 1,60E-06 |
| CDHR2 | 118,11 | 0,42 | 0,09 | 4,79 | 1,63E-06 |
| POLR2B | 4332,34 | -0,24 | 0,05 | -4,78 | 1,72E-06 |
| TMEM178B | 42,46 | 1,11 | 0,23 | 4,78 | 1,72E-06 |
| ZNF808 | 1067,76 | -0,20 | 0,04 | -4,79 | 1,71E-06 |
| ZNF397 | 2412,64 | -0,13 | 0,03 | -4,78 | 1,78E-06 |
| IDUA | 211,82 | 0,43 | 0,09 | 4,77 | 1,84E-06 |
| SLC39A4 | 57,17 | 0,47 | 0,10 | 4,75 | 1,99E-06 |
| COL4A1 | 62,42 | 1,21 | 0,25 | 4,76 | 1,97E-06 |
| ANKLE1 | 36,52 | 0,46 | 0,10 | 4,75 | 1,99E-06 |
| TNXB | 129,33 | 0,70 | 0,15 | 4,72 | 2,32E-06 |
| GPRC5B | 46,21 | 1,09 | 0,23 | 4,73 | 2,26E-06 |
| LRRC2 | 38,01 | 0,82 | 0,17 | 4,73 | 2,26E-06 |
| PTPN11 | 2253,99 | -0,14 | 0,03 | -4,73 | 2,25E-06 |
| ZNF579 | 35,02 | 0,70 | 0,15 | 4,73 | 2,23E-06 |
| ITGB4 | 52,95 | 0,87 | 0,18 | 4,73 | 2,29E-06 |
| TOP2B | 5668,50 | -0,25 | 0,05 | -4,75 | 2,08E-06 |
| DNM1 | 55,36 | 0,87 | 0,18 | 4,73 | 2,29E-06 |
| PROB1 | 30,49 | 0,86 | 0,18 | 4,72 | 2,32E-06 |
| LMNTD2 | 45,52 | 0,51 | 0,11 | 4,74 | 2,12E-06 |
| RAD21 | 4499,43 | -0,20 | 0,04 | -4,72 | 2,41E-06 |
| SLC9A5 | 37,45 | 0,71 | 0,15 | 4,71 | 2,52E-06 |
| GJC2 | 46,46 | 0,42 | 0,09 | 4,71 | 2,53E-06 |
| CRYBG2 | 58,09 | 0,55 | 0,12 | 4,71 | 2,51E-06 |
| ACOT11 | 110,31 | 0,65 | 0,14 | 4,70 | 2,58E-06 |
| DNTTIP2 | 900,58 | -0,34 | 0,07 | -4,70 | 2,60E-06 |
| TPR | 8562,40 | -0,21 | 0,05 | -4,70 | 2,64E-06 |
| FAM241B | 29,62 | 0,55 | 0,12 | 4,69 | 2,71E-06 |
| P3H3 | 38,85 | 0,60 | 0,13 | 4,66 | 3,16E-06 |
| RAB3IL1 | 77,33 | 0,64 | 0,14 | 4,68 | 2,90E-06 |
| CORO6 | 38,76 | 0,57 | 0,12 | 4,65 | 3,24E-06 |
| COL7A1 | 98,85 | 0,77 | 0,17 | 4,66 | 3,10E-06 |
| FASN | 186,46 | 0,64 | 0,14 | 4,66 | 3,20E-06 |
| CHPF | 53,30 | 0,51 | 0,11 | 4,66 | 3,15E-06 |
| C1S | 35,39 | 0,86 | 0,18 | 4,66 | 3,15E-06 |
| ARHGEF4 | 61,56 | 1,04 | 0,22 | 4,66 | 3,20E-06 |

| | | | | | |
|---|---|---|---|---|---|
| ZSCAN9 | 397,72 | -0,18 | 0,04 | -4,68 | 2,88E-06 |
| HTATSF1 | 819,98 | -0,26 | 0,06 | -4,66 | 3,22E-06 |
| BAG5 | 1494,27 | -0,17 | 0,04 | -4,67 | 3,03E-06 |
| C21orf58 | 84,57 | 0,41 | 0,09 | 4,68 | 2,90E-06 |
| R3HCC1L | 1475,44 | -0,20 | 0,04 | -4,68 | 2,88E-06 |
| TRIM46 | 54,09 | 0,58 | 0,12 | 4,66 | 3,15E-06 |
| LZTS3 | 34,76 | 0,95 | 0,20 | 4,67 | 3,03E-06 |
| RANBP2 | 5657,53 | -0,21 | 0,05 | -4,65 | 3,29E-06 |
| ETV3 | 1887,64 | -0,11 | 0,02 | -4,64 | 3,46E-06 |
| CWC22 | 1108,54 | -0,26 | 0,06 | -4,64 | 3,42E-06 |
| PITPNM3 | 63,15 | 1,02 | 0,22 | 4,65 | 3,33E-06 |
| EPRS1 | 2177,67 | -0,21 | 0,05 | -4,64 | 3,41E-06 |
| SLC5A2 | 33,99 | 0,83 | 0,18 | 4,64 | 3,45E-06 |
| TERF1 | 957,58 | -0,14 | 0,03 | -4,63 | 3,60E-06 |
| SKOR1 | 46,80 | 0,95 | 0,21 | 4,63 | 3,65E-06 |
| PRRT3 | 61,23 | 0,49 | 0,11 | 4,63 | 3,65E-06 |
| KCNC3 | 59,28 | 0,67 | 0,14 | 4,63 | 3,58E-06 |
| TFR2 | 94,75 | 0,53 | 0,11 | 4,63 | 3,65E-06 |
| SNW1 | 1511,60 | -0,20 | 0,04 | -4,62 | 3,83E-06 |
| LARP7 | 677,92 | -0,32 | 0,07 | -4,60 | 4,19E-06 |
| RFC1 | 1774,75 | -0,16 | 0,03 | -4,60 | 4,16E-06 |
| FAM83F | 76,20 | 0,85 | 0,18 | 4,60 | 4,14E-06 |
| FARP1 | 106,66 | 1,32 | 0,29 | 4,60 | 4,20E-06 |
| YPEL4 | 47,13 | 0,52 | 0,11 | 4,60 | 4,17E-06 |
| PTK6 | 40,93 | 0,56 | 0,12 | 4,61 | 4,09E-06 |
| CCNE1 | 40,20 | 0,52 | 0,11 | 4,62 | 3,84E-06 |
| TMEM161A | 112,96 | 0,28 | 0,06 | 4,62 | 3,86E-06 |
| SLC29A1 | 135,43 | 0,63 | 0,14 | 4,62 | 3,84E-06 |
| ADGRV1 | 107,35 | 1,11 | 0,24 | 4,61 | 4,06E-06 |
| TDRD12 | 42,19 | 0,99 | 0,21 | 4,62 | 3,77E-06 |
| PTDSS2 | 192,81 | 0,30 | 0,07 | 4,60 | 4,15E-06 |
| ASAP3 | 46,12 | 0,80 | 0,17 | 4,60 | 4,13E-06 |
| FBXL8 | 124,58 | 0,25 | 0,05 | 4,61 | 3,97E-06 |
| VARS2 | 224,60 | 0,36 | 0,08 | 4,61 | 4,08E-06 |
| E2F1 | 62,41 | 0,45 | 0,10 | 4,60 | 4,26E-06 |
| CEP350 | 7329,23 | -0,16 | 0,03 | -4,60 | 4,28E-06 |
| PROSER2 | 38,38 | 0,60 | 0,13 | 4,58 | 4,59E-06 |
| TREX2 | 26,44 | 0,52 | 0,11 | 4,59 | 4,53E-06 |
| NRL | 71,01 | 0,27 | 0,06 | 4,58 | 4,65E-06 |
| SMU1 | 2552,67 | -0,13 | 0,03 | -4,59 | 4,48E-06 |
| MYO1C | 192,85 | 0,42 | 0,09 | 4,58 | 4,60E-06 |
| MAGOH | 438,21 | -0,28 | 0,06 | -4,58 | 4,64E-06 |
| ADGRB2 | 57,41 | 1,01 | 0,22 | 4,58 | 4,68E-06 |
| MAP3K6 | 128,35 | 0,37 | 0,08 | 4,57 | 4,85E-06 |
| PARVA | 47,56 | 0,96 | 0,21 | 4,57 | 4,85E-06 |
| TONSL | 73,90 | 0,59 | 0,13 | 4,57 | 4,79E-06 |

| | | | | | |
|---|---|---|---|---|---|
| GFUS | 747,41 | 0,43 | 0,09 | 4,57 | 4,82E-06 |
| CES4A | 146,45 | 0,29 | 0,06 | 4,57 | 4,92E-06 |
| NOS3 | 53,78 | 0,68 | 0,15 | 4,57 | 4,89E-06 |
| PLEKHA7 | 105,51 | 0,64 | 0,14 | 4,56 | 5,04E-06 |
| CBX1 | 1471,23 | -0,22 | 0,05 | -4,56 | 5,09E-06 |
| HMBS | 207,77 | 0,41 | 0,09 | 4,56 | 5,22E-06 |
| JAKMIP3 | 54,81 | 0,86 | 0,19 | 4,56 | 5,19E-06 |
| FLT3LG | 81,80 | 0,51 | 0,11 | 4,54 | 5,50E-06 |
| SLC5A9 | 35,25 | 0,75 | 0,16 | 4,55 | 5,44E-06 |
| HSD3B7 | 32,17 | 0,50 | 0,11 | 4,55 | 5,34E-06 |
| ALDH7A1 | 57,62 | 0,89 | 0,20 | 4,55 | 5,31E-06 |
| GNA11 | 63,72 | 0,58 | 0,13 | 4,55 | 5,42E-06 |
| KIFC1 | 35,19 | 0,62 | 0,14 | 4,55 | 5,48E-06 |
| PLEKHG5 | 96,70 | 0,74 | 0,16 | 4,54 | 5,52E-06 |
| DNMT3B | 43,76 | 0,83 | 0,18 | 4,55 | 5,39E-06 |
| MAGI2 | 65,11 | 1,03 | 0,23 | 4,54 | 5,62E-06 |
| CRNKL1 | 2106,61 | -0,21 | 0,05 | -4,54 | 5,67E-06 |
| TRIM47 | 44,09 | 0,43 | 0,10 | 4,52 | 6,17E-06 |
| RHPN1 | 108,24 | 0,44 | 0,10 | 4,52 | 6,19E-06 |
| ZC3H13 | 2755,05 | -0,15 | 0,03 | -4,52 | 6,19E-06 |
| PDE2A | 70,86 | 0,76 | 0,17 | 4,52 | 6,13E-06 |
| ZNF700 | 882,92 | -0,18 | 0,04 | -4,53 | 5,80E-06 |
| HSP90AA1 | 3236,47 | -0,41 | 0,09 | -4,52 | 6,17E-06 |
| LIPE | 81,60 | 0,42 | 0,09 | 4,53 | 5,94E-06 |
| ECSIT | 241,29 | 0,28 | 0,06 | 4,53 | 5,89E-06 |
| GGT5 | 30,76 | 0,90 | 0,20 | 4,53 | 5,84E-06 |
| WDR48 | 2198,14 | -0,15 | 0,03 | -4,52 | 6,08E-06 |
| XRCC5 | 8599,47 | -0,20 | 0,04 | -4,53 | 6,02E-06 |
| TRPM4 | 48,86 | 0,86 | 0,19 | 4,53 | 5,98E-06 |
| ZNF146 | 1199,34 | -0,30 | 0,07 | -4,53 | 6,00E-06 |
| PHF3 | 6902,81 | -0,23 | 0,05 | -4,52 | 6,24E-06 |
| LRP5 | 70,08 | 0,78 | 0,17 | 4,50 | 6,64E-06 |
| FAM20C | 100,62 | 0,56 | 0,13 | 4,51 | 6,63E-06 |
| RRP36 | 707,87 | -0,11 | 0,02 | -4,50 | 6,70E-06 |
| USP16 | 1742,13 | -0,25 | 0,05 | -4,50 | 6,73E-06 |
| MAPK10 | 90,80 | 1,11 | 0,25 | 4,50 | 6,71E-06 |
| DNAH10 | 122,27 | 0,92 | 0,20 | 4,50 | 6,67E-06 |
| SERINC2 | 47,27 | 0,89 | 0,20 | 4,51 | 6,51E-06 |
| LRFN3 | 34,09 | 0,72 | 0,16 | 4,50 | 6,93E-06 |
| DAGLA | 52,54 | 0,72 | 0,16 | 4,49 | 6,96E-06 |
| EIF4B | 14567,29 | -0,21 | 0,05 | -4,50 | 6,95E-06 |
| SMARCA5 | 3010,09 | -0,27 | 0,06 | -4,50 | 6,94E-06 |
| CERCAM | 34,93 | 0,90 | 0,20 | 4,50 | 6,89E-06 |
| MGLL | 195,02 | 0,47 | 0,10 | 4,49 | 6,99E-06 |
| CCDC157 | 78,88 | 0,40 | 0,09 | 4,49 | 7,20E-06 |
| CLBA1 | 48,97 | 0,48 | 0,11 | 4,49 | 7,09E-06 |

| | | | | | |
|---|---|---|---|---|---|
| WDR31 | 35,58 | 0,60 | 0,13 | 4,49 | 7,22E-06 |
| COL8A2 | 39,58 | 0,58 | 0,13 | 4,49 | 7,16E-06 |
| SEPTIN7 | 6843,13 | -0,25 | 0,05 | -4,49 | 7,23E-06 |
| PDCD4 | 7005,18 | -0,20 | 0,05 | -4,48 | 7,49E-06 |
| CCDC154 | 58,66 | 0,69 | 0,15 | 4,48 | 7,57E-06 |
| NOL3 | 61,01 | 0,40 | 0,09 | 4,47 | 7,69E-06 |
| KCNK12 | 35,78 | 0,78 | 0,17 | 4,47 | 7,76E-06 |
| ADGRB1 | 96,53 | 0,92 | 0,20 | 4,47 | 7,85E-06 |
| PHKG1 | 41,69 | 0,42 | 0,09 | 4,46 | 8,14E-06 |
| ACE | 57,43 | 0,71 | 0,16 | 4,46 | 8,12E-06 |
| MOAP1 | 1001,26 | -0,16 | 0,04 | -4,46 | 8,19E-06 |
| FLT4 | 114,78 | 0,67 | 0,15 | 4,46 | 8,35E-06 |
| KLHDC8A | 28,91 | 0,78 | 0,18 | 4,45 | 8,46E-06 |
| RAPGEF3 | 48,26 | 1,10 | 0,25 | 4,45 | 8,42E-06 |
| KLHDC7B | 55,58 | 0,53 | 0,12 | 4,45 | 8,63E-06 |
| RTEL1 | 77,79 | 0,31 | 0,07 | 4,45 | 8,76E-06 |
| SLC22A23 | 240,00 | 0,41 | 0,09 | 4,45 | 8,77E-06 |
| AEBP1 | 54,14 | 0,74 | 0,17 | 4,44 | 8,82E-06 |
| SCARF2 | 34,51 | 0,75 | 0,17 | 4,44 | 8,97E-06 |
| ZSCAN32 | 773,13 | -0,10 | 0,02 | -4,44 | 8,92E-06 |
| AGBL5 | 118,53 | 0,34 | 0,08 | 4,44 | 8,98E-06 |
| ZNF462 | 59,28 | 0,97 | 0,22 | 4,43 | 9,25E-06 |
| ADCY6 | 44,41 | 0,93 | 0,21 | 4,43 | 9,24E-06 |
| EDA | 94,06 | 0,51 | 0,12 | 4,43 | 9,51E-06 |
| RNF215 | 152,77 | 0,31 | 0,07 | 4,43 | 9,51E-06 |
| SET | 5510,91 | -0,24 | 0,05 | -4,43 | 9,52E-06 |
| SLC48A1 | 356,90 | 0,45 | 0,10 | 4,42 | 9,74E-06 |
| TNK1 | 32,24 | 0,54 | 0,12 | 4,42 | 9,72E-06 |
| TYMS | 34,15 | 0,71 | 0,16 | 4,42 | 9,67E-06 |
| SNAPC2 | 60,00 | 0,32 | 0,07 | 4,42 | 9,77E-06 |
| RPP25 | 26,22 | 0,51 | 0,12 | 4,42 | 9,96E-06 |
| MCOLN1 | 340,69 | 0,40 | 0,09 | 4,42 | 9,97E-06 |
| BCL2L15 | 102,79 | 0,28 | 0,06 | 4,42 | 9,94E-06 |
| PC | 84,11 | 0,55 | 0,13 | 4,42 | 1,01E-05 |
| DNAJC8 | 1198,97 | -0,24 | 0,05 | -4,41 | 1,02E-05 |
| SLU7 | 1952,02 | -0,27 | 0,06 | -4,41 | 1,02E-05 |
| NR1D1 | 33,62 | 0,52 | 0,12 | 4,41 | 1,04E-05 |
| PARG | 1228,27 | -0,16 | 0,04 | -4,41 | 1,06E-05 |
| CCDC85C | 110,21 | 0,63 | 0,14 | 4,40 | 1,07E-05 |
| RUSC1 | 82,37 | 0,37 | 0,08 | 4,40 | 1,08E-05 |
| TGS1 | 959,39 | -0,14 | 0,03 | -4,40 | 1,08E-05 |
| SEMA6C | 46,29 | 0,69 | 0,16 | 4,40 | 1,08E-05 |
| KIF2C | 34,60 | 0,65 | 0,15 | 4,40 | 1,09E-05 |
| NBL1 | 59,48 | 0,60 | 0,14 | 4,40 | 1,10E-05 |
| METTL14 | 1084,99 | -0,20 | 0,05 | -4,39 | 1,12E-05 |
| SHARPIN | 570,98 | 0,32 | 0,07 | 4,39 | 1,11E-05 |

| | | | | | |
|---|---|---|---|---|---|
| TF | 70,53 | 1,09 | 0,25 | 4,39 | 1,12E-05 |
| ZSWIM4 | 81,15 | 0,47 | 0,11 | 4,39 | 1,16E-05 |
| ZNF628 | 61,60 | 0,54 | 0,12 | 4,38 | 1,17E-05 |
| HSPA4 | 2076,09 | -0,19 | 0,04 | -4,38 | 1,16E-05 |
| NAV2 | 85,13 | 0,97 | 0,22 | 4,39 | 1,15E-05 |
| KATNA1 | 383,64 | -0,13 | 0,03 | -4,38 | 1,17E-05 |
| CYB5R3 | 302,31 | 0,39 | 0,09 | 4,39 | 1,16E-05 |
| TXN2 | 152,79 | 0,23 | 0,05 | 4,38 | 1,20E-05 |
| HECTD1 | 6056,83 | -0,15 | 0,03 | -4,38 | 1,19E-05 |
| FHDC1 | 96,55 | 0,47 | 0,11 | 4,38 | 1,19E-05 |
| GAS6 | 123,08 | 0,48 | 0,11 | 4,38 | 1,20E-05 |
| UPF2 | 3022,50 | -0,19 | 0,04 | -4,38 | 1,21E-05 |
| MYRF | 47,83 | 1,03 | 0,24 | 4,38 | 1,21E-05 |
| ZNF219 | 71,68 | 0,53 | 0,12 | 4,37 | 1,22E-05 |
| TANC1 | 61,02 | 0,85 | 0,19 | 4,37 | 1,24E-05 |
| KMT5C | 132,29 | 0,36 | 0,08 | 4,37 | 1,24E-05 |
| MGAT3 | 109,77 | 0,45 | 0,10 | 4,37 | 1,25E-05 |
| MT-ND6 | 202,61 | 0,83 | 0,19 | 4,37 | 1,25E-05 |
| CDCA3 | 26,84 | 0,55 | 0,13 | 4,37 | 1,27E-05 |
| RECQL4 | 42,75 | 0,61 | 0,14 | 4,37 | 1,26E-05 |
| CCDC61 | 116,52 | 0,35 | 0,08 | 4,37 | 1,27E-05 |
| AKAP9 | 4924,66 | -0,20 | 0,05 | -4,36 | 1,28E-05 |
| MPHOSPH10 | 646,06 | -0,21 | 0,05 | -4,36 | 1,28E-05 |
| KCNH2 | 97,61 | 0,57 | 0,13 | 4,36 | 1,29E-05 |
| EAPP | 1004,95 | -0,25 | 0,06 | -4,36 | 1,29E-05 |
| NCL | 3818,98 | -0,19 | 0,04 | -4,36 | 1,31E-05 |
| PDHB | 1104,47 | -0,14 | 0,03 | -4,36 | 1,31E-05 |
| C3 | 49,17 | 0,71 | 0,16 | 4,36 | 1,31E-05 |
| GPSM1 | 103,91 | 0,47 | 0,11 | 4,35 | 1,34E-05 |
| ADCY3 | 212,49 | 0,34 | 0,08 | 4,35 | 1,33E-05 |
| EGFL7 | 34,36 | 0,57 | 0,13 | 4,35 | 1,34E-05 |
| MFSD13A | 113,66 | 0,35 | 0,08 | 4,35 | 1,34E-05 |
| PBRM1 | 3468,10 | -0,12 | 0,03 | -4,36 | 1,33E-05 |
| PLXNA1 | 215,65 | 0,54 | 0,12 | 4,35 | 1,37E-05 |
| CKB | 65,24 | 0,42 | 0,10 | 4,35 | 1,36E-05 |
| IL4I1 | 28,64 | 0,61 | 0,14 | 4,35 | 1,36E-05 |
| ZNF414 | 52,26 | 0,51 | 0,12 | 4,35 | 1,37E-05 |
| RNASE3 | 35,13 | 0,74 | 0,17 | 4,35 | 1,38E-05 |
| ATP5F1D | 90,95 | 0,44 | 0,10 | 4,35 | 1,39E-05 |
| ATMIN | 1268,59 | -0,12 | 0,03 | -4,35 | 1,39E-05 |
| AMN | 60,57 | 0,67 | 0,15 | 4,34 | 1,42E-05 |
| UGP2 | 2445,48 | -0,15 | 0,03 | -4,34 | 1,43E-05 |
| DNAH12 | 65,74 | 0,94 | 0,22 | 4,34 | 1,44E-05 |
| TRMT9B | 45,93 | 1,00 | 0,23 | 4,34 | 1,45E-05 |
| CACNB3 | 74,38 | 0,49 | 0,11 | 4,34 | 1,44E-05 |
| COL5A2 | 49,34 | 0,62 | 0,14 | 4,34 | 1,43E-05 |

| | | | | | |
|---|---|---|---|---|---|
| SNAPC5 | 435,48 | -0,15 | 0,04 | -4,34 | 1,44E-05 |
| CELSR3 | 112,71 | 0,54 | 0,12 | 4,34 | 1,45E-05 |
| TRIB3 | 104,51 | 0,36 | 0,08 | 4,33 | 1,46E-05 |
| PHF7 | 185,48 | 0,14 | 0,03 | 4,33 | 1,48E-05 |
| ABCB9 | 94,40 | 0,53 | 0,12 | 4,33 | 1,52E-05 |
| BMAL2 | 67,89 | 0,50 | 0,11 | 4,32 | 1,55E-05 |
| CCDC47 | 1375,33 | -0,15 | 0,03 | -4,32 | 1,56E-05 |
| KLHL29 | 76,18 | 0,82 | 0,19 | 4,32 | 1,57E-05 |
| PKN3 | 30,82 | 0,78 | 0,18 | 4,32 | 1,59E-05 |
| NFIX | 309,29 | 0,50 | 0,12 | 4,32 | 1,60E-05 |
| GPR162 | 136,04 | 0,44 | 0,10 | 4,31 | 1,60E-05 |
| DDR1 | 72,14 | 0,66 | 0,15 | 4,31 | 1,62E-05 |
| CABYR | 36,45 | 0,60 | 0,14 | 4,31 | 1,62E-05 |
| ZNF182 | 365,17 | -0,14 | 0,03 | -4,31 | 1,62E-05 |
| NEMF | 2727,68 | -0,15 | 0,03 | -4,31 | 1,65E-05 |
| KLHL35 | 33,87 | 0,74 | 0,17 | 4,31 | 1,64E-05 |
| SLC12A3 | 35,84 | 0,81 | 0,19 | 4,31 | 1,64E-05 |
| RAB15 | 43,63 | 0,53 | 0,12 | 4,31 | 1,66E-05 |
| ZNRF1 | 155,76 | 0,36 | 0,08 | 4,30 | 1,68E-05 |
| DENR | 1147,69 | -0,18 | 0,04 | -4,30 | 1,69E-05 |
| ZNF350 | 1045,78 | -0,21 | 0,05 | -4,30 | 1,73E-05 |
| SIAE | 184,31 | 0,26 | 0,06 | 4,30 | 1,72E-05 |
| MEIS2 | 47,31 | 0,97 | 0,22 | 4,30 | 1,72E-05 |
| ARHGAP33 | 55,68 | 0,63 | 0,15 | 4,30 | 1,71E-05 |
| NECAB3 | 126,07 | 0,31 | 0,07 | 4,30 | 1,73E-05 |
| SMAD6 | 63,78 | 0,86 | 0,20 | 4,30 | 1,73E-05 |
| NPTXR | 38,40 | 0,70 | 0,16 | 4,30 | 1,72E-05 |
| PSD | 55,73 | 0,65 | 0,15 | 4,29 | 1,77E-05 |
| ADISSP | 150,81 | 0,34 | 0,08 | 4,29 | 1,79E-05 |
| RAD23A | 316,46 | 0,45 | 0,11 | 4,29 | 1,79E-05 |
| RAPGEF4 | 43,31 | 0,90 | 0,21 | 4,29 | 1,81E-05 |
| TRIP6 | 127,49 | 0,29 | 0,07 | 4,29 | 1,81E-05 |
| MICALL2 | 78,00 | 0,67 | 0,16 | 4,29 | 1,83E-05 |
| DDX18 | 2204,51 | -0,18 | 0,04 | -4,28 | 1,84E-05 |
| PTPRF | 98,99 | 0,79 | 0,19 | 4,28 | 1,84E-05 |
| MROH6 | 80,41 | 0,50 | 0,12 | 4,28 | 1,87E-05 |
| SDK1 | 84,43 | 0,97 | 0,23 | 4,27 | 1,92E-05 |
| PDZK1IP1 | 663,87 | 0,50 | 0,12 | 4,27 | 1,93E-05 |
| FRS3 | 75,89 | 0,32 | 0,07 | 4,27 | 1,93E-05 |
| PPFIA3 | 55,94 | 0,73 | 0,17 | 4,27 | 1,95E-05 |
| DLGAP3 | 54,20 | 0,90 | 0,21 | 4,27 | 1,97E-05 |
| FAM20A | 35,51 | 0,80 | 0,19 | 4,27 | 1,98E-05 |
| SYN3 | 45,78 | 0,97 | 0,23 | 4,27 | 1,96E-05 |
| TNKS1BP1 | 117,28 | 0,50 | 0,12 | 4,27 | 1,98E-05 |
| UAP1 | 466,93 | -0,21 | 0,05 | -4,27 | 1,94E-05 |
| ITGA3 | 99,05 | 0,51 | 0,12 | 4,27 | 1,97E-05 |

| | | | | | |
|---|---|---|---|---|---|
| DNAJC21 | 1219,62 | -0,17 | 0,04 | -4,27 | 1,96E-05 |
| SLC8A3 | 57,87 | 0,82 | 0,19 | 4,27 | 2,00E-05 |
| MYO7A | 118,08 | 0,82 | 0,19 | 4,27 | 1,99E-05 |
| GOLGA7B | 233,62 | 0,27 | 0,06 | 4,26 | 2,01E-05 |
| E2F7 | 36,96 | 0,69 | 0,16 | 4,26 | 2,05E-05 |
| TSPO2 | 64,60 | 0,60 | 0,14 | 4,26 | 2,05E-05 |
| MLH1 | 995,86 | -0,12 | 0,03 | -4,26 | 2,04E-05 |
| CEBPZ | 1000,20 | -0,33 | 0,08 | -4,26 | 2,05E-05 |
| MARVELD2 | 30,25 | 0,51 | 0,12 | 4,26 | 2,04E-05 |
| PDIA6 | 2194,42 | -0,18 | 0,04 | -4,25 | 2,14E-05 |
| ELF3 | 42,17 | 0,61 | 0,14 | 4,25 | 2,16E-05 |
| NKD1 | 112,01 | 0,60 | 0,14 | 4,25 | 2,15E-05 |
| HNRNPH2 | 2479,38 | -0,32 | 0,08 | -4,25 | 2,15E-05 |
| SH3PXD2B | 45,53 | 0,82 | 0,19 | 4,25 | 2,14E-05 |
| STRIP2 | 55,10 | 0,56 | 0,13 | 4,25 | 2,14E-05 |
| LRRC56 | 47,58 | 0,46 | 0,11 | 4,25 | 2,16E-05 |
| ADAM32 | 43,19 | 0,80 | 0,19 | 4,24 | 2,19E-05 |
| EPS8L2 | 90,20 | 0,36 | 0,09 | 4,24 | 2,19E-05 |
| RSF1 | 3481,45 | -0,15 | 0,04 | -4,24 | 2,19E-05 |
| DLC1 | 48,96 | 0,86 | 0,20 | 4,24 | 2,22E-05 |
| NFIB | 46,36 | 0,89 | 0,21 | 4,24 | 2,21E-05 |
| USP8 | 3625,37 | -0,12 | 0,03 | -4,24 | 2,22E-05 |
| PLLP | 32,76 | 0,72 | 0,17 | 4,23 | 2,32E-05 |
| LTV1 | 222,32 | -0,21 | 0,05 | -4,23 | 2,33E-05 |
| NT5DC4 | 38,11 | 0,59 | 0,14 | 4,23 | 2,30E-05 |
| PTK7 | 67,05 | 0,54 | 0,13 | 4,23 | 2,31E-05 |
| SDCBP2 | 36,15 | 0,46 | 0,11 | 4,24 | 2,27E-05 |
| BICRA | 144,90 | 0,63 | 0,15 | 4,23 | 2,32E-05 |
| LAMB3 | 42,09 | 0,74 | 0,18 | 4,24 | 2,26E-05 |
| INTU | 56,96 | 0,87 | 0,20 | 4,23 | 2,30E-05 |
| ZNF160 | 1480,72 | -0,12 | 0,03 | -4,24 | 2,29E-05 |
| CC2D2A | 51,78 | 0,68 | 0,16 | 4,23 | 2,31E-05 |
| INIP | 961,62 | -0,12 | 0,03 | -4,24 | 2,28E-05 |
| PIGQ | 215,89 | 0,38 | 0,09 | 4,24 | 2,27E-05 |
| CFAP251 | 78,10 | 0,58 | 0,14 | 4,23 | 2,31E-05 |
| BAIAP2 | 116,61 | 0,64 | 0,15 | 4,23 | 2,32E-05 |
| RHCE | 61,32 | 0,58 | 0,14 | 4,23 | 2,33E-05 |
| MAMLD1 | 41,57 | 0,55 | 0,13 | 4,23 | 2,38E-05 |
| ENAH | 48,91 | 0,90 | 0,21 | 4,23 | 2,38E-05 |
| TRPV1 | 72,03 | 0,43 | 0,10 | 4,22 | 2,39E-05 |
| GNPTAB | 2571,13 | -0,16 | 0,04 | -4,22 | 2,41E-05 |
| C1R | 36,89 | 0,56 | 0,13 | 4,22 | 2,42E-05 |
| RYR3 | 144,79 | 1,01 | 0,24 | 4,22 | 2,50E-05 |
| TTC7B | 213,16 | 0,52 | 0,12 | 4,22 | 2,49E-05 |
| MYO1A | 37,04 | 0,62 | 0,15 | 4,22 | 2,50E-05 |
| ABLIM3 | 90,41 | 0,64 | 0,15 | 4,21 | 2,52E-05 |

| | | | | | |
|---|---|---|---|---|---|
| CWF19L2 | 1618,98 | -0,27 | 0,06 | -4,21 | 2,55E-05 |
| ZNF134 | 846,56 | -0,16 | 0,04 | -4,21 | 2,59E-05 |
| SRP54 | 1555,22 | -0,17 | 0,04 | -4,20 | 2,62E-05 |
| NAALADL2 | 30,82 | 0,67 | 0,16 | 4,20 | 2,65E-05 |
| SSB | 793,80 | -0,32 | 0,08 | -4,20 | 2,69E-05 |
| SYF2 | 1668,28 | -0,28 | 0,07 | -4,20 | 2,71E-05 |
| ZFX | 3545,12 | -0,13 | 0,03 | -4,20 | 2,70E-05 |
| CHST13 | 45,06 | 0,62 | 0,15 | 4,20 | 2,73E-05 |
| GNL2 | 716,81 | -0,12 | 0,03 | -4,19 | 2,77E-05 |
| MED1 | 1646,19 | -0,07 | 0,02 | -4,19 | 2,78E-05 |
| PWP1 | 827,35 | -0,18 | 0,04 | -4,19 | 2,81E-05 |
| SMC6 | 1449,22 | -0,25 | 0,06 | -4,19 | 2,82E-05 |
| CRACD | 52,73 | 0,86 | 0,20 | 4,18 | 2,86E-05 |
| HES6 | 63,79 | 0,42 | 0,10 | 4,18 | 2,86E-05 |
| RAB9A | 619,29 | -0,17 | 0,04 | -4,18 | 2,87E-05 |
| DDX50 | 878,07 | -0,23 | 0,06 | -4,18 | 2,88E-05 |
| KALRN | 201,96 | 0,62 | 0,15 | 4,18 | 2,91E-05 |
| STARD10 | 130,35 | 0,36 | 0,09 | 4,18 | 2,90E-05 |
| FAM53A | 43,14 | 0,61 | 0,14 | 4,18 | 2,91E-05 |
| RIPOR3 | 104,64 | 0,52 | 0,12 | 4,18 | 2,92E-05 |
| KCNA2 | 126,56 | 0,52 | 0,12 | 4,18 | 2,93E-05 |
| LRFN4 | 60,86 | 0,50 | 0,12 | 4,17 | 2,99E-05 |
| PDZD2 | 80,86 | 0,81 | 0,19 | 4,17 | 2,98E-05 |
| KCNG1 | 65,15 | 0,76 | 0,18 | 4,17 | 3,03E-05 |
| IGSF11 | 36,60 | 0,81 | 0,19 | 4,17 | 3,03E-05 |
| AGAP3 | 286,37 | 0,42 | 0,10 | 4,17 | 3,02E-05 |
| USP6 | 54,41 | 0,85 | 0,20 | 4,17 | 3,01E-05 |
| ADNP | 3278,58 | -0,12 | 0,03 | -4,17 | 3,05E-05 |
| SLC22A17 | 38,95 | 0,57 | 0,14 | 4,17 | 3,05E-05 |
| TTC39A | 78,52 | 0,75 | 0,18 | 4,17 | 3,11E-05 |
| UTP14A | 376,88 | -0,15 | 0,04 | -4,17 | 3,11E-05 |
| SDC3 | 39,50 | 0,90 | 0,22 | 4,17 | 3,11E-05 |
| MPST | 453,33 | 0,21 | 0,05 | 4,16 | 3,13E-05 |
| GNG2 | 3251,79 | -0,17 | 0,04 | -4,16 | 3,13E-05 |
| DNLZ | 29,68 | 0,39 | 0,09 | 4,16 | 3,14E-05 |
| HBQ1 | 57,53 | 0,56 | 0,13 | 4,16 | 3,17E-05 |
| NUDT17 | 73,98 | 0,21 | 0,05 | 4,16 | 3,16E-05 |
| BIRC5 | 33,67 | 0,56 | 0,13 | 4,16 | 3,20E-05 |
| MYO18B | 81,62 | 1,18 | 0,28 | 4,16 | 3,23E-05 |
| PEAR1 | 55,92 | 0,79 | 0,19 | 4,16 | 3,23E-05 |
| ENTREP3 | 47,54 | 0,55 | 0,13 | 4,15 | 3,29E-05 |
| ILDR2 | 52,07 | 0,90 | 0,22 | 4,15 | 3,33E-05 |
| ITGA10 | 93,40 | 0,39 | 0,09 | 4,15 | 3,34E-05 |
| SORBS1 | 84,29 | 0,60 | 0,14 | 4,15 | 3,34E-05 |
| ARMC5 | 99,84 | 0,36 | 0,09 | 4,15 | 3,31E-05 |
| RSRC2 | 3359,11 | -0,17 | 0,04 | -4,15 | 3,31E-05 |

| | | | | | |
|---|---|---|---|---|---|
| MFSD6L | 71,82 | 0,44 | 0,11 | 4,15 | 3,32E-05 |
| DLL1 | 59,22 | 0,57 | 0,14 | 4,15 | 3,35E-05 |
| DNAH8 | 103,22 | 0,84 | 0,20 | 4,15 | 3,36E-05 |
| LSM12 | 205,25 | 0,13 | 0,03 | 4,15 | 3,37E-05 |
| BOP1 | 99,06 | 0,35 | 0,08 | 4,14 | 3,41E-05 |
| MATN2 | 39,94 | 0,91 | 0,22 | 4,14 | 3,40E-05 |
| C19orf47 | 87,67 | 0,43 | 0,10 | 4,14 | 3,41E-05 |
| CENPM | 28,95 | 0,48 | 0,11 | 4,14 | 3,43E-05 |
| HMG20A | 1100,44 | -0,12 | 0,03 | -4,14 | 3,47E-05 |
| PPIL4 | 1140,95 | -0,21 | 0,05 | -4,14 | 3,46E-05 |
| DNAH14 | 74,23 | 0,94 | 0,23 | 4,14 | 3,47E-05 |
| PAPLN | 65,62 | 0,70 | 0,17 | 4,14 | 3,45E-05 |
| PRPF38A | 1439,05 | -0,12 | 0,03 | -4,14 | 3,49E-05 |
| ZNF511 | 98,97 | 0,20 | 0,05 | 4,14 | 3,48E-05 |
| NT5DC2 | 38,09 | 0,65 | 0,16 | 4,14 | 3,50E-05 |
| EPS8L1 | 63,03 | 0,69 | 0,17 | 4,14 | 3,52E-05 |
| SECISBP2L | 2393,18 | -0,13 | 0,03 | -4,14 | 3,53E-05 |
| GPC2 | 107,82 | 0,28 | 0,07 | 4,14 | 3,54E-05 |
| COPB2 | 3347,56 | -0,15 | 0,04 | -4,14 | 3,55E-05 |
| MYO7B | 130,48 | 0,49 | 0,12 | 4,13 | 3,59E-05 |
| ZNRF3 | 54,32 | 0,62 | 0,15 | 4,13 | 3,58E-05 |
| PPFIA4 | 91,32 | 0,64 | 0,15 | 4,13 | 3,61E-05 |
| ABCA3 | 105,33 | 0,49 | 0,12 | 4,13 | 3,60E-05 |
| TBC1D24 | 113,07 | 0,42 | 0,10 | 4,13 | 3,60E-05 |
| NUP133 | 1464,55 | -0,10 | 0,02 | -4,13 | 3,62E-05 |
| PUSL1 | 98,26 | 0,25 | 0,06 | 4,13 | 3,69E-05 |
| NPR3 | 41,45 | 0,77 | 0,19 | 4,13 | 3,68E-05 |
| PSMD9 | 318,85 | 0,14 | 0,03 | 4,13 | 3,69E-05 |
| SLC2A5 | 86,57 | 0,48 | 0,12 | 4,13 | 3,68E-05 |
| BMP8B | 55,92 | 0,48 | 0,12 | 4,13 | 3,68E-05 |
| EFEMP2 | 98,37 | 0,27 | 0,07 | 4,13 | 3,70E-05 |
| BOD1L1 | 14216,34 | -0,21 | 0,05 | -4,12 | 3,74E-05 |
| GIPR | 94,72 | 0,40 | 0,10 | 4,12 | 3,76E-05 |
| RELL2 | 241,42 | 0,21 | 0,05 | 4,12 | 3,78E-05 |
| SH2D3A | 137,33 | 0,43 | 0,11 | 4,12 | 3,78E-05 |
| CYP2E1 | 50,32 | 0,86 | 0,21 | 4,12 | 3,85E-05 |
| TECTA | 69,76 | 0,62 | 0,15 | 4,11 | 3,88E-05 |
| CPS1 | 41,12 | 0,80 | 0,19 | 4,11 | 3,88E-05 |
| ENSG00000284976 | 121,41 | 0,37 | 0,09 | 4,11 | 3,90E-05 |
| ZNF385D | 40,62 | 0,69 | 0,17 | 4,11 | 3,96E-05 |
| PTGIR | 177,12 | 0,30 | 0,07 | 4,11 | 4,00E-05 |
| SDAD1 | 933,71 | -0,18 | 0,04 | -4,11 | 3,94E-05 |
| SPTBN5 | 161,05 | 0,61 | 0,15 | 4,11 | 3,98E-05 |
| AK4 | 51,72 | 0,56 | 0,14 | 4,11 | 3,96E-05 |
| CCDC124 | 135,53 | 0,28 | 0,07 | 4,11 | 3,95E-05 |
| NUMBL | 78,96 | 0,46 | 0,11 | 4,11 | 3,99E-05 |

| | | | | | |
|---|---|---|---|---|---|
| SCNN1D | 61,26 | 0,47 | 0,11 | 4,11 | 3,98E-05 |
| ETF1 | 1997,29 | -0,15 | 0,04 | -4,11 | 3,97E-05 |
| STK32B | 48,56 | 0,85 | 0,21 | 4,11 | 3,94E-05 |
| DIPK1B | 43,08 | 0,52 | 0,13 | 4,11 | 4,02E-05 |
| ADSS1 | 43,62 | 0,55 | 0,13 | 4,11 | 4,03E-05 |
| PARP3 | 68,29 | 0,34 | 0,08 | 4,10 | 4,08E-05 |
| ZNF44 | 1234,24 | -0,16 | 0,04 | -4,10 | 4,08E-05 |
| TLK2 | 2471,85 | -0,10 | 0,02 | -4,10 | 4,09E-05 |
| HSPA9 | 2706,01 | -0,15 | 0,04 | -4,10 | 4,08E-05 |
| RIN1 | 83,47 | 0,39 | 0,10 | 4,10 | 4,08E-05 |
| PRX | 93,05 | 0,35 | 0,09 | 4,10 | 4,06E-05 |
| ATRX | 6566,28 | -0,21 | 0,05 | -4,10 | 4,10E-05 |
| IGF2BP3 | 112,29 | 0,46 | 0,11 | 4,10 | 4,10E-05 |
| BBS5 | 27,44 | 0,63 | 0,15 | 4,10 | 4,13E-05 |
| LEO1 | 513,03 | -0,16 | 0,04 | -4,10 | 4,13E-05 |
| GOLGA6L10 | 46,14 | 0,69 | 0,17 | 4,10 | 4,16E-05 |
| MAPK11 | 62,61 | 0,35 | 0,09 | 4,10 | 4,16E-05 |
| PTH2R | 54,98 | 0,39 | 0,10 | 4,10 | 4,15E-05 |
| DBNDD2 | 52,33 | 0,38 | 0,09 | 4,09 | 4,26E-05 |
| ATXN7L2 | 55,70 | 0,50 | 0,12 | 4,09 | 4,29E-05 |
| COL17A1 | 50,70 | 0,93 | 0,23 | 4,09 | 4,34E-05 |
| SORBS3 | 167,36 | 0,39 | 0,10 | 4,09 | 4,35E-05 |
| STAG3 | 44,99 | 0,54 | 0,13 | 4,09 | 4,38E-05 |
| SGPP2 | 39,64 | 0,48 | 0,12 | 4,08 | 4,42E-05 |
| LRPPRC | 2129,57 | -0,17 | 0,04 | -4,08 | 4,48E-05 |
| HPS1 | 1789,24 | 0,27 | 0,07 | 4,08 | 4,45E-05 |
| GOLGA5 | 1049,25 | -0,13 | 0,03 | -4,08 | 4,47E-05 |
| COL6A3 | 159,59 | 0,62 | 0,15 | 4,08 | 4,48E-05 |
| PLEKHD1 | 43,10 | 0,88 | 0,22 | 4,08 | 4,48E-05 |
| CDV3 | 6116,64 | -0,18 | 0,04 | -4,08 | 4,50E-05 |
| PIN4 | 253,52 | -0,20 | 0,05 | -4,08 | 4,53E-05 |
| ZNF766 | 730,45 | -0,16 | 0,04 | -4,08 | 4,56E-05 |
| EPOR | 127,13 | 0,34 | 0,08 | 4,08 | 4,57E-05 |
| MMP11 | 37,26 | 0,60 | 0,15 | 4,08 | 4,58E-05 |
| MAMDC4 | 49,48 | 0,62 | 0,15 | 4,07 | 4,61E-05 |
| AHRR | 53,69 | 0,90 | 0,22 | 4,07 | 4,64E-05 |
| CCDC120 | 33,92 | 0,60 | 0,15 | 4,07 | 4,71E-05 |
| HBM | 209,07 | 0,58 | 0,14 | 4,07 | 4,70E-05 |
| FAM89B | 103,01 | 0,22 | 0,05 | 4,07 | 4,71E-05 |
| CROCC | 669,44 | 0,31 | 0,08 | 4,07 | 4,76E-05 |
| PPP1R13L | 37,05 | 0,68 | 0,17 | 4,07 | 4,80E-05 |
| YME1L1 | 4493,39 | -0,15 | 0,04 | -4,07 | 4,78E-05 |
| PLOD2 | 49,90 | 0,61 | 0,15 | 4,07 | 4,80E-05 |
| ZBTB46 | 55,89 | 0,50 | 0,12 | 4,06 | 4,85E-05 |
| NRXN3 | 95,37 | 0,76 | 0,19 | 4,06 | 4,85E-05 |
| ZC3H15 | 1021,53 | -0,32 | 0,08 | -4,06 | 4,85E-05 |

| | | | | | |
|---|---|---|---|---|---|
| CCDC78 | 29,39 | 0,57 | 0,14 | 4,06 | 4,89E-05 |
| PHLDA3 | 28,50 | 0,63 | 0,16 | 4,06 | 4,88E-05 |
| SLC28A3 | 30,82 | 0,69 | 0,17 | 4,06 | 4,90E-05 |
| ATG9B | 76,06 | 0,49 | 0,12 | 4,06 | 4,92E-05 |
| P2RY6 | 35,20 | 0,61 | 0,15 | 4,06 | 4,92E-05 |
| ZNF227 | 983,60 | -0,19 | 0,05 | -4,06 | 4,94E-05 |
| VIRMA | 2085,42 | -0,12 | 0,03 | -4,06 | 4,96E-05 |
| NARS1 | 2336,29 | -0,18 | 0,04 | -4,05 | 5,04E-05 |
| IQCK | 34,96 | 0,68 | 0,17 | 4,05 | 5,07E-05 |
| AKR1C1 | 63,29 | 0,53 | 0,13 | 4,06 | 5,01E-05 |
| NR4A1 | 61,98 | 0,65 | 0,16 | 4,05 | 5,04E-05 |
| RNF168 | 1566,77 | -0,13 | 0,03 | -4,05 | 5,08E-05 |
| ROCK1 | 7252,50 | -0,21 | 0,05 | -4,06 | 5,00E-05 |
| SCN1B | 72,82 | 0,53 | 0,13 | 4,05 | 5,06E-05 |
| WDR24 | 89,27 | 0,35 | 0,09 | 4,05 | 5,04E-05 |
| KCNIP2 | 97,94 | 0,40 | 0,10 | 4,05 | 5,07E-05 |
| TMEM201 | 52,90 | 0,63 | 0,15 | 4,05 | 5,01E-05 |
| DOT1L | 328,41 | 0,47 | 0,12 | 4,05 | 5,10E-05 |
| KIF18B | 27,87 | 0,72 | 0,18 | 4,05 | 5,13E-05 |
| ZNF14 | 521,20 | -0,31 | 0,08 | -4,04 | 5,28E-05 |
| ZDHHC1 | 53,65 | 0,43 | 0,11 | 4,04 | 5,30E-05 |
| DOCK3 | 128,65 | 0,41 | 0,10 | 4,04 | 5,30E-05 |
| RNF207 | 78,75 | 0,58 | 0,14 | 4,04 | 5,29E-05 |
| OLFM2 | 96,89 | 0,30 | 0,07 | 4,04 | 5,32E-05 |
| GNAO1 | 108,86 | 0,69 | 0,17 | 4,04 | 5,35E-05 |
| AJAP1 | 93,29 | 0,78 | 0,19 | 4,04 | 5,34E-05 |
| GAREM2 | 44,64 | 0,55 | 0,14 | 4,04 | 5,43E-05 |
| PPP1R16A | 103,18 | 0,38 | 0,09 | 4,04 | 5,42E-05 |
| PIWIL2 | 52,01 | 0,62 | 0,15 | 4,04 | 5,40E-05 |
| RASAL2 | 82,53 | 0,64 | 0,16 | 4,04 | 5,43E-05 |
| BCCIP | 610,14 | -0,17 | 0,04 | -4,04 | 5,39E-05 |
| SPC24 | 26,01 | 0,47 | 0,12 | 4,04 | 5,45E-05 |
| FBXW9 | 54,97 | 0,26 | 0,06 | 4,03 | 5,46E-05 |
| SH3GL1 | 512,58 | 0,28 | 0,07 | 4,04 | 5,45E-05 |
| PHAX | 680,81 | -0,21 | 0,05 | -4,03 | 5,53E-05 |
| MYLK3 | 42,70 | 0,78 | 0,19 | 4,03 | 5,56E-05 |
| WDR86 | 76,38 | 0,49 | 0,12 | 4,03 | 5,57E-05 |
| ZMYND10 | 50,16 | 0,34 | 0,08 | 4,03 | 5,59E-05 |
| AJUBA | 40,61 | 0,47 | 0,12 | 4,03 | 5,68E-05 |
| MRFAP1L1 | 1717,32 | -0,25 | 0,06 | -4,02 | 5,76E-05 |
| MIER2 | 180,91 | 0,44 | 0,11 | 4,02 | 5,77E-05 |
| TRIP13 | 34,08 | 0,67 | 0,17 | 4,02 | 5,83E-05 |
| AFAP1L2 | 78,33 | 0,45 | 0,11 | 4,02 | 5,83E-05 |
| PRSS36 | 63,07 | 0,33 | 0,08 | 4,02 | 5,87E-05 |
| SLC25A22 | 159,68 | 0,28 | 0,07 | 4,02 | 5,92E-05 |
| RNF20 | 2215,17 | -0,17 | 0,04 | -4,02 | 5,90E-05 |

| | | | | | |
|---|---|---|---|---|---|
| MYEOV | 37,08 | 0,65 | 0,16 | 4,01 | 5,95E-05 |
| TMPRSS13 | 52,75 | 0,52 | 0,13 | 4,02 | 5,90E-05 |
| MTREX | 2280,46 | -0,24 | 0,06 | -4,01 | 5,95E-05 |
| SYTL2 | 2309,97 | -0,33 | 0,08 | -4,02 | 5,94E-05 |
| WDR97 | 77,66 | 0,50 | 0,12 | 4,02 | 5,91E-05 |
| CDCA2 | 32,13 | 0,58 | 0,14 | 4,01 | 6,02E-05 |
| SPNS2 | 57,11 | 0,52 | 0,13 | 4,01 | 6,00E-05 |
| ZFPM1 | 166,64 | 0,46 | 0,11 | 4,01 | 6,01E-05 |
| CHID1 | 477,82 | 0,20 | 0,05 | 4,01 | 6,03E-05 |
| SH2D6 | 32,61 | 0,52 | 0,13 | 4,01 | 6,07E-05 |
| SLC39A6 | 696,40 | -0,15 | 0,04 | -4,01 | 6,10E-05 |
| DMPK | 114,03 | 0,48 | 0,12 | 4,01 | 6,13E-05 |
| FBXL16 | 46,79 | 0,52 | 0,13 | 4,01 | 6,13E-05 |
| ICAM5 | 50,18 | 0,44 | 0,11 | 4,01 | 6,16E-05 |
| CTIF | 152,34 | 0,47 | 0,12 | 4,01 | 6,19E-05 |
| ZDHHC11 | 61,47 | 0,70 | 0,18 | 4,00 | 6,25E-05 |
| CWF19L1 | 1323,37 | -0,10 | 0,02 | -4,00 | 6,33E-05 |
| TOMM70 | 1248,64 | -0,20 | 0,05 | -4,00 | 6,38E-05 |
| ABHD8 | 132,94 | 0,19 | 0,05 | 4,00 | 6,39E-05 |
| ELMO3 | 98,23 | 0,33 | 0,08 | 4,00 | 6,40E-05 |
| CUX2 | 54,78 | 0,79 | 0,20 | 3,99 | 6,48E-05 |
| B3GNT9 | 111,18 | 0,24 | 0,06 | 3,99 | 6,50E-05 |
| SELENON | 111,63 | 0,48 | 0,12 | 3,99 | 6,51E-05 |
| RUFY4 | 67,75 | 0,54 | 0,13 | 3,99 | 6,54E-05 |
| SLCO5A1 | 50,33 | 0,77 | 0,19 | 3,99 | 6,54E-05 |
| CRAT | 121,53 | 0,43 | 0,11 | 3,99 | 6,53E-05 |
| PLSCR4 | 37,93 | 0,52 | 0,13 | 3,99 | 6,57E-05 |
| UHRF1 | 75,87 | 0,48 | 0,12 | 3,99 | 6,64E-05 |
| MORF4L2 | 1227,26 | -0,19 | 0,05 | -3,99 | 6,66E-05 |
| MYL5 | 90,72 | 0,35 | 0,09 | 3,99 | 6,68E-05 |
| EVC | 73,27 | 0,69 | 0,17 | 3,99 | 6,75E-05 |
| CCDC174 | 1168,47 | -0,10 | 0,02 | -3,99 | 6,74E-05 |
| ISOC2 | 52,14 | 0,36 | 0,09 | 3,99 | 6,72E-05 |
| MMP17 | 61,42 | 0,64 | 0,16 | 3,99 | 6,75E-05 |
| RGS2 | 14430,56 | -0,35 | 0,09 | -3,98 | 6,80E-05 |
| STXBP1 | 101,22 | 0,44 | 0,11 | 3,98 | 6,80E-05 |
| PLEKHG2 | 223,01 | 0,36 | 0,09 | 3,98 | 6,80E-05 |
| ABCB1 | 662,80 | -0,26 | 0,07 | -3,98 | 6,82E-05 |
| APCDD1 | 42,82 | 0,72 | 0,18 | 3,98 | 6,84E-05 |
| NPHP1 | 62,19 | 0,73 | 0,18 | 3,98 | 6,93E-05 |
| FSCN2 | 25,40 | 0,48 | 0,12 | 3,98 | 6,95E-05 |
| TBC1D16 | 106,94 | 0,51 | 0,13 | 3,98 | 6,94E-05 |
| ASPSCR1 | 304,27 | 0,41 | 0,10 | 3,98 | 6,92E-05 |
| FAM133B | 709,03 | -0,21 | 0,05 | -3,98 | 7,00E-05 |
| GAMT | 43,90 | 0,33 | 0,08 | 3,98 | 7,01E-05 |
| IGSF9B | 141,49 | 0,52 | 0,13 | 3,97 | 7,07E-05 |

| | | | | | |
|---|---|---|---|---|---|
| KLHL24 | 4235,72 | -0,14 | 0,04 | -3,97 | 7,06E-05 |
| ABCB6 | 23,61 | 0,37 | 0,09 | 3,97 | 7,05E-05 |
| DHX9 | 4959,54 | -0,14 | 0,04 | -3,97 | 7,08E-05 |
| NOTCH4 | 77,97 | 0,51 | 0,13 | 3,97 | 7,08E-05 |
| ADPRHL1 | 82,14 | 0,63 | 0,16 | 3,97 | 7,12E-05 |
| ABLIM2 | 80,52 | 0,54 | 0,14 | 3,97 | 7,16E-05 |
| KLC2 | 168,68 | 0,32 | 0,08 | 3,97 | 7,20E-05 |
| SAR1A | 3668,22 | -0,13 | 0,03 | -3,97 | 7,23E-05 |
| FBRSL1 | 325,60 | 0,54 | 0,14 | 3,97 | 7,23E-05 |
| MBD2 | 2211,03 | -0,14 | 0,04 | -3,97 | 7,28E-05 |
| STX1A | 39,45 | 0,51 | 0,13 | 3,97 | 7,30E-05 |
| RFNG | 121,34 | 0,37 | 0,09 | 3,97 | 7,31E-05 |
| AP3M1 | 1423,46 | -0,11 | 0,03 | -3,97 | 7,33E-05 |
| CDS1 | 32,15 | 0,52 | 0,13 | 3,96 | 7,35E-05 |
| B4GALNT4 | 33,80 | 0,79 | 0,20 | 3,96 | 7,38E-05 |
| MYO16 | 88,38 | 0,78 | 0,20 | 3,96 | 7,42E-05 |
| TRIM4 | 1394,28 | -0,10 | 0,03 | -3,96 | 7,51E-05 |
| PPP2R5B | 571,01 | 0,31 | 0,08 | 3,96 | 7,59E-05 |
| C1orf159 | 181,33 | 0,32 | 0,08 | 3,96 | 7,60E-05 |
| SMC5 | 2346,58 | -0,22 | 0,06 | -3,96 | 7,59E-05 |
| ETV5 | 38,05 | 0,71 | 0,18 | 3,96 | 7,61E-05 |
| CUL3 | 3749,14 | -0,17 | 0,04 | -3,96 | 7,58E-05 |
| NCKIPSD | 130,44 | 0,29 | 0,07 | 3,96 | 7,64E-05 |
| HLF | 54,66 | 0,57 | 0,14 | 3,96 | 7,64E-05 |
| SRGAP1 | 79,03 | 0,93 | 0,24 | 3,95 | 7,67E-05 |
| PDE4A | 103,66 | 0,47 | 0,12 | 3,95 | 7,67E-05 |
| SCUBE2 | 70,63 | 0,56 | 0,14 | 3,95 | 7,71E-05 |
| SRC | 145,80 | 0,52 | 0,13 | 3,95 | 7,73E-05 |
| DLG4 | 170,01 | 0,41 | 0,10 | 3,95 | 7,84E-05 |
| FOXD2 | 41,43 | 0,79 | 0,20 | 3,95 | 7,84E-05 |
| MYCBPAP | 55,51 | 0,66 | 0,17 | 3,95 | 7,84E-05 |
| GAL3ST4 | 32,56 | 0,52 | 0,13 | 3,95 | 7,96E-05 |
| MSH2 | 764,79 | -0,20 | 0,05 | -3,94 | 7,99E-05 |
| RBMS3 | 55,01 | 0,69 | 0,17 | 3,94 | 8,00E-05 |
| COX15 | 741,40 | -0,10 | 0,03 | -3,94 | 8,04E-05 |
| VSIG10 | 179,95 | 0,45 | 0,11 | 3,94 | 8,07E-05 |
| FXYD6 | 31,12 | 0,70 | 0,18 | 3,94 | 8,11E-05 |
| SNRPD3 | 955,40 | -0,13 | 0,03 | -3,94 | 8,11E-05 |
| EPHB2 | 81,21 | 0,81 | 0,20 | 3,94 | 8,17E-05 |
| RTEL1-TNFRSF6B | 46,55 | 0,52 | 0,13 | 3,94 | 8,21E-05 |
| TYW3 | 519,30 | -0,26 | 0,07 | -3,94 | 8,23E-05 |
| LTBP4 | 271,08 | 0,43 | 0,11 | 3,94 | 8,31E-05 |
| SRM | 78,86 | 0,37 | 0,09 | 3,93 | 8,33E-05 |
| MEGF8 | 235,04 | 0,40 | 0,10 | 3,94 | 8,29E-05 |
| TSPAN17 | 530,02 | 0,16 | 0,04 | 3,94 | 8,28E-05 |
| PHETA1 | 94,29 | 0,36 | 0,09 | 3,94 | 8,31E-05 |

| | | | | | |
|---|---|---|---|---|---|
| MIA3 | 2858,41 | -0,12 | 0,03 | -3,93 | 8,36E-05 |
| KIFBP | 368,86 | -0,12 | 0,03 | -3,93 | 8,36E-05 |
| SLC25A34 | 31,82 | 0,47 | 0,12 | 3,93 | 8,37E-05 |
| ISYNA1 | 80,14 | 0,37 | 0,09 | 3,93 | 8,40E-05 |
| ACSM1 | 39,65 | 0,57 | 0,15 | 3,93 | 8,44E-05 |
| SIGLEC15 | 52,80 | 0,47 | 0,12 | 3,93 | 8,43E-05 |
| LYNX1 | 66,43 | 0,51 | 0,13 | 3,93 | 8,42E-05 |
| ATP6V1C1 | 1619,53 | -0,18 | 0,04 | -3,93 | 8,48E-05 |
| GRSF1 | 1741,57 | -0,13 | 0,03 | -3,93 | 8,51E-05 |
| WDR70 | 1138,56 | -0,14 | 0,04 | -3,93 | 8,53E-05 |
| RPUSD1 | 150,97 | 0,25 | 0,06 | 3,93 | 8,55E-05 |
| TUBA8 | 49,37 | 0,34 | 0,09 | 3,93 | 8,58E-05 |
| PPP1R37 | 192,56 | 0,34 | 0,09 | 3,93 | 8,64E-05 |
| PLCB3 | 163,96 | 0,33 | 0,08 | 3,93 | 8,63E-05 |
| FBN1 | 159,81 | 0,65 | 0,17 | 3,93 | 8,65E-05 |
| DDX46 | 2690,64 | -0,12 | 0,03 | -3,92 | 8,67E-05 |
| PIP5K1C | 337,98 | 0,42 | 0,11 | 3,92 | 8,72E-05 |
| RTCB | 1018,14 | -0,16 | 0,04 | -3,92 | 8,72E-05 |
| NOC4L | 87,43 | 0,36 | 0,09 | 3,92 | 8,70E-05 |
| BCAS2 | 433,67 | -0,23 | 0,06 | -3,92 | 8,75E-05 |
| SPNS1 | 27,26 | 0,59 | 0,15 | 3,92 | 8,75E-05 |
| AJM1 | 73,87 | 0,45 | 0,12 | 3,92 | 8,78E-05 |
| PGBD2 | 502,81 | -0,11 | 0,03 | -3,92 | 8,83E-05 |
| YTHDC1 | 5501,53 | -0,20 | 0,05 | -3,92 | 8,82E-05 |
| CWC15 | 584,58 | -0,20 | 0,05 | -3,92 | 8,84E-05 |
| TRIP11 | 2373,16 | -0,20 | 0,05 | -3,92 | 8,88E-05 |
| CACNA1I | 324,72 | 0,47 | 0,12 | 3,92 | 8,97E-05 |
| CTU2 | 93,20 | 0,26 | 0,07 | 3,91 | 9,07E-05 |
| DIS3 | 2284,88 | -0,18 | 0,05 | -3,91 | 9,12E-05 |
| EVPL | 66,07 | 0,65 | 0,17 | 3,91 | 9,22E-05 |
| TMEM120B | 133,15 | 0,31 | 0,08 | 3,91 | 9,25E-05 |
| AQR | 3378,24 | -0,14 | 0,04 | -3,91 | 9,39E-05 |
| KCTD15 | 314,98 | 0,24 | 0,06 | 3,91 | 9,30E-05 |
| METAP2 | 789,93 | -0,24 | 0,06 | -3,91 | 9,33E-05 |
| ADGRA2 | 71,13 | 0,57 | 0,15 | 3,91 | 9,39E-05 |
| EXD3 | 239,72 | 0,32 | 0,08 | 3,91 | 9,39E-05 |
| FNDC11 | 30,22 | 0,55 | 0,14 | 3,91 | 9,38E-05 |
| RGS12 | 295,09 | 0,35 | 0,09 | 3,91 | 9,36E-05 |
| MYH10 | 70,77 | 0,62 | 0,16 | 3,91 | 9,29E-05 |
| TXLNG | 615,58 | -0,13 | 0,03 | -3,91 | 9,32E-05 |
| LCN2 | 107,38 | 0,47 | 0,12 | 3,91 | 9,38E-05 |
| POLR2J3.1 | 110,68 | 0,47 | 0,12 | 3,91 | 9,42E-05 |
| MISP3 | 28,27 | 0,40 | 0,10 | 3,90 | 9,43E-05 |
| EFCAB6 | 54,97 | 0,81 | 0,21 | 3,90 | 9,46E-05 |
| ROM1 | 30,28 | 0,40 | 0,10 | 3,90 | 9,51E-05 |
| THOP1 | 222,09 | 0,27 | 0,07 | 3,90 | 9,57E-05 |

| | | | | | |
|---|---|---|---|---|---|
| USO1 | 2448,46 | -0,17 | 0,04 | -3,90 | 9,56E-05 |
| FBXO15 | 65,32 | 0,57 | 0,15 | 3,90 | 9,58E-05 |
| TCF3 | 513,05 | 0,28 | 0,07 | 3,90 | 9,58E-05 |
| POLD2 | 125,12 | 0,25 | 0,06 | 3,90 | 9,57E-05 |
| PAK4 | 81,95 | 0,52 | 0,13 | 3,90 | 9,60E-05 |
| SNX21 | 145,08 | 0,25 | 0,06 | 3,90 | 9,68E-05 |
| DEK | 2907,40 | -0,31 | 0,08 | -3,90 | 9,70E-05 |
| AHDC1 | 123,63 | 0,46 | 0,12 | 3,90 | 9,68E-05 |
| CELSR2 | 91,80 | 0,52 | 0,13 | 3,90 | 9,69E-05 |
| CTTNBP2 | 36,72 | 0,79 | 0,20 | 3,90 | 9,78E-05 |
| OSBPL6 | 50,13 | 0,71 | 0,18 | 3,90 | 9,77E-05 |
| TMCC2 | 434,54 | 0,49 | 0,13 | 3,90 | 9,79E-05 |
| SLTM | 4278,14 | -0,17 | 0,04 | -3,89 | 9,83E-05 |
| RNF152 | 61,20 | 0,64 | 0,16 | 3,89 | 9,83E-05 |
| HBP1 | 6205,10 | -0,20 | 0,05 | -3,89 | 9,90E-05 |
| SLC16A13 | 49,29 | 0,35 | 0,09 | 3,89 | 9,95E-05 |
| HACD1 | 58,30 | 0,35 | 0,09 | 3,89 | 9,95E-05 |
| SHROOM1 | 66,40 | 0,57 | 0,15 | 3,89 | 9,87E-05 |
| KPTN | 69,82 | 0,37 | 0,09 | 3,89 | 9,88E-05 |
| RHBDL1 | 25,97 | 0,51 | 0,13 | 3,89 | 9,96E-05 |
| SNRNP40 | 1042,82 | -0,13 | 0,03 | -3,89 | 9,93E-05 |
| MTCL1 | 103,71 | 0,79 | 0,20 | 3,89 | 9,95E-05 |
| FCSK | 204,64 | 0,38 | 0,10 | 3,89 | 1,00E-04 |
| RBM48 | 813,97 | -0,15 | 0,04 | -3,89 | 1,00E-04 |
| SETX | 18949,69 | -0,17 | 0,04 | -3,89 | 1,01E-04 |
| MCOLN3 | 45,65 | 0,60 | 0,15 | 3,89 | 1,01E-04 |
| ACTR2 | 17601,46 | -0,18 | 0,05 | -3,89 | 1,01E-04 |
| ENOX2 | 450,46 | -0,11 | 0,03 | -3,89 | 1,02E-04 |
| UPP2 | 26,03 | 0,44 | 0,11 | 3,88 | 1,02E-04 |
| SENP5 | 3139,51 | -0,11 | 0,03 | -3,89 | 1,02E-04 |
| IL17RC | 59,81 | 0,38 | 0,10 | 3,88 | 1,03E-04 |
| ADAMTSL4 | 191,16 | 0,56 | 0,14 | 3,88 | 1,03E-04 |
| PRPF18 | 805,42 | -0,21 | 0,05 | -3,88 | 1,03E-04 |
| MMACHC | 59,06 | 0,28 | 0,07 | 3,88 | 1,04E-04 |
| CHTF18 | 188,59 | 0,33 | 0,08 | 3,88 | 1,04E-04 |
| IFT140 | 158,46 | 0,34 | 0,09 | 3,88 | 1,04E-04 |
| RAI14 | 45,64 | 0,84 | 0,22 | 3,88 | 1,04E-04 |
| OLFM1 | 65,46 | 0,76 | 0,20 | 3,88 | 1,05E-04 |
| PTMS | 167,53 | 0,42 | 0,11 | 3,88 | 1,05E-04 |
| SOX8 | 48,71 | 0,64 | 0,16 | 3,88 | 1,05E-04 |
| SV2A | 41,41 | 0,48 | 0,12 | 3,88 | 1,05E-04 |
| MMEL1 | 36,65 | 0,69 | 0,18 | 3,88 | 1,05E-04 |
| LRRC24 | 33,85 | 0,39 | 0,10 | 3,88 | 1,06E-04 |
| KRBA1 | 131,90 | 0,36 | 0,09 | 3,88 | 1,06E-04 |
| MAP3K10 | 137,59 | 0,37 | 0,10 | 3,87 | 1,07E-04 |
| IKZF4 | 63,22 | 0,41 | 0,11 | 3,87 | 1,08E-04 |

| | | | | | |
|---|---|---|---|---|---|
| CNOT6L | 7149,66 | -0,20 | 0,05 | -3,87 | 1,08E-04 |
| ATAT1 | 45,25 | 0,34 | 0,09 | 3,87 | 1,08E-04 |
| SH3D21 | 113,45 | 0,34 | 0,09 | 3,87 | 1,08E-04 |
| KCNH3 | 82,03 | 0,54 | 0,14 | 3,87 | 1,09E-04 |
| CPTP | 78,51 | 0,39 | 0,10 | 3,87 | 1,10E-04 |
| STEEP1 | 448,91 | -0,16 | 0,04 | -3,87 | 1,10E-04 |
| LRP3 | 68,94 | 0,46 | 0,12 | 3,87 | 1,11E-04 |
| MRPL4 | 90,45 | 0,31 | 0,08 | 3,87 | 1,11E-04 |
| FN3K | 47,00 | 0,47 | 0,12 | 3,86 | 1,11E-04 |
| ZNF131 | 1317,21 | -0,11 | 0,03 | -3,86 | 1,11E-04 |
| BEGAIN | 81,14 | 0,79 | 0,20 | 3,86 | 1,12E-04 |
| CRHBP | 27,12 | 0,49 | 0,13 | 3,86 | 1,12E-04 |
| PPOX | 407,16 | 0,15 | 0,04 | 3,86 | 1,12E-04 |
| EPHX1 | 27,11 | 0,51 | 0,13 | 3,86 | 1,12E-04 |
| TM7SF2 | 202,91 | 0,18 | 0,05 | 3,86 | 1,13E-04 |
| POMT2 | 138,60 | 0,33 | 0,09 | 3,86 | 1,13E-04 |
| LRP11 | 29,01 | 0,56 | 0,15 | 3,86 | 1,14E-04 |
| ESCO1 | 1311,54 | -0,30 | 0,08 | -3,86 | 1,15E-04 |
| ZNF33A | 5365,08 | -0,18 | 0,05 | -3,86 | 1,15E-04 |
| RTL5 | 30,64 | 0,48 | 0,12 | 3,85 | 1,16E-04 |
| NPAS2 | 54,78 | 0,78 | 0,20 | 3,86 | 1,15E-04 |
| ANKMY2 | 461,00 | -0,16 | 0,04 | -3,85 | 1,16E-04 |
| FAM83E | 37,40 | 0,43 | 0,11 | 3,85 | 1,16E-04 |
| SIRT1 | 1467,70 | -0,15 | 0,04 | -3,85 | 1,16E-04 |
| RAB22A | 1916,16 | -0,10 | 0,03 | -3,86 | 1,16E-04 |
| AIMP1 | 499,74 | -0,28 | 0,07 | -3,85 | 1,16E-04 |
| INF2 | 333,51 | 0,43 | 0,11 | 3,86 | 1,15E-04 |
| ACOXL | 60,33 | 0,60 | 0,16 | 3,85 | 1,17E-04 |
| LONRF2 | 61,40 | 0,89 | 0,23 | 3,85 | 1,18E-04 |
| C1orf116 | 42,37 | 0,52 | 0,14 | 3,85 | 1,18E-04 |
| NELFA | 259,28 | 0,31 | 0,08 | 3,85 | 1,19E-04 |
| TMC4 | 43,14 | 0,50 | 0,13 | 3,85 | 1,19E-04 |
| KTN1 | 2964,85 | -0,21 | 0,06 | -3,85 | 1,19E-04 |
| KLHL11 | 423,44 | -0,13 | 0,03 | -3,85 | 1,19E-04 |
| PIK3C3 | 4100,57 | -0,17 | 0,04 | -3,85 | 1,20E-04 |
| NAPSA | 35,88 | 0,51 | 0,13 | 3,85 | 1,20E-04 |
| CCT6A | 1787,24 | -0,17 | 0,04 | -3,85 | 1,20E-04 |
| NIPBL | 8378,89 | -0,17 | 0,04 | -3,85 | 1,19E-04 |
| AGRN | 123,01 | 0,77 | 0,20 | 3,85 | 1,20E-04 |
| MGA | 4417,34 | -0,11 | 0,03 | -3,85 | 1,20E-04 |
| AFTPH | 3531,81 | -0,17 | 0,05 | -3,84 | 1,21E-04 |
| NRP2 | 98,59 | 0,63 | 0,16 | 3,84 | 1,21E-04 |
| KISS1R | 27,50 | 0,45 | 0,12 | 3,84 | 1,22E-04 |
| KY | 39,76 | 0,51 | 0,13 | 3,84 | 1,22E-04 |
| SMS | 883,67 | -0,17 | 0,05 | -3,84 | 1,23E-04 |
| SPIRE2 | 43,38 | 0,68 | 0,18 | 3,84 | 1,24E-04 |

| | | | | | |
|---|---|---|---|---|---|
| LSM3 | 271,77 | -0,26 | 0,07 | -3,84 | 1,24E-04 |
| ANKRD33B | 344,43 | 0,28 | 0,07 | 3,84 | 1,23E-04 |
| FBXO2 | 39,50 | 0,56 | 0,15 | 3,84 | 1,23E-04 |
| ACBD5 | 1402,99 | -0,16 | 0,04 | -3,84 | 1,24E-04 |
| SNX2 | 2829,17 | -0,22 | 0,06 | -3,84 | 1,24E-04 |
| CHERP | 227,16 | 0,31 | 0,08 | 3,83 | 1,26E-04 |
| MSH6 | 1348,85 | -0,13 | 0,03 | -3,84 | 1,26E-04 |
| TMEM143 | 105,08 | 0,19 | 0,05 | 3,84 | 1,25E-04 |
| CALM1 | 8870,39 | -0,11 | 0,03 | -3,84 | 1,25E-04 |
| MYO10 | 103,63 | 0,62 | 0,16 | 3,84 | 1,25E-04 |
| SARDH | 112,49 | 0,50 | 0,13 | 3,83 | 1,26E-04 |
| NTRK1 | 42,64 | 0,81 | 0,21 | 3,83 | 1,26E-04 |
| IWS1 | 1906,36 | -0,12 | 0,03 | -3,83 | 1,27E-04 |
| CLUH | 255,51 | 0,36 | 0,09 | 3,83 | 1,27E-04 |
| CDC42EP4 | 119,54 | 0,38 | 0,10 | 3,83 | 1,27E-04 |
| TMPRSS5 | 48,94 | 0,49 | 0,13 | 3,83 | 1,27E-04 |
| ADCK5 | 154,26 | 0,21 | 0,06 | 3,83 | 1,28E-04 |
| HAS3 | 43,90 | 0,56 | 0,15 | 3,83 | 1,28E-04 |
| EIF3A | 6174,32 | -0,14 | 0,04 | -3,83 | 1,29E-04 |
| CIZ1 | 339,71 | 0,31 | 0,08 | 3,83 | 1,30E-04 |
| PKD1L3 | 34,02 | 0,65 | 0,17 | 3,82 | 1,31E-04 |
| MFGE8 | 175,36 | 0,36 | 0,09 | 3,82 | 1,31E-04 |
| FBXO11 | 3417,77 | -0,16 | 0,04 | -3,82 | 1,31E-04 |
| RTKN | 35,11 | 0,62 | 0,16 | 3,82 | 1,32E-04 |
| VPS35 | 3410,88 | -0,13 | 0,03 | -3,82 | 1,33E-04 |
| KCNK10 | 48,21 | 0,81 | 0,21 | 3,82 | 1,34E-04 |
| NAV1 | 227,67 | 0,39 | 0,10 | 3,82 | 1,35E-04 |
| ZFP64 | 239,28 | 0,23 | 0,06 | 3,82 | 1,33E-04 |
| HAGHL | 105,25 | 0,34 | 0,09 | 3,82 | 1,33E-04 |
| LIMK1 | 254,85 | 0,35 | 0,09 | 3,82 | 1,34E-04 |
| MAP2K7 | 949,94 | 0,25 | 0,06 | 3,82 | 1,34E-04 |
| RPS6KL1 | 39,17 | 0,62 | 0,16 | 3,82 | 1,33E-04 |
| KLHL25 | 111,70 | 0,36 | 0,10 | 3,82 | 1,35E-04 |
| SLC25A23 | 63,51 | 0,38 | 0,10 | 3,82 | 1,35E-04 |
| ZBTB11 | 2232,83 | -0,17 | 0,04 | -3,82 | 1,35E-04 |
| ATP8A2 | 98,30 | 0,59 | 0,15 | 3,82 | 1,35E-04 |
| UBR1 | 3017,97 | -0,13 | 0,03 | -3,82 | 1,36E-04 |
| SMC2 | 481,62 | -0,23 | 0,06 | -3,82 | 1,36E-04 |
| KLHL17 | 124,19 | 0,32 | 0,08 | 3,82 | 1,36E-04 |
| MED12L | 142,60 | 0,36 | 0,10 | 3,81 | 1,37E-04 |
| PCBP3 | 106,46 | 0,55 | 0,14 | 3,81 | 1,37E-04 |
| CMTM8 | 26,60 | 0,45 | 0,12 | 3,81 | 1,37E-04 |
| OSBPL9 | 2452,99 | -0,13 | 0,03 | -3,81 | 1,38E-04 |
| TMPRSS3 | 52,77 | 0,52 | 0,14 | 3,81 | 1,39E-04 |
| TELO2 | 308,58 | 0,31 | 0,08 | 3,81 | 1,39E-04 |
| PRR12 | 237,65 | 0,59 | 0,16 | 3,81 | 1,39E-04 |

| | | | | | |
|---|---|---|---|---|---|
| FSCN3 | 42,79 | 0,35 | 0,09 | 3,81 | 1,39E-04 |
| PDE9A | 64,99 | 0,48 | 0,13 | 3,81 | 1,41E-04 |
| HYKK | 39,56 | 0,46 | 0,12 | 3,81 | 1,41E-04 |
| HAT1 | 653,78 | -0,27 | 0,07 | -3,81 | 1,41E-04 |
| DNAI2 | 40,36 | 0,52 | 0,14 | 3,81 | 1,41E-04 |
| ZNF503 | 37,75 | 0,65 | 0,17 | 3,80 | 1,42E-04 |
| PSMD13 | 1849,16 | -0,11 | 0,03 | -3,80 | 1,42E-04 |
| MCM10 | 47,24 | 0,65 | 0,17 | 3,81 | 1,41E-04 |
| MACROD1 | 58,90 | 0,33 | 0,09 | 3,80 | 1,42E-04 |
| RRN3 | 1047,33 | -0,16 | 0,04 | -3,80 | 1,42E-04 |
| CCDC28A | 836,09 | -0,16 | 0,04 | -3,80 | 1,42E-04 |
| NDUFS7 | 376,21 | 0,17 | 0,04 | 3,80 | 1,43E-04 |
| SLFNL1 | 34,04 | 0,58 | 0,15 | 3,80 | 1,43E-04 |
| CSE1L | 1294,74 | -0,15 | 0,04 | -3,80 | 1,43E-04 |
| LCAT | 37,87 | 0,41 | 0,11 | 3,80 | 1,43E-04 |
| CDH1 | 149,51 | 0,48 | 0,13 | 3,80 | 1,44E-04 |
| TMC5 | 73,06 | 0,60 | 0,16 | 3,80 | 1,44E-04 |
| IFI27 | 545,72 | 0,96 | 0,25 | 3,80 | 1,44E-04 |
| NEDD4L | 373,26 | 0,36 | 0,09 | 3,80 | 1,44E-04 |
| MLST8 | 146,15 | 0,20 | 0,05 | 3,80 | 1,45E-04 |
| ANO8 | 198,31 | 0,30 | 0,08 | 3,80 | 1,44E-04 |
| CASZ1 | 247,81 | 0,43 | 0,11 | 3,80 | 1,45E-04 |
| ATP7B | 132,55 | 0,38 | 0,10 | 3,80 | 1,46E-04 |
| CACNB1 | 148,88 | 0,31 | 0,08 | 3,80 | 1,47E-04 |
| SLC47A1 | 57,29 | 0,63 | 0,17 | 3,80 | 1,47E-04 |
| NUTM2D | 39,53 | 0,39 | 0,10 | 3,79 | 1,48E-04 |
| MTA1 | 382,08 | 0,33 | 0,09 | 3,79 | 1,48E-04 |
| HSPD1 | 1481,51 | -0,17 | 0,04 | -3,80 | 1,47E-04 |
| FCF1 | 923,73 | -0,17 | 0,04 | -3,79 | 1,48E-04 |
| ERCC2 | 162,66 | 0,27 | 0,07 | 3,79 | 1,48E-04 |
| SLC43A1 | 216,69 | 0,23 | 0,06 | 3,79 | 1,49E-04 |
| C17orf49 | 39,13 | 0,27 | 0,07 | 3,79 | 1,49E-04 |
| DAB1 | 58,65 | 0,55 | 0,15 | 3,79 | 1,49E-04 |
| NOP58 | 1382,54 | -0,23 | 0,06 | -3,79 | 1,50E-04 |
| PTPRG | 69,91 | 0,77 | 0,20 | 3,79 | 1,50E-04 |
| CTSE | 69,07 | 0,56 | 0,15 | 3,79 | 1,52E-04 |
| UBLCP1 | 1006,49 | -0,21 | 0,05 | -3,79 | 1,52E-04 |
| ZNF512B | 148,93 | 0,43 | 0,11 | 3,79 | 1,51E-04 |
| GCC1 | 1921,68 | -0,13 | 0,03 | -3,79 | 1,52E-04 |
| DNAAF5 | 201,21 | 0,30 | 0,08 | 3,79 | 1,52E-04 |
| MIEN1 | 95,25 | 0,26 | 0,07 | 3,79 | 1,51E-04 |
| GTPBP3 | 148,59 | 0,24 | 0,06 | 3,79 | 1,52E-04 |
| ZBTB42 | 129,94 | 0,29 | 0,08 | 3,79 | 1,53E-04 |
| MSANTD1 | 85,01 | 0,54 | 0,14 | 3,79 | 1,53E-04 |
| KCND3 | 44,50 | 0,69 | 0,18 | 3,78 | 1,54E-04 |
| CYP26C1 | 89,68 | 0,38 | 0,10 | 3,78 | 1,56E-04 |

| | | | | | |
|---|---|---|---|---|---|
| STAG1 | 3497,33 | -0,15 | 0,04 | -3,78 | 1,56E-04 |
| ARMH1 | 111,64 | 0,30 | 0,08 | 3,78 | 1,57E-04 |
| MPHOSPH8 | 2930,40 | -0,09 | 0,03 | -3,78 | 1,57E-04 |
| CATIP | 28,15 | 0,40 | 0,11 | 3,78 | 1,57E-04 |
| INPP5E | 172,42 | 0,37 | 0,10 | 3,78 | 1,58E-04 |
| PSEN2 | 172,29 | 0,21 | 0,06 | 3,78 | 1,58E-04 |
| LYG1 | 45,38 | 0,23 | 0,06 | 3,78 | 1,58E-04 |
| SCRIB | 323,01 | 0,37 | 0,10 | 3,78 | 1,58E-04 |
| ACKR2 | 37,44 | 0,55 | 0,15 | 3,78 | 1,59E-04 |
| SH3BP5 | 3060,93 | -0,12 | 0,03 | -3,78 | 1,59E-04 |
| RNF6 | 1387,28 | -0,20 | 0,05 | -3,78 | 1,59E-04 |
| TFAP2E | 79,19 | 0,31 | 0,08 | 3,78 | 1,60E-04 |
| PSMC6 | 1401,11 | -0,23 | 0,06 | -3,77 | 1,60E-04 |
| ZNF497 | 28,14 | 0,42 | 0,11 | 3,77 | 1,61E-04 |
| BCL2A1 | 617,17 | -0,51 | 0,13 | -3,77 | 1,61E-04 |
| USP35 | 186,90 | 0,28 | 0,07 | 3,77 | 1,61E-04 |
| SLIT1 | 90,70 | 0,65 | 0,17 | 3,77 | 1,60E-04 |
| EGLN3 | 131,83 | 0,39 | 0,10 | 3,77 | 1,61E-04 |
| FGFRL1 | 170,74 | 0,28 | 0,07 | 3,77 | 1,61E-04 |
| ZNF737 | 821,73 | -0,24 | 0,06 | -3,77 | 1,62E-04 |
| AOC1 | 25,51 | 0,79 | 0,21 | 3,77 | 1,63E-04 |
| AP4M1 | 123,18 | 0,21 | 0,06 | 3,77 | 1,65E-04 |
| MGAT4B | 844,28 | 0,21 | 0,05 | 3,77 | 1,65E-04 |
| GMPPA | 192,50 | 0,23 | 0,06 | 3,77 | 1,65E-04 |
| RECQL | 1807,31 | -0,28 | 0,08 | -3,77 | 1,64E-04 |
| ARFGAP3 | 1516,57 | -0,12 | 0,03 | -3,77 | 1,65E-04 |
| BCAM | 45,68 | 0,75 | 0,20 | 3,77 | 1,65E-04 |
| EXO1 | 41,14 | 0,51 | 0,14 | 3,77 | 1,64E-04 |
| USE1 | 68,46 | 0,25 | 0,07 | 3,77 | 1,65E-04 |
| PIK3R6 | 336,97 | 0,37 | 0,10 | 3,77 | 1,65E-04 |
| APBA1 | 70,79 | 0,61 | 0,16 | 3,77 | 1,65E-04 |
| LYAR | 678,29 | -0,21 | 0,06 | -3,77 | 1,66E-04 |
| CELSR1 | 164,52 | 0,62 | 0,16 | 3,77 | 1,66E-04 |
| ZSCAN12 | 399,40 | -0,23 | 0,06 | -3,76 | 1,67E-04 |
| SPHK1 | 149,59 | 0,26 | 0,07 | 3,76 | 1,67E-04 |
| TGM1 | 49,89 | 0,43 | 0,11 | 3,76 | 1,68E-04 |
| MCRIP2 | 188,22 | 0,22 | 0,06 | 3,76 | 1,68E-04 |
| PKD1L1 | 92,26 | 0,56 | 0,15 | 3,76 | 1,69E-04 |
| NDUFAF3 | 597,82 | 0,14 | 0,04 | 3,76 | 1,70E-04 |
| ST18 | 57,30 | 0,84 | 0,22 | 3,76 | 1,69E-04 |
| RAB21 | 3509,73 | -0,13 | 0,03 | -3,76 | 1,69E-04 |
| BCAS1 | 39,29 | 0,70 | 0,19 | 3,76 | 1,70E-04 |
| FAM234A | 74,57 | 0,34 | 0,09 | 3,76 | 1,70E-04 |
| MSH5 | 58,64 | 0,32 | 0,09 | 3,76 | 1,71E-04 |
| SLC1A3 | 47,81 | 0,56 | 0,15 | 3,76 | 1,72E-04 |
| PRR7 | 190,74 | 0,22 | 0,06 | 3,76 | 1,73E-04 |

| | | | | | |
|---|---|---|---|---|---|
| WNT2B | 64,09 | 0,65 | 0,17 | 3,76 | 1,72E-04 |
| RBBP5 | 1788,27 | -0,11 | 0,03 | -3,76 | 1,73E-04 |
| ALOX12 | 50,08 | 0,56 | 0,15 | 3,76 | 1,72E-04 |
| GZMB | 1277,91 | -0,33 | 0,09 | -3,76 | 1,73E-04 |
| ZRSR2 | 705,47 | -0,11 | 0,03 | -3,76 | 1,72E-04 |
| DMAC2 | 107,07 | 0,22 | 0,06 | 3,76 | 1,73E-04 |
| RPL7 | 6990,19 | -0,69 | 0,18 | -3,76 | 1,73E-04 |
| TUBG1 | 79,84 | 0,19 | 0,05 | 3,75 | 1,74E-04 |
| FAM81A | 38,25 | 0,62 | 0,16 | 3,75 | 1,75E-04 |
| TAF6 | 180,27 | 0,30 | 0,08 | 3,75 | 1,76E-04 |
| LTBP3 | 427,04 | 0,40 | 0,11 | 3,75 | 1,76E-04 |
| DRC1 | 42,98 | 0,60 | 0,16 | 3,75 | 1,75E-04 |
| NCDN | 105,85 | 0,42 | 0,11 | 3,75 | 1,76E-04 |
| WAPL | 5007,94 | -0,15 | 0,04 | -3,75 | 1,76E-04 |
| ENG | 48,30 | 0,58 | 0,15 | 3,75 | 1,76E-04 |
| UBE2Q1 | 1260,56 | -0,10 | 0,03 | -3,75 | 1,76E-04 |
| NRIP3 | 46,38 | 0,49 | 0,13 | 3,75 | 1,77E-04 |
| ERG | 52,76 | 0,67 | 0,18 | 3,75 | 1,78E-04 |
| IGSF22 | 43,49 | 0,64 | 0,17 | 3,75 | 1,78E-04 |
| AKT1S1 | 372,34 | 0,19 | 0,05 | 3,75 | 1,78E-04 |
| CEP295 | 1642,27 | -0,14 | 0,04 | -3,75 | 1,79E-04 |
| RMND5B | 513,93 | 0,15 | 0,04 | 3,75 | 1,80E-04 |
| ST6GALNAC4 | 341,52 | 0,42 | 0,11 | 3,75 | 1,80E-04 |
| CLEC18B | 51,36 | 0,64 | 0,17 | 3,75 | 1,80E-04 |
| FZD5 | 138,93 | 0,31 | 0,08 | 3,75 | 1,79E-04 |
| ZNF197 | 715,94 | -0,19 | 0,05 | -3,75 | 1,80E-04 |
| TPRA1 | 440,16 | 0,13 | 0,04 | 3,74 | 1,82E-04 |
| PLOD3 | 271,86 | 0,24 | 0,06 | 3,74 | 1,83E-04 |
| CUL2 | 726,67 | -0,14 | 0,04 | -3,74 | 1,84E-04 |
| CIT | 130,21 | 0,57 | 0,15 | 3,74 | 1,85E-04 |
| STEAP3 | 50,08 | 0,50 | 0,13 | 3,74 | 1,85E-04 |
| NSMAF | 1977,16 | -0,11 | 0,03 | -3,74 | 1,85E-04 |
| PSMA3 | 783,80 | -0,21 | 0,06 | -3,74 | 1,86E-04 |
| ARMCX3 | 863,42 | -0,13 | 0,03 | -3,74 | 1,86E-04 |
| TMEM40 | 70,50 | 0,38 | 0,10 | 3,74 | 1,87E-04 |
| APBB2 | 54,59 | 0,77 | 0,21 | 3,74 | 1,87E-04 |
| PRKD3 | 1808,17 | -0,14 | 0,04 | -3,74 | 1,87E-04 |
| DTX3 | 60,09 | 0,47 | 0,13 | 3,73 | 1,88E-04 |
| SYNGR1 | 53,70 | 0,53 | 0,14 | 3,73 | 1,89E-04 |
| ECD | 1084,61 | -0,09 | 0,02 | -3,73 | 1,90E-04 |
| ZUP1 | 453,43 | -0,20 | 0,05 | -3,73 | 1,90E-04 |
| ENY2 | 795,76 | -0,19 | 0,05 | -3,73 | 1,90E-04 |
| CAND1 | 3351,87 | -0,17 | 0,05 | -3,73 | 1,90E-04 |
| THOC2 | 2804,83 | -0,17 | 0,05 | -3,73 | 1,91E-04 |
| CMYA5 | 74,53 | 0,53 | 0,14 | 3,73 | 1,91E-04 |
| SETD2 | 4793,79 | -0,11 | 0,03 | -3,73 | 1,91E-04 |

| | | | | | |
|---|---|---|---|---|---|
| PBX4 | 103,29 | 0,28 | 0,08 | 3,73 | 1,91E-04 |
| SLC24A3 | 135,87 | 0,42 | 0,11 | 3,73 | 1,91E-04 |
| TAX1BP1 | 4689,05 | -0,25 | 0,07 | -3,73 | 1,93E-04 |
| SARAF | 10111,88 | -0,17 | 0,04 | -3,73 | 1,93E-04 |
| BPTF | 5705,32 | -0,11 | 0,03 | -3,73 | 1,94E-04 |
| DOCK1 | 72,44 | 0,67 | 0,18 | 3,73 | 1,94E-04 |
| NAA50 | 3950,36 | -0,17 | 0,04 | -3,73 | 1,94E-04 |
| CCDC25 | 475,37 | -0,16 | 0,04 | -3,72 | 1,96E-04 |
| ALKBH6 | 101,46 | 0,18 | 0,05 | 3,72 | 1,97E-04 |
| DMGDH | 40,79 | 0,64 | 0,17 | 3,72 | 1,98E-04 |
| POLR3GL | 250,33 | -0,24 | 0,06 | -3,72 | 1,98E-04 |
| COA8 | 264,54 | 0,11 | 0,03 | 3,72 | 1,98E-04 |
| FIGNL2 | 63,41 | 0,50 | 0,13 | 3,72 | 1,99E-04 |
| ZDHHC11B | 48,23 | 0,60 | 0,16 | 3,72 | 1,99E-04 |
| WNT5B | 47,65 | 0,56 | 0,15 | 3,72 | 2,02E-04 |
| LGMN | 51,85 | 0,38 | 0,10 | 3,72 | 2,02E-04 |
| TMEM163 | 42,52 | 0,70 | 0,19 | 3,72 | 2,03E-04 |
| TMEM39B | 116,17 | 0,29 | 0,08 | 3,71 | 2,04E-04 |
| GCC2 | 3317,73 | -0,22 | 0,06 | -3,71 | 2,05E-04 |
| CAPN5 | 161,75 | 0,42 | 0,11 | 3,71 | 2,05E-04 |
| MDK | 29,14 | 0,49 | 0,13 | 3,71 | 2,04E-04 |
| PTP4A2 | 6672,83 | -0,13 | 0,04 | -3,71 | 2,06E-04 |
| TSPAN18 | 195,89 | 0,33 | 0,09 | 3,71 | 2,05E-04 |
| PSMD1 | 1878,33 | -0,13 | 0,03 | -3,71 | 2,06E-04 |
| ERAL1 | 181,47 | 0,13 | 0,04 | 3,71 | 2,06E-04 |
| TMEM8B | 154,66 | 0,40 | 0,11 | 3,71 | 2,07E-04 |
| TMEM44 | 64,85 | 0,46 | 0,12 | 3,71 | 2,08E-04 |
| BMP1 | 92,55 | 0,42 | 0,11 | 3,71 | 2,08E-04 |
| CCNT1 | 2416,59 | -0,12 | 0,03 | -3,71 | 2,09E-04 |
| GPR137 | 190,14 | 0,19 | 0,05 | 3,71 | 2,09E-04 |
| FSTL3 | 66,96 | 0,31 | 0,08 | 3,71 | 2,09E-04 |
| WDR18 | 165,46 | 0,20 | 0,05 | 3,71 | 2,10E-04 |
| RPH3AL | 87,96 | 0,49 | 0,13 | 3,71 | 2,09E-04 |
| PLBD2 | 193,60 | 0,35 | 0,09 | 3,71 | 2,09E-04 |
| SRP72 | 1485,54 | -0,17 | 0,05 | -3,71 | 2,10E-04 |
| ATM | 27911,31 | -0,17 | 0,05 | -3,71 | 2,10E-04 |
| FBXO10 | 31,77 | 0,47 | 0,13 | 3,71 | 2,10E-04 |
| ACBD4 | 121,56 | 0,28 | 0,08 | 3,71 | 2,10E-04 |
| OSGIN1 | 45,63 | 0,40 | 0,11 | 3,71 | 2,11E-04 |
| PDK2 | 244,27 | 0,25 | 0,07 | 3,70 | 2,12E-04 |
| GPR146 | 667,51 | 0,47 | 0,13 | 3,70 | 2,12E-04 |
| GTF2E1 | 739,05 | -0,19 | 0,05 | -3,70 | 2,13E-04 |
| MTFR1L | 734,46 | 0,11 | 0,03 | 3,70 | 2,12E-04 |
| FAAP20 | 261,47 | 0,25 | 0,07 | 3,70 | 2,12E-04 |
| TMEM150B | 79,93 | 0,37 | 0,10 | 3,70 | 2,14E-04 |
| GTF2B | 831,56 | -0,22 | 0,06 | -3,70 | 2,14E-04 |

| | | | | | |
|---|---|---|---|---|---|
| TRIM7 | 61,56 | 0,49 | 0,13 | 3,70 | 2,13E-04 |
| ANO9 | 448,55 | 0,28 | 0,08 | 3,70 | 2,14E-04 |
| PXDC1 | 47,15 | 0,45 | 0,12 | 3,70 | 2,15E-04 |
| HCN3 | 36,62 | 0,41 | 0,11 | 3,70 | 2,16E-04 |
| TGM2 | 170,62 | 0,49 | 0,13 | 3,70 | 2,16E-04 |
| GRM2 | 43,44 | 0,69 | 0,19 | 3,70 | 2,16E-04 |
| PIK3R1 | 5323,85 | -0,16 | 0,04 | -3,70 | 2,17E-04 |
| CHAF1B | 77,20 | 0,34 | 0,09 | 3,70 | 2,17E-04 |
| ARHGEF17 | 55,64 | 0,80 | 0,22 | 3,70 | 2,17E-04 |
| ACBD3 | 1655,64 | -0,15 | 0,04 | -3,70 | 2,17E-04 |
| PACS2 | 443,13 | 0,37 | 0,10 | 3,70 | 2,17E-04 |
| ARFGEF1 | 5907,97 | -0,13 | 0,04 | -3,70 | 2,19E-04 |
| RNF212 | 42,03 | 0,49 | 0,13 | 3,70 | 2,19E-04 |
| OCIAD1 | 1582,27 | -0,12 | 0,03 | -3,70 | 2,20E-04 |
| P3H1 | 494,87 | 0,12 | 0,03 | 3,69 | 2,21E-04 |
| TBX6 | 26,26 | 0,39 | 0,11 | 3,69 | 2,21E-04 |
| GTF2E2 | 865,14 | -0,10 | 0,03 | -3,69 | 2,22E-04 |
| PINLYP | 25,19 | 0,60 | 0,16 | 3,69 | 2,22E-04 |
| KCNMB1 | 124,58 | 0,31 | 0,08 | 3,69 | 2,22E-04 |
| ACOT7 | 62,64 | 0,52 | 0,14 | 3,69 | 2,23E-04 |
| MFHAS1 | 840,53 | 0,17 | 0,05 | 3,69 | 2,24E-04 |
| EVI5L | 151,30 | 0,37 | 0,10 | 3,69 | 2,25E-04 |
| SLC6A12 | 126,10 | 0,43 | 0,12 | 3,69 | 2,25E-04 |
| ZDHHC8 | 222,70 | 0,37 | 0,10 | 3,69 | 2,26E-04 |
| ATF5 | 80,80 | 0,33 | 0,09 | 3,69 | 2,26E-04 |
| ELK1 | 255,96 | 0,15 | 0,04 | 3,69 | 2,27E-04 |
| PNPLA7 | 116,20 | 0,57 | 0,16 | 3,69 | 2,28E-04 |
| TMCO1 | 1201,50 | -0,28 | 0,08 | -3,68 | 2,29E-04 |
| CTSO | 829,72 | -0,17 | 0,05 | -3,68 | 2,30E-04 |
| PLAAT5 | 45,94 | 0,54 | 0,15 | 3,68 | 2,29E-04 |
| TMEM236 | 27,78 | 0,45 | 0,12 | 3,68 | 2,30E-04 |
| TMIGD3 | 32,84 | 0,45 | 0,12 | 3,68 | 2,30E-04 |
| ALS2CL | 326,45 | 0,32 | 0,09 | 3,68 | 2,30E-04 |
| PMPCB | 1437,35 | -0,15 | 0,04 | -3,68 | 2,30E-04 |
| UCN | 33,26 | 0,29 | 0,08 | 3,68 | 2,30E-04 |
| ZNF771 | 61,88 | 0,29 | 0,08 | 3,68 | 2,31E-04 |
| KDM5A | 13325,36 | -0,13 | 0,04 | -3,68 | 2,32E-04 |
| ASAP2 | 173,13 | 0,37 | 0,10 | 3,68 | 2,32E-04 |
| TJP3 | 85,33 | 0,35 | 0,10 | 3,68 | 2,32E-04 |
| RAB3C | 35,45 | 0,58 | 0,16 | 3,68 | 2,33E-04 |
| NOCT | 69,60 | 0,24 | 0,06 | 3,68 | 2,33E-04 |
| HEPACAM2 | 58,11 | 0,47 | 0,13 | 3,68 | 2,34E-04 |
| GGT7 | 223,40 | 0,22 | 0,06 | 3,68 | 2,34E-04 |
| PMM1 | 163,18 | 0,17 | 0,05 | 3,68 | 2,35E-04 |
| CORO2A | 277,04 | 0,21 | 0,06 | 3,68 | 2,35E-04 |
| DPF3 | 109,16 | 0,45 | 0,12 | 3,68 | 2,35E-04 |

| | | | | | |
|---|---|---|---|---|---|
| BDP1 | 3596,34 | -0,16 | 0,04 | -3,68 | 2,36E-04 |
| B9D1 | 50,12 | 0,33 | 0,09 | 3,68 | 2,37E-04 |
| INHBA | 43,39 | 0,76 | 0,21 | 3,68 | 2,38E-04 |
| MYBBP1A | 335,18 | 0,26 | 0,07 | 3,67 | 2,40E-04 |
| ZFTA | 144,75 | 0,27 | 0,07 | 3,67 | 2,40E-04 |
| TGFB3 | 56,26 | 0,46 | 0,13 | 3,67 | 2,40E-04 |
| ADAMTS4 | 55,89 | 0,44 | 0,12 | 3,67 | 2,40E-04 |
| PTGER3 | 37,39 | 0,66 | 0,18 | 3,67 | 2,41E-04 |
| DNAH6 | 151,77 | 0,48 | 0,13 | 3,67 | 2,42E-04 |
| GPBP1 | 4650,65 | -0,23 | 0,06 | -3,67 | 2,44E-04 |
| EIF2S2 | 1526,85 | -0,20 | 0,05 | -3,67 | 2,44E-04 |
| FYTTD1 | 1713,12 | -0,16 | 0,04 | -3,67 | 2,45E-04 |
| KLLN | 79,87 | 0,25 | 0,07 | 3,67 | 2,45E-04 |
| CACNA1F | 28,94 | 0,63 | 0,17 | 3,67 | 2,46E-04 |
| CFAP119 | 118,80 | 0,21 | 0,06 | 3,67 | 2,47E-04 |
| CDKN1A | 243,40 | 0,36 | 0,10 | 3,66 | 2,47E-04 |
| MVB12A | 81,07 | 0,38 | 0,10 | 3,66 | 2,48E-04 |
| LY6G5C | 31,28 | 0,40 | 0,11 | 3,66 | 2,48E-04 |
| PIF1 | 41,60 | 0,43 | 0,12 | 3,66 | 2,48E-04 |
| XRCC6 | 4707,02 | -0,17 | 0,05 | -3,66 | 2,49E-04 |
| H2AX | 157,56 | 0,29 | 0,08 | 3,66 | 2,49E-04 |
| EIF5 | 4282,40 | -0,14 | 0,04 | -3,66 | 2,49E-04 |
| PDE6B | 154,11 | 0,26 | 0,07 | 3,66 | 2,52E-04 |
| SLC7A8 | 41,75 | 0,61 | 0,17 | 3,66 | 2,52E-04 |
| PAK3 | 48,50 | 0,58 | 0,16 | 3,66 | 2,52E-04 |
| NUCKS1 | 4072,45 | -0,18 | 0,05 | -3,66 | 2,52E-04 |
| ZBTB2 | 416,66 | -0,12 | 0,03 | -3,66 | 2,53E-04 |
| IARS2 | 1393,14 | -0,12 | 0,03 | -3,66 | 2,53E-04 |
| ADORA2B | 49,67 | 0,34 | 0,09 | 3,66 | 2,54E-04 |
| POLE | 880,15 | 0,19 | 0,05 | 3,66 | 2,54E-04 |
| NHSL1 | 86,74 | 0,47 | 0,13 | 3,66 | 2,53E-04 |
| RHAG | 124,86 | 0,45 | 0,12 | 3,66 | 2,54E-04 |
| GINS2 | 36,32 | 0,43 | 0,12 | 3,66 | 2,56E-04 |
| SBNO1 | 3601,91 | -0,11 | 0,03 | -3,66 | 2,56E-04 |
| SRPRB | 852,46 | -0,16 | 0,04 | -3,66 | 2,56E-04 |
| IPO5 | 1451,85 | -0,12 | 0,03 | -3,66 | 2,57E-04 |
| BRCC3 | 582,13 | -0,12 | 0,03 | -3,65 | 2,57E-04 |
| DNAJA1 | 2497,31 | -0,30 | 0,08 | -3,65 | 2,58E-04 |
| ACRBP | 243,28 | 0,27 | 0,07 | 3,66 | 2,57E-04 |
| SKIC3 | 2912,98 | -0,20 | 0,05 | -3,65 | 2,58E-04 |
| BZW1 | 3695,17 | -0,19 | 0,05 | -3,65 | 2,58E-04 |
| ENSG00000284691 | 231,02 | 0,21 | 0,06 | 3,65 | 2,58E-04 |
| PRPF4 | 796,85 | -0,11 | 0,03 | -3,65 | 2,59E-04 |
| DZIP1L | 57,30 | 0,58 | 0,16 | 3,65 | 2,59E-04 |
| THBS4 | 69,24 | 0,42 | 0,11 | 3,65 | 2,59E-04 |
| ZNF837 | 60,63 | 0,30 | 0,08 | 3,65 | 2,59E-04 |

| | | | | | |
|---|---|---|---|---|---|
| ZNF189 | 845,29 | -0,21 | 0,06 | -3,65 | 2,60E-04 |
| MICU2 | 1452,37 | -0,19 | 0,05 | -3,65 | 2,60E-04 |
| PKD1 | 639,49 | 0,43 | 0,12 | 3,65 | 2,60E-04 |
| KIAA0586 | 1386,47 | -0,14 | 0,04 | -3,65 | 2,61E-04 |
| ZNF35 | 229,63 | -0,18 | 0,05 | -3,65 | 2,62E-04 |
| GPATCH11 | 581,40 | -0,24 | 0,06 | -3,65 | 2,62E-04 |
| DUSP4 | 56,23 | 0,58 | 0,16 | 3,65 | 2,63E-04 |
| CLDN7 | 29,84 | 0,43 | 0,12 | 3,65 | 2,63E-04 |
| NGDN | 538,76 | -0,16 | 0,04 | -3,65 | 2,63E-04 |
| CUTC | 678,79 | -0,17 | 0,05 | -3,65 | 2,65E-04 |
| PLAU | 28,85 | 0,49 | 0,13 | 3,65 | 2,64E-04 |
| SORBS2 | 66,37 | 0,87 | 0,24 | 3,65 | 2,64E-04 |
| PALD1 | 43,50 | 0,73 | 0,20 | 3,65 | 2,64E-04 |
| SBDS | 447,50 | -0,23 | 0,06 | -3,65 | 2,64E-04 |
| RCCD1 | 185,44 | 0,27 | 0,07 | 3,65 | 2,65E-04 |
| ZNF436 | 457,42 | -0,12 | 0,03 | -3,65 | 2,67E-04 |
| WDR47 | 805,44 | -0,12 | 0,03 | -3,65 | 2,67E-04 |
| KIZ | 857,86 | -0,11 | 0,03 | -3,64 | 2,69E-04 |
| ADAMTS17 | 186,10 | 0,36 | 0,10 | 3,64 | 2,71E-04 |
| SNRK | 8578,12 | -0,16 | 0,04 | -3,64 | 2,71E-04 |
| CARMIL2 | 297,60 | 0,38 | 0,10 | 3,64 | 2,71E-04 |
| MRPL38 | 36,98 | 0,39 | 0,11 | 3,64 | 2,74E-04 |
| ZC3HAV1L | 81,21 | 0,29 | 0,08 | 3,64 | 2,75E-04 |
| ITGA2 | 69,03 | 0,45 | 0,12 | 3,64 | 2,76E-04 |
| SFPQ | 10278,52 | -0,13 | 0,03 | -3,64 | 2,77E-04 |
| ADGRL1 | 118,11 | 0,48 | 0,13 | 3,63 | 2,79E-04 |
| RBBP7 | 1276,23 | -0,12 | 0,03 | -3,63 | 2,79E-04 |
| PSD3 | 145,12 | 0,42 | 0,12 | 3,63 | 2,79E-04 |
| NRM | 61,35 | 0,23 | 0,06 | 3,63 | 2,79E-04 |
| STK11IP | 322,37 | 0,27 | 0,07 | 3,63 | 2,79E-04 |
| DDX52 | 1067,56 | -0,09 | 0,03 | -3,63 | 2,82E-04 |
| FXYD2 | 37,73 | 0,54 | 0,15 | 3,63 | 2,83E-04 |
| KANK2 | 174,21 | 0,52 | 0,14 | 3,63 | 2,84E-04 |
| DENND4C | 3164,68 | -0,14 | 0,04 | -3,63 | 2,84E-04 |
| TRMT10C | 334,91 | -0,32 | 0,09 | -3,63 | 2,84E-04 |
| ARHGEF10 | 46,87 | 0,55 | 0,15 | 3,63 | 2,85E-04 |
| JOSD2 | 133,43 | 0,18 | 0,05 | 3,63 | 2,85E-04 |
| KMT5B | 3074,98 | -0,12 | 0,03 | -3,63 | 2,85E-04 |
| RB1CC1 | 5036,94 | -0,23 | 0,06 | -3,63 | 2,86E-04 |
| HES1 | 46,28 | 0,40 | 0,11 | 3,63 | 2,87E-04 |
| IPO7 | 1948,29 | -0,16 | 0,04 | -3,62 | 2,90E-04 |
| TAF1B | 597,88 | -0,16 | 0,04 | -3,62 | 2,89E-04 |
| GPLD1 | 89,06 | 0,40 | 0,11 | 3,62 | 2,90E-04 |
| ZNF45 | 722,09 | -0,11 | 0,03 | -3,62 | 2,91E-04 |
| HMCN1 | 70,06 | 0,71 | 0,19 | 3,62 | 2,92E-04 |
| WASF3 | 56,25 | 0,56 | 0,15 | 3,62 | 2,93E-04 |

| | | | | | |
|---|---|---|---|---|---|
| SMPD3 | 267,80 | 0,52 | 0,14 | 3,62 | 2,93E-04 |
| FBXO41 | 237,86 | 0,39 | 0,11 | 3,62 | 2,95E-04 |
| C3orf52 | 36,47 | 0,53 | 0,15 | 3,62 | 2,95E-04 |
| C17orf113 | 34,08 | 0,43 | 0,12 | 3,62 | 2,95E-04 |
| ZFAND2B | 516,57 | 0,12 | 0,03 | 3,62 | 2,96E-04 |
| STARD5 | 85,27 | 0,24 | 0,07 | 3,62 | 2,96E-04 |
| RBMX2 | 408,96 | -0,15 | 0,04 | -3,62 | 2,96E-04 |
| ZNF341 | 115,18 | 0,40 | 0,11 | 3,62 | 2,97E-04 |
| MCF2L | 297,37 | 0,44 | 0,12 | 3,62 | 2,97E-04 |
| BBLN | 288,49 | 0,16 | 0,05 | 3,62 | 2,97E-04 |
| RBFOX2 | 86,66 | 0,64 | 0,18 | 3,62 | 2,98E-04 |
| DPH1 | 186,71 | 0,26 | 0,07 | 3,62 | 2,99E-04 |
| SAXO1 | 61,09 | 0,53 | 0,15 | 3,62 | 3,00E-04 |
| ZFYVE21 | 423,61 | 0,13 | 0,04 | 3,62 | 2,99E-04 |
| CAB39 | 7530,09 | -0,15 | 0,04 | -3,62 | 2,99E-04 |
| IQCH | 49,93 | 0,61 | 0,17 | 3,61 | 3,01E-04 |
| HOMER2 | 201,91 | 0,26 | 0,07 | 3,61 | 3,01E-04 |
| FBLN7 | 151,15 | 0,29 | 0,08 | 3,61 | 3,02E-04 |
| SUCLA2 | 695,09 | -0,14 | 0,04 | -3,61 | 3,02E-04 |
| FBXL18 | 118,09 | 0,40 | 0,11 | 3,61 | 3,03E-04 |
| HIC1 | 142,79 | 0,38 | 0,11 | 3,61 | 3,04E-04 |
| RAB3GAP2 | 2996,77 | -0,12 | 0,03 | -3,61 | 3,04E-04 |
| HBA2 | 212,79 | 0,68 | 0,19 | 3,61 | 3,04E-04 |
| PODXL | 42,53 | 0,62 | 0,17 | 3,61 | 3,04E-04 |
| HJURP | 39,29 | 0,57 | 0,16 | 3,61 | 3,05E-04 |
| PEMT | 97,23 | 0,21 | 0,06 | 3,61 | 3,06E-04 |
| SEC63 | 1572,63 | -0,10 | 0,03 | -3,61 | 3,07E-04 |
| AMZ1 | 60,06 | 0,53 | 0,15 | 3,61 | 3,08E-04 |
| GATD1 | 335,56 | 0,27 | 0,07 | 3,61 | 3,08E-04 |
| EME2 | 602,48 | 0,24 | 0,07 | 3,61 | 3,08E-04 |
| SETD1A | 154,16 | 0,42 | 0,12 | 3,61 | 3,08E-04 |
| RARS1 | 1252,07 | -0,17 | 0,05 | -3,61 | 3,09E-04 |
| SPATA9 | 34,37 | 0,46 | 0,13 | 3,61 | 3,11E-04 |
| MTIF2 | 655,14 | -0,15 | 0,04 | -3,60 | 3,13E-04 |
| IRF6 | 37,37 | 0,60 | 0,17 | 3,60 | 3,13E-04 |
| TRPV3 | 69,64 | 0,66 | 0,18 | 3,60 | 3,14E-04 |
| UNC5CL | 96,62 | 0,24 | 0,07 | 3,60 | 3,15E-04 |
| TMEM63C | 74,35 | 0,56 | 0,16 | 3,60 | 3,15E-04 |
| BTBD2 | 287,69 | 0,31 | 0,09 | 3,60 | 3,14E-04 |
| TEDC1 | 312,15 | 0,20 | 0,06 | 3,60 | 3,14E-04 |
| CCT5 | 2985,04 | -0,14 | 0,04 | -3,60 | 3,14E-04 |
| HYI | 143,00 | 0,23 | 0,06 | 3,60 | 3,14E-04 |
| TMEM79 | 82,85 | 0,23 | 0,07 | 3,60 | 3,15E-04 |
| RNFT2 | 66,08 | 0,41 | 0,11 | 3,60 | 3,16E-04 |
| MYL6B | 41,85 | 0,32 | 0,09 | 3,60 | 3,16E-04 |
| RPAP2 | 1283,82 | -0,16 | 0,04 | -3,60 | 3,18E-04 |

| | | | | | |
|---|---|---|---|---|---|
| RBMXL1 | 1058,47 | -0,17 | 0,05 | -3,60 | 3,18E-04 |
| CYB561 | 366,73 | 0,25 | 0,07 | 3,60 | 3,18E-04 |
| TSPAN9 | 109,89 | 0,56 | 0,16 | 3,60 | 3,20E-04 |
| IFRD2 | 392,00 | 0,21 | 0,06 | 3,60 | 3,21E-04 |
| CARM1 | 459,54 | 0,41 | 0,12 | 3,60 | 3,21E-04 |
| PNPLA6 | 934,83 | 0,31 | 0,09 | 3,60 | 3,23E-04 |
| CCDC150 | 46,37 | 0,68 | 0,19 | 3,60 | 3,23E-04 |
| PTPRS | 148,34 | 0,58 | 0,16 | 3,60 | 3,24E-04 |
| ZNF17 | 387,54 | -0,22 | 0,06 | -3,59 | 3,25E-04 |
| TLN2 | 169,85 | 0,63 | 0,18 | 3,59 | 3,25E-04 |
| MTIF3 | 624,92 | -0,13 | 0,04 | -3,59 | 3,26E-04 |
| KIFC3 | 232,78 | 0,33 | 0,09 | 3,59 | 3,27E-04 |
| PIBF1 | 1073,67 | -0,23 | 0,06 | -3,59 | 3,28E-04 |
| CNTNAP1 | 86,54 | 0,45 | 0,13 | 3,59 | 3,30E-04 |
| CNOT8 | 3166,31 | -0,13 | 0,04 | -3,59 | 3,30E-04 |
| PPEF1 | 26,32 | 0,43 | 0,12 | 3,59 | 3,30E-04 |
| YWHAB | 8409,79 | -0,12 | 0,03 | -3,59 | 3,30E-04 |
| ENSG00000283486 | 23,67 | 0,48 | 0,13 | 3,59 | 3,31E-04 |
| PRKAR1B | 145,80 | 0,35 | 0,10 | 3,59 | 3,32E-04 |
| GZMA | 854,62 | -0,41 | 0,11 | -3,59 | 3,32E-04 |
| GSDME | 49,76 | 0,67 | 0,19 | 3,59 | 3,33E-04 |
| CCDC9B | 120,59 | 0,34 | 0,09 | 3,59 | 3,36E-04 |
| MBD4 | 1917,48 | -0,15 | 0,04 | -3,59 | 3,36E-04 |
| BCL2L12 | 60,45 | 0,20 | 0,06 | 3,59 | 3,36E-04 |
| RAB40B | 43,37 | 0,46 | 0,13 | 3,58 | 3,37E-04 |
| ASB2 | 125,97 | 0,36 | 0,10 | 3,58 | 3,39E-04 |
| GAS8 | 128,92 | 0,26 | 0,07 | 3,58 | 3,40E-04 |
| NAA15 | 1836,32 | -0,18 | 0,05 | -3,58 | 3,40E-04 |
| S1PR2 | 53,78 | 0,32 | 0,09 | 3,58 | 3,41E-04 |
| TSPEAR | 38,69 | 0,69 | 0,19 | 3,58 | 3,43E-04 |
| HNRNPA3 | 4855,07 | -0,12 | 0,03 | -3,58 | 3,46E-04 |
| RNASE6 | 776,34 | -0,30 | 0,08 | -3,58 | 3,47E-04 |
| CCT7 | 2115,86 | -0,12 | 0,03 | -3,58 | 3,47E-04 |
| FXR1 | 2919,76 | -0,19 | 0,05 | -3,58 | 3,48E-04 |
| RNF175 | 79,45 | 0,38 | 0,11 | 3,58 | 3,48E-04 |
| DTX1 | 92,52 | 0,43 | 0,12 | 3,58 | 3,50E-04 |
| MRAS | 94,03 | 0,42 | 0,12 | 3,57 | 3,51E-04 |
| EIF2AK3 | 1145,73 | -0,14 | 0,04 | -3,57 | 3,53E-04 |
| REXO1 | 314,63 | 0,40 | 0,11 | 3,57 | 3,54E-04 |
| SLC2A11 | 183,56 | 0,25 | 0,07 | 3,57 | 3,55E-04 |
| BAZ1B | 2454,76 | -0,10 | 0,03 | -3,57 | 3,55E-04 |
| SLC5A6 | 215,82 | 0,22 | 0,06 | 3,57 | 3,57E-04 |
| SYNGAP1 | 226,44 | 0,41 | 0,12 | 3,57 | 3,57E-04 |
| GSE1 | 1589,30 | 0,22 | 0,06 | 3,57 | 3,57E-04 |
| ENTPD5 | 483,63 | 0,21 | 0,06 | 3,57 | 3,58E-04 |
| KLHL26 | 105,36 | 0,30 | 0,08 | 3,57 | 3,59E-04 |

| | | | | | |
|---|---|---|---|---|---|
| WWTR1 | 52,91 | 0,48 | 0,13 | 3,57 | 3,59E-04 |
| NFIC | 324,03 | 0,40 | 0,11 | 3,57 | 3,60E-04 |
| ACSF2 | 308,45 | 0,20 | 0,06 | 3,57 | 3,60E-04 |
| PIK3R4 | 1713,80 | -0,10 | 0,03 | -3,57 | 3,60E-04 |
| VPS41 | 3943,77 | -0,14 | 0,04 | -3,57 | 3,61E-04 |
| RNMT | 1144,40 | -0,12 | 0,03 | -3,57 | 3,61E-04 |
| RFX1 | 208,06 | 0,41 | 0,12 | 3,57 | 3,63E-04 |
| ESR1 | 96,52 | 0,45 | 0,13 | 3,57 | 3,63E-04 |
| EMID1 | 61,98 | 0,54 | 0,15 | 3,56 | 3,65E-04 |
| LDLR | 271,15 | 0,27 | 0,08 | 3,56 | 3,65E-04 |
| EIF4A2 | 8925,98 | -0,23 | 0,07 | -3,56 | 3,65E-04 |
| LIG4 | 577,24 | -0,29 | 0,08 | -3,56 | 3,66E-04 |
| FRMD4A | 186,02 | 0,54 | 0,15 | 3,56 | 3,67E-04 |
| RELB | 341,62 | 0,23 | 0,06 | 3,56 | 3,68E-04 |
| SELENOS | 488,66 | -0,11 | 0,03 | -3,56 | 3,68E-04 |
| ATP6AP2 | 2958,00 | -0,16 | 0,04 | -3,56 | 3,69E-04 |
| ESF1 | 349,38 | -0,34 | 0,10 | -3,56 | 3,70E-04 |
| PUM3 | 733,86 | -0,18 | 0,05 | -3,56 | 3,70E-04 |
| ZNF180 | 599,70 | -0,17 | 0,05 | -3,56 | 3,70E-04 |
| PIK3C2A | 2437,18 | -0,12 | 0,03 | -3,56 | 3,74E-04 |
| C4BPA | 118,23 | 0,98 | 0,28 | 3,56 | 3,75E-04 |
| OCEL1 | 41,33 | 0,36 | 0,10 | 3,56 | 3,76E-04 |
| THUMPD1 | 2233,59 | -0,19 | 0,05 | -3,56 | 3,76E-04 |
| CDADC1 | 384,19 | -0,14 | 0,04 | -3,56 | 3,76E-04 |
| KYAT1 | 154,88 | 0,24 | 0,07 | 3,56 | 3,77E-04 |
| HAUS6 | 1199,07 | -0,14 | 0,04 | -3,56 | 3,78E-04 |
| TOPBP1 | 2263,09 | -0,12 | 0,03 | -3,56 | 3,78E-04 |
| NR1H3 | 104,03 | 0,22 | 0,06 | 3,55 | 3,78E-04 |
| SIRT3 | 264,33 | 0,16 | 0,04 | 3,55 | 3,78E-04 |
| SLC2A4RG | 182,74 | 0,30 | 0,08 | 3,55 | 3,80E-04 |
| HPS3 | 2146,84 | -0,12 | 0,03 | -3,55 | 3,84E-04 |
| MTHFD1L | 133,41 | 0,35 | 0,10 | 3,55 | 3,85E-04 |
| GRINA | 685,26 | 0,49 | 0,14 | 3,55 | 3,88E-04 |
| ZMYND12 | 24,79 | 0,41 | 0,11 | 3,55 | 3,89E-04 |
| SCAMP4 | 108,37 | 0,44 | 0,12 | 3,55 | 3,90E-04 |
| DCAF13 | 806,83 | -0,20 | 0,06 | -3,55 | 3,90E-04 |
| APPL1 | 2172,38 | -0,15 | 0,04 | -3,55 | 3,91E-04 |
| SMIM10L2A | 38,29 | 0,45 | 0,13 | 3,55 | 3,91E-04 |
| PPP6R3 | 6744,47 | -0,10 | 0,03 | -3,55 | 3,92E-04 |
| EIF5B | 1036,04 | -0,12 | 0,03 | -3,55 | 3,92E-04 |
| TTC34 | 97,73 | 0,49 | 0,14 | 3,55 | 3,92E-04 |
| AP2A1 | 1681,48 | 0,35 | 0,10 | 3,55 | 3,92E-04 |
| NEURL4 | 506,54 | 0,15 | 0,04 | 3,55 | 3,93E-04 |
| KANK3 | 55,68 | 0,40 | 0,11 | 3,54 | 3,95E-04 |
| OPRL1 | 254,80 | 0,24 | 0,07 | 3,54 | 3,96E-04 |
| BCAT2 | 100,69 | 0,26 | 0,07 | 3,54 | 3,96E-04 |

| | | | | | |
|---|---|---|---|---|---|
| HSPA12A | 72,74 | 0,67 | 0,19 | 3,54 | 3,97E-04 |
| NPM1 | 4667,88 | -0,27 | 0,08 | -3,54 | 4,00E-04 |
| GFI1B | 293,54 | 0,32 | 0,09 | 3,54 | 3,99E-04 |
| CDK20 | 88,68 | 0,30 | 0,08 | 3,54 | 4,00E-04 |
| MOK | 81,30 | 0,53 | 0,15 | 3,54 | 4,00E-04 |
| EMILIN1 | 55,51 | 0,42 | 0,12 | 3,54 | 4,01E-04 |
| SLC12A4 | 491,33 | 0,22 | 0,06 | 3,54 | 4,02E-04 |
| DNAJC7 | 2194,21 | -0,07 | 0,02 | -3,54 | 4,02E-04 |
| ITCH | 5733,78 | -0,13 | 0,04 | -3,54 | 4,02E-04 |
| CABP4 | 43,61 | 0,42 | 0,12 | 3,54 | 4,03E-04 |
| CSTF2T | 1371,49 | -0,15 | 0,04 | -3,54 | 4,05E-04 |
| PMP22 | 46,02 | 0,61 | 0,17 | 3,54 | 4,05E-04 |
| SGTA | 184,67 | 0,31 | 0,09 | 3,54 | 4,05E-04 |
| PTGFRN | 72,03 | 0,45 | 0,13 | 3,54 | 4,05E-04 |
| CEP120 | 2042,26 | -0,15 | 0,04 | -3,54 | 4,06E-04 |
| GPT2 | 58,16 | 0,60 | 0,17 | 3,54 | 4,08E-04 |
| PIDD1 | 150,03 | 0,30 | 0,08 | 3,53 | 4,08E-04 |
| OSBP2 | 1241,66 | 0,46 | 0,13 | 3,53 | 4,14E-04 |
| IBTK | 1900,38 | -0,16 | 0,05 | -3,53 | 4,16E-04 |
| SNTG2 | 66,83 | 0,57 | 0,16 | 3,53 | 4,17E-04 |
| LSR | 113,03 | 0,28 | 0,08 | 3,53 | 4,17E-04 |
| TTLL12 | 229,14 | 0,24 | 0,07 | 3,53 | 4,18E-04 |
| SLC2A8 | 68,89 | 0,40 | 0,11 | 3,53 | 4,19E-04 |
| LPCAT4 | 355,65 | 0,19 | 0,06 | 3,53 | 4,20E-04 |
| SLC29A2 | 52,58 | 0,40 | 0,11 | 3,53 | 4,20E-04 |
| ADSS2 | 2443,61 | -0,15 | 0,04 | -3,53 | 4,20E-04 |
| ZNF830 | 724,52 | -0,21 | 0,06 | -3,53 | 4,20E-04 |
| PRPS1 | 761,40 | -0,16 | 0,05 | -3,53 | 4,22E-04 |
| ULK3 | 295,21 | 0,25 | 0,07 | 3,52 | 4,25E-04 |
| RGS7 | 31,55 | 0,44 | 0,13 | 3,52 | 4,26E-04 |
| NPEPL1 | 342,71 | 0,29 | 0,08 | 3,52 | 4,28E-04 |
| CCDC65 | 139,41 | -0,27 | 0,08 | -3,52 | 4,30E-04 |
| USP47 | 3352,25 | -0,20 | 0,06 | -3,52 | 4,30E-04 |
| TTLL11 | 148,31 | 0,21 | 0,06 | 3,52 | 4,30E-04 |
| NRBP2 | 120,85 | 0,32 | 0,09 | 3,52 | 4,32E-04 |
| DHX37 | 299,69 | 0,25 | 0,07 | 3,52 | 4,33E-04 |
| BCL7B | 299,73 | 0,13 | 0,04 | 3,52 | 4,33E-04 |
| DPM1 | 749,34 | -0,25 | 0,07 | -3,52 | 4,36E-04 |
| NPHP4 | 269,10 | 0,41 | 0,12 | 3,52 | 4,37E-04 |
| SUMO1 | 1517,06 | -0,21 | 0,06 | -3,52 | 4,37E-04 |
| CCND1 | 58,76 | 0,55 | 0,16 | 3,52 | 4,37E-04 |
| SNIP1 | 1256,03 | -0,10 | 0,03 | -3,52 | 4,38E-04 |
| SBK1 | 138,26 | 0,45 | 0,13 | 3,52 | 4,39E-04 |
| XRCC4 | 342,15 | -0,33 | 0,09 | -3,52 | 4,39E-04 |
| CERS4 | 374,33 | 0,19 | 0,05 | 3,51 | 4,40E-04 |
| SAMD10 | 50,67 | 0,36 | 0,10 | 3,51 | 4,41E-04 |

| | | | | | |
|---|---|---|---|---|---|
| SRGAP3 | 254,57 | 0,42 | 0,12 | 3,51 | 4,42E-04 |
| FAM114A1 | 109,43 | 0,23 | 0,06 | 3,51 | 4,43E-04 |
| ARHGEF19 | 98,48 | 0,31 | 0,09 | 3,51 | 4,45E-04 |
| LMF1 | 312,80 | 0,27 | 0,08 | 3,51 | 4,46E-04 |
| NOXA1 | 117,76 | 0,28 | 0,08 | 3,51 | 4,45E-04 |
| DCTN5 | 2068,37 | -0,12 | 0,04 | -3,51 | 4,45E-04 |
| FRMD6 | 57,85 | 0,62 | 0,18 | 3,51 | 4,47E-04 |
| LZIC | 649,88 | -0,13 | 0,04 | -3,51 | 4,47E-04 |
| OBI1 | 538,78 | -0,28 | 0,08 | -3,51 | 4,46E-04 |
| UTP18 | 930,12 | -0,14 | 0,04 | -3,51 | 4,46E-04 |
| WIZ | 263,53 | 0,39 | 0,11 | 3,51 | 4,47E-04 |
| PODN | 52,74 | 0,64 | 0,18 | 3,51 | 4,49E-04 |
| SLC1A5 | 1336,33 | 0,38 | 0,11 | 3,51 | 4,49E-04 |
| FAM171A1 | 68,62 | 0,32 | 0,09 | 3,51 | 4,49E-04 |
| PDS5A | 3952,98 | -0,11 | 0,03 | -3,51 | 4,51E-04 |
| LZTR1 | 445,49 | 0,18 | 0,05 | 3,51 | 4,51E-04 |
| HDDC2 | 571,75 | -0,19 | 0,06 | -3,51 | 4,51E-04 |
| PER1 | 168,20 | 0,42 | 0,12 | 3,51 | 4,52E-04 |
| HNRNPU | 24366,11 | -0,13 | 0,04 | -3,51 | 4,54E-04 |
| KIF5B | 4575,67 | -0,19 | 0,06 | -3,51 | 4,54E-04 |
| GP5 | 55,50 | 0,39 | 0,11 | 3,51 | 4,56E-04 |
| DDX51 | 172,82 | 0,34 | 0,10 | 3,51 | 4,56E-04 |
| RNF103 | 1370,18 | -0,13 | 0,04 | -3,50 | 4,57E-04 |
| CC2D1A | 140,42 | 0,42 | 0,12 | 3,50 | 4,58E-04 |
| WHRN | 73,69 | 0,48 | 0,14 | 3,50 | 4,58E-04 |
| FHL2 | 55,01 | 0,48 | 0,14 | 3,50 | 4,59E-04 |
| TPRN | 93,91 | 0,30 | 0,09 | 3,50 | 4,58E-04 |
| DYRK3 | 137,19 | 0,41 | 0,12 | 3,50 | 4,60E-04 |
| PNLDC1 | 40,07 | 0,37 | 0,11 | 3,50 | 4,62E-04 |
| MNAT1 | 399,36 | -0,18 | 0,05 | -3,50 | 4,62E-04 |
| DNAJC9 | 898,31 | -0,14 | 0,04 | -3,50 | 4,64E-04 |
| KCNK5 | 35,37 | 0,55 | 0,16 | 3,50 | 4,63E-04 |
| FDXR | 112,87 | 0,25 | 0,07 | 3,50 | 4,63E-04 |
| ADD3 | 7301,78 | -0,15 | 0,04 | -3,50 | 4,64E-04 |
| RRM2 | 73,73 | 0,38 | 0,11 | 3,50 | 4,65E-04 |
| CHD9 | 3527,98 | -0,13 | 0,04 | -3,50 | 4,65E-04 |
| HPS5 | 1068,32 | -0,09 | 0,03 | -3,50 | 4,67E-04 |
| ARG2 | 46,13 | 0,48 | 0,14 | 3,50 | 4,67E-04 |
| SRFBP1 | 377,81 | -0,17 | 0,05 | -3,50 | 4,67E-04 |
| COX6C | 445,20 | -0,25 | 0,07 | -3,50 | 4,69E-04 |
| CCDC186 | 2666,82 | -0,22 | 0,06 | -3,50 | 4,69E-04 |
| PPP1R8 | 1077,60 | -0,14 | 0,04 | -3,50 | 4,71E-04 |
| GATA1 | 542,57 | 0,39 | 0,11 | 3,50 | 4,72E-04 |
| TTYH2 | 263,33 | 0,30 | 0,09 | 3,50 | 4,74E-04 |
| ALKBH8 | 549,80 | -0,18 | 0,05 | -3,50 | 4,74E-04 |
| PRKG1 | 67,92 | 0,55 | 0,16 | 3,50 | 4,73E-04 |

| | | | | | |
|---|---|---|---|---|---|
| MROH1 | 485,29 | 0,38 | 0,11 | 3,49 | 4,74E-04 |
| PLAA | 753,44 | -0,11 | 0,03 | -3,49 | 4,75E-04 |
| CATSPERG | 265,45 | 0,24 | 0,07 | 3,49 | 4,76E-04 |
| ING3 | 992,31 | -0,18 | 0,05 | -3,49 | 4,76E-04 |
| DLG2 | 92,27 | 0,49 | 0,14 | 3,49 | 4,79E-04 |
| HMGCS1 | 734,15 | -0,15 | 0,04 | -3,49 | 4,82E-04 |
| ZNF653 | 124,93 | 0,21 | 0,06 | 3,49 | 4,81E-04 |
| DKC1 | 840,25 | -0,11 | 0,03 | -3,49 | 4,82E-04 |
| RSL1D1 | 1626,73 | -0,16 | 0,05 | -3,49 | 4,82E-04 |
| NEURL2 | 36,31 | 0,24 | 0,07 | 3,49 | 4,82E-04 |
| RGS5 | 28,64 | 0,51 | 0,15 | 3,49 | 4,83E-04 |
| SPOP | 2833,16 | -0,10 | 0,03 | -3,49 | 4,85E-04 |
| CHCHD7 | 849,24 | -0,14 | 0,04 | -3,49 | 4,86E-04 |
| AIFM3 | 131,67 | 0,36 | 0,10 | 3,49 | 4,87E-04 |
| AACS | 340,19 | 0,28 | 0,08 | 3,49 | 4,87E-04 |
| TEX22 | 65,38 | 0,27 | 0,08 | 3,49 | 4,88E-04 |
| PNN | 2584,24 | -0,15 | 0,04 | -3,49 | 4,88E-04 |
| TCP1 | 2542,60 | -0,12 | 0,04 | -3,49 | 4,89E-04 |
| TAF1C | 208,58 | 0,37 | 0,11 | 3,49 | 4,90E-04 |
| ZNF841 | 892,30 | -0,13 | 0,04 | -3,48 | 4,92E-04 |
| COMTD1 | 101,40 | 0,20 | 0,06 | 3,48 | 4,94E-04 |
| POMZP3 | 61,30 | 0,26 | 0,08 | 3,48 | 4,96E-04 |
| SNRPE | 338,58 | -0,27 | 0,08 | -3,48 | 4,96E-04 |
| HSPA8 | 12437,01 | -0,19 | 0,05 | -3,48 | 4,98E-04 |
| OPA1 | 3339,86 | -0,17 | 0,05 | -3,48 | 4,98E-04 |
| C19orf44 | 45,97 | 0,36 | 0,10 | 3,48 | 4,99E-04 |
| TM9SF3 | 4427,59 | -0,20 | 0,06 | -3,48 | 5,02E-04 |
| SUB1 | 2524,40 | -0,25 | 0,07 | -3,48 | 5,03E-04 |
| ZNF417 | 1310,48 | -0,12 | 0,04 | -3,48 | 5,05E-04 |
| KIF2A | 2691,46 | -0,16 | 0,05 | -3,48 | 5,05E-04 |
| OTULINL | 1724,25 | -0,16 | 0,05 | -3,48 | 5,06E-04 |
| MINAR1 | 66,14 | 0,41 | 0,12 | 3,48 | 5,07E-04 |
| ZNF37A | 1052,96 | -0,22 | 0,06 | -3,48 | 5,08E-04 |
| ZNF764 | 128,56 | 0,24 | 0,07 | 3,48 | 5,09E-04 |
| ACSBG1 | 66,85 | 0,57 | 0,16 | 3,48 | 5,10E-04 |
| GIMAP7 | 2888,73 | -0,28 | 0,08 | -3,48 | 5,10E-04 |
| EIF3D | 3031,13 | -0,13 | 0,04 | -3,47 | 5,14E-04 |
| TTLL7 | 85,20 | 0,47 | 0,14 | 3,47 | 5,16E-04 |
| GIT1 | 348,20 | 0,27 | 0,08 | 3,47 | 5,18E-04 |
| SNORC | 54,19 | 0,40 | 0,11 | 3,47 | 5,19E-04 |
| ZDHHC12 | 93,23 | 0,28 | 0,08 | 3,47 | 5,20E-04 |
| ARMCX5 | 354,55 | -0,14 | 0,04 | -3,47 | 5,22E-04 |
| TMEM175 | 268,21 | 0,28 | 0,08 | 3,47 | 5,23E-04 |
| RABGGTA | 93,94 | 0,32 | 0,09 | 3,47 | 5,23E-04 |
| ZNF776 | 1280,11 | -0,18 | 0,05 | -3,47 | 5,24E-04 |
| CCNB2 | 30,98 | 0,43 | 0,12 | 3,47 | 5,24E-04 |

| | | | | | |
|---|---|---|---|---|---|
| SLC25A27 | 38,05 | 0,41 | 0,12 | 3,47 | 5,25E-04 |
| UTP23 | 807,02 | -0,17 | 0,05 | -3,47 | 5,28E-04 |
| PROSER3 | 99,95 | 0,38 | 0,11 | 3,46 | 5,31E-04 |
| SGIP1 | 54,76 | 0,66 | 0,19 | 3,46 | 5,32E-04 |
| BACE1 | 74,19 | 0,40 | 0,12 | 3,46 | 5,33E-04 |
| RPF1 | 648,69 | -0,21 | 0,06 | -3,46 | 5,34E-04 |
| SPEN | 6107,18 | -0,11 | 0,03 | -3,46 | 5,35E-04 |
| XPA | 590,62 | -0,20 | 0,06 | -3,46 | 5,40E-04 |
| ANK1 | 1360,99 | 0,44 | 0,13 | 3,46 | 5,40E-04 |
| ALDH4A1 | 33,06 | 0,43 | 0,12 | 3,46 | 5,40E-04 |
| RPL6 | 5249,55 | -0,31 | 0,09 | -3,46 | 5,42E-04 |
| FIZ1 | 111,09 | 0,31 | 0,09 | 3,46 | 5,43E-04 |
| CUL4B | 2262,86 | -0,16 | 0,05 | -3,46 | 5,44E-04 |
| PTS | 445,29 | -0,11 | 0,03 | -3,46 | 5,44E-04 |
| IVNS1ABP | 7481,85 | -0,17 | 0,05 | -3,46 | 5,45E-04 |
| PROX2 | 24,96 | 0,47 | 0,14 | 3,46 | 5,45E-04 |
| ACAA2 | 621,51 | -0,15 | 0,04 | -3,46 | 5,46E-04 |
| CD163L1 | 53,05 | 0,66 | 0,19 | 3,46 | 5,46E-04 |
| ZNF598 | 255,91 | 0,26 | 0,08 | 3,45 | 5,52E-04 |
| GSTO2 | 75,54 | 0,34 | 0,10 | 3,45 | 5,52E-04 |
| INTS10 | 1642,65 | -0,09 | 0,03 | -3,45 | 5,53E-04 |
| CAV2 | 63,51 | 0,57 | 0,16 | 3,45 | 5,54E-04 |
| FAH | 238,37 | 0,21 | 0,06 | 3,45 | 5,54E-04 |
| LRIF1 | 790,43 | -0,19 | 0,05 | -3,45 | 5,55E-04 |
| TXNRD2 | 708,56 | 0,21 | 0,06 | 3,45 | 5,55E-04 |
| ZNF777 | 84,44 | 0,34 | 0,10 | 3,45 | 5,56E-04 |
| GNAI1 | 91,20 | 0,49 | 0,14 | 3,45 | 5,56E-04 |
| PNRC2 | 3766,78 | -0,24 | 0,07 | -3,45 | 5,56E-04 |
| DMWD | 205,32 | 0,29 | 0,08 | 3,45 | 5,59E-04 |
| RAP1GAP | 167,99 | 0,87 | 0,25 | 3,45 | 5,59E-04 |
| TRMT2A | 534,12 | 0,14 | 0,04 | 3,45 | 5,59E-04 |
| PYGM | 158,65 | 0,33 | 0,09 | 3,45 | 5,60E-04 |
| TMX4 | 6882,88 | -0,17 | 0,05 | -3,45 | 5,60E-04 |
| TTC1 | 1053,31 | -0,14 | 0,04 | -3,45 | 5,61E-04 |
| ZNF430 | 1310,88 | -0,12 | 0,03 | -3,45 | 5,62E-04 |
| UBE2C | 35,16 | 0,34 | 0,10 | 3,45 | 5,63E-04 |
| UBXN4 | 3612,41 | -0,15 | 0,04 | -3,45 | 5,65E-04 |
| GRIN3A | 42,48 | 0,60 | 0,18 | 3,45 | 5,67E-04 |
| PTP4A3 | 533,70 | 0,22 | 0,06 | 3,45 | 5,67E-04 |
| DCLK2 | 53,58 | 0,69 | 0,20 | 3,45 | 5,67E-04 |
| NASP | 1709,97 | -0,11 | 0,03 | -3,45 | 5,69E-04 |
| DLGAP4 | 236,21 | 0,48 | 0,14 | 3,45 | 5,70E-04 |
| TBC1D15 | 2102,25 | -0,19 | 0,05 | -3,44 | 5,74E-04 |
| C20orf204 | 85,01 | 0,33 | 0,10 | 3,44 | 5,74E-04 |
| ARHGAP12 | 1439,30 | -0,18 | 0,05 | -3,44 | 5,74E-04 |
| MPO | 142,86 | 0,41 | 0,12 | 3,44 | 5,75E-04 |

| | | | | | |
|---|---|---|---|---|---|
| ATP8A1 | 5254,92 | -0,20 | 0,06 | -3,44 | 5,75E-04 |
| GALNT1 | 1759,29 | -0,23 | 0,07 | -3,44 | 5,78E-04 |
| P4HA1 | 1001,02 | -0,16 | 0,05 | -3,44 | 5,80E-04 |
| GARIN1A | 63,45 | 0,42 | 0,12 | 3,44 | 5,80E-04 |
| CHDH | 54,88 | 0,72 | 0,21 | 3,44 | 5,81E-04 |
| SETD3 | 1590,43 | -0,11 | 0,03 | -3,44 | 5,82E-04 |
| PNPLA8 | 1797,74 | -0,26 | 0,08 | -3,44 | 5,82E-04 |
| DNAJB11 | 727,12 | -0,11 | 0,03 | -3,44 | 5,84E-04 |
| LARS1 | 2678,25 | -0,13 | 0,04 | -3,44 | 5,88E-04 |
| NAA35 | 860,34 | -0,12 | 0,04 | -3,44 | 5,87E-04 |
| PTPN22 | 1426,31 | -0,19 | 0,06 | -3,44 | 5,86E-04 |
| MYO19 | 778,35 | 0,17 | 0,05 | 3,44 | 5,87E-04 |
| MPIG6B | 58,40 | 0,59 | 0,17 | 3,44 | 5,87E-04 |
| ITIH4 | 28,61 | 0,36 | 0,11 | 3,44 | 5,88E-04 |
| SMIM5 | 569,37 | 0,37 | 0,11 | 3,44 | 5,91E-04 |
| CACFD1 | 110,32 | 0,24 | 0,07 | 3,44 | 5,90E-04 |
| LTC4S | 133,56 | 0,24 | 0,07 | 3,44 | 5,92E-04 |
| ATN1 | 74,77 | 0,48 | 0,14 | 3,44 | 5,92E-04 |
| FAAP100 | 281,24 | 0,31 | 0,09 | 3,43 | 5,93E-04 |
| PPWD1 | 1998,76 | -0,19 | 0,06 | -3,43 | 5,94E-04 |
| POLL | 803,95 | 0,24 | 0,07 | 3,43 | 5,95E-04 |
| NME6 | 1063,83 | -0,08 | 0,02 | -3,43 | 5,97E-04 |
| RIOX2 | 1063,78 | -0,15 | 0,04 | -3,43 | 5,99E-04 |
| PBDC1 | 242,76 | -0,16 | 0,05 | -3,43 | 6,03E-04 |
| BEND3 | 57,43 | 0,44 | 0,13 | 3,43 | 6,03E-04 |
| IK | 3129,77 | -0,14 | 0,04 | -3,43 | 6,05E-04 |
| SCN8A | 101,01 | 0,61 | 0,18 | 3,43 | 6,05E-04 |
| CCT8 | 2147,10 | -0,18 | 0,05 | -3,43 | 6,06E-04 |
| HOOK2 | 222,79 | 0,19 | 0,05 | 3,43 | 6,09E-04 |
| PRP4K | 3511,24 | -0,13 | 0,04 | -3,43 | 6,10E-04 |
| SPPL2B | 326,13 | 0,32 | 0,09 | 3,43 | 6,10E-04 |
| NEURL1 | 185,10 | 0,32 | 0,09 | 3,43 | 6,11E-04 |
| ABCA13 | 416,93 | 0,56 | 0,16 | 3,43 | 6,11E-04 |
| LRRC20 | 61,82 | 0,29 | 0,09 | 3,42 | 6,15E-04 |
| SEZ6L | 74,30 | 0,53 | 0,16 | 3,42 | 6,17E-04 |
| PSMD6 | 1954,69 | -0,12 | 0,03 | -3,42 | 6,17E-04 |
| CFDP1 | 503,06 | -0,11 | 0,03 | -3,42 | 6,17E-04 |
| ACADM | 800,59 | -0,20 | 0,06 | -3,42 | 6,19E-04 |
| ZNF121 | 1956,82 | -0,15 | 0,04 | -3,42 | 6,19E-04 |
| YJU2B | 419,30 | 0,19 | 0,05 | 3,42 | 6,22E-04 |
| ABCC6 | 99,39 | 0,46 | 0,14 | 3,42 | 6,22E-04 |
| TRPT1 | 99,98 | 0,17 | 0,05 | 3,42 | 6,24E-04 |
| CDPF1 | 107,80 | 0,16 | 0,05 | 3,42 | 6,32E-04 |
| MYH3 | 234,77 | 0,27 | 0,08 | 3,42 | 6,32E-04 |
| YPEL3 | 2399,18 | 0,30 | 0,09 | 3,42 | 6,31E-04 |
| PCM1 | 7376,17 | -0,14 | 0,04 | -3,42 | 6,32E-04 |

| | | | | | |
|---|---|---|---|---|---|
| STARD13 | 54,66 | 0,60 | 0,17 | 3,42 | 6,33E-04 |
| PLCH2 | 229,48 | 0,41 | 0,12 | 3,42 | 6,32E-04 |
| TMEM184C | 1257,06 | -0,13 | 0,04 | -3,42 | 6,33E-04 |
| POLR2E | 510,41 | 0,17 | 0,05 | 3,42 | 6,34E-04 |
| ZNF33B | 1465,44 | -0,16 | 0,05 | -3,42 | 6,34E-04 |
| TRIM3 | 139,98 | 0,29 | 0,09 | 3,42 | 6,35E-04 |
| ABCB7 | 773,95 | -0,13 | 0,04 | -3,42 | 6,38E-04 |
| AIFM2 | 121,03 | 0,28 | 0,08 | 3,41 | 6,39E-04 |
| ETFDH | 657,86 | -0,13 | 0,04 | -3,41 | 6,40E-04 |
| CBLN3 | 41,67 | 0,39 | 0,11 | 3,41 | 6,40E-04 |
| MYPOP | 61,13 | 0,37 | 0,11 | 3,41 | 6,39E-04 |
| KATNAL2 | 44,85 | 0,57 | 0,17 | 3,41 | 6,41E-04 |
| SOX12 | 50,18 | 0,44 | 0,13 | 3,41 | 6,42E-04 |
| ATAD3A | 153,60 | 0,21 | 0,06 | 3,41 | 6,42E-04 |
| MED4 | 1031,46 | -0,14 | 0,04 | -3,41 | 6,42E-04 |
| MAP2K2 | 1044,20 | 0,22 | 0,07 | 3,41 | 6,44E-04 |
| RIF1 | 4889,23 | -0,17 | 0,05 | -3,41 | 6,44E-04 |
| RAMAC | 501,46 | -0,19 | 0,06 | -3,41 | 6,44E-04 |
| KLHL20 | 1048,70 | -0,13 | 0,04 | -3,41 | 6,44E-04 |
| SLC6A9 | 123,04 | 0,50 | 0,15 | 3,41 | 6,45E-04 |
| DNAJC4 | 138,57 | 0,23 | 0,07 | 3,41 | 6,47E-04 |
| RGPD3 | 35,24 | 0,48 | 0,14 | 3,41 | 6,51E-04 |
| PLPP7 | 35,99 | 0,46 | 0,14 | 3,41 | 6,53E-04 |
| SLC6A13 | 37,91 | 0,58 | 0,17 | 3,41 | 6,53E-04 |
| PKP2 | 57,14 | 0,47 | 0,14 | 3,41 | 6,52E-04 |
| WDR53 | 221,07 | -0,11 | 0,03 | -3,41 | 6,54E-04 |
| GINS1 | 44,85 | 0,40 | 0,12 | 3,41 | 6,55E-04 |
| PJA2 | 8043,88 | -0,18 | 0,05 | -3,41 | 6,55E-04 |
| CEP135 | 1125,92 | -0,15 | 0,04 | -3,41 | 6,56E-04 |
| MEAF6 | 1147,20 | -0,11 | 0,03 | -3,41 | 6,59E-04 |
| TADA3 | 910,16 | 0,18 | 0,05 | 3,41 | 6,61E-04 |
| USP54 | 144,39 | 0,32 | 0,09 | 3,40 | 6,65E-04 |
| KBTBD4 | 719,22 | -0,10 | 0,03 | -3,40 | 6,64E-04 |
| STPG1 | 102,01 | 0,30 | 0,09 | 3,40 | 6,65E-04 |
| CANX | 7690,09 | -0,12 | 0,04 | -3,40 | 6,64E-04 |
| WDR5 | 361,30 | 0,12 | 0,04 | 3,40 | 6,68E-04 |
| CKAP2L | 23,64 | 0,43 | 0,13 | 3,40 | 6,68E-04 |
| OLIG1 | 144,55 | 0,32 | 0,10 | 3,40 | 6,68E-04 |
| SMPD1 | 162,14 | 0,25 | 0,07 | 3,40 | 6,67E-04 |
| FASTKD5 | 737,19 | -0,11 | 0,03 | -3,40 | 6,68E-04 |
| BRIX1 | 353,36 | -0,21 | 0,06 | -3,40 | 6,69E-04 |
| PRR11 | 128,82 | 0,21 | 0,06 | 3,40 | 6,75E-04 |
| CPSF2 | 2753,18 | -0,11 | 0,03 | -3,40 | 6,75E-04 |
| DOK4 | 74,93 | 0,40 | 0,12 | 3,40 | 6,76E-04 |
| PMFBP1 | 61,84 | 0,45 | 0,13 | 3,40 | 6,79E-04 |
| RPH3A | 61,89 | 0,77 | 0,23 | 3,40 | 6,79E-04 |

| | | | | | |
|---|---|---|---|---|---|
| TRAIP | 40,55 | 0,27 | 0,08 | 3,40 | 6,80E-04 |
| TAF7 | 1262,59 | -0,29 | 0,09 | -3,40 | 6,85E-04 |
| KCNMA1 | 177,60 | 0,79 | 0,23 | 3,40 | 6,85E-04 |
| TMF1 | 3791,58 | -0,18 | 0,05 | -3,40 | 6,86E-04 |
| CLCN7 | 650,28 | 0,35 | 0,10 | 3,39 | 6,87E-04 |
| PSMB1 | 1872,26 | -0,16 | 0,05 | -3,39 | 6,87E-04 |
| CYTIP | 10316,32 | -0,15 | 0,04 | -3,39 | 6,88E-04 |
| ZCCHC24 | 106,96 | 0,28 | 0,08 | 3,39 | 6,89E-04 |
| ZNF615 | 454,25 | -0,20 | 0,06 | -3,39 | 6,89E-04 |
| PPP6R1 | 1128,68 | 0,32 | 0,10 | 3,39 | 6,91E-04 |
| TNFRSF11A | 73,00 | 0,51 | 0,15 | 3,39 | 6,97E-04 |
| SLC2A1 | 2006,56 | 0,28 | 0,08 | 3,39 | 6,97E-04 |
| VEZT | 1255,17 | -0,13 | 0,04 | -3,39 | 6,98E-04 |
| NAGS | 59,30 | 0,25 | 0,07 | 3,39 | 6,98E-04 |
| CARNS1 | 87,22 | 0,44 | 0,13 | 3,39 | 6,99E-04 |
| TSN | 1345,76 | -0,15 | 0,04 | -3,39 | 7,00E-04 |
| ADCY4 | 123,39 | 0,38 | 0,11 | 3,39 | 7,00E-04 |
| U2AF1L4 | 33,25 | 0,27 | 0,08 | 3,39 | 7,01E-04 |
| MFAP1 | 964,59 | -0,17 | 0,05 | -3,39 | 7,02E-04 |
| CCSER2 | 2864,10 | -0,16 | 0,05 | -3,39 | 7,03E-04 |
| RNF126 | 473,40 | 0,20 | 0,06 | 3,39 | 7,04E-04 |
| MIS18BP1 | 2515,09 | -0,20 | 0,06 | -3,39 | 7,08E-04 |
| PRMT9 | 503,53 | -0,13 | 0,04 | -3,39 | 7,08E-04 |
| ZNF224 | 2181,72 | -0,12 | 0,04 | -3,39 | 7,10E-04 |
| CALD1 | 95,57 | 0,51 | 0,15 | 3,39 | 7,09E-04 |
| YIF1B | 147,83 | 0,21 | 0,06 | 3,38 | 7,15E-04 |
| HEXIM2 | 70,24 | 0,20 | 0,06 | 3,38 | 7,16E-04 |
| DNAJC2 | 553,26 | -0,12 | 0,04 | -3,38 | 7,17E-04 |
| SLC39A13 | 172,57 | 0,24 | 0,07 | 3,38 | 7,17E-04 |
| SPOCK1 | 51,71 | 0,52 | 0,15 | 3,38 | 7,17E-04 |
| NDUFA13 | 65,37 | 0,28 | 0,08 | 3,38 | 7,19E-04 |
| C2CD2L | 382,27 | 0,27 | 0,08 | 3,38 | 7,19E-04 |
| AP3B1 | 4215,74 | -0,15 | 0,04 | -3,38 | 7,22E-04 |
| MFSD12 | 466,69 | 0,27 | 0,08 | 3,38 | 7,24E-04 |
| ORC1 | 40,90 | 0,38 | 0,11 | 3,38 | 7,24E-04 |
| CBX8 | 143,75 | 0,21 | 0,06 | 3,38 | 7,25E-04 |
| CEP57 | 1454,97 | -0,21 | 0,06 | -3,38 | 7,25E-04 |
| PSMC1 | 1125,73 | -0,11 | 0,03 | -3,38 | 7,27E-04 |
| POU6F1 | 182,56 | 0,27 | 0,08 | 3,38 | 7,30E-04 |
| DIAPH3 | 59,40 | 0,47 | 0,14 | 3,38 | 7,31E-04 |
| TMEM150A | 59,17 | 0,31 | 0,09 | 3,38 | 7,32E-04 |
| EFCAB8 | 105,05 | 0,38 | 0,11 | 3,38 | 7,33E-04 |
| DUSP11 | 1199,10 | -0,11 | 0,03 | -3,38 | 7,34E-04 |
| SAC3D1 | 55,74 | 0,23 | 0,07 | 3,38 | 7,36E-04 |
| ZNF3 | 826,02 | -0,07 | 0,02 | -3,38 | 7,36E-04 |
| HSDL2 | 2096,61 | -0,21 | 0,06 | -3,38 | 7,35E-04 |

| | | | | | |
|---|---|---|---|---|---|
| POC5 | 600,10 | -0,14 | 0,04 | -3,38 | 7,36E-04 |
| OTX1 | 49,79 | 0,57 | 0,17 | 3,38 | 7,37E-04 |
| NCBP2 | 1333,18 | -0,14 | 0,04 | -3,38 | 7,37E-04 |
| PTPN2 | 2067,34 | -0,11 | 0,03 | -3,38 | 7,37E-04 |
| CROT | 438,16 | -0,16 | 0,05 | -3,37 | 7,46E-04 |
| ARID4B | 6598,63 | -0,17 | 0,05 | -3,37 | 7,46E-04 |
| RLIM | 5528,49 | -0,17 | 0,05 | -3,37 | 7,46E-04 |
| PLK1 | 81,26 | 0,30 | 0,09 | 3,37 | 7,48E-04 |
| HMGN3 | 468,50 | -0,17 | 0,05 | -3,37 | 7,52E-04 |
| PDE1B | 235,13 | 0,28 | 0,08 | 3,37 | 7,52E-04 |
| ILF2 | 1374,05 | -0,15 | 0,05 | -3,37 | 7,55E-04 |
| PARN | 1227,89 | -0,08 | 0,02 | -3,37 | 7,58E-04 |
| KLRB1 | 667,19 | -0,36 | 0,11 | -3,37 | 7,59E-04 |
| SDC4 | 75,05 | 0,31 | 0,09 | 3,37 | 7,59E-04 |
| ABTB2 | 146,40 | 0,40 | 0,12 | 3,37 | 7,62E-04 |
| C1orf56 | 31,95 | 0,24 | 0,07 | 3,37 | 7,64E-04 |
| USP42 | 399,35 | 0,19 | 0,06 | 3,37 | 7,64E-04 |
| RPAP3 | 1396,46 | -0,19 | 0,06 | -3,36 | 7,67E-04 |
| MMUT | 532,30 | -0,18 | 0,05 | -3,36 | 7,68E-04 |
| MUC20 | 31,56 | 0,50 | 0,15 | 3,36 | 7,69E-04 |
| MAST2 | 301,96 | 0,22 | 0,07 | 3,36 | 7,68E-04 |
| FANK1 | 46,36 | 0,48 | 0,14 | 3,36 | 7,69E-04 |
| SSNA1 | 491,19 | 0,14 | 0,04 | 3,36 | 7,70E-04 |
| MFSD2B | 263,20 | 0,39 | 0,12 | 3,36 | 7,71E-04 |
| TBK1 | 1532,14 | -0,21 | 0,06 | -3,36 | 7,73E-04 |
| METTL18 | 135,94 | -0,36 | 0,11 | -3,36 | 7,73E-04 |
| ATP13A2 | 229,01 | 0,33 | 0,10 | 3,36 | 7,75E-04 |
| NDST2 | 94,88 | 0,24 | 0,07 | 3,36 | 7,82E-04 |
| MARK4 | 255,90 | 0,29 | 0,09 | 3,36 | 7,84E-04 |
| DTHD1 | 847,10 | -0,32 | 0,09 | -3,36 | 7,84E-04 |
| MCAT | 142,33 | 0,12 | 0,04 | 3,36 | 7,85E-04 |
| G6PC3 | 126,86 | 0,22 | 0,06 | 3,36 | 7,89E-04 |
| HECTD3 | 617,44 | 0,18 | 0,05 | 3,36 | 7,89E-04 |
| TNPO2 | 470,73 | 0,22 | 0,07 | 3,36 | 7,90E-04 |
| SMPD4 | 405,48 | 0,20 | 0,06 | 3,36 | 7,91E-04 |
| PNPLA2 | 850,94 | 0,31 | 0,09 | 3,36 | 7,93E-04 |
| CENPV | 90,96 | 0,21 | 0,06 | 3,36 | 7,93E-04 |
| UBA2 | 1601,63 | -0,15 | 0,05 | -3,36 | 7,93E-04 |
| FCGBP | 247,38 | 0,44 | 0,13 | 3,36 | 7,92E-04 |
| FAM107B | 4654,94 | -0,10 | 0,03 | -3,35 | 7,94E-04 |
| ZNF143 | 1380,02 | -0,12 | 0,04 | -3,35 | 8,02E-04 |
| NOL8 | 1485,31 | -0,12 | 0,04 | -3,35 | 8,02E-04 |
| SHQ1 | 595,71 | -0,11 | 0,03 | -3,35 | 8,04E-04 |
| RAB11B | 447,31 | 0,31 | 0,09 | 3,35 | 8,04E-04 |
| SMOX | 317,54 | 0,35 | 0,11 | 3,35 | 8,05E-04 |
| MRPS35 | 537,24 | -0,17 | 0,05 | -3,35 | 8,08E-04 |

| | | | | | |
|---|---|---|---|---|---|
| INTS1 | 966,72 | 0,34 | 0,10 | 3,35 | 8,08E-04 |
| SLC6A4 | 51,22 | 0,54 | 0,16 | 3,35 | 8,14E-04 |
| ACER2 | 100,34 | 0,21 | 0,06 | 3,35 | 8,15E-04 |
| GNPAT | 1273,12 | -0,09 | 0,03 | -3,35 | 8,21E-04 |
| MYB | 249,45 | 0,19 | 0,06 | 3,35 | 8,21E-04 |
| RLF | 2242,75 | -0,19 | 0,06 | -3,34 | 8,23E-04 |
| CWC27 | 495,87 | -0,17 | 0,05 | -3,34 | 8,24E-04 |
| TVP23B | 971,10 | -0,18 | 0,06 | -3,34 | 8,31E-04 |
| RNF157 | 406,43 | 0,25 | 0,08 | 3,34 | 8,30E-04 |
| KRR1 | 1325,84 | -0,22 | 0,07 | -3,34 | 8,31E-04 |
| TAF13 | 298,94 | -0,24 | 0,07 | -3,34 | 8,32E-04 |
| CD109 | 96,28 | 0,38 | 0,11 | 3,34 | 8,33E-04 |
| GDI2 | 4738,57 | -0,17 | 0,05 | -3,34 | 8,35E-04 |
| SLC4A2 | 396,28 | 0,25 | 0,07 | 3,34 | 8,36E-04 |
| KATNB1 | 273,38 | 0,20 | 0,06 | 3,34 | 8,39E-04 |
| PRPF40A | 4455,05 | -0,18 | 0,05 | -3,34 | 8,39E-04 |
| SLK | 4328,88 | -0,21 | 0,06 | -3,34 | 8,41E-04 |
| GPN1 | 426,33 | -0,12 | 0,04 | -3,34 | 8,42E-04 |
| CLDN15 | 159,69 | 0,31 | 0,09 | 3,34 | 8,43E-04 |
| TOR4A | 374,93 | 0,29 | 0,09 | 3,34 | 8,46E-04 |
| COPB1 | 2799,79 | -0,15 | 0,05 | -3,34 | 8,46E-04 |
| RNF26 | 69,90 | 0,31 | 0,09 | 3,34 | 8,47E-04 |
| EXOSC9 | 691,73 | -0,16 | 0,05 | -3,34 | 8,49E-04 |
| SMURF2 | 1909,06 | -0,10 | 0,03 | -3,34 | 8,51E-04 |
| PLEKHG4 | 188,90 | 0,24 | 0,07 | 3,34 | 8,51E-04 |
| STAP1 | 259,26 | -0,30 | 0,09 | -3,34 | 8,51E-04 |
| MMP14 | 51,03 | 0,39 | 0,12 | 3,34 | 8,52E-04 |
| DPH2 | 126,48 | 0,20 | 0,06 | 3,34 | 8,53E-04 |
| RPA2 | 1348,05 | -0,14 | 0,04 | -3,33 | 8,56E-04 |
| FBXL19 | 327,05 | 0,17 | 0,05 | 3,33 | 8,56E-04 |
| AHSA1 | 1039,95 | -0,11 | 0,03 | -3,33 | 8,55E-04 |
| STAG2 | 10083,91 | -0,22 | 0,07 | -3,33 | 8,61E-04 |
| TRMO | 1546,54 | -0,13 | 0,04 | -3,33 | 8,60E-04 |
| PHGDH | 83,41 | 0,54 | 0,16 | 3,33 | 8,59E-04 |
| MOSPD3 | 49,73 | 0,22 | 0,07 | 3,33 | 8,60E-04 |
| PODXL2 | 40,87 | 0,35 | 0,11 | 3,33 | 8,63E-04 |
| ARID4A | 3838,94 | -0,20 | 0,06 | -3,33 | 8,63E-04 |
| ABO | 62,99 | 0,62 | 0,19 | 3,33 | 8,65E-04 |
| SLF2 | 2008,01 | -0,10 | 0,03 | -3,33 | 8,66E-04 |
| DSN1 | 268,50 | -0,10 | 0,03 | -3,33 | 8,67E-04 |
| GGNBP2 | 2956,20 | -0,10 | 0,03 | -3,33 | 8,66E-04 |
| ALPK3 | 112,88 | 0,44 | 0,13 | 3,33 | 8,72E-04 |
| NUCB1 | 1088,00 | 0,28 | 0,08 | 3,33 | 8,72E-04 |
| SEMA7A | 301,29 | 0,25 | 0,07 | 3,33 | 8,73E-04 |
| HDAC10 | 57,56 | 0,31 | 0,09 | 3,33 | 8,75E-04 |
| DDX42 | 2814,42 | -0,09 | 0,03 | -3,33 | 8,77E-04 |

| | | | | | |
|---|---|---|---|---|---|
| SLC7A1 | 514,91 | 0,20 | 0,06 | 3,33 | 8,77E-04 |
| PWWP3A | 384,18 | 0,18 | 0,05 | 3,33 | 8,79E-04 |
| LARGE2 | 58,99 | 0,40 | 0,12 | 3,33 | 8,82E-04 |
| CENPT | 168,10 | 0,30 | 0,09 | 3,33 | 8,80E-04 |
| CCDC73 | 33,43 | 0,55 | 0,17 | 3,33 | 8,81E-04 |
| ESAM | 105,46 | 0,34 | 0,10 | 3,33 | 8,82E-04 |
| B3GNT7 | 36,86 | 0,50 | 0,15 | 3,33 | 8,82E-04 |
| RNF32 | 42,78 | 0,47 | 0,14 | 3,33 | 8,84E-04 |
| PCF11 | 6993,01 | -0,12 | 0,04 | -3,33 | 8,84E-04 |
| OXTR | 42,01 | 0,59 | 0,18 | 3,33 | 8,84E-04 |
| ADCY10 | 63,04 | 0,37 | 0,11 | 3,32 | 8,85E-04 |
| WRAP73 | 591,15 | 0,15 | 0,04 | 3,32 | 8,89E-04 |
| PRDX2 | 1083,68 | 0,35 | 0,11 | 3,32 | 8,88E-04 |
| SLC26A6 | 259,80 | 0,20 | 0,06 | 3,32 | 8,89E-04 |
| ZNF468 | 647,20 | -0,19 | 0,06 | -3,32 | 8,88E-04 |
| MPPED2 | 40,98 | 0,68 | 0,20 | 3,32 | 8,90E-04 |
| ZBTB38 | 2409,77 | -0,16 | 0,05 | -3,32 | 8,95E-04 |
| PELP1 | 222,95 | 0,25 | 0,07 | 3,32 | 8,96E-04 |
| BBS1 | 57,19 | 0,22 | 0,06 | 3,32 | 8,96E-04 |
| ATG5 | 970,25 | -0,22 | 0,07 | -3,32 | 8,99E-04 |
| LDHD | 38,45 | 0,48 | 0,14 | 3,32 | 8,99E-04 |
| COIL | 494,43 | -0,14 | 0,04 | -3,32 | 8,99E-04 |
| SLC13A3 | 46,78 | 0,56 | 0,17 | 3,32 | 9,00E-04 |
| KIFC2 | 324,83 | 0,22 | 0,07 | 3,32 | 9,00E-04 |
| AFG3L2 | 1118,95 | -0,08 | 0,02 | -3,32 | 9,01E-04 |
| MUTYH | 294,55 | 0,12 | 0,04 | 3,32 | 9,02E-04 |
| CHMP6 | 255,71 | 0,20 | 0,06 | 3,32 | 9,02E-04 |
| HSF1 | 348,71 | 0,26 | 0,08 | 3,32 | 9,05E-04 |
| ESR2 | 57,06 | 0,55 | 0,17 | 3,32 | 9,06E-04 |
| UNC13B | 113,65 | 0,54 | 0,16 | 3,32 | 9,09E-04 |
| SNED1 | 345,86 | 0,34 | 0,10 | 3,32 | 9,08E-04 |
| SCFD1 | 1956,30 | -0,16 | 0,05 | -3,32 | 9,10E-04 |
| ENO3 | 130,35 | 0,15 | 0,05 | 3,32 | 9,10E-04 |
| HENMT1 | 488,29 | -0,15 | 0,05 | -3,32 | 9,11E-04 |
| IGHMBP2 | 422,96 | 0,24 | 0,07 | 3,32 | 9,13E-04 |
| RNPEPL1 | 778,10 | 0,35 | 0,11 | 3,32 | 9,16E-04 |
| G3BP1 | 5230,45 | -0,10 | 0,03 | -3,31 | 9,17E-04 |
| CDK16 | 413,54 | 0,16 | 0,05 | 3,31 | 9,20E-04 |
| ZNF622 | 1119,07 | -0,16 | 0,05 | -3,31 | 9,20E-04 |
| MAN1A2 | 3108,48 | -0,18 | 0,05 | -3,31 | 9,21E-04 |
| E2F4 | 876,21 | 0,18 | 0,05 | 3,31 | 9,22E-04 |
| BAG6 | 2128,20 | 0,29 | 0,09 | 3,31 | 9,22E-04 |
| SCAF1 | 322,16 | 0,33 | 0,10 | 3,31 | 9,25E-04 |
| DDX3X | 24007,26 | -0,16 | 0,05 | -3,31 | 9,25E-04 |
| ICE1 | 1983,99 | -0,11 | 0,03 | -3,31 | 9,28E-04 |
| SLC35G5 | 25,87 | 0,31 | 0,09 | 3,31 | 9,28E-04 |

| | | | | | |
|---|---|---|---|---|---|
| SMARCD3 | 393,88 | 0,20 | 0,06 | 3,31 | 9,29E-04 |
| CCDC43 | 447,45 | -0,17 | 0,05 | -3,31 | 9,29E-04 |
| CAD | 482,78 | 0,25 | 0,07 | 3,31 | 9,31E-04 |
| LRFN2 | 39,02 | 0,62 | 0,19 | 3,31 | 9,31E-04 |
| EVC2 | 75,03 | 0,52 | 0,16 | 3,31 | 9,31E-04 |
| CDON | 75,62 | 0,48 | 0,15 | 3,31 | 9,33E-04 |
| MARCKSL1 | 100,36 | 0,30 | 0,09 | 3,31 | 9,34E-04 |
| ZBTB1 | 2077,33 | -0,22 | 0,07 | -3,31 | 9,34E-04 |
| TNNC2 | 30,71 | 0,43 | 0,13 | 3,31 | 9,36E-04 |
| RANBP6 | 991,60 | -0,28 | 0,09 | -3,31 | 9,38E-04 |
| SH2D1A | 904,71 | -0,29 | 0,09 | -3,31 | 9,38E-04 |
| RBM18 | 757,31 | -0,13 | 0,04 | -3,31 | 9,39E-04 |
| GBGT1 | 573,93 | 0,19 | 0,06 | 3,31 | 9,40E-04 |
| ELF1 | 7254,87 | -0,13 | 0,04 | -3,31 | 9,41E-04 |
| MFSD4A | 35,32 | 0,46 | 0,14 | 3,31 | 9,43E-04 |
| PPP2CA | 2585,28 | -0,14 | 0,04 | -3,31 | 9,45E-04 |
| NFS1 | 392,67 | 0,09 | 0,03 | 3,31 | 9,46E-04 |
| NIN | 16050,86 | -0,13 | 0,04 | -3,31 | 9,47E-04 |
| CNOT10 | 1192,68 | -0,08 | 0,02 | -3,31 | 9,48E-04 |
| BCLAF1 | 9390,76 | -0,19 | 0,06 | -3,30 | 9,51E-04 |
| SLC49A3 | 299,20 | 0,24 | 0,07 | 3,30 | 9,52E-04 |
| RASL11A | 67,70 | 0,26 | 0,08 | 3,30 | 9,52E-04 |
| TLCD2 | 55,88 | 0,42 | 0,13 | 3,30 | 9,54E-04 |
| ECHDC1 | 1483,50 | -0,19 | 0,06 | -3,30 | 9,55E-04 |
| SEC23B | 1694,64 | -0,10 | 0,03 | -3,30 | 9,56E-04 |
| NIFK | 500,36 | -0,19 | 0,06 | -3,30 | 9,59E-04 |
| LRSAM1 | 482,49 | 0,22 | 0,07 | 3,30 | 9,60E-04 |
| CACNA1D | 238,38 | 0,46 | 0,14 | 3,30 | 9,60E-04 |
| TRIP10 | 40,47 | 0,41 | 0,12 | 3,30 | 9,61E-04 |
| UROS | 666,22 | 0,09 | 0,03 | 3,30 | 9,63E-04 |
| ACTR6 | 499,38 | -0,25 | 0,07 | -3,30 | 9,67E-04 |
| TMED10 | 3674,61 | -0,12 | 0,04 | -3,30 | 9,68E-04 |
| ZDHHC9 | 52,48 | 0,28 | 0,09 | 3,30 | 9,69E-04 |
| NKIRAS2 | 448,27 | 0,14 | 0,04 | 3,30 | 9,73E-04 |
| CIC | 572,15 | 0,39 | 0,12 | 3,30 | 9,75E-04 |
| PWWP2A | 1862,36 | -0,08 | 0,02 | -3,30 | 9,77E-04 |
| PIK3CA | 2957,65 | -0,18 | 0,06 | -3,30 | 9,78E-04 |
| MATCAP1 | 87,22 | 0,31 | 0,09 | 3,30 | 9,79E-04 |
| DNAJC3 | 4057,36 | -0,17 | 0,05 | -3,30 | 9,80E-04 |
| PPP4R3B | 5288,11 | -0,16 | 0,05 | -3,30 | 9,84E-04 |
| GNB1L | 155,75 | 0,22 | 0,07 | 3,29 | 9,85E-04 |
| TCEA2 | 154,45 | 0,21 | 0,06 | 3,29 | 9,85E-04 |
| FURIN | 1385,79 | 0,34 | 0,10 | 3,29 | 9,89E-04 |
| TMEM87A | 1940,40 | -0,10 | 0,03 | -3,29 | 9,89E-04 |
| ASAH2 | 37,10 | 0,54 | 0,16 | 3,29 | 9,90E-04 |
| COPS4 | 440,35 | -0,23 | 0,07 | -3,29 | 9,92E-04 |

| | | | | | |
|---|---|---|---|---|---|
| ERP44 | 1664,60 | -0,15 | 0,04 | -3,29 | 9,95E-04 |
| RSPH9 | 25,27 | 0,51 | 0,15 | 3,29 | 9,95E-04 |
| PSPH | 73,78 | 0,22 | 0,07 | 3,29 | 9,96E-04 |
| C2 | 93,52 | 0,20 | 0,06 | 3,29 | 1,00E-03 |
| IQSEC2 | 141,81 | 0,36 | 0,11 | 3,29 | 1,01E-03 |
| CEP112 | 77,86 | 0,51 | 0,16 | 3,29 | 1,01E-03 |
| AUH | 842,41 | -0,11 | 0,03 | -3,29 | 1,01E-03 |
| CHML | 985,95 | -0,29 | 0,09 | -3,29 | 1,02E-03 |
| UBALD1 | 238,18 | 0,24 | 0,07 | 3,29 | 1,02E-03 |
| SMC4 | 1362,03 | -0,18 | 0,05 | -3,28 | 1,02E-03 |
| AAGAB | 1073,87 | -0,11 | 0,03 | -3,28 | 1,03E-03 |
| PPHLN1 | 2168,18 | -0,09 | 0,03 | -3,28 | 1,03E-03 |
| DCLRE1C | 1294,90 | -0,08 | 0,02 | -3,28 | 1,03E-03 |
| P2RY10 | 1080,21 | -0,20 | 0,06 | -3,28 | 1,03E-03 |
| ANKS3 | 256,64 | 0,18 | 0,06 | 3,28 | 1,03E-03 |
| TENM4 | 107,02 | 0,83 | 0,25 | 3,28 | 1,03E-03 |
| ELOF1 | 569,00 | 0,28 | 0,09 | 3,28 | 1,04E-03 |
| ARAP2 | 5810,14 | -0,20 | 0,06 | -3,28 | 1,04E-03 |
| OCM | 21,47 | 0,32 | 0,10 | 3,28 | 1,04E-03 |
| EDRF1 | 1903,89 | -0,09 | 0,03 | -3,28 | 1,04E-03 |
| CCDC59 | 653,23 | -0,27 | 0,08 | -3,28 | 1,04E-03 |
| EEA1 | 1212,12 | -0,18 | 0,05 | -3,28 | 1,04E-03 |
| ZNF426 | 790,72 | -0,15 | 0,05 | -3,28 | 1,05E-03 |
| WRAP53 | 130,55 | 0,13 | 0,04 | 3,28 | 1,05E-03 |
| TMEM25 | 70,19 | 0,28 | 0,09 | 3,28 | 1,05E-03 |
| ZMYND11 | 1401,68 | -0,14 | 0,04 | -3,28 | 1,05E-03 |
| ZNF135 | 84,23 | 0,37 | 0,11 | 3,28 | 1,05E-03 |
| LTN1 | 3313,72 | -0,16 | 0,05 | -3,28 | 1,05E-03 |
| ZNF525 | 517,98 | -0,30 | 0,09 | -3,28 | 1,05E-03 |
| JADE1 | 2651,66 | -0,10 | 0,03 | -3,28 | 1,05E-03 |
| PRXL2B | 156,61 | 0,19 | 0,06 | 3,28 | 1,05E-03 |
| TIMM17A | 588,62 | -0,15 | 0,05 | -3,28 | 1,05E-03 |
| CIAO3 | 290,53 | 0,21 | 0,06 | 3,28 | 1,05E-03 |
| PCDH9 | 1198,91 | -0,40 | 0,12 | -3,28 | 1,05E-03 |
| GNL3 | 794,59 | -0,22 | 0,07 | -3,28 | 1,06E-03 |
| P2RY2 | 278,37 | 0,32 | 0,10 | 3,27 | 1,06E-03 |
| SLC35E4 | 62,87 | 0,38 | 0,12 | 3,27 | 1,06E-03 |
| WDCP | 315,71 | -0,12 | 0,04 | -3,27 | 1,06E-03 |
| MIB2 | 699,45 | 0,15 | 0,04 | 3,27 | 1,06E-03 |
| BAZ1A | 6712,38 | -0,19 | 0,06 | -3,27 | 1,06E-03 |
| USP48 | 3585,27 | -0,10 | 0,03 | -3,27 | 1,06E-03 |
| GFM1 | 1369,05 | -0,12 | 0,04 | -3,27 | 1,07E-03 |
| DND1 | 50,69 | 0,25 | 0,08 | 3,27 | 1,07E-03 |
| CDC23 | 627,91 | -0,10 | 0,03 | -3,27 | 1,07E-03 |
| ACVRL1 | 46,88 | 0,53 | 0,16 | 3,27 | 1,07E-03 |
| PBX1 | 360,64 | 0,36 | 0,11 | 3,27 | 1,07E-03 |

| | | | | | |
|---|---|---|---|---|---|
| PCSK6 | 200,25 | 0,42 | 0,13 | 3,27 | 1,07E-03 |
| CDHR1 | 57,14 | 0,50 | 0,15 | 3,27 | 1,08E-03 |
| NPR2 | 88,46 | 0,28 | 0,09 | 3,27 | 1,08E-03 |
| OBSCN | 1524,96 | 0,31 | 0,10 | 3,27 | 1,08E-03 |
| C1orf198 | 254,02 | 0,31 | 0,09 | 3,27 | 1,08E-03 |
| RFPL2 | 39,03 | 0,47 | 0,14 | 3,27 | 1,08E-03 |
| GRB10 | 388,74 | 0,31 | 0,10 | 3,27 | 1,09E-03 |
| ZNF644 | 2750,62 | -0,23 | 0,07 | -3,27 | 1,09E-03 |
| THRB | 64,14 | 0,55 | 0,17 | 3,27 | 1,09E-03 |
| SIGLEC11 | 24,75 | 0,54 | 0,17 | 3,27 | 1,09E-03 |
| SFRP2 | 38,83 | 0,48 | 0,15 | 3,27 | 1,09E-03 |
| TMEM134 | 170,79 | 0,22 | 0,07 | 3,27 | 1,09E-03 |
| P3H4 | 78,84 | 0,28 | 0,09 | 3,26 | 1,10E-03 |
| MAP3K12 | 230,22 | 0,22 | 0,07 | 3,26 | 1,10E-03 |
| GRK4 | 73,03 | 0,29 | 0,09 | 3,26 | 1,10E-03 |
| NUP107 | 1166,54 | -0,15 | 0,05 | -3,26 | 1,11E-03 |
| DBN1 | 127,16 | 0,34 | 0,10 | 3,26 | 1,11E-03 |
| DLEC1 | 166,32 | 0,30 | 0,09 | 3,26 | 1,11E-03 |
| MOXD1 | 30,05 | 0,54 | 0,16 | 3,26 | 1,11E-03 |
| CD24 | 563,44 | 0,34 | 0,10 | 3,26 | 1,11E-03 |
| CDC40 | 2085,29 | -0,16 | 0,05 | -3,26 | 1,11E-03 |
| EIF4E | 1386,71 | -0,14 | 0,04 | -3,26 | 1,12E-03 |
| PCYT1B | 49,21 | 0,46 | 0,14 | 3,26 | 1,12E-03 |
| ZNF346 | 504,03 | 0,14 | 0,04 | 3,26 | 1,12E-03 |
| CHD1 | 6348,17 | -0,18 | 0,05 | -3,26 | 1,13E-03 |
| KLHL33 | 33,35 | 0,53 | 0,16 | 3,26 | 1,13E-03 |
| LHPP | 181,59 | 0,24 | 0,07 | 3,26 | 1,13E-03 |
| IFI27L1 | 74,47 | 0,20 | 0,06 | 3,26 | 1,13E-03 |
| SNAPC4 | 272,05 | 0,27 | 0,08 | 3,26 | 1,13E-03 |
| ANK2 | 52,19 | 0,84 | 0,26 | 3,26 | 1,13E-03 |
| AHCTF1 | 7856,45 | -0,16 | 0,05 | -3,26 | 1,13E-03 |
| TBL3 | 323,43 | 0,15 | 0,05 | 3,26 | 1,13E-03 |
| FBXO44 | 229,92 | 0,19 | 0,06 | 3,26 | 1,13E-03 |
| COA5 | 1134,26 | -0,14 | 0,04 | -3,26 | 1,13E-03 |
| BRAP | 1406,33 | -0,09 | 0,03 | -3,26 | 1,13E-03 |
| TJAP1 | 273,24 | 0,22 | 0,07 | 3,26 | 1,13E-03 |
| F12 | 35,07 | 0,37 | 0,11 | 3,25 | 1,14E-03 |
| CNPPD1 | 843,85 | 0,26 | 0,08 | 3,25 | 1,14E-03 |
| CCT2 | 1215,29 | -0,23 | 0,07 | -3,25 | 1,14E-03 |
| UQCRB | 889,42 | -0,42 | 0,13 | -3,25 | 1,14E-03 |
| FAM167A | 46,35 | 0,54 | 0,17 | 3,25 | 1,15E-03 |
| GTF2F2 | 618,01 | -0,14 | 0,04 | -3,25 | 1,15E-03 |
| HMGA1 | 186,77 | 0,29 | 0,09 | 3,25 | 1,15E-03 |
| ZSWIM1 | 136,49 | 0,18 | 0,05 | 3,25 | 1,15E-03 |
| TMEM255B | 143,69 | 0,29 | 0,09 | 3,25 | 1,15E-03 |
| PPIL3 | 335,70 | -0,19 | 0,06 | -3,25 | 1,15E-03 |

| | | | | | |
|---|---|---|---|---|---|
| ZMYND15 | 34,52 | 0,50 | 0,15 | 3,25 | 1,15E-03 |
| ODR4 | 835,28 | -0,18 | 0,06 | -3,25 | 1,15E-03 |
| SCARF1 | 1058,96 | 0,32 | 0,10 | 3,25 | 1,15E-03 |
| C3orf20 | 61,51 | 0,38 | 0,12 | 3,25 | 1,16E-03 |
| HK1 | 2206,33 | 0,24 | 0,07 | 3,25 | 1,16E-03 |
| VPS4B | 4715,91 | -0,13 | 0,04 | -3,25 | 1,16E-03 |
| MCM9 | 1899,78 | -0,10 | 0,03 | -3,25 | 1,17E-03 |
| TUT4 | 3150,67 | -0,12 | 0,04 | -3,25 | 1,17E-03 |
| LAMB2 | 53,67 | 0,49 | 0,15 | 3,25 | 1,17E-03 |
| GABBR1 | 645,21 | 0,32 | 0,10 | 3,25 | 1,17E-03 |
| ZNF559 | 636,20 | -0,15 | 0,05 | -3,24 | 1,17E-03 |
| ACTMAP | 40,89 | 0,35 | 0,11 | 3,24 | 1,18E-03 |
| CEP162 | 617,34 | -0,19 | 0,06 | -3,24 | 1,18E-03 |
| C3orf33 | 31,84 | 0,26 | 0,08 | 3,24 | 1,18E-03 |
| ARVCF | 133,12 | 0,36 | 0,11 | 3,24 | 1,18E-03 |
| ATF3 | 44,39 | 0,47 | 0,15 | 3,24 | 1,19E-03 |
| FAM83G | 121,23 | 0,31 | 0,10 | 3,24 | 1,18E-03 |
| ATPSCKMT | 433,91 | -0,14 | 0,04 | -3,24 | 1,19E-03 |
| ATL2 | 1190,07 | -0,16 | 0,05 | -3,24 | 1,19E-03 |
| SERINC1 | 8452,08 | -0,23 | 0,07 | -3,24 | 1,19E-03 |
| BLOC1S2 | 636,41 | -0,24 | 0,08 | -3,24 | 1,20E-03 |
| SERPINE2 | 55,36 | 0,49 | 0,15 | 3,24 | 1,20E-03 |
| RTRAF | 1055,78 | -0,17 | 0,05 | -3,24 | 1,20E-03 |
| FGFR1 | 148,50 | 0,42 | 0,13 | 3,24 | 1,20E-03 |
| RGS6 | 190,45 | 0,40 | 0,12 | 3,24 | 1,20E-03 |
| SAMD1 | 75,68 | 0,32 | 0,10 | 3,24 | 1,20E-03 |
| DDX24 | 2970,55 | -0,10 | 0,03 | -3,24 | 1,20E-03 |
| EIF3E | 3821,48 | -0,29 | 0,09 | -3,24 | 1,21E-03 |
| IFITM10 | 82,28 | 0,30 | 0,09 | 3,24 | 1,21E-03 |
| KCNAB3 | 54,99 | 0,39 | 0,12 | 3,24 | 1,21E-03 |
| COL24A1 | 88,49 | 0,44 | 0,13 | 3,24 | 1,21E-03 |
| IFT25 | 543,45 | -0,13 | 0,04 | -3,24 | 1,21E-03 |
| GRHL1 | 108,47 | 0,30 | 0,09 | 3,23 | 1,22E-03 |
| PXMP2 | 25,81 | 0,25 | 0,08 | 3,23 | 1,22E-03 |
| PPP4R3A | 5077,37 | -0,10 | 0,03 | -3,23 | 1,22E-03 |
| DOHH | 187,95 | 0,19 | 0,06 | 3,23 | 1,22E-03 |
| NOC3L | 720,56 | -0,20 | 0,06 | -3,23 | 1,22E-03 |
| CBFA2T3 | 338,37 | 0,33 | 0,10 | 3,23 | 1,23E-03 |
| SPTY2D1 | 1112,68 | -0,09 | 0,03 | -3,23 | 1,23E-03 |
| MGAT4A | 6509,21 | -0,16 | 0,05 | -3,23 | 1,23E-03 |
| CRLF3 | 5296,43 | -0,13 | 0,04 | -3,23 | 1,23E-03 |
| NPDC1 | 97,15 | 0,39 | 0,12 | 3,23 | 1,23E-03 |
| SLX4 | 163,15 | 0,38 | 0,12 | 3,23 | 1,23E-03 |
| L3MBTL3 | 1157,66 | -0,14 | 0,04 | -3,23 | 1,23E-03 |
| USP1 | 1542,99 | -0,25 | 0,08 | -3,23 | 1,23E-03 |
| DNAJC10 | 2647,10 | -0,18 | 0,06 | -3,23 | 1,24E-03 |

| | | | | | |
|---|---|---|---|---|---|
| MSANTD4 | 766,15 | -0,19 | 0,06 | -3,23 | 1,24E-03 |
| MLLT1 | 325,44 | 0,35 | 0,11 | 3,23 | 1,24E-03 |
| NFKB2 | 1283,96 | 0,21 | 0,06 | 3,23 | 1,24E-03 |
| SPSB3 | 1471,35 | 0,13 | 0,04 | 3,23 | 1,25E-03 |
| YWHAQ | 2060,72 | -0,13 | 0,04 | -3,23 | 1,25E-03 |
| LILRB4 | 121,76 | 0,33 | 0,10 | 3,22 | 1,26E-03 |
| PCBP4 | 122,66 | 0,21 | 0,07 | 3,22 | 1,26E-03 |
| PIAS3 | 158,80 | 0,28 | 0,09 | 3,22 | 1,26E-03 |
| XRCC3 | 163,03 | 0,27 | 0,08 | 3,22 | 1,27E-03 |
| SLC26A11 | 285,51 | 0,23 | 0,07 | 3,22 | 1,27E-03 |
| SHLD2 | 738,61 | -0,20 | 0,06 | -3,22 | 1,27E-03 |
| UQCRC2 | 2630,59 | -0,12 | 0,04 | -3,22 | 1,27E-03 |
| ZNF28 | 556,80 | -0,19 | 0,06 | -3,22 | 1,28E-03 |
| PLA2G15 | 92,05 | 0,30 | 0,09 | 3,22 | 1,28E-03 |
| KIF9 | 68,47 | 0,29 | 0,09 | 3,22 | 1,28E-03 |
| SPHK2 | 184,83 | 0,23 | 0,07 | 3,22 | 1,28E-03 |
| LMBRD1 | 3096,96 | -0,18 | 0,06 | -3,22 | 1,28E-03 |
| TTI1 | 1017,30 | -0,08 | 0,02 | -3,22 | 1,29E-03 |
| BDH2 | 160,57 | -0,23 | 0,07 | -3,22 | 1,29E-03 |
| NEDD1 | 1006,05 | -0,17 | 0,05 | -3,22 | 1,29E-03 |
| G3BP2 | 4223,20 | -0,13 | 0,04 | -3,22 | 1,29E-03 |
| KCNC4 | 250,68 | 0,37 | 0,12 | 3,22 | 1,29E-03 |
| LLGL1 | 298,55 | 0,21 | 0,06 | 3,22 | 1,29E-03 |
| RAD9A | 177,13 | 0,25 | 0,08 | 3,22 | 1,29E-03 |
| PPIG | 2238,80 | -0,27 | 0,08 | -3,22 | 1,29E-03 |
| NCOR2 | 1121,03 | 0,30 | 0,09 | 3,22 | 1,29E-03 |
| ZNF668 | 113,92 | 0,18 | 0,06 | 3,22 | 1,29E-03 |
| PAQR4 | 74,18 | 0,25 | 0,08 | 3,22 | 1,29E-03 |
| FAM13B | 2809,05 | -0,11 | 0,03 | -3,22 | 1,30E-03 |
| STON1 | 87,76 | 0,25 | 0,08 | 3,22 | 1,30E-03 |
| TKFC | 187,82 | 0,21 | 0,07 | 3,22 | 1,30E-03 |
| CLPTM1 | 371,81 | 0,24 | 0,07 | 3,22 | 1,30E-03 |
| KCTD18 | 839,32 | -0,08 | 0,02 | -3,21 | 1,30E-03 |
| DHFR2 | 420,91 | -0,17 | 0,05 | -3,21 | 1,31E-03 |
| SCYL3 | 1530,99 | -0,11 | 0,04 | -3,21 | 1,31E-03 |
| SMIM38 | 41,23 | 0,48 | 0,15 | 3,21 | 1,32E-03 |
| DDX21 | 2779,42 | -0,14 | 0,04 | -3,21 | 1,32E-03 |
| PPID | 382,92 | -0,11 | 0,03 | -3,21 | 1,32E-03 |
| DEPDC1B | 43,54 | 0,38 | 0,12 | 3,21 | 1,32E-03 |
| AGPAT1 | 187,35 | 0,28 | 0,09 | 3,21 | 1,32E-03 |
| ANKRD12 | 6307,94 | -0,21 | 0,07 | -3,21 | 1,32E-03 |
| NDUFS4 | 236,43 | -0,21 | 0,07 | -3,21 | 1,32E-03 |
| ENGASE | 1147,91 | 0,17 | 0,05 | 3,21 | 1,32E-03 |
| SNRPA | 194,03 | 0,20 | 0,06 | 3,21 | 1,32E-03 |
| CALHM5 | 39,70 | 0,43 | 0,14 | 3,21 | 1,33E-03 |
| RIOK2 | 548,51 | -0,22 | 0,07 | -3,21 | 1,33E-03 |

| | | | | | |
|---|---|---|---|---|---|
| RORC | 104,37 | 0,36 | 0,11 | 3,21 | 1,33E-03 |
| PTPRB | 60,68 | 0,59 | 0,19 | 3,21 | 1,33E-03 |
| TCF7L2 | 564,33 | 0,23 | 0,07 | 3,21 | 1,33E-03 |
| ACACB | 571,79 | 0,24 | 0,08 | 3,21 | 1,34E-03 |
| RIOK1 | 519,89 | -0,12 | 0,04 | -3,21 | 1,34E-03 |
| IQGAP2 | 7364,28 | -0,15 | 0,05 | -3,21 | 1,34E-03 |
| GSTK1 | 2610,86 | 0,12 | 0,04 | 3,21 | 1,34E-03 |
| UBE2S | 347,61 | 0,17 | 0,05 | 3,21 | 1,35E-03 |
| GPR153 | 52,01 | 0,50 | 0,16 | 3,21 | 1,34E-03 |
| TMEM19 | 845,44 | -0,14 | 0,04 | -3,21 | 1,34E-03 |
| SIRT6 | 189,55 | 0,15 | 0,05 | 3,21 | 1,35E-03 |
| SF3A2 | 336,77 | 0,20 | 0,06 | 3,21 | 1,35E-03 |
| GP6 | 43,74 | 0,41 | 0,13 | 3,21 | 1,35E-03 |
| ADD2 | 282,68 | 0,29 | 0,09 | 3,20 | 1,36E-03 |
| TMEM63B | 877,81 | 0,36 | 0,11 | 3,20 | 1,36E-03 |
| ALDOA | 253,85 | 0,14 | 0,05 | 3,20 | 1,36E-03 |
| ZNF701 | 1057,43 | -0,15 | 0,05 | -3,20 | 1,36E-03 |
| PRICKLE2 | 63,13 | 0,64 | 0,20 | 3,20 | 1,37E-03 |
| FKBP9 | 135,76 | 0,30 | 0,09 | 3,20 | 1,37E-03 |
| DDX59 | 839,64 | -0,10 | 0,03 | -3,20 | 1,37E-03 |
| BRF2 | 409,00 | -0,08 | 0,03 | -3,20 | 1,37E-03 |
| IGBP1 | 897,45 | -0,19 | 0,06 | -3,20 | 1,37E-03 |
| DTNA | 66,14 | 0,53 | 0,17 | 3,20 | 1,37E-03 |
| SLC41A1 | 187,77 | 0,25 | 0,08 | 3,20 | 1,38E-03 |
| FAM110C | 55,14 | 0,40 | 0,13 | 3,20 | 1,38E-03 |
| ALKBH3 | 526,09 | -0,11 | 0,04 | -3,20 | 1,38E-03 |
| CABLES2 | 67,39 | 0,34 | 0,11 | 3,20 | 1,38E-03 |
| AARSD1 | 68,16 | 0,21 | 0,06 | 3,20 | 1,38E-03 |
| KAT14 | 513,53 | -0,11 | 0,03 | -3,20 | 1,38E-03 |
| HDAC7 | 1770,74 | 0,26 | 0,08 | 3,20 | 1,38E-03 |
| FAM220A | 314,38 | -0,11 | 0,03 | -3,20 | 1,38E-03 |
| WBP4 | 441,79 | -0,17 | 0,05 | -3,20 | 1,39E-03 |
| NOC2L | 392,61 | 0,22 | 0,07 | 3,20 | 1,39E-03 |
| R3HDM4 | 9550,54 | 0,32 | 0,10 | 3,20 | 1,39E-03 |
| ZFR | 3217,28 | -0,11 | 0,03 | -3,20 | 1,39E-03 |
| CD86 | 1030,54 | -0,17 | 0,05 | -3,20 | 1,39E-03 |
| FRG1 | 533,41 | -0,19 | 0,06 | -3,20 | 1,39E-03 |
| TCTN2 | 91,63 | 0,26 | 0,08 | 3,19 | 1,40E-03 |
| SH3BP4 | 68,60 | 0,48 | 0,15 | 3,19 | 1,40E-03 |
| ZNF836 | 440,78 | -0,19 | 0,06 | -3,19 | 1,40E-03 |
| OLR1 | 33,68 | 0,55 | 0,17 | 3,19 | 1,41E-03 |
| NID1 | 141,15 | 0,36 | 0,11 | 3,19 | 1,41E-03 |
| ATOSA | 3909,23 | -0,14 | 0,04 | -3,19 | 1,41E-03 |
| RBBP9 | 313,94 | -0,11 | 0,03 | -3,19 | 1,41E-03 |
| REV3L | 2658,47 | -0,11 | 0,03 | -3,19 | 1,41E-03 |
| IZUMO4 | 46,16 | 0,25 | 0,08 | 3,19 | 1,42E-03 |

| | | | | | |
|---|---|---|---|---|---|
| KIF20B | 445,80 | -0,29 | 0,09 | -3,19 | 1,41E-03 |
| TOPORS | 2775,70 | -0,23 | 0,07 | -3,19 | 1,42E-03 |
| BCS1L | 233,40 | 0,13 | 0,04 | 3,19 | 1,42E-03 |
| ERGIC2 | 1719,41 | -0,23 | 0,07 | -3,19 | 1,42E-03 |
| EXOC3L1 | 61,49 | 0,51 | 0,16 | 3,19 | 1,42E-03 |
| CBLL1 | 1123,78 | -0,16 | 0,05 | -3,19 | 1,43E-03 |
| ANLN | 68,33 | 0,37 | 0,12 | 3,19 | 1,43E-03 |
| WASF1 | 45,40 | 0,30 | 0,09 | 3,19 | 1,43E-03 |
| FOXD4L1 | 25,38 | 0,30 | 0,09 | 3,19 | 1,43E-03 |
| CCDC81 | 38,65 | 0,44 | 0,14 | 3,19 | 1,43E-03 |
| ZBTB44 | 4176,19 | -0,13 | 0,04 | -3,19 | 1,44E-03 |
| H2AJ | 865,75 | 0,26 | 0,08 | 3,19 | 1,44E-03 |
| API5 | 2673,90 | -0,14 | 0,04 | -3,19 | 1,44E-03 |
| CTSA | 789,46 | 0,26 | 0,08 | 3,19 | 1,44E-03 |
| MRPS18C | 705,40 | -0,20 | 0,06 | -3,19 | 1,44E-03 |
| DCHS1 | 123,30 | 0,44 | 0,14 | 3,19 | 1,44E-03 |
| MARVELD1 | 48,95 | 0,45 | 0,14 | 3,19 | 1,44E-03 |
| MSANTD7 | 914,64 | -0,09 | 0,03 | -3,19 | 1,44E-03 |
| EEFSEC | 185,34 | 0,17 | 0,05 | 3,19 | 1,45E-03 |
| C1orf131 | 512,81 | -0,09 | 0,03 | -3,19 | 1,45E-03 |
| ZCCHC17 | 625,22 | -0,12 | 0,04 | -3,18 | 1,45E-03 |
| EIF2S1 | 1390,97 | -0,18 | 0,06 | -3,18 | 1,45E-03 |
| TSPAN15 | 67,05 | 0,37 | 0,12 | 3,18 | 1,45E-03 |
| CNBD2 | 58,09 | 0,35 | 0,11 | 3,18 | 1,45E-03 |
| BSCL2 | 105,92 | 0,13 | 0,04 | 3,18 | 1,46E-03 |
| RNF123 | 1673,61 | 0,22 | 0,07 | 3,18 | 1,46E-03 |
| SLC14A1 | 1351,35 | 0,27 | 0,09 | 3,18 | 1,46E-03 |
| KRCC1 | 710,62 | -0,25 | 0,08 | -3,18 | 1,46E-03 |
| CERS3 | 61,26 | 0,41 | 0,13 | 3,18 | 1,46E-03 |
| ABCE1 | 1158,80 | -0,21 | 0,07 | -3,18 | 1,46E-03 |
| PDGFA | 134,93 | 0,35 | 0,11 | 3,18 | 1,47E-03 |
| NSA2 | 1000,76 | -0,27 | 0,08 | -3,18 | 1,47E-03 |
| NEK8 | 80,90 | 0,29 | 0,09 | 3,18 | 1,47E-03 |
| PPM1N | 94,69 | 0,21 | 0,07 | 3,18 | 1,47E-03 |
| SMYD5 | 166,19 | 0,23 | 0,07 | 3,18 | 1,47E-03 |
| MYBL1 | 2371,44 | -0,26 | 0,08 | -3,18 | 1,47E-03 |
| RABL2B | 234,19 | 0,24 | 0,08 | 3,18 | 1,47E-03 |
| ATP5PB | 2146,42 | -0,19 | 0,06 | -3,18 | 1,47E-03 |
| FGF13 | 72,68 | 0,63 | 0,20 | 3,18 | 1,48E-03 |
| SYTL1 | 1838,48 | 0,17 | 0,05 | 3,18 | 1,48E-03 |
| ZNF92 | 372,38 | -0,30 | 0,10 | -3,18 | 1,48E-03 |
| ACAP3 | 798,99 | 0,24 | 0,08 | 3,18 | 1,48E-03 |
| SMARCAD1 | 1189,31 | -0,19 | 0,06 | -3,18 | 1,49E-03 |
| ASTE1 | 685,95 | -0,13 | 0,04 | -3,18 | 1,49E-03 |
| APPBP2 | 2136,09 | -0,11 | 0,03 | -3,18 | 1,50E-03 |
| PCYT2 | 305,18 | 0,14 | 0,04 | 3,18 | 1,50E-03 |

| | | | | | |
|---|---|---|---|---|---|
| TOR2A | 116,45 | 0,26 | 0,08 | 3,18 | 1,50E-03 |
| ASH1L | 6577,89 | -0,08 | 0,03 | -3,18 | 1,50E-03 |
| KIF19 | 68,00 | 0,57 | 0,18 | 3,17 | 1,50E-03 |
| UPF3B | 411,42 | -0,11 | 0,03 | -3,17 | 1,50E-03 |
| CETN3 | 172,78 | -0,31 | 0,10 | -3,17 | 1,50E-03 |
| KIAA0930 | 963,96 | 0,27 | 0,09 | 3,17 | 1,51E-03 |
| EPHB6 | 195,65 | 0,30 | 0,09 | 3,17 | 1,51E-03 |
| TTK | 46,21 | 0,45 | 0,14 | 3,17 | 1,51E-03 |
| TASL | 929,65 | -0,21 | 0,07 | -3,17 | 1,51E-03 |
| FHIP2B | 283,58 | 0,30 | 0,09 | 3,17 | 1,51E-03 |
| TXNDC12 | 445,25 | -0,13 | 0,04 | -3,17 | 1,52E-03 |
| STARD7 | 3103,56 | -0,10 | 0,03 | -3,17 | 1,52E-03 |
| TBC1D23 | 2307,07 | -0,16 | 0,05 | -3,17 | 1,52E-03 |
| PFAS | 160,06 | 0,29 | 0,09 | 3,17 | 1,53E-03 |
| STAT4 | 2680,63 | -0,16 | 0,05 | -3,17 | 1,53E-03 |
| CYB5B | 1104,87 | -0,11 | 0,04 | -3,17 | 1,53E-03 |
| STPG4 | 34,93 | 0,30 | 0,09 | 3,17 | 1,53E-03 |
| SAMD3 | 1753,89 | -0,22 | 0,07 | -3,17 | 1,54E-03 |
| PYCR3 | 69,19 | 0,24 | 0,07 | 3,17 | 1,54E-03 |
| CTSG | 35,76 | 0,55 | 0,17 | 3,17 | 1,54E-03 |
| SGSM3 | 88,12 | 0,19 | 0,06 | 3,17 | 1,55E-03 |
| CSNK2A1 | 3464,14 | -0,07 | 0,02 | -3,17 | 1,55E-03 |
| MEST | 138,39 | 0,27 | 0,09 | 3,17 | 1,55E-03 |
| ABCF2 | 72,42 | 0,18 | 0,06 | 3,17 | 1,55E-03 |
| SEM1 | 817,25 | -0,16 | 0,05 | -3,17 | 1,55E-03 |
| HAPLN3 | 579,72 | 0,20 | 0,06 | 3,16 | 1,55E-03 |
| TAF2 | 1857,32 | -0,11 | 0,04 | -3,16 | 1,56E-03 |
| TLCD4 | 524,00 | 0,34 | 0,11 | 3,16 | 1,56E-03 |
| MYO5C | 91,86 | 0,46 | 0,14 | 3,16 | 1,57E-03 |
| SLC25A35 | 68,97 | 0,29 | 0,09 | 3,16 | 1,57E-03 |
| POLN | 77,27 | 0,32 | 0,10 | 3,16 | 1,57E-03 |
| SLC66A1 | 126,10 | 0,20 | 0,06 | 3,16 | 1,57E-03 |
| ZNF205 | 27,83 | 0,46 | 0,14 | 3,16 | 1,57E-03 |
| SLC4A8 | 534,52 | 0,21 | 0,07 | 3,16 | 1,58E-03 |
| FHAD1 | 62,59 | 0,74 | 0,23 | 3,16 | 1,58E-03 |
| ZNF226 | 1297,76 | -0,19 | 0,06 | -3,16 | 1,58E-03 |
| AP3B2 | 52,99 | 0,78 | 0,25 | 3,16 | 1,58E-03 |
| TRAPPC11 | 2152,19 | -0,09 | 0,03 | -3,16 | 1,58E-03 |
| TGFB1 | 853,78 | 0,38 | 0,12 | 3,16 | 1,59E-03 |
| HELQ | 857,23 | -0,13 | 0,04 | -3,16 | 1,59E-03 |
| OTULIN | 4047,08 | -0,10 | 0,03 | -3,16 | 1,59E-03 |
| ARHGEF37 | 128,52 | 0,40 | 0,13 | 3,16 | 1,59E-03 |
| WDR89 | 461,29 | -0,30 | 0,10 | -3,16 | 1,59E-03 |
| UNC45B | 52,66 | 0,41 | 0,13 | 3,16 | 1,59E-03 |
| MAP1S | 259,42 | 0,23 | 0,07 | 3,16 | 1,60E-03 |
| EIF4E2 | 2520,68 | -0,09 | 0,03 | -3,16 | 1,60E-03 |

| | | | | | |
|---|---|---|---|---|---|
| TJP2 | 183,69 | 0,28 | 0,09 | 3,16 | 1,60E-03 |
| QDPR | 120,01 | 0,21 | 0,07 | 3,16 | 1,60E-03 |
| ENSG00000285589 | 65,07 | 0,36 | 0,11 | 3,15 | 1,62E-03 |
| DMTN | 5055,16 | 0,42 | 0,13 | 3,15 | 1,62E-03 |
| ISM1 | 53,35 | 0,38 | 0,12 | 3,15 | 1,62E-03 |
| USP37 | 1247,80 | -0,10 | 0,03 | -3,15 | 1,62E-03 |
| INSC | 36,85 | 0,54 | 0,17 | 3,15 | 1,62E-03 |
| AZU1 | 34,23 | 0,43 | 0,14 | 3,15 | 1,62E-03 |
| C2orf49 | 959,40 | -0,13 | 0,04 | -3,15 | 1,62E-03 |
| ANKRD9 | 431,35 | 0,37 | 0,12 | 3,15 | 1,63E-03 |
| CLN3 | 374,20 | 0,20 | 0,06 | 3,15 | 1,63E-03 |
| DHCR24 | 121,72 | 0,34 | 0,11 | 3,15 | 1,63E-03 |
| SCNM1 | 241,81 | 0,16 | 0,05 | 3,15 | 1,64E-03 |
| MPHOSPH6 | 437,34 | -0,16 | 0,05 | -3,15 | 1,65E-03 |
| SART3 | 2286,65 | -0,08 | 0,03 | -3,15 | 1,65E-03 |
| KEL | 166,86 | 0,39 | 0,13 | 3,15 | 1,66E-03 |
| EEIG2 | 1075,90 | -0,12 | 0,04 | -3,15 | 1,66E-03 |
| ZKSCAN4 | 406,21 | -0,10 | 0,03 | -3,15 | 1,66E-03 |
| ZNF444 | 227,33 | 0,27 | 0,09 | 3,15 | 1,66E-03 |
| RNF187 | 487,14 | 0,30 | 0,09 | 3,14 | 1,67E-03 |
| ZNF782 | 830,19 | -0,11 | 0,04 | -3,14 | 1,67E-03 |
| ZNF184 | 400,22 | -0,21 | 0,07 | -3,14 | 1,67E-03 |
| PRR5 | 511,84 | 0,23 | 0,07 | 3,14 | 1,66E-03 |
| MTERF2 | 377,51 | -0,14 | 0,04 | -3,14 | 1,67E-03 |
| SPATA33 | 131,98 | 0,14 | 0,05 | 3,14 | 1,67E-03 |
| SLC22A18AS | 28,55 | 0,44 | 0,14 | 3,14 | 1,67E-03 |
| SLC35D2 | 212,23 | 0,09 | 0,03 | 3,14 | 1,67E-03 |
| BLOC1S5 | 479,93 | -0,13 | 0,04 | -3,14 | 1,67E-03 |
| RNGTT | 1456,53 | -0,12 | 0,04 | -3,14 | 1,67E-03 |
| LAT | 64,98 | 0,37 | 0,12 | 3,14 | 1,67E-03 |
| LIG1 | 330,46 | 0,23 | 0,07 | 3,14 | 1,68E-03 |
| PLVAP | 156,73 | 0,43 | 0,14 | 3,14 | 1,68E-03 |
| EPN1 | 830,01 | 0,29 | 0,09 | 3,14 | 1,69E-03 |
| OLIG2 | 115,93 | 0,44 | 0,14 | 3,14 | 1,69E-03 |
| WNT10A | 50,52 | 0,38 | 0,12 | 3,14 | 1,69E-03 |
| CLPX | 1850,84 | -0,10 | 0,03 | -3,14 | 1,70E-03 |
| ZNF484 | 922,89 | -0,23 | 0,07 | -3,14 | 1,70E-03 |
| NR3C1 | 8506,47 | -0,13 | 0,04 | -3,14 | 1,70E-03 |
| SSR1 | 4076,18 | -0,10 | 0,03 | -3,14 | 1,70E-03 |
| UTP6 | 1827,27 | -0,10 | 0,03 | -3,14 | 1,71E-03 |
| GTF3C3 | 1167,70 | -0,14 | 0,05 | -3,14 | 1,72E-03 |
| DHX15 | 5128,14 | -0,16 | 0,05 | -3,13 | 1,72E-03 |
| MANEAL | 39,93 | 0,37 | 0,12 | 3,13 | 1,72E-03 |
| SCD | 87,61 | 0,26 | 0,08 | 3,13 | 1,72E-03 |
| ZNF627 | 350,74 | -0,16 | 0,05 | -3,13 | 1,73E-03 |
| TMEM191C | 25,94 | 0,33 | 0,10 | 3,13 | 1,73E-03 |

| | | | | | |
|---|---|---|---|---|---|
| MIB1 | 3607,99 | -0,15 | 0,05 | -3,13 | 1,73E-03 |
| PSMA4 | 1273,56 | -0,26 | 0,08 | -3,13 | 1,73E-03 |
| DYNLRB1 | 395,56 | 0,12 | 0,04 | 3,13 | 1,73E-03 |
| MKLN1 | 8240,30 | -0,14 | 0,04 | -3,13 | 1,74E-03 |
| SLC4A1AP | 913,93 | -0,09 | 0,03 | -3,13 | 1,73E-03 |
| ELANE | 32,84 | 0,45 | 0,14 | 3,13 | 1,74E-03 |
| PLXNA3 | 332,36 | 0,33 | 0,10 | 3,13 | 1,74E-03 |
| STOML1 | 208,75 | 0,21 | 0,07 | 3,13 | 1,74E-03 |
| PPP2R5C | 8939,32 | -0,10 | 0,03 | -3,13 | 1,75E-03 |
| NDUFAF1 | 407,33 | -0,13 | 0,04 | -3,13 | 1,75E-03 |
| ARL1 | 749,96 | -0,15 | 0,05 | -3,13 | 1,75E-03 |
| ZNF319 | 338,04 | 0,24 | 0,08 | 3,13 | 1,75E-03 |
| L3MBTL1 | 434,62 | 0,19 | 0,06 | 3,13 | 1,76E-03 |
| MORF4L1 | 4271,80 | -0,11 | 0,04 | -3,13 | 1,76E-03 |
| WDR43 | 1330,02 | -0,15 | 0,05 | -3,13 | 1,77E-03 |
| WDR4 | 272,80 | 0,19 | 0,06 | 3,13 | 1,77E-03 |
| ZC3H8 | 533,92 | -0,12 | 0,04 | -3,13 | 1,78E-03 |
| ZNF330 | 874,49 | -0,13 | 0,04 | -3,13 | 1,78E-03 |
| NUP42 | 920,99 | -0,11 | 0,03 | -3,13 | 1,78E-03 |
| RESF1 | 33304,37 | -0,21 | 0,07 | -3,12 | 1,78E-03 |
| VBP1 | 650,85 | -0,29 | 0,09 | -3,12 | 1,79E-03 |
| ARF1 | 3950,40 | 0,16 | 0,05 | 3,12 | 1,79E-03 |
| CHKA | 193,11 | 0,18 | 0,06 | 3,12 | 1,79E-03 |
| ZFP91 | 1465,31 | -0,09 | 0,03 | -3,12 | 1,79E-03 |
| MRPS22 | 568,53 | -0,14 | 0,05 | -3,12 | 1,80E-03 |
| DARS2 | 421,37 | -0,08 | 0,02 | -3,12 | 1,80E-03 |
| ABCA9 | 57,83 | 0,59 | 0,19 | 3,12 | 1,80E-03 |
| RRAS2 | 763,37 | -0,21 | 0,07 | -3,12 | 1,80E-03 |
| KLHL12 | 1365,16 | -0,10 | 0,03 | -3,12 | 1,81E-03 |
| ARMC1 | 683,50 | -0,14 | 0,05 | -3,12 | 1,81E-03 |
| MAFG | 292,88 | 0,26 | 0,08 | 3,12 | 1,82E-03 |
| CDAN1 | 328,03 | 0,22 | 0,07 | 3,12 | 1,82E-03 |
| STK33 | 68,14 | 0,52 | 0,17 | 3,12 | 1,82E-03 |
| REEP4 | 113,80 | 0,31 | 0,10 | 3,12 | 1,82E-03 |
| TOP1MT | 226,76 | 0,21 | 0,07 | 3,12 | 1,82E-03 |
| SUCLG2 | 1519,35 | -0,13 | 0,04 | -3,12 | 1,83E-03 |
| SH3TC2 | 134,08 | 0,43 | 0,14 | 3,12 | 1,83E-03 |
| THSD1 | 47,65 | 0,30 | 0,10 | 3,12 | 1,83E-03 |
| NRBF2 | 1407,39 | -0,21 | 0,07 | -3,12 | 1,83E-03 |
| LLPH | 882,01 | -0,11 | 0,03 | -3,12 | 1,84E-03 |
| SYNJ2BP | 1025,93 | -0,14 | 0,04 | -3,12 | 1,84E-03 |
| RPTOR | 858,06 | 0,23 | 0,07 | 3,11 | 1,84E-03 |
| VLDLR | 91,65 | 0,45 | 0,14 | 3,11 | 1,84E-03 |
| MANF | 447,09 | -0,16 | 0,05 | -3,11 | 1,84E-03 |
| GSTM2 | 165,63 | 0,31 | 0,10 | 3,11 | 1,84E-03 |
| CASP8AP2 | 1412,49 | -0,23 | 0,07 | -3,11 | 1,85E-03 |

| | | | | | |
|---|---|---|---|---|---|
| CFAP97 | 1072,30 | -0,18 | 0,06 | -3,11 | 1,85E-03 |
| PTCD1 | 104,16 | 0,16 | 0,05 | 3,11 | 1,86E-03 |
| CCDC163 | 36,76 | 0,44 | 0,14 | 3,11 | 1,87E-03 |
| POU5F1B | 46,65 | 0,32 | 0,10 | 3,11 | 1,87E-03 |
| THUMPD3 | 536,00 | -0,11 | 0,04 | -3,11 | 1,87E-03 |
| APOM | 31,86 | 0,19 | 0,06 | 3,11 | 1,87E-03 |
| TMEM53 | 48,00 | 0,24 | 0,08 | 3,11 | 1,87E-03 |
| PLXNB2 | 888,22 | 0,38 | 0,12 | 3,11 | 1,87E-03 |
| FAM222B | 114,25 | 0,23 | 0,07 | 3,11 | 1,88E-03 |
| FAAH | 189,10 | 0,26 | 0,08 | 3,11 | 1,88E-03 |
| NDOR1 | 244,47 | 0,24 | 0,08 | 3,11 | 1,88E-03 |
| TAF1A | 306,99 | -0,15 | 0,05 | -3,11 | 1,88E-03 |
| DDX1 | 1053,04 | -0,14 | 0,05 | -3,11 | 1,89E-03 |
| RPS6KA2 | 330,88 | 0,27 | 0,09 | 3,11 | 1,89E-03 |
| PSMC2 | 797,16 | -0,19 | 0,06 | -3,11 | 1,89E-03 |
| FAXDC2 | 1404,19 | 0,38 | 0,12 | 3,11 | 1,89E-03 |
| STK26 | 2203,31 | -0,15 | 0,05 | -3,11 | 1,89E-03 |
| SPDYA | 173,07 | -0,21 | 0,07 | -3,11 | 1,89E-03 |
| BCAR3 | 80,11 | 0,43 | 0,14 | 3,11 | 1,90E-03 |
| FAM200A | 300,41 | -0,17 | 0,05 | -3,10 | 1,90E-03 |
| SNRPB2 | 641,62 | -0,19 | 0,06 | -3,10 | 1,90E-03 |
| ROCK2 | 2032,52 | -0,13 | 0,04 | -3,10 | 1,91E-03 |
| APC | 4124,27 | -0,15 | 0,05 | -3,10 | 1,91E-03 |
| ALG11 | 981,42 | -0,10 | 0,03 | -3,10 | 1,91E-03 |
| ATG4D | 323,68 | 0,14 | 0,05 | 3,10 | 1,91E-03 |
| HNRNPR | 4655,46 | -0,15 | 0,05 | -3,10 | 1,92E-03 |
| LOXHD1 | 408,89 | 0,43 | 0,14 | 3,10 | 1,92E-03 |
| SELENOW | 395,17 | 0,13 | 0,04 | 3,10 | 1,92E-03 |
| LMLN | 162,49 | 0,18 | 0,06 | 3,10 | 1,93E-03 |
| PITPNM2 | 280,04 | 0,31 | 0,10 | 3,10 | 1,93E-03 |
| ZNF561 | 764,28 | -0,11 | 0,04 | -3,10 | 1,93E-03 |
| BRWD1 | 7881,20 | -0,16 | 0,05 | -3,10 | 1,94E-03 |
| CBX3 | 2079,85 | -0,21 | 0,07 | -3,10 | 1,94E-03 |
| ZFYVE19 | 381,98 | 0,12 | 0,04 | 3,10 | 1,95E-03 |
| C18orf21 | 206,64 | -0,17 | 0,05 | -3,10 | 1,95E-03 |
| SCP2 | 1679,62 | -0,15 | 0,05 | -3,10 | 1,95E-03 |
| ANKRD54 | 210,32 | 0,14 | 0,05 | 3,10 | 1,95E-03 |
| RAN | 2110,78 | -0,17 | 0,05 | -3,10 | 1,95E-03 |
| LSS | 887,06 | 0,16 | 0,05 | 3,10 | 1,95E-03 |
| PCDH1 | 88,40 | 0,36 | 0,12 | 3,10 | 1,95E-03 |
| LYPD6B | 30,34 | 0,64 | 0,21 | 3,10 | 1,97E-03 |
| FHIP1A | 80,34 | 0,35 | 0,11 | 3,09 | 1,97E-03 |
| FBXW2 | 4275,30 | -0,09 | 0,03 | -3,09 | 1,97E-03 |
| ZNF75D | 1421,96 | -0,09 | 0,03 | -3,09 | 1,97E-03 |
| DPAGT1 | 172,41 | 0,14 | 0,05 | 3,09 | 1,97E-03 |
| SNX8 | 252,68 | 0,19 | 0,06 | 3,09 | 1,97E-03 |

| | | | | | |
|---|---|---|---|---|---|
| PAK1IP1 | 162,73 | -0,17 | 0,06 | -3,09 | 1,98E-03 |
| ZNF605 | 541,48 | -0,19 | 0,06 | -3,09 | 1,98E-03 |
| ETFBKMT | 99,83 | 0,16 | 0,05 | 3,09 | 1,98E-03 |
| UROD | 694,32 | 0,15 | 0,05 | 3,09 | 1,99E-03 |
| GALNTL6 | 38,05 | 0,64 | 0,21 | 3,09 | 1,99E-03 |
| LMNA | 308,01 | 0,28 | 0,09 | 3,09 | 1,99E-03 |
| CEP290 | 920,74 | -0,23 | 0,07 | -3,09 | 1,99E-03 |
| DHX29 | 1043,47 | -0,13 | 0,04 | -3,09 | 1,99E-03 |
| ZNF721 | 726,16 | -0,27 | 0,09 | -3,09 | 2,00E-03 |
| LSM2 | 51,10 | 0,17 | 0,06 | 3,09 | 2,00E-03 |
| DESI1 | 677,29 | 0,14 | 0,04 | 3,09 | 2,00E-03 |
| MRPL47 | 197,07 | -0,26 | 0,08 | -3,09 | 2,01E-03 |
| TFAM | 1495,37 | -0,19 | 0,06 | -3,09 | 2,01E-03 |
| SYNCRIP | 3252,88 | -0,15 | 0,05 | -3,09 | 2,01E-03 |
| COPS5 | 1479,35 | -0,09 | 0,03 | -3,09 | 2,01E-03 |
| SLC23A3 | 27,18 | 0,36 | 0,12 | 3,09 | 2,01E-03 |
| SLFN5 | 6880,77 | -0,14 | 0,05 | -3,09 | 2,02E-03 |
| ZNF25 | 538,87 | -0,15 | 0,05 | -3,09 | 2,02E-03 |
| PITPNM1 | 875,89 | 0,25 | 0,08 | 3,09 | 2,02E-03 |
| PUS3 | 177,88 | -0,16 | 0,05 | -3,09 | 2,02E-03 |
| B4GALT7 | 284,69 | 0,12 | 0,04 | 3,09 | 2,03E-03 |
| METTL26 | 129,90 | 0,18 | 0,06 | 3,09 | 2,03E-03 |
| IRAG2 | 6183,70 | -0,16 | 0,05 | -3,09 | 2,03E-03 |
| ABCB8 | 296,97 | 0,23 | 0,07 | 3,08 | 2,04E-03 |
| TOLLIP | 881,69 | 0,19 | 0,06 | 3,08 | 2,04E-03 |
| MFSD3 | 104,71 | 0,16 | 0,05 | 3,08 | 2,05E-03 |
| ZNF517 | 202,83 | 0,15 | 0,05 | 3,08 | 2,04E-03 |
| SCAF11 | 10144,98 | -0,13 | 0,04 | -3,08 | 2,06E-03 |
| PARD3B | 64,92 | 0,52 | 0,17 | 3,08 | 2,07E-03 |
| RIMS3 | 91,19 | 0,29 | 0,10 | 3,08 | 2,07E-03 |
| MRPS10 | 704,36 | -0,14 | 0,05 | -3,08 | 2,08E-03 |
| SPINDOC | 84,23 | 0,20 | 0,07 | 3,08 | 2,08E-03 |
| RPL36A | 172,55 | -0,12 | 0,04 | -3,08 | 2,08E-03 |
| RPAP1 | 277,01 | 0,20 | 0,07 | 3,08 | 2,08E-03 |
| MIGA2 | 616,98 | 0,14 | 0,04 | 3,08 | 2,08E-03 |
| BRD7 | 1886,73 | -0,09 | 0,03 | -3,08 | 2,09E-03 |
| EEF2K | 534,99 | 0,21 | 0,07 | 3,08 | 2,09E-03 |
| ZNF567 | 319,60 | -0,18 | 0,06 | -3,08 | 2,09E-03 |
| ZFP62 | 1235,05 | -0,18 | 0,06 | -3,08 | 2,09E-03 |
| LIN7B | 42,19 | 0,24 | 0,08 | 3,08 | 2,09E-03 |
| SPNS3 | 193,60 | 0,27 | 0,09 | 3,08 | 2,09E-03 |
| TAOK3 | 4054,62 | -0,09 | 0,03 | -3,08 | 2,09E-03 |
| STK4 | 22057,61 | -0,11 | 0,04 | -3,08 | 2,09E-03 |
| CAPN15 | 498,20 | 0,40 | 0,13 | 3,08 | 2,10E-03 |
| PAQR9 | 34,58 | 0,58 | 0,19 | 3,08 | 2,10E-03 |
| CCAR1 | 1919,97 | -0,08 | 0,03 | -3,08 | 2,10E-03 |

| | | | | | |
|---|---|---|---|---|---|
| ZNF696 | 277,48 | 0,14 | 0,04 | 3,07 | 2,11E-03 |
| CEP78 | 2193,40 | -0,21 | 0,07 | -3,07 | 2,11E-03 |
| DDX20 | 686,99 | -0,13 | 0,04 | -3,07 | 2,11E-03 |
| DPP8 | 3900,80 | -0,09 | 0,03 | -3,07 | 2,11E-03 |
| KCNJ5-AS1 | 29,27 | 0,41 | 0,13 | 3,07 | 2,11E-03 |
| MROH8 | 59,34 | 0,27 | 0,09 | 3,07 | 2,11E-03 |
| EXOC1 | 2571,07 | -0,15 | 0,05 | -3,07 | 2,12E-03 |
| PLCH1 | 44,70 | 0,51 | 0,16 | 3,07 | 2,12E-03 |
| JOSD1 | 1208,19 | -0,09 | 0,03 | -3,07 | 2,13E-03 |
| FANCM | 547,17 | -0,17 | 0,06 | -3,07 | 2,13E-03 |
| FEN1 | 216,60 | -0,14 | 0,05 | -3,07 | 2,14E-03 |
| TRPM7 | 3738,78 | -0,13 | 0,04 | -3,07 | 2,14E-03 |
| ZNF213 | 226,67 | 0,20 | 0,07 | 3,07 | 2,14E-03 |
| ZNF761 | 325,25 | -0,19 | 0,06 | -3,07 | 2,14E-03 |
| SLC38A5 | 286,31 | 0,37 | 0,12 | 3,07 | 2,15E-03 |
| DDA1 | 254,93 | 0,18 | 0,06 | 3,07 | 2,15E-03 |
| MYO18A | 1774,58 | 0,20 | 0,06 | 3,07 | 2,15E-03 |
| DBT | 835,58 | -0,07 | 0,02 | -3,07 | 2,15E-03 |
| SLC25A39 | 10114,03 | 0,37 | 0,12 | 3,07 | 2,16E-03 |
| TRPM2 | 297,54 | 0,31 | 0,10 | 3,07 | 2,16E-03 |
| HSPA14 | 347,06 | -0,19 | 0,06 | -3,07 | 2,17E-03 |
| APBA3 | 406,00 | 0,14 | 0,05 | 3,07 | 2,17E-03 |
| CNOT7 | 2556,45 | -0,16 | 0,05 | -3,07 | 2,17E-03 |
| FBXO46 | 127,71 | 0,29 | 0,09 | 3,06 | 2,18E-03 |
| ZFP92 | 59,64 | 0,34 | 0,11 | 3,06 | 2,18E-03 |
| COMMD2 | 913,68 | -0,15 | 0,05 | -3,06 | 2,19E-03 |
| ZNF518B | 1798,59 | -0,15 | 0,05 | -3,06 | 2,19E-03 |
| VPS9D1 | 470,89 | 0,25 | 0,08 | 3,06 | 2,20E-03 |
| LRFN1 | 125,13 | 0,37 | 0,12 | 3,06 | 2,20E-03 |
| GTPBP4 | 809,65 | -0,09 | 0,03 | -3,06 | 2,20E-03 |
| USP20 | 1287,99 | 0,13 | 0,04 | 3,06 | 2,20E-03 |
| FAM114A2 | 764,28 | -0,10 | 0,03 | -3,06 | 2,20E-03 |
| C6orf136 | 121,28 | 0,16 | 0,05 | 3,06 | 2,20E-03 |
| CLIP2 | 228,41 | 0,29 | 0,09 | 3,06 | 2,21E-03 |
| CEP85 | 602,03 | 0,09 | 0,03 | 3,06 | 2,21E-03 |
| FAM217B | 1272,83 | -0,11 | 0,03 | -3,06 | 2,21E-03 |
| BTBD10 | 1881,74 | -0,16 | 0,05 | -3,06 | 2,21E-03 |
| ATP2B1 | 7524,51 | -0,14 | 0,05 | -3,06 | 2,21E-03 |
| PSIP1 | 3300,86 | -0,16 | 0,05 | -3,06 | 2,21E-03 |
| SNX6 | 2427,23 | -0,17 | 0,06 | -3,06 | 2,22E-03 |
| ITGA9 | 105,33 | 0,44 | 0,14 | 3,06 | 2,23E-03 |
| MED7 | 503,78 | -0,18 | 0,06 | -3,06 | 2,23E-03 |
| VIL1 | 90,20 | 0,39 | 0,13 | 3,06 | 2,24E-03 |
| CISH | 501,01 | 0,28 | 0,09 | 3,06 | 2,24E-03 |
| PIWIL4 | 157,68 | 0,17 | 0,05 | 3,06 | 2,24E-03 |
| REST | 2940,13 | -0,12 | 0,04 | -3,06 | 2,24E-03 |

| | | | | | |
|---|---|---|---|---|---|
| NME2 | 41,49 | 0,34 | 0,11 | 3,06 | 2,24E-03 |
| KANSL2 | 1160,89 | -0,09 | 0,03 | -3,06 | 2,24E-03 |
| ABHD12 | 339,92 | 0,14 | 0,05 | 3,06 | 2,25E-03 |
| SLC18A1 | 35,80 | 0,56 | 0,18 | 3,06 | 2,25E-03 |
| GFRA2 | 57,77 | 0,51 | 0,17 | 3,05 | 2,25E-03 |
| ST6GALNAC1 | 43,89 | 0,37 | 0,12 | 3,05 | 2,26E-03 |
| EMC2 | 1046,14 | -0,22 | 0,07 | -3,05 | 2,26E-03 |
| PPP1R26 | 220,45 | 0,21 | 0,07 | 3,05 | 2,27E-03 |
| ZNF638 | 8692,99 | -0,14 | 0,05 | -3,05 | 2,27E-03 |
| JMJD8 | 133,02 | 0,21 | 0,07 | 3,05 | 2,28E-03 |
| SNRPD1 | 410,96 | -0,16 | 0,05 | -3,05 | 2,28E-03 |
| MORC3 | 4123,15 | -0,21 | 0,07 | -3,05 | 2,28E-03 |
| GDPD5 | 429,86 | 0,29 | 0,09 | 3,05 | 2,28E-03 |
| OAF | 134,54 | 0,29 | 0,09 | 3,05 | 2,29E-03 |
| CACYBP | 901,22 | -0,17 | 0,06 | -3,05 | 2,29E-03 |
| CYBC1 | 2976,93 | 0,24 | 0,08 | 3,05 | 2,29E-03 |
| LZTS2 | 284,27 | 0,18 | 0,06 | 3,05 | 2,29E-03 |
| ARHGAP29 | 90,05 | 0,42 | 0,14 | 3,05 | 2,30E-03 |
| PLD2 | 196,53 | 0,27 | 0,09 | 3,05 | 2,30E-03 |
| ARMC3 | 53,34 | 0,44 | 0,15 | 3,05 | 2,30E-03 |
| DHRS7 | 2724,52 | -0,15 | 0,05 | -3,05 | 2,30E-03 |
| TRIM36 | 75,82 | 0,33 | 0,11 | 3,05 | 2,31E-03 |
| GPATCH2L | 6096,06 | -0,08 | 0,03 | -3,05 | 2,31E-03 |
| NCAPH | 64,88 | 0,27 | 0,09 | 3,05 | 2,31E-03 |
| PPDPF | 207,94 | 0,36 | 0,12 | 3,05 | 2,31E-03 |
| POLK | 1777,17 | -0,17 | 0,06 | -3,05 | 2,32E-03 |
| SMAD4 | 4565,87 | -0,10 | 0,03 | -3,05 | 2,32E-03 |
| ZNF888 | 279,03 | -0,19 | 0,06 | -3,05 | 2,32E-03 |
| WBP2 | 1172,69 | 0,25 | 0,08 | 3,05 | 2,32E-03 |
| LGALS3BP | 100,40 | 0,35 | 0,12 | 3,05 | 2,32E-03 |
| NDUFS1 | 2691,52 | -0,07 | 0,02 | -3,04 | 2,33E-03 |
| ALG5 | 374,71 | -0,14 | 0,05 | -3,05 | 2,33E-03 |
| MVK | 191,43 | 0,20 | 0,06 | 3,04 | 2,33E-03 |
| ZZZ3 | 1853,68 | -0,16 | 0,05 | -3,04 | 2,33E-03 |
| UBALD2 | 943,70 | 0,19 | 0,06 | 3,04 | 2,33E-03 |
| ARHGAP6 | 135,27 | 0,29 | 0,09 | 3,04 | 2,33E-03 |
| PSMD7 | 1571,10 | -0,12 | 0,04 | -3,04 | 2,33E-03 |
| GIN1 | 343,90 | -0,20 | 0,07 | -3,04 | 2,34E-03 |
| SGSM2 | 817,47 | 0,22 | 0,07 | 3,04 | 2,35E-03 |
| TRIM9 | 77,42 | 0,38 | 0,13 | 3,04 | 2,35E-03 |
| KCTD1 | 125,80 | 0,28 | 0,09 | 3,04 | 2,35E-03 |
| CEP15 | 40,89 | 0,34 | 0,11 | 3,04 | 2,36E-03 |
| ZNF629 | 153,78 | 0,18 | 0,06 | 3,04 | 2,36E-03 |
| GNA12 | 576,28 | 0,28 | 0,09 | 3,04 | 2,37E-03 |
| BTBD19 | 86,61 | 0,34 | 0,11 | 3,04 | 2,36E-03 |
| SLC35C1 | 165,50 | 0,24 | 0,08 | 3,04 | 2,36E-03 |

| | | | | | |
|---|---|---|---|---|---|
| RNF19A | 4884,65 | -0,14 | 0,04 | -3,04 | 2,37E-03 |
| PDIA5 | 106,98 | 0,22 | 0,07 | 3,04 | 2,37E-03 |
| TGFBR1 | 5653,84 | -0,16 | 0,05 | -3,04 | 2,37E-03 |
| IRS1 | 94,67 | 0,34 | 0,11 | 3,04 | 2,37E-03 |
| TRIM52 | 1700,88 | -0,11 | 0,03 | -3,04 | 2,37E-03 |
| MBNL2 | 1371,07 | -0,16 | 0,05 | -3,04 | 2,38E-03 |
| DAG1 | 278,31 | 0,17 | 0,05 | 3,04 | 2,38E-03 |
| SH3GLB2 | 1115,36 | 0,15 | 0,05 | 3,04 | 2,38E-03 |
| C1orf21 | 812,98 | -0,26 | 0,09 | -3,04 | 2,39E-03 |
| ZMAT5 | 292,42 | 0,10 | 0,03 | 3,04 | 2,39E-03 |
| HMGB1 | 5335,43 | -0,25 | 0,08 | -3,04 | 2,39E-03 |
| DHX36 | 2699,77 | -0,12 | 0,04 | -3,04 | 2,39E-03 |
| RAB6B | 102,87 | 0,40 | 0,13 | 3,04 | 2,39E-03 |
| TSFM | 257,64 | -0,09 | 0,03 | -3,04 | 2,39E-03 |
| OSBP | 1370,33 | -0,07 | 0,02 | -3,04 | 2,40E-03 |
| HECA | 6327,94 | -0,11 | 0,03 | -3,04 | 2,40E-03 |
| GHDC | 151,28 | 0,22 | 0,07 | 3,04 | 2,40E-03 |
| POLR1F | 348,56 | -0,22 | 0,07 | -3,04 | 2,40E-03 |
| NECAP2 | 1931,54 | 0,08 | 0,03 | 3,04 | 2,40E-03 |
| HSD17B4 | 2536,66 | -0,14 | 0,05 | -3,03 | 2,41E-03 |
| ARPC3 | 5232,58 | -0,19 | 0,06 | -3,03 | 2,41E-03 |
| BUB3 | 2787,40 | -0,11 | 0,04 | -3,03 | 2,41E-03 |
| RCL1 | 592,17 | 0,16 | 0,05 | 3,03 | 2,42E-03 |
| MICALL1 | 212,80 | 0,29 | 0,10 | 3,03 | 2,42E-03 |
| CPSF3 | 701,99 | -0,10 | 0,03 | -3,03 | 2,42E-03 |
| IPO13 | 218,75 | 0,22 | 0,07 | 3,03 | 2,42E-03 |
| ZCCHC9 | 744,36 | -0,14 | 0,05 | -3,03 | 2,43E-03 |
| CCNH | 4183,48 | -0,15 | 0,05 | -3,03 | 2,43E-03 |
| CHCHD5 | 45,05 | 0,24 | 0,08 | 3,03 | 2,43E-03 |
| DDR2 | 74,43 | 0,51 | 0,17 | 3,03 | 2,44E-03 |
| CYB5RL | 144,61 | 0,24 | 0,08 | 3,03 | 2,44E-03 |
| RCE1 | 189,43 | 0,13 | 0,04 | 3,03 | 2,44E-03 |
| MCRS1 | 403,65 | 0,16 | 0,05 | 3,03 | 2,44E-03 |
| EIF5A | 1792,77 | 0,13 | 0,04 | 3,03 | 2,45E-03 |
| DOCK11 | 10517,82 | -0,12 | 0,04 | -3,03 | 2,45E-03 |
| CCDC183 | 28,92 | 0,24 | 0,08 | 3,03 | 2,45E-03 |
| REXO5 | 71,54 | 0,30 | 0,10 | 3,03 | 2,45E-03 |
| CRBN | 3273,78 | -0,16 | 0,05 | -3,03 | 2,45E-03 |
| ASNSD1 | 920,92 | -0,27 | 0,09 | -3,03 | 2,46E-03 |
| SRSF10 | 3973,40 | -0,17 | 0,05 | -3,03 | 2,46E-03 |
| TECR | 238,92 | 0,20 | 0,07 | 3,03 | 2,47E-03 |
| TRAF7 | 436,94 | 0,21 | 0,07 | 3,03 | 2,47E-03 |
| SHROOM4 | 42,03 | 0,50 | 0,17 | 3,03 | 2,46E-03 |
| FBXO34 | 1324,40 | -0,09 | 0,03 | -3,03 | 2,47E-03 |
| SPON1 | 72,28 | 0,35 | 0,11 | 3,03 | 2,46E-03 |
| BLZF1 | 870,87 | -0,20 | 0,06 | -3,03 | 2,47E-03 |

| | | | | | |
|---|---|---|---|---|---|
| CEP131 | 182,55 | 0,23 | 0,07 | 3,03 | 2,47E-03 |
| TMEM179B | 270,70 | 0,14 | 0,05 | 3,03 | 2,48E-03 |
| GLIS3 | 86,54 | 0,48 | 0,16 | 3,02 | 2,49E-03 |
| LYL1 | 2553,67 | 0,24 | 0,08 | 3,02 | 2,50E-03 |
| HEXD | 392,68 | 0,24 | 0,08 | 3,02 | 2,51E-03 |
| ITSN2 | 9379,33 | -0,11 | 0,04 | -3,02 | 2,51E-03 |
| NCK1 | 1116,52 | -0,14 | 0,05 | -3,02 | 2,51E-03 |
| CALM2 | 10738,78 | -0,20 | 0,07 | -3,02 | 2,51E-03 |
| GSKIP | 620,69 | -0,23 | 0,08 | -3,02 | 2,52E-03 |
| SPAG5 | 95,40 | 0,21 | 0,07 | 3,02 | 2,52E-03 |
| ZNF521 | 41,82 | 0,53 | 0,17 | 3,02 | 2,52E-03 |
| KIAA0232 | 6083,83 | -0,15 | 0,05 | -3,02 | 2,52E-03 |
| PLD4 | 225,93 | 0,24 | 0,08 | 3,02 | 2,53E-03 |
| PRTFDC1 | 36,30 | 0,32 | 0,11 | 3,02 | 2,53E-03 |
| SLC25A42 | 225,57 | 0,20 | 0,07 | 3,02 | 2,53E-03 |
| PLK2 | 47,60 | 0,30 | 0,10 | 3,02 | 2,54E-03 |
| COPZ2 | 31,82 | 0,30 | 0,10 | 3,02 | 2,55E-03 |
| POLRMT | 518,56 | 0,16 | 0,05 | 3,02 | 2,56E-03 |
| C2CD2 | 351,99 | 0,15 | 0,05 | 3,02 | 2,56E-03 |
| DYTN | 33,40 | 0,42 | 0,14 | 3,02 | 2,56E-03 |
| GDPGP1 | 74,67 | 0,21 | 0,07 | 3,01 | 2,57E-03 |
| DBI | 414,88 | -0,22 | 0,07 | -3,01 | 2,57E-03 |
| MRPS9 | 442,80 | -0,15 | 0,05 | -3,01 | 2,57E-03 |
| SUGCT | 69,89 | 0,31 | 0,10 | 3,01 | 2,57E-03 |
| PLEKHH2 | 61,61 | 0,52 | 0,17 | 3,01 | 2,58E-03 |
| SUOX | 146,93 | 0,21 | 0,07 | 3,01 | 2,58E-03 |
| POU2F2 | 746,08 | 0,29 | 0,10 | 3,01 | 2,58E-03 |
| GIMAP4 | 11182,34 | -0,19 | 0,06 | -3,01 | 2,59E-03 |
| VPS45 | 887,93 | -0,11 | 0,04 | -3,01 | 2,59E-03 |
| RBM42 | 158,63 | 0,21 | 0,07 | 3,01 | 2,58E-03 |
| VWF | 144,90 | 0,53 | 0,18 | 3,01 | 2,59E-03 |
| PCK2 | 198,34 | 0,24 | 0,08 | 3,01 | 2,59E-03 |
| MEF2C | 3327,04 | -0,16 | 0,05 | -3,01 | 2,60E-03 |
| FIP1L1 | 1294,10 | -0,12 | 0,04 | -3,01 | 2,60E-03 |
| CCS | 913,85 | 0,18 | 0,06 | 3,01 | 2,60E-03 |
| LSG1 | 1007,71 | -0,11 | 0,04 | -3,01 | 2,61E-03 |
| ADGRD1 | 187,96 | 0,41 | 0,14 | 3,01 | 2,62E-03 |
| SLC8B1 | 1321,01 | 0,15 | 0,05 | 3,01 | 2,62E-03 |
| SIGLEC8 | 134,30 | 0,43 | 0,14 | 3,01 | 2,62E-03 |
| SRSF3 | 7636,25 | -0,15 | 0,05 | -3,01 | 2,62E-03 |
| PRSS53 | 59,86 | 0,25 | 0,08 | 3,01 | 2,62E-03 |
| LGALS12 | 183,73 | 0,31 | 0,10 | 3,01 | 2,63E-03 |
| NCKAP5L | 261,64 | 0,28 | 0,09 | 3,01 | 2,63E-03 |
| AQP1 | 79,81 | 0,39 | 0,13 | 3,01 | 2,63E-03 |
| ANXA1 | 4894,94 | -0,27 | 0,09 | -3,01 | 2,64E-03 |
| CLASRP | 388,14 | 0,24 | 0,08 | 3,01 | 2,64E-03 |

| | | | | | |
|---|---|---|---|---|---|
| ADGRG6 | 47,34 | 0,46 | 0,15 | 3,01 | 2,64E-03 |
| HOXB3 | 121,31 | 0,34 | 0,11 | 3,01 | 2,64E-03 |
| RNF214 | 312,85 | 0,15 | 0,05 | 3,01 | 2,64E-03 |
| TIPRL | 1008,35 | -0,16 | 0,05 | -3,01 | 2,64E-03 |
| RDH14 | 237,54 | -0,21 | 0,07 | -3,01 | 2,65E-03 |
| TMEM88 | 47,62 | 0,36 | 0,12 | 3,00 | 2,66E-03 |
| ITGAD | 52,21 | 0,53 | 0,18 | 3,00 | 2,66E-03 |
| RSBN1L | 3872,31 | -0,12 | 0,04 | -3,00 | 2,67E-03 |
| TRMT6 | 444,82 | -0,11 | 0,04 | -3,00 | 2,67E-03 |
| PGGHG | 2227,13 | 0,33 | 0,11 | 3,00 | 2,67E-03 |
| AASDHPPT | 1025,29 | -0,16 | 0,05 | -3,00 | 2,68E-03 |
| TPCN1 | 421,52 | 0,32 | 0,11 | 3,00 | 2,68E-03 |
| PIGZ | 34,65 | 0,32 | 0,11 | 3,00 | 2,68E-03 |
| POLD1 | 374,69 | 0,13 | 0,04 | 3,00 | 2,69E-03 |
| SLC27A1 | 145,23 | 0,31 | 0,10 | 3,00 | 2,69E-03 |
| FKBP3 | 462,17 | -0,18 | 0,06 | -3,00 | 2,69E-03 |
| MYNN | 1053,91 | -0,21 | 0,07 | -3,00 | 2,69E-03 |
| GOLGA8K | 23,19 | 0,43 | 0,14 | 3,00 | 2,70E-03 |
| USP28 | 1365,43 | -0,16 | 0,05 | -3,00 | 2,70E-03 |
| RHOF | 566,68 | 0,21 | 0,07 | 3,00 | 2,71E-03 |
| AHCY | 169,98 | 0,19 | 0,06 | 3,00 | 2,71E-03 |
| AGBL2 | 65,36 | 0,24 | 0,08 | 3,00 | 2,71E-03 |
| MTERF1 | 713,58 | -0,18 | 0,06 | -3,00 | 2,72E-03 |
| SRSF1 | 1861,45 | -0,16 | 0,05 | -3,00 | 2,72E-03 |
| SNPH | 161,21 | 0,27 | 0,09 | 3,00 | 2,73E-03 |
| EIF3M | 2256,60 | -0,15 | 0,05 | -3,00 | 2,73E-03 |
| ARHGAP15 | 8440,22 | -0,17 | 0,06 | -3,00 | 2,73E-03 |
| PLA2G4C | 89,50 | 0,50 | 0,17 | 3,00 | 2,73E-03 |
| BRICD5 | 142,63 | 0,18 | 0,06 | 3,00 | 2,73E-03 |
| FASTKD2 | 848,62 | -0,15 | 0,05 | -3,00 | 2,74E-03 |
| ADAM15 | 427,90 | 0,20 | 0,07 | 2,99 | 2,75E-03 |
| PPP2CB | 653,85 | -0,11 | 0,04 | -2,99 | 2,75E-03 |
| SMN1 | 108,31 | -0,16 | 0,05 | -2,99 | 2,75E-03 |
| QTRT1 | 215,67 | 0,14 | 0,05 | 2,99 | 2,76E-03 |
| UTP4 | 310,85 | 0,12 | 0,04 | 2,99 | 2,76E-03 |
| CTDSPL | 225,07 | 0,33 | 0,11 | 2,99 | 2,76E-03 |
| ZCCHC7 | 1820,74 | -0,15 | 0,05 | -2,99 | 2,76E-03 |
| TPM1 | 903,80 | 0,19 | 0,07 | 2,99 | 2,77E-03 |
| CHUK | 1702,42 | -0,16 | 0,05 | -2,99 | 2,77E-03 |
| NFIA | 450,60 | 0,18 | 0,06 | 2,99 | 2,78E-03 |
| ZNF670 | 43,55 | -0,23 | 0,08 | -2,99 | 2,78E-03 |
| PYHIN1 | 1600,67 | -0,19 | 0,06 | -2,99 | 2,78E-03 |
| TNKS2 | 5441,54 | -0,10 | 0,03 | -2,99 | 2,78E-03 |
| TNRC18 | 1958,96 | 0,37 | 0,12 | 2,99 | 2,78E-03 |
| VDAC2 | 926,15 | -0,07 | 0,02 | -2,99 | 2,79E-03 |
| SPTLC1 | 2035,88 | -0,09 | 0,03 | -2,99 | 2,79E-03 |

| | | | | | |
|---|---|---|---|---|---|
| ARHGAP22 | 162,28 | 0,34 | 0,11 | 2,99 | 2,79E-03 |
| AP2A2 | 758,92 | 0,22 | 0,07 | 2,99 | 2,80E-03 |
| MSTO1 | 148,20 | 0,15 | 0,05 | 2,99 | 2,80E-03 |
| ZFHX3 | 421,61 | 0,30 | 0,10 | 2,99 | 2,80E-03 |
| HAUS2 | 489,01 | -0,10 | 0,03 | -2,99 | 2,81E-03 |
| SENP6 | 5708,40 | -0,14 | 0,05 | -2,99 | 2,81E-03 |
| MMADHC | 1756,31 | -0,17 | 0,06 | -2,99 | 2,81E-03 |
| TTC28 | 147,13 | 0,42 | 0,14 | 2,99 | 2,81E-03 |
| UTP15 | 526,42 | -0,14 | 0,05 | -2,99 | 2,82E-03 |
| JMJD1C | 13718,67 | -0,15 | 0,05 | -2,99 | 2,82E-03 |
| MRPS31 | 621,20 | -0,15 | 0,05 | -2,99 | 2,83E-03 |
| GLS2 | 35,79 | 0,29 | 0,10 | 2,99 | 2,82E-03 |
| KLF1 | 1183,81 | 0,38 | 0,13 | 2,99 | 2,83E-03 |
| RPS24 | 3184,70 | -0,27 | 0,09 | -2,99 | 2,83E-03 |
| RAI1 | 341,00 | 0,27 | 0,09 | 2,99 | 2,84E-03 |
| TMEM160 | 177,41 | 0,13 | 0,04 | 2,98 | 2,85E-03 |
| NOL10 | 714,99 | -0,08 | 0,03 | -2,98 | 2,85E-03 |
| CREBRF | 8144,23 | -0,14 | 0,05 | -2,98 | 2,85E-03 |
| ZNF675 | 586,09 | -0,15 | 0,05 | -2,98 | 2,85E-03 |
| NCLN | 457,58 | 0,27 | 0,09 | 2,98 | 2,86E-03 |
| CBR4 | 1550,05 | -0,13 | 0,04 | -2,98 | 2,87E-03 |
| MRPS23 | 435,30 | -0,13 | 0,05 | -2,98 | 2,87E-03 |
| MEX3C | 2102,85 | -0,15 | 0,05 | -2,98 | 2,88E-03 |
| RBM8A | 2629,60 | -0,11 | 0,04 | -2,98 | 2,88E-03 |
| PPP2R3A | 43,02 | 0,44 | 0,15 | 2,98 | 2,88E-03 |
| TCEAL8 | 250,69 | -0,25 | 0,08 | -2,98 | 2,88E-03 |
| SAXO4 | 33,52 | 0,42 | 0,14 | 2,98 | 2,88E-03 |
| GMPS | 1126,73 | -0,10 | 0,03 | -2,98 | 2,89E-03 |
| FTL | 45344,38 | 0,21 | 0,07 | 2,98 | 2,89E-03 |
| CCDC134 | 128,93 | 0,21 | 0,07 | 2,98 | 2,90E-03 |
| ING4 | 875,81 | -0,10 | 0,03 | -2,98 | 2,90E-03 |
| CRTC1 | 324,42 | 0,27 | 0,09 | 2,98 | 2,91E-03 |
| CIR1 | 1583,75 | -0,16 | 0,06 | -2,98 | 2,91E-03 |
| LIN54 | 1258,46 | -0,10 | 0,03 | -2,98 | 2,91E-03 |
| PLXNA4 | 235,06 | 0,35 | 0,12 | 2,98 | 2,91E-03 |
| PROCA1 | 117,38 | 0,26 | 0,09 | 2,98 | 2,92E-03 |
| NFYB | 443,51 | -0,22 | 0,08 | -2,98 | 2,91E-03 |
| CDYL2 | 359,73 | 0,15 | 0,05 | 2,98 | 2,92E-03 |
| STK31 | 48,69 | 0,43 | 0,14 | 2,98 | 2,92E-03 |
| MSL2 | 4443,83 | -0,07 | 0,02 | -2,98 | 2,92E-03 |
| ATG9A | 460,33 | 0,18 | 0,06 | 2,98 | 2,93E-03 |
| CLEC2B | 1631,99 | -0,27 | 0,09 | -2,98 | 2,93E-03 |
| MEGF6 | 1571,99 | 0,22 | 0,07 | 2,97 | 2,93E-03 |
| POLR2J2 | 101,06 | 0,32 | 0,11 | 2,97 | 2,93E-03 |
| CCDC93 | 3556,51 | -0,08 | 0,03 | -2,97 | 2,94E-03 |
| RHOT2 | 1100,33 | 0,13 | 0,04 | 2,97 | 2,94E-03 |

| | | | | | |
|---|---|---|---|---|---|
| PER2 | 263,61 | 0,20 | 0,07 | 2,97 | 2,94E-03 |
| ATP2C2 | 56,75 | 0,59 | 0,20 | 2,97 | 2,96E-03 |
| NSMF | 377,09 | 0,21 | 0,07 | 2,97 | 2,96E-03 |
| FZR1 | 1360,68 | 0,16 | 0,05 | 2,97 | 2,96E-03 |
| PCNA | 582,32 | -0,15 | 0,05 | -2,97 | 2,97E-03 |
| IBA57 | 583,53 | 0,16 | 0,06 | 2,97 | 2,97E-03 |
| ATP6V1G2 | 44,59 | 0,20 | 0,07 | 2,97 | 2,97E-03 |
| PLRG1 | 1069,40 | -0,17 | 0,06 | -2,97 | 2,97E-03 |
| ATP6V1F | 152,18 | 0,12 | 0,04 | 2,97 | 2,97E-03 |
| GALNT8 | 36,05 | 0,46 | 0,16 | 2,97 | 2,97E-03 |
| TMUB2 | 670,10 | 0,15 | 0,05 | 2,97 | 2,97E-03 |
| MIOS | 1115,31 | -0,16 | 0,05 | -2,97 | 2,98E-03 |
| UFSP2 | 372,68 | -0,14 | 0,05 | -2,97 | 2,99E-03 |
| FAM171B | 28,23 | 0,37 | 0,12 | 2,97 | 2,99E-03 |
| CNBP | 8703,83 | -0,14 | 0,05 | -2,97 | 2,99E-03 |
| FAM3A | 262,24 | 0,14 | 0,05 | 2,97 | 2,99E-03 |
| NFYA | 1916,53 | -0,09 | 0,03 | -2,97 | 2,99E-03 |
| DNAJC13 | 3965,66 | -0,11 | 0,04 | -2,97 | 3,00E-03 |
| ZNF618 | 139,90 | 0,34 | 0,12 | 2,97 | 3,00E-03 |
| ENDOV | 218,44 | 0,18 | 0,06 | 2,97 | 3,00E-03 |
| DDX27 | 1032,75 | -0,09 | 0,03 | -2,97 | 3,01E-03 |
| BRI3BP | 1107,64 | 0,11 | 0,04 | 2,97 | 3,02E-03 |
| CAMTA2 | 353,90 | 0,36 | 0,12 | 2,97 | 3,02E-03 |
| RRM1 | 623,42 | -0,12 | 0,04 | -2,97 | 3,02E-03 |
| MACROH2A2 | 60,61 | 0,18 | 0,06 | 2,97 | 3,02E-03 |
| ZNF707 | 374,84 | 0,16 | 0,05 | 2,97 | 3,02E-03 |
| EEIG1 | 673,30 | 0,31 | 0,10 | 2,96 | 3,03E-03 |
| HAUS3 | 996,61 | -0,14 | 0,05 | -2,96 | 3,03E-03 |
| SNTB1 | 1091,98 | 0,11 | 0,04 | 2,96 | 3,03E-03 |
| GPX1 | 7252,73 | 0,32 | 0,11 | 2,96 | 3,04E-03 |
| AMOTL1 | 112,84 | 0,36 | 0,12 | 2,96 | 3,04E-03 |
| SELENOO | 690,52 | 0,14 | 0,05 | 2,96 | 3,04E-03 |
| CRACDL | 85,53 | 0,37 | 0,13 | 2,96 | 3,04E-03 |
| PGGT1B | 3065,26 | -0,17 | 0,06 | -2,96 | 3,04E-03 |
| SLC39A14 | 119,58 | 0,21 | 0,07 | 2,96 | 3,05E-03 |
| RNASEL | 3008,16 | -0,13 | 0,04 | -2,96 | 3,05E-03 |
| CEP152 | 649,28 | -0,14 | 0,05 | -2,96 | 3,05E-03 |
| ENO2 | 308,26 | 0,18 | 0,06 | 2,96 | 3,05E-03 |
| HLTF | 662,83 | -0,18 | 0,06 | -2,96 | 3,05E-03 |
| RUBCNL | 2539,43 | -0,23 | 0,08 | -2,96 | 3,06E-03 |
| SENP7 | 3370,66 | -0,18 | 0,06 | -2,96 | 3,06E-03 |
| IPO8 | 2443,25 | -0,10 | 0,03 | -2,96 | 3,06E-03 |
| P2RX5-TAX1BP3 | 35,89 | 0,33 | 0,11 | 2,96 | 3,06E-03 |
| DEAF1 | 309,55 | 0,18 | 0,06 | 2,96 | 3,06E-03 |
| UBE2O | 1091,99 | 0,31 | 0,11 | 2,96 | 3,06E-03 |
| GFM2 | 825,69 | -0,13 | 0,05 | -2,96 | 3,07E-03 |

| | | | | | |
|---|---|---|---|---|---|
| HPF1 | 186,87 | -0,15 | 0,05 | -2,96 | 3,07E-03 |
| CACNA1E | 423,16 | 0,36 | 0,12 | 2,96 | 3,07E-03 |
| HDAC2 | 925,05 | -0,14 | 0,05 | -2,96 | 3,07E-03 |
| INSIG2 | 844,10 | -0,16 | 0,05 | -2,96 | 3,08E-03 |
| YIPF5 | 951,60 | -0,15 | 0,05 | -2,96 | 3,08E-03 |
| GPI | 656,61 | 0,20 | 0,07 | 2,96 | 3,09E-03 |
| LOXL2 | 149,42 | 0,22 | 0,07 | 2,96 | 3,09E-03 |
| C10orf143 | 173,60 | 0,17 | 0,06 | 2,96 | 3,10E-03 |
| STT3B | 5348,16 | -0,16 | 0,05 | -2,96 | 3,10E-03 |
| AP5Z1 | 766,32 | 0,19 | 0,06 | 2,96 | 3,10E-03 |
| THRA | 381,88 | 0,18 | 0,06 | 2,96 | 3,11E-03 |
| FARSA | 284,75 | 0,12 | 0,04 | 2,96 | 3,11E-03 |
| EFCAB12 | 69,81 | 0,35 | 0,12 | 2,96 | 3,11E-03 |
| PGAP3 | 181,15 | 0,17 | 0,06 | 2,96 | 3,12E-03 |
| TMEM94 | 325,29 | 0,27 | 0,09 | 2,96 | 3,12E-03 |
| VRK1 | 777,16 | -0,25 | 0,08 | -2,96 | 3,12E-03 |
| BSG | 14800,91 | 0,34 | 0,11 | 2,96 | 3,12E-03 |
| MUS81 | 635,18 | 0,11 | 0,04 | 2,96 | 3,12E-03 |
| SPRTN | 869,90 | -0,12 | 0,04 | -2,96 | 3,12E-03 |
| SERBP1 | 5732,24 | -0,12 | 0,04 | -2,96 | 3,12E-03 |
| UFL1 | 1441,94 | -0,20 | 0,07 | -2,95 | 3,13E-03 |
| TOP3B | 776,93 | 0,12 | 0,04 | 2,95 | 3,13E-03 |
| BRD10 | 3593,78 | -0,08 | 0,03 | -2,95 | 3,14E-03 |
| CNNM4 | 139,80 | 0,26 | 0,09 | 2,95 | 3,14E-03 |
| CEP97 | 1021,99 | -0,14 | 0,05 | -2,95 | 3,14E-03 |
| SELENOF | 3801,55 | -0,17 | 0,06 | -2,95 | 3,15E-03 |
| OSTC | 697,81 | -0,21 | 0,07 | -2,95 | 3,15E-03 |
| CEP89 | 246,17 | 0,21 | 0,07 | 2,95 | 3,15E-03 |
| SGK3 | 1140,05 | -0,09 | 0,03 | -2,95 | 3,15E-03 |
| TMEM167B | 1952,57 | -0,17 | 0,06 | -2,95 | 3,18E-03 |
| OXER1 | 60,61 | 0,38 | 0,13 | 2,95 | 3,18E-03 |
| HIBADH | 341,94 | -0,12 | 0,04 | -2,95 | 3,18E-03 |
| KMT2E | 7245,13 | -0,09 | 0,03 | -2,95 | 3,18E-03 |
| NEPRO | 1414,80 | -0,12 | 0,04 | -2,95 | 3,18E-03 |
| RARG | 241,29 | 0,19 | 0,07 | 2,95 | 3,19E-03 |
| ZNF613 | 275,87 | -0,20 | 0,07 | -2,95 | 3,19E-03 |
| LEMD3 | 2328,05 | -0,12 | 0,04 | -2,95 | 3,19E-03 |
| GINM1 | 691,74 | -0,11 | 0,04 | -2,95 | 3,20E-03 |
| EPS15 | 4795,03 | -0,09 | 0,03 | -2,95 | 3,20E-03 |
| GLI4 | 158,62 | 0,27 | 0,09 | 2,95 | 3,20E-03 |
| ZNF12 | 1764,65 | -0,17 | 0,06 | -2,95 | 3,20E-03 |
| GIMAP6 | 6924,59 | -0,12 | 0,04 | -2,95 | 3,21E-03 |
| DXO | 104,23 | 0,14 | 0,05 | 2,95 | 3,21E-03 |
| PLEKHB1 | 167,41 | 0,23 | 0,08 | 2,95 | 3,22E-03 |
| ATP10D | 1636,54 | -0,12 | 0,04 | -2,95 | 3,22E-03 |
| RAPSN | 40,02 | 0,34 | 0,12 | 2,94 | 3,23E-03 |

| | | | | | |
|---|---|---|---|---|---|
| KBTBD2 | 3916,19 | -0,13 | 0,05 | -2,94 | 3,23E-03 |
| IDI1 | 1198,31 | -0,22 | 0,08 | -2,94 | 3,23E-03 |
| NDUFS5 | 315,25 | -0,28 | 0,09 | -2,94 | 3,24E-03 |
| FAM117A | 1527,06 | 0,24 | 0,08 | 2,94 | 3,24E-03 |
| TMEM169 | 129,67 | 0,12 | 0,04 | 2,94 | 3,24E-03 |
| BCL9 | 141,77 | 0,25 | 0,09 | 2,94 | 3,25E-03 |
| FANCE | 150,48 | 0,12 | 0,04 | 2,94 | 3,25E-03 |
| ASCC3 | 3165,78 | -0,18 | 0,06 | -2,94 | 3,25E-03 |
| CEACAM8 | 109,56 | 0,46 | 0,16 | 2,94 | 3,25E-03 |
| ABCC3 | 441,54 | 0,31 | 0,11 | 2,94 | 3,26E-03 |
| PNP | 1600,51 | 0,17 | 0,06 | 2,94 | 3,26E-03 |
| PUM2 | 7768,72 | -0,10 | 0,03 | -2,94 | 3,27E-03 |
| TRIM62 | 169,84 | 0,31 | 0,11 | 2,94 | 3,28E-03 |
| ADORA3 | 69,55 | 0,34 | 0,12 | 2,94 | 3,28E-03 |
| ZNF624 | 301,77 | -0,20 | 0,07 | -2,94 | 3,28E-03 |
| PARVB | 229,49 | 0,27 | 0,09 | 2,94 | 3,28E-03 |
| PGAP6 | 627,68 | 0,26 | 0,09 | 2,94 | 3,28E-03 |
| SBF1 | 1566,11 | 0,28 | 0,10 | 2,94 | 3,29E-03 |
| NAE1 | 685,85 | -0,14 | 0,05 | -2,94 | 3,30E-03 |
| RBM27 | 941,76 | -0,15 | 0,05 | -2,94 | 3,30E-03 |
| ZNF84 | 972,35 | -0,17 | 0,06 | -2,94 | 3,30E-03 |
| DLD | 1687,49 | -0,14 | 0,05 | -2,94 | 3,31E-03 |
| GREM2 | 34,82 | 0,39 | 0,13 | 2,94 | 3,31E-03 |
| CDC123 | 1855,24 | -0,12 | 0,04 | -2,94 | 3,31E-03 |
| ISY1 | 513,04 | -0,09 | 0,03 | -2,94 | 3,32E-03 |
| PJVK | 47,18 | 0,26 | 0,09 | 2,94 | 3,33E-03 |
| SPSB1 | 44,96 | 0,32 | 0,11 | 2,94 | 3,33E-03 |
| ACTL6A | 704,43 | -0,13 | 0,04 | -2,94 | 3,33E-03 |
| SSR3 | 2682,15 | -0,14 | 0,05 | -2,93 | 3,34E-03 |
| ZNF446 | 126,51 | 0,18 | 0,06 | 2,93 | 3,34E-03 |
| FBXL12 | 293,49 | 0,11 | 0,04 | 2,93 | 3,34E-03 |
| GLB1L | 50,63 | 0,32 | 0,11 | 2,93 | 3,34E-03 |
| ZNF649 | 588,29 | -0,14 | 0,05 | -2,93 | 3,35E-03 |
| TXNRD1 | 4604,47 | -0,14 | 0,05 | -2,93 | 3,35E-03 |
| THOC1 | 1195,26 | -0,13 | 0,04 | -2,93 | 3,36E-03 |
| RANBP10 | 1177,14 | 0,28 | 0,10 | 2,93 | 3,37E-03 |
| TTYH3 | 440,36 | 0,35 | 0,12 | 2,93 | 3,37E-03 |
| STRAP | 527,91 | -0,14 | 0,05 | -2,93 | 3,38E-03 |
| HYAL2 | 165,23 | 0,13 | 0,04 | 2,93 | 3,38E-03 |
| FMR1 | 3316,23 | -0,14 | 0,05 | -2,93 | 3,39E-03 |
| TLK1 | 2323,28 | -0,12 | 0,04 | -2,93 | 3,39E-03 |
| UTP11 | 497,75 | -0,10 | 0,04 | -2,93 | 3,40E-03 |
| ZNF611 | 1089,41 | -0,10 | 0,03 | -2,93 | 3,40E-03 |
| HDAC1 | 3072,35 | -0,09 | 0,03 | -2,93 | 3,40E-03 |
| PRKAG1 | 895,81 | -0,09 | 0,03 | -2,93 | 3,40E-03 |
| FBF1 | 135,70 | 0,12 | 0,04 | 2,93 | 3,41E-03 |

| | | | | | |
|---|---|---|---|---|---|
| ERCC6L2 | 2162,15 | -0,14 | 0,05 | -2,93 | 3,41E-03 |
| SASH1 | 245,11 | 0,29 | 0,10 | 2,93 | 3,41E-03 |
| LRRC27 | 215,21 | 0,16 | 0,05 | 2,93 | 3,43E-03 |
| CDKAL1 | 1221,85 | -0,12 | 0,04 | -2,93 | 3,43E-03 |
| STK36 | 242,20 | 0,16 | 0,06 | 2,93 | 3,43E-03 |
| CATSPERE | 68,43 | 0,25 | 0,08 | 2,93 | 3,43E-03 |
| LSM10 | 432,66 | 0,13 | 0,05 | 2,93 | 3,44E-03 |
| PORCN | 49,02 | 0,30 | 0,10 | 2,93 | 3,43E-03 |
| KIN | 1086,57 | -0,10 | 0,03 | -2,93 | 3,44E-03 |
| SH2B1 | 484,51 | 0,26 | 0,09 | 2,93 | 3,44E-03 |
| CCNI2 | 83,08 | 0,20 | 0,07 | 2,93 | 3,44E-03 |
| ATP5F1A | 5429,08 | -0,11 | 0,04 | -2,93 | 3,44E-03 |
| POLE3 | 841,45 | -0,13 | 0,04 | -2,93 | 3,44E-03 |
| RPL23 | 3050,42 | -0,43 | 0,15 | -2,92 | 3,46E-03 |
| IFT122 | 404,20 | 0,15 | 0,05 | 2,92 | 3,46E-03 |
| DRG1 | 694,08 | -0,09 | 0,03 | -2,92 | 3,48E-03 |
| NDUFA5 | 575,19 | -0,24 | 0,08 | -2,92 | 3,49E-03 |
| GPS1 | 621,34 | 0,11 | 0,04 | 2,92 | 3,50E-03 |
| ENOPH1 | 481,44 | -0,13 | 0,04 | -2,92 | 3,50E-03 |
| PCNX3 | 819,63 | 0,26 | 0,09 | 2,92 | 3,50E-03 |
| MRPL39 | 285,65 | -0,16 | 0,05 | -2,92 | 3,50E-03 |
| EEF1B2 | 1501,21 | -0,35 | 0,12 | -2,92 | 3,51E-03 |
| FIGNL1 | 395,89 | -0,18 | 0,06 | -2,92 | 3,51E-03 |
| ESPN | 206,62 | 0,53 | 0,18 | 2,92 | 3,51E-03 |
| HSPA2 | 35,79 | 0,34 | 0,12 | 2,92 | 3,51E-03 |
| GGCT | 122,80 | -0,17 | 0,06 | -2,92 | 3,52E-03 |
| MCTS1 | 417,42 | -0,14 | 0,05 | -2,92 | 3,53E-03 |
| TRIM23 | 1721,47 | -0,20 | 0,07 | -2,92 | 3,53E-03 |
| ZFPL1 | 224,34 | 0,12 | 0,04 | 2,92 | 3,53E-03 |
| CTNNB1 | 6985,29 | -0,08 | 0,03 | -2,92 | 3,53E-03 |
| EIF4G2 | 8403,81 | -0,10 | 0,03 | -2,92 | 3,53E-03 |
| FBXL3 | 3999,75 | -0,15 | 0,05 | -2,92 | 3,53E-03 |
| ATG4A | 330,30 | -0,12 | 0,04 | -2,92 | 3,54E-03 |
| CHM | 871,01 | -0,15 | 0,05 | -2,92 | 3,55E-03 |
| CTSS | 51151,88 | -0,18 | 0,06 | -2,92 | 3,56E-03 |
| ZBTB5 | 512,87 | -0,10 | 0,03 | -2,91 | 3,56E-03 |
| AK6 | 82,84 | -0,23 | 0,08 | -2,91 | 3,57E-03 |
| SYAP1 | 1310,13 | -0,10 | 0,03 | -2,91 | 3,57E-03 |
| YIPF2 | 335,98 | 0,12 | 0,04 | 2,91 | 3,60E-03 |
| NUDT4 | 2800,09 | 0,22 | 0,07 | 2,91 | 3,60E-03 |
| NUP54 | 812,02 | -0,15 | 0,05 | -2,91 | 3,61E-03 |
| SCARB1 | 171,56 | 0,29 | 0,10 | 2,91 | 3,61E-03 |
| FAM72B | 25,08 | 0,27 | 0,09 | 2,91 | 3,61E-03 |
| ENSG00000280433 | 145,29 | -0,21 | 0,07 | -2,91 | 3,61E-03 |
| ZNF91 | 1747,92 | -0,19 | 0,06 | -2,91 | 3,61E-03 |
| CPAMD8 | 148,18 | 0,37 | 0,13 | 2,91 | 3,62E-03 |

| | | | | | |
|---|---|---|---|---|---|
| AGPAT3 | 1380,08 | 0,17 | 0,06 | 2,91 | 3,62E-03 |
| POC1A | 65,39 | 0,19 | 0,07 | 2,91 | 3,63E-03 |
| MIIP | 922,88 | 0,15 | 0,05 | 2,91 | 3,63E-03 |
| INTS12 | 348,38 | -0,10 | 0,04 | -2,91 | 3,63E-03 |
| EMC10 | 1051,85 | 0,11 | 0,04 | 2,91 | 3,63E-03 |
| CYCS | 1504,21 | -0,17 | 0,06 | -2,91 | 3,64E-03 |
| TSNARE1 | 288,84 | 0,23 | 0,08 | 2,91 | 3,65E-03 |
| SETDB2 | 1363,33 | -0,10 | 0,04 | -2,91 | 3,65E-03 |
| DERL3 | 37,57 | 0,38 | 0,13 | 2,91 | 3,65E-03 |
| NIPA2 | 1372,72 | -0,08 | 0,03 | -2,91 | 3,65E-03 |
| TSC2 | 937,97 | 0,30 | 0,10 | 2,91 | 3,66E-03 |
| DTL | 57,11 | 0,34 | 0,12 | 2,91 | 3,66E-03 |
| ARFGAP1 | 674,22 | 0,23 | 0,08 | 2,91 | 3,66E-03 |
| OARD1 | 1104,13 | -0,12 | 0,04 | -2,91 | 3,67E-03 |
| TPPP | 70,63 | 0,42 | 0,14 | 2,90 | 3,68E-03 |
| SF3B6 | 473,83 | -0,27 | 0,09 | -2,90 | 3,68E-03 |
| VAV2 | 290,42 | 0,28 | 0,10 | 2,90 | 3,68E-03 |
| TRA2B | 6595,92 | -0,13 | 0,04 | -2,90 | 3,69E-03 |
| ACTR8 | 1239,01 | -0,09 | 0,03 | -2,90 | 3,69E-03 |
| STPG2 | 23,33 | 0,32 | 0,11 | 2,90 | 3,69E-03 |
| GPATCH2 | 937,27 | -0,15 | 0,05 | -2,90 | 3,69E-03 |
| ZNF510 | 855,47 | -0,18 | 0,06 | -2,90 | 3,69E-03 |
| SKIC8 | 831,15 | -0,08 | 0,03 | -2,90 | 3,71E-03 |
| CILP | 39,48 | 0,36 | 0,12 | 2,90 | 3,71E-03 |
| PRDM8 | 377,23 | 0,15 | 0,05 | 2,90 | 3,71E-03 |
| RBM41 | 768,24 | -0,14 | 0,05 | -2,90 | 3,72E-03 |
| TIGD7 | 464,15 | -0,22 | 0,08 | -2,90 | 3,72E-03 |
| DYNLT3 | 381,49 | -0,30 | 0,10 | -2,90 | 3,72E-03 |
| PPP2R3C | 1396,67 | -0,17 | 0,06 | -2,90 | 3,72E-03 |
| NECAB2 | 54,33 | 0,41 | 0,14 | 2,90 | 3,73E-03 |
| TESK1 | 142,15 | 0,21 | 0,07 | 2,90 | 3,73E-03 |
| F2RL3 | 38,22 | 0,37 | 0,13 | 2,90 | 3,74E-03 |
| PHIP | 9164,68 | -0,16 | 0,06 | -2,90 | 3,74E-03 |
| RPL7L1 | 2200,91 | -0,10 | 0,03 | -2,90 | 3,74E-03 |
| COMT | 626,24 | 0,11 | 0,04 | 2,90 | 3,74E-03 |
| PAIP2 | 2449,76 | -0,17 | 0,06 | -2,90 | 3,75E-03 |
| TMEM59 | 3964,73 | -0,15 | 0,05 | -2,90 | 3,75E-03 |
| STING1 | 497,86 | 0,18 | 0,06 | 2,90 | 3,75E-03 |
| PWWP2B | 280,70 | 0,14 | 0,05 | 2,90 | 3,76E-03 |
| BTLA | 979,86 | -0,24 | 0,08 | -2,90 | 3,76E-03 |
| UEVLD | 750,60 | -0,10 | 0,04 | -2,90 | 3,76E-03 |
| P2RX7 | 671,75 | 0,17 | 0,06 | 2,90 | 3,76E-03 |
| AAAS | 150,30 | 0,16 | 0,06 | 2,90 | 3,76E-03 |
| PEX11G | 31,03 | 0,37 | 0,13 | 2,90 | 3,76E-03 |
| MAZ | 547,46 | 0,26 | 0,09 | 2,90 | 3,76E-03 |
| DCAF15 | 357,24 | 0,21 | 0,07 | 2,90 | 3,76E-03 |

| | | | | | |
|---|---|---|---|---|---|
| AZIN1 | 4190,11 | -0,14 | 0,05 | -2,90 | 3,77E-03 |
| BMS1 | 1326,98 | -0,11 | 0,04 | -2,90 | 3,77E-03 |
| GLOD4 | 1153,59 | -0,11 | 0,04 | -2,90 | 3,79E-03 |
| TMEM200A | 57,74 | 0,34 | 0,12 | 2,90 | 3,79E-03 |
| UMPS | 660,39 | -0,10 | 0,04 | -2,89 | 3,80E-03 |
| HRAS | 90,20 | 0,19 | 0,07 | 2,89 | 3,80E-03 |
| TTC13 | 2115,25 | -0,10 | 0,03 | -2,89 | 3,80E-03 |
| FASTK | 584,99 | 0,13 | 0,05 | 2,89 | 3,80E-03 |
| RABL3 | 563,70 | -0,14 | 0,05 | -2,89 | 3,81E-03 |
| COMMD6 | 279,18 | -0,40 | 0,14 | -2,89 | 3,81E-03 |
| ZNF549 | 654,55 | -0,11 | 0,04 | -2,89 | 3,81E-03 |
| ITGB1BP2 | 49,46 | 0,22 | 0,07 | 2,89 | 3,82E-03 |
| SDK2 | 129,23 | 0,49 | 0,17 | 2,89 | 3,82E-03 |
| DNAJC25 | 152,02 | -0,14 | 0,05 | -2,89 | 3,82E-03 |
| DTYMK | 181,30 | 0,11 | 0,04 | 2,89 | 3,82E-03 |
| ATP13A1 | 570,60 | 0,22 | 0,07 | 2,89 | 3,82E-03 |
| HINFP | 441,65 | 0,13 | 0,05 | 2,89 | 3,83E-03 |
| CLTA | 1110,92 | -0,09 | 0,03 | -2,89 | 3,84E-03 |
| CABP5 | 48,58 | 0,39 | 0,14 | 2,89 | 3,85E-03 |
| NOL6 | 328,30 | 0,20 | 0,07 | 2,89 | 3,85E-03 |
| CENPC | 2596,78 | -0,14 | 0,05 | -2,89 | 3,85E-03 |
| OR2W3 | 404,47 | 0,40 | 0,14 | 2,89 | 3,86E-03 |
| PPP2R5E | 1367,86 | -0,10 | 0,04 | -2,89 | 3,86E-03 |
| PCNX4 | 3209,19 | -0,10 | 0,03 | -2,89 | 3,86E-03 |
| AGTPBP1 | 7405,00 | -0,16 | 0,06 | -2,89 | 3,87E-03 |
| ZNF322 | 518,28 | -0,20 | 0,07 | -2,89 | 3,88E-03 |
| SEC14L4 | 86,10 | 0,38 | 0,13 | 2,89 | 3,88E-03 |
| CCT3 | 2022,36 | -0,11 | 0,04 | -2,89 | 3,88E-03 |
| TTC21B | 1479,36 | -0,13 | 0,04 | -2,89 | 3,88E-03 |
| A3GALT2 | 30,86 | 0,38 | 0,13 | 2,89 | 3,89E-03 |
| CLTC | 7146,67 | -0,11 | 0,04 | -2,89 | 3,89E-03 |
| KMT5A | 343,19 | 0,14 | 0,05 | 2,89 | 3,90E-03 |
| LTK | 157,07 | 0,31 | 0,11 | 2,89 | 3,90E-03 |
| SREBF2 | 616,50 | 0,26 | 0,09 | 2,89 | 3,90E-03 |
| UAP1L1 | 141,78 | 0,30 | 0,11 | 2,89 | 3,91E-03 |
| MAP2K3 | 2852,23 | 0,27 | 0,09 | 2,89 | 3,91E-03 |
| DCTN6 | 190,20 | -0,14 | 0,05 | -2,88 | 3,92E-03 |
| GARNL3 | 156,19 | 0,30 | 0,10 | 2,88 | 3,92E-03 |
| RBM12 | 3503,27 | -0,13 | 0,05 | -2,88 | 3,94E-03 |
| ZNF845 | 588,11 | -0,26 | 0,09 | -2,88 | 3,94E-03 |
| COL13A1 | 90,64 | 0,51 | 0,18 | 2,88 | 3,94E-03 |
| ENSG00000258311 | 36,84 | 0,29 | 0,10 | 2,88 | 3,95E-03 |
| BAMBI | 26,77 | 0,44 | 0,15 | 2,88 | 3,97E-03 |
| AFG2A | 1037,98 | -0,13 | 0,05 | -2,88 | 3,98E-03 |
| ZNF500 | 238,01 | 0,24 | 0,08 | 2,88 | 3,98E-03 |
| ZNF22 | 499,49 | -0,26 | 0,09 | -2,88 | 3,98E-03 |

| | | | | | |
|---|---|---|---|---|---|
| AP4E1 | 1288,66 | -0,10 | 0,04 | -2,88 | 3,99E-03 |
| GAS2 | 53,03 | 0,27 | 0,09 | 2,88 | 4,00E-03 |
| TSNAX | 1342,31 | -0,18 | 0,06 | -2,88 | 4,00E-03 |
| UCKL1 | 267,67 | 0,15 | 0,05 | 2,88 | 4,00E-03 |
| SCLY | 209,78 | 0,17 | 0,06 | 2,88 | 4,02E-03 |
| BACE2 | 356,80 | 0,24 | 0,08 | 2,88 | 4,02E-03 |
| ZMYM3 | 244,50 | 0,21 | 0,07 | 2,88 | 4,02E-03 |
| ZMYM5 | 860,74 | -0,13 | 0,04 | -2,88 | 4,03E-03 |
| UCP2 | 1447,38 | 0,28 | 0,10 | 2,88 | 4,03E-03 |
| TMEM214 | 275,05 | 0,20 | 0,07 | 2,88 | 4,03E-03 |
| RPS23 | 8134,53 | -0,29 | 0,10 | -2,87 | 4,04E-03 |
| CUL1 | 1511,98 | -0,09 | 0,03 | -2,87 | 4,05E-03 |
| CDK15 | 44,79 | 0,53 | 0,18 | 2,87 | 4,06E-03 |
| ZDHHC6 | 1388,46 | -0,09 | 0,03 | -2,87 | 4,06E-03 |
| WBP11 | 427,90 | -0,12 | 0,04 | -2,87 | 4,06E-03 |
| ABRACL | 653,12 | -0,19 | 0,07 | -2,87 | 4,06E-03 |
| ADH5 | 1239,61 | -0,16 | 0,05 | -2,87 | 4,07E-03 |
| NOL11 | 1068,46 | -0,13 | 0,04 | -2,87 | 4,07E-03 |
| SOCS1 | 66,50 | 0,33 | 0,12 | 2,87 | 4,07E-03 |
| C1QTNF9 | 39,85 | 0,17 | 0,06 | 2,87 | 4,08E-03 |
| RILPL1 | 144,34 | 0,25 | 0,09 | 2,87 | 4,10E-03 |
| UPB1 | 96,01 | 0,30 | 0,11 | 2,87 | 4,10E-03 |
| UBE2V2 | 597,26 | -0,13 | 0,05 | -2,87 | 4,10E-03 |
| JAK1 | 14428,73 | -0,11 | 0,04 | -2,87 | 4,10E-03 |
| WDR45 | 2530,01 | 0,19 | 0,07 | 2,87 | 4,11E-03 |
| TRABD | 648,14 | 0,23 | 0,08 | 2,87 | 4,11E-03 |
| HNRNPC | 14194,41 | -0,11 | 0,04 | -2,87 | 4,15E-03 |
| GRIP1 | 70,58 | 0,39 | 0,13 | 2,87 | 4,16E-03 |
| TREML1 | 172,00 | 0,35 | 0,12 | 2,87 | 4,16E-03 |
| REC8 | 183,05 | 0,34 | 0,12 | 2,87 | 4,16E-03 |
| ERCC6 | 2289,32 | -0,09 | 0,03 | -2,87 | 4,16E-03 |
| CNIH3 | 192,96 | 0,19 | 0,07 | 2,86 | 4,17E-03 |
| DPM2 | 1279,32 | 0,29 | 0,10 | 2,86 | 4,17E-03 |
| N4BP3 | 93,31 | 0,31 | 0,11 | 2,86 | 4,17E-03 |
| ZBTB43 | 1085,33 | -0,06 | 0,02 | -2,86 | 4,17E-03 |
| TPPP3 | 60,95 | 0,28 | 0,10 | 2,86 | 4,18E-03 |
| CCDC159 | 275,22 | 0,15 | 0,05 | 2,86 | 4,18E-03 |
| PDE3A | 92,58 | 0,35 | 0,12 | 2,86 | 4,18E-03 |
| INKA1 | 42,55 | 0,20 | 0,07 | 2,86 | 4,18E-03 |
| EIF2S3 | 5545,83 | -0,10 | 0,04 | -2,86 | 4,18E-03 |
| NOP2 | 57,45 | 0,14 | 0,05 | 2,86 | 4,18E-03 |
| MAP3K15 | 44,25 | 0,47 | 0,16 | 2,86 | 4,19E-03 |
| SGSM1 | 167,28 | 0,32 | 0,11 | 2,86 | 4,19E-03 |
| CCDC112 | 260,93 | -0,18 | 0,06 | -2,86 | 4,19E-03 |
| PTPRC | 72301,63 | -0,14 | 0,05 | -2,86 | 4,20E-03 |
| CRYBG1 | 5437,32 | -0,09 | 0,03 | -2,86 | 4,20E-03 |

| | | | | | |
|---|---|---|---|---|---|
| RABGGTB | 1725,68 | -0,14 | 0,05 | -2,86 | 4,22E-03 |
| LTBP2 | 226,27 | 0,29 | 0,10 | 2,86 | 4,22E-03 |
| TMEM30B | 47,49 | 0,31 | 0,11 | 2,86 | 4,23E-03 |
| TAF11 | 687,31 | -0,08 | 0,03 | -2,86 | 4,24E-03 |
| NARS2 | 606,58 | -0,12 | 0,04 | -2,86 | 4,25E-03 |
| NFKBID | 344,25 | 0,18 | 0,06 | 2,86 | 4,25E-03 |
| STK11 | 3087,72 | 0,18 | 0,06 | 2,86 | 4,27E-03 |
| ZNF853 | 64,45 | 0,26 | 0,09 | 2,86 | 4,27E-03 |
| MMGT1 | 784,85 | -0,10 | 0,03 | -2,86 | 4,27E-03 |
| KIF3A | 552,77 | -0,14 | 0,05 | -2,86 | 4,28E-03 |
| PPM1K | 3445,02 | -0,14 | 0,05 | -2,86 | 4,28E-03 |
| NSF | 803,70 | -0,09 | 0,03 | -2,86 | 4,29E-03 |
| TNFSF15 | 53,83 | 0,35 | 0,12 | 2,86 | 4,30E-03 |
| CHEK1 | 85,04 | 0,29 | 0,10 | 2,85 | 4,30E-03 |
| TANGO2 | 2374,59 | 0,22 | 0,08 | 2,85 | 4,31E-03 |
| SLC17A9 | 130,05 | 0,28 | 0,10 | 2,85 | 4,32E-03 |
| CPNE2 | 348,51 | 0,21 | 0,07 | 2,85 | 4,32E-03 |
| CEBPG | 1001,40 | -0,15 | 0,05 | -2,85 | 4,32E-03 |
| MRPL3 | 805,04 | -0,18 | 0,06 | -2,85 | 4,33E-03 |
| CHST14 | 72,45 | 0,23 | 0,08 | 2,85 | 4,33E-03 |
| BCL2L1 | 12468,22 | 0,33 | 0,12 | 2,85 | 4,34E-03 |
| ZNF354A | 297,53 | -0,22 | 0,08 | -2,85 | 4,36E-03 |
| TLR4 | 15494,06 | -0,20 | 0,07 | -2,85 | 4,36E-03 |
| POLA1 | 629,86 | -0,12 | 0,04 | -2,85 | 4,37E-03 |
| VEGFA | 409,33 | 0,20 | 0,07 | 2,85 | 4,37E-03 |
| MYSM1 | 3447,09 | -0,13 | 0,04 | -2,85 | 4,37E-03 |
| MAGIX | 77,10 | 0,18 | 0,06 | 2,85 | 4,38E-03 |
| MKS1 | 172,22 | 0,13 | 0,05 | 2,85 | 4,38E-03 |
| EFCAB5 | 72,34 | 0,32 | 0,11 | 2,85 | 4,38E-03 |
| RPL34 | 1910,33 | -0,43 | 0,15 | -2,85 | 4,38E-03 |
| STAB1 | 1410,90 | 0,28 | 0,10 | 2,85 | 4,39E-03 |
| P4HTM | 617,15 | 0,12 | 0,04 | 2,85 | 4,40E-03 |
| DDX23 | 3105,09 | -0,09 | 0,03 | -2,85 | 4,40E-03 |
| AP5S1 | 224,14 | 0,13 | 0,04 | 2,85 | 4,40E-03 |
| ATOSB | 1375,91 | 0,19 | 0,07 | 2,85 | 4,41E-03 |
| ADCK1 | 158,37 | 0,25 | 0,09 | 2,85 | 4,41E-03 |
| SLC30A9 | 1750,20 | -0,14 | 0,05 | -2,85 | 4,41E-03 |
| PEX2 | 1399,51 | -0,15 | 0,05 | -2,85 | 4,42E-03 |
| APOL4 | 47,13 | 0,52 | 0,18 | 2,85 | 4,43E-03 |
| JMY | 1336,96 | -0,11 | 0,04 | -2,85 | 4,43E-03 |
| PAXBP1 | 1983,34 | -0,13 | 0,05 | -2,85 | 4,44E-03 |
| VAV3 | 2765,59 | -0,12 | 0,04 | -2,85 | 4,44E-03 |
| DARS1 | 1315,11 | -0,15 | 0,05 | -2,84 | 4,46E-03 |
| GLE1 | 1349,62 | -0,07 | 0,02 | -2,84 | 4,46E-03 |
| RIMBP3 | 65,00 | 0,23 | 0,08 | 2,84 | 4,46E-03 |
| NACA | 9049,82 | -0,18 | 0,06 | -2,84 | 4,47E-03 |

| | | | | | |
|---|---|---|---|---|---|
| CEP83 | 549,33 | -0,13 | 0,05 | -2,84 | 4,47E-03 |
| FBXO28 | 1592,43 | -0,11 | 0,04 | -2,84 | 4,47E-03 |
| COA1 | 2635,36 | -0,14 | 0,05 | -2,84 | 4,48E-03 |
| LYRM2 | 654,40 | -0,18 | 0,06 | -2,84 | 4,48E-03 |
| SCAPER | 1608,32 | -0,13 | 0,05 | -2,84 | 4,49E-03 |
| MSRB3 | 201,33 | 0,21 | 0,07 | 2,84 | 4,49E-03 |
| NDRG2 | 149,67 | 0,21 | 0,07 | 2,84 | 4,49E-03 |
| CR1L | 347,50 | 0,31 | 0,11 | 2,84 | 4,49E-03 |
| CUEDC1 | 336,27 | 0,23 | 0,08 | 2,84 | 4,49E-03 |
| SHISA4 | 120,33 | 0,41 | 0,15 | 2,84 | 4,49E-03 |
| USP21 | 286,38 | 0,13 | 0,05 | 2,84 | 4,50E-03 |
| C1GALT1C1 | 463,15 | -0,19 | 0,07 | -2,84 | 4,50E-03 |
| SLC5A3 | 4519,06 | -0,15 | 0,05 | -2,84 | 4,51E-03 |
| TNFAIP8L1 | 290,85 | 0,14 | 0,05 | 2,84 | 4,51E-03 |
| TRIM65 | 394,38 | 0,15 | 0,05 | 2,84 | 4,52E-03 |
| ABL1 | 363,14 | 0,19 | 0,07 | 2,84 | 4,53E-03 |
| RAB2A | 2907,98 | -0,13 | 0,05 | -2,84 | 4,53E-03 |
| SHF | 86,71 | 0,22 | 0,08 | 2,84 | 4,55E-03 |
| SUMO3 | 360,78 | 0,11 | 0,04 | 2,84 | 4,56E-03 |
| OVGP1 | 78,24 | 0,22 | 0,08 | 2,84 | 4,56E-03 |
| LBR | 9515,32 | -0,16 | 0,06 | -2,84 | 4,57E-03 |
| SCAND1 | 470,72 | 0,11 | 0,04 | 2,84 | 4,57E-03 |
| ZSWIM9 | 143,42 | 0,21 | 0,08 | 2,84 | 4,58E-03 |
| CDK7 | 500,26 | -0,10 | 0,03 | -2,83 | 4,58E-03 |
| MIA2 | 2381,70 | -0,11 | 0,04 | -2,83 | 4,59E-03 |
| ZNF581 | 125,65 | 0,15 | 0,05 | 2,83 | 4,60E-03 |
| VTA1 | 942,80 | -0,15 | 0,05 | -2,83 | 4,61E-03 |
| SLC12A7 | 707,45 | 0,33 | 0,12 | 2,83 | 4,61E-03 |
| ADPGK | 5680,70 | -0,08 | 0,03 | -2,83 | 4,61E-03 |
| SLC30A5 | 1358,73 | -0,09 | 0,03 | -2,83 | 4,62E-03 |
| CLNS1A | 1346,21 | -0,11 | 0,04 | -2,83 | 4,63E-03 |
| ATAD1 | 1426,83 | -0,19 | 0,07 | -2,83 | 4,62E-03 |
| RPL23A | 12110,58 | -0,18 | 0,06 | -2,83 | 4,64E-03 |
| TRAF6 | 1416,46 | -0,08 | 0,03 | -2,83 | 4,65E-03 |
| UPP1 | 1177,98 | 0,13 | 0,05 | 2,83 | 4,65E-03 |
| FAM3C | 709,65 | -0,20 | 0,07 | -2,83 | 4,65E-03 |
| MAPKBP1 | 613,21 | 0,16 | 0,06 | 2,83 | 4,65E-03 |
| PIGA | 583,15 | -0,13 | 0,05 | -2,83 | 4,67E-03 |
| NKAPD1 | 758,08 | -0,15 | 0,05 | -2,83 | 4,68E-03 |
| TDRD9 | 250,43 | 0,33 | 0,12 | 2,83 | 4,68E-03 |
| PTPN23 | 322,05 | 0,24 | 0,08 | 2,83 | 4,70E-03 |
| MAPK7 | 180,57 | 0,21 | 0,07 | 2,83 | 4,70E-03 |
| ZNF816 | 313,10 | -0,22 | 0,08 | -2,83 | 4,72E-03 |
| KIAA1191 | 1074,92 | 0,10 | 0,03 | 2,83 | 4,72E-03 |
| TES | 3388,49 | -0,11 | 0,04 | -2,83 | 4,73E-03 |
| HSPH1 | 1091,17 | -0,17 | 0,06 | -2,83 | 4,73E-03 |

| | | | | | |
|---|---|---|---|---|---|
| MAP1A | 121,95 | 0,34 | 0,12 | 2,82 | 4,75E-03 |
| CARD9 | 716,54 | 0,16 | 0,06 | 2,82 | 4,76E-03 |
| DYRK1B | 275,91 | 0,18 | 0,06 | 2,82 | 4,76E-03 |
| RGCC | 941,49 | 0,19 | 0,07 | 2,82 | 4,76E-03 |
| PEDS1 | 243,22 | 0,17 | 0,06 | 2,82 | 4,76E-03 |
| ZNHIT6 | 729,42 | -0,14 | 0,05 | -2,82 | 4,76E-03 |
| CDC14B | 370,69 | 0,18 | 0,06 | 2,82 | 4,77E-03 |
| KIF1C | 495,72 | 0,20 | 0,07 | 2,82 | 4,77E-03 |
| KCNK13 | 37,77 | 0,30 | 0,11 | 2,82 | 4,77E-03 |
| KBTBD8 | 348,09 | -0,27 | 0,10 | -2,82 | 4,78E-03 |
| SPR | 34,29 | 0,31 | 0,11 | 2,82 | 4,79E-03 |
| ERLEC1 | 670,23 | -0,15 | 0,05 | -2,82 | 4,79E-03 |
| SENP8 | 33,98 | 0,30 | 0,11 | 2,82 | 4,79E-03 |
| SERPINH1 | 190,69 | 0,17 | 0,06 | 2,82 | 4,80E-03 |
| DUS1L | 767,07 | 0,12 | 0,04 | 2,82 | 4,83E-03 |
| CCAR2 | 657,87 | 0,19 | 0,07 | 2,82 | 4,84E-03 |
| CLEC11A | 59,92 | 0,35 | 0,12 | 2,82 | 4,85E-03 |
| ZBTB3 | 171,48 | 0,13 | 0,05 | 2,82 | 4,86E-03 |
| MED24 | 387,85 | 0,17 | 0,06 | 2,82 | 4,86E-03 |
| PTGR2 | 91,16 | 0,21 | 0,07 | 2,82 | 4,88E-03 |
| C20orf96 | 29,13 | 0,33 | 0,12 | 2,81 | 4,88E-03 |
| SLC6A8 | 909,73 | 0,43 | 0,15 | 2,81 | 4,90E-03 |
| ZFYVE28 | 375,14 | 0,26 | 0,09 | 2,81 | 4,90E-03 |
| RBAK | 822,54 | -0,23 | 0,08 | -2,81 | 4,90E-03 |
| EID1 | 1900,13 | -0,21 | 0,08 | -2,81 | 4,90E-03 |
| PMS1 | 779,63 | -0,15 | 0,05 | -2,81 | 4,90E-03 |
| IL31RA | 58,07 | 0,42 | 0,15 | 2,81 | 4,91E-03 |
| PRKX | 1946,30 | -0,08 | 0,03 | -2,81 | 4,91E-03 |
| TNNT3 | 71,68 | 0,33 | 0,12 | 2,81 | 4,91E-03 |
| SRSF5 | 11183,83 | -0,11 | 0,04 | -2,81 | 4,93E-03 |
| C7orf50 | 419,83 | 0,14 | 0,05 | 2,81 | 4,94E-03 |
| HRK | 68,79 | 0,53 | 0,19 | 2,81 | 4,95E-03 |
| CEP72 | 223,48 | 0,19 | 0,07 | 2,81 | 4,95E-03 |
| SRR | 60,46 | 0,18 | 0,07 | 2,81 | 4,96E-03 |
| KIAA1671 | 170,26 | 0,44 | 0,16 | 2,81 | 4,96E-03 |
| RAPGEF5 | 70,46 | 0,48 | 0,17 | 2,81 | 4,96E-03 |
| CEP104 | 868,81 | 0,14 | 0,05 | 2,81 | 4,96E-03 |
| HGD | 104,78 | 0,18 | 0,06 | 2,81 | 4,96E-03 |
| ZNF316 | 1626,79 | 0,15 | 0,05 | 2,81 | 4,97E-03 |
| MYLK4 | 94,89 | 0,22 | 0,08 | 2,81 | 4,98E-03 |
| RUNDC3A | 640,02 | 0,38 | 0,13 | 2,81 | 5,00E-03 |
| DHRS11 | 73,23 | 0,25 | 0,09 | 2,81 | 5,00E-03 |
| NAPA | 1040,76 | 0,22 | 0,08 | 2,81 | 5,02E-03 |
| TTC3 | 5199,78 | -0,12 | 0,04 | -2,81 | 5,02E-03 |
| AP1B1 | 559,31 | 0,22 | 0,08 | 2,81 | 5,02E-03 |
| MARCHF5 | 761,55 | -0,10 | 0,04 | -2,81 | 5,02E-03 |

| | | | | | |
|---|---|---|---|---|---|
| SOX13 | 106,65 | 0,38 | 0,14 | 2,80 | 5,04E-03 |
| RAB18 | 3525,67 | -0,16 | 0,06 | -2,80 | 5,05E-03 |
| SKIC2 | 894,44 | 0,17 | 0,06 | 2,80 | 5,05E-03 |
| HSD17B11 | 5849,27 | -0,16 | 0,06 | -2,80 | 5,06E-03 |
| CD96 | 2842,11 | -0,15 | 0,05 | -2,80 | 5,06E-03 |
| COMMD8 | 312,92 | -0,33 | 0,12 | -2,80 | 5,07E-03 |
| UBA5 | 844,30 | -0,17 | 0,06 | -2,80 | 5,07E-03 |
| ZNF699 | 475,46 | -0,12 | 0,04 | -2,80 | 5,08E-03 |
| GSPT2 | 129,62 | -0,16 | 0,06 | -2,80 | 5,08E-03 |
| WDR3 | 890,14 | -0,12 | 0,04 | -2,80 | 5,08E-03 |
| COL18A1 | 1685,36 | 0,30 | 0,11 | 2,80 | 5,08E-03 |
| WTAP | 4709,83 | -0,13 | 0,04 | -2,80 | 5,08E-03 |
| ZSCAN16 | 300,10 | -0,13 | 0,05 | -2,80 | 5,08E-03 |
| TIMM50 | 352,75 | 0,09 | 0,03 | 2,80 | 5,10E-03 |
| LRGUK | 44,48 | 0,45 | 0,16 | 2,80 | 5,10E-03 |
| NAA20 | 476,64 | -0,10 | 0,04 | -2,80 | 5,11E-03 |
| ARRDC3 | 10155,55 | -0,22 | 0,08 | -2,80 | 5,11E-03 |
| RIPK3 | 236,68 | 0,20 | 0,07 | 2,80 | 5,12E-03 |
| UBXN7 | 4115,03 | -0,09 | 0,03 | -2,80 | 5,12E-03 |
| PHF6 | 1047,18 | -0,16 | 0,06 | -2,80 | 5,13E-03 |
| OMA1 | 1283,71 | -0,13 | 0,05 | -2,80 | 5,13E-03 |
| FAHD2A | 299,14 | 0,12 | 0,04 | 2,80 | 5,13E-03 |
| TAMALIN | 101,77 | 0,22 | 0,08 | 2,80 | 5,13E-03 |
| CCDC89 | 56,98 | -0,26 | 0,09 | -2,80 | 5,13E-03 |
| ABCA10 | 161,37 | 0,22 | 0,08 | 2,80 | 5,14E-03 |
| KIF4A | 41,57 | 0,40 | 0,14 | 2,80 | 5,14E-03 |
| PHF19 | 732,83 | 0,14 | 0,05 | 2,80 | 5,14E-03 |
| SLBP | 1151,64 | -0,11 | 0,04 | -2,80 | 5,14E-03 |
| THAP12 | 1731,94 | -0,18 | 0,06 | -2,80 | 5,14E-03 |
| ZNF337 | 1097,30 | -0,10 | 0,04 | -2,80 | 5,15E-03 |
| FRMD8 | 860,54 | 0,18 | 0,06 | 2,80 | 5,15E-03 |
| SLC2A6 | 639,41 | 0,17 | 0,06 | 2,80 | 5,15E-03 |
| PPARGC1B | 457,51 | 0,20 | 0,07 | 2,80 | 5,16E-03 |
| KLC3 | 352,07 | 0,38 | 0,14 | 2,80 | 5,17E-03 |
| EDEM3 | 4704,04 | -0,13 | 0,05 | -2,80 | 5,18E-03 |
| TMEM232 | 46,98 | 0,33 | 0,12 | 2,80 | 5,18E-03 |
| THRAP3 | 6022,71 | -0,10 | 0,04 | -2,80 | 5,18E-03 |
| KDM1A | 1227,15 | -0,08 | 0,03 | -2,80 | 5,19E-03 |
| ACSM3 | 362,61 | 0,23 | 0,08 | 2,79 | 5,19E-03 |
| NDUFAF7 | 761,34 | -0,11 | 0,04 | -2,79 | 5,20E-03 |
| ADAM10 | 9290,73 | -0,13 | 0,05 | -2,79 | 5,20E-03 |
| CYP4F12 | 195,99 | 0,27 | 0,10 | 2,79 | 5,21E-03 |
| SAMSN1 | 1327,31 | -0,17 | 0,06 | -2,79 | 5,24E-03 |
| TAF5 | 284,07 | -0,08 | 0,03 | -2,79 | 5,24E-03 |
| CUL9 | 1057,29 | 0,18 | 0,07 | 2,79 | 5,26E-03 |
| CUL7 | 254,85 | 0,20 | 0,07 | 2,79 | 5,27E-03 |

| | | | | | |
|---|---|---|---|---|---|
| BTNL9 | 45,93 | 0,57 | 0,20 | 2,79 | 5,27E-03 |
| MDH1 | 1131,68 | -0,14 | 0,05 | -2,79 | 5,27E-03 |
| EVA1B | 165,39 | 0,17 | 0,06 | 2,79 | 5,27E-03 |
| ZNF283 | 530,13 | -0,13 | 0,05 | -2,79 | 5,28E-03 |
| SATB2 | 52,00 | 0,47 | 0,17 | 2,79 | 5,28E-03 |
| ZHX1 | 831,50 | -0,18 | 0,06 | -2,79 | 5,29E-03 |
| LDLRAD3 | 91,05 | 0,27 | 0,10 | 2,79 | 5,29E-03 |
| BRD3OS | 119,69 | 0,18 | 0,06 | 2,79 | 5,29E-03 |
| ZNF780B | 2315,03 | -0,18 | 0,06 | -2,79 | 5,29E-03 |
| ACAD10 | 752,73 | 0,13 | 0,05 | 2,79 | 5,29E-03 |
| SLC16A14 | 60,67 | 0,40 | 0,14 | 2,79 | 5,29E-03 |
| RIBC1 | 26,17 | 0,28 | 0,10 | 2,79 | 5,30E-03 |
| UBR3 | 3783,97 | -0,11 | 0,04 | -2,79 | 5,30E-03 |
| ZNF780A | 2010,08 | -0,17 | 0,06 | -2,79 | 5,32E-03 |
| EED | 1120,78 | -0,15 | 0,05 | -2,79 | 5,32E-03 |
| HNRNPF | 4781,86 | -0,10 | 0,04 | -2,79 | 5,32E-03 |
| SORD | 328,22 | 0,18 | 0,07 | 2,79 | 5,32E-03 |
| AKR1E2 | 175,95 | 0,31 | 0,11 | 2,79 | 5,32E-03 |
| ZNF10 | 269,28 | -0,16 | 0,06 | -2,79 | 5,33E-03 |
| RHOBTB1 | 212,81 | 0,28 | 0,10 | 2,79 | 5,33E-03 |
| IRF5 | 598,00 | 0,19 | 0,07 | 2,79 | 5,34E-03 |
| CCDC77 | 264,99 | -0,10 | 0,03 | -2,79 | 5,35E-03 |
| SLC25A53 | 579,56 | -0,11 | 0,04 | -2,79 | 5,35E-03 |
| WRN | 871,47 | -0,12 | 0,04 | -2,78 | 5,36E-03 |
| XRN2 | 4476,85 | -0,11 | 0,04 | -2,78 | 5,36E-03 |
| CASP8 | 9565,51 | -0,11 | 0,04 | -2,78 | 5,36E-03 |
| MRPL50 | 464,15 | -0,26 | 0,09 | -2,78 | 5,37E-03 |
| MAPRE1 | 3535,89 | -0,10 | 0,04 | -2,78 | 5,37E-03 |
| TBKBP1 | 283,67 | 0,27 | 0,10 | 2,78 | 5,36E-03 |
| BAIAP3 | 250,18 | 0,33 | 0,12 | 2,78 | 5,37E-03 |
| FAT1 | 84,22 | 0,52 | 0,19 | 2,78 | 5,37E-03 |
| PRSS33 | 103,05 | 0,45 | 0,16 | 2,78 | 5,38E-03 |
| LRRC42 | 116,42 | 0,14 | 0,05 | 2,78 | 5,39E-03 |
| AMD1 | 5424,99 | -0,13 | 0,05 | -2,78 | 5,39E-03 |
| PREB | 437,92 | 0,11 | 0,04 | 2,78 | 5,40E-03 |
| RPP38 | 338,56 | -0,09 | 0,03 | -2,78 | 5,40E-03 |
| CMC1 | 969,62 | -0,20 | 0,07 | -2,78 | 5,41E-03 |
| NUDT21 | 1864,37 | -0,17 | 0,06 | -2,78 | 5,41E-03 |
| RIT1 | 2850,23 | -0,16 | 0,06 | -2,78 | 5,41E-03 |
| ASGR2 | 112,04 | 0,29 | 0,11 | 2,78 | 5,41E-03 |
| PRNP | 1288,43 | -0,12 | 0,04 | -2,78 | 5,41E-03 |
| GLO1 | 652,46 | -0,16 | 0,06 | -2,78 | 5,42E-03 |
| ATXN10 | 880,41 | -0,09 | 0,03 | -2,78 | 5,42E-03 |
| GPR157 | 182,43 | 0,18 | 0,06 | 2,78 | 5,42E-03 |
| ANKRD27 | 1271,34 | 0,06 | 0,02 | 2,78 | 5,43E-03 |
| KAZN | 247,16 | 0,41 | 0,15 | 2,78 | 5,43E-03 |

| | | | | | |
|---|---|---|---|---|---|
| BTAF1 | 5706,32 | -0,11 | 0,04 | -2,78 | 5,43E-03 |
| MTDH | 2872,18 | -0,10 | 0,04 | -2,78 | 5,43E-03 |
| SUCLG1 | 1133,64 | -0,10 | 0,04 | -2,78 | 5,44E-03 |
| GPR35 | 179,06 | 0,22 | 0,08 | 2,78 | 5,44E-03 |
| QRICH2 | 143,40 | 0,18 | 0,06 | 2,78 | 5,45E-03 |
| CLK1 | 7478,55 | -0,15 | 0,06 | -2,78 | 5,47E-03 |
| KCNMB4 | 59,34 | 0,27 | 0,10 | 2,78 | 5,47E-03 |
| LIPC | 59,12 | 0,36 | 0,13 | 2,78 | 5,48E-03 |
| ANAPC2 | 660,22 | 0,18 | 0,06 | 2,78 | 5,49E-03 |
| SPX | 87,48 | 0,33 | 0,12 | 2,78 | 5,49E-03 |
| KIDINS220 | 7651,87 | -0,11 | 0,04 | -2,78 | 5,50E-03 |
| ZC3H12A | 531,66 | 0,12 | 0,04 | 2,78 | 5,51E-03 |
| HIF1A | 4686,89 | -0,14 | 0,05 | -2,78 | 5,51E-03 |
| LRRC14 | 386,29 | 0,20 | 0,07 | 2,78 | 5,51E-03 |
| BBX | 4347,36 | -0,10 | 0,04 | -2,77 | 5,52E-03 |
| VARS1 | 294,50 | 0,21 | 0,08 | 2,77 | 5,52E-03 |
| SLC25A45 | 396,68 | 0,24 | 0,09 | 2,77 | 5,53E-03 |
| MAK16 | 532,74 | -0,13 | 0,05 | -2,77 | 5,54E-03 |
| PYCR2 | 432,01 | 0,14 | 0,05 | 2,77 | 5,54E-03 |
| HAAO | 49,87 | 0,27 | 0,10 | 2,77 | 5,55E-03 |
| PGAP2 | 172,51 | 0,15 | 0,05 | 2,77 | 5,55E-03 |
| TUBB2A | 721,55 | 0,75 | 0,27 | 2,77 | 5,57E-03 |
| CHURC1 | 2062,66 | -0,14 | 0,05 | -2,77 | 5,57E-03 |
| CNOT3 | 279,00 | 0,30 | 0,11 | 2,77 | 5,58E-03 |
| GLUD1 | 3176,87 | -0,10 | 0,04 | -2,77 | 5,58E-03 |
| LRRC61 | 127,01 | 0,16 | 0,06 | 2,77 | 5,59E-03 |
| EPAS1 | 143,53 | 0,28 | 0,10 | 2,77 | 5,59E-03 |
| ZBTB7A | 636,85 | 0,23 | 0,08 | 2,77 | 5,60E-03 |
| ZNF41 | 542,38 | -0,07 | 0,03 | -2,77 | 5,60E-03 |
| MAGT1 | 2435,45 | -0,12 | 0,04 | -2,77 | 5,61E-03 |
| TATDN3 | 563,49 | -0,10 | 0,04 | -2,77 | 5,63E-03 |
| RBM25 | 7494,61 | -0,12 | 0,04 | -2,77 | 5,63E-03 |
| CDC37L1 | 618,43 | -0,19 | 0,07 | -2,77 | 5,62E-03 |
| MEN1 | 172,54 | 0,21 | 0,07 | 2,77 | 5,63E-03 |
| FGFBP2 | 821,30 | -0,28 | 0,10 | -2,77 | 5,63E-03 |
| CDKN3 | 29,24 | 0,35 | 0,13 | 2,77 | 5,63E-03 |
| PARD3 | 113,31 | 0,33 | 0,12 | 2,77 | 5,63E-03 |
| ABCC10 | 921,11 | 0,16 | 0,06 | 2,77 | 5,63E-03 |
| TSSK6 | 90,06 | 0,13 | 0,05 | 2,77 | 5,64E-03 |
| MED16 | 382,94 | 0,26 | 0,09 | 2,77 | 5,64E-03 |
| LONP2 | 3782,68 | -0,09 | 0,03 | -2,77 | 5,65E-03 |
| SNAPC3 | 2214,26 | -0,10 | 0,04 | -2,77 | 5,66E-03 |
| CMPK1 | 2371,97 | -0,18 | 0,06 | -2,77 | 5,66E-03 |
| SRP14 | 1578,72 | -0,24 | 0,09 | -2,77 | 5,66E-03 |
| SFXN2 | 252,93 | 0,19 | 0,07 | 2,77 | 5,66E-03 |
| COQ4 | 247,78 | 0,18 | 0,07 | 2,77 | 5,66E-03 |

| | | | | | |
|---|---|---|---|---|---|
| HBS1L | 2017,62 | -0,15 | 0,05 | -2,77 | 5,67E-03 |
| ZBTB17 | 612,89 | 0,08 | 0,03 | 2,77 | 5,67E-03 |
| PRIM2 | 604,36 | -0,12 | 0,04 | -2,77 | 5,67E-03 |
| PPP1R12A | 8625,28 | -0,15 | 0,05 | -2,77 | 5,68E-03 |
| KIF3C | 181,75 | 0,22 | 0,08 | 2,77 | 5,68E-03 |
| UTP14C | 183,53 | -0,15 | 0,05 | -2,77 | 5,68E-03 |
| KMT2B | 600,41 | 0,37 | 0,14 | 2,77 | 5,68E-03 |
| YWHAZ | 16958,51 | -0,09 | 0,03 | -2,77 | 5,69E-03 |
| CEACAM6 | 28,74 | 0,47 | 0,17 | 2,77 | 5,69E-03 |
| SEC14L5 | 83,47 | 0,47 | 0,17 | 2,76 | 5,70E-03 |
| ATG4C | 631,93 | -0,18 | 0,06 | -2,76 | 5,70E-03 |
| BLOC1S3 | 207,35 | 0,19 | 0,07 | 2,76 | 5,72E-03 |
| TPRKB | 307,64 | -0,13 | 0,05 | -2,76 | 5,73E-03 |
| DNAI3 | 40,20 | 0,43 | 0,15 | 2,76 | 5,73E-03 |
| NMD3 | 782,92 | -0,21 | 0,08 | -2,76 | 5,73E-03 |
| CCDC90B | 886,42 | -0,12 | 0,04 | -2,76 | 5,74E-03 |
| DOP1A | 2660,82 | -0,11 | 0,04 | -2,76 | 5,74E-03 |
| GK5 | 1916,24 | -0,14 | 0,05 | -2,76 | 5,76E-03 |
| POP4 | 616,11 | -0,07 | 0,03 | -2,76 | 5,79E-03 |
| TESMIN | 205,05 | 0,16 | 0,06 | 2,76 | 5,79E-03 |
| PIEZO1 | 1540,55 | 0,29 | 0,11 | 2,76 | 5,79E-03 |
| MLH3 | 1242,21 | -0,12 | 0,05 | -2,76 | 5,80E-03 |
| OSBPL11 | 2470,94 | -0,12 | 0,04 | -2,76 | 5,80E-03 |
| SIAH2 | 3357,40 | 0,29 | 0,10 | 2,76 | 5,80E-03 |
| NABP2 | 121,04 | 0,11 | 0,04 | 2,76 | 5,80E-03 |
| ZNF24 | 5368,02 | -0,15 | 0,05 | -2,76 | 5,81E-03 |
| RAP1A | 3809,38 | -0,12 | 0,04 | -2,76 | 5,81E-03 |
| UBR7 | 507,63 | -0,07 | 0,03 | -2,76 | 5,81E-03 |
| ZNF791 | 1422,58 | -0,13 | 0,05 | -2,76 | 5,81E-03 |
| TNS1 | 3841,09 | 0,38 | 0,14 | 2,76 | 5,83E-03 |
| ZNF124 | 395,28 | -0,14 | 0,05 | -2,76 | 5,83E-03 |
| LRRC37A | 126,08 | 0,25 | 0,09 | 2,76 | 5,83E-03 |
| DNHD1 | 943,78 | 0,22 | 0,08 | 2,76 | 5,85E-03 |
| MCTP2 | 10377,33 | -0,17 | 0,06 | -2,76 | 5,85E-03 |
| RUSC2 | 121,60 | 0,28 | 0,10 | 2,76 | 5,85E-03 |
| INCENP | 289,35 | 0,15 | 0,05 | 2,76 | 5,86E-03 |
| HACL1 | 821,89 | -0,11 | 0,04 | -2,75 | 5,87E-03 |
| ARHGEF10L | 625,61 | 0,20 | 0,07 | 2,75 | 5,88E-03 |
| PPM1H | 179,84 | 0,24 | 0,09 | 2,75 | 5,89E-03 |
| ZMYM2 | 5522,27 | -0,12 | 0,04 | -2,75 | 5,89E-03 |
| MAN1C1 | 374,38 | 0,25 | 0,09 | 2,75 | 5,89E-03 |
| RAB27A | 3359,15 | -0,12 | 0,04 | -2,75 | 5,89E-03 |
| FNTA | 2599,71 | -0,11 | 0,04 | -2,75 | 5,89E-03 |
| RILP | 1313,12 | 0,29 | 0,10 | 2,75 | 5,89E-03 |
| ZER1 | 1529,23 | 0,28 | 0,10 | 2,75 | 5,89E-03 |
| PKN2 | 5205,84 | -0,17 | 0,06 | -2,75 | 5,90E-03 |

| | | | | | |
|---|---|---|---|---|---|
| VNN3P | 3696,65 | -0,24 | 0,09 | -2,75 | 5,90E-03 |
| DENND1C | 469,91 | 0,26 | 0,09 | 2,75 | 5,90E-03 |
| THAP4 | 181,20 | 0,15 | 0,06 | 2,75 | 5,90E-03 |
| EID3 | 245,14 | -0,11 | 0,04 | -2,75 | 5,91E-03 |
| ZNF671 | 974,76 | -0,11 | 0,04 | -2,75 | 5,91E-03 |
| NDUFB3 | 256,28 | -0,32 | 0,12 | -2,75 | 5,91E-03 |
| FBLN2 | 59,50 | 0,46 | 0,17 | 2,75 | 5,92E-03 |
| STK38 | 6296,44 | -0,09 | 0,03 | -2,75 | 5,94E-03 |
| CLASP2 | 3095,39 | -0,09 | 0,03 | -2,75 | 5,94E-03 |
| RPS6KB2 | 839,46 | 0,09 | 0,03 | 2,75 | 5,95E-03 |
| GBE1 | 1104,40 | -0,14 | 0,05 | -2,75 | 5,95E-03 |
| ESCO2 | 50,42 | 0,38 | 0,14 | 2,75 | 5,95E-03 |
| AP1G2 | 1232,79 | 0,17 | 0,06 | 2,75 | 5,96E-03 |
| FEZ1 | 111,24 | 0,34 | 0,12 | 2,75 | 5,96E-03 |
| BUD13 | 496,22 | -0,06 | 0,02 | -2,75 | 5,96E-03 |
| ATP6V1D | 1000,19 | -0,14 | 0,05 | -2,75 | 5,98E-03 |
| FITM1 | 26,63 | 0,19 | 0,07 | 2,75 | 6,00E-03 |
| MFAP3 | 1281,14 | -0,10 | 0,04 | -2,75 | 6,01E-03 |
| HPCAL1 | 842,90 | 0,21 | 0,08 | 2,75 | 6,01E-03 |
| GPAT2 | 46,16 | 0,47 | 0,17 | 2,75 | 6,01E-03 |
| TXNIP | 77703,64 | -0,13 | 0,05 | -2,75 | 6,02E-03 |
| DHCR7 | 91,17 | 0,25 | 0,09 | 2,75 | 6,02E-03 |
| SELENBP1 | 3560,91 | 0,38 | 0,14 | 2,75 | 6,02E-03 |
| DHX40 | 1473,66 | -0,14 | 0,05 | -2,75 | 6,02E-03 |
| ALDH9A1 | 1406,93 | -0,10 | 0,04 | -2,75 | 6,03E-03 |
| CLP1 | 414,93 | -0,10 | 0,04 | -2,75 | 6,03E-03 |
| HMGN1 | 2875,22 | -0,13 | 0,05 | -2,75 | 6,05E-03 |
| MRPL51 | 447,05 | -0,20 | 0,07 | -2,75 | 6,05E-03 |
| ANXA6 | 7347,05 | -0,11 | 0,04 | -2,75 | 6,05E-03 |
| TASP1 | 529,32 | -0,12 | 0,04 | -2,74 | 6,05E-03 |
| ZNF664 | 1192,43 | -0,09 | 0,03 | -2,74 | 6,06E-03 |
| ZNF692 | 464,06 | 0,12 | 0,04 | 2,74 | 6,06E-03 |
| TAB1 | 349,66 | 0,17 | 0,06 | 2,74 | 6,06E-03 |
| SYMPK | 804,85 | 0,24 | 0,09 | 2,74 | 6,07E-03 |
| RSRC1 | 986,41 | -0,13 | 0,05 | -2,74 | 6,07E-03 |
| CAAP1 | 660,88 | -0,13 | 0,05 | -2,74 | 6,07E-03 |
| SF3B1 | 20851,39 | -0,14 | 0,05 | -2,74 | 6,07E-03 |
| TOMM20 | 1891,06 | -0,15 | 0,06 | -2,74 | 6,08E-03 |
| SEC31B | 305,26 | 0,27 | 0,10 | 2,74 | 6,08E-03 |
| TMEM138 | 610,10 | 0,09 | 0,03 | 2,74 | 6,08E-03 |
| CHIT1 | 171,57 | 0,21 | 0,08 | 2,74 | 6,09E-03 |
| BICDL1 | 311,31 | 0,20 | 0,07 | 2,74 | 6,10E-03 |
| PRXL2A | 116,50 | 0,20 | 0,07 | 2,74 | 6,10E-03 |
| FRYL | 8050,62 | -0,09 | 0,03 | -2,74 | 6,10E-03 |
| ITFG1 | 1647,77 | -0,09 | 0,03 | -2,74 | 6,11E-03 |
| SEC23A | 2217,63 | -0,09 | 0,03 | -2,74 | 6,11E-03 |

| | | | | | |
|---|---|---|---|---|---|
| SLC39A9 | 1834,76 | -0,06 | 0,02 | -2,74 | 6,11E-03 |
| ZNF320 | 112,01 | 0,17 | 0,06 | 2,74 | 6,13E-03 |
| PGM3 | 733,50 | -0,13 | 0,05 | -2,74 | 6,13E-03 |
| EAF1 | 1333,42 | -0,08 | 0,03 | -2,74 | 6,15E-03 |
| HCFC1 | 645,22 | 0,26 | 0,09 | 2,74 | 6,15E-03 |
| POP7 | 189,71 | 0,13 | 0,05 | 2,74 | 6,16E-03 |
| PCIF1 | 967,09 | 0,14 | 0,05 | 2,74 | 6,17E-03 |
| DDB2 | 353,16 | 0,18 | 0,07 | 2,74 | 6,17E-03 |
| BPI | 116,52 | 0,38 | 0,14 | 2,74 | 6,19E-03 |
| LMNB1 | 2025,24 | -0,22 | 0,08 | -2,74 | 6,20E-03 |
| TPP2 | 4522,97 | -0,12 | 0,04 | -2,74 | 6,22E-03 |
| MED6 | 976,07 | -0,10 | 0,04 | -2,74 | 6,22E-03 |
| ZBTB21 | 696,38 | -0,10 | 0,04 | -2,74 | 6,23E-03 |
| MMP24OS | 539,10 | 0,17 | 0,06 | 2,74 | 6,23E-03 |
| PLCL2 | 5131,66 | -0,11 | 0,04 | -2,73 | 6,24E-03 |
| PFKFB1 | 29,60 | 0,30 | 0,11 | 2,73 | 6,25E-03 |
| STK17A | 4335,55 | -0,13 | 0,05 | -2,73 | 6,25E-03 |
| PANX2 | 129,67 | 0,37 | 0,14 | 2,73 | 6,26E-03 |
| CDK10 | 510,09 | 0,19 | 0,07 | 2,73 | 6,26E-03 |
| YBX3 | 13704,60 | 0,30 | 0,11 | 2,73 | 6,27E-03 |
| PDCD10 | 1829,90 | -0,20 | 0,07 | -2,73 | 6,27E-03 |
| GSTA4 | 31,83 | 0,28 | 0,10 | 2,73 | 6,27E-03 |
| CAPN10 | 1040,98 | 0,11 | 0,04 | 2,73 | 6,28E-03 |
| CDC26 | 342,50 | -0,15 | 0,05 | -2,73 | 6,28E-03 |
| ZW10 | 457,14 | -0,09 | 0,03 | -2,73 | 6,29E-03 |
| SIGLEC10 | 2077,45 | 0,21 | 0,08 | 2,73 | 6,29E-03 |
| MAP2K4 | 2859,60 | -0,14 | 0,05 | -2,73 | 6,29E-03 |
| MYBPC3 | 213,08 | 0,28 | 0,10 | 2,73 | 6,30E-03 |
| UBE3A | 3376,56 | -0,12 | 0,04 | -2,73 | 6,31E-03 |
| GMPR | 2131,03 | 0,35 | 0,13 | 2,73 | 6,31E-03 |
| FADS1 | 347,14 | 0,19 | 0,07 | 2,73 | 6,31E-03 |
| AMACR | 149,53 | -0,17 | 0,06 | -2,73 | 6,31E-03 |
| CCDC24 | 56,91 | 0,30 | 0,11 | 2,73 | 6,33E-03 |
| ZNF441 | 558,43 | -0,28 | 0,10 | -2,73 | 6,33E-03 |
| LY6G6F | 26,80 | 0,34 | 0,12 | 2,73 | 6,34E-03 |
| VDAC3 | 1432,44 | -0,12 | 0,05 | -2,73 | 6,34E-03 |
| ANKRD34A | 50,14 | 0,20 | 0,07 | 2,73 | 6,35E-03 |
| HDAC4 | 3076,94 | 0,17 | 0,06 | 2,73 | 6,35E-03 |
| SUSD4 | 131,54 | 0,29 | 0,10 | 2,73 | 6,36E-03 |
| HTRA4 | 61,17 | 0,21 | 0,08 | 2,73 | 6,36E-03 |
| MCF2L2 | 303,49 | 0,25 | 0,09 | 2,73 | 6,36E-03 |
| SAMD9 | 4917,97 | -0,36 | 0,13 | -2,73 | 6,37E-03 |
| TNFAIP8 | 5201,40 | -0,11 | 0,04 | -2,73 | 6,37E-03 |
| ZNF222 | 114,37 | -0,16 | 0,06 | -2,73 | 6,37E-03 |
| LINGO3 | 50,23 | 0,21 | 0,08 | 2,73 | 6,37E-03 |
| CACNA2D4 | 231,93 | 0,19 | 0,07 | 2,73 | 6,39E-03 |

| | | | | | |
|---|---|---|---|---|---|
| LRRC40 | 545,62 | -0,18 | 0,07 | -2,73 | 6,40E-03 |
| ABCA2 | 826,33 | 0,30 | 0,11 | 2,73 | 6,41E-03 |
| USP7 | 4874,29 | -0,07 | 0,03 | -2,73 | 6,42E-03 |
| MNT | 730,12 | 0,22 | 0,08 | 2,73 | 6,43E-03 |
| FAM76A | 460,56 | -0,08 | 0,03 | -2,73 | 6,42E-03 |
| CDNF | 36,45 | 0,22 | 0,08 | 2,72 | 6,43E-03 |
| ZNF669 | 277,23 | -0,15 | 0,06 | -2,72 | 6,43E-03 |
| TRAF3IP2 | 147,04 | 0,17 | 0,06 | 2,72 | 6,44E-03 |
| ITGB1 | 7660,99 | -0,16 | 0,06 | -2,72 | 6,44E-03 |
| CHL1 | 69,14 | 0,43 | 0,16 | 2,72 | 6,45E-03 |
| SHCBP1 | 62,71 | 0,29 | 0,11 | 2,72 | 6,46E-03 |
| STXBP3 | 4356,87 | -0,16 | 0,06 | -2,72 | 6,46E-03 |
| U2SURP | 4319,56 | -0,15 | 0,06 | -2,72 | 6,48E-03 |
| RCHY1 | 1295,53 | -0,15 | 0,05 | -2,72 | 6,48E-03 |
| SNX4 | 488,01 | -0,21 | 0,08 | -2,72 | 6,48E-03 |
| OPLAH | 129,72 | 0,32 | 0,12 | 2,72 | 6,49E-03 |
| SLC38A7 | 141,53 | 0,21 | 0,08 | 2,72 | 6,49E-03 |
| ENSG00000256591 | 415,39 | 0,16 | 0,06 | 2,72 | 6,51E-03 |
| EXOC2 | 1950,69 | -0,10 | 0,04 | -2,72 | 6,51E-03 |
| SSBP4 | 388,64 | 0,20 | 0,07 | 2,72 | 6,52E-03 |
| SYP | 41,73 | 0,32 | 0,12 | 2,72 | 6,52E-03 |
| COQ10B | 1502,53 | -0,13 | 0,05 | -2,72 | 6,52E-03 |
| LRRC8A | 610,16 | 0,15 | 0,06 | 2,72 | 6,52E-03 |
| MED10 | 543,79 | -0,08 | 0,03 | -2,72 | 6,53E-03 |
| SPRYD3 | 385,57 | 0,15 | 0,06 | 2,72 | 6,54E-03 |
| CXCL8 | 590,61 | -0,34 | 0,12 | -2,72 | 6,54E-03 |
| RAB4B | 157,37 | 0,17 | 0,06 | 2,72 | 6,56E-03 |
| NRROS | 190,95 | 0,28 | 0,10 | 2,72 | 6,57E-03 |
| PJA1 | 362,23 | -0,13 | 0,05 | -2,72 | 6,57E-03 |
| CDC6 | 49,52 | 0,28 | 0,10 | 2,72 | 6,57E-03 |
| DVL3 | 367,67 | 0,23 | 0,09 | 2,72 | 6,57E-03 |
| PLEKHM2 | 581,13 | 0,21 | 0,08 | 2,72 | 6,58E-03 |
| REDIC1 | 34,87 | 0,34 | 0,12 | 2,72 | 6,58E-03 |
| DDX5 | 26984,63 | -0,10 | 0,04 | -2,72 | 6,59E-03 |
| LCP1 | 46157,17 | -0,13 | 0,05 | -2,72 | 6,59E-03 |
| ALKBH1 | 377,82 | -0,06 | 0,02 | -2,72 | 6,59E-03 |
| MKI67 | 297,58 | 0,26 | 0,10 | 2,72 | 6,60E-03 |
| ZNF667 | 85,94 | 0,38 | 0,14 | 2,72 | 6,60E-03 |
| ZNF277 | 623,71 | -0,15 | 0,05 | -2,72 | 6,60E-03 |
| PABPC1L | 353,38 | 0,26 | 0,10 | 2,72 | 6,60E-03 |
| HCCS | 291,17 | -0,10 | 0,04 | -2,72 | 6,61E-03 |
| UBAP1 | 1522,78 | 0,16 | 0,06 | 2,72 | 6,62E-03 |
| TRIM37 | 932,90 | -0,10 | 0,04 | -2,72 | 6,62E-03 |
| LRRK1 | 1039,79 | 0,20 | 0,07 | 2,72 | 6,62E-03 |
| TBC1D22B | 621,07 | 0,19 | 0,07 | 2,72 | 6,63E-03 |
| RPS6KA4 | 620,05 | 0,14 | 0,05 | 2,71 | 6,63E-03 |

| | | | | | |
|---|---|---|---|---|---|
| VENTX | 93,82 | 0,30 | 0,11 | 2,71 | 6,64E-03 |
| MXD4 | 1977,58 | 0,12 | 0,04 | 2,71 | 6,64E-03 |
| PRPF31 | 819,53 | -0,10 | 0,04 | -2,71 | 6,69E-03 |
| SPRED2 | 68,25 | 0,28 | 0,10 | 2,71 | 6,69E-03 |
| GATB | 350,30 | 0,12 | 0,05 | 2,71 | 6,70E-03 |
| MAPRE3 | 79,57 | 0,30 | 0,11 | 2,71 | 6,70E-03 |
| TTC14 | 3746,58 | -0,17 | 0,06 | -2,71 | 6,71E-03 |
| UBASH3B | 1217,18 | 0,11 | 0,04 | 2,71 | 6,71E-03 |
| SLC37A1 | 679,17 | 0,09 | 0,03 | 2,71 | 6,71E-03 |
| ETFB | 327,27 | 0,08 | 0,03 | 2,71 | 6,72E-03 |
| RAB5A | 1901,29 | -0,11 | 0,04 | -2,71 | 6,72E-03 |
| RPL22 | 2687,17 | -0,20 | 0,07 | -2,71 | 6,73E-03 |
| MYO1B | 91,24 | 0,40 | 0,15 | 2,71 | 6,73E-03 |
| PRDM15 | 477,10 | 0,16 | 0,06 | 2,71 | 6,73E-03 |
| CD1C | 290,69 | -0,20 | 0,08 | -2,71 | 6,74E-03 |
| SLX9 | 238,94 | 0,09 | 0,03 | 2,71 | 6,73E-03 |
| OPA3 | 256,57 | 0,14 | 0,05 | 2,71 | 6,74E-03 |
| SPRYD4 | 339,49 | 0,13 | 0,05 | 2,71 | 6,77E-03 |
| PPAT | 353,01 | -0,14 | 0,05 | -2,71 | 6,77E-03 |
| TRMT112 | 1392,80 | -0,14 | 0,05 | -2,71 | 6,77E-03 |
| FAM3B | 33,19 | 0,96 | 0,35 | 2,71 | 6,77E-03 |
| KLF12 | 3881,86 | -0,15 | 0,05 | -2,71 | 6,78E-03 |
| CDC42BPB | 404,09 | 0,19 | 0,07 | 2,71 | 6,80E-03 |
| FUNDC1 | 138,47 | -0,16 | 0,06 | -2,71 | 6,81E-03 |
| ZNF736 | 1990,36 | -0,13 | 0,05 | -2,70 | 6,85E-03 |
| ATR | 3314,90 | -0,12 | 0,04 | -2,70 | 6,85E-03 |
| WDR35 | 526,49 | -0,13 | 0,05 | -2,70 | 6,86E-03 |
| CC2D1B | 988,57 | 0,15 | 0,05 | 2,70 | 6,87E-03 |
| SSH3 | 489,38 | 0,17 | 0,06 | 2,70 | 6,87E-03 |
| TATDN1 | 287,18 | -0,12 | 0,04 | -2,70 | 6,87E-03 |
| ICAM4 | 198,86 | 0,23 | 0,09 | 2,70 | 6,88E-03 |
| ANKRD6 | 306,39 | 0,13 | 0,05 | 2,70 | 6,89E-03 |
| TRAPPC8 | 5194,93 | -0,11 | 0,04 | -2,70 | 6,91E-03 |
| CLEC2D | 7824,17 | -0,14 | 0,05 | -2,70 | 6,91E-03 |
| DLG1 | 2157,21 | -0,10 | 0,04 | -2,70 | 6,92E-03 |
| ANKRD36C | 2132,27 | -0,15 | 0,05 | -2,70 | 6,92E-03 |
| MIGA1 | 1054,29 | -0,10 | 0,04 | -2,70 | 6,93E-03 |
| RABL6 | 505,54 | 0,18 | 0,07 | 2,70 | 6,93E-03 |
| PARS2 | 52,46 | 0,15 | 0,06 | 2,70 | 6,93E-03 |
| KLRF1 | 1031,01 | -0,27 | 0,10 | -2,70 | 6,93E-03 |
| UCP3 | 106,43 | 0,17 | 0,06 | 2,70 | 6,95E-03 |
| SLC66A2 | 1521,89 | 0,17 | 0,06 | 2,70 | 6,97E-03 |
| PTMA | 6953,54 | -0,13 | 0,05 | -2,70 | 6,98E-03 |
| ZFP41 | 130,13 | 0,22 | 0,08 | 2,70 | 6,98E-03 |
| NAA10 | 261,50 | 0,12 | 0,05 | 2,70 | 6,98E-03 |
| UCHL5 | 1240,49 | -0,18 | 0,07 | -2,70 | 7,01E-03 |

| | | | | | |
|---|---|---|---|---|---|
| PSMD5 | 910,78 | -0,07 | 0,03 | -2,70 | 7,03E-03 |
| ATOH8 | 56,81 | 0,51 | 0,19 | 2,69 | 7,04E-03 |
| FH | 600,06 | -0,11 | 0,04 | -2,69 | 7,04E-03 |
| OTUD7A | 193,55 | 0,25 | 0,09 | 2,69 | 7,05E-03 |
| ZNF326 | 1269,43 | -0,13 | 0,05 | -2,69 | 7,05E-03 |
| METTL25B | 319,93 | 0,10 | 0,04 | 2,69 | 7,05E-03 |
| RSL24D1 | 1002,65 | -0,34 | 0,13 | -2,69 | 7,06E-03 |
| HSPA13 | 936,88 | -0,22 | 0,08 | -2,69 | 7,06E-03 |
| GBA1 | 280,31 | 0,19 | 0,07 | 2,69 | 7,07E-03 |
| HGS | 863,20 | 0,22 | 0,08 | 2,69 | 7,07E-03 |
| IFT22 | 36,86 | 0,26 | 0,10 | 2,69 | 7,08E-03 |
| IKBIP | 1390,82 | -0,19 | 0,07 | -2,69 | 7,08E-03 |
| TNFAIP1 | 401,19 | 0,10 | 0,04 | 2,69 | 7,09E-03 |
| PALS2 | 688,93 | -0,17 | 0,06 | -2,69 | 7,10E-03 |
| MIS12 | 612,75 | -0,15 | 0,06 | -2,69 | 7,10E-03 |
| RAB1A | 2899,86 | -0,12 | 0,05 | -2,69 | 7,11E-03 |
| CTSK | 156,89 | 0,16 | 0,06 | 2,69 | 7,12E-03 |
| RTF1 | 1217,47 | -0,08 | 0,03 | -2,69 | 7,12E-03 |
| MAST4 | 585,63 | 0,15 | 0,06 | 2,69 | 7,13E-03 |
| GRAMD1B | 478,42 | 0,26 | 0,10 | 2,69 | 7,14E-03 |
| AGGF1 | 667,03 | -0,14 | 0,05 | -2,69 | 7,14E-03 |
| AKAP5 | 162,76 | -0,28 | 0,10 | -2,69 | 7,14E-03 |
| NME4 | 278,70 | 0,19 | 0,07 | 2,69 | 7,14E-03 |
| C17orf58 | 31,99 | 0,26 | 0,10 | 2,69 | 7,14E-03 |
| RANBP9 | 1200,57 | -0,12 | 0,04 | -2,69 | 7,15E-03 |
| GBA2 | 1297,64 | 0,13 | 0,05 | 2,69 | 7,16E-03 |
| C6orf47 | 1076,38 | -0,08 | 0,03 | -2,69 | 7,16E-03 |
| ELF2 | 5431,84 | -0,12 | 0,05 | -2,69 | 7,16E-03 |
| PPP1R12C | 1007,96 | 0,24 | 0,09 | 2,69 | 7,16E-03 |
| GEMIN7 | 244,44 | 0,11 | 0,04 | 2,69 | 7,16E-03 |
| RGPD8 | 295,46 | -0,23 | 0,08 | -2,69 | 7,16E-03 |
| IL11RA | 406,91 | 0,21 | 0,08 | 2,69 | 7,15E-03 |
| HAUS5 | 152,30 | 0,18 | 0,07 | 2,69 | 7,17E-03 |
| FGF2 | 30,51 | 0,50 | 0,19 | 2,69 | 7,17E-03 |
| ZNF260 | 374,23 | -0,21 | 0,08 | -2,69 | 7,18E-03 |
| MICAL1 | 1131,77 | 0,28 | 0,10 | 2,69 | 7,18E-03 |
| NECTIN1 | 141,38 | 0,31 | 0,12 | 2,69 | 7,19E-03 |
| MARCHF9 | 219,50 | 0,18 | 0,07 | 2,69 | 7,20E-03 |
| MRPS2 | 178,95 | 0,15 | 0,06 | 2,69 | 7,20E-03 |
| NVL | 740,73 | -0,08 | 0,03 | -2,69 | 7,20E-03 |
| SRF | 622,09 | 0,12 | 0,05 | 2,69 | 7,21E-03 |
| PAQR6 | 69,22 | 0,31 | 0,11 | 2,69 | 7,21E-03 |
| ZNF574 | 206,32 | 0,14 | 0,05 | 2,69 | 7,23E-03 |
| XK | 1785,34 | 0,34 | 0,13 | 2,69 | 7,24E-03 |
| ULK1 | 724,25 | 0,33 | 0,12 | 2,68 | 7,27E-03 |
| AHNAK | 61397,00 | -0,10 | 0,04 | -2,68 | 7,27E-03 |

| | | | | | |
|---|---|---|---|---|---|
| NRP1 | 117,71 | 0,35 | 0,13 | 2,68 | 7,27E-03 |
| ANP32A | 5775,46 | -0,11 | 0,04 | -2,68 | 7,28E-03 |
| SLC4A1 | 13186,76 | 0,38 | 0,14 | 2,68 | 7,29E-03 |
| MMP28 | 47,10 | 0,40 | 0,15 | 2,68 | 7,31E-03 |
| MARCHF2 | 2409,81 | 0,24 | 0,09 | 2,68 | 7,31E-03 |
| COG7 | 377,66 | 0,11 | 0,04 | 2,68 | 7,33E-03 |
| TRMU | 908,75 | 0,09 | 0,03 | 2,68 | 7,33E-03 |
| ZNF253 | 415,39 | -0,17 | 0,06 | -2,68 | 7,34E-03 |
| SACS | 2078,31 | -0,21 | 0,08 | -2,68 | 7,34E-03 |
| PHAF1 | 344,48 | 0,15 | 0,06 | 2,68 | 7,35E-03 |
| DNM1L | 1570,50 | -0,10 | 0,04 | -2,68 | 7,36E-03 |
| ZNF75A | 1093,59 | -0,11 | 0,04 | -2,68 | 7,36E-03 |
| ACOT13 | 294,30 | -0,10 | 0,04 | -2,68 | 7,36E-03 |
| AKAP11 | 3030,05 | -0,12 | 0,05 | -2,68 | 7,38E-03 |
| PLEC | 9492,09 | 0,20 | 0,07 | 2,68 | 7,37E-03 |
| PIGK | 757,55 | -0,20 | 0,08 | -2,68 | 7,38E-03 |
| CCDC17 | 242,22 | 0,21 | 0,08 | 2,68 | 7,40E-03 |
| SMIM15 | 701,68 | -0,18 | 0,07 | -2,68 | 7,41E-03 |
| FKBP8 | 21789,24 | 0,31 | 0,11 | 2,68 | 7,42E-03 |
| MCC | 125,34 | 0,32 | 0,12 | 2,68 | 7,42E-03 |
| ME2 | 3025,69 | -0,08 | 0,03 | -2,68 | 7,42E-03 |
| UHRF2 | 3504,42 | -0,10 | 0,04 | -2,68 | 7,43E-03 |
| TBCE | 236,97 | 0,13 | 0,05 | 2,68 | 7,43E-03 |
| ZNF282 | 497,89 | 0,11 | 0,04 | 2,68 | 7,43E-03 |
| DEDD | 295,92 | 0,13 | 0,05 | 2,68 | 7,44E-03 |
| ZNF234 | 648,34 | -0,09 | 0,03 | -2,68 | 7,46E-03 |
| TMX2 | 637,01 | -0,07 | 0,02 | -2,68 | 7,47E-03 |
| CEP85L | 3413,48 | -0,14 | 0,05 | -2,67 | 7,48E-03 |
| LGALS9 | 481,93 | 0,27 | 0,10 | 2,67 | 7,48E-03 |
| BTBD1 | 1608,74 | -0,11 | 0,04 | -2,67 | 7,48E-03 |
| PRKCZ | 572,27 | 0,13 | 0,05 | 2,67 | 7,48E-03 |
| MAP2 | 48,66 | 0,49 | 0,18 | 2,67 | 7,49E-03 |
| RABGAP1L | 9564,55 | -0,11 | 0,04 | -2,67 | 7,49E-03 |
| PGM1 | 826,09 | -0,10 | 0,04 | -2,67 | 7,49E-03 |
| COASY | 208,69 | 0,25 | 0,09 | 2,67 | 7,50E-03 |
| CDK2AP1 | 298,71 | 0,15 | 0,05 | 2,67 | 7,51E-03 |
| EPM2AIP1 | 2213,31 | -0,13 | 0,05 | -2,67 | 7,52E-03 |
| FADS3 | 88,01 | 0,26 | 0,10 | 2,67 | 7,52E-03 |
| FBXO8 | 710,81 | -0,16 | 0,06 | -2,67 | 7,53E-03 |
| TFAP4 | 173,46 | 0,21 | 0,08 | 2,67 | 7,53E-03 |
| GTF2H3 | 497,06 | -0,11 | 0,04 | -2,67 | 7,54E-03 |
| SNRPF | 282,40 | -0,17 | 0,06 | -2,67 | 7,54E-03 |
| PFDN6 | 310,82 | -0,13 | 0,05 | -2,67 | 7,54E-03 |
| URI1 | 1709,30 | -0,13 | 0,05 | -2,67 | 7,56E-03 |
| UBXN6 | 8355,17 | 0,29 | 0,11 | 2,67 | 7,56E-03 |
| ENSG00000256029 | 78,09 | 0,23 | 0,09 | 2,67 | 7,56E-03 |

| | | | | | |
|---|---|---|---|---|---|
| TTC31 | 369,19 | 0,15 | 0,06 | 2,67 | 7,58E-03 |
| VPS28 | 1425,72 | 0,15 | 0,05 | 2,67 | 7,58E-03 |
| RRM2B | 3046,90 | -0,18 | 0,07 | -2,67 | 7,59E-03 |
| EBAG9 | 457,38 | -0,15 | 0,05 | -2,67 | 7,59E-03 |
| HLA-DMB | 2939,86 | -0,14 | 0,05 | -2,67 | 7,60E-03 |
| TSR2 | 456,03 | -0,09 | 0,03 | -2,67 | 7,60E-03 |
| TRNT1 | 1146,98 | -0,11 | 0,04 | -2,67 | 7,63E-03 |
| DUSP12 | 375,07 | -0,11 | 0,04 | -2,67 | 7,64E-03 |
| TCEA3 | 140,31 | 0,24 | 0,09 | 2,67 | 7,65E-03 |
| BORCS8 | 311,62 | 0,23 | 0,09 | 2,67 | 7,65E-03 |
| RAVER1 | 224,08 | 0,24 | 0,09 | 2,67 | 7,66E-03 |
| SSPN | 92,13 | 0,29 | 0,11 | 2,67 | 7,67E-03 |
| POFUT2 | 425,13 | 0,18 | 0,07 | 2,67 | 7,68E-03 |
| TUBB4B | 1456,57 | 0,12 | 0,05 | 2,67 | 7,68E-03 |
| GPATCH4 | 340,42 | -0,12 | 0,04 | -2,67 | 7,70E-03 |
| GOPC | 1645,02 | -0,15 | 0,06 | -2,66 | 7,70E-03 |
| HSPE1 | 208,53 | -0,16 | 0,06 | -2,66 | 7,71E-03 |
| ARF6 | 4538,52 | -0,10 | 0,04 | -2,66 | 7,71E-03 |
| ST8SIA4 | 9708,19 | -0,16 | 0,06 | -2,66 | 7,71E-03 |
| NDUFAF2 | 105,41 | -0,15 | 0,05 | -2,66 | 7,71E-03 |
| PHF1 | 641,93 | 0,15 | 0,06 | 2,66 | 7,72E-03 |
| DHRS1 | 199,83 | 0,15 | 0,06 | 2,66 | 7,74E-03 |
| TNFRSF21 | 45,91 | 0,35 | 0,13 | 2,66 | 7,74E-03 |
| PLEKHF2 | 1619,81 | -0,17 | 0,06 | -2,66 | 7,76E-03 |
| ITPR3 | 949,73 | 0,23 | 0,09 | 2,66 | 7,76E-03 |
| ENKD1 | 157,71 | 0,13 | 0,05 | 2,66 | 7,76E-03 |
| PSMF1 | 2530,36 | 0,24 | 0,09 | 2,66 | 7,77E-03 |
| ZNF844 | 906,95 | -0,17 | 0,06 | -2,66 | 7,78E-03 |
| ALG1L2 | 64,98 | 0,30 | 0,11 | 2,66 | 7,79E-03 |
| PLA2G6 | 455,71 | 0,18 | 0,07 | 2,66 | 7,79E-03 |
| DVL2 | 179,58 | 0,20 | 0,08 | 2,66 | 7,79E-03 |
| PNISR | 9704,70 | -0,13 | 0,05 | -2,66 | 7,80E-03 |
| CITED2 | 948,07 | 0,08 | 0,03 | 2,66 | 7,79E-03 |
| HSF2 | 392,58 | -0,16 | 0,06 | -2,66 | 7,80E-03 |
| ACSL3 | 1463,58 | -0,11 | 0,04 | -2,66 | 7,80E-03 |
| PPCS | 847,81 | -0,09 | 0,04 | -2,66 | 7,80E-03 |
| EIF4EBP1 | 71,33 | 0,16 | 0,06 | 2,66 | 7,81E-03 |
| LSM6 | 1058,52 | -0,13 | 0,05 | -2,66 | 7,83E-03 |
| ZNF181 | 415,55 | -0,22 | 0,08 | -2,66 | 7,83E-03 |
| ST8SIA1 | 478,53 | 0,20 | 0,07 | 2,66 | 7,83E-03 |
| MRPS14 | 563,48 | -0,12 | 0,05 | -2,66 | 7,83E-03 |
| UNC13D | 2171,97 | 0,25 | 0,10 | 2,66 | 7,83E-03 |
| ACSS2 | 602,75 | 0,14 | 0,05 | 2,66 | 7,84E-03 |
| SOS2 | 4517,65 | -0,14 | 0,05 | -2,66 | 7,84E-03 |
| PHC1 | 293,08 | 0,14 | 0,05 | 2,66 | 7,84E-03 |
| OGFRL1 | 18721,78 | -0,19 | 0,07 | -2,66 | 7,85E-03 |

| | | | | | |
|---|---|---|---|---|---|
| LGALS3 | 1393,09 | 0,25 | 0,09 | 2,66 | 7,85E-03 |
| BAG3 | 97,88 | 0,18 | 0,07 | 2,66 | 7,85E-03 |
| ZNF420 | 363,47 | -0,16 | 0,06 | -2,66 | 7,85E-03 |
| CD5 | 635,48 | 0,20 | 0,07 | 2,66 | 7,86E-03 |
| ZG16B | 25,39 | 0,31 | 0,11 | 2,66 | 7,87E-03 |
| TRAPPC4 | 948,69 | -0,09 | 0,03 | -2,66 | 7,90E-03 |
| EHD3 | 300,22 | 0,21 | 0,08 | 2,66 | 7,90E-03 |
| RPL36AL | 2185,50 | -0,21 | 0,08 | -2,66 | 7,90E-03 |
| ASTN2 | 182,29 | 0,23 | 0,09 | 2,66 | 7,92E-03 |
| TENT4B | 1179,48 | -0,08 | 0,03 | -2,66 | 7,93E-03 |
| HEXB | 1486,43 | -0,10 | 0,04 | -2,66 | 7,93E-03 |
| PTRH2 | 512,42 | -0,11 | 0,04 | -2,65 | 7,94E-03 |
| TAPT1 | 1925,11 | -0,10 | 0,04 | -2,65 | 7,94E-03 |
| KRT18 | 56,51 | 0,26 | 0,10 | 2,65 | 7,95E-03 |
| RASA2 | 4672,39 | -0,12 | 0,05 | -2,65 | 7,95E-03 |
| KSR1 | 1342,38 | 0,22 | 0,08 | 2,65 | 7,97E-03 |
| BNIP2 | 6034,74 | -0,14 | 0,05 | -2,65 | 7,97E-03 |
| SH3RF1 | 134,99 | 0,20 | 0,08 | 2,65 | 7,98E-03 |
| C13orf46 | 271,37 | 0,28 | 0,11 | 2,65 | 7,98E-03 |
| SH2B2 | 266,89 | 0,24 | 0,09 | 2,65 | 7,98E-03 |
| ZNF18 | 896,37 | -0,07 | 0,03 | -2,65 | 7,99E-03 |
| EXOC5 | 2376,48 | -0,15 | 0,06 | -2,65 | 8,00E-03 |
| RASA1 | 2441,23 | -0,10 | 0,04 | -2,65 | 8,00E-03 |
| PCDHGC3 | 31,37 | 0,33 | 0,12 | 2,65 | 8,01E-03 |
| CD6 | 1430,40 | 0,20 | 0,07 | 2,65 | 8,03E-03 |
| NECTIN2 | 89,61 | 0,57 | 0,22 | 2,65 | 8,04E-03 |
| CAPRIN1 | 3947,21 | -0,06 | 0,02 | -2,65 | 8,05E-03 |
| VCP | 4781,29 | -0,10 | 0,04 | -2,65 | 8,07E-03 |
| CHMP5 | 798,10 | -0,24 | 0,09 | -2,65 | 8,07E-03 |
| WASHC5 | 1680,65 | -0,07 | 0,03 | -2,65 | 8,07E-03 |
| SP3 | 9789,23 | -0,15 | 0,06 | -2,65 | 8,08E-03 |
| SLC35A4 | 245,26 | 0,19 | 0,07 | 2,65 | 8,09E-03 |
| IQCB1 | 1027,46 | -0,14 | 0,05 | -2,65 | 8,09E-03 |
| TMED2 | 2450,64 | -0,15 | 0,06 | -2,65 | 8,09E-03 |
| AATF | 1631,55 | -0,09 | 0,03 | -2,65 | 8,10E-03 |
| MCM3 | 992,84 | -0,10 | 0,04 | -2,65 | 8,10E-03 |
| HNRNPK | 15917,47 | -0,11 | 0,04 | -2,65 | 8,11E-03 |
| CAPS | 377,57 | 0,15 | 0,06 | 2,65 | 8,11E-03 |
| SARM1 | 538,83 | 0,19 | 0,07 | 2,65 | 8,11E-03 |
| CRYZ | 307,38 | -0,23 | 0,09 | -2,65 | 8,11E-03 |
| PRKACB | 3365,58 | -0,21 | 0,08 | -2,65 | 8,12E-03 |
| DDX10 | 808,94 | -0,11 | 0,04 | -2,65 | 8,15E-03 |
| MCMBP | 3179,00 | -0,10 | 0,04 | -2,65 | 8,15E-03 |
| ZNF473 | 307,44 | -0,09 | 0,03 | -2,64 | 8,17E-03 |
| RPS6KC1 | 1174,97 | -0,10 | 0,04 | -2,64 | 8,18E-03 |
| ZNF394 | 2511,78 | -0,08 | 0,03 | -2,64 | 8,19E-03 |

| | | | | | |
|---|---|---|---|---|---|
| SDF4 | 871,31 | 0,14 | 0,05 | 2,64 | 8,20E-03 |
| TIGD2 | 156,97 | -0,21 | 0,08 | -2,64 | 8,21E-03 |
| TBC1D13 | 255,43 | 0,21 | 0,08 | 2,64 | 8,23E-03 |
| ALPK2 | 55,49 | 0,52 | 0,20 | 2,64 | 8,26E-03 |
| FAM98C | 365,44 | 0,12 | 0,05 | 2,64 | 8,26E-03 |
| DEPDC5 | 763,21 | 0,14 | 0,05 | 2,64 | 8,27E-03 |
| HDAC11 | 138,51 | 0,17 | 0,07 | 2,64 | 8,27E-03 |
| ABCC2 | 218,45 | 0,17 | 0,06 | 2,64 | 8,28E-03 |
| ZNF440 | 676,87 | -0,09 | 0,03 | -2,64 | 8,30E-03 |
| SPTLC3 | 54,59 | 0,39 | 0,15 | 2,64 | 8,30E-03 |
| SMARCA4 | 967,08 | 0,21 | 0,08 | 2,64 | 8,31E-03 |
| CORO7 | 568,18 | 0,19 | 0,07 | 2,64 | 8,31E-03 |
| SMAD1 | 139,95 | 0,25 | 0,09 | 2,64 | 8,31E-03 |
| MAP3K21 | 109,05 | 0,23 | 0,09 | 2,64 | 8,31E-03 |
| CHCHD3 | 979,68 | -0,09 | 0,03 | -2,64 | 8,32E-03 |
| RNF34 | 2245,82 | -0,06 | 0,02 | -2,64 | 8,34E-03 |
| GCM1 | 37,03 | 0,26 | 0,10 | 2,64 | 8,34E-03 |
| GDPD3 | 108,73 | 0,28 | 0,11 | 2,64 | 8,34E-03 |
| AP3S2 | 577,19 | 0,14 | 0,05 | 2,64 | 8,35E-03 |
| PDIK1L | 403,00 | -0,20 | 0,08 | -2,64 | 8,35E-03 |
| COG3 | 2213,82 | -0,08 | 0,03 | -2,64 | 8,36E-03 |
| ZC3H7B | 794,53 | 0,17 | 0,07 | 2,64 | 8,37E-03 |
| SNRPG | 405,13 | -0,20 | 0,08 | -2,64 | 8,37E-03 |
| NTAN1 | 427,67 | 0,15 | 0,06 | 2,64 | 8,38E-03 |
| DNAJC30 | 93,18 | 0,17 | 0,06 | 2,64 | 8,38E-03 |
| ZNF408 | 260,41 | 0,08 | 0,03 | 2,64 | 8,38E-03 |
| EXOSC3 | 842,64 | -0,11 | 0,04 | -2,64 | 8,40E-03 |
| ZC3H3 | 418,59 | 0,21 | 0,08 | 2,64 | 8,41E-03 |
| GOLPH3 | 2441,09 | -0,07 | 0,03 | -2,64 | 8,41E-03 |
| UBFD1 | 712,12 | 0,10 | 0,04 | 2,63 | 8,42E-03 |
| GNAI3 | 8745,46 | -0,11 | 0,04 | -2,63 | 8,42E-03 |
| HIC2 | 393,01 | 0,11 | 0,04 | 2,63 | 8,42E-03 |
| TMEM38A | 87,04 | 0,17 | 0,06 | 2,63 | 8,42E-03 |
| MFN1 | 2527,76 | -0,12 | 0,05 | -2,63 | 8,43E-03 |
| COMMD1 | 334,97 | -0,08 | 0,03 | -2,63 | 8,43E-03 |
| USP34 | 12171,20 | -0,10 | 0,04 | -2,63 | 8,44E-03 |
| ZNHIT1 | 742,50 | 0,08 | 0,03 | 2,63 | 8,47E-03 |
| GINS3 | 68,10 | 0,20 | 0,08 | 2,63 | 8,47E-03 |
| NFATC1 | 514,50 | 0,22 | 0,08 | 2,63 | 8,49E-03 |
| NEK10 | 96,17 | 0,29 | 0,11 | 2,63 | 8,49E-03 |
| RAD17 | 646,89 | -0,11 | 0,04 | -2,63 | 8,49E-03 |
| FAM32A | 1490,79 | -0,12 | 0,05 | -2,63 | 8,50E-03 |
| PDS5B | 3234,31 | -0,12 | 0,05 | -2,63 | 8,50E-03 |
| GXYLT1 | 1040,10 | -0,11 | 0,04 | -2,63 | 8,50E-03 |
| CACHD1 | 68,91 | 0,41 | 0,15 | 2,63 | 8,52E-03 |
| RRAGB | 546,00 | -0,09 | 0,03 | -2,63 | 8,53E-03 |

| | | | | | |
|---|---|---|---|---|---|
| PANK4 | 862,65 | 0,09 | 0,03 | 2,63 | 8,55E-03 |
| METAP1D | 141,91 | 0,16 | 0,06 | 2,63 | 8,55E-03 |
| DCP2 | 12045,98 | -0,15 | 0,06 | -2,63 | 8,56E-03 |
| TBCD | 1406,80 | 0,15 | 0,06 | 2,63 | 8,57E-03 |
| SLC46A3 | 761,24 | -0,08 | 0,03 | -2,63 | 8,58E-03 |
| ATP6V1H | 1243,79 | -0,08 | 0,03 | -2,63 | 8,58E-03 |
| SOX6 | 810,50 | 0,28 | 0,11 | 2,63 | 8,58E-03 |
| PCDH12 | 42,29 | 0,38 | 0,14 | 2,63 | 8,58E-03 |
| CLDN23 | 41,86 | 0,31 | 0,12 | 2,63 | 8,59E-03 |
| SLC37A4 | 187,30 | 0,12 | 0,05 | 2,63 | 8,59E-03 |
| VAT1 | 158,58 | 0,18 | 0,07 | 2,63 | 8,60E-03 |
| ADAP2 | 230,37 | 0,18 | 0,07 | 2,63 | 8,60E-03 |
| IMPACT | 522,90 | -0,12 | 0,05 | -2,63 | 8,62E-03 |
| DNAJC14 | 796,91 | -0,07 | 0,03 | -2,63 | 8,62E-03 |
| TRAF3 | 1360,32 | 0,11 | 0,04 | 2,63 | 8,62E-03 |
| SHOC2 | 4821,86 | -0,16 | 0,06 | -2,63 | 8,64E-03 |
| MORN1 | 294,31 | 0,14 | 0,05 | 2,63 | 8,65E-03 |
| OXSR1 | 2607,32 | -0,09 | 0,03 | -2,63 | 8,66E-03 |
| RAB39B | 362,58 | -0,12 | 0,05 | -2,63 | 8,66E-03 |
| FBXO33 | 1976,08 | -0,11 | 0,04 | -2,62 | 8,67E-03 |
| ACTR3 | 17734,27 | -0,13 | 0,05 | -2,62 | 8,68E-03 |
| ZBTB37 | 7642,23 | -0,10 | 0,04 | -2,62 | 8,70E-03 |
| USP38 | 1825,91 | -0,09 | 0,03 | -2,62 | 8,70E-03 |
| KLHL13 | 35,74 | 0,30 | 0,12 | 2,62 | 8,70E-03 |
| ZNF770 | 2522,77 | -0,14 | 0,05 | -2,62 | 8,76E-03 |
| PSMC4 | 978,63 | -0,08 | 0,03 | -2,62 | 8,78E-03 |
| PAPOLG | 1973,65 | -0,08 | 0,03 | -2,62 | 8,79E-03 |
| ARAF | 497,77 | 0,16 | 0,06 | 2,62 | 8,82E-03 |
| MAOA | 54,94 | 0,47 | 0,18 | 2,62 | 8,81E-03 |
| ZNF76 | 550,83 | 0,13 | 0,05 | 2,62 | 8,81E-03 |
| RIPOR1 | 785,78 | 0,18 | 0,07 | 2,62 | 8,81E-03 |
| PRDM11 | 236,91 | 0,23 | 0,09 | 2,62 | 8,81E-03 |
| GALE | 151,40 | 0,11 | 0,04 | 2,62 | 8,81E-03 |
| PRAG1 | 150,00 | 0,24 | 0,09 | 2,62 | 8,83E-03 |
| PRCD | 50,38 | 0,33 | 0,13 | 2,62 | 8,84E-03 |
| POLR3F | 263,75 | -0,10 | 0,04 | -2,62 | 8,84E-03 |
| ZNF235 | 579,25 | -0,08 | 0,03 | -2,62 | 8,85E-03 |
| DCBLD2 | 103,10 | 0,28 | 0,11 | 2,62 | 8,85E-03 |
| DPP9 | 700,17 | 0,20 | 0,07 | 2,62 | 8,85E-03 |
| COQ7 | 432,44 | -0,08 | 0,03 | -2,62 | 8,85E-03 |
| IST1 | 4685,78 | -0,07 | 0,03 | -2,62 | 8,86E-03 |
| PDGFD | 322,65 | -0,22 | 0,09 | -2,62 | 8,87E-03 |
| CBLB | 2874,14 | -0,14 | 0,05 | -2,62 | 8,87E-03 |
| SLFN12 | 769,90 | -0,15 | 0,06 | -2,62 | 8,89E-03 |
| BRAT1 | 800,23 | 0,11 | 0,04 | 2,62 | 8,90E-03 |
| REPIN1 | 828,46 | 0,12 | 0,05 | 2,62 | 8,90E-03 |

| | | | | | |
|---|---|---|---|---|---|
| MPHOSPH9 | 1368,30 | -0,09 | 0,04 | -2,62 | 8,91E-03 |
| ZMPSTE24 | 1674,80 | -0,13 | 0,05 | -2,62 | 8,92E-03 |
| ZNF860 | 185,94 | -0,31 | 0,12 | -2,62 | 8,92E-03 |
| CTNS | 273,09 | 0,10 | 0,04 | 2,61 | 8,93E-03 |
| RPL22L1 | 230,00 | -0,24 | 0,09 | -2,61 | 8,94E-03 |
| CCNF | 97,80 | 0,20 | 0,07 | 2,61 | 8,95E-03 |
| ARHGEF3 | 2718,66 | -0,08 | 0,03 | -2,61 | 8,96E-03 |
| SENP2 | 1376,61 | -0,07 | 0,03 | -2,61 | 8,95E-03 |
| PPIP5K2 | 4704,38 | -0,16 | 0,06 | -2,61 | 8,97E-03 |
| ZNF490 | 248,19 | -0,07 | 0,03 | -2,61 | 8,97E-03 |
| PRICKLE3 | 98,97 | 0,21 | 0,08 | 2,61 | 8,98E-03 |
| BDH1 | 306,30 | 0,17 | 0,07 | 2,61 | 8,99E-03 |
| DNAI4 | 88,51 | 0,24 | 0,09 | 2,61 | 8,99E-03 |
| GAS2L3 | 86,86 | 0,22 | 0,08 | 2,61 | 8,99E-03 |
| PPRC1 | 217,36 | 0,21 | 0,08 | 2,61 | 9,00E-03 |
| ERH | 801,11 | -0,25 | 0,10 | -2,61 | 9,00E-03 |
| ANKIB1 | 2144,21 | -0,08 | 0,03 | -2,61 | 9,02E-03 |
| DALRD3 | 395,77 | 0,08 | 0,03 | 2,61 | 9,02E-03 |
| CSF1 | 185,25 | 0,25 | 0,10 | 2,61 | 9,03E-03 |
| FHOD1 | 322,84 | 0,23 | 0,09 | 2,61 | 9,04E-03 |
| RPN1 | 4465,32 | -0,09 | 0,04 | -2,61 | 9,04E-03 |
| RGS19 | 1784,71 | 0,15 | 0,06 | 2,61 | 9,04E-03 |
| NMB | 34,33 | 0,18 | 0,07 | 2,61 | 9,03E-03 |
| MAP3K1 | 12079,02 | -0,09 | 0,04 | -2,61 | 9,04E-03 |
| EIF4A1 | 572,45 | 0,11 | 0,04 | 2,61 | 9,04E-03 |
| C9orf85 | 326,35 | -0,07 | 0,03 | -2,61 | 9,05E-03 |
| CEP170 | 3201,21 | -0,11 | 0,04 | -2,61 | 9,06E-03 |
| GABARAPL1 | 3247,56 | -0,15 | 0,06 | -2,61 | 9,07E-03 |
| QTRT2 | 1140,92 | -0,07 | 0,03 | -2,61 | 9,07E-03 |
| TP53BP2 | 1378,53 | -0,06 | 0,02 | -2,61 | 9,07E-03 |
| ELMOD2 | 1433,31 | -0,19 | 0,07 | -2,61 | 9,09E-03 |
| H2AZ1 | 3189,45 | -0,17 | 0,06 | -2,61 | 9,12E-03 |
| RPS27A | 5979,29 | -0,26 | 0,10 | -2,61 | 9,13E-03 |
| NIF3L1 | 425,84 | -0,10 | 0,04 | -2,61 | 9,13E-03 |
| MMP8 | 148,50 | 0,49 | 0,19 | 2,61 | 9,14E-03 |
| TMEM259 | 1228,78 | 0,19 | 0,07 | 2,61 | 9,14E-03 |
| ZMAT1 | 1134,97 | -0,12 | 0,05 | -2,61 | 9,15E-03 |
| ARPP19 | 4760,31 | -0,11 | 0,04 | -2,61 | 9,15E-03 |
| BCL9L | 822,07 | 0,28 | 0,11 | 2,61 | 9,16E-03 |
| FLNB | 1317,62 | 0,14 | 0,05 | 2,61 | 9,16E-03 |
| EPDR1 | 39,52 | 0,27 | 0,10 | 2,60 | 9,19E-03 |
| UBAC1 | 879,98 | 0,10 | 0,04 | 2,60 | 9,19E-03 |
| SMIM29 | 211,68 | 0,10 | 0,04 | 2,60 | 9,21E-03 |
| MARCHF7 | 8599,24 | -0,14 | 0,05 | -2,60 | 9,20E-03 |
| GALNT5 | 87,63 | 0,32 | 0,12 | 2,60 | 9,21E-03 |
| ACO2 | 609,65 | 0,14 | 0,05 | 2,60 | 9,23E-03 |

| | | | | | |
|---|---|---|---|---|---|
| PRKAR1A | 11433,57 | -0,14 | 0,06 | -2,60 | 9,23E-03 |
| FOXO3B | 92,21 | 0,15 | 0,06 | 2,60 | 9,23E-03 |
| ZSCAN30 | 664,10 | -0,09 | 0,04 | -2,60 | 9,23E-03 |
| MTF2 | 1719,96 | -0,12 | 0,05 | -2,60 | 9,23E-03 |
| ATP13A4 | 115,01 | 0,28 | 0,11 | 2,60 | 9,24E-03 |
| NBN | 3485,06 | -0,17 | 0,07 | -2,60 | 9,25E-03 |
| ALAS2 | 40357,46 | 0,41 | 0,16 | 2,60 | 9,26E-03 |
| SCMH1 | 384,08 | 0,11 | 0,04 | 2,60 | 9,27E-03 |
| RBMX | 3453,25 | -0,09 | 0,03 | -2,60 | 9,30E-03 |
| RASGRP4 | 886,01 | 0,32 | 0,12 | 2,60 | 9,30E-03 |
| GLB1L2 | 68,68 | 0,51 | 0,20 | 2,60 | 9,29E-03 |
| TROAP | 185,32 | 0,29 | 0,11 | 2,60 | 9,30E-03 |
| ATP6V0E2 | 288,35 | 0,14 | 0,06 | 2,60 | 9,31E-03 |
| PHLDB3 | 175,20 | 0,16 | 0,06 | 2,60 | 9,31E-03 |
| OGA | 17671,67 | -0,10 | 0,04 | -2,60 | 9,31E-03 |
| SPATA20 | 214,77 | 0,25 | 0,10 | 2,60 | 9,31E-03 |
| CDCA8 | 35,43 | 0,27 | 0,10 | 2,60 | 9,31E-03 |
| LAPTM4B | 107,43 | 0,20 | 0,08 | 2,60 | 9,32E-03 |
| CSTPP1 | 140,08 | 0,19 | 0,07 | 2,60 | 9,33E-03 |
| PTPN20 | 37,08 | 0,43 | 0,17 | 2,60 | 9,33E-03 |
| SLC41A3 | 412,90 | 0,11 | 0,04 | 2,60 | 9,32E-03 |
| HSPBP1 | 98,25 | 0,16 | 0,06 | 2,60 | 9,33E-03 |
| DSTYK | 1699,73 | -0,06 | 0,02 | -2,60 | 9,34E-03 |
| PLCXD2 | 1001,49 | -0,13 | 0,05 | -2,60 | 9,34E-03 |
| HECTD4 | 3729,01 | 0,12 | 0,05 | 2,60 | 9,34E-03 |
| KIF23 | 95,41 | 0,26 | 0,10 | 2,60 | 9,35E-03 |
| NAP1L1 | 3771,66 | -0,13 | 0,05 | -2,60 | 9,35E-03 |
| NRXN1 | 168,99 | 0,34 | 0,13 | 2,60 | 9,36E-03 |
| TTC27 | 646,43 | -0,11 | 0,04 | -2,60 | 9,37E-03 |
| NUFIP2 | 9931,82 | -0,10 | 0,04 | -2,60 | 9,37E-03 |
| FBXO3 | 2981,69 | -0,11 | 0,04 | -2,60 | 9,38E-03 |
| MTG1 | 208,90 | 0,16 | 0,06 | 2,60 | 9,39E-03 |
| VIPR1 | 430,25 | 0,21 | 0,08 | 2,60 | 9,39E-03 |
| TAF9B | 159,91 | -0,19 | 0,07 | -2,60 | 9,40E-03 |
| ZNF805 | 693,20 | -0,09 | 0,03 | -2,60 | 9,44E-03 |
| DNAH7 | 99,32 | 0,40 | 0,15 | 2,60 | 9,44E-03 |
| GET4 | 158,68 | 0,19 | 0,07 | 2,60 | 9,43E-03 |
| PPP1R13B | 229,76 | 0,19 | 0,07 | 2,60 | 9,44E-03 |
| DACH1 | 162,37 | 0,20 | 0,08 | 2,60 | 9,44E-03 |
| CENPN | 79,78 | 0,18 | 0,07 | 2,60 | 9,45E-03 |
| ME3 | 91,88 | 0,26 | 0,10 | 2,60 | 9,45E-03 |
| BOD1 | 490,86 | -0,10 | 0,04 | -2,60 | 9,45E-03 |
| ZNF785 | 226,82 | 0,17 | 0,07 | 2,60 | 9,46E-03 |
| CLTCL1 | 345,35 | 0,19 | 0,07 | 2,59 | 9,47E-03 |
| ARHGEF6 | 4950,20 | -0,08 | 0,03 | -2,59 | 9,46E-03 |
| ERMAP | 664,92 | 0,12 | 0,05 | 2,59 | 9,48E-03 |

| | | | | | |
|---|---|---|---|---|---|
| ALG2 | 621,60 | -0,08 | 0,03 | -2,59 | 9,49E-03 |
| CDK2AP2 | 89,85 | 0,24 | 0,09 | 2,59 | 9,49E-03 |
| GAPT | 1299,71 | -0,20 | 0,08 | -2,59 | 9,50E-03 |
| SLC9A1 | 201,73 | 0,31 | 0,12 | 2,59 | 9,52E-03 |
| NDUFAF4 | 181,85 | -0,20 | 0,08 | -2,59 | 9,52E-03 |
| RPS25 | 6088,00 | -0,25 | 0,10 | -2,59 | 9,53E-03 |
| ST14 | 261,42 | 0,22 | 0,08 | 2,59 | 9,53E-03 |
| PUS7L | 1838,15 | -0,16 | 0,06 | -2,59 | 9,54E-03 |
| PTPA | 779,73 | 0,11 | 0,04 | 2,59 | 9,55E-03 |
| SLC22A18 | 366,00 | 0,21 | 0,08 | 2,59 | 9,56E-03 |
| CAPZA1 | 8248,96 | -0,19 | 0,07 | -2,59 | 9,58E-03 |
| ZNF711 | 75,12 | -0,25 | 0,09 | -2,59 | 9,58E-03 |
| C22orf39 | 794,40 | -0,05 | 0,02 | -2,59 | 9,57E-03 |
| SNX15 | 55,01 | 0,18 | 0,07 | 2,59 | 9,59E-03 |
| MRPL33 | 505,00 | -0,13 | 0,05 | -2,59 | 9,61E-03 |
| ACVR1 | 441,34 | 0,10 | 0,04 | 2,59 | 9,61E-03 |
| TIPARP | 1898,67 | -0,11 | 0,04 | -2,59 | 9,62E-03 |
| EIF3J | 994,81 | -0,12 | 0,05 | -2,59 | 9,62E-03 |
| RMDN1 | 1297,02 | -0,10 | 0,04 | -2,59 | 9,64E-03 |
| XYLT2 | 409,02 | 0,12 | 0,05 | 2,59 | 9,63E-03 |
| BLTP3B | 3309,03 | -0,13 | 0,05 | -2,59 | 9,63E-03 |
| GGT1 | 249,69 | 0,17 | 0,07 | 2,59 | 9,65E-03 |
| EXOSC7 | 639,74 | -0,09 | 0,03 | -2,59 | 9,66E-03 |
| RFX2 | 388,87 | 0,25 | 0,10 | 2,59 | 9,66E-03 |
| OSTF1 | 6249,11 | -0,13 | 0,05 | -2,59 | 9,67E-03 |
| CMTR2 | 1373,10 | -0,17 | 0,06 | -2,59 | 9,69E-03 |
| NUDT22 | 443,54 | 0,09 | 0,04 | 2,59 | 9,69E-03 |
| GLIPR1 | 8333,94 | -0,16 | 0,06 | -2,59 | 9,70E-03 |
| RGS14 | 1490,31 | 0,19 | 0,07 | 2,59 | 9,70E-03 |
| HLA-DOA | 881,77 | -0,19 | 0,07 | -2,59 | 9,72E-03 |
| LRMDA | 236,65 | 0,28 | 0,11 | 2,59 | 9,73E-03 |
| SUGT1 | 2348,47 | -0,08 | 0,03 | -2,58 | 9,75E-03 |
| NATD1 | 702,89 | 0,29 | 0,11 | 2,58 | 9,76E-03 |
| SLC35F3 | 55,60 | 0,34 | 0,13 | 2,58 | 9,76E-03 |
| GABPA | 1821,67 | -0,17 | 0,06 | -2,58 | 9,76E-03 |
| PPP1R7 | 879,51 | -0,07 | 0,03 | -2,58 | 9,76E-03 |
| MITF | 152,80 | 0,19 | 0,07 | 2,58 | 9,77E-03 |
| HDAC6 | 954,94 | 0,11 | 0,04 | 2,58 | 9,78E-03 |
| TASOR | 4781,53 | -0,11 | 0,04 | -2,58 | 9,77E-03 |
| MAFF | 188,90 | 0,16 | 0,06 | 2,58 | 9,78E-03 |
| SMG9 | 307,39 | 0,18 | 0,07 | 2,58 | 9,79E-03 |
| TMCO6 | 420,46 | 0,11 | 0,04 | 2,58 | 9,79E-03 |
| HDHD5 | 498,61 | 0,08 | 0,03 | 2,58 | 9,80E-03 |
| LRP8 | 338,47 | 0,14 | 0,05 | 2,58 | 9,81E-03 |
| FANCA | 334,15 | 0,25 | 0,10 | 2,58 | 9,82E-03 |
| PSMD14 | 649,79 | -0,18 | 0,07 | -2,58 | 9,85E-03 |

| | | | | | |
|---|---|---|---|---|---|
| DDX54 | 559,59 | 0,12 | 0,04 | 2,58 | 9,87E-03 |
| WDR81 | 982,11 | 0,15 | 0,06 | 2,58 | 9,87E-03 |
| SH3BP1 | 248,04 | 0,20 | 0,08 | 2,58 | 9,89E-03 |
| SEC22B | 3611,02 | -0,12 | 0,05 | -2,58 | 9,91E-03 |
| SRSF4 | 4158,28 | -0,07 | 0,03 | -2,58 | 9,92E-03 |
| ZFP14 | 819,03 | -0,14 | 0,05 | -2,58 | 9,93E-03 |
| TBX19 | 144,15 | 0,20 | 0,08 | 2,58 | 9,94E-03 |
| GGPS1 | 316,98 | -0,11 | 0,04 | -2,58 | 9,94E-03 |
| TNFRSF25 | 1085,50 | 0,16 | 0,06 | 2,58 | 9,95E-03 |
| ZNF347 | 519,20 | -0,14 | 0,05 | -2,58 | 9,97E-03 |
| MAP3K7 | 2357,06 | -0,11 | 0,04 | -2,58 | 9,97E-03 |
| MKNK2 | 2688,23 | 0,22 | 0,08 | 2,58 | 9,97E-03 |
| ZYX | 1383,68 | 0,30 | 0,12 | 2,58 | 9,98E-03 |
| LRBA | 7691,62 | -0,12 | 0,04 | -2,58 | 9,98E-03 |
| CCDC18 | 586,69 | -0,18 | 0,07 | -2,58 | 9,99E-03 |
| JRKL | 316,39 | -0,19 | 0,07 | -2,58 | 1,00E-02 |
| ENSG00000260729 | 19,59 | 0,25 | 0,10 | 2,58 | 1,00E-02 |
| TARBP2 | 172,25 | 0,11 | 0,04 | 2,58 | 1,00E-02 |
| RBM39 | 18047,95 | -0,12 | 0,05 | -2,57 | 1,00E-02 |
| CEBPE | 132,87 | 0,23 | 0,09 | 2,57 | 1,00E-02 |
| RNF8 | 436,24 | -0,09 | 0,04 | -2,57 | 1,00E-02 |
| ATP2A3 | 3765,07 | 0,19 | 0,07 | 2,57 | 1,01E-02 |
| MIX23 | 132,76 | -0,13 | 0,05 | -2,57 | 1,01E-02 |
| TNF | 85,39 | 0,16 | 0,06 | 2,57 | 1,01E-02 |
| CCL4 | 467,29 | -0,21 | 0,08 | -2,57 | 1,01E-02 |
| RANGAP1 | 684,02 | 0,14 | 0,05 | 2,57 | 1,01E-02 |
| SLC46A1 | 157,07 | 0,19 | 0,07 | 2,57 | 1,01E-02 |
| ZNF148 | 4436,48 | -0,13 | 0,05 | -2,57 | 1,01E-02 |
| CRIP2 | 109,74 | 0,30 | 0,12 | 2,57 | 1,01E-02 |
| RRP15 | 443,22 | -0,13 | 0,05 | -2,57 | 1,01E-02 |
| ENSG00000279765 | 909,44 | -0,11 | 0,04 | -2,57 | 1,01E-02 |
| DFFA | 1120,43 | -0,07 | 0,03 | -2,57 | 1,01E-02 |
| VEPH1 | 68,81 | 0,35 | 0,14 | 2,57 | 1,01E-02 |
| AKR1C8 | 32,99 | 0,34 | 0,13 | 2,57 | 1,02E-02 |
| SRP9 | 1847,86 | -0,21 | 0,08 | -2,57 | 1,02E-02 |
| ZNF880 | 227,48 | -0,14 | 0,06 | -2,57 | 1,02E-02 |
| RDX | 625,56 | -0,11 | 0,04 | -2,57 | 1,02E-02 |
| SP2 | 193,94 | 0,17 | 0,07 | 2,57 | 1,02E-02 |
| FAM72A | 31,79 | 0,23 | 0,09 | 2,57 | 1,02E-02 |
| CPSF1 | 643,99 | 0,23 | 0,09 | 2,57 | 1,02E-02 |
| ALDH8A1 | 72,23 | 0,23 | 0,09 | 2,57 | 1,02E-02 |
| GNPDA2 | 486,70 | -0,19 | 0,07 | -2,57 | 1,02E-02 |
| CLU | 377,40 | 0,31 | 0,12 | 2,57 | 1,02E-02 |
| S1PR5 | 1131,14 | -0,22 | 0,09 | -2,57 | 1,02E-02 |
| CSNK1G3 | 1174,38 | -0,18 | 0,07 | -2,57 | 1,02E-02 |
| SIRT2 | 347,48 | 0,18 | 0,07 | 2,57 | 1,02E-02 |

| | | | | | |
|---|---|---|---|---|---|
| PLD6 | 161,68 | 0,15 | 0,06 | 2,57 | 1,03E-02 |
| HADHA | 3747,38 | -0,09 | 0,03 | -2,57 | 1,03E-02 |
| ESD | 923,58 | -0,13 | 0,05 | -2,57 | 1,03E-02 |
| NDUFS8 | 256,20 | 0,13 | 0,05 | 2,57 | 1,03E-02 |
| LY6E | 478,25 | 0,35 | 0,14 | 2,57 | 1,03E-02 |
| SLC27A5 | 144,00 | 0,15 | 0,06 | 2,57 | 1,03E-02 |
| LDB2 | 46,20 | 0,42 | 0,16 | 2,57 | 1,03E-02 |
| ZNF292 | 7098,86 | -0,16 | 0,06 | -2,57 | 1,03E-02 |
| RANBP17 | 91,07 | 0,40 | 0,15 | 2,57 | 1,03E-02 |
| RTF2 | 4385,76 | -0,13 | 0,05 | -2,57 | 1,03E-02 |
| FCRLB | 38,09 | 0,26 | 0,10 | 2,57 | 1,03E-02 |
| DCTN3 | 682,71 | 0,08 | 0,03 | 2,56 | 1,03E-02 |
| METRN | 76,94 | 0,21 | 0,08 | 2,56 | 1,03E-02 |
| ITM2C | 196,51 | 0,18 | 0,07 | 2,56 | 1,03E-02 |
| IFNGR1 | 6205,26 | -0,18 | 0,07 | -2,56 | 1,03E-02 |
| CAPN1 | 1389,50 | 0,15 | 0,06 | 2,56 | 1,03E-02 |
| DIP2C | 315,30 | 0,23 | 0,09 | 2,56 | 1,04E-02 |
| JAK3 | 3232,44 | 0,24 | 0,09 | 2,56 | 1,04E-02 |
| CRYBG3 | 1190,95 | -0,12 | 0,05 | -2,56 | 1,04E-02 |
| POLR1E | 399,38 | -0,10 | 0,04 | -2,56 | 1,04E-02 |
| TMEM168 | 1845,26 | -0,13 | 0,05 | -2,56 | 1,04E-02 |
| ZNF302 | 937,40 | -0,17 | 0,07 | -2,56 | 1,04E-02 |
| CLIC5 | 184,32 | 0,20 | 0,08 | 2,56 | 1,04E-02 |
| SPATA6L | 95,34 | 0,26 | 0,10 | 2,56 | 1,04E-02 |
| WDR59 | 912,81 | 0,10 | 0,04 | 2,56 | 1,04E-02 |
| PPARA | 565,51 | 0,12 | 0,05 | 2,56 | 1,04E-02 |
| PCGF3 | 1637,18 | 0,12 | 0,05 | 2,56 | 1,04E-02 |
| CALU | 829,43 | -0,08 | 0,03 | -2,56 | 1,04E-02 |
| CLEC16A | 1360,73 | 0,14 | 0,06 | 2,56 | 1,05E-02 |
| MRPL32 | 470,28 | -0,19 | 0,08 | -2,56 | 1,05E-02 |
| WDR82 | 6138,37 | -0,08 | 0,03 | -2,56 | 1,05E-02 |
| WWP1 | 2266,55 | -0,11 | 0,04 | -2,56 | 1,05E-02 |
| WDR75 | 1350,88 | -0,13 | 0,05 | -2,56 | 1,05E-02 |
| TTC19 | 1117,88 | -0,08 | 0,03 | -2,56 | 1,05E-02 |
| CARD19 | 730,76 | 0,12 | 0,05 | 2,56 | 1,05E-02 |
| TEPSIN | 645,26 | 0,20 | 0,08 | 2,56 | 1,05E-02 |
| AK2 | 2870,85 | -0,07 | 0,03 | -2,56 | 1,05E-02 |
| ICE2 | 1966,63 | -0,11 | 0,04 | -2,56 | 1,05E-02 |
| EFR3A | 3585,66 | -0,12 | 0,05 | -2,56 | 1,05E-02 |
| DTX2 | 285,13 | 0,25 | 0,10 | 2,56 | 1,05E-02 |
| ZNF175 | 384,25 | -0,09 | 0,04 | -2,56 | 1,05E-02 |
| PROC | 34,73 | 0,36 | 0,14 | 2,56 | 1,05E-02 |
| UPK3A | 37,83 | 0,25 | 0,10 | 2,56 | 1,05E-02 |
| MRFAP1L2 | 661,00 | -0,12 | 0,05 | -2,56 | 1,05E-02 |
| PHOSPHO2 | 93,89 | -0,16 | 0,06 | -2,56 | 1,05E-02 |
| IRF3 | 1134,43 | 0,10 | 0,04 | 2,56 | 1,05E-02 |

| | | | | | |
|---|---|---|---|---|---|
| PPP1R9B | 2547,95 | 0,18 | 0,07 | 2,56 | 1,05E-02 |
| POMGNT2 | 83,08 | 0,16 | 0,06 | 2,56 | 1,05E-02 |
| ZNF557 | 1250,47 | -0,10 | 0,04 | -2,56 | 1,06E-02 |
| SZRD1 | 1493,74 | 0,11 | 0,04 | 2,56 | 1,06E-02 |
| MYL12A | 8298,44 | -0,13 | 0,05 | -2,56 | 1,06E-02 |
| FER | 794,70 | -0,12 | 0,05 | -2,56 | 1,06E-02 |
| PBLD | 103,80 | 0,15 | 0,06 | 2,56 | 1,06E-02 |
| THAP6 | 775,23 | -0,11 | 0,04 | -2,56 | 1,06E-02 |
| EHBP1 | 737,84 | -0,10 | 0,04 | -2,55 | 1,06E-02 |
| ARHGEF5 | 58,09 | 0,27 | 0,11 | 2,55 | 1,06E-02 |
| TFE3 | 945,75 | 0,19 | 0,07 | 2,55 | 1,07E-02 |
| TAS2R19 | 245,68 | -0,18 | 0,07 | -2,55 | 1,07E-02 |
| C17orf107 | 31,16 | 0,33 | 0,13 | 2,55 | 1,07E-02 |
| EXOSC2 | 906,72 | -0,08 | 0,03 | -2,55 | 1,07E-02 |
| RAP1GDS1 | 1243,35 | -0,11 | 0,04 | -2,55 | 1,07E-02 |
| PDLIM7 | 596,36 | 0,28 | 0,11 | 2,55 | 1,07E-02 |
| ORC5 | 650,45 | -0,15 | 0,06 | -2,55 | 1,07E-02 |
| AFAP1 | 369,63 | 0,28 | 0,11 | 2,55 | 1,07E-02 |
| CA1 | 2217,27 | 0,42 | 0,16 | 2,55 | 1,07E-02 |
| MTNAP1 | 470,86 | -0,10 | 0,04 | -2,55 | 1,07E-02 |
| WASHC4 | 10403,25 | -0,16 | 0,06 | -2,55 | 1,07E-02 |
| NEK1 | 774,89 | -0,11 | 0,04 | -2,55 | 1,07E-02 |
| ZBTB33 | 1112,93 | -0,18 | 0,07 | -2,55 | 1,08E-02 |
| BTG1 | 5381,01 | -0,13 | 0,05 | -2,55 | 1,08E-02 |
| SYNJ2 | 576,58 | 0,16 | 0,06 | 2,55 | 1,08E-02 |
| GADD45A | 289,28 | 0,16 | 0,06 | 2,55 | 1,08E-02 |
| BROX | 3320,16 | -0,12 | 0,05 | -2,55 | 1,08E-02 |
| USF2 | 818,55 | 0,19 | 0,07 | 2,55 | 1,08E-02 |
| ZDHHC17 | 4233,08 | -0,15 | 0,06 | -2,55 | 1,08E-02 |
| PPP2R1B | 1360,65 | -0,08 | 0,03 | -2,55 | 1,08E-02 |
| CARNMT1 | 786,02 | -0,15 | 0,06 | -2,55 | 1,08E-02 |
| TBC1D10B | 433,80 | 0,25 | 0,10 | 2,55 | 1,08E-02 |
| BNIPL | 78,12 | 0,17 | 0,07 | 2,55 | 1,08E-02 |
| ZNF432 | 457,28 | -0,17 | 0,07 | -2,55 | 1,08E-02 |
| ERBIN | 11728,57 | -0,13 | 0,05 | -2,55 | 1,09E-02 |
| PRRC2A | 1130,90 | 0,31 | 0,12 | 2,55 | 1,09E-02 |
| ACSL4 | 4730,59 | -0,16 | 0,06 | -2,55 | 1,09E-02 |
| SRSF7 | 3784,71 | -0,12 | 0,05 | -2,55 | 1,09E-02 |
| ENSG00000257341 | 22,15 | 0,21 | 0,08 | 2,55 | 1,09E-02 |
| ITGA2B | 696,70 | 0,36 | 0,14 | 2,55 | 1,09E-02 |
| TUG1 | 8090,19 | -0,09 | 0,04 | -2,55 | 1,09E-02 |
| EXOSC1 | 684,55 | -0,09 | 0,03 | -2,55 | 1,09E-02 |
| SPART | 1607,41 | -0,13 | 0,05 | -2,55 | 1,09E-02 |
| ABHD5 | 7136,83 | -0,19 | 0,07 | -2,55 | 1,09E-02 |
| GP1BA | 28,14 | 0,42 | 0,16 | 2,55 | 1,09E-02 |
| ATP9A | 317,08 | 0,22 | 0,09 | 2,54 | 1,09E-02 |

| | | | | | |
|---|---|---|---|---|---|
| PLA2G4B | 54,92 | 0,23 | 0,09 | 2,54 | 1,09E-02 |
| CPT1B | 56,14 | 0,28 | 0,11 | 2,54 | 1,10E-02 |
| LDHA | 2203,26 | -0,09 | 0,04 | -2,54 | 1,10E-02 |
| CIMAP1B | 570,87 | 0,26 | 0,10 | 2,54 | 1,10E-02 |
| TOR1AIP2 | 4053,47 | -0,07 | 0,03 | -2,54 | 1,10E-02 |
| PTGES2 | 387,00 | 0,11 | 0,04 | 2,54 | 1,10E-02 |
| IFRD1 | 6150,14 | -0,17 | 0,07 | -2,54 | 1,10E-02 |
| SURF1 | 496,65 | 0,07 | 0,03 | 2,54 | 1,11E-02 |
| TMEM126B | 523,18 | -0,15 | 0,06 | -2,54 | 1,11E-02 |
| CISD1 | 231,60 | -0,18 | 0,07 | -2,54 | 1,11E-02 |
| UTP25 | 846,61 | -0,09 | 0,04 | -2,54 | 1,11E-02 |
| ATXN2L | 720,85 | 0,24 | 0,09 | 2,54 | 1,11E-02 |
| CD69 | 379,43 | -0,27 | 0,11 | -2,54 | 1,11E-02 |
| CFAP20 | 382,02 | -0,09 | 0,03 | -2,54 | 1,11E-02 |
| ANKH | 1574,71 | 0,18 | 0,07 | 2,54 | 1,11E-02 |
| HDHD2 | 600,40 | -0,10 | 0,04 | -2,54 | 1,11E-02 |
| CCDC180 | 103,46 | 0,27 | 0,11 | 2,54 | 1,11E-02 |
| ABCG2 | 149,12 | 0,29 | 0,11 | 2,54 | 1,12E-02 |
| ZNF70 | 204,30 | 0,17 | 0,07 | 2,54 | 1,12E-02 |
| CLYBL | 226,35 | 0,15 | 0,06 | 2,54 | 1,12E-02 |
| ZNF443 | 145,17 | -0,20 | 0,08 | -2,54 | 1,12E-02 |
| TRMT12 | 542,58 | -0,14 | 0,06 | -2,54 | 1,12E-02 |
| ENPP5 | 126,70 | -0,22 | 0,09 | -2,54 | 1,12E-02 |
| TG | 353,32 | 0,24 | 0,10 | 2,54 | 1,12E-02 |
| SGSH | 378,05 | 0,30 | 0,12 | 2,54 | 1,12E-02 |
| PRELID3B | 1291,71 | -0,15 | 0,06 | -2,54 | 1,12E-02 |
| KLHL7 | 690,50 | -0,09 | 0,04 | -2,54 | 1,12E-02 |
| TTC24 | 52,14 | 0,27 | 0,11 | 2,54 | 1,12E-02 |
| SLC24A1 | 161,25 | 0,19 | 0,07 | 2,53 | 1,12E-02 |
| ABCC1 | 1230,37 | 0,19 | 0,08 | 2,53 | 1,13E-02 |
| GABRR2 | 77,79 | 0,24 | 0,10 | 2,53 | 1,13E-02 |
| GTPBP2 | 937,19 | 0,18 | 0,07 | 2,53 | 1,13E-02 |
| SYNPO2 | 79,33 | 0,33 | 0,13 | 2,53 | 1,13E-02 |
| CAPZA2 | 4687,08 | -0,22 | 0,09 | -2,53 | 1,13E-02 |
| CSNK1E | 300,35 | 0,14 | 0,05 | 2,53 | 1,13E-02 |
| RASAL3 | 877,39 | 0,23 | 0,09 | 2,53 | 1,14E-02 |
| ZNF142 | 350,69 | 0,21 | 0,08 | 2,53 | 1,14E-02 |
| HAPSTR1 | 10289,65 | -0,10 | 0,04 | -2,53 | 1,14E-02 |
| ATP2C1 | 2240,41 | -0,09 | 0,04 | -2,53 | 1,14E-02 |
| TAF9 | 393,69 | -0,14 | 0,05 | -2,53 | 1,14E-02 |
| SOAT1 | 2931,14 | -0,09 | 0,03 | -2,53 | 1,14E-02 |
| SUFU | 246,63 | 0,21 | 0,08 | 2,53 | 1,14E-02 |
| DBF4 | 382,56 | -0,14 | 0,05 | -2,53 | 1,14E-02 |
| SIPA1L3 | 582,13 | 0,22 | 0,09 | 2,53 | 1,14E-02 |
| NBR1 | 5195,17 | -0,09 | 0,03 | -2,53 | 1,14E-02 |
| C4orf50 | 177,83 | 0,39 | 0,16 | 2,53 | 1,14E-02 |

| | | | | | |
|---|---|---|---|---|---|
| PRKAR2A | 1490,55 | -0,05 | 0,02 | -2,53 | 1,14E-02 |
| PIM1 | 12663,54 | 0,22 | 0,09 | 2,53 | 1,14E-02 |
| NUP50 | 5786,31 | -0,11 | 0,04 | -2,53 | 1,14E-02 |
| TUT1 | 94,29 | 0,16 | 0,06 | 2,53 | 1,15E-02 |
| MGST1 | 166,96 | 0,17 | 0,07 | 2,53 | 1,15E-02 |
| LMF2 | 616,82 | 0,17 | 0,07 | 2,53 | 1,15E-02 |
| PLEK2 | 452,76 | 0,30 | 0,12 | 2,53 | 1,15E-02 |
| SPIDR | 1973,76 | -0,08 | 0,03 | -2,53 | 1,15E-02 |
| NAA16 | 1072,76 | -0,11 | 0,04 | -2,53 | 1,15E-02 |
| TBC1D17 | 462,69 | 0,18 | 0,07 | 2,53 | 1,15E-02 |
| YEATS4 | 223,02 | -0,19 | 0,07 | -2,53 | 1,15E-02 |
| ENPP4 | 749,97 | -0,23 | 0,09 | -2,53 | 1,16E-02 |
| DFFB | 439,36 | 0,07 | 0,03 | 2,53 | 1,16E-02 |
| ACP5 | 330,67 | 0,18 | 0,07 | 2,52 | 1,16E-02 |
| MYCT1 | 73,64 | 0,22 | 0,09 | 2,52 | 1,16E-02 |
| SETD1B | 779,98 | 0,30 | 0,12 | 2,52 | 1,16E-02 |
| AGTRAP | 896,24 | 0,17 | 0,07 | 2,52 | 1,16E-02 |
| ARSA | 234,11 | 0,32 | 0,13 | 2,52 | 1,16E-02 |
| PIM2 | 1546,58 | 0,14 | 0,06 | 2,52 | 1,16E-02 |
| KRTCAP3 | 42,57 | 0,23 | 0,09 | 2,52 | 1,16E-02 |
| EMC1 | 606,46 | 0,12 | 0,05 | 2,52 | 1,16E-02 |
| OTUD6B | 709,87 | -0,17 | 0,07 | -2,52 | 1,16E-02 |
| RECQL5 | 517,94 | 0,11 | 0,04 | 2,52 | 1,16E-02 |
| RMND1 | 371,69 | -0,09 | 0,03 | -2,52 | 1,16E-02 |
| USPL1 | 1238,68 | -0,10 | 0,04 | -2,52 | 1,17E-02 |
| RAB14 | 2745,45 | -0,10 | 0,04 | -2,52 | 1,17E-02 |
| SLC25A37 | 72992,32 | 0,22 | 0,09 | 2,52 | 1,17E-02 |
| NT5E | 231,56 | -0,23 | 0,09 | -2,52 | 1,17E-02 |
| LRRC7 | 67,59 | 0,51 | 0,20 | 2,52 | 1,17E-02 |
| MAP3K9 | 176,87 | 0,25 | 0,10 | 2,52 | 1,17E-02 |
| CCT4 | 1766,51 | -0,12 | 0,05 | -2,52 | 1,17E-02 |
| PLD3 | 479,59 | 0,18 | 0,07 | 2,52 | 1,17E-02 |
| RNF145 | 3725,05 | -0,09 | 0,04 | -2,52 | 1,17E-02 |
| MYL12B | 4058,29 | -0,16 | 0,06 | -2,52 | 1,18E-02 |
| ZFP90 | 1119,57 | -0,11 | 0,04 | -2,52 | 1,18E-02 |
| SNRNP27 | 301,63 | -0,12 | 0,05 | -2,52 | 1,18E-02 |
| FBXL6 | 155,01 | 0,17 | 0,07 | 2,52 | 1,18E-02 |
| DPY30 | 330,37 | -0,15 | 0,06 | -2,52 | 1,18E-02 |
| KHK | 64,52 | 0,19 | 0,08 | 2,52 | 1,18E-02 |
| SERPINF2 | 45,93 | 0,25 | 0,10 | 2,52 | 1,18E-02 |
| CAMSAP1 | 844,43 | 0,07 | 0,03 | 2,52 | 1,18E-02 |
| ADA | 325,16 | 0,12 | 0,05 | 2,52 | 1,18E-02 |
| CACTIN | 436,70 | 0,16 | 0,06 | 2,52 | 1,18E-02 |
| NDUFC2 | 390,22 | -0,13 | 0,05 | -2,52 | 1,18E-02 |
| GPBAR1 | 193,44 | 0,16 | 0,07 | 2,52 | 1,18E-02 |
| AXIN2 | 234,78 | 0,19 | 0,08 | 2,52 | 1,18E-02 |

| | | | | | |
|---|---|---|---|---|---|
| FIG4 | 1490,58 | -0,11 | 0,04 | -2,52 | 1,19E-02 |
| TAPBP | 2832,95 | 0,23 | 0,09 | 2,52 | 1,19E-02 |
| MAPK8IP3 | 2248,43 | 0,18 | 0,07 | 2,52 | 1,19E-02 |
| COL9A2 | 185,70 | 0,26 | 0,10 | 2,52 | 1,19E-02 |
| AGER | 248,15 | 0,26 | 0,10 | 2,52 | 1,19E-02 |
| GSS | 308,02 | 0,09 | 0,04 | 2,52 | 1,19E-02 |
| SRSF6 | 4028,75 | -0,09 | 0,04 | -2,52 | 1,19E-02 |
| KLHL28 | 2075,39 | -0,14 | 0,06 | -2,52 | 1,19E-02 |
| FERMT3 | 1640,60 | 0,18 | 0,07 | 2,51 | 1,19E-02 |
| PPP3CA | 3424,31 | -0,10 | 0,04 | -2,51 | 1,19E-02 |
| SNX14 | 2399,09 | -0,11 | 0,04 | -2,51 | 1,19E-02 |
| CDKN2C | 134,23 | 0,17 | 0,07 | 2,51 | 1,19E-02 |
| ATP6V1E1 | 1778,51 | -0,13 | 0,05 | -2,51 | 1,19E-02 |
| PPIL1 | 181,25 | -0,11 | 0,05 | -2,51 | 1,20E-02 |
| GEMIN6 | 276,37 | -0,12 | 0,05 | -2,51 | 1,20E-02 |
| MFNG | 965,42 | 0,15 | 0,06 | 2,51 | 1,20E-02 |
| SLC38A1 | 10313,58 | -0,12 | 0,05 | -2,51 | 1,20E-02 |
| DNAJA2 | 2265,98 | -0,13 | 0,05 | -2,51 | 1,20E-02 |
| DGKZ | 2059,40 | 0,22 | 0,09 | 2,51 | 1,20E-02 |
| TCN2 | 71,84 | 0,27 | 0,11 | 2,51 | 1,20E-02 |
| FBXO31 | 354,70 | 0,16 | 0,06 | 2,51 | 1,21E-02 |
| POLQ | 140,96 | 0,23 | 0,09 | 2,51 | 1,21E-02 |
| FBXO39 | 33,69 | 0,36 | 0,14 | 2,51 | 1,21E-02 |
| ZNF141 | 1058,85 | -0,19 | 0,08 | -2,51 | 1,21E-02 |
| C11orf65 | 32,39 | 0,28 | 0,11 | 2,51 | 1,21E-02 |
| ACKR1 | 37,49 | 0,37 | 0,15 | 2,51 | 1,21E-02 |
| TMEM129 | 323,99 | 0,15 | 0,06 | 2,51 | 1,21E-02 |
| POT1 | 668,48 | -0,17 | 0,07 | -2,51 | 1,21E-02 |
| DENND2C | 120,67 | 0,24 | 0,10 | 2,51 | 1,21E-02 |
| TECPR1 | 1898,17 | 0,13 | 0,05 | 2,51 | 1,21E-02 |
| DSTN | 542,93 | -0,09 | 0,04 | -2,51 | 1,21E-02 |
| NRIP1 | 1649,10 | -0,14 | 0,05 | -2,51 | 1,22E-02 |
| ERGIC3 | 755,84 | 0,11 | 0,04 | 2,51 | 1,22E-02 |
| SNX18 | 6032,38 | -0,12 | 0,05 | -2,51 | 1,22E-02 |
| ARHGAP5 | 1916,98 | -0,19 | 0,08 | -2,51 | 1,22E-02 |
| PMPCA | 425,90 | 0,10 | 0,04 | 2,51 | 1,22E-02 |
| MYO9A | 2311,43 | -0,09 | 0,04 | -2,51 | 1,22E-02 |
| TMEM241 | 125,03 | 0,20 | 0,08 | 2,51 | 1,22E-02 |
| TBC1D9B | 844,02 | 0,21 | 0,09 | 2,51 | 1,22E-02 |
| FAM174C | 187,38 | 0,11 | 0,05 | 2,51 | 1,22E-02 |
| KRBOX5 | 888,79 | -0,12 | 0,05 | -2,51 | 1,22E-02 |
| SPI1 | 1117,36 | 0,24 | 0,10 | 2,51 | 1,22E-02 |
| ANKRD13D | 2885,15 | 0,17 | 0,07 | 2,50 | 1,23E-02 |
| PRLR | 136,72 | 0,24 | 0,10 | 2,51 | 1,22E-02 |
| SREBF1 | 404,91 | 0,25 | 0,10 | 2,50 | 1,22E-02 |
| FAM110A | 374,71 | 0,16 | 0,06 | 2,50 | 1,22E-02 |

| | | | | | |
|---|---|---|---|---|---|
| SHPRH | 3245,34 | -0,11 | 0,05 | -2,50 | 1,23E-02 |
| DNAJC1 | 697,61 | -0,10 | 0,04 | -2,50 | 1,23E-02 |
| KIFAP3 | 921,29 | -0,14 | 0,05 | -2,50 | 1,23E-02 |
| RMI2 | 60,15 | 0,34 | 0,13 | 2,50 | 1,23E-02 |
| LTBR | 574,54 | 0,19 | 0,08 | 2,50 | 1,23E-02 |
| LRIT3 | 24,63 | 0,26 | 0,10 | 2,50 | 1,23E-02 |
| MTMR11 | 330,20 | 0,14 | 0,06 | 2,50 | 1,23E-02 |
| MAGI3 | 170,47 | 0,18 | 0,07 | 2,50 | 1,23E-02 |
| MELK | 54,27 | 0,31 | 0,12 | 2,50 | 1,23E-02 |
| THEM6 | 235,32 | 0,12 | 0,05 | 2,50 | 1,23E-02 |
| COMMD7 | 98,48 | 0,12 | 0,05 | 2,50 | 1,23E-02 |
| NIP7 | 619,35 | -0,10 | 0,04 | -2,50 | 1,24E-02 |
| FANCG | 167,56 | 0,12 | 0,05 | 2,50 | 1,24E-02 |
| NACC1 | 267,21 | 0,22 | 0,09 | 2,50 | 1,24E-02 |
| GTF2H1 | 1426,41 | -0,08 | 0,03 | -2,50 | 1,24E-02 |
| MALSU1 | 431,40 | -0,08 | 0,03 | -2,50 | 1,24E-02 |
| FAM204A | 2044,65 | -0,09 | 0,04 | -2,50 | 1,24E-02 |
| TMEM11 | 242,82 | 0,11 | 0,04 | 2,50 | 1,24E-02 |
| RNF44 | 879,28 | 0,20 | 0,08 | 2,50 | 1,25E-02 |
| WDR41 | 1155,75 | -0,10 | 0,04 | -2,50 | 1,25E-02 |
| VMAC | 317,97 | 0,13 | 0,05 | 2,50 | 1,25E-02 |
| DMTF1 | 4786,79 | -0,08 | 0,03 | -2,50 | 1,25E-02 |
| ALOX15 | 300,52 | 0,41 | 0,17 | 2,50 | 1,25E-02 |
| CERT1 | 4699,77 | -0,09 | 0,04 | -2,50 | 1,25E-02 |
| LOXL3 | 210,02 | 0,17 | 0,07 | 2,50 | 1,25E-02 |
| DNAJB14 | 4680,04 | -0,18 | 0,07 | -2,50 | 1,24E-02 |
| B3GALNT2 | 368,38 | -0,13 | 0,05 | -2,50 | 1,25E-02 |
| CSTB | 433,81 | 0,12 | 0,05 | 2,50 | 1,25E-02 |
| SLC10A3 | 181,76 | 0,15 | 0,06 | 2,50 | 1,25E-02 |
| LUC7L3 | 5665,24 | -0,12 | 0,05 | -2,50 | 1,25E-02 |
| TMEM222 | 324,60 | 0,12 | 0,05 | 2,50 | 1,25E-02 |
| CIPC | 471,51 | -0,07 | 0,03 | -2,50 | 1,25E-02 |
| SEC61B | 986,79 | -0,12 | 0,05 | -2,50 | 1,25E-02 |
| ZNF526 | 163,87 | 0,19 | 0,07 | 2,50 | 1,25E-02 |
| SAMD4A | 308,23 | 0,24 | 0,10 | 2,50 | 1,26E-02 |
| RHBDD3 | 183,88 | 0,11 | 0,04 | 2,50 | 1,26E-02 |
| MED8 | 772,47 | -0,06 | 0,02 | -2,50 | 1,26E-02 |
| AP3S1 | 781,70 | -0,13 | 0,05 | -2,50 | 1,26E-02 |
| TP53 | 481,29 | 0,14 | 0,06 | 2,50 | 1,26E-02 |
| HMGB2 | 2025,05 | -0,24 | 0,10 | -2,50 | 1,26E-02 |
| VPS72 | 495,63 | -0,06 | 0,03 | -2,50 | 1,26E-02 |
| LAMP5 | 29,01 | 0,31 | 0,13 | 2,49 | 1,26E-02 |
| SUPV3L1 | 565,29 | -0,08 | 0,03 | -2,49 | 1,26E-02 |
| RHBDD2 | 258,21 | 0,15 | 0,06 | 2,49 | 1,26E-02 |
| GSTZ1 | 304,13 | 0,10 | 0,04 | 2,49 | 1,26E-02 |
| TINF2 | 1998,01 | -0,06 | 0,03 | -2,49 | 1,26E-02 |

| | | | | | |
|---|---|---|---|---|---|
| RO60 | 4571,32 | -0,14 | 0,06 | -2,49 | 1,27E-02 |
| PDE6D | 529,74 | -0,05 | 0,02 | -2,49 | 1,27E-02 |
| POM121C | 540,54 | 0,20 | 0,08 | 2,49 | 1,27E-02 |
| REV1 | 1843,85 | -0,08 | 0,03 | -2,49 | 1,27E-02 |
| CHORDC1 | 2629,79 | -0,14 | 0,06 | -2,49 | 1,27E-02 |
| SBNO2 | 2850,96 | 0,25 | 0,10 | 2,49 | 1,27E-02 |
| ODAD4 | 55,79 | 0,32 | 0,13 | 2,49 | 1,27E-02 |
| CFAP53 | 36,36 | 0,27 | 0,11 | 2,49 | 1,27E-02 |
| LYSET | 158,71 | -0,12 | 0,05 | -2,49 | 1,27E-02 |
| EPN2 | 295,55 | 0,19 | 0,08 | 2,49 | 1,28E-02 |
| ASCC2 | 4705,46 | 0,26 | 0,10 | 2,49 | 1,28E-02 |
| SUZ12 | 1270,68 | -0,15 | 0,06 | -2,49 | 1,28E-02 |
| NFIL3 | 784,23 | -0,18 | 0,07 | -2,49 | 1,28E-02 |
| ARPC2 | 10312,01 | -0,10 | 0,04 | -2,49 | 1,28E-02 |
| MAPK1IP1L | 1643,09 | -0,07 | 0,03 | -2,49 | 1,28E-02 |
| ABRAXAS1 | 827,24 | -0,12 | 0,05 | -2,49 | 1,28E-02 |
| NUDCD1 | 446,73 | -0,23 | 0,09 | -2,49 | 1,28E-02 |
| PSME3IP1 | 3181,16 | -0,07 | 0,03 | -2,49 | 1,28E-02 |
| AP2S1 | 1087,41 | 0,15 | 0,06 | 2,49 | 1,29E-02 |
| RAG1 | 52,33 | 0,30 | 0,12 | 2,49 | 1,29E-02 |
| ORC3 | 618,13 | -0,10 | 0,04 | -2,49 | 1,29E-02 |
| RNF11 | 6324,94 | 0,26 | 0,11 | 2,49 | 1,29E-02 |
| NSD2 | 1511,70 | 0,10 | 0,04 | 2,49 | 1,29E-02 |
| CBFB | 2229,25 | -0,10 | 0,04 | -2,49 | 1,29E-02 |
| GOLGA8H | 34,26 | 0,27 | 0,11 | 2,49 | 1,29E-02 |
| SKA2 | 263,64 | -0,15 | 0,06 | -2,49 | 1,29E-02 |
| ALG12 | 403,09 | 0,10 | 0,04 | 2,48 | 1,30E-02 |
| ABI1 | 5310,62 | -0,12 | 0,05 | -2,48 | 1,30E-02 |
| CALHM2 | 400,34 | 0,15 | 0,06 | 2,48 | 1,30E-02 |
| EPC1 | 4399,80 | -0,09 | 0,04 | -2,48 | 1,30E-02 |
| TIGD1 | 459,78 | -0,19 | 0,08 | -2,48 | 1,30E-02 |
| MNDA | 30718,36 | -0,19 | 0,08 | -2,48 | 1,30E-02 |
| TNIP1 | 3944,83 | 0,14 | 0,06 | 2,48 | 1,30E-02 |
| EBLN2 | 313,87 | -0,09 | 0,03 | -2,48 | 1,30E-02 |
| TPX2 | 74,23 | 0,25 | 0,10 | 2,48 | 1,30E-02 |
| ARL4C | 4218,10 | -0,11 | 0,05 | -2,48 | 1,30E-02 |
| ICA1 | 130,28 | 0,16 | 0,07 | 2,48 | 1,31E-02 |
| PNKP | 996,76 | 0,16 | 0,06 | 2,48 | 1,31E-02 |
| THBS3 | 497,22 | 0,12 | 0,05 | 2,48 | 1,31E-02 |
| CDCA7 | 74,79 | 0,24 | 0,10 | 2,48 | 1,31E-02 |
| BCLAF3 | 1226,49 | -0,10 | 0,04 | -2,48 | 1,31E-02 |
| DAP | 653,11 | 0,18 | 0,07 | 2,48 | 1,31E-02 |
| ZNF81 | 1132,93 | -0,08 | 0,03 | -2,48 | 1,31E-02 |
| LAMP1 | 2706,14 | -0,12 | 0,05 | -2,48 | 1,31E-02 |
| PIAS4 | 477,72 | 0,12 | 0,05 | 2,48 | 1,31E-02 |
| RNF141 | 4024,15 | -0,15 | 0,06 | -2,48 | 1,32E-02 |

| | | | | | |
|---|---|---|---|---|---|
| STK38L | 2875,02 | -0,14 | 0,06 | -2,48 | 1,32E-02 |
| LAMTOR3 | 1096,38 | -0,18 | 0,07 | -2,48 | 1,32E-02 |
| OR4D1 | 50,08 | 0,22 | 0,09 | 2,48 | 1,32E-02 |
| ALDH3B1 | 545,08 | 0,17 | 0,07 | 2,48 | 1,32E-02 |
| LAX1 | 738,30 | -0,12 | 0,05 | -2,48 | 1,32E-02 |
| MAPKAP1 | 2059,13 | -0,06 | 0,02 | -2,48 | 1,32E-02 |
| RNF169 | 5688,46 | -0,10 | 0,04 | -2,48 | 1,32E-02 |
| SOCS4 | 1387,57 | -0,16 | 0,07 | -2,48 | 1,32E-02 |
| TP53I3 | 130,02 | 0,16 | 0,07 | 2,48 | 1,32E-02 |
| ZKSCAN1 | 2798,02 | -0,05 | 0,02 | -2,48 | 1,33E-02 |
| EDARADD | 57,14 | 0,25 | 0,10 | 2,48 | 1,33E-02 |
| SUPT7L | 1194,51 | -0,09 | 0,04 | -2,48 | 1,33E-02 |
| C11orf24 | 170,03 | 0,13 | 0,05 | 2,48 | 1,33E-02 |
| YPEL5 | 3556,71 | -0,15 | 0,06 | -2,47 | 1,33E-02 |
| IREB2 | 2914,17 | -0,08 | 0,03 | -2,47 | 1,33E-02 |
| RAPGEFL1 | 132,49 | 0,31 | 0,13 | 2,47 | 1,33E-02 |
| RPL5 | 7512,87 | -0,20 | 0,08 | -2,47 | 1,33E-02 |
| NUP205 | 2263,48 | -0,09 | 0,03 | -2,47 | 1,34E-02 |
| RAB8B | 7032,27 | -0,16 | 0,07 | -2,47 | 1,34E-02 |
| PCED1A | 568,10 | 0,10 | 0,04 | 2,47 | 1,34E-02 |
| FNDC3A | 2836,89 | -0,09 | 0,04 | -2,47 | 1,34E-02 |
| HGH1 | 140,55 | 0,15 | 0,06 | 2,47 | 1,34E-02 |
| FAM118B | 575,05 | -0,10 | 0,04 | -2,47 | 1,34E-02 |
| TNPO3 | 3053,84 | -0,08 | 0,03 | -2,47 | 1,34E-02 |
| WDR11 | 2945,23 | -0,09 | 0,04 | -2,47 | 1,34E-02 |
| COG6 | 833,01 | -0,15 | 0,06 | -2,47 | 1,34E-02 |
| BUB1B | 57,36 | 0,25 | 0,10 | 2,47 | 1,34E-02 |
| ZGRF1 | 461,04 | -0,13 | 0,05 | -2,47 | 1,34E-02 |
| METTL25 | 600,95 | -0,12 | 0,05 | -2,47 | 1,34E-02 |
| SZT2 | 2148,96 | 0,17 | 0,07 | 2,47 | 1,34E-02 |
| VPS50 | 1660,19 | -0,16 | 0,07 | -2,47 | 1,34E-02 |
| FASTKD3 | 270,93 | -0,10 | 0,04 | -2,47 | 1,35E-02 |
| UQCRHL | 42,03 | 0,18 | 0,07 | 2,47 | 1,35E-02 |
| ZNF225 | 435,30 | -0,10 | 0,04 | -2,47 | 1,35E-02 |
| TNK2 | 1993,25 | 0,24 | 0,10 | 2,47 | 1,35E-02 |
| GIGYF2 | 4765,92 | -0,08 | 0,03 | -2,47 | 1,35E-02 |
| SUMO2 | 2536,67 | -0,13 | 0,05 | -2,47 | 1,35E-02 |
| ENSG00000261732 | 50,71 | 0,15 | 0,06 | 2,47 | 1,35E-02 |
| PER3 | 223,84 | 0,16 | 0,07 | 2,47 | 1,35E-02 |
| TLE5 | 1158,81 | 0,16 | 0,07 | 2,47 | 1,35E-02 |
| COPS8 | 764,44 | -0,13 | 0,05 | -2,47 | 1,35E-02 |
| TRAF2 | 425,71 | 0,10 | 0,04 | 2,47 | 1,36E-02 |
| CTDSPL2 | 2398,46 | -0,12 | 0,05 | -2,47 | 1,36E-02 |
| PLXDC1 | 162,64 | 0,26 | 0,10 | 2,47 | 1,36E-02 |
| CASP3 | 1145,07 | -0,18 | 0,07 | -2,47 | 1,36E-02 |
| PCBP2 | 4686,55 | 0,12 | 0,05 | 2,47 | 1,36E-02 |

| | | | | | |
|---|---|---|---|---|---|
| EIF2A | 1500,15 | -0,16 | 0,06 | -2,47 | 1,36E-02 |
| PPP1R18 | 1510,99 | 0,25 | 0,10 | 2,47 | 1,37E-02 |
| ITGB5 | 382,46 | 0,27 | 0,11 | 2,47 | 1,37E-02 |
| TSTD2 | 620,28 | -0,07 | 0,03 | -2,47 | 1,37E-02 |
| PAIP1 | 750,44 | -0,12 | 0,05 | -2,47 | 1,37E-02 |
| ZBTB26 | 431,21 | -0,11 | 0,04 | -2,46 | 1,37E-02 |
| ZNF570 | 390,96 | -0,14 | 0,06 | -2,46 | 1,38E-02 |
| SRRT | 498,85 | 0,19 | 0,08 | 2,46 | 1,38E-02 |
| DHODH | 244,12 | 0,12 | 0,05 | 2,46 | 1,38E-02 |
| PEX11B | 414,34 | -0,08 | 0,03 | -2,46 | 1,38E-02 |
| RELA | 615,29 | 0,16 | 0,06 | 2,46 | 1,38E-02 |
| NR1D2 | 1619,20 | -0,14 | 0,06 | -2,46 | 1,38E-02 |
| RPS20 | 8839,63 | -0,24 | 0,10 | -2,46 | 1,38E-02 |
| ARF5 | 515,76 | 0,11 | 0,04 | 2,46 | 1,38E-02 |
| ZNF317 | 1313,12 | -0,05 | 0,02 | -2,46 | 1,38E-02 |
| ECI2 | 554,43 | -0,08 | 0,03 | -2,46 | 1,39E-02 |
| ZNF439 | 726,24 | -0,13 | 0,05 | -2,46 | 1,39E-02 |
| FBXW11 | 2226,12 | -0,07 | 0,03 | -2,46 | 1,39E-02 |
| CFAP161 | 67,30 | 0,21 | 0,08 | 2,46 | 1,39E-02 |
| ADAM12 | 106,95 | 0,37 | 0,15 | 2,46 | 1,39E-02 |
| MAPRE2 | 1743,18 | -0,07 | 0,03 | -2,46 | 1,39E-02 |
| SLC29A3 | 117,96 | 0,17 | 0,07 | 2,46 | 1,39E-02 |
| CLK4 | 4025,91 | -0,14 | 0,05 | -2,46 | 1,39E-02 |
| THOC7 | 316,71 | -0,13 | 0,05 | -2,46 | 1,39E-02 |
| C2orf92 | 141,58 | 0,18 | 0,07 | 2,46 | 1,39E-02 |
| IRGQ | 542,02 | 0,09 | 0,04 | 2,46 | 1,40E-02 |
| SYCE1L | 42,99 | 0,19 | 0,08 | 2,46 | 1,40E-02 |
| SPCS3 | 7485,81 | -0,14 | 0,06 | -2,46 | 1,40E-02 |
| BPHL | 178,01 | 0,11 | 0,05 | 2,46 | 1,40E-02 |
| MINK1 | 634,42 | 0,27 | 0,11 | 2,46 | 1,40E-02 |
| GFOD2 | 889,88 | 0,14 | 0,06 | 2,46 | 1,40E-02 |
| EIF4H | 5210,45 | -0,07 | 0,03 | -2,46 | 1,40E-02 |
| TMA7 | 753,19 | -0,32 | 0,13 | -2,46 | 1,40E-02 |
| CHD1L | 1020,68 | -0,07 | 0,03 | -2,46 | 1,40E-02 |
| EEF1A1 | 123252,90 | -0,21 | 0,09 | -2,46 | 1,40E-02 |
| SMARCD1 | 604,88 | 0,09 | 0,04 | 2,46 | 1,40E-02 |
| EPHX2 | 432,37 | 0,18 | 0,08 | 2,46 | 1,41E-02 |
| MIEF1 | 570,34 | 0,10 | 0,04 | 2,46 | 1,41E-02 |
| MFSD10 | 441,93 | 0,14 | 0,06 | 2,45 | 1,41E-02 |
| INTS13 | 590,11 | -0,11 | 0,04 | -2,45 | 1,42E-02 |
| MAP4K2 | 789,05 | 0,18 | 0,07 | 2,45 | 1,42E-02 |
| ZNF267 | 2972,95 | -0,17 | 0,07 | -2,45 | 1,42E-02 |
| ENSG00000124593 | 28,46 | 0,18 | 0,07 | 2,45 | 1,42E-02 |
| SEC23IP | 978,54 | -0,06 | 0,02 | -2,45 | 1,42E-02 |
| ARL14EP | 1283,40 | -0,17 | 0,07 | -2,45 | 1,42E-02 |
| NOTCH1 | 3930,87 | 0,28 | 0,11 | 2,45 | 1,42E-02 |

| | | | | | |
|---|---|---|---|---|---|
| RHOBTB2 | 224,53 | 0,19 | 0,08 | 2,45 | 1,43E-02 |
| UFM1 | 1754,15 | -0,20 | 0,08 | -2,45 | 1,43E-02 |
| SSBP1 | 1327,33 | -0,11 | 0,05 | -2,45 | 1,43E-02 |
| PIGS | 569,14 | 0,10 | 0,04 | 2,45 | 1,43E-02 |
| CTNNBL1 | 1225,30 | -0,07 | 0,03 | -2,45 | 1,43E-02 |
| ARHGAP1 | 609,79 | 0,21 | 0,08 | 2,45 | 1,43E-02 |
| NLRC5 | 5578,75 | 0,21 | 0,08 | 2,45 | 1,43E-02 |
| PPP6C | 1797,75 | -0,09 | 0,04 | -2,45 | 1,43E-02 |
| BOLA3 | 79,88 | 0,15 | 0,06 | 2,45 | 1,43E-02 |
| TMX1 | 1690,45 | -0,21 | 0,08 | -2,45 | 1,44E-02 |
| MAP3K13 | 173,46 | 0,18 | 0,07 | 2,45 | 1,44E-02 |
| APOBEC3F | 153,86 | 0,20 | 0,08 | 2,45 | 1,44E-02 |
| GINS4 | 90,19 | 0,17 | 0,07 | 2,45 | 1,44E-02 |
| PHF10 | 1365,78 | -0,12 | 0,05 | -2,45 | 1,44E-02 |
| SRSF9 | 2525,24 | -0,07 | 0,03 | -2,45 | 1,45E-02 |
| C8orf58 | 90,67 | 0,14 | 0,06 | 2,44 | 1,45E-02 |
| TMCC3 | 3180,49 | -0,16 | 0,07 | -2,44 | 1,45E-02 |
| ARHGEF11 | 1206,63 | 0,20 | 0,08 | 2,44 | 1,45E-02 |
| PEBP1 | 990,96 | -0,12 | 0,05 | -2,44 | 1,45E-02 |
| BCKDK | 537,17 | 0,13 | 0,05 | 2,44 | 1,45E-02 |
| RNF13 | 5972,54 | -0,14 | 0,06 | -2,44 | 1,45E-02 |
| HS3ST1 | 58,63 | 0,35 | 0,14 | 2,44 | 1,45E-02 |
| TNFAIP2 | 8027,99 | 0,22 | 0,09 | 2,44 | 1,45E-02 |
| TRA2A | 3746,30 | -0,09 | 0,04 | -2,44 | 1,45E-02 |
| CNN3 | 33,12 | 0,31 | 0,13 | 2,44 | 1,46E-02 |
| NRN1 | 49,36 | 0,45 | 0,18 | 2,44 | 1,46E-02 |
| PELI3 | 207,05 | 0,12 | 0,05 | 2,44 | 1,46E-02 |
| XXYLT1 | 245,65 | 0,13 | 0,05 | 2,44 | 1,46E-02 |
| SLC35E1 | 782,09 | 0,13 | 0,05 | 2,44 | 1,46E-02 |
| NUP35 | 442,71 | -0,11 | 0,05 | -2,44 | 1,46E-02 |
| PKN1 | 1046,60 | 0,20 | 0,08 | 2,44 | 1,46E-02 |
| KLHL34 | 33,35 | 0,31 | 0,13 | 2,44 | 1,46E-02 |
| ABHD14B | 195,49 | 0,16 | 0,06 | 2,44 | 1,46E-02 |
| RSKR | 226,74 | 0,19 | 0,08 | 2,44 | 1,46E-02 |
| ETAA1 | 451,45 | -0,22 | 0,09 | -2,44 | 1,47E-02 |
| SLC1A4 | 165,37 | 0,16 | 0,06 | 2,44 | 1,47E-02 |
| SINHCAF | 1561,15 | -0,11 | 0,04 | -2,44 | 1,47E-02 |
| CHRNA10 | 119,92 | 0,17 | 0,07 | 2,44 | 1,47E-02 |
| NAP1L2 | 170,54 | -0,22 | 0,09 | -2,44 | 1,47E-02 |
| SLC35C2 | 1665,43 | 0,08 | 0,03 | 2,44 | 1,47E-02 |
| D2HGDH | 434,54 | 0,18 | 0,07 | 2,44 | 1,47E-02 |
| SF3A3 | 1644,58 | -0,08 | 0,03 | -2,44 | 1,47E-02 |
| MAN2C1 | 1679,77 | 0,17 | 0,07 | 2,44 | 1,47E-02 |
| DBR1 | 625,41 | -0,10 | 0,04 | -2,44 | 1,47E-02 |
| RAB44 | 352,99 | 0,24 | 0,10 | 2,44 | 1,48E-02 |
| SEMA3C | 472,51 | -0,16 | 0,07 | -2,44 | 1,48E-02 |

| | | | | | |
|---|---|---|---|---|---|
| GTPBP8 | 462,20 | -0,13 | 0,05 | -2,44 | 1,48E-02 |
| EVI2A | 1952,92 | -0,35 | 0,14 | -2,44 | 1,48E-02 |
| INPP5K | 1330,77 | 0,12 | 0,05 | 2,44 | 1,48E-02 |
| ZNF268 | 648,05 | -0,18 | 0,07 | -2,44 | 1,48E-02 |
| H2AZ2 | 2719,12 | -0,06 | 0,03 | -2,44 | 1,48E-02 |
| TP53RK | 994,19 | -0,11 | 0,05 | -2,44 | 1,48E-02 |
| BAZ2B | 13030,84 | -0,15 | 0,06 | -2,44 | 1,48E-02 |
| AMFR | 2675,50 | 0,14 | 0,06 | 2,44 | 1,49E-02 |
| DNAJB9 | 525,76 | -0,19 | 0,08 | -2,44 | 1,49E-02 |
| SMPDL3B | 30,26 | 0,29 | 0,12 | 2,44 | 1,49E-02 |
| PDGFB | 77,27 | 0,25 | 0,10 | 2,44 | 1,49E-02 |
| PHETA2 | 126,96 | 0,12 | 0,05 | 2,44 | 1,49E-02 |
| CRISP3 | 99,10 | 0,40 | 0,16 | 2,43 | 1,49E-02 |
| FEM1B | 2874,91 | -0,12 | 0,05 | -2,43 | 1,49E-02 |
| CAPN7 | 2511,65 | -0,10 | 0,04 | -2,43 | 1,49E-02 |
| RALGAPA1 | 2916,25 | -0,10 | 0,04 | -2,43 | 1,49E-02 |
| ST3GAL2 | 2510,16 | 0,14 | 0,06 | 2,43 | 1,49E-02 |
| LAIR2 | 74,14 | -0,28 | 0,11 | -2,43 | 1,50E-02 |
| ZMIZ2 | 708,42 | 0,23 | 0,09 | 2,43 | 1,50E-02 |
| ACP2 | 151,36 | 0,16 | 0,06 | 2,43 | 1,50E-02 |
| OSGEPL1 | 303,95 | -0,13 | 0,06 | -2,43 | 1,50E-02 |
| CLSTN3 | 564,01 | 0,20 | 0,08 | 2,43 | 1,50E-02 |
| NMI | 2581,41 | -0,15 | 0,06 | -2,43 | 1,50E-02 |
| CHKB-CPT1B | 44,54 | 0,19 | 0,08 | 2,43 | 1,50E-02 |
| RBL2 | 12409,08 | -0,09 | 0,04 | -2,43 | 1,51E-02 |
| STARD8 | 128,59 | 0,23 | 0,09 | 2,43 | 1,51E-02 |
| EOMES | 762,80 | -0,20 | 0,08 | -2,43 | 1,51E-02 |
| PABIR3 | 324,89 | 0,10 | 0,04 | 2,43 | 1,51E-02 |
| TMC8 | 2611,77 | 0,20 | 0,08 | 2,43 | 1,51E-02 |
| DHPS | 140,36 | 0,14 | 0,06 | 2,43 | 1,51E-02 |
| TFCP2L1 | 68,32 | 0,49 | 0,20 | 2,43 | 1,51E-02 |
| AZIN2 | 144,92 | 0,18 | 0,07 | 2,43 | 1,52E-02 |
| SLC25A1 | 241,91 | 0,12 | 0,05 | 2,43 | 1,52E-02 |
| ZKSCAN5 | 627,09 | -0,06 | 0,03 | -2,43 | 1,52E-02 |
| SLC39A7 | 938,62 | -0,08 | 0,03 | -2,43 | 1,52E-02 |
| RPS6KB1 | 1539,46 | -0,06 | 0,03 | -2,43 | 1,52E-02 |
| CYSTM1 | 546,07 | 0,19 | 0,08 | 2,43 | 1,52E-02 |
| TMEM156 | 376,83 | -0,13 | 0,05 | -2,43 | 1,53E-02 |
| OLA1 | 492,74 | -0,11 | 0,04 | -2,43 | 1,53E-02 |
| VILL | 639,39 | 0,12 | 0,05 | 2,43 | 1,53E-02 |
| MRPL22 | 349,31 | -0,10 | 0,04 | -2,43 | 1,53E-02 |
| ASGR1 | 230,74 | 0,14 | 0,06 | 2,43 | 1,53E-02 |
| UBTD1 | 85,36 | 0,20 | 0,08 | 2,43 | 1,53E-02 |
| GFPT2 | 39,78 | 0,56 | 0,23 | 2,42 | 1,53E-02 |
| PDLIM2 | 1137,94 | 0,18 | 0,07 | 2,42 | 1,54E-02 |
| PRSS21 | 27,77 | 0,40 | 0,17 | 2,42 | 1,54E-02 |

| | | | | | |
|---|---|---|---|---|---|
| RNF115 | 1750,30 | -0,06 | 0,02 | -2,42 | 1,54E-02 |
| SYPL1 | 726,53 | -0,11 | 0,05 | -2,42 | 1,54E-02 |
| SLC12A9 | 3615,08 | 0,17 | 0,07 | 2,42 | 1,54E-02 |
| LRWD1 | 396,14 | 0,18 | 0,08 | 2,42 | 1,54E-02 |
| SPPL3 | 849,77 | 0,12 | 0,05 | 2,42 | 1,54E-02 |
| TRIM28 | 925,17 | 0,15 | 0,06 | 2,42 | 1,55E-02 |
| ZNF654 | 1396,23 | -0,19 | 0,08 | -2,42 | 1,55E-02 |
| LGR4 | 79,81 | 0,23 | 0,10 | 2,42 | 1,55E-02 |
| HNRNPD | 3947,48 | -0,08 | 0,03 | -2,42 | 1,55E-02 |
| ACCS | 583,90 | 0,27 | 0,11 | 2,42 | 1,55E-02 |
| ETFRF1 | 315,95 | -0,16 | 0,07 | -2,42 | 1,55E-02 |
| C1D | 512,09 | -0,17 | 0,07 | -2,42 | 1,56E-02 |
| FCHSD1 | 744,82 | 0,16 | 0,07 | 2,42 | 1,56E-02 |
| FLT1 | 139,95 | 0,34 | 0,14 | 2,42 | 1,56E-02 |
| HMGCR | 2180,83 | -0,11 | 0,05 | -2,42 | 1,56E-02 |
| IARS1 | 2041,30 | -0,08 | 0,03 | -2,42 | 1,56E-02 |
| CDC42SE2 | 9942,04 | -0,11 | 0,05 | -2,42 | 1,56E-02 |
| TOR3A | 682,79 | 0,08 | 0,03 | 2,42 | 1,56E-02 |
| TXNRD3 | 79,42 | 0,24 | 0,10 | 2,42 | 1,57E-02 |
| TSC22D4 | 738,35 | 0,26 | 0,11 | 2,42 | 1,57E-02 |
| MRPS11 | 624,06 | -0,05 | 0,02 | -2,42 | 1,57E-02 |
| PEX14 | 208,09 | 0,14 | 0,06 | 2,42 | 1,57E-02 |
| GLMN | 426,61 | -0,12 | 0,05 | -2,42 | 1,57E-02 |
| NLGN3 | 64,16 | 0,30 | 0,12 | 2,42 | 1,57E-02 |
| LSM8 | 1385,92 | -0,13 | 0,05 | -2,42 | 1,57E-02 |
| SPATA13 | 3938,06 | -0,09 | 0,04 | -2,41 | 1,57E-02 |
| ZSCAN31 | 36,40 | 0,31 | 0,13 | 2,42 | 1,57E-02 |
| TUBGCP6 | 1708,90 | 0,15 | 0,06 | 2,41 | 1,57E-02 |
| TTC23 | 50,93 | 0,33 | 0,14 | 2,41 | 1,57E-02 |
| HPS4 | 929,09 | 0,08 | 0,03 | 2,41 | 1,58E-02 |
| CCDC62 | 59,57 | 0,17 | 0,07 | 2,41 | 1,58E-02 |
| PFKM | 298,85 | 0,11 | 0,04 | 2,41 | 1,58E-02 |
| PNKD | 268,23 | 0,12 | 0,05 | 2,41 | 1,58E-02 |
| TTC22 | 325,07 | 0,16 | 0,06 | 2,41 | 1,58E-02 |
| PRPF38B | 3111,42 | -0,12 | 0,05 | -2,41 | 1,58E-02 |
| TBC1D2 | 404,54 | 0,18 | 0,07 | 2,41 | 1,58E-02 |
| TFB2M | 201,94 | -0,11 | 0,05 | -2,41 | 1,58E-02 |
| INPPL1 | 950,58 | 0,20 | 0,08 | 2,41 | 1,58E-02 |
| TMEM234 | 361,16 | 0,14 | 0,06 | 2,41 | 1,58E-02 |
| MAP3K2 | 12730,64 | -0,13 | 0,06 | -2,41 | 1,58E-02 |
| VPS4A | 512,93 | 0,06 | 0,03 | 2,41 | 1,59E-02 |
| GALT | 827,66 | 0,11 | 0,05 | 2,41 | 1,59E-02 |
| PDCD2 | 1609,33 | -0,10 | 0,04 | -2,41 | 1,59E-02 |
| CAPN14 | 61,45 | 0,30 | 0,12 | 2,41 | 1,59E-02 |
| RPL17 | 455,25 | -0,16 | 0,06 | -2,41 | 1,59E-02 |
| OXR1 | 2234,96 | -0,16 | 0,07 | -2,41 | 1,59E-02 |

| | | | | | |
|---|---|---|---|---|---|
| EVA1C | 207,79 | 0,16 | 0,07 | 2,41 | 1,59E-02 |
| FASLG | 211,67 | -0,17 | 0,07 | -2,41 | 1,59E-02 |
| MYOCOS | 28,62 | 0,23 | 0,09 | 2,41 | 1,60E-02 |
| EPS8 | 135,82 | 0,25 | 0,10 | 2,41 | 1,60E-02 |
| TYW5 | 795,39 | -0,09 | 0,04 | -2,41 | 1,60E-02 |
| ZNF713 | 54,81 | 0,19 | 0,08 | 2,41 | 1,60E-02 |
| DCAKD | 306,33 | 0,09 | 0,04 | 2,41 | 1,60E-02 |
| PDLIM1 | 381,25 | 0,17 | 0,07 | 2,41 | 1,61E-02 |
| WDR36 | 1673,01 | -0,13 | 0,05 | -2,41 | 1,61E-02 |
| CYP20A1 | 896,34 | -0,06 | 0,03 | -2,41 | 1,61E-02 |
| EIF4A3 | 1134,01 | -0,08 | 0,03 | -2,41 | 1,61E-02 |
| MTMR6 | 2786,05 | -0,13 | 0,05 | -2,41 | 1,61E-02 |
| S100A8 | 10460,80 | -0,36 | 0,15 | -2,41 | 1,61E-02 |
| ZNF555 | 403,59 | -0,12 | 0,05 | -2,41 | 1,61E-02 |
| MAGED1 | 150,74 | 0,17 | 0,07 | 2,41 | 1,61E-02 |
| SCRN3 | 507,02 | -0,10 | 0,04 | -2,41 | 1,62E-02 |
| PSMA1 | 869,81 | -0,09 | 0,04 | -2,40 | 1,62E-02 |
| ARHGAP19 | 1450,25 | -0,12 | 0,05 | -2,40 | 1,62E-02 |
| EDC4 | 476,03 | 0,18 | 0,07 | 2,40 | 1,62E-02 |
| TRAPPC2 | 1050,85 | -0,07 | 0,03 | -2,40 | 1,63E-02 |
| LARP1 | 2171,10 | 0,14 | 0,06 | 2,40 | 1,63E-02 |
| NLGN4Y | 31,58 | 0,25 | 0,11 | 2,40 | 1,63E-02 |
| FPGS | 507,73 | 0,11 | 0,05 | 2,40 | 1,63E-02 |
| PDE3B | 6916,11 | -0,13 | 0,05 | -2,40 | 1,63E-02 |
| OCLN | 37,91 | 0,46 | 0,19 | 2,40 | 1,63E-02 |
| STRN | 3411,03 | -0,09 | 0,04 | -2,40 | 1,63E-02 |
| TMC6 | 3407,16 | 0,16 | 0,06 | 2,40 | 1,63E-02 |
| ZNF528 | 796,44 | -0,12 | 0,05 | -2,40 | 1,64E-02 |
| MOSPD2 | 2514,73 | -0,15 | 0,06 | -2,40 | 1,64E-02 |
| IER3IP1 | 277,55 | -0,18 | 0,07 | -2,40 | 1,64E-02 |
| MRE11 | 2040,37 | -0,09 | 0,04 | -2,40 | 1,64E-02 |
| UBA6 | 2777,29 | -0,13 | 0,05 | -2,40 | 1,64E-02 |
| DENND4B | 2088,66 | 0,19 | 0,08 | 2,40 | 1,64E-02 |
| SLC25A33 | 130,66 | 0,09 | 0,04 | 2,40 | 1,65E-02 |
| PDCL | 744,37 | -0,09 | 0,04 | -2,40 | 1,65E-02 |
| TMEM219 | 443,24 | 0,08 | 0,03 | 2,40 | 1,65E-02 |
| SYTL3 | 1542,56 | -0,15 | 0,06 | -2,40 | 1,65E-02 |
| MLC1 | 190,23 | 0,21 | 0,09 | 2,40 | 1,65E-02 |
| MYCBP | 410,70 | -0,12 | 0,05 | -2,40 | 1,65E-02 |
| SMAGP | 64,06 | 0,17 | 0,07 | 2,40 | 1,66E-02 |
| TMEM167A | 2889,64 | -0,17 | 0,07 | -2,40 | 1,66E-02 |
| ARMT1 | 738,63 | -0,16 | 0,06 | -2,40 | 1,66E-02 |
| IGSF8 | 245,45 | 0,19 | 0,08 | 2,40 | 1,66E-02 |
| ZNF775 | 64,19 | 0,14 | 0,06 | 2,40 | 1,66E-02 |
| ARRDC1 | 635,82 | 0,15 | 0,06 | 2,40 | 1,66E-02 |
| BRD4 | 1685,53 | 0,22 | 0,09 | 2,40 | 1,66E-02 |

| | | | | | |
|---|---|---|---|---|---|
| POFUT1 | 499,07 | 0,10 | 0,04 | 2,40 | 1,66E-02 |
| ADIPOR1 | 29040,58 | 0,26 | 0,11 | 2,39 | 1,66E-02 |
| PPIA | 5314,73 | -0,14 | 0,06 | -2,39 | 1,67E-02 |
| HIVEP2 | 3321,74 | -0,08 | 0,03 | -2,39 | 1,67E-02 |
| MRPL19 | 649,43 | -0,09 | 0,04 | -2,39 | 1,67E-02 |
| ATP11B | 8544,43 | -0,14 | 0,06 | -2,39 | 1,67E-02 |
| ATP5PD | 914,40 | -0,12 | 0,05 | -2,39 | 1,67E-02 |
| SLC11A2 | 1274,16 | 0,07 | 0,03 | 2,39 | 1,67E-02 |
| PSMC3 | 951,25 | -0,11 | 0,05 | -2,39 | 1,67E-02 |
| CCDC9 | 494,73 | 0,14 | 0,06 | 2,39 | 1,68E-02 |
| AP5M1 | 2589,72 | -0,14 | 0,06 | -2,39 | 1,68E-02 |
| ARMC10 | 772,23 | -0,07 | 0,03 | -2,39 | 1,68E-02 |
| TMEM177 | 138,56 | 0,12 | 0,05 | 2,39 | 1,68E-02 |
| RAP1B | 7093,90 | -0,09 | 0,04 | -2,39 | 1,69E-02 |
| ZFAND6 | 3293,67 | -0,11 | 0,05 | -2,39 | 1,69E-02 |
| SYT17 | 96,32 | 0,28 | 0,12 | 2,39 | 1,69E-02 |
| STK39 | 920,71 | -0,12 | 0,05 | -2,39 | 1,69E-02 |
| PARP8 | 15106,00 | -0,13 | 0,05 | -2,39 | 1,69E-02 |
| SH3BP2 | 4374,55 | 0,21 | 0,09 | 2,39 | 1,69E-02 |
| AVPR1A | 88,19 | 0,24 | 0,10 | 2,39 | 1,69E-02 |
| SIKE1 | 1418,76 | -0,13 | 0,06 | -2,39 | 1,69E-02 |
| ENSG00000241962 | 21,13 | 0,21 | 0,09 | 2,39 | 1,69E-02 |
| SRD5A1 | 561,01 | 0,10 | 0,04 | 2,39 | 1,70E-02 |
| PPP6R2 | 1405,28 | 0,14 | 0,06 | 2,39 | 1,70E-02 |
| HOXB4 | 79,89 | 0,18 | 0,08 | 2,39 | 1,70E-02 |
| PNMA1 | 382,55 | 0,10 | 0,04 | 2,39 | 1,70E-02 |
| CWC25 | 1145,45 | -0,07 | 0,03 | -2,39 | 1,70E-02 |
| RALGPS2 | 3145,55 | -0,19 | 0,08 | -2,39 | 1,70E-02 |
| GPATCH8 | 2656,30 | -0,07 | 0,03 | -2,39 | 1,70E-02 |
| METTL5 | 241,28 | -0,11 | 0,05 | -2,39 | 1,70E-02 |
| ACTA2 | 70,77 | 0,27 | 0,11 | 2,39 | 1,70E-02 |
| TSPAN3 | 687,46 | -0,11 | 0,05 | -2,39 | 1,70E-02 |
| SNX25 | 496,32 | -0,13 | 0,05 | -2,39 | 1,70E-02 |
| STBD1 | 43,14 | 0,20 | 0,08 | 2,39 | 1,70E-02 |
| PARD6A | 41,92 | 0,14 | 0,06 | 2,39 | 1,70E-02 |
| PHRF1 | 991,80 | 0,16 | 0,07 | 2,39 | 1,70E-02 |
| DCAF16 | 1920,74 | -0,07 | 0,03 | -2,38 | 1,71E-02 |
| MIER1 | 5011,81 | -0,14 | 0,06 | -2,38 | 1,71E-02 |
| DHX30 | 932,45 | 0,10 | 0,04 | 2,38 | 1,71E-02 |
| GPAA1 | 388,56 | 0,11 | 0,05 | 2,38 | 1,71E-02 |
| URM1 | 138,03 | 0,14 | 0,06 | 2,38 | 1,71E-02 |
| SLC27A4 | 218,19 | 0,11 | 0,05 | 2,38 | 1,72E-02 |
| STARD3 | 744,89 | 0,14 | 0,06 | 2,38 | 1,72E-02 |
| TSEN15 | 926,72 | -0,12 | 0,05 | -2,38 | 1,72E-02 |
| DUSP18 | 376,37 | 0,14 | 0,06 | 2,38 | 1,72E-02 |
| GPD1L | 791,96 | -0,08 | 0,03 | -2,38 | 1,72E-02 |

| | | | | | |
|---|---|---|---|---|---|
| RNF146 | 2347,17 | -0,13 | 0,05 | -2,38 | 1,72E-02 |
| POLR2F | 496,02 | 0,06 | 0,03 | 2,38 | 1,72E-02 |
| PHKG2 | 361,53 | 0,11 | 0,05 | 2,38 | 1,72E-02 |
| PIK3C2B | 501,70 | 0,17 | 0,07 | 2,38 | 1,72E-02 |
| FAM229A | 196,08 | 0,21 | 0,09 | 2,38 | 1,72E-02 |
| ARHGAP4 | 3338,22 | 0,12 | 0,05 | 2,38 | 1,72E-02 |
| HMG20B | 375,78 | 0,11 | 0,05 | 2,38 | 1,72E-02 |
| GSTM4 | 191,90 | 0,22 | 0,09 | 2,38 | 1,72E-02 |
| DRG2 | 616,66 | 0,11 | 0,05 | 2,38 | 1,72E-02 |
| PTX3 | 51,03 | 0,22 | 0,09 | 2,38 | 1,72E-02 |
| FAM209A | 28,63 | 0,21 | 0,09 | 2,38 | 1,73E-02 |
| ALAD | 715,63 | 0,09 | 0,04 | 2,38 | 1,73E-02 |
| ORAI1 | 462,17 | 0,12 | 0,05 | 2,38 | 1,73E-02 |
| ELMOD3 | 818,76 | 0,06 | 0,03 | 2,38 | 1,74E-02 |
| RPS6KA3 | 8239,06 | -0,09 | 0,04 | -2,38 | 1,74E-02 |
| GMPR2 | 2036,37 | -0,09 | 0,04 | -2,38 | 1,74E-02 |
| TSHZ2 | 470,28 | 0,21 | 0,09 | 2,38 | 1,74E-02 |
| TAFAZZIN | 624,02 | 0,12 | 0,05 | 2,38 | 1,74E-02 |
| TCIRG1 | 3865,40 | 0,18 | 0,08 | 2,38 | 1,74E-02 |
| AKT2 | 2757,40 | 0,09 | 0,04 | 2,38 | 1,74E-02 |
| KANTR | 888,72 | -0,11 | 0,05 | -2,38 | 1,74E-02 |
| PICK1 | 285,97 | 0,10 | 0,04 | 2,38 | 1,74E-02 |
| ENTPD6 | 641,12 | 0,12 | 0,05 | 2,38 | 1,75E-02 |
| RNF2 | 754,40 | -0,11 | 0,05 | -2,38 | 1,75E-02 |
| GZMM | 1027,78 | -0,16 | 0,07 | -2,38 | 1,75E-02 |
| ANTXR2 | 7778,27 | -0,11 | 0,05 | -2,38 | 1,75E-02 |
| ABHD17B | 672,91 | -0,08 | 0,03 | -2,38 | 1,75E-02 |
| DCAF12 | 28214,62 | 0,27 | 0,11 | 2,38 | 1,75E-02 |
| OGFOD3 | 356,88 | 0,10 | 0,04 | 2,38 | 1,75E-02 |
| NBEAL1 | 1209,16 | -0,09 | 0,04 | -2,38 | 1,75E-02 |
| ITPKC | 294,91 | 0,09 | 0,04 | 2,37 | 1,76E-02 |
| PRPF40B | 192,55 | 0,18 | 0,08 | 2,37 | 1,76E-02 |
| PRUNE2 | 103,98 | 0,44 | 0,18 | 2,37 | 1,76E-02 |
| RAB36 | 68,64 | 0,28 | 0,12 | 2,37 | 1,76E-02 |
| THAP1 | 312,23 | -0,13 | 0,05 | -2,37 | 1,76E-02 |
| RBIS | 463,51 | -0,16 | 0,07 | -2,37 | 1,76E-02 |
| ZFYVE16 | 5806,77 | -0,14 | 0,06 | -2,37 | 1,76E-02 |
| BLVRB | 8842,36 | 0,28 | 0,12 | 2,37 | 1,76E-02 |
| CBFA2T2 | 599,57 | 0,17 | 0,07 | 2,37 | 1,76E-02 |
| GLIPR1L2 | 33,48 | 0,30 | 0,13 | 2,37 | 1,76E-02 |
| VPS54 | 1160,81 | -0,10 | 0,04 | -2,37 | 1,76E-02 |
| MYLK | 580,69 | 0,24 | 0,10 | 2,37 | 1,76E-02 |
| CREM | 425,97 | -0,08 | 0,03 | -2,37 | 1,77E-02 |
| EMG1 | 1002,50 | -0,09 | 0,04 | -2,37 | 1,77E-02 |
| PRPF39 | 973,53 | -0,13 | 0,06 | -2,37 | 1,77E-02 |
| ATF2 | 2518,76 | -0,12 | 0,05 | -2,37 | 1,77E-02 |

| | | | | | |
|---|---|---|---|---|---|
| TUBG2 | 107,13 | 0,13 | 0,05 | 2,37 | 1,77E-02 |
| HIBCH | 975,80 | -0,09 | 0,04 | -2,37 | 1,77E-02 |
| PAPOLA | 9869,39 | -0,10 | 0,04 | -2,37 | 1,77E-02 |
| CYLD | 12606,47 | -0,11 | 0,04 | -2,37 | 1,77E-02 |
| ZNF140 | 656,30 | -0,08 | 0,03 | -2,37 | 1,77E-02 |
| CEP19 | 848,32 | -0,18 | 0,08 | -2,37 | 1,78E-02 |
| RNF167 | 388,21 | 0,16 | 0,07 | 2,37 | 1,78E-02 |
| CABIN1 | 1784,79 | 0,16 | 0,07 | 2,37 | 1,78E-02 |
| DYNC1LI1 | 2421,23 | -0,12 | 0,05 | -2,37 | 1,78E-02 |
| HAGH | 4662,69 | 0,24 | 0,10 | 2,37 | 1,78E-02 |
| RAB1B | 733,07 | 0,18 | 0,08 | 2,37 | 1,78E-02 |
| IMPA1 | 935,31 | -0,12 | 0,05 | -2,37 | 1,79E-02 |
| INTS5 | 217,72 | 0,15 | 0,06 | 2,37 | 1,79E-02 |
| RMDN2 | 286,80 | -0,11 | 0,05 | -2,37 | 1,79E-02 |
| CYSLTR1 | 1009,38 | -0,12 | 0,05 | -2,37 | 1,79E-02 |
| CAMTA1 | 467,65 | 0,11 | 0,05 | 2,37 | 1,79E-02 |
| DGKQ | 619,64 | 0,18 | 0,08 | 2,37 | 1,79E-02 |
| TXNDC9 | 490,13 | -0,18 | 0,08 | -2,37 | 1,80E-02 |
| ZNF710 | 734,86 | 0,19 | 0,08 | 2,37 | 1,80E-02 |
| EIF3H | 3389,64 | -0,11 | 0,05 | -2,36 | 1,80E-02 |
| SCAMP1 | 1264,86 | -0,13 | 0,06 | -2,36 | 1,81E-02 |
| CD247 | 4228,75 | -0,12 | 0,05 | -2,36 | 1,81E-02 |
| TMEM71 | 5325,60 | -0,13 | 0,05 | -2,36 | 1,81E-02 |
| ENPP2 | 105,60 | 0,24 | 0,10 | 2,36 | 1,81E-02 |
| TMEM229B | 515,94 | 0,14 | 0,06 | 2,36 | 1,81E-02 |
| MAL | 314,90 | 0,17 | 0,07 | 2,36 | 1,81E-02 |
| ANKRD26 | 627,00 | -0,14 | 0,06 | -2,36 | 1,81E-02 |
| CCDC86 | 95,02 | 0,15 | 0,06 | 2,36 | 1,82E-02 |
| YDJC | 265,89 | 0,10 | 0,04 | 2,36 | 1,82E-02 |
| EML3 | 690,36 | 0,21 | 0,09 | 2,36 | 1,82E-02 |
| MTM1 | 1267,73 | -0,09 | 0,04 | -2,36 | 1,82E-02 |
| EMB | 7529,60 | -0,13 | 0,06 | -2,36 | 1,82E-02 |
| ILVBL | 271,75 | 0,10 | 0,04 | 2,36 | 1,82E-02 |
| INTS14 | 684,30 | -0,07 | 0,03 | -2,36 | 1,82E-02 |
| SIGIRR | 769,85 | 0,18 | 0,08 | 2,36 | 1,82E-02 |
| EHMT2 | 530,69 | 0,13 | 0,06 | 2,36 | 1,83E-02 |
| PPP4R2 | 3127,69 | -0,17 | 0,07 | -2,36 | 1,83E-02 |
| TSPAN33 | 380,52 | 0,18 | 0,08 | 2,36 | 1,83E-02 |
| TP53I13 | 588,55 | 0,11 | 0,05 | 2,36 | 1,83E-02 |
| SDCBP | 13764,84 | -0,15 | 0,06 | -2,36 | 1,83E-02 |
| SLC27A2 | 36,83 | 0,27 | 0,12 | 2,36 | 1,83E-02 |
| CLINT1 | 3072,50 | -0,09 | 0,04 | -2,36 | 1,83E-02 |
| CUL5 | 1610,04 | -0,13 | 0,06 | -2,36 | 1,84E-02 |
| USP33 | 3805,90 | -0,09 | 0,04 | -2,36 | 1,84E-02 |
| CORIN | 97,12 | 0,41 | 0,17 | 2,36 | 1,84E-02 |
| NUCB2 | 680,54 | -0,11 | 0,05 | -2,36 | 1,84E-02 |

| | | | | | |
|---|---|---|---|---|---|
| TMBIM6 | 12829,51 | -0,08 | 0,03 | -2,36 | 1,84E-02 |
| PHLPP2 | 789,29 | 0,12 | 0,05 | 2,36 | 1,85E-02 |
| EFCAB14 | 3233,02 | -0,05 | 0,02 | -2,36 | 1,85E-02 |
| ARIH1 | 4296,09 | -0,06 | 0,02 | -2,36 | 1,85E-02 |
| ARPC5L | 630,87 | -0,09 | 0,04 | -2,36 | 1,85E-02 |
| HINT3 | 971,07 | -0,11 | 0,05 | -2,35 | 1,86E-02 |
| FAM89A | 103,04 | 0,14 | 0,06 | 2,35 | 1,86E-02 |
| ZNF614 | 369,82 | -0,14 | 0,06 | -2,35 | 1,86E-02 |
| SAP30BP | 1958,84 | -0,06 | 0,02 | -2,35 | 1,87E-02 |
| THG1L | 242,60 | -0,10 | 0,04 | -2,35 | 1,87E-02 |
| ZNF594 | 574,79 | -0,19 | 0,08 | -2,35 | 1,87E-02 |
| SS18L1 | 490,09 | 0,08 | 0,03 | 2,35 | 1,87E-02 |
| CLPB | 767,04 | 0,09 | 0,04 | 2,35 | 1,87E-02 |
| ANKMY1 | 1155,45 | 0,13 | 0,05 | 2,35 | 1,87E-02 |
| ZNF79 | 481,86 | -0,06 | 0,03 | -2,35 | 1,88E-02 |
| RIOX1 | 604,42 | -0,08 | 0,03 | -2,35 | 1,88E-02 |
| ORC2 | 1262,09 | -0,07 | 0,03 | -2,35 | 1,88E-02 |
| VPS36 | 2191,77 | -0,07 | 0,03 | -2,35 | 1,88E-02 |
| RPL3L | 29,05 | 0,58 | 0,25 | 2,35 | 1,88E-02 |
| RAB3GAP1 | 2374,67 | -0,09 | 0,04 | -2,35 | 1,88E-02 |
| PTK2 | 446,58 | 0,13 | 0,06 | 2,35 | 1,88E-02 |
| MRPL35 | 512,93 | -0,12 | 0,05 | -2,35 | 1,88E-02 |
| ZNF480 | 427,60 | -0,18 | 0,08 | -2,35 | 1,88E-02 |
| TMEM63A | 2994,03 | 0,20 | 0,08 | 2,35 | 1,89E-02 |
| SPATA2L | 143,35 | 0,15 | 0,06 | 2,35 | 1,89E-02 |
| CASP4 | 7654,09 | -0,15 | 0,06 | -2,35 | 1,89E-02 |
| TAL1 | 1497,49 | 0,24 | 0,10 | 2,35 | 1,89E-02 |
| IFT74 | 270,13 | -0,12 | 0,05 | -2,35 | 1,89E-02 |
| PML | 684,76 | 0,28 | 0,12 | 2,35 | 1,90E-02 |
| MTHFR | 895,95 | 0,18 | 0,08 | 2,35 | 1,90E-02 |
| GMFG | 5057,96 | -0,17 | 0,07 | -2,35 | 1,90E-02 |
| FBXO38 | 4984,39 | -0,10 | 0,04 | -2,34 | 1,90E-02 |
| CACUL1 | 6504,92 | -0,10 | 0,04 | -2,34 | 1,90E-02 |
| CSTF1 | 714,45 | -0,05 | 0,02 | -2,34 | 1,90E-02 |
| GTF2A2 | 756,97 | -0,10 | 0,04 | -2,34 | 1,91E-02 |
| FRRS1 | 254,41 | 0,20 | 0,08 | 2,34 | 1,91E-02 |
| ZNF211 | 761,72 | -0,07 | 0,03 | -2,34 | 1,91E-02 |
| FERRY3 | 559,41 | -0,11 | 0,05 | -2,34 | 1,91E-02 |
| YES1 | 354,33 | -0,12 | 0,05 | -2,34 | 1,91E-02 |
| MECR | 163,15 | 0,11 | 0,05 | 2,34 | 1,91E-02 |
| PGLS | 471,18 | 0,15 | 0,06 | 2,34 | 1,92E-02 |
| TUBB6 | 170,83 | 0,21 | 0,09 | 2,34 | 1,92E-02 |
| SLC9A6 | 907,57 | -0,07 | 0,03 | -2,34 | 1,92E-02 |
| YTHDF2 | 1197,92 | -0,05 | 0,02 | -2,34 | 1,92E-02 |
| SPTB | 1692,82 | 0,35 | 0,15 | 2,34 | 1,92E-02 |
| ZNF138 | 401,43 | -0,17 | 0,07 | -2,34 | 1,92E-02 |

| | | | | | |
|---|---|---|---|---|---|
| PIGC | 1434,22 | -0,23 | 0,10 | -2,34 | 1,92E-02 |
| COQ5 | 319,60 | -0,13 | 0,06 | -2,34 | 1,93E-02 |
| RHD | 91,81 | 0,52 | 0,22 | 2,34 | 1,93E-02 |
| UBQLN4 | 172,77 | 0,16 | 0,07 | 2,34 | 1,93E-02 |
| PEX10 | 201,97 | 0,09 | 0,04 | 2,34 | 1,93E-02 |
| RPS4X | 10571,97 | -0,15 | 0,06 | -2,34 | 1,93E-02 |
| CD83 | 289,46 | -0,14 | 0,06 | -2,34 | 1,93E-02 |
| SMG8 | 622,43 | -0,07 | 0,03 | -2,34 | 1,93E-02 |
| TTI2 | 451,35 | -0,06 | 0,03 | -2,34 | 1,93E-02 |
| DAP3 | 1445,99 | -0,06 | 0,03 | -2,34 | 1,94E-02 |
| ATG12 | 3063,12 | -0,12 | 0,05 | -2,34 | 1,94E-02 |
| MAP4K1 | 440,71 | 0,21 | 0,09 | 2,34 | 1,94E-02 |
| LIN7C | 1785,32 | -0,17 | 0,07 | -2,34 | 1,94E-02 |
| PTCD3 | 2560,00 | -0,08 | 0,04 | -2,34 | 1,94E-02 |
| FAM210B | 8250,21 | 0,29 | 0,12 | 2,34 | 1,94E-02 |
| RPAIN | 1451,20 | -0,08 | 0,03 | -2,34 | 1,95E-02 |
| RGL1 | 138,34 | 0,17 | 0,07 | 2,34 | 1,95E-02 |
| ATXN3 | 1888,28 | -0,07 | 0,03 | -2,34 | 1,95E-02 |
| ZMAT4 | 42,83 | 0,45 | 0,19 | 2,34 | 1,95E-02 |
| STX6 | 2578,09 | -0,08 | 0,04 | -2,33 | 1,96E-02 |
| COMMD10 | 799,78 | -0,11 | 0,05 | -2,33 | 1,96E-02 |
| STK25 | 880,48 | 0,11 | 0,05 | 2,33 | 1,96E-02 |
| IL18RAP | 2487,52 | -0,22 | 0,10 | -2,33 | 1,96E-02 |
| ZNF507 | 1661,36 | -0,10 | 0,04 | -2,33 | 1,96E-02 |
| ANKS6 | 244,52 | 0,15 | 0,07 | 2,33 | 1,97E-02 |
| ZNF304 | 487,55 | -0,10 | 0,04 | -2,33 | 1,97E-02 |
| SDF2 | 1229,65 | -0,09 | 0,04 | -2,33 | 1,97E-02 |
| CEMIP2 | 6364,45 | -0,09 | 0,04 | -2,33 | 1,97E-02 |
| MPV17 | 729,53 | 0,06 | 0,03 | 2,33 | 1,98E-02 |
| UBAP1L | 408,61 | 0,20 | 0,09 | 2,33 | 1,98E-02 |
| RARS2 | 1223,93 | -0,08 | 0,04 | -2,33 | 1,98E-02 |
| SUGP1 | 327,01 | 0,15 | 0,06 | 2,33 | 1,99E-02 |
| OSBPL7 | 274,94 | 0,17 | 0,08 | 2,33 | 1,99E-02 |
| KRBA2 | 128,98 | -0,12 | 0,05 | -2,33 | 1,99E-02 |
| FBXO4 | 266,95 | -0,14 | 0,06 | -2,33 | 1,99E-02 |
| NFU1 | 244,40 | -0,09 | 0,04 | -2,33 | 1,99E-02 |
| COX7B | 421,34 | -0,18 | 0,08 | -2,33 | 1,99E-02 |
| ZBTB45 | 265,47 | 0,09 | 0,04 | 2,33 | 1,99E-02 |
| SLC35G1 | 179,54 | 0,13 | 0,06 | 2,33 | 1,99E-02 |
| NCBP1 | 1777,94 | -0,08 | 0,03 | -2,33 | 2,00E-02 |
| NARF | 1452,10 | 0,15 | 0,07 | 2,33 | 2,01E-02 |
| ZKSCAN8 | 1741,31 | -0,08 | 0,03 | -2,33 | 2,01E-02 |
| AGPS | 1872,95 | -0,12 | 0,05 | -2,33 | 2,01E-02 |
| MSS51 | 275,24 | 0,10 | 0,04 | 2,32 | 2,01E-02 |
| MRGBP | 374,44 | -0,08 | 0,03 | -2,32 | 2,01E-02 |
| C17orf100 | 25,81 | 0,25 | 0,11 | 2,32 | 2,01E-02 |

| | | | | | |
|---|---|---|---|---|---|
| PAN2 | 851,63 | 0,11 | 0,05 | 2,32 | 2,01E-02 |
| MRM3 | 319,37 | -0,08 | 0,03 | -2,32 | 2,01E-02 |
| CSAD | 834,65 | 0,20 | 0,09 | 2,32 | 2,01E-02 |
| TSPOAP1 | 478,01 | 0,28 | 0,12 | 2,32 | 2,02E-02 |
| STOML2 | 982,47 | 0,13 | 0,06 | 2,32 | 2,02E-02 |
| RNF113A | 441,44 | -0,09 | 0,04 | -2,32 | 2,02E-02 |
| MRPS33 | 286,22 | -0,16 | 0,07 | -2,32 | 2,02E-02 |
| ZNF229 | 54,01 | 0,26 | 0,11 | 2,32 | 2,02E-02 |
| ZNF576 | 178,18 | 0,07 | 0,03 | 2,32 | 2,02E-02 |
| RAB11FIP3 | 397,45 | 0,16 | 0,07 | 2,32 | 2,02E-02 |
| STK17B | 20509,02 | -0,13 | 0,06 | -2,32 | 2,03E-02 |
| SGO1 | 32,76 | 0,23 | 0,10 | 2,32 | 2,03E-02 |
| FOXK1 | 1253,70 | 0,17 | 0,07 | 2,32 | 2,03E-02 |
| SAR1B | 1451,35 | -0,07 | 0,03 | -2,32 | 2,03E-02 |
| IFT88 | 549,88 | -0,12 | 0,05 | -2,32 | 2,03E-02 |
| CHD2 | 13849,27 | -0,07 | 0,03 | -2,32 | 2,04E-02 |
| RAB40C | 337,05 | 0,13 | 0,06 | 2,32 | 2,04E-02 |
| ARCN1 | 2349,24 | -0,07 | 0,03 | -2,32 | 2,04E-02 |
| ZNF324B | 175,06 | 0,10 | 0,04 | 2,32 | 2,04E-02 |
| ARMC8 | 2687,96 | -0,07 | 0,03 | -2,32 | 2,04E-02 |
| LRP1B | 49,72 | 0,46 | 0,20 | 2,32 | 2,04E-02 |
| CNIH1 | 688,17 | -0,13 | 0,06 | -2,32 | 2,04E-02 |
| FEZ2 | 1879,31 | -0,09 | 0,04 | -2,32 | 2,04E-02 |
| EIF3I | 1158,39 | -0,10 | 0,04 | -2,32 | 2,04E-02 |
| GOLM2 | 3348,52 | -0,11 | 0,05 | -2,32 | 2,05E-02 |
| RALGPS1 | 605,79 | 0,12 | 0,05 | 2,32 | 2,05E-02 |
| PTPN4 | 5679,93 | -0,14 | 0,06 | -2,32 | 2,05E-02 |
| RBM4B | 401,69 | -0,06 | 0,02 | -2,32 | 2,05E-02 |
| TUT7 | 13183,44 | -0,13 | 0,06 | -2,32 | 2,05E-02 |
| CREB1 | 6695,86 | -0,11 | 0,05 | -2,32 | 2,05E-02 |
| BLM | 326,92 | -0,09 | 0,04 | -2,32 | 2,06E-02 |
| EPM2A | 431,84 | 0,12 | 0,05 | 2,32 | 2,06E-02 |
| EMC7 | 485,58 | -0,08 | 0,03 | -2,32 | 2,06E-02 |
| N4BP2 | 1570,95 | -0,15 | 0,06 | -2,32 | 2,06E-02 |
| EMC8 | 243,13 | 0,10 | 0,04 | 2,31 | 2,06E-02 |
| NPAT | 2689,13 | -0,09 | 0,04 | -2,31 | 2,06E-02 |
| NBPF14 | 1737,95 | 0,14 | 0,06 | 2,31 | 2,07E-02 |
| G2E3 | 1447,81 | -0,10 | 0,04 | -2,31 | 2,07E-02 |
| CYP7B1 | 31,71 | 0,30 | 0,13 | 2,31 | 2,07E-02 |
| WDR55 | 1186,46 | 0,06 | 0,03 | 2,31 | 2,07E-02 |
| GSK3A | 451,98 | 0,14 | 0,06 | 2,31 | 2,07E-02 |
| FBRS | 1573,17 | 0,16 | 0,07 | 2,31 | 2,07E-02 |
| ABTB1 | 4307,42 | 0,22 | 0,09 | 2,31 | 2,07E-02 |
| GFPT1 | 1130,54 | -0,08 | 0,03 | -2,31 | 2,07E-02 |
| POLE2 | 128,12 | 0,26 | 0,11 | 2,31 | 2,07E-02 |
| BIRC3 | 5900,73 | -0,20 | 0,09 | -2,31 | 2,07E-02 |

| | | | | | |
|---|---|---|---|---|---|
| SPAG9 | 9703,66 | -0,11 | 0,05 | -2,31 | 2,07E-02 |
| DBP | 403,04 | 0,12 | 0,05 | 2,31 | 2,07E-02 |
| ABCD3 | 1157,74 | -0,10 | 0,04 | -2,31 | 2,07E-02 |
| SLMAP | 3091,74 | -0,08 | 0,03 | -2,31 | 2,07E-02 |
| PIAS2 | 2065,96 | -0,10 | 0,05 | -2,31 | 2,08E-02 |
| RPL10A | 7936,18 | -0,17 | 0,07 | -2,31 | 2,08E-02 |
| TMEM104 | 324,90 | 0,17 | 0,07 | 2,31 | 2,08E-02 |
| ZBTB14 | 966,69 | -0,11 | 0,05 | -2,31 | 2,08E-02 |
| ZCCHC4 | 467,29 | -0,09 | 0,04 | -2,31 | 2,08E-02 |
| CCP110 | 952,07 | -0,10 | 0,04 | -2,31 | 2,09E-02 |
| CLEC5A | 165,28 | 0,22 | 0,09 | 2,31 | 2,09E-02 |
| GUK1 | 23938,07 | 0,24 | 0,10 | 2,31 | 2,09E-02 |
| POLR1B | 755,65 | -0,08 | 0,03 | -2,31 | 2,09E-02 |
| SPARC | 792,70 | 0,27 | 0,12 | 2,31 | 2,09E-02 |
| FXYD5 | 643,31 | 0,11 | 0,05 | 2,31 | 2,10E-02 |
| NLRP6 | 1122,31 | 0,20 | 0,09 | 2,31 | 2,10E-02 |
| NDUFA12 | 637,32 | -0,13 | 0,06 | -2,31 | 2,10E-02 |
| ZNF281 | 2393,68 | -0,14 | 0,06 | -2,31 | 2,10E-02 |
| FBLN5 | 105,80 | 0,25 | 0,11 | 2,31 | 2,10E-02 |
| RAB11A | 1978,83 | -0,11 | 0,05 | -2,31 | 2,10E-02 |
| DPPA4 | 29,07 | 0,31 | 0,14 | 2,31 | 2,10E-02 |
| AMBRA1 | 983,91 | 0,12 | 0,05 | 2,31 | 2,10E-02 |
| SAP18 | 2359,29 | -0,11 | 0,05 | -2,31 | 2,10E-02 |
| KIF21A | 545,78 | -0,16 | 0,07 | -2,31 | 2,10E-02 |
| CPSF4 | 401,26 | 0,07 | 0,03 | 2,31 | 2,10E-02 |
| MRPL37 | 382,39 | 0,07 | 0,03 | 2,31 | 2,11E-02 |
| C11orf58 | 4460,24 | -0,13 | 0,06 | -2,31 | 2,11E-02 |
| ZNF77 | 108,42 | -0,09 | 0,04 | -2,31 | 2,11E-02 |
| GCFC2 | 606,02 | -0,12 | 0,05 | -2,31 | 2,11E-02 |
| ALDOC | 61,62 | 0,18 | 0,08 | 2,31 | 2,11E-02 |
| C11orf54 | 754,02 | -0,08 | 0,03 | -2,31 | 2,11E-02 |
| ELK4 | 4504,07 | -0,08 | 0,03 | -2,31 | 2,11E-02 |
| L1TD1 | 41,07 | 0,44 | 0,19 | 2,31 | 2,11E-02 |
| FOXK2 | 791,82 | 0,11 | 0,05 | 2,31 | 2,12E-02 |
| PPP1R2 | 3333,55 | -0,11 | 0,05 | -2,31 | 2,11E-02 |
| MRC2 | 781,08 | 0,27 | 0,12 | 2,31 | 2,12E-02 |
| DDHD2 | 1519,41 | -0,09 | 0,04 | -2,31 | 2,11E-02 |
| MTCL2 | 300,61 | 0,23 | 0,10 | 2,31 | 2,11E-02 |
| MYD88 | 1948,72 | 0,18 | 0,08 | 2,31 | 2,12E-02 |
| NAA40 | 201,95 | 0,24 | 0,11 | 2,31 | 2,12E-02 |
| MAPK8 | 1618,19 | -0,10 | 0,04 | -2,30 | 2,12E-02 |
| RNF139 | 1343,08 | -0,14 | 0,06 | -2,30 | 2,12E-02 |
| C10orf105 | 162,29 | 0,27 | 0,12 | 2,30 | 2,12E-02 |
| CIITA | 2627,40 | 0,23 | 0,10 | 2,30 | 2,12E-02 |
| WDR25 | 216,95 | 0,09 | 0,04 | 2,30 | 2,13E-02 |
| EPB42 | 3042,01 | 0,31 | 0,14 | 2,30 | 2,13E-02 |

| | | | | | |
|---|---|---|---|---|---|
| PCBP1 | 2187,74 | 0,19 | 0,08 | 2,30 | 2,13E-02 |
| NTNG2 | 960,27 | 0,26 | 0,11 | 2,30 | 2,13E-02 |
| EPC2 | 2348,77 | -0,08 | 0,04 | -2,30 | 2,13E-02 |
| RHBDF2 | 1075,56 | 0,19 | 0,08 | 2,30 | 2,13E-02 |
| CENPP | 145,20 | 0,19 | 0,08 | 2,30 | 2,14E-02 |
| ZNF578 | 61,14 | 0,22 | 0,10 | 2,30 | 2,14E-02 |
| PSMA5 | 1932,43 | -0,10 | 0,04 | -2,30 | 2,14E-02 |
| MMD | 1214,92 | 0,18 | 0,08 | 2,30 | 2,14E-02 |
| FHL1 | 368,36 | 0,16 | 0,07 | 2,30 | 2,14E-02 |
| GYPC | 18324,38 | 0,25 | 0,11 | 2,30 | 2,14E-02 |
| STOM | 4475,57 | 0,17 | 0,08 | 2,30 | 2,14E-02 |
| NOL12 | 182,51 | 0,15 | 0,07 | 2,30 | 2,14E-02 |
| SRXN1 | 625,87 | 0,20 | 0,09 | 2,30 | 2,15E-02 |
| FAM98B | 684,22 | -0,11 | 0,05 | -2,30 | 2,16E-02 |
| PLAGL2 | 597,37 | 0,15 | 0,07 | 2,30 | 2,16E-02 |
| SEL1L | 5827,48 | -0,09 | 0,04 | -2,30 | 2,16E-02 |
| AP1AR | 610,34 | -0,16 | 0,07 | -2,30 | 2,16E-02 |
| CIAO1 | 1839,95 | -0,04 | 0,02 | -2,30 | 2,16E-02 |
| S1PR1 | 3262,77 | -0,11 | 0,05 | -2,30 | 2,17E-02 |
| TRPV2 | 1158,93 | 0,11 | 0,05 | 2,30 | 2,17E-02 |
| IFT57 | 459,02 | -0,17 | 0,07 | -2,30 | 2,17E-02 |
| DNAJC6 | 436,88 | 0,27 | 0,12 | 2,30 | 2,17E-02 |
| TRIM13 | 1127,34 | -0,11 | 0,05 | -2,30 | 2,17E-02 |
| DCAF17 | 873,95 | -0,09 | 0,04 | -2,30 | 2,17E-02 |
| MINDY3 | 1140,45 | -0,12 | 0,05 | -2,30 | 2,17E-02 |
| NPIPA1 | 52,60 | 0,22 | 0,10 | 2,29 | 2,18E-02 |
| CHAF1A | 179,54 | 0,13 | 0,06 | 2,29 | 2,18E-02 |
| CDK1 | 42,17 | 0,27 | 0,12 | 2,29 | 2,18E-02 |
| ZNF431 | 2291,11 | -0,08 | 0,04 | -2,29 | 2,18E-02 |
| CD55 | 6554,83 | -0,15 | 0,07 | -2,29 | 2,18E-02 |
| PKHD1L1 | 243,04 | 0,26 | 0,11 | 2,29 | 2,18E-02 |
| EXOC4 | 2818,59 | -0,08 | 0,03 | -2,29 | 2,18E-02 |
| CHRAC1 | 242,03 | 0,07 | 0,03 | 2,29 | 2,19E-02 |
| CFAP54 | 142,39 | 0,26 | 0,11 | 2,29 | 2,19E-02 |
| ENSG00000285053 | 222,51 | -0,09 | 0,04 | -2,29 | 2,19E-02 |
| SLC7A6OS | 1311,51 | -0,04 | 0,02 | -2,29 | 2,19E-02 |
| MID1IP1 | 2876,97 | -0,12 | 0,05 | -2,29 | 2,19E-02 |
| RBM38 | 6679,24 | 0,26 | 0,11 | 2,29 | 2,19E-02 |
| DPH5 | 426,38 | -0,13 | 0,06 | -2,29 | 2,20E-02 |
| ZNF366 | 53,75 | 0,25 | 0,11 | 2,29 | 2,20E-02 |
| ACYP2 | 359,39 | 0,07 | 0,03 | 2,29 | 2,20E-02 |
| ZNF362 | 315,26 | 0,16 | 0,07 | 2,29 | 2,20E-02 |
| FKRP | 224,55 | 0,17 | 0,07 | 2,29 | 2,20E-02 |
| UBOX5 | 199,82 | 0,17 | 0,07 | 2,29 | 2,20E-02 |
| FTSJ1 | 218,95 | 0,11 | 0,05 | 2,29 | 2,20E-02 |
| DYRK1A | 11607,12 | -0,07 | 0,03 | -2,29 | 2,21E-02 |

| | | | | | |
|---|---|---|---|---|---|
| FGFR2 | 142,30 | 0,37 | 0,16 | 2,29 | 2,21E-02 |
| FZD1 | 227,47 | 0,16 | 0,07 | 2,29 | 2,21E-02 |
| PTOV1 | 858,72 | 0,12 | 0,05 | 2,29 | 2,21E-02 |
| FBXO45 | 384,08 | -0,11 | 0,05 | -2,29 | 2,21E-02 |
| METTL22 | 1013,80 | 0,11 | 0,05 | 2,29 | 2,21E-02 |
| IFI30 | 249,04 | 0,15 | 0,07 | 2,29 | 2,22E-02 |
| MSANTD2 | 618,02 | -0,08 | 0,04 | -2,29 | 2,22E-02 |
| SPAG8 | 55,85 | 0,14 | 0,06 | 2,29 | 2,22E-02 |
| HMGN4 | 1571,67 | -0,10 | 0,04 | -2,29 | 2,22E-02 |
| CLSTN1 | 638,02 | 0,19 | 0,08 | 2,29 | 2,22E-02 |
| TM2D2 | 513,97 | -0,09 | 0,04 | -2,29 | 2,22E-02 |
| ATAD2 | 779,00 | -0,11 | 0,05 | -2,29 | 2,23E-02 |
| IDH3A | 796,88 | -0,06 | 0,03 | -2,29 | 2,23E-02 |
| FAF2 | 1259,85 | -0,05 | 0,02 | -2,29 | 2,23E-02 |
| ARHGAP27 | 3348,31 | 0,19 | 0,08 | 2,29 | 2,23E-02 |
| ENSG00000182230 | 71,55 | 0,30 | 0,13 | 2,29 | 2,23E-02 |
| COL4A4 | 168,46 | 0,31 | 0,14 | 2,28 | 2,23E-02 |
| SNX33 | 180,54 | 0,18 | 0,08 | 2,28 | 2,23E-02 |
| TMEM242 | 438,82 | -0,07 | 0,03 | -2,28 | 2,23E-02 |
| PEX1 | 1094,68 | -0,08 | 0,03 | -2,28 | 2,24E-02 |
| COX20 | 748,93 | -0,09 | 0,04 | -2,28 | 2,24E-02 |
| SLC44A2 | 3410,91 | 0,18 | 0,08 | 2,28 | 2,24E-02 |
| PCGF1 | 144,20 | 0,07 | 0,03 | 2,28 | 2,24E-02 |
| PXDNL | 58,52 | 0,24 | 0,11 | 2,28 | 2,24E-02 |
| GALNS | 769,54 | 0,23 | 0,10 | 2,28 | 2,25E-02 |
| HEATR3 | 991,87 | -0,09 | 0,04 | -2,28 | 2,25E-02 |
| ABCD1 | 261,86 | 0,19 | 0,08 | 2,28 | 2,25E-02 |
| PPIB | 2556,24 | -0,10 | 0,04 | -2,28 | 2,25E-02 |
| RPRD1A | 1912,91 | -0,09 | 0,04 | -2,28 | 2,25E-02 |
| CPD | 8867,28 | -0,14 | 0,06 | -2,28 | 2,25E-02 |
| PLPP6 | 356,59 | -0,10 | 0,05 | -2,28 | 2,25E-02 |
| CRIP3 | 30,63 | 0,25 | 0,11 | 2,28 | 2,25E-02 |
| SFXN3 | 617,54 | 0,09 | 0,04 | 2,28 | 2,25E-02 |
| CLDN5 | 198,71 | 0,25 | 0,11 | 2,28 | 2,25E-02 |
| CSNK1A1 | 3237,72 | -0,07 | 0,03 | -2,28 | 2,25E-02 |
| CD101 | 604,25 | 0,18 | 0,08 | 2,28 | 2,26E-02 |
| ANKRD49 | 740,11 | -0,17 | 0,08 | -2,28 | 2,26E-02 |
| INHBB | 39,46 | 0,31 | 0,14 | 2,28 | 2,27E-02 |
| AXIN1 | 1077,40 | 0,13 | 0,06 | 2,28 | 2,27E-02 |
| LENG8 | 2129,70 | 0,27 | 0,12 | 2,28 | 2,27E-02 |
| RDH16 | 51,70 | 0,21 | 0,09 | 2,28 | 2,27E-02 |
| CYB5R4 | 6537,34 | -0,12 | 0,05 | -2,28 | 2,27E-02 |
| MKRN2 | 747,51 | -0,05 | 0,02 | -2,28 | 2,27E-02 |
| GRAMD1C | 674,37 | -0,20 | 0,09 | -2,28 | 2,28E-02 |
| MOB3A | 2629,80 | 0,21 | 0,09 | 2,28 | 2,28E-02 |
| ZPR1 | 862,28 | -0,06 | 0,02 | -2,28 | 2,28E-02 |

| | | | | | |
|---|---|---|---|---|---|
| SEPTIN8 | 176,22 | 0,16 | 0,07 | 2,28 | 2,28E-02 |
| SPTSSB | 54,98 | 0,26 | 0,11 | 2,28 | 2,28E-02 |
| ME1 | 49,41 | 0,24 | 0,10 | 2,28 | 2,28E-02 |
| ZDHHC15 | 26,90 | 0,31 | 0,14 | 2,28 | 2,28E-02 |
| ASB6 | 346,31 | 0,15 | 0,07 | 2,28 | 2,29E-02 |
| PLCB1 | 1443,47 | -0,12 | 0,05 | -2,28 | 2,29E-02 |
| FAM219A | 233,50 | 0,17 | 0,08 | 2,27 | 2,29E-02 |
| ATF1 | 691,73 | -0,09 | 0,04 | -2,27 | 2,29E-02 |
| STX7 | 10844,51 | -0,11 | 0,05 | -2,27 | 2,30E-02 |
| WASF2 | 3473,99 | 0,18 | 0,08 | 2,27 | 2,30E-02 |
| TSPAN32 | 1118,92 | 0,16 | 0,07 | 2,27 | 2,30E-02 |
| CSMD1 | 76,42 | 0,62 | 0,27 | 2,27 | 2,30E-02 |
| POLR2K | 233,03 | -0,28 | 0,12 | -2,27 | 2,30E-02 |
| ADGRG5 | 468,28 | 0,21 | 0,09 | 2,27 | 2,30E-02 |
| KLC4 | 295,82 | 0,13 | 0,06 | 2,27 | 2,30E-02 |
| CTDSP1 | 1540,33 | 0,14 | 0,06 | 2,27 | 2,30E-02 |
| CMTM6 | 12731,95 | -0,15 | 0,06 | -2,27 | 2,31E-02 |
| KXD1 | 738,73 | 0,11 | 0,05 | 2,27 | 2,31E-02 |
| FGFR1OP2 | 3367,82 | -0,16 | 0,07 | -2,27 | 2,31E-02 |
| SRCAP | 439,06 | 0,13 | 0,06 | 2,27 | 2,31E-02 |
| MPND | 222,26 | 0,09 | 0,04 | 2,27 | 2,31E-02 |
| ZNF26 | 1456,42 | -0,07 | 0,03 | -2,27 | 2,31E-02 |
| GLRX5 | 4708,95 | 0,28 | 0,12 | 2,27 | 2,31E-02 |
| PITHD1 | 4131,90 | 0,26 | 0,11 | 2,27 | 2,31E-02 |
| GIGYF1 | 1048,80 | 0,23 | 0,10 | 2,27 | 2,32E-02 |
| FBXW7 | 3928,13 | -0,07 | 0,03 | -2,27 | 2,32E-02 |
| M6PR | 3991,30 | -0,09 | 0,04 | -2,27 | 2,32E-02 |
| PTAR1 | 6547,46 | -0,14 | 0,06 | -2,27 | 2,32E-02 |
| ZNF551 | 500,42 | -0,09 | 0,04 | -2,27 | 2,32E-02 |
| XPOT | 1614,00 | -0,09 | 0,04 | -2,27 | 2,33E-02 |
| TFIP11 | 1778,93 | -0,09 | 0,04 | -2,27 | 2,33E-02 |
| TMEM86A | 91,07 | 0,16 | 0,07 | 2,27 | 2,33E-02 |
| ADGRF3 | 80,95 | 0,20 | 0,09 | 2,27 | 2,34E-02 |
| TBCA | 425,58 | -0,11 | 0,05 | -2,27 | 2,34E-02 |
| NOMO1 | 360,69 | 0,14 | 0,06 | 2,27 | 2,34E-02 |
| ZNF335 | 828,15 | 0,14 | 0,06 | 2,27 | 2,34E-02 |
| FAM174A | 835,46 | -0,14 | 0,06 | -2,27 | 2,34E-02 |
| SEC11A | 1135,92 | -0,07 | 0,03 | -2,27 | 2,34E-02 |
| SLCO4C1 | 1185,92 | -0,14 | 0,06 | -2,27 | 2,34E-02 |
| ABCD4 | 598,57 | 0,12 | 0,05 | 2,27 | 2,35E-02 |
| ACSL6 | 472,07 | 0,14 | 0,06 | 2,27 | 2,35E-02 |
| FMNL3 | 933,69 | 0,16 | 0,07 | 2,27 | 2,35E-02 |
| GRAMD1A | 1433,72 | 0,18 | 0,08 | 2,27 | 2,35E-02 |
| LIPH | 39,25 | 0,34 | 0,15 | 2,27 | 2,35E-02 |
| MTRF1L | 539,61 | -0,13 | 0,06 | -2,27 | 2,35E-02 |
| PSMD10 | 665,83 | -0,16 | 0,07 | -2,26 | 2,36E-02 |

| | | | | | |
|---|---|---|---|---|---|
| CMTM5 | 183,23 | 0,26 | 0,12 | 2,26 | 2,36E-02 |
| BTBD7 | 1520,66 | -0,06 | 0,03 | -2,26 | 2,36E-02 |
| CA2 | 475,11 | 0,22 | 0,10 | 2,26 | 2,36E-02 |
| VAC14 | 642,83 | 0,15 | 0,07 | 2,26 | 2,37E-02 |
| ZNF155 | 313,77 | -0,16 | 0,07 | -2,26 | 2,37E-02 |
| TMEM14C | 409,80 | 0,14 | 0,06 | 2,26 | 2,37E-02 |
| KLF16 | 398,29 | 0,15 | 0,07 | 2,26 | 2,37E-02 |
| ADPRS | 700,69 | -0,09 | 0,04 | -2,26 | 2,37E-02 |
| MTAP | 1191,00 | -0,08 | 0,04 | -2,26 | 2,37E-02 |
| ZDHHC14 | 178,57 | 0,21 | 0,09 | 2,26 | 2,37E-02 |
| TMEM203 | 570,37 | -0,08 | 0,04 | -2,26 | 2,37E-02 |
| SLC52A2 | 195,44 | 0,10 | 0,04 | 2,26 | 2,37E-02 |
| FAM199X | 2447,15 | -0,10 | 0,04 | -2,26 | 2,38E-02 |
| ARMC6 | 308,22 | 0,08 | 0,04 | 2,26 | 2,38E-02 |
| C12orf60 | 28,86 | 0,26 | 0,12 | 2,26 | 2,38E-02 |
| QSOX2 | 478,45 | 0,14 | 0,06 | 2,26 | 2,38E-02 |
| SCAF8 | 3190,68 | -0,06 | 0,03 | -2,26 | 2,38E-02 |
| THAP5 | 1257,30 | -0,16 | 0,07 | -2,26 | 2,38E-02 |
| SNX5 | 2277,49 | -0,07 | 0,03 | -2,26 | 2,38E-02 |
| TAB3 | 1928,56 | -0,07 | 0,03 | -2,26 | 2,39E-02 |
| TMEM170A | 1666,96 | -0,06 | 0,03 | -2,26 | 2,39E-02 |
| MIDEAS | 2224,07 | 0,14 | 0,06 | 2,26 | 2,39E-02 |
| XBP1 | 2541,63 | -0,09 | 0,04 | -2,26 | 2,39E-02 |
| MOV10 | 548,65 | 0,19 | 0,08 | 2,26 | 2,40E-02 |
| ZNG1A | 841,12 | -0,12 | 0,06 | -2,26 | 2,40E-02 |
| GSDMB | 1229,17 | 0,13 | 0,06 | 2,26 | 2,40E-02 |
| CLECL1P | 131,34 | -0,25 | 0,11 | -2,26 | 2,41E-02 |
| CNTROB | 279,11 | 0,12 | 0,05 | 2,26 | 2,41E-02 |
| CTSC | 6730,50 | -0,10 | 0,05 | -2,26 | 2,41E-02 |
| DNMT3A | 1679,13 | 0,12 | 0,05 | 2,25 | 2,41E-02 |
| TSPAN4 | 175,76 | 0,16 | 0,07 | 2,26 | 2,41E-02 |
| ZNF250 | 408,06 | -0,07 | 0,03 | -2,25 | 2,42E-02 |
| TIMMDC1 | 1106,43 | -0,08 | 0,04 | -2,25 | 2,42E-02 |
| MON1A | 168,04 | 0,09 | 0,04 | 2,25 | 2,42E-02 |
| STXBP2 | 779,32 | 0,19 | 0,08 | 2,25 | 2,42E-02 |
| IPO11 | 596,99 | -0,10 | 0,04 | -2,25 | 2,43E-02 |
| NAA30 | 1126,83 | -0,11 | 0,05 | -2,25 | 2,43E-02 |
| ATF6B | 574,93 | 0,18 | 0,08 | 2,25 | 2,42E-02 |
| TPM2 | 65,09 | 0,25 | 0,11 | 2,25 | 2,43E-02 |
| CSK | 2519,38 | 0,18 | 0,08 | 2,25 | 2,43E-02 |
| RB1 | 2223,06 | -0,10 | 0,05 | -2,25 | 2,43E-02 |
| ZNF835 | 93,69 | 0,21 | 0,09 | 2,25 | 2,44E-02 |
| SACM1L | 3905,78 | -0,10 | 0,04 | -2,25 | 2,44E-02 |
| RDH5 | 70,72 | 0,21 | 0,09 | 2,25 | 2,44E-02 |
| SEC22A | 517,63 | -0,08 | 0,03 | -2,25 | 2,44E-02 |
| E4F1 | 852,44 | 0,07 | 0,03 | 2,25 | 2,44E-02 |

| | | | | | |
|---|---|---|---|---|---|
| CD1A | 41,17 | 0,25 | 0,11 | 2,25 | 2,44E-02 |
| SDHD | 1326,56 | -0,09 | 0,04 | -2,25 | 2,44E-02 |
| TNFRSF4 | 113,98 | 0,19 | 0,08 | 2,25 | 2,45E-02 |
| SIAH1 | 956,88 | -0,08 | 0,03 | -2,25 | 2,45E-02 |
| CBX4 | 498,49 | 0,11 | 0,05 | 2,25 | 2,45E-02 |
| ZDHHC4 | 241,60 | 0,06 | 0,03 | 2,25 | 2,45E-02 |
| NUDT14 | 85,67 | 0,14 | 0,06 | 2,25 | 2,45E-02 |
| SLC4A5 | 90,56 | 0,14 | 0,06 | 2,25 | 2,45E-02 |
| GNA15 | 376,54 | 0,12 | 0,06 | 2,25 | 2,46E-02 |
| SYVN1 | 365,47 | 0,16 | 0,07 | 2,25 | 2,46E-02 |
| RWDD4 | 318,80 | -0,10 | 0,04 | -2,25 | 2,46E-02 |
| SEMA5A | 98,59 | 0,31 | 0,14 | 2,25 | 2,46E-02 |
| DYNC1I2 | 993,17 | -0,09 | 0,04 | -2,25 | 2,46E-02 |
| SLC22A16 | 103,63 | 0,23 | 0,10 | 2,25 | 2,46E-02 |
| SEC14L2 | 70,74 | 0,22 | 0,10 | 2,25 | 2,46E-02 |
| MGRN1 | 1284,11 | 0,23 | 0,10 | 2,25 | 2,46E-02 |
| LYRM9 | 226,61 | 0,09 | 0,04 | 2,25 | 2,47E-02 |
| KDELR1 | 156,59 | 0,18 | 0,08 | 2,25 | 2,46E-02 |
| HMCES | 891,14 | -0,07 | 0,03 | -2,25 | 2,46E-02 |
| UBE2Q2 | 780,32 | -0,10 | 0,04 | -2,25 | 2,47E-02 |
| MTMR9 | 1623,42 | -0,07 | 0,03 | -2,25 | 2,46E-02 |
| ENSG00000261884 | 369,42 | 0,11 | 0,05 | 2,25 | 2,47E-02 |
| GLT8D1 | 638,40 | -0,06 | 0,02 | -2,25 | 2,47E-02 |
| CRTC2 | 524,75 | 0,15 | 0,07 | 2,25 | 2,47E-02 |
| NOMO2 | 442,81 | 0,12 | 0,05 | 2,25 | 2,47E-02 |
| ATP9B | 2993,85 | -0,08 | 0,04 | -2,25 | 2,47E-02 |
| STIM2 | 1886,48 | -0,07 | 0,03 | -2,25 | 2,47E-02 |
| AFMID | 104,58 | 0,13 | 0,06 | 2,25 | 2,47E-02 |
| DRICH1 | 51,85 | 0,20 | 0,09 | 2,25 | 2,48E-02 |
| RNF111 | 2459,65 | -0,08 | 0,04 | -2,24 | 2,48E-02 |
| MAP7D1 | 997,59 | 0,19 | 0,08 | 2,24 | 2,49E-02 |
| TDRP | 110,53 | 0,26 | 0,12 | 2,24 | 2,49E-02 |
| FAM83D | 41,73 | 0,21 | 0,10 | 2,24 | 2,49E-02 |
| GABPB1 | 753,32 | -0,10 | 0,05 | -2,24 | 2,49E-02 |
| JAG1 | 263,32 | 0,16 | 0,07 | 2,24 | 2,49E-02 |
| MIER3 | 770,92 | -0,15 | 0,07 | -2,24 | 2,50E-02 |
| SLC26A2 | 921,06 | -0,07 | 0,03 | -2,24 | 2,50E-02 |
| TRMT11 | 592,17 | -0,11 | 0,05 | -2,24 | 2,51E-02 |
| C2CD5 | 2609,46 | -0,09 | 0,04 | -2,24 | 2,51E-02 |
| NMNAT3 | 176,42 | 0,18 | 0,08 | 2,24 | 2,51E-02 |
| OPHN1 | 108,99 | 0,19 | 0,09 | 2,24 | 2,51E-02 |
| UTRN | 22135,53 | -0,10 | 0,04 | -2,24 | 2,51E-02 |
| TSPAN5 | 3650,67 | 0,23 | 0,10 | 2,24 | 2,51E-02 |
| TCERG1 | 2138,34 | -0,10 | 0,04 | -2,24 | 2,51E-02 |
| SHC1 | 687,24 | 0,13 | 0,06 | 2,24 | 2,52E-02 |
| AP2M1 | 2041,52 | 0,16 | 0,07 | 2,24 | 2,52E-02 |

| | | | | | |
|---|---|---|---|---|---|
| ARB2A | 1420,25 | -0,11 | 0,05 | -2,24 | 2,52E-02 |
| DHX16 | 1765,29 | -0,06 | 0,03 | -2,24 | 2,52E-02 |
| TEKTIP1 | 39,31 | 0,31 | 0,14 | 2,24 | 2,53E-02 |
| AGFG2 | 353,49 | 0,14 | 0,06 | 2,24 | 2,53E-02 |
| MRPS18B | 573,49 | -0,09 | 0,04 | -2,24 | 2,53E-02 |
| PREPL | 1403,96 | -0,08 | 0,04 | -2,24 | 2,54E-02 |
| RALGDS | 497,05 | 0,16 | 0,07 | 2,24 | 2,54E-02 |
| RRP7A | 1290,14 | 0,12 | 0,05 | 2,24 | 2,54E-02 |
| BMF | 546,44 | 0,15 | 0,07 | 2,23 | 2,54E-02 |
| SH2B3 | 2572,25 | 0,14 | 0,06 | 2,23 | 2,54E-02 |
| DNAJB2 | 2625,41 | 0,19 | 0,08 | 2,23 | 2,54E-02 |
| PPP1R15B | 5200,32 | -0,07 | 0,03 | -2,23 | 2,54E-02 |
| KCTD20 | 3837,69 | -0,08 | 0,03 | -2,23 | 2,54E-02 |
| EXOSC4 | 171,98 | 0,10 | 0,04 | 2,23 | 2,55E-02 |
| IDH3G | 616,03 | 0,07 | 0,03 | 2,23 | 2,55E-02 |
| EDF1 | 2220,67 | -0,09 | 0,04 | -2,23 | 2,55E-02 |
| FHL3 | 405,11 | 0,19 | 0,08 | 2,23 | 2,56E-02 |
| LTF | 536,55 | 0,38 | 0,17 | 2,23 | 2,56E-02 |
| ZNF286A | 279,54 | -0,11 | 0,05 | -2,23 | 2,56E-02 |
| SS18L2 | 264,33 | -0,17 | 0,08 | -2,23 | 2,56E-02 |
| SEPTIN9 | 3348,33 | 0,16 | 0,07 | 2,23 | 2,57E-02 |
| RPL31 | 2889,25 | -0,34 | 0,15 | -2,23 | 2,57E-02 |
| SP4 | 1833,84 | -0,09 | 0,04 | -2,23 | 2,57E-02 |
| ECH1 | 309,69 | 0,11 | 0,05 | 2,23 | 2,58E-02 |
| TMEM126A | 138,73 | -0,18 | 0,08 | -2,23 | 2,58E-02 |
| MOCS2 | 313,75 | -0,12 | 0,06 | -2,23 | 2,58E-02 |
| IMPDH2 | 836,28 | -0,09 | 0,04 | -2,23 | 2,58E-02 |
| USP9X | 7807,97 | -0,09 | 0,04 | -2,23 | 2,58E-02 |
| BMI1 | 727,19 | -0,15 | 0,07 | -2,23 | 2,58E-02 |
| SHLD3 | 130,03 | -0,18 | 0,08 | -2,23 | 2,59E-02 |
| ACD | 120,54 | 0,14 | 0,06 | 2,23 | 2,59E-02 |
| C16orf74 | 135,95 | 0,15 | 0,07 | 2,23 | 2,59E-02 |
| RPL14 | 7859,15 | -0,14 | 0,06 | -2,23 | 2,59E-02 |
| GPHN | 193,13 | 0,13 | 0,06 | 2,23 | 2,59E-02 |
| GCN1 | 2078,38 | 0,11 | 0,05 | 2,23 | 2,59E-02 |
| MBD6 | 428,61 | 0,28 | 0,13 | 2,23 | 2,60E-02 |
| ABCA6 | 148,48 | 0,40 | 0,18 | 2,23 | 2,60E-02 |
| MLLT11 | 97,70 | 0,10 | 0,05 | 2,23 | 2,60E-02 |
| NAMPT | 66056,86 | -0,19 | 0,08 | -2,23 | 2,60E-02 |
| GATA3 | 175,63 | 0,15 | 0,07 | 2,23 | 2,60E-02 |
| IKBKG | 323,99 | 0,13 | 0,06 | 2,23 | 2,60E-02 |
| CCNA2 | 71,37 | 0,19 | 0,08 | 2,23 | 2,60E-02 |
| GALNT2 | 852,70 | 0,10 | 0,04 | 2,23 | 2,60E-02 |
| DERL1 | 2296,33 | -0,05 | 0,02 | -2,23 | 2,60E-02 |
| CHEK2 | 198,26 | 0,10 | 0,05 | 2,22 | 2,61E-02 |
| PLCB2 | 3889,58 | 0,19 | 0,09 | 2,22 | 2,61E-02 |

| | | | | | |
|---|---|---|---|---|---|
| HLA-A | 41509,28 | 0,19 | 0,08 | 2,22 | 2,61E-02 |
| XAB2 | 652,21 | 0,11 | 0,05 | 2,22 | 2,61E-02 |
| ABHD11 | 130,41 | 0,12 | 0,05 | 2,22 | 2,61E-02 |
| ADRM1 | 411,27 | 0,10 | 0,04 | 2,22 | 2,61E-02 |
| TEP1 | 3691,27 | 0,12 | 0,05 | 2,22 | 2,62E-02 |
| PTGER4 | 2135,16 | -0,08 | 0,04 | -2,22 | 2,62E-02 |
| ZNF778 | 688,42 | -0,05 | 0,02 | -2,22 | 2,62E-02 |
| GRPEL1 | 699,33 | -0,08 | 0,03 | -2,22 | 2,62E-02 |
| NSUN6 | 724,52 | -0,08 | 0,04 | -2,22 | 2,63E-02 |
| SPAST | 2056,78 | -0,10 | 0,05 | -2,22 | 2,63E-02 |
| KDM8 | 197,37 | 0,10 | 0,05 | 2,22 | 2,63E-02 |
| TMCO4 | 461,37 | 0,11 | 0,05 | 2,22 | 2,63E-02 |
| THAP3 | 279,35 | 0,07 | 0,03 | 2,22 | 2,63E-02 |
| TRAPPC6B | 1352,48 | -0,12 | 0,05 | -2,22 | 2,63E-02 |
| GNPDA1 | 525,96 | -0,07 | 0,03 | -2,22 | 2,64E-02 |
| CCDC66 | 1206,09 | -0,11 | 0,05 | -2,22 | 2,64E-02 |
| ANAPC13 | 707,24 | -0,12 | 0,05 | -2,22 | 2,64E-02 |
| NHLRC4 | 81,90 | 0,13 | 0,06 | 2,22 | 2,64E-02 |
| PLEKHG7 | 50,30 | 0,30 | 0,14 | 2,22 | 2,64E-02 |
| SLC25A38 | 822,53 | 0,10 | 0,04 | 2,22 | 2,64E-02 |
| ZNF518A | 2829,24 | -0,14 | 0,06 | -2,22 | 2,64E-02 |
| ANKRD52 | 591,79 | 0,17 | 0,08 | 2,22 | 2,64E-02 |
| EIF3L | 2555,70 | -0,08 | 0,04 | -2,22 | 2,64E-02 |
| PGP | 446,80 | 0,08 | 0,04 | 2,22 | 2,64E-02 |
| DOLPP1 | 162,69 | 0,08 | 0,03 | 2,22 | 2,65E-02 |
| KAT2A | 190,77 | 0,18 | 0,08 | 2,22 | 2,65E-02 |
| GNMT | 33,23 | 0,24 | 0,11 | 2,22 | 2,66E-02 |
| ARHGEF28 | 105,84 | 0,38 | 0,17 | 2,22 | 2,66E-02 |
| USP36 | 731,91 | 0,16 | 0,07 | 2,22 | 2,66E-02 |
| IGF2BP2 | 825,94 | 0,26 | 0,12 | 2,22 | 2,66E-02 |
| TRMT13 | 875,94 | -0,15 | 0,07 | -2,22 | 2,66E-02 |
| DESI2 | 1061,29 | -0,08 | 0,04 | -2,22 | 2,67E-02 |
| DHX8 | 2930,07 | -0,07 | 0,03 | -2,22 | 2,67E-02 |
| ZNF684 | 148,11 | -0,13 | 0,06 | -2,22 | 2,67E-02 |
| PLK3 | 401,27 | 0,17 | 0,08 | 2,22 | 2,67E-02 |
| STEAP1B | 118,74 | 0,27 | 0,12 | 2,22 | 2,67E-02 |
| KDM3A | 4579,42 | -0,07 | 0,03 | -2,22 | 2,67E-02 |
| CAMLG | 592,80 | -0,11 | 0,05 | -2,22 | 2,68E-02 |
| STRBP | 1072,97 | -0,15 | 0,07 | -2,21 | 2,68E-02 |
| GAA | 1003,00 | 0,20 | 0,09 | 2,21 | 2,68E-02 |
| SKAP2 | 3302,32 | -0,17 | 0,08 | -2,21 | 2,68E-02 |
| DGCR8 | 635,23 | 0,13 | 0,06 | 2,21 | 2,69E-02 |
| ITPKB | 3492,75 | 0,12 | 0,05 | 2,21 | 2,69E-02 |
| AOC3 | 104,75 | 0,31 | 0,14 | 2,21 | 2,69E-02 |
| NSMCE2 | 637,70 | -0,08 | 0,04 | -2,21 | 2,69E-02 |
| C5 | 239,66 | 0,12 | 0,06 | 2,21 | 2,69E-02 |

| | | | | | |
|---|---|---|---|---|---|
| EXOSC8 | 549,77 | -0,11 | 0,05 | -2,21 | 2,70E-02 |
| LSM14A | 4096,70 | -0,06 | 0,03 | -2,21 | 2,70E-02 |
| CCND3 | 3205,02 | 0,11 | 0,05 | 2,21 | 2,70E-02 |
| PLPP1 | 124,22 | 0,13 | 0,06 | 2,21 | 2,70E-02 |
| PRPSAP1 | 393,14 | 0,08 | 0,03 | 2,21 | 2,71E-02 |
| FAM161A | 45,38 | 0,22 | 0,10 | 2,21 | 2,71E-02 |
| SAPCD2 | 46,70 | 0,35 | 0,16 | 2,21 | 2,71E-02 |
| ITSN1 | 504,13 | 0,17 | 0,08 | 2,21 | 2,71E-02 |
| LCOR | 6495,30 | -0,07 | 0,03 | -2,21 | 2,72E-02 |
| ICAM1 | 654,92 | 0,15 | 0,07 | 2,21 | 2,72E-02 |
| CDK13 | 7074,57 | -0,05 | 0,02 | -2,21 | 2,72E-02 |
| PYGO2 | 384,02 | 0,15 | 0,07 | 2,21 | 2,73E-02 |
| IFT70B | 141,86 | -0,14 | 0,06 | -2,21 | 2,73E-02 |
| TAF12 | 707,09 | -0,09 | 0,04 | -2,21 | 2,73E-02 |
| FLCN | 1381,86 | 0,11 | 0,05 | 2,21 | 2,73E-02 |
| TMOD1 | 944,90 | 0,26 | 0,12 | 2,21 | 2,73E-02 |
| NUP37 | 309,58 | -0,11 | 0,05 | -2,21 | 2,73E-02 |
| MALT1 | 3725,95 | -0,09 | 0,04 | -2,21 | 2,73E-02 |
| PRKAB2 | 1039,09 | -0,07 | 0,03 | -2,21 | 2,73E-02 |
| CHRFAM7A | 25,19 | 0,28 | 0,13 | 2,21 | 2,73E-02 |
| FUNDC2 | 778,72 | 0,12 | 0,05 | 2,21 | 2,73E-02 |
| RCSD1 | 7421,53 | -0,08 | 0,04 | -2,21 | 2,74E-02 |
| TMED5 | 3018,24 | -0,11 | 0,05 | -2,20 | 2,75E-02 |
| NAPRT | 592,62 | 0,13 | 0,06 | 2,20 | 2,75E-02 |
| BLOC1S6 | 2396,40 | -0,12 | 0,06 | -2,20 | 2,75E-02 |
| DNASE1L3 | 32,69 | 0,27 | 0,12 | 2,20 | 2,75E-02 |
| SNRNP48 | 1081,35 | -0,12 | 0,06 | -2,20 | 2,75E-02 |
| PYROXD1 | 1090,55 | -0,13 | 0,06 | -2,20 | 2,75E-02 |
| PRR14 | 410,42 | 0,14 | 0,06 | 2,20 | 2,76E-02 |
| RBL1 | 961,36 | -0,10 | 0,04 | -2,20 | 2,76E-02 |
| GIMAP2 | 2244,63 | -0,18 | 0,08 | -2,20 | 2,76E-02 |
| CRYZL1 | 836,37 | -0,07 | 0,03 | -2,20 | 2,76E-02 |
| CDKN2AIP | 1027,13 | -0,13 | 0,06 | -2,20 | 2,76E-02 |
| LDHB | 754,97 | -0,17 | 0,08 | -2,20 | 2,77E-02 |
| SENP1 | 979,23 | -0,05 | 0,02 | -2,20 | 2,77E-02 |
| C15orf39 | 1823,02 | 0,24 | 0,11 | 2,20 | 2,77E-02 |
| DYNLT2B | 23,15 | 0,18 | 0,08 | 2,20 | 2,78E-02 |
| LYRM7 | 890,26 | -0,11 | 0,05 | -2,20 | 2,78E-02 |
| RGMB | 201,31 | 0,19 | 0,09 | 2,20 | 2,78E-02 |
| SH3PXD2A | 311,86 | 0,20 | 0,09 | 2,20 | 2,78E-02 |
| GSN | 2186,75 | 0,15 | 0,07 | 2,20 | 2,78E-02 |
| ZBTB7B | 2152,08 | 0,14 | 0,06 | 2,20 | 2,78E-02 |
| TOGARAM1 | 871,27 | -0,07 | 0,03 | -2,20 | 2,79E-02 |
| CSRNP2 | 541,09 | 0,08 | 0,03 | 2,20 | 2,79E-02 |
| COA3 | 368,15 | -0,10 | 0,04 | -2,20 | 2,79E-02 |
| CHPT1 | 2237,73 | 0,15 | 0,07 | 2,20 | 2,79E-02 |

| | | | | | |
|---|---|---|---|---|---|
| TFPI | 116,36 | 0,21 | 0,09 | 2,20 | 2,79E-02 |
| SIGMAR1 | 327,05 | 0,07 | 0,03 | 2,20 | 2,80E-02 |
| SCML2 | 45,22 | 0,18 | 0,08 | 2,20 | 2,80E-02 |
| ASF1B | 67,59 | 0,23 | 0,11 | 2,20 | 2,80E-02 |
| SLC25A20 | 1026,14 | -0,12 | 0,05 | -2,20 | 2,80E-02 |
| MLXIP | 1254,35 | 0,22 | 0,10 | 2,20 | 2,80E-02 |
| AP1S2 | 3140,77 | -0,19 | 0,09 | -2,20 | 2,81E-02 |
| PRCC | 427,42 | 0,09 | 0,04 | 2,20 | 2,81E-02 |
| PARP11 | 1179,97 | -0,10 | 0,04 | -2,20 | 2,81E-02 |
| SPATA25 | 25,07 | 0,16 | 0,07 | 2,20 | 2,81E-02 |
| ABR | 1824,00 | 0,19 | 0,09 | 2,20 | 2,81E-02 |
| NLRP3 | 1508,22 | -0,12 | 0,05 | -2,20 | 2,82E-02 |
| SUPT4H1 | 133,12 | -0,12 | 0,06 | -2,20 | 2,82E-02 |
| C14orf119 | 956,00 | -0,08 | 0,04 | -2,20 | 2,82E-02 |
| AMPD3 | 1890,69 | 0,10 | 0,05 | 2,19 | 2,82E-02 |
| H1-10 | 539,92 | 0,16 | 0,07 | 2,19 | 2,82E-02 |
| RBM17 | 1648,35 | -0,06 | 0,03 | -2,19 | 2,82E-02 |
| SPDL1 | 200,07 | -0,11 | 0,05 | -2,19 | 2,82E-02 |
| CPNE5 | 201,22 | 0,20 | 0,09 | 2,19 | 2,82E-02 |
| CRAMP1 | 338,33 | 0,16 | 0,07 | 2,19 | 2,82E-02 |
| MTERF3 | 269,22 | -0,08 | 0,04 | -2,19 | 2,82E-02 |
| SEC61G | 622,61 | -0,14 | 0,06 | -2,19 | 2,82E-02 |
| GALNT12 | 179,71 | 0,12 | 0,05 | 2,19 | 2,82E-02 |
| PSAT1 | 67,39 | 0,18 | 0,08 | 2,19 | 2,83E-02 |
| TRAF5 | 2309,41 | -0,10 | 0,05 | -2,19 | 2,83E-02 |
| PSMB5 | 437,44 | -0,08 | 0,04 | -2,19 | 2,83E-02 |
| APBB3 | 416,09 | 0,11 | 0,05 | 2,19 | 2,83E-02 |
| GTPBP10 | 1268,23 | -0,10 | 0,05 | -2,19 | 2,84E-02 |
| ERCC1 | 583,36 | 0,08 | 0,04 | 2,19 | 2,84E-02 |
| PDK4 | 987,56 | -0,21 | 0,10 | -2,19 | 2,84E-02 |
| PSPC1 | 3717,58 | -0,07 | 0,03 | -2,19 | 2,84E-02 |
| LEKR1 | 53,40 | 0,26 | 0,12 | 2,19 | 2,84E-02 |
| PLEKHA1 | 2896,92 | -0,12 | 0,05 | -2,19 | 2,84E-02 |
| GKAP1 | 212,26 | -0,12 | 0,05 | -2,19 | 2,84E-02 |
| ZNF485 | 141,79 | -0,11 | 0,05 | -2,19 | 2,84E-02 |
| ELF4 | 880,54 | 0,21 | 0,10 | 2,19 | 2,84E-02 |
| HS2ST1 | 976,86 | -0,10 | 0,05 | -2,19 | 2,85E-02 |
| FANCB | 221,82 | -0,13 | 0,06 | -2,19 | 2,85E-02 |
| IL5RA | 772,02 | 0,25 | 0,12 | 2,19 | 2,86E-02 |
| SIDT2 | 578,12 | 0,23 | 0,10 | 2,19 | 2,86E-02 |
| SLC6A16 | 168,38 | 0,16 | 0,07 | 2,19 | 2,86E-02 |
| FMNL1 | 3558,58 | 0,29 | 0,13 | 2,19 | 2,86E-02 |
| TBC1D25 | 303,81 | 0,15 | 0,07 | 2,19 | 2,86E-02 |
| NEMP1 | 899,67 | -0,09 | 0,04 | -2,19 | 2,86E-02 |
| SHISAL2A | 277,69 | 0,17 | 0,08 | 2,19 | 2,86E-02 |
| EXOC8 | 2538,82 | -0,10 | 0,04 | -2,19 | 2,87E-02 |

| | | | | | |
|---|---|---|---|---|---|
| KPNA3 | 860,67 | -0,09 | 0,04 | -2,19 | 2,87E-02 |
| NDUFB5 | 870,32 | -0,11 | 0,05 | -2,19 | 2,87E-02 |
| THNSL1 | 153,99 | -0,18 | 0,08 | -2,19 | 2,87E-02 |
| QRSL1 | 692,62 | -0,10 | 0,05 | -2,19 | 2,88E-02 |
| GGCX | 671,85 | 0,09 | 0,04 | 2,19 | 2,88E-02 |
| TSPAN14 | 3948,95 | 0,12 | 0,05 | 2,19 | 2,88E-02 |
| TTLL3 | 799,69 | 0,21 | 0,10 | 2,19 | 2,88E-02 |
| ABRAXAS2 | 282,73 | -0,07 | 0,03 | -2,19 | 2,88E-02 |
| DYRK2 | 5631,23 | -0,11 | 0,05 | -2,19 | 2,88E-02 |
| GUCD1 | 7036,24 | 0,20 | 0,09 | 2,19 | 2,88E-02 |
| WNT7A | 140,21 | 0,19 | 0,09 | 2,19 | 2,88E-02 |
| ZBTB47 | 344,73 | 0,13 | 0,06 | 2,19 | 2,88E-02 |
| CCZ1B | 598,88 | -0,10 | 0,05 | -2,19 | 2,89E-02 |
| ELP2 | 2540,23 | -0,08 | 0,04 | -2,19 | 2,89E-02 |
| SEMA3A | 36,83 | 0,34 | 0,15 | 2,18 | 2,89E-02 |
| PAOX | 174,29 | 0,11 | 0,05 | 2,18 | 2,89E-02 |
| PARM1 | 51,34 | 0,24 | 0,11 | 2,18 | 2,89E-02 |
| PDHA1 | 814,86 | -0,05 | 0,02 | -2,18 | 2,89E-02 |
| USP14 | 838,70 | -0,10 | 0,04 | -2,18 | 2,89E-02 |
| GYS1 | 351,42 | 0,13 | 0,06 | 2,18 | 2,89E-02 |
| TIMM29 | 690,11 | -0,07 | 0,03 | -2,18 | 2,90E-02 |
| MFSD14A | 286,47 | -0,10 | 0,05 | -2,18 | 2,90E-02 |
| CEP164 | 969,51 | 0,13 | 0,06 | 2,18 | 2,90E-02 |
| ZNF32 | 261,48 | -0,14 | 0,07 | -2,18 | 2,90E-02 |
| NUP160 | 1608,66 | -0,06 | 0,03 | -2,18 | 2,91E-02 |
| BBS7 | 305,61 | -0,10 | 0,05 | -2,18 | 2,91E-02 |
| SSBP3 | 575,21 | 0,16 | 0,08 | 2,18 | 2,91E-02 |
| NAGPA | 383,89 | 0,09 | 0,04 | 2,18 | 2,91E-02 |
| FGD2 | 1393,68 | 0,19 | 0,09 | 2,18 | 2,92E-02 |
| NNT | 2110,78 | -0,07 | 0,03 | -2,18 | 2,92E-02 |
| MAF1 | 3367,06 | 0,17 | 0,08 | 2,18 | 2,92E-02 |
| ATP5IF1 | 363,33 | -0,13 | 0,06 | -2,18 | 2,92E-02 |
| MCRIP1 | 369,65 | 0,09 | 0,04 | 2,18 | 2,92E-02 |
| TARS1 | 1080,55 | -0,09 | 0,04 | -2,18 | 2,92E-02 |
| ZNF467 | 548,99 | 0,20 | 0,09 | 2,18 | 2,92E-02 |
| STAU1 | 1363,26 | 0,12 | 0,06 | 2,18 | 2,93E-02 |
| ILK | 943,14 | 0,12 | 0,05 | 2,18 | 2,93E-02 |
| CD4 | 1231,12 | 0,15 | 0,07 | 2,18 | 2,93E-02 |
| NUS1 | 762,67 | -0,12 | 0,06 | -2,18 | 2,93E-02 |
| HPCAL4 | 132,17 | 0,20 | 0,09 | 2,18 | 2,94E-02 |
| ARHGEF1 | 4150,12 | 0,20 | 0,09 | 2,18 | 2,94E-02 |
| FBXO5 | 193,07 | -0,14 | 0,06 | -2,18 | 2,95E-02 |
| RORA | 8281,35 | -0,14 | 0,06 | -2,18 | 2,95E-02 |
| CTTN | 500,61 | 0,23 | 0,11 | 2,18 | 2,95E-02 |
| CHSY1 | 2625,39 | -0,13 | 0,06 | -2,18 | 2,95E-02 |
| UBE2A | 855,15 | -0,09 | 0,04 | -2,18 | 2,95E-02 |

| | | | | | |
|---|---|---|---|---|---|
| RAB12 | 684,59 | -0,08 | 0,04 | -2,18 | 2,95E-02 |
| TCOF1 | 656,33 | 0,13 | 0,06 | 2,18 | 2,96E-02 |
| NFKBIE | 457,56 | 0,09 | 0,04 | 2,18 | 2,96E-02 |
| CENPQ | 105,54 | -0,21 | 0,10 | -2,18 | 2,96E-02 |
| ZNF799 | 325,10 | -0,09 | 0,04 | -2,18 | 2,96E-02 |
| TADA1 | 315,88 | -0,09 | 0,04 | -2,17 | 2,96E-02 |
| EP400 | 1507,36 | 0,20 | 0,09 | 2,17 | 2,96E-02 |
| PEX13 | 1160,77 | -0,09 | 0,04 | -2,17 | 2,97E-02 |
| PAIP2B | 401,60 | -0,12 | 0,05 | -2,17 | 2,97E-02 |
| GRK2 | 4978,10 | 0,15 | 0,07 | 2,17 | 2,97E-02 |
| VPS13C | 19145,12 | -0,11 | 0,05 | -2,17 | 2,97E-02 |
| KLRD1 | 5402,25 | -0,18 | 0,08 | -2,17 | 2,98E-02 |
| RUSF1 | 528,77 | 0,10 | 0,05 | 2,17 | 2,98E-02 |
| PHF14 | 2148,01 | -0,13 | 0,06 | -2,17 | 2,98E-02 |
| PCP2 | 31,91 | 0,18 | 0,08 | 2,17 | 2,99E-02 |
| H3-3B | 29505,56 | -0,13 | 0,06 | -2,17 | 2,99E-02 |
| ARFIP1 | 2074,05 | -0,13 | 0,06 | -2,17 | 2,99E-02 |
| NUP43 | 1373,53 | -0,08 | 0,04 | -2,17 | 2,99E-02 |
| RARA | 1498,26 | 0,19 | 0,09 | 2,17 | 2,99E-02 |
| ATF7IP | 7904,56 | -0,08 | 0,04 | -2,17 | 3,00E-02 |
| IQCE | 750,57 | 0,13 | 0,06 | 2,17 | 3,01E-02 |
| EEF2KMT | 121,34 | 0,10 | 0,05 | 2,17 | 3,01E-02 |
| MRPS30 | 1450,17 | -0,11 | 0,05 | -2,17 | 3,02E-02 |
| WDTC1 | 2218,85 | 0,14 | 0,07 | 2,17 | 3,02E-02 |
| CASD1 | 1546,65 | -0,14 | 0,07 | -2,17 | 3,03E-02 |
| ZC3H4 | 947,95 | 0,11 | 0,05 | 2,17 | 3,03E-02 |
| PHB1 | 1055,42 | -0,11 | 0,05 | -2,17 | 3,03E-02 |
| RBCK1 | 1355,33 | 0,15 | 0,07 | 2,17 | 3,03E-02 |
| KCTD13 | 778,32 | 0,12 | 0,06 | 2,17 | 3,03E-02 |
| MED13 | 7444,55 | -0,07 | 0,03 | -2,17 | 3,03E-02 |
| AUP1 | 1298,08 | 0,06 | 0,03 | 2,17 | 3,03E-02 |
| PI4KB | 1284,03 | 0,11 | 0,05 | 2,17 | 3,04E-02 |
| ZBTB22 | 357,15 | 0,09 | 0,04 | 2,17 | 3,04E-02 |
| LRRC63 | 31,15 | 0,19 | 0,09 | 2,17 | 3,04E-02 |
| TRAPPC13 | 689,96 | -0,12 | 0,06 | -2,16 | 3,04E-02 |
| SOS1 | 3069,68 | -0,10 | 0,04 | -2,16 | 3,04E-02 |
| KDM6B | 2364,53 | 0,26 | 0,12 | 2,16 | 3,04E-02 |
| UGCG | 1549,04 | -0,12 | 0,06 | -2,16 | 3,04E-02 |
| FAM136A | 387,37 | -0,08 | 0,04 | -2,16 | 3,05E-02 |
| SLC39A3 | 197,81 | 0,10 | 0,05 | 2,16 | 3,05E-02 |
| PSMD12 | 1410,54 | -0,07 | 0,03 | -2,16 | 3,05E-02 |
| RAP2A | 1625,37 | -0,17 | 0,08 | -2,16 | 3,05E-02 |
| UVRAG | 2246,41 | -0,07 | 0,03 | -2,16 | 3,05E-02 |
| NOB1 | 148,88 | 0,11 | 0,05 | 2,16 | 3,05E-02 |
| VPS26A | 842,89 | -0,09 | 0,04 | -2,16 | 3,05E-02 |
| PDP1 | 1605,53 | -0,11 | 0,05 | -2,16 | 3,05E-02 |

| | | | | | |
|---|---|---|---|---|---|
| MAP7 | 140,78 | 0,18 | 0,08 | 2,16 | 3,05E-02 |
| TACC3 | 1728,26 | 0,17 | 0,08 | 2,16 | 3,05E-02 |
| SPATS2 | 154,63 | 0,14 | 0,07 | 2,16 | 3,05E-02 |
| POGLUT1 | 1042,36 | -0,07 | 0,03 | -2,16 | 3,06E-02 |
| NUDT5 | 1739,37 | -0,08 | 0,04 | -2,16 | 3,07E-02 |
| PIGF | 675,96 | -0,11 | 0,05 | -2,16 | 3,08E-02 |
| MTCH2 | 702,55 | -0,07 | 0,03 | -2,16 | 3,08E-02 |
| HEATR5B | 3791,61 | -0,08 | 0,04 | -2,16 | 3,08E-02 |
| KCNQ1 | 705,27 | 0,16 | 0,07 | 2,16 | 3,09E-02 |
| PICALM | 21362,68 | -0,11 | 0,05 | -2,16 | 3,09E-02 |
| DHX57 | 839,31 | -0,06 | 0,03 | -2,16 | 3,10E-02 |
| ATPAF2 | 392,38 | 0,09 | 0,04 | 2,16 | 3,10E-02 |
| PIK3CG | 4226,67 | -0,08 | 0,04 | -2,16 | 3,10E-02 |
| ARHGEF18 | 259,38 | 0,15 | 0,07 | 2,16 | 3,10E-02 |
| DEPDC7 | 33,79 | 0,24 | 0,11 | 2,16 | 3,10E-02 |
| RBM7 | 704,70 | -0,10 | 0,05 | -2,16 | 3,10E-02 |
| MBTD1 | 2224,41 | -0,07 | 0,03 | -2,16 | 3,10E-02 |
| PPM1B | 1868,42 | -0,14 | 0,06 | -2,16 | 3,10E-02 |
| ZNF416 | 231,03 | -0,10 | 0,05 | -2,16 | 3,10E-02 |
| SNX3 | 3854,62 | 0,18 | 0,08 | 2,16 | 3,11E-02 |
| SON | 14313,61 | -0,06 | 0,03 | -2,16 | 3,11E-02 |
| RPL24 | 4474,61 | -0,21 | 0,10 | -2,16 | 3,11E-02 |
| RBM12B | 3719,45 | -0,14 | 0,07 | -2,16 | 3,11E-02 |
| JMJD7-PLA2G4B | 318,83 | 0,18 | 0,08 | 2,16 | 3,11E-02 |
| DYNC2I2 | 98,70 | 0,16 | 0,07 | 2,15 | 3,12E-02 |
| ARID5B | 2072,76 | -0,10 | 0,05 | -2,15 | 3,12E-02 |
| NCALD | 762,66 | -0,14 | 0,07 | -2,15 | 3,12E-02 |
| PIM3 | 1147,69 | 0,12 | 0,06 | 2,15 | 3,13E-02 |
| AQP10 | 61,79 | 0,29 | 0,14 | 2,15 | 3,13E-02 |
| TNFRSF14 | 2190,52 | 0,12 | 0,05 | 2,15 | 3,13E-02 |
| KLF3 | 6628,97 | -0,08 | 0,03 | -2,15 | 3,13E-02 |
| TPMT | 385,15 | -0,13 | 0,06 | -2,15 | 3,13E-02 |
| NPIPB3 | 69,44 | 0,27 | 0,12 | 2,15 | 3,14E-02 |
| STRN4 | 1144,85 | 0,15 | 0,07 | 2,15 | 3,14E-02 |
| RNF10 | 10155,33 | 0,22 | 0,10 | 2,15 | 3,14E-02 |
| TEC | 429,13 | 0,12 | 0,06 | 2,15 | 3,15E-02 |
| CDKL3 | 39,59 | 0,14 | 0,07 | 2,15 | 3,15E-02 |
| PTGDR2 | 441,97 | 0,23 | 0,11 | 2,15 | 3,15E-02 |
| P2RX4 | 630,59 | 0,08 | 0,04 | 2,15 | 3,15E-02 |
| ENSG00000278384 | 142,15 | 0,42 | 0,19 | 2,15 | 3,16E-02 |
| PRIM1 | 216,16 | -0,11 | 0,05 | -2,15 | 3,17E-02 |
| MYO15B | 2110,68 | 0,20 | 0,10 | 2,15 | 3,17E-02 |
| ELOC | 473,81 | -0,08 | 0,04 | -2,15 | 3,17E-02 |
| C1orf52 | 575,80 | -0,08 | 0,04 | -2,15 | 3,17E-02 |
| CHN1 | 81,05 | 0,21 | 0,10 | 2,15 | 3,18E-02 |
| CDK4 | 175,02 | 0,09 | 0,04 | 2,15 | 3,18E-02 |

| | | | | | |
|---|---|---|---|---|---|
| SIGLEC1 | 203,01 | 0,47 | 0,22 | 2,15 | 3,18E-02 |
| NBEAL2 | 5600,72 | 0,27 | 0,12 | 2,15 | 3,18E-02 |
| ENSG00000112096 | 52483,24 | -0,15 | 0,07 | -2,15 | 3,19E-02 |
| NDUFA8 | 244,28 | -0,10 | 0,05 | -2,15 | 3,19E-02 |
| GMIP | 2396,66 | 0,23 | 0,11 | 2,15 | 3,19E-02 |
| EAF2 | 411,69 | -0,15 | 0,07 | -2,15 | 3,19E-02 |
| CDKN1B | 6875,37 | -0,09 | 0,04 | -2,15 | 3,19E-02 |
| GTF2A1 | 2499,39 | -0,10 | 0,05 | -2,15 | 3,20E-02 |
| MLX | 1399,31 | 0,06 | 0,03 | 2,14 | 3,20E-02 |
| CRYL1 | 344,47 | 0,08 | 0,04 | 2,14 | 3,20E-02 |
| ARL6IP5 | 4326,03 | -0,11 | 0,05 | -2,14 | 3,20E-02 |
| PRDX6 | 5640,01 | 0,22 | 0,10 | 2,14 | 3,20E-02 |
| GLCCI1 | 1189,03 | -0,09 | 0,04 | -2,14 | 3,20E-02 |
| INTS11 | 810,94 | 0,11 | 0,05 | 2,14 | 3,21E-02 |
| YTHDF3 | 4005,84 | -0,09 | 0,04 | -2,14 | 3,21E-02 |
| PRMT7 | 822,10 | 0,09 | 0,04 | 2,14 | 3,21E-02 |
| MCM7 | 932,41 | 0,09 | 0,04 | 2,14 | 3,21E-02 |
| LRCH4 | 1088,77 | 0,12 | 0,06 | 2,14 | 3,22E-02 |
| NLRX1 | 356,26 | 0,16 | 0,08 | 2,14 | 3,22E-02 |
| ZNF407 | 3418,67 | -0,09 | 0,04 | -2,14 | 3,22E-02 |
| FAR1 | 5182,49 | -0,15 | 0,07 | -2,14 | 3,23E-02 |
| PCNT | 1886,92 | 0,11 | 0,05 | 2,14 | 3,23E-02 |
| LYSMD3 | 1453,53 | -0,18 | 0,08 | -2,14 | 3,24E-02 |
| TPGS2 | 1698,06 | 0,10 | 0,05 | 2,14 | 3,24E-02 |
| ERCC5 | 649,17 | -0,07 | 0,03 | -2,14 | 3,25E-02 |
| ANKRD55 | 164,13 | 0,20 | 0,09 | 2,14 | 3,25E-02 |
| TAF4 | 433,54 | 0,18 | 0,08 | 2,14 | 3,25E-02 |
| CCPG1 | 2514,41 | -0,12 | 0,05 | -2,14 | 3,25E-02 |
| POP5 | 341,23 | -0,09 | 0,04 | -2,14 | 3,25E-02 |
| SAMTOR | 986,53 | -0,12 | 0,06 | -2,14 | 3,26E-02 |
| TOMM40 | 280,38 | 0,10 | 0,05 | 2,14 | 3,26E-02 |
| FAM131B | 60,81 | 0,23 | 0,11 | 2,14 | 3,26E-02 |
| TTC17 | 5067,99 | -0,07 | 0,03 | -2,14 | 3,26E-02 |
| ST7 | 196,20 | 0,10 | 0,05 | 2,14 | 3,27E-02 |
| PARK7 | 1774,84 | -0,11 | 0,05 | -2,14 | 3,27E-02 |
| FBXO36 | 33,42 | 0,18 | 0,08 | 2,14 | 3,27E-02 |
| ERI1 | 747,99 | -0,10 | 0,05 | -2,14 | 3,27E-02 |
| PIGO | 402,69 | 0,11 | 0,05 | 2,14 | 3,27E-02 |
| PRKCSH | 971,39 | 0,14 | 0,07 | 2,13 | 3,28E-02 |
| RYK | 1385,17 | -0,08 | 0,04 | -2,13 | 3,28E-02 |
| DGKA | 3567,31 | 0,12 | 0,06 | 2,13 | 3,28E-02 |
| NUP85 | 596,49 | 0,10 | 0,05 | 2,13 | 3,28E-02 |
| PTPN18 | 1434,79 | 0,13 | 0,06 | 2,13 | 3,28E-02 |
| ATP11C | 1960,01 | -0,09 | 0,04 | -2,13 | 3,29E-02 |
| GUF1 | 1011,58 | -0,12 | 0,05 | -2,13 | 3,29E-02 |
| NUDT15 | 109,92 | -0,11 | 0,05 | -2,13 | 3,29E-02 |

| | | | | | |
|---|---|---|---|---|---|
| DNAJC15 | 681,02 | -0,15 | 0,07 | -2,13 | 3,30E-02 |
| KL | 49,00 | 0,26 | 0,12 | 2,13 | 3,30E-02 |
| CLSPN | 84,50 | 0,23 | 0,11 | 2,13 | 3,30E-02 |
| ZBTB6 | 1000,99 | -0,15 | 0,07 | -2,13 | 3,30E-02 |
| SLC16A4 | 36,35 | 0,18 | 0,08 | 2,13 | 3,30E-02 |
| SECTM1 | 1821,82 | 0,19 | 0,09 | 2,13 | 3,30E-02 |
| TMA16 | 551,30 | -0,11 | 0,05 | -2,13 | 3,30E-02 |
| GNA13 | 6240,47 | -0,09 | 0,04 | -2,13 | 3,30E-02 |
| EZH2 | 323,54 | 0,11 | 0,05 | 2,13 | 3,30E-02 |
| CACNA2D2 | 116,20 | 0,31 | 0,15 | 2,13 | 3,30E-02 |
| MBIP | 307,46 | -0,13 | 0,06 | -2,13 | 3,30E-02 |
| HDAC5 | 951,06 | 0,16 | 0,07 | 2,13 | 3,30E-02 |
| C4orf3 | 3197,20 | -0,14 | 0,07 | -2,13 | 3,31E-02 |
| NFKBIL1 | 90,62 | 0,11 | 0,05 | 2,13 | 3,31E-02 |
| CAPNS1 | 473,43 | 0,13 | 0,06 | 2,13 | 3,31E-02 |
| CSGALNACT2 | 2441,39 | -0,09 | 0,04 | -2,13 | 3,31E-02 |
| PDIA3 | 1287,20 | -0,08 | 0,04 | -2,13 | 3,31E-02 |
| OSCAR | 349,06 | 0,18 | 0,08 | 2,13 | 3,32E-02 |
| ARRDC4 | 563,05 | -0,18 | 0,08 | -2,13 | 3,32E-02 |
| RPL11 | 4967,57 | -0,23 | 0,11 | -2,13 | 3,32E-02 |
| AEBP2 | 1463,68 | -0,09 | 0,04 | -2,13 | 3,33E-02 |
| PPFIBP1 | 165,04 | 0,18 | 0,08 | 2,13 | 3,33E-02 |
| ZNF655 | 4346,91 | -0,06 | 0,03 | -2,13 | 3,33E-02 |
| ELP1 | 1841,60 | -0,07 | 0,03 | -2,13 | 3,33E-02 |
| CCNI | 6298,68 | 0,14 | 0,07 | 2,13 | 3,33E-02 |
| SLFN12L | 4125,79 | -0,11 | 0,05 | -2,13 | 3,33E-02 |
| CFAP410 | 319,40 | 0,12 | 0,06 | 2,13 | 3,34E-02 |
| RETREG2 | 2514,63 | 0,11 | 0,05 | 2,13 | 3,34E-02 |
| APTX | 537,58 | -0,05 | 0,02 | -2,13 | 3,34E-02 |
| PDP2 | 291,82 | 0,09 | 0,04 | 2,13 | 3,34E-02 |
| NEK7 | 5697,55 | -0,13 | 0,06 | -2,13 | 3,35E-02 |
| DBF4B | 134,70 | 0,17 | 0,08 | 2,13 | 3,34E-02 |
| PSKH1 | 341,97 | 0,12 | 0,06 | 2,13 | 3,35E-02 |
| PANX1 | 528,88 | -0,09 | 0,04 | -2,13 | 3,35E-02 |
| C3orf38 | 1083,39 | -0,14 | 0,07 | -2,13 | 3,35E-02 |
| SASS6 | 232,56 | -0,10 | 0,05 | -2,13 | 3,35E-02 |
| CD3D | 1088,00 | -0,17 | 0,08 | -2,13 | 3,35E-02 |
| JUP | 108,43 | 0,36 | 0,17 | 2,13 | 3,35E-02 |
| SRRM1 | 1999,19 | -0,06 | 0,03 | -2,13 | 3,35E-02 |
| CEP44 | 971,54 | -0,10 | 0,05 | -2,12 | 3,36E-02 |
| DIS3L | 835,88 | -0,06 | 0,03 | -2,12 | 3,36E-02 |
| RPE | 2395,72 | -0,11 | 0,05 | -2,12 | 3,36E-02 |
| ZNF678 | 824,05 | -0,16 | 0,07 | -2,12 | 3,36E-02 |
| WNT10B | 67,17 | 0,19 | 0,09 | 2,12 | 3,36E-02 |
| ZBTB41 | 1037,44 | -0,20 | 0,10 | -2,12 | 3,36E-02 |
| ZNF425 | 84,35 | 0,12 | 0,05 | 2,12 | 3,37E-02 |

| | | | | | |
|---|---|---|---|---|---|
| ZNG1B | 1393,66 | -0,12 | 0,05 | -2,12 | 3,37E-02 |
| LGR6 | 118,97 | 0,27 | 0,13 | 2,12 | 3,37E-02 |
| CISD2 | 1125,30 | 0,19 | 0,09 | 2,12 | 3,37E-02 |
| AGPAT5 | 736,29 | -0,13 | 0,06 | -2,12 | 3,37E-02 |
| HAVCR2 | 777,62 | -0,10 | 0,05 | -2,12 | 3,37E-02 |
| TOR1AIP1 | 3410,88 | -0,07 | 0,04 | -2,12 | 3,37E-02 |
| GGH | 54,53 | 0,20 | 0,09 | 2,12 | 3,37E-02 |
| TDP2 | 2886,68 | -0,12 | 0,05 | -2,12 | 3,37E-02 |
| POMP | 728,75 | -0,12 | 0,06 | -2,12 | 3,38E-02 |
| ZNF626 | 383,52 | -0,14 | 0,07 | -2,12 | 3,38E-02 |
| ATP13A3 | 3225,23 | -0,08 | 0,04 | -2,12 | 3,39E-02 |
| CTSB | 10048,66 | 0,16 | 0,07 | 2,12 | 3,39E-02 |
| SPRY1 | 53,51 | 0,24 | 0,11 | 2,12 | 3,39E-02 |
| ATG4B | 722,44 | 0,12 | 0,05 | 2,12 | 3,39E-02 |
| UBA7 | 1383,25 | 0,14 | 0,07 | 2,12 | 3,40E-02 |
| ALG3 | 174,81 | 0,07 | 0,03 | 2,12 | 3,40E-02 |
| PEX26 | 1330,56 | 0,09 | 0,04 | 2,12 | 3,40E-02 |
| ARRB2 | 4287,54 | 0,16 | 0,07 | 2,12 | 3,40E-02 |
| VPS33B | 103,01 | 0,08 | 0,04 | 2,12 | 3,40E-02 |
| KLHL15 | 639,59 | -0,09 | 0,04 | -2,12 | 3,41E-02 |
| PRKCI | 693,02 | -0,10 | 0,05 | -2,12 | 3,41E-02 |
| KIAA1143 | 1354,25 | -0,09 | 0,04 | -2,12 | 3,41E-02 |
| GPR65 | 2420,03 | -0,15 | 0,07 | -2,12 | 3,42E-02 |
| CDC34 | 5495,57 | 0,22 | 0,11 | 2,12 | 3,42E-02 |
| PDHX | 460,96 | -0,09 | 0,04 | -2,12 | 3,42E-02 |
| USP24 | 7048,28 | -0,07 | 0,03 | -2,12 | 3,42E-02 |
| PANK3 | 3592,78 | -0,09 | 0,04 | -2,12 | 3,42E-02 |
| WBP2NL | 124,94 | 0,12 | 0,06 | 2,12 | 3,43E-02 |
| ASAH1 | 11113,96 | -0,12 | 0,06 | -2,12 | 3,43E-02 |
| GMDS | 360,78 | 0,13 | 0,06 | 2,12 | 3,43E-02 |
| MRTFA | 1536,78 | 0,13 | 0,06 | 2,12 | 3,43E-02 |
| CSKMT | 134,86 | -0,10 | 0,05 | -2,12 | 3,43E-02 |
| NT5DC1 | 712,26 | -0,08 | 0,04 | -2,12 | 3,43E-02 |
| AKT3 | 2188,25 | -0,10 | 0,05 | -2,12 | 3,43E-02 |
| ZNF496 | 221,39 | 0,21 | 0,10 | 2,12 | 3,44E-02 |
| SEMA4F | 324,05 | 0,10 | 0,05 | 2,11 | 3,45E-02 |
| MED21 | 883,63 | -0,09 | 0,04 | -2,11 | 3,45E-02 |
| LRRN1 | 97,86 | 0,21 | 0,10 | 2,11 | 3,45E-02 |
| THADA | 3060,74 | -0,07 | 0,03 | -2,11 | 3,45E-02 |
| C8orf82 | 214,53 | 0,10 | 0,05 | 2,11 | 3,45E-02 |
| IQGAP1 | 24338,52 | -0,11 | 0,05 | -2,11 | 3,46E-02 |
| DLEU7 | 49,65 | 0,20 | 0,09 | 2,11 | 3,46E-02 |
| MXI1 | 6472,54 | 0,22 | 0,10 | 2,11 | 3,46E-02 |
| KRT1 | 1181,01 | 0,37 | 0,17 | 2,11 | 3,46E-02 |
| EDC3 | 449,09 | 0,10 | 0,05 | 2,11 | 3,46E-02 |
| ZNF107 | 1360,13 | -0,15 | 0,07 | -2,11 | 3,46E-02 |

| | | | | | |
|---|---|---|---|---|---|
| ZNF616 | 291,08 | -0,12 | 0,06 | -2,11 | 3,48E-02 |
| ACAP2 | 13517,04 | -0,10 | 0,05 | -2,11 | 3,48E-02 |
| PPP2R5D | 206,01 | 0,15 | 0,07 | 2,11 | 3,48E-02 |
| FAM184A | 125,65 | 0,19 | 0,09 | 2,11 | 3,49E-02 |
| CSRP1 | 818,57 | 0,06 | 0,03 | 2,11 | 3,49E-02 |
| YWHAG | 1438,60 | -0,06 | 0,03 | -2,11 | 3,49E-02 |
| MZF1 | 644,54 | 0,11 | 0,05 | 2,11 | 3,50E-02 |
| NAA60 | 551,69 | 0,09 | 0,04 | 2,11 | 3,50E-02 |
| POLR2J3 | 418,21 | 0,17 | 0,08 | 2,11 | 3,50E-02 |
| NUDT19 | 312,16 | 0,07 | 0,03 | 2,11 | 3,50E-02 |
| RAB29 | 1852,60 | -0,08 | 0,04 | -2,11 | 3,50E-02 |
| KCNA3 | 1078,04 | 0,11 | 0,05 | 2,11 | 3,50E-02 |
| NLE1 | 320,79 | 0,10 | 0,05 | 2,11 | 3,51E-02 |
| INAFM1 | 135,09 | 0,12 | 0,06 | 2,11 | 3,51E-02 |
| TESK2 | 440,97 | 0,09 | 0,04 | 2,11 | 3,51E-02 |
| DDOST | 2233,19 | -0,07 | 0,03 | -2,11 | 3,51E-02 |
| GNG7 | 171,02 | 0,18 | 0,09 | 2,11 | 3,52E-02 |
| ARSB | 896,98 | 0,10 | 0,05 | 2,11 | 3,52E-02 |
| HERPUD2 | 2192,72 | -0,06 | 0,03 | -2,11 | 3,52E-02 |
| CD7 | 143,51 | 0,20 | 0,09 | 2,11 | 3,53E-02 |
| ANKRD37 | 78,47 | 0,13 | 0,06 | 2,10 | 3,53E-02 |
| ZDHHC24 | 406,06 | 0,12 | 0,06 | 2,10 | 3,53E-02 |
| ATP5PO | 427,73 | -0,12 | 0,06 | -2,10 | 3,53E-02 |
| CCNC | 1116,79 | -0,14 | 0,07 | -2,10 | 3,54E-02 |
| SFI1 | 2133,89 | 0,12 | 0,06 | 2,10 | 3,55E-02 |
| CNOT4 | 1217,97 | -0,07 | 0,03 | -2,10 | 3,55E-02 |
| B3GALNT1 | 47,72 | 0,24 | 0,11 | 2,10 | 3,55E-02 |
| OXCT1 | 867,05 | -0,10 | 0,05 | -2,10 | 3,55E-02 |
| RIN3 | 1325,32 | 0,22 | 0,10 | 2,10 | 3,55E-02 |
| LDB1 | 1263,26 | 0,09 | 0,04 | 2,10 | 3,55E-02 |
| RELCH | 5736,34 | -0,09 | 0,04 | -2,10 | 3,56E-02 |
| BORA | 290,75 | -0,11 | 0,05 | -2,10 | 3,57E-02 |
| TMEM208 | 409,18 | -0,08 | 0,04 | -2,10 | 3,57E-02 |
| FGF9 | 50,93 | 0,25 | 0,12 | 2,10 | 3,57E-02 |
| HIVEP3 | 361,01 | 0,21 | 0,10 | 2,10 | 3,57E-02 |
| MIF4GD | 205,37 | 0,08 | 0,04 | 2,10 | 3,57E-02 |
| ZFP36 | 1479,47 | 0,14 | 0,07 | 2,10 | 3,58E-02 |
| TRIM41 | 1034,88 | 0,06 | 0,03 | 2,10 | 3,58E-02 |
| GFER | 186,59 | 0,08 | 0,04 | 2,10 | 3,58E-02 |
| RPS6 | 11296,14 | -0,19 | 0,09 | -2,10 | 3,58E-02 |
| FANCD2 | 765,88 | -0,06 | 0,03 | -2,10 | 3,58E-02 |
| EIF3G | 3164,27 | -0,09 | 0,04 | -2,10 | 3,58E-02 |
| MRPL40 | 140,67 | -0,15 | 0,07 | -2,10 | 3,58E-02 |
| ZNF106 | 9162,36 | -0,09 | 0,05 | -2,10 | 3,58E-02 |
| OSTM1 | 2598,56 | -0,12 | 0,06 | -2,10 | 3,58E-02 |
| KIAA0319 | 302,31 | 0,21 | 0,10 | 2,10 | 3,59E-02 |

| | | | | | |
|---|---|---|---|---|---|
| AATK | 1098,05 | 0,24 | 0,11 | 2,10 | 3,59E-02 |
| MRPL1 | 218,94 | -0,16 | 0,07 | -2,10 | 3,60E-02 |
| TTC8 | 106,35 | 0,15 | 0,07 | 2,10 | 3,60E-02 |
| ECPAS | 3204,95 | -0,06 | 0,03 | -2,10 | 3,60E-02 |
| AGL | 1134,44 | -0,09 | 0,04 | -2,10 | 3,60E-02 |
| N4BP1 | 3663,57 | -0,10 | 0,05 | -2,10 | 3,60E-02 |
| CS | 978,21 | 0,10 | 0,05 | 2,10 | 3,60E-02 |
| LRP10 | 3345,85 | 0,20 | 0,10 | 2,10 | 3,61E-02 |
| ATP5MC3 | 1161,08 | -0,10 | 0,05 | -2,10 | 3,61E-02 |
| LMAN1 | 1744,73 | -0,11 | 0,05 | -2,10 | 3,61E-02 |
| RAB28 | 667,78 | -0,09 | 0,04 | -2,10 | 3,61E-02 |
| USP25 | 5373,00 | -0,10 | 0,05 | -2,10 | 3,62E-02 |
| SLC16A5 | 339,02 | 0,18 | 0,08 | 2,09 | 3,62E-02 |
| RANBP3 | 757,66 | 0,15 | 0,07 | 2,09 | 3,62E-02 |
| SLC38A2 | 7013,85 | -0,12 | 0,06 | -2,09 | 3,62E-02 |
| AMZ2 | 844,24 | -0,07 | 0,03 | -2,09 | 3,63E-02 |
| DTD2 | 227,78 | -0,09 | 0,04 | -2,09 | 3,63E-02 |
| AASDH | 626,97 | -0,09 | 0,05 | -2,09 | 3,63E-02 |
| KLHL9 | 2187,09 | -0,13 | 0,06 | -2,09 | 3,63E-02 |
| MGME1 | 670,92 | -0,09 | 0,04 | -2,09 | 3,63E-02 |
| ITGAX | 6519,62 | 0,26 | 0,12 | 2,09 | 3,63E-02 |
| MCM3AP | 2320,97 | 0,11 | 0,05 | 2,09 | 3,64E-02 |
| MIS18A | 65,91 | 0,12 | 0,06 | 2,09 | 3,64E-02 |
| TMEM67 | 176,06 | 0,12 | 0,06 | 2,09 | 3,64E-02 |
| TMPRSS9 | 405,13 | 0,30 | 0,15 | 2,09 | 3,64E-02 |
| HEATR9 | 43,93 | 0,30 | 0,14 | 2,09 | 3,64E-02 |
| NR4A2 | 95,54 | 0,12 | 0,06 | 2,09 | 3,65E-02 |
| MS4A3 | 169,41 | 0,30 | 0,14 | 2,09 | 3,65E-02 |
| ZNF680 | 532,05 | -0,16 | 0,07 | -2,09 | 3,64E-02 |
| KAT6A | 11381,61 | -0,08 | 0,04 | -2,09 | 3,64E-02 |
| ASF1A | 794,29 | -0,14 | 0,07 | -2,09 | 3,65E-02 |
| TNPO1 | 7800,18 | -0,11 | 0,05 | -2,09 | 3,65E-02 |
| PIGBOS1 | 245,10 | -0,08 | 0,04 | -2,09 | 3,65E-02 |
| NSUN2 | 2284,56 | -0,07 | 0,03 | -2,09 | 3,65E-02 |
| LANCL3 | 145,46 | 0,16 | 0,08 | 2,09 | 3,65E-02 |
| ABT1 | 1429,28 | -0,08 | 0,04 | -2,09 | 3,65E-02 |
| VNN2 | 19651,11 | -0,19 | 0,09 | -2,09 | 3,66E-02 |
| KYAT3 | 577,05 | -0,08 | 0,04 | -2,09 | 3,66E-02 |
| ST3GAL3 | 485,40 | 0,15 | 0,07 | 2,09 | 3,66E-02 |
| SLC25A24 | 1486,68 | -0,10 | 0,05 | -2,09 | 3,66E-02 |
| C7orf25 | 136,75 | -0,12 | 0,06 | -2,09 | 3,67E-02 |
| CMTM2 | 1892,05 | -0,20 | 0,10 | -2,09 | 3,68E-02 |
| BCL2L13 | 1390,11 | 0,09 | 0,04 | 2,09 | 3,68E-02 |
| STIMATE | 89,62 | 0,13 | 0,06 | 2,09 | 3,68E-02 |
| ACAA1 | 1778,92 | 0,10 | 0,05 | 2,09 | 3,68E-02 |
| CEP126 | 175,28 | -0,14 | 0,07 | -2,09 | 3,68E-02 |

| | | | | | |
|---|---|---|---|---|---|
| RIC8B | 827,25 | -0,08 | 0,04 | -2,09 | 3,68E-02 |
| THAP9 | 145,10 | -0,15 | 0,07 | -2,09 | 3,68E-02 |
| HDAC3 | 655,47 | 0,07 | 0,03 | 2,09 | 3,69E-02 |
| RAVER2 | 157,17 | 0,24 | 0,11 | 2,09 | 3,69E-02 |
| FIS1 | 2254,95 | 0,17 | 0,08 | 2,09 | 3,69E-02 |
| HNRNPA0 | 1433,88 | -0,09 | 0,04 | -2,09 | 3,70E-02 |
| ZBTB10 | 795,23 | -0,14 | 0,07 | -2,09 | 3,71E-02 |
| HEATR1 | 2142,57 | -0,08 | 0,04 | -2,08 | 3,71E-02 |
| FAM169A | 644,37 | -0,14 | 0,07 | -2,08 | 3,71E-02 |
| ATE1 | 988,53 | -0,05 | 0,02 | -2,08 | 3,71E-02 |
| N4BP2L2 | 12885,98 | -0,08 | 0,04 | -2,08 | 3,72E-02 |
| BANK1 | 1564,37 | -0,19 | 0,09 | -2,08 | 3,72E-02 |
| SIPA1 | 2247,79 | 0,19 | 0,09 | 2,08 | 3,73E-02 |
| CFAP70 | 210,81 | 0,14 | 0,07 | 2,08 | 3,73E-02 |
| DNAJC24 | 741,93 | -0,11 | 0,05 | -2,08 | 3,73E-02 |
| SMCHD1 | 60239,34 | -0,14 | 0,07 | -2,08 | 3,73E-02 |
| IL12RB1 | 619,31 | 0,12 | 0,06 | 2,08 | 3,74E-02 |
| ZNF879 | 282,41 | -0,14 | 0,07 | -2,08 | 3,74E-02 |
| FAHD2B | 68,96 | 0,14 | 0,07 | 2,08 | 3,74E-02 |
| KANK1 | 378,78 | 0,19 | 0,09 | 2,08 | 3,74E-02 |
| OSGIN2 | 1860,40 | -0,15 | 0,07 | -2,08 | 3,75E-02 |
| GLRX2 | 613,28 | -0,09 | 0,04 | -2,08 | 3,74E-02 |
| CMTM7 | 978,79 | 0,09 | 0,04 | 2,08 | 3,75E-02 |
| DLG5 | 255,63 | 0,25 | 0,12 | 2,08 | 3,75E-02 |
| MYOM1 | 246,34 | 0,14 | 0,07 | 2,08 | 3,76E-02 |
| CDIP1 | 245,03 | 0,15 | 0,07 | 2,08 | 3,76E-02 |
| SESN1 | 1371,55 | -0,11 | 0,05 | -2,08 | 3,77E-02 |
| PIKFYVE | 5580,64 | -0,07 | 0,04 | -2,08 | 3,77E-02 |
| SRI | 1130,48 | -0,09 | 0,05 | -2,08 | 3,77E-02 |
| SMAD2 | 8569,23 | -0,07 | 0,03 | -2,08 | 3,78E-02 |
| BACH1 | 15562,79 | -0,12 | 0,06 | -2,08 | 3,78E-02 |
| PPIE | 771,02 | -0,11 | 0,05 | -2,08 | 3,79E-02 |
| BZW2 | 549,78 | -0,08 | 0,04 | -2,08 | 3,79E-02 |
| DYNC1LI2 | 4593,79 | -0,06 | 0,03 | -2,07 | 3,80E-02 |
| PPP2R2A | 1934,61 | -0,09 | 0,04 | -2,07 | 3,81E-02 |
| ZNF527 | 314,16 | -0,10 | 0,05 | -2,07 | 3,81E-02 |
| KLRG1 | 1712,30 | -0,19 | 0,09 | -2,07 | 3,82E-02 |
| IFIT1B | 911,17 | 0,28 | 0,14 | 2,07 | 3,82E-02 |
| TEX261 | 328,47 | 0,14 | 0,07 | 2,07 | 3,82E-02 |
| FGL2 | 21965,30 | -0,17 | 0,08 | -2,07 | 3,82E-02 |
| SPOPL | 6743,97 | -0,14 | 0,07 | -2,07 | 3,82E-02 |
| EFL1 | 609,47 | -0,07 | 0,03 | -2,07 | 3,82E-02 |
| CASTOR1 | 54,93 | 0,11 | 0,05 | 2,07 | 3,83E-02 |
| ZNF383 | 792,58 | -0,06 | 0,03 | -2,07 | 3,83E-02 |
| CCDC50 | 1294,63 | -0,13 | 0,07 | -2,07 | 3,83E-02 |
| PRKAA1 | 3790,15 | -0,08 | 0,04 | -2,07 | 3,84E-02 |

| | | | | | |
|---|---|---|---|---|---|
| POLR3E | 1121,49 | 0,07 | 0,04 | 2,07 | 3,84E-02 |
| UQCRFS1 | 920,81 | -0,08 | 0,04 | -2,07 | 3,84E-02 |
| INTS7 | 725,36 | -0,08 | 0,04 | -2,07 | 3,85E-02 |
| SLC9A9 | 971,17 | -0,08 | 0,04 | -2,07 | 3,85E-02 |
| BAG2 | 268,19 | -0,13 | 0,06 | -2,07 | 3,86E-02 |
| ETHE1 | 192,93 | 0,07 | 0,04 | 2,07 | 3,86E-02 |
| SEPTIN2 | 5823,75 | -0,07 | 0,03 | -2,07 | 3,85E-02 |
| WDR13 | 1233,84 | 0,08 | 0,04 | 2,07 | 3,86E-02 |
| SCART1 | 385,65 | 0,21 | 0,10 | 2,07 | 3,85E-02 |
| GLMP | 113,40 | 0,16 | 0,08 | 2,07 | 3,86E-02 |
| NKRF | 364,06 | -0,08 | 0,04 | -2,07 | 3,86E-02 |
| GMFB | 1941,65 | -0,17 | 0,08 | -2,07 | 3,86E-02 |
| NOL7 | 798,85 | -0,08 | 0,04 | -2,07 | 3,87E-02 |
| WDR76 | 314,50 | -0,10 | 0,05 | -2,07 | 3,87E-02 |
| ZNF529 | 685,09 | -0,09 | 0,05 | -2,07 | 3,87E-02 |
| ITGB3BP | 365,39 | -0,14 | 0,07 | -2,07 | 3,87E-02 |
| MTCH1 | 2208,14 | 0,07 | 0,03 | 2,07 | 3,87E-02 |
| UNK | 544,00 | 0,10 | 0,05 | 2,07 | 3,88E-02 |
| EFCAB2 | 223,88 | 0,15 | 0,07 | 2,07 | 3,89E-02 |
| APEH | 1328,64 | 0,05 | 0,02 | 2,07 | 3,89E-02 |
| UBE2D4 | 141,53 | 0,09 | 0,05 | 2,07 | 3,89E-02 |
| OLFM4 | 135,57 | 0,46 | 0,22 | 2,07 | 3,89E-02 |
| PIAS1 | 6059,84 | -0,09 | 0,04 | -2,06 | 3,90E-02 |
| RIMOC1 | 880,03 | -0,06 | 0,03 | -2,06 | 3,90E-02 |
| TMEM184B | 530,87 | 0,20 | 0,10 | 2,06 | 3,90E-02 |
| ZNF600 | 715,02 | -0,15 | 0,07 | -2,06 | 3,90E-02 |
| IL20RB | 58,11 | 0,18 | 0,09 | 2,06 | 3,90E-02 |
| NDUFA9 | 902,23 | -0,07 | 0,03 | -2,06 | 3,90E-02 |
| CADM4 | 73,27 | 0,19 | 0,09 | 2,06 | 3,90E-02 |
| CD164 | 8862,17 | -0,11 | 0,06 | -2,06 | 3,91E-02 |
| CYB5D2 | 178,46 | 0,07 | 0,03 | 2,06 | 3,91E-02 |
| ADPRM | 456,92 | -0,15 | 0,07 | -2,06 | 3,92E-02 |
| CSTF2 | 304,76 | -0,06 | 0,03 | -2,06 | 3,92E-02 |
| GORASP1 | 722,83 | 0,08 | 0,04 | 2,06 | 3,93E-02 |
| DUSP2 | 320,05 | -0,19 | 0,09 | -2,06 | 3,93E-02 |
| ENSG00000284773 | 56,81 | 0,15 | 0,07 | 2,06 | 3,93E-02 |
| NENF | 131,65 | 0,10 | 0,05 | 2,06 | 3,93E-02 |
| PDE4B | 3893,56 | -0,10 | 0,05 | -2,06 | 3,94E-02 |
| SLC12A2 | 757,78 | -0,10 | 0,05 | -2,06 | 3,94E-02 |
| JMJD7 | 45,47 | 0,19 | 0,09 | 2,06 | 3,94E-02 |
| KRAS | 2803,85 | -0,09 | 0,04 | -2,06 | 3,94E-02 |
| OGFOD2 | 66,54 | 0,09 | 0,05 | 2,06 | 3,94E-02 |
| LTBP1 | 525,63 | 0,22 | 0,11 | 2,06 | 3,95E-02 |
| OPN3 | 265,32 | -0,08 | 0,04 | -2,06 | 3,96E-02 |
| OAZ1 | 43506,53 | 0,16 | 0,08 | 2,06 | 3,96E-02 |
| LANCL1 | 1050,27 | -0,09 | 0,04 | -2,06 | 3,96E-02 |

| | | | | | |
|---|---|---|---|---|---|
| ZKSCAN2 | 189,35 | 0,12 | 0,06 | 2,06 | 3,96E-02 |
| DNAJA4 | 1080,12 | 0,19 | 0,09 | 2,06 | 3,97E-02 |
| ZCCHC10 | 213,92 | -0,17 | 0,09 | -2,06 | 3,97E-02 |
| BLOC1S4 | 610,92 | -0,08 | 0,04 | -2,06 | 3,97E-02 |
| VCF1 | 4909,10 | 0,19 | 0,09 | 2,06 | 3,97E-02 |
| IMPDH1 | 1761,49 | 0,16 | 0,08 | 2,06 | 3,97E-02 |
| SEPHS2 | 575,97 | 0,09 | 0,05 | 2,06 | 3,97E-02 |
| MGAT2 | 1693,81 | -0,09 | 0,04 | -2,06 | 3,98E-02 |
| FKBP1B | 72,45 | 0,21 | 0,10 | 2,06 | 3,98E-02 |
| GRK6 | 3794,74 | 0,11 | 0,05 | 2,06 | 3,98E-02 |
| SMIM30 | 143,22 | -0,20 | 0,10 | -2,06 | 3,98E-02 |
| SMNDC1 | 1076,61 | -0,11 | 0,05 | -2,06 | 3,98E-02 |
| C14orf28 | 199,66 | -0,11 | 0,06 | -2,06 | 3,98E-02 |
| SLIRP | 213,49 | -0,09 | 0,05 | -2,06 | 3,98E-02 |
| CAPN3 | 156,43 | 0,17 | 0,08 | 2,05 | 3,99E-02 |
| DAPK1 | 1701,40 | 0,15 | 0,07 | 2,05 | 4,00E-02 |
| B2M | 118149,52 | -0,19 | 0,09 | -2,05 | 4,00E-02 |
| CLOCK | 971,44 | -0,08 | 0,04 | -2,05 | 4,00E-02 |
| UBL7 | 1004,60 | 0,11 | 0,05 | 2,05 | 4,00E-02 |
| KIF18A | 155,67 | 0,19 | 0,09 | 2,05 | 4,00E-02 |
| UBE2N | 1850,40 | -0,08 | 0,04 | -2,05 | 4,00E-02 |
| GBP3 | 1168,21 | -0,16 | 0,08 | -2,05 | 4,00E-02 |
| SEL1L3 | 2477,63 | -0,11 | 0,05 | -2,05 | 4,01E-02 |
| OAT | 986,01 | -0,11 | 0,06 | -2,05 | 4,01E-02 |
| WDR5B | 577,92 | -0,13 | 0,06 | -2,05 | 4,01E-02 |
| GPR135 | 141,85 | 0,14 | 0,07 | 2,05 | 4,01E-02 |
| POLM | 617,76 | 0,12 | 0,06 | 2,05 | 4,02E-02 |
| MED15 | 1895,16 | 0,15 | 0,07 | 2,05 | 4,03E-02 |
| NCOA7 | 1501,14 | -0,09 | 0,04 | -2,05 | 4,03E-02 |
| TULP3 | 87,82 | 0,12 | 0,06 | 2,05 | 4,03E-02 |
| TMEM80 | 365,94 | 0,10 | 0,05 | 2,05 | 4,03E-02 |
| ECHDC2 | 897,42 | 0,13 | 0,06 | 2,05 | 4,03E-02 |
| AP1M1 | 1260,96 | 0,10 | 0,05 | 2,05 | 4,03E-02 |
| RALA | 450,37 | -0,07 | 0,04 | -2,05 | 4,03E-02 |
| N4BP2L1 | 1329,73 | -0,06 | 0,03 | -2,05 | 4,04E-02 |
| LSM7 | 506,37 | -0,12 | 0,06 | -2,05 | 4,04E-02 |
| TMTC3 | 795,62 | -0,14 | 0,07 | -2,05 | 4,04E-02 |
| RCN3 | 195,68 | 0,30 | 0,15 | 2,05 | 4,04E-02 |
| LCORL | 1470,99 | -0,15 | 0,07 | -2,05 | 4,04E-02 |
| AFDN | 855,29 | 0,19 | 0,09 | 2,05 | 4,06E-02 |
| C14orf132 | 135,72 | 0,25 | 0,12 | 2,05 | 4,06E-02 |
| POM121 | 738,15 | 0,13 | 0,06 | 2,05 | 4,06E-02 |
| IPP | 406,22 | -0,06 | 0,03 | -2,05 | 4,06E-02 |
| AIF1 | 1544,85 | -0,19 | 0,09 | -2,05 | 4,06E-02 |
| C9orf40 | 251,82 | 0,13 | 0,07 | 2,05 | 4,06E-02 |
| NADSYN1 | 2738,29 | 0,09 | 0,05 | 2,05 | 4,06E-02 |

| | | | | | |
|---|---|---|---|---|---|
| PUS1 | 439,86 | 0,08 | 0,04 | 2,05 | 4,07E-02 |
| DIAPH2 | 1560,17 | -0,09 | 0,04 | -2,05 | 4,07E-02 |
| MAPK6 | 697,50 | -0,09 | 0,04 | -2,05 | 4,07E-02 |
| TMEM216 | 222,05 | 0,07 | 0,04 | 2,05 | 4,07E-02 |
| PCYOX1 | 662,78 | -0,06 | 0,03 | -2,05 | 4,08E-02 |
| SIN3B | 636,17 | 0,22 | 0,11 | 2,05 | 4,08E-02 |
| LPAR2 | 653,40 | 0,24 | 0,12 | 2,05 | 4,08E-02 |
| TSC22D1 | 1202,39 | 0,09 | 0,04 | 2,04 | 4,09E-02 |
| TRIM58 | 13293,98 | 0,27 | 0,13 | 2,04 | 4,09E-02 |
| PEX12 | 141,19 | -0,09 | 0,04 | -2,04 | 4,09E-02 |
| IRAK2 | 230,28 | 0,10 | 0,05 | 2,04 | 4,09E-02 |
| SELP | 377,20 | 0,17 | 0,08 | 2,04 | 4,10E-02 |
| VAMP2 | 383,22 | 0,16 | 0,08 | 2,04 | 4,10E-02 |
| PDZD8 | 3602,56 | -0,13 | 0,06 | -2,04 | 4,10E-02 |
| E2F2 | 1185,59 | 0,25 | 0,12 | 2,04 | 4,10E-02 |
| RNASEH2B | 4298,29 | -0,08 | 0,04 | -2,04 | 4,10E-02 |
| CAMP | 407,58 | 0,27 | 0,13 | 2,04 | 4,11E-02 |
| ADAM9 | 978,88 | -0,10 | 0,05 | -2,04 | 4,11E-02 |
| TAF15 | 2031,86 | -0,06 | 0,03 | -2,04 | 4,11E-02 |
| CHCHD6 | 152,91 | 0,12 | 0,06 | 2,04 | 4,11E-02 |
| ZNF599 | 159,95 | 0,10 | 0,05 | 2,04 | 4,13E-02 |
| LETM1 | 791,72 | 0,08 | 0,04 | 2,04 | 4,13E-02 |
| MAP1LC3A | 84,61 | 0,18 | 0,09 | 2,04 | 4,13E-02 |
| CEACAM1 | 553,62 | 0,20 | 0,10 | 2,04 | 4,13E-02 |
| SUCO | 2422,14 | -0,11 | 0,05 | -2,04 | 4,14E-02 |
| PPP1R35 | 328,09 | 0,10 | 0,05 | 2,04 | 4,16E-02 |
| DMXL1 | 4313,98 | -0,09 | 0,04 | -2,04 | 4,16E-02 |
| INSR | 538,74 | 0,13 | 0,06 | 2,04 | 4,16E-02 |
| AP3D1 | 1332,39 | 0,13 | 0,07 | 2,04 | 4,16E-02 |
| ATXN7L3 | 1876,77 | 0,08 | 0,04 | 2,04 | 4,17E-02 |
| BCL6-AS1 | 122,45 | -0,26 | 0,13 | -2,04 | 4,17E-02 |
| TOB2 | 336,91 | 0,13 | 0,07 | 2,04 | 4,17E-02 |
| ENOSF1 | 948,34 | 0,13 | 0,06 | 2,04 | 4,17E-02 |
| NETO2 | 152,84 | 0,13 | 0,06 | 2,04 | 4,18E-02 |
| STIP1 | 1342,82 | -0,07 | 0,04 | -2,04 | 4,18E-02 |
| IQCC | 27,47 | 0,17 | 0,08 | 2,04 | 4,18E-02 |
| OTUB1 | 608,79 | 0,06 | 0,03 | 2,04 | 4,18E-02 |
| FKBP4 | 380,10 | 0,09 | 0,04 | 2,04 | 4,18E-02 |
| PI4KA | 2168,89 | 0,14 | 0,07 | 2,03 | 4,19E-02 |
| METAP1 | 921,93 | -0,07 | 0,03 | -2,03 | 4,20E-02 |
| ZNF587B | 1014,88 | -0,06 | 0,03 | -2,03 | 4,20E-02 |
| TIMM21 | 337,20 | -0,08 | 0,04 | -2,03 | 4,20E-02 |
| TMEM192 | 504,49 | -0,05 | 0,02 | -2,03 | 4,20E-02 |
| ZNF550 | 661,17 | -0,10 | 0,05 | -2,03 | 4,20E-02 |
| GALK1 | 340,05 | 0,09 | 0,04 | 2,03 | 4,20E-02 |
| OSBPL8 | 14604,50 | -0,15 | 0,08 | -2,03 | 4,20E-02 |

| | | | | | |
|---|---|---|---|---|---|
| UNC50 | 1052,19 | -0,06 | 0,03 | -2,03 | 4,21E-02 |
| PRAF2 | 35,18 | 0,18 | 0,09 | 2,03 | 4,21E-02 |
| MBLAC1 | 65,05 | 0,11 | 0,06 | 2,03 | 4,21E-02 |
| RPL4 | 13999,89 | -0,15 | 0,07 | -2,03 | 4,22E-02 |
| STX12 | 1467,50 | -0,07 | 0,03 | -2,03 | 4,22E-02 |
| KCTD6 | 261,42 | -0,07 | 0,03 | -2,03 | 4,23E-02 |
| RICTOR | 14240,23 | -0,12 | 0,06 | -2,03 | 4,23E-02 |
| CNOT2 | 4444,82 | -0,09 | 0,04 | -2,03 | 4,23E-02 |
| HAUS8 | 187,69 | 0,07 | 0,04 | 2,03 | 4,23E-02 |
| SYDE2 | 139,07 | 0,18 | 0,09 | 2,03 | 4,23E-02 |
| CACNG6 | 47,75 | 0,30 | 0,15 | 2,03 | 4,23E-02 |
| ACAT2 | 390,58 | -0,10 | 0,05 | -2,03 | 4,23E-02 |
| CCDC82 | 1941,98 | -0,12 | 0,06 | -2,03 | 4,24E-02 |
| RSPH14 | 47,97 | 0,24 | 0,12 | 2,03 | 4,24E-02 |
| GATAD2A | 1456,70 | 0,17 | 0,08 | 2,03 | 4,24E-02 |
| ANO5 | 109,06 | 0,21 | 0,10 | 2,03 | 4,24E-02 |
| ROGDI | 417,54 | 0,15 | 0,07 | 2,03 | 4,24E-02 |
| MBLAC2 | 247,61 | -0,14 | 0,07 | -2,03 | 4,24E-02 |
| ANKRA2 | 936,59 | -0,09 | 0,05 | -2,03 | 4,25E-02 |
| LPGAT1 | 8132,55 | -0,10 | 0,05 | -2,03 | 4,25E-02 |
| ZNF672 | 752,03 | 0,07 | 0,04 | 2,03 | 4,25E-02 |
| ATAD2B | 4598,01 | -0,07 | 0,04 | -2,03 | 4,26E-02 |
| POLR3H | 411,90 | 0,12 | 0,06 | 2,03 | 4,26E-02 |
| MYLIP | 2939,59 | -0,09 | 0,05 | -2,03 | 4,27E-02 |
| ZNF486 | 802,85 | -0,12 | 0,06 | -2,03 | 4,27E-02 |
| RCC1 | 266,00 | 0,08 | 0,04 | 2,03 | 4,27E-02 |
| COQ8B | 658,01 | 0,08 | 0,04 | 2,03 | 4,28E-02 |
| CAMKK1 | 399,38 | 0,17 | 0,09 | 2,03 | 4,28E-02 |
| PCMTD1 | 6229,38 | -0,13 | 0,06 | -2,02 | 4,29E-02 |
| LRP1 | 2520,54 | 0,24 | 0,12 | 2,02 | 4,29E-02 |
| SNX13 | 5579,90 | -0,11 | 0,05 | -2,02 | 4,30E-02 |
| ERO1A | 3794,10 | -0,10 | 0,05 | -2,02 | 4,30E-02 |
| SOD1 | 1259,12 | -0,13 | 0,06 | -2,02 | 4,30E-02 |
| SREK1IP1 | 912,24 | -0,11 | 0,05 | -2,02 | 4,30E-02 |
| WDR7 | 2915,16 | -0,07 | 0,03 | -2,02 | 4,30E-02 |
| DNAH1 | 2223,84 | 0,15 | 0,08 | 2,02 | 4,30E-02 |
| LAPTM4A | 1446,62 | -0,09 | 0,05 | -2,02 | 4,30E-02 |
| MAU2 | 1661,06 | 0,16 | 0,08 | 2,02 | 4,30E-02 |
| PTBP1 | 2036,57 | 0,10 | 0,05 | 2,02 | 4,31E-02 |
| RABGAP1 | 1740,58 | -0,06 | 0,03 | -2,02 | 4,32E-02 |
| ZNF48 | 147,38 | 0,10 | 0,05 | 2,02 | 4,32E-02 |
| CD37 | 2040,39 | 0,16 | 0,08 | 2,02 | 4,33E-02 |
| SRRM2 | 6001,49 | 0,20 | 0,10 | 2,02 | 4,33E-02 |
| AGA | 528,46 | -0,10 | 0,05 | -2,02 | 4,33E-02 |
| RNF170 | 650,87 | -0,07 | 0,04 | -2,02 | 4,33E-02 |
| ANXA5 | 3227,85 | -0,11 | 0,06 | -2,02 | 4,34E-02 |

| | | | | | |
|---|---|---|---|---|---|
| RXRB | 978,94 | 0,10 | 0,05 | 2,02 | 4,34E-02 |
| TLNRD1 | 544,03 | 0,10 | 0,05 | 2,02 | 4,34E-02 |
| TUBGCP4 | 564,90 | 0,09 | 0,05 | 2,02 | 4,34E-02 |
| SAT2 | 155,78 | 0,11 | 0,05 | 2,02 | 4,35E-02 |
| ZNF43 | 777,21 | -0,08 | 0,04 | -2,02 | 4,35E-02 |
| C16orf86 | 86,69 | 0,09 | 0,04 | 2,02 | 4,35E-02 |
| AKTIP | 1586,06 | -0,09 | 0,05 | -2,02 | 4,35E-02 |
| MAP1LC3B2 | 55,33 | 0,22 | 0,11 | 2,02 | 4,36E-02 |
| BRCA1 | 528,22 | -0,10 | 0,05 | -2,02 | 4,36E-02 |
| MAX | 3704,70 | 0,07 | 0,04 | 2,02 | 4,36E-02 |
| SUN1 | 1545,90 | 0,08 | 0,04 | 2,02 | 4,37E-02 |
| ZNF765 | 537,51 | -0,17 | 0,08 | -2,02 | 4,37E-02 |
| SMARCE1 | 1124,55 | -0,08 | 0,04 | -2,02 | 4,38E-02 |
| IDE | 986,64 | -0,05 | 0,03 | -2,02 | 4,38E-02 |
| ZWINT | 72,71 | 0,18 | 0,09 | 2,02 | 4,39E-02 |
| CNR1 | 80,63 | 0,25 | 0,13 | 2,01 | 4,40E-02 |
| TP53I11 | 413,73 | 0,23 | 0,11 | 2,01 | 4,40E-02 |
| PTRH1 | 139,21 | 0,11 | 0,06 | 2,01 | 4,40E-02 |
| STAM2 | 2444,21 | -0,10 | 0,05 | -2,01 | 4,40E-02 |
| SAMD8 | 2816,43 | -0,09 | 0,05 | -2,01 | 4,41E-02 |
| CHMP1A | 690,58 | 0,11 | 0,06 | 2,01 | 4,41E-02 |
| SKI | 1068,12 | 0,19 | 0,09 | 2,01 | 4,41E-02 |
| CHMP2B | 874,32 | -0,11 | 0,06 | -2,01 | 4,42E-02 |
| PLEKHM1 | 1341,20 | 0,17 | 0,09 | 2,01 | 4,42E-02 |
| CEP63 | 3172,16 | -0,10 | 0,05 | -2,01 | 4,42E-02 |
| MRPL9 | 392,32 | -0,07 | 0,03 | -2,01 | 4,43E-02 |
| TRIM10 | 538,75 | 0,24 | 0,12 | 2,01 | 4,43E-02 |
| PIGT | 993,59 | 0,08 | 0,04 | 2,01 | 4,43E-02 |
| YTHDC2 | 2444,72 | -0,09 | 0,04 | -2,01 | 4,44E-02 |
| NXT2 | 300,62 | -0,17 | 0,09 | -2,01 | 4,44E-02 |
| GET3 | 911,52 | 0,12 | 0,06 | 2,01 | 4,45E-02 |
| EXO5 | 277,74 | -0,08 | 0,04 | -2,01 | 4,46E-02 |
| RUFY3 | 1478,48 | -0,07 | 0,03 | -2,01 | 4,47E-02 |
| CBX6 | 1131,99 | 0,12 | 0,06 | 2,01 | 4,47E-02 |
| NUBP1 | 666,72 | -0,06 | 0,03 | -2,01 | 4,47E-02 |
| EGF | 192,11 | 0,23 | 0,12 | 2,01 | 4,48E-02 |
| B3GAT3 | 340,55 | 0,09 | 0,04 | 2,01 | 4,48E-02 |
| SLC25A25 | 556,09 | 0,06 | 0,03 | 2,01 | 4,48E-02 |
| PIN1 | 895,58 | 0,05 | 0,03 | 2,01 | 4,48E-02 |
| STIL | 447,32 | 0,19 | 0,09 | 2,01 | 4,49E-02 |
| ANKRD46 | 610,41 | -0,09 | 0,05 | -2,01 | 4,49E-02 |
| CCDC88A | 2846,91 | -0,11 | 0,05 | -2,01 | 4,49E-02 |
| BRF1 | 975,04 | 0,10 | 0,05 | 2,01 | 4,49E-02 |
| NSMCE1 | 522,35 | -0,07 | 0,03 | -2,00 | 4,50E-02 |
| DIXDC1 | 115,63 | 0,22 | 0,11 | 2,00 | 4,50E-02 |
| MACF1 | 28111,26 | -0,07 | 0,04 | -2,00 | 4,50E-02 |

| | | | | | |
|---|---|---|---|---|---|
| CNTNAP2 | 102,24 | 0,38 | 0,19 | 2,00 | 4,50E-02 |
| IQCN | 268,63 | 0,23 | 0,11 | 2,00 | 4,50E-02 |
| MRPS28 | 240,67 | -0,09 | 0,05 | -2,00 | 4,50E-02 |
| FUT11 | 1110,62 | -0,08 | 0,04 | -2,00 | 4,51E-02 |
| XPO1 | 5292,66 | -0,08 | 0,04 | -2,00 | 4,51E-02 |
| PEX6 | 779,57 | 0,10 | 0,05 | 2,00 | 4,51E-02 |
| RHOG | 716,23 | 0,16 | 0,08 | 2,00 | 4,51E-02 |
| METTL6 | 467,03 | -0,05 | 0,03 | -2,00 | 4,51E-02 |
| ALDH1A2 | 199,15 | 0,25 | 0,12 | 2,00 | 4,51E-02 |
| ABHD10 | 809,97 | -0,14 | 0,07 | -2,00 | 4,51E-02 |
| UBXN11 | 683,21 | 0,18 | 0,09 | 2,00 | 4,52E-02 |
| FBXO22 | 1304,53 | -0,10 | 0,05 | -2,00 | 4,52E-02 |
| CD244 | 1678,88 | -0,09 | 0,04 | -2,00 | 4,53E-02 |
| MSH3 | 840,86 | -0,07 | 0,04 | -2,00 | 4,53E-02 |
| FAM76B | 1620,59 | -0,13 | 0,07 | -2,00 | 4,53E-02 |
| GUCY1B1 | 234,14 | 0,18 | 0,09 | 2,00 | 4,54E-02 |
| ARID2 | 2806,71 | -0,06 | 0,03 | -2,00 | 4,54E-02 |
| FLI1 | 7318,77 | -0,06 | 0,03 | -2,00 | 4,54E-02 |
| CZIB | 650,72 | -0,05 | 0,02 | -2,00 | 4,54E-02 |
| VAPA | 4699,67 | -0,09 | 0,04 | -2,00 | 4,55E-02 |
| TUSC2 | 317,31 | 0,07 | 0,03 | 2,00 | 4,55E-02 |
| PFKL | 1971,44 | 0,10 | 0,05 | 2,00 | 4,55E-02 |
| LDLRAP1 | 1290,36 | 0,14 | 0,07 | 2,00 | 4,55E-02 |
| RPA3 | 246,65 | -0,12 | 0,06 | -2,00 | 4,55E-02 |
| MME | 15895,47 | -0,18 | 0,09 | -2,00 | 4,56E-02 |
| HDGF | 5444,06 | 0,18 | 0,09 | 2,00 | 4,56E-02 |
| C12orf76 | 495,08 | 0,06 | 0,03 | 2,00 | 4,56E-02 |
| ZNF586 | 1817,80 | -0,07 | 0,04 | -2,00 | 4,56E-02 |
| PTPN6 | 2874,52 | 0,17 | 0,09 | 2,00 | 4,57E-02 |
| SRGN | 32869,03 | -0,19 | 0,10 | -2,00 | 4,57E-02 |
| GAK | 2318,94 | 0,15 | 0,08 | 2,00 | 4,57E-02 |
| LRP6 | 201,81 | 0,20 | 0,10 | 2,00 | 4,58E-02 |
| SLC35G2 | 29,88 | 0,34 | 0,17 | 2,00 | 4,58E-02 |
| HLA-DPB1 | 4683,17 | -0,11 | 0,05 | -2,00 | 4,58E-02 |
| IDH3B | 944,17 | -0,06 | 0,03 | -2,00 | 4,60E-02 |
| PRSS23 | 500,75 | -0,19 | 0,10 | -2,00 | 4,60E-02 |
| PDCL3 | 176,59 | -0,10 | 0,05 | -1,99 | 4,61E-02 |
| KLRC4-KLRK1 | 256,20 | -0,24 | 0,12 | -1,99 | 4,61E-02 |
| ANKAR | 637,00 | -0,08 | 0,04 | -1,99 | 4,61E-02 |
| PDCD6IP | 4834,96 | -0,07 | 0,03 | -1,99 | 4,62E-02 |
| EIF1AX | 1038,85 | -0,14 | 0,07 | -1,99 | 4,62E-02 |
| NPLOC4 | 2431,85 | 0,09 | 0,05 | 1,99 | 4,62E-02 |
| SOCS5 | 740,88 | -0,09 | 0,04 | -1,99 | 4,62E-02 |
| MRPL24 | 339,63 | -0,08 | 0,04 | -1,99 | 4,62E-02 |
| KDM2B | 805,06 | 0,12 | 0,06 | 1,99 | 4,62E-02 |
| PHF2 | 917,69 | 0,13 | 0,07 | 1,99 | 4,63E-02 |

| | | | | | |
|---|---|---|---|---|---|
| TOMM5 | 201,71 | -0,09 | 0,05 | -1,99 | 4,63E-02 |
| CSNK1G2 | 2617,17 | 0,10 | 0,05 | 1,99 | 4,63E-02 |
| TESC | 2035,45 | 0,20 | 0,10 | 1,99 | 4,64E-02 |
| VWA5A | 243,28 | 0,09 | 0,04 | 1,99 | 4,64E-02 |
| PAFAH1B3 | 122,37 | 0,09 | 0,04 | 1,99 | 4,64E-02 |
| PDE7A | 6996,20 | -0,09 | 0,04 | -1,99 | 4,65E-02 |
| MYG1 | 413,52 | 0,09 | 0,04 | 1,99 | 4,65E-02 |
| PTP4A1 | 74,63 | 0,12 | 0,06 | 1,99 | 4,66E-02 |
| MBD3 | 697,15 | 0,07 | 0,03 | 1,99 | 4,66E-02 |
| DPCD | 178,74 | 0,21 | 0,11 | 1,99 | 4,67E-02 |
| MPI | 404,24 | 0,09 | 0,05 | 1,99 | 4,67E-02 |
| POMC | 64,14 | 0,22 | 0,11 | 1,99 | 4,66E-02 |
| LARP4 | 1086,05 | -0,09 | 0,04 | -1,99 | 4,66E-02 |
| TPK1 | 1152,29 | -0,08 | 0,04 | -1,99 | 4,66E-02 |
| TMEM106A | 361,04 | 0,18 | 0,09 | 1,99 | 4,67E-02 |
| PRDM4 | 965,68 | -0,04 | 0,02 | -1,99 | 4,67E-02 |
| CXorf38 | 1996,59 | -0,06 | 0,03 | -1,99 | 4,67E-02 |
| DIABLO | 325,39 | 0,06 | 0,03 | 1,99 | 4,68E-02 |
| ATPAF1 | 693,44 | -0,05 | 0,03 | -1,99 | 4,68E-02 |
| SORCS3 | 145,26 | 0,27 | 0,14 | 1,99 | 4,68E-02 |
| NT5C3B | 179,40 | 0,15 | 0,08 | 1,99 | 4,68E-02 |
| ARRB1 | 1561,37 | 0,15 | 0,07 | 1,99 | 4,69E-02 |
| MRPS18A | 316,08 | -0,06 | 0,03 | -1,99 | 4,69E-02 |
| ZNF641 | 3792,42 | -0,11 | 0,06 | -1,99 | 4,69E-02 |
| CTH | 53,34 | 0,16 | 0,08 | 1,99 | 4,70E-02 |
| ACADS | 313,16 | 0,08 | 0,04 | 1,99 | 4,70E-02 |
| MRM1 | 162,41 | 0,09 | 0,05 | 1,99 | 4,70E-02 |
| POLR3K | 196,38 | 0,07 | 0,03 | 1,99 | 4,70E-02 |
| RAB11FIP2 | 1667,42 | -0,10 | 0,05 | -1,99 | 4,70E-02 |
| ZNF852 | 603,84 | -0,06 | 0,03 | -1,99 | 4,71E-02 |
| SLC35A5 | 1440,25 | -0,08 | 0,04 | -1,99 | 4,71E-02 |
| ARMCX4 | 311,28 | 0,12 | 0,06 | 1,98 | 4,72E-02 |
| ARMC7 | 224,11 | 0,12 | 0,06 | 1,98 | 4,72E-02 |
| STON2 | 556,05 | 0,16 | 0,08 | 1,98 | 4,72E-02 |
| DNM3 | 467,47 | 0,14 | 0,07 | 1,98 | 4,72E-02 |
| GZF1 | 627,81 | -0,07 | 0,04 | -1,98 | 4,72E-02 |
| PF4V1 | 108,99 | 0,34 | 0,17 | 1,98 | 4,73E-02 |
| ATF7 | 464,10 | 0,18 | 0,09 | 1,98 | 4,73E-02 |
| ORC4 | 1676,57 | -0,10 | 0,05 | -1,98 | 4,73E-02 |
| IL15RA | 269,78 | 0,10 | 0,05 | 1,98 | 4,73E-02 |
| RNF227 | 112,89 | 0,07 | 0,04 | 1,98 | 4,73E-02 |
| ZKSCAN7 | 220,06 | -0,09 | 0,04 | -1,98 | 4,74E-02 |
| FDX1 | 487,51 | -0,09 | 0,05 | -1,98 | 4,75E-02 |
| POLA2 | 113,70 | 0,13 | 0,07 | 1,98 | 4,76E-02 |
| FNIP1 | 5413,79 | -0,10 | 0,05 | -1,98 | 4,76E-02 |
| RAD23B | 3894,95 | -0,08 | 0,04 | -1,98 | 4,77E-02 |

| | | | | | |
|---|---|---|---|---|---|
| SSTR3 | 170,38 | 0,17 | 0,09 | 1,98 | 4,78E-02 |
| HNRNPAB | 1255,33 | -0,06 | 0,03 | -1,98 | 4,78E-02 |
| FKBP11 | 294,11 | 0,16 | 0,08 | 1,98 | 4,79E-02 |
| XPO7 | 3220,93 | 0,12 | 0,06 | 1,98 | 4,79E-02 |
| CIP2A | 267,01 | -0,13 | 0,07 | -1,98 | 4,79E-02 |
| NRAS | 1569,39 | -0,09 | 0,05 | -1,98 | 4,79E-02 |
| TMED3 | 894,28 | 0,08 | 0,04 | 1,98 | 4,80E-02 |
| LDAH | 535,39 | -0,08 | 0,04 | -1,98 | 4,80E-02 |
| BPNT2 | 1815,94 | -0,07 | 0,04 | -1,98 | 4,81E-02 |
| STMN3 | 657,87 | 0,10 | 0,05 | 1,98 | 4,81E-02 |
| RBM15 | 2476,08 | -0,06 | 0,03 | -1,98 | 4,81E-02 |
| RPS8 | 9478,96 | -0,16 | 0,08 | -1,98 | 4,82E-02 |
| MRPL27 | 415,79 | -0,07 | 0,04 | -1,98 | 4,82E-02 |
| CNTNAP3B | 45,89 | 0,39 | 0,20 | 1,98 | 4,82E-02 |
| CTNNAL1 | 220,22 | 0,24 | 0,12 | 1,98 | 4,83E-02 |
| TRIM33 | 5460,72 | -0,07 | 0,04 | -1,97 | 4,83E-02 |
| KHSRP | 646,77 | 0,14 | 0,07 | 1,97 | 4,83E-02 |
| HBEGF | 40,19 | 0,22 | 0,11 | 1,97 | 4,84E-02 |
| CXXC1 | 873,56 | 0,06 | 0,03 | 1,97 | 4,85E-02 |
| TRIM11 | 944,23 | 0,11 | 0,06 | 1,97 | 4,85E-02 |
| MRPL2 | 120,75 | 0,09 | 0,05 | 1,97 | 4,86E-02 |
| H3C10 | 2441,64 | 0,17 | 0,09 | 1,97 | 4,86E-02 |
| SIRPD | 173,59 | 0,15 | 0,08 | 1,97 | 4,86E-02 |
| DVL1 | 537,42 | 0,07 | 0,03 | 1,97 | 4,87E-02 |
| RAB11FIP5 | 146,62 | 0,21 | 0,11 | 1,97 | 4,87E-02 |
| REL | 2909,18 | -0,06 | 0,03 | -1,97 | 4,88E-02 |
| ADGRE1 | 1334,88 | 0,17 | 0,09 | 1,97 | 4,88E-02 |
| PHYKPL | 3236,89 | 0,11 | 0,06 | 1,97 | 4,88E-02 |
| PCNX1 | 15917,49 | -0,09 | 0,05 | -1,97 | 4,89E-02 |
| XYLB | 116,44 | 0,17 | 0,09 | 1,97 | 4,90E-02 |
| ZBTB24 | 1248,40 | -0,07 | 0,03 | -1,97 | 4,90E-02 |
| LMO4 | 941,16 | -0,06 | 0,03 | -1,97 | 4,91E-02 |
| HIKESHI | 518,10 | -0,08 | 0,04 | -1,97 | 4,91E-02 |
| PDCD1LG2 | 42,30 | 0,29 | 0,15 | 1,97 | 4,92E-02 |
| CYRIB | 9169,28 | -0,09 | 0,05 | -1,97 | 4,92E-02 |
| RBM5 | 9811,84 | -0,05 | 0,03 | -1,97 | 4,92E-02 |
| SOX4 | 240,80 | 0,12 | 0,06 | 1,97 | 4,92E-02 |
| DPH7 | 540,84 | 0,06 | 0,03 | 1,97 | 4,92E-02 |
| SFR1 | 153,35 | -0,13 | 0,07 | -1,97 | 4,93E-02 |
| KIF21B | 2697,66 | 0,16 | 0,08 | 1,97 | 4,93E-02 |
| CCDC97 | 375,56 | 0,16 | 0,08 | 1,97 | 4,93E-02 |
| TDRD3 | 835,52 | -0,09 | 0,04 | -1,97 | 4,93E-02 |
| UBE3C | 3197,39 | -0,05 | 0,02 | -1,97 | 4,94E-02 |
| RALGAPB | 5303,30 | -0,06 | 0,03 | -1,97 | 4,94E-02 |
| ANPEP | 1304,59 | 0,26 | 0,13 | 1,97 | 4,94E-02 |
| SEMA4B | 1187,96 | 0,16 | 0,08 | 1,97 | 4,94E-02 |

| | | | | | |
|---|---|---|---|---|---|
| UBB | 57939,39 | 0,24 | 0,12 | 1,96 | 4,94E-02 |
| DENND1A | 1390,00 | 0,12 | 0,06 | 1,96 | 4,95E-02 |
| MCEE | 313,01 | -0,07 | 0,04 | -1,96 | 4,96E-02 |
| LSM5 | 272,10 | -0,13 | 0,07 | -1,96 | 4,96E-02 |
| ZMYND19 | 210,30 | 0,07 | 0,04 | 1,96 | 4,97E-02 |
| HERPUD1 | 2034,42 | -0,06 | 0,03 | -1,96 | 4,97E-02 |
| BMX | 265,49 | -0,22 | 0,11 | -1,96 | 4,97E-02 |
| FLNA | 19308,27 | 0,17 | 0,09 | 1,96 | 4,97E-02 |
| ASPHD2 | 131,14 | 0,13 | 0,06 | 1,96 | 4,98E-02 |
| KMT2D | 4594,36 | 0,20 | 0,10 | 1,96 | 4,98E-02 |
| COPS2 | 1252,49 | -0,16 | 0,08 | -1,96 | 4,98E-02 |
| KPNA4 | 4373,79 | -0,07 | 0,04 | -1,96 | 4,98E-02 |
| L3MBTL2 | 473,50 | 0,08 | 0,04 | 1,96 | 4,99E-02 |
| MPP1 | 10006,87 | 0,20 | 0,10 | 1,96 | 4,99E-02 |
| NTN4 | 50,37 | 0,30 | 0,15 | 1,96 | 4,99E-02 |
| AGAP2 | 1008,17 | 0,15 | 0,08 | 1,96 | 4,99E-02 |
| SUMF2 | 1606,98 | -0,07 | 0,04 | -1,96 | 5,00E-02 |
| KARS1 | 2142,05 | -0,08 | 0,04 | -1,96 | 5,01E-02 |
| PTPRN2 | 315,42 | 0,17 | 0,09 | 1,96 | 5,02E-02 |
| TPCN2 | 891,48 | 0,17 | 0,09 | 1,96 | 5,02E-02 |
| TBC1D10A | 487,82 | 0,12 | 0,06 | 1,96 | 5,02E-02 |
| NOD2 | 1788,06 | 0,18 | 0,09 | 1,96 | 5,02E-02 |
| KIF5C | 379,85 | 0,14 | 0,07 | 1,96 | 5,03E-02 |
| ZNF30 | 111,66 | -0,12 | 0,06 | -1,96 | 5,03E-02 |
| PEX3 | 407,38 | -0,12 | 0,06 | -1,96 | 5,03E-02 |
| SPIN1 | 758,31 | -0,05 | 0,03 | -1,96 | 5,03E-02 |
| SEC16A | 1532,66 | 0,18 | 0,09 | 1,96 | 5,04E-02 |
| TCF7 | 2651,28 | 0,17 | 0,09 | 1,96 | 5,04E-02 |
| GPS2 | 133,10 | 0,11 | 0,06 | 1,96 | 5,04E-02 |
| PSMA7 | 2052,27 | -0,08 | 0,04 | -1,96 | 5,05E-02 |
| ANAPC7 | 948,09 | -0,04 | 0,02 | -1,96 | 5,05E-02 |
| MAT2B | 5291,44 | -0,09 | 0,05 | -1,95 | 5,06E-02 |
| PMEPA1 | 94,31 | 0,19 | 0,10 | 1,95 | 5,07E-02 |
| YY1 | 3527,42 | -0,04 | 0,02 | -1,95 | 5,08E-02 |
| BCAP29 | 610,97 | -0,09 | 0,04 | -1,95 | 5,08E-02 |
| PDCD5 | 418,14 | -0,08 | 0,04 | -1,95 | 5,08E-02 |
| PRKCD | 1356,39 | 0,20 | 0,10 | 1,95 | 5,08E-02 |
| DRAM2 | 1742,76 | -0,10 | 0,05 | -1,95 | 5,08E-02 |
| KPNA1 | 2859,07 | -0,07 | 0,03 | -1,95 | 5,08E-02 |
| PLXND1 | 607,99 | 0,22 | 0,11 | 1,95 | 5,09E-02 |
| IRF8 | 2175,16 | -0,09 | 0,05 | -1,95 | 5,09E-02 |
| ADAM8 | 6752,05 | 0,14 | 0,07 | 1,95 | 5,09E-02 |
| SH3GLB1 | 5059,44 | -0,10 | 0,05 | -1,95 | 5,09E-02 |
| UBE2J2 | 1356,57 | 0,06 | 0,03 | 1,95 | 5,09E-02 |
| FBXO21 | 1155,62 | -0,06 | 0,03 | -1,95 | 5,09E-02 |
| ZYG11B | 2441,75 | -0,08 | 0,04 | -1,95 | 5,09E-02 |

| | | | | | |
|---|---|---|---|---|---|
| TRRAP | 3511,28 | 0,12 | 0,06 | 1,95 | 5,09E-02 |
| RADX | 226,88 | -0,13 | 0,06 | -1,95 | 5,10E-02 |
| YARS2 | 542,07 | -0,07 | 0,04 | -1,95 | 5,10E-02 |
| LNPEP | 12208,97 | -0,08 | 0,04 | -1,95 | 5,10E-02 |
| MAP3K5 | 6652,54 | -0,09 | 0,05 | -1,95 | 5,10E-02 |
| LRRC25 | 1483,40 | 0,17 | 0,09 | 1,95 | 5,11E-02 |
| NAV3 | 114,63 | 0,29 | 0,15 | 1,95 | 5,11E-02 |
| CD300C | 283,00 | 0,13 | 0,06 | 1,95 | 5,11E-02 |
| RNF31 | 745,97 | 0,10 | 0,05 | 1,95 | 5,11E-02 |
| ATXN7L1 | 818,65 | 0,08 | 0,04 | 1,95 | 5,12E-02 |
| PRKCH | 6663,00 | -0,09 | 0,04 | -1,95 | 5,12E-02 |
| PURA | 2868,95 | -0,11 | 0,06 | -1,95 | 5,12E-02 |
| BATF3 | 71,54 | 0,14 | 0,07 | 1,95 | 5,12E-02 |
| IFFO1 | 700,99 | 0,13 | 0,06 | 1,95 | 5,13E-02 |
| CREG1 | 2496,02 | 0,16 | 0,08 | 1,95 | 5,13E-02 |
| LIN52 | 296,82 | -0,06 | 0,03 | -1,95 | 5,13E-02 |
| XRCC1 | 228,12 | 0,12 | 0,06 | 1,95 | 5,14E-02 |
| RTCA | 1055,37 | -0,05 | 0,03 | -1,95 | 5,14E-02 |
| MED22 | 1299,28 | 0,07 | 0,04 | 1,95 | 5,14E-02 |
| EXOC3 | 1280,33 | 0,13 | 0,07 | 1,95 | 5,14E-02 |
| ZFP82 | 253,06 | -0,13 | 0,07 | -1,95 | 5,14E-02 |
| SHFL | 555,24 | 0,12 | 0,06 | 1,95 | 5,14E-02 |
| STAU2 | 1497,89 | -0,07 | 0,04 | -1,95 | 5,15E-02 |
| ZNF687 | 702,54 | 0,14 | 0,07 | 1,95 | 5,15E-02 |
| CASP2 | 1641,86 | 0,09 | 0,05 | 1,95 | 5,15E-02 |
| RAB2B | 4034,60 | 0,18 | 0,09 | 1,95 | 5,16E-02 |
| WDFY1 | 2685,11 | -0,10 | 0,05 | -1,95 | 5,17E-02 |
| SNRNP70 | 1818,98 | 0,15 | 0,08 | 1,94 | 5,18E-02 |
| SLC25A26 | 642,89 | -0,07 | 0,04 | -1,94 | 5,19E-02 |
| UIMC1 | 2841,85 | -0,08 | 0,04 | -1,94 | 5,19E-02 |
| ZNF681 | 237,16 | -0,16 | 0,08 | -1,94 | 5,19E-02 |
| MANBAL | 361,66 | 0,07 | 0,04 | 1,94 | 5,20E-02 |
| INTS2 | 955,27 | -0,07 | 0,03 | -1,94 | 5,20E-02 |
| EPHB4 | 252,34 | 0,21 | 0,11 | 1,94 | 5,20E-02 |
| EXPH5 | 145,35 | 0,16 | 0,08 | 1,94 | 5,20E-02 |
| CCDC92 | 549,04 | 0,08 | 0,04 | 1,94 | 5,21E-02 |
| LGALS9C | 45,42 | 0,32 | 0,16 | 1,94 | 5,21E-02 |
| IRAK1 | 778,12 | 0,13 | 0,07 | 1,94 | 5,21E-02 |
| HIPK3 | 11446,39 | -0,10 | 0,05 | -1,94 | 5,21E-02 |
| ETFA | 1695,24 | -0,08 | 0,04 | -1,94 | 5,22E-02 |
| ZNF230 | 813,92 | -0,12 | 0,06 | -1,94 | 5,22E-02 |
| MEIS1 | 212,24 | 0,18 | 0,09 | 1,94 | 5,23E-02 |
| RAD1 | 641,89 | -0,06 | 0,03 | -1,94 | 5,23E-02 |
| OGDH | 1526,07 | 0,11 | 0,06 | 1,94 | 5,23E-02 |
| WASHC3 | 884,17 | -0,08 | 0,04 | -1,94 | 5,23E-02 |
| SRSF11 | 6105,55 | -0,09 | 0,05 | -1,94 | 5,23E-02 |

| | | | | | |
|---|---|---|---|---|---|
| ZNF493 | 2392,71 | -0,09 | 0,05 | -1,94 | 5,24E-02 |
| VSIG4 | 54,69 | -0,21 | 0,11 | -1,94 | 5,24E-02 |
| TBC1D12 | 294,53 | 0,12 | 0,06 | 1,94 | 5,24E-02 |
| ZNF461 | 307,58 | -0,09 | 0,05 | -1,94 | 5,25E-02 |
| MRS2 | 614,13 | -0,07 | 0,04 | -1,94 | 5,25E-02 |
| ZBTB48 | 694,60 | 0,08 | 0,04 | 1,94 | 5,25E-02 |
| H2AC25 | 326,59 | 0,17 | 0,09 | 1,94 | 5,25E-02 |
| TASOR2 | 3605,34 | -0,06 | 0,03 | -1,94 | 5,25E-02 |
| DENND11 | 3150,02 | -0,08 | 0,04 | -1,94 | 5,26E-02 |
| USP31 | 424,77 | 0,08 | 0,04 | 1,94 | 5,26E-02 |
| FEM1A | 1989,12 | 0,08 | 0,04 | 1,94 | 5,26E-02 |
| CDK2 | 212,40 | 0,06 | 0,03 | 1,94 | 5,26E-02 |
| RNF220 | 1205,60 | 0,05 | 0,03 | 1,94 | 5,27E-02 |
| RASGRP2 | 5539,77 | 0,07 | 0,04 | 1,94 | 5,27E-02 |
| GPALPP1 | 877,68 | -0,06 | 0,03 | -1,94 | 5,28E-02 |
| POLR1G | 46,18 | 0,14 | 0,07 | 1,94 | 5,28E-02 |
| ENSG00000188897 | 2504,40 | 0,26 | 0,13 | 1,94 | 5,28E-02 |
| SLC4A7 | 3174,05 | -0,11 | 0,06 | -1,94 | 5,28E-02 |
| NSD3 | 7678,77 | -0,06 | 0,03 | -1,94 | 5,29E-02 |
| VEZF1 | 2359,29 | -0,07 | 0,03 | -1,94 | 5,29E-02 |
| RTN1 | 550,73 | 0,13 | 0,06 | 1,94 | 5,29E-02 |
| CTTNBP2NL | 123,20 | 0,13 | 0,07 | 1,93 | 5,30E-02 |
| TICAM2 | 133,21 | 0,10 | 0,05 | 1,93 | 5,31E-02 |
| EFHD2 | 3854,83 | 0,14 | 0,07 | 1,93 | 5,31E-02 |
| DOP1B | 928,59 | 0,13 | 0,07 | 1,93 | 5,32E-02 |
| TRMT10A | 155,49 | -0,14 | 0,07 | -1,93 | 5,32E-02 |
| UBAP2 | 940,13 | 0,10 | 0,05 | 1,93 | 5,33E-02 |
| ZC3H12D | 826,15 | 0,12 | 0,06 | 1,93 | 5,33E-02 |
| PRKRA | 617,53 | -0,07 | 0,03 | -1,93 | 5,33E-02 |
| MAPK3 | 546,52 | 0,11 | 0,06 | 1,93 | 5,33E-02 |
| COQ9 | 565,67 | -0,04 | 0,02 | -1,93 | 5,34E-02 |
| PRPF19 | 1334,51 | -0,06 | 0,03 | -1,93 | 5,34E-02 |
| IFT27 | 358,98 | 0,08 | 0,04 | 1,93 | 5,36E-02 |
| SARS2 | 56,94 | 0,12 | 0,06 | 1,93 | 5,36E-02 |
| KIF27 | 1271,82 | -0,14 | 0,07 | -1,93 | 5,36E-02 |
| ZNF385A | 209,45 | 0,21 | 0,11 | 1,93 | 5,36E-02 |
| TMX3 | 2540,37 | -0,15 | 0,08 | -1,93 | 5,36E-02 |
| HYOU1 | 584,00 | 0,12 | 0,06 | 1,93 | 5,37E-02 |
| TMEM218 | 668,27 | -0,05 | 0,03 | -1,93 | 5,36E-02 |
| TEX10 | 900,81 | -0,07 | 0,04 | -1,93 | 5,36E-02 |
| FGD3 | 3953,35 | 0,18 | 0,09 | 1,93 | 5,38E-02 |
| PLB1 | 909,42 | 0,22 | 0,11 | 1,93 | 5,38E-02 |
| USB1 | 986,61 | 0,13 | 0,07 | 1,93 | 5,38E-02 |
| HEBP2 | 4044,61 | -0,09 | 0,04 | -1,93 | 5,38E-02 |
| AP1G1 | 4512,27 | -0,06 | 0,03 | -1,93 | 5,38E-02 |
| TNFRSF13B | 80,92 | 0,25 | 0,13 | 1,93 | 5,39E-02 |

| | | | | | |
|---|---|---|---|---|---|
| ACAP1 | 2677,36 | 0,12 | 0,06 | 1,93 | 5,39E-02 |
| ATAD3B | 689,74 | 0,10 | 0,05 | 1,93 | 5,39E-02 |
| TATDN2 | 929,77 | 0,07 | 0,04 | 1,93 | 5,39E-02 |
| ZC3H18 | 682,73 | 0,13 | 0,07 | 1,93 | 5,40E-02 |
| PRKG2 | 71,25 | 0,26 | 0,13 | 1,93 | 5,40E-02 |
| NCKAP1 | 350,02 | 0,13 | 0,07 | 1,93 | 5,40E-02 |
| ZNF552 | 803,48 | -0,08 | 0,04 | -1,93 | 5,41E-02 |
| GTF3C6 | 325,66 | -0,10 | 0,05 | -1,93 | 5,41E-02 |
| TDG | 984,73 | -0,07 | 0,03 | -1,93 | 5,41E-02 |
| PGRMC2 | 500,98 | -0,06 | 0,03 | -1,93 | 5,42E-02 |
| MARCHF3 | 291,12 | 0,12 | 0,06 | 1,92 | 5,43E-02 |
| CCR5 | 693,17 | -0,17 | 0,09 | -1,92 | 5,43E-02 |
| CCDC91 | 1523,74 | -0,09 | 0,05 | -1,92 | 5,43E-02 |
| ORC6 | 56,06 | 0,16 | 0,08 | 1,92 | 5,43E-02 |
| ATP5PF | 200,08 | -0,13 | 0,07 | -1,92 | 5,43E-02 |
| LASP1NB | 178,31 | 0,10 | 0,05 | 1,92 | 5,44E-02 |
| CDCA7L | 631,67 | -0,13 | 0,07 | -1,92 | 5,44E-02 |
| GYPE | 72,45 | 0,19 | 0,10 | 1,92 | 5,44E-02 |
| NT5M | 224,45 | 0,22 | 0,12 | 1,92 | 5,44E-02 |
| RBBP4 | 2894,89 | -0,05 | 0,03 | -1,92 | 5,45E-02 |
| RPS3A | 5408,90 | -0,31 | 0,16 | -1,92 | 5,45E-02 |
| GZMH | 1263,19 | -0,26 | 0,14 | -1,92 | 5,45E-02 |
| SANBR | 353,11 | 0,06 | 0,03 | 1,92 | 5,45E-02 |
| TMEM14A | 128,48 | -0,13 | 0,07 | -1,92 | 5,45E-02 |
| C5orf22 | 719,87 | -0,06 | 0,03 | -1,92 | 5,46E-02 |
| DGKD | 2632,98 | 0,14 | 0,07 | 1,92 | 5,46E-02 |
| TBC1D2B | 3126,61 | 0,07 | 0,04 | 1,92 | 5,46E-02 |
| ZNF610 | 36,79 | 0,20 | 0,10 | 1,92 | 5,48E-02 |
| TAOK1 | 8494,87 | -0,07 | 0,03 | -1,92 | 5,48E-02 |
| MAP3K11 | 2866,60 | 0,12 | 0,06 | 1,92 | 5,48E-02 |
| ENTR1 | 510,10 | 0,06 | 0,03 | 1,92 | 5,48E-02 |
| CCNT2 | 4307,13 | -0,11 | 0,06 | -1,92 | 5,49E-02 |
| HEMK1 | 1201,43 | 0,09 | 0,05 | 1,92 | 5,49E-02 |
| MRPS27 | 803,40 | -0,06 | 0,03 | -1,92 | 5,50E-02 |
| PRKCQ | 1513,88 | -0,08 | 0,04 | -1,92 | 5,51E-02 |
| COX17 | 413,60 | -0,08 | 0,04 | -1,92 | 5,51E-02 |
| SREK1 | 4285,19 | -0,10 | 0,05 | -1,92 | 5,52E-02 |
| DAGLB | 609,29 | 0,10 | 0,05 | 1,92 | 5,53E-02 |
| ACADSB | 603,36 | -0,07 | 0,04 | -1,92 | 5,53E-02 |
| EFHC1 | 430,89 | 0,11 | 0,06 | 1,92 | 5,53E-02 |
| BCO2 | 78,63 | 0,17 | 0,09 | 1,92 | 5,54E-02 |
| SIRT7 | 1655,43 | 0,07 | 0,04 | 1,92 | 5,55E-02 |
| NOL4L | 1346,16 | 0,12 | 0,06 | 1,92 | 5,55E-02 |
| RWDD1 | 352,51 | -0,11 | 0,06 | -1,91 | 5,56E-02 |
| NSUN5 | 513,96 | 0,06 | 0,03 | 1,91 | 5,56E-02 |
| ZFAND1 | 598,79 | -0,14 | 0,07 | -1,91 | 5,56E-02 |

| | | | | | |
|---|---|---|---|---|---|
| THUMPD2 | 381,50 | -0,09 | 0,05 | -1,91 | 5,56E-02 |
| STRADB | 18046,13 | 0,23 | 0,12 | 1,91 | 5,57E-02 |
| MYCBP2 | 14479,72 | -0,07 | 0,04 | -1,91 | 5,56E-02 |
| WSB2 | 996,58 | 0,09 | 0,04 | 1,91 | 5,57E-02 |
| ZNF449 | 338,74 | -0,09 | 0,05 | -1,91 | 5,57E-02 |
| FANCC | 301,58 | 0,15 | 0,08 | 1,91 | 5,57E-02 |
| STK32C | 351,66 | 0,09 | 0,05 | 1,91 | 5,58E-02 |
| SMIM3 | 335,01 | 0,11 | 0,06 | 1,91 | 5,59E-02 |
| BRWD3 | 4658,37 | -0,08 | 0,04 | -1,91 | 5,59E-02 |
| FTCDNL1 | 107,82 | 0,11 | 0,06 | 1,91 | 5,60E-02 |
| SLFN11 | 1994,54 | -0,07 | 0,04 | -1,91 | 5,60E-02 |
| CAMK1 | 557,74 | 0,13 | 0,07 | 1,91 | 5,60E-02 |
| LILRA6 | 772,76 | 0,21 | 0,11 | 1,91 | 5,61E-02 |
| NAAA | 2192,93 | 0,11 | 0,06 | 1,91 | 5,61E-02 |
| CSNK1D | 2427,29 | 0,15 | 0,08 | 1,91 | 5,61E-02 |
| PRIMPOL | 445,53 | -0,07 | 0,04 | -1,91 | 5,62E-02 |
| ITPR1 | 6488,68 | -0,08 | 0,04 | -1,91 | 5,62E-02 |
| PRR3 | 358,16 | -0,05 | 0,03 | -1,91 | 5,62E-02 |
| VDAC1 | 793,45 | -0,05 | 0,03 | -1,91 | 5,62E-02 |
| COL5A3 | 120,83 | 0,40 | 0,21 | 1,91 | 5,62E-02 |
| KIAA1586 | 575,79 | -0,18 | 0,09 | -1,91 | 5,62E-02 |
| ZC3H12B | 64,29 | 0,16 | 0,08 | 1,91 | 5,63E-02 |
| HSPA5 | 2569,44 | -0,07 | 0,04 | -1,91 | 5,63E-02 |
| PTGR3 | 2323,40 | -0,06 | 0,03 | -1,91 | 5,64E-02 |
| DDX6 | 18438,77 | -0,07 | 0,04 | -1,91 | 5,64E-02 |
| PHOSPHO1 | 3198,78 | 0,18 | 0,09 | 1,91 | 5,64E-02 |
| RASSF6 | 43,86 | 0,26 | 0,13 | 1,91 | 5,65E-02 |
| ITGAE | 390,18 | 0,06 | 0,03 | 1,91 | 5,65E-02 |
| SKIL | 1753,84 | -0,12 | 0,07 | -1,91 | 5,65E-02 |
| STT3A | 1406,16 | -0,04 | 0,02 | -1,91 | 5,65E-02 |
| TPM3 | 13234,48 | -0,08 | 0,04 | -1,91 | 5,65E-02 |
| PPIH | 357,70 | -0,06 | 0,03 | -1,91 | 5,65E-02 |
| SEC11C | 490,63 | -0,08 | 0,04 | -1,91 | 5,66E-02 |
| PXN | 2614,81 | 0,23 | 0,12 | 1,91 | 5,66E-02 |
| NIPAL1 | 57,54 | 0,26 | 0,14 | 1,91 | 5,67E-02 |
| MRPL15 | 197,09 | -0,11 | 0,06 | -1,91 | 5,67E-02 |
| PON2 | 245,48 | -0,09 | 0,05 | -1,91 | 5,67E-02 |
| DDX39B | 1591,94 | 0,17 | 0,09 | 1,91 | 5,68E-02 |
| CCDC14 | 1843,14 | -0,10 | 0,05 | -1,91 | 5,68E-02 |
| SORT1 | 1326,45 | 0,14 | 0,07 | 1,90 | 5,68E-02 |
| RPGR | 1483,68 | -0,11 | 0,06 | -1,90 | 5,68E-02 |
| HOPX | 594,29 | -0,16 | 0,08 | -1,90 | 5,69E-02 |
| AARS1 | 756,84 | 0,09 | 0,05 | 1,90 | 5,69E-02 |
| MIDN | 1374,55 | 0,17 | 0,09 | 1,90 | 5,69E-02 |
| FRA10AC1 | 969,81 | -0,11 | 0,06 | -1,90 | 5,69E-02 |
| PCLAF | 26,75 | 0,21 | 0,11 | 1,90 | 5,70E-02 |

| | | | | | |
|---|---|---|---|---|---|
| TGIF2 | 379,48 | 0,09 | 0,05 | 1,90 | 5,71E-02 |
| URB1 | 977,04 | 0,09 | 0,05 | 1,90 | 5,71E-02 |
| GHRL | 156,96 | 0,15 | 0,08 | 1,90 | 5,71E-02 |
| ZNF548 | 851,70 | -0,07 | 0,04 | -1,90 | 5,72E-02 |
| TGDS | 478,06 | -0,10 | 0,06 | -1,90 | 5,72E-02 |
| ESM1 | 30,48 | 0,29 | 0,15 | 1,90 | 5,72E-02 |
| TRIM32 | 426,21 | -0,07 | 0,04 | -1,90 | 5,73E-02 |
| ZNF254 | 626,90 | -0,13 | 0,07 | -1,90 | 5,74E-02 |
| S100A13 | 34,10 | 0,13 | 0,07 | 1,90 | 5,74E-02 |
| VCPIP1 | 7467,46 | -0,11 | 0,06 | -1,90 | 5,74E-02 |
| EPHA1 | 193,49 | 0,16 | 0,08 | 1,90 | 5,74E-02 |
| AGAP9 | 132,28 | 0,22 | 0,12 | 1,90 | 5,75E-02 |
| SLA | 14953,13 | -0,09 | 0,05 | -1,90 | 5,75E-02 |
| ZNF266 | 2020,29 | -0,08 | 0,04 | -1,90 | 5,75E-02 |
| PTER | 592,26 | -0,09 | 0,05 | -1,90 | 5,75E-02 |
| RBFA | 196,56 | 0,08 | 0,04 | 1,90 | 5,75E-02 |
| GID4 | 549,02 | 0,12 | 0,06 | 1,90 | 5,76E-02 |
| C6orf89 | 2468,46 | -0,06 | 0,03 | -1,90 | 5,76E-02 |
| RAP2B | 4150,62 | -0,08 | 0,04 | -1,90 | 5,77E-02 |
| TFG | 1475,76 | -0,08 | 0,04 | -1,90 | 5,77E-02 |
| SCYL1 | 866,09 | 0,09 | 0,05 | 1,90 | 5,77E-02 |
| NUTF2 | 654,72 | 0,07 | 0,04 | 1,90 | 5,78E-02 |
| NUP155 | 1249,80 | -0,05 | 0,02 | -1,90 | 5,79E-02 |
| GGA1 | 667,25 | 0,13 | 0,07 | 1,90 | 5,80E-02 |
| IPMK | 3233,49 | -0,15 | 0,08 | -1,90 | 5,80E-02 |
| SIGLEC6 | 47,39 | 0,22 | 0,12 | 1,90 | 5,80E-02 |
| APRT | 488,52 | 0,08 | 0,04 | 1,90 | 5,80E-02 |
| GRN | 1774,93 | 0,16 | 0,09 | 1,89 | 5,81E-02 |
| POLR3D | 415,67 | 0,08 | 0,04 | 1,89 | 5,81E-02 |
| SPG7 | 3745,33 | 0,09 | 0,05 | 1,89 | 5,81E-02 |
| CCDC117 | 1130,99 | -0,05 | 0,03 | -1,89 | 5,82E-02 |
| CLIP4 | 2161,93 | -0,08 | 0,04 | -1,89 | 5,82E-02 |
| TMEM181 | 2032,73 | -0,07 | 0,04 | -1,89 | 5,82E-02 |
| SAE1 | 820,21 | -0,06 | 0,03 | -1,89 | 5,82E-02 |
| ZNF451 | 4794,28 | -0,08 | 0,04 | -1,89 | 5,82E-02 |
| TRAP1 | 592,65 | 0,09 | 0,05 | 1,89 | 5,83E-02 |
| PRKD2 | 1492,59 | 0,16 | 0,08 | 1,89 | 5,83E-02 |
| PPP1CC | 3815,02 | -0,08 | 0,04 | -1,89 | 5,83E-02 |
| ZSWIM3 | 96,52 | 0,08 | 0,04 | 1,89 | 5,83E-02 |
| PPA1 | 583,30 | -0,15 | 0,08 | -1,89 | 5,84E-02 |
| EXTL2 | 148,49 | -0,10 | 0,05 | -1,89 | 5,85E-02 |
| WASHC1 | 151,19 | 0,21 | 0,11 | 1,89 | 5,85E-02 |
| QNG1 | 354,24 | -0,07 | 0,03 | -1,89 | 5,85E-02 |
| ATP6V1G1 | 1815,55 | -0,14 | 0,07 | -1,89 | 5,85E-02 |
| EEF1AKMT2 | 350,26 | -0,10 | 0,05 | -1,89 | 5,85E-02 |
| XPO5 | 654,69 | 0,08 | 0,04 | 1,89 | 5,85E-02 |

| | | | | | |
|---|---|---|---|---|---|
| NMRAL1 | 602,83 | 0,08 | 0,04 | 1,89 | 5,86E-02 |
| GTF3C2 | 809,72 | 0,10 | 0,05 | 1,89 | 5,87E-02 |
| FBXL5 | 16289,89 | -0,13 | 0,07 | -1,89 | 5,87E-02 |
| MRPL13 | 267,42 | -0,14 | 0,07 | -1,89 | 5,87E-02 |
| KCTD21 | 270,90 | 0,11 | 0,06 | 1,89 | 5,88E-02 |
| DNAJB1 | 1431,51 | -0,06 | 0,03 | -1,89 | 5,88E-02 |
| SCN9A | 209,48 | 0,18 | 0,10 | 1,89 | 5,89E-02 |
| ENSG00000285476 | 53,31 | 0,28 | 0,15 | 1,89 | 5,89E-02 |
| H6PD | 1053,60 | 0,15 | 0,08 | 1,89 | 5,90E-02 |
| ZCRB1 | 207,22 | -0,10 | 0,05 | -1,89 | 5,89E-02 |
| APIP | 395,56 | -0,11 | 0,06 | -1,89 | 5,90E-02 |
| PLD5P1 | 167,13 | -0,12 | 0,06 | -1,89 | 5,90E-02 |
| PGLYRP1 | 107,66 | 0,21 | 0,11 | 1,89 | 5,90E-02 |
| EIF2AK1 | 3856,95 | 0,16 | 0,09 | 1,89 | 5,90E-02 |
| GON7 | 172,82 | -0,14 | 0,07 | -1,89 | 5,91E-02 |
| CREB3L4 | 104,32 | 0,08 | 0,04 | 1,89 | 5,93E-02 |
| DIPK1A | 281,09 | -0,10 | 0,06 | -1,89 | 5,93E-02 |
| RMI1 | 370,60 | -0,10 | 0,05 | -1,89 | 5,93E-02 |
| TMEM263 | 641,37 | -0,11 | 0,06 | -1,89 | 5,93E-02 |
| NBPF9 | 359,31 | 0,11 | 0,06 | 1,89 | 5,94E-02 |
| STRIP1 | 1101,72 | 0,08 | 0,04 | 1,88 | 5,94E-02 |
| ADAM28 | 1624,34 | -0,14 | 0,08 | -1,88 | 5,95E-02 |
| MTX3 | 909,56 | -0,09 | 0,05 | -1,88 | 5,95E-02 |
| AKAP10 | 2754,53 | -0,06 | 0,03 | -1,88 | 5,95E-02 |
| LMBR1L | 1450,61 | 0,06 | 0,03 | 1,88 | 5,96E-02 |
| AAR2 | 245,16 | 0,07 | 0,03 | 1,88 | 5,96E-02 |
| TFDP1 | 1644,43 | 0,14 | 0,08 | 1,88 | 5,97E-02 |
| ZNF573 | 396,96 | -0,07 | 0,04 | -1,88 | 5,97E-02 |
| UBR5 | 7692,48 | -0,06 | 0,03 | -1,88 | 5,97E-02 |
| TFEB | 1038,87 | 0,13 | 0,07 | 1,88 | 5,97E-02 |
| RABL2A | 229,84 | 0,13 | 0,07 | 1,88 | 5,97E-02 |
| ARAP3 | 622,57 | 0,24 | 0,13 | 1,88 | 5,97E-02 |
| DPH3 | 496,63 | -0,12 | 0,06 | -1,88 | 5,98E-02 |
| FPGT | 603,23 | -0,16 | 0,08 | -1,88 | 5,99E-02 |
| KCNK6 | 605,45 | 0,08 | 0,04 | 1,88 | 5,99E-02 |
| GMCL1 | 1905,22 | -0,08 | 0,04 | -1,88 | 6,00E-02 |
| PHF5A | 259,35 | -0,11 | 0,06 | -1,88 | 6,00E-02 |
| BTBD8 | 314,95 | -0,10 | 0,06 | -1,88 | 6,00E-02 |
| IL18BP | 240,27 | 0,15 | 0,08 | 1,88 | 6,01E-02 |
| LACTB2 | 273,69 | -0,09 | 0,05 | -1,88 | 6,02E-02 |
| DICER1 | 11600,82 | -0,09 | 0,05 | -1,88 | 6,04E-02 |
| TTC21A | 200,01 | 0,20 | 0,10 | 1,88 | 6,04E-02 |
| MMAA | 344,30 | -0,08 | 0,04 | -1,88 | 6,04E-02 |
| GGA3 | 867,79 | 0,13 | 0,07 | 1,88 | 6,04E-02 |
| CCDC149 | 324,84 | 0,15 | 0,08 | 1,88 | 6,04E-02 |
| EXOC7 | 2268,59 | 0,08 | 0,04 | 1,88 | 6,05E-02 |

| | | | | | |
|---|---|---|---|---|---|
| VPS13A | 3900,05 | -0,10 | 0,05 | -1,88 | 6,05E-02 |
| ZBED6 | 45,95 | 0,14 | 0,07 | 1,88 | 6,05E-02 |
| NDUFA6 | 405,58 | -0,15 | 0,08 | -1,88 | 6,06E-02 |
| ITPR2 | 7888,43 | -0,08 | 0,04 | -1,88 | 6,06E-02 |
| CPLANE1 | 1011,83 | -0,08 | 0,04 | -1,88 | 6,07E-02 |
| CD151 | 502,49 | 0,16 | 0,09 | 1,88 | 6,07E-02 |
| MDM1 | 939,91 | -0,09 | 0,05 | -1,87 | 6,09E-02 |
| TRUB1 | 521,92 | -0,12 | 0,06 | -1,87 | 6,09E-02 |
| ADSL | 386,72 | 0,06 | 0,03 | 1,87 | 6,09E-02 |
| BBS9 | 867,44 | -0,08 | 0,04 | -1,87 | 6,10E-02 |
| NOD1 | 746,37 | 0,10 | 0,05 | 1,87 | 6,10E-02 |
| NEIL2 | 311,53 | 0,10 | 0,05 | 1,87 | 6,10E-02 |
| SLC35A1 | 201,07 | -0,16 | 0,09 | -1,87 | 6,10E-02 |
| TRIM44 | 2616,13 | -0,05 | 0,03 | -1,87 | 6,11E-02 |
| BBS10 | 625,07 | -0,12 | 0,07 | -1,87 | 6,11E-02 |
| RGL2 | 1893,69 | 0,13 | 0,07 | 1,87 | 6,12E-02 |
| ABCA7 | 2339,95 | 0,20 | 0,10 | 1,87 | 6,12E-02 |
| FSIP1 | 55,95 | 0,20 | 0,11 | 1,87 | 6,13E-02 |
| BEND7 | 103,80 | 0,25 | 0,14 | 1,87 | 6,13E-02 |
| TPGS1 | 564,39 | 0,06 | 0,03 | 1,87 | 6,14E-02 |
| FAM91A1 | 5062,14 | -0,09 | 0,05 | -1,87 | 6,14E-02 |
| HYLS1 | 158,10 | -0,09 | 0,05 | -1,87 | 6,14E-02 |
| KLRK1 | 447,98 | -0,15 | 0,08 | -1,87 | 6,15E-02 |
| RPL26L1 | 67,02 | -0,20 | 0,10 | -1,87 | 6,16E-02 |
| LEPROTL1 | 3088,78 | -0,08 | 0,04 | -1,87 | 6,16E-02 |
| CD47 | 9644,58 | -0,07 | 0,04 | -1,87 | 6,16E-02 |
| CENPO | 168,08 | 0,11 | 0,06 | 1,87 | 6,16E-02 |
| SLC50A1 | 375,84 | 0,07 | 0,04 | 1,87 | 6,16E-02 |
| PRKN | 176,61 | 0,13 | 0,07 | 1,87 | 6,16E-02 |
| ARIH2 | 3046,25 | -0,04 | 0,02 | -1,87 | 6,17E-02 |
| HELZ2 | 2506,76 | 0,21 | 0,11 | 1,87 | 6,17E-02 |
| CDC73 | 3741,23 | -0,08 | 0,04 | -1,87 | 6,17E-02 |
| CCT6B | 80,04 | 0,13 | 0,07 | 1,87 | 6,17E-02 |
| PKD2 | 706,68 | -0,06 | 0,03 | -1,87 | 6,17E-02 |
| TMEM115 | 325,83 | 0,07 | 0,04 | 1,87 | 6,18E-02 |
| PTPN13 | 358,32 | 0,14 | 0,08 | 1,87 | 6,18E-02 |
| RNF125 | 1730,49 | -0,06 | 0,03 | -1,87 | 6,18E-02 |
| RFXANK | 663,53 | 0,07 | 0,04 | 1,87 | 6,18E-02 |
| LRRC37A3 | 313,97 | 0,18 | 0,10 | 1,87 | 6,18E-02 |
| TAF5L | 653,30 | -0,04 | 0,02 | -1,87 | 6,19E-02 |
| CNOT11 | 621,21 | -0,04 | 0,02 | -1,87 | 6,19E-02 |
| WAS | 880,82 | 0,19 | 0,10 | 1,87 | 6,19E-02 |
| TEFM | 420,51 | -0,08 | 0,04 | -1,87 | 6,20E-02 |
| CTDNEP1 | 599,17 | 0,13 | 0,07 | 1,87 | 6,20E-02 |
| ADORA2A | 167,07 | 0,09 | 0,05 | 1,87 | 6,20E-02 |
| MRPL16 | 415,23 | -0,06 | 0,03 | -1,87 | 6,20E-02 |

| | | | | | |
|---|---|---|---|---|---|
| TAOK2 | 1078,83 | 0,09 | 0,05 | 1,87 | 6,20E-02 |
| TARDBP | 1553,87 | -0,06 | 0,03 | -1,87 | 6,21E-02 |
| TMEM250 | 1190,67 | 0,06 | 0,03 | 1,87 | 6,21E-02 |
| CLEC1B | 115,90 | 0,21 | 0,11 | 1,86 | 6,22E-02 |
| GLRX3 | 907,60 | -0,05 | 0,03 | -1,86 | 6,22E-02 |
| SLC35E2B | 1034,04 | 0,16 | 0,09 | 1,86 | 6,22E-02 |
| TFB1M | 605,62 | -0,08 | 0,04 | -1,86 | 6,23E-02 |
| UCK1 | 406,93 | 0,06 | 0,03 | 1,86 | 6,23E-02 |
| JRK | 1422,36 | 0,08 | 0,04 | 1,86 | 6,23E-02 |
| STARD4 | 620,67 | -0,14 | 0,07 | -1,86 | 6,24E-02 |
| RELT | 3691,22 | 0,12 | 0,07 | 1,86 | 6,24E-02 |
| CCDC15 | 234,04 | -0,13 | 0,07 | -1,86 | 6,24E-02 |
| ANKRD10 | 5744,59 | -0,09 | 0,05 | -1,86 | 6,25E-02 |
| RNASEK | 129,78 | 0,11 | 0,06 | 1,86 | 6,26E-02 |
| RNF166 | 1494,49 | 0,17 | 0,09 | 1,86 | 6,26E-02 |
| ZNF329 | 364,19 | -0,08 | 0,05 | -1,86 | 6,27E-02 |
| IDH1 | 1244,36 | -0,08 | 0,05 | -1,86 | 6,27E-02 |
| SMIM35 | 47,32 | 0,24 | 0,13 | 1,86 | 6,26E-02 |
| MYO6 | 357,21 | -0,18 | 0,10 | -1,86 | 6,27E-02 |
| GRIK4 | 93,19 | 0,47 | 0,25 | 1,86 | 6,27E-02 |
| C10orf88 | 325,26 | -0,09 | 0,05 | -1,86 | 6,27E-02 |
| CD1E | 34,17 | 0,26 | 0,14 | 1,86 | 6,27E-02 |
| ZNF821 | 159,16 | 0,08 | 0,04 | 1,86 | 6,27E-02 |
| ZNF639 | 735,92 | -0,08 | 0,04 | -1,86 | 6,27E-02 |
| EIF2B3 | 352,13 | -0,05 | 0,03 | -1,86 | 6,30E-02 |
| HIVEP1 | 2136,16 | -0,07 | 0,04 | -1,86 | 6,30E-02 |
| SLC35F6 | 379,70 | 0,13 | 0,07 | 1,86 | 6,30E-02 |
| FOXJ2 | 571,20 | 0,13 | 0,07 | 1,86 | 6,31E-02 |
| MRPL10 | 230,14 | 0,08 | 0,04 | 1,86 | 6,31E-02 |
| DAZAP2 | 9481,44 | -0,08 | 0,04 | -1,86 | 6,31E-02 |
| RAB5B | 1484,99 | 0,15 | 0,08 | 1,86 | 6,31E-02 |
| C2orf69 | 406,03 | -0,10 | 0,06 | -1,86 | 6,31E-02 |
| RGS1 | 60,02 | -0,24 | 0,13 | -1,86 | 6,31E-02 |
| MARCHF1 | 3789,41 | -0,12 | 0,07 | -1,86 | 6,32E-02 |
| ZNF83 | 1030,54 | -0,11 | 0,06 | -1,86 | 6,32E-02 |
| ECHS1 | 500,39 | -0,07 | 0,04 | -1,86 | 6,33E-02 |
| EFNB1 | 123,54 | 0,10 | 0,06 | 1,86 | 6,33E-02 |
| CFL1 | 2612,74 | 0,11 | 0,06 | 1,86 | 6,33E-02 |
| ZBED5 | 1778,36 | -0,12 | 0,07 | -1,86 | 6,34E-02 |
| SEPHS1 | 627,79 | -0,05 | 0,03 | -1,86 | 6,34E-02 |
| TBC1D19 | 456,74 | -0,12 | 0,06 | -1,86 | 6,34E-02 |
| ZC3H12C | 206,40 | 0,13 | 0,07 | 1,86 | 6,35E-02 |
| RTN4 | 2410,45 | -0,08 | 0,04 | -1,86 | 6,35E-02 |
| TDP1 | 911,51 | -0,06 | 0,03 | -1,85 | 6,36E-02 |
| TNS3 | 771,22 | 0,15 | 0,08 | 1,85 | 6,36E-02 |
| PLAAT4 | 807,68 | -0,11 | 0,06 | -1,85 | 6,36E-02 |

| | | | | | |
|---|---|---|---|---|---|
| TMEM220 | 73,62 | 0,16 | 0,09 | 1,85 | 6,36E-02 |
| TAF1 | 3578,59 | -0,06 | 0,03 | -1,85 | 6,36E-02 |
| ANKRD18A | 128,49 | 0,16 | 0,08 | 1,85 | 6,38E-02 |
| NBPF20 | 100,61 | 0,14 | 0,07 | 1,85 | 6,37E-02 |
| MSH4 | 94,45 | 0,11 | 0,06 | 1,85 | 6,38E-02 |
| ELP3 | 1200,73 | -0,07 | 0,04 | -1,85 | 6,38E-02 |
| ANAPC16 | 3447,78 | -0,07 | 0,04 | -1,85 | 6,38E-02 |
| ITGB1BP1 | 1071,41 | -0,05 | 0,03 | -1,85 | 6,39E-02 |
| ACTR10 | 1205,30 | -0,08 | 0,04 | -1,85 | 6,39E-02 |
| FLYWCH1 | 583,89 | 0,13 | 0,07 | 1,85 | 6,40E-02 |
| KLHL23 | 41,46 | 0,18 | 0,10 | 1,85 | 6,41E-02 |
| RBM26 | 3950,37 | -0,08 | 0,04 | -1,85 | 6,41E-02 |
| GUSB | 505,59 | 0,08 | 0,04 | 1,85 | 6,41E-02 |
| HCAR3 | 811,47 | 0,16 | 0,09 | 1,85 | 6,42E-02 |
| GNAS | 5763,44 | 0,12 | 0,07 | 1,85 | 6,42E-02 |
| STX17 | 1383,70 | -0,07 | 0,04 | -1,85 | 6,43E-02 |
| CLEC4E | 2897,14 | -0,16 | 0,09 | -1,85 | 6,43E-02 |
| LRRC45 | 312,72 | 0,08 | 0,04 | 1,85 | 6,43E-02 |
| USP40 | 583,62 | 0,08 | 0,04 | 1,85 | 6,43E-02 |
| AGAP4 | 324,41 | 0,09 | 0,05 | 1,85 | 6,43E-02 |
| SYNJ1 | 2105,40 | -0,08 | 0,04 | -1,85 | 6,44E-02 |
| MLLT6 | 2768,78 | 0,15 | 0,08 | 1,85 | 6,44E-02 |
| MINDY2 | 1751,46 | -0,09 | 0,05 | -1,85 | 6,44E-02 |
| ABHD14A | 51,00 | 0,10 | 0,06 | 1,85 | 6,45E-02 |
| FAF1 | 1141,19 | -0,05 | 0,03 | -1,85 | 6,45E-02 |
| UBR2 | 14891,47 | -0,09 | 0,05 | -1,85 | 6,46E-02 |
| PUS10 | 507,49 | -0,08 | 0,04 | -1,85 | 6,46E-02 |
| ZNF571 | 209,84 | -0,12 | 0,06 | -1,85 | 6,46E-02 |
| SMPD2 | 222,58 | 0,08 | 0,05 | 1,85 | 6,46E-02 |
| SMIM14 | 1852,14 | -0,07 | 0,04 | -1,85 | 6,47E-02 |
| VAMP4 | 903,71 | -0,12 | 0,06 | -1,85 | 6,47E-02 |
| FAM200B | 992,52 | -0,13 | 0,07 | -1,85 | 6,47E-02 |
| TM9SF4 | 821,99 | 0,12 | 0,06 | 1,85 | 6,47E-02 |
| METTL15 | 743,36 | -0,09 | 0,05 | -1,85 | 6,49E-02 |
| PLCG1 | 2786,17 | 0,09 | 0,05 | 1,85 | 6,49E-02 |
| ARHGAP30 | 14737,32 | -0,07 | 0,04 | -1,85 | 6,49E-02 |
| EEF1E1 | 114,34 | -0,13 | 0,07 | -1,85 | 6,50E-02 |
| PRELID2 | 88,59 | 0,15 | 0,08 | 1,84 | 6,51E-02 |
| TSG101 | 1559,36 | -0,06 | 0,03 | -1,84 | 6,52E-02 |
| ELOVL7 | 440,87 | 0,18 | 0,10 | 1,84 | 6,52E-02 |
| NUP58 | 3930,53 | -0,08 | 0,04 | -1,84 | 6,53E-02 |
| ENSG00000221995 | 185,03 | 0,16 | 0,09 | 1,84 | 6,54E-02 |
| ZNF595 | 360,05 | -0,15 | 0,08 | -1,84 | 6,55E-02 |
| BRAF | 4221,09 | -0,07 | 0,04 | -1,84 | 6,55E-02 |
| SPICE1-CFAP44 | 24,96 | 0,14 | 0,08 | 1,84 | 6,55E-02 |
| BUD31 | 821,71 | -0,07 | 0,04 | -1,84 | 6,56E-02 |

| | | | | | |
|---|---|---|---|---|---|
| DDHD1 | 3639,96 | -0,08 | 0,04 | -1,84 | 6,56E-02 |
| MBNL3 | 17502,15 | 0,20 | 0,11 | 1,84 | 6,57E-02 |
| STX8 | 476,30 | -0,06 | 0,03 | -1,84 | 6,57E-02 |
| CORO1B | 966,29 | 0,06 | 0,03 | 1,84 | 6,57E-02 |
| RERE | 2729,14 | 0,12 | 0,07 | 1,84 | 6,58E-02 |
| XPNPEP3 | 506,40 | 0,08 | 0,04 | 1,84 | 6,58E-02 |
| LTB4R2 | 326,27 | 0,09 | 0,05 | 1,84 | 6,58E-02 |
| OST4 | 3027,25 | 0,12 | 0,07 | 1,84 | 6,58E-02 |
| ENSG00000239920 | 70,63 | 0,18 | 0,10 | 1,84 | 6,58E-02 |
| TERF2 | 569,02 | 0,06 | 0,03 | 1,84 | 6,59E-02 |
| SUDS3 | 2888,11 | -0,06 | 0,03 | -1,84 | 6,59E-02 |
| MRNIP | 488,39 | 0,13 | 0,07 | 1,84 | 6,59E-02 |
| SERF2 | 21256,94 | 0,16 | 0,09 | 1,84 | 6,59E-02 |
| NADK2 | 508,28 | -0,08 | 0,05 | -1,84 | 6,60E-02 |
| PSMG2 | 772,34 | -0,06 | 0,03 | -1,84 | 6,60E-02 |
| NR6A1 | 273,71 | 0,15 | 0,08 | 1,84 | 6,60E-02 |
| TSHZ3 | 340,47 | 0,14 | 0,08 | 1,84 | 6,61E-02 |
| TBCEL-TECTA | 27,28 | 0,22 | 0,12 | 1,84 | 6,61E-02 |
| EIF4EBP3 | 25,24 | 0,14 | 0,08 | 1,84 | 6,61E-02 |
| UTS2 | 40,66 | 0,33 | 0,18 | 1,84 | 6,63E-02 |
| ZNF813 | 302,41 | -0,13 | 0,07 | -1,84 | 6,64E-02 |
| UBL3 | 2209,77 | -0,08 | 0,04 | -1,84 | 6,64E-02 |
| ANGEL2 | 2780,42 | -0,08 | 0,05 | -1,84 | 6,64E-02 |
| THSD7A | 58,44 | 0,34 | 0,19 | 1,84 | 6,64E-02 |
| TMEM187 | 109,00 | 0,08 | 0,04 | 1,84 | 6,64E-02 |
| IMPA2 | 1114,50 | 0,11 | 0,06 | 1,84 | 6,65E-02 |
| EME1 | 26,32 | 0,20 | 0,11 | 1,84 | 6,65E-02 |
| ZNF117 | 3255,69 | -0,18 | 0,10 | -1,84 | 6,65E-02 |
| TRAF1 | 1483,04 | 0,11 | 0,06 | 1,84 | 6,65E-02 |
| RPL19 | 10201,93 | -0,10 | 0,06 | -1,83 | 6,66E-02 |
| PRRC1 | 1208,42 | -0,06 | 0,04 | -1,83 | 6,66E-02 |
| MAIP1 | 119,02 | -0,09 | 0,05 | -1,83 | 6,67E-02 |
| ZNF354C | 488,81 | -0,14 | 0,08 | -1,83 | 6,67E-02 |
| RUBCN | 1162,61 | 0,15 | 0,08 | 1,83 | 6,68E-02 |
| TMEM30A | 4996,47 | -0,10 | 0,06 | -1,83 | 6,68E-02 |
| HTT | 3096,61 | 0,10 | 0,06 | 1,83 | 6,69E-02 |
| RSBN1 | 2156,63 | -0,07 | 0,04 | -1,83 | 6,69E-02 |
| SFXN4 | 198,35 | 0,08 | 0,04 | 1,83 | 6,69E-02 |
| SERINC3 | 4358,20 | -0,06 | 0,03 | -1,83 | 6,69E-02 |
| ATP1B3 | 498,42 | -0,06 | 0,03 | -1,83 | 6,70E-02 |
| TMEM161B | 1757,15 | -0,10 | 0,05 | -1,83 | 6,70E-02 |
| IFT80 | 444,33 | -0,09 | 0,05 | -1,83 | 6,70E-02 |
| KBTBD7 | 1404,47 | -0,13 | 0,07 | -1,83 | 6,71E-02 |
| MICAL3 | 912,04 | 0,12 | 0,07 | 1,83 | 6,71E-02 |
| TSR1 | 773,86 | -0,05 | 0,03 | -1,83 | 6,72E-02 |
| CHCHD4 | 175,48 | -0,07 | 0,04 | -1,83 | 6,73E-02 |

| | | | | | |
|---|---|---|---|---|---|
| CAPZB | 1208,02 | 0,13 | 0,07 | 1,83 | 6,73E-02 |
| PRDX3 | 1587,81 | -0,08 | 0,04 | -1,83 | 6,73E-02 |
| VRK2 | 955,41 | -0,11 | 0,06 | -1,83 | 6,73E-02 |
| ZNF790 | 240,97 | -0,11 | 0,06 | -1,83 | 6,74E-02 |
| APEX2 | 178,98 | 0,09 | 0,05 | 1,83 | 6,74E-02 |
| RBM45 | 658,20 | -0,09 | 0,05 | -1,83 | 6,74E-02 |
| HEBP1 | 194,39 | 0,17 | 0,09 | 1,83 | 6,75E-02 |
| TMEM60 | 332,27 | -0,10 | 0,05 | -1,83 | 6,75E-02 |
| CDIPT | 1561,96 | 0,10 | 0,05 | 1,83 | 6,76E-02 |
| CDK17 | 3682,19 | -0,08 | 0,04 | -1,83 | 6,77E-02 |
| CHMP1B | 3847,40 | -0,10 | 0,05 | -1,83 | 6,77E-02 |
| EMD | 206,13 | 0,10 | 0,05 | 1,83 | 6,77E-02 |
| RBM33 | 7411,70 | -0,06 | 0,04 | -1,83 | 6,77E-02 |
| CMIP | 1399,05 | 0,19 | 0,10 | 1,83 | 6,77E-02 |
| DCUN1D5 | 272,30 | -0,10 | 0,06 | -1,83 | 6,78E-02 |
| MRPS25 | 1639,10 | -0,06 | 0,03 | -1,83 | 6,78E-02 |
| KREMEN1 | 682,36 | 0,22 | 0,12 | 1,83 | 6,78E-02 |
| RUVBL1 | 685,57 | -0,07 | 0,04 | -1,83 | 6,78E-02 |
| HACD3 | 621,83 | -0,07 | 0,04 | -1,83 | 6,80E-02 |
| PCMT1 | 1477,25 | -0,10 | 0,06 | -1,82 | 6,80E-02 |
| MCM8 | 490,92 | -0,06 | 0,04 | -1,82 | 6,80E-02 |
| CHST12 | 997,79 | 0,18 | 0,10 | 1,82 | 6,82E-02 |
| ENTPD4 | 4216,48 | -0,05 | 0,03 | -1,82 | 6,84E-02 |
| POMT1 | 511,57 | 0,08 | 0,04 | 1,82 | 6,84E-02 |
| ICA1L | 135,14 | 0,14 | 0,08 | 1,82 | 6,85E-02 |
| CAMK2D | 2098,06 | -0,09 | 0,05 | -1,82 | 6,85E-02 |
| NOA1 | 485,61 | -0,04 | 0,02 | -1,82 | 6,85E-02 |
| TTPAL | 873,79 | 0,06 | 0,03 | 1,82 | 6,85E-02 |
| NOLC1 | 581,90 | -0,06 | 0,03 | -1,82 | 6,85E-02 |
| ENSG00000259529 | 102,80 | 0,21 | 0,12 | 1,82 | 6,86E-02 |
| BAHD1 | 540,85 | 0,09 | 0,05 | 1,82 | 6,86E-02 |
| SLC25A46 | 2030,16 | -0,11 | 0,06 | -1,82 | 6,87E-02 |
| AKAP1 | 541,14 | 0,07 | 0,04 | 1,82 | 6,89E-02 |
| HLA-DRB1 | 4844,10 | -0,12 | 0,07 | -1,82 | 6,89E-02 |
| PLIN4 | 358,68 | 0,21 | 0,12 | 1,82 | 6,90E-02 |
| NFE2L2 | 2454,41 | -0,08 | 0,04 | -1,82 | 6,90E-02 |
| ARK2C | 167,38 | 0,25 | 0,14 | 1,82 | 6,91E-02 |
| LRRCC1 | 127,74 | -0,14 | 0,08 | -1,82 | 6,91E-02 |
| KDSR | 515,50 | 0,06 | 0,03 | 1,82 | 6,91E-02 |
| GLTP | 1023,84 | -0,06 | 0,03 | -1,82 | 6,92E-02 |
| SNX20 | 1621,38 | 0,11 | 0,06 | 1,82 | 6,93E-02 |
| BPGM | 3617,24 | 0,23 | 0,13 | 1,82 | 6,94E-02 |
| TCAIM | 693,09 | -0,06 | 0,03 | -1,82 | 6,94E-02 |
| SLC25A3 | 3158,63 | -0,04 | 0,02 | -1,82 | 6,94E-02 |
| YJU2 | 177,25 | 0,08 | 0,04 | 1,81 | 6,95E-02 |
| SLC9A7 | 886,72 | -0,12 | 0,06 | -1,81 | 6,95E-02 |

| | | | | | |
|---|---|---|---|---|---|
| LILRB3 | 2880,22 | 0,17 | 0,09 | 1,81 | 6,95E-02 |
| FAM193B | 831,52 | 0,18 | 0,10 | 1,81 | 6,96E-02 |
| PIGV | 490,39 | -0,05 | 0,03 | -1,81 | 6,97E-02 |
| PGM5 | 218,88 | 0,30 | 0,16 | 1,81 | 6,97E-02 |
| MOB2 | 1405,19 | 0,06 | 0,03 | 1,81 | 6,98E-02 |
| CES2 | 497,89 | 0,07 | 0,04 | 1,81 | 6,98E-02 |
| EFHC2 | 211,19 | -0,11 | 0,06 | -1,81 | 6,98E-02 |
| MARCHF8 | 11473,47 | 0,19 | 0,10 | 1,81 | 6,98E-02 |
| FBXO7 | 19249,32 | 0,18 | 0,10 | 1,81 | 6,99E-02 |
| BABAM1 | 706,97 | 0,13 | 0,07 | 1,81 | 6,99E-02 |
| MAST3 | 2584,96 | 0,18 | 0,10 | 1,81 | 7,00E-02 |
| EEF1AKMT3 | 48,23 | 0,15 | 0,08 | 1,81 | 7,00E-02 |
| EFCAB7 | 236,14 | -0,12 | 0,07 | -1,81 | 7,00E-02 |
| LENG1 | 516,63 | -0,05 | 0,03 | -1,81 | 7,00E-02 |
| WDR91 | 810,31 | 0,08 | 0,04 | 1,81 | 7,00E-02 |
| IKBKE | 1286,49 | 0,07 | 0,04 | 1,81 | 7,01E-02 |
| NFE2 | 5709,43 | 0,12 | 0,07 | 1,81 | 7,02E-02 |
| MRPL44 | 791,10 | -0,08 | 0,05 | -1,81 | 7,02E-02 |
| CCNL1 | 9738,25 | -0,09 | 0,05 | -1,81 | 7,03E-02 |
| GLS | 7534,12 | -0,11 | 0,06 | -1,81 | 7,04E-02 |
| FBL | 957,16 | -0,10 | 0,06 | -1,81 | 7,04E-02 |
| RNF130 | 6371,50 | -0,09 | 0,05 | -1,81 | 7,05E-02 |
| SERP1 | 3939,72 | -0,10 | 0,06 | -1,81 | 7,05E-02 |
| SLC36A1 | 1451,11 | 0,12 | 0,07 | 1,81 | 7,05E-02 |
| RBMS1 | 4570,79 | -0,06 | 0,04 | -1,81 | 7,06E-02 |
| ADGRA3 | 137,79 | 0,19 | 0,11 | 1,81 | 7,07E-02 |
| UNKL | 1353,06 | 0,13 | 0,07 | 1,81 | 7,07E-02 |
| ARRDC5 | 180,15 | 0,14 | 0,08 | 1,81 | 7,09E-02 |
| ELOVL5 | 6906,74 | -0,08 | 0,04 | -1,81 | 7,09E-02 |
| PRAM1 | 558,64 | 0,21 | 0,12 | 1,81 | 7,10E-02 |
| HELZ | 6882,65 | -0,06 | 0,03 | -1,81 | 7,10E-02 |
| DOCK10 | 7807,64 | -0,08 | 0,04 | -1,80 | 7,11E-02 |
| ZNF66 | 634,26 | -0,14 | 0,08 | -1,80 | 7,11E-02 |
| GCOM1 | 89,60 | 0,27 | 0,15 | 1,80 | 7,12E-02 |
| NGLY1 | 2740,59 | -0,07 | 0,04 | -1,80 | 7,12E-02 |
| NIPSNAP2 | 806,07 | -0,05 | 0,03 | -1,80 | 7,13E-02 |
| ZNF597 | 408,03 | -0,08 | 0,05 | -1,80 | 7,13E-02 |
| OGG1 | 586,87 | 0,10 | 0,06 | 1,80 | 7,14E-02 |
| LLGL2 | 499,32 | 0,14 | 0,08 | 1,80 | 7,14E-02 |
| DNAJC19 | 715,76 | -0,10 | 0,06 | -1,80 | 7,15E-02 |
| MTMR10 | 5011,73 | -0,08 | 0,05 | -1,80 | 7,16E-02 |
| TYK2 | 5457,84 | 0,09 | 0,05 | 1,80 | 7,16E-02 |
| RGP1 | 824,63 | 0,08 | 0,05 | 1,80 | 7,17E-02 |
| AKIRIN1 | 2745,65 | -0,06 | 0,03 | -1,80 | 7,17E-02 |
| HSBP1 | 1193,24 | -0,11 | 0,06 | -1,80 | 7,19E-02 |
| UNC45A | 808,54 | 0,06 | 0,03 | 1,80 | 7,19E-02 |

| | | | | | |
|---|---|---|---|---|---|
| RAC2 | 2508,66 | 0,14 | 0,08 | 1,80 | 7,19E-02 |
| ZNF540 | 318,04 | -0,12 | 0,06 | -1,80 | 7,19E-02 |
| CCNL2 | 4144,19 | 0,05 | 0,03 | 1,80 | 7,20E-02 |
| NHEJ1 | 60,99 | 0,11 | 0,06 | 1,80 | 7,21E-02 |
| C21orf91 | 2075,13 | -0,14 | 0,08 | -1,80 | 7,21E-02 |
| AMDHD1 | 53,97 | 0,14 | 0,08 | 1,80 | 7,21E-02 |
| DCTN2 | 605,24 | 0,08 | 0,04 | 1,80 | 7,22E-02 |
| ZNF747 | 192,20 | 0,06 | 0,03 | 1,80 | 7,22E-02 |
| FCRLA | 317,43 | -0,19 | 0,11 | -1,80 | 7,22E-02 |
| DYNLL2 | 3204,11 | -0,07 | 0,04 | -1,80 | 7,22E-02 |
| CSDE1 | 15085,90 | -0,09 | 0,05 | -1,80 | 7,23E-02 |
| SERPINB1 | 4125,28 | -0,09 | 0,05 | -1,80 | 7,23E-02 |
| ZNF280B | 67,15 | 0,16 | 0,09 | 1,80 | 7,23E-02 |
| MAVS | 2219,95 | 0,12 | 0,07 | 1,80 | 7,23E-02 |
| CDKN2A | 139,07 | 0,14 | 0,08 | 1,80 | 7,23E-02 |
| YOD1 | 11129,96 | 0,20 | 0,11 | 1,80 | 7,24E-02 |
| RPL13A | 29304,80 | -0,14 | 0,08 | -1,80 | 7,24E-02 |
| WWC2 | 324,63 | 0,10 | 0,06 | 1,80 | 7,24E-02 |
| CD2 | 1188,85 | -0,11 | 0,06 | -1,80 | 7,24E-02 |
| LARGE1 | 231,45 | 0,20 | 0,11 | 1,80 | 7,24E-02 |
| IRF9 | 529,77 | 0,14 | 0,08 | 1,80 | 7,24E-02 |
| TIMD4 | 35,99 | 0,17 | 0,09 | 1,80 | 7,25E-02 |
| DHRS9 | 1093,19 | -0,17 | 0,10 | -1,80 | 7,25E-02 |
| PAPSS1 | 1635,02 | 0,12 | 0,07 | 1,80 | 7,25E-02 |
| MAP3K8 | 1682,92 | -0,06 | 0,03 | -1,80 | 7,26E-02 |
| AMDHD2 | 751,01 | 0,08 | 0,04 | 1,79 | 7,27E-02 |
| ACTB | 27397,85 | 0,16 | 0,09 | 1,79 | 7,27E-02 |
| MDH2 | 492,30 | 0,08 | 0,04 | 1,79 | 7,28E-02 |
| AGAP5 | 59,96 | 0,10 | 0,06 | 1,79 | 7,28E-02 |
| MED18 | 640,58 | -0,07 | 0,04 | -1,79 | 7,28E-02 |
| TMEM199 | 640,64 | -0,04 | 0,02 | -1,79 | 7,29E-02 |
| PPARD | 469,25 | 0,11 | 0,06 | 1,79 | 7,30E-02 |
| FBXW5 | 1739,79 | 0,07 | 0,04 | 1,79 | 7,31E-02 |
| ZNF200 | 1267,10 | -0,07 | 0,04 | -1,79 | 7,31E-02 |
| ZSWIM7 | 349,79 | -0,05 | 0,03 | -1,79 | 7,32E-02 |
| POLR1D | 2082,93 | 0,10 | 0,06 | 1,79 | 7,32E-02 |
| PHF20L1 | 9977,97 | -0,11 | 0,06 | -1,79 | 7,32E-02 |
| PARP6 | 840,03 | 0,08 | 0,04 | 1,79 | 7,33E-02 |
| SEPSECS | 957,50 | -0,08 | 0,04 | -1,79 | 7,33E-02 |
| S1PR3 | 468,43 | 0,14 | 0,08 | 1,79 | 7,34E-02 |
| BCDIN3D | 324,18 | -0,08 | 0,04 | -1,79 | 7,35E-02 |
| ARL8A | 421,68 | 0,11 | 0,06 | 1,79 | 7,36E-02 |
| PTBP3 | 10482,35 | -0,07 | 0,04 | -1,79 | 7,36E-02 |
| ATG3 | 2778,61 | -0,10 | 0,06 | -1,79 | 7,36E-02 |
| XIAP | 4074,90 | -0,06 | 0,03 | -1,79 | 7,37E-02 |
| DPY19L2 | 81,38 | 0,21 | 0,12 | 1,79 | 7,37E-02 |

| | | | | | |
|---|---|---|---|---|---|
| DUSP7 | 628,40 | 0,08 | 0,04 | 1,79 | 7,37E-02 |
| SSX2IP | 558,76 | -0,12 | 0,07 | -1,79 | 7,37E-02 |
| TRAPPC9 | 1808,97 | 0,10 | 0,06 | 1,79 | 7,37E-02 |
| TMEM33 | 4653,86 | -0,09 | 0,05 | -1,79 | 7,37E-02 |
| PTK2B | 4435,53 | 0,13 | 0,08 | 1,79 | 7,38E-02 |
| ERCC8 | 472,08 | -0,10 | 0,05 | -1,79 | 7,38E-02 |
| MAP3K14 | 615,77 | 0,13 | 0,08 | 1,79 | 7,39E-02 |
| TANK | 6077,85 | -0,09 | 0,05 | -1,79 | 7,40E-02 |
| SULF2 | 4152,78 | -0,12 | 0,07 | -1,79 | 7,39E-02 |
| PLEKHA3 | 2253,38 | -0,06 | 0,03 | -1,79 | 7,40E-02 |
| TM2D3 | 1636,53 | -0,05 | 0,03 | -1,79 | 7,40E-02 |
| TGFBR3 | 1448,79 | -0,14 | 0,08 | -1,79 | 7,40E-02 |
| PLEKHO2 | 1066,88 | 0,16 | 0,09 | 1,79 | 7,41E-02 |
| TBC1D20 | 1346,71 | 0,06 | 0,03 | 1,78 | 7,43E-02 |
| ZNF20 | 77,63 | 0,09 | 0,05 | 1,78 | 7,44E-02 |
| TLR5 | 1066,56 | 0,15 | 0,08 | 1,78 | 7,45E-02 |
| HMMR | 33,70 | 0,26 | 0,15 | 1,78 | 7,45E-02 |
| TRAT1 | 1333,27 | -0,17 | 0,09 | -1,78 | 7,46E-02 |
| CHD4 | 446,66 | 0,08 | 0,04 | 1,78 | 7,46E-02 |
| TMEM106B | 2162,80 | -0,12 | 0,07 | -1,78 | 7,46E-02 |
| KCTD11 | 122,26 | 0,09 | 0,05 | 1,78 | 7,47E-02 |
| ZFP2 | 34,68 | 0,20 | 0,11 | 1,78 | 7,47E-02 |
| MRC1 | 65,73 | 0,25 | 0,14 | 1,78 | 7,47E-02 |
| ZNF623 | 1018,80 | -0,05 | 0,03 | -1,78 | 7,47E-02 |
| DHX33 | 372,76 | 0,06 | 0,03 | 1,78 | 7,47E-02 |
| BIRC6 | 15906,70 | -0,07 | 0,04 | -1,78 | 7,48E-02 |
| ZSCAN18 | 614,23 | 0,11 | 0,06 | 1,78 | 7,48E-02 |
| FAM13A | 1052,35 | 0,11 | 0,06 | 1,78 | 7,49E-02 |
| GTF3C1 | 1381,31 | 0,08 | 0,05 | 1,78 | 7,48E-02 |
| NLRP2 | 146,96 | 0,22 | 0,13 | 1,78 | 7,49E-02 |
| HSP90AB1 | 2201,88 | -0,07 | 0,04 | -1,78 | 7,49E-02 |
| ANAPC4 | 1528,41 | -0,07 | 0,04 | -1,78 | 7,50E-02 |
| GCDH | 460,82 | 0,06 | 0,03 | 1,78 | 7,50E-02 |
| ANKRD44 | 23663,03 | -0,06 | 0,04 | -1,78 | 7,50E-02 |
| MTR | 3431,30 | -0,06 | 0,03 | -1,78 | 7,50E-02 |
| RPUSD4 | 510,71 | -0,05 | 0,03 | -1,78 | 7,50E-02 |
| PRR14L | 4200,05 | -0,05 | 0,03 | -1,78 | 7,51E-02 |
| ARID5A | 652,43 | 0,13 | 0,07 | 1,78 | 7,51E-02 |
| SCAP | 1243,84 | 0,14 | 0,08 | 1,78 | 7,51E-02 |
| TMEM70 | 540,55 | -0,09 | 0,05 | -1,78 | 7,51E-02 |
| STRN3 | 1419,80 | -0,06 | 0,04 | -1,78 | 7,52E-02 |
| TAS2R41 | 52,30 | 0,23 | 0,13 | 1,78 | 7,52E-02 |
| TPRG1L | 1234,30 | 0,08 | 0,05 | 1,78 | 7,53E-02 |
| CYTH2 | 890,56 | 0,08 | 0,05 | 1,78 | 7,53E-02 |
| THYN1 | 395,06 | -0,06 | 0,03 | -1,78 | 7,53E-02 |
| GALNT14 | 182,50 | 0,22 | 0,13 | 1,78 | 7,55E-02 |

| | | | | | |
|---|---|---|---|---|---|
| RDH13 | 261,31 | 0,10 | 0,06 | 1,78 | 7,55E-02 |
| PRKCA | 1494,46 | 0,11 | 0,06 | 1,78 | 7,55E-02 |
| FLVCR2 | 357,88 | 0,11 | 0,06 | 1,78 | 7,56E-02 |
| RNF5 | 707,34 | 0,10 | 0,06 | 1,78 | 7,57E-02 |
| NUP62CL | 36,88 | 0,17 | 0,09 | 1,78 | 7,57E-02 |
| MS4A1 | 2067,81 | -0,17 | 0,10 | -1,78 | 7,58E-02 |
| NAP1L3 | 56,97 | -0,16 | 0,09 | -1,78 | 7,58E-02 |
| USP15 | 23396,70 | -0,12 | 0,07 | -1,78 | 7,58E-02 |
| COLQ | 243,78 | 0,14 | 0,08 | 1,78 | 7,58E-02 |
| GPR19 | 74,09 | 0,11 | 0,06 | 1,78 | 7,59E-02 |
| UPRT | 603,47 | -0,08 | 0,04 | -1,77 | 7,59E-02 |
| RAD9B | 35,49 | 0,13 | 0,07 | 1,77 | 7,60E-02 |
| EMP1 | 82,02 | 0,23 | 0,13 | 1,77 | 7,62E-02 |
| CCNB1IP1 | 361,07 | -0,09 | 0,05 | -1,77 | 7,62E-02 |
| HNRNPM | 2587,94 | -0,05 | 0,03 | -1,77 | 7,62E-02 |
| EIF2B2 | 536,76 | -0,05 | 0,03 | -1,77 | 7,62E-02 |
| HOXB2 | 126,36 | 0,15 | 0,08 | 1,77 | 7,63E-02 |
| RBPJ | 5520,89 | -0,08 | 0,05 | -1,77 | 7,64E-02 |
| HS6ST1 | 236,51 | 0,11 | 0,06 | 1,77 | 7,64E-02 |
| MANEA | 510,23 | -0,14 | 0,08 | -1,77 | 7,64E-02 |
| GMEB2 | 428,87 | 0,12 | 0,07 | 1,77 | 7,65E-02 |
| GIMAP1 | 2473,30 | -0,06 | 0,04 | -1,77 | 7,66E-02 |
| ZC3H6 | 1667,58 | -0,06 | 0,04 | -1,77 | 7,66E-02 |
| ZMYM1 | 560,44 | -0,09 | 0,05 | -1,77 | 7,66E-02 |
| SPACA6 | 257,07 | 0,16 | 0,09 | 1,77 | 7,67E-02 |
| NT5C2 | 7038,49 | -0,10 | 0,06 | -1,77 | 7,67E-02 |
| PTGS1 | 1190,23 | 0,15 | 0,09 | 1,77 | 7,68E-02 |
| NISCH | 1648,03 | 0,09 | 0,05 | 1,77 | 7,68E-02 |
| SRP19 | 632,58 | -0,05 | 0,03 | -1,77 | 7,68E-02 |
| TENT5C | 25059,21 | 0,23 | 0,13 | 1,77 | 7,69E-02 |
| RWDD3 | 245,40 | -0,08 | 0,05 | -1,77 | 7,70E-02 |
| PDPK1 | 1770,55 | 0,11 | 0,06 | 1,77 | 7,69E-02 |
| ZEB1 | 2434,80 | -0,08 | 0,04 | -1,77 | 7,69E-02 |
| CFAP36 | 307,31 | -0,09 | 0,05 | -1,77 | 7,71E-02 |
| LENG9 | 104,06 | 0,09 | 0,05 | 1,77 | 7,71E-02 |
| SDR39U1 | 402,67 | 0,07 | 0,04 | 1,77 | 7,71E-02 |
| EHD1 | 2546,72 | 0,11 | 0,06 | 1,77 | 7,72E-02 |
| SCCPDH | 327,10 | -0,11 | 0,06 | -1,77 | 7,72E-02 |
| EMC4 | 913,80 | -0,07 | 0,04 | -1,77 | 7,73E-02 |
| MOB3B | 273,61 | 0,10 | 0,06 | 1,77 | 7,73E-02 |
| ZNF677 | 172,66 | 0,14 | 0,08 | 1,77 | 7,73E-02 |
| L2HGDH | 217,42 | 0,08 | 0,04 | 1,77 | 7,73E-02 |
| FECH | 5071,06 | 0,21 | 0,12 | 1,77 | 7,73E-02 |
| VASH1 | 340,46 | 0,17 | 0,10 | 1,77 | 7,74E-02 |
| WDR44 | 566,57 | -0,06 | 0,03 | -1,77 | 7,74E-02 |
| SCAMP2 | 916,96 | 0,10 | 0,05 | 1,77 | 7,75E-02 |

| | | | | | |
|---|---|---|---|---|---|
| CMTM3 | 912,25 | 0,07 | 0,04 | 1,77 | 7,75E-02 |
| AKR1C3 | 189,23 | -0,18 | 0,10 | -1,77 | 7,75E-02 |
| NSUN4 | 443,69 | 0,07 | 0,04 | 1,77 | 7,76E-02 |
| TBC1D3L | 44,16 | 0,29 | 0,17 | 1,76 | 7,78E-02 |
| VAPB | 1686,12 | -0,06 | 0,03 | -1,76 | 7,78E-02 |
| MPZL3 | 5096,54 | -0,11 | 0,06 | -1,76 | 7,78E-02 |
| CEP20 | 485,82 | -0,10 | 0,05 | -1,76 | 7,80E-02 |
| PSMA2 | 394,88 | -0,05 | 0,03 | -1,76 | 7,80E-02 |
| HCFC2 | 996,75 | -0,10 | 0,05 | -1,76 | 7,81E-02 |
| ADAM22 | 359,29 | -0,12 | 0,07 | -1,76 | 7,81E-02 |
| ARID3B | 621,24 | 0,14 | 0,08 | 1,76 | 7,82E-02 |
| RABGEF1 | 254,47 | 0,05 | 0,03 | 1,76 | 7,82E-02 |
| XPO4 | 1984,87 | -0,07 | 0,04 | -1,76 | 7,83E-02 |
| SIRT5 | 553,40 | 0,04 | 0,02 | 1,76 | 7,85E-02 |
| YIPF3 | 466,84 | 0,13 | 0,07 | 1,76 | 7,85E-02 |
| MTUS1 | 139,61 | 0,25 | 0,14 | 1,76 | 7,85E-02 |
| NCAPG | 82,25 | 0,23 | 0,13 | 1,76 | 7,86E-02 |
| EID2 | 237,38 | -0,09 | 0,05 | -1,76 | 7,86E-02 |
| FHIT | 533,39 | 0,16 | 0,09 | 1,76 | 7,86E-02 |
| CDC42EP2 | 56,84 | 0,15 | 0,09 | 1,76 | 7,86E-02 |
| SMARCB1 | 503,84 | 0,13 | 0,07 | 1,76 | 7,87E-02 |
| YAE1 | 181,91 | -0,11 | 0,06 | -1,76 | 7,87E-02 |
| SLC35B3 | 1025,21 | -0,10 | 0,05 | -1,76 | 7,87E-02 |
| HYCC1 | 1941,52 | -0,09 | 0,05 | -1,76 | 7,88E-02 |
| HNRNPH1 | 16842,69 | -0,08 | 0,04 | -1,76 | 7,88E-02 |
| ZMIZ1 | 1542,85 | 0,14 | 0,08 | 1,76 | 7,89E-02 |
| ARID1A | 3228,74 | 0,12 | 0,07 | 1,76 | 7,89E-02 |
| NBAS | 3127,19 | -0,06 | 0,04 | -1,76 | 7,89E-02 |
| ATF6 | 4991,12 | -0,08 | 0,04 | -1,76 | 7,89E-02 |
| BTG2 | 1703,04 | 0,13 | 0,07 | 1,76 | 7,89E-02 |
| ANXA7 | 2525,93 | -0,06 | 0,03 | -1,76 | 7,89E-02 |
| PTCRA | 164,41 | 0,17 | 0,10 | 1,76 | 7,89E-02 |
| NTMT1 | 219,08 | 0,07 | 0,04 | 1,76 | 7,90E-02 |
| TTC7A | 1793,76 | 0,09 | 0,05 | 1,76 | 7,90E-02 |
| GNAZ | 416,57 | 0,18 | 0,10 | 1,76 | 7,92E-02 |
| SUPT5H | 1416,50 | 0,10 | 0,06 | 1,76 | 7,92E-02 |
| ECT2 | 303,57 | -0,12 | 0,07 | -1,76 | 7,92E-02 |
| THTPA | 157,10 | 0,08 | 0,05 | 1,76 | 7,92E-02 |
| NHP2 | 514,89 | -0,08 | 0,04 | -1,76 | 7,92E-02 |
| TIRAP | 371,81 | 0,11 | 0,06 | 1,76 | 7,92E-02 |
| IFI16 | 6046,19 | -0,11 | 0,06 | -1,75 | 7,93E-02 |
| TMEM135 | 436,05 | -0,09 | 0,05 | -1,75 | 7,93E-02 |
| RCBTB2 | 3098,77 | -0,09 | 0,05 | -1,75 | 7,93E-02 |
| H1-0 | 758,26 | 0,22 | 0,13 | 1,75 | 7,93E-02 |
| MCUR1 | 422,05 | 0,07 | 0,04 | 1,75 | 7,94E-02 |
| TOP1 | 2443,10 | -0,07 | 0,04 | -1,75 | 7,95E-02 |

| | | | | | |
|---|---|---|---|---|---|
| ZBTB4 | 949,42 | 0,11 | 0,06 | 1,75 | 7,95E-02 |
| DTD1 | 386,54 | -0,06 | 0,03 | -1,75 | 7,95E-02 |
| MBOAT7 | 1947,87 | 0,18 | 0,10 | 1,75 | 7,95E-02 |
| SRPK2 | 5097,69 | -0,08 | 0,04 | -1,75 | 7,96E-02 |
| ENSG00000282034 | 39,41 | 0,10 | 0,06 | 1,75 | 7,97E-02 |
| CDC25B | 912,86 | 0,16 | 0,09 | 1,75 | 7,98E-02 |
| SMYD2 | 1064,05 | -0,06 | 0,03 | -1,75 | 7,99E-02 |
| MAPK13 | 1161,55 | 0,17 | 0,10 | 1,75 | 7,99E-02 |
| METTL16 | 575,00 | 0,09 | 0,05 | 1,75 | 7,99E-02 |
| RPL3 | 11700,70 | -0,11 | 0,07 | -1,75 | 8,00E-02 |
| BCORL1 | 248,82 | 0,17 | 0,09 | 1,75 | 8,00E-02 |
| LIPT1 | 136,77 | -0,08 | 0,05 | -1,75 | 8,02E-02 |
| TWF1 | 1071,13 | -0,14 | 0,08 | -1,75 | 8,02E-02 |
| CMTM4 | 286,50 | 0,09 | 0,05 | 1,75 | 8,02E-02 |
| CRY2 | 852,27 | 0,06 | 0,03 | 1,75 | 8,02E-02 |
| BRD1 | 1433,10 | 0,08 | 0,05 | 1,75 | 8,03E-02 |
| CCHCR1 | 267,49 | 0,08 | 0,04 | 1,75 | 8,03E-02 |
| HSPBAP1 | 1866,17 | -0,08 | 0,05 | -1,75 | 8,03E-02 |
| SLC2A14 | 56,63 | 0,33 | 0,19 | 1,75 | 8,04E-02 |
| FASTKD1 | 692,34 | -0,09 | 0,05 | -1,75 | 8,05E-02 |
| TCEAL4 | 154,51 | -0,12 | 0,07 | -1,75 | 8,05E-02 |
| TM9SF2 | 3461,82 | -0,08 | 0,04 | -1,75 | 8,05E-02 |
| FAM168B | 2078,29 | -0,05 | 0,03 | -1,75 | 8,05E-02 |
| RHOT1 | 2384,18 | -0,09 | 0,05 | -1,75 | 8,06E-02 |
| NAT1 | 264,73 | -0,10 | 0,06 | -1,75 | 8,07E-02 |
| PREP | 817,42 | -0,06 | 0,03 | -1,75 | 8,07E-02 |
| C19orf25 | 849,26 | 0,07 | 0,04 | 1,75 | 8,08E-02 |
| EHBP1L1 | 8228,46 | 0,11 | 0,06 | 1,75 | 8,08E-02 |
| CYB561A3 | 1057,74 | 0,08 | 0,05 | 1,75 | 8,09E-02 |
| MFF | 1529,47 | -0,08 | 0,05 | -1,75 | 8,09E-02 |
| GIPC1 | 181,67 | 0,10 | 0,06 | 1,75 | 8,09E-02 |
| LRP12 | 146,46 | 0,13 | 0,07 | 1,74 | 8,10E-02 |
| CRIPT | 729,71 | -0,06 | 0,04 | -1,74 | 8,10E-02 |
| IL17RA | 6570,96 | 0,15 | 0,09 | 1,74 | 8,10E-02 |
| CNNM3 | 657,58 | 0,10 | 0,06 | 1,74 | 8,11E-02 |
| TOMM22 | 1071,44 | -0,09 | 0,05 | -1,74 | 8,11E-02 |
| HSD17B12 | 658,66 | -0,07 | 0,04 | -1,74 | 8,12E-02 |
| GPRIN3 | 4466,63 | -0,10 | 0,06 | -1,74 | 8,13E-02 |
| RINL | 932,64 | 0,06 | 0,03 | 1,74 | 8,13E-02 |
| CYFIP1 | 989,16 | 0,10 | 0,06 | 1,74 | 8,14E-02 |
| ZNF543 | 390,86 | -0,06 | 0,04 | -1,74 | 8,14E-02 |
| DAZAP1 | 1749,31 | 0,09 | 0,05 | 1,74 | 8,14E-02 |
| KPNA5 | 1238,65 | -0,14 | 0,08 | -1,74 | 8,14E-02 |
| INTS3 | 1520,33 | 0,11 | 0,06 | 1,74 | 8,14E-02 |
| CALCB | 55,08 | 0,19 | 0,11 | 1,74 | 8,16E-02 |
| COG5 | 2469,16 | -0,07 | 0,04 | -1,74 | 8,17E-02 |

| | | | | | |
|---|---|---|---|---|---|
| CEP41 | 228,49 | 0,10 | 0,06 | 1,74 | 8,17E-02 |
| ARF3 | 1184,65 | 0,14 | 0,08 | 1,74 | 8,17E-02 |
| ATF7IP2 | 989,88 | -0,08 | 0,05 | -1,74 | 8,18E-02 |
| TMEM260 | 2281,64 | -0,07 | 0,04 | -1,74 | 8,18E-02 |
| USP10 | 3483,72 | -0,09 | 0,05 | -1,74 | 8,19E-02 |
| TMTC2 | 903,90 | 0,08 | 0,05 | 1,74 | 8,19E-02 |
| ATG16L1 | 1842,87 | -0,05 | 0,03 | -1,74 | 8,20E-02 |
| RPN2 | 1931,96 | -0,05 | 0,03 | -1,74 | 8,20E-02 |
| BYSL | 103,30 | 0,08 | 0,05 | 1,74 | 8,21E-02 |
| MDGA1 | 256,14 | 0,40 | 0,23 | 1,74 | 8,21E-02 |
| POLR2A | 1538,68 | 0,18 | 0,10 | 1,74 | 8,22E-02 |
| MED12 | 864,73 | 0,18 | 0,10 | 1,74 | 8,22E-02 |
| STARD3NL | 817,31 | -0,08 | 0,04 | -1,74 | 8,24E-02 |
| SUMO4 | 375,10 | -0,12 | 0,07 | -1,74 | 8,24E-02 |
| PURB | 3912,42 | -0,08 | 0,04 | -1,74 | 8,25E-02 |
| TNNI2 | 85,73 | 0,12 | 0,07 | 1,74 | 8,26E-02 |
| ATP5MJ | 430,45 | -0,13 | 0,08 | -1,74 | 8,26E-02 |
| NOP16 | 128,62 | 0,07 | 0,04 | 1,74 | 8,27E-02 |
| EMSY | 2260,44 | -0,06 | 0,03 | -1,73 | 8,28E-02 |
| AOAH | 7577,24 | -0,10 | 0,06 | -1,73 | 8,28E-02 |
| PELI2 | 3960,84 | -0,09 | 0,05 | -1,73 | 8,29E-02 |
| SLC2A3 | 10074,81 | -0,12 | 0,07 | -1,73 | 8,29E-02 |
| PCYOX1L | 482,51 | 0,08 | 0,04 | 1,73 | 8,30E-02 |
| CNR2 | 329,42 | 0,14 | 0,08 | 1,73 | 8,30E-02 |
| LTO1 | 953,70 | 0,06 | 0,03 | 1,73 | 8,31E-02 |
| STAMBP | 1339,79 | -0,05 | 0,03 | -1,73 | 8,31E-02 |
| FITM2 | 235,56 | 0,09 | 0,05 | 1,73 | 8,32E-02 |
| SLC38A10 | 2003,51 | 0,12 | 0,07 | 1,73 | 8,33E-02 |
| GPN3 | 268,57 | -0,11 | 0,06 | -1,73 | 8,35E-02 |
| ITGA4 | 12351,34 | -0,09 | 0,05 | -1,73 | 8,36E-02 |
| NDUFAF5 | 499,56 | -0,06 | 0,03 | -1,73 | 8,37E-02 |
| SEMA4D | 6931,76 | 0,15 | 0,09 | 1,73 | 8,37E-02 |
| CTNNBIP1 | 186,68 | 0,12 | 0,07 | 1,73 | 8,37E-02 |
| ARSK | 212,71 | -0,09 | 0,05 | -1,73 | 8,38E-02 |
| MTRES1 | 198,71 | -0,07 | 0,04 | -1,73 | 8,39E-02 |
| TBC1D5 | 3541,95 | -0,05 | 0,03 | -1,73 | 8,41E-02 |
| SPATA7 | 141,10 | 0,10 | 0,06 | 1,73 | 8,41E-02 |
| DNAAF11 | 446,43 | -0,17 | 0,10 | -1,73 | 8,42E-02 |
| MFSD6 | 1294,58 | -0,06 | 0,03 | -1,73 | 8,42E-02 |
| DZANK1 | 161,98 | 0,13 | 0,07 | 1,73 | 8,45E-02 |
| TNNT1 | 73,36 | 0,27 | 0,16 | 1,73 | 8,45E-02 |
| MITD1 | 891,10 | -0,07 | 0,04 | -1,72 | 8,46E-02 |
| ABHD13 | 2171,66 | -0,14 | 0,08 | -1,72 | 8,47E-02 |
| PLPPR2 | 393,09 | 0,18 | 0,10 | 1,72 | 8,47E-02 |
| CEP95 | 1295,75 | -0,06 | 0,03 | -1,72 | 8,47E-02 |
| SLC26A8 | 241,97 | 0,20 | 0,12 | 1,72 | 8,48E-02 |

| | | | | | |
|---|---|---|---|---|---|
| CSTF3 | 808,50 | -0,08 | 0,05 | -1,72 | 8,48E-02 |
| DCAF11 | 2962,26 | 0,07 | 0,04 | 1,72 | 8,48E-02 |
| CYB561D1 | 1023,71 | 0,08 | 0,05 | 1,72 | 8,48E-02 |
| RPP14 | 684,47 | -0,04 | 0,02 | -1,72 | 8,48E-02 |
| SNAI3 | 185,39 | 0,11 | 0,06 | 1,72 | 8,48E-02 |
| NHLRC1 | 37,93 | 0,13 | 0,07 | 1,72 | 8,48E-02 |
| SMAD5 | 1471,44 | -0,09 | 0,05 | -1,72 | 8,49E-02 |
| TEK | 71,72 | 0,30 | 0,17 | 1,72 | 8,49E-02 |
| DCLRE1A | 275,31 | -0,11 | 0,06 | -1,72 | 8,49E-02 |
| FGD4 | 3306,98 | -0,11 | 0,06 | -1,72 | 8,50E-02 |
| CLCN6 | 560,12 | 0,10 | 0,06 | 1,72 | 8,50E-02 |
| SGF29 | 258,90 | 0,06 | 0,04 | 1,72 | 8,51E-02 |
| PHYH | 115,53 | 0,10 | 0,06 | 1,72 | 8,51E-02 |
| CHD6 | 3517,32 | -0,06 | 0,03 | -1,72 | 8,52E-02 |
| NUP210 | 3284,04 | 0,11 | 0,07 | 1,72 | 8,52E-02 |
| SIRPG | 160,37 | 0,12 | 0,07 | 1,72 | 8,53E-02 |
| ARPC5 | 17499,42 | -0,10 | 0,06 | -1,72 | 8,53E-02 |
| GOLGA7 | 1487,07 | -0,07 | 0,04 | -1,72 | 8,53E-02 |
| FSTL1 | 193,06 | 0,22 | 0,13 | 1,72 | 8,53E-02 |
| VPS37A | 1223,01 | -0,06 | 0,04 | -1,72 | 8,55E-02 |
| INTS8 | 2667,70 | -0,05 | 0,03 | -1,72 | 8,57E-02 |
| NUSAP1 | 35,37 | 0,13 | 0,08 | 1,72 | 8,57E-02 |
| KCNAB2 | 3040,72 | 0,11 | 0,06 | 1,72 | 8,58E-02 |
| PIK3R3 | 112,21 | 0,14 | 0,08 | 1,72 | 8,59E-02 |
| ZNF256 | 168,85 | -0,09 | 0,05 | -1,72 | 8,59E-02 |
| CEP128 | 684,23 | -0,09 | 0,05 | -1,72 | 8,59E-02 |
| SWAP70 | 1598,82 | -0,10 | 0,06 | -1,72 | 8,60E-02 |
| FKBP1A | 1826,00 | 0,11 | 0,06 | 1,72 | 8,61E-02 |
| RNPC3 | 1983,04 | -0,08 | 0,05 | -1,72 | 8,61E-02 |
| IER5L | 186,38 | 0,11 | 0,06 | 1,72 | 8,61E-02 |
| FABP5 | 41,97 | 0,18 | 0,11 | 1,72 | 8,62E-02 |
| FTSJ3 | 1018,37 | -0,05 | 0,03 | -1,72 | 8,63E-02 |
| UBE3B | 1861,27 | 0,06 | 0,03 | 1,71 | 8,65E-02 |
| APH1A | 2170,93 | -0,05 | 0,03 | -1,71 | 8,66E-02 |
| DONSON | 160,46 | 0,05 | 0,03 | 1,71 | 8,66E-02 |
| FBXO30 | 1637,04 | -0,11 | 0,06 | -1,71 | 8,66E-02 |
| PDGFRB | 110,48 | 0,24 | 0,14 | 1,71 | 8,66E-02 |
| UQCRH | 851,20 | -0,14 | 0,08 | -1,71 | 8,67E-02 |
| PRMT2 | 5273,06 | -0,06 | 0,03 | -1,71 | 8,68E-02 |
| PFN2 | 98,33 | 0,16 | 0,09 | 1,71 | 8,70E-02 |
| CD320 | 86,83 | 0,15 | 0,09 | 1,71 | 8,70E-02 |
| IMP3 | 566,09 | -0,08 | 0,04 | -1,71 | 8,70E-02 |
| TAS2R30 | 403,34 | -0,11 | 0,07 | -1,71 | 8,71E-02 |
| CYP2U1 | 341,49 | -0,08 | 0,05 | -1,71 | 8,71E-02 |
| SNU13 | 1094,71 | -0,05 | 0,03 | -1,71 | 8,71E-02 |
| SLC35F5 | 2914,59 | -0,07 | 0,04 | -1,71 | 8,71E-02 |

| | | | | | |
|---|---|---|---|---|---|
| TLE1 | 324,95 | 0,11 | 0,06 | 1,71 | 8,71E-02 |
| FUCA2 | 503,34 | -0,07 | 0,04 | -1,71 | 8,72E-02 |
| VPS37C | 318,71 | 0,12 | 0,07 | 1,71 | 8,72E-02 |
| CCDC57 | 1005,05 | 0,12 | 0,07 | 1,71 | 8,72E-02 |
| SLF1 | 1045,76 | -0,10 | 0,06 | -1,71 | 8,74E-02 |
| SYCP2L | 49,08 | 0,35 | 0,20 | 1,71 | 8,74E-02 |
| MTO1 | 1257,64 | -0,05 | 0,03 | -1,71 | 8,74E-02 |
| PASK | 1132,97 | 0,10 | 0,06 | 1,71 | 8,75E-02 |
| DACT1 | 121,92 | 0,24 | 0,14 | 1,71 | 8,75E-02 |
| COP1 | 7681,58 | -0,09 | 0,05 | -1,71 | 8,76E-02 |
| BET1 | 664,30 | -0,11 | 0,06 | -1,71 | 8,78E-02 |
| MDC1 | 414,85 | 0,09 | 0,06 | 1,71 | 8,79E-02 |
| UBC | 11873,78 | -0,08 | 0,05 | -1,71 | 8,80E-02 |
| PHF20 | 2961,35 | -0,05 | 0,03 | -1,71 | 8,80E-02 |
| FYN | 7360,07 | -0,07 | 0,04 | -1,71 | 8,80E-02 |
| GPR137B | 193,23 | 0,10 | 0,06 | 1,71 | 8,81E-02 |
| LBH | 5656,76 | -0,09 | 0,05 | -1,70 | 8,82E-02 |
| SCOC | 331,51 | -0,16 | 0,10 | -1,70 | 8,83E-02 |
| CSNK2B | 205,65 | 0,09 | 0,05 | 1,70 | 8,83E-02 |
| ABCD2 | 1269,25 | -0,15 | 0,09 | -1,70 | 8,83E-02 |
| SLC19A1 | 2010,27 | 0,17 | 0,10 | 1,70 | 8,83E-02 |
| HLA-DPA1 | 4978,31 | -0,09 | 0,06 | -1,70 | 8,85E-02 |
| CRTAP | 1768,76 | -0,06 | 0,03 | -1,70 | 8,85E-02 |
| ATG16L2 | 10924,74 | 0,16 | 0,09 | 1,70 | 8,85E-02 |
| VTI1A | 2643,96 | -0,06 | 0,03 | -1,70 | 8,87E-02 |
| ADRB2 | 1336,60 | -0,09 | 0,06 | -1,70 | 8,88E-02 |
| ITGAV | 927,49 | -0,08 | 0,04 | -1,70 | 8,88E-02 |
| PATZ1 | 584,46 | 0,08 | 0,04 | 1,70 | 8,89E-02 |
| IGFBP4 | 59,46 | 0,19 | 0,11 | 1,70 | 8,89E-02 |
| ARHGAP25 | 10690,94 | -0,07 | 0,04 | -1,70 | 8,89E-02 |
| MAN2B1 | 2013,18 | 0,08 | 0,05 | 1,70 | 8,89E-02 |
| UBE2G1 | 1295,03 | -0,04 | 0,02 | -1,70 | 8,89E-02 |
| HERC4 | 6440,22 | -0,06 | 0,04 | -1,70 | 8,90E-02 |
| INPP5A | 1021,72 | 0,08 | 0,05 | 1,70 | 8,90E-02 |
| ASRGL1 | 260,46 | 0,10 | 0,06 | 1,70 | 8,91E-02 |
| FAM131A | 559,55 | 0,09 | 0,05 | 1,70 | 8,91E-02 |
| CDK14 | 1221,14 | -0,12 | 0,07 | -1,70 | 8,92E-02 |
| RRP12 | 858,36 | 0,14 | 0,09 | 1,70 | 8,93E-02 |
| KDM4A | 1148,63 | -0,05 | 0,03 | -1,70 | 8,93E-02 |
| PLEKHA8 | 792,59 | -0,06 | 0,03 | -1,70 | 8,93E-02 |
| CHFR | 2686,30 | 0,07 | 0,04 | 1,70 | 8,94E-02 |
| SUPT20H | 3832,34 | -0,08 | 0,05 | -1,70 | 8,94E-02 |
| HTRA2 | 762,07 | 0,05 | 0,03 | 1,70 | 8,94E-02 |
| FPGT-TNNI3K | 41,48 | 0,16 | 0,09 | 1,70 | 8,95E-02 |
| USP30 | 290,99 | 0,05 | 0,03 | 1,70 | 8,95E-02 |
| VPS13B | 10662,09 | -0,06 | 0,04 | -1,70 | 8,96E-02 |

| | | | | | |
|---|---|---|---|---|---|
| PSMB7 | 1141,71 | -0,06 | 0,04 | -1,70 | 8,96E-02 |
| ST6GALNAC6 | 278,32 | 0,11 | 0,06 | 1,70 | 8,96E-02 |
| BORCS7 | 359,25 | -0,12 | 0,07 | -1,70 | 8,97E-02 |
| ADCY7 | 7228,15 | 0,08 | 0,04 | 1,70 | 8,97E-02 |
| PPIL2 | 935,49 | 0,09 | 0,05 | 1,70 | 8,97E-02 |
| ABAT | 661,40 | 0,12 | 0,07 | 1,70 | 8,98E-02 |
| NAT9 | 450,90 | 0,05 | 0,03 | 1,70 | 8,98E-02 |
| WIPI2 | 2127,84 | 0,08 | 0,05 | 1,70 | 8,98E-02 |
| CNNM2 | 481,69 | 0,09 | 0,05 | 1,70 | 9,00E-02 |
| HM13 | 1242,80 | 0,13 | 0,08 | 1,70 | 9,00E-02 |
| SENP3 | 303,84 | 0,05 | 0,03 | 1,69 | 9,01E-02 |
| SLC30A4 | 270,15 | -0,08 | 0,05 | -1,69 | 9,02E-02 |
| WDR33 | 3365,99 | -0,05 | 0,03 | -1,69 | 9,03E-02 |
| TMEM18 | 448,97 | -0,04 | 0,03 | -1,69 | 9,03E-02 |
| DTNB | 380,94 | 0,09 | 0,05 | 1,69 | 9,04E-02 |
| ADAMTS10 | 301,44 | 0,15 | 0,09 | 1,69 | 9,04E-02 |
| FCHO1 | 1154,27 | 0,10 | 0,06 | 1,69 | 9,05E-02 |
| ACACA | 837,95 | 0,09 | 0,05 | 1,69 | 9,05E-02 |
| STK35 | 869,49 | 0,09 | 0,06 | 1,69 | 9,06E-02 |
| SLC1A7 | 65,27 | 0,29 | 0,17 | 1,69 | 9,06E-02 |
| TOM1L2 | 699,00 | 0,11 | 0,06 | 1,69 | 9,07E-02 |
| METTL21A | 497,56 | -0,05 | 0,03 | -1,69 | 9,09E-02 |
| SS18 | 2449,92 | -0,07 | 0,04 | -1,69 | 9,10E-02 |
| SPTA1 | 459,94 | 0,21 | 0,12 | 1,69 | 9,11E-02 |
| PWP2 | 95,22 | -0,36 | 0,21 | -1,69 | 9,11E-02 |
| FMC1 | 51,31 | 0,08 | 0,05 | 1,69 | 9,12E-02 |
| HILPDA | 67,90 | 0,09 | 0,05 | 1,69 | 9,14E-02 |
| UNC119 | 1572,89 | 0,08 | 0,05 | 1,69 | 9,14E-02 |
| TIMM44 | 305,25 | 0,07 | 0,04 | 1,69 | 9,16E-02 |
| IL10RB | 356,17 | 0,10 | 0,06 | 1,69 | 9,16E-02 |
| ARL8B | 1611,55 | -0,06 | 0,04 | -1,69 | 9,17E-02 |
| RAB9B | 39,47 | 0,13 | 0,08 | 1,69 | 9,17E-02 |
| TIGD5 | 250,23 | 0,08 | 0,04 | 1,69 | 9,18E-02 |
| INTS6 | 6222,13 | -0,07 | 0,04 | -1,69 | 9,19E-02 |
| IRAK1BP1 | 148,82 | -0,12 | 0,07 | -1,69 | 9,20E-02 |
| CIRBP | 8745,44 | -0,06 | 0,03 | -1,68 | 9,22E-02 |
| TTLL1 | 105,65 | 0,08 | 0,05 | 1,68 | 9,23E-02 |
| PF4 | 2285,17 | 0,18 | 0,11 | 1,68 | 9,24E-02 |
| ZFTRAF1 | 992,52 | 0,11 | 0,07 | 1,68 | 9,25E-02 |
| MED30 | 503,39 | -0,09 | 0,05 | -1,68 | 9,25E-02 |
| ZNF718 | 641,63 | -0,09 | 0,05 | -1,68 | 9,25E-02 |
| LINC02218 | 338,21 | -0,15 | 0,09 | -1,68 | 9,27E-02 |
| COX19 | 1245,05 | -0,04 | 0,03 | -1,68 | 9,28E-02 |
| CHPF2 | 1247,53 | 0,11 | 0,06 | 1,68 | 9,28E-02 |
| ZNF395 | 302,76 | 0,13 | 0,08 | 1,68 | 9,30E-02 |
| RREB1 | 2377,18 | 0,09 | 0,05 | 1,68 | 9,30E-02 |

| | | | | | |
|---|---|---|---|---|---|
| NECAP1 | 1782,97 | -0,04 | 0,03 | -1,68 | 9,31E-02 |
| PSTPIP1 | 4095,07 | 0,10 | 0,06 | 1,68 | 9,31E-02 |
| SMARCC2 | 1196,36 | 0,10 | 0,06 | 1,68 | 9,31E-02 |
| ENSG00000284431 | 23,43 | 0,10 | 0,06 | 1,68 | 9,32E-02 |
| NDUFS2 | 1293,89 | -0,04 | 0,03 | -1,68 | 9,32E-02 |
| SCYL2 | 2771,28 | -0,07 | 0,04 | -1,68 | 9,32E-02 |
| APMAP | 4968,88 | -0,10 | 0,06 | -1,68 | 9,33E-02 |
| CYP2R1 | 625,72 | -0,07 | 0,04 | -1,68 | 9,33E-02 |
| ZNF862 | 1072,10 | 0,07 | 0,04 | 1,68 | 9,34E-02 |
| MCCC1 | 807,39 | -0,05 | 0,03 | -1,68 | 9,34E-02 |
| SMARCD2 | 2299,35 | -0,05 | 0,03 | -1,68 | 9,35E-02 |
| TGFBRAP1 | 652,05 | 0,06 | 0,04 | 1,68 | 9,38E-02 |
| ALG14 | 106,79 | 0,10 | 0,06 | 1,68 | 9,38E-02 |
| NHLRC2 | 2044,78 | -0,07 | 0,04 | -1,68 | 9,38E-02 |
| DOK3 | 1985,45 | 0,14 | 0,09 | 1,68 | 9,39E-02 |
| CDCA4 | 259,78 | 0,07 | 0,04 | 1,67 | 9,41E-02 |
| NSG1 | 135,23 | 0,19 | 0,12 | 1,67 | 9,43E-02 |
| IP6K2 | 1240,94 | -0,04 | 0,02 | -1,67 | 9,43E-02 |
| MAN2B2 | 1230,76 | 0,10 | 0,06 | 1,67 | 9,43E-02 |
| SH2D3C | 1555,30 | 0,13 | 0,07 | 1,67 | 9,43E-02 |
| ZC3H10 | 696,43 | -0,04 | 0,02 | -1,67 | 9,44E-02 |
| KYNU | 1853,22 | -0,11 | 0,06 | -1,67 | 9,45E-02 |
| TRIP12 | 12476,55 | -0,06 | 0,04 | -1,67 | 9,46E-02 |
| HNRNPDL | 3699,89 | -0,06 | 0,04 | -1,67 | 9,46E-02 |
| CHP1 | 1262,69 | 0,10 | 0,06 | 1,67 | 9,47E-02 |
| NEO1 | 271,05 | 0,13 | 0,08 | 1,67 | 9,47E-02 |
| IRF7 | 509,89 | 0,18 | 0,11 | 1,67 | 9,49E-02 |
| ARSD | 453,39 | 0,08 | 0,05 | 1,67 | 9,49E-02 |
| VRK3 | 1721,53 | 0,06 | 0,04 | 1,67 | 9,51E-02 |
| CALM3 | 1260,60 | 0,10 | 0,06 | 1,67 | 9,51E-02 |
| SPATA6 | 359,21 | 0,11 | 0,07 | 1,67 | 9,52E-02 |
| GSAP | 3381,18 | -0,08 | 0,05 | -1,67 | 9,53E-02 |
| SLC25A43 | 168,01 | 0,07 | 0,04 | 1,67 | 9,55E-02 |
| ZFP1 | 210,04 | -0,09 | 0,05 | -1,67 | 9,55E-02 |
| SLC27A3 | 600,75 | 0,09 | 0,05 | 1,67 | 9,55E-02 |
| PSME1 | 5003,89 | -0,09 | 0,05 | -1,67 | 9,56E-02 |
| TAS2R60 | 49,21 | 0,24 | 0,15 | 1,67 | 9,58E-02 |
| PYURF | 694,22 | -0,12 | 0,07 | -1,67 | 9,58E-02 |
| RPL35A | 3810,04 | -0,17 | 0,10 | -1,66 | 9,59E-02 |
| MED25 | 1508,21 | 0,11 | 0,07 | 1,66 | 9,60E-02 |
| FAM43A | 437,82 | -0,08 | 0,05 | -1,66 | 9,60E-02 |
| PYGB | 1250,39 | 0,07 | 0,04 | 1,66 | 9,60E-02 |
| JAZF1 | 1829,51 | 0,12 | 0,07 | 1,66 | 9,61E-02 |
| LXN | 194,13 | -0,11 | 0,07 | -1,66 | 9,63E-02 |
| DYSF | 5805,74 | 0,19 | 0,11 | 1,66 | 9,64E-02 |
| CCDC12 | 837,29 | -0,05 | 0,03 | -1,66 | 9,64E-02 |

| | | | | | |
|---|---|---|---|---|---|
| TIMM23B | 566,93 | -0,06 | 0,03 | -1,66 | 9,65E-02 |
| PLIN5 | 337,39 | 0,16 | 0,09 | 1,66 | 9,65E-02 |
| TAS2R31 | 291,93 | -0,11 | 0,07 | -1,66 | 9,65E-02 |
| ATRIP | 226,21 | 0,06 | 0,03 | 1,66 | 9,66E-02 |
| UCHL3 | 101,90 | -0,09 | 0,06 | -1,66 | 9,66E-02 |
| TCF12 | 1997,89 | -0,05 | 0,03 | -1,66 | 9,66E-02 |
| DNPEP | 614,94 | 0,06 | 0,03 | 1,66 | 9,67E-02 |
| SEPTIN10 | 110,65 | 0,14 | 0,08 | 1,66 | 9,67E-02 |
| RCN2 | 451,92 | -0,08 | 0,05 | -1,66 | 9,67E-02 |
| ZNF8 | 378,06 | 0,09 | 0,05 | 1,66 | 9,68E-02 |
| MAP3K7CL | 992,19 | -0,13 | 0,08 | -1,66 | 9,68E-02 |
| PA2G4 | 807,93 | 0,08 | 0,05 | 1,66 | 9,68E-02 |
| AEN | 846,58 | 0,07 | 0,04 | 1,66 | 9,68E-02 |
| REX1BD | 681,98 | 0,06 | 0,04 | 1,66 | 9,70E-02 |
| ZNF708 | 1376,36 | -0,11 | 0,07 | -1,66 | 9,71E-02 |
| TCL1A | 549,40 | -0,21 | 0,12 | -1,66 | 9,70E-02 |
| PMS2 | 449,72 | -0,04 | 0,02 | -1,66 | 9,72E-02 |
| KATNIP | 907,30 | 0,10 | 0,06 | 1,66 | 9,72E-02 |
| JAKMIP2 | 457,96 | -0,15 | 0,09 | -1,66 | 9,74E-02 |
| GRPEL2 | 759,54 | -0,09 | 0,05 | -1,66 | 9,74E-02 |
| GUCY1A1 | 276,45 | 0,13 | 0,08 | 1,66 | 9,76E-02 |
| ARHGDIB | 9415,41 | -0,06 | 0,04 | -1,66 | 9,76E-02 |
| ABCA5 | 2237,44 | -0,09 | 0,05 | -1,66 | 9,76E-02 |
| AHI1 | 365,20 | -0,13 | 0,08 | -1,66 | 9,76E-02 |
| STX18 | 664,33 | -0,03 | 0,02 | -1,66 | 9,78E-02 |
| NEK11 | 162,15 | 0,11 | 0,07 | 1,66 | 9,79E-02 |
| MTA3 | 350,31 | 0,08 | 0,05 | 1,66 | 9,79E-02 |
| LHFPL2 | 587,86 | 0,14 | 0,09 | 1,65 | 9,79E-02 |
| HARS1 | 1035,98 | -0,04 | 0,02 | -1,65 | 9,81E-02 |
| MTRF1 | 519,90 | -0,05 | 0,03 | -1,65 | 9,81E-02 |
| ZNF69 | 160,23 | -0,08 | 0,05 | -1,65 | 9,82E-02 |
| OFD1 | 2043,28 | -0,07 | 0,04 | -1,65 | 9,83E-02 |
| CTSD | 1042,77 | 0,11 | 0,06 | 1,65 | 9,83E-02 |
| TMEM107 | 103,17 | 0,06 | 0,04 | 1,65 | 9,84E-02 |
| SYNE2 | 23868,68 | -0,08 | 0,05 | -1,65 | 9,86E-02 |
| DPEP3 | 130,04 | 0,18 | 0,11 | 1,65 | 9,86E-02 |
| IQSEC1 | 5969,04 | 0,12 | 0,07 | 1,65 | 9,87E-02 |
| TPD52 | 648,03 | -0,10 | 0,06 | -1,65 | 9,87E-02 |
| OXSM | 176,05 | -0,07 | 0,04 | -1,65 | 9,89E-02 |
| PIP5K1B | 526,84 | 0,09 | 0,05 | 1,65 | 9,90E-02 |
| GDI1 | 919,00 | 0,10 | 0,06 | 1,65 | 9,91E-02 |
| ARL6 | 185,39 | -0,12 | 0,07 | -1,65 | 9,91E-02 |
| BAZ2A | 3256,47 | 0,13 | 0,08 | 1,65 | 9,91E-02 |
| ARHGEF2 | 4620,15 | 0,10 | 0,06 | 1,65 | 9,91E-02 |
| ITGAL | 5263,04 | 0,12 | 0,07 | 1,65 | 9,92E-02 |
| POPDC2 | 86,33 | 0,09 | 0,06 | 1,65 | 9,92E-02 |

| | | | | | |
|---|---|---|---|---|---|
| RAB5C | 711,97 | 0,08 | 0,05 | 1,65 | 9,93E-02 |
| B3GNT2 | 862,81 | -0,08 | 0,05 | -1,65 | 9,93E-02 |
| SH3YL1 | 1588,83 | -0,08 | 0,05 | -1,65 | 9,94E-02 |
| CXCR4 | 6405,59 | -0,08 | 0,05 | -1,65 | 9,95E-02 |
| RSPRY1 | 1279,35 | -0,04 | 0,02 | -1,65 | 9,95E-02 |
| HIP1R | 905,62 | 0,09 | 0,06 | 1,65 | 9,96E-02 |
| TUFM | 1468,79 | -0,07 | 0,04 | -1,65 | 9,96E-02 |
| ETV6 | 1307,12 | 0,13 | 0,08 | 1,65 | 9,98E-02 |
| PAPSS2 | 146,49 | 0,13 | 0,08 | 1,65 | 9,98E-02 |
| NFX1 | 1370,09 | -0,05 | 0,03 | -1,65 | 9,99E-02 |
| TLR1 | 11524,48 | -0,11 | 0,07 | -1,65 | 9,99E-02 |
| PCID2 | 1487,82 | -0,05 | 0,03 | -1,65 | 9,99E-02 |
| RCAN1 | 327,46 | -0,05 | 0,03 | -1,64 | 1,00E-01 |
| G0S2 | 43,54 | 0,21 | 0,13 | 1,64 | 1,00E-01 |
| TRAM2 | 524,24 | 0,09 | 0,05 | 1,64 | 1,00E-01 |
| STUB1 | 538,87 | 0,05 | 0,03 | 1,64 | 1,00E-01 |
| NDUFB6 | 611,61 | -0,09 | 0,06 | -1,64 | 1,00E-01 |
| NUDT13 | 86,84 | 0,09 | 0,05 | 1,64 | 1,00E-01 |
| BPNT1 | 442,88 | -0,07 | 0,04 | -1,64 | 1,00E-01 |
| ARL5B | 1237,32 | -0,11 | 0,07 | -1,64 | 1,01E-01 |
| GCLC | 1276,61 | 0,09 | 0,05 | 1,64 | 1,01E-01 |
| SNX30 | 2219,65 | -0,06 | 0,04 | -1,64 | 1,01E-01 |
| MZT2B | 1138,60 | 0,12 | 0,07 | 1,64 | 1,01E-01 |
| TSPYL1 | 2564,46 | -0,06 | 0,04 | -1,64 | 1,01E-01 |
| UBE2I | 1676,70 | 0,05 | 0,03 | 1,64 | 1,01E-01 |
| ZNF607 | 203,78 | -0,09 | 0,06 | -1,64 | 1,01E-01 |
| KRTCAP2 | 160,80 | 0,07 | 0,04 | 1,64 | 1,01E-01 |
| AMIGO1 | 311,17 | 0,10 | 0,06 | 1,64 | 1,01E-01 |
| KDM7A | 9331,69 | -0,07 | 0,04 | -1,64 | 1,01E-01 |
| RNPS1 | 1040,77 | 0,07 | 0,04 | 1,64 | 1,01E-01 |
| TBC1D32 | 614,25 | -0,09 | 0,06 | -1,64 | 1,01E-01 |
| GPA33 | 192,01 | 0,11 | 0,07 | 1,64 | 1,01E-01 |
| CD79B | 854,50 | -0,14 | 0,09 | -1,64 | 1,01E-01 |
| SOCS2 | 302,77 | 0,09 | 0,06 | 1,64 | 1,01E-01 |
| EXOC6 | 1667,54 | -0,09 | 0,06 | -1,64 | 1,01E-01 |
| SC5D | 394,93 | -0,10 | 0,06 | -1,64 | 1,01E-01 |
| GRHPR | 813,55 | 0,06 | 0,04 | 1,64 | 1,01E-01 |
| JKAMP | 1378,56 | -0,08 | 0,05 | -1,64 | 1,01E-01 |
| CHMP3 | 2247,87 | -0,06 | 0,03 | -1,64 | 1,01E-01 |
| RFC3 | 293,47 | -0,09 | 0,05 | -1,64 | 1,01E-01 |
| GALK2 | 1194,82 | -0,05 | 0,03 | -1,64 | 1,02E-01 |
| TRIM8 | 1849,94 | 0,12 | 0,07 | 1,64 | 1,02E-01 |
| ELOVL4 | 54,39 | 0,18 | 0,11 | 1,64 | 1,02E-01 |
| TMEM243 | 1281,72 | -0,07 | 0,04 | -1,64 | 1,02E-01 |
| GIMAP8 | 2061,04 | -0,07 | 0,04 | -1,64 | 1,02E-01 |
| ABITRAM | 201,20 | -0,05 | 0,03 | -1,64 | 1,02E-01 |

| | | | | | |
|---|---|---|---|---|---|
| NUP153 | 4273,97 | -0,05 | 0,03 | -1,64 | 1,02E-01 |
| SH3TC1 | 1494,59 | 0,08 | 0,05 | 1,63 | 1,02E-01 |
| SLAIN2 | 1781,63 | -0,05 | 0,03 | -1,63 | 1,02E-01 |
| PHC3 | 8531,76 | -0,05 | 0,03 | -1,63 | 1,02E-01 |
| DCUN1D4 | 867,18 | -0,08 | 0,05 | -1,63 | 1,03E-01 |
| CPNE3 | 2833,92 | -0,06 | 0,04 | -1,63 | 1,03E-01 |
| ZNF275 | 552,23 | 0,09 | 0,05 | 1,63 | 1,03E-01 |
| PXYLP1 | 507,60 | 0,09 | 0,05 | 1,63 | 1,03E-01 |
| TBL1XR1 | 8364,89 | -0,08 | 0,05 | -1,63 | 1,03E-01 |
| UFC1 | 633,78 | -0,09 | 0,06 | -1,63 | 1,03E-01 |
| DZIP3 | 691,53 | -0,12 | 0,07 | -1,63 | 1,03E-01 |
| DDX56 | 888,83 | 0,05 | 0,03 | 1,63 | 1,03E-01 |
| EIF6 | 824,75 | -0,07 | 0,04 | -1,63 | 1,03E-01 |
| NDUFA10 | 2243,07 | -0,04 | 0,03 | -1,63 | 1,03E-01 |
| PHKB | 3099,34 | -0,05 | 0,03 | -1,63 | 1,03E-01 |
| KBTBD11 | 1064,15 | -0,08 | 0,05 | -1,63 | 1,03E-01 |
| SMDT1 | 200,91 | 0,07 | 0,04 | 1,63 | 1,03E-01 |
| FFAR2 | 3044,05 | 0,14 | 0,08 | 1,63 | 1,04E-01 |
| LAS1L | 742,02 | -0,05 | 0,03 | -1,63 | 1,04E-01 |
| THEMIS | 2104,63 | -0,12 | 0,07 | -1,63 | 1,04E-01 |
| GPR18 | 200,46 | -0,12 | 0,07 | -1,63 | 1,04E-01 |
| SPIN2B | 87,11 | 0,07 | 0,04 | 1,63 | 1,04E-01 |
| ERO1B | 1249,23 | -0,07 | 0,04 | -1,63 | 1,04E-01 |
| PSMB4 | 2440,39 | -0,07 | 0,04 | -1,63 | 1,04E-01 |
| ETS1 | 11595,24 | -0,07 | 0,04 | -1,63 | 1,04E-01 |
| UGGT1 | 5025,58 | -0,05 | 0,03 | -1,63 | 1,04E-01 |
| LIN37 | 181,35 | 0,06 | 0,04 | 1,62 | 1,04E-01 |
| AMY2B | 696,64 | -0,07 | 0,04 | -1,62 | 1,04E-01 |
| ZNF513 | 323,68 | 0,05 | 0,03 | 1,62 | 1,04E-01 |
| ARL5A | 1906,87 | -0,09 | 0,06 | -1,62 | 1,05E-01 |
| WARS2 | 268,76 | -0,06 | 0,04 | -1,62 | 1,05E-01 |
| FAM193A | 1078,92 | 0,11 | 0,07 | 1,62 | 1,05E-01 |
| XPC | 2952,53 | -0,06 | 0,03 | -1,62 | 1,05E-01 |
| CHRNE | 77,33 | 0,17 | 0,10 | 1,62 | 1,05E-01 |
| MAGOHB | 446,74 | -0,09 | 0,05 | -1,62 | 1,05E-01 |
| GPR63 | 45,24 | 0,20 | 0,12 | 1,62 | 1,05E-01 |
| CCR2 | 3776,59 | -0,10 | 0,06 | -1,62 | 1,05E-01 |
| FREY1 | 24,31 | 0,11 | 0,07 | 1,62 | 1,05E-01 |
| PRKDC | 7247,37 | -0,07 | 0,04 | -1,62 | 1,05E-01 |
| CLC | 2012,41 | 0,23 | 0,14 | 1,62 | 1,05E-01 |
| CALHM6 | 279,08 | 0,19 | 0,12 | 1,62 | 1,05E-01 |
| QKI | 10992,83 | -0,09 | 0,06 | -1,62 | 1,05E-01 |
| PCDHGB2 | 33,04 | 0,17 | 0,10 | 1,62 | 1,05E-01 |
| POLR1A | 938,03 | 0,08 | 0,05 | 1,62 | 1,06E-01 |
| LEAP2 | 88,11 | 0,07 | 0,04 | 1,62 | 1,06E-01 |
| STK40 | 2311,41 | 0,13 | 0,08 | 1,62 | 1,06E-01 |

| | | | | | |
|---|---|---|---|---|---|
| STAC3 | 126,03 | 0,10 | 0,06 | 1,62 | 1,06E-01 |
| ANKRD34B | 70,44 | 0,24 | 0,15 | 1,62 | 1,06E-01 |
| TIMM8A | 122,01 | -0,07 | 0,04 | -1,62 | 1,06E-01 |
| TRMT1L | 770,00 | -0,08 | 0,05 | -1,62 | 1,06E-01 |
| EWSR1 | 6772,00 | -0,04 | 0,02 | -1,62 | 1,06E-01 |
| LAMA2 | 124,36 | 0,17 | 0,11 | 1,62 | 1,06E-01 |
| ZNF688 | 348,55 | 0,06 | 0,04 | 1,62 | 1,06E-01 |
| FILIP1L | 36,32 | 0,29 | 0,18 | 1,61 | 1,06E-01 |
| ZNF433 | 34,94 | 0,16 | 0,10 | 1,61 | 1,06E-01 |
| MRPL43 | 440,16 | 0,04 | 0,03 | 1,61 | 1,06E-01 |
| TTC33 | 545,13 | -0,13 | 0,08 | -1,61 | 1,06E-01 |
| FAM162A | 274,50 | -0,07 | 0,04 | -1,61 | 1,06E-01 |
| MPC1 | 640,63 | -0,07 | 0,04 | -1,61 | 1,06E-01 |
| LMNB2 | 404,40 | 0,07 | 0,04 | 1,61 | 1,07E-01 |
| ZSCAN22 | 145,17 | 0,06 | 0,04 | 1,61 | 1,07E-01 |
| TTC32 | 723,40 | -0,10 | 0,06 | -1,61 | 1,07E-01 |
| FUBP3 | 1338,23 | -0,04 | 0,02 | -1,61 | 1,07E-01 |
| LIG3 | 914,18 | 0,06 | 0,04 | 1,61 | 1,07E-01 |
| CEACAM3 | 1052,14 | 0,15 | 0,09 | 1,61 | 1,07E-01 |
| MEPCE | 768,63 | 0,06 | 0,04 | 1,61 | 1,07E-01 |
| ETNK1 | 4130,55 | -0,11 | 0,07 | -1,61 | 1,07E-01 |
| HERC1 | 14994,67 | -0,06 | 0,04 | -1,61 | 1,07E-01 |
| CHIC1 | 704,34 | -0,11 | 0,07 | -1,61 | 1,07E-01 |
| ACTR1A | 1524,47 | 0,06 | 0,04 | 1,61 | 1,07E-01 |
| DGLUCY | 5266,86 | 0,10 | 0,06 | 1,61 | 1,07E-01 |
| TNFRSF1A | 2876,02 | 0,13 | 0,08 | 1,61 | 1,07E-01 |
| SLC30A6 | 1277,46 | -0,06 | 0,04 | -1,61 | 1,07E-01 |
| LAMP2 | 9460,71 | -0,09 | 0,06 | -1,61 | 1,07E-01 |
| MAFK | 1243,50 | 0,06 | 0,04 | 1,61 | 1,07E-01 |
| EHMT1 | 2731,80 | 0,09 | 0,06 | 1,61 | 1,07E-01 |
| PDGFC | 182,14 | 0,11 | 0,07 | 1,61 | 1,07E-01 |
| LRRC41 | 665,56 | 0,05 | 0,03 | 1,61 | 1,07E-01 |
| AQP9 | 15696,29 | -0,12 | 0,07 | -1,61 | 1,07E-01 |
| CEP192 | 4093,79 | -0,05 | 0,03 | -1,61 | 1,07E-01 |
| TPP1 | 1050,43 | 0,11 | 0,07 | 1,61 | 1,07E-01 |
| SSBP2 | 1993,70 | -0,07 | 0,04 | -1,61 | 1,07E-01 |
| FZD2 | 79,87 | 0,13 | 0,08 | 1,61 | 1,07E-01 |
| AGO2 | 4834,43 | 0,11 | 0,07 | 1,61 | 1,08E-01 |
| TMEM128 | 387,51 | -0,07 | 0,05 | -1,61 | 1,08E-01 |
| NAGLU | 245,42 | 0,08 | 0,05 | 1,61 | 1,08E-01 |
| PPP1R3B | 4919,02 | -0,11 | 0,07 | -1,61 | 1,08E-01 |
| SLC39A10 | 2571,73 | -0,09 | 0,06 | -1,61 | 1,08E-01 |
| BTBD3 | 197,50 | 0,08 | 0,05 | 1,61 | 1,08E-01 |
| GCAT | 135,95 | 0,23 | 0,14 | 1,61 | 1,08E-01 |
| SUSD3 | 426,02 | 0,08 | 0,05 | 1,61 | 1,08E-01 |
| ANKRD20A4P | 36,21 | 0,36 | 0,22 | 1,61 | 1,08E-01 |

| | | | | | |
|---|---|---|---|---|---|
| FARSB | 581,92 | -0,06 | 0,04 | -1,61 | 1,08E-01 |
| MTFR2 | 39,90 | 0,12 | 0,08 | 1,61 | 1,08E-01 |
| GATC | 621,59 | -0,05 | 0,03 | -1,61 | 1,08E-01 |
| FAM153A | 363,46 | 0,17 | 0,11 | 1,61 | 1,08E-01 |
| SAT1 | 10266,62 | -0,11 | 0,07 | -1,61 | 1,08E-01 |
| SLC25A19 | 180,73 | 0,07 | 0,04 | 1,61 | 1,08E-01 |
| WLS | 1952,06 | -0,18 | 0,11 | -1,61 | 1,08E-01 |
| CTDSP2 | 4602,67 | 0,12 | 0,07 | 1,61 | 1,08E-01 |
| BCL7A | 121,20 | 0,14 | 0,09 | 1,61 | 1,08E-01 |
| NAALADL1 | 154,48 | 0,08 | 0,05 | 1,61 | 1,08E-01 |
| PPP1R3F | 463,77 | 0,06 | 0,04 | 1,60 | 1,09E-01 |
| ASB8 | 1385,99 | -0,04 | 0,03 | -1,60 | 1,09E-01 |
| HMGXB3 | 1133,28 | 0,05 | 0,03 | 1,60 | 1,09E-01 |
| SMARCA2 | 7857,92 | -0,05 | 0,03 | -1,60 | 1,09E-01 |
| WDR27 | 887,48 | 0,09 | 0,06 | 1,60 | 1,09E-01 |
| ZNF501 | 134,32 | -0,13 | 0,08 | -1,60 | 1,09E-01 |
| NSD1 | 6627,43 | -0,05 | 0,03 | -1,60 | 1,09E-01 |
| EML6 | 198,37 | 0,18 | 0,11 | 1,60 | 1,09E-01 |
| PRMT3 | 814,83 | -0,06 | 0,04 | -1,60 | 1,09E-01 |
| HSPA1A | 1494,49 | -0,12 | 0,07 | -1,60 | 1,09E-01 |
| FAM229B | 31,94 | 0,18 | 0,11 | 1,60 | 1,09E-01 |
| AZI2 | 923,78 | -0,12 | 0,08 | -1,60 | 1,09E-01 |
| HAX1 | 927,59 | -0,06 | 0,04 | -1,60 | 1,09E-01 |
| KDM4B | 3095,97 | 0,14 | 0,08 | 1,60 | 1,10E-01 |
| ATP5F1C | 1086,18 | -0,10 | 0,06 | -1,60 | 1,10E-01 |
| SVBP | 375,76 | 0,12 | 0,07 | 1,60 | 1,10E-01 |
| AOC2 | 220,14 | 0,16 | 0,10 | 1,60 | 1,10E-01 |
| SRBD1 | 1538,25 | -0,08 | 0,05 | -1,60 | 1,10E-01 |
| CENPB | 1442,17 | -0,07 | 0,04 | -1,60 | 1,10E-01 |
| TMEM50B | 1166,60 | -0,08 | 0,05 | -1,60 | 1,10E-01 |
| TMPO | 5722,19 | -0,08 | 0,05 | -1,60 | 1,10E-01 |
| IL16 | 14051,10 | -0,06 | 0,04 | -1,60 | 1,10E-01 |
| TST | 450,62 | 0,08 | 0,05 | 1,60 | 1,10E-01 |
| ZC3H7A | 1778,42 | -0,04 | 0,03 | -1,60 | 1,10E-01 |
| FLACC1 | 343,11 | 0,12 | 0,07 | 1,60 | 1,10E-01 |
| BCKDHB | 393,13 | -0,07 | 0,05 | -1,60 | 1,11E-01 |
| UNC93B1 | 1951,37 | 0,10 | 0,06 | 1,60 | 1,11E-01 |
| FAM111A | 3806,95 | -0,06 | 0,04 | -1,60 | 1,11E-01 |
| RNF144A | 1270,06 | 0,10 | 0,06 | 1,60 | 1,11E-01 |
| SUSD6 | 2284,78 | 0,14 | 0,09 | 1,60 | 1,11E-01 |
| HDHD3 | 156,27 | 0,09 | 0,05 | 1,59 | 1,11E-01 |
| DDX17 | 51393,38 | -0,05 | 0,03 | -1,59 | 1,11E-01 |
| SLC25A36 | 1869,62 | -0,11 | 0,07 | -1,59 | 1,11E-01 |
| CCDC6 | 1143,01 | -0,05 | 0,03 | -1,59 | 1,11E-01 |
| ACOX1 | 6066,61 | -0,10 | 0,06 | -1,59 | 1,11E-01 |
| LIMD1 | 1607,70 | 0,08 | 0,05 | 1,59 | 1,11E-01 |

| | | | | | |
|---|---|---|---|---|---|
| BLTP1 | 15164,38 | -0,08 | 0,05 | -1,59 | 1,11E-01 |
| MARK2 | 1325,47 | 0,14 | 0,09 | 1,59 | 1,11E-01 |
| ZSCAN29 | 979,94 | -0,04 | 0,03 | -1,59 | 1,11E-01 |
| TIMELESS | 216,40 | 0,09 | 0,06 | 1,59 | 1,11E-01 |
| MKRN1 | 28311,98 | 0,16 | 0,10 | 1,59 | 1,11E-01 |
| MKKS | 469,57 | -0,07 | 0,04 | -1,59 | 1,11E-01 |
| RAPH1 | 227,34 | 0,12 | 0,07 | 1,59 | 1,11E-01 |
| PTGDR | 1152,85 | -0,13 | 0,08 | -1,59 | 1,11E-01 |
| MCCC2 | 686,00 | -0,05 | 0,03 | -1,59 | 1,11E-01 |
| KLHL32 | 78,77 | 0,12 | 0,07 | 1,59 | 1,11E-01 |
| NDRG3 | 730,89 | -0,05 | 0,03 | -1,59 | 1,12E-01 |
| GPAT3 | 1460,41 | -0,11 | 0,07 | -1,59 | 1,12E-01 |
| WAC | 7147,50 | -0,05 | 0,03 | -1,59 | 1,12E-01 |
| PRKACA | 568,50 | 0,16 | 0,10 | 1,59 | 1,12E-01 |
| CLCC1 | 960,78 | -0,05 | 0,03 | -1,59 | 1,12E-01 |
| FNBP4 | 3856,24 | -0,05 | 0,03 | -1,59 | 1,12E-01 |
| SEMA4C | 517,73 | 0,08 | 0,05 | 1,59 | 1,12E-01 |
| CRKL | 4019,46 | -0,05 | 0,03 | -1,59 | 1,12E-01 |
| TMBIM1 | 3786,21 | 0,06 | 0,04 | 1,59 | 1,12E-01 |
| PLPBP | 878,31 | -0,04 | 0,03 | -1,59 | 1,12E-01 |
| LPCAT3 | 525,54 | 0,08 | 0,05 | 1,59 | 1,12E-01 |
| ALDH3A2 | 1107,05 | -0,04 | 0,02 | -1,59 | 1,12E-01 |
| SPATA1 | 64,09 | 0,13 | 0,08 | 1,59 | 1,12E-01 |
| SIT1 | 936,87 | -0,09 | 0,06 | -1,59 | 1,12E-01 |
| RPP30 | 548,51 | -0,06 | 0,04 | -1,59 | 1,12E-01 |
| NSMCE3 | 476,62 | -0,06 | 0,04 | -1,59 | 1,13E-01 |
| TNFSF4 | 330,02 | 0,10 | 0,07 | 1,59 | 1,13E-01 |
| NAB1 | 2035,46 | -0,08 | 0,05 | -1,59 | 1,13E-01 |
| IFT172 | 384,11 | 0,08 | 0,05 | 1,59 | 1,13E-01 |
| WWC3 | 2079,67 | 0,11 | 0,07 | 1,59 | 1,13E-01 |
| DHX34 | 1956,39 | 0,12 | 0,07 | 1,59 | 1,13E-01 |
| ZSWIM8 | 1802,66 | 0,12 | 0,07 | 1,59 | 1,13E-01 |
| SHPK | 33,34 | 0,10 | 0,06 | 1,59 | 1,13E-01 |
| RAB34 | 185,57 | 0,09 | 0,05 | 1,59 | 1,13E-01 |
| CAVIN3 | 26,88 | 0,25 | 0,16 | 1,59 | 1,13E-01 |
| MSR1 | 203,04 | -0,20 | 0,13 | -1,58 | 1,13E-01 |
| ZNF773 | 321,09 | -0,07 | 0,04 | -1,58 | 1,13E-01 |
| ING2 | 123,24 | -0,09 | 0,06 | -1,58 | 1,13E-01 |
| BCR | 1572,72 | 0,10 | 0,06 | 1,58 | 1,13E-01 |
| EGLN1 | 6251,11 | -0,09 | 0,06 | -1,58 | 1,13E-01 |
| ZNF763 | 174,78 | -0,07 | 0,05 | -1,58 | 1,13E-01 |
| LMBRD2 | 1624,01 | -0,09 | 0,06 | -1,58 | 1,13E-01 |
| PHF13 | 245,56 | 0,07 | 0,04 | 1,58 | 1,13E-01 |
| COX7C | 1032,88 | -0,18 | 0,11 | -1,58 | 1,14E-01 |
| NRDC | 6268,09 | -0,07 | 0,04 | -1,58 | 1,14E-01 |
| RNF217 | 409,52 | 0,10 | 0,06 | 1,58 | 1,14E-01 |

| | | | | | |
|---|---|---|---|---|---|
| RANBP1 | 545,37 | -0,06 | 0,04 | -1,58 | 1,14E-01 |
| MEA1 | 423,96 | -0,05 | 0,03 | -1,58 | 1,14E-01 |
| GALNT3 | 1330,32 | -0,09 | 0,06 | -1,58 | 1,14E-01 |
| CAMKK2 | 3002,44 | 0,10 | 0,06 | 1,58 | 1,14E-01 |
| YIPF4 | 6021,75 | -0,06 | 0,04 | -1,58 | 1,14E-01 |
| VDR | 348,14 | 0,13 | 0,08 | 1,58 | 1,14E-01 |
| PPP1R3E | 673,06 | 0,06 | 0,04 | 1,58 | 1,14E-01 |
| KHDRBS1 | 1779,25 | -0,05 | 0,03 | -1,58 | 1,14E-01 |
| NT5C3A | 1902,00 | -0,11 | 0,07 | -1,58 | 1,14E-01 |
| BRD9 | 1672,04 | 0,04 | 0,03 | 1,58 | 1,14E-01 |
| RASSF7 | 292,09 | 0,07 | 0,04 | 1,58 | 1,14E-01 |
| GYG1 | 810,78 | -0,07 | 0,05 | -1,58 | 1,14E-01 |
| CNPY4 | 396,34 | -0,06 | 0,04 | -1,58 | 1,14E-01 |
| CSPP1 | 658,17 | -0,06 | 0,04 | -1,58 | 1,14E-01 |
| TMEM267 | 371,22 | -0,08 | 0,05 | -1,58 | 1,15E-01 |
| ARRDC2 | 1572,56 | 0,06 | 0,04 | 1,58 | 1,15E-01 |
| PPTC7 | 2980,80 | -0,06 | 0,04 | -1,58 | 1,15E-01 |
| DLGAP5 | 58,10 | 0,22 | 0,14 | 1,58 | 1,15E-01 |
| PLEKHJ1 | 764,52 | 0,05 | 0,03 | 1,58 | 1,15E-01 |
| EBP | 309,41 | -0,07 | 0,04 | -1,58 | 1,15E-01 |
| BRI3 | 1232,25 | 0,08 | 0,05 | 1,58 | 1,15E-01 |
| OTUD1 | 713,28 | -0,08 | 0,05 | -1,58 | 1,15E-01 |
| HPRT1 | 121,08 | -0,10 | 0,06 | -1,58 | 1,15E-01 |
| CKAP5 | 1231,65 | -0,05 | 0,03 | -1,58 | 1,15E-01 |
| BEX2 | 88,87 | -0,15 | 0,09 | -1,57 | 1,15E-01 |
| PTPRA | 1602,54 | 0,07 | 0,04 | 1,57 | 1,15E-01 |
| NUDCD2 | 406,32 | -0,08 | 0,05 | -1,57 | 1,15E-01 |
| THOC5 | 1805,12 | 0,09 | 0,06 | 1,57 | 1,16E-01 |
| CRIP1 | 260,41 | 0,08 | 0,05 | 1,57 | 1,16E-01 |
| PDE4D | 1487,25 | -0,08 | 0,05 | -1,57 | 1,16E-01 |
| ARAP1 | 7825,28 | 0,15 | 0,10 | 1,57 | 1,16E-01 |
| PGAM5 | 349,36 | 0,06 | 0,04 | 1,57 | 1,16E-01 |
| CD81 | 757,60 | 0,10 | 0,06 | 1,57 | 1,16E-01 |
| OTUD3 | 784,12 | -0,06 | 0,04 | -1,57 | 1,16E-01 |
| STAMBPL1 | 1158,06 | -0,09 | 0,05 | -1,57 | 1,16E-01 |
| NKAPL | 51,31 | -0,14 | 0,09 | -1,57 | 1,16E-01 |
| GNB2 | 3261,78 | 0,11 | 0,07 | 1,57 | 1,16E-01 |
| INPP5F | 727,09 | -0,10 | 0,06 | -1,57 | 1,16E-01 |
| APLP2 | 6883,11 | -0,08 | 0,05 | -1,57 | 1,16E-01 |
| ADARB2 | 126,66 | 0,45 | 0,29 | 1,57 | 1,16E-01 |
| UVSSA | 2062,59 | 0,11 | 0,07 | 1,57 | 1,16E-01 |
| UTP20 | 1004,06 | -0,06 | 0,04 | -1,57 | 1,16E-01 |
| DNAJC17 | 508,04 | 0,05 | 0,03 | 1,57 | 1,16E-01 |
| SLC66A3 | 876,70 | -0,07 | 0,04 | -1,57 | 1,17E-01 |
| ADAM23 | 240,43 | 0,19 | 0,12 | 1,57 | 1,17E-01 |
| STIMATE-MUSTN1 | 32,34 | 0,15 | 0,10 | 1,57 | 1,17E-01 |

| | | | | | |
|---|---|---|---|---|---|
| TXNL4B | 609,83 | -0,05 | 0,03 | -1,57 | 1,17E-01 |
| TTL | 775,45 | 0,06 | 0,04 | 1,57 | 1,17E-01 |
| TMEM170B | 3444,65 | -0,12 | 0,08 | -1,57 | 1,17E-01 |
| SPINT2 | 1027,12 | 0,06 | 0,04 | 1,57 | 1,17E-01 |
| ATP6V1A | 5566,34 | -0,08 | 0,05 | -1,57 | 1,17E-01 |
| NDUFS6 | 471,97 | -0,06 | 0,04 | -1,57 | 1,17E-01 |
| MAN2A1 | 4711,47 | -0,06 | 0,04 | -1,57 | 1,17E-01 |
| SFXN5 | 1004,18 | 0,09 | 0,06 | 1,57 | 1,17E-01 |
| CDRT4 | 79,43 | 0,16 | 0,10 | 1,57 | 1,17E-01 |
| INTS15 | 698,32 | 0,05 | 0,03 | 1,57 | 1,17E-01 |
| N6AMT1 | 503,52 | -0,08 | 0,05 | -1,57 | 1,17E-01 |
| ZNF276 | 3965,10 | 0,06 | 0,04 | 1,56 | 1,18E-01 |
| CDKN2D | 2437,60 | 0,10 | 0,06 | 1,56 | 1,18E-01 |
| TLR10 | 708,78 | -0,09 | 0,06 | -1,56 | 1,18E-01 |
| IRAG1 | 1812,17 | -0,12 | 0,08 | -1,56 | 1,18E-01 |
| C1orf50 | 298,71 | 0,06 | 0,04 | 1,56 | 1,18E-01 |
| PTPN7 | 566,78 | 0,10 | 0,07 | 1,56 | 1,18E-01 |
| ATP7A | 2576,66 | -0,06 | 0,04 | -1,56 | 1,18E-01 |
| TRPS1 | 2521,20 | -0,06 | 0,04 | -1,56 | 1,18E-01 |
| LUC7L2 | 1569,65 | -0,06 | 0,04 | -1,56 | 1,18E-01 |
| RHOA | 8761,24 | -0,06 | 0,04 | -1,56 | 1,18E-01 |
| LRRC58 | 2020,85 | -0,07 | 0,05 | -1,56 | 1,19E-01 |
| DENND3 | 13031,52 | 0,13 | 0,08 | 1,56 | 1,19E-01 |
| SLC7A6 | 2086,83 | 0,08 | 0,05 | 1,56 | 1,19E-01 |
| TCEA1 | 2199,90 | -0,08 | 0,05 | -1,56 | 1,19E-01 |
| P2RY11 | 151,87 | 0,09 | 0,06 | 1,56 | 1,19E-01 |
| PPP4R4 | 69,59 | 0,22 | 0,14 | 1,56 | 1,19E-01 |
| NKTR | 13684,37 | -0,07 | 0,04 | -1,56 | 1,19E-01 |
| DDIT4 | 1210,46 | -0,13 | 0,08 | -1,56 | 1,19E-01 |
| PPP2R1A | 1634,35 | 0,06 | 0,04 | 1,56 | 1,19E-01 |
| BORCS5 | 589,28 | 0,09 | 0,05 | 1,56 | 1,19E-01 |
| ABHD4 | 643,18 | 0,08 | 0,05 | 1,56 | 1,19E-01 |
| ZDHHC3 | 1673,08 | 0,05 | 0,03 | 1,56 | 1,19E-01 |
| ECE1 | 3020,30 | 0,13 | 0,08 | 1,56 | 1,19E-01 |
| CYB5A | 382,24 | 0,07 | 0,05 | 1,56 | 1,19E-01 |
| RBBP6 | 2754,56 | -0,05 | 0,03 | -1,56 | 1,19E-01 |
| SDHB | 1211,23 | -0,06 | 0,04 | -1,56 | 1,20E-01 |
| DNM2 | 4352,45 | 0,10 | 0,06 | 1,56 | 1,20E-01 |
| TIMM22 | 376,96 | -0,04 | 0,03 | -1,56 | 1,20E-01 |
| TAS2R20 | 183,16 | -0,13 | 0,08 | -1,56 | 1,20E-01 |
| RAD52 | 331,49 | 0,06 | 0,04 | 1,56 | 1,20E-01 |
| DMAC1 | 567,38 | -0,11 | 0,07 | -1,55 | 1,20E-01 |
| SP140 | 2939,37 | -0,09 | 0,06 | -1,55 | 1,20E-01 |
| KLF2 | 3395,13 | 0,08 | 0,05 | 1,55 | 1,20E-01 |
| ZNF839 | 486,30 | 0,06 | 0,04 | 1,55 | 1,20E-01 |
| JARID2 | 4082,89 | -0,08 | 0,05 | -1,55 | 1,20E-01 |

| | | | | | |
|---|---|---|---|---|---|
| ZNF19 | 124,12 | 0,12 | 0,08 | 1,55 | 1,20E-01 |
| TCEAL1 | 80,10 | -0,11 | 0,07 | -1,55 | 1,20E-01 |
| EIF4G1 | 2185,29 | 0,09 | 0,06 | 1,55 | 1,20E-01 |
| ENSG00000215271 | 395,83 | 0,04 | 0,02 | 1,55 | 1,20E-01 |
| ARHGDIA | 617,28 | 0,13 | 0,08 | 1,55 | 1,20E-01 |
| CCN3 | 355,14 | -0,15 | 0,10 | -1,55 | 1,20E-01 |
| MCPH1 | 1100,20 | -0,05 | 0,03 | -1,55 | 1,21E-01 |
| COX4I1 | 3168,82 | -0,08 | 0,05 | -1,55 | 1,21E-01 |
| IFNAR1 | 5323,63 | -0,07 | 0,04 | -1,55 | 1,21E-01 |
| AK9 | 507,21 | -0,07 | 0,05 | -1,55 | 1,21E-01 |
| FLOT2 | 2600,40 | 0,13 | 0,09 | 1,55 | 1,21E-01 |
| ANXA2 | 5369,56 | -0,07 | 0,04 | -1,55 | 1,21E-01 |
| POLR3G | 43,96 | 0,14 | 0,09 | 1,55 | 1,21E-01 |
| SH2D1B | 491,64 | -0,14 | 0,09 | -1,55 | 1,21E-01 |
| ZNG1E | 1086,81 | -0,09 | 0,06 | -1,55 | 1,21E-01 |
| LCK | 4067,72 | -0,08 | 0,05 | -1,55 | 1,21E-01 |
| IL12RB2 | 171,20 | 0,15 | 0,09 | 1,55 | 1,21E-01 |
| MMS19 | 1123,28 | 0,05 | 0,03 | 1,55 | 1,22E-01 |
| DDB1 | 4025,48 | 0,07 | 0,05 | 1,55 | 1,22E-01 |
| MMP25 | 4243,21 | 0,18 | 0,12 | 1,55 | 1,22E-01 |
| EIF2D | 802,47 | 0,04 | 0,03 | 1,55 | 1,22E-01 |
| ABHD17C | 56,97 | 0,17 | 0,11 | 1,55 | 1,22E-01 |
| ERBB2 | 223,73 | 0,16 | 0,10 | 1,55 | 1,22E-01 |
| RFX7 | 1156,83 | -0,07 | 0,05 | -1,55 | 1,22E-01 |
| SOX5 | 73,58 | 0,24 | 0,15 | 1,55 | 1,22E-01 |
| TXNL1 | 975,59 | -0,05 | 0,03 | -1,55 | 1,22E-01 |
| BAG1 | 6436,21 | 0,15 | 0,10 | 1,54 | 1,22E-01 |
| FHIP1B | 835,42 | 0,10 | 0,07 | 1,54 | 1,22E-01 |
| EIF3F | 2741,22 | -0,06 | 0,04 | -1,54 | 1,22E-01 |
| SUGP2 | 2119,78 | 0,07 | 0,04 | 1,54 | 1,22E-01 |
| NFKBIB | 309,51 | 0,06 | 0,04 | 1,54 | 1,23E-01 |
| OTUD4 | 2161,44 | -0,05 | 0,03 | -1,54 | 1,23E-01 |
| GTDC1 | 1488,20 | -0,06 | 0,04 | -1,54 | 1,23E-01 |
| FBXO6 | 297,12 | 0,13 | 0,08 | 1,54 | 1,23E-01 |
| PNPO | 285,50 | 0,05 | 0,03 | 1,54 | 1,23E-01 |
| ARL2BP | 1018,83 | -0,06 | 0,04 | -1,54 | 1,23E-01 |
| CRK | 2506,89 | -0,06 | 0,04 | -1,54 | 1,23E-01 |
| PALB2 | 317,21 | -0,05 | 0,03 | -1,54 | 1,23E-01 |
| GLYCTK | 401,80 | 0,09 | 0,06 | 1,54 | 1,23E-01 |
| RTL6 | 374,42 | 0,10 | 0,06 | 1,54 | 1,23E-01 |
| IFIT2 | 8023,48 | -0,20 | 0,13 | -1,54 | 1,23E-01 |
| GCNT4 | 729,75 | -0,12 | 0,08 | -1,54 | 1,23E-01 |
| COPS7A | 602,57 | 0,05 | 0,03 | 1,54 | 1,24E-01 |
| MAGEH1 | 207,09 | -0,09 | 0,06 | -1,54 | 1,24E-01 |
| ACAT1 | 329,02 | -0,10 | 0,06 | -1,54 | 1,24E-01 |
| NIBAN2 | 581,62 | 0,09 | 0,06 | 1,54 | 1,24E-01 |

| | | | | | |
|---|---|---|---|---|---|
| PLXNA2 | 447,92 | 0,14 | 0,09 | 1,54 | 1,24E-01 |
| SYNGR2 | 1230,69 | 0,06 | 0,04 | 1,54 | 1,24E-01 |
| FRMD3 | 313,93 | 0,13 | 0,08 | 1,54 | 1,24E-01 |
| USP49 | 465,50 | 0,06 | 0,04 | 1,54 | 1,24E-01 |
| SLC39A11 | 648,61 | 0,07 | 0,05 | 1,54 | 1,24E-01 |
| TBCC | 1604,35 | -0,06 | 0,04 | -1,54 | 1,25E-01 |
| ING5 | 1131,36 | 0,05 | 0,03 | 1,54 | 1,25E-01 |
| NCOA3 | 5388,88 | -0,06 | 0,04 | -1,54 | 1,25E-01 |
| CD58 | 1513,20 | -0,07 | 0,05 | -1,54 | 1,25E-01 |
| USP45 | 566,83 | -0,09 | 0,06 | -1,54 | 1,25E-01 |
| SLC25A12 | 773,10 | -0,05 | 0,03 | -1,54 | 1,25E-01 |
| POMGNT1 | 417,07 | 0,06 | 0,04 | 1,53 | 1,25E-01 |
| LRCH1 | 983,11 | -0,05 | 0,03 | -1,53 | 1,25E-01 |
| OCIAD2 | 232,55 | -0,09 | 0,06 | -1,53 | 1,25E-01 |
| NUF2 | 53,86 | 0,16 | 0,11 | 1,53 | 1,25E-01 |
| CNOT6 | 1777,76 | -0,05 | 0,03 | -1,53 | 1,25E-01 |
| REM2 | 288,57 | 0,13 | 0,08 | 1,53 | 1,25E-01 |
| TTC5 | 1042,81 | -0,05 | 0,03 | -1,53 | 1,25E-01 |
| AGO3 | 3898,79 | -0,05 | 0,03 | -1,53 | 1,25E-01 |
| TMEM268 | 446,47 | 0,12 | 0,08 | 1,53 | 1,25E-01 |
| CIB1 | 1031,11 | 0,06 | 0,04 | 1,53 | 1,25E-01 |
| PDXK | 1877,54 | 0,09 | 0,06 | 1,53 | 1,25E-01 |
| ALDH1A1 | 770,60 | -0,12 | 0,08 | -1,53 | 1,25E-01 |
| GPR183 | 1360,89 | -0,13 | 0,09 | -1,53 | 1,25E-01 |
| DPH6 | 432,67 | -0,10 | 0,06 | -1,53 | 1,26E-01 |
| KLHL42 | 883,58 | -0,05 | 0,03 | -1,53 | 1,26E-01 |
| B4GALNT3 | 173,36 | 0,25 | 0,17 | 1,53 | 1,26E-01 |
| ATP1B1 | 96,84 | 0,14 | 0,09 | 1,53 | 1,26E-01 |
| ZNF345 | 322,78 | -0,06 | 0,04 | -1,53 | 1,26E-01 |
| TMED1 | 376,53 | 0,07 | 0,05 | 1,53 | 1,26E-01 |
| MYO9B | 5465,91 | 0,13 | 0,08 | 1,53 | 1,26E-01 |
| TNFAIP8L2 | 1611,57 | -0,06 | 0,04 | -1,53 | 1,26E-01 |
| C6orf120 | 910,34 | -0,08 | 0,05 | -1,53 | 1,26E-01 |
| MFSD2A | 113,94 | 0,11 | 0,07 | 1,53 | 1,26E-01 |
| UBA3 | 3220,08 | -0,08 | 0,05 | -1,53 | 1,26E-01 |
| DPP3 | 243,80 | 0,07 | 0,05 | 1,53 | 1,26E-01 |
| BBOF1 | 508,44 | 0,18 | 0,11 | 1,53 | 1,26E-01 |
| CD27 | 298,24 | 0,10 | 0,07 | 1,53 | 1,26E-01 |
| LAIR1 | 1669,57 | 0,06 | 0,04 | 1,53 | 1,26E-01 |
| CNTF | 103,56 | -0,06 | 0,04 | -1,53 | 1,26E-01 |
| CFAP92 | 417,64 | 0,14 | 0,09 | 1,53 | 1,26E-01 |
| SHKBP1 | 2126,30 | 0,14 | 0,09 | 1,53 | 1,26E-01 |
| LYPLAL1 | 967,58 | -0,11 | 0,07 | -1,53 | 1,27E-01 |
| G6PD | 695,94 | 0,13 | 0,09 | 1,53 | 1,27E-01 |
| TBRG1 | 2394,32 | -0,04 | 0,03 | -1,53 | 1,27E-01 |
| SUCNR1 | 53,19 | 0,16 | 0,10 | 1,53 | 1,27E-01 |

| | | | | | |
|---|---|---|---|---|---|
| ACER3 | 1086,44 | 0,06 | 0,04 | 1,53 | 1,27E-01 |
| NUAK1 | 70,82 | 0,27 | 0,18 | 1,53 | 1,27E-01 |
| ZHX3 | 292,63 | 0,10 | 0,06 | 1,53 | 1,27E-01 |
| NME8 | 382,16 | -0,12 | 0,08 | -1,53 | 1,27E-01 |
| PHTF2 | 1928,49 | -0,08 | 0,05 | -1,53 | 1,27E-01 |
| MEGF9 | 11189,33 | -0,10 | 0,07 | -1,53 | 1,27E-01 |
| MCFD2 | 955,08 | -0,03 | 0,02 | -1,53 | 1,27E-01 |
| PSD4 | 5332,15 | 0,10 | 0,06 | 1,53 | 1,27E-01 |
| HACE1 | 740,43 | -0,06 | 0,04 | -1,53 | 1,27E-01 |
| PRDX1 | 1805,42 | -0,10 | 0,06 | -1,53 | 1,27E-01 |
| LIMD2 | 6457,42 | 0,06 | 0,04 | 1,53 | 1,27E-01 |
| NMT2 | 689,64 | -0,07 | 0,04 | -1,53 | 1,27E-01 |
| FBXO9 | 2804,65 | 0,09 | 0,06 | 1,53 | 1,27E-01 |
| ENC1 | 912,66 | -0,16 | 0,10 | -1,53 | 1,27E-01 |
| LRP2BP | 58,36 | 0,15 | 0,10 | 1,53 | 1,27E-01 |
| MAP2K6 | 2424,76 | -0,07 | 0,05 | -1,53 | 1,27E-01 |
| YTHDF1 | 812,05 | 0,04 | 0,03 | 1,52 | 1,27E-01 |
| LIMA1 | 263,77 | -0,08 | 0,05 | -1,52 | 1,27E-01 |
| SLC46A2 | 258,87 | 0,10 | 0,06 | 1,52 | 1,27E-01 |
| OCRL | 247,79 | 0,07 | 0,05 | 1,52 | 1,27E-01 |
| SMG6 | 1067,51 | 0,09 | 0,06 | 1,52 | 1,27E-01 |
| FN3KRP | 342,26 | 0,06 | 0,04 | 1,52 | 1,28E-01 |
| SKP2 | 751,11 | -0,05 | 0,03 | -1,52 | 1,28E-01 |
| PTPN9 | 708,40 | 0,05 | 0,03 | 1,52 | 1,28E-01 |
| DNAL1 | 254,83 | -0,05 | 0,04 | -1,52 | 1,28E-01 |
| CD46 | 15238,70 | -0,08 | 0,05 | -1,52 | 1,28E-01 |
| DLAT | 376,54 | -0,05 | 0,03 | -1,52 | 1,28E-01 |
| BEX4 | 307,42 | -0,09 | 0,06 | -1,52 | 1,28E-01 |
| RPS18 | 13203,79 | -0,16 | 0,11 | -1,52 | 1,28E-01 |
| CD70 | 27,50 | 0,16 | 0,11 | 1,52 | 1,28E-01 |
| SIVA1 | 692,25 | 0,05 | 0,03 | 1,52 | 1,28E-01 |
| CIAO2A | 670,78 | -0,09 | 0,06 | -1,52 | 1,28E-01 |
| TRIM68 | 319,57 | -0,04 | 0,03 | -1,52 | 1,28E-01 |
| CTC1 | 2002,80 | 0,09 | 0,06 | 1,52 | 1,28E-01 |
| EIF3B | 1826,95 | 0,07 | 0,05 | 1,52 | 1,28E-01 |
| UBQLN2 | 821,72 | 0,06 | 0,04 | 1,52 | 1,28E-01 |
| HNRNPLL | 2091,19 | -0,08 | 0,05 | -1,52 | 1,29E-01 |
| HPSE | 923,94 | 0,10 | 0,07 | 1,52 | 1,29E-01 |
| STX16-NPEPL1 | 133,92 | 0,11 | 0,07 | 1,52 | 1,29E-01 |
| LONP1 | 870,96 | 0,05 | 0,04 | 1,52 | 1,29E-01 |
| EEF1D | 7096,10 | -0,08 | 0,05 | -1,52 | 1,29E-01 |
| FAM78A | 1787,27 | 0,07 | 0,05 | 1,52 | 1,29E-01 |
| MAP3K4 | 1803,39 | -0,06 | 0,04 | -1,52 | 1,29E-01 |
| RNF138 | 1205,13 | -0,10 | 0,07 | -1,52 | 1,29E-01 |
| SLC25A14 | 265,88 | -0,05 | 0,03 | -1,52 | 1,29E-01 |
| KLF5 | 107,61 | 0,13 | 0,08 | 1,52 | 1,29E-01 |

| | | | | | |
|---|---|---|---|---|---|
| UBR4 | 7306,59 | 0,09 | 0,06 | 1,52 | 1,29E-01 |
| FAM177B | 117,93 | -0,12 | 0,08 | -1,52 | 1,29E-01 |
| CCDC22 | 396,36 | 0,06 | 0,04 | 1,52 | 1,29E-01 |
| TCAF2 | 185,20 | 0,09 | 0,06 | 1,52 | 1,30E-01 |
| PDAP1 | 781,32 | 0,05 | 0,03 | 1,52 | 1,30E-01 |
| MYO5A | 5358,95 | -0,06 | 0,04 | -1,52 | 1,30E-01 |
| METTL2A | 346,77 | -0,05 | 0,03 | -1,52 | 1,30E-01 |
| WNK1 | 8741,58 | 0,10 | 0,07 | 1,51 | 1,30E-01 |
| CHTF8 | 1094,09 | 0,05 | 0,03 | 1,51 | 1,30E-01 |
| DDIT3 | 394,24 | -0,09 | 0,06 | -1,51 | 1,30E-01 |
| KBTBD3 | 221,78 | -0,10 | 0,06 | -1,51 | 1,30E-01 |
| ANKDD1B | 76,58 | 0,12 | 0,08 | 1,51 | 1,30E-01 |
| ITGB3 | 1453,77 | 0,22 | 0,15 | 1,51 | 1,30E-01 |
| CREBZF | 4105,70 | -0,08 | 0,05 | -1,51 | 1,30E-01 |
| PDE12 | 1172,20 | -0,04 | 0,02 | -1,51 | 1,31E-01 |
| CCNK | 1251,29 | -0,04 | 0,03 | -1,51 | 1,31E-01 |
| GTF2H5 | 232,26 | -0,09 | 0,06 | -1,51 | 1,31E-01 |
| TPT1 | 11240,99 | -0,16 | 0,11 | -1,51 | 1,31E-01 |
| TBC1D31 | 1121,68 | -0,06 | 0,04 | -1,51 | 1,31E-01 |
| MAML1 | 1645,37 | 0,10 | 0,07 | 1,51 | 1,31E-01 |
| LRRC39 | 26,04 | 0,14 | 0,09 | 1,51 | 1,31E-01 |
| AQP3 | 306,93 | 0,10 | 0,06 | 1,51 | 1,31E-01 |
| PFN1 | 3625,63 | 0,07 | 0,05 | 1,51 | 1,31E-01 |
| HNRNPUL2 | 671,87 | -0,04 | 0,03 | -1,51 | 1,32E-01 |
| KGD4 | 349,85 | -0,08 | 0,05 | -1,51 | 1,32E-01 |
| IL2RA | 337,62 | 0,09 | 0,06 | 1,51 | 1,32E-01 |
| TNFRSF10D | 326,01 | 0,06 | 0,04 | 1,51 | 1,32E-01 |
| KAT5 | 894,90 | -0,04 | 0,03 | -1,51 | 1,32E-01 |
| ADARB1 | 868,93 | 0,09 | 0,06 | 1,51 | 1,32E-01 |
| RMDN3 | 504,52 | 0,05 | 0,04 | 1,50 | 1,32E-01 |
| SRRD | 908,18 | 0,12 | 0,08 | 1,50 | 1,32E-01 |
| C12orf43 | 263,79 | 0,05 | 0,03 | 1,50 | 1,32E-01 |
| DNAJC18 | 128,00 | 0,07 | 0,05 | 1,50 | 1,32E-01 |
| MID2 | 150,80 | 0,09 | 0,06 | 1,50 | 1,32E-01 |
| ANAPC1 | 1568,88 | -0,05 | 0,04 | -1,50 | 1,32E-01 |
| MED23 | 3391,36 | -0,05 | 0,04 | -1,50 | 1,33E-01 |
| NAPG | 1464,68 | -0,06 | 0,04 | -1,50 | 1,33E-01 |
| HTR7 | 38,85 | 0,15 | 0,10 | 1,50 | 1,33E-01 |
| DECR1 | 4248,21 | -0,07 | 0,05 | -1,50 | 1,33E-01 |
| MOGS | 540,97 | 0,08 | 0,05 | 1,50 | 1,33E-01 |
| RBKS | 98,07 | 0,08 | 0,05 | 1,50 | 1,33E-01 |
| PPP2R5A | 4292,75 | -0,08 | 0,05 | -1,50 | 1,33E-01 |
| HP1BP3 | 6525,13 | -0,04 | 0,02 | -1,50 | 1,33E-01 |
| SERTAD3 | 279,23 | 0,07 | 0,04 | 1,50 | 1,33E-01 |
| CLDN12 | 72,11 | 0,11 | 0,07 | 1,50 | 1,34E-01 |
| LRRC34 | 40,18 | 0,14 | 0,10 | 1,50 | 1,34E-01 |

| | | | | | |
|---|---|---|---|---|---|
| BTN3A2 | 4274,26 | 0,11 | 0,07 | 1,50 | 1,34E-01 |
| SLITRK5 | 45,60 | 0,22 | 0,14 | 1,50 | 1,34E-01 |
| NFAM1 | 3041,37 | 0,18 | 0,12 | 1,50 | 1,34E-01 |
| IL2RB | 656,99 | 0,15 | 0,10 | 1,50 | 1,34E-01 |
| COPZ1 | 940,92 | 0,04 | 0,03 | 1,50 | 1,34E-01 |
| TBC1D10C | 3614,04 | 0,06 | 0,04 | 1,50 | 1,34E-01 |
| TACSTD2 | 46,94 | 0,30 | 0,20 | 1,50 | 1,34E-01 |
| ZNF530 | 267,28 | -0,06 | 0,04 | -1,50 | 1,34E-01 |
| CREBL2 | 1911,70 | -0,07 | 0,04 | -1,50 | 1,34E-01 |
| SIRPA | 2682,86 | 0,12 | 0,08 | 1,50 | 1,34E-01 |
| LNP1 | 48,78 | 0,12 | 0,08 | 1,50 | 1,34E-01 |
| FOXO3 | 10016,93 | 0,11 | 0,07 | 1,50 | 1,34E-01 |
| PAFAH1B2 | 2572,52 | -0,04 | 0,03 | -1,50 | 1,35E-01 |
| GPR174 | 2530,65 | -0,10 | 0,07 | -1,50 | 1,35E-01 |
| RRH | 29,36 | 0,11 | 0,08 | 1,50 | 1,35E-01 |
| IFT46 | 143,85 | 0,05 | 0,04 | 1,50 | 1,35E-01 |
| NPIPB2 | 43,95 | 0,14 | 0,09 | 1,50 | 1,35E-01 |
| UTY | 4645,31 | 0,29 | 0,19 | 1,49 | 1,35E-01 |
| DYNC2H1 | 472,51 | -0,10 | 0,06 | -1,49 | 1,35E-01 |
| NDUFAB1 | 496,53 | -0,06 | 0,04 | -1,49 | 1,35E-01 |
| LCMT1 | 248,61 | 0,06 | 0,04 | 1,49 | 1,35E-01 |
| KDM5D | 2520,05 | 0,31 | 0,21 | 1,49 | 1,35E-01 |
| METTL1 | 45,35 | 0,10 | 0,07 | 1,49 | 1,35E-01 |
| PCNP | 1968,21 | -0,09 | 0,06 | -1,49 | 1,36E-01 |
| ACSS1 | 1487,40 | 0,06 | 0,04 | 1,49 | 1,36E-01 |
| SLC25A40 | 1291,66 | -0,08 | 0,05 | -1,49 | 1,36E-01 |
| SDE2 | 1971,63 | -0,06 | 0,04 | -1,49 | 1,36E-01 |
| WDR6 | 1346,54 | 0,08 | 0,05 | 1,49 | 1,36E-01 |
| ARMH3 | 1022,58 | 0,05 | 0,03 | 1,49 | 1,36E-01 |
| CKLF | 977,00 | -0,08 | 0,05 | -1,49 | 1,36E-01 |
| QSOX1 | 719,70 | 0,11 | 0,07 | 1,49 | 1,37E-01 |
| PHKA2 | 1367,50 | 0,08 | 0,05 | 1,49 | 1,37E-01 |
| TP53INP1 | 6311,46 | -0,08 | 0,05 | -1,49 | 1,37E-01 |
| RAP1GAP2 | 2722,81 | 0,12 | 0,08 | 1,49 | 1,37E-01 |
| BBIP1 | 1212,09 | -0,05 | 0,04 | -1,49 | 1,37E-01 |
| ZNF442 | 89,71 | 0,14 | 0,09 | 1,49 | 1,37E-01 |
| ZNF502 | 139,29 | -0,14 | 0,10 | -1,49 | 1,37E-01 |
| NXF1 | 1758,39 | 0,07 | 0,05 | 1,49 | 1,37E-01 |
| TACC1 | 8258,07 | -0,06 | 0,04 | -1,49 | 1,37E-01 |
| ACVR1B | 696,79 | 0,08 | 0,05 | 1,49 | 1,37E-01 |
| SCD5 | 80,11 | 0,18 | 0,12 | 1,49 | 1,37E-01 |
| RNPEP | 880,83 | 0,10 | 0,07 | 1,49 | 1,37E-01 |
| ZNF584 | 141,53 | 0,06 | 0,04 | 1,49 | 1,37E-01 |
| ABI3 | 438,60 | 0,10 | 0,07 | 1,49 | 1,37E-01 |
| L3MBTL4 | 289,85 | 0,11 | 0,07 | 1,49 | 1,38E-01 |
| PRKCB | 11386,31 | -0,06 | 0,04 | -1,48 | 1,38E-01 |

| | | | | | |
|---|---|---|---|---|---|
| ATAD5 | 319,15 | -0,07 | 0,05 | -1,48 | 1,38E-01 |
| CSF1R | 1331,00 | 0,13 | 0,09 | 1,48 | 1,38E-01 |
| ARL17B | 106,44 | 0,19 | 0,13 | 1,48 | 1,38E-01 |
| ABCB10 | 1419,55 | 0,08 | 0,05 | 1,48 | 1,38E-01 |
| DGCR2 | 2949,74 | 0,10 | 0,07 | 1,48 | 1,38E-01 |
| AVEN | 164,32 | -0,06 | 0,04 | -1,48 | 1,39E-01 |
| ITM2A | 870,17 | -0,14 | 0,09 | -1,48 | 1,39E-01 |
| HPS6 | 446,97 | 0,06 | 0,04 | 1,48 | 1,39E-01 |
| INO80E | 1117,78 | 0,07 | 0,04 | 1,48 | 1,39E-01 |
| MFSD1 | 4335,22 | -0,08 | 0,05 | -1,48 | 1,39E-01 |
| STXBP5 | 6384,58 | -0,09 | 0,06 | -1,48 | 1,39E-01 |
| HK3 | 5521,50 | 0,10 | 0,06 | 1,48 | 1,39E-01 |
| PARVG | 3912,59 | 0,07 | 0,05 | 1,48 | 1,39E-01 |
| LYPLA2 | 396,68 | 0,06 | 0,04 | 1,48 | 1,39E-01 |
| NFRKB | 949,53 | 0,07 | 0,05 | 1,48 | 1,39E-01 |
| GABARAPL2 | 2228,70 | 0,10 | 0,07 | 1,48 | 1,39E-01 |
| VKORC1L1 | 643,30 | -0,04 | 0,03 | -1,48 | 1,39E-01 |
| SLC44A1 | 1462,94 | -0,05 | 0,03 | -1,48 | 1,39E-01 |
| CEP57L1 | 252,58 | -0,08 | 0,05 | -1,48 | 1,39E-01 |
| SPATS2L | 215,27 | 0,19 | 0,13 | 1,48 | 1,39E-01 |
| SLC35D1 | 1294,11 | -0,05 | 0,04 | -1,48 | 1,40E-01 |
| GNGT2 | 125,42 | -0,08 | 0,05 | -1,48 | 1,40E-01 |
| SCAMP3 | 304,99 | 0,05 | 0,04 | 1,48 | 1,40E-01 |
| USP53 | 1445,70 | -0,13 | 0,09 | -1,48 | 1,40E-01 |
| CD8B | 741,74 | -0,16 | 0,11 | -1,48 | 1,40E-01 |
| TCTA | 578,18 | 0,05 | 0,03 | 1,48 | 1,40E-01 |
| SFT2D3 | 560,57 | -0,07 | 0,05 | -1,48 | 1,40E-01 |
| MICAL2 | 7580,28 | 0,11 | 0,08 | 1,48 | 1,40E-01 |
| TRIM38 | 5823,99 | -0,07 | 0,05 | -1,48 | 1,40E-01 |
| MYOF | 794,97 | 0,14 | 0,09 | 1,47 | 1,40E-01 |
| LINS1 | 2102,92 | -0,07 | 0,04 | -1,47 | 1,40E-01 |
| LFNG | 3152,55 | 0,07 | 0,05 | 1,47 | 1,40E-01 |
| NUP62 | 639,55 | 0,09 | 0,06 | 1,47 | 1,40E-01 |
| ZNF248 | 486,25 | -0,07 | 0,04 | -1,47 | 1,41E-01 |
| RPS6KA5 | 5940,98 | -0,07 | 0,05 | -1,47 | 1,41E-01 |
| COMMD3 | 289,62 | -0,10 | 0,07 | -1,47 | 1,41E-01 |
| TPH1 | 73,52 | 0,14 | 0,10 | 1,47 | 1,41E-01 |
| DTWD1 | 805,69 | -0,06 | 0,04 | -1,47 | 1,41E-01 |
| IP6K1 | 1570,30 | 0,09 | 0,06 | 1,47 | 1,41E-01 |
| SYTL4 | 110,76 | 0,17 | 0,11 | 1,47 | 1,41E-01 |
| AKAP7 | 433,97 | -0,09 | 0,06 | -1,47 | 1,41E-01 |
| CD226 | 4526,04 | -0,08 | 0,06 | -1,47 | 1,41E-01 |
| BRPF3 | 706,54 | 0,08 | 0,05 | 1,47 | 1,41E-01 |
| NEIL1 | 227,59 | 0,12 | 0,08 | 1,47 | 1,42E-01 |
| U2AF2 | 1420,21 | 0,07 | 0,05 | 1,47 | 1,42E-01 |
| ZNF572 | 135,40 | -0,11 | 0,07 | -1,47 | 1,42E-01 |

| | | | | | |
|---|---|---|---|---|---|
| ALAS1 | 480,10 | 0,08 | 0,05 | 1,47 | 1,42E-01 |
| NEK3 | 197,65 | 0,07 | 0,04 | 1,47 | 1,42E-01 |
| FAM228B | 410,27 | -0,04 | 0,03 | -1,47 | 1,42E-01 |
| SMURF1 | 1220,37 | 0,05 | 0,03 | 1,47 | 1,42E-01 |
| ALCAM | 984,17 | -0,09 | 0,06 | -1,47 | 1,42E-01 |
| RBBP8 | 297,24 | -0,10 | 0,07 | -1,47 | 1,42E-01 |
| ZNF772 | 416,03 | -0,10 | 0,07 | -1,47 | 1,42E-01 |
| TMEM68 | 434,35 | -0,05 | 0,03 | -1,47 | 1,42E-01 |
| TMEM69 | 366,05 | -0,04 | 0,03 | -1,47 | 1,42E-01 |
| XKR8 | 630,28 | 0,10 | 0,07 | 1,47 | 1,43E-01 |
| PIGL | 575,86 | 0,06 | 0,04 | 1,47 | 1,43E-01 |
| EMP3 | 2684,74 | -0,07 | 0,05 | -1,47 | 1,43E-01 |
| UBE2G2 | 2996,60 | -0,04 | 0,03 | -1,47 | 1,43E-01 |
| SIRPB1 | 1736,47 | 0,13 | 0,09 | 1,47 | 1,43E-01 |
| GALNT7 | 1523,24 | -0,06 | 0,04 | -1,47 | 1,43E-01 |
| MPEG1 | 7341,63 | -0,09 | 0,06 | -1,47 | 1,43E-01 |
| ZDHHC16 | 501,89 | 0,05 | 0,03 | 1,46 | 1,43E-01 |
| LSM11 | 338,65 | -0,05 | 0,03 | -1,46 | 1,43E-01 |
| NUP210L | 98,32 | 0,27 | 0,19 | 1,46 | 1,43E-01 |
| SOWAHC | 113,44 | 0,11 | 0,08 | 1,46 | 1,43E-01 |
| SYNM | 318,19 | -0,19 | 0,13 | -1,46 | 1,43E-01 |
| APAF1 | 8957,93 | -0,08 | 0,05 | -1,46 | 1,43E-01 |
| PGK1 | 9309,97 | -0,05 | 0,04 | -1,46 | 1,43E-01 |
| ZFAND5 | 5357,24 | -0,07 | 0,05 | -1,46 | 1,43E-01 |
| TXN | 751,94 | -0,14 | 0,10 | -1,46 | 1,44E-01 |
| GEMIN2 | 154,71 | -0,08 | 0,05 | -1,46 | 1,44E-01 |
| C17orf75 | 358,41 | -0,06 | 0,04 | -1,46 | 1,44E-01 |
| PROSER1 | 1075,65 | -0,04 | 0,03 | -1,46 | 1,44E-01 |
| MMAB | 231,55 | 0,07 | 0,05 | 1,46 | 1,44E-01 |
| GTPBP1 | 2292,09 | 0,10 | 0,07 | 1,46 | 1,44E-01 |
| TBRG4 | 1046,63 | 0,05 | 0,04 | 1,46 | 1,44E-01 |
| UHMK1 | 8697,18 | -0,07 | 0,05 | -1,46 | 1,44E-01 |
| TRMT5 | 219,92 | -0,07 | 0,05 | -1,46 | 1,44E-01 |
| PIK3CD | 5183,91 | 0,12 | 0,08 | 1,46 | 1,44E-01 |
| ISL2 | 122,10 | 0,16 | 0,11 | 1,46 | 1,45E-01 |
| PSEN1 | 5483,98 | -0,06 | 0,04 | -1,46 | 1,45E-01 |
| MDM2 | 6851,46 | -0,07 | 0,05 | -1,46 | 1,45E-01 |
| PHF24 | 34,94 | 0,26 | 0,18 | 1,46 | 1,45E-01 |
| HLA-DQA1 | 1831,61 | -0,11 | 0,08 | -1,46 | 1,45E-01 |
| FAM120AOS | 1866,27 | -0,04 | 0,03 | -1,46 | 1,45E-01 |
| DNAAF9 | 690,31 | 0,07 | 0,05 | 1,46 | 1,45E-01 |
| ARHGAP31 | 640,24 | 0,08 | 0,05 | 1,46 | 1,45E-01 |
| TMEM165 | 3138,31 | 0,06 | 0,04 | 1,46 | 1,45E-01 |
| P4HA2 | 218,47 | 0,11 | 0,08 | 1,46 | 1,45E-01 |
| RAD51D | 633,84 | 0,06 | 0,04 | 1,46 | 1,45E-01 |
| COL9A3 | 252,51 | 0,19 | 0,13 | 1,46 | 1,45E-01 |

| | | | | | |
|---|---|---|---|---|---|
| SLC43A2 | 3296,42 | 0,11 | 0,08 | 1,46 | 1,45E-01 |
| RHOQ | 2669,31 | -0,06 | 0,04 | -1,46 | 1,46E-01 |
| TRMT61B | 312,71 | -0,05 | 0,04 | -1,46 | 1,46E-01 |
| ORAI2 | 3544,36 | 0,10 | 0,07 | 1,46 | 1,46E-01 |
| CCDC138 | 210,36 | -0,08 | 0,05 | -1,46 | 1,46E-01 |
| ZDHHC21 | 1655,25 | -0,11 | 0,08 | -1,46 | 1,46E-01 |
| EML4 | 5133,63 | -0,05 | 0,04 | -1,45 | 1,46E-01 |
| NAF1 | 442,73 | -0,06 | 0,04 | -1,46 | 1,46E-01 |
| BNIP1 | 105,01 | 0,07 | 0,05 | 1,45 | 1,46E-01 |
| DAD1 | 1314,44 | -0,07 | 0,05 | -1,45 | 1,46E-01 |
| ARHGAP32 | 346,46 | 0,08 | 0,06 | 1,45 | 1,46E-01 |
| AFF4 | 4072,05 | -0,04 | 0,03 | -1,45 | 1,46E-01 |
| CCDC126 | 558,87 | -0,10 | 0,07 | -1,45 | 1,46E-01 |
| EPS15L1 | 1168,05 | 0,13 | 0,09 | 1,45 | 1,46E-01 |
| BTBD6 | 411,10 | 0,06 | 0,04 | 1,45 | 1,46E-01 |
| CENPJ | 283,40 | -0,06 | 0,04 | -1,45 | 1,46E-01 |
| TUBGCP2 | 1531,41 | 0,05 | 0,04 | 1,45 | 1,46E-01 |
| SNAP23 | 6051,48 | -0,07 | 0,05 | -1,45 | 1,46E-01 |
| FAM185A | 316,27 | -0,06 | 0,04 | -1,45 | 1,46E-01 |
| SAV1 | 394,07 | -0,06 | 0,04 | -1,45 | 1,46E-01 |
| ALOX5AP | 3598,41 | -0,11 | 0,07 | -1,45 | 1,46E-01 |
| LYSMD4 | 470,32 | 0,06 | 0,04 | 1,45 | 1,46E-01 |
| ARHGEF9 | 965,44 | -0,06 | 0,04 | -1,45 | 1,46E-01 |
| LARP1B | 456,25 | -0,07 | 0,05 | -1,45 | 1,46E-01 |
| TMEM39A | 1133,50 | -0,04 | 0,03 | -1,45 | 1,47E-01 |
| NOTCH2 | 12798,24 | 0,09 | 0,06 | 1,45 | 1,47E-01 |
| MTHFSD | 601,58 | 0,04 | 0,03 | 1,45 | 1,47E-01 |
| LRIG1 | 1114,98 | 0,06 | 0,04 | 1,45 | 1,47E-01 |
| EFCAB11 | 122,69 | 0,09 | 0,06 | 1,45 | 1,47E-01 |
| CBL | 4812,81 | 0,12 | 0,09 | 1,45 | 1,47E-01 |
| KLHL8 | 1585,56 | -0,07 | 0,05 | -1,45 | 1,47E-01 |
| GZMK | 671,48 | -0,16 | 0,11 | -1,45 | 1,47E-01 |
| PGM2 | 2219,44 | -0,07 | 0,05 | -1,45 | 1,47E-01 |
| ERF | 353,15 | 0,10 | 0,07 | 1,45 | 1,47E-01 |
| KRBOX4 | 389,27 | -0,04 | 0,03 | -1,45 | 1,47E-01 |
| LUZP1 | 855,26 | -0,04 | 0,03 | -1,45 | 1,47E-01 |
| TRERF1 | 1710,19 | 0,07 | 0,05 | 1,45 | 1,47E-01 |
| CBX7 | 1834,50 | 0,09 | 0,06 | 1,45 | 1,47E-01 |
| CTNND1 | 750,28 | 0,09 | 0,06 | 1,45 | 1,47E-01 |
| LPL | 38,59 | 0,26 | 0,18 | 1,45 | 1,47E-01 |
| C1QA | 119,12 | -0,18 | 0,13 | -1,45 | 1,47E-01 |
| RDH11 | 1096,65 | -0,05 | 0,04 | -1,45 | 1,48E-01 |
| ZBTB20 | 3792,09 | -0,06 | 0,04 | -1,45 | 1,48E-01 |
| GOLGA3 | 2677,24 | 0,06 | 0,04 | 1,45 | 1,48E-01 |
| PPIP5K1 | 151,90 | 0,07 | 0,05 | 1,45 | 1,48E-01 |
| SMIM19 | 209,99 | -0,07 | 0,05 | -1,45 | 1,48E-01 |

| | | | | | |
|---|---|---|---|---|---|
| SRGAP2 | 1112,85 | 0,10 | 0,07 | 1,45 | 1,48E-01 |
| CCDC191 | 462,95 | -0,07 | 0,05 | -1,45 | 1,48E-01 |
| TOR1B | 1207,67 | -0,10 | 0,07 | -1,45 | 1,48E-01 |
| PTDSS1 | 2052,95 | -0,04 | 0,03 | -1,45 | 1,48E-01 |
| GALNT11 | 999,68 | -0,04 | 0,03 | -1,45 | 1,48E-01 |
| CDK5RAP3 | 1168,55 | 0,06 | 0,04 | 1,45 | 1,48E-01 |
| FADD | 1313,55 | -0,06 | 0,04 | -1,45 | 1,48E-01 |
| CENPI | 91,28 | 0,11 | 0,08 | 1,45 | 1,48E-01 |
| ZNF483 | 330,36 | -0,09 | 0,06 | -1,44 | 1,49E-01 |
| KIT | 123,44 | 0,13 | 0,09 | 1,44 | 1,49E-01 |
| MYO1G | 6177,21 | 0,06 | 0,04 | 1,44 | 1,49E-01 |
| CARMIL1 | 439,21 | 0,12 | 0,08 | 1,44 | 1,49E-01 |
| TUBB1 | 3015,13 | 0,14 | 0,10 | 1,44 | 1,49E-01 |
| FAM111B | 73,17 | -0,19 | 0,13 | -1,44 | 1,49E-01 |
| TENT4A | 877,78 | 0,07 | 0,05 | 1,44 | 1,49E-01 |
| SLC9B1 | 94,99 | 0,09 | 0,06 | 1,44 | 1,49E-01 |
| PMAIP1 | 432,51 | -0,12 | 0,08 | -1,44 | 1,49E-01 |
| VSTM1 | 223,87 | 0,15 | 0,11 | 1,44 | 1,49E-01 |
| DIMT1 | 500,65 | -0,07 | 0,05 | -1,44 | 1,50E-01 |
| ZNF16 | 184,05 | -0,06 | 0,04 | -1,44 | 1,50E-01 |
| FUBP1 | 5394,89 | -0,07 | 0,05 | -1,44 | 1,50E-01 |
| ATL1 | 125,40 | 0,10 | 0,07 | 1,44 | 1,50E-01 |
| PSTPIP2 | 1566,31 | 0,10 | 0,07 | 1,44 | 1,50E-01 |
| COCH | 83,71 | 0,21 | 0,15 | 1,44 | 1,50E-01 |
| ZNF662 | 74,29 | 0,14 | 0,10 | 1,44 | 1,50E-01 |
| YAF2 | 2007,90 | -0,05 | 0,04 | -1,44 | 1,50E-01 |
| RBM4 | 1537,61 | -0,04 | 0,03 | -1,44 | 1,50E-01 |
| ENO1 | 7717,95 | -0,06 | 0,04 | -1,44 | 1,50E-01 |
| STAT5B | 6121,84 | 0,09 | 0,06 | 1,44 | 1,50E-01 |
| CLEC4A | 310,03 | -0,11 | 0,07 | -1,44 | 1,50E-01 |
| SLC36A4 | 1228,27 | -0,09 | 0,06 | -1,44 | 1,51E-01 |
| CD177 | 79,79 | 0,30 | 0,21 | 1,44 | 1,51E-01 |
| TUBA1C | 629,76 | 0,06 | 0,04 | 1,44 | 1,51E-01 |
| AKR7A2 | 377,18 | -0,05 | 0,03 | -1,44 | 1,51E-01 |
| WDR83 | 190,19 | 0,04 | 0,03 | 1,44 | 1,51E-01 |
| AK3 | 689,72 | -0,06 | 0,04 | -1,44 | 1,51E-01 |
| FAM234B | 216,17 | 0,07 | 0,05 | 1,44 | 1,51E-01 |
| RPP25L | 359,39 | -0,05 | 0,04 | -1,44 | 1,51E-01 |
| DHRS4L2 | 174,03 | 0,06 | 0,04 | 1,44 | 1,51E-01 |
| AMMECR1 | 639,25 | -0,06 | 0,04 | -1,44 | 1,51E-01 |
| ELP4 | 1697,31 | -0,08 | 0,05 | -1,44 | 1,51E-01 |
| PLAAT3 | 157,93 | -0,10 | 0,07 | -1,44 | 1,51E-01 |
| ANKRD31 | 61,52 | 0,16 | 0,11 | 1,44 | 1,51E-01 |
| GAPVD1 | 4138,84 | -0,04 | 0,03 | -1,44 | 1,51E-01 |
| PIP4K2A | 13409,00 | 0,11 | 0,08 | 1,44 | 1,51E-01 |
| ATP6AP1 | 779,91 | 0,07 | 0,05 | 1,43 | 1,51E-01 |

| | | | | | |
|---|---|---|---|---|---|
| AGK | 1187,83 | -0,06 | 0,04 | -1,43 | 1,51E-01 |
| UBAC2 | 1769,78 | 0,04 | 0,03 | 1,43 | 1,51E-01 |
| CDHR3 | 430,03 | 0,10 | 0,07 | 1,43 | 1,52E-01 |
| NINL | 126,75 | 0,21 | 0,15 | 1,43 | 1,52E-01 |
| ZBTB49 | 470,85 | -0,04 | 0,03 | -1,43 | 1,52E-01 |
| PPCDC | 995,21 | 0,11 | 0,08 | 1,43 | 1,52E-01 |
| RPL21 | 7763,72 | -0,19 | 0,14 | -1,43 | 1,52E-01 |
| ZNF471 | 74,55 | 0,17 | 0,12 | 1,43 | 1,52E-01 |
| ZNF274 | 1690,47 | -0,04 | 0,03 | -1,43 | 1,52E-01 |
| PPIF | 1066,28 | 0,07 | 0,05 | 1,43 | 1,52E-01 |
| TLE4 | 4904,54 | -0,06 | 0,04 | -1,43 | 1,52E-01 |
| BRD8 | 1007,55 | -0,05 | 0,04 | -1,43 | 1,52E-01 |
| BUB1 | 112,16 | 0,15 | 0,11 | 1,43 | 1,52E-01 |
| RRP1B | 659,97 | 0,07 | 0,05 | 1,43 | 1,52E-01 |
| APBB1 | 490,20 | 0,08 | 0,06 | 1,43 | 1,52E-01 |
| GORAB | 368,46 | -0,08 | 0,05 | -1,43 | 1,52E-01 |
| MAN2A2 | 4018,65 | 0,08 | 0,06 | 1,43 | 1,53E-01 |
| FAM8A1 | 4145,99 | -0,10 | 0,07 | -1,43 | 1,53E-01 |
| LRRC1 | 135,05 | 0,10 | 0,07 | 1,43 | 1,53E-01 |
| HCLS1 | 19320,77 | -0,07 | 0,05 | -1,43 | 1,53E-01 |
| AKNA | 8154,77 | 0,13 | 0,09 | 1,43 | 1,53E-01 |
| CANT1 | 1452,76 | 0,09 | 0,07 | 1,43 | 1,53E-01 |
| SELENOI | 1080,69 | -0,05 | 0,04 | -1,43 | 1,53E-01 |
| INSIG1 | 1263,02 | -0,06 | 0,04 | -1,43 | 1,53E-01 |
| AOPEP | 678,83 | 0,07 | 0,05 | 1,43 | 1,53E-01 |
| MDFIC | 2584,14 | -0,07 | 0,05 | -1,43 | 1,53E-01 |
| KLHDC4 | 1350,44 | 0,10 | 0,07 | 1,43 | 1,53E-01 |
| FAM135A | 806,95 | -0,08 | 0,05 | -1,43 | 1,53E-01 |
| FAM120C | 309,06 | 0,06 | 0,04 | 1,43 | 1,53E-01 |
| TSPYL5 | 102,48 | 0,12 | 0,09 | 1,43 | 1,53E-01 |
| ZFY | 547,26 | -0,16 | 0,11 | -1,43 | 1,53E-01 |
| RGS18 | 4860,52 | -0,15 | 0,10 | -1,43 | 1,53E-01 |
| ESYT1 | 2469,24 | 0,07 | 0,05 | 1,43 | 1,54E-01 |
| RRAGC | 476,41 | -0,05 | 0,03 | -1,43 | 1,54E-01 |
| FMO5 | 338,45 | -0,07 | 0,05 | -1,43 | 1,54E-01 |
| TEF | 209,32 | 0,07 | 0,05 | 1,43 | 1,54E-01 |
| KNSTRN | 308,02 | -0,06 | 0,05 | -1,43 | 1,54E-01 |
| CSNK1G1 | 713,04 | 0,07 | 0,05 | 1,43 | 1,54E-01 |
| VWDE | 163,91 | -0,22 | 0,15 | -1,43 | 1,54E-01 |
| ST20 | 882,81 | -0,10 | 0,07 | -1,43 | 1,54E-01 |
| ZNF658 | 160,12 | -0,11 | 0,08 | -1,43 | 1,54E-01 |
| EHHADH | 84,30 | 0,11 | 0,08 | 1,43 | 1,54E-01 |
| PRUNE1 | 681,40 | 0,05 | 0,04 | 1,43 | 1,54E-01 |
| MBTPS2 | 345,20 | -0,06 | 0,04 | -1,42 | 1,54E-01 |
| APOOL | 301,98 | -0,05 | 0,03 | -1,42 | 1,54E-01 |
| E2F6 | 244,03 | -0,05 | 0,03 | -1,42 | 1,54E-01 |

| | | | | | |
|---|---|---|---|---|---|
| CEBPB | 4484,21 | 0,09 | 0,06 | 1,42 | 1,54E-01 |
| RPP21 | 52,60 | 0,08 | 0,06 | 1,42 | 1,55E-01 |
| CNN2 | 4579,68 | 0,12 | 0,08 | 1,42 | 1,55E-01 |
| MVD | 379,60 | 0,06 | 0,05 | 1,42 | 1,55E-01 |
| MYH9 | 16788,57 | 0,12 | 0,09 | 1,42 | 1,55E-01 |
| FKTN | 674,44 | -0,06 | 0,04 | -1,42 | 1,55E-01 |
| PINK1 | 3853,77 | 0,11 | 0,08 | 1,42 | 1,55E-01 |
| TAS2R43 | 89,71 | 0,27 | 0,19 | 1,42 | 1,55E-01 |
| VWA8 | 1517,12 | -0,05 | 0,04 | -1,42 | 1,55E-01 |
| NCSTN | 1147,39 | 0,08 | 0,06 | 1,42 | 1,55E-01 |
| TRPC6 | 79,85 | 0,19 | 0,14 | 1,42 | 1,55E-01 |
| FAM117B | 1175,46 | -0,07 | 0,05 | -1,42 | 1,55E-01 |
| POLG2 | 652,15 | -0,05 | 0,03 | -1,42 | 1,55E-01 |
| SLC43A3 | 273,78 | 0,07 | 0,05 | 1,42 | 1,55E-01 |
| SNRPB | 660,70 | -0,04 | 0,03 | -1,42 | 1,55E-01 |
| PARP4 | 5308,72 | -0,06 | 0,04 | -1,42 | 1,55E-01 |
| NCK2 | 2163,18 | 0,04 | 0,03 | 1,42 | 1,56E-01 |
| BFSP1 | 60,37 | 0,21 | 0,15 | 1,42 | 1,56E-01 |
| ZNF280D | 927,08 | -0,09 | 0,07 | -1,42 | 1,56E-01 |
| PLS1 | 58,78 | 0,15 | 0,11 | 1,42 | 1,56E-01 |
| SETD6 | 322,38 | 0,05 | 0,03 | 1,42 | 1,56E-01 |
| NINJ2 | 616,42 | 0,12 | 0,09 | 1,42 | 1,56E-01 |
| CCNG1 | 1521,84 | -0,09 | 0,06 | -1,42 | 1,56E-01 |
| ANGPT1 | 352,03 | -0,15 | 0,11 | -1,42 | 1,56E-01 |
| RNF40 | 2417,05 | 0,07 | 0,05 | 1,42 | 1,56E-01 |
| GPM6A | 101,53 | 0,28 | 0,20 | 1,42 | 1,56E-01 |
| SKA3 | 99,03 | 0,18 | 0,13 | 1,42 | 1,56E-01 |
| C1RL | 2421,49 | 0,08 | 0,06 | 1,42 | 1,56E-01 |
| UBE2M | 702,47 | 0,07 | 0,05 | 1,42 | 1,56E-01 |
| ZNF829 | 208,64 | -0,09 | 0,06 | -1,42 | 1,57E-01 |
| ZSCAN5A | 283,72 | 0,07 | 0,05 | 1,42 | 1,57E-01 |
| WASL | 630,48 | -0,07 | 0,05 | -1,42 | 1,57E-01 |
| RASGEF1B | 736,25 | -0,07 | 0,05 | -1,42 | 1,57E-01 |
| PSMB6 | 653,47 | -0,07 | 0,05 | -1,42 | 1,57E-01 |
| PRKAB1 | 930,01 | 0,04 | 0,03 | 1,42 | 1,57E-01 |
| ATP23 | 142,22 | 0,09 | 0,06 | 1,41 | 1,57E-01 |
| ZNF74 | 242,86 | 0,06 | 0,05 | 1,41 | 1,57E-01 |
| VCPKMT | 1045,39 | -0,07 | 0,05 | -1,41 | 1,57E-01 |
| H2AC15 | 328,30 | -0,10 | 0,07 | -1,41 | 1,57E-01 |
| UBE2D3 | 16576,99 | -0,06 | 0,05 | -1,41 | 1,58E-01 |
| ANGEL1 | 568,53 | 0,05 | 0,04 | 1,41 | 1,58E-01 |
| APOBEC3B | 61,59 | 0,37 | 0,26 | 1,41 | 1,58E-01 |
| ZC3H14 | 1336,90 | -0,04 | 0,03 | -1,41 | 1,58E-01 |
| SEMA4A | 1700,19 | 0,12 | 0,08 | 1,41 | 1,58E-01 |
| SNAPIN | 402,36 | -0,05 | 0,04 | -1,41 | 1,58E-01 |
| ALPL | 1309,93 | 0,18 | 0,13 | 1,41 | 1,58E-01 |

| | | | | | |
|---|---|---|---|---|---|
| PGPEP1 | 784,01 | 0,07 | 0,05 | 1,41 | 1,58E-01 |
| AKR1B1 | 397,77 | -0,05 | 0,04 | -1,41 | 1,58E-01 |
| EOLA2 | 327,38 | 0,05 | 0,03 | 1,41 | 1,58E-01 |
| ATG2A | 986,43 | 0,16 | 0,11 | 1,41 | 1,58E-01 |
| NBDY | 112,96 | 0,07 | 0,05 | 1,41 | 1,58E-01 |
| ADAMTS1 | 230,69 | -0,14 | 0,10 | -1,41 | 1,59E-01 |
| COL4A3 | 204,47 | 0,19 | 0,13 | 1,41 | 1,59E-01 |
| KMO | 346,19 | -0,09 | 0,06 | -1,41 | 1,59E-01 |
| ISCA2 | 550,61 | -0,06 | 0,04 | -1,41 | 1,59E-01 |
| GPR89B | 553,71 | -0,06 | 0,04 | -1,41 | 1,59E-01 |
| TRMT61A | 219,62 | 0,06 | 0,04 | 1,41 | 1,59E-01 |
| POLR2G | 917,54 | -0,04 | 0,03 | -1,41 | 1,59E-01 |
| TMEM50A | 3674,53 | -0,07 | 0,05 | -1,41 | 1,59E-01 |
| ENSG00000250644 | 107,63 | 0,12 | 0,09 | 1,41 | 1,59E-01 |
| MZT1 | 223,91 | -0,12 | 0,08 | -1,41 | 1,60E-01 |
| PNO1 | 295,08 | -0,05 | 0,04 | -1,41 | 1,60E-01 |
| CNTRL | 3742,47 | -0,05 | 0,04 | -1,41 | 1,60E-01 |
| ZFP37 | 94,22 | -0,13 | 0,09 | -1,41 | 1,60E-01 |
| TIMM23 | 557,22 | -0,06 | 0,04 | -1,41 | 1,60E-01 |
| EIF2B4 | 402,18 | 0,04 | 0,03 | 1,41 | 1,60E-01 |
| SF3B3 | 3421,75 | -0,04 | 0,03 | -1,40 | 1,60E-01 |
| TARBP1 | 1543,99 | -0,06 | 0,04 | -1,40 | 1,60E-01 |
| PARP1 | 2342,77 | -0,06 | 0,04 | -1,40 | 1,61E-01 |
| ADAP1 | 694,19 | 0,07 | 0,05 | 1,40 | 1,61E-01 |
| RBMS2 | 237,43 | 0,13 | 0,09 | 1,40 | 1,61E-01 |
| JCHAIN | 1540,21 | -0,23 | 0,16 | -1,40 | 1,61E-01 |
| UACA | 157,42 | 0,16 | 0,11 | 1,40 | 1,61E-01 |
| PEA15 | 530,10 | 0,06 | 0,05 | 1,40 | 1,61E-01 |
| TMPPE | 705,65 | 0,09 | 0,06 | 1,40 | 1,62E-01 |
| PET117 | 62,84 | -0,09 | 0,06 | -1,40 | 1,62E-01 |
| GADD45GIP1 | 200,99 | 0,07 | 0,05 | 1,40 | 1,62E-01 |
| TREM1 | 6534,30 | -0,11 | 0,08 | -1,40 | 1,62E-01 |
| KHDRBS2 | 120,25 | -0,14 | 0,10 | -1,40 | 1,62E-01 |
| MPDU1 | 402,23 | 0,05 | 0,04 | 1,40 | 1,62E-01 |
| SLC38A11 | 162,21 | 0,19 | 0,13 | 1,40 | 1,62E-01 |
| INO80 | 1525,40 | -0,04 | 0,03 | -1,40 | 1,62E-01 |
| C14orf93 | 308,13 | 0,04 | 0,03 | 1,40 | 1,62E-01 |
| RPS7 | 2468,36 | -0,19 | 0,13 | -1,40 | 1,62E-01 |
| MTERF4 | 1971,73 | -0,05 | 0,04 | -1,40 | 1,62E-01 |
| ACOT2 | 136,98 | 0,07 | 0,05 | 1,40 | 1,63E-01 |
| NAA25 | 1213,55 | -0,04 | 0,03 | -1,40 | 1,63E-01 |
| PNPLA1 | 123,01 | 0,12 | 0,09 | 1,40 | 1,63E-01 |
| RFX3 | 1816,25 | -0,06 | 0,05 | -1,40 | 1,63E-01 |
| MRPL58 | 170,91 | -0,06 | 0,04 | -1,40 | 1,63E-01 |
| RAB31 | 5637,95 | -0,08 | 0,06 | -1,40 | 1,63E-01 |
| CAMK1D | 2366,80 | 0,10 | 0,07 | 1,40 | 1,63E-01 |

| | | | | | |
|---|---|---|---|---|---|
| PYROXD2 | 455,77 | 0,07 | 0,05 | 1,40 | 1,63E-01 |
| ANKRD50 | 847,08 | -0,08 | 0,06 | -1,39 | 1,63E-01 |
| RPS19 | 10185,50 | -0,09 | 0,06 | -1,39 | 1,63E-01 |
| PIGU | 250,37 | 0,05 | 0,04 | 1,39 | 1,63E-01 |
| ITLN1 | 99,77 | 0,23 | 0,17 | 1,39 | 1,64E-01 |
| RPSA | 9507,28 | -0,11 | 0,08 | -1,39 | 1,64E-01 |
| PAM16 | 208,98 | 0,13 | 0,09 | 1,39 | 1,64E-01 |
| RCN1 | 151,75 | 0,08 | 0,06 | 1,39 | 1,64E-01 |
| EIF3K | 2400,29 | -0,06 | 0,04 | -1,39 | 1,64E-01 |
| TMEM237 | 105,23 | 0,11 | 0,08 | 1,39 | 1,64E-01 |
| MED17 | 1466,68 | -0,03 | 0,02 | -1,39 | 1,64E-01 |
| C16orf87 | 320,44 | -0,05 | 0,04 | -1,39 | 1,64E-01 |
| KAT2B | 6720,69 | 0,09 | 0,06 | 1,39 | 1,64E-01 |
| GPR171 | 452,54 | -0,11 | 0,08 | -1,39 | 1,64E-01 |
| PKM | 2700,73 | 0,09 | 0,06 | 1,39 | 1,64E-01 |
| MYO3B | 85,71 | 0,28 | 0,20 | 1,39 | 1,65E-01 |
| CERS5 | 932,00 | 0,04 | 0,03 | 1,39 | 1,65E-01 |
| RAB30 | 763,04 | -0,09 | 0,06 | -1,39 | 1,65E-01 |
| INTS4 | 1419,85 | -0,03 | 0,02 | -1,39 | 1,65E-01 |
| SHISA5 | 2204,40 | 0,10 | 0,08 | 1,39 | 1,65E-01 |
| UGDH | 324,16 | -0,06 | 0,04 | -1,39 | 1,65E-01 |
| H3C1 | 266,25 | -0,11 | 0,08 | -1,39 | 1,65E-01 |
| BEND2 | 259,20 | 0,15 | 0,10 | 1,39 | 1,65E-01 |
| TMEM45B | 240,02 | 0,12 | 0,09 | 1,39 | 1,65E-01 |
| GRIPAP1 | 1047,18 | 0,09 | 0,06 | 1,39 | 1,65E-01 |
| METTL2B | 688,12 | -0,04 | 0,03 | -1,39 | 1,65E-01 |
| MADD | 2041,08 | 0,07 | 0,05 | 1,39 | 1,66E-01 |
| RUNDC1 | 561,68 | 0,07 | 0,05 | 1,39 | 1,66E-01 |
| MRPL54 | 400,63 | -0,08 | 0,06 | -1,39 | 1,66E-01 |
| PSMG1 | 269,93 | -0,06 | 0,04 | -1,39 | 1,66E-01 |
| IL1RL1 | 495,94 | 0,19 | 0,14 | 1,39 | 1,66E-01 |
| PIGB | 2320,65 | -0,09 | 0,06 | -1,39 | 1,66E-01 |
| UPF1 | 3403,33 | 0,08 | 0,06 | 1,39 | 1,66E-01 |
| LAMTOR4 | 2119,18 | -0,07 | 0,05 | -1,39 | 1,66E-01 |
| GBF1 | 1272,95 | 0,08 | 0,06 | 1,39 | 1,66E-01 |
| NIPSNAP1 | 196,79 | 0,06 | 0,04 | 1,39 | 1,66E-01 |
| PABPC4 | 1349,13 | 0,05 | 0,03 | 1,39 | 1,66E-01 |
| MRPL18 | 483,95 | -0,06 | 0,04 | -1,39 | 1,66E-01 |
| WDR17 | 89,28 | 0,12 | 0,09 | 1,38 | 1,66E-01 |
| NPL | 4625,54 | -0,09 | 0,07 | -1,38 | 1,66E-01 |
| CD160 | 273,82 | -0,13 | 0,10 | -1,38 | 1,66E-01 |
| CCDC7 | 877,55 | -0,09 | 0,06 | -1,38 | 1,66E-01 |
| TAP1 | 6754,07 | 0,09 | 0,06 | 1,38 | 1,66E-01 |
| H1-3 | 3451,90 | -0,10 | 0,07 | -1,38 | 1,66E-01 |
| MARS1 | 2104,74 | 0,07 | 0,05 | 1,38 | 1,67E-01 |
| VPS8 | 8649,68 | -0,07 | 0,05 | -1,38 | 1,67E-01 |

| | | | | | |
|---|---|---|---|---|---|
| RASA3 | 3559,16 | 0,08 | 0,06 | 1,38 | 1,67E-01 |
| FCRL5 | 804,98 | -0,22 | 0,16 | -1,38 | 1,67E-01 |
| SPRING1 | 1207,69 | -0,03 | 0,02 | -1,38 | 1,67E-01 |
| RXRA | 5255,57 | 0,10 | 0,07 | 1,38 | 1,67E-01 |
| ZAP70 | 3864,53 | 0,08 | 0,06 | 1,38 | 1,67E-01 |
| ANKRD36 | 3107,14 | -0,07 | 0,05 | -1,38 | 1,67E-01 |
| GOLT1B | 1004,38 | -0,10 | 0,07 | -1,38 | 1,67E-01 |
| NFXL1 | 1113,84 | -0,14 | 0,10 | -1,38 | 1,67E-01 |
| MXD3 | 1084,68 | 0,09 | 0,07 | 1,38 | 1,67E-01 |
| TIFAB | 46,81 | 0,13 | 0,09 | 1,38 | 1,68E-01 |
| ZNF892 | 139,40 | -0,06 | 0,05 | -1,38 | 1,68E-01 |
| ILRUN | 2025,47 | 0,05 | 0,04 | 1,38 | 1,68E-01 |
| VANGL1 | 207,56 | 0,09 | 0,07 | 1,38 | 1,68E-01 |
| MORC2 | 1106,55 | 0,04 | 0,03 | 1,38 | 1,68E-01 |
| BTG3 | 132,09 | -0,08 | 0,06 | -1,38 | 1,68E-01 |
| C4orf33 | 591,32 | -0,07 | 0,05 | -1,38 | 1,68E-01 |
| ZNF846 | 336,91 | -0,05 | 0,04 | -1,38 | 1,68E-01 |
| CPNE1 | 2330,86 | 0,04 | 0,03 | 1,38 | 1,68E-01 |
| NQO2 | 1586,78 | 0,11 | 0,08 | 1,38 | 1,68E-01 |
| SH2D2A | 157,94 | 0,16 | 0,12 | 1,38 | 1,68E-01 |
| ZNF563 | 126,27 | -0,09 | 0,07 | -1,38 | 1,69E-01 |
| ATP5MK | 539,58 | -0,09 | 0,07 | -1,38 | 1,69E-01 |
| SPAG7 | 248,63 | -0,07 | 0,05 | -1,38 | 1,69E-01 |
| IGSF6 | 8199,23 | -0,09 | 0,06 | -1,38 | 1,69E-01 |
| METTL13 | 750,01 | -0,04 | 0,03 | -1,38 | 1,69E-01 |
| CNST | 2381,48 | -0,05 | 0,03 | -1,38 | 1,69E-01 |
| ORMDL1 | 1912,72 | -0,07 | 0,05 | -1,38 | 1,69E-01 |
| FRMPD3 | 88,33 | 0,18 | 0,13 | 1,37 | 1,69E-01 |
| STK10 | 7495,92 | 0,10 | 0,07 | 1,37 | 1,69E-01 |
| PSMG3 | 269,59 | 0,05 | 0,04 | 1,37 | 1,69E-01 |
| MRPS7 | 445,57 | -0,06 | 0,05 | -1,37 | 1,69E-01 |
| SLC37A3 | 1318,87 | -0,09 | 0,06 | -1,37 | 1,69E-01 |
| PTTG1 | 93,96 | 0,10 | 0,07 | 1,37 | 1,70E-01 |
| NUP188 | 667,20 | 0,07 | 0,05 | 1,37 | 1,70E-01 |
| CCND2 | 3341,59 | -0,06 | 0,04 | -1,37 | 1,70E-01 |
| CPSF7 | 1103,14 | 0,09 | 0,07 | 1,37 | 1,70E-01 |
| MED9 | 254,80 | 0,05 | 0,04 | 1,37 | 1,70E-01 |
| DDX49 | 563,22 | 0,04 | 0,03 | 1,37 | 1,70E-01 |
| CEPT1 | 2571,61 | -0,06 | 0,05 | -1,37 | 1,70E-01 |
| TFPT | 247,59 | -0,06 | 0,04 | -1,37 | 1,70E-01 |
| SUV39H2 | 102,06 | -0,09 | 0,06 | -1,37 | 1,70E-01 |
| NME7 | 236,08 | -0,07 | 0,05 | -1,37 | 1,71E-01 |
| GANAB | 1723,76 | 0,07 | 0,05 | 1,37 | 1,71E-01 |
| ENSG00000258461 | 110,39 | 0,10 | 0,07 | 1,37 | 1,71E-01 |
| ZC4H2 | 120,54 | 0,08 | 0,06 | 1,37 | 1,71E-01 |
| NUDT9 | 277,22 | -0,05 | 0,03 | -1,37 | 1,71E-01 |

| | | | | | |
|---|---|---|---|---|---|
| DUSP10 | 307,94 | -0,04 | 0,03 | -1,37 | 1,71E-01 |
| FOXRED1 | 289,52 | 0,07 | 0,05 | 1,37 | 1,71E-01 |
| ZNF544 | 498,22 | -0,05 | 0,04 | -1,37 | 1,71E-01 |
| MRTFB | 624,56 | 0,07 | 0,05 | 1,37 | 1,72E-01 |
| SDHC | 1710,33 | -0,03 | 0,03 | -1,37 | 1,72E-01 |
| LIAS | 491,11 | 0,03 | 0,03 | 1,37 | 1,72E-01 |
| NEBL | 203,33 | 0,39 | 0,28 | 1,37 | 1,72E-01 |
| BRD3 | 896,21 | 0,08 | 0,05 | 1,37 | 1,72E-01 |
| RNH1 | 1363,39 | 0,06 | 0,04 | 1,37 | 1,72E-01 |
| DERL2 | 1209,08 | -0,04 | 0,03 | -1,36 | 1,72E-01 |
| MRPL30 | 493,60 | -0,05 | 0,04 | -1,36 | 1,72E-01 |
| UFD1 | 917,28 | 0,04 | 0,03 | 1,36 | 1,72E-01 |
| EXOSC6 | 1232,63 | -0,05 | 0,04 | -1,36 | 1,72E-01 |
| KIF3B | 988,34 | -0,04 | 0,03 | -1,36 | 1,72E-01 |
| BMP6 | 207,56 | 0,11 | 0,08 | 1,36 | 1,73E-01 |
| TIMM17B | 519,48 | 0,05 | 0,04 | 1,36 | 1,73E-01 |
| TCN1 | 133,94 | 0,16 | 0,12 | 1,36 | 1,73E-01 |
| ZNF232 | 203,83 | -0,05 | 0,04 | -1,36 | 1,73E-01 |
| UBN2 | 2840,86 | -0,04 | 0,03 | -1,36 | 1,73E-01 |
| ZSCAN21 | 152,47 | -0,04 | 0,03 | -1,36 | 1,73E-01 |
| BEST1 | 1555,61 | 0,11 | 0,08 | 1,36 | 1,73E-01 |
| RGS9 | 135,94 | 0,16 | 0,12 | 1,36 | 1,73E-01 |
| STX16 | 3090,60 | 0,06 | 0,04 | 1,36 | 1,73E-01 |
| NANP | 129,00 | -0,06 | 0,05 | -1,36 | 1,73E-01 |
| RPRD2 | 1023,57 | -0,04 | 0,03 | -1,36 | 1,73E-01 |
| ARHGAP9 | 3413,81 | 0,12 | 0,08 | 1,36 | 1,73E-01 |
| LITAF | 15175,79 | -0,09 | 0,06 | -1,36 | 1,73E-01 |
| NFATC2IP | 1520,92 | 0,05 | 0,04 | 1,36 | 1,74E-01 |
| UQCRQ | 446,90 | -0,10 | 0,07 | -1,36 | 1,74E-01 |
| MORN3 | 137,66 | 0,09 | 0,07 | 1,36 | 1,74E-01 |
| LILRB1 | 681,52 | 0,11 | 0,08 | 1,36 | 1,74E-01 |
| GNB1 | 3786,83 | 0,06 | 0,05 | 1,36 | 1,74E-01 |
| METTL23 | 535,97 | -0,05 | 0,04 | -1,36 | 1,74E-01 |
| CARD11 | 1121,24 | 0,10 | 0,07 | 1,36 | 1,74E-01 |
| DNMT1 | 1759,16 | 0,06 | 0,04 | 1,36 | 1,74E-01 |
| COX7A2 | 968,03 | -0,11 | 0,08 | -1,36 | 1,74E-01 |
| RABEP2 | 546,20 | 0,07 | 0,05 | 1,36 | 1,74E-01 |
| MERTK | 183,34 | 0,13 | 0,10 | 1,36 | 1,74E-01 |
| AVL9 | 4574,52 | -0,04 | 0,03 | -1,36 | 1,74E-01 |
| ENSG00000258539 | 59,99 | 0,16 | 0,12 | 1,36 | 1,74E-01 |
| KPNB1 | 6340,21 | -0,07 | 0,05 | -1,36 | 1,74E-01 |
| RC3H2 | 4369,51 | -0,04 | 0,03 | -1,36 | 1,74E-01 |
| TAB2 | 4249,37 | -0,05 | 0,03 | -1,36 | 1,74E-01 |
| TMED4 | 1982,60 | -0,03 | 0,02 | -1,36 | 1,75E-01 |
| TSEN54 | 323,71 | 0,09 | 0,07 | 1,36 | 1,75E-01 |
| CD200R1 | 687,20 | -0,09 | 0,06 | -1,36 | 1,75E-01 |

| | | | | | |
|---|---|---|---|---|---|
| SGMS1 | 1905,07 | -0,04 | 0,03 | -1,36 | 1,75E-01 |
| UBXN10 | 124,63 | 0,13 | 0,09 | 1,36 | 1,75E-01 |
| ULK2 | 833,27 | 0,05 | 0,03 | 1,36 | 1,75E-01 |
| SPTLC2 | 4948,98 | -0,05 | 0,04 | -1,36 | 1,75E-01 |
| MOSMO | 801,49 | -0,06 | 0,05 | -1,35 | 1,75E-01 |
| CBR3 | 56,05 | 0,14 | 0,10 | 1,35 | 1,76E-01 |
| ERG28 | 167,72 | 0,05 | 0,03 | 1,35 | 1,76E-01 |
| ENSG00000281593 | 38,00 | 0,12 | 0,09 | 1,35 | 1,76E-01 |
| ZFP30 | 526,52 | -0,10 | 0,07 | -1,35 | 1,76E-01 |
| CNP | 435,32 | 0,08 | 0,06 | 1,35 | 1,76E-01 |
| WBP1 | 330,06 | 0,04 | 0,03 | 1,35 | 1,76E-01 |
| RACK1 | 13939,18 | -0,06 | 0,05 | -1,35 | 1,76E-01 |
| TC2N | 5642,19 | -0,09 | 0,07 | -1,35 | 1,76E-01 |
| PPM1F | 4587,35 | 0,09 | 0,07 | 1,35 | 1,76E-01 |
| INVS | 918,51 | -0,04 | 0,03 | -1,35 | 1,76E-01 |
| OAZ2 | 4385,10 | 0,08 | 0,06 | 1,35 | 1,76E-01 |
| EOGT | 277,58 | 0,04 | 0,03 | 1,35 | 1,76E-01 |
| IFIT5 | 1902,09 | -0,15 | 0,11 | -1,35 | 1,76E-01 |
| ISOC1 | 368,89 | -0,07 | 0,05 | -1,35 | 1,77E-01 |
| PHF23 | 1017,83 | -0,04 | 0,03 | -1,35 | 1,77E-01 |
| SLAMF7 | 1117,93 | -0,09 | 0,07 | -1,35 | 1,77E-01 |
| INO80D | 5138,54 | -0,05 | 0,03 | -1,35 | 1,77E-01 |
| SGCE | 98,52 | 0,20 | 0,15 | 1,35 | 1,77E-01 |
| ZNF683 | 67,44 | 0,27 | 0,20 | 1,35 | 1,77E-01 |
| SPOCK2 | 2987,14 | 0,09 | 0,06 | 1,35 | 1,77E-01 |
| LRRC37B | 732,29 | 0,06 | 0,05 | 1,35 | 1,77E-01 |
| SESN2 | 196,44 | 0,10 | 0,07 | 1,35 | 1,77E-01 |
| TMEM101 | 670,88 | -0,05 | 0,04 | -1,35 | 1,77E-01 |
| THEM4 | 475,73 | 0,08 | 0,06 | 1,35 | 1,77E-01 |
| C1orf216 | 245,87 | -0,06 | 0,05 | -1,35 | 1,77E-01 |
| CLDND1 | 1945,28 | -0,07 | 0,05 | -1,35 | 1,77E-01 |
| IMMT | 1128,79 | -0,04 | 0,03 | -1,35 | 1,78E-01 |
| SLC25A5 | 1548,18 | -0,05 | 0,04 | -1,35 | 1,78E-01 |
| C2CD3 | 1469,17 | 0,05 | 0,04 | 1,35 | 1,78E-01 |
| LIX1L | 1186,16 | 0,04 | 0,03 | 1,35 | 1,78E-01 |
| NHLRC3 | 1199,95 | -0,05 | 0,04 | -1,35 | 1,78E-01 |
| LILRB2 | 2056,80 | 0,12 | 0,09 | 1,35 | 1,78E-01 |
| CCDC171 | 151,23 | 0,12 | 0,09 | 1,35 | 1,78E-01 |
| EVI5 | 1461,60 | -0,07 | 0,05 | -1,35 | 1,79E-01 |
| CAMK2G | 2824,38 | 0,09 | 0,06 | 1,34 | 1,79E-01 |
| ANKDD1A | 230,61 | 0,15 | 0,11 | 1,34 | 1,79E-01 |
| COA6 | 168,42 | -0,09 | 0,06 | -1,34 | 1,79E-01 |
| MEI1 | 403,64 | 0,06 | 0,05 | 1,34 | 1,79E-01 |
| RNF114 | 2446,02 | -0,04 | 0,03 | -1,34 | 1,79E-01 |
| FAN1 | 1049,56 | -0,06 | 0,05 | -1,34 | 1,79E-01 |
| DCAF1 | 1227,31 | -0,04 | 0,03 | -1,34 | 1,79E-01 |

| | | | | | |
|---|---|---|---|---|---|
| FANCL | 219,81 | -0,10 | 0,07 | -1,34 | 1,79E-01 |
| GNAL | 100,94 | 0,16 | 0,12 | 1,34 | 1,79E-01 |
| PSMC5 | 524,96 | 0,05 | 0,04 | 1,34 | 1,79E-01 |
| FEM1C | 1756,72 | -0,07 | 0,05 | -1,34 | 1,79E-01 |
| DUT | 1500,49 | -0,07 | 0,05 | -1,34 | 1,80E-01 |
| FMNL2 | 190,68 | 0,12 | 0,09 | 1,34 | 1,80E-01 |
| CLK2 | 938,78 | 0,05 | 0,04 | 1,34 | 1,80E-01 |
| PADI2 | 2048,31 | 0,17 | 0,12 | 1,34 | 1,80E-01 |
| GTF2H2C | 419,62 | -0,06 | 0,05 | -1,34 | 1,80E-01 |
| ZNF592 | 1927,38 | 0,09 | 0,07 | 1,34 | 1,80E-01 |
| ITGAM | 2807,11 | 0,12 | 0,09 | 1,34 | 1,80E-01 |
| SLC35A2 | 207,74 | 0,05 | 0,04 | 1,34 | 1,80E-01 |
| GTF3C5 | 725,59 | 0,03 | 0,03 | 1,34 | 1,80E-01 |
| REXO4 | 628,24 | -0,04 | 0,03 | -1,34 | 1,80E-01 |
| SF1 | 3964,76 | 0,07 | 0,05 | 1,34 | 1,80E-01 |
| DTX4 | 278,95 | 0,10 | 0,07 | 1,34 | 1,80E-01 |
| AKAP8L | 1425,40 | 0,05 | 0,04 | 1,34 | 1,80E-01 |
| TRIAP1 | 148,03 | -0,12 | 0,09 | -1,34 | 1,81E-01 |
| ASAP1 | 3584,95 | 0,08 | 0,06 | 1,34 | 1,81E-01 |
| CD300LF | 1373,10 | 0,08 | 0,06 | 1,34 | 1,81E-01 |
| PISD | 2500,25 | 0,10 | 0,08 | 1,34 | 1,82E-01 |
| FRS2 | 1179,96 | -0,05 | 0,04 | -1,34 | 1,82E-01 |
| KLHL3 | 613,23 | 0,08 | 0,06 | 1,34 | 1,82E-01 |
| ODF2L | 2418,41 | -0,08 | 0,06 | -1,33 | 1,82E-01 |
| ZFC3H1 | 5906,24 | -0,06 | 0,04 | -1,33 | 1,82E-01 |
| PSMA6 | 607,85 | -0,07 | 0,05 | -1,33 | 1,82E-01 |
| LPAR1 | 309,23 | 0,12 | 0,09 | 1,33 | 1,83E-01 |
| MTMR1 | 2012,24 | -0,04 | 0,03 | -1,33 | 1,83E-01 |
| MAP9 | 218,71 | -0,11 | 0,08 | -1,33 | 1,83E-01 |
| TCF4 | 1377,72 | -0,10 | 0,08 | -1,33 | 1,83E-01 |
| PPP1R12B | 6105,89 | -0,10 | 0,07 | -1,33 | 1,83E-01 |
| IGIP | 647,98 | -0,07 | 0,06 | -1,33 | 1,83E-01 |
| BLMH | 501,80 | -0,04 | 0,03 | -1,33 | 1,83E-01 |
| NSL1 | 1073,05 | -0,06 | 0,04 | -1,33 | 1,83E-01 |
| RENBP | 493,72 | 0,08 | 0,06 | 1,33 | 1,84E-01 |
| AKT1 | 3558,07 | 0,06 | 0,05 | 1,33 | 1,84E-01 |
| RHEB | 830,40 | -0,04 | 0,03 | -1,33 | 1,84E-01 |
| CPNE8 | 621,62 | -0,07 | 0,06 | -1,33 | 1,84E-01 |
| KIF24 | 57,80 | 0,11 | 0,08 | 1,33 | 1,84E-01 |
| IL1RAP | 4802,86 | -0,09 | 0,07 | -1,33 | 1,84E-01 |
| BCL10 | 1284,85 | -0,05 | 0,03 | -1,33 | 1,84E-01 |
| FMN1 | 686,40 | 0,22 | 0,17 | 1,33 | 1,84E-01 |
| MAGED2 | 1095,79 | -0,05 | 0,04 | -1,33 | 1,85E-01 |
| SMIM27 | 187,70 | 0,07 | 0,05 | 1,33 | 1,85E-01 |
| TRAPPC12 | 2413,16 | 0,06 | 0,04 | 1,33 | 1,85E-01 |
| CLEC1A | 102,15 | 0,11 | 0,08 | 1,33 | 1,85E-01 |

| | | | | | |
|---|---|---|---|---|---|
| PSMB3 | 2813,80 | -0,07 | 0,05 | -1,33 | 1,85E-01 |
| FBXW8 | 352,94 | 0,07 | 0,06 | 1,33 | 1,85E-01 |
| ABCC5 | 2929,52 | -0,08 | 0,06 | -1,33 | 1,85E-01 |
| RMC1 | 1363,61 | 0,04 | 0,03 | 1,32 | 1,85E-01 |
| REEP5 | 1661,50 | -0,07 | 0,05 | -1,32 | 1,86E-01 |
| DPP7 | 721,60 | 0,07 | 0,05 | 1,32 | 1,86E-01 |
| CA13 | 226,56 | 0,06 | 0,05 | 1,32 | 1,86E-01 |
| TIFA | 770,94 | -0,10 | 0,08 | -1,32 | 1,86E-01 |
| NKAP | 492,86 | -0,04 | 0,03 | -1,32 | 1,86E-01 |
| PTTG1IP | 5076,54 | 0,05 | 0,04 | 1,32 | 1,86E-01 |
| EZR | 3575,81 | -0,06 | 0,04 | -1,32 | 1,86E-01 |
| RNF7 | 789,73 | -0,05 | 0,04 | -1,32 | 1,86E-01 |
| ADK | 894,98 | -0,07 | 0,05 | -1,32 | 1,86E-01 |
| GPR89A | 371,01 | -0,06 | 0,04 | -1,32 | 1,86E-01 |
| DNAJC11 | 620,50 | 0,05 | 0,04 | 1,32 | 1,86E-01 |
| AGPAT2 | 515,53 | 0,06 | 0,05 | 1,32 | 1,87E-01 |
| ALDH6A1 | 384,00 | 0,05 | 0,04 | 1,32 | 1,87E-01 |
| SNX11 | 1232,21 | 0,06 | 0,05 | 1,32 | 1,87E-01 |
| CLUAP1 | 351,95 | 0,06 | 0,05 | 1,32 | 1,87E-01 |
| MAFB | 561,85 | 0,10 | 0,08 | 1,32 | 1,87E-01 |
| TGFBR2 | 10806,17 | -0,05 | 0,03 | -1,32 | 1,87E-01 |
| VIPAS39 | 910,67 | -0,03 | 0,02 | -1,32 | 1,87E-01 |
| IL32 | 666,67 | 0,10 | 0,08 | 1,32 | 1,87E-01 |
| UBE2E1 | 1060,35 | -0,05 | 0,04 | -1,32 | 1,87E-01 |
| TRIM14 | 1074,13 | 0,10 | 0,08 | 1,32 | 1,87E-01 |
| HACD2 | 784,83 | -0,04 | 0,03 | -1,32 | 1,87E-01 |
| PPME1 | 398,67 | 0,04 | 0,03 | 1,32 | 1,87E-01 |
| TGIF1 | 741,40 | -0,05 | 0,04 | -1,32 | 1,87E-01 |
| KLF4 | 475,53 | 0,08 | 0,06 | 1,32 | 1,87E-01 |
| UBE2W | 3402,90 | -0,08 | 0,06 | -1,32 | 1,87E-01 |
| GUCA1B | 65,60 | 0,08 | 0,06 | 1,32 | 1,87E-01 |
| ZNF781 | 79,44 | -0,12 | 0,09 | -1,32 | 1,87E-01 |
| CRTAM | 195,71 | -0,11 | 0,08 | -1,32 | 1,87E-01 |
| TSPO | 3876,66 | 0,07 | 0,05 | 1,32 | 1,88E-01 |
| RAB10 | 4109,56 | -0,05 | 0,04 | -1,32 | 1,88E-01 |
| HTD2 | 21,90 | 0,08 | 0,06 | 1,32 | 1,88E-01 |
| ADGRE2 | 3380,26 | 0,11 | 0,09 | 1,32 | 1,88E-01 |
| BARD1 | 534,86 | -0,07 | 0,05 | -1,32 | 1,88E-01 |
| IL7R | 19362,60 | -0,08 | 0,06 | -1,32 | 1,88E-01 |
| PPM1M | 1812,44 | 0,05 | 0,04 | 1,32 | 1,88E-01 |
| ENSG00000272916 | 34,39 | 0,13 | 0,10 | 1,32 | 1,88E-01 |
| ASB7 | 1118,98 | -0,04 | 0,03 | -1,32 | 1,88E-01 |
| MATK | 722,51 | -0,10 | 0,07 | -1,32 | 1,88E-01 |
| SIRPB2 | 2555,36 | 0,12 | 0,09 | 1,32 | 1,88E-01 |
| SELENOT | 2724,32 | -0,08 | 0,06 | -1,31 | 1,89E-01 |
| ATP8B2 | 3973,59 | 0,06 | 0,04 | 1,31 | 1,89E-01 |

| | | | | | |
|---|---|---|---|---|---|
| ZNF217 | 9321,28 | -0,05 | 0,04 | -1,31 | 1,89E-01 |
| POLR2C | 863,78 | -0,04 | 0,03 | -1,31 | 1,89E-01 |
| TNFSF13 | 490,83 | 0,07 | 0,05 | 1,31 | 1,89E-01 |
| ENTPD7 | 334,14 | 0,04 | 0,03 | 1,31 | 1,89E-01 |
| STMP1 | 1566,93 | 0,09 | 0,07 | 1,31 | 1,89E-01 |
| SOCS3 | 1056,11 | 0,12 | 0,09 | 1,31 | 1,89E-01 |
| KLKB1 | 67,64 | 0,14 | 0,10 | 1,31 | 1,89E-01 |
| HSBP1L1 | 135,79 | 0,08 | 0,06 | 1,31 | 1,89E-01 |
| BAD | 351,62 | 0,05 | 0,04 | 1,31 | 1,89E-01 |
| MICOS10 | 495,91 | -0,04 | 0,03 | -1,31 | 1,89E-01 |
| TSSK4 | 24,87 | 0,12 | 0,09 | 1,31 | 1,90E-01 |
| MTA2 | 2200,31 | -0,04 | 0,03 | -1,31 | 1,90E-01 |
| FOXJ3 | 2154,60 | -0,04 | 0,03 | -1,31 | 1,90E-01 |
| ZNF783 | 946,87 | 0,05 | 0,04 | 1,31 | 1,90E-01 |
| CDK5R1 | 550,88 | 0,10 | 0,07 | 1,31 | 1,90E-01 |
| NCF4 | 1725,33 | 0,10 | 0,08 | 1,31 | 1,90E-01 |
| AIP | 376,97 | 0,06 | 0,05 | 1,31 | 1,90E-01 |
| ELAPOR2 | 386,16 | -0,13 | 0,10 | -1,31 | 1,90E-01 |
| SNAPC1 | 135,34 | -0,08 | 0,06 | -1,31 | 1,90E-01 |
| MTMR12 | 1178,81 | -0,04 | 0,03 | -1,31 | 1,90E-01 |
| MACIR | 210,19 | 0,06 | 0,05 | 1,31 | 1,90E-01 |
| HLA-E | 69373,45 | -0,06 | 0,04 | -1,31 | 1,91E-01 |
| POLR3B | 487,94 | -0,05 | 0,04 | -1,31 | 1,91E-01 |
| PHACTR4 | 465,17 | 0,08 | 0,06 | 1,31 | 1,91E-01 |
| GLA | 488,25 | -0,06 | 0,05 | -1,31 | 1,91E-01 |
| GNG5 | 330,79 | 0,06 | 0,05 | 1,31 | 1,91E-01 |
| TMOD3 | 4569,91 | -0,04 | 0,03 | -1,31 | 1,91E-01 |
| RFXAP | 278,76 | -0,06 | 0,04 | -1,31 | 1,91E-01 |
| MLLT3 | 1197,26 | -0,07 | 0,06 | -1,31 | 1,91E-01 |
| SMAD3 | 1804,60 | 0,08 | 0,06 | 1,31 | 1,91E-01 |
| PIGX | 1511,93 | -0,07 | 0,05 | -1,31 | 1,91E-01 |
| ZNF800 | 2242,81 | -0,06 | 0,05 | -1,31 | 1,92E-01 |
| CILK1 | 707,10 | -0,03 | 0,02 | -1,31 | 1,92E-01 |
| EIF5A2 | 241,56 | -0,09 | 0,07 | -1,31 | 1,92E-01 |
| BSDC1 | 2708,71 | 0,04 | 0,03 | 1,31 | 1,92E-01 |
| NAPB | 989,62 | -0,05 | 0,04 | -1,30 | 1,92E-01 |
| VTI1B | 2571,94 | 0,07 | 0,06 | 1,30 | 1,92E-01 |
| ZNF569 | 263,66 | -0,08 | 0,06 | -1,30 | 1,92E-01 |
| LRRC59 | 1219,91 | -0,04 | 0,03 | -1,30 | 1,92E-01 |
| PLEKHF1 | 517,52 | -0,08 | 0,06 | -1,30 | 1,92E-01 |
| TTLL4 | 479,14 | 0,10 | 0,07 | 1,30 | 1,93E-01 |
| RIGI | 5068,82 | -0,12 | 0,09 | -1,30 | 1,93E-01 |
| DNAJB4 | 398,92 | -0,07 | 0,05 | -1,30 | 1,93E-01 |
| TRAF3IP3 | 5445,00 | -0,04 | 0,03 | -1,30 | 1,93E-01 |
| IL15 | 630,72 | 0,07 | 0,06 | 1,30 | 1,93E-01 |
| LMBR1 | 2526,76 | -0,04 | 0,03 | -1,30 | 1,93E-01 |

| | | | | | |
|---|---|---|---|---|---|
| CCL4L2 | 97,99 | 0,17 | 0,13 | 1,30 | 1,93E-01 |
| CGAS | 792,96 | 0,05 | 0,04 | 1,30 | 1,93E-01 |
| CUX1 | 1988,71 | 0,09 | 0,07 | 1,30 | 1,93E-01 |
| LEPROT | 1321,19 | 0,05 | 0,04 | 1,30 | 1,94E-01 |
| EYS | 95,52 | 0,13 | 0,10 | 1,30 | 1,94E-01 |
| MOCS3 | 473,90 | -0,05 | 0,04 | -1,30 | 1,94E-01 |
| HSD17B10 | 490,05 | -0,06 | 0,04 | -1,30 | 1,94E-01 |
| ZDHHC18 | 5873,52 | 0,10 | 0,07 | 1,30 | 1,94E-01 |
| ANKRD16 | 131,36 | 0,06 | 0,05 | 1,30 | 1,94E-01 |
| LBHD1 | 226,78 | 0,06 | 0,05 | 1,30 | 1,94E-01 |
| GPAM | 465,46 | -0,06 | 0,05 | -1,30 | 1,94E-01 |
| XRCC2 | 66,88 | 0,12 | 0,10 | 1,30 | 1,94E-01 |
| METTL17 | 623,75 | 0,04 | 0,03 | 1,30 | 1,94E-01 |
| TOMM40L | 287,45 | 0,04 | 0,03 | 1,30 | 1,94E-01 |
| RNF122 | 236,44 | 0,09 | 0,07 | 1,30 | 1,94E-01 |
| WIPF2 | 1307,75 | 0,09 | 0,07 | 1,30 | 1,94E-01 |
| ADGRE5 | 9586,09 | 0,09 | 0,07 | 1,30 | 1,94E-01 |
| CYB5D1 | 43,94 | -0,07 | 0,06 | -1,30 | 1,95E-01 |
| BIRC2 | 4637,12 | -0,09 | 0,07 | -1,30 | 1,95E-01 |
| PSTK | 131,55 | 0,08 | 0,06 | 1,30 | 1,95E-01 |
| PEAK1 | 1011,07 | -0,05 | 0,04 | -1,30 | 1,95E-01 |
| BECN1 | 2943,66 | -0,04 | 0,03 | -1,30 | 1,95E-01 |
| PITPNB | 882,02 | -0,05 | 0,04 | -1,30 | 1,95E-01 |
| HS1BP3 | 624,89 | 0,09 | 0,07 | 1,30 | 1,95E-01 |
| MB21D2 | 124,31 | 0,09 | 0,07 | 1,30 | 1,95E-01 |
| MTOR | 1908,63 | 0,06 | 0,05 | 1,30 | 1,95E-01 |
| ZRANB2 | 4083,74 | -0,09 | 0,07 | -1,30 | 1,95E-01 |
| NF2 | 837,98 | 0,06 | 0,04 | 1,29 | 1,95E-01 |
| ZNF132 | 304,29 | -0,09 | 0,07 | -1,29 | 1,95E-01 |
| LPAR6 | 1144,39 | -0,16 | 0,12 | -1,29 | 1,95E-01 |
| CFH | 264,56 | -0,16 | 0,12 | -1,29 | 1,95E-01 |
| LACTB | 1227,34 | -0,07 | 0,06 | -1,29 | 1,95E-01 |
| GPR27 | 583,21 | -0,10 | 0,08 | -1,29 | 1,96E-01 |
| FCGR3B | 43423,73 | -0,12 | 0,09 | -1,29 | 1,96E-01 |
| ZFYVE26 | 1942,18 | 0,06 | 0,04 | 1,29 | 1,96E-01 |
| RPIA | 1972,90 | 0,12 | 0,09 | 1,29 | 1,96E-01 |
| NDUFB9 | 978,48 | -0,05 | 0,04 | -1,29 | 1,96E-01 |
| NDEL1 | 7074,55 | -0,08 | 0,06 | -1,29 | 1,96E-01 |
| RFK | 425,75 | -0,07 | 0,05 | -1,29 | 1,96E-01 |
| CRELD1 | 341,13 | 0,05 | 0,04 | 1,29 | 1,96E-01 |
| KDM5C | 3335,67 | 0,05 | 0,04 | 1,29 | 1,96E-01 |
| SLC35B1 | 501,53 | 0,03 | 0,02 | 1,29 | 1,96E-01 |
| IFT43 | 166,87 | 0,05 | 0,04 | 1,29 | 1,97E-01 |
| ALG9 | 206,22 | 0,06 | 0,04 | 1,29 | 1,97E-01 |
| LY6G5B | 75,19 | 0,13 | 0,10 | 1,29 | 1,97E-01 |
| ZFYVE1 | 726,73 | 0,05 | 0,04 | 1,29 | 1,97E-01 |

| | | | | | |
|---|---|---|---|---|---|
| CD8A | 1676,48 | -0,14 | 0,11 | -1,29 | 1,97E-01 |
| DYNLT2 | 92,30 | -0,10 | 0,07 | -1,29 | 1,97E-01 |
| TMCO3 | 2035,99 | -0,06 | 0,04 | -1,29 | 1,97E-01 |
| TRIOBP | 1730,58 | 0,08 | 0,06 | 1,29 | 1,97E-01 |
| EML2 | 696,89 | 0,04 | 0,03 | 1,29 | 1,97E-01 |
| ACOX3 | 514,51 | 0,04 | 0,03 | 1,29 | 1,97E-01 |
| ZNF71 | 290,85 | 0,05 | 0,04 | 1,29 | 1,97E-01 |
| TTC16 | 303,75 | 0,14 | 0,10 | 1,29 | 1,97E-01 |
| SLC30A1 | 1243,38 | -0,10 | 0,08 | -1,29 | 1,97E-01 |
| UBE2H | 6460,45 | 0,09 | 0,07 | 1,29 | 1,97E-01 |
| LRRC28 | 553,83 | 0,05 | 0,04 | 1,29 | 1,97E-01 |
| KCNN4 | 281,47 | 0,11 | 0,08 | 1,29 | 1,97E-01 |
| NDUFV2 | 465,50 | -0,04 | 0,03 | -1,29 | 1,98E-01 |
| ELP5 | 264,92 | -0,04 | 0,03 | -1,29 | 1,98E-01 |
| RP2 | 3431,61 | -0,08 | 0,06 | -1,29 | 1,98E-01 |
| ST13 | 1961,50 | 0,07 | 0,06 | 1,29 | 1,98E-01 |
| ZNF583 | 224,57 | -0,10 | 0,08 | -1,29 | 1,98E-01 |
| SASH3 | 3159,76 | 0,06 | 0,04 | 1,29 | 1,98E-01 |
| TIA1 | 4057,11 | -0,07 | 0,05 | -1,29 | 1,98E-01 |
| LEF1 | 3168,10 | 0,09 | 0,07 | 1,29 | 1,98E-01 |
| RPL30 | 9048,03 | -0,13 | 0,10 | -1,29 | 1,98E-01 |
| PFKFB2 | 1059,29 | 0,08 | 0,06 | 1,29 | 1,98E-01 |
| NFE2L3 | 276,91 | -0,09 | 0,07 | -1,29 | 1,99E-01 |
| RAP2C | 1466,88 | -0,08 | 0,07 | -1,29 | 1,99E-01 |
| CHST10 | 93,76 | 0,13 | 0,10 | 1,29 | 1,99E-01 |
| MED31 | 472,24 | -0,06 | 0,05 | -1,29 | 1,98E-01 |
| NINJ1 | 939,54 | 0,11 | 0,08 | 1,29 | 1,99E-01 |
| GARRE1 | 1069,49 | 0,05 | 0,04 | 1,29 | 1,99E-01 |
| CTSF | 120,05 | 0,08 | 0,06 | 1,29 | 1,99E-01 |
| YWHAH | 412,85 | 0,07 | 0,05 | 1,28 | 1,99E-01 |
| GPR84 | 30,11 | 0,15 | 0,11 | 1,28 | 1,99E-01 |
| MAP7D3 | 311,49 | -0,05 | 0,04 | -1,28 | 1,99E-01 |
| BAG4 | 845,30 | -0,06 | 0,05 | -1,28 | 1,99E-01 |
| ANKS1A | 1125,77 | 0,07 | 0,06 | 1,28 | 1,99E-01 |
| TMED7 | 1827,94 | -0,09 | 0,07 | -1,28 | 1,99E-01 |
| LUC7L | 1396,45 | 0,05 | 0,04 | 1,28 | 2,00E-01 |
| CMKLR1 | 561,70 | -0,10 | 0,08 | -1,28 | 2,00E-01 |
| RAPGEF1 | 2387,25 | 0,11 | 0,09 | 1,28 | 2,00E-01 |
| WIPF1 | 12724,19 | -0,05 | 0,04 | -1,28 | 2,00E-01 |
| RPS6KA1 | 5117,74 | 0,06 | 0,05 | 1,28 | 2,00E-01 |
| ADAM17 | 3038,83 | -0,05 | 0,04 | -1,28 | 2,00E-01 |
| DR1 | 4244,93 | -0,05 | 0,04 | -1,28 | 2,00E-01 |
| ZBTB40 | 2054,83 | 0,05 | 0,04 | 1,28 | 2,00E-01 |
| VAMP1 | 1011,74 | 0,06 | 0,05 | 1,28 | 2,01E-01 |
| ITPK1 | 2039,99 | 0,10 | 0,08 | 1,28 | 2,01E-01 |
| CPLANE2 | 61,42 | 0,08 | 0,06 | 1,28 | 2,01E-01 |

| | | | | | |
|---|---|---|---|---|---|
| RTL10 | 454,81 | 0,06 | 0,05 | 1,28 | 2,02E-01 |
| JAM3 | 437,63 | 0,12 | 0,10 | 1,28 | 2,02E-01 |
| TNFSF10 | 5715,36 | -0,10 | 0,08 | -1,28 | 2,02E-01 |
| TOGARAM2 | 337,08 | 0,12 | 0,09 | 1,28 | 2,02E-01 |
| COL19A1 | 677,05 | -0,13 | 0,10 | -1,28 | 2,02E-01 |
| CSF2RB | 8131,96 | 0,10 | 0,08 | 1,28 | 2,02E-01 |
| SAMD4B | 758,23 | 0,11 | 0,09 | 1,27 | 2,02E-01 |
| ZNF691 | 286,80 | -0,03 | 0,03 | -1,27 | 2,02E-01 |
| TMEM38B | 305,30 | -0,07 | 0,05 | -1,27 | 2,03E-01 |
| NIPA1 | 612,25 | -0,05 | 0,04 | -1,27 | 2,03E-01 |
| FXR2 | 595,24 | 0,05 | 0,04 | 1,27 | 2,03E-01 |
| PPP2R2D | 1984,91 | -0,04 | 0,03 | -1,27 | 2,03E-01 |
| DCTN1 | 1591,85 | 0,06 | 0,05 | 1,27 | 2,04E-01 |
| ENSG00000273748 | 427,91 | -0,16 | 0,13 | -1,27 | 2,04E-01 |
| TYSND1 | 655,17 | 0,05 | 0,04 | 1,27 | 2,04E-01 |
| TAGLN2 | 12625,33 | 0,07 | 0,06 | 1,27 | 2,04E-01 |
| ZNF746 | 2414,82 | 0,09 | 0,07 | 1,27 | 2,04E-01 |
| ZNF554 | 175,82 | 0,06 | 0,04 | 1,27 | 2,05E-01 |
| HAUS7 | 63,96 | 0,10 | 0,07 | 1,27 | 2,05E-01 |
| ATOX1 | 569,95 | 0,05 | 0,04 | 1,27 | 2,05E-01 |
| ANKRD23 | 93,32 | 0,06 | 0,04 | 1,27 | 2,05E-01 |
| ARMH2 | 60,24 | -0,08 | 0,06 | -1,27 | 2,05E-01 |
| NCOA2 | 8251,57 | -0,05 | 0,04 | -1,27 | 2,05E-01 |
| PRR4 | 25,99 | 0,12 | 0,10 | 1,27 | 2,05E-01 |
| FNBP1L | 281,47 | 0,08 | 0,06 | 1,27 | 2,06E-01 |
| SMAD7 | 566,67 | 0,07 | 0,05 | 1,27 | 2,06E-01 |
| TMEM164 | 2452,48 | 0,14 | 0,11 | 1,26 | 2,06E-01 |
| ZNF415 | 43,66 | 0,17 | 0,13 | 1,26 | 2,06E-01 |
| ORMDL2 | 682,48 | -0,03 | 0,02 | -1,26 | 2,06E-01 |
| STIM1 | 1934,41 | 0,07 | 0,06 | 1,26 | 2,06E-01 |
| MAD2L2 | 289,96 | 0,05 | 0,04 | 1,26 | 2,07E-01 |
| ADAT2 | 460,16 | -0,06 | 0,04 | -1,26 | 2,07E-01 |
| SNX19 | 3362,41 | -0,04 | 0,03 | -1,26 | 2,07E-01 |
| MPG | 516,13 | 0,04 | 0,03 | 1,26 | 2,07E-01 |
| TAS2R50 | 93,88 | -0,10 | 0,08 | -1,26 | 2,07E-01 |
| CD53 | 13479,26 | -0,04 | 0,03 | -1,26 | 2,07E-01 |
| SLC18A2 | 106,74 | 0,12 | 0,09 | 1,26 | 2,07E-01 |
| METTL4 | 829,13 | -0,06 | 0,05 | -1,26 | 2,07E-01 |
| SPTAN1 | 3843,30 | 0,08 | 0,06 | 1,26 | 2,07E-01 |
| BHLHE40 | 876,30 | 0,07 | 0,06 | 1,26 | 2,07E-01 |
| C19orf38 | 930,59 | 0,07 | 0,05 | 1,26 | 2,07E-01 |
| FKBP14 | 199,75 | -0,06 | 0,05 | -1,26 | 2,08E-01 |
| FBXL15 | 370,95 | 0,04 | 0,03 | 1,26 | 2,08E-01 |
| DCXR | 343,13 | 0,06 | 0,05 | 1,26 | 2,08E-01 |
| KLHL21 | 1948,86 | 0,09 | 0,07 | 1,26 | 2,08E-01 |
| BMPR1A | 462,78 | -0,07 | 0,05 | -1,26 | 2,08E-01 |

| | | | | | |
|---|---|---|---|---|---|
| PIK3R5 | 4087,54 | 0,09 | 0,07 | 1,26 | 2,08E-01 |
| STX2 | 557,14 | 0,08 | 0,06 | 1,26 | 2,08E-01 |
| CRTC3 | 1581,16 | 0,05 | 0,04 | 1,26 | 2,09E-01 |
| AFF2 | 261,93 | 0,10 | 0,08 | 1,26 | 2,09E-01 |
| GYPA | 296,86 | 0,16 | 0,13 | 1,26 | 2,09E-01 |
| TMEM121B | 192,68 | 0,14 | 0,11 | 1,26 | 2,09E-01 |
| RBM44 | 185,33 | 0,08 | 0,07 | 1,26 | 2,09E-01 |
| C5orf15 | 879,13 | -0,06 | 0,05 | -1,26 | 2,09E-01 |
| PILRA | 570,67 | 0,14 | 0,11 | 1,26 | 2,09E-01 |
| CDC37 | 2226,49 | -0,04 | 0,03 | -1,26 | 2,09E-01 |
| CCDC115 | 855,84 | -0,04 | 0,03 | -1,25 | 2,10E-01 |
| DOCK4 | 1595,97 | 0,13 | 0,10 | 1,25 | 2,10E-01 |
| ITM2B | 28412,80 | -0,06 | 0,05 | -1,25 | 2,10E-01 |
| ZNF90 | 132,77 | -0,08 | 0,07 | -1,25 | 2,10E-01 |
| LYSMD1 | 42,88 | 0,08 | 0,07 | 1,25 | 2,10E-01 |
| AFG2B | 728,87 | -0,05 | 0,04 | -1,25 | 2,10E-01 |
| TUFT1 | 164,22 | 0,08 | 0,06 | 1,25 | 2,10E-01 |
| SETMAR | 125,37 | -0,06 | 0,04 | -1,25 | 2,10E-01 |
| FCRL6 | 1082,88 | -0,13 | 0,11 | -1,25 | 2,11E-01 |
| ENPP3 | 94,92 | 0,16 | 0,13 | 1,25 | 2,11E-01 |
| WDSUB1 | 260,52 | -0,05 | 0,04 | -1,25 | 2,11E-01 |
| CC2D2B | 212,19 | 0,10 | 0,08 | 1,25 | 2,11E-01 |
| ANXA11 | 5309,52 | 0,08 | 0,07 | 1,25 | 2,11E-01 |
| ADGRB3 | 44,06 | 0,21 | 0,17 | 1,25 | 2,11E-01 |
| LSM14B | 916,12 | 0,05 | 0,04 | 1,25 | 2,11E-01 |
| CLIC1 | 8660,50 | -0,07 | 0,06 | -1,25 | 2,11E-01 |
| SLC38A9 | 1056,86 | -0,04 | 0,03 | -1,25 | 2,11E-01 |
| CHD8 | 3991,76 | -0,04 | 0,04 | -1,25 | 2,11E-01 |
| GPKOW | 445,45 | -0,04 | 0,03 | -1,25 | 2,11E-01 |
| PGAP1 | 449,11 | 0,08 | 0,06 | 1,25 | 2,11E-01 |
| UBA1 | 1987,13 | 0,09 | 0,07 | 1,25 | 2,12E-01 |
| ZSWIM5 | 135,90 | 0,10 | 0,08 | 1,25 | 2,12E-01 |
| MRM2 | 450,08 | -0,04 | 0,03 | -1,25 | 2,12E-01 |
| GAS7 | 2530,43 | 0,10 | 0,08 | 1,25 | 2,12E-01 |
| KAT8 | 1724,06 | -0,03 | 0,03 | -1,25 | 2,12E-01 |
| R3HCC1 | 442,35 | -0,03 | 0,03 | -1,25 | 2,12E-01 |
| IRAK4 | 3270,53 | -0,04 | 0,03 | -1,25 | 2,12E-01 |
| TNFAIP3 | 1071,71 | 0,07 | 0,05 | 1,25 | 2,12E-01 |
| UXT | 790,38 | -0,07 | 0,06 | -1,25 | 2,12E-01 |
| KLRC4 | 225,65 | -0,14 | 0,12 | -1,25 | 2,13E-01 |
| RGS3 | 1488,16 | 0,06 | 0,05 | 1,25 | 2,13E-01 |
| CFAP298 | 263,14 | -0,04 | 0,03 | -1,25 | 2,13E-01 |
| ZNF23 | 176,49 | 0,06 | 0,05 | 1,24 | 2,13E-01 |
| UBXN8 | 94,84 | -0,08 | 0,06 | -1,24 | 2,13E-01 |
| RPS13 | 6002,25 | -0,09 | 0,08 | -1,24 | 2,13E-01 |
| RAB24 | 793,66 | 0,07 | 0,06 | 1,24 | 2,14E-01 |

| | | | | | |
|---|---|---|---|---|---|
| RAD54L2 | 1495,58 | 0,04 | 0,04 | 1,24 | 2,14E-01 |
| SLC39A8 | 577,16 | -0,07 | 0,05 | -1,24 | 2,14E-01 |
| CASP1 | 4926,79 | -0,08 | 0,06 | -1,24 | 2,14E-01 |
| MED28 | 5289,88 | -0,03 | 0,03 | -1,24 | 2,14E-01 |
| PPP3CC | 1653,32 | -0,06 | 0,05 | -1,24 | 2,14E-01 |
| RNF4 | 2259,86 | -0,03 | 0,03 | -1,24 | 2,14E-01 |
| ITPRID2 | 3716,91 | -0,06 | 0,05 | -1,24 | 2,15E-01 |
| JUNB | 4229,50 | 0,07 | 0,06 | 1,24 | 2,15E-01 |
| DGAT1 | 865,13 | 0,06 | 0,05 | 1,24 | 2,15E-01 |
| ATG2B | 3519,61 | -0,04 | 0,03 | -1,24 | 2,15E-01 |
| MC1R | 221,79 | 0,08 | 0,06 | 1,24 | 2,15E-01 |
| GRWD1 | 309,98 | 0,06 | 0,05 | 1,24 | 2,15E-01 |
| MCUB | 1463,65 | -0,05 | 0,04 | -1,24 | 2,15E-01 |
| TSPAN31 | 511,13 | -0,03 | 0,03 | -1,24 | 2,15E-01 |
| EP300 | 5711,94 | 0,06 | 0,05 | 1,24 | 2,15E-01 |
| TYMP | 5885,13 | 0,10 | 0,08 | 1,24 | 2,16E-01 |
| HYCC2 | 9205,06 | -0,08 | 0,07 | -1,24 | 2,16E-01 |
| SQLE | 207,98 | 0,07 | 0,06 | 1,24 | 2,16E-01 |
| IVD | 864,43 | 0,06 | 0,05 | 1,24 | 2,16E-01 |
| EPSTI1 | 1945,23 | 0,18 | 0,15 | 1,24 | 2,16E-01 |
| STMN1 | 345,10 | -0,08 | 0,06 | -1,24 | 2,16E-01 |
| FCER1A | 469,17 | -0,16 | 0,13 | -1,24 | 2,16E-01 |
| GAN | 972,15 | -0,03 | 0,03 | -1,24 | 2,16E-01 |
| B3GLCT | 250,87 | -0,05 | 0,04 | -1,24 | 2,16E-01 |
| KLF9 | 990,53 | -0,07 | 0,05 | -1,24 | 2,17E-01 |
| ABCC4 | 1349,17 | 0,09 | 0,07 | 1,24 | 2,17E-01 |
| YBEY | 224,01 | -0,07 | 0,05 | -1,23 | 2,17E-01 |
| TRIM16 | 202,93 | 0,06 | 0,05 | 1,23 | 2,17E-01 |
| MAD1L1 | 1409,82 | 0,05 | 0,04 | 1,23 | 2,17E-01 |
| BABAM2 | 616,82 | -0,05 | 0,04 | -1,23 | 2,17E-01 |
| RBM3 | 4270,69 | -0,06 | 0,05 | -1,23 | 2,17E-01 |
| TENT5A | 1961,21 | -0,07 | 0,06 | -1,23 | 2,17E-01 |
| UBL4A | 646,01 | 0,05 | 0,04 | 1,23 | 2,17E-01 |
| SMG5 | 1195,65 | 0,08 | 0,07 | 1,23 | 2,17E-01 |
| ACYP1 | 221,37 | 0,06 | 0,05 | 1,23 | 2,17E-01 |
| GALM | 466,58 | -0,07 | 0,06 | -1,23 | 2,17E-01 |
| CTBP1 | 3997,43 | -0,03 | 0,03 | -1,23 | 2,18E-01 |
| SMIM13 | 320,89 | -0,06 | 0,05 | -1,23 | 2,18E-01 |
| BTF3L4 | 1110,90 | -0,06 | 0,05 | -1,23 | 2,18E-01 |
| ST6GALNAC2 | 522,05 | 0,10 | 0,08 | 1,23 | 2,18E-01 |
| GPAT4 | 2005,56 | -0,03 | 0,02 | -1,23 | 2,19E-01 |
| PTPRO | 469,57 | -0,08 | 0,07 | -1,23 | 2,19E-01 |
| TMEM87B | 1236,55 | -0,05 | 0,04 | -1,23 | 2,19E-01 |
| DNAJC16 | 738,23 | 0,03 | 0,03 | 1,23 | 2,19E-01 |
| TRMT44 | 574,78 | 0,04 | 0,03 | 1,23 | 2,19E-01 |
| MPP7 | 1915,35 | -0,06 | 0,05 | -1,23 | 2,19E-01 |

| | | | | | |
|---|---|---|---|---|---|
| ZNF195 | 1547,65 | -0,05 | 0,04 | -1,23 | 2,19E-01 |
| MON2 | 6529,44 | -0,05 | 0,04 | -1,23 | 2,19E-01 |
| TMEM123 | 7156,09 | -0,10 | 0,08 | -1,23 | 2,19E-01 |
| CLEC17A | 187,94 | -0,13 | 0,10 | -1,23 | 2,20E-01 |
| IMPG2 | 203,46 | 0,07 | 0,06 | 1,23 | 2,20E-01 |
| RCBTB1 | 994,77 | -0,04 | 0,04 | -1,23 | 2,20E-01 |
| RAB3IP | 872,40 | -0,05 | 0,04 | -1,23 | 2,20E-01 |
| LLCFC1 | 20,49 | 0,15 | 0,12 | 1,23 | 2,20E-01 |
| KEAP1 | 517,17 | 0,04 | 0,03 | 1,23 | 2,20E-01 |
| SLFN14 | 227,56 | 0,13 | 0,11 | 1,23 | 2,20E-01 |
| BBS12 | 108,81 | -0,08 | 0,06 | -1,23 | 2,20E-01 |
| TMEM65 | 3469,03 | -0,05 | 0,04 | -1,23 | 2,20E-01 |
| OTUD5 | 1140,58 | 0,07 | 0,06 | 1,23 | 2,21E-01 |
| ARHGAP45 | 9804,51 | 0,07 | 0,06 | 1,22 | 2,21E-01 |
| PRICKLE1 | 190,32 | 0,10 | 0,08 | 1,22 | 2,21E-01 |
| GPR180 | 521,18 | -0,08 | 0,06 | -1,22 | 2,21E-01 |
| MRPL17 | 419,40 | -0,06 | 0,05 | -1,22 | 2,21E-01 |
| DEDD2 | 2520,03 | 0,06 | 0,05 | 1,22 | 2,21E-01 |
| NEMP2 | 754,02 | -0,06 | 0,05 | -1,22 | 2,21E-01 |
| COX16 | 67,83 | -0,06 | 0,05 | -1,22 | 2,22E-01 |
| GNPNAT1 | 295,81 | -0,06 | 0,05 | -1,22 | 2,22E-01 |
| POLR2M | 656,21 | -0,08 | 0,06 | -1,22 | 2,22E-01 |
| APPL2 | 1798,59 | -0,05 | 0,04 | -1,22 | 2,22E-01 |
| ATL3 | 2126,13 | -0,04 | 0,03 | -1,22 | 2,22E-01 |
| EDAR | 167,38 | 0,13 | 0,11 | 1,22 | 2,22E-01 |
| XRN1 | 8214,16 | -0,07 | 0,06 | -1,22 | 2,22E-01 |
| ERCC4 | 544,22 | -0,03 | 0,03 | -1,22 | 2,22E-01 |
| GOLGA1 | 771,96 | 0,05 | 0,04 | 1,22 | 2,22E-01 |
| NDUFB4 | 825,37 | -0,05 | 0,04 | -1,22 | 2,22E-01 |
| HMOX1 | 316,38 | 0,09 | 0,07 | 1,22 | 2,22E-01 |
| FMO4 | 140,37 | -0,05 | 0,04 | -1,22 | 2,22E-01 |
| GNB4 | 3746,79 | -0,08 | 0,06 | -1,22 | 2,22E-01 |
| SNCA | 35143,78 | 0,14 | 0,12 | 1,22 | 2,22E-01 |
| EPPK1 | 269,96 | 0,13 | 0,10 | 1,22 | 2,22E-01 |
| AMMECR1L | 714,78 | 0,04 | 0,03 | 1,22 | 2,22E-01 |
| FBH1 | 1856,69 | 0,04 | 0,04 | 1,22 | 2,22E-01 |
| IFT81 | 82,74 | 0,10 | 0,08 | 1,22 | 2,23E-01 |
| PHC2 | 2394,51 | 0,12 | 0,10 | 1,22 | 2,23E-01 |
| SLC15A2 | 665,60 | 0,07 | 0,06 | 1,22 | 2,23E-01 |
| WWP2 | 2613,98 | 0,08 | 0,07 | 1,22 | 2,24E-01 |
| ENSG00000260007 | 64,89 | 0,07 | 0,06 | 1,22 | 2,24E-01 |
| NRBP1 | 1182,71 | 0,05 | 0,04 | 1,22 | 2,24E-01 |
| ADGRG1 | 675,73 | 0,14 | 0,12 | 1,22 | 2,24E-01 |
| DDX19A | 315,71 | 0,04 | 0,03 | 1,22 | 2,24E-01 |
| ALDH5A1 | 723,49 | 0,10 | 0,08 | 1,22 | 2,24E-01 |
| VNN1 | 751,99 | -0,14 | 0,11 | -1,22 | 2,24E-01 |

| | | | | | |
|---|---|---|---|---|---|
| ZNF660 | 69,03 | 0,10 | 0,09 | 1,21 | 2,24E-01 |
| MCL1 | 60466,77 | -0,06 | 0,05 | -1,21 | 2,24E-01 |
| NEB | 319,01 | 0,13 | 0,11 | 1,21 | 2,25E-01 |
| TRAM1 | 4166,22 | -0,06 | 0,05 | -1,21 | 2,25E-01 |
| KCTD10 | 795,35 | 0,05 | 0,04 | 1,21 | 2,25E-01 |
| IDNK | 276,87 | -0,04 | 0,04 | -1,21 | 2,25E-01 |
| ZNF566 | 339,44 | -0,06 | 0,05 | -1,21 | 2,25E-01 |
| DENND6A | 2149,75 | -0,04 | 0,03 | -1,21 | 2,25E-01 |
| CDK5 | 151,04 | 0,05 | 0,04 | 1,21 | 2,25E-01 |
| F2RL1 | 970,20 | -0,10 | 0,08 | -1,21 | 2,26E-01 |
| CRY1 | 370,59 | -0,04 | 0,04 | -1,21 | 2,26E-01 |
| MAPK1 | 12484,40 | -0,05 | 0,04 | -1,21 | 2,26E-01 |
| IKBKB | 2984,23 | 0,06 | 0,05 | 1,21 | 2,26E-01 |
| SH3BGRL | 4111,26 | -0,06 | 0,05 | -1,21 | 2,26E-01 |
| DDRGK1 | 731,16 | -0,03 | 0,03 | -1,21 | 2,26E-01 |
| TIMM10 | 236,49 | -0,09 | 0,08 | -1,21 | 2,26E-01 |
| POGK | 1376,09 | -0,03 | 0,02 | -1,21 | 2,26E-01 |
| FAM241A | 441,72 | 0,06 | 0,05 | 1,21 | 2,26E-01 |
| MCU | 819,71 | 0,04 | 0,04 | 1,21 | 2,26E-01 |
| PTBP2 | 1545,81 | -0,07 | 0,05 | -1,21 | 2,27E-01 |
| CHTOP | 3120,92 | -0,03 | 0,03 | -1,21 | 2,27E-01 |
| HERC2 | 2184,68 | 0,07 | 0,06 | 1,21 | 2,27E-01 |
| FAM221A | 185,81 | 0,05 | 0,04 | 1,21 | 2,27E-01 |
| NCAM1 | 526,84 | -0,11 | 0,09 | -1,21 | 2,27E-01 |
| SLC9A8 | 1426,53 | 0,08 | 0,07 | 1,21 | 2,27E-01 |
| RHEBL1 | 43,93 | 0,09 | 0,07 | 1,21 | 2,27E-01 |
| KLHL2 | 3306,99 | -0,07 | 0,06 | -1,21 | 2,27E-01 |
| ZNF706 | 1636,16 | -0,05 | 0,04 | -1,21 | 2,27E-01 |
| MRPL45 | 196,28 | -0,05 | 0,04 | -1,21 | 2,28E-01 |
| EBNA1BP2 | 367,07 | -0,05 | 0,04 | -1,21 | 2,28E-01 |
| UQCC1 | 699,49 | 0,04 | 0,04 | 1,21 | 2,28E-01 |
| MECP2 | 2565,18 | 0,09 | 0,07 | 1,21 | 2,28E-01 |
| P2RX5 | 160,83 | 0,11 | 0,09 | 1,21 | 2,28E-01 |
| TAF8 | 1606,86 | 0,04 | 0,03 | 1,21 | 2,28E-01 |
| RPL9 | 4731,77 | -0,17 | 0,14 | -1,21 | 2,28E-01 |
| ZNF823 | 102,68 | -0,08 | 0,06 | -1,20 | 2,28E-01 |
| PPA2 | 1192,01 | -0,05 | 0,04 | -1,20 | 2,28E-01 |
| MRPS5 | 1273,33 | -0,03 | 0,02 | -1,20 | 2,29E-01 |
| ZNF34 | 161,65 | -0,04 | 0,04 | -1,20 | 2,29E-01 |
| TMSB4X | 73267,17 | -0,07 | 0,06 | -1,20 | 2,29E-01 |
| SLC11A1 | 6974,08 | 0,11 | 0,09 | 1,20 | 2,29E-01 |
| MRPL57 | 414,18 | -0,06 | 0,05 | -1,20 | 2,29E-01 |
| EMC9 | 89,59 | 0,06 | 0,05 | 1,20 | 2,29E-01 |
| STEAP4 | 12851,63 | -0,09 | 0,08 | -1,20 | 2,29E-01 |
| NEXMIF | 85,69 | 0,12 | 0,10 | 1,20 | 2,29E-01 |
| VHL | 2967,11 | -0,03 | 0,02 | -1,20 | 2,29E-01 |

| | | | | | |
|---|---|---|---|---|---|
| TBC1D7 | 552,33 | -0,06 | 0,05 | -1,20 | 2,29E-01 |
| MCF2 | 61,60 | 0,11 | 0,09 | 1,20 | 2,29E-01 |
| SERINC5 | 3204,25 | 0,07 | 0,06 | 1,20 | 2,29E-01 |
| FBXW4 | 1512,14 | 0,04 | 0,03 | 1,20 | 2,29E-01 |
| BASP1 | 6762,37 | -0,10 | 0,08 | -1,20 | 2,29E-01 |
| PNOC | 34,83 | 0,13 | 0,11 | 1,20 | 2,29E-01 |
| PHACTR1 | 470,39 | -0,09 | 0,08 | -1,20 | 2,29E-01 |
| PYGL | 8480,21 | -0,09 | 0,07 | -1,20 | 2,30E-01 |
| MON1B | 1182,87 | 0,05 | 0,04 | 1,20 | 2,30E-01 |
| ZNF263 | 1011,39 | -0,03 | 0,02 | -1,20 | 2,30E-01 |
| ENSG00000263464 | 72,81 | -0,13 | 0,11 | -1,20 | 2,30E-01 |
| HIPK2 | 1696,79 | 0,10 | 0,08 | 1,20 | 2,30E-01 |
| FANCF | 313,02 | -0,06 | 0,05 | -1,20 | 2,30E-01 |
| VPS26B | 1339,80 | 0,04 | 0,04 | 1,20 | 2,30E-01 |
| LASP1 | 3786,52 | 0,09 | 0,07 | 1,20 | 2,30E-01 |
| TNFSF8 | 1102,32 | -0,05 | 0,04 | -1,20 | 2,31E-01 |
| FOXO1 | 2140,28 | -0,04 | 0,04 | -1,20 | 2,31E-01 |
| F11R | 745,45 | 0,10 | 0,09 | 1,20 | 2,31E-01 |
| IL21R | 668,86 | 0,06 | 0,05 | 1,20 | 2,31E-01 |
| TEX30 | 150,97 | -0,08 | 0,07 | -1,20 | 2,31E-01 |
| COX5B | 1835,63 | -0,06 | 0,05 | -1,20 | 2,31E-01 |
| UBE2Z | 1328,89 | 0,06 | 0,05 | 1,20 | 2,32E-01 |
| RAB37 | 1827,42 | 0,06 | 0,05 | 1,20 | 2,32E-01 |
| CASS4 | 1659,18 | -0,07 | 0,06 | -1,20 | 2,32E-01 |
| ATP6V0E1 | 3596,96 | -0,07 | 0,06 | -1,20 | 2,32E-01 |
| ZNF564 | 143,80 | 0,06 | 0,05 | 1,20 | 2,32E-01 |
| KCTD17 | 151,20 | 0,05 | 0,05 | 1,20 | 2,32E-01 |
| GMEB1 | 737,23 | -0,03 | 0,02 | -1,20 | 2,32E-01 |
| HSDL1 | 642,07 | -0,04 | 0,03 | -1,19 | 2,32E-01 |
| BIN2 | 6509,70 | -0,04 | 0,03 | -1,19 | 2,32E-01 |
| COTL1 | 5125,29 | 0,09 | 0,07 | 1,19 | 2,32E-01 |
| ECHDC3 | 189,83 | 0,11 | 0,09 | 1,19 | 2,32E-01 |
| BID | 2483,56 | 0,05 | 0,04 | 1,19 | 2,32E-01 |
| NAGA | 987,54 | 0,05 | 0,05 | 1,19 | 2,33E-01 |
| MBOAT1 | 1181,03 | 0,05 | 0,04 | 1,19 | 2,33E-01 |
| XPR1 | 1587,77 | -0,03 | 0,03 | -1,19 | 2,33E-01 |
| RNF24 | 4555,83 | 0,12 | 0,10 | 1,19 | 2,33E-01 |
| CTPS1 | 692,56 | 0,05 | 0,04 | 1,19 | 2,33E-01 |
| USP22 | 1985,83 | 0,07 | 0,06 | 1,19 | 2,33E-01 |
| IDO1 | 215,95 | 0,18 | 0,15 | 1,19 | 2,33E-01 |
| TSPAN2 | 1247,46 | -0,09 | 0,08 | -1,19 | 2,33E-01 |
| FAM98A | 518,47 | -0,04 | 0,04 | -1,19 | 2,34E-01 |
| SLC25A29 | 620,74 | 0,11 | 0,09 | 1,19 | 2,34E-01 |
| CCDC136 | 267,61 | 0,07 | 0,06 | 1,19 | 2,34E-01 |
| EVL | 7432,25 | 0,07 | 0,06 | 1,19 | 2,34E-01 |
| PHF12 | 2082,73 | 0,09 | 0,07 | 1,19 | 2,34E-01 |

| | | | | | |
|---|---|---|---|---|---|
| IL18R1 | 1576,38 | -0,07 | 0,06 | -1,19 | 2,34E-01 |
| PGBD4 | 294,09 | -0,07 | 0,06 | -1,19 | 2,34E-01 |
| CLCN3 | 2672,38 | -0,04 | 0,04 | -1,19 | 2,34E-01 |
| FKBP2 | 472,42 | -0,04 | 0,04 | -1,19 | 2,34E-01 |
| ELAPOR1 | 1244,38 | 0,17 | 0,14 | 1,19 | 2,34E-01 |
| PRPS2 | 263,56 | -0,05 | 0,04 | -1,19 | 2,34E-01 |
| PRCP | 2709,06 | -0,05 | 0,04 | -1,19 | 2,35E-01 |
| CATSPER2 | 289,55 | 0,08 | 0,07 | 1,19 | 2,35E-01 |
| FUZ | 309,37 | 0,06 | 0,05 | 1,19 | 2,35E-01 |
| CCDC88C | 4744,87 | 0,07 | 0,06 | 1,19 | 2,35E-01 |
| VMO1 | 30,26 | -0,17 | 0,14 | -1,19 | 2,35E-01 |
| NSRP1 | 840,16 | -0,03 | 0,02 | -1,19 | 2,35E-01 |
| ZNF568 | 459,67 | -0,06 | 0,05 | -1,19 | 2,35E-01 |
| ACTR3B | 262,86 | 0,05 | 0,04 | 1,19 | 2,35E-01 |
| ID2 | 1116,62 | -0,06 | 0,05 | -1,19 | 2,36E-01 |
| SYT11 | 681,79 | -0,06 | 0,05 | -1,19 | 2,36E-01 |
| ZNF492 | 29,87 | 0,15 | 0,12 | 1,19 | 2,36E-01 |
| IAH1 | 690,25 | -0,03 | 0,03 | -1,19 | 2,36E-01 |
| NUAK2 | 1730,89 | 0,09 | 0,08 | 1,19 | 2,36E-01 |
| SLC16A7 | 2991,81 | -0,06 | 0,05 | -1,19 | 2,36E-01 |
| TEX9 | 102,50 | 0,14 | 0,12 | 1,19 | 2,36E-01 |
| MESD | 1088,88 | -0,03 | 0,03 | -1,18 | 2,36E-01 |
| PRPSAP2 | 1044,04 | -0,03 | 0,03 | -1,18 | 2,37E-01 |
| ST3GAL4 | 718,13 | 0,09 | 0,08 | 1,18 | 2,37E-01 |
| IER5 | 2110,08 | -0,05 | 0,04 | -1,18 | 2,37E-01 |
| SARS1 | 2289,13 | -0,04 | 0,03 | -1,18 | 2,37E-01 |
| NBPF3 | 405,20 | -0,09 | 0,07 | -1,18 | 2,37E-01 |
| FUOM | 227,50 | 0,06 | 0,05 | 1,18 | 2,37E-01 |
| SCML4 | 1768,94 | 0,06 | 0,05 | 1,18 | 2,37E-01 |
| KIF15 | 109,96 | 0,14 | 0,12 | 1,18 | 2,38E-01 |
| MMS22L | 974,84 | -0,05 | 0,04 | -1,18 | 2,38E-01 |
| ACTR1B | 1031,25 | 0,03 | 0,03 | 1,18 | 2,38E-01 |
| ZNHIT2 | 112,44 | 0,05 | 0,04 | 1,18 | 2,38E-01 |
| POLDIP3 | 1855,29 | 0,04 | 0,03 | 1,18 | 2,38E-01 |
| CMBL | 48,72 | 0,23 | 0,20 | 1,18 | 2,38E-01 |
| GMPPB | 552,04 | 0,04 | 0,03 | 1,18 | 2,38E-01 |
| LRR1 | 357,28 | -0,05 | 0,04 | -1,18 | 2,39E-01 |
| FIRRM | 461,49 | 0,04 | 0,03 | 1,18 | 2,39E-01 |
| CPQ | 4073,13 | -0,07 | 0,06 | -1,18 | 2,39E-01 |
| BICRAL | 1831,17 | 0,05 | 0,05 | 1,18 | 2,39E-01 |
| PTPN12 | 2916,50 | -0,06 | 0,05 | -1,18 | 2,39E-01 |
| TARS2 | 384,61 | 0,04 | 0,03 | 1,18 | 2,39E-01 |
| MAPKAPK2 | 1419,18 | 0,08 | 0,07 | 1,18 | 2,39E-01 |
| CAPS2 | 236,37 | -0,07 | 0,06 | -1,18 | 2,40E-01 |
| GATA2 | 257,10 | 0,20 | 0,17 | 1,18 | 2,40E-01 |
| CD63 | 3152,53 | -0,06 | 0,05 | -1,18 | 2,40E-01 |

| | | | | | |
|---|---|---|---|---|---|
| ZBED10P | 192,04 | 0,07 | 0,06 | 1,18 | 2,40E-01 |
| CMPK2 | 948,67 | 0,19 | 0,16 | 1,18 | 2,40E-01 |
| ARMCX2 | 86,61 | 0,09 | 0,07 | 1,17 | 2,40E-01 |
| ZXDA | 286,27 | -0,05 | 0,05 | -1,17 | 2,40E-01 |
| SPAG1 | 264,36 | -0,08 | 0,06 | -1,17 | 2,40E-01 |
| ERMP1 | 1227,70 | -0,05 | 0,04 | -1,17 | 2,40E-01 |
| COPE | 1061,50 | 0,04 | 0,04 | 1,17 | 2,40E-01 |
| TSHZ1 | 533,43 | 0,05 | 0,04 | 1,17 | 2,40E-01 |
| H2BC8 | 705,76 | -0,09 | 0,07 | -1,17 | 2,40E-01 |
| B3GNT8 | 500,71 | 0,08 | 0,07 | 1,17 | 2,41E-01 |
| HRH4 | 453,21 | 0,13 | 0,11 | 1,17 | 2,41E-01 |
| B4GALT4 | 821,28 | -0,05 | 0,04 | -1,17 | 2,41E-01 |
| GBP2 | 10782,79 | -0,08 | 0,07 | -1,17 | 2,41E-01 |
| ZNF236 | 1442,44 | 0,04 | 0,03 | 1,17 | 2,41E-01 |
| GHITM | 4913,96 | -0,06 | 0,05 | -1,17 | 2,41E-01 |
| HEXIM1 | 1368,23 | 0,05 | 0,04 | 1,17 | 2,42E-01 |
| FAM124B | 84,30 | 0,09 | 0,08 | 1,17 | 2,42E-01 |
| CYP4F3 | 6396,06 | -0,10 | 0,09 | -1,17 | 2,42E-01 |
| LYZ | 78088,20 | -0,10 | 0,09 | -1,17 | 2,42E-01 |
| ZMYM4 | 1244,01 | -0,03 | 0,03 | -1,17 | 2,42E-01 |
| DIP2A | 2971,42 | 0,08 | 0,07 | 1,17 | 2,42E-01 |
| FOXO4 | 3613,63 | 0,12 | 0,10 | 1,17 | 2,42E-01 |
| PLSCR2 | 55,20 | 0,13 | 0,11 | 1,17 | 2,42E-01 |
| MOB4 | 699,70 | -0,06 | 0,05 | -1,17 | 2,42E-01 |
| KCNE3 | 5471,65 | -0,07 | 0,06 | -1,17 | 2,42E-01 |
| ARGLU1 | 8764,36 | -0,06 | 0,05 | -1,17 | 2,42E-01 |
| SERTAD2 | 994,50 | -0,05 | 0,04 | -1,17 | 2,42E-01 |
| PNPT1 | 697,76 | -0,08 | 0,06 | -1,17 | 2,42E-01 |
| DDX28 | 335,25 | -0,03 | 0,03 | -1,17 | 2,43E-01 |
| TPI1 | 3259,55 | -0,04 | 0,04 | -1,17 | 2,43E-01 |
| LY9 | 906,63 | 0,07 | 0,06 | 1,17 | 2,43E-01 |
| CAPN12 | 150,17 | 0,10 | 0,08 | 1,17 | 2,43E-01 |
| ENSG00000241489 | 150,28 | 0,10 | 0,09 | 1,17 | 2,43E-01 |
| LARS2 | 826,78 | -0,04 | 0,03 | -1,17 | 2,43E-01 |
| STN1 | 1195,15 | -0,04 | 0,04 | -1,17 | 2,43E-01 |
| CD200 | 152,23 | -0,13 | 0,11 | -1,17 | 2,43E-01 |
| ATP6V1B2 | 15994,09 | -0,06 | 0,05 | -1,17 | 2,43E-01 |
| PI4K2B | 491,16 | -0,09 | 0,07 | -1,17 | 2,44E-01 |
| FAM53C | 2183,74 | 0,09 | 0,07 | 1,17 | 2,44E-01 |
| TMEM120A | 443,62 | 0,06 | 0,05 | 1,17 | 2,44E-01 |
| FNTB | 207,30 | 0,05 | 0,04 | 1,17 | 2,44E-01 |
| NUDT3 | 2267,60 | -0,04 | 0,04 | -1,17 | 2,44E-01 |
| COX14 | 312,46 | -0,06 | 0,05 | -1,16 | 2,44E-01 |
| PRKCE | 842,42 | 0,09 | 0,07 | 1,17 | 2,44E-01 |
| ACSS3 | 829,48 | -0,09 | 0,08 | -1,16 | 2,44E-01 |
| ATG13 | 1261,46 | 0,05 | 0,04 | 1,16 | 2,44E-01 |

| | | | | | |
|---|---|---|---|---|---|
| NEIL3 | 273,11 | 0,15 | 0,13 | 1,16 | 2,44E-01 |
| PCSK7 | 4638,38 | 0,05 | 0,04 | 1,16 | 2,44E-01 |
| TERF2IP | 3014,69 | -0,05 | 0,04 | -1,16 | 2,45E-01 |
| FAS | 2464,21 | -0,08 | 0,07 | -1,16 | 2,45E-01 |
| YIF1A | 331,05 | 0,04 | 0,03 | 1,16 | 2,45E-01 |
| RAB4A | 813,38 | -0,04 | 0,04 | -1,16 | 2,45E-01 |
| NLRC4 | 1684,02 | -0,07 | 0,06 | -1,16 | 2,45E-01 |
| RNF213 | 31008,40 | 0,11 | 0,09 | 1,16 | 2,45E-01 |
| RPF2 | 222,47 | -0,06 | 0,05 | -1,16 | 2,45E-01 |
| UBE2F-SCLY | 20,38 | 0,08 | 0,07 | 1,16 | 2,45E-01 |
| ST6GALNAC3 | 268,50 | -0,07 | 0,06 | -1,16 | 2,45E-01 |
| DMAP1 | 899,24 | 0,04 | 0,03 | 1,16 | 2,45E-01 |
| DEPTOR | 45,79 | 0,11 | 0,10 | 1,16 | 2,46E-01 |
| RFTN1 | 1245,51 | -0,04 | 0,04 | -1,16 | 2,46E-01 |
| ZNF384 | 881,83 | 0,05 | 0,04 | 1,16 | 2,46E-01 |
| VGLL4 | 619,67 | 0,05 | 0,04 | 1,16 | 2,46E-01 |
| MGST3 | 880,71 | -0,04 | 0,04 | -1,16 | 2,46E-01 |
| NOL9 | 910,44 | 0,05 | 0,04 | 1,16 | 2,46E-01 |
| SSR2 | 2620,27 | -0,05 | 0,04 | -1,16 | 2,46E-01 |
| YY2 | 53,12 | -0,06 | 0,05 | -1,16 | 2,46E-01 |
| TXK | 2866,06 | -0,07 | 0,06 | -1,16 | 2,47E-01 |
| MUL1 | 516,27 | -0,03 | 0,03 | -1,16 | 2,47E-01 |
| ARV1 | 178,21 | -0,05 | 0,04 | -1,16 | 2,47E-01 |
| ORAI3 | 674,73 | 0,04 | 0,04 | 1,16 | 2,47E-01 |
| SNX10 | 3304,01 | -0,07 | 0,06 | -1,16 | 2,47E-01 |
| ZNF101 | 2766,65 | -0,05 | 0,04 | -1,16 | 2,48E-01 |
| MVB12B | 227,67 | 0,11 | 0,10 | 1,16 | 2,48E-01 |
| PRDX5 | 3893,87 | 0,11 | 0,09 | 1,16 | 2,48E-01 |
| RAB23 | 69,98 | 0,11 | 0,09 | 1,16 | 2,48E-01 |
| RPUSD3 | 489,47 | 0,04 | 0,03 | 1,16 | 2,48E-01 |
| ZDHHC5 | 1970,99 | 0,03 | 0,03 | 1,16 | 2,48E-01 |
| EIF1AD | 709,80 | -0,03 | 0,02 | -1,16 | 2,48E-01 |
| BCL11A | 2362,37 | -0,08 | 0,07 | -1,16 | 2,48E-01 |
| ASNS | 68,61 | 0,07 | 0,06 | 1,16 | 2,48E-01 |
| SP100 | 9749,26 | -0,06 | 0,05 | -1,15 | 2,48E-01 |
| MASTL | 445,93 | -0,07 | 0,06 | -1,15 | 2,48E-01 |
| ALG13 | 2346,43 | -0,05 | 0,05 | -1,15 | 2,49E-01 |
| STAT2 | 3983,25 | 0,10 | 0,09 | 1,15 | 2,49E-01 |
| LYST | 40427,37 | -0,06 | 0,05 | -1,15 | 2,49E-01 |
| CDK19 | 2492,69 | -0,06 | 0,05 | -1,15 | 2,49E-01 |
| LIMK2 | 5260,25 | 0,09 | 0,08 | 1,15 | 2,49E-01 |
| TRIM39 | 558,52 | -0,03 | 0,03 | -1,15 | 2,49E-01 |
| TRIO | 1840,42 | 0,07 | 0,06 | 1,15 | 2,49E-01 |
| NHS | 432,51 | -0,08 | 0,07 | -1,15 | 2,49E-01 |
| WDR73 | 987,49 | 0,03 | 0,02 | 1,15 | 2,49E-01 |
| FAM174B | 355,88 | 0,09 | 0,07 | 1,15 | 2,49E-01 |

| | | | | | |
|---|---|---|---|---|---|
| HUS1B | 80,50 | 0,08 | 0,07 | 1,15 | 2,49E-01 |
| PELI1 | 9588,11 | -0,07 | 0,06 | -1,15 | 2,50E-01 |
| SNX12 | 641,64 | -0,03 | 0,02 | -1,15 | 2,50E-01 |
| LILRA1 | 1573,41 | 0,08 | 0,07 | 1,15 | 2,50E-01 |
| SPN | 2629,32 | 0,07 | 0,06 | 1,15 | 2,50E-01 |
| GTF3A | 1447,10 | -0,05 | 0,04 | -1,15 | 2,50E-01 |
| SVIP | 808,77 | -0,11 | 0,09 | -1,15 | 2,50E-01 |
| TAS2R5 | 61,08 | 0,08 | 0,07 | 1,15 | 2,50E-01 |
| ZFAND3 | 2348,13 | 0,07 | 0,06 | 1,15 | 2,50E-01 |
| PPP5C | 606,14 | 0,05 | 0,05 | 1,15 | 2,50E-01 |
| SPIRE1 | 163,52 | 0,08 | 0,07 | 1,15 | 2,50E-01 |
| ZBTB16 | 444,71 | 0,11 | 0,10 | 1,15 | 2,50E-01 |
| CA5B | 1281,42 | -0,04 | 0,03 | -1,15 | 2,51E-01 |
| HLA-DMA | 933,95 | -0,06 | 0,05 | -1,15 | 2,51E-01 |
| CLNK | 83,40 | 0,20 | 0,18 | 1,15 | 2,51E-01 |
| DEF8 | 3472,00 | 0,07 | 0,06 | 1,15 | 2,51E-01 |
| PGS1 | 4391,91 | 0,07 | 0,06 | 1,15 | 2,51E-01 |
| UBAP2L | 1694,29 | 0,05 | 0,04 | 1,15 | 2,51E-01 |
| ARL13B | 252,24 | -0,04 | 0,04 | -1,15 | 2,51E-01 |
| TRIT1 | 589,48 | -0,04 | 0,03 | -1,15 | 2,51E-01 |
| MICU1 | 1086,73 | -0,05 | 0,04 | -1,15 | 2,52E-01 |
| RPS14 | 23587,67 | -0,10 | 0,09 | -1,15 | 2,52E-01 |
| RRP9 | 218,03 | 0,05 | 0,04 | 1,15 | 2,52E-01 |
| PHF8 | 1606,06 | 0,05 | 0,04 | 1,15 | 2,52E-01 |
| TDRD7 | 1419,99 | -0,07 | 0,06 | -1,15 | 2,52E-01 |
| PDE7B | 91,21 | 0,11 | 0,09 | 1,14 | 2,52E-01 |
| NPIPB11 | 37,73 | 0,14 | 0,12 | 1,14 | 2,52E-01 |
| SPATA2 | 210,67 | 0,07 | 0,06 | 1,14 | 2,53E-01 |
| CLCN4 | 460,47 | -0,11 | 0,09 | -1,14 | 2,53E-01 |
| TBPL1 | 936,21 | -0,05 | 0,05 | -1,14 | 2,53E-01 |
| UBTD2 | 244,72 | -0,04 | 0,03 | -1,14 | 2,53E-01 |
| MTMR2 | 458,88 | -0,05 | 0,04 | -1,14 | 2,53E-01 |
| NEFL | 84,39 | 0,15 | 0,13 | 1,14 | 2,53E-01 |
| VSIR | 11242,57 | 0,07 | 0,07 | 1,14 | 2,53E-01 |
| BET1L | 496,34 | 0,08 | 0,07 | 1,14 | 2,54E-01 |
| COPG1 | 2356,29 | -0,04 | 0,03 | -1,14 | 2,54E-01 |
| MAP3K3 | 3233,10 | 0,08 | 0,07 | 1,14 | 2,54E-01 |
| KLF10 | 328,35 | -0,06 | 0,06 | -1,14 | 2,54E-01 |
| LRRC75A | 242,22 | 0,08 | 0,07 | 1,14 | 2,54E-01 |
| ZNF207 | 12384,17 | -0,04 | 0,04 | -1,14 | 2,54E-01 |
| GRAMD4 | 1111,96 | 0,05 | 0,04 | 1,14 | 2,54E-01 |
| PCDHGA6 | 63,47 | 0,12 | 0,11 | 1,14 | 2,54E-01 |
| CASP7 | 616,21 | -0,05 | 0,05 | -1,14 | 2,55E-01 |
| CLEC18A | 71,86 | 0,18 | 0,16 | 1,14 | 2,55E-01 |
| CEP70 | 220,28 | -0,08 | 0,07 | -1,14 | 2,55E-01 |
| RINT1 | 554,32 | -0,03 | 0,03 | -1,14 | 2,55E-01 |

| | | | | | |
|---|---|---|---|---|---|
| AGMAT | 155,32 | 0,08 | 0,07 | 1,14 | 2,55E-01 |
| NUDT1 | 282,71 | 0,06 | 0,06 | 1,14 | 2,55E-01 |
| ENSG00000269825 | 268,91 | -0,06 | 0,05 | -1,14 | 2,55E-01 |
| RPL18 | 11960,56 | -0,06 | 0,05 | -1,14 | 2,55E-01 |
| CCDC125 | 1580,74 | -0,06 | 0,05 | -1,14 | 2,55E-01 |
| MYO1F | 15488,41 | 0,09 | 0,08 | 1,14 | 2,55E-01 |
| DAPK3 | 766,97 | 0,04 | 0,04 | 1,14 | 2,55E-01 |
| DDO | 40,16 | 0,13 | 0,12 | 1,14 | 2,56E-01 |
| SSH1 | 1485,87 | 0,08 | 0,07 | 1,14 | 2,56E-01 |
| ATG101 | 551,45 | -0,04 | 0,04 | -1,14 | 2,56E-01 |
| CCSAP | 1711,21 | -0,06 | 0,05 | -1,14 | 2,56E-01 |
| MYO1D | 300,50 | 0,08 | 0,07 | 1,13 | 2,56E-01 |
| C5AR1 | 2180,46 | 0,11 | 0,10 | 1,13 | 2,56E-01 |
| RAD50 | 148,77 | 0,04 | 0,04 | 1,13 | 2,57E-01 |
| F8 | 118,80 | 0,08 | 0,07 | 1,13 | 2,57E-01 |
| FBP1 | 241,49 | 0,06 | 0,05 | 1,13 | 2,57E-01 |
| TNFSF13B | 3510,46 | -0,10 | 0,09 | -1,13 | 2,57E-01 |
| GALNT6 | 679,20 | 0,06 | 0,05 | 1,13 | 2,57E-01 |
| RNF121 | 470,37 | 0,03 | 0,03 | 1,13 | 2,57E-01 |
| GDAP1 | 270,80 | -0,05 | 0,04 | -1,13 | 2,57E-01 |
| SFT2D2 | 4929,39 | 0,06 | 0,05 | 1,13 | 2,57E-01 |
| USP39 | 1053,98 | -0,02 | 0,02 | -1,13 | 2,57E-01 |
| PCMTD2 | 3226,10 | -0,04 | 0,04 | -1,13 | 2,57E-01 |
| ABCB4 | 279,58 | -0,11 | 0,10 | -1,13 | 2,58E-01 |
| IPPK | 536,40 | 0,03 | 0,02 | 1,13 | 2,58E-01 |
| INO80C | 207,37 | 0,04 | 0,04 | 1,13 | 2,58E-01 |
| GART | 1204,65 | -0,03 | 0,03 | -1,13 | 2,58E-01 |
| ATXN2 | 1227,91 | 0,06 | 0,05 | 1,13 | 2,58E-01 |
| ZNF2 | 228,66 | -0,03 | 0,02 | -1,13 | 2,59E-01 |
| IFNG | 37,28 | -0,16 | 0,14 | -1,13 | 2,59E-01 |
| POR | 663,21 | 0,07 | 0,06 | 1,13 | 2,59E-01 |
| CKAP2 | 434,46 | -0,06 | 0,05 | -1,13 | 2,59E-01 |
| ERMARD | 938,49 | -0,04 | 0,03 | -1,13 | 2,59E-01 |
| SNX16 | 463,36 | -0,09 | 0,08 | -1,13 | 2,59E-01 |
| S100Z | 311,14 | 0,07 | 0,06 | 1,13 | 2,59E-01 |
| CCDC137 | 257,79 | 0,04 | 0,04 | 1,13 | 2,59E-01 |
| ZDBF2 | 379,39 | -0,11 | 0,09 | -1,13 | 2,59E-01 |
| BRMS1 | 731,63 | -0,04 | 0,04 | -1,13 | 2,59E-01 |
| PABIR2 | 827,49 | -0,04 | 0,04 | -1,13 | 2,59E-01 |
| DCAF4 | 194,59 | 0,08 | 0,07 | 1,13 | 2,59E-01 |
| ZNF460 | 2749,52 | -0,04 | 0,03 | -1,13 | 2,60E-01 |
| PMVK | 232,85 | 0,05 | 0,04 | 1,13 | 2,60E-01 |
| ATP11A | 4232,30 | 0,08 | 0,07 | 1,13 | 2,60E-01 |
| CD72 | 289,48 | -0,10 | 0,09 | -1,13 | 2,60E-01 |
| TRIM2 | 190,44 | 0,09 | 0,08 | 1,13 | 2,60E-01 |
| ZFP69 | 174,61 | -0,04 | 0,04 | -1,13 | 2,60E-01 |

| | | | | | |
|---|---|---|---|---|---|
| CD163 | 1100,64 | -0,08 | 0,07 | -1,13 | 2,60E-01 |
| PALS1 | 457,09 | -0,05 | 0,04 | -1,13 | 2,60E-01 |
| ZNF738 | 291,49 | -0,04 | 0,04 | -1,13 | 2,60E-01 |
| FLII | 3604,14 | 0,06 | 0,05 | 1,13 | 2,60E-01 |
| EPHA4 | 746,53 | -0,07 | 0,06 | -1,12 | 2,61E-01 |
| CLMN | 883,28 | 0,08 | 0,07 | 1,12 | 2,61E-01 |
| ALG1 | 406,43 | 0,03 | 0,03 | 1,12 | 2,61E-01 |
| PYCARD | 1666,49 | 0,06 | 0,05 | 1,12 | 2,61E-01 |
| POU2AF1 | 367,78 | 0,12 | 0,10 | 1,12 | 2,61E-01 |
| RPUSD2 | 182,62 | -0,05 | 0,04 | -1,12 | 2,61E-01 |
| OSER1 | 1699,75 | -0,06 | 0,05 | -1,12 | 2,61E-01 |
| PRORP | 475,34 | -0,05 | 0,05 | -1,12 | 2,61E-01 |
| STAM | 967,44 | -0,04 | 0,04 | -1,12 | 2,62E-01 |
| TXNDC15 | 965,62 | -0,03 | 0,03 | -1,12 | 2,62E-01 |
| POLR2H | 402,68 | 0,03 | 0,03 | 1,12 | 2,62E-01 |
| GPRASP1 | 1212,71 | -0,07 | 0,06 | -1,12 | 2,62E-01 |
| MCM2 | 232,54 | 0,08 | 0,07 | 1,12 | 2,62E-01 |
| CD1D | 1309,89 | -0,06 | 0,06 | -1,12 | 2,62E-01 |
| SLC25A30 | 640,56 | -0,03 | 0,02 | -1,12 | 2,62E-01 |
| RABEPK | 365,86 | -0,04 | 0,03 | -1,12 | 2,63E-01 |
| USP11 | 314,59 | 0,07 | 0,06 | 1,12 | 2,63E-01 |
| GTF2F1 | 938,51 | -0,04 | 0,03 | -1,12 | 2,63E-01 |
| RAPGEF6 | 1834,36 | -0,05 | 0,04 | -1,12 | 2,63E-01 |
| NLRP1 | 5867,03 | 0,11 | 0,09 | 1,12 | 2,63E-01 |
| PRPF6 | 1807,52 | -0,04 | 0,03 | -1,12 | 2,63E-01 |
| ADCY9 | 402,77 | 0,08 | 0,07 | 1,12 | 2,63E-01 |
| SGTB | 1717,40 | -0,05 | 0,05 | -1,12 | 2,63E-01 |
| TTLL5 | 682,58 | 0,06 | 0,05 | 1,12 | 2,63E-01 |
| ZRANB1 | 2515,28 | -0,05 | 0,04 | -1,12 | 2,64E-01 |
| NELFE | 450,49 | 0,03 | 0,03 | 1,12 | 2,64E-01 |
| ZNF804A | 91,18 | 0,07 | 0,06 | 1,12 | 2,64E-01 |
| CHAC2 | 52,99 | -0,09 | 0,08 | -1,12 | 2,65E-01 |
| DAPK2 | 2531,54 | -0,09 | 0,08 | -1,12 | 2,65E-01 |
| ZNF438 | 1053,58 | -0,08 | 0,07 | -1,12 | 2,65E-01 |
| ASL | 302,59 | 0,04 | 0,04 | 1,12 | 2,65E-01 |
| ANKRD13C | 802,96 | -0,05 | 0,04 | -1,11 | 2,65E-01 |
| OXA1L | 1637,15 | 0,03 | 0,03 | 1,11 | 2,65E-01 |
| FICD | 265,84 | -0,03 | 0,03 | -1,11 | 2,65E-01 |
| C17orf67 | 231,06 | 0,04 | 0,04 | 1,11 | 2,65E-01 |
| KBTBD6 | 501,96 | -0,06 | 0,05 | -1,11 | 2,65E-01 |
| RFC2 | 449,32 | -0,03 | 0,03 | -1,11 | 2,65E-01 |
| VKORC1 | 158,97 | 0,04 | 0,04 | 1,11 | 2,66E-01 |
| MEMO1 | 642,74 | 0,04 | 0,04 | 1,11 | 2,66E-01 |
| USF3 | 2833,69 | -0,04 | 0,04 | -1,11 | 2,66E-01 |
| DCUN1D2 | 757,46 | -0,03 | 0,03 | -1,11 | 2,66E-01 |
| ARMCX6 | 306,67 | -0,05 | 0,04 | -1,11 | 2,67E-01 |

| | | | | | |
|---|---|---|---|---|---|
| NDUFS3 | 377,31 | -0,05 | 0,04 | -1,11 | 2,67E-01 |
| ACTN1 | 3916,11 | 0,09 | 0,09 | 1,11 | 2,67E-01 |
| NPIPB12 | 109,22 | 0,11 | 0,10 | 1,11 | 2,67E-01 |
| LATS1 | 1746,37 | -0,03 | 0,03 | -1,11 | 2,67E-01 |
| CDK5RAP2 | 2399,01 | -0,04 | 0,04 | -1,11 | 2,67E-01 |
| TMCC1 | 3072,62 | -0,06 | 0,06 | -1,11 | 2,67E-01 |
| BANP | 2517,57 | 0,05 | 0,05 | 1,11 | 2,67E-01 |
| JADE3 | 87,99 | 0,11 | 0,10 | 1,11 | 2,67E-01 |
| PTPMT1 | 177,80 | -0,04 | 0,03 | -1,11 | 2,68E-01 |
| COX11 | 1086,93 | -0,05 | 0,04 | -1,11 | 2,68E-01 |
| TM2D1 | 1443,31 | -0,03 | 0,03 | -1,11 | 2,68E-01 |
| SAP130 | 653,04 | 0,05 | 0,04 | 1,11 | 2,68E-01 |
| MSI2 | 2563,23 | 0,04 | 0,04 | 1,11 | 2,68E-01 |
| RFX5 | 1674,58 | -0,03 | 0,03 | -1,11 | 2,68E-01 |
| SMARCC1 | 2029,80 | -0,04 | 0,03 | -1,11 | 2,68E-01 |
| DOK1 | 1046,38 | 0,04 | 0,04 | 1,11 | 2,68E-01 |
| IMMP1L | 362,75 | -0,06 | 0,05 | -1,11 | 2,68E-01 |
| IGFLR1 | 187,77 | 0,06 | 0,05 | 1,11 | 2,68E-01 |
| DCAF4L1 | 87,96 | 0,08 | 0,07 | 1,11 | 2,69E-01 |
| TRDMT1 | 1180,15 | -0,05 | 0,04 | -1,11 | 2,69E-01 |
| PPP4R1 | 7437,51 | -0,07 | 0,06 | -1,11 | 2,69E-01 |
| BCL2L2 | 269,31 | 0,05 | 0,04 | 1,10 | 2,69E-01 |
| NUDC | 638,68 | -0,04 | 0,04 | -1,10 | 2,69E-01 |
| ZNF582 | 127,38 | -0,06 | 0,06 | -1,10 | 2,70E-01 |
| TRMT1 | 520,92 | 0,04 | 0,04 | 1,10 | 2,70E-01 |
| HNMT | 470,43 | -0,08 | 0,07 | -1,10 | 2,70E-01 |
| NIPSNAP3A | 397,18 | -0,08 | 0,07 | -1,10 | 2,70E-01 |
| ENSG00000272305 | 33,58 | 0,10 | 0,09 | 1,10 | 2,70E-01 |
| ECI1 | 340,84 | 0,04 | 0,03 | 1,10 | 2,70E-01 |
| WDR19 | 482,47 | -0,04 | 0,04 | -1,10 | 2,70E-01 |
| RTTN | 1789,49 | -0,05 | 0,04 | -1,10 | 2,70E-01 |
| ZBP1 | 1896,15 | 0,10 | 0,09 | 1,10 | 2,70E-01 |
| CCNE2 | 55,80 | 0,10 | 0,09 | 1,10 | 2,70E-01 |
| CD2AP | 1462,99 | -0,07 | 0,06 | -1,10 | 2,71E-01 |
| PROS1 | 194,79 | 0,14 | 0,13 | 1,10 | 2,71E-01 |
| SLC25A2 | 27,73 | 0,10 | 0,09 | 1,10 | 2,71E-01 |
| ZNF287 | 234,39 | -0,06 | 0,06 | -1,10 | 2,71E-01 |
| PIP4K2B | 1627,26 | -0,03 | 0,03 | -1,10 | 2,71E-01 |
| NACC2 | 1658,35 | 0,08 | 0,07 | 1,10 | 2,71E-01 |
| LIN7A | 2475,96 | -0,09 | 0,08 | -1,10 | 2,72E-01 |
| GGTA1 | 145,94 | 0,11 | 0,10 | 1,10 | 2,72E-01 |
| SERPINI1 | 168,71 | -0,08 | 0,07 | -1,10 | 2,72E-01 |
| PLEKHB2 | 4049,28 | -0,03 | 0,03 | -1,10 | 2,72E-01 |
| FAM219B | 1092,47 | 0,03 | 0,02 | 1,10 | 2,72E-01 |
| COQ6 | 505,07 | 0,03 | 0,03 | 1,10 | 2,72E-01 |
| SCARB2 | 1647,50 | -0,05 | 0,04 | -1,10 | 2,73E-01 |

| | | | | | |
|---|---|---|---|---|---|
| HUWE1 | 4836,25 | 0,06 | 0,06 | 1,10 | 2,73E-01 |
| NR2C1 | 682,48 | -0,04 | 0,04 | -1,10 | 2,73E-01 |
| PPT1 | 4458,39 | -0,07 | 0,06 | -1,10 | 2,73E-01 |
| RAB43 | 406,53 | 0,05 | 0,05 | 1,10 | 2,74E-01 |
| AP4B1 | 1240,63 | -0,03 | 0,03 | -1,09 | 2,74E-01 |
| HNRNPL | 3015,96 | -0,02 | 0,02 | -1,09 | 2,74E-01 |
| MTF1 | 1330,53 | 0,06 | 0,05 | 1,09 | 2,74E-01 |
| ZNRD2 | 67,74 | 0,05 | 0,05 | 1,09 | 2,74E-01 |
| HHAT | 225,22 | 0,07 | 0,07 | 1,09 | 2,74E-01 |
| AGPAT4 | 902,12 | -0,06 | 0,06 | -1,09 | 2,74E-01 |
| DPY19L4 | 615,74 | -0,06 | 0,06 | -1,09 | 2,74E-01 |
| IGF1R | 4351,10 | 0,08 | 0,07 | 1,09 | 2,74E-01 |
| KLHDC8B | 249,39 | 0,09 | 0,08 | 1,09 | 2,75E-01 |
| KRIT1 | 876,48 | 0,05 | 0,05 | 1,09 | 2,75E-01 |
| ZNF487 | 442,28 | -0,05 | 0,05 | -1,09 | 2,75E-01 |
| SH3RF3 | 224,69 | 0,11 | 0,10 | 1,09 | 2,75E-01 |
| ABHD16A | 296,65 | 0,04 | 0,04 | 1,09 | 2,75E-01 |
| ENSG00000285976 | 63,45 | 0,07 | 0,06 | 1,09 | 2,75E-01 |
| PSRC1 | 119,94 | 0,06 | 0,06 | 1,09 | 2,75E-01 |
| PM20D2 | 617,54 | -0,04 | 0,04 | -1,09 | 2,75E-01 |
| BMERB1 | 48,05 | 0,11 | 0,10 | 1,09 | 2,75E-01 |
| TRAK2 | 5033,48 | 0,08 | 0,08 | 1,09 | 2,75E-01 |
| TRAK1 | 1293,96 | 0,06 | 0,06 | 1,09 | 2,76E-01 |
| ZNF264 | 2869,62 | -0,03 | 0,03 | -1,09 | 2,76E-01 |
| USP9Y | 2031,98 | 0,21 | 0,19 | 1,09 | 2,76E-01 |
| MSRB1 | 3436,95 | 0,07 | 0,07 | 1,09 | 2,77E-01 |
| PMM2 | 745,27 | -0,04 | 0,04 | -1,09 | 2,77E-01 |
| TMA7B | 229,90 | 0,07 | 0,06 | 1,09 | 2,77E-01 |
| CLPTM1L | 1569,06 | 0,03 | 0,02 | 1,09 | 2,77E-01 |
| KIAA1958 | 276,02 | 0,08 | 0,08 | 1,09 | 2,77E-01 |
| GFOD1 | 1322,87 | 0,06 | 0,06 | 1,09 | 2,77E-01 |
| FOLR3 | 721,82 | 0,16 | 0,15 | 1,09 | 2,77E-01 |
| RALY | 2267,56 | 0,04 | 0,04 | 1,09 | 2,78E-01 |
| PSAP | 14316,24 | 0,08 | 0,07 | 1,08 | 2,78E-01 |
| ALMS1 | 2403,39 | -0,04 | 0,04 | -1,08 | 2,78E-01 |
| CD3G | 1615,65 | -0,06 | 0,06 | -1,08 | 2,78E-01 |
| TAMM41 | 439,63 | 0,07 | 0,06 | 1,08 | 2,78E-01 |
| MFSD8 | 1457,75 | -0,03 | 0,03 | -1,08 | 2,79E-01 |
| COX18 | 869,42 | -0,04 | 0,03 | -1,08 | 2,79E-01 |
| STYX | 1754,15 | -0,06 | 0,05 | -1,08 | 2,79E-01 |
| ARHGEF12 | 3679,77 | 0,10 | 0,09 | 1,08 | 2,79E-01 |
| ARHGAP17 | 1203,92 | 0,07 | 0,07 | 1,08 | 2,79E-01 |
| MCM4 | 262,15 | 0,06 | 0,06 | 1,08 | 2,79E-01 |
| TUBGCP5 | 589,65 | -0,04 | 0,04 | -1,08 | 2,79E-01 |
| UNG | 268,79 | -0,05 | 0,04 | -1,08 | 2,79E-01 |
| TAF3 | 371,71 | 0,04 | 0,04 | 1,08 | 2,79E-01 |

| | | | | | |
|---|---|---|---|---|---|
| NQO1 | 40,76 | 0,09 | 0,08 | 1,08 | 2,79E-01 |
| HLA-DQA2 | 149,58 | 0,33 | 0,31 | 1,08 | 2,79E-01 |
| TMEM205 | 414,53 | -0,04 | 0,04 | -1,08 | 2,80E-01 |
| CATSPERB | 240,88 | 0,07 | 0,07 | 1,08 | 2,80E-01 |
| PLXDC2 | 5509,97 | -0,06 | 0,06 | -1,08 | 2,80E-01 |
| TYW1 | 611,45 | -0,03 | 0,03 | -1,08 | 2,80E-01 |
| DSC1 | 125,72 | 0,15 | 0,13 | 1,08 | 2,80E-01 |
| HAUS1 | 253,22 | -0,06 | 0,05 | -1,08 | 2,80E-01 |
| CREBBP | 4552,69 | 0,08 | 0,08 | 1,08 | 2,80E-01 |
| MTBP | 382,55 | -0,06 | 0,05 | -1,08 | 2,80E-01 |
| BBS4 | 393,13 | -0,03 | 0,03 | -1,08 | 2,81E-01 |
| MR1 | 2065,39 | -0,04 | 0,03 | -1,08 | 2,81E-01 |
| UNC119B | 711,38 | 0,04 | 0,04 | 1,08 | 2,81E-01 |
| SERPINF1 | 73,89 | 0,10 | 0,09 | 1,08 | 2,81E-01 |
| MTRR | 1221,47 | -0,04 | 0,04 | -1,08 | 2,81E-01 |
| DDI2 | 2254,59 | -0,03 | 0,03 | -1,08 | 2,81E-01 |
| NMNAT1 | 470,44 | -0,04 | 0,04 | -1,08 | 2,82E-01 |
| TBC1D30 | 241,84 | 0,08 | 0,07 | 1,08 | 2,82E-01 |
| ALG10B | 641,20 | -0,07 | 0,07 | -1,08 | 2,82E-01 |
| CDKL1 | 498,05 | 0,06 | 0,05 | 1,08 | 2,82E-01 |
| ABHD3 | 3203,61 | -0,06 | 0,05 | -1,07 | 2,82E-01 |
| IRF2BP1 | 430,39 | 0,04 | 0,04 | 1,07 | 2,83E-01 |
| KCTD7 | 1264,81 | -0,04 | 0,04 | -1,07 | 2,83E-01 |
| ABCF3 | 839,61 | 0,03 | 0,03 | 1,07 | 2,83E-01 |
| PARPBP | 141,77 | 0,07 | 0,07 | 1,07 | 2,83E-01 |
| AIM2 | 514,98 | -0,11 | 0,10 | -1,07 | 2,83E-01 |
| CHAMP1 | 258,72 | -0,03 | 0,03 | -1,07 | 2,83E-01 |
| MRPL20 | 659,91 | -0,04 | 0,04 | -1,07 | 2,83E-01 |
| TXNL4A | 1249,96 | -0,02 | 0,02 | -1,07 | 2,83E-01 |
| PRDM2 | 3626,76 | -0,04 | 0,04 | -1,07 | 2,83E-01 |
| FAM50B | 185,69 | 0,06 | 0,05 | 1,07 | 2,83E-01 |
| ANXA2R | 562,57 | -0,06 | 0,05 | -1,07 | 2,83E-01 |
| POLD3 | 783,57 | -0,04 | 0,04 | -1,07 | 2,83E-01 |
| CRPPA | 119,99 | 0,09 | 0,08 | 1,07 | 2,83E-01 |
| STS | 269,01 | -0,06 | 0,05 | -1,07 | 2,83E-01 |
| DIS3L2 | 923,20 | 0,04 | 0,04 | 1,07 | 2,84E-01 |
| SYK | 9782,26 | -0,05 | 0,05 | -1,07 | 2,84E-01 |
| TRIM35 | 853,30 | 0,03 | 0,03 | 1,07 | 2,84E-01 |
| RMND5A | 7599,03 | -0,05 | 0,05 | -1,07 | 2,84E-01 |
| AP5B1 | 2763,54 | 0,11 | 0,10 | 1,07 | 2,84E-01 |
| CD300E | 1282,36 | 0,08 | 0,07 | 1,07 | 2,84E-01 |
| CNEP1R1 | 1471,02 | -0,05 | 0,05 | -1,07 | 2,84E-01 |
| YY1AP1 | 1960,99 | 0,04 | 0,04 | 1,07 | 2,85E-01 |
| ARFRP1 | 1405,09 | 0,04 | 0,03 | 1,07 | 2,85E-01 |
| GCHFR | 178,47 | 0,06 | 0,05 | 1,07 | 2,85E-01 |
| RTN3 | 3457,55 | -0,07 | 0,06 | -1,07 | 2,85E-01 |

| | | | | | |
|---|---|---|---|---|---|
| CPVL | 2842,86 | -0,07 | 0,06 | -1,07 | 2,85E-01 |
| NOPCHAP1 | 903,26 | 0,05 | 0,05 | 1,07 | 2,85E-01 |
| MTSS1 | 1798,24 | -0,06 | 0,05 | -1,07 | 2,85E-01 |
| ELL2 | 599,66 | 0,04 | 0,04 | 1,07 | 2,86E-01 |
| NADK | 6323,85 | 0,08 | 0,08 | 1,07 | 2,86E-01 |
| TTC12 | 856,01 | 0,05 | 0,05 | 1,07 | 2,86E-01 |
| PEX11A | 77,60 | 0,07 | 0,07 | 1,07 | 2,86E-01 |
| RHOBTB3 | 478,34 | 0,07 | 0,07 | 1,07 | 2,87E-01 |
| MRPL42 | 1355,91 | -0,04 | 0,03 | -1,07 | 2,87E-01 |
| IKZF5 | 990,85 | -0,05 | 0,05 | -1,07 | 2,87E-01 |
| GOLGA8B | 1430,42 | -0,08 | 0,08 | -1,07 | 2,87E-01 |
| SIGLEC5 | 89,87 | 0,11 | 0,10 | 1,07 | 2,87E-01 |
| TXNDC16 | 791,52 | -0,05 | 0,05 | -1,06 | 2,87E-01 |
| MAPK14 | 12270,09 | -0,06 | 0,06 | -1,06 | 2,87E-01 |
| RBM14 | 1755,96 | 0,05 | 0,05 | 1,06 | 2,87E-01 |
| SERPINA1 | 6026,12 | 0,09 | 0,08 | 1,06 | 2,87E-01 |
| ALKBH2 | 156,20 | -0,05 | 0,05 | -1,06 | 2,88E-01 |
| CPSF6 | 3110,29 | -0,07 | 0,07 | -1,06 | 2,88E-01 |
| ENSG00000284952 | 41,42 | 0,05 | 0,05 | 1,06 | 2,89E-01 |
| CHI3L1 | 2317,16 | -0,15 | 0,14 | -1,06 | 2,89E-01 |
| CD180 | 610,98 | -0,07 | 0,07 | -1,06 | 2,89E-01 |
| MED26 | 203,82 | 0,05 | 0,05 | 1,06 | 2,89E-01 |
| SDC2 | 89,29 | 0,13 | 0,12 | 1,06 | 2,89E-01 |
| CKS2 | 63,17 | -0,11 | 0,10 | -1,06 | 2,89E-01 |
| OR2L2 | 25,41 | 0,17 | 0,16 | 1,06 | 2,90E-01 |
| FAM161B | 156,12 | 0,04 | 0,04 | 1,06 | 2,90E-01 |
| RNF135 | 703,12 | 0,05 | 0,04 | 1,06 | 2,90E-01 |
| MINDY1 | 1881,80 | 0,08 | 0,08 | 1,06 | 2,90E-01 |
| DOCK2 | 16003,76 | -0,05 | 0,05 | -1,06 | 2,90E-01 |
| GNAI2 | 4531,45 | 0,07 | 0,07 | 1,06 | 2,90E-01 |
| TRIM26 | 839,60 | 0,05 | 0,05 | 1,06 | 2,90E-01 |
| CD300LB | 860,00 | 0,08 | 0,07 | 1,06 | 2,90E-01 |
| CHI3L2 | 237,80 | 0,09 | 0,08 | 1,06 | 2,90E-01 |
| GNG11 | 548,89 | 0,13 | 0,12 | 1,06 | 2,91E-01 |
| HDGFL2 | 613,61 | 0,05 | 0,04 | 1,06 | 2,91E-01 |
| NBPF1 | 564,89 | 0,08 | 0,07 | 1,06 | 2,91E-01 |
| RNFT1 | 677,44 | -0,05 | 0,05 | -1,06 | 2,91E-01 |
| IRAK3 | 6892,74 | 0,07 | 0,07 | 1,06 | 2,91E-01 |
| TP53BP1 | 1598,00 | -0,04 | 0,03 | -1,06 | 2,91E-01 |
| C1orf43 | 1356,80 | 0,04 | 0,03 | 1,06 | 2,91E-01 |
| PPP2R2B | 406,54 | -0,09 | 0,08 | -1,06 | 2,91E-01 |
| SCAF4 | 798,34 | 0,04 | 0,04 | 1,06 | 2,91E-01 |
| RAB38 | 30,74 | 0,10 | 0,09 | 1,05 | 2,91E-01 |
| KIAA2013 | 1145,87 | 0,05 | 0,04 | 1,05 | 2,92E-01 |
| AIG1 | 398,78 | 0,04 | 0,04 | 1,05 | 2,92E-01 |
| SMAP1 | 838,20 | -0,03 | 0,02 | -1,05 | 2,92E-01 |

| | | | | | |
|---|---|---|---|---|---|
| GDF7 | 148,84 | 0,17 | 0,17 | 1,05 | 2,92E-01 |
| PCDHGA5 | 45,37 | 0,11 | 0,10 | 1,05 | 2,93E-01 |
| TMEM248 | 2507,15 | -0,02 | 0,02 | -1,05 | 2,93E-01 |
| DPEP2 | 3255,67 | 0,08 | 0,08 | 1,05 | 2,93E-01 |
| TIMP2 | 850,21 | 0,11 | 0,11 | 1,05 | 2,93E-01 |
| SF3B4 | 186,00 | 0,06 | 0,06 | 1,05 | 2,94E-01 |
| ACTG1 | 21460,96 | 0,04 | 0,04 | 1,05 | 2,94E-01 |
| ERVK3-1 | 850,15 | -0,04 | 0,03 | -1,05 | 2,94E-01 |
| DPYD | 11067,66 | -0,06 | 0,05 | -1,05 | 2,94E-01 |
| SGO2 | 112,65 | -0,10 | 0,10 | -1,05 | 2,94E-01 |
| SLC35E3 | 959,69 | 0,06 | 0,06 | 1,05 | 2,94E-01 |
| EIF4EBP2 | 4143,45 | 0,05 | 0,05 | 1,05 | 2,94E-01 |
| GDAP2 | 2661,92 | -0,04 | 0,04 | -1,05 | 2,94E-01 |
| KLHL6 | 2579,24 | 0,04 | 0,03 | 1,05 | 2,95E-01 |
| C8orf33 | 554,90 | -0,03 | 0,03 | -1,05 | 2,95E-01 |
| PLA2G4A | 312,67 | -0,07 | 0,06 | -1,05 | 2,95E-01 |
| CUBN | 856,98 | 0,07 | 0,07 | 1,05 | 2,95E-01 |
| P2RX1 | 797,03 | 0,10 | 0,09 | 1,05 | 2,95E-01 |
| PSME4 | 4457,61 | -0,04 | 0,04 | -1,05 | 2,95E-01 |
| GCA | 17593,09 | -0,09 | 0,08 | -1,05 | 2,95E-01 |
| APH1B | 1251,43 | -0,03 | 0,03 | -1,05 | 2,95E-01 |
| MYC | 495,92 | 0,06 | 0,05 | 1,05 | 2,96E-01 |
| GSPT1 | 10007,24 | 0,10 | 0,10 | 1,05 | 2,96E-01 |
| ACBD6 | 1287,32 | -0,03 | 0,03 | -1,04 | 2,96E-01 |
| SNX22 | 501,95 | 0,08 | 0,08 | 1,04 | 2,96E-01 |
| PRKAR2B | 2153,27 | 0,10 | 0,10 | 1,04 | 2,96E-01 |
| THNSL2 | 69,37 | 0,19 | 0,18 | 1,04 | 2,96E-01 |
| RAB11FIP1 | 10793,20 | -0,06 | 0,06 | -1,04 | 2,97E-01 |
| MBOAT2 | 2768,16 | 0,07 | 0,06 | 1,04 | 2,97E-01 |
| PPP1CB | 8614,88 | -0,08 | 0,07 | -1,04 | 2,97E-01 |
| HPGD | 169,71 | -0,09 | 0,09 | -1,04 | 2,97E-01 |
| ESRRA | 1544,87 | 0,04 | 0,04 | 1,04 | 2,97E-01 |
| LGSN | 35,44 | 0,22 | 0,21 | 1,04 | 2,97E-01 |
| VPS53 | 1852,51 | 0,05 | 0,05 | 1,04 | 2,97E-01 |
| ALG10 | 561,75 | -0,05 | 0,05 | -1,04 | 2,97E-01 |
| CCDC152 | 145,50 | -0,09 | 0,09 | -1,04 | 2,97E-01 |
| RITA1 | 215,49 | 0,03 | 0,03 | 1,04 | 2,98E-01 |
| MLYCD | 994,13 | 0,05 | 0,05 | 1,04 | 2,98E-01 |
| FAM151B | 213,71 | 0,06 | 0,06 | 1,04 | 2,98E-01 |
| FKBP5 | 3815,44 | 0,08 | 0,08 | 1,04 | 2,98E-01 |
| PI16 | 120,42 | 0,09 | 0,09 | 1,04 | 2,98E-01 |
| NEXN | 123,47 | 0,09 | 0,09 | 1,04 | 2,98E-01 |
| PARD6B | 212,61 | -0,04 | 0,04 | -1,04 | 2,99E-01 |
| ADIPOR2 | 1551,89 | -0,03 | 0,03 | -1,04 | 2,99E-01 |
| MCM6 | 476,11 | -0,04 | 0,04 | -1,04 | 2,99E-01 |
| NDUFC1 | 338,46 | -0,04 | 0,04 | -1,04 | 2,99E-01 |

| | | | | | |
|---|---|---|---|---|---|
| BLTP3A | 797,65 | 0,05 | 0,05 | 1,04 | 2,99E-01 |
| ARPIN | 275,87 | 0,13 | 0,12 | 1,04 | 2,99E-01 |
| TPRG1 | 307,12 | -0,10 | 0,10 | -1,04 | 2,99E-01 |
| TNFRSF18 | 102,93 | -0,10 | 0,09 | -1,04 | 2,99E-01 |
| SNRPN | 306,67 | 0,04 | 0,04 | 1,04 | 3,00E-01 |
| BCOR | 642,73 | 0,05 | 0,05 | 1,04 | 3,00E-01 |
| IL13RA1 | 3219,33 | -0,06 | 0,06 | -1,04 | 3,00E-01 |
| STX4 | 990,51 | 0,04 | 0,04 | 1,04 | 3,00E-01 |
| LMAN2L | 320,83 | -0,02 | 0,02 | -1,04 | 3,00E-01 |
| TAFA1 | 86,26 | -0,09 | 0,09 | -1,04 | 3,00E-01 |
| CENPL | 204,40 | -0,04 | 0,04 | -1,04 | 3,00E-01 |
| PARP10 | 2082,95 | 0,07 | 0,07 | 1,04 | 3,01E-01 |
| RUFY2 | 714,28 | -0,03 | 0,03 | -1,03 | 3,01E-01 |
| POLR2D | 770,81 | -0,02 | 0,02 | -1,03 | 3,01E-01 |
| CPPED1 | 8009,93 | -0,06 | 0,06 | -1,03 | 3,01E-01 |
| TOP2A | 131,94 | 0,11 | 0,10 | 1,03 | 3,01E-01 |
| GPX4 | 1242,33 | 0,05 | 0,05 | 1,03 | 3,01E-01 |
| H2BC15 | 346,38 | 0,06 | 0,06 | 1,03 | 3,01E-01 |
| DAPP1 | 4758,44 | -0,06 | 0,06 | -1,03 | 3,01E-01 |
| ITGA1 | 293,45 | 0,09 | 0,09 | 1,03 | 3,01E-01 |
| RASSF5 | 5841,50 | 0,05 | 0,04 | 1,03 | 3,01E-01 |
| TAF6L | 237,74 | 0,03 | 0,03 | 1,03 | 3,01E-01 |
| CARD6 | 475,38 | -0,06 | 0,06 | -1,03 | 3,02E-01 |
| TSPAN13 | 175,33 | -0,08 | 0,08 | -1,03 | 3,02E-01 |
| ESS2 | 574,88 | 0,04 | 0,04 | 1,03 | 3,02E-01 |
| GPR75 | 100,12 | 0,06 | 0,06 | 1,03 | 3,02E-01 |
| AMN1 | 623,74 | -0,05 | 0,05 | -1,03 | 3,03E-01 |
| COG1 | 664,48 | 0,03 | 0,03 | 1,03 | 3,03E-01 |
| TOP3A | 1568,43 | 0,05 | 0,05 | 1,03 | 3,03E-01 |
| SLC24A4 | 1346,31 | 0,10 | 0,10 | 1,03 | 3,03E-01 |
| ZNF100 | 831,16 | 0,09 | 0,09 | 1,03 | 3,03E-01 |
| ZNF239 | 100,17 | -0,07 | 0,07 | -1,03 | 3,03E-01 |
| KIAA0513 | 3169,34 | 0,08 | 0,07 | 1,03 | 3,04E-01 |
| MRPL49 | 686,27 | -0,02 | 0,02 | -1,03 | 3,04E-01 |
| RGPD2 | 217,70 | -0,18 | 0,18 | -1,03 | 3,04E-01 |
| TMEM183A | 2072,99 | 0,04 | 0,04 | 1,03 | 3,04E-01 |
| ILF3 | 3707,95 | 0,04 | 0,04 | 1,03 | 3,04E-01 |
| SESN3 | 11082,81 | 0,07 | 0,07 | 1,03 | 3,05E-01 |
| NRCAM | 86,73 | 0,18 | 0,18 | 1,03 | 3,05E-01 |
| PARP15 | 2170,36 | -0,06 | 0,06 | -1,03 | 3,05E-01 |
| DNTTIP1 | 799,69 | 0,06 | 0,05 | 1,03 | 3,05E-01 |
| COPS3 | 1154,92 | -0,05 | 0,05 | -1,03 | 3,05E-01 |
| KDM4C | 4363,99 | -0,04 | 0,04 | -1,03 | 3,05E-01 |
| ASXL2 | 3169,35 | -0,03 | 0,03 | -1,03 | 3,05E-01 |
| VCF2 | 109,32 | 0,08 | 0,08 | 1,03 | 3,05E-01 |
| APOO | 70,18 | 0,05 | 0,05 | 1,03 | 3,05E-01 |

| | | | | | |
|---|---|---|---|---|---|
| JADE2 | 2451,50 | 0,05 | 0,05 | 1,02 | 3,05E-01 |
| COPA | 4869,84 | -0,04 | 0,03 | -1,02 | 3,06E-01 |
| NPTN | 1830,11 | -0,04 | 0,04 | -1,02 | 3,06E-01 |
| NAP1L4 | 5426,53 | -0,03 | 0,03 | -1,02 | 3,06E-01 |
| PDSS2 | 229,25 | 0,03 | 0,03 | 1,02 | 3,06E-01 |
| C18orf54 | 189,48 | -0,07 | 0,06 | -1,02 | 3,06E-01 |
| CATSPER1 | 99,13 | 0,06 | 0,06 | 1,02 | 3,06E-01 |
| LDLRAD4 | 1725,28 | 0,04 | 0,04 | 1,02 | 3,07E-01 |
| EID2B | 157,24 | -0,05 | 0,05 | -1,02 | 3,07E-01 |
| ELOVL6 | 210,70 | -0,06 | 0,06 | -1,02 | 3,07E-01 |
| INTS9 | 457,16 | 0,03 | 0,03 | 1,02 | 3,07E-01 |
| DRAP1 | 360,62 | 0,06 | 0,06 | 1,02 | 3,07E-01 |
| UBE2T | 37,54 | 0,08 | 0,08 | 1,02 | 3,07E-01 |
| GDE1 | 1756,40 | 0,05 | 0,05 | 1,02 | 3,07E-01 |
| CLEC12B | 298,07 | 0,14 | 0,14 | 1,02 | 3,07E-01 |
| MSL3 | 2443,53 | -0,04 | 0,04 | -1,02 | 3,07E-01 |
| TNRC6C | 1859,45 | 0,05 | 0,05 | 1,02 | 3,07E-01 |
| NDUFB1 | 780,80 | -0,05 | 0,05 | -1,02 | 3,08E-01 |
| PLD1 | 757,59 | 0,06 | 0,06 | 1,02 | 3,08E-01 |
| DNAJC5 | 3184,30 | 0,06 | 0,06 | 1,02 | 3,08E-01 |
| MTG2 | 631,19 | 0,06 | 0,06 | 1,02 | 3,08E-01 |
| BTNL8 | 752,79 | -0,09 | 0,09 | -1,02 | 3,08E-01 |
| P2RY1 | 202,46 | 0,07 | 0,07 | 1,02 | 3,08E-01 |
| LEPR | 414,58 | 0,09 | 0,09 | 1,02 | 3,08E-01 |
| ZNF717 | 325,18 | -0,04 | 0,04 | -1,02 | 3,08E-01 |
| SLC35B4 | 626,99 | -0,03 | 0,03 | -1,02 | 3,08E-01 |
| ZNF740 | 2368,37 | -0,02 | 0,02 | -1,02 | 3,08E-01 |
| ICAM3 | 8676,19 | -0,06 | 0,06 | -1,02 | 3,08E-01 |
| RAB20 | 383,14 | -0,08 | 0,08 | -1,02 | 3,09E-01 |
| CASC3 | 2533,51 | 0,07 | 0,07 | 1,02 | 3,09E-01 |
| ADO | 561,63 | -0,04 | 0,04 | -1,02 | 3,09E-01 |
| SPG11 | 8864,38 | -0,04 | 0,04 | -1,02 | 3,09E-01 |
| PAQR3 | 454,66 | -0,05 | 0,05 | -1,02 | 3,09E-01 |
| NAP1L5 | 37,78 | 0,09 | 0,09 | 1,02 | 3,09E-01 |
| PYM1 | 407,85 | 0,03 | 0,03 | 1,02 | 3,10E-01 |
| PTCH1 | 561,09 | 0,09 | 0,09 | 1,02 | 3,10E-01 |
| TMED9 | 1314,37 | -0,04 | 0,04 | -1,02 | 3,10E-01 |
| SYCP3 | 55,28 | 0,09 | 0,08 | 1,02 | 3,10E-01 |
| LANCL2 | 303,91 | -0,03 | 0,03 | -1,01 | 3,10E-01 |
| HDDC3 | 320,18 | 0,04 | 0,04 | 1,01 | 3,10E-01 |
| PAN3 | 5478,98 | -0,04 | 0,04 | -1,01 | 3,10E-01 |
| TXNDC17 | 350,58 | -0,06 | 0,06 | -1,01 | 3,11E-01 |
| IFNAR2 | 2477,71 | -0,04 | 0,04 | -1,01 | 3,11E-01 |
| KANSL3 | 1456,62 | 0,05 | 0,05 | 1,01 | 3,11E-01 |
| TMLHE | 894,56 | 0,04 | 0,04 | 1,01 | 3,11E-01 |
| DDIAS | 166,21 | 0,06 | 0,06 | 1,01 | 3,11E-01 |

| | | | | | |
|---|---|---|---|---|---|
| CD80 | 55,26 | 0,09 | 0,09 | 1,01 | 3,11E-01 |
| MSN | 6138,32 | 0,08 | 0,08 | 1,01 | 3,11E-01 |
| RPL27A | 9252,95 | -0,07 | 0,07 | -1,01 | 3,11E-01 |
| NUDT18 | 263,18 | 0,04 | 0,04 | 1,01 | 3,12E-01 |
| ERAP1 | 3520,14 | -0,03 | 0,03 | -1,01 | 3,12E-01 |
| TMEM9 | 254,44 | 0,05 | 0,05 | 1,01 | 3,12E-01 |
| ZCCHC14 | 557,19 | 0,04 | 0,03 | 1,01 | 3,12E-01 |
| EIF4G3 | 2182,84 | -0,05 | 0,05 | -1,01 | 3,12E-01 |
| HAVCR1 | 81,30 | 0,08 | 0,08 | 1,01 | 3,12E-01 |
| PACS1 | 2833,07 | 0,07 | 0,07 | 1,01 | 3,12E-01 |
| HTATIP2 | 1226,90 | -0,05 | 0,05 | -1,01 | 3,13E-01 |
| PI4K2A | 294,19 | 0,04 | 0,04 | 1,01 | 3,13E-01 |
| PIPOX | 219,41 | 0,04 | 0,04 | 1,01 | 3,13E-01 |
| GPN2 | 709,62 | 0,03 | 0,03 | 1,01 | 3,13E-01 |
| PABIR1 | 715,53 | 0,02 | 0,02 | 1,01 | 3,14E-01 |
| STARD9 | 738,31 | 0,07 | 0,07 | 1,01 | 3,14E-01 |
| PAFAH2 | 766,19 | -0,04 | 0,04 | -1,01 | 3,14E-01 |
| MBNL1 | 26783,30 | -0,04 | 0,04 | -1,01 | 3,14E-01 |
| TIGIT | 1316,65 | -0,07 | 0,07 | -1,01 | 3,14E-01 |
| LRRC8D | 1198,59 | -0,03 | 0,03 | -1,01 | 3,14E-01 |
| FAM210A | 322,02 | -0,04 | 0,04 | -1,01 | 3,14E-01 |
| TAPBPL | 1070,67 | 0,05 | 0,05 | 1,01 | 3,14E-01 |
| ENSG00000257524 | 85,40 | 0,07 | 0,07 | 1,01 | 3,15E-01 |
| SMIM10L1 | 423,73 | -0,07 | 0,06 | -1,01 | 3,15E-01 |
| CD59 | 1599,31 | -0,05 | 0,05 | -1,01 | 3,15E-01 |
| MICA | 508,35 | 0,04 | 0,04 | 1,01 | 3,15E-01 |
| DERA | 577,31 | -0,04 | 0,04 | -1,00 | 3,15E-01 |
| SYNE3 | 4565,46 | 0,04 | 0,04 | 1,00 | 3,15E-01 |
| TOM1 | 1027,26 | 0,06 | 0,06 | 1,00 | 3,15E-01 |
| AIFM1 | 378,48 | -0,03 | 0,03 | -1,00 | 3,16E-01 |
| CD48 | 5517,99 | -0,06 | 0,06 | -1,00 | 3,16E-01 |
| ADNP2 | 586,83 | 0,03 | 0,03 | 1,00 | 3,16E-01 |
| CAST | 5876,75 | -0,03 | 0,03 | -1,00 | 3,16E-01 |
| VASP | 5765,51 | 0,06 | 0,06 | 1,00 | 3,16E-01 |
| DSP | 98,17 | 0,22 | 0,22 | 1,00 | 3,16E-01 |
| TFDP2 | 1438,63 | -0,04 | 0,04 | -1,00 | 3,16E-01 |
| SIDT1 | 1663,98 | -0,05 | 0,05 | -1,00 | 3,17E-01 |
| TIMM8B | 314,43 | -0,09 | 0,09 | -1,00 | 3,16E-01 |
| PEPD | 673,80 | 0,03 | 0,03 | 1,00 | 3,17E-01 |
| ALYREF | 644,15 | -0,04 | 0,04 | -1,00 | 3,17E-01 |
| HDX | 111,49 | -0,06 | 0,06 | -1,00 | 3,17E-01 |
| ST3GAL5 | 1612,12 | 0,04 | 0,04 | 1,00 | 3,17E-01 |
| SEC22C | 1292,46 | -0,03 | 0,03 | -1,00 | 3,17E-01 |
| RHEX | 41,85 | 0,15 | 0,15 | 1,00 | 3,17E-01 |
| SLC16A6 | 1196,24 | -0,04 | 0,04 | -1,00 | 3,17E-01 |
| GPX7 | 137,52 | 0,05 | 0,05 | 1,00 | 3,18E-01 |

| | | | | | |
|---|---|---|---|---|---|
| TBCK | 2145,53 | -0,05 | 0,05 | -1,00 | 3,18E-01 |
| TADA2B | 1955,13 | -0,03 | 0,03 | -1,00 | 3,18E-01 |
| TET3 | 2902,63 | 0,06 | 0,06 | 1,00 | 3,18E-01 |
| CHIC2 | 2245,82 | -0,05 | 0,05 | -1,00 | 3,18E-01 |
| DDX19B | 224,16 | 0,03 | 0,03 | 1,00 | 3,19E-01 |
| CFAP418 | 71,69 | 0,06 | 0,06 | 1,00 | 3,19E-01 |
| SLC45A3 | 160,68 | 0,14 | 0,14 | 1,00 | 3,19E-01 |
| ZNF620 | 115,13 | 0,04 | 0,04 | 1,00 | 3,19E-01 |
| TNFRSF13C | 170,45 | 0,12 | 0,12 | 1,00 | 3,19E-01 |
| PIGN | 1758,74 | -0,04 | 0,04 | -1,00 | 3,19E-01 |
| PCBD1 | 126,97 | 0,05 | 0,05 | 1,00 | 3,19E-01 |
| PCED1B | 613,01 | 0,06 | 0,06 | 1,00 | 3,19E-01 |
| HNRNPUL1 | 2601,40 | 0,06 | 0,06 | 0,99 | 3,20E-01 |
| TMEM91 | 721,56 | 0,06 | 0,06 | 0,99 | 3,20E-01 |
| NETO1 | 55,89 | 0,13 | 0,13 | 0,99 | 3,20E-01 |
| TMEM14B | 775,74 | 0,04 | 0,04 | 0,99 | 3,20E-01 |
| NDUFB10 | 553,44 | -0,04 | 0,04 | -0,99 | 3,20E-01 |
| SIGLEC9 | 1590,60 | 0,07 | 0,07 | 0,99 | 3,20E-01 |
| SLC22A15 | 2490,07 | -0,06 | 0,06 | -0,99 | 3,20E-01 |
| VMA21 | 1322,04 | -0,05 | 0,05 | -0,99 | 3,20E-01 |
| F13A1 | 5597,36 | 0,09 | 0,09 | 0,99 | 3,20E-01 |
| PHB2 | 1527,25 | -0,04 | 0,04 | -0,99 | 3,21E-01 |
| CARS2 | 2938,57 | 0,04 | 0,05 | 0,99 | 3,21E-01 |
| AASS | 154,91 | 0,08 | 0,08 | 0,99 | 3,21E-01 |
| MAPK9 | 1015,98 | -0,02 | 0,03 | -0,99 | 3,21E-01 |
| BEX3 | 270,66 | 0,09 | 0,09 | 0,99 | 3,21E-01 |
| TLN1 | 15145,86 | 0,09 | 0,09 | 0,99 | 3,21E-01 |
| BRD2 | 3776,97 | 0,04 | 0,04 | 0,99 | 3,22E-01 |
| RUNX3 | 1148,61 | 0,07 | 0,07 | 0,99 | 3,22E-01 |
| PKNOX1 | 614,72 | 0,05 | 0,05 | 0,99 | 3,22E-01 |
| ASB13 | 259,71 | 0,04 | 0,04 | 0,99 | 3,22E-01 |
| CRLS1 | 924,95 | -0,04 | 0,04 | -0,99 | 3,22E-01 |
| PCGF6 | 250,54 | -0,05 | 0,05 | -0,99 | 3,22E-01 |
| GPRASP2 | 64,95 | 0,07 | 0,07 | 0,99 | 3,22E-01 |
| KLHDC10 | 1547,37 | -0,02 | 0,02 | -0,99 | 3,22E-01 |
| NDUFA4 | 786,29 | -0,10 | 0,10 | -0,99 | 3,23E-01 |
| ZNF318 | 1366,96 | -0,04 | 0,04 | -0,99 | 3,23E-01 |
| SURF2 | 162,26 | 0,04 | 0,04 | 0,99 | 3,23E-01 |
| LSM1 | 434,92 | -0,07 | 0,07 | -0,99 | 3,23E-01 |
| CLCN5 | 476,71 | 0,05 | 0,05 | 0,99 | 3,23E-01 |
| HFE | 214,76 | 0,04 | 0,04 | 0,99 | 3,23E-01 |
| ZCCHC8 | 986,90 | -0,03 | 0,03 | -0,99 | 3,23E-01 |
| TBCCD1 | 473,17 | -0,03 | 0,03 | -0,99 | 3,23E-01 |
| ENSG00000282988 | 330,33 | 0,09 | 0,09 | 0,99 | 3,23E-01 |
| CTCF | 1187,18 | 0,04 | 0,04 | 0,99 | 3,23E-01 |
| MFAP3L | 887,55 | 0,08 | 0,08 | 0,99 | 3,23E-01 |

| | | | | | |
|---|---|---|---|---|---|
| FBXO25 | 761,54 | -0,03 | 0,03 | -0,99 | 3,24E-01 |
| CCNY | 3735,64 | -0,03 | 0,03 | -0,99 | 3,24E-01 |
| RASGRP1 | 2984,16 | -0,04 | 0,04 | -0,99 | 3,24E-01 |
| AP1S1 | 177,75 | 0,05 | 0,05 | 0,98 | 3,25E-01 |
| SF3A1 | 1327,25 | 0,06 | 0,06 | 0,98 | 3,25E-01 |
| UMAD1 | 687,03 | -0,04 | 0,04 | -0,98 | 3,25E-01 |
| BLNK | 304,11 | -0,08 | 0,08 | -0,98 | 3,25E-01 |
| MS4A6A | 4935,72 | -0,06 | 0,06 | -0,98 | 3,25E-01 |
| DYNLT1 | 2018,97 | -0,06 | 0,06 | -0,98 | 3,25E-01 |
| RNF14 | 2317,27 | 0,07 | 0,07 | 0,98 | 3,26E-01 |
| LRRK2 | 47433,68 | -0,07 | 0,07 | -0,98 | 3,26E-01 |
| AHCYL1 | 1616,35 | -0,02 | 0,02 | -0,98 | 3,27E-01 |
| UQCRC1 | 1395,27 | -0,03 | 0,03 | -0,98 | 3,27E-01 |
| PRPF3 | 1855,24 | -0,03 | 0,03 | -0,98 | 3,27E-01 |
| POLR2J | 549,06 | 0,04 | 0,04 | 0,98 | 3,27E-01 |
| SMIM1 | 215,49 | 0,21 | 0,21 | 0,98 | 3,28E-01 |
| NPC2 | 1778,59 | -0,05 | 0,05 | -0,98 | 3,28E-01 |
| NLRC3 | 3031,11 | 0,05 | 0,05 | 0,98 | 3,28E-01 |
| SMIM7 | 1105,03 | -0,03 | 0,03 | -0,98 | 3,28E-01 |
| ACADVL | 2344,46 | 0,04 | 0,04 | 0,98 | 3,28E-01 |
| EEF2 | 24360,25 | 0,04 | 0,04 | 0,98 | 3,28E-01 |
| CLK3 | 3750,35 | 0,03 | 0,03 | 0,98 | 3,28E-01 |
| SLCO3A1 | 3111,78 | 0,05 | 0,05 | 0,98 | 3,28E-01 |
| EOLA1 | 359,30 | -0,03 | 0,03 | -0,98 | 3,28E-01 |
| SLC16A10 | 304,86 | 0,08 | 0,08 | 0,98 | 3,28E-01 |
| RTKN2 | 468,06 | -0,09 | 0,10 | -0,98 | 3,28E-01 |
| PUM1 | 3080,17 | -0,03 | 0,03 | -0,98 | 3,29E-01 |
| EYA3 | 1094,43 | 0,04 | 0,04 | 0,98 | 3,29E-01 |
| DDTL | 31,30 | 0,18 | 0,19 | 0,98 | 3,29E-01 |
| SMG7 | 2039,91 | 0,04 | 0,04 | 0,98 | 3,29E-01 |
| AHR | 2054,03 | 0,06 | 0,06 | 0,98 | 3,29E-01 |
| H1-5 | 774,19 | 0,10 | 0,10 | 0,98 | 3,29E-01 |
| NR2C2AP | 222,61 | -0,04 | 0,04 | -0,98 | 3,29E-01 |
| FGR | 10090,66 | 0,05 | 0,05 | 0,98 | 3,29E-01 |
| FYB1 | 17323,08 | -0,05 | 0,05 | -0,97 | 3,30E-01 |
| PFKP | 1051,85 | 0,05 | 0,05 | 0,97 | 3,30E-01 |
| MAB21L3 | 77,42 | 0,11 | 0,12 | 0,97 | 3,30E-01 |
| XYLT1 | 1657,19 | 0,06 | 0,06 | 0,97 | 3,30E-01 |
| TIGAR | 152,26 | 0,03 | 0,04 | 0,97 | 3,31E-01 |
| TLR6 | 5677,69 | -0,06 | 0,06 | -0,97 | 3,31E-01 |
| FOCAD | 1438,24 | -0,05 | 0,05 | -0,97 | 3,31E-01 |
| TSGA10 | 414,06 | -0,07 | 0,07 | -0,97 | 3,31E-01 |
| MAML3 | 687,77 | 0,07 | 0,07 | 0,97 | 3,31E-01 |
| TTF2 | 659,17 | 0,04 | 0,04 | 0,97 | 3,31E-01 |
| CCNYL1 | 474,79 | 0,03 | 0,03 | 0,97 | 3,31E-01 |
| AURKA | 126,36 | 0,08 | 0,08 | 0,97 | 3,32E-01 |

| | | | | | |
|---|---|---|---|---|---|
| SERPINB9 | 2484,83 | 0,04 | 0,04 | 0,97 | 3,32E-01 |
| ZNF850 | 186,12 | -0,05 | 0,05 | -0,97 | 3,32E-01 |
| CCDC146 | 520,69 | -0,06 | 0,06 | -0,97 | 3,32E-01 |
| UBE2L3 | 1083,45 | -0,03 | 0,03 | -0,97 | 3,32E-01 |
| CD99L2 | 503,52 | 0,03 | 0,03 | 0,97 | 3,32E-01 |
| C2orf88 | 819,97 | 0,09 | 0,09 | 0,97 | 3,32E-01 |
| C18orf32 | 345,66 | 0,03 | 0,03 | 0,97 | 3,32E-01 |
| ARHGAP21 | 938,71 | 0,04 | 0,04 | 0,97 | 3,33E-01 |
| RNF182 | 455,89 | 0,25 | 0,25 | 0,97 | 3,33E-01 |
| NEK4 | 905,37 | -0,02 | 0,02 | -0,97 | 3,33E-01 |
| MACROH2A1 | 11122,04 | -0,04 | 0,04 | -0,97 | 3,33E-01 |
| CCDC88B | 6204,15 | 0,06 | 0,06 | 0,97 | 3,33E-01 |
| ALG6 | 1057,21 | -0,04 | 0,04 | -0,97 | 3,34E-01 |
| RAPGEF2 | 5102,35 | -0,05 | 0,05 | -0,97 | 3,34E-01 |
| KCTD9 | 979,88 | -0,04 | 0,04 | -0,97 | 3,34E-01 |
| GRAMD2B | 151,58 | 0,05 | 0,05 | 0,97 | 3,34E-01 |
| ZNF506 | 2558,76 | -0,04 | 0,04 | -0,97 | 3,34E-01 |
| JAKMIP1 | 46,63 | 0,11 | 0,11 | 0,97 | 3,34E-01 |
| UBXN1 | 1915,91 | -0,04 | 0,04 | -0,97 | 3,34E-01 |
| KCNMB3 | 98,72 | 0,05 | 0,06 | 0,97 | 3,34E-01 |
| PLIN2 | 825,74 | -0,04 | 0,05 | -0,97 | 3,34E-01 |
| NOXRED1 | 101,51 | 0,04 | 0,05 | 0,97 | 3,34E-01 |
| MRPL53 | 158,75 | -0,03 | 0,03 | -0,97 | 3,34E-01 |
| RIC1 | 3446,01 | -0,03 | 0,03 | -0,97 | 3,34E-01 |
| VPS35L | 789,11 | 0,03 | 0,03 | 0,97 | 3,34E-01 |
| PIGP | 244,81 | 0,05 | 0,05 | 0,96 | 3,35E-01 |
| SLC44A5 | 59,28 | 0,26 | 0,27 | 0,96 | 3,35E-01 |
| PDIA4 | 1086,42 | -0,04 | 0,04 | -0,96 | 3,35E-01 |
| LRRIQ3 | 42,51 | 0,13 | 0,14 | 0,96 | 3,35E-01 |
| AAMP | 913,97 | -0,03 | 0,03 | -0,96 | 3,35E-01 |
| PRKAG2 | 1042,29 | 0,03 | 0,04 | 0,96 | 3,35E-01 |
| ST3GAL1 | 4713,76 | 0,05 | 0,05 | 0,96 | 3,36E-01 |
| DENND5B | 283,54 | 0,08 | 0,09 | 0,96 | 3,36E-01 |
| HARBI1 | 147,19 | 0,03 | 0,04 | 0,96 | 3,36E-01 |
| ZNF514 | 918,26 | -0,05 | 0,05 | -0,96 | 3,36E-01 |
| PTGDS | 92,26 | 0,12 | 0,12 | 0,96 | 3,37E-01 |
| NCAPD2 | 1089,09 | 0,04 | 0,04 | 0,96 | 3,37E-01 |
| LACC1 | 158,71 | -0,07 | 0,07 | -0,96 | 3,37E-01 |
| SDR42E1 | 350,49 | 0,05 | 0,06 | 0,96 | 3,37E-01 |
| BRPF1 | 838,05 | 0,04 | 0,04 | 0,96 | 3,38E-01 |
| CCDC28B | 124,20 | 0,05 | 0,05 | 0,96 | 3,38E-01 |
| SPOUT1 | 587,67 | 0,03 | 0,03 | 0,96 | 3,38E-01 |
| SQSTM1 | 5411,71 | 0,04 | 0,04 | 0,96 | 3,38E-01 |
| AKIP1 | 190,97 | -0,04 | 0,04 | -0,96 | 3,38E-01 |
| AIRIM | 302,33 | 0,03 | 0,03 | 0,96 | 3,38E-01 |
| TLR8 | 5690,06 | -0,06 | 0,06 | -0,96 | 3,38E-01 |

| | | | | | |
|---|---|---|---|---|---|
| CDK11B | 723,91 | -0,04 | 0,04 | -0,96 | 3,38E-01 |
| ZDHHC20 | 4739,96 | -0,05 | 0,06 | -0,96 | 3,39E-01 |
| DNASE1L1 | 776,66 | 0,04 | 0,04 | 0,96 | 3,39E-01 |
| C2orf76 | 157,86 | -0,04 | 0,04 | -0,96 | 3,40E-01 |
| SIMC1 | 281,68 | 0,04 | 0,05 | 0,95 | 3,40E-01 |
| ZKSCAN3 | 418,90 | 0,03 | 0,03 | 0,95 | 3,40E-01 |
| SCPEP1 | 2339,58 | -0,05 | 0,05 | -0,95 | 3,40E-01 |
| FUT7 | 1216,84 | -0,07 | 0,07 | -0,95 | 3,40E-01 |
| KDM1B | 1676,81 | 0,05 | 0,06 | 0,95 | 3,41E-01 |
| EEF1AKMT1 | 92,11 | -0,05 | 0,05 | -0,95 | 3,41E-01 |
| MT1F | 79,98 | 0,06 | 0,06 | 0,95 | 3,41E-01 |
| WDR12 | 444,49 | -0,03 | 0,03 | -0,95 | 3,41E-01 |
| NUDT12 | 72,36 | 0,12 | 0,12 | 0,95 | 3,41E-01 |
| WDR46 | 662,20 | -0,03 | 0,03 | -0,95 | 3,42E-01 |
| CDYL | 1362,26 | 0,07 | 0,07 | 0,95 | 3,42E-01 |
| RIOK3 | 10561,41 | 0,09 | 0,09 | 0,95 | 3,42E-01 |
| PDPR | 1135,54 | 0,09 | 0,09 | 0,95 | 3,42E-01 |
| MRPL46 | 340,61 | -0,04 | 0,04 | -0,95 | 3,42E-01 |
| LAMTOR1 | 1869,81 | 0,04 | 0,04 | 0,95 | 3,42E-01 |
| DST | 1032,25 | 0,07 | 0,07 | 0,95 | 3,42E-01 |
| PERP | 57,09 | 0,10 | 0,11 | 0,95 | 3,42E-01 |
| GATAD2B | 1605,61 | 0,08 | 0,08 | 0,95 | 3,43E-01 |
| PDXDC1 | 2175,35 | -0,03 | 0,03 | -0,95 | 3,43E-01 |
| RRP1 | 319,24 | 0,04 | 0,04 | 0,95 | 3,43E-01 |
| COG2 | 1220,99 | -0,03 | 0,03 | -0,95 | 3,43E-01 |
| PSMB2 | 621,70 | 0,03 | 0,03 | 0,95 | 3,43E-01 |
| P2RY14 | 492,94 | 0,12 | 0,12 | 0,95 | 3,44E-01 |
| CBY1 | 228,16 | 0,03 | 0,03 | 0,95 | 3,44E-01 |
| DCLRE1B | 147,02 | 0,04 | 0,04 | 0,95 | 3,44E-01 |
| PRDM1 | 1312,06 | -0,04 | 0,05 | -0,95 | 3,45E-01 |
| RHNO1 | 271,22 | -0,04 | 0,04 | -0,94 | 3,45E-01 |
| LIME1 | 132,63 | 0,04 | 0,04 | 0,94 | 3,45E-01 |
| DHDDS | 447,32 | 0,03 | 0,03 | 0,94 | 3,45E-01 |
| TMEM185A | 384,09 | -0,03 | 0,03 | -0,94 | 3,45E-01 |
| SERTAD1 | 388,05 | -0,04 | 0,04 | -0,94 | 3,45E-01 |
| PGBD1 | 58,29 | 0,06 | 0,06 | 0,94 | 3,45E-01 |
| NLK | 957,19 | -0,02 | 0,02 | -0,94 | 3,45E-01 |
| KIR2DL1 | 23,61 | 0,16 | 0,17 | 0,94 | 3,45E-01 |
| TRIM24 | 1307,17 | -0,03 | 0,03 | -0,94 | 3,46E-01 |
| VPS29 | 2080,59 | -0,04 | 0,05 | -0,94 | 3,46E-01 |
| MGMT | 423,27 | -0,04 | 0,04 | -0,94 | 3,46E-01 |
| RGS10 | 1685,39 | 0,08 | 0,09 | 0,94 | 3,46E-01 |
| HK2 | 2185,17 | 0,06 | 0,07 | 0,94 | 3,46E-01 |
| CD28 | 2817,37 | -0,06 | 0,06 | -0,94 | 3,47E-01 |
| QPCT | 2586,64 | -0,08 | 0,08 | -0,94 | 3,47E-01 |
| UBE2E3 | 477,15 | 0,03 | 0,04 | 0,94 | 3,47E-01 |

| | | | | | |
|---|---|---|---|---|---|
| NSFL1C | 3006,00 | 0,05 | 0,06 | 0,94 | 3,47E-01 |
| PFDN5 | 1972,08 | -0,10 | 0,11 | -0,94 | 3,47E-01 |
| RASSF2 | 23237,37 | -0,05 | 0,06 | -0,94 | 3,47E-01 |
| CORO1A | 12652,33 | 0,05 | 0,05 | 0,94 | 3,48E-01 |
| UBE2E2 | 215,89 | 0,04 | 0,05 | 0,94 | 3,48E-01 |
| GNS | 5951,44 | -0,05 | 0,05 | -0,94 | 3,48E-01 |
| ARPC1B | 1161,28 | 0,04 | 0,04 | 0,94 | 3,48E-01 |
| NEK6 | 1852,72 | 0,06 | 0,06 | 0,94 | 3,48E-01 |
| HIP1 | 1444,41 | 0,11 | 0,11 | 0,94 | 3,49E-01 |
| GNL1 | 890,13 | -0,02 | 0,03 | -0,94 | 3,49E-01 |
| NDC1 | 671,52 | -0,03 | 0,03 | -0,94 | 3,49E-01 |
| SQOR | 1319,38 | -0,05 | 0,05 | -0,94 | 3,49E-01 |
| SOCS7 | 680,90 | 0,03 | 0,03 | 0,94 | 3,49E-01 |
| BST2 | 1683,34 | -0,07 | 0,07 | -0,94 | 3,49E-01 |
| IDH2 | 905,16 | 0,04 | 0,04 | 0,94 | 3,50E-01 |
| NONO | 2134,98 | 0,03 | 0,03 | 0,94 | 3,50E-01 |
| RPGRIP1 | 319,10 | -0,07 | 0,08 | -0,94 | 3,50E-01 |
| LRCH3 | 2267,29 | 0,03 | 0,03 | 0,94 | 3,50E-01 |
| SCLT1 | 5178,67 | -0,06 | 0,06 | -0,93 | 3,50E-01 |
| ANAPC5 | 3650,00 | -0,02 | 0,02 | -0,93 | 3,50E-01 |
| LILRA2 | 2252,09 | 0,07 | 0,07 | 0,93 | 3,50E-01 |
| PPP1CA | 2010,60 | 0,04 | 0,04 | 0,93 | 3,50E-01 |
| ERICH1 | 3644,87 | -0,05 | 0,05 | -0,93 | 3,50E-01 |
| IFIH1 | 1783,80 | -0,09 | 0,09 | -0,93 | 3,50E-01 |
| JAK2 | 5991,68 | -0,06 | 0,07 | -0,93 | 3,51E-01 |
| C5orf24 | 1192,90 | -0,04 | 0,04 | -0,93 | 3,51E-01 |
| CLIP1 | 2932,83 | -0,04 | 0,04 | -0,93 | 3,51E-01 |
| PRPF8 | 4394,38 | 0,05 | 0,06 | 0,93 | 3,51E-01 |
| CXCL1 | 108,57 | -0,08 | 0,08 | -0,93 | 3,51E-01 |
| PRMT1 | 444,85 | 0,03 | 0,03 | 0,93 | 3,51E-01 |
| PES1 | 787,21 | -0,03 | 0,03 | -0,93 | 3,51E-01 |
| CENPF | 183,13 | 0,09 | 0,10 | 0,93 | 3,51E-01 |
| MRPS6 | 356,56 | -0,05 | 0,05 | -0,93 | 3,52E-01 |
| TRIM25 | 7737,72 | 0,07 | 0,08 | 0,93 | 3,52E-01 |
| CCNG2 | 2559,53 | -0,04 | 0,05 | -0,93 | 3,53E-01 |
| USP32 | 8537,15 | -0,05 | 0,05 | -0,93 | 3,53E-01 |
| AHCYL2 | 723,90 | 0,03 | 0,03 | 0,93 | 3,53E-01 |
| RAB3D | 3164,12 | 0,08 | 0,09 | 0,93 | 3,53E-01 |
| H2BK1 | 23,17 | 0,06 | 0,07 | 0,93 | 3,53E-01 |
| CARS1 | 1797,00 | 0,03 | 0,03 | 0,93 | 3,53E-01 |
| SETDB1 | 880,72 | 0,05 | 0,05 | 0,93 | 3,54E-01 |
| CHCHD1 | 278,85 | -0,05 | 0,05 | -0,93 | 3,54E-01 |
| MAP4K4 | 6973,41 | -0,04 | 0,05 | -0,93 | 3,54E-01 |
| NUBP2 | 788,71 | 0,03 | 0,03 | 0,93 | 3,54E-01 |
| DELE1 | 1153,25 | 0,05 | 0,05 | 0,93 | 3,54E-01 |
| PATL1 | 1080,19 | 0,05 | 0,06 | 0,93 | 3,54E-01 |

| | | | | | |
|---|---|---|---|---|---|
| DAAM2 | 257,26 | 0,18 | 0,19 | 0,93 | 3,54E-01 |
| MAP1LC3B | 1862,21 | 0,04 | 0,04 | 0,93 | 3,55E-01 |
| ATP10A | 1073,40 | 0,05 | 0,06 | 0,93 | 3,55E-01 |
| UBE2D1 | 2905,23 | -0,08 | 0,08 | -0,93 | 3,55E-01 |
| RAMACL | 58,58 | 0,08 | 0,09 | 0,92 | 3,56E-01 |
| PRRC2C | 9590,06 | -0,04 | 0,04 | -0,92 | 3,56E-01 |
| HINT1 | 839,86 | -0,09 | 0,09 | -0,92 | 3,56E-01 |
| ZNF646 | 1622,74 | 0,05 | 0,06 | 0,92 | 3,56E-01 |
| SLC37A2 | 744,40 | 0,06 | 0,06 | 0,92 | 3,56E-01 |
| MKRN3 | 84,13 | 0,13 | 0,14 | 0,92 | 3,56E-01 |
| CEBPZOS | 706,09 | -0,03 | 0,03 | -0,92 | 3,57E-01 |
| DNAJB5 | 88,38 | 0,10 | 0,11 | 0,92 | 3,57E-01 |
| APOL3 | 4166,34 | -0,04 | 0,04 | -0,92 | 3,58E-01 |
| ZNF331 | 517,12 | 0,04 | 0,04 | 0,92 | 3,58E-01 |
| FCGR2B | 635,60 | -0,08 | 0,09 | -0,92 | 3,58E-01 |
| RRAGD | 1097,28 | -0,05 | 0,06 | -0,92 | 3,58E-01 |
| CNOT1 | 7801,11 | -0,04 | 0,04 | -0,92 | 3,58E-01 |
| MFSD11 | 791,37 | 0,02 | 0,02 | 0,92 | 3,58E-01 |
| DOCK9 | 2161,99 | 0,04 | 0,05 | 0,92 | 3,59E-01 |
| PRELID1 | 2171,45 | -0,04 | 0,04 | -0,92 | 3,59E-01 |
| KLRC3 | 133,53 | -0,13 | 0,14 | -0,92 | 3,59E-01 |
| ZNF174 | 243,19 | 0,02 | 0,02 | 0,92 | 3,59E-01 |
| RPS3 | 9524,40 | -0,06 | 0,07 | -0,92 | 3,59E-01 |
| KDM3B | 3637,28 | 0,05 | 0,06 | 0,92 | 3,59E-01 |
| MOB1A | 9316,93 | -0,04 | 0,04 | -0,92 | 3,59E-01 |
| OSBPL5 | 763,83 | 0,09 | 0,10 | 0,92 | 3,59E-01 |
| LRRFIP1 | 14544,35 | -0,03 | 0,04 | -0,92 | 3,59E-01 |
| TRAPPC6A | 353,23 | -0,04 | 0,04 | -0,92 | 3,60E-01 |
| YKT6 | 1326,32 | -0,02 | 0,02 | -0,92 | 3,60E-01 |
| HRH2 | 2541,46 | 0,07 | 0,08 | 0,92 | 3,60E-01 |
| CDC42BPA | 375,39 | 0,09 | 0,10 | 0,92 | 3,60E-01 |
| ABHD18 | 1522,46 | -0,02 | 0,03 | -0,91 | 3,61E-01 |
| SWT1 | 835,06 | -0,08 | 0,09 | -0,91 | 3,61E-01 |
| GRAP2 | 2684,64 | 0,04 | 0,04 | 0,91 | 3,61E-01 |
| COL6A2 | 366,04 | 0,10 | 0,11 | 0,91 | 3,61E-01 |
| DCP1A | 936,90 | -0,03 | 0,03 | -0,91 | 3,61E-01 |
| SH3BGRL2 | 1389,41 | 0,10 | 0,10 | 0,91 | 3,61E-01 |
| EDEM1 | 3926,37 | 0,03 | 0,04 | 0,91 | 3,61E-01 |
| NBPF11 | 717,92 | 0,04 | 0,05 | 0,91 | 3,61E-01 |
| LMAN2 | 2645,79 | -0,03 | 0,03 | -0,91 | 3,61E-01 |
| RNF41 | 2199,51 | -0,03 | 0,03 | -0,91 | 3,62E-01 |
| GNPTG | 825,23 | 0,03 | 0,03 | 0,91 | 3,62E-01 |
| DUSP3 | 917,94 | 0,04 | 0,05 | 0,91 | 3,62E-01 |
| CCDC34 | 48,95 | 0,09 | 0,10 | 0,91 | 3,62E-01 |
| NCF1 | 4230,00 | 0,07 | 0,08 | 0,91 | 3,62E-01 |
| SAXO2 | 61,86 | -0,18 | 0,20 | -0,91 | 3,62E-01 |

| | | | | | |
|---|---|---|---|---|---|
| S100A10 | 751,42 | 0,04 | 0,04 | 0,91 | 3,62E-01 |
| STX5 | 775,26 | 0,03 | 0,03 | 0,91 | 3,63E-01 |
| VPS33A | 728,09 | -0,02 | 0,02 | -0,91 | 3,63E-01 |
| ZFAND4 | 1037,68 | 0,03 | 0,03 | 0,91 | 3,63E-01 |
| ZNF524 | 311,87 | 0,04 | 0,04 | 0,91 | 3,63E-01 |
| PFKFB4 | 2409,08 | 0,06 | 0,07 | 0,91 | 3,63E-01 |
| AMER1 | 261,89 | 0,04 | 0,04 | 0,91 | 3,63E-01 |
| CRISPLD2 | 3264,67 | -0,09 | 0,10 | -0,91 | 3,64E-01 |
| MPC2 | 929,25 | 0,05 | 0,05 | 0,91 | 3,64E-01 |
| ARPC4 | 5164,68 | -0,04 | 0,04 | -0,91 | 3,64E-01 |
| CPEB3 | 417,06 | 0,04 | 0,04 | 0,91 | 3,64E-01 |
| NBPF10 | 448,79 | 0,05 | 0,06 | 0,91 | 3,65E-01 |
| HYPK | 336,12 | 0,04 | 0,04 | 0,91 | 3,65E-01 |
| NCAPG2 | 515,83 | 0,10 | 0,11 | 0,91 | 3,65E-01 |
| MAP4 | 1176,92 | 0,05 | 0,06 | 0,91 | 3,65E-01 |
| ELK3 | 749,62 | -0,03 | 0,03 | -0,91 | 3,65E-01 |
| UBXN2B | 3273,93 | -0,05 | 0,06 | -0,90 | 3,65E-01 |
| NIT1 | 606,81 | 0,03 | 0,03 | 0,90 | 3,66E-01 |
| TRIM69 | 1743,84 | -0,05 | 0,05 | -0,90 | 3,66E-01 |
| UQCC4 | 106,80 | -0,04 | 0,05 | -0,90 | 3,66E-01 |
| KANSL1L | 1154,19 | -0,04 | 0,04 | -0,90 | 3,66E-01 |
| TRIM59 | 401,49 | -0,05 | 0,05 | -0,90 | 3,66E-01 |
| ZNF285 | 77,42 | 0,09 | 0,10 | 0,90 | 3,66E-01 |
| ZNF85 | 231,33 | -0,05 | 0,05 | -0,90 | 3,67E-01 |
| AKAP6 | 235,64 | 0,10 | 0,11 | 0,90 | 3,67E-01 |
| TOP6BL | 257,96 | 0,06 | 0,07 | 0,90 | 3,67E-01 |
| CLEC9A | 80,40 | -0,10 | 0,11 | -0,90 | 3,67E-01 |
| RPA1 | 1067,66 | 0,04 | 0,05 | 0,90 | 3,67E-01 |
| RFC4 | 164,63 | -0,04 | 0,05 | -0,90 | 3,67E-01 |
| NSMCE4A | 900,21 | -0,03 | 0,03 | -0,90 | 3,67E-01 |
| TMUB1 | 238,86 | 0,04 | 0,04 | 0,90 | 3,68E-01 |
| PPP1R16B | 1403,68 | 0,05 | 0,05 | 0,90 | 3,68E-01 |
| ACSF3 | 1585,95 | 0,04 | 0,05 | 0,90 | 3,68E-01 |
| TK2 | 972,39 | 0,05 | 0,05 | 0,90 | 3,69E-01 |
| TRIR | 1150,29 | 0,03 | 0,04 | 0,90 | 3,69E-01 |
| METTL3 | 1408,39 | -0,02 | 0,03 | -0,90 | 3,69E-01 |
| CDR2 | 750,23 | -0,04 | 0,05 | -0,90 | 3,70E-01 |
| OGFOD1 | 460,64 | -0,02 | 0,03 | -0,90 | 3,70E-01 |
| IPCEF1 | 3310,88 | -0,03 | 0,04 | -0,90 | 3,70E-01 |
| ABHD6 | 256,54 | 0,03 | 0,03 | 0,90 | 3,71E-01 |
| PPIAL4H | 82,53 | -0,11 | 0,12 | -0,89 | 3,71E-01 |
| ALDH16A1 | 728,44 | 0,04 | 0,04 | 0,89 | 3,71E-01 |
| TMEM127 | 2735,25 | 0,07 | 0,07 | 0,89 | 3,71E-01 |
| SPPL2A | 2431,11 | -0,03 | 0,03 | -0,89 | 3,72E-01 |
| LONRF3 | 406,69 | -0,04 | 0,04 | -0,89 | 3,72E-01 |
| CAMK4 | 4126,37 | -0,07 | 0,07 | -0,89 | 3,72E-01 |

| | | | | | |
|---|---|---|---|---|---|
| TIMM10B | 574,77 | -0,03 | 0,03 | -0,89 | 3,73E-01 |
| PARL | 184,22 | -0,03 | 0,03 | -0,89 | 3,73E-01 |
| KPNA6 | 1885,01 | 0,03 | 0,04 | 0,89 | 3,73E-01 |
| CLIC2 | 238,35 | 0,08 | 0,09 | 0,89 | 3,73E-01 |
| GPD2 | 1521,69 | -0,04 | 0,05 | -0,89 | 3,73E-01 |
| FOXP1 | 8031,70 | -0,04 | 0,04 | -0,89 | 3,73E-01 |
| TAS2R46 | 131,13 | -0,07 | 0,08 | -0,89 | 3,73E-01 |
| CPT1A | 1403,73 | 0,07 | 0,08 | 0,89 | 3,73E-01 |
| ADHFE1 | 146,33 | 0,04 | 0,05 | 0,89 | 3,74E-01 |
| FCN1 | 21435,41 | -0,06 | 0,06 | -0,89 | 3,74E-01 |
| AKAP13 | 22248,33 | -0,04 | 0,05 | -0,89 | 3,74E-01 |
| NECTIN3 | 109,82 | 0,07 | 0,08 | 0,89 | 3,74E-01 |
| USP12 | 3021,91 | 0,07 | 0,08 | 0,89 | 3,74E-01 |
| TRIP4 | 512,23 | -0,03 | 0,03 | -0,89 | 3,74E-01 |
| TMEM185B | 563,62 | 0,04 | 0,05 | 0,89 | 3,75E-01 |
| ODF2 | 709,29 | 0,03 | 0,04 | 0,89 | 3,75E-01 |
| WDHD1 | 182,33 | -0,06 | 0,07 | -0,89 | 3,75E-01 |
| IKZF2 | 2826,79 | -0,07 | 0,08 | -0,89 | 3,75E-01 |
| FYCO1 | 1351,83 | 0,05 | 0,05 | 0,89 | 3,76E-01 |
| MGAT5 | 2684,31 | -0,03 | 0,03 | -0,89 | 3,76E-01 |
| TSC22D3 | 5338,26 | 0,06 | 0,07 | 0,89 | 3,76E-01 |
| MSMO1 | 445,66 | -0,05 | 0,06 | -0,88 | 3,76E-01 |
| FGD6 | 370,13 | 0,06 | 0,06 | 0,88 | 3,76E-01 |
| SPON2 | 690,05 | 0,09 | 0,10 | 0,88 | 3,76E-01 |
| TMTC1 | 483,94 | -0,15 | 0,17 | -0,88 | 3,77E-01 |
| CALCRL | 106,80 | -0,08 | 0,09 | -0,88 | 3,77E-01 |
| DENND1B | 2045,27 | -0,05 | 0,06 | -0,88 | 3,77E-01 |
| H2BC13 | 199,20 | 0,09 | 0,10 | 0,88 | 3,77E-01 |
| E2F5 | 246,62 | -0,07 | 0,08 | -0,88 | 3,78E-01 |
| RASGRP3 | 417,78 | 0,07 | 0,08 | 0,88 | 3,78E-01 |
| ZEB2 | 17084,04 | -0,04 | 0,05 | -0,88 | 3,78E-01 |
| MTHFD2L | 483,86 | -0,05 | 0,06 | -0,88 | 3,78E-01 |
| WDR77 | 424,00 | 0,03 | 0,04 | 0,88 | 3,78E-01 |
| CENPK | 448,20 | -0,12 | 0,13 | -0,88 | 3,78E-01 |
| CXCR5 | 283,73 | -0,08 | 0,09 | -0,88 | 3,79E-01 |
| TNFAIP6 | 784,56 | -0,12 | 0,14 | -0,88 | 3,79E-01 |
| CCL3 | 84,44 | 0,09 | 0,10 | 0,88 | 3,79E-01 |
| MACC1 | 93,03 | 0,08 | 0,09 | 0,88 | 3,79E-01 |
| TMEM254 | 189,94 | 0,03 | 0,03 | 0,88 | 3,79E-01 |
| TTC9 | 497,54 | 0,06 | 0,07 | 0,88 | 3,79E-01 |
| GPR108 | 720,17 | 0,04 | 0,04 | 0,88 | 3,79E-01 |
| LSMEM1 | 364,63 | -0,07 | 0,08 | -0,88 | 3,79E-01 |
| URGCP | 657,96 | 0,03 | 0,03 | 0,88 | 3,80E-01 |
| CDK12 | 3679,60 | -0,02 | 0,03 | -0,88 | 3,80E-01 |
| TMEM144 | 395,50 | 0,07 | 0,08 | 0,88 | 3,81E-01 |
| RASSF4 | 1246,85 | 0,05 | 0,06 | 0,88 | 3,82E-01 |

| | | | | | |
|---|---|---|---|---|---|
| GOLGA8R | 60,17 | -0,09 | 0,10 | -0,87 | 3,82E-01 |
| DDX60 | 2860,11 | -0,09 | 0,11 | -0,87 | 3,82E-01 |
| GPR141 | 1055,42 | -0,08 | 0,09 | -0,87 | 3,82E-01 |
| WDR20 | 1917,09 | -0,03 | 0,03 | -0,87 | 3,82E-01 |
| DNASE1 | 2129,17 | 0,03 | 0,03 | 0,87 | 3,82E-01 |
| TULP4 | 1309,91 | -0,03 | 0,03 | -0,87 | 3,82E-01 |
| CFL2 | 231,68 | -0,06 | 0,06 | -0,87 | 3,82E-01 |
| ZDHHC23 | 280,00 | -0,05 | 0,06 | -0,87 | 3,82E-01 |
| LPIN2 | 4746,89 | 0,05 | 0,06 | 0,87 | 3,83E-01 |
| KIAA0040 | 4022,69 | -0,04 | 0,05 | -0,87 | 3,83E-01 |
| ZNF251 | 429,16 | 0,03 | 0,03 | 0,87 | 3,83E-01 |
| APOLD1 | 271,31 | 0,05 | 0,06 | 0,87 | 3,83E-01 |
| TNFRSF10B | 2470,37 | 0,04 | 0,05 | 0,87 | 3,84E-01 |
| ANKRD11 | 4612,54 | 0,06 | 0,07 | 0,87 | 3,84E-01 |
| PHPT1 | 257,79 | 0,04 | 0,04 | 0,87 | 3,84E-01 |
| MRPL34 | 278,86 | 0,04 | 0,04 | 0,87 | 3,84E-01 |
| TRIQK | 708,91 | -0,06 | 0,07 | -0,87 | 3,84E-01 |
| CYP27A1 | 751,65 | -0,09 | 0,10 | -0,87 | 3,85E-01 |
| SP140L | 1989,85 | -0,03 | 0,04 | -0,87 | 3,85E-01 |
| SERPINB8 | 586,45 | -0,04 | 0,04 | -0,87 | 3,85E-01 |
| GSDMD | 2608,03 | 0,04 | 0,04 | 0,87 | 3,85E-01 |
| ACOT8 | 676,03 | 0,02 | 0,02 | 0,87 | 3,85E-01 |
| B3GALT9 | 119,94 | -0,04 | 0,05 | -0,87 | 3,86E-01 |
| SATB1 | 3980,06 | -0,04 | 0,04 | -0,87 | 3,86E-01 |
| TXLNA | 1054,82 | 0,03 | 0,04 | 0,87 | 3,86E-01 |
| MEF2D | 1437,04 | 0,06 | 0,07 | 0,87 | 3,86E-01 |
| PIP5K1A | 1275,37 | 0,02 | 0,03 | 0,87 | 3,87E-01 |
| KLRC2 | 194,76 | -0,15 | 0,17 | -0,87 | 3,87E-01 |
| CCDC102A | 200,99 | 0,05 | 0,06 | 0,86 | 3,87E-01 |
| ZNF532 | 419,11 | 0,06 | 0,07 | 0,86 | 3,87E-01 |
| ENTPD1 | 5952,89 | -0,05 | 0,06 | -0,86 | 3,87E-01 |
| ZSCAN25 | 1147,83 | 0,03 | 0,03 | 0,86 | 3,88E-01 |
| IL6ST | 6051,92 | -0,06 | 0,07 | -0,86 | 3,88E-01 |
| POLR3A | 839,60 | 0,03 | 0,03 | 0,86 | 3,88E-01 |
| SLC22A5 | 295,38 | 0,06 | 0,07 | 0,86 | 3,88E-01 |
| AGO4 | 9829,17 | -0,05 | 0,05 | -0,86 | 3,88E-01 |
| ZNF516 | 5352,04 | 0,06 | 0,07 | 0,86 | 3,88E-01 |
| RLN1 | 52,47 | -0,07 | 0,08 | -0,86 | 3,88E-01 |
| TUBE1 | 601,28 | -0,05 | 0,06 | -0,86 | 3,89E-01 |
| FAAH2 | 262,33 | -0,07 | 0,08 | -0,86 | 3,88E-01 |
| AP2B1 | 4019,13 | 0,03 | 0,04 | 0,86 | 3,89E-01 |
| MEAK7 | 427,87 | 0,04 | 0,05 | 0,86 | 3,89E-01 |
| KNTC1 | 1049,95 | -0,03 | 0,04 | -0,86 | 3,89E-01 |
| DNAJB12 | 1399,33 | 0,04 | 0,04 | 0,86 | 3,90E-01 |
| TBC1D8B | 93,04 | 0,07 | 0,08 | 0,86 | 3,90E-01 |
| RNF181 | 593,21 | -0,04 | 0,04 | -0,86 | 3,90E-01 |

| | | | | | |
|---|---|---|---|---|---|
| MAP4K5 | 1649,06 | -0,04 | 0,04 | -0,86 | 3,90E-01 |
| REEP3 | 820,55 | -0,05 | 0,05 | -0,86 | 3,90E-01 |
| PANK2 | 3227,98 | -0,03 | 0,04 | -0,86 | 3,90E-01 |
| MAEA | 2043,03 | 0,05 | 0,05 | 0,86 | 3,90E-01 |
| ARHGAP18 | 881,59 | -0,04 | 0,05 | -0,86 | 3,90E-01 |
| MICB | 802,77 | 0,05 | 0,05 | 0,86 | 3,90E-01 |
| ACSL1 | 29093,46 | -0,08 | 0,09 | -0,86 | 3,90E-01 |
| NIPAL2 | 530,37 | -0,06 | 0,07 | -0,86 | 3,90E-01 |
| FGFBP3 | 113,97 | 0,05 | 0,05 | 0,86 | 3,90E-01 |
| EFNA5 | 44,37 | 0,13 | 0,15 | 0,86 | 3,91E-01 |
| HSF5 | 102,13 | 0,05 | 0,06 | 0,86 | 3,91E-01 |
| ALKBH7 | 585,58 | 0,04 | 0,05 | 0,86 | 3,91E-01 |
| SLC4A10 | 335,52 | -0,13 | 0,15 | -0,86 | 3,91E-01 |
| ORMDL3 | 3644,43 | 0,05 | 0,06 | 0,86 | 3,91E-01 |
| CNPY3 | 1449,50 | 0,05 | 0,05 | 0,86 | 3,91E-01 |
| NPIPB4 | 238,29 | 0,09 | 0,11 | 0,86 | 3,91E-01 |
| DNAAF2 | 191,91 | -0,05 | 0,05 | -0,86 | 3,91E-01 |
| MVP | 6291,87 | 0,04 | 0,05 | 0,86 | 3,92E-01 |
| ZFP3 | 263,60 | -0,04 | 0,05 | -0,86 | 3,92E-01 |
| GARS1 | 764,43 | -0,03 | 0,03 | -0,86 | 3,92E-01 |
| ILKAP | 707,56 | 0,04 | 0,04 | 0,86 | 3,92E-01 |
| TNFRSF17 | 33,40 | -0,12 | 0,14 | -0,85 | 3,93E-01 |
| EIF2AK4 | 935,68 | 0,03 | 0,03 | 0,85 | 3,93E-01 |
| SAFB | 2756,97 | -0,03 | 0,03 | -0,85 | 3,93E-01 |
| SURF4 | 1994,28 | 0,02 | 0,02 | 0,85 | 3,93E-01 |
| ARFGEF2 | 2282,49 | -0,02 | 0,03 | -0,85 | 3,93E-01 |
| RAB39A | 124,89 | 0,06 | 0,07 | 0,85 | 3,93E-01 |
| RC3H1 | 9096,13 | -0,03 | 0,04 | -0,85 | 3,93E-01 |
| TUBA4A | 2326,96 | 0,04 | 0,04 | 0,85 | 3,94E-01 |
| YLPM1 | 3195,08 | -0,03 | 0,04 | -0,85 | 3,94E-01 |
| MSRB2 | 362,00 | -0,05 | 0,06 | -0,85 | 3,94E-01 |
| XNDC1N | 546,03 | -0,04 | 0,04 | -0,85 | 3,94E-01 |
| ZNF202 | 391,45 | -0,03 | 0,03 | -0,85 | 3,94E-01 |
| FCRL1 | 1989,27 | -0,09 | 0,10 | -0,85 | 3,94E-01 |
| NDFIP1 | 2956,84 | -0,03 | 0,03 | -0,85 | 3,94E-01 |
| DHRS4 | 320,42 | -0,03 | 0,04 | -0,85 | 3,95E-01 |
| SETD7 | 1166,74 | -0,03 | 0,03 | -0,85 | 3,95E-01 |
| CETN2 | 239,42 | -0,06 | 0,07 | -0,85 | 3,95E-01 |
| MED11 | 455,20 | -0,03 | 0,04 | -0,85 | 3,95E-01 |
| POLD4 | 877,77 | 0,03 | 0,04 | 0,85 | 3,95E-01 |
| MTURN | 3176,54 | 0,05 | 0,06 | 0,85 | 3,95E-01 |
| PPP1R21 | 2285,24 | -0,03 | 0,04 | -0,85 | 3,96E-01 |
| KDM6A | 3049,36 | -0,03 | 0,04 | -0,85 | 3,96E-01 |
| RALB | 4207,37 | -0,05 | 0,06 | -0,85 | 3,96E-01 |
| GPR107 | 1900,45 | 0,05 | 0,05 | 0,85 | 3,96E-01 |
| CYBB | 19898,61 | -0,05 | 0,06 | -0,85 | 3,96E-01 |

| | | | | | |
|---|---|---|---|---|---|
| UQCC5 | 146,98 | 0,03 | 0,04 | 0,85 | 3,96E-01 |
| FCRL2 | 786,26 | -0,10 | 0,11 | -0,85 | 3,96E-01 |
| RETSAT | 463,96 | 0,03 | 0,03 | 0,85 | 3,97E-01 |
| WSB1 | 7987,24 | -0,04 | 0,05 | -0,85 | 3,97E-01 |
| RHOU | 595,13 | -0,05 | 0,05 | -0,85 | 3,97E-01 |
| FES | 2584,55 | 0,05 | 0,06 | 0,85 | 3,97E-01 |
| PRDX4 | 224,68 | -0,06 | 0,07 | -0,85 | 3,97E-01 |
| CSF3R | 34606,93 | 0,07 | 0,08 | 0,85 | 3,97E-01 |
| TMEM131L | 4777,53 | 0,03 | 0,04 | 0,85 | 3,98E-01 |
| KRI1 | 611,49 | 0,04 | 0,05 | 0,85 | 3,98E-01 |
| PPP1R15A | 996,75 | 0,06 | 0,07 | 0,85 | 3,98E-01 |
| PPM1D | 828,99 | -0,03 | 0,03 | -0,84 | 3,98E-01 |
| ATXN7L3B | 1830,13 | -0,02 | 0,03 | -0,84 | 3,98E-01 |
| EPB41 | 46321,67 | 0,08 | 0,10 | 0,84 | 3,98E-01 |
| NUP98 | 6837,20 | -0,03 | 0,04 | -0,84 | 3,99E-01 |
| PVR | 325,45 | 0,06 | 0,07 | 0,84 | 3,99E-01 |
| ZNF333 | 1422,98 | 0,03 | 0,04 | 0,84 | 3,99E-01 |
| DCPS | 327,42 | 0,03 | 0,03 | 0,84 | 3,99E-01 |
| TFRC | 2795,79 | 0,06 | 0,07 | 0,84 | 3,99E-01 |
| CXCL5 | 324,82 | 0,10 | 0,12 | 0,84 | 3,99E-01 |
| KCTD2 | 643,44 | 0,03 | 0,04 | 0,84 | 3,99E-01 |
| LY75 | 1030,03 | -0,04 | 0,05 | -0,84 | 4,00E-01 |
| ZZEF1 | 5860,28 | 0,04 | 0,05 | 0,84 | 4,00E-01 |
| TIMM13 | 407,57 | 0,04 | 0,04 | 0,84 | 4,00E-01 |
| CELF1 | 6465,81 | 0,03 | 0,04 | 0,84 | 4,00E-01 |
| CPEB2 | 2145,10 | -0,05 | 0,06 | -0,84 | 4,00E-01 |
| DIP2B | 7264,50 | -0,04 | 0,05 | -0,84 | 4,00E-01 |
| ZBTB18 | 2673,19 | -0,04 | 0,04 | -0,84 | 4,00E-01 |
| PRR5L | 2533,85 | -0,05 | 0,06 | -0,84 | 4,00E-01 |
| PARP16 | 223,19 | 0,05 | 0,06 | 0,84 | 4,01E-01 |
| EPB41L2 | 474,83 | -0,05 | 0,05 | -0,84 | 4,01E-01 |
| MSL1 | 9096,34 | -0,05 | 0,05 | -0,84 | 4,01E-01 |
| APOBEC3G | 1171,06 | -0,05 | 0,06 | -0,84 | 4,01E-01 |
| ASPM | 187,69 | 0,08 | 0,10 | 0,84 | 4,01E-01 |
| DRAXIN | 139,93 | 0,08 | 0,09 | 0,84 | 4,01E-01 |
| CCR7 | 1983,41 | 0,06 | 0,08 | 0,84 | 4,02E-01 |
| TRADD | 1590,86 | 0,03 | 0,03 | 0,84 | 4,02E-01 |
| TMEM81 | 152,60 | -0,03 | 0,04 | -0,84 | 4,03E-01 |
| COMMD4 | 271,78 | 0,03 | 0,04 | 0,84 | 4,03E-01 |
| NDFIP2 | 163,58 | 0,07 | 0,08 | 0,84 | 4,03E-01 |
| KDM2A | 5448,47 | 0,04 | 0,05 | 0,84 | 4,03E-01 |
| PLCD1 | 372,52 | 0,03 | 0,04 | 0,84 | 4,03E-01 |
| APOL2 | 4384,69 | -0,05 | 0,06 | -0,84 | 4,03E-01 |
| ZNF831 | 1861,81 | 0,05 | 0,05 | 0,84 | 4,03E-01 |
| IL18 | 178,91 | -0,05 | 0,06 | -0,84 | 4,03E-01 |
| PCDHGA7 | 40,91 | 0,09 | 0,11 | 0,84 | 4,03E-01 |

| | | | | | |
|---|---|---|---|---|---|
| APOBEC3D | 62,43 | 0,06 | 0,07 | 0,84 | 4,04E-01 |
| TAS2R13 | 135,83 | -0,07 | 0,08 | -0,83 | 4,04E-01 |
| ENDOG | 117,61 | 0,05 | 0,06 | 0,83 | 4,04E-01 |
| ZNF724 | 73,53 | -0,04 | 0,05 | -0,83 | 4,04E-01 |
| ANK3 | 2165,13 | 0,06 | 0,07 | 0,83 | 4,04E-01 |
| TECPR2 | 3900,28 | 0,06 | 0,08 | 0,83 | 4,04E-01 |
| OSM | 162,41 | 0,07 | 0,09 | 0,83 | 4,04E-01 |
| COG4 | 1421,55 | -0,02 | 0,02 | -0,83 | 4,04E-01 |
| C1GALT1 | 3184,65 | -0,05 | 0,06 | -0,83 | 4,04E-01 |
| NTHL1 | 153,95 | 0,04 | 0,04 | 0,83 | 4,05E-01 |
| RING1 | 443,01 | -0,02 | 0,03 | -0,83 | 4,05E-01 |
| FAAP24 | 89,69 | 0,05 | 0,06 | 0,83 | 4,05E-01 |
| SWSAP1 | 99,56 | 0,04 | 0,04 | 0,83 | 4,05E-01 |
| SMARCAL1 | 493,61 | 0,03 | 0,04 | 0,83 | 4,05E-01 |
| HSPA6 | 4112,71 | 0,07 | 0,08 | 0,83 | 4,05E-01 |
| LONRF1 | 953,42 | -0,03 | 0,04 | -0,83 | 4,05E-01 |
| UBE2D2 | 1498,15 | -0,02 | 0,03 | -0,83 | 4,05E-01 |
| KCNE1 | 803,81 | 0,06 | 0,08 | 0,83 | 4,06E-01 |
| LAT2 | 3758,13 | -0,04 | 0,05 | -0,83 | 4,06E-01 |
| NDC80 | 148,84 | -0,05 | 0,06 | -0,83 | 4,06E-01 |
| GALNT10 | 2004,33 | 0,04 | 0,04 | 0,83 | 4,07E-01 |
| PPP4C | 2129,37 | -0,03 | 0,04 | -0,83 | 4,07E-01 |
| CLPP | 251,99 | 0,03 | 0,04 | 0,83 | 4,07E-01 |
| SECISBP2 | 5049,17 | -0,03 | 0,03 | -0,83 | 4,07E-01 |
| LGALSL | 610,47 | 0,07 | 0,08 | 0,83 | 4,07E-01 |
| SHMT1 | 230,27 | 0,05 | 0,06 | 0,83 | 4,08E-01 |
| CDK9 | 1366,11 | 0,02 | 0,03 | 0,83 | 4,08E-01 |
| TBC1D22A | 2537,34 | 0,04 | 0,04 | 0,83 | 4,08E-01 |
| KATNAL1 | 498,03 | -0,04 | 0,05 | -0,83 | 4,08E-01 |
| FUT8 | 1002,08 | -0,04 | 0,05 | -0,83 | 4,08E-01 |
| CCZ1 | 574,26 | 0,03 | 0,04 | 0,83 | 4,08E-01 |
| ACTN4 | 2585,10 | 0,06 | 0,08 | 0,83 | 4,08E-01 |
| MRPS26 | 349,10 | -0,03 | 0,04 | -0,83 | 4,08E-01 |
| CST3 | 1697,90 | 0,06 | 0,07 | 0,83 | 4,09E-01 |
| ZNF608 | 169,36 | 0,08 | 0,10 | 0,83 | 4,09E-01 |
| CCNDBP1 | 5666,58 | 0,04 | 0,05 | 0,83 | 4,09E-01 |
| S100P | 1961,15 | -0,13 | 0,16 | -0,83 | 4,09E-01 |
| STK16 | 634,52 | 0,03 | 0,04 | 0,83 | 4,09E-01 |
| CLCF1 | 54,53 | 0,07 | 0,09 | 0,83 | 4,09E-01 |
| ULK4 | 1054,17 | -0,04 | 0,04 | -0,83 | 4,09E-01 |
| PTCD2 | 437,59 | 0,03 | 0,04 | 0,82 | 4,10E-01 |
| IFITM1 | 3364,97 | 0,06 | 0,08 | 0,82 | 4,10E-01 |
| CTSZ | 4820,78 | -0,04 | 0,05 | -0,82 | 4,10E-01 |
| KAT6B | 2338,22 | -0,03 | 0,04 | -0,82 | 4,10E-01 |
| CPA3 | 141,60 | -0,14 | 0,17 | -0,82 | 4,10E-01 |
| PACRGL | 425,22 | -0,03 | 0,04 | -0,82 | 4,10E-01 |

| | | | | | |
|---|---|---|---|---|---|
| CCR8 | 52,02 | -0,08 | 0,10 | -0,82 | 4,11E-01 |
| RPS27L | 832,83 | -0,05 | 0,06 | -0,82 | 4,11E-01 |
| KLHL18 | 1294,03 | 0,03 | 0,04 | 0,82 | 4,11E-01 |
| TTC9C | 575,56 | -0,02 | 0,02 | -0,82 | 4,11E-01 |
| THAP2 | 301,95 | -0,05 | 0,06 | -0,82 | 4,11E-01 |
| SDHA | 2207,48 | -0,02 | 0,03 | -0,82 | 4,11E-01 |
| LRRC8B | 1208,19 | 0,04 | 0,05 | 0,82 | 4,12E-01 |
| POLDIP2 | 853,34 | 0,02 | 0,03 | 0,82 | 4,12E-01 |
| KLF11 | 480,23 | -0,03 | 0,04 | -0,82 | 4,12E-01 |
| NFAT5 | 5439,96 | 0,04 | 0,05 | 0,82 | 4,13E-01 |
| ARL6IP4 | 54,68 | 0,04 | 0,05 | 0,82 | 4,13E-01 |
| NCBP2AS2 | 253,55 | -0,04 | 0,05 | -0,82 | 4,14E-01 |
| SLC33A1 | 694,55 | -0,03 | 0,04 | -0,82 | 4,14E-01 |
| ATRN | 1251,34 | -0,02 | 0,03 | -0,82 | 4,14E-01 |
| METTL8 | 717,67 | -0,05 | 0,06 | -0,82 | 4,14E-01 |
| HECTD2 | 133,53 | 0,08 | 0,09 | 0,82 | 4,14E-01 |
| CIAPIN1 | 491,93 | -0,02 | 0,02 | -0,82 | 4,14E-01 |
| NAIF1 | 391,61 | 0,02 | 0,03 | 0,82 | 4,15E-01 |
| RBM6 | 3051,14 | -0,03 | 0,04 | -0,82 | 4,15E-01 |
| CTSH | 1442,86 | 0,04 | 0,05 | 0,82 | 4,15E-01 |
| RACGAP1 | 299,90 | 0,03 | 0,04 | 0,82 | 4,15E-01 |
| TTBK2 | 1789,13 | -0,02 | 0,02 | -0,81 | 4,15E-01 |
| HADHB | 2390,95 | -0,02 | 0,03 | -0,81 | 4,15E-01 |
| RPA4 | 62,22 | -0,04 | 0,05 | -0,81 | 4,15E-01 |
| POP1 | 277,81 | -0,03 | 0,04 | -0,81 | 4,15E-01 |
| ARL17A | 258,39 | 0,11 | 0,13 | 0,81 | 4,16E-01 |
| ISG20 | 2911,79 | 0,05 | 0,06 | 0,81 | 4,16E-01 |
| NDE1 | 2670,41 | 0,06 | 0,07 | 0,81 | 4,16E-01 |
| LYSMD2 | 480,36 | 0,04 | 0,05 | 0,81 | 4,16E-01 |
| FAR2 | 1563,12 | 0,05 | 0,06 | 0,81 | 4,16E-01 |
| CD82 | 1853,59 | 0,04 | 0,05 | 0,81 | 4,16E-01 |
| ZNF793 | 286,57 | 0,05 | 0,06 | 0,81 | 4,16E-01 |
| PDK3 | 1994,33 | -0,04 | 0,05 | -0,81 | 4,17E-01 |
| ZNF273 | 323,89 | -0,05 | 0,06 | -0,81 | 4,17E-01 |
| C9orf72 | 7461,23 | -0,06 | 0,07 | -0,81 | 4,17E-01 |
| COBLL1 | 623,17 | -0,07 | 0,09 | -0,81 | 4,17E-01 |
| LST1 | 5337,31 | 0,04 | 0,05 | 0,81 | 4,17E-01 |
| TSTD1 | 579,36 | -0,03 | 0,04 | -0,81 | 4,17E-01 |
| NCAPD3 | 916,26 | 0,03 | 0,03 | 0,81 | 4,18E-01 |
| EIF2B1 | 971,79 | -0,02 | 0,02 | -0,81 | 4,18E-01 |
| P2RY13 | 1593,93 | 0,06 | 0,07 | 0,81 | 4,18E-01 |
| HIGD1A | 402,73 | -0,07 | 0,08 | -0,81 | 4,18E-01 |
| HECW2 | 570,84 | -0,07 | 0,09 | -0,81 | 4,18E-01 |
| PCCA | 574,47 | -0,04 | 0,04 | -0,81 | 4,18E-01 |
| SMYD4 | 737,44 | 0,03 | 0,03 | 0,81 | 4,18E-01 |
| CGGBP1 | 4809,94 | -0,04 | 0,06 | -0,81 | 4,19E-01 |

| | | | | | |
|---|---|---|---|---|---|
| ARFIP2 | 443,77 | 0,02 | 0,02 | 0,81 | 4,19E-01 |
| PCDHGB5 | 37,08 | 0,09 | 0,11 | 0,81 | 4,19E-01 |
| STAT6 | 8094,19 | 0,05 | 0,06 | 0,81 | 4,19E-01 |
| GTF3C4 | 647,97 | -0,02 | 0,02 | -0,81 | 4,19E-01 |
| SNRPA1 | 790,08 | -0,03 | 0,03 | -0,81 | 4,19E-01 |
| USP27X | 103,69 | -0,04 | 0,05 | -0,81 | 4,20E-01 |
| HKDC1 | 176,19 | 0,08 | 0,10 | 0,81 | 4,20E-01 |
| H3C11 | 484,87 | 0,05 | 0,07 | 0,81 | 4,20E-01 |
| TUBA1A | 4402,02 | -0,04 | 0,06 | -0,81 | 4,20E-01 |
| TRAPPC3 | 1090,12 | -0,02 | 0,03 | -0,81 | 4,20E-01 |
| COLGALT1 | 1544,89 | 0,03 | 0,04 | 0,81 | 4,21E-01 |
| SEC31A | 2226,37 | -0,02 | 0,03 | -0,81 | 4,21E-01 |
| DSC2 | 1402,61 | 0,09 | 0,11 | 0,80 | 4,21E-01 |
| DNAL4 | 131,29 | -0,03 | 0,04 | -0,80 | 4,21E-01 |
| RTN4IP1 | 223,32 | -0,03 | 0,04 | -0,80 | 4,21E-01 |
| PAAF1 | 370,73 | 0,03 | 0,04 | 0,80 | 4,21E-01 |
| CD74 | 4548,65 | 0,08 | 0,10 | 0,80 | 4,22E-01 |
| NTAQ1 | 195,01 | 0,04 | 0,05 | 0,80 | 4,22E-01 |
| EMC3 | 3001,06 | 0,06 | 0,08 | 0,80 | 4,22E-01 |
| ZGPAT | 593,61 | 0,03 | 0,03 | 0,80 | 4,22E-01 |
| SLC20A1 | 3238,11 | -0,03 | 0,04 | -0,80 | 4,22E-01 |
| B3GNT5 | 614,57 | 0,06 | 0,07 | 0,80 | 4,22E-01 |
| PCBD2 | 464,60 | 0,03 | 0,03 | 0,80 | 4,23E-01 |
| PDE5A | 931,26 | 0,07 | 0,09 | 0,80 | 4,23E-01 |
| TSEN2 | 425,18 | 0,03 | 0,04 | 0,80 | 4,23E-01 |
| MTFP1 | 34,19 | -0,05 | 0,07 | -0,80 | 4,23E-01 |
| NAPEPLD | 551,57 | -0,03 | 0,03 | -0,80 | 4,23E-01 |
| TOR1A | 935,34 | 0,03 | 0,03 | 0,80 | 4,23E-01 |
| BRIP1 | 235,81 | 0,05 | 0,07 | 0,80 | 4,24E-01 |
| SCRN1 | 907,71 | -0,04 | 0,05 | -0,80 | 4,24E-01 |
| CERS2 | 2376,08 | 0,03 | 0,04 | 0,80 | 4,24E-01 |
| USF1 | 3564,03 | -0,03 | 0,04 | -0,80 | 4,24E-01 |
| NUP214 | 3474,68 | 0,03 | 0,04 | 0,80 | 4,24E-01 |
| BTNL3 | 85,35 | 0,28 | 0,35 | 0,80 | 4,24E-01 |
| SLC49A4 | 1405,78 | -0,04 | 0,05 | -0,80 | 4,24E-01 |
| PDLIM5 | 1677,72 | 0,03 | 0,03 | 0,80 | 4,25E-01 |
| FPR2 | 6642,87 | -0,06 | 0,08 | -0,80 | 4,25E-01 |
| SELENOK | 1761,54 | 0,07 | 0,09 | 0,80 | 4,25E-01 |
| RPS19BP1 | 676,91 | 0,03 | 0,03 | 0,80 | 4,25E-01 |
| H2AC11 | 1451,58 | -0,07 | 0,08 | -0,80 | 4,25E-01 |
| EFNA4 | 114,35 | 0,03 | 0,04 | 0,80 | 4,25E-01 |
| UST | 294,00 | 0,05 | 0,06 | 0,80 | 4,25E-01 |
| DNAJB6 | 4245,80 | -0,04 | 0,05 | -0,80 | 4,25E-01 |
| ATG14 | 1008,06 | -0,02 | 0,03 | -0,80 | 4,25E-01 |
| FBXO42 | 1472,39 | 0,02 | 0,03 | 0,80 | 4,26E-01 |
| PDRG1 | 301,46 | -0,03 | 0,03 | -0,80 | 4,26E-01 |

| | | | | | |
|---|---|---|---|---|---|
| PRR13 | 659,65 | 0,05 | 0,07 | 0,80 | 4,26E-01 |
| NCAPH2 | 597,56 | 0,03 | 0,03 | 0,80 | 4,26E-01 |
| SGPL1 | 1570,44 | 0,03 | 0,04 | 0,80 | 4,26E-01 |
| NABP1 | 21763,67 | -0,05 | 0,06 | -0,80 | 4,27E-01 |
| C19orf12 | 776,71 | -0,03 | 0,03 | -0,79 | 4,27E-01 |
| RAF1 | 8015,85 | -0,04 | 0,05 | -0,79 | 4,27E-01 |
| PRADC1 | 125,66 | 0,04 | 0,05 | 0,79 | 4,27E-01 |
| PUF60 | 1056,74 | -0,02 | 0,03 | -0,79 | 4,27E-01 |
| TACO1 | 270,38 | -0,02 | 0,03 | -0,79 | 4,28E-01 |
| RETREG3 | 2586,52 | -0,02 | 0,02 | -0,79 | 4,28E-01 |
| SIN3A | 2101,96 | -0,03 | 0,04 | -0,79 | 4,28E-01 |
| MFSD9 | 624,16 | -0,06 | 0,08 | -0,79 | 4,28E-01 |
| DYNC2LI1 | 98,68 | 0,06 | 0,07 | 0,79 | 4,28E-01 |
| CCR9 | 44,86 | 0,09 | 0,11 | 0,79 | 4,29E-01 |
| UGT8 | 87,30 | -0,08 | 0,11 | -0,79 | 4,29E-01 |
| UBQLN1 | 4669,50 | -0,03 | 0,03 | -0,79 | 4,29E-01 |
| TIAM1 | 2168,71 | 0,04 | 0,05 | 0,79 | 4,29E-01 |
| TAP2 | 1945,83 | 0,05 | 0,06 | 0,79 | 4,29E-01 |
| EGLN2 | 1588,87 | 0,04 | 0,05 | 0,79 | 4,29E-01 |
| SGK1 | 3003,13 | -0,05 | 0,06 | -0,79 | 4,29E-01 |
| LIPN | 525,82 | -0,09 | 0,11 | -0,79 | 4,29E-01 |
| DNAJA3 | 840,89 | 0,02 | 0,03 | 0,79 | 4,30E-01 |
| C1orf74 | 104,73 | -0,04 | 0,05 | -0,79 | 4,30E-01 |
| PLPP5 | 491,31 | -0,05 | 0,06 | -0,79 | 4,31E-01 |
| ELAVL1 | 689,60 | 0,03 | 0,04 | 0,79 | 4,31E-01 |
| SESTD1 | 1171,93 | -0,05 | 0,06 | -0,79 | 4,31E-01 |
| EXD2 | 294,90 | 0,03 | 0,04 | 0,79 | 4,31E-01 |
| SNRNP35 | 750,57 | -0,02 | 0,02 | -0,79 | 4,31E-01 |
| IL7 | 99,60 | -0,07 | 0,09 | -0,79 | 4,31E-01 |
| ADPRH | 257,29 | 0,04 | 0,05 | 0,79 | 4,31E-01 |
| LSM4 | 668,87 | -0,03 | 0,04 | -0,79 | 4,32E-01 |
| ARMCX1 | 52,15 | 0,06 | 0,08 | 0,79 | 4,32E-01 |
| RIN2 | 547,78 | 0,05 | 0,07 | 0,79 | 4,32E-01 |
| TUBB | 3193,07 | 0,03 | 0,03 | 0,79 | 4,32E-01 |
| ACAD9 | 998,69 | 0,03 | 0,03 | 0,79 | 4,32E-01 |
| STX11 | 2086,68 | 0,06 | 0,07 | 0,79 | 4,32E-01 |
| PCDHGB6 | 69,60 | 0,08 | 0,11 | 0,79 | 4,32E-01 |
| L3HYPDH | 551,06 | -0,03 | 0,04 | -0,78 | 4,33E-01 |
| NOTCH2NLA | 549,37 | -0,06 | 0,07 | -0,78 | 4,32E-01 |
| BLCAP | 1712,26 | 0,04 | 0,05 | 0,78 | 4,33E-01 |
| ABI2 | 896,73 | 0,04 | 0,05 | 0,78 | 4,33E-01 |
| CHMP4B | 796,22 | 0,04 | 0,05 | 0,78 | 4,33E-01 |
| PSME2 | 492,78 | 0,05 | 0,06 | 0,78 | 4,33E-01 |
| DHX32 | 347,19 | -0,03 | 0,04 | -0,78 | 4,34E-01 |
| SLC35F2 | 152,85 | 0,05 | 0,06 | 0,78 | 4,34E-01 |
| MS4A14 | 389,56 | 0,06 | 0,07 | 0,78 | 4,34E-01 |

| | | | | | |
|---|---|---|---|---|---|
| DYM | 1936,85 | -0,02 | 0,03 | -0,78 | 4,35E-01 |
| ISCA1 | 2643,41 | 0,07 | 0,09 | 0,78 | 4,35E-01 |
| USP18 | 119,54 | 0,12 | 0,15 | 0,78 | 4,35E-01 |
| C9orf153 | 42,11 | 0,06 | 0,08 | 0,78 | 4,35E-01 |
| CLEC7A | 11984,02 | -0,05 | 0,06 | -0,78 | 4,35E-01 |
| LMO2 | 2063,79 | -0,04 | 0,06 | -0,78 | 4,35E-01 |
| ADD1 | 4495,27 | 0,05 | 0,06 | 0,78 | 4,35E-01 |
| ZNF621 | 814,95 | -0,03 | 0,03 | -0,78 | 4,35E-01 |
| NDUFV3 | 1110,65 | 0,02 | 0,03 | 0,78 | 4,36E-01 |
| LATS2 | 690,72 | 0,04 | 0,05 | 0,78 | 4,36E-01 |
| EMILIN2 | 2160,66 | 0,04 | 0,06 | 0,78 | 4,36E-01 |
| PACSIN2 | 5850,63 | -0,04 | 0,06 | -0,78 | 4,36E-01 |
| TTC4 | 79,51 | -0,04 | 0,05 | -0,78 | 4,36E-01 |
| DENND5A | 6694,70 | 0,05 | 0,06 | 0,78 | 4,36E-01 |
| SPMIP4 | 78,43 | 0,05 | 0,06 | 0,78 | 4,36E-01 |
| NDUFB2 | 654,40 | -0,04 | 0,05 | -0,78 | 4,36E-01 |
| VPS18 | 1070,99 | -0,03 | 0,03 | -0,78 | 4,36E-01 |
| OS9 | 2230,77 | 0,04 | 0,06 | 0,78 | 4,36E-01 |
| TFEC | 1775,52 | -0,08 | 0,10 | -0,78 | 4,37E-01 |
| TBC1D9 | 1331,25 | -0,04 | 0,05 | -0,78 | 4,36E-01 |
| CST7 | 402,20 | 0,07 | 0,09 | 0,78 | 4,37E-01 |
| NAXD | 728,00 | 0,02 | 0,03 | 0,78 | 4,37E-01 |
| ANKFY1 | 2835,24 | 0,04 | 0,05 | 0,78 | 4,37E-01 |
| LYRM4 | 336,12 | 0,04 | 0,05 | 0,78 | 4,37E-01 |
| SOWAHD | 283,43 | 0,04 | 0,06 | 0,78 | 4,37E-01 |
| GOLGA8O | 35,38 | -0,11 | 0,14 | -0,78 | 4,37E-01 |
| LRIG2 | 1309,28 | 0,03 | 0,04 | 0,78 | 4,37E-01 |
| R3HDM2 | 802,88 | 0,05 | 0,06 | 0,78 | 4,37E-01 |
| FCGR3A | 5365,47 | -0,05 | 0,07 | -0,78 | 4,37E-01 |
| HMGN5 | 35,37 | 0,07 | 0,09 | 0,78 | 4,38E-01 |
| ZMAT2 | 2898,81 | -0,07 | 0,09 | -0,77 | 4,38E-01 |
| NBPF15 | 1283,80 | 0,05 | 0,06 | 0,77 | 4,39E-01 |
| ACSL5 | 1695,79 | -0,02 | 0,03 | -0,77 | 4,39E-01 |
| TAF10 | 902,97 | 0,02 | 0,03 | 0,77 | 4,39E-01 |
| ANKEF1 | 206,74 | -0,05 | 0,06 | -0,77 | 4,39E-01 |
| TMEM41B | 1142,26 | -0,03 | 0,04 | -0,77 | 4,39E-01 |
| NIBAN3 | 1233,88 | -0,07 | 0,09 | -0,77 | 4,39E-01 |
| SEC13 | 1201,93 | -0,02 | 0,02 | -0,77 | 4,40E-01 |
| CAP1 | 7880,62 | -0,04 | 0,05 | -0,77 | 4,40E-01 |
| GALC | 2017,57 | -0,03 | 0,04 | -0,77 | 4,40E-01 |
| BCL7C | 455,22 | -0,03 | 0,04 | -0,77 | 4,40E-01 |
| APBA2 | 879,52 | 0,06 | 0,07 | 0,77 | 4,40E-01 |
| RASGEF1A | 234,58 | 0,05 | 0,06 | 0,77 | 4,40E-01 |
| VCL | 4875,51 | 0,05 | 0,06 | 0,77 | 4,41E-01 |
| BCAT1 | 467,53 | -0,08 | 0,10 | -0,77 | 4,41E-01 |
| B4GALT1 | 1917,03 | 0,04 | 0,06 | 0,77 | 4,41E-01 |

| | | | | | |
|---|---|---|---|---|---|
| RBM15B | 1433,47 | -0,02 | 0,02 | -0,77 | 4,41E-01 |
| ZNRF2 | 1792,17 | -0,04 | 0,05 | -0,77 | 4,41E-01 |
| SMPDL3A | 197,12 | -0,05 | 0,06 | -0,77 | 4,41E-01 |
| APOBEC3A | 5299,77 | 0,07 | 0,09 | 0,77 | 4,42E-01 |
| CNPY2 | 253,17 | -0,03 | 0,03 | -0,77 | 4,42E-01 |
| C12orf57 | 831,93 | -0,06 | 0,08 | -0,77 | 4,42E-01 |
| IFIT3 | 5135,45 | -0,12 | 0,16 | -0,77 | 4,42E-01 |
| LAMC1 | 190,03 | 0,08 | 0,10 | 0,77 | 4,42E-01 |
| TRIM21 | 1938,98 | -0,04 | 0,06 | -0,77 | 4,42E-01 |
| ARL2 | 162,09 | -0,03 | 0,04 | -0,77 | 4,43E-01 |
| REELD1 | 49,36 | 0,09 | 0,12 | 0,77 | 4,43E-01 |
| IL1RN | 650,73 | 0,07 | 0,09 | 0,77 | 4,44E-01 |
| PSMD3 | 792,00 | 0,02 | 0,03 | 0,77 | 4,44E-01 |
| KIAA1328 | 533,80 | -0,03 | 0,04 | -0,77 | 4,44E-01 |
| H3-3A | 11142,78 | -0,04 | 0,06 | -0,77 | 4,44E-01 |
| C19orf53 | 308,54 | 0,04 | 0,05 | 0,77 | 4,44E-01 |
| OXNAD1 | 5381,66 | -0,05 | 0,07 | -0,76 | 4,44E-01 |
| CHD3 | 5041,95 | 0,03 | 0,04 | 0,76 | 4,45E-01 |
| MOB1B | 1440,31 | -0,04 | 0,05 | -0,76 | 4,45E-01 |
| ISCU | 3141,49 | 0,04 | 0,05 | 0,76 | 4,45E-01 |
| PARP2 | 121,43 | -0,04 | 0,05 | -0,76 | 4,46E-01 |
| NLN | 308,64 | 0,05 | 0,06 | 0,76 | 4,46E-01 |
| HIRIP3 | 216,76 | -0,03 | 0,04 | -0,76 | 4,46E-01 |
| FCGR1A | 637,51 | 0,11 | 0,14 | 0,76 | 4,46E-01 |
| GAB1 | 1871,87 | -0,04 | 0,05 | -0,76 | 4,46E-01 |
| TMIGD2 | 186,91 | -0,07 | 0,09 | -0,76 | 4,46E-01 |
| PIGW | 141,29 | 0,03 | 0,04 | 0,76 | 4,46E-01 |
| RAB19 | 23,51 | 0,06 | 0,08 | 0,76 | 4,46E-01 |
| ERI3 | 498,78 | 0,03 | 0,04 | 0,76 | 4,47E-01 |
| PARP12 | 2555,93 | 0,06 | 0,08 | 0,76 | 4,48E-01 |
| ANXA3 | 1107,13 | -0,09 | 0,11 | -0,76 | 4,48E-01 |
| TMEM9B | 1257,12 | -0,04 | 0,05 | -0,76 | 4,48E-01 |
| BMP2K | 4595,24 | -0,04 | 0,05 | -0,76 | 4,49E-01 |
| RRS1 | 250,68 | -0,03 | 0,04 | -0,76 | 4,49E-01 |
| SPAG16 | 150,42 | -0,05 | 0,07 | -0,76 | 4,49E-01 |
| PPBP | 4381,73 | 0,10 | 0,13 | 0,76 | 4,49E-01 |
| ZMYM6 | 900,37 | -0,03 | 0,03 | -0,76 | 4,49E-01 |
| TUBGCP3 | 2302,31 | 0,03 | 0,04 | 0,76 | 4,49E-01 |
| RPL27 | 3469,09 | -0,08 | 0,11 | -0,76 | 4,50E-01 |
| ZNF891 | 991,59 | -0,03 | 0,04 | -0,76 | 4,50E-01 |
| XPO6 | 22415,25 | 0,05 | 0,06 | 0,75 | 4,50E-01 |
| ABCF1 | 811,61 | 0,03 | 0,04 | 0,75 | 4,50E-01 |
| OSBPL2 | 3938,84 | -0,04 | 0,05 | -0,75 | 4,50E-01 |
| SELENOM | 112,03 | 0,05 | 0,07 | 0,75 | 4,50E-01 |
| ATG10 | 284,88 | 0,04 | 0,05 | 0,75 | 4,50E-01 |
| OXLD1 | 755,67 | -0,03 | 0,04 | -0,75 | 4,50E-01 |

| | | | | | |
|---|---|---|---|---|---|
| CCM2 | 4326,68 | -0,03 | 0,04 | -0,75 | 4,51E-01 |
| RBM23 | 4228,64 | -0,03 | 0,04 | -0,75 | 4,51E-01 |
| CRCP | 835,05 | 0,02 | 0,03 | 0,75 | 4,51E-01 |
| TRAPPC14 | 1062,74 | 0,05 | 0,06 | 0,75 | 4,51E-01 |
| ENSG00000148362 | 443,11 | 0,04 | 0,05 | 0,75 | 4,51E-01 |
| GEMIN4 | 440,67 | -0,03 | 0,04 | -0,75 | 4,52E-01 |
| HACD4 | 3974,36 | -0,03 | 0,05 | -0,75 | 4,52E-01 |
| HOMER3 | 225,54 | 0,05 | 0,06 | 0,75 | 4,52E-01 |
| BTK | 2092,36 | -0,03 | 0,04 | -0,75 | 4,52E-01 |
| NRG1 | 271,17 | 0,11 | 0,14 | 0,75 | 4,52E-01 |
| RAD51AP1 | 50,65 | 0,07 | 0,10 | 0,75 | 4,52E-01 |
| STYK1 | 94,75 | 0,09 | 0,12 | 0,75 | 4,52E-01 |
| PDE4DIP | 1092,18 | -0,03 | 0,04 | -0,75 | 4,52E-01 |
| LILRA5 | 3232,93 | -0,06 | 0,08 | -0,75 | 4,52E-01 |
| RBM19 | 1298,20 | 0,03 | 0,04 | 0,75 | 4,52E-01 |
| ARL3 | 167,00 | -0,03 | 0,04 | -0,75 | 4,53E-01 |
| PLK4 | 85,86 | 0,06 | 0,08 | 0,75 | 4,53E-01 |
| NT5DC3 | 292,10 | 0,06 | 0,08 | 0,75 | 4,54E-01 |
| TNKS | 2552,84 | -0,02 | 0,03 | -0,75 | 4,54E-01 |
| GAB3 | 1708,27 | -0,04 | 0,05 | -0,75 | 4,54E-01 |
| LRRC70 | 45,16 | -0,07 | 0,09 | -0,75 | 4,54E-01 |
| CYREN | 863,68 | -0,02 | 0,02 | -0,75 | 4,54E-01 |
| TRAF3IP1 | 267,25 | 0,03 | 0,04 | 0,75 | 4,54E-01 |
| VCAN | 27496,78 | -0,05 | 0,07 | -0,75 | 4,55E-01 |
| GCSAM | 394,79 | -0,05 | 0,06 | -0,75 | 4,55E-01 |
| YBX1 | 12655,64 | 0,07 | 0,09 | 0,75 | 4,55E-01 |
| CD33 | 507,22 | 0,05 | 0,07 | 0,75 | 4,55E-01 |
| OAS1 | 2017,31 | 0,10 | 0,13 | 0,75 | 4,55E-01 |
| ZNF354B | 395,83 | -0,04 | 0,05 | -0,75 | 4,55E-01 |
| GEN1 | 609,25 | -0,05 | 0,07 | -0,75 | 4,55E-01 |
| AURKAIP1 | 1231,04 | -0,03 | 0,04 | -0,75 | 4,55E-01 |
| WEE1 | 307,95 | 0,05 | 0,07 | 0,75 | 4,55E-01 |
| MBTPS1 | 3022,30 | 0,02 | 0,03 | 0,75 | 4,56E-01 |
| DDX3Y | 2399,68 | 0,13 | 0,18 | 0,75 | 4,56E-01 |
| TRAF4 | 171,92 | 0,04 | 0,06 | 0,74 | 4,56E-01 |
| POLG | 3337,36 | 0,02 | 0,03 | 0,74 | 4,57E-01 |
| ERMN | 433,86 | -0,04 | 0,06 | -0,74 | 4,57E-01 |
| ATP5MC2 | 1433,49 | -0,02 | 0,03 | -0,74 | 4,57E-01 |
| WWOX | 458,81 | 0,04 | 0,05 | 0,74 | 4,58E-01 |
| KPNA2 | 102,53 | 0,05 | 0,06 | 0,74 | 4,57E-01 |
| ICOS | 585,47 | -0,05 | 0,07 | -0,74 | 4,58E-01 |
| TTN | 6721,19 | 0,06 | 0,08 | 0,74 | 4,58E-01 |
| KLHDC2 | 1539,30 | -0,03 | 0,05 | -0,74 | 4,59E-01 |
| NAXE | 250,45 | -0,03 | 0,04 | -0,74 | 4,59E-01 |
| NHSL2 | 4788,40 | 0,06 | 0,08 | 0,74 | 4,59E-01 |
| P2RY12 | 211,80 | 0,07 | 0,09 | 0,74 | 4,59E-01 |

| | | | | | |
|---|---|---|---|---|---|
| HELLS | 475,12 | 0,04 | 0,06 | 0,74 | 4,59E-01 |
| RNF149 | 22971,42 | -0,04 | 0,06 | -0,74 | 4,59E-01 |
| ACTR5 | 426,75 | -0,02 | 0,03 | -0,74 | 4,60E-01 |
| SPTSSA | 271,79 | -0,04 | 0,06 | -0,74 | 4,60E-01 |
| AMPD2 | 2930,69 | 0,06 | 0,08 | 0,74 | 4,60E-01 |
| CDKN2AIPNL | 208,80 | -0,03 | 0,05 | -0,74 | 4,60E-01 |
| SCRN2 | 235,19 | 0,03 | 0,05 | 0,74 | 4,60E-01 |
| MXRA7 | 256,37 | -0,09 | 0,12 | -0,74 | 4,60E-01 |
| ABHD17A | 3696,53 | 0,03 | 0,04 | 0,74 | 4,61E-01 |
| ZNF682 | 193,70 | -0,03 | 0,05 | -0,74 | 4,61E-01 |
| ZNF786 | 347,07 | 0,02 | 0,03 | 0,74 | 4,61E-01 |
| CYTH4 | 11967,64 | 0,03 | 0,05 | 0,74 | 4,61E-01 |
| DEXI | 543,22 | -0,02 | 0,03 | -0,74 | 4,61E-01 |
| C11orf21 | 3135,22 | 0,03 | 0,05 | 0,74 | 4,61E-01 |
| PRDM5 | 611,39 | -0,08 | 0,11 | -0,74 | 4,61E-01 |
| GPCPD1 | 11547,69 | -0,04 | 0,05 | -0,74 | 4,62E-01 |
| PSMD2 | 2133,40 | 0,03 | 0,04 | 0,74 | 4,62E-01 |
| PECAM1 | 10497,18 | -0,04 | 0,05 | -0,74 | 4,62E-01 |
| TSPYL2 | 366,56 | 0,04 | 0,05 | 0,74 | 4,62E-01 |
| SH3KBP1 | 3284,58 | -0,03 | 0,05 | -0,74 | 4,62E-01 |
| PIGM | 615,15 | -0,02 | 0,03 | -0,74 | 4,62E-01 |
| TMEM176A | 166,60 | -0,16 | 0,22 | -0,74 | 4,62E-01 |
| SLC31A1 | 936,46 | -0,04 | 0,05 | -0,74 | 4,62E-01 |
| MDN1 | 3734,76 | 0,04 | 0,05 | 0,74 | 4,62E-01 |
| TAGLN | 436,98 | 0,08 | 0,11 | 0,74 | 4,62E-01 |
| ENSG00000249141 | 27,83 | -0,07 | 0,10 | -0,74 | 4,62E-01 |
| TOB1 | 615,71 | -0,03 | 0,04 | -0,73 | 4,63E-01 |
| H2AC14 | 266,98 | 0,09 | 0,13 | 0,73 | 4,63E-01 |
| DIAPH1 | 4078,74 | 0,05 | 0,07 | 0,73 | 4,64E-01 |
| DDX55 | 621,05 | -0,02 | 0,03 | -0,73 | 4,64E-01 |
| SSU72 | 2182,14 | 0,03 | 0,04 | 0,73 | 4,65E-01 |
| ALPK1 | 6536,73 | 0,05 | 0,07 | 0,73 | 4,66E-01 |
| OPTN | 3337,43 | 0,06 | 0,08 | 0,73 | 4,66E-01 |
| ADCK2 | 297,98 | 0,03 | 0,04 | 0,73 | 4,66E-01 |
| DGCR6L | 364,94 | 0,03 | 0,04 | 0,73 | 4,66E-01 |
| FKBPL | 60,40 | 0,04 | 0,05 | 0,73 | 4,66E-01 |
| MTMR3 | 6586,68 | 0,05 | 0,06 | 0,73 | 4,66E-01 |
| JAML | 26519,45 | -0,04 | 0,05 | -0,73 | 4,67E-01 |
| FNDC3B | 6561,17 | -0,04 | 0,05 | -0,73 | 4,67E-01 |
| ROPN1L | 441,40 | 0,05 | 0,07 | 0,73 | 4,67E-01 |
| SAMD9L | 6432,93 | -0,08 | 0,10 | -0,73 | 4,67E-01 |
| NEU1 | 853,47 | -0,03 | 0,04 | -0,73 | 4,67E-01 |
| HEMGN | 3038,73 | -0,09 | 0,12 | -0,73 | 4,68E-01 |
| RNASEH2C | 781,83 | 0,03 | 0,04 | 0,73 | 4,68E-01 |
| TLE3 | 5686,43 | 0,05 | 0,08 | 0,72 | 4,68E-01 |
| ASXL1 | 2374,99 | 0,03 | 0,04 | 0,72 | 4,69E-01 |

| | | | | | |
|---|---|---|---|---|---|
| KIF16B | 683,41 | -0,04 | 0,06 | -0,72 | 4,69E-01 |
| PAFAH1B1 | 2665,03 | -0,03 | 0,04 | -0,72 | 4,69E-01 |
| NIBAN1 | 23826,75 | -0,05 | 0,07 | -0,72 | 4,69E-01 |
| SPICE1 | 295,26 | -0,03 | 0,04 | -0,72 | 4,69E-01 |
| FIBP | 358,97 | 0,02 | 0,03 | 0,72 | 4,69E-01 |
| SEC24B | 1787,71 | -0,02 | 0,02 | -0,72 | 4,70E-01 |
| ZNF546 | 384,89 | -0,03 | 0,05 | -0,72 | 4,70E-01 |
| EIF4ENIF1 | 702,04 | 0,02 | 0,03 | 0,72 | 4,70E-01 |
| TCTN1 | 382,14 | 0,04 | 0,06 | 0,72 | 4,70E-01 |
| NSDHL | 135,35 | -0,03 | 0,04 | -0,72 | 4,71E-01 |
| CEP295NL | 437,21 | 0,08 | 0,12 | 0,72 | 4,71E-01 |
| HDAC9 | 1577,55 | -0,05 | 0,07 | -0,72 | 4,72E-01 |
| ZBTB34 | 3823,97 | -0,04 | 0,05 | -0,72 | 4,72E-01 |
| POC1B | 1230,65 | 0,03 | 0,04 | 0,72 | 4,72E-01 |
| DUSP14 | 75,44 | 0,05 | 0,07 | 0,72 | 4,72E-01 |
| COX10 | 349,45 | -0,02 | 0,03 | -0,72 | 4,73E-01 |
| LMO7 | 493,15 | 0,04 | 0,05 | 0,72 | 4,73E-01 |
| MSRA | 443,31 | 0,05 | 0,07 | 0,72 | 4,73E-01 |
| KIF13A | 3812,11 | -0,05 | 0,06 | -0,72 | 4,73E-01 |
| HEXA | 1569,68 | 0,02 | 0,03 | 0,72 | 4,73E-01 |
| B3GNTL1 | 881,21 | 0,06 | 0,08 | 0,72 | 4,73E-01 |
| ZNF714 | 536,20 | -0,04 | 0,05 | -0,72 | 4,73E-01 |
| TRAFD1 | 2359,65 | 0,04 | 0,06 | 0,72 | 4,74E-01 |
| EMC6 | 186,14 | 0,03 | 0,04 | 0,72 | 4,74E-01 |
| FUT4 | 652,00 | 0,02 | 0,03 | 0,72 | 4,74E-01 |
| CENATAC | 1106,83 | 0,03 | 0,04 | 0,72 | 4,74E-01 |
| GSTM1 | 71,08 | 0,40 | 0,57 | 0,71 | 4,75E-01 |
| XCL2 | 113,72 | -0,09 | 0,12 | -0,71 | 4,75E-01 |
| MZB1 | 135,37 | 0,09 | 0,12 | 0,71 | 4,75E-01 |
| ZNF324 | 584,44 | 0,02 | 0,03 | 0,71 | 4,75E-01 |
| GMNN | 96,24 | -0,04 | 0,06 | -0,71 | 4,76E-01 |
| UBE2K | 1949,03 | -0,03 | 0,04 | -0,71 | 4,76E-01 |
| GSTM3 | 96,44 | -0,12 | 0,17 | -0,71 | 4,76E-01 |
| PLBD1 | 4418,69 | -0,05 | 0,07 | -0,71 | 4,76E-01 |
| H3C6 | 167,58 | -0,06 | 0,08 | -0,71 | 4,76E-01 |
| LGALS8 | 3140,02 | -0,02 | 0,03 | -0,71 | 4,76E-01 |
| VPS39 | 2149,31 | 0,03 | 0,05 | 0,71 | 4,76E-01 |
| PGAM1 | 1054,84 | 0,03 | 0,04 | 0,71 | 4,78E-01 |
| RSAD1 | 567,13 | 0,02 | 0,03 | 0,71 | 4,78E-01 |
| RNF185 | 870,38 | 0,03 | 0,04 | 0,71 | 4,78E-01 |
| TCP11L1 | 572,60 | 0,03 | 0,04 | 0,71 | 4,78E-01 |
| FCRL3 | 1504,12 | -0,06 | 0,08 | -0,71 | 4,78E-01 |
| DNAAF10 | 169,96 | -0,03 | 0,04 | -0,71 | 4,79E-01 |
| NOP53 | 5882,67 | -0,04 | 0,06 | -0,71 | 4,79E-01 |
| DNPH1 | 180,68 | 0,04 | 0,06 | 0,71 | 4,79E-01 |
| NELFCD | 882,01 | -0,02 | 0,03 | -0,71 | 4,79E-01 |

| | | | | | |
|---|---|---|---|---|---|
| ZFP28 | 208,12 | -0,05 | 0,07 | -0,71 | 4,79E-01 |
| DUS4L | 213,13 | 0,03 | 0,04 | 0,71 | 4,79E-01 |
| DAAM1 | 515,05 | -0,03 | 0,04 | -0,71 | 4,80E-01 |
| CIMAP2 | 88,63 | 0,08 | 0,11 | 0,71 | 4,80E-01 |
| RETREG1 | 577,41 | 0,04 | 0,06 | 0,71 | 4,80E-01 |
| CD36 | 4903,88 | -0,05 | 0,07 | -0,71 | 4,80E-01 |
| ENSG00000257390 | 47,94 | 0,03 | 0,04 | 0,71 | 4,80E-01 |
| PPP3CB | 2046,89 | -0,03 | 0,04 | -0,71 | 4,80E-01 |
| CHN2 | 464,10 | 0,05 | 0,07 | 0,71 | 4,80E-01 |
| SFMBT2 | 1536,19 | 0,03 | 0,04 | 0,71 | 4,81E-01 |
| BCAS3 | 1961,59 | 0,03 | 0,04 | 0,71 | 4,81E-01 |
| ELMO1 | 5161,73 | -0,03 | 0,05 | -0,70 | 4,81E-01 |
| FOXN2 | 3843,79 | -0,04 | 0,06 | -0,70 | 4,81E-01 |
| CLN6 | 328,37 | 0,05 | 0,06 | 0,70 | 4,81E-01 |
| SLC35A3 | 1418,37 | -0,02 | 0,03 | -0,70 | 4,82E-01 |
| RSAD2 | 2598,43 | 0,16 | 0,22 | 0,70 | 4,82E-01 |
| DHTKD1 | 1529,12 | 0,03 | 0,04 | 0,70 | 4,82E-01 |
| COPS7B | 601,61 | 0,03 | 0,05 | 0,70 | 4,82E-01 |
| MED20 | 412,92 | -0,02 | 0,02 | -0,70 | 4,82E-01 |
| KLRC1 | 372,95 | -0,09 | 0,13 | -0,70 | 4,82E-01 |
| ADAR | 13327,45 | 0,05 | 0,07 | 0,70 | 4,83E-01 |
| SGCD | 69,35 | 0,13 | 0,19 | 0,70 | 4,83E-01 |
| ZXDB | 262,74 | 0,03 | 0,04 | 0,70 | 4,83E-01 |
| PLA2G12A | 591,64 | 0,03 | 0,05 | 0,70 | 4,83E-01 |
| TTC38 | 963,42 | -0,04 | 0,06 | -0,70 | 4,84E-01 |
| RAB7A | 1126,66 | 0,05 | 0,07 | 0,70 | 4,84E-01 |
| SPEF2 | 495,79 | -0,04 | 0,06 | -0,70 | 4,84E-01 |
| NUFIP1 | 91,98 | 0,04 | 0,06 | 0,70 | 4,84E-01 |
| SERPINB6 | 1574,04 | 0,03 | 0,05 | 0,70 | 4,85E-01 |
| RBSN | 1397,45 | 0,02 | 0,02 | 0,70 | 4,85E-01 |
| RBM10 | 1236,93 | -0,02 | 0,03 | -0,70 | 4,85E-01 |
| TNFRSF1B | 8824,68 | 0,04 | 0,06 | 0,70 | 4,85E-01 |
| PTGES | 31,69 | 0,11 | 0,16 | 0,70 | 4,85E-01 |
| VMP1 | 17171,29 | -0,04 | 0,06 | -0,70 | 4,85E-01 |
| SHMT2 | 680,39 | -0,03 | 0,04 | -0,70 | 4,85E-01 |
| GSK3B | 3085,72 | -0,03 | 0,04 | -0,70 | 4,85E-01 |
| PTPRE | 9584,63 | -0,03 | 0,05 | -0,70 | 4,85E-01 |
| UBE4A | 3901,64 | -0,02 | 0,03 | -0,70 | 4,86E-01 |
| CLEC4C | 219,72 | -0,10 | 0,15 | -0,70 | 4,86E-01 |
| TET2 | 13062,10 | -0,04 | 0,05 | -0,70 | 4,87E-01 |
| ARHGEF40 | 1903,96 | 0,06 | 0,09 | 0,70 | 4,87E-01 |
| ACIN1 | 4822,30 | 0,03 | 0,04 | 0,69 | 4,87E-01 |
| ID3 | 119,24 | -0,06 | 0,08 | -0,69 | 4,87E-01 |
| TMEM176B | 207,64 | -0,16 | 0,23 | -0,69 | 4,87E-01 |
| PIGH | 343,13 | -0,02 | 0,03 | -0,69 | 4,88E-01 |
| GAPDH | 10864,02 | 0,03 | 0,04 | 0,69 | 4,88E-01 |

| | | | | | |
|---|---|---|---|---|---|
| SND1 | 2720,18 | -0,02 | 0,04 | -0,69 | 4,88E-01 |
| DENND4A | 6595,99 | -0,03 | 0,04 | -0,69 | 4,88E-01 |
| HIPK1 | 8148,66 | 0,03 | 0,04 | 0,69 | 4,88E-01 |
| KIAA0319L | 2659,45 | 0,04 | 0,06 | 0,69 | 4,89E-01 |
| ALG8 | 406,04 | 0,02 | 0,03 | 0,69 | 4,88E-01 |
| ZNF154 | 558,43 | 0,06 | 0,09 | 0,69 | 4,89E-01 |
| IGF2R | 35958,47 | -0,06 | 0,08 | -0,69 | 4,89E-01 |
| MTARC1 | 1409,30 | -0,06 | 0,08 | -0,69 | 4,89E-01 |
| GSTP1 | 1540,43 | -0,03 | 0,05 | -0,69 | 4,89E-01 |
| GEMIN8 | 268,62 | -0,02 | 0,04 | -0,69 | 4,90E-01 |
| TMEM62 | 782,57 | -0,03 | 0,04 | -0,69 | 4,90E-01 |
| OR52K2 | 113,33 | 0,09 | 0,13 | 0,69 | 4,90E-01 |
| SMCR8 | 2251,13 | 0,03 | 0,05 | 0,69 | 4,90E-01 |
| ZNF792 | 753,88 | 0,02 | 0,03 | 0,69 | 4,90E-01 |
| C1orf174 | 581,58 | 0,02 | 0,02 | 0,69 | 4,90E-01 |
| RNF144B | 4400,43 | -0,04 | 0,06 | -0,69 | 4,90E-01 |
| MRPS12 | 241,93 | -0,03 | 0,04 | -0,69 | 4,91E-01 |
| NUDT16L1 | 472,12 | 0,02 | 0,03 | 0,69 | 4,91E-01 |
| NREP | 232,51 | 0,03 | 0,05 | 0,69 | 4,91E-01 |
| CLEC4F | 75,48 | 0,15 | 0,22 | 0,69 | 4,92E-01 |
| MED29 | 688,37 | 0,02 | 0,03 | 0,69 | 4,92E-01 |
| SRPK1 | 2931,38 | -0,04 | 0,06 | -0,69 | 4,92E-01 |
| JHY | 91,90 | 0,05 | 0,07 | 0,69 | 4,92E-01 |
| PRSS57 | 33,64 | 0,09 | 0,14 | 0,69 | 4,92E-01 |
| TET1 | 172,36 | 0,04 | 0,05 | 0,69 | 4,92E-01 |
| KMT2A | 4084,53 | 0,04 | 0,05 | 0,69 | 4,92E-01 |
| S1PR4 | 2543,97 | 0,03 | 0,05 | 0,69 | 4,93E-01 |
| H2BC21 | 3007,53 | 0,04 | 0,05 | 0,69 | 4,93E-01 |
| ENSG00000260861 | 89,59 | 0,05 | 0,07 | 0,69 | 4,93E-01 |
| H2BC17 | 205,96 | 0,06 | 0,09 | 0,69 | 4,93E-01 |
| CREB3L2 | 885,42 | 0,04 | 0,06 | 0,69 | 4,93E-01 |
| ACTR3C | 499,02 | 0,03 | 0,04 | 0,68 | 4,93E-01 |
| STYXL1 | 437,18 | -0,02 | 0,04 | -0,68 | 4,94E-01 |
| ELOVL1 | 886,89 | 0,02 | 0,03 | 0,68 | 4,94E-01 |
| PCDHGA8 | 37,86 | 0,09 | 0,12 | 0,68 | 4,94E-01 |
| LPP | 6272,90 | -0,03 | 0,04 | -0,68 | 4,94E-01 |
| CCR3 | 1072,33 | 0,07 | 0,10 | 0,68 | 4,95E-01 |
| TNIK | 2264,94 | -0,03 | 0,04 | -0,68 | 4,95E-01 |
| RASGRF2 | 1233,99 | 0,04 | 0,06 | 0,68 | 4,96E-01 |
| APBB1IP | 4249,82 | -0,04 | 0,06 | -0,68 | 4,97E-01 |
| GOLGA8N | 269,80 | -0,05 | 0,07 | -0,68 | 4,97E-01 |
| EEPD1 | 804,79 | 0,05 | 0,08 | 0,68 | 4,97E-01 |
| SRGAP2C | 573,57 | 0,04 | 0,06 | 0,68 | 4,97E-01 |
| HP | 102,58 | 0,10 | 0,15 | 0,68 | 4,97E-01 |
| IFI35 | 894,39 | 0,06 | 0,10 | 0,68 | 4,97E-01 |
| SNX9 | 614,16 | 0,03 | 0,04 | 0,68 | 4,97E-01 |

| | | | | | |
|---|---|---|---|---|---|
| RRAGA | 540,56 | -0,02 | 0,03 | -0,68 | 4,97E-01 |
| DLST | 1647,17 | 0,02 | 0,03 | 0,68 | 4,97E-01 |
| VPS52 | 915,67 | 0,02 | 0,03 | 0,68 | 4,98E-01 |
| MORC4 | 209,92 | -0,04 | 0,05 | -0,68 | 4,98E-01 |
| H2AC20 | 5948,21 | -0,05 | 0,07 | -0,68 | 4,98E-01 |
| FBXL14 | 413,86 | 0,02 | 0,03 | 0,68 | 4,98E-01 |
| PFKFB3 | 2117,66 | 0,06 | 0,09 | 0,68 | 4,98E-01 |
| LNPK | 1487,44 | -0,03 | 0,05 | -0,68 | 4,98E-01 |
| RIDA | 83,24 | -0,05 | 0,07 | -0,68 | 4,99E-01 |
| CDA | 507,82 | 0,07 | 0,10 | 0,68 | 4,99E-01 |
| CA8 | 107,30 | 0,09 | 0,14 | 0,68 | 4,99E-01 |
| CLEC4D | 732,39 | -0,07 | 0,10 | -0,68 | 4,99E-01 |
| RBP7 | 159,63 | -0,05 | 0,08 | -0,68 | 4,99E-01 |
| ANXA4 | 821,60 | 0,02 | 0,03 | 0,68 | 4,99E-01 |
| MYCL | 430,78 | 0,04 | 0,06 | 0,67 | 5,00E-01 |
| PRMT6 | 474,13 | -0,04 | 0,05 | -0,67 | 5,00E-01 |
| CLEC6A | 97,25 | 0,07 | 0,10 | 0,67 | 5,01E-01 |
| IKZF1 | 11841,15 | 0,03 | 0,04 | 0,67 | 5,01E-01 |
| PCDHGB3 | 53,43 | 0,06 | 0,09 | 0,67 | 5,01E-01 |
| FUS | 5479,26 | 0,04 | 0,06 | 0,67 | 5,01E-01 |
| LPAR5 | 201,78 | 0,04 | 0,06 | 0,67 | 5,01E-01 |
| CHMP4A | 163,21 | 0,03 | 0,04 | 0,67 | 5,01E-01 |
| HSH2D | 1693,76 | 0,06 | 0,09 | 0,67 | 5,02E-01 |
| AFF1 | 5645,57 | 0,04 | 0,05 | 0,67 | 5,02E-01 |
| TNFRSF12A | 233,99 | 0,04 | 0,06 | 0,67 | 5,02E-01 |
| SEC24C | 2121,35 | 0,03 | 0,04 | 0,67 | 5,02E-01 |
| SFMBT1 | 643,47 | 0,03 | 0,05 | 0,67 | 5,03E-01 |
| RPL12 | 3551,23 | -0,04 | 0,06 | -0,67 | 5,03E-01 |
| ZCCHC3 | 1074,14 | -0,02 | 0,03 | -0,67 | 5,03E-01 |
| DEGS1 | 1584,95 | -0,03 | 0,04 | -0,67 | 5,03E-01 |
| PABPC1 | 8770,56 | -0,03 | 0,04 | -0,67 | 5,04E-01 |
| USP51 | 47,06 | 0,05 | 0,08 | 0,67 | 5,04E-01 |
| PHF21A | 4213,78 | -0,04 | 0,05 | -0,67 | 5,04E-01 |
| HERC6 | 1020,25 | -0,05 | 0,08 | -0,67 | 5,04E-01 |
| DPF2 | 983,12 | 0,03 | 0,05 | 0,67 | 5,04E-01 |
| ITGA5 | 4288,26 | 0,04 | 0,06 | 0,67 | 5,04E-01 |
| CCNB1 | 96,18 | 0,06 | 0,08 | 0,67 | 5,04E-01 |
| NPRL3 | 2189,20 | 0,12 | 0,18 | 0,67 | 5,05E-01 |
| ELOA | 621,93 | 0,02 | 0,03 | 0,67 | 5,05E-01 |
| CX3CR1 | 2517,57 | -0,04 | 0,07 | -0,67 | 5,05E-01 |
| LAGE3 | 264,35 | -0,03 | 0,05 | -0,67 | 5,05E-01 |
| SMIM12 | 1308,16 | -0,01 | 0,02 | -0,67 | 5,06E-01 |
| CASK | 1421,84 | -0,03 | 0,04 | -0,67 | 5,06E-01 |
| FDXACB1 | 51,96 | -0,03 | 0,05 | -0,67 | 5,06E-01 |
| SOCS6 | 226,57 | -0,04 | 0,06 | -0,67 | 5,06E-01 |
| GTF2H2 | 345,22 | -0,09 | 0,13 | -0,67 | 5,06E-01 |

| | | | | | |
|---|---|---|---|---|---|
| TUBA1B | 3365,43 | 0,02 | 0,04 | 0,67 | 5,06E-01 |
| WDFY4 | 4224,98 | 0,05 | 0,08 | 0,66 | 5,06E-01 |
| MBD1 | 1568,71 | 0,02 | 0,03 | 0,66 | 5,06E-01 |
| RPL15 | 8229,19 | -0,04 | 0,06 | -0,66 | 5,06E-01 |
| PLIN3 | 1158,62 | 0,04 | 0,06 | 0,66 | 5,07E-01 |
| ITPRIPL1 | 226,44 | -0,05 | 0,07 | -0,66 | 5,07E-01 |
| MCM5 | 1120,37 | 0,03 | 0,04 | 0,66 | 5,07E-01 |
| HLA-DOB | 212,46 | -0,06 | 0,10 | -0,66 | 5,07E-01 |
| ASPH | 3002,92 | 0,05 | 0,07 | 0,66 | 5,07E-01 |
| NCF2 | 24513,09 | -0,04 | 0,06 | -0,66 | 5,08E-01 |
| FAM50A | 436,12 | -0,02 | 0,03 | -0,66 | 5,08E-01 |
| E2F3 | 1225,10 | 0,03 | 0,05 | 0,66 | 5,08E-01 |
| CYTH3 | 320,97 | 0,04 | 0,05 | 0,66 | 5,08E-01 |
| TPM4 | 5486,31 | -0,03 | 0,04 | -0,66 | 5,08E-01 |
| NUP93 | 992,59 | 0,02 | 0,03 | 0,66 | 5,08E-01 |
| ASPRV1 | 319,60 | 0,07 | 0,11 | 0,66 | 5,09E-01 |
| COA7 | 186,98 | 0,03 | 0,05 | 0,66 | 5,09E-01 |
| ENSG00000275464 | 245,73 | 0,07 | 0,11 | 0,66 | 5,09E-01 |
| CENPE | 139,30 | -0,07 | 0,11 | -0,66 | 5,09E-01 |
| SLC30A7 | 2696,62 | -0,02 | 0,04 | -0,66 | 5,09E-01 |
| ZCWPW1 | 195,77 | 0,02 | 0,03 | 0,66 | 5,10E-01 |
| ERP29 | 751,86 | -0,03 | 0,04 | -0,66 | 5,10E-01 |
| FARP2 | 754,56 | 0,03 | 0,05 | 0,66 | 5,10E-01 |
| PTEN | 24919,34 | -0,04 | 0,06 | -0,66 | 5,11E-01 |
| DGUOK | 489,27 | 0,02 | 0,03 | 0,66 | 5,11E-01 |
| SYCP2 | 399,77 | -0,05 | 0,07 | -0,66 | 5,11E-01 |
| CENPH | 83,66 | -0,05 | 0,07 | -0,66 | 5,12E-01 |
| TAF1D | 4010,35 | -0,05 | 0,08 | -0,66 | 5,12E-01 |
| COPS9 | 334,34 | -0,04 | 0,06 | -0,66 | 5,12E-01 |
| SLC25A44 | 1694,81 | 0,04 | 0,06 | 0,66 | 5,12E-01 |
| KANSL1 | 7173,39 | 0,04 | 0,06 | 0,66 | 5,12E-01 |
| HMGXB4 | 466,54 | -0,02 | 0,02 | -0,65 | 5,13E-01 |
| UBE3D | 409,41 | -0,03 | 0,05 | -0,65 | 5,13E-01 |
| C1QBP | 907,74 | -0,03 | 0,05 | -0,65 | 5,13E-01 |
| PKIA | 287,22 | -0,05 | 0,08 | -0,65 | 5,13E-01 |
| KNL1 | 242,43 | 0,06 | 0,09 | 0,65 | 5,13E-01 |
| LIMS1 | 4349,90 | 0,04 | 0,06 | 0,65 | 5,14E-01 |
| IKZF3 | 4914,00 | -0,04 | 0,06 | -0,65 | 5,14E-01 |
| ZNF630 | 150,86 | -0,04 | 0,06 | -0,65 | 5,15E-01 |
| SLC12A6 | 10965,56 | 0,03 | 0,05 | 0,65 | 5,15E-01 |
| GRAP | 745,91 | -0,03 | 0,05 | -0,65 | 5,15E-01 |
| HSD17B13 | 59,26 | 0,05 | 0,07 | 0,65 | 5,15E-01 |
| CYRIA | 3858,89 | -0,03 | 0,05 | -0,65 | 5,16E-01 |
| WASHC2A | 1208,28 | -0,02 | 0,04 | -0,65 | 5,16E-01 |
| ABLIM1 | 4100,97 | -0,03 | 0,05 | -0,65 | 5,18E-01 |
| RHOH | 2992,06 | -0,03 | 0,05 | -0,65 | 5,18E-01 |

| | | | | | |
|---|---|---|---|---|---|
| PTPRM | 430,98 | -0,07 | 0,10 | -0,65 | 5,18E-01 |
| DENND2D | 3489,96 | -0,03 | 0,04 | -0,65 | 5,18E-01 |
| CDC42EP3 | 3827,75 | -0,03 | 0,05 | -0,65 | 5,18E-01 |
| GPR160 | 765,84 | -0,04 | 0,07 | -0,65 | 5,18E-01 |
| PCCB | 513,59 | 0,01 | 0,02 | 0,65 | 5,18E-01 |
| MAP2K1 | 1614,42 | -0,02 | 0,03 | -0,65 | 5,19E-01 |
| DHFR | 442,30 | 0,02 | 0,03 | 0,65 | 5,19E-01 |
| HS3ST3B1 | 573,96 | -0,03 | 0,04 | -0,64 | 5,19E-01 |
| UFSP1 | 112,24 | -0,03 | 0,05 | -0,64 | 5,20E-01 |
| ZNF749 | 161,76 | -0,04 | 0,06 | -0,64 | 5,20E-01 |
| ENSG00000280071 | 133,35 | 0,08 | 0,12 | 0,64 | 5,20E-01 |
| MAP4K3 | 686,62 | -0,03 | 0,04 | -0,64 | 5,21E-01 |
| ZNF585A | 337,18 | -0,02 | 0,04 | -0,64 | 5,21E-01 |
| SLC3A2 | 1231,47 | 0,02 | 0,04 | 0,64 | 5,22E-01 |
| DHRS12 | 501,67 | 0,05 | 0,07 | 0,64 | 5,22E-01 |
| POLI | 1165,96 | -0,03 | 0,05 | -0,64 | 5,23E-01 |
| GOLGA8A | 1231,69 | 0,04 | 0,07 | 0,64 | 5,23E-01 |
| NFATC3 | 3515,49 | -0,02 | 0,03 | -0,64 | 5,23E-01 |
| IGLL5 | 84,04 | -0,09 | 0,14 | -0,64 | 5,23E-01 |
| DMXL2 | 11153,45 | -0,04 | 0,06 | -0,64 | 5,23E-01 |
| GVQW3 | 220,82 | 0,03 | 0,05 | 0,64 | 5,23E-01 |
| CCL28 | 175,21 | 0,03 | 0,04 | 0,64 | 5,24E-01 |
| CCL5 | 4538,47 | -0,05 | 0,08 | -0,64 | 5,24E-01 |
| DGKE | 1211,72 | -0,03 | 0,05 | -0,64 | 5,24E-01 |
| ATP8B4 | 1713,13 | -0,04 | 0,06 | -0,64 | 5,24E-01 |
| WDR1 | 7273,56 | 0,03 | 0,04 | 0,64 | 5,25E-01 |
| TBC1D14 | 5532,24 | -0,03 | 0,05 | -0,64 | 5,25E-01 |
| ERLIN1 | 821,20 | 0,03 | 0,04 | 0,64 | 5,24E-01 |
| DOCK7 | 576,85 | 0,04 | 0,06 | 0,64 | 5,25E-01 |
| SPTBN1 | 4763,02 | 0,03 | 0,05 | 0,63 | 5,26E-01 |
| ERRFI1 | 38,49 | 0,06 | 0,10 | 0,63 | 5,26E-01 |
| ACP6 | 549,09 | 0,03 | 0,05 | 0,63 | 5,26E-01 |
| PLXNC1 | 18218,35 | -0,04 | 0,06 | -0,63 | 5,27E-01 |
| ZNF577 | 462,49 | -0,03 | 0,04 | -0,63 | 5,27E-01 |
| MAD2L1 | 120,92 | -0,05 | 0,08 | -0,63 | 5,27E-01 |
| SMC1A | 2172,92 | -0,02 | 0,04 | -0,63 | 5,27E-01 |
| SLAIN1 | 599,61 | -0,03 | 0,04 | -0,63 | 5,28E-01 |
| RPS9 | 12909,85 | -0,04 | 0,07 | -0,63 | 5,28E-01 |
| ITGB7 | 1834,74 | 0,03 | 0,05 | 0,63 | 5,28E-01 |
| SMCO4 | 273,20 | 0,04 | 0,07 | 0,63 | 5,29E-01 |
| APLF | 292,21 | -0,03 | 0,06 | -0,63 | 5,29E-01 |
| TRAPPC5 | 592,68 | -0,03 | 0,05 | -0,63 | 5,29E-01 |
| TNFRSF10A | 955,44 | 0,02 | 0,04 | 0,63 | 5,29E-01 |
| SLC19A2 | 222,96 | 0,03 | 0,04 | 0,63 | 5,29E-01 |
| C15orf40 | 1194,71 | 0,02 | 0,03 | 0,63 | 5,29E-01 |
| AVIL | 768,60 | 0,05 | 0,08 | 0,63 | 5,30E-01 |

| | | | | | |
|---|---|---|---|---|---|
| CGRRF1 | 374,74 | -0,04 | 0,06 | -0,63 | 5,30E-01 |
| SMYD3 | 636,64 | 0,04 | 0,06 | 0,63 | 5,30E-01 |
| EIF1AY | 879,96 | 0,11 | 0,18 | 0,63 | 5,30E-01 |
| PLAC8 | 3897,38 | -0,03 | 0,04 | -0,63 | 5,30E-01 |
| ZDHHC2 | 2963,61 | 0,04 | 0,06 | 0,63 | 5,30E-01 |
| MRPL23 | 148,77 | 0,06 | 0,09 | 0,63 | 5,30E-01 |
| GASK1B | 1319,22 | 0,05 | 0,07 | 0,63 | 5,30E-01 |
| NCOA6 | 1306,19 | 0,03 | 0,05 | 0,63 | 5,30E-01 |
| RBM43 | 454,08 | -0,03 | 0,05 | -0,63 | 5,31E-01 |
| EIF4E3 | 5512,63 | 0,04 | 0,06 | 0,63 | 5,31E-01 |
| SERF1B | 45,41 | -0,05 | 0,08 | -0,63 | 5,31E-01 |
| C2orf42 | 361,57 | 0,01 | 0,02 | 0,63 | 5,31E-01 |
| NCR1 | 335,81 | -0,04 | 0,07 | -0,63 | 5,31E-01 |
| IQCG | 318,74 | -0,02 | 0,04 | -0,63 | 5,32E-01 |
| INPP5D | 4850,87 | 0,04 | 0,07 | 0,63 | 5,31E-01 |
| SLC8A1 | 4245,65 | -0,04 | 0,07 | -0,63 | 5,31E-01 |
| POU5F2 | 1153,90 | 0,04 | 0,07 | 0,63 | 5,32E-01 |
| CALR | 1589,03 | 0,03 | 0,05 | 0,63 | 5,32E-01 |
| DOCK5 | 16458,29 | -0,05 | 0,07 | -0,62 | 5,32E-01 |
| GAB2 | 2715,90 | 0,05 | 0,08 | 0,62 | 5,32E-01 |
| XPNPEP1 | 1675,76 | -0,02 | 0,03 | -0,62 | 5,32E-01 |
| BBS2 | 1014,85 | -0,03 | 0,04 | -0,62 | 5,32E-01 |
| EXOC6B | 689,74 | -0,02 | 0,04 | -0,62 | 5,33E-01 |
| RAD18 | 481,97 | -0,02 | 0,03 | -0,62 | 5,33E-01 |
| SVIL | 4372,63 | 0,04 | 0,07 | 0,62 | 5,33E-01 |
| AREL1 | 4603,23 | -0,03 | 0,05 | -0,62 | 5,33E-01 |
| IFFO2 | 995,62 | -0,03 | 0,04 | -0,62 | 5,33E-01 |
| MRPL11 | 280,94 | -0,03 | 0,05 | -0,62 | 5,33E-01 |
| ENSG00000285816 | 122,69 | 0,06 | 0,09 | 0,62 | 5,33E-01 |
| GPR52 | 134,63 | 0,05 | 0,09 | 0,62 | 5,34E-01 |
| GLT1D1 | 3301,12 | 0,05 | 0,07 | 0,62 | 5,34E-01 |
| SRP68 | 653,17 | 0,02 | 0,04 | 0,62 | 5,35E-01 |
| CCDC167 | 105,69 | 0,05 | 0,07 | 0,62 | 5,35E-01 |
| EPB41L5 | 330,83 | 0,03 | 0,04 | 0,62 | 5,35E-01 |
| ATF4 | 2574,72 | -0,02 | 0,04 | -0,62 | 5,35E-01 |
| LTA4H | 6971,55 | -0,04 | 0,06 | -0,62 | 5,35E-01 |
| AIDA | 1380,98 | 0,03 | 0,05 | 0,62 | 5,35E-01 |
| PIP4P2 | 1867,26 | -0,04 | 0,07 | -0,62 | 5,35E-01 |
| EXTL3 | 2699,28 | -0,04 | 0,06 | -0,62 | 5,36E-01 |
| ZNF875 | 1038,83 | 0,03 | 0,04 | 0,62 | 5,36E-01 |
| PSMD4 | 3002,44 | -0,03 | 0,05 | -0,62 | 5,36E-01 |
| ANKRD28 | 1919,50 | -0,03 | 0,05 | -0,62 | 5,36E-01 |
| MANSC1 | 591,58 | 0,05 | 0,08 | 0,62 | 5,36E-01 |
| PFDN4 | 133,00 | -0,05 | 0,09 | -0,62 | 5,36E-01 |
| SELL | 21006,04 | -0,04 | 0,06 | -0,62 | 5,36E-01 |
| BRCA2 | 274,81 | -0,05 | 0,07 | -0,62 | 5,36E-01 |

| | | | | | |
|---|---|---|---|---|---|
| SCFD2 | 765,43 | -0,02 | 0,03 | -0,62 | 5,36E-01 |
| GON4L | 2176,39 | -0,03 | 0,04 | -0,62 | 5,37E-01 |
| CD19 | 167,90 | 0,07 | 0,11 | 0,62 | 5,37E-01 |
| DUS3L | 388,89 | 0,02 | 0,04 | 0,62 | 5,37E-01 |
| KLF8 | 183,00 | 0,05 | 0,08 | 0,62 | 5,37E-01 |
| ST8SIA6 | 152,52 | -0,04 | 0,07 | -0,62 | 5,37E-01 |
| HSD17B8 | 69,65 | 0,03 | 0,05 | 0,62 | 5,37E-01 |
| F2R | 628,96 | -0,04 | 0,07 | -0,62 | 5,37E-01 |
| RFFL | 2767,38 | 0,03 | 0,05 | 0,62 | 5,38E-01 |
| HEG1 | 752,62 | 0,03 | 0,06 | 0,62 | 5,38E-01 |
| TNFRSF10C | 5705,71 | -0,04 | 0,07 | -0,61 | 5,39E-01 |
| VPS37B | 1128,58 | 0,04 | 0,06 | 0,61 | 5,39E-01 |
| TWF2 | 1717,27 | 0,02 | 0,03 | 0,61 | 5,39E-01 |
| GGA2 | 2415,44 | -0,03 | 0,04 | -0,61 | 5,39E-01 |
| ZNF789 | 482,88 | -0,02 | 0,03 | -0,61 | 5,39E-01 |
| GLIPR2 | 5125,34 | -0,03 | 0,05 | -0,61 | 5,39E-01 |
| CDH23 | 2098,76 | 0,04 | 0,06 | 0,61 | 5,39E-01 |
| PBX3 | 717,40 | 0,02 | 0,04 | 0,61 | 5,40E-01 |
| CBX5 | 1752,87 | 0,03 | 0,06 | 0,61 | 5,40E-01 |
| ZNF562 | 818,59 | -0,02 | 0,03 | -0,61 | 5,40E-01 |
| BOLA1 | 158,58 | -0,02 | 0,04 | -0,61 | 5,40E-01 |
| SPIN4 | 169,03 | -0,03 | 0,05 | -0,61 | 5,40E-01 |
| ITGB8 | 168,42 | 0,11 | 0,17 | 0,61 | 5,40E-01 |
| LGALS2 | 456,44 | -0,09 | 0,14 | -0,61 | 5,40E-01 |
| PRF1 | 1962,01 | -0,06 | 0,10 | -0,61 | 5,41E-01 |
| VOPP1 | 2258,93 | 0,03 | 0,04 | 0,61 | 5,41E-01 |
| TAS2R10 | 30,38 | -0,05 | 0,09 | -0,61 | 5,41E-01 |
| SLC25A32 | 321,82 | -0,03 | 0,05 | -0,61 | 5,41E-01 |
| NF1 | 3232,13 | -0,02 | 0,04 | -0,61 | 5,41E-01 |
| DIPK2A | 1164,14 | -0,03 | 0,04 | -0,61 | 5,41E-01 |
| FCER2 | 105,68 | -0,07 | 0,12 | -0,61 | 5,42E-01 |
| IFIT1 | 2157,19 | -0,12 | 0,20 | -0,61 | 5,42E-01 |
| TBC1D4 | 1796,81 | -0,04 | 0,06 | -0,61 | 5,42E-01 |
| ZNF726 | 95,46 | 0,04 | 0,06 | 0,61 | 5,42E-01 |
| HLCS | 506,11 | 0,03 | 0,05 | 0,61 | 5,43E-01 |
| RSRP1 | 12810,41 | -0,03 | 0,05 | -0,61 | 5,43E-01 |
| ZNF300 | 128,48 | 0,06 | 0,10 | 0,61 | 5,43E-01 |
| FOLR2 | 49,92 | 0,05 | 0,08 | 0,61 | 5,43E-01 |
| TVP23C | 519,07 | -0,03 | 0,04 | -0,61 | 5,44E-01 |
| SELPLG | 4674,42 | 0,03 | 0,06 | 0,61 | 5,44E-01 |
| NKG7 | 612,15 | 0,06 | 0,11 | 0,61 | 5,45E-01 |
| RP9 | 148,56 | -0,02 | 0,04 | -0,61 | 5,45E-01 |
| ZNF768 | 184,41 | 0,02 | 0,04 | 0,61 | 5,45E-01 |
| GCSAML | 93,73 | 0,08 | 0,13 | 0,61 | 5,45E-01 |
| SLA2 | 746,27 | 0,03 | 0,05 | 0,61 | 5,45E-01 |
| AARS2 | 591,94 | 0,02 | 0,03 | 0,61 | 5,45E-01 |

| | | | | | |
|---|---|---|---|---|---|
| SMN2 | 106,90 | -0,06 | 0,10 | -0,61 | 5,45E-01 |
| DUSP23 | 230,04 | 0,02 | 0,04 | 0,60 | 5,47E-01 |
| PEX7 | 199,72 | -0,02 | 0,04 | -0,60 | 5,48E-01 |
| PSMB8 | 3482,92 | -0,03 | 0,05 | -0,60 | 5,48E-01 |
| GPRASP3 | 98,65 | 0,04 | 0,06 | 0,60 | 5,49E-01 |
| PHLPP1 | 580,13 | 0,03 | 0,05 | 0,60 | 5,49E-01 |
| SRA1 | 601,08 | 0,02 | 0,04 | 0,60 | 5,49E-01 |
| SLC25A4 | 120,03 | 0,04 | 0,07 | 0,60 | 5,49E-01 |
| RAB33B | 1128,78 | -0,04 | 0,06 | -0,60 | 5,49E-01 |
| CRYGS | 181,83 | -0,02 | 0,04 | -0,60 | 5,50E-01 |
| TOX4 | 2252,52 | -0,02 | 0,03 | -0,60 | 5,50E-01 |
| EIPR1 | 689,06 | 0,02 | 0,03 | 0,60 | 5,50E-01 |
| SIK2 | 684,56 | 0,02 | 0,04 | 0,60 | 5,50E-01 |
| DYNLL1 | 681,61 | 0,03 | 0,05 | 0,60 | 5,51E-01 |
| CYFIP2 | 7711,11 | 0,02 | 0,04 | 0,60 | 5,51E-01 |
| EXT2 | 1102,27 | -0,02 | 0,03 | -0,60 | 5,51E-01 |
| IMP4 | 750,52 | -0,02 | 0,03 | -0,60 | 5,52E-01 |
| ST3GAL6 | 1456,87 | -0,04 | 0,06 | -0,60 | 5,52E-01 |
| LZTFL1 | 419,69 | -0,03 | 0,04 | -0,59 | 5,52E-01 |
| TMEM186 | 185,37 | -0,02 | 0,03 | -0,59 | 5,52E-01 |
| DRC12 | 111,36 | -0,05 | 0,08 | -0,59 | 5,52E-01 |
| TMEM109 | 327,69 | 0,03 | 0,05 | 0,59 | 5,53E-01 |
| TNFRSF9 | 408,27 | 0,05 | 0,08 | 0,59 | 5,54E-01 |
| SKP1 | 3156,57 | -0,02 | 0,04 | -0,59 | 5,54E-01 |
| ZNF689 | 333,56 | 0,02 | 0,03 | 0,59 | 5,54E-01 |
| CPM | 183,28 | 0,04 | 0,06 | 0,59 | 5,54E-01 |
| LINGO2 | 52,66 | 0,08 | 0,14 | 0,59 | 5,54E-01 |
| MTLN | 121,67 | 0,04 | 0,06 | 0,59 | 5,55E-01 |
| SLC40A1 | 3210,51 | 0,04 | 0,06 | 0,59 | 5,55E-01 |
| DISP1 | 134,54 | 0,03 | 0,05 | 0,59 | 5,55E-01 |
| MGAM2 | 607,89 | 0,06 | 0,11 | 0,59 | 5,55E-01 |
| YWHAE | 832,21 | -0,02 | 0,04 | -0,59 | 5,55E-01 |
| XKRX | 68,13 | 0,07 | 0,11 | 0,59 | 5,55E-01 |
| H2AC6 | 11570,55 | -0,03 | 0,06 | -0,59 | 5,55E-01 |
| NCEH1 | 269,44 | 0,02 | 0,04 | 0,59 | 5,56E-01 |
| ARL6IP6 | 1034,59 | -0,03 | 0,04 | -0,59 | 5,56E-01 |
| FSBP | 29,19 | -0,06 | 0,11 | -0,59 | 5,56E-01 |
| CCDC144A | 699,30 | -0,16 | 0,27 | -0,59 | 5,56E-01 |
| PALLD | 199,33 | -0,05 | 0,08 | -0,59 | 5,57E-01 |
| ZC3HC1 | 271,31 | -0,02 | 0,03 | -0,59 | 5,57E-01 |
| FZD6 | 121,60 | -0,05 | 0,09 | -0,59 | 5,57E-01 |
| ATRAID | 895,27 | -0,02 | 0,04 | -0,59 | 5,57E-01 |
| H2AC8 | 1506,01 | -0,04 | 0,08 | -0,59 | 5,57E-01 |
| COG8 | 938,95 | 0,01 | 0,02 | 0,59 | 5,57E-01 |
| DTNBP1 | 462,45 | 0,02 | 0,04 | 0,59 | 5,57E-01 |
| ZFAT | 986,51 | 0,02 | 0,04 | 0,59 | 5,57E-01 |

| | | | | | |
|---|---|---|---|---|---|
| NIPSNAP3B | 86,45 | 0,05 | 0,08 | 0,59 | 5,57E-01 |
| CYP1B1 | 939,96 | -0,05 | 0,08 | -0,59 | 5,58E-01 |
| COQ10A | 180,40 | 0,02 | 0,04 | 0,59 | 5,58E-01 |
| SERPINI2 | 58,03 | 0,06 | 0,10 | 0,59 | 5,58E-01 |
| CKS1B | 95,25 | -0,04 | 0,08 | -0,59 | 5,58E-01 |
| CCDC71 | 440,99 | 0,02 | 0,03 | 0,59 | 5,58E-01 |
| IFT20 | 319,00 | -0,02 | 0,04 | -0,59 | 5,59E-01 |
| DPY19L1 | 805,21 | -0,03 | 0,05 | -0,59 | 5,59E-01 |
| RNF216 | 2256,07 | 0,02 | 0,04 | 0,58 | 5,59E-01 |
| MPRIP | 4577,08 | 0,03 | 0,05 | 0,58 | 5,59E-01 |
| PREX1 | 16313,50 | 0,04 | 0,07 | 0,58 | 5,59E-01 |
| ATF7-NPFF | 30,88 | 0,04 | 0,07 | 0,58 | 5,60E-01 |
| ZHX2 | 596,46 | 0,03 | 0,06 | 0,58 | 5,60E-01 |
| CERS6 | 1271,05 | -0,02 | 0,04 | -0,58 | 5,60E-01 |
| TRIM66 | 941,41 | 0,04 | 0,07 | 0,58 | 5,60E-01 |
| ENSG00000284526 | 178,40 | 0,06 | 0,10 | 0,58 | 5,60E-01 |
| ADI1 | 991,14 | -0,02 | 0,04 | -0,58 | 5,60E-01 |
| PHLDB2 | 380,74 | -0,04 | 0,07 | -0,58 | 5,60E-01 |
| KHNYN | 5194,56 | -0,02 | 0,03 | -0,58 | 5,61E-01 |
| ASCL2 | 304,69 | -0,04 | 0,07 | -0,58 | 5,61E-01 |
| CDC42 | 806,35 | -0,02 | 0,03 | -0,58 | 5,61E-01 |
| EIF2B5 | 1204,70 | -0,01 | 0,02 | -0,58 | 5,61E-01 |
| XKR3 | 60,81 | 0,12 | 0,20 | 0,58 | 5,61E-01 |
| GPX3 | 29,86 | 0,09 | 0,15 | 0,58 | 5,61E-01 |
| ZNF398 | 1163,56 | -0,02 | 0,03 | -0,58 | 5,61E-01 |
| DAB2 | 467,15 | 0,03 | 0,06 | 0,58 | 5,61E-01 |
| ENSG00000259316 | 47,63 | 0,04 | 0,07 | 0,58 | 5,62E-01 |
| BCAS4 | 417,68 | 0,04 | 0,07 | 0,58 | 5,63E-01 |
| SAP30L | 1948,49 | 0,02 | 0,03 | 0,58 | 5,63E-01 |
| EIF1B | 936,99 | 0,03 | 0,05 | 0,58 | 5,65E-01 |
| REPS1 | 1864,18 | -0,02 | 0,03 | -0,58 | 5,65E-01 |
| DDX41 | 936,36 | -0,02 | 0,03 | -0,58 | 5,65E-01 |
| ABCG1 | 1186,07 | -0,05 | 0,09 | -0,57 | 5,65E-01 |
| NCOA1 | 6279,39 | -0,03 | 0,05 | -0,57 | 5,67E-01 |
| EARS2 | 383,20 | 0,02 | 0,03 | 0,57 | 5,67E-01 |
| MZT2A | 616,06 | 0,03 | 0,05 | 0,57 | 5,67E-01 |
| CDC7 | 286,48 | -0,03 | 0,06 | -0,57 | 5,67E-01 |
| BMPR2 | 987,87 | -0,02 | 0,04 | -0,57 | 5,67E-01 |
| NAA38 | 508,86 | 0,03 | 0,05 | 0,57 | 5,67E-01 |
| SEH1L | 674,39 | 0,02 | 0,03 | 0,57 | 5,68E-01 |
| BEND5 | 88,82 | 0,05 | 0,09 | 0,57 | 5,68E-01 |
| DTWD2 | 330,31 | -0,02 | 0,04 | -0,57 | 5,68E-01 |
| LCMT2 | 382,73 | -0,03 | 0,04 | -0,57 | 5,68E-01 |
| ZC3H11A | 121,34 | -0,02 | 0,04 | -0,57 | 5,68E-01 |
| BLK | 592,73 | -0,05 | 0,10 | -0,57 | 5,68E-01 |
| ZNF784 | 141,63 | 0,02 | 0,04 | 0,57 | 5,69E-01 |

| | | | | | |
|---|---|---|---|---|---|
| GNE | 531,89 | -0,01 | 0,02 | -0,57 | 5,69E-01 |
| TBC1D8 | 883,27 | 0,04 | 0,06 | 0,57 | 5,69E-01 |
| SLC2A13 | 432,56 | -0,02 | 0,04 | -0,57 | 5,69E-01 |
| BAX | 813,78 | 0,02 | 0,03 | 0,57 | 5,69E-01 |
| PKIG | 174,20 | -0,05 | 0,09 | -0,57 | 5,70E-01 |
| POLB | 1044,95 | -0,03 | 0,05 | -0,57 | 5,70E-01 |
| KLHL14 | 164,19 | -0,07 | 0,12 | -0,57 | 5,70E-01 |
| ZNF428 | 408,32 | 0,03 | 0,05 | 0,57 | 5,71E-01 |
| DISC1 | 1527,20 | 0,04 | 0,06 | 0,57 | 5,71E-01 |
| ARHGAP42 | 126,15 | 0,04 | 0,08 | 0,57 | 5,70E-01 |
| DNASE2 | 751,27 | -0,03 | 0,05 | -0,57 | 5,70E-01 |
| TDRKH | 212,05 | 0,03 | 0,05 | 0,57 | 5,71E-01 |
| HMGB3 | 51,16 | 0,06 | 0,10 | 0,57 | 5,71E-01 |
| HLA-C | 51346,61 | 0,04 | 0,06 | 0,57 | 5,71E-01 |
| ZSCAN20 | 161,45 | 0,03 | 0,06 | 0,57 | 5,71E-01 |
| NCOA4 | 39066,47 | 0,04 | 0,07 | 0,57 | 5,71E-01 |
| RAB11FIP4 | 4161,41 | 0,03 | 0,05 | 0,57 | 5,72E-01 |
| CENPX | 267,70 | -0,03 | 0,05 | -0,57 | 5,72E-01 |
| FLT3 | 228,62 | -0,05 | 0,09 | -0,57 | 5,72E-01 |
| CCR6 | 141,93 | 0,04 | 0,07 | 0,56 | 5,72E-01 |
| KIR3DL2 | 34,28 | 0,08 | 0,14 | 0,56 | 5,73E-01 |
| CFAP58 | 154,68 | 0,05 | 0,09 | 0,56 | 5,73E-01 |
| ABTB3 | 594,98 | 0,03 | 0,06 | 0,56 | 5,73E-01 |
| AMOT | 120,98 | 0,04 | 0,06 | 0,56 | 5,73E-01 |
| SLC23A2 | 1832,60 | -0,03 | 0,04 | -0,56 | 5,74E-01 |
| JMJD4 | 556,18 | 0,02 | 0,04 | 0,56 | 5,74E-01 |
| FCHSD2 | 1479,99 | -0,03 | 0,05 | -0,56 | 5,74E-01 |
| IL23A | 135,35 | -0,04 | 0,07 | -0,56 | 5,75E-01 |
| TENT2 | 4468,76 | -0,02 | 0,04 | -0,56 | 5,75E-01 |
| MYZAP | 28,43 | 0,08 | 0,14 | 0,56 | 5,75E-01 |
| SFT2D1 | 1452,45 | -0,02 | 0,04 | -0,56 | 5,75E-01 |
| ERC1 | 1589,08 | -0,03 | 0,05 | -0,56 | 5,75E-01 |
| RNLS | 91,17 | 0,03 | 0,05 | 0,56 | 5,75E-01 |
| LIPA | 1582,53 | 0,03 | 0,06 | 0,56 | 5,76E-01 |
| IL27RA | 1047,44 | 0,02 | 0,04 | 0,56 | 5,76E-01 |
| NDUFAF8 | 218,94 | 0,03 | 0,05 | 0,56 | 5,76E-01 |
| GCNT2 | 491,17 | -0,03 | 0,06 | -0,56 | 5,76E-01 |
| ITPRIP | 3467,28 | 0,04 | 0,07 | 0,56 | 5,77E-01 |
| ZDHHC13 | 992,41 | -0,02 | 0,03 | -0,56 | 5,77E-01 |
| PKP4 | 636,54 | 0,03 | 0,05 | 0,56 | 5,77E-01 |
| DCAF7 | 1733,81 | 0,03 | 0,06 | 0,56 | 5,77E-01 |
| MRPS17 | 98,84 | -0,03 | 0,05 | -0,56 | 5,78E-01 |
| FXN | 321,34 | 0,03 | 0,05 | 0,56 | 5,78E-01 |
| GTSF1 | 56,98 | -0,05 | 0,10 | -0,56 | 5,79E-01 |
| AFF3 | 1064,49 | -0,05 | 0,09 | -0,56 | 5,79E-01 |
| TMEM117 | 153,48 | -0,03 | 0,06 | -0,55 | 5,79E-01 |

| | | | | | |
|---|---|---|---|---|---|
| H1-2 | 5992,79 | -0,03 | 0,06 | -0,55 | 5,80E-01 |
| TRAPPC2L | 358,65 | 0,02 | 0,04 | 0,55 | 5,80E-01 |
| CFLAR | 38817,70 | -0,03 | 0,05 | -0,55 | 5,81E-01 |
| VPS13D | 5943,39 | 0,03 | 0,05 | 0,55 | 5,81E-01 |
| SPSB2 | 396,06 | -0,03 | 0,05 | -0,55 | 5,82E-01 |
| HLA-DQB1 | 2869,64 | 0,08 | 0,15 | 0,55 | 5,82E-01 |
| PCNX2 | 1662,25 | 0,03 | 0,05 | 0,55 | 5,82E-01 |
| RPS4Y1 | 2137,97 | 0,11 | 0,20 | 0,55 | 5,83E-01 |
| IL23R | 48,91 | 0,08 | 0,14 | 0,55 | 5,83E-01 |
| ATP2B4 | 4056,20 | 0,03 | 0,06 | 0,55 | 5,83E-01 |
| KCNJ15 | 8121,04 | -0,05 | 0,09 | -0,55 | 5,84E-01 |
| DCBLD1 | 227,29 | -0,04 | 0,07 | -0,55 | 5,84E-01 |
| ARHGAP24 | 1091,17 | -0,04 | 0,07 | -0,55 | 5,84E-01 |
| KRT72 | 120,44 | 0,10 | 0,18 | 0,55 | 5,84E-01 |
| NRF1 | 1041,66 | -0,02 | 0,03 | -0,55 | 5,84E-01 |
| LY86 | 513,19 | -0,03 | 0,06 | -0,55 | 5,84E-01 |
| R3HDM1 | 780,23 | -0,02 | 0,03 | -0,55 | 5,85E-01 |
| MTHFS | 319,72 | -0,03 | 0,05 | -0,55 | 5,85E-01 |
| CRELD2 | 417,99 | 0,02 | 0,04 | 0,55 | 5,85E-01 |
| YARS1 | 953,83 | 0,02 | 0,04 | 0,55 | 5,85E-01 |
| RPL13 | 19728,79 | -0,03 | 0,06 | -0,55 | 5,85E-01 |
| USP44 | 150,73 | -0,05 | 0,10 | -0,55 | 5,85E-01 |
| CDK8 | 428,82 | 0,02 | 0,04 | 0,55 | 5,86E-01 |
| CTLA4 | 244,57 | -0,04 | 0,07 | -0,55 | 5,86E-01 |
| C4orf46 | 163,94 | -0,02 | 0,04 | -0,54 | 5,86E-01 |
| POLR1H | 431,07 | -0,02 | 0,03 | -0,54 | 5,86E-01 |
| ACAD8 | 731,26 | 0,02 | 0,03 | 0,54 | 5,86E-01 |
| SUMF1 | 1407,24 | -0,02 | 0,03 | -0,54 | 5,86E-01 |
| TEX2 | 591,65 | 0,02 | 0,03 | 0,54 | 5,87E-01 |
| SLC25A11 | 728,22 | 0,01 | 0,03 | 0,54 | 5,87E-01 |
| PATJ | 2549,47 | -0,03 | 0,05 | -0,54 | 5,87E-01 |
| PTGES3 | 1699,85 | -0,03 | 0,06 | -0,54 | 5,87E-01 |
| DNAJC27 | 230,53 | -0,02 | 0,04 | -0,54 | 5,87E-01 |
| APOL6 | 10620,42 | -0,04 | 0,08 | -0,54 | 5,88E-01 |
| ARHGAP26 | 15106,04 | -0,04 | 0,07 | -0,54 | 5,88E-01 |
| TTC39B | 2015,60 | -0,03 | 0,05 | -0,54 | 5,88E-01 |
| NME1 | 102,38 | 0,04 | 0,07 | 0,54 | 5,88E-01 |
| TMSB15B | 45,23 | 0,04 | 0,07 | 0,54 | 5,87E-01 |
| PPFIA1 | 1915,86 | 0,03 | 0,06 | 0,54 | 5,88E-01 |
| GCH1 | 1769,45 | 0,03 | 0,06 | 0,54 | 5,88E-01 |
| KCTD12 | 3510,75 | -0,03 | 0,06 | -0,54 | 5,89E-01 |
| TREML2 | 1686,74 | -0,03 | 0,06 | -0,54 | 5,89E-01 |
| SPIB | 165,16 | -0,05 | 0,09 | -0,54 | 5,89E-01 |
| SPIN3 | 586,70 | 0,03 | 0,05 | 0,54 | 5,89E-01 |
| DUSP19 | 105,80 | 0,03 | 0,06 | 0,54 | 5,89E-01 |
| SPECC1L | 336,58 | 0,03 | 0,05 | 0,54 | 5,89E-01 |

| | | | | | |
|---|---|---|---|---|---|
| NMRK1 | 970,10 | -0,03 | 0,05 | -0,54 | 5,90E-01 |
| DOK2 | 1367,53 | 0,02 | 0,04 | 0,54 | 5,90E-01 |
| PPFIBP2 | 791,26 | 0,03 | 0,06 | 0,54 | 5,90E-01 |
| HLA-F | 6126,69 | 0,03 | 0,05 | 0,54 | 5,90E-01 |
| KIAA0825 | 1435,74 | 0,04 | 0,07 | 0,54 | 5,91E-01 |
| ZNF418 | 228,05 | 0,04 | 0,07 | 0,54 | 5,91E-01 |
| LETMD1 | 1115,44 | 0,01 | 0,03 | 0,54 | 5,91E-01 |
| KLHL22 | 1017,11 | 0,02 | 0,04 | 0,54 | 5,91E-01 |
| GEMIN5 | 575,42 | -0,02 | 0,03 | -0,54 | 5,91E-01 |
| SMUG1 | 767,43 | -0,01 | 0,02 | -0,54 | 5,91E-01 |
| PIK3CB | 2113,69 | -0,02 | 0,04 | -0,54 | 5,91E-01 |
| SLAMF1 | 623,59 | -0,03 | 0,05 | -0,54 | 5,92E-01 |
| HIF1AN | 2883,10 | -0,02 | 0,04 | -0,54 | 5,92E-01 |
| UBIAD1 | 508,80 | 0,02 | 0,03 | 0,54 | 5,93E-01 |
| NSUN7 | 361,79 | 0,04 | 0,08 | 0,53 | 5,93E-01 |
| STAT5A | 2904,64 | 0,02 | 0,04 | 0,53 | 5,93E-01 |
| CHD7 | 3089,04 | -0,02 | 0,04 | -0,53 | 5,93E-01 |
| TMBIM4 | 1585,47 | 0,02 | 0,05 | 0,53 | 5,93E-01 |
| CD44 | 9784,15 | -0,02 | 0,03 | -0,53 | 5,93E-01 |
| BACH2 | 1470,20 | -0,04 | 0,07 | -0,53 | 5,93E-01 |
| BIVM | 185,26 | -0,03 | 0,05 | -0,53 | 5,94E-01 |
| ZNF609 | 2105,87 | 0,02 | 0,04 | 0,53 | 5,94E-01 |
| DENND10 | 1742,51 | 0,02 | 0,04 | 0,53 | 5,94E-01 |
| SLC17A5 | 580,64 | -0,01 | 0,02 | -0,53 | 5,94E-01 |
| PANK1 | 107,81 | 0,04 | 0,08 | 0,53 | 5,95E-01 |
| ELAC2 | 1250,29 | 0,01 | 0,02 | 0,53 | 5,95E-01 |
| LRRC37A2 | 516,75 | 0,04 | 0,07 | 0,53 | 5,95E-01 |
| PABPN1 | 1317,31 | -0,01 | 0,03 | -0,53 | 5,95E-01 |
| FCGRT | 4300,26 | 0,03 | 0,06 | 0,53 | 5,96E-01 |
| MS4A2 | 165,60 | 0,09 | 0,17 | 0,53 | 5,96E-01 |
| GPR15 | 174,82 | 0,07 | 0,12 | 0,53 | 5,96E-01 |
| KLF13 | 6568,68 | 0,02 | 0,05 | 0,53 | 5,97E-01 |
| SUV39H1 | 142,17 | -0,02 | 0,04 | -0,53 | 5,97E-01 |
| RNF25 | 251,97 | -0,01 | 0,02 | -0,53 | 5,97E-01 |
| LMTK2 | 2193,44 | -0,03 | 0,07 | -0,53 | 5,97E-01 |
| GNL3L | 1417,28 | -0,02 | 0,05 | -0,53 | 5,97E-01 |
| UQCC2 | 316,22 | 0,02 | 0,03 | 0,53 | 5,97E-01 |
| TESPA1 | 1567,16 | 0,03 | 0,06 | 0,53 | 5,97E-01 |
| GPBP1L1 | 3376,09 | 0,02 | 0,03 | 0,53 | 5,98E-01 |
| PPM1L | 947,88 | 0,03 | 0,05 | 0,53 | 5,98E-01 |
| ZNF665 | 147,92 | 0,04 | 0,07 | 0,53 | 5,98E-01 |
| NUDT2 | 212,57 | -0,02 | 0,05 | -0,53 | 5,99E-01 |
| PIGG | 2049,18 | 0,02 | 0,04 | 0,53 | 5,98E-01 |
| KIR3DL1 | 46,46 | 0,08 | 0,15 | 0,53 | 5,99E-01 |
| SEPTIN1 | 1429,53 | -0,02 | 0,04 | -0,53 | 5,99E-01 |
| TCF19 | 114,60 | 0,03 | 0,07 | 0,52 | 6,00E-01 |

| | | | | | |
|---|---|---|---|---|---|
| CYB5R1 | 770,69 | 0,02 | 0,04 | 0,52 | 6,00E-01 |
| MYDGF | 598,76 | -0,02 | 0,05 | -0,52 | 6,00E-01 |
| HOOK1 | 285,74 | -0,05 | 0,09 | -0,52 | 6,00E-01 |
| USP3 | 6644,87 | -0,02 | 0,05 | -0,52 | 6,00E-01 |
| PAX5 | 525,57 | 0,06 | 0,11 | 0,52 | 6,01E-01 |
| ITPA | 567,09 | 0,02 | 0,03 | 0,52 | 6,02E-01 |
| DGAT2 | 3727,46 | -0,04 | 0,07 | -0,52 | 6,01E-01 |
| MISFA | 42,86 | 0,04 | 0,07 | 0,52 | 6,02E-01 |
| EPB41L3 | 1082,40 | 0,03 | 0,06 | 0,52 | 6,02E-01 |
| WBP1L | 2060,43 | 0,02 | 0,04 | 0,52 | 6,02E-01 |
| AGAP1 | 380,26 | 0,06 | 0,11 | 0,52 | 6,02E-01 |
| ARL4A | 899,75 | 0,05 | 0,10 | 0,52 | 6,02E-01 |
| CHRNB1 | 212,77 | -0,03 | 0,05 | -0,52 | 6,02E-01 |
| CEP43 | 975,75 | -0,02 | 0,05 | -0,52 | 6,03E-01 |
| RSU1 | 2014,71 | 0,03 | 0,05 | 0,52 | 6,04E-01 |
| NANS | 540,98 | -0,02 | 0,03 | -0,52 | 6,04E-01 |
| NUDCD3 | 2490,69 | -0,01 | 0,03 | -0,52 | 6,04E-01 |
| CBR1 | 402,30 | -0,03 | 0,05 | -0,52 | 6,05E-01 |
| SIK3 | 2911,00 | 0,03 | 0,06 | 0,52 | 6,05E-01 |
| LAMTOR2 | 576,55 | -0,02 | 0,04 | -0,52 | 6,05E-01 |
| CAPG | 999,17 | -0,03 | 0,06 | -0,52 | 6,05E-01 |
| ADGRG3 | 1473,64 | 0,06 | 0,11 | 0,52 | 6,05E-01 |
| PROCR | 34,17 | 0,05 | 0,09 | 0,52 | 6,06E-01 |
| KAT7 | 3038,29 | -0,02 | 0,04 | -0,52 | 6,06E-01 |
| PEX19 | 496,54 | 0,01 | 0,02 | 0,52 | 6,06E-01 |
| THEMIS2 | 9815,93 | 0,03 | 0,06 | 0,52 | 6,07E-01 |
| LYVE1 | 64,04 | -0,05 | 0,09 | -0,51 | 6,07E-01 |
| PTPRK | 208,30 | -0,05 | 0,09 | -0,52 | 6,07E-01 |
| FUT10 | 286,35 | -0,05 | 0,09 | -0,51 | 6,07E-01 |
| GPANK1 | 677,91 | -0,02 | 0,03 | -0,51 | 6,07E-01 |
| NLRP12 | 5231,39 | -0,03 | 0,06 | -0,51 | 6,07E-01 |
| MRPS21 | 585,06 | -0,03 | 0,05 | -0,51 | 6,07E-01 |
| SRSF8 | 1532,83 | -0,02 | 0,04 | -0,51 | 6,07E-01 |
| GCNA | 255,39 | 0,02 | 0,04 | 0,51 | 6,07E-01 |
| PDZD11 | 100,79 | 0,02 | 0,04 | 0,51 | 6,07E-01 |
| HOMER1 | 101,93 | 0,04 | 0,08 | 0,51 | 6,08E-01 |
| CCR4 | 1253,98 | -0,04 | 0,07 | -0,51 | 6,08E-01 |
| SLC20A2 | 762,65 | 0,02 | 0,03 | 0,51 | 6,08E-01 |
| SPCS1 | 371,01 | -0,02 | 0,04 | -0,51 | 6,09E-01 |
| FNIP2 | 922,35 | -0,02 | 0,04 | -0,51 | 6,09E-01 |
| NIPAL3 | 2188,55 | -0,02 | 0,04 | -0,51 | 6,10E-01 |
| MTHFD2 | 865,30 | -0,03 | 0,06 | -0,51 | 6,10E-01 |
| ZFAND2A | 343,02 | -0,02 | 0,04 | -0,51 | 6,10E-01 |
| CHST2 | 1269,64 | -0,03 | 0,06 | -0,51 | 6,10E-01 |
| TPST1 | 510,41 | -0,06 | 0,11 | -0,51 | 6,11E-01 |
| NSUN3 | 1926,03 | -0,03 | 0,06 | -0,51 | 6,11E-01 |

| | | | | | |
|---|---|---|---|---|---|
| ENSG00000285253 | 38,84 | 0,03 | 0,06 | 0,51 | 6,11E-01 |
| FBXO48 | 251,62 | 0,02 | 0,03 | 0,51 | 6,11E-01 |
| RXYLT1 | 287,46 | 0,02 | 0,04 | 0,51 | 6,11E-01 |
| ENSG00000284906 | 41,57 | 0,03 | 0,06 | 0,51 | 6,11E-01 |
| NPHP3 | 538,32 | 0,02 | 0,04 | 0,51 | 6,11E-01 |
| CMC2 | 615,42 | 0,02 | 0,03 | 0,51 | 6,11E-01 |
| CXCR6 | 300,48 | -0,06 | 0,11 | -0,51 | 6,11E-01 |
| TMEM258 | 664,03 | -0,02 | 0,05 | -0,51 | 6,12E-01 |
| H2BC9 | 600,44 | -0,04 | 0,07 | -0,51 | 6,12E-01 |
| PPP3R1 | 3912,78 | 0,03 | 0,06 | 0,51 | 6,12E-01 |
| TMEM245 | 2167,28 | 0,02 | 0,04 | 0,51 | 6,12E-01 |
| SAFB2 | 2636,35 | -0,01 | 0,03 | -0,51 | 6,12E-01 |
| MRI1 | 632,57 | 0,03 | 0,05 | 0,51 | 6,12E-01 |
| ZNF93 | 208,70 | -0,02 | 0,04 | -0,51 | 6,12E-01 |
| RNF19B | 1603,19 | 0,03 | 0,07 | 0,51 | 6,12E-01 |
| RPL10 | 15549,47 | -0,03 | 0,05 | -0,51 | 6,12E-01 |
| H1-6 | 52,51 | -0,05 | 0,09 | -0,51 | 6,12E-01 |
| H2AC4 | 185,92 | 0,06 | 0,12 | 0,51 | 6,13E-01 |
| GPR68 | 266,72 | -0,03 | 0,07 | -0,50 | 6,14E-01 |
| JMJD6 | 994,11 | -0,02 | 0,03 | -0,50 | 6,14E-01 |
| DSEL | 156,29 | 0,05 | 0,10 | 0,50 | 6,14E-01 |
| DCAF5 | 1831,87 | -0,02 | 0,03 | -0,50 | 6,15E-01 |
| CFAP68 | 83,13 | 0,03 | 0,07 | 0,50 | 6,15E-01 |
| RLIG1 | 559,92 | -0,03 | 0,07 | -0,50 | 6,16E-01 |
| PLA2G7 | 251,49 | -0,04 | 0,08 | -0,50 | 6,16E-01 |
| STRADA | 859,45 | 0,02 | 0,04 | 0,50 | 6,16E-01 |
| PMF1 | 282,53 | 0,02 | 0,04 | 0,50 | 6,17E-01 |
| SMIM8 | 775,81 | -0,02 | 0,05 | -0,50 | 6,17E-01 |
| EFTUD2 | 1295,88 | 0,02 | 0,04 | 0,50 | 6,17E-01 |
| CCR1 | 1652,92 | 0,04 | 0,09 | 0,50 | 6,17E-01 |
| ZNF7 | 571,48 | 0,01 | 0,03 | 0,50 | 6,17E-01 |
| VSIG1 | 289,13 | 0,03 | 0,06 | 0,50 | 6,17E-01 |
| PDCD6 | 2093,78 | 0,01 | 0,02 | 0,50 | 6,18E-01 |
| ZNF133 | 574,60 | -0,01 | 0,03 | -0,50 | 6,18E-01 |
| BTRC | 508,83 | 0,02 | 0,04 | 0,50 | 6,18E-01 |
| ANG | 27,71 | -0,04 | 0,09 | -0,50 | 6,18E-01 |
| NAGK | 4905,17 | -0,03 | 0,06 | -0,50 | 6,18E-01 |
| NPEPPS | 3867,21 | -0,02 | 0,04 | -0,50 | 6,19E-01 |
| COPS6 | 408,61 | 0,01 | 0,03 | 0,50 | 6,19E-01 |
| SLC7A7 | 2477,13 | 0,03 | 0,05 | 0,50 | 6,19E-01 |
| ZSCAN2 | 278,94 | 0,01 | 0,03 | 0,50 | 6,19E-01 |
| SPRYD7 | 144,40 | 0,02 | 0,04 | 0,50 | 6,19E-01 |
| CAPRIN2 | 1251,65 | -0,02 | 0,03 | -0,50 | 6,20E-01 |
| FCGR1BP | 774,24 | 0,06 | 0,12 | 0,50 | 6,20E-01 |
| NAT10 | 1067,98 | 0,02 | 0,04 | 0,50 | 6,20E-01 |
| PXK | 1449,79 | -0,02 | 0,05 | -0,50 | 6,20E-01 |

| | | | | | |
|---|---|---|---|---|---|
| GPR132 | 1239,86 | 0,02 | 0,04 | 0,49 | 6,21E-01 |
| RPS2 | 16882,17 | 0,02 | 0,05 | 0,49 | 6,21E-01 |
| SPINT1 | 232,70 | 0,04 | 0,08 | 0,49 | 6,21E-01 |
| MTCP1 | 53,19 | 0,02 | 0,05 | 0,49 | 6,21E-01 |
| CALCOCO1 | 3197,85 | 0,02 | 0,04 | 0,49 | 6,21E-01 |
| SLC31A2 | 2994,41 | -0,03 | 0,07 | -0,49 | 6,21E-01 |
| ACP1 | 1782,48 | 0,03 | 0,05 | 0,49 | 6,22E-01 |
| HMGN2 | 3932,27 | -0,02 | 0,03 | -0,49 | 6,22E-01 |
| PDSS1 | 152,20 | 0,02 | 0,04 | 0,49 | 6,22E-01 |
| PCDHGB7 | 60,55 | 0,05 | 0,11 | 0,49 | 6,22E-01 |
| PRRG4 | 1401,75 | -0,04 | 0,09 | -0,49 | 6,22E-01 |
| SUPT6H | 2405,32 | 0,03 | 0,06 | 0,49 | 6,22E-01 |
| THBS1 | 1163,97 | -0,06 | 0,12 | -0,49 | 6,23E-01 |
| EXOG | 563,06 | -0,02 | 0,05 | -0,49 | 6,23E-01 |
| BICD1 | 434,48 | -0,03 | 0,05 | -0,49 | 6,23E-01 |
| CYB561D2 | 409,10 | -0,01 | 0,03 | -0,49 | 6,23E-01 |
| CAT | 6754,75 | 0,03 | 0,07 | 0,49 | 6,23E-01 |
| PITPNC1 | 1761,62 | -0,01 | 0,03 | -0,49 | 6,24E-01 |
| ZFP57 | 42,97 | 0,19 | 0,38 | 0,49 | 6,24E-01 |
| MREG | 206,31 | 0,02 | 0,04 | 0,49 | 6,24E-01 |
| TCEANC | 593,49 | 0,02 | 0,05 | 0,49 | 6,24E-01 |
| UBE4B | 1986,40 | 0,03 | 0,05 | 0,49 | 6,25E-01 |
| MAML2 | 1524,95 | 0,02 | 0,04 | 0,49 | 6,25E-01 |
| RBM22 | 908,37 | -0,01 | 0,03 | -0,49 | 6,25E-01 |
| NDST1 | 1731,09 | 0,05 | 0,09 | 0,49 | 6,25E-01 |
| FLYWCH2 | 166,73 | -0,02 | 0,04 | -0,49 | 6,25E-01 |
| KLHL36 | 2850,36 | 0,02 | 0,03 | 0,49 | 6,25E-01 |
| CCRL2 | 148,92 | 0,03 | 0,06 | 0,49 | 6,25E-01 |
| RIPK2 | 634,28 | -0,02 | 0,05 | -0,49 | 6,26E-01 |
| NEU3 | 675,51 | 0,02 | 0,04 | 0,49 | 6,26E-01 |
| DUSP22 | 2375,54 | -0,02 | 0,03 | -0,49 | 6,26E-01 |
| ATP2A2 | 2632,02 | 0,02 | 0,03 | 0,49 | 6,26E-01 |
| COLGALT2 | 155,22 | 0,06 | 0,13 | 0,49 | 6,27E-01 |
| MCTP1 | 3344,36 | -0,03 | 0,06 | -0,49 | 6,27E-01 |
| PAQR8 | 618,79 | 0,02 | 0,05 | 0,49 | 6,27E-01 |
| CASP5 | 472,11 | -0,06 | 0,11 | -0,48 | 6,28E-01 |
| WDR49 | 244,78 | 0,04 | 0,08 | 0,48 | 6,28E-01 |
| SLC7A11 | 138,22 | -0,04 | 0,08 | -0,48 | 6,28E-01 |
| BTBD9 | 1315,19 | 0,02 | 0,03 | 0,48 | 6,28E-01 |
| LCLAT1 | 764,23 | -0,02 | 0,05 | -0,48 | 6,29E-01 |
| POLE4 | 314,72 | -0,02 | 0,04 | -0,48 | 6,29E-01 |
| TUBD1 | 351,92 | -0,02 | 0,03 | -0,48 | 6,29E-01 |
| ACVR2A | 471,35 | -0,02 | 0,05 | -0,48 | 6,29E-01 |
| CD2BP2 | 1539,71 | -0,01 | 0,03 | -0,48 | 6,30E-01 |
| RBX1 | 1149,96 | 0,03 | 0,07 | 0,48 | 6,30E-01 |
| ERCC3 | 1164,05 | 0,01 | 0,03 | 0,48 | 6,30E-01 |

| | | | | | |
|---|---|---|---|---|---|
| BCL2L11 | 1771,97 | 0,02 | 0,05 | 0,48 | 6,30E-01 |
| USP5 | 812,82 | -0,02 | 0,04 | -0,48 | 6,31E-01 |
| MLKL | 2533,73 | 0,03 | 0,05 | 0,48 | 6,31E-01 |
| ZNF257 | 222,31 | -0,04 | 0,09 | -0,48 | 6,31E-01 |
| TRANK1 | 13376,87 | -0,03 | 0,06 | -0,48 | 6,31E-01 |
| TPD52L2 | 3175,19 | -0,03 | 0,05 | -0,48 | 6,31E-01 |
| KLHDC3 | 1193,20 | 0,01 | 0,02 | 0,48 | 6,32E-01 |
| MS4A4A | 165,83 | 0,04 | 0,09 | 0,48 | 6,32E-01 |
| MAF | 878,82 | -0,03 | 0,06 | -0,48 | 6,32E-01 |
| SYS1 | 547,25 | 0,02 | 0,04 | 0,48 | 6,33E-01 |
| PCYT1A | 3013,72 | -0,02 | 0,04 | -0,48 | 6,33E-01 |
| MTRFR | 554,33 | 0,02 | 0,03 | 0,48 | 6,33E-01 |
| TNIP3 | 83,12 | -0,06 | 0,12 | -0,48 | 6,34E-01 |
| CTBS | 4023,08 | -0,03 | 0,06 | -0,48 | 6,34E-01 |
| SHTN1 | 569,83 | -0,04 | 0,07 | -0,48 | 6,34E-01 |
| TP53INP2 | 167,97 | 0,05 | 0,11 | 0,48 | 6,34E-01 |
| RAB8A | 1385,91 | 0,02 | 0,05 | 0,48 | 6,34E-01 |
| WDR26 | 10213,01 | -0,02 | 0,05 | -0,48 | 6,35E-01 |
| HSCB | 151,85 | -0,02 | 0,04 | -0,48 | 6,35E-01 |
| DDX39A | 780,97 | 0,02 | 0,03 | 0,47 | 6,35E-01 |
| MYEF2 | 510,42 | 0,04 | 0,08 | 0,47 | 6,35E-01 |
| CFAP45 | 241,34 | 0,04 | 0,09 | 0,47 | 6,35E-01 |
| IRF1 | 18230,82 | 0,03 | 0,05 | 0,47 | 6,36E-01 |
| TAF4B | 341,22 | 0,02 | 0,05 | 0,47 | 6,36E-01 |
| MARF1 | 5843,83 | 0,03 | 0,06 | 0,47 | 6,36E-01 |
| MARCO | 228,05 | -0,05 | 0,12 | -0,47 | 6,37E-01 |
| ANKZF1 | 2159,73 | 0,02 | 0,03 | 0,47 | 6,38E-01 |
| SLC16A3 | 8804,65 | 0,03 | 0,06 | 0,47 | 6,38E-01 |
| KIF1B | 2761,29 | -0,03 | 0,07 | -0,47 | 6,38E-01 |
| HEATR5A | 1042,40 | -0,02 | 0,04 | -0,47 | 6,38E-01 |
| SNUPN | 248,71 | -0,01 | 0,03 | -0,47 | 6,38E-01 |
| ERV3-1 | 3177,12 | -0,03 | 0,06 | -0,47 | 6,39E-01 |
| KIF14 | 120,24 | 0,06 | 0,12 | 0,47 | 6,39E-01 |
| DHX58 | 651,86 | 0,04 | 0,09 | 0,47 | 6,39E-01 |
| ENSG00000275993 | 25,81 | 0,06 | 0,13 | 0,47 | 6,39E-01 |
| TCFL5 | 308,08 | -0,02 | 0,03 | -0,47 | 6,39E-01 |
| CYBRD1 | 1386,03 | -0,03 | 0,07 | -0,47 | 6,39E-01 |
| IL6R | 11473,97 | -0,03 | 0,06 | -0,47 | 6,40E-01 |
| SPECC1 | 4824,47 | 0,04 | 0,08 | 0,47 | 6,40E-01 |
| PSMD11 | 994,60 | 0,01 | 0,02 | 0,47 | 6,40E-01 |
| SELENOH | 864,65 | 0,02 | 0,05 | 0,47 | 6,40E-01 |
| IRF2BP2 | 3323,46 | -0,01 | 0,03 | -0,47 | 6,40E-01 |
| AKIRIN2 | 1067,36 | -0,02 | 0,04 | -0,47 | 6,40E-01 |
| ZNF212 | 393,79 | 0,02 | 0,03 | 0,47 | 6,40E-01 |
| PLEKHO1 | 3413,26 | -0,02 | 0,04 | -0,47 | 6,41E-01 |
| PIH1D1 | 734,82 | 0,01 | 0,03 | 0,47 | 6,41E-01 |

| | | | | | |
|---|---|---|---|---|---|
| PPP1R9A | 125,37 | 0,06 | 0,12 | 0,47 | 6,41E-01 |
| CD9 | 692,50 | 0,05 | 0,10 | 0,47 | 6,41E-01 |
| DYNC1H1 | 7490,90 | 0,02 | 0,05 | 0,47 | 6,42E-01 |
| PROK2 | 2928,09 | -0,04 | 0,09 | -0,46 | 6,42E-01 |
| KATNBL1 | 3279,60 | 0,03 | 0,06 | 0,46 | 6,42E-01 |
| ENSG00000266302 | 119,53 | -0,14 | 0,30 | -0,46 | 6,43E-01 |
| ALOX5 | 6311,59 | 0,03 | 0,06 | 0,46 | 6,43E-01 |
| GM2A | 1424,27 | 0,03 | 0,07 | 0,46 | 6,42E-01 |
| LPCAT1 | 2381,56 | -0,03 | 0,06 | -0,46 | 6,43E-01 |
| CKAP4 | 930,19 | 0,03 | 0,07 | 0,46 | 6,43E-01 |
| ADAMTS6 | 137,36 | 0,05 | 0,10 | 0,46 | 6,43E-01 |
| TCF20 | 3163,05 | -0,02 | 0,04 | -0,46 | 6,43E-01 |
| HGF | 518,05 | -0,03 | 0,07 | -0,46 | 6,44E-01 |
| IFTAP | 35,32 | -0,04 | 0,09 | -0,46 | 6,44E-01 |
| ARK2N | 3461,93 | -0,02 | 0,04 | -0,46 | 6,44E-01 |
| SCML1 | 324,00 | 0,04 | 0,08 | 0,46 | 6,44E-01 |
| CD38 | 720,24 | 0,03 | 0,07 | 0,46 | 6,44E-01 |
| CCDC69 | 2285,70 | 0,03 | 0,06 | 0,46 | 6,44E-01 |
| SLFN13 | 1471,90 | -0,02 | 0,05 | -0,46 | 6,45E-01 |
| POLR3C | 438,31 | 0,01 | 0,02 | 0,46 | 6,45E-01 |
| USP19 | 2388,74 | 0,02 | 0,04 | 0,46 | 6,45E-01 |
| TMEM42 | 387,86 | 0,02 | 0,04 | 0,46 | 6,45E-01 |
| CENPW | 26,12 | 0,04 | 0,09 | 0,46 | 6,45E-01 |
| TIMM9 | 81,76 | -0,02 | 0,05 | -0,46 | 6,45E-01 |
| ELL | 1585,93 | 0,03 | 0,07 | 0,46 | 6,46E-01 |
| GATM | 106,33 | 0,05 | 0,10 | 0,46 | 6,46E-01 |
| MARK3 | 5387,75 | 0,02 | 0,04 | 0,46 | 6,46E-01 |
| TOE1 | 253,48 | 0,01 | 0,03 | 0,46 | 6,47E-01 |
| MAD2L1BP | 364,08 | 0,02 | 0,04 | 0,46 | 6,47E-01 |
| SGPP1 | 876,42 | -0,03 | 0,06 | -0,46 | 6,47E-01 |
| CD14 | 4559,61 | -0,03 | 0,07 | -0,46 | 6,47E-01 |
| EXT1 | 835,90 | 0,02 | 0,05 | 0,46 | 6,47E-01 |
| BMAL1 | 2713,83 | -0,02 | 0,04 | -0,46 | 6,47E-01 |
| HLA-DQB2 | 67,58 | 0,10 | 0,22 | 0,46 | 6,47E-01 |
| ANKRD39 | 148,04 | 0,01 | 0,03 | 0,46 | 6,47E-01 |
| SIGLEC7 | 336,48 | 0,03 | 0,07 | 0,46 | 6,48E-01 |
| LILRA4 | 56,92 | -0,05 | 0,11 | -0,46 | 6,48E-01 |
| C16orf54 | 3051,58 | 0,03 | 0,06 | 0,46 | 6,48E-01 |
| DCK | 2047,61 | -0,03 | 0,06 | -0,46 | 6,48E-01 |
| IER2 | 1988,59 | 0,02 | 0,05 | 0,46 | 6,48E-01 |
| ADTRP | 404,06 | 0,05 | 0,11 | 0,46 | 6,49E-01 |
| RPGRIP1L | 197,68 | 0,03 | 0,07 | 0,46 | 6,49E-01 |
| FDPS | 1164,94 | -0,01 | 0,02 | -0,46 | 6,49E-01 |
| ADA2 | 3343,78 | 0,03 | 0,06 | 0,45 | 6,49E-01 |
| PSMC3IP | 65,66 | 0,02 | 0,05 | 0,45 | 6,50E-01 |
| RASSF3 | 6159,09 | -0,02 | 0,05 | -0,45 | 6,50E-01 |

| | | | | | |
|---|---|---|---|---|---|
| IDS | 4178,32 | 0,03 | 0,06 | 0,45 | 6,50E-01 |
| ENSG00000285444 | 334,83 | 0,04 | 0,10 | 0,45 | 6,50E-01 |
| TMEM204 | 645,10 | -0,03 | 0,07 | -0,45 | 6,51E-01 |
| TCP11L2 | 10253,76 | 0,02 | 0,05 | 0,45 | 6,51E-01 |
| ARID1B | 6785,91 | -0,02 | 0,04 | -0,45 | 6,51E-01 |
| AGAP6 | 453,69 | 0,02 | 0,05 | 0,45 | 6,51E-01 |
| ACKR3 | 131,38 | 0,02 | 0,05 | 0,45 | 6,52E-01 |
| HADH | 479,28 | 0,02 | 0,04 | 0,45 | 6,52E-01 |
| PLGLB2 | 51,33 | -0,07 | 0,16 | -0,45 | 6,52E-01 |
| NDUFAF6 | 391,03 | 0,02 | 0,03 | 0,45 | 6,53E-01 |
| TGFBI | 1836,95 | 0,03 | 0,06 | 0,45 | 6,53E-01 |
| SRPRA | 1662,45 | 0,02 | 0,05 | 0,45 | 6,53E-01 |
| RAD54B | 72,10 | 0,04 | 0,08 | 0,45 | 6,54E-01 |
| CNTLN | 422,22 | -0,03 | 0,06 | -0,45 | 6,54E-01 |
| DUSP28 | 290,64 | 0,01 | 0,03 | 0,45 | 6,55E-01 |
| CPT2 | 656,74 | 0,01 | 0,02 | 0,45 | 6,55E-01 |
| BLVRA | 469,13 | -0,03 | 0,06 | -0,45 | 6,55E-01 |
| NOP14 | 776,92 | 0,02 | 0,03 | 0,45 | 6,55E-01 |
| RIPK1 | 1431,92 | -0,02 | 0,04 | -0,45 | 6,55E-01 |
| AP1S3 | 125,31 | 0,03 | 0,08 | 0,45 | 6,56E-01 |
| MAP3K20 | 958,59 | 0,02 | 0,05 | 0,45 | 6,56E-01 |
| BRK1 | 1813,15 | -0,02 | 0,06 | -0,44 | 6,57E-01 |
| PLEKHA5 | 277,02 | -0,04 | 0,09 | -0,44 | 6,57E-01 |
| BTN2A1 | 1536,41 | 0,03 | 0,06 | 0,44 | 6,57E-01 |
| JPT1 | 1707,66 | -0,02 | 0,05 | -0,44 | 6,57E-01 |
| EIF1 | 3090,77 | 0,02 | 0,05 | 0,44 | 6,57E-01 |
| SCAI | 718,40 | -0,02 | 0,04 | -0,44 | 6,57E-01 |
| ARID3A | 1506,84 | 0,03 | 0,07 | 0,44 | 6,57E-01 |
| ZCWPW2 | 41,37 | 0,03 | 0,07 | 0,44 | 6,59E-01 |
| PRXL2C | 310,66 | -0,02 | 0,04 | -0,44 | 6,59E-01 |
| TSPYL4 | 459,66 | 0,01 | 0,03 | 0,44 | 6,59E-01 |
| ARMC2 | 383,07 | 0,02 | 0,05 | 0,44 | 6,59E-01 |
| INTS6L | 1853,54 | -0,02 | 0,04 | -0,44 | 6,59E-01 |
| TMOD2 | 2684,44 | -0,02 | 0,05 | -0,44 | 6,59E-01 |
| ZNF365 | 75,74 | -0,05 | 0,10 | -0,44 | 6,59E-01 |
| B3GAT1 | 267,10 | 0,06 | 0,14 | 0,44 | 6,60E-01 |
| THAP7 | 406,02 | 0,02 | 0,04 | 0,44 | 6,60E-01 |
| NEDD9 | 6034,30 | -0,03 | 0,06 | -0,44 | 6,60E-01 |
| SEPTIN6 | 2457,81 | -0,02 | 0,04 | -0,44 | 6,60E-01 |
| SLC35B2 | 429,62 | 0,01 | 0,03 | 0,44 | 6,61E-01 |
| GPATCH3 | 260,59 | 0,01 | 0,03 | 0,44 | 6,61E-01 |
| NPRL2 | 670,35 | -0,01 | 0,03 | -0,44 | 6,61E-01 |
| MRPL55 | 259,69 | 0,01 | 0,03 | 0,44 | 6,62E-01 |
| MS4A4E | 416,96 | -0,03 | 0,07 | -0,44 | 6,62E-01 |
| TWNK | 257,04 | 0,02 | 0,04 | 0,44 | 6,62E-01 |
| IRF2 | 2856,15 | -0,02 | 0,05 | -0,44 | 6,62E-01 |

| | | | | | |
|---|---|---|---|---|---|
| MRPL28 | 608,38 | -0,02 | 0,03 | -0,44 | 6,63E-01 |
| RHBDD1 | 1567,89 | 0,02 | 0,05 | 0,44 | 6,63E-01 |
| GOT1 | 195,06 | 0,02 | 0,04 | 0,44 | 6,63E-01 |
| ZNF565 | 120,85 | 0,02 | 0,05 | 0,44 | 6,63E-01 |
| ZSCAN23 | 66,77 | -0,05 | 0,12 | -0,43 | 6,64E-01 |
| FBXL20 | 2478,05 | 0,02 | 0,05 | 0,44 | 6,64E-01 |
| CALML4 | 98,26 | -0,02 | 0,05 | -0,43 | 6,64E-01 |
| CLDND2 | 179,32 | 0,04 | 0,09 | 0,43 | 6,64E-01 |
| TRABD2A | 2457,15 | 0,03 | 0,07 | 0,43 | 6,64E-01 |
| INKA2 | 386,78 | -0,03 | 0,07 | -0,43 | 6,64E-01 |
| PRH1 | 362,07 | -0,02 | 0,06 | -0,43 | 6,65E-01 |
| STXBP4 | 515,27 | -0,02 | 0,05 | -0,43 | 6,65E-01 |
| TMEM252 | 135,33 | -0,04 | 0,10 | -0,43 | 6,65E-01 |
| MACROD2 | 116,27 | 0,06 | 0,15 | 0,43 | 6,65E-01 |
| ANKRD13A | 8661,24 | -0,02 | 0,04 | -0,43 | 6,65E-01 |
| CNDP2 | 2193,99 | -0,02 | 0,04 | -0,43 | 6,66E-01 |
| PAK2 | 4892,37 | -0,02 | 0,05 | -0,43 | 6,66E-01 |
| CA4 | 371,94 | -0,04 | 0,10 | -0,43 | 6,66E-01 |
| AAMDC | 92,27 | 0,02 | 0,05 | 0,43 | 6,67E-01 |
| RNASEH2A | 24,51 | 0,03 | 0,08 | 0,43 | 6,67E-01 |
| CDS2 | 5518,64 | -0,02 | 0,05 | -0,43 | 6,67E-01 |
| DHX35 | 612,17 | -0,01 | 0,03 | -0,43 | 6,67E-01 |
| CCDC122 | 166,55 | -0,04 | 0,10 | -0,43 | 6,67E-01 |
| NIT2 | 658,32 | 0,01 | 0,03 | 0,43 | 6,67E-01 |
| RALBP1 | 3384,38 | -0,02 | 0,05 | -0,43 | 6,67E-01 |
| MAPKAPK3 | 2947,48 | 0,02 | 0,04 | 0,43 | 6,68E-01 |
| TSC1 | 2043,07 | 0,01 | 0,03 | 0,43 | 6,68E-01 |
| SLC4A4 | 226,67 | -0,04 | 0,09 | -0,43 | 6,68E-01 |
| MPZL2 | 392,26 | -0,03 | 0,07 | -0,43 | 6,68E-01 |
| LRRC23 | 94,76 | 0,03 | 0,06 | 0,43 | 6,69E-01 |
| SDCCAG8 | 1138,92 | -0,01 | 0,03 | -0,43 | 6,69E-01 |
| SARNP | 196,08 | 0,01 | 0,03 | 0,43 | 6,69E-01 |
| TCF25 | 2521,65 | 0,02 | 0,04 | 0,43 | 6,69E-01 |
| C1QTNF3 | 120,64 | 0,02 | 0,06 | 0,43 | 6,69E-01 |
| PITPNA | 2118,90 | -0,02 | 0,06 | -0,43 | 6,69E-01 |
| ZNF827 | 304,00 | 0,03 | 0,07 | 0,43 | 6,69E-01 |
| RELL1 | 1610,07 | -0,03 | 0,06 | -0,43 | 6,70E-01 |
| BATF | 188,16 | -0,02 | 0,05 | -0,43 | 6,70E-01 |
| GBP4 | 2732,59 | 0,04 | 0,09 | 0,43 | 6,71E-01 |
| NBPF26 | 2111,94 | -0,04 | 0,11 | -0,43 | 6,71E-01 |
| GABPB2 | 1176,75 | 0,01 | 0,03 | 0,42 | 6,71E-01 |
| CD300A | 3386,25 | 0,02 | 0,05 | 0,42 | 6,71E-01 |
| LRRC57 | 704,56 | 0,01 | 0,03 | 0,42 | 6,72E-01 |
| TKT | 9800,23 | 0,02 | 0,06 | 0,42 | 6,73E-01 |
| HGSNAT | 1186,54 | 0,02 | 0,05 | 0,42 | 6,73E-01 |
| RILPL2 | 2195,95 | 0,02 | 0,04 | 0,42 | 6,73E-01 |

| | | | | | |
|---|---|---|---|---|---|
| TGOLN2 | 8392,70 | 0,02 | 0,05 | 0,42 | 6,73E-01 |
| GPR155 | 3212,03 | -0,02 | 0,05 | -0,42 | 6,73E-01 |
| LRRN3 | 274,18 | -0,06 | 0,14 | -0,42 | 6,73E-01 |
| PIP4P1 | 942,70 | 0,01 | 0,02 | 0,42 | 6,74E-01 |
| METRNL | 1483,27 | -0,02 | 0,06 | -0,42 | 6,74E-01 |
| SNRNP200 | 4890,86 | -0,02 | 0,04 | -0,42 | 6,74E-01 |
| DCP1B | 377,00 | -0,02 | 0,04 | -0,42 | 6,74E-01 |
| UQCR10 | 525,44 | -0,02 | 0,06 | -0,42 | 6,74E-01 |
| FANCI | 397,21 | 0,02 | 0,04 | 0,42 | 6,74E-01 |
| FAM200C | 64,61 | -0,03 | 0,07 | -0,42 | 6,74E-01 |
| USP4 | 4580,44 | 0,02 | 0,05 | 0,42 | 6,75E-01 |
| JDP2 | 1060,32 | 0,02 | 0,06 | 0,42 | 6,75E-01 |
| CMSS1 | 271,69 | 0,02 | 0,05 | 0,42 | 6,76E-01 |
| TMEM116 | 733,47 | 0,02 | 0,04 | 0,42 | 6,77E-01 |
| WDR74 | 467,12 | 0,01 | 0,03 | 0,42 | 6,77E-01 |
| LRRFIP2 | 2567,41 | -0,02 | 0,04 | -0,42 | 6,77E-01 |
| PDCD2L | 49,11 | 0,02 | 0,05 | 0,42 | 6,77E-01 |
| ZNHIT3 | 603,27 | -0,02 | 0,04 | -0,42 | 6,78E-01 |
| ZSWIM6 | 2298,29 | -0,02 | 0,04 | -0,41 | 6,78E-01 |
| MRPS16 | 1365,64 | -0,01 | 0,03 | -0,41 | 6,79E-01 |
| TMT1A | 2531,52 | -0,02 | 0,05 | -0,41 | 6,79E-01 |
| KDELR2 | 793,23 | 0,01 | 0,02 | 0,41 | 6,79E-01 |
| QRICH1 | 1610,69 | 0,01 | 0,03 | 0,41 | 6,79E-01 |
| CD22 | 627,99 | -0,04 | 0,10 | -0,41 | 6,80E-01 |
| TENM1 | 591,14 | -0,04 | 0,10 | -0,41 | 6,79E-01 |
| PPP1R11 | 347,16 | 0,02 | 0,04 | 0,41 | 6,80E-01 |
| WIPI1 | 940,93 | 0,02 | 0,05 | 0,41 | 6,80E-01 |
| NUDT7 | 71,64 | -0,03 | 0,07 | -0,41 | 6,80E-01 |
| EML5 | 390,99 | 0,03 | 0,07 | 0,41 | 6,80E-01 |
| SF3B2 | 1461,87 | 0,02 | 0,05 | 0,41 | 6,80E-01 |
| ZC2HC1A | 291,61 | -0,04 | 0,09 | -0,41 | 6,80E-01 |
| DYRK4 | 299,09 | 0,02 | 0,05 | 0,41 | 6,81E-01 |
| CYP51A1 | 85,65 | 0,02 | 0,05 | 0,41 | 6,81E-01 |
| TRMT10B | 352,60 | 0,01 | 0,03 | 0,41 | 6,81E-01 |
| SWI5 | 97,70 | -0,02 | 0,04 | -0,41 | 6,81E-01 |
| DUSP16 | 709,53 | 0,02 | 0,05 | 0,41 | 6,81E-01 |
| TAS2R14 | 302,51 | -0,03 | 0,08 | -0,41 | 6,81E-01 |
| NUB1 | 2254,99 | -0,02 | 0,05 | -0,41 | 6,82E-01 |
| ATP6V0A1 | 1649,22 | 0,02 | 0,05 | 0,41 | 6,82E-01 |
| GNAQ | 6086,45 | -0,02 | 0,05 | -0,41 | 6,83E-01 |
| FGGY | 481,64 | -0,02 | 0,06 | -0,41 | 6,83E-01 |
| THOC3 | 707,91 | 0,02 | 0,05 | 0,41 | 6,83E-01 |
| RFC5 | 297,27 | 0,01 | 0,03 | 0,41 | 6,84E-01 |
| ZNF787 | 936,81 | 0,02 | 0,04 | 0,41 | 6,84E-01 |
| SLC18B1 | 671,75 | 0,02 | 0,05 | 0,41 | 6,84E-01 |
| ZNF404 | 43,11 | -0,04 | 0,09 | -0,41 | 6,84E-01 |

| | | | | | |
|---|---|---|---|---|---|
| VPS11 | 936,73 | 0,01 | 0,03 | 0,41 | 6,84E-01 |
| EHD4 | 698,84 | 0,02 | 0,04 | 0,41 | 6,85E-01 |
| GPSM2 | 420,17 | -0,02 | 0,06 | -0,41 | 6,85E-01 |
| SLC22A4 | 1369,65 | 0,03 | 0,07 | 0,41 | 6,85E-01 |
| CACNA2D3 | 188,81 | 0,03 | 0,08 | 0,40 | 6,86E-01 |
| COMMD5 | 565,02 | -0,01 | 0,02 | -0,40 | 6,86E-01 |
| MYOM2 | 434,32 | -0,09 | 0,23 | -0,40 | 6,87E-01 |
| CYBA | 9778,45 | -0,02 | 0,05 | -0,40 | 6,87E-01 |
| DOLK | 129,83 | 0,01 | 0,03 | 0,40 | 6,88E-01 |
| SEC61A2 | 496,97 | 0,02 | 0,05 | 0,40 | 6,88E-01 |
| CEBPA | 1262,16 | 0,02 | 0,05 | 0,40 | 6,88E-01 |
| DDX31 | 477,12 | 0,02 | 0,04 | 0,40 | 6,88E-01 |
| CD3E | 657,77 | 0,03 | 0,07 | 0,40 | 6,88E-01 |
| LYPLA1 | 1822,29 | -0,02 | 0,05 | -0,40 | 6,88E-01 |
| FPR1 | 13363,46 | 0,03 | 0,07 | 0,40 | 6,88E-01 |
| FAM216A | 235,47 | -0,02 | 0,05 | -0,40 | 6,88E-01 |
| SERGEF | 590,29 | -0,02 | 0,04 | -0,40 | 6,89E-01 |
| SNAP47 | 490,15 | 0,01 | 0,03 | 0,40 | 6,89E-01 |
| UBASH3A | 989,49 | 0,02 | 0,06 | 0,40 | 6,89E-01 |
| RCAN3 | 2787,14 | 0,02 | 0,06 | 0,40 | 6,89E-01 |
| PLGRKT | 284,67 | -0,02 | 0,04 | -0,40 | 6,89E-01 |
| CMTR1 | 2313,72 | -0,02 | 0,05 | -0,40 | 6,89E-01 |
| GBP1 | 3332,16 | 0,05 | 0,12 | 0,40 | 6,90E-01 |
| CLHC1 | 241,17 | 0,03 | 0,08 | 0,40 | 6,91E-01 |
| SCN3A | 184,04 | 0,05 | 0,13 | 0,40 | 6,91E-01 |
| TNRC6A | 3068,96 | -0,02 | 0,04 | -0,40 | 6,92E-01 |
| MTMR14 | 2874,01 | 0,01 | 0,04 | 0,40 | 6,92E-01 |
| ABCA1 | 4894,58 | -0,04 | 0,09 | -0,40 | 6,93E-01 |
| NXPE3 | 1569,17 | -0,01 | 0,03 | -0,40 | 6,93E-01 |
| MAGEF1 | 207,22 | -0,02 | 0,05 | -0,39 | 6,93E-01 |
| RHOB | 2932,83 | -0,02 | 0,06 | -0,39 | 6,93E-01 |
| INPP1 | 306,30 | 0,02 | 0,04 | 0,39 | 6,93E-01 |
| FAM168A | 1240,68 | 0,02 | 0,04 | 0,39 | 6,93E-01 |
| ZNF57 | 64,52 | -0,02 | 0,05 | -0,39 | 6,94E-01 |
| MGAT1 | 4768,67 | 0,02 | 0,04 | 0,39 | 6,94E-01 |
| SMG1 | 12583,10 | -0,02 | 0,04 | -0,39 | 6,94E-01 |
| MTPAP | 692,44 | -0,01 | 0,02 | -0,39 | 6,94E-01 |
| GID8 | 1599,29 | 0,01 | 0,03 | 0,39 | 6,94E-01 |
| BNIP3 | 415,93 | 0,02 | 0,04 | 0,39 | 6,94E-01 |
| UPF3A | 1001,90 | -0,01 | 0,03 | -0,39 | 6,95E-01 |
| TRAPPC2B | 60,26 | 0,03 | 0,07 | 0,39 | 6,95E-01 |
| RUNX2 | 1420,26 | -0,02 | 0,05 | -0,39 | 6,95E-01 |
| GET1 | 441,97 | 0,02 | 0,05 | 0,39 | 6,95E-01 |
| CCDC71L | 878,00 | 0,02 | 0,06 | 0,39 | 6,95E-01 |
| DYNC2I1 | 501,99 | -0,02 | 0,04 | -0,39 | 6,96E-01 |
| PNMA3 | 210,23 | 0,04 | 0,11 | 0,39 | 6,96E-01 |

| | | | | | |
|---|---|---|---|---|---|
| CSNK2A2 | 1062,37 | -0,01 | 0,03 | -0,39 | 6,96E-01 |
| TRIM34 | 44,52 | -0,02 | 0,06 | -0,39 | 6,96E-01 |
| GADD45B | 665,53 | 0,02 | 0,06 | 0,39 | 6,96E-01 |
| GPR55 | 117,19 | 0,03 | 0,08 | 0,39 | 6,97E-01 |
| ATXN7 | 1021,73 | -0,02 | 0,04 | -0,39 | 6,97E-01 |
| DNMBP | 413,28 | 0,03 | 0,07 | 0,39 | 6,98E-01 |
| ALKBH5 | 2136,56 | 0,01 | 0,03 | 0,39 | 6,98E-01 |
| POU2F1 | 2633,07 | -0,02 | 0,04 | -0,39 | 6,99E-01 |
| GATAD1 | 1166,17 | -0,01 | 0,03 | -0,39 | 6,99E-01 |
| TM9SF1 | 132,43 | -0,01 | 0,03 | -0,39 | 6,99E-01 |
| SLC10A7 | 519,91 | -0,01 | 0,04 | -0,39 | 7,00E-01 |
| SMAP2 | 10464,05 | 0,03 | 0,07 | 0,38 | 7,01E-01 |
| CDKN2B | 154,68 | -0,03 | 0,07 | -0,38 | 7,01E-01 |
| BTN3A3 | 3005,51 | 0,02 | 0,04 | 0,38 | 7,01E-01 |
| CCDC85B | 499,24 | 0,02 | 0,05 | 0,38 | 7,01E-01 |
| SLC9B2 | 281,51 | 0,02 | 0,04 | 0,38 | 7,01E-01 |
| FAM53B | 2457,03 | 0,02 | 0,05 | 0,38 | 7,01E-01 |
| LEMD2 | 2470,28 | -0,01 | 0,02 | -0,38 | 7,02E-01 |
| GTF2IRD2B | 448,44 | 0,01 | 0,03 | 0,38 | 7,02E-01 |
| NHERF1 | 2188,64 | 0,02 | 0,06 | 0,38 | 7,03E-01 |
| HOOK3 | 3607,11 | -0,01 | 0,03 | -0,38 | 7,03E-01 |
| TRAPPC3L | 43,13 | -0,05 | 0,13 | -0,38 | 7,03E-01 |
| SLC22A1 | 346,87 | 0,04 | 0,10 | 0,38 | 7,03E-01 |
| IFNGR2 | 4362,30 | 0,02 | 0,05 | 0,38 | 7,03E-01 |
| VPS26C | 2076,02 | 0,01 | 0,02 | 0,38 | 7,03E-01 |
| MARS2 | 438,57 | -0,02 | 0,04 | -0,38 | 7,03E-01 |
| TNRC6B | 11327,22 | -0,02 | 0,05 | -0,38 | 7,03E-01 |
| ADAT1 | 1473,36 | 0,01 | 0,03 | 0,38 | 7,03E-01 |
| FLVCR1 | 1022,67 | 0,02 | 0,05 | 0,38 | 7,03E-01 |
| APP | 1706,11 | -0,02 | 0,05 | -0,38 | 7,03E-01 |
| UQCC3 | 177,36 | -0,02 | 0,05 | -0,38 | 7,04E-01 |
| MYL6 | 9706,90 | -0,04 | 0,10 | -0,38 | 7,05E-01 |
| AKR1A1 | 668,40 | 0,01 | 0,03 | 0,38 | 7,05E-01 |
| BST1 | 1865,86 | -0,02 | 0,07 | -0,38 | 7,05E-01 |
| JAGN1 | 344,87 | -0,01 | 0,03 | -0,38 | 7,05E-01 |
| UBE2J1 | 2606,45 | -0,02 | 0,06 | -0,38 | 7,05E-01 |
| MICU3 | 221,20 | -0,04 | 0,09 | -0,38 | 7,05E-01 |
| DMAC2L | 1204,25 | 0,01 | 0,03 | 0,38 | 7,06E-01 |
| PTGER2 | 788,75 | -0,02 | 0,04 | -0,38 | 7,06E-01 |
| PLCL1 | 616,89 | 0,02 | 0,06 | 0,38 | 7,06E-01 |
| PITRM1 | 1007,55 | 0,01 | 0,04 | 0,38 | 7,06E-01 |
| JPT2 | 525,78 | -0,01 | 0,03 | -0,38 | 7,06E-01 |
| RPS11 | 18790,39 | -0,03 | 0,07 | -0,38 | 7,07E-01 |
| PLEK | 4505,45 | -0,02 | 0,06 | -0,38 | 7,07E-01 |
| KCMF1 | 2295,52 | -0,01 | 0,03 | -0,38 | 7,07E-01 |
| DLG3 | 253,84 | 0,02 | 0,07 | 0,38 | 7,07E-01 |

| | | | | | |
|---|---|---|---|---|---|
| LRRC8C | 2327,71 | -0,02 | 0,04 | -0,38 | 7,07E-01 |
| ZNF491 | 102,45 | 0,02 | 0,06 | 0,38 | 7,07E-01 |
| FAM177A1 | 796,29 | 0,01 | 0,03 | 0,37 | 7,08E-01 |
| IL1R1 | 979,43 | 0,04 | 0,11 | 0,37 | 7,09E-01 |
| CFAP44 | 362,92 | 0,02 | 0,05 | 0,37 | 7,09E-01 |
| TMEM230 | 1220,18 | -0,01 | 0,03 | -0,37 | 7,09E-01 |
| MED13L | 15030,59 | -0,02 | 0,04 | -0,37 | 7,10E-01 |
| GK | 5894,56 | -0,03 | 0,08 | -0,37 | 7,10E-01 |
| CMTM1 | 181,24 | 0,02 | 0,07 | 0,37 | 7,10E-01 |
| TRIB2 | 1954,70 | -0,02 | 0,05 | -0,37 | 7,11E-01 |
| TNIP2 | 1057,59 | -0,01 | 0,03 | -0,37 | 7,11E-01 |
| NXT1 | 126,72 | -0,02 | 0,04 | -0,37 | 7,11E-01 |
| PLGLB1 | 131,58 | -0,03 | 0,08 | -0,37 | 7,12E-01 |
| CPEB4 | 5835,00 | -0,02 | 0,05 | -0,37 | 7,12E-01 |
| DCTD | 581,59 | -0,01 | 0,02 | -0,37 | 7,12E-01 |
| CDC27 | 2854,84 | -0,02 | 0,04 | -0,37 | 7,12E-01 |
| CXCL16 | 567,01 | 0,03 | 0,08 | 0,37 | 7,12E-01 |
| PDE8A | 1144,49 | 0,01 | 0,03 | 0,37 | 7,12E-01 |
| SERAC1 | 447,22 | -0,02 | 0,04 | -0,37 | 7,13E-01 |
| CD93 | 10129,26 | -0,03 | 0,08 | -0,37 | 7,14E-01 |
| ATP1A1 | 3361,82 | 0,01 | 0,03 | 0,37 | 7,14E-01 |
| PID1 | 349,22 | 0,03 | 0,08 | 0,37 | 7,14E-01 |
| MTPN | 9735,43 | 0,02 | 0,05 | 0,37 | 7,14E-01 |
| BORCS6 | 489,03 | 0,01 | 0,03 | 0,37 | 7,15E-01 |
| EREG | 69,85 | 0,05 | 0,14 | 0,37 | 7,15E-01 |
| RPL8 | 10195,19 | 0,02 | 0,06 | 0,37 | 7,15E-01 |
| NOP56 | 1334,96 | -0,01 | 0,03 | -0,36 | 7,15E-01 |
| ZNF221 | 84,85 | -0,03 | 0,07 | -0,36 | 7,16E-01 |
| RPL18A | 9119,11 | 0,02 | 0,06 | 0,36 | 7,16E-01 |
| KIF13B | 1619,56 | 0,02 | 0,04 | 0,36 | 7,16E-01 |
| NME3 | 557,57 | 0,01 | 0,04 | 0,36 | 7,16E-01 |
| ATP5F1B | 6188,64 | -0,02 | 0,04 | -0,36 | 7,16E-01 |
| BTF3 | 3324,21 | -0,03 | 0,07 | -0,36 | 7,16E-01 |
| DCAF10 | 1859,81 | 0,01 | 0,03 | 0,36 | 7,17E-01 |
| DGKK | 111,92 | 0,06 | 0,16 | 0,36 | 7,17E-01 |
| SDHAF3 | 142,77 | -0,02 | 0,05 | -0,36 | 7,18E-01 |
| C12orf75 | 441,22 | -0,02 | 0,07 | -0,36 | 7,18E-01 |
| FAM20B | 1404,82 | 0,02 | 0,04 | 0,36 | 7,18E-01 |
| MCOLN2 | 535,64 | -0,03 | 0,08 | -0,36 | 7,18E-01 |
| INAFM2 | 1241,76 | -0,02 | 0,05 | -0,36 | 7,19E-01 |
| GNRH1 | 116,29 | -0,02 | 0,04 | -0,36 | 7,19E-01 |
| CDKN1C | 447,45 | 0,04 | 0,10 | 0,36 | 7,19E-01 |
| CCDC170 | 275,91 | 0,02 | 0,07 | 0,36 | 7,19E-01 |
| UBE2V1 | 719,47 | 0,02 | 0,04 | 0,36 | 7,19E-01 |
| ABHD15 | 596,76 | -0,01 | 0,04 | -0,36 | 7,20E-01 |
| NUDT6 | 118,88 | 0,02 | 0,06 | 0,36 | 7,20E-01 |

| | | | | | |
|---|---|---|---|---|---|
| VPS25 | 478,22 | -0,01 | 0,03 | -0,36 | 7,20E-01 |
| CDH26 | 264,36 | 0,02 | 0,06 | 0,36 | 7,20E-01 |
| SLC38A6 | 518,32 | 0,02 | 0,04 | 0,36 | 7,20E-01 |
| RASSF1 | 3083,98 | -0,02 | 0,04 | -0,36 | 7,20E-01 |
| CNTNAP3C | 472,53 | 0,05 | 0,14 | 0,36 | 7,20E-01 |
| CD52 | 4061,24 | -0,03 | 0,09 | -0,36 | 7,20E-01 |
| GOLM1 | 211,19 | 0,03 | 0,08 | 0,36 | 7,20E-01 |
| HSPA1L | 226,78 | -0,02 | 0,05 | -0,36 | 7,20E-01 |
| LRPAP1 | 2997,29 | 0,01 | 0,03 | 0,36 | 7,21E-01 |
| AAK1 | 5569,48 | 0,02 | 0,05 | 0,36 | 7,21E-01 |
| IRGM | 41,09 | 0,03 | 0,07 | 0,36 | 7,21E-01 |
| SPRED1 | 345,29 | 0,03 | 0,08 | 0,36 | 7,21E-01 |
| RPS17 | 297,76 | -0,01 | 0,03 | -0,36 | 7,21E-01 |
| PIK3AP1 | 7196,19 | -0,02 | 0,06 | -0,36 | 7,21E-01 |
| SUN2 | 8710,54 | 0,02 | 0,04 | 0,36 | 7,21E-01 |
| NCBP3 | 2062,83 | -0,01 | 0,03 | -0,36 | 7,21E-01 |
| SAMD12 | 421,46 | -0,03 | 0,09 | -0,36 | 7,22E-01 |
| ZBED4 | 1028,94 | -0,01 | 0,03 | -0,36 | 7,22E-01 |
| SP110 | 5570,31 | 0,02 | 0,06 | 0,36 | 7,22E-01 |
| MTFR1 | 186,53 | 0,01 | 0,04 | 0,36 | 7,22E-01 |
| PUDP | 247,89 | 0,01 | 0,03 | 0,35 | 7,23E-01 |
| IFT52 | 368,08 | 0,02 | 0,04 | 0,35 | 7,23E-01 |
| DCUN1D3 | 374,93 | 0,01 | 0,04 | 0,35 | 7,24E-01 |
| C8orf76 | 192,44 | 0,01 | 0,03 | 0,35 | 7,24E-01 |
| TARS3 | 872,32 | -0,01 | 0,04 | -0,35 | 7,24E-01 |
| ICMT | 811,82 | -0,01 | 0,03 | -0,35 | 7,24E-01 |
| HLA-B | 75476,35 | -0,02 | 0,05 | -0,35 | 7,24E-01 |
| MDM4 | 11351,05 | -0,01 | 0,03 | -0,35 | 7,24E-01 |
| TMEM272 | 1047,08 | 0,03 | 0,08 | 0,35 | 7,25E-01 |
| TALDO1 | 11231,78 | -0,02 | 0,06 | -0,35 | 7,25E-01 |
| ZNF512 | 711,79 | -0,01 | 0,03 | -0,35 | 7,25E-01 |
| NGRN | 758,32 | 0,01 | 0,03 | 0,35 | 7,26E-01 |
| ACVR2B | 671,57 | 0,02 | 0,06 | 0,35 | 7,25E-01 |
| ZIK1 | 262,55 | -0,02 | 0,05 | -0,35 | 7,26E-01 |
| DDX47 | 179,87 | 0,01 | 0,04 | 0,35 | 7,26E-01 |
| CCNJ | 462,79 | -0,01 | 0,03 | -0,35 | 7,26E-01 |
| SLC2A9 | 323,48 | 0,03 | 0,07 | 0,35 | 7,26E-01 |
| SH3BP5L | 2300,66 | -0,02 | 0,04 | -0,35 | 7,26E-01 |
| CDK5RAP1 | 795,70 | -0,01 | 0,02 | -0,35 | 7,26E-01 |
| SNX27 | 3953,96 | 0,02 | 0,07 | 0,35 | 7,27E-01 |
| COA4 | 471,47 | -0,01 | 0,03 | -0,35 | 7,27E-01 |
| PIK3IP1 | 4379,71 | -0,02 | 0,05 | -0,35 | 7,27E-01 |
| YIPF6 | 1946,23 | 0,02 | 0,04 | 0,35 | 7,27E-01 |
| TXNDC11 | 817,05 | 0,01 | 0,03 | 0,35 | 7,28E-01 |
| ZFYVE9 | 256,54 | 0,02 | 0,06 | 0,35 | 7,28E-01 |
| TSR3 | 637,59 | 0,01 | 0,04 | 0,35 | 7,28E-01 |

| | | | | | |
|---|---|---|---|---|---|
| EPB41L4A | 200,89 | 0,03 | 0,10 | 0,35 | 7,28E-01 |
| PIP4K2C | 635,20 | -0,01 | 0,03 | -0,35 | 7,28E-01 |
| ARG1 | 289,35 | 0,05 | 0,13 | 0,35 | 7,28E-01 |
| FAIM | 90,04 | 0,02 | 0,06 | 0,35 | 7,29E-01 |
| NET1 | 284,46 | 0,02 | 0,07 | 0,35 | 7,29E-01 |
| SDF2L1 | 239,90 | 0,02 | 0,05 | 0,35 | 7,29E-01 |
| YIPF1 | 663,86 | -0,01 | 0,04 | -0,35 | 7,29E-01 |
| IL10RA | 8071,20 | -0,01 | 0,04 | -0,35 | 7,29E-01 |
| LRATD2 | 517,36 | 0,02 | 0,05 | 0,35 | 7,30E-01 |
| TSC22D2 | 1078,15 | -0,01 | 0,03 | -0,35 | 7,30E-01 |
| RPRD1B | 711,91 | 0,02 | 0,05 | 0,35 | 7,30E-01 |
| NKIRAS1 | 107,31 | -0,02 | 0,05 | -0,35 | 7,30E-01 |
| CLTB | 1143,00 | -0,02 | 0,05 | -0,34 | 7,30E-01 |
| MBP | 10096,03 | -0,02 | 0,05 | -0,34 | 7,31E-01 |
| F5 | 1767,81 | 0,03 | 0,08 | 0,34 | 7,31E-01 |
| MLLT10 | 1743,27 | -0,01 | 0,03 | -0,34 | 7,31E-01 |
| WDR45B | 1318,33 | -0,01 | 0,02 | -0,34 | 7,31E-01 |
| TRMT2B | 1195,26 | -0,01 | 0,03 | -0,34 | 7,32E-01 |
| TSSC4 | 1032,90 | -0,01 | 0,04 | -0,34 | 7,32E-01 |
| LCP2 | 9995,93 | -0,01 | 0,04 | -0,34 | 7,32E-01 |
| FAHD1 | 459,89 | 0,02 | 0,06 | 0,34 | 7,32E-01 |
| DGKG | 726,15 | 0,02 | 0,07 | 0,34 | 7,32E-01 |
| ARFGAP2 | 2481,10 | 0,01 | 0,03 | 0,34 | 7,32E-01 |
| LAP3 | 2142,52 | 0,04 | 0,11 | 0,34 | 7,32E-01 |
| GLUL | 29214,98 | 0,02 | 0,07 | 0,34 | 7,32E-01 |
| XRRA1 | 757,40 | 0,04 | 0,11 | 0,34 | 7,32E-01 |
| BCL6 | 6318,43 | 0,03 | 0,09 | 0,34 | 7,32E-01 |
| BEND4 | 146,35 | 0,04 | 0,11 | 0,34 | 7,32E-01 |
| CFD | 3946,45 | 0,05 | 0,14 | 0,34 | 7,33E-01 |
| KLC1 | 3284,23 | 0,02 | 0,05 | 0,34 | 7,33E-01 |
| ST7L | 466,38 | 0,01 | 0,03 | 0,34 | 7,34E-01 |
| ELAC1 | 201,50 | -0,01 | 0,04 | -0,34 | 7,34E-01 |
| DHX38 | 2165,26 | -0,01 | 0,03 | -0,34 | 7,34E-01 |
| PNPLA4 | 96,37 | -0,02 | 0,05 | -0,34 | 7,34E-01 |
| MRPL36 | 153,13 | -0,02 | 0,06 | -0,34 | 7,35E-01 |
| CTBP2 | 2301,50 | -0,02 | 0,05 | -0,34 | 7,36E-01 |
| NOP10 | 1139,06 | -0,02 | 0,06 | -0,34 | 7,36E-01 |
| NCOR1 | 6228,11 | -0,01 | 0,04 | -0,34 | 7,36E-01 |
| TMEM140 | 4697,92 | -0,02 | 0,07 | -0,34 | 7,36E-01 |
| ALS2 | 1015,37 | 0,01 | 0,04 | 0,34 | 7,37E-01 |
| B3GALT6 | 511,26 | -0,01 | 0,03 | -0,34 | 7,37E-01 |
| S100B | 63,10 | -0,08 | 0,23 | -0,33 | 7,38E-01 |
| HARS2 | 1097,25 | -0,01 | 0,03 | -0,33 | 7,38E-01 |
| CSRNP1 | 985,47 | 0,02 | 0,07 | 0,33 | 7,38E-01 |
| RAD51C | 272,24 | -0,02 | 0,06 | -0,33 | 7,39E-01 |
| CLEC10A | 184,64 | 0,03 | 0,08 | 0,33 | 7,40E-01 |

| | | | | | |
|---|---|---|---|---|---|
| COQ3 | 70,92 | -0,02 | 0,05 | -0,33 | 7,41E-01 |
| SNX24 | 170,31 | 0,01 | 0,04 | 0,33 | 7,41E-01 |
| PPM1A | 9043,77 | 0,02 | 0,06 | 0,33 | 7,42E-01 |
| RIPOR2 | 27620,80 | 0,02 | 0,05 | 0,33 | 7,42E-01 |
| AGBL3 | 207,36 | 0,02 | 0,07 | 0,33 | 7,42E-01 |
| MICOS13 | 259,31 | -0,02 | 0,05 | -0,33 | 7,42E-01 |
| REXO2 | 721,68 | 0,02 | 0,06 | 0,33 | 7,43E-01 |
| CENPU | 77,76 | -0,02 | 0,07 | -0,33 | 7,43E-01 |
| SORL1 | 78617,78 | -0,02 | 0,07 | -0,33 | 7,44E-01 |
| KDM5B | 2417,36 | -0,01 | 0,04 | -0,33 | 7,44E-01 |
| AMIGO2 | 242,03 | -0,01 | 0,05 | -0,33 | 7,44E-01 |
| ZCCHC2 | 2833,52 | -0,03 | 0,09 | -0,33 | 7,44E-01 |
| RBM34 | 94,13 | 0,01 | 0,04 | 0,33 | 7,44E-01 |
| HMBOX1 | 1933,42 | 0,01 | 0,04 | 0,33 | 7,44E-01 |
| STAT3 | 7354,66 | 0,02 | 0,06 | 0,33 | 7,45E-01 |
| ZNF589 | 1008,56 | -0,01 | 0,04 | -0,33 | 7,45E-01 |
| SYNE1 | 19858,25 | -0,02 | 0,06 | -0,33 | 7,45E-01 |
| RCC1L | 535,90 | -0,01 | 0,03 | -0,33 | 7,45E-01 |
| FARS2 | 677,20 | 0,01 | 0,04 | 0,33 | 7,45E-01 |
| SPG21 | 2383,71 | -0,01 | 0,03 | -0,32 | 7,45E-01 |
| SUPT3H | 287,92 | -0,02 | 0,06 | -0,32 | 7,46E-01 |
| ACOT4 | 36,42 | 0,03 | 0,10 | 0,32 | 7,46E-01 |
| ALDH1B1 | 125,32 | 0,02 | 0,05 | 0,32 | 7,46E-01 |
| FCMR | 1238,58 | 0,02 | 0,07 | 0,32 | 7,47E-01 |
| GANC | 1713,46 | -0,01 | 0,03 | -0,32 | 7,47E-01 |
| TBL2 | 550,50 | -0,01 | 0,02 | -0,32 | 7,47E-01 |
| CDKL5 | 946,92 | -0,02 | 0,05 | -0,32 | 7,47E-01 |
| ERLIN2 | 905,94 | 0,01 | 0,02 | 0,32 | 7,47E-01 |
| SLITRK4 | 154,42 | -0,03 | 0,08 | -0,32 | 7,47E-01 |
| PAWR | 306,51 | -0,03 | 0,09 | -0,32 | 7,47E-01 |
| CREB3 | 271,62 | -0,01 | 0,03 | -0,32 | 7,47E-01 |
| ZNF280C | 220,22 | -0,01 | 0,04 | -0,32 | 7,48E-01 |
| TDRD15 | 32,86 | 0,06 | 0,18 | 0,32 | 7,48E-01 |
| BNIP3L | 36795,47 | 0,03 | 0,10 | 0,32 | 7,48E-01 |
| MRTO4 | 340,94 | -0,01 | 0,05 | -0,32 | 7,48E-01 |
| GOLGA6L9 | 131,34 | 0,03 | 0,08 | 0,32 | 7,49E-01 |
| FPR3 | 60,87 | -0,03 | 0,10 | -0,32 | 7,49E-01 |
| TRPC4AP | 2959,01 | 0,01 | 0,04 | 0,32 | 7,49E-01 |
| UBE2B | 2559,18 | 0,02 | 0,05 | 0,32 | 7,50E-01 |
| PLEKHG1 | 469,50 | -0,03 | 0,09 | -0,32 | 7,50E-01 |
| TKTL1 | 119,58 | -0,04 | 0,11 | -0,32 | 7,50E-01 |
| MEF2A | 2188,04 | 0,01 | 0,04 | 0,32 | 7,51E-01 |
| CEP76 | 248,84 | -0,01 | 0,04 | -0,32 | 7,51E-01 |
| ERP27 | 488,31 | 0,02 | 0,05 | 0,32 | 7,51E-01 |
| MMRN1 | 356,70 | 0,04 | 0,12 | 0,32 | 7,51E-01 |
| HERC5 | 1716,36 | -0,05 | 0,16 | -0,32 | 7,52E-01 |

| | | | | | |
|---|---|---|---|---|---|
| OTUD7B | 593,81 | 0,01 | 0,04 | 0,32 | 7,52E-01 |
| NELFB | 939,67 | 0,01 | 0,03 | 0,32 | 7,52E-01 |
| ATIC | 425,68 | 0,01 | 0,04 | 0,32 | 7,52E-01 |
| SLC25A28 | 637,59 | 0,01 | 0,03 | 0,32 | 7,52E-01 |
| MATR3 | 1813,01 | -0,01 | 0,04 | -0,32 | 7,52E-01 |
| RABIF | 458,54 | -0,01 | 0,03 | -0,31 | 7,53E-01 |
| IFNLR1 | 153,96 | 0,03 | 0,09 | 0,31 | 7,53E-01 |
| NOG | 207,35 | -0,04 | 0,12 | -0,31 | 7,53E-01 |
| NDUFV1 | 1011,65 | 0,01 | 0,03 | 0,31 | 7,53E-01 |
| GRK3 | 2970,40 | 0,01 | 0,05 | 0,31 | 7,54E-01 |
| MTX1 | 1194,08 | -0,02 | 0,06 | -0,31 | 7,54E-01 |
| REPS2 | 3289,12 | -0,02 | 0,08 | -0,31 | 7,55E-01 |
| FSD1L | 318,32 | 0,02 | 0,07 | 0,31 | 7,55E-01 |
| SART1 | 1478,75 | 0,01 | 0,05 | 0,31 | 7,56E-01 |
| ARHGEF7 | 2160,58 | 0,01 | 0,03 | 0,31 | 7,56E-01 |
| SSR4 | 823,00 | 0,01 | 0,05 | 0,31 | 7,57E-01 |
| WNT16 | 51,88 | -0,05 | 0,16 | -0,31 | 7,58E-01 |
| CHMP2A | 1316,31 | -0,02 | 0,05 | -0,31 | 7,58E-01 |
| PAICS | 383,00 | -0,01 | 0,04 | -0,31 | 7,58E-01 |
| PARP14 | 16351,30 | -0,03 | 0,09 | -0,31 | 7,58E-01 |
| FCAR | 1166,79 | 0,03 | 0,08 | 0,31 | 7,59E-01 |
| BICD2 | 3182,35 | 0,01 | 0,04 | 0,31 | 7,59E-01 |
| ANKLE2 | 2940,54 | -0,01 | 0,03 | -0,31 | 7,59E-01 |
| RGL4 | 662,67 | 0,02 | 0,07 | 0,31 | 7,59E-01 |
| EPHB1 | 3833,06 | -0,02 | 0,07 | -0,31 | 7,60E-01 |
| INO80B | 43,70 | -0,02 | 0,06 | -0,31 | 7,60E-01 |
| BEX5 | 31,33 | -0,03 | 0,10 | -0,30 | 7,60E-01 |
| CHST7 | 591,94 | 0,02 | 0,06 | 0,30 | 7,60E-01 |
| ZDHHC7 | 3816,64 | -0,01 | 0,04 | -0,30 | 7,61E-01 |
| FAM24B | 130,16 | -0,02 | 0,06 | -0,30 | 7,61E-01 |
| ITGB2 | 20517,76 | 0,02 | 0,06 | 0,30 | 7,61E-01 |
| C1QTNF6 | 412,70 | 0,01 | 0,04 | 0,30 | 7,61E-01 |
| KCNQ5 | 203,78 | -0,02 | 0,07 | -0,30 | 7,61E-01 |
| PPAN | 84,82 | -0,01 | 0,05 | -0,30 | 7,61E-01 |
| HDC | 296,41 | 0,05 | 0,18 | 0,30 | 7,62E-01 |
| LETM2 | 242,20 | 0,02 | 0,07 | 0,30 | 7,62E-01 |
| CADM1 | 104,17 | 0,04 | 0,14 | 0,30 | 7,62E-01 |
| WHAMM | 960,61 | 0,01 | 0,03 | 0,30 | 7,62E-01 |
| NELL2 | 1943,44 | -0,02 | 0,08 | -0,30 | 7,63E-01 |
| CELF2 | 15811,02 | -0,01 | 0,04 | -0,30 | 7,63E-01 |
| SNF8 | 865,13 | 0,01 | 0,02 | 0,30 | 7,63E-01 |
| ENSG00000267179 | 27,47 | 0,02 | 0,06 | 0,30 | 7,63E-01 |
| SERPINB2 | 62,61 | 0,03 | 0,10 | 0,30 | 7,65E-01 |
| CNTNAP3 | 2606,61 | -0,04 | 0,13 | -0,30 | 7,65E-01 |
| CLASP1 | 3900,04 | -0,01 | 0,04 | -0,30 | 7,66E-01 |
| KRT23 | 2348,08 | -0,03 | 0,10 | -0,30 | 7,66E-01 |

| | | | | | |
|---|---|---|---|---|---|
| KIF22 | 658,24 | 0,01 | 0,04 | 0,30 | 7,66E-01 |
| LPXN | 1384,46 | -0,01 | 0,03 | -0,30 | 7,66E-01 |
| ATP6V1E2 | 199,21 | 0,01 | 0,05 | 0,30 | 7,66E-01 |
| RAB6A | 3034,65 | -0,01 | 0,05 | -0,30 | 7,67E-01 |
| COPRS | 87,59 | 0,01 | 0,05 | 0,30 | 7,68E-01 |
| INPP4B | 5026,97 | -0,02 | 0,07 | -0,29 | 7,68E-01 |
| SEC62 | 4090,98 | -0,02 | 0,07 | -0,29 | 7,69E-01 |
| TMEM154 | 20721,59 | -0,02 | 0,05 | -0,29 | 7,69E-01 |
| NFKBIZ | 3946,71 | -0,01 | 0,04 | -0,29 | 7,69E-01 |
| TCTN3 | 232,05 | 0,01 | 0,04 | 0,29 | 7,69E-01 |
| TRIM22 | 11011,94 | -0,02 | 0,08 | -0,29 | 7,70E-01 |
| ZNF606 | 585,83 | 0,01 | 0,03 | 0,29 | 7,70E-01 |
| PSME3 | 1567,48 | 0,02 | 0,05 | 0,29 | 7,70E-01 |
| MIEF2 | 360,18 | 0,01 | 0,03 | 0,29 | 7,71E-01 |
| ISG20L2 | 1003,48 | 0,01 | 0,03 | 0,29 | 7,71E-01 |
| MSANTD3 | 201,91 | -0,01 | 0,05 | -0,29 | 7,71E-01 |
| TWSG1 | 307,67 | 0,02 | 0,05 | 0,29 | 7,71E-01 |
| VAV1 | 4907,47 | -0,01 | 0,04 | -0,29 | 7,72E-01 |
| TIAM2 | 594,12 | 0,02 | 0,07 | 0,29 | 7,72E-01 |
| TADA2A | 597,64 | 0,01 | 0,03 | 0,29 | 7,72E-01 |
| SLAMF6 | 1104,56 | -0,02 | 0,05 | -0,29 | 7,72E-01 |
| B3GALT2 | 69,71 | -0,04 | 0,13 | -0,29 | 7,72E-01 |
| CORO1C | 2236,10 | 0,02 | 0,05 | 0,29 | 7,73E-01 |
| TAGAP | 5419,92 | 0,01 | 0,05 | 0,29 | 7,73E-01 |
| CCDC30 | 416,53 | 0,02 | 0,06 | 0,29 | 7,73E-01 |
| TNFRSF8 | 295,88 | 0,02 | 0,06 | 0,29 | 7,73E-01 |
| HSPA1B | 331,59 | -0,02 | 0,07 | -0,29 | 7,73E-01 |
| UBE2F | 611,81 | 0,01 | 0,03 | 0,29 | 7,73E-01 |
| EFCAB13 | 352,19 | -0,02 | 0,08 | -0,29 | 7,74E-01 |
| YEATS2 | 1703,21 | 0,01 | 0,03 | 0,29 | 7,74E-01 |
| OMG | 41,25 | 0,02 | 0,09 | 0,29 | 7,74E-01 |
| ICAM2 | 1094,87 | 0,01 | 0,03 | 0,29 | 7,74E-01 |
| CD40LG | 319,64 | -0,02 | 0,06 | -0,29 | 7,75E-01 |
| KLF7 | 2932,53 | 0,02 | 0,06 | 0,29 | 7,75E-01 |
| TCEANC2 | 1069,50 | -0,01 | 0,02 | -0,29 | 7,76E-01 |
| FAM118A | 1372,20 | 0,03 | 0,11 | 0,28 | 7,76E-01 |
| DSE | 2150,46 | -0,02 | 0,05 | -0,28 | 7,76E-01 |
| PRMT5 | 516,44 | -0,01 | 0,05 | -0,28 | 7,76E-01 |
| TFCP2 | 718,14 | 0,01 | 0,04 | 0,28 | 7,76E-01 |
| BTN2A2 | 509,22 | 0,01 | 0,05 | 0,28 | 7,76E-01 |
| MYO1E | 430,44 | 0,02 | 0,07 | 0,28 | 7,77E-01 |
| COQ8A | 1366,62 | 0,01 | 0,05 | 0,28 | 7,77E-01 |
| SAYSD1 | 418,90 | -0,01 | 0,04 | -0,28 | 7,77E-01 |
| C1orf35 | 373,27 | -0,01 | 0,03 | -0,28 | 7,77E-01 |
| CDH2 | 68,69 | -0,04 | 0,14 | -0,28 | 7,77E-01 |
| TMTC4 | 555,34 | 0,01 | 0,05 | 0,28 | 7,78E-01 |

| | | | | | |
|---|---|---|---|---|---|
| TMEM209 | 345,32 | 0,01 | 0,04 | 0,28 | 7,78E-01 |
| GPATCH1 | 374,31 | 0,01 | 0,03 | 0,28 | 7,78E-01 |
| PNRC1 | 5166,41 | 0,01 | 0,05 | 0,28 | 7,78E-01 |
| ARL10 | 540,45 | 0,01 | 0,05 | 0,28 | 7,78E-01 |
| RYBP | 3284,57 | -0,01 | 0,05 | -0,28 | 7,79E-01 |
| NAB2 | 223,91 | 0,01 | 0,04 | 0,28 | 7,79E-01 |
| QARS1 | 1912,96 | 0,01 | 0,03 | 0,28 | 7,79E-01 |
| FUCA1 | 600,41 | 0,01 | 0,05 | 0,28 | 7,80E-01 |
| HDLBP | 4779,28 | 0,01 | 0,05 | 0,28 | 7,80E-01 |
| ZNF470 | 204,39 | -0,02 | 0,07 | -0,28 | 7,80E-01 |
| KMT2C | 13298,72 | -0,01 | 0,05 | -0,28 | 7,81E-01 |
| FDFT1 | 2048,26 | 0,01 | 0,03 | 0,28 | 7,81E-01 |
| CTNNA1 | 2561,76 | -0,02 | 0,06 | -0,28 | 7,81E-01 |
| C5AR2 | 4751,35 | 0,02 | 0,06 | 0,28 | 7,81E-01 |
| P4HB | 2278,48 | 0,01 | 0,04 | 0,28 | 7,81E-01 |
| CCDC142 | 1121,18 | 0,01 | 0,03 | 0,28 | 7,82E-01 |
| PRDM10 | 756,87 | -0,01 | 0,03 | -0,28 | 7,82E-01 |
| ACP3 | 1221,70 | 0,02 | 0,07 | 0,28 | 7,82E-01 |
| ZNF585B | 325,65 | -0,01 | 0,04 | -0,28 | 7,82E-01 |
| FLOT1 | 2425,60 | 0,02 | 0,06 | 0,28 | 7,82E-01 |
| OSBPL1A | 410,64 | 0,02 | 0,06 | 0,28 | 7,82E-01 |
| UQCC6 | 185,29 | -0,01 | 0,05 | -0,28 | 7,83E-01 |
| FOSL2 | 4855,39 | -0,02 | 0,08 | -0,28 | 7,83E-01 |
| HAUS4 | 799,97 | -0,02 | 0,07 | -0,27 | 7,83E-01 |
| TYW1B | 296,65 | -0,01 | 0,04 | -0,27 | 7,84E-01 |
| PAG1 | 8236,56 | -0,01 | 0,04 | -0,27 | 7,84E-01 |
| PDZD4 | 503,44 | 0,02 | 0,08 | 0,27 | 7,84E-01 |
| MACO1 | 741,29 | -0,01 | 0,03 | -0,27 | 7,84E-01 |
| H2BC12 | 2266,80 | 0,01 | 0,05 | 0,27 | 7,85E-01 |
| MAK | 732,64 | 0,02 | 0,09 | 0,27 | 7,85E-01 |
| TMED8 | 1876,42 | -0,01 | 0,04 | -0,27 | 7,85E-01 |
| BAK1 | 656,66 | 0,01 | 0,05 | 0,27 | 7,86E-01 |
| SAAL1 | 306,55 | 0,01 | 0,04 | 0,27 | 7,86E-01 |
| ERN1 | 3349,30 | -0,02 | 0,07 | -0,27 | 7,87E-01 |
| LPIN1 | 1927,01 | 0,01 | 0,03 | 0,27 | 7,87E-01 |
| MAN1A1 | 3963,47 | -0,02 | 0,06 | -0,27 | 7,87E-01 |
| UCK2 | 382,23 | -0,01 | 0,05 | -0,27 | 7,88E-01 |
| DET1 | 212,61 | 0,01 | 0,05 | 0,27 | 7,88E-01 |
| XKR9 | 39,72 | 0,03 | 0,13 | 0,27 | 7,88E-01 |
| SEC24D | 1142,06 | 0,01 | 0,04 | 0,27 | 7,89E-01 |
| IGFBP3 | 161,82 | 0,03 | 0,10 | 0,27 | 7,89E-01 |
| ZNF80 | 55,61 | -0,04 | 0,13 | -0,27 | 7,89E-01 |
| RPLP0 | 12181,60 | -0,02 | 0,07 | -0,27 | 7,89E-01 |
| ZMAT3 | 1484,30 | -0,01 | 0,04 | -0,27 | 7,89E-01 |
| ASB3 | 359,67 | -0,01 | 0,03 | -0,27 | 7,90E-01 |
| CIAO2B | 540,80 | 0,01 | 0,04 | 0,27 | 7,90E-01 |

| | | | | | |
|---|---|---|---|---|---|
| ENSG00000271254 | 281,30 | 0,02 | 0,08 | 0,27 | 7,90E-01 |
| MRPL52 | 369,03 | -0,01 | 0,03 | -0,27 | 7,90E-01 |
| ALDH2 | 490,24 | 0,02 | 0,07 | 0,27 | 7,91E-01 |
| UBXN2A | 486,08 | -0,01 | 0,03 | -0,27 | 7,91E-01 |
| RUNX1 | 3009,65 | 0,01 | 0,04 | 0,26 | 7,91E-01 |
| NUMB | 4673,71 | 0,02 | 0,06 | 0,26 | 7,92E-01 |
| SNN | 3304,93 | 0,01 | 0,05 | 0,26 | 7,92E-01 |
| PTPRJ | 10865,33 | 0,02 | 0,06 | 0,26 | 7,92E-01 |
| ZNF208 | 92,06 | -0,03 | 0,12 | -0,26 | 7,92E-01 |
| ANKRD36B | 1313,23 | -0,02 | 0,07 | -0,26 | 7,92E-01 |
| MOB3C | 1497,74 | 0,01 | 0,04 | 0,26 | 7,92E-01 |
| RPL39L | 34,83 | 0,02 | 0,09 | 0,26 | 7,92E-01 |
| ZNF705EP | 24,12 | 0,02 | 0,09 | 0,26 | 7,92E-01 |
| NBPF19 | 3464,52 | 0,01 | 0,06 | 0,26 | 7,92E-01 |
| CXCR1 | 8779,52 | -0,02 | 0,07 | -0,26 | 7,92E-01 |
| EXOSC5 | 188,37 | 0,01 | 0,04 | 0,26 | 7,93E-01 |
| TMSB4Y | 73,27 | 0,02 | 0,06 | 0,26 | 7,93E-01 |
| BRMS1L | 106,79 | -0,02 | 0,06 | -0,26 | 7,93E-01 |
| ERI2 | 184,59 | 0,01 | 0,05 | 0,26 | 7,94E-01 |
| TMEM97 | 69,66 | -0,02 | 0,07 | -0,26 | 7,94E-01 |
| H2AC13 | 866,71 | 0,02 | 0,09 | 0,26 | 7,94E-01 |
| LSP1 | 9614,92 | -0,02 | 0,07 | -0,26 | 7,94E-01 |
| DEF6 | 3849,87 | -0,01 | 0,03 | -0,26 | 7,94E-01 |
| ENSG00000275895 | 242,91 | -0,06 | 0,24 | -0,26 | 7,94E-01 |
| HLX | 567,46 | 0,02 | 0,07 | 0,26 | 7,94E-01 |
| NKX3-1 | 238,40 | -0,06 | 0,23 | -0,26 | 7,95E-01 |
| ARSG | 1789,46 | 0,01 | 0,05 | 0,26 | 7,95E-01 |
| MPPE1 | 4197,38 | 0,01 | 0,05 | 0,26 | 7,95E-01 |
| GIT2 | 5209,01 | 0,01 | 0,05 | 0,26 | 7,95E-01 |
| AFG1L | 211,41 | -0,01 | 0,04 | -0,26 | 7,95E-01 |
| CMAS | 522,05 | 0,02 | 0,06 | 0,26 | 7,95E-01 |
| NBPF12 | 1253,61 | 0,01 | 0,05 | 0,26 | 7,96E-01 |
| RAD51B | 709,03 | -0,01 | 0,04 | -0,26 | 7,96E-01 |
| THAP11 | 556,04 | 0,01 | 0,03 | 0,26 | 7,97E-01 |
| PPP1R14B | 188,83 | -0,01 | 0,05 | -0,26 | 7,97E-01 |
| TMEM102 | 288,39 | 0,01 | 0,04 | 0,26 | 7,97E-01 |
| RNF38 | 1940,60 | 0,01 | 0,03 | 0,26 | 7,98E-01 |
| C5orf34 | 75,28 | -0,02 | 0,06 | -0,26 | 7,98E-01 |
| CASP9 | 442,77 | 0,01 | 0,04 | 0,26 | 7,98E-01 |
| UBP1 | 1939,06 | 0,01 | 0,03 | 0,25 | 7,99E-01 |
| TMEM41A | 703,90 | -0,01 | 0,03 | -0,25 | 7,99E-01 |
| ATP6V0A2 | 1611,40 | 0,01 | 0,03 | 0,25 | 7,99E-01 |
| CEACAM4 | 1144,91 | 0,02 | 0,08 | 0,25 | 7,99E-01 |
| CCDC141 | 817,76 | 0,02 | 0,08 | 0,25 | 7,99E-01 |
| TRIM56 | 5166,15 | -0,01 | 0,04 | -0,25 | 8,00E-01 |
| PADI4 | 1785,03 | 0,03 | 0,10 | 0,25 | 8,01E-01 |

| | | | | | |
|---|---|---|---|---|---|
| BAP1 | 1306,98 | 0,01 | 0,03 | 0,25 | 8,01E-01 |
| CYTH1 | 10241,54 | 0,01 | 0,04 | 0,25 | 8,02E-01 |
| LPCAT2 | 4323,51 | 0,02 | 0,07 | 0,25 | 8,02E-01 |
| FBXO32 | 434,62 | 0,01 | 0,05 | 0,25 | 8,02E-01 |
| B4GALT5 | 3689,73 | -0,02 | 0,07 | -0,25 | 8,02E-01 |
| TOX | 220,43 | -0,02 | 0,06 | -0,25 | 8,02E-01 |
| BNC2 | 223,91 | 0,03 | 0,11 | 0,25 | 8,02E-01 |
| ZCCHC18 | 110,56 | -0,02 | 0,07 | -0,25 | 8,02E-01 |
| NFATC2 | 1455,46 | 0,01 | 0,05 | 0,25 | 8,02E-01 |
| PPP1R10 | 2122,10 | 0,01 | 0,04 | 0,25 | 8,03E-01 |
| CACNB4 | 692,67 | 0,03 | 0,11 | 0,25 | 8,03E-01 |
| ZNF697 | 330,42 | 0,02 | 0,06 | 0,25 | 8,03E-01 |
| TBCB | 944,30 | -0,01 | 0,03 | -0,25 | 8,04E-01 |
| MRPS34 | 434,01 | 0,01 | 0,04 | 0,25 | 8,04E-01 |
| PGM2L1 | 608,84 | 0,01 | 0,06 | 0,25 | 8,04E-01 |
| ZNF410 | 408,88 | 0,01 | 0,02 | 0,25 | 8,04E-01 |
| DOCK8 | 21800,54 | 0,01 | 0,05 | 0,25 | 8,04E-01 |
| LAPTM5 | 8311,20 | 0,02 | 0,06 | 0,25 | 8,05E-01 |
| PTTG2 | 175,01 | -0,02 | 0,08 | -0,25 | 8,05E-01 |
| TM6SF1 | 1433,37 | -0,01 | 0,06 | -0,25 | 8,05E-01 |
| PLOD1 | 645,91 | 0,02 | 0,07 | 0,25 | 8,05E-01 |
| ITK | 6311,82 | -0,01 | 0,05 | -0,25 | 8,05E-01 |
| NTPCR | 726,64 | -0,01 | 0,05 | -0,25 | 8,05E-01 |
| ARHGAP35 | 1546,09 | 0,01 | 0,04 | 0,25 | 8,05E-01 |
| NFE2L1 | 1662,47 | 0,01 | 0,03 | 0,25 | 8,06E-01 |
| RDH10 | 189,72 | 0,01 | 0,05 | 0,24 | 8,07E-01 |
| C2orf68 | 1886,06 | 0,01 | 0,03 | 0,24 | 8,07E-01 |
| ATXN1 | 5596,17 | -0,01 | 0,05 | -0,24 | 8,07E-01 |
| FRAT1 | 2563,69 | 0,02 | 0,06 | 0,24 | 8,07E-01 |
| GCLM | 683,25 | 0,01 | 0,05 | 0,24 | 8,08E-01 |
| PSMD8 | 592,95 | 0,01 | 0,03 | 0,24 | 8,07E-01 |
| CD84 | 3285,43 | 0,01 | 0,04 | 0,24 | 8,08E-01 |
| LRRC69 | 206,74 | 0,01 | 0,04 | 0,24 | 8,08E-01 |
| H2BC6 | 632,08 | 0,02 | 0,07 | 0,24 | 8,09E-01 |
| WDR54 | 138,98 | 0,01 | 0,05 | 0,24 | 8,09E-01 |
| CERK | 4154,26 | 0,01 | 0,03 | 0,24 | 8,09E-01 |
| RNASEH1 | 365,07 | 0,01 | 0,02 | 0,24 | 8,09E-01 |
| LRRC47 | 942,50 | 0,01 | 0,02 | 0,24 | 8,09E-01 |
| PUS7 | 165,52 | -0,01 | 0,05 | -0,24 | 8,09E-01 |
| ENSG00000213204 | 117,88 | -0,01 | 0,05 | -0,24 | 8,10E-01 |
| RTP4 | 410,03 | 0,03 | 0,11 | 0,24 | 8,10E-01 |
| NMUR1 | 220,18 | 0,02 | 0,10 | 0,24 | 8,10E-01 |
| DCAF6 | 1943,06 | -0,01 | 0,04 | -0,24 | 8,10E-01 |
| ENSG00000284292 | 50,04 | -0,01 | 0,05 | -0,24 | 8,10E-01 |
| S100A6 | 2707,60 | 0,01 | 0,05 | 0,24 | 8,11E-01 |
| POGLUT3 | 227,40 | -0,01 | 0,05 | -0,24 | 8,12E-01 |

| | | | | | |
|---|---|---|---|---|---|
| XCL1 | 50,92 | 0,03 | 0,12 | 0,24 | 8,13E-01 |
| GFI1 | 329,38 | 0,02 | 0,08 | 0,24 | 8,13E-01 |
| ARL6IP1 | 2850,81 | -0,01 | 0,06 | -0,24 | 8,13E-01 |
| BTD | 525,63 | -0,01 | 0,03 | -0,24 | 8,13E-01 |
| KCTD3 | 234,35 | -0,01 | 0,05 | -0,24 | 8,13E-01 |
| H2BC5 | 1996,03 | -0,01 | 0,06 | -0,24 | 8,13E-01 |
| MINPP1 | 279,35 | -0,01 | 0,06 | -0,24 | 8,13E-01 |
| UGGT2 | 305,71 | 0,02 | 0,07 | 0,23 | 8,14E-01 |
| FNBP1 | 3753,07 | -0,01 | 0,04 | -0,23 | 8,15E-01 |
| XAF1 | 3275,18 | 0,03 | 0,12 | 0,23 | 8,15E-01 |
| INPP5B | 1670,64 | 0,01 | 0,04 | 0,23 | 8,15E-01 |
| RPS5 | 6351,79 | -0,02 | 0,07 | -0,23 | 8,15E-01 |
| MRRF | 548,22 | -0,01 | 0,03 | -0,23 | 8,15E-01 |
| PSMG4 | 1246,10 | -0,01 | 0,04 | -0,23 | 8,15E-01 |
| RER1 | 2859,94 | -0,01 | 0,03 | -0,23 | 8,15E-01 |
| U2AF1 | 393,76 | -0,04 | 0,19 | -0,23 | 8,15E-01 |
| RIC8A | 1696,77 | -0,01 | 0,03 | -0,23 | 8,16E-01 |
| NMT1 | 1506,83 | 0,01 | 0,03 | 0,23 | 8,16E-01 |
| DUSP1 | 10310,37 | 0,02 | 0,07 | 0,23 | 8,16E-01 |
| ACOT9 | 1479,21 | -0,01 | 0,05 | -0,23 | 8,16E-01 |
| H2BC18 | 2866,35 | -0,01 | 0,06 | -0,23 | 8,17E-01 |
| EXOSC10 | 1090,09 | -0,01 | 0,03 | -0,23 | 8,17E-01 |
| SURF6 | 927,79 | -0,01 | 0,03 | -0,23 | 8,17E-01 |
| POLR1C | 372,50 | -0,01 | 0,03 | -0,23 | 8,17E-01 |
| LYRM1 | 650,37 | 0,01 | 0,03 | 0,23 | 8,18E-01 |
| CRADD | 450,54 | 0,01 | 0,04 | 0,23 | 8,19E-01 |
| CYSLTR2 | 709,90 | 0,02 | 0,07 | 0,23 | 8,19E-01 |
| GRB2 | 8924,91 | -0,01 | 0,04 | -0,23 | 8,19E-01 |
| IFI44 | 1575,09 | 0,04 | 0,18 | 0,23 | 8,20E-01 |
| OAS2 | 2374,01 | 0,03 | 0,11 | 0,23 | 8,20E-01 |
| RFLNB | 1878,05 | 0,02 | 0,08 | 0,23 | 8,21E-01 |
| GDF11 | 415,29 | -0,01 | 0,04 | -0,23 | 8,21E-01 |
| DUSP5 | 363,81 | -0,01 | 0,06 | -0,23 | 8,21E-01 |
| TIPIN | 86,68 | 0,01 | 0,06 | 0,23 | 8,21E-01 |
| AP3M2 | 887,15 | -0,01 | 0,04 | -0,23 | 8,21E-01 |
| SPATA24 | 30,17 | 0,01 | 0,06 | 0,23 | 8,22E-01 |
| PHF11 | 1731,06 | -0,01 | 0,05 | -0,23 | 8,22E-01 |
| FRY | 9468,09 | 0,01 | 0,06 | 0,22 | 8,22E-01 |
| CARF | 694,13 | -0,01 | 0,04 | -0,22 | 8,22E-01 |
| FBXL4 | 2319,02 | -0,01 | 0,04 | -0,22 | 8,22E-01 |
| TRIM27 | 2901,07 | 0,01 | 0,04 | 0,22 | 8,23E-01 |
| IFI6 | 2663,14 | 0,04 | 0,17 | 0,22 | 8,23E-01 |
| SGCB | 148,78 | 0,01 | 0,06 | 0,22 | 8,24E-01 |
| PAF1 | 364,56 | 0,01 | 0,04 | 0,22 | 8,24E-01 |
| C5orf63 | 411,61 | -0,01 | 0,05 | -0,22 | 8,24E-01 |
| SLC25A16 | 926,29 | 0,00 | 0,02 | -0,22 | 8,24E-01 |

| | | | | | |
|---|---|---|---|---|---|
| DHRS7B | 708,90 | 0,01 | 0,03 | 0,22 | 8,25E-01 |
| MBD5 | 3693,56 | -0,01 | 0,05 | -0,22 | 8,25E-01 |
| IL4R | 6106,69 | 0,01 | 0,06 | 0,22 | 8,25E-01 |
| PLAUR | 1536,58 | 0,02 | 0,07 | 0,22 | 8,25E-01 |
| SEPTIN11 | 1100,22 | -0,01 | 0,04 | -0,22 | 8,25E-01 |
| SCIMP | 863,08 | -0,01 | 0,06 | -0,22 | 8,25E-01 |
| PTPN1 | 1984,61 | 0,01 | 0,03 | 0,22 | 8,26E-01 |
| ODC1 | 5206,70 | 0,02 | 0,09 | 0,22 | 8,26E-01 |
| ZC3HAV1 | 7321,86 | 0,01 | 0,05 | 0,22 | 8,26E-01 |
| CNOT9 | 1322,01 | 0,01 | 0,03 | 0,22 | 8,26E-01 |
| ANKHD1-EIF4EBP3 | 279,05 | -0,01 | 0,05 | -0,22 | 8,27E-01 |
| TAS2R4 | 213,76 | 0,01 | 0,06 | 0,22 | 8,28E-01 |
| ARL15 | 1067,11 | -0,01 | 0,05 | -0,22 | 8,28E-01 |
| MRPL48 | 213,08 | -0,01 | 0,05 | -0,22 | 8,28E-01 |
| DDX43 | 55,82 | 0,03 | 0,15 | 0,22 | 8,29E-01 |
| LIN9 | 131,30 | 0,01 | 0,07 | 0,21 | 8,30E-01 |
| MAP10 | 110,52 | -0,02 | 0,07 | -0,21 | 8,30E-01 |
| TEX264 | 1074,00 | 0,01 | 0,02 | 0,21 | 8,30E-01 |
| ALKBH4 | 182,67 | 0,01 | 0,03 | 0,21 | 8,31E-01 |
| CUZD1 | 137,00 | 0,01 | 0,06 | 0,21 | 8,31E-01 |
| MANBA | 4300,69 | -0,01 | 0,04 | -0,21 | 8,31E-01 |
| TIMM23B-AGAP6 | 68,46 | 0,02 | 0,07 | 0,21 | 8,31E-01 |
| TAFA2 | 1096,08 | 0,01 | 0,07 | 0,21 | 8,32E-01 |
| HAL | 2813,13 | -0,02 | 0,07 | -0,21 | 8,32E-01 |
| ARF4 | 1173,66 | -0,01 | 0,03 | -0,21 | 8,32E-01 |
| DCTN4 | 2111,30 | -0,01 | 0,03 | -0,21 | 8,32E-01 |
| DDAH2 | 719,20 | -0,01 | 0,05 | -0,21 | 8,33E-01 |
| MFSD14B | 2597,47 | 0,01 | 0,05 | 0,21 | 8,33E-01 |
| GOT2 | 395,48 | 0,01 | 0,04 | 0,21 | 8,33E-01 |
| AGO1 | 2700,21 | 0,01 | 0,05 | 0,21 | 8,33E-01 |
| ENSG00000283761 | 134,99 | 0,01 | 0,04 | 0,21 | 8,33E-01 |
| TMEM106C | 296,98 | 0,01 | 0,04 | 0,21 | 8,33E-01 |
| TMEM182 | 150,99 | 0,01 | 0,06 | 0,21 | 8,34E-01 |
| ADAM20 | 117,88 | 0,02 | 0,08 | 0,21 | 8,34E-01 |
| NR3C2 | 906,30 | -0,01 | 0,06 | -0,21 | 8,34E-01 |
| KNOP1 | 543,32 | 0,01 | 0,04 | 0,21 | 8,34E-01 |
| PTRHD1 | 225,81 | 0,01 | 0,07 | 0,21 | 8,35E-01 |
| SETD5 | 3766,39 | 0,01 | 0,05 | 0,21 | 8,35E-01 |
| CUTA | 329,34 | 0,01 | 0,04 | 0,21 | 8,35E-01 |
| CCDC121 | 73,30 | -0,02 | 0,07 | -0,21 | 8,36E-01 |
| TBC1D1 | 4806,09 | -0,01 | 0,05 | -0,21 | 8,36E-01 |
| ZNF185 | 1200,29 | 0,02 | 0,07 | 0,21 | 8,36E-01 |
| FAM120B | 2282,34 | -0,01 | 0,04 | -0,21 | 8,36E-01 |
| ZNF814 | 706,79 | 0,01 | 0,05 | 0,21 | 8,36E-01 |
| EPG5 | 4296,24 | 0,01 | 0,04 | 0,21 | 8,37E-01 |
| ANKRD42 | 401,08 | -0,01 | 0,03 | -0,20 | 8,38E-01 |

| | | | | | |
|---|---|---|---|---|---|
| RAB35 | 1127,70 | -0,01 | 0,03 | -0,21 | 8,38E-01 |
| CLIC4 | 977,18 | 0,01 | 0,06 | 0,20 | 8,38E-01 |
| BIN3 | 1376,79 | 0,01 | 0,05 | 0,20 | 8,38E-01 |
| FLAD1 | 636,33 | 0,01 | 0,03 | 0,20 | 8,38E-01 |
| RAC1 | 2999,87 | -0,01 | 0,04 | -0,20 | 8,39E-01 |
| CAMKMT | 606,15 | -0,01 | 0,04 | -0,20 | 8,39E-01 |
| ABL2 | 1206,16 | 0,01 | 0,04 | 0,20 | 8,39E-01 |
| GSTCD | 249,08 | 0,01 | 0,04 | 0,20 | 8,39E-01 |
| SRGAP2B | 422,16 | 0,01 | 0,06 | 0,20 | 8,39E-01 |
| RCOR1 | 2322,74 | -0,01 | 0,06 | -0,20 | 8,40E-01 |
| DDX60L | 11548,16 | -0,02 | 0,09 | -0,20 | 8,40E-01 |
| SAMM50 | 672,23 | 0,00 | 0,02 | -0,20 | 8,40E-01 |
| PEF1 | 725,63 | 0,01 | 0,03 | 0,20 | 8,41E-01 |
| CASP6 | 304,34 | -0,01 | 0,04 | -0,20 | 8,42E-01 |
| ZNF391 | 44,33 | 0,02 | 0,10 | 0,20 | 8,42E-01 |
| GPR82 | 85,02 | 0,02 | 0,11 | 0,20 | 8,43E-01 |
| LIPT2 | 82,35 | -0,01 | 0,06 | -0,20 | 8,43E-01 |
| NCR3 | 124,31 | -0,02 | 0,08 | -0,20 | 8,43E-01 |
| CD79A | 177,21 | 0,02 | 0,12 | 0,20 | 8,43E-01 |
| GBP5 | 12558,74 | 0,02 | 0,12 | 0,20 | 8,44E-01 |
| ANAPC15 | 135,08 | 0,01 | 0,04 | 0,20 | 8,44E-01 |
| UBN1 | 4544,44 | -0,02 | 0,08 | -0,20 | 8,44E-01 |
| DPP4 | 1056,99 | 0,01 | 0,06 | 0,20 | 8,44E-01 |
| ADM | 1098,05 | -0,02 | 0,09 | -0,20 | 8,44E-01 |
| PCDHGA11 | 67,39 | 0,02 | 0,11 | 0,20 | 8,44E-01 |
| DROSHA | 959,45 | 0,01 | 0,04 | 0,20 | 8,45E-01 |
| BCKDHA | 32,47 | -0,01 | 0,05 | -0,20 | 8,45E-01 |
| ZBTB25 | 2455,82 | -0,01 | 0,04 | -0,20 | 8,45E-01 |
| PGD | 6712,98 | -0,01 | 0,06 | -0,20 | 8,45E-01 |
| ZNF445 | 2863,12 | 0,01 | 0,06 | 0,20 | 8,45E-01 |
| NFKB1 | 3410,81 | 0,01 | 0,03 | 0,20 | 8,45E-01 |
| BCL11B | 4219,47 | -0,01 | 0,05 | -0,19 | 8,46E-01 |
| RSPH3 | 594,49 | -0,01 | 0,06 | -0,19 | 8,46E-01 |
| ZNF284 | 86,96 | 0,01 | 0,04 | 0,19 | 8,46E-01 |
| TBP | 409,94 | 0,00 | 0,03 | -0,19 | 8,46E-01 |
| BLTP2 | 4790,01 | 0,01 | 0,04 | 0,19 | 8,46E-01 |
| NDUFB7 | 536,54 | -0,01 | 0,05 | -0,19 | 8,46E-01 |
| ATXN1L | 1523,67 | 0,01 | 0,04 | 0,19 | 8,47E-01 |
| PSENEN | 344,33 | 0,01 | 0,05 | 0,19 | 8,47E-01 |
| MAPDA | 184,68 | 0,01 | 0,05 | 0,19 | 8,47E-01 |
| STAT1 | 13318,39 | -0,02 | 0,08 | -0,19 | 8,47E-01 |
| GDPD1 | 103,93 | 0,01 | 0,05 | 0,19 | 8,48E-01 |
| CLEC12A | 4649,86 | 0,03 | 0,14 | 0,19 | 8,48E-01 |
| SNX29 | 2225,13 | 0,01 | 0,04 | 0,19 | 8,48E-01 |
| SBF2 | 4091,32 | 0,01 | 0,05 | 0,19 | 8,49E-01 |
| ZNF396 | 158,80 | -0,01 | 0,06 | -0,19 | 8,49E-01 |

| | | | | | |
|---|---|---|---|---|---|
| BFAR | 665,77 | 0,00 | 0,03 | 0,19 | 8,49E-01 |
| PTAFR | 7860,98 | -0,01 | 0,06 | -0,19 | 8,50E-01 |
| TBX21 | 699,76 | -0,02 | 0,09 | -0,19 | 8,50E-01 |
| CRIM1 | 312,64 | 0,01 | 0,06 | 0,19 | 8,51E-01 |
| PAQR7 | 220,95 | -0,01 | 0,06 | -0,19 | 8,51E-01 |
| ZNFX1 | 6307,96 | 0,01 | 0,05 | 0,19 | 8,51E-01 |
| CXCL6 | 31,13 | 0,02 | 0,11 | 0,19 | 8,51E-01 |
| GOSR2 | 2089,65 | -0,01 | 0,03 | -0,19 | 8,52E-01 |
| ZNF343 | 267,12 | -0,01 | 0,03 | -0,19 | 8,52E-01 |
| PLAGL1 | 816,17 | 0,01 | 0,06 | 0,19 | 8,53E-01 |
| FBXL17 | 647,96 | -0,01 | 0,03 | -0,18 | 8,53E-01 |
| TBL1X | 2943,66 | -0,01 | 0,07 | -0,18 | 8,53E-01 |
| CYP4V2 | 1233,00 | 0,01 | 0,05 | 0,18 | 8,54E-01 |
| PAK1 | 3469,81 | -0,01 | 0,06 | -0,18 | 8,54E-01 |
| UBTF | 1979,77 | 0,01 | 0,03 | 0,18 | 8,55E-01 |
| CLN5 | 757,83 | -0,01 | 0,04 | -0,18 | 8,55E-01 |
| HSD17B7 | 537,56 | 0,01 | 0,03 | 0,18 | 8,55E-01 |
| TCHP | 1170,73 | 0,00 | 0,03 | -0,18 | 8,55E-01 |
| SLC25A17 | 398,05 | -0,01 | 0,03 | -0,18 | 8,55E-01 |
| CES1 | 285,30 | -0,02 | 0,13 | -0,18 | 8,55E-01 |
| PCGF5 | 4798,76 | -0,01 | 0,06 | -0,18 | 8,56E-01 |
| TMEM147 | 277,29 | -0,01 | 0,04 | -0,18 | 8,56E-01 |
| MAN1B1 | 1334,44 | 0,01 | 0,03 | 0,18 | 8,56E-01 |
| LARP4B | 4157,35 | 0,01 | 0,04 | 0,18 | 8,56E-01 |
| ZRANB3 | 211,10 | -0,01 | 0,05 | -0,18 | 8,57E-01 |
| TRAPPC10 | 3217,25 | -0,01 | 0,04 | -0,18 | 8,58E-01 |
| PHACTR2 | 1708,18 | -0,01 | 0,04 | -0,18 | 8,58E-01 |
| OSBPL3 | 1316,66 | -0,01 | 0,04 | -0,18 | 8,58E-01 |
| PEX5 | 530,16 | 0,01 | 0,03 | 0,18 | 8,58E-01 |
| TAS2R3 | 137,69 | 0,01 | 0,07 | 0,18 | 8,59E-01 |
| TLR3 | 84,41 | 0,02 | 0,10 | 0,18 | 8,59E-01 |
| TRPM6 | 1467,35 | -0,01 | 0,08 | -0,18 | 8,59E-01 |
| RAB32 | 919,58 | -0,01 | 0,06 | -0,18 | 8,59E-01 |
| FOXN3 | 4728,72 | 0,01 | 0,05 | 0,18 | 8,59E-01 |
| CR2 | 164,97 | 0,02 | 0,10 | 0,18 | 8,59E-01 |
| PELO | 354,61 | 0,01 | 0,05 | 0,18 | 8,59E-01 |
| SLC6A6 | 15543,61 | 0,01 | 0,06 | 0,18 | 8,59E-01 |
| PGRMC1 | 1047,41 | 0,01 | 0,07 | 0,18 | 8,60E-01 |
| VPS51 | 2761,31 | 0,01 | 0,04 | 0,18 | 8,61E-01 |
| CTSL | 295,76 | -0,02 | 0,09 | -0,18 | 8,61E-01 |
| TNFSF14 | 2233,75 | -0,01 | 0,05 | -0,18 | 8,61E-01 |
| CINP | 577,34 | 0,00 | 0,03 | -0,18 | 8,61E-01 |
| CYC1 | 733,69 | -0,01 | 0,04 | -0,17 | 8,61E-01 |
| ZFP69B | 99,68 | 0,01 | 0,04 | 0,17 | 8,62E-01 |
| CFP | 4386,97 | -0,01 | 0,06 | -0,17 | 8,62E-01 |
| CASP10 | 2258,58 | 0,01 | 0,04 | 0,17 | 8,62E-01 |

| | | | | | |
|---|---|---|---|---|---|
| AGFG1 | 4585,46 | -0,01 | 0,04 | -0,17 | 8,62E-01 |
| SEC14L1 | 10455,13 | 0,01 | 0,08 | 0,17 | 8,62E-01 |
| SIL1 | 569,14 | 0,00 | 0,03 | -0,17 | 8,62E-01 |
| ASCC1 | 255,84 | 0,01 | 0,04 | 0,17 | 8,62E-01 |
| FAM227B | 145,09 | 0,01 | 0,08 | 0,17 | 8,63E-01 |
| ARL11 | 1874,41 | -0,01 | 0,05 | -0,17 | 8,63E-01 |
| SRD5A3 | 155,02 | 0,01 | 0,07 | 0,17 | 8,63E-01 |
| C8orf88 | 74,44 | 0,02 | 0,09 | 0,17 | 8,63E-01 |
| SETD9 | 112,19 | 0,01 | 0,08 | 0,17 | 8,64E-01 |
| C3orf18 | 310,35 | 0,01 | 0,04 | 0,17 | 8,64E-01 |
| TRNAU1AP | 645,77 | 0,00 | 0,02 | 0,17 | 8,64E-01 |
| PAM | 1773,74 | 0,02 | 0,09 | 0,17 | 8,64E-01 |
| ARHGAP11A | 190,79 | -0,01 | 0,08 | -0,17 | 8,64E-01 |
| WDPCP | 683,09 | 0,01 | 0,04 | 0,17 | 8,65E-01 |
| ZNF547 | 101,31 | -0,01 | 0,06 | -0,17 | 8,65E-01 |
| ASAH2B | 100,41 | 0,01 | 0,09 | 0,17 | 8,65E-01 |
| NDUFA2 | 738,21 | -0,01 | 0,06 | -0,17 | 8,65E-01 |
| TOMM34 | 224,13 | -0,01 | 0,04 | -0,17 | 8,65E-01 |
| TLR2 | 13056,61 | -0,01 | 0,07 | -0,17 | 8,65E-01 |
| ERGIC1 | 5642,35 | -0,01 | 0,06 | -0,17 | 8,65E-01 |
| STX10 | 1848,95 | -0,01 | 0,05 | -0,17 | 8,65E-01 |
| SIPA1L1 | 5783,16 | -0,01 | 0,07 | -0,17 | 8,65E-01 |
| FRMD4B | 729,09 | -0,01 | 0,05 | -0,17 | 8,66E-01 |
| ARL16 | 371,28 | 0,01 | 0,05 | 0,17 | 8,66E-01 |
| PTGS2 | 1376,96 | -0,01 | 0,07 | -0,17 | 8,67E-01 |
| PHEX | 88,68 | -0,02 | 0,11 | -0,17 | 8,67E-01 |
| MPLKIP | 643,37 | 0,00 | 0,03 | 0,17 | 8,67E-01 |
| TXLNB | 79,38 | 0,01 | 0,07 | 0,17 | 8,67E-01 |
| THBD | 902,64 | 0,02 | 0,09 | 0,17 | 8,67E-01 |
| CTPS2 | 295,26 | -0,01 | 0,04 | -0,17 | 8,67E-01 |
| SRSF2 | 2928,78 | 0,00 | 0,03 | -0,17 | 8,68E-01 |
| AIMP2 | 164,23 | 0,01 | 0,03 | 0,17 | 8,68E-01 |
| NCR3LG1 | 150,10 | 0,01 | 0,07 | 0,17 | 8,68E-01 |
| HABP4 | 443,09 | -0,01 | 0,05 | -0,17 | 8,68E-01 |
| FCHO2 | 1962,44 | -0,01 | 0,06 | -0,17 | 8,68E-01 |
| NUP88 | 735,65 | -0,01 | 0,04 | -0,17 | 8,69E-01 |
| ENSG00000269693 | 54,61 | -0,01 | 0,06 | -0,16 | 8,69E-01 |
| PCSK5 | 748,87 | -0,01 | 0,05 | -0,16 | 8,69E-01 |
| GOLGA2 | 1073,67 | 0,01 | 0,05 | 0,16 | 8,70E-01 |
| C1orf162 | 1290,29 | -0,01 | 0,04 | -0,16 | 8,70E-01 |
| C6orf62 | 11825,51 | -0,01 | 0,04 | -0,16 | 8,71E-01 |
| ANTKMT | 323,37 | -0,01 | 0,04 | -0,16 | 8,71E-01 |
| CAVIN2 | 2179,67 | 0,02 | 0,09 | 0,16 | 8,71E-01 |
| ACLY | 2258,27 | -0,01 | 0,04 | -0,16 | 8,71E-01 |
| AP4S1 | 509,25 | 0,01 | 0,03 | 0,16 | 8,71E-01 |
| ZFYVE27 | 1982,31 | 0,00 | 0,03 | -0,16 | 8,72E-01 |

| | | | | | |
|---|---|---|---|---|---|
| HUS1 | 897,76 | 0,00 | 0,03 | -0,16 | 8,72E-01 |
| MRPS24 | 32,54 | 0,01 | 0,07 | 0,16 | 8,72E-01 |
| MMP9 | 912,06 | 0,02 | 0,13 | 0,16 | 8,72E-01 |
| COX7A2L | 1486,96 | -0,01 | 0,05 | -0,16 | 8,72E-01 |
| ITPRIPL2 | 783,17 | 0,01 | 0,05 | 0,16 | 8,72E-01 |
| AK5 | 419,70 | 0,01 | 0,09 | 0,16 | 8,72E-01 |
| GTF2IRD2 | 183,29 | 0,01 | 0,07 | 0,16 | 8,73E-01 |
| SLC25A15 | 169,07 | 0,01 | 0,04 | 0,16 | 8,73E-01 |
| NRDE2 | 1884,80 | -0,01 | 0,03 | -0,16 | 8,74E-01 |
| XKR6 | 329,66 | -0,01 | 0,06 | -0,16 | 8,74E-01 |
| C11orf71 | 300,97 | 0,01 | 0,05 | 0,16 | 8,74E-01 |
| TGFA | 903,84 | -0,01 | 0,07 | -0,16 | 8,74E-01 |
| SCO1 | 993,47 | 0,00 | 0,03 | -0,16 | 8,74E-01 |
| CD302 | 2447,85 | 0,01 | 0,06 | 0,16 | 8,75E-01 |
| OSBPL10 | 591,37 | 0,02 | 0,11 | 0,16 | 8,75E-01 |
| PLSCR1 | 3622,23 | -0,02 | 0,11 | -0,16 | 8,75E-01 |
| ELMO2 | 1542,63 | 0,01 | 0,04 | 0,16 | 8,75E-01 |
| ALDH18A1 | 455,32 | 0,01 | 0,04 | 0,16 | 8,75E-01 |
| PSMB10 | 1629,04 | -0,01 | 0,05 | -0,16 | 8,76E-01 |
| GOLPH3L | 461,40 | 0,00 | 0,03 | -0,16 | 8,76E-01 |
| MRPL41 | 440,90 | 0,01 | 0,05 | 0,16 | 8,76E-01 |
| ANO10 | 500,24 | 0,01 | 0,05 | 0,16 | 8,76E-01 |
| SLC15A4 | 3839,57 | 0,01 | 0,05 | 0,16 | 8,77E-01 |
| PBXIP1 | 4928,43 | 0,01 | 0,05 | 0,16 | 8,77E-01 |
| OASL | 830,41 | -0,02 | 0,14 | -0,15 | 8,77E-01 |
| PATL2 | 1280,40 | 0,01 | 0,07 | 0,15 | 8,78E-01 |
| TMEM43 | 2937,98 | -0,01 | 0,04 | -0,15 | 8,78E-01 |
| CARHSP1 | 999,27 | -0,01 | 0,04 | -0,15 | 8,78E-01 |
| LTB | 4338,19 | -0,01 | 0,05 | -0,15 | 8,78E-01 |
| PAXIP1 | 718,97 | 0,00 | 0,03 | -0,15 | 8,78E-01 |
| TRPC1 | 220,48 | 0,01 | 0,08 | 0,15 | 8,79E-01 |
| APOBEC3H | 39,28 | 0,01 | 0,10 | 0,15 | 8,79E-01 |
| DHRS13 | 911,20 | 0,01 | 0,06 | 0,15 | 8,79E-01 |
| YPEL1 | 428,25 | 0,01 | 0,06 | 0,15 | 8,80E-01 |
| ZNF596 | 97,92 | -0,01 | 0,08 | -0,15 | 8,80E-01 |
| IFT56 | 188,35 | 0,01 | 0,10 | 0,15 | 8,80E-01 |
| ZNF674 | 520,41 | -0,01 | 0,04 | -0,15 | 8,80E-01 |
| FOS | 1296,94 | -0,01 | 0,08 | -0,15 | 8,80E-01 |
| ADGRE3 | 2710,51 | -0,01 | 0,07 | -0,15 | 8,80E-01 |
| RFESD | 164,21 | -0,01 | 0,07 | -0,15 | 8,81E-01 |
| CAB39L | 184,33 | 0,01 | 0,04 | 0,15 | 8,81E-01 |
| POLH | 550,58 | 0,00 | 0,03 | -0,15 | 8,81E-01 |
| TANGO6 | 1138,18 | 0,00 | 0,03 | -0,15 | 8,81E-01 |
| ASB1 | 1156,60 | 0,00 | 0,03 | 0,15 | 8,81E-01 |
| TANC2 | 1459,68 | -0,01 | 0,05 | -0,15 | 8,82E-01 |
| DDX11 | 505,31 | -0,02 | 0,13 | -0,15 | 8,81E-01 |

| | | | | | |
|---|---|---|---|---|---|
| COPG2 | 412,63 | -0,01 | 0,04 | -0,15 | 8,82E-01 |
| CTDP1 | 2413,37 | -0,01 | 0,04 | -0,15 | 8,82E-01 |
| CLN8 | 750,93 | 0,01 | 0,04 | 0,15 | 8,82E-01 |
| ACAD11 | 248,19 | -0,01 | 0,04 | -0,15 | 8,82E-01 |
| ANKHD1 | 986,84 | -0,01 | 0,04 | -0,15 | 8,82E-01 |
| CPED1 | 354,14 | 0,01 | 0,07 | 0,15 | 8,82E-01 |
| OGFR | 3264,35 | 0,01 | 0,05 | 0,15 | 8,83E-01 |
| ZWILCH | 208,07 | -0,01 | 0,05 | -0,15 | 8,83E-01 |
| PARP9 | 5386,23 | -0,01 | 0,09 | -0,15 | 8,84E-01 |
| CAPNS2 | 211,88 | -0,01 | 0,08 | -0,15 | 8,84E-01 |
| ENSG00000285708 | 31,67 | 0,01 | 0,08 | 0,15 | 8,84E-01 |
| RGS17 | 32,87 | 0,02 | 0,16 | 0,15 | 8,84E-01 |
| NAIP | 1580,46 | 0,01 | 0,10 | 0,15 | 8,84E-01 |
| TCAF1 | 246,98 | -0,01 | 0,04 | -0,15 | 8,84E-01 |
| GCNT1 | 828,66 | -0,01 | 0,04 | -0,15 | 8,84E-01 |
| RBM47 | 5016,58 | 0,01 | 0,07 | 0,14 | 8,85E-01 |
| LRRC4 | 1341,74 | 0,01 | 0,07 | 0,14 | 8,86E-01 |
| IL1B | 1065,25 | 0,01 | 0,07 | 0,14 | 8,86E-01 |
| EBPL | 220,05 | 0,01 | 0,05 | 0,14 | 8,87E-01 |
| TRIM5 | 1036,88 | -0,01 | 0,06 | -0,14 | 8,87E-01 |
| MIPEP | 277,10 | 0,01 | 0,04 | 0,14 | 8,87E-01 |
| KHDC4 | 1564,88 | 0,00 | 0,03 | 0,14 | 8,87E-01 |
| SLC16A1 | 344,56 | 0,01 | 0,04 | 0,14 | 8,88E-01 |
| H1-4 | 5141,56 | 0,01 | 0,06 | 0,14 | 8,88E-01 |
| LYN | 11278,71 | -0,01 | 0,06 | -0,14 | 8,88E-01 |
| NBEA | 366,85 | 0,01 | 0,07 | 0,14 | 8,89E-01 |
| CEACAM21 | 449,85 | 0,01 | 0,04 | 0,14 | 8,89E-01 |
| HMGCL | 619,26 | 0,00 | 0,02 | 0,14 | 8,89E-01 |
| GLB1 | 1722,60 | 0,00 | 0,03 | -0,14 | 8,89E-01 |
| OR52K1 | 373,11 | -0,01 | 0,10 | -0,14 | 8,90E-01 |
| CHST11 | 5222,57 | 0,01 | 0,05 | 0,14 | 8,90E-01 |
| SLX4IP | 514,46 | 0,00 | 0,04 | 0,14 | 8,90E-01 |
| RCOR3 | 2485,71 | 0,00 | 0,03 | 0,14 | 8,90E-01 |
| GLYR1 | 2194,14 | 0,00 | 0,03 | -0,14 | 8,91E-01 |
| ADAM19 | 3948,21 | 0,01 | 0,06 | 0,14 | 8,91E-01 |
| ZNG1C | 594,89 | -0,01 | 0,05 | -0,14 | 8,92E-01 |
| HMOX2 | 919,39 | 0,00 | 0,03 | -0,14 | 8,92E-01 |
| MRPL21 | 215,94 | 0,01 | 0,05 | 0,14 | 8,92E-01 |
| CCNQ | 317,13 | 0,00 | 0,04 | 0,14 | 8,93E-01 |
| HVCN1 | 1185,84 | -0,01 | 0,05 | -0,13 | 8,93E-01 |
| KCNH8 | 112,42 | 0,01 | 0,11 | 0,13 | 8,94E-01 |
| B4GALT6 | 113,68 | 0,01 | 0,08 | 0,13 | 8,94E-01 |
| IFT70A | 103,98 | 0,01 | 0,07 | 0,13 | 8,94E-01 |
| ZNF519 | 441,78 | -0,01 | 0,05 | -0,13 | 8,96E-01 |
| RBM14-RBM4 | 272,15 | 0,00 | 0,04 | -0,13 | 8,96E-01 |
| THOC6 | 326,57 | 0,00 | 0,04 | 0,13 | 8,96E-01 |

| | | | | | |
|---|---|---|---|---|---|
| ERAP2 | 3312,42 | -0,01 | 0,08 | -0,13 | 8,97E-01 |
| SNAP29 | 772,14 | 0,00 | 0,03 | 0,13 | 8,97E-01 |
| SLC15A3 | 1374,90 | 0,01 | 0,06 | 0,13 | 8,97E-01 |
| SF3B5 | 795,19 | -0,01 | 0,05 | -0,13 | 8,97E-01 |
| MAP2K5 | 679,70 | -0,01 | 0,05 | -0,13 | 8,97E-01 |
| IRF2BPL | 1277,35 | 0,01 | 0,06 | 0,13 | 8,98E-01 |
| NOMO3 | 249,35 | 0,01 | 0,08 | 0,13 | 8,98E-01 |
| CALCOCO2 | 2249,57 | -0,01 | 0,05 | -0,13 | 8,98E-01 |
| ST6GAL1 | 3735,44 | -0,01 | 0,05 | -0,13 | 8,98E-01 |
| GIMAP5 | 2252,80 | -0,01 | 0,05 | -0,13 | 8,98E-01 |
| CUL4A | 3087,37 | 0,00 | 0,03 | -0,13 | 8,98E-01 |
| CDIN1 | 316,09 | -0,01 | 0,05 | -0,13 | 8,98E-01 |
| MTFMT | 518,98 | 0,00 | 0,03 | -0,13 | 8,99E-01 |
| DPY19L3 | 1377,69 | -0,01 | 0,04 | -0,13 | 8,99E-01 |
| GRK5 | 964,68 | 0,00 | 0,04 | 0,13 | 8,99E-01 |
| NOSIP | 1975,23 | -0,01 | 0,04 | -0,13 | 9,00E-01 |
| SEC24A | 824,67 | 0,00 | 0,04 | -0,13 | 9,00E-01 |
| NOP9 | 1461,28 | 0,00 | 0,04 | -0,12 | 9,01E-01 |
| IGFBP7 | 331,37 | 0,01 | 0,07 | 0,12 | 9,01E-01 |
| CREB5 | 7104,05 | -0,01 | 0,08 | -0,12 | 9,01E-01 |
| FBN2 | 573,45 | -0,01 | 0,10 | -0,12 | 9,01E-01 |
| ITGA6 | 2641,86 | -0,01 | 0,06 | -0,12 | 9,02E-01 |
| C15orf61 | 177,37 | 0,01 | 0,06 | 0,12 | 9,02E-01 |
| LNX2 | 549,99 | 0,00 | 0,03 | -0,12 | 9,02E-01 |
| ZMYND8 | 2165,68 | 0,00 | 0,03 | 0,12 | 9,03E-01 |
| MPZL1 | 3092,35 | -0,01 | 0,08 | -0,12 | 9,03E-01 |
| LTB4R | 1535,51 | -0,01 | 0,06 | -0,12 | 9,03E-01 |
| FADS2 | 342,00 | 0,02 | 0,17 | 0,12 | 9,03E-01 |
| RIMKLB | 627,22 | -0,01 | 0,04 | -0,12 | 9,04E-01 |
| PCTP | 1309,69 | 0,01 | 0,07 | 0,12 | 9,04E-01 |
| SNRNP25 | 152,13 | 0,01 | 0,05 | 0,12 | 9,04E-01 |
| RPP40 | 87,39 | 0,01 | 0,06 | 0,12 | 9,04E-01 |
| PPM1G | 1290,80 | -0,01 | 0,04 | -0,12 | 9,05E-01 |
| SFSWAP | 2569,22 | 0,00 | 0,03 | -0,12 | 9,05E-01 |
| GGACT | 198,01 | 0,01 | 0,05 | 0,12 | 9,05E-01 |
| SAMHD1 | 9316,78 | -0,01 | 0,04 | -0,12 | 9,05E-01 |
| KCTD5 | 882,34 | 0,00 | 0,03 | 0,12 | 9,05E-01 |
| SSH2 | 17829,36 | 0,01 | 0,07 | 0,12 | 9,05E-01 |
| NUBPL | 306,51 | -0,01 | 0,05 | -0,12 | 9,06E-01 |
| EIF2AK2 | 3301,78 | 0,01 | 0,10 | 0,12 | 9,06E-01 |
| RAB33A | 71,46 | 0,01 | 0,06 | 0,12 | 9,06E-01 |
| MGST2 | 201,97 | 0,01 | 0,05 | 0,12 | 9,06E-01 |
| LRG1 | 1382,01 | -0,01 | 0,08 | -0,12 | 9,06E-01 |
| GSTO1 | 1273,05 | 0,01 | 0,06 | 0,12 | 9,07E-01 |
| GOSR1 | 1277,81 | 0,00 | 0,03 | -0,12 | 9,07E-01 |
| ANAPC10 | 170,57 | 0,01 | 0,05 | 0,12 | 9,07E-01 |

| | | | | | |
|---|---|---|---|---|---|
| URB2 | 481,05 | 0,00 | 0,03 | -0,12 | 9,07E-01 |
| NDRG1 | 1971,55 | 0,00 | 0,04 | -0,12 | 9,08E-01 |
| HHEX | 715,56 | -0,01 | 0,05 | -0,12 | 9,08E-01 |
| RUVBL2 | 478,21 | 0,00 | 0,03 | -0,12 | 9,08E-01 |
| RWDD2A | 90,92 | 0,01 | 0,05 | 0,11 | 9,08E-01 |
| CHST15 | 10449,97 | -0,01 | 0,07 | -0,11 | 9,09E-01 |
| ENDOD1 | 1362,53 | 0,01 | 0,06 | 0,11 | 9,09E-01 |
| NEDD4 | 314,45 | 0,01 | 0,07 | 0,11 | 9,09E-01 |
| MTMR4 | 2042,77 | 0,00 | 0,03 | -0,11 | 9,09E-01 |
| CXXC5 | 473,37 | -0,01 | 0,06 | -0,11 | 9,10E-01 |
| MEFV | 7185,53 | 0,01 | 0,07 | 0,11 | 9,10E-01 |
| TRUB2 | 402,98 | 0,00 | 0,03 | 0,11 | 9,10E-01 |
| ZNF587 | 2020,00 | 0,00 | 0,04 | -0,11 | 9,11E-01 |
| CRACR2A | 1374,51 | 0,00 | 0,04 | -0,11 | 9,11E-01 |
| ATG7 | 3095,29 | -0,01 | 0,05 | -0,11 | 9,11E-01 |
| HELB | 4496,83 | 0,00 | 0,04 | 0,11 | 9,11E-01 |
| TMEM131 | 2778,39 | 0,00 | 0,04 | -0,11 | 9,11E-01 |
| TMEM64 | 615,57 | -0,01 | 0,06 | -0,11 | 9,11E-01 |
| KLHL5 | 1352,39 | -0,01 | 0,05 | -0,11 | 9,12E-01 |
| ENSG00000264324 | 24,91 | -0,01 | 0,07 | -0,11 | 9,12E-01 |
| DPYSL2 | 2302,08 | 0,01 | 0,05 | 0,11 | 9,13E-01 |
| MED14 | 1537,08 | 0,00 | 0,04 | 0,11 | 9,13E-01 |
| ITFG2 | 1329,10 | 0,00 | 0,03 | -0,11 | 9,13E-01 |
| SYNRG | 4729,95 | 0,00 | 0,04 | -0,11 | 9,13E-01 |
| WASHC2C | 1762,19 | 0,00 | 0,05 | -0,11 | 9,14E-01 |
| RFWD3 | 930,43 | 0,00 | 0,03 | 0,11 | 9,14E-01 |
| SAP30 | 221,97 | 0,01 | 0,06 | 0,11 | 9,14E-01 |
| MAGEE1 | 80,66 | -0,01 | 0,06 | -0,11 | 9,14E-01 |
| MAPKAPK5 | 1502,64 | 0,00 | 0,03 | -0,11 | 9,14E-01 |
| PCDHGA9 | 35,41 | -0,01 | 0,11 | -0,11 | 9,14E-01 |
| INPP4A | 7529,33 | 0,00 | 0,04 | -0,11 | 9,15E-01 |
| GTF2I | 938,81 | 0,00 | 0,04 | 0,11 | 9,15E-01 |
| SLC25A13 | 665,10 | 0,00 | 0,04 | -0,11 | 9,15E-01 |
| CD40 | 327,34 | -0,01 | 0,06 | -0,11 | 9,15E-01 |
| DCAF8 | 2629,62 | 0,00 | 0,03 | 0,11 | 9,15E-01 |
| CUEDC2 | 283,33 | 0,00 | 0,03 | -0,11 | 9,15E-01 |
| SMIM20 | 162,08 | -0,01 | 0,06 | -0,11 | 9,16E-01 |
| ATP5MC1 | 299,17 | 0,00 | 0,04 | -0,11 | 9,16E-01 |
| MED27 | 290,48 | 0,00 | 0,03 | 0,11 | 9,16E-01 |
| BIN1 | 1584,04 | 0,01 | 0,05 | 0,10 | 9,16E-01 |
| MTHFD1 | 602,94 | 0,00 | 0,03 | 0,10 | 9,16E-01 |
| BUD23 | 485,49 | 0,00 | 0,03 | 0,10 | 9,17E-01 |
| ARPC1A | 282,43 | 0,00 | 0,03 | -0,10 | 9,17E-01 |
| PFDN1 | 165,31 | 0,00 | 0,03 | -0,10 | 9,17E-01 |
| SETBP1 | 607,35 | -0,01 | 0,07 | -0,10 | 9,17E-01 |
| ZNF419 | 360,45 | 0,00 | 0,03 | -0,10 | 9,18E-01 |

| | | | | | |
|---|---|---|---|---|---|
| EI24 | 591,86 | 0,00 | 0,03 | 0,10 | 9,18E-01 |
| AKAP12 | 246,62 | 0,02 | 0,17 | 0,10 | 9,18E-01 |
| C6orf52 | 28,45 | 0,01 | 0,09 | 0,10 | 9,18E-01 |
| KIF11 | 122,14 | 0,01 | 0,08 | 0,10 | 9,19E-01 |
| ANKRD17 | 3811,65 | 0,00 | 0,04 | 0,10 | 9,19E-01 |
| NPC1 | 1699,51 | 0,00 | 0,05 | -0,10 | 9,19E-01 |
| C3orf62 | 1245,90 | 0,01 | 0,06 | 0,10 | 9,20E-01 |
| TPST2 | 2125,13 | 0,00 | 0,04 | 0,10 | 9,20E-01 |
| RAB27B | 1430,32 | 0,01 | 0,08 | 0,10 | 9,21E-01 |
| COMMD9 | 653,05 | 0,00 | 0,03 | -0,10 | 9,22E-01 |
| HERC3 | 6145,41 | 0,00 | 0,04 | -0,10 | 9,22E-01 |
| IMMP2L | 519,02 | -0,01 | 0,06 | -0,10 | 9,22E-01 |
| CHCHD2 | 2251,10 | -0,01 | 0,06 | -0,10 | 9,23E-01 |
| MILR1 | 425,62 | -0,01 | 0,07 | -0,10 | 9,23E-01 |
| RBM28 | 1077,88 | 0,00 | 0,05 | 0,10 | 9,23E-01 |
| PBX2 | 3655,49 | -0,01 | 0,05 | -0,10 | 9,23E-01 |
| MS4A7 | 2659,38 | -0,01 | 0,07 | -0,10 | 9,23E-01 |
| NFYC | 1718,52 | 0,00 | 0,04 | -0,10 | 9,24E-01 |
| ACO1 | 384,41 | 0,00 | 0,04 | -0,10 | 9,24E-01 |
| MX2 | 6786,24 | 0,01 | 0,09 | 0,10 | 9,24E-01 |
| B3GALT4 | 806,75 | 0,00 | 0,03 | -0,09 | 9,25E-01 |
| C6orf163 | 365,60 | 0,00 | 0,05 | 0,09 | 9,26E-01 |
| WDFY2 | 4680,99 | 0,00 | 0,04 | -0,09 | 9,26E-01 |
| PXMP4 | 390,98 | 0,00 | 0,03 | 0,09 | 9,26E-01 |
| ANKRD40 | 523,35 | 0,00 | 0,03 | 0,09 | 9,26E-01 |
| PTPDC1 | 285,93 | 0,00 | 0,05 | -0,09 | 9,27E-01 |
| UBE2R2 | 1868,22 | -0,01 | 0,06 | -0,09 | 9,27E-01 |
| FAU | 4644,71 | 0,01 | 0,07 | 0,09 | 9,27E-01 |
| ZNF169 | 654,64 | 0,00 | 0,03 | 0,09 | 9,27E-01 |
| PDK1 | 3852,76 | 0,00 | 0,05 | 0,09 | 9,28E-01 |
| B9D2 | 377,13 | 0,00 | 0,05 | 0,09 | 9,28E-01 |
| SNTB2 | 1598,59 | 0,00 | 0,03 | -0,09 | 9,28E-01 |
| BBC3 | 508,68 | 0,00 | 0,05 | 0,09 | 9,29E-01 |
| NEK9 | 2777,32 | 0,00 | 0,02 | -0,09 | 9,28E-01 |
| STK3 | 655,32 | 0,01 | 0,06 | 0,09 | 9,29E-01 |
| MRPL14 | 304,82 | 0,00 | 0,04 | -0,09 | 9,29E-01 |
| FKBP15 | 4088,80 | 0,00 | 0,04 | 0,09 | 9,29E-01 |
| ANAPC11 | 458,91 | 0,00 | 0,04 | -0,09 | 9,29E-01 |
| POMK | 530,78 | 0,00 | 0,03 | 0,09 | 9,30E-01 |
| ZNF223 | 58,04 | 0,00 | 0,05 | -0,09 | 9,30E-01 |
| NFKBIA | 2660,94 | 0,00 | 0,04 | 0,09 | 9,30E-01 |
| NUMA1 | 5981,86 | 0,00 | 0,04 | 0,09 | 9,30E-01 |
| DIDO1 | 4416,88 | 0,00 | 0,03 | 0,09 | 9,30E-01 |
| SULT1B1 | 4419,49 | -0,01 | 0,08 | -0,09 | 9,31E-01 |
| PSMB9 | 5599,17 | -0,01 | 0,06 | -0,09 | 9,31E-01 |
| PLEKHG3 | 6391,75 | 0,01 | 0,07 | 0,09 | 9,32E-01 |

| | | | | | |
|---|---|---|---|---|---|
| HCAR2 | 737,47 | -0,01 | 0,08 | -0,08 | 9,33E-01 |
| CR1 | 6650,30 | -0,01 | 0,08 | -0,08 | 9,34E-01 |
| GLCE | 475,68 | 0,00 | 0,04 | 0,08 | 9,34E-01 |
| VAMP3 | 1918,12 | 0,00 | 0,04 | -0,08 | 9,34E-01 |
| ZNF367 | 191,45 | 0,00 | 0,06 | 0,08 | 9,35E-01 |
| ASH2L | 981,37 | 0,00 | 0,04 | -0,08 | 9,35E-01 |
| SNX17 | 2283,22 | 0,00 | 0,03 | -0,08 | 9,35E-01 |
| CEP68 | 674,58 | 0,00 | 0,05 | -0,08 | 9,35E-01 |
| KRT73 | 181,73 | -0,01 | 0,17 | -0,08 | 9,36E-01 |
| DGKH | 1271,43 | 0,00 | 0,04 | 0,08 | 9,36E-01 |
| TIAL1 | 1923,64 | 0,00 | 0,05 | -0,08 | 9,36E-01 |
| CNKSR2 | 560,49 | 0,01 | 0,07 | 0,08 | 9,36E-01 |
| PLEKHA2 | 3805,12 | 0,00 | 0,03 | -0,08 | 9,36E-01 |
| DTX3L | 4262,05 | -0,01 | 0,07 | -0,08 | 9,36E-01 |
| APOL1 | 1033,13 | 0,01 | 0,09 | 0,08 | 9,37E-01 |
| ZNF382 | 263,86 | 0,00 | 0,06 | 0,08 | 9,37E-01 |
| C1orf115 | 74,72 | 0,01 | 0,09 | 0,08 | 9,38E-01 |
| NUGGC | 279,50 | 0,01 | 0,09 | 0,08 | 9,38E-01 |
| SLC39A1 | 113,32 | 0,00 | 0,04 | -0,08 | 9,38E-01 |
| TTC39C | 2004,80 | 0,00 | 0,04 | 0,08 | 9,38E-01 |
| CAMSAP2 | 220,01 | 0,01 | 0,07 | 0,08 | 9,38E-01 |
| MGAM | 10795,44 | 0,01 | 0,09 | 0,08 | 9,38E-01 |
| JUN | 303,32 | 0,00 | 0,06 | 0,08 | 9,38E-01 |
| NT5C | 493,86 | 0,00 | 0,04 | 0,08 | 9,39E-01 |
| TMEM223 | 233,89 | 0,00 | 0,04 | -0,08 | 9,40E-01 |
| AVPI1 | 35,42 | 0,01 | 0,08 | 0,08 | 9,40E-01 |
| WARS1 | 3995,09 | -0,01 | 0,09 | -0,07 | 9,40E-01 |
| CPOX | 831,78 | 0,00 | 0,04 | -0,07 | 9,40E-01 |
| STK19 | 337,02 | 0,00 | 0,03 | -0,07 | 9,41E-01 |
| FAM120A | 3908,60 | 0,00 | 0,04 | -0,07 | 9,41E-01 |
| ETS2 | 2317,89 | 0,00 | 0,05 | -0,07 | 9,41E-01 |
| CCDC51 | 138,58 | 0,00 | 0,04 | 0,07 | 9,41E-01 |
| CDK6 | 2298,27 | 0,00 | 0,04 | 0,07 | 9,42E-01 |
| HIRA | 319,27 | 0,00 | 0,04 | 0,07 | 9,42E-01 |
| RWDD2B | 85,68 | 0,00 | 0,05 | 0,07 | 9,42E-01 |
| LDAF1 | 346,28 | 0,00 | 0,04 | -0,07 | 9,43E-01 |
| CSGALNACT1 | 1387,18 | 0,00 | 0,07 | -0,07 | 9,43E-01 |
| CCDC127 | 507,58 | 0,00 | 0,04 | -0,07 | 9,43E-01 |
| NUDT16 | 1709,70 | 0,00 | 0,04 | 0,07 | 9,43E-01 |
| TM7SF3 | 1082,34 | 0,00 | 0,03 | 0,07 | 9,43E-01 |
| RAE1 | 762,38 | 0,00 | 0,02 | -0,07 | 9,43E-01 |
| DUSP6 | 2629,12 | 0,00 | 0,06 | -0,07 | 9,44E-01 |
| PPP1R3D | 1214,41 | 0,00 | 0,04 | -0,07 | 9,44E-01 |
| ZXDC | 2755,36 | 0,00 | 0,04 | 0,07 | 9,44E-01 |
| DNA2 | 304,90 | 0,00 | 0,06 | -0,07 | 9,44E-01 |
| MARCHF6 | 6814,62 | 0,00 | 0,04 | -0,07 | 9,45E-01 |

| | | | | | |
|---|---|---|---|---|---|
| MX1 | 3908,27 | 0,01 | 0,16 | 0,07 | 9,45E-01 |
| SHLD1 | 289,80 | 0,00 | 0,06 | 0,07 | 9,45E-01 |
| VPS16 | 1219,95 | 0,00 | 0,03 | -0,07 | 9,46E-01 |
| EDEM2 | 1384,44 | 0,00 | 0,03 | 0,07 | 9,46E-01 |
| HLA-DRB5 | 1507,01 | 0,01 | 0,20 | 0,07 | 9,46E-01 |
| ATP6V0B | 4646,79 | 0,00 | 0,05 | -0,07 | 9,46E-01 |
| MFSD5 | 448,84 | 0,00 | 0,03 | 0,07 | 9,46E-01 |
| KIAA0753 | 812,29 | 0,00 | 0,04 | 0,07 | 9,46E-01 |
| MAT2A | 2353,60 | 0,00 | 0,03 | -0,07 | 9,47E-01 |
| DUS2 | 709,99 | 0,00 | 0,04 | 0,07 | 9,47E-01 |
| RECK | 341,10 | 0,00 | 0,03 | -0,07 | 9,47E-01 |
| CNIH4 | 1240,28 | 0,00 | 0,05 | -0,07 | 9,47E-01 |
| MLEC | 1994,41 | 0,00 | 0,03 | -0,07 | 9,47E-01 |
| H2AC12 | 344,02 | 0,01 | 0,11 | 0,07 | 9,47E-01 |
| NCOA5 | 679,21 | 0,00 | 0,03 | -0,06 | 9,49E-01 |
| ANO6 | 2922,29 | 0,00 | 0,04 | -0,06 | 9,49E-01 |
| HEATR6 | 546,16 | 0,00 | 0,04 | 0,06 | 9,49E-01 |
| RFT1 | 649,44 | 0,00 | 0,03 | 0,06 | 9,49E-01 |
| OGT | 19928,03 | 0,00 | 0,04 | -0,06 | 9,49E-01 |
| IRF4 | 661,02 | 0,00 | 0,06 | -0,06 | 9,49E-01 |
| KCNJ2 | 5138,20 | 0,00 | 0,08 | 0,06 | 9,50E-01 |
| OSGEP | 1019,65 | 0,00 | 0,03 | 0,06 | 9,50E-01 |
| TBCEL | 1261,54 | 0,00 | 0,08 | 0,06 | 9,50E-01 |
| CCNJL | 1441,01 | 0,01 | 0,09 | 0,06 | 9,50E-01 |
| MRPS15 | 302,08 | 0,00 | 0,03 | 0,06 | 9,50E-01 |
| DRAM1 | 488,12 | 0,00 | 0,05 | -0,06 | 9,51E-01 |
| C2orf74 | 59,18 | 0,00 | 0,05 | 0,06 | 9,51E-01 |
| CCDC102B | 128,09 | 0,00 | 0,06 | -0,06 | 9,51E-01 |
| EBF1 | 292,73 | 0,01 | 0,10 | 0,06 | 9,52E-01 |
| RAB5IF | 1273,64 | 0,00 | 0,05 | -0,06 | 9,52E-01 |
| TCEAL3 | 104,18 | 0,00 | 0,06 | 0,06 | 9,53E-01 |
| MED19 | 113,58 | 0,00 | 0,04 | 0,06 | 9,53E-01 |
| MXD1 | 26536,97 | 0,00 | 0,07 | 0,06 | 9,53E-01 |
| COQ2 | 557,29 | 0,00 | 0,04 | -0,06 | 9,54E-01 |
| RHOC | 449,82 | 0,00 | 0,05 | 0,06 | 9,54E-01 |
| SLC25A51 | 659,30 | 0,00 | 0,03 | 0,06 | 9,54E-01 |
| CAPN2 | 3800,51 | 0,00 | 0,04 | -0,06 | 9,54E-01 |
| TSEN34 | 2194,78 | 0,00 | 0,06 | 0,06 | 9,54E-01 |
| PHLDA1 | 154,68 | 0,00 | 0,06 | -0,06 | 9,54E-01 |
| NPM3 | 137,31 | 0,00 | 0,05 | -0,06 | 9,55E-01 |
| B4GAT1 | 73,66 | 0,00 | 0,06 | 0,06 | 9,55E-01 |
| HINT2 | 46,39 | 0,00 | 0,06 | -0,06 | 9,55E-01 |
| YPEL2 | 2152,20 | 0,00 | 0,03 | 0,06 | 9,56E-01 |
| MFSD4B | 1226,85 | 0,00 | 0,03 | 0,06 | 9,56E-01 |
| MLF2 | 1306,30 | 0,00 | 0,05 | 0,05 | 9,56E-01 |
| GSR | 2550,25 | 0,00 | 0,03 | 0,05 | 9,56E-01 |

| | | | | | |
|---|---|---|---|---|---|
| WDR37 | 2666,83 | 0,00 | 0,03 | 0,05 | 9,57E-01 |
| CTSW | 384,68 | -0,01 | 0,11 | -0,05 | 9,57E-01 |
| USP46 | 493,08 | 0,00 | 0,05 | 0,05 | 9,57E-01 |
| ELP6 | 395,00 | 0,00 | 0,03 | -0,05 | 9,57E-01 |
| SETD4 | 696,68 | 0,00 | 0,03 | 0,05 | 9,57E-01 |
| DBNL | 11039,38 | 0,00 | 0,04 | 0,05 | 9,58E-01 |
| MFN2 | 3337,80 | 0,00 | 0,07 | 0,05 | 9,58E-01 |
| MPZ | 246,13 | 0,00 | 0,05 | 0,05 | 9,58E-01 |
| POGZ | 2044,36 | 0,00 | 0,04 | -0,05 | 9,58E-01 |
| RPS10 | 114,33 | 0,00 | 0,03 | 0,05 | 9,59E-01 |
| GLRX | 2751,00 | 0,00 | 0,05 | -0,05 | 9,59E-01 |
| CARD8 | 6901,70 | 0,00 | 0,05 | 0,05 | 9,60E-01 |
| PHTF1 | 1157,57 | 0,00 | 0,04 | -0,05 | 9,60E-01 |
| MPV17L2 | 63,01 | 0,00 | 0,05 | 0,05 | 9,60E-01 |
| ACVR1C | 206,04 | 0,00 | 0,09 | 0,05 | 9,60E-01 |
| GNB5 | 1194,63 | 0,00 | 0,04 | 0,05 | 9,61E-01 |
| ZNF429 | 662,63 | 0,00 | 0,07 | 0,05 | 9,61E-01 |
| IRS2 | 1531,82 | 0,00 | 0,07 | 0,05 | 9,61E-01 |
| MOSPD1 | 376,17 | 0,00 | 0,07 | 0,05 | 9,61E-01 |
| ABHD2 | 8936,24 | 0,00 | 0,07 | -0,05 | 9,61E-01 |
| MARCKS | 2707,36 | 0,00 | 0,07 | 0,05 | 9,63E-01 |
| STX3 | 5754,46 | 0,00 | 0,07 | 0,05 | 9,63E-01 |
| ZBTB8OS | 462,14 | 0,00 | 0,03 | -0,05 | 9,64E-01 |
| TGM3 | 98,04 | 0,01 | 0,13 | 0,05 | 9,64E-01 |
| IPO9 | 1682,71 | 0,00 | 0,03 | 0,04 | 9,65E-01 |
| SDHAF2 | 602,41 | 0,00 | 0,03 | 0,04 | 9,65E-01 |
| CDC42SE1 | 16081,73 | 0,00 | 0,03 | 0,04 | 9,65E-01 |
| EZH1 | 2498,13 | 0,00 | 0,04 | 0,04 | 9,66E-01 |
| LY96 | 576,38 | 0,00 | 0,09 | -0,04 | 9,66E-01 |
| TRIB1 | 797,28 | 0,00 | 0,06 | 0,04 | 9,66E-01 |
| CCDC107 | 365,32 | 0,00 | 0,04 | 0,04 | 9,66E-01 |
| PACC1 | 471,65 | 0,00 | 0,03 | -0,04 | 9,67E-01 |
| SPRY2 | 63,64 | 0,00 | 0,08 | 0,04 | 9,67E-01 |
| RRBP1 | 1646,69 | 0,00 | 0,05 | -0,04 | 9,68E-01 |
| RRP8 | 920,39 | 0,00 | 0,03 | 0,04 | 9,68E-01 |
| NOM1 | 944,47 | 0,00 | 0,03 | 0,04 | 9,68E-01 |
| RUFY1 | 2472,43 | 0,00 | 0,05 | -0,04 | 9,68E-01 |
| VIM | 19423,24 | 0,00 | 0,05 | -0,04 | 9,69E-01 |
| PINX1 | 156,01 | 0,00 | 0,04 | 0,04 | 9,69E-01 |
| SNX1 | 4063,33 | 0,00 | 0,03 | -0,04 | 9,69E-01 |
| GORASP2 | 543,58 | 0,00 | 0,03 | -0,04 | 9,69E-01 |
| RBM11 | 56,50 | 0,00 | 0,10 | -0,04 | 9,70E-01 |
| CIDEB | 639,06 | 0,00 | 0,05 | -0,04 | 9,70E-01 |
| METTL9 | 3845,00 | 0,00 | 0,06 | 0,04 | 9,71E-01 |
| IL1R2 | 2640,17 | 0,00 | 0,08 | -0,04 | 9,71E-01 |
| SIPA1L2 | 2052,31 | 0,00 | 0,09 | 0,03 | 9,72E-01 |

| | | | | | |
|---|---|---|---|---|---|
| KLHDC1 | 298,72 | 0,00 | 0,05 | 0,03 | 9,72E-01 |
| PEX16 | 888,28 | 0,00 | 0,03 | -0,03 | 9,73E-01 |
| RIC3 | 695,34 | 0,00 | 0,07 | 0,03 | 9,73E-01 |
| SDHAF4 | 213,83 | 0,00 | 0,04 | -0,03 | 9,73E-01 |
| FTO | 1522,26 | 0,00 | 0,05 | -0,03 | 9,73E-01 |
| KLF6 | 8423,44 | 0,00 | 0,04 | -0,03 | 9,73E-01 |
| SFXN1 | 1120,43 | 0,00 | 0,04 | 0,03 | 9,74E-01 |
| SULT1A2 | 39,79 | 0,00 | 0,14 | 0,03 | 9,74E-01 |
| SKAP1 | 821,38 | 0,00 | 0,06 | 0,03 | 9,74E-01 |
| USP6NL | 1122,32 | 0,00 | 0,04 | 0,03 | 9,75E-01 |
| NR2C2 | 3572,63 | 0,00 | 0,04 | -0,03 | 9,76E-01 |
| ADAMTS5 | 119,08 | 0,00 | 0,10 | 0,03 | 9,76E-01 |
| CHMP7 | 3108,62 | 0,00 | 0,04 | 0,03 | 9,76E-01 |
| HDAC8 | 874,72 | 0,00 | 0,05 | 0,03 | 9,76E-01 |
| FHIP2A | 2545,50 | 0,00 | 0,05 | -0,03 | 9,76E-01 |
| RNASET2 | 9567,64 | 0,00 | 0,06 | -0,03 | 9,76E-01 |
| CCDC32 | 701,67 | 0,00 | 0,04 | 0,03 | 9,76E-01 |
| ZNF619 | 392,41 | 0,00 | 0,03 | 0,03 | 9,77E-01 |
| BCAP31 | 1174,03 | 0,00 | 0,03 | 0,03 | 9,77E-01 |
| ARHGAP10 | 496,09 | 0,00 | 0,06 | -0,03 | 9,77E-01 |
| COX5A | 149,05 | 0,00 | 0,05 | 0,03 | 9,79E-01 |
| TIGD6 | 138,38 | 0,00 | 0,06 | -0,03 | 9,79E-01 |
| DCTPP1 | 195,71 | 0,00 | 0,04 | 0,03 | 9,79E-01 |
| RGPD1 | 36,56 | 0,00 | 0,16 | 0,03 | 9,80E-01 |
| GPR34 | 146,99 | 0,00 | 0,11 | -0,03 | 9,80E-01 |
| CEP250 | 1857,80 | 0,00 | 0,04 | 0,02 | 9,80E-01 |
| STK24 | 3812,09 | 0,00 | 0,03 | 0,02 | 9,81E-01 |
| FBXL13 | 885,20 | 0,00 | 0,09 | -0,02 | 9,81E-01 |
| HDGFL3 | 258,28 | 0,00 | 0,09 | 0,02 | 9,81E-01 |
| UXS1 | 1370,95 | 0,00 | 0,04 | -0,02 | 9,81E-01 |
| HORMAD1 | 261,00 | 0,00 | 0,09 | 0,02 | 9,81E-01 |
| ANKUB1 | 126,29 | 0,00 | 0,09 | -0,02 | 9,81E-01 |
| ARNT | 1816,94 | 0,00 | 0,03 | 0,02 | 9,82E-01 |
| ZNG1F | 558,27 | 0,00 | 0,07 | -0,02 | 9,82E-01 |
| ESYT2 | 4538,42 | 0,00 | 0,03 | -0,02 | 9,82E-01 |
| CCSER1 | 250,49 | 0,00 | 0,08 | -0,02 | 9,83E-01 |
| GLG1 | 5257,93 | 0,00 | 0,04 | -0,02 | 9,84E-01 |
| BCL2 | 5448,51 | 0,00 | 0,06 | 0,02 | 9,85E-01 |
| ENSG00000285897 | 40,83 | 0,00 | 0,05 | 0,02 | 9,86E-01 |
| PLCG2 | 5833,91 | 0,00 | 0,06 | 0,02 | 9,86E-01 |
| SIRT4 | 37,95 | 0,00 | 0,07 | -0,02 | 9,86E-01 |
| AUTS2 | 624,25 | 0,00 | 0,09 | 0,02 | 9,86E-01 |
| WRNIP1 | 1342,95 | 0,00 | 0,02 | -0,02 | 9,86E-01 |
| PCDHGA10 | 76,32 | 0,00 | 0,11 | 0,02 | 9,86E-01 |
| BTN3A1 | 7389,57 | 0,00 | 0,05 | 0,02 | 9,87E-01 |
| DCUN1D1 | 4415,17 | 0,00 | 0,07 | 0,02 | 9,87E-01 |

| | | | | | |
|---|---|---|---|---|---|
| TLR7 | 482,16 | 0,00 | 0,06 | -0,02 | 9,87E-01 |
| GOLIM4 | 475,58 | 0,00 | 0,04 | -0,02 | 9,88E-01 |
| SULT1A1 | 1455,70 | 0,00 | 0,09 | 0,02 | 9,88E-01 |
| FCGR2A | 19660,78 | 0,00 | 0,07 | -0,01 | 9,88E-01 |
| GNLY | 2516,91 | 0,00 | 0,11 | -0,01 | 9,88E-01 |
| KICS2 | 393,44 | 0,00 | 0,04 | 0,01 | 9,89E-01 |
| SDHAF1 | 286,98 | 0,00 | 0,04 | -0,01 | 9,89E-01 |
| NCKAP1L | 8392,43 | 0,00 | 0,03 | 0,01 | 9,89E-01 |
| PECR | 523,10 | 0,00 | 0,03 | 0,01 | 9,89E-01 |
| SEC61A1 | 2923,34 | 0,00 | 0,02 | -0,01 | 9,89E-01 |
| WDFY3 | 11704,46 | 0,00 | 0,07 | 0,01 | 9,89E-01 |
| RALGAPA2 | 5901,26 | 0,00 | 0,06 | -0,01 | 9,89E-01 |
| ZNF652 | 4527,34 | 0,00 | 0,05 | 0,01 | 9,89E-01 |
| PLEKHM3 | 1843,54 | 0,00 | 0,04 | 0,01 | 9,90E-01 |
| FAT4 | 137,91 | 0,00 | 0,12 | -0,01 | 9,90E-01 |
| H2BC4 | 3923,76 | 0,00 | 0,07 | -0,01 | 9,90E-01 |
| HCK | 12361,03 | 0,00 | 0,06 | -0,01 | 9,90E-01 |
| ZFP36L2 | 3902,81 | 0,00 | 0,05 | -0,01 | 9,90E-01 |
| CDC16 | 1404,34 | 0,00 | 0,04 | -0,01 | 9,91E-01 |
| ENKUR | 126,85 | 0,00 | 0,10 | -0,01 | 9,91E-01 |
| SUSD1 | 1292,28 | 0,00 | 0,04 | 0,01 | 9,91E-01 |
| SLC41A2 | 90,00 | 0,00 | 0,07 | 0,01 | 9,92E-01 |
| ZFP36L1 | 2746,03 | 0,00 | 0,06 | 0,01 | 9,92E-01 |
| SGMS2 | 823,83 | 0,00 | 0,06 | 0,01 | 9,92E-01 |
| FAM156A | 46,51 | 0,00 | 0,05 | 0,01 | 9,92E-01 |
| CDK11A | 200,38 | 0,00 | 0,06 | -0,01 | 9,92E-01 |
| TAF11L2 | 35,24 | 0,00 | 0,12 | 0,01 | 9,94E-01 |
| TMEM273 | 1305,61 | 0,00 | 0,08 | -0,01 | 9,94E-01 |
| C12orf42 | 248,13 | 0,00 | 0,07 | -0,01 | 9,94E-01 |
| ENSA | 1461,22 | 0,00 | 0,03 | 0,01 | 9,94E-01 |
| C9orf78 | 6990,15 | 0,00 | 0,11 | 0,01 | 9,94E-01 |
| S100PBP | 2052,08 | 0,00 | 0,03 | 0,01 | 9,94E-01 |
| ZNF558 | 441,92 | 0,00 | 0,04 | 0,01 | 9,95E-01 |
| CDC14A | 2523,03 | 0,00 | 0,04 | -0,01 | 9,95E-01 |
| PILRB | 503,04 | 0,00 | 0,10 | -0,01 | 9,95E-01 |
| DHRS3 | 356,21 | 0,00 | 0,05 | 0,01 | 9,95E-01 |
| RCC2 | 1929,41 | 0,00 | 0,04 | 0,01 | 9,95E-01 |
| FZD3 | 251,62 | 0,00 | 0,08 | -0,01 | 9,96E-01 |
| ZBTB39 | 392,09 | 0,00 | 0,03 | 0,00 | 9,96E-01 |
| MKNK1 | 2670,85 | 0,00 | 0,06 | 0,00 | 9,96E-01 |
| FAM149B1 | 523,51 | 0,00 | 0,02 | 0,00 | 9,97E-01 |
| C3AR1 | 332,04 | 0,00 | 0,08 | 0,00 | 9,97E-01 |
| ING1 | 929,77 | 0,00 | 0,03 | 0,00 | 9,97E-01 |
| SLC45A4 | 3959,34 | 0,00 | 0,08 | 0,00 | 9,97E-01 |
| MTX2 | 318,22 | 0,00 | 0,06 | 0,00 | 9,97E-01 |
| ATP6V0D1 | 3765,95 | 0,00 | 0,05 | 0,00 | 9,98E-01 |

| | | | | | |
|---|---|---|---|---|---|
| PDCD7 | 646,00 | 0,00 | 0,03 | 0,00 | 9,98E-01 |
| TBXAS1 | 7666,25 | 0,00 | 0,06 | 0,00 | 9,98E-01 |
| MTMR8 | 230,95 | 0,00 | 0,05 | 0,00 | 9,98E-01 |
| SSRP1 | 730,07 | 0,00 | 0,03 | 0,00 | 9,98E-01 |
| SP1 | 6974,16 | 0,00 | 0,05 | 0,00 | 9,99E-01 |
| PDCD11 | 1172,11 | 0,00 | 0,03 | 0,00 | 9,99E-01 |
| PLAG1 | 360,02 | 0,00 | 0,08 | 0,00 | 9,99E-01 |
| AKAP8 | 1271,18 | 0,00 | 0,02 | 0,00 | 9,99E-01 |
| FRAT2 | 8608,28 | 0,00 | 0,07 | 0,00 | 9,99E-01 |
| PRKRIP1 | 785,40 | 0,00 | 0,02 | 0,00 | 9,99E-01 |
| USP13 | 334,69 | 0,00 | 0,04 | 0,00 | 1,00E+00 |
| QSER1 | 1200,52 | 0,00 | 0,03 | 0,00 | 1,00E+00 |

| padj |
| --- |
| 1,11E-04 |
| 1,11E-04 |
| 1,11E-04 |
| 1,11E-04 |
| 1,11E-04 |
| 1,11E-04 |
| 1,11E-04 |
| 1,17E-04 |
| 1,17E-04 |
| 1,17E-04 |
| 1,24E-04 |
| 1,59E-04 |
| 1,59E-04 |
| 1,73E-04 |
| 1,73E-04 |
| 1,73E-04 |
| 1,94E-04 |
| 1,94E-04 |
| 1,94E-04 |
| 1,94E-04 |
| 1,94E-04 |
| 1,94E-04 |
| 1,94E-04 |
| 1,94E-04 |
| 1,95E-04 |
| 1,97E-04 |
| 1,97E-04 |
| 1,97E-04 |
| 2,02E-04 |
| 2,02E-04 |
| 2,02E-04 |
| 2,05E-04 |
| 2,19E-04 |
| 2,23E-04 |
| 2,23E-04 |
| 2,23E-04 |
| 2,37E-04 |
| 2,37E-04 |
| 2,37E-04 |
| 2,42E-04 |
| 2,42E-04 |
| 2,42E-04 |
| 2,42E-04 |
| 2,42E-04 |
| 2,45E-04 |

2,58E-04
3,12E-04
3,16E-04
3,16E-04
3,16E-04
3,21E-04
3,25E-04
3,42E-04
3,48E-04
3,48E-04
3,52E-04
3,52E-04
3,54E-04
3,54E-04
3,54E-04
3,59E-04
3,66E-04
3,78E-04
3,78E-04
3,78E-04
3,82E-04
3,82E-04
3,82E-04
3,82E-04
3,82E-04
3,82E-04
3,82E-04
3,82E-04
3,82E-04
3,82E-04
3,91E-04
3,95E-04
3,95E-04
3,95E-04
3,96E-04
3,96E-04
3,98E-04
4,03E-04
4,08E-04
4,08E-04
4,08E-04
4,08E-04
4,08E-04
4,08E-04
4,08E-04
4,08E-04

4,08E-04
4,08E-04
4,08E-04
4,08E-04
4,08E-04
4,08E-04
4,08E-04
4,10E-04
4,11E-04
4,11E-04
4,11E-04
4,11E-04
4,11E-04
4,14E-04
4,14E-04
4,14E-04
4,14E-04
4,14E-04
4,15E-04
4,15E-04
4,15E-04
4,15E-04
4,15E-04
4,15E-04
4,15E-04
4,15E-04
4,15E-04
4,15E-04
4,15E-04
4,15E-04
4,15E-04
4,15E-04
4,15E-04
4,15E-04
4,15E-04
4,15E-04
4,31E-04
4,31E-04
4,31E-04
4,31E-04
4,31E-04
4,31E-04
4,31E-04
4,34E-04
4,34E-04
4,34E-04

4,34E-04
4,34E-04
4,34E-04
4,41E-04
4,43E-04
4,47E-04
4,47E-04
4,48E-04
4,48E-04
4,48E-04
4,48E-04
4,48E-04
4,48E-04
4,48E-04
4,48E-04
4,54E-04
4,55E-04
4,57E-04
4,57E-04
4,57E-04
4,57E-04
4,57E-04
4,57E-04
4,57E-04
4,57E-04
4,57E-04
4,57E-04
4,57E-04
4,57E-04
4,57E-04
4,58E-04
4,74E-04
4,74E-04
4,74E-04
4,74E-04
4,74E-04
4,74E-04
4,74E-04
4,76E-04
4,76E-04
4,76E-04
4,76E-04
4,76E-04
4,76E-04
4,79E-04
4,79E-04

4,79E-04
4,79E-04
4,79E-04
4,94E-04
4,97E-04
5,02E-04
5,04E-04
5,07E-04
5,21E-04
5,21E-04
5,21E-04
5,28E-04
5,30E-04
5,30E-04
5,38E-04
5,41E-04
5,41E-04
5,41E-04
5,43E-04
5,43E-04
5,43E-04
5,54E-04
5,54E-04
5,62E-04
5,62E-04
5,62E-04
5,66E-04
5,66E-04
5,66E-04
5,66E-04
5,70E-04
5,70E-04
5,70E-04
5,74E-04
5,75E-04
5,75E-04
5,82E-04
5,90E-04
5,93E-04
5,94E-04
5,94E-04
5,94E-04
5,94E-04
5,96E-04
6,02E-04
6,02E-04

6,03E-04
6,11E-04
6,11E-04
6,11E-04
6,11E-04
6,11E-04
6,11E-04
6,15E-04
6,15E-04
6,15E-04
6,15E-04
6,18E-04
6,18E-04
6,21E-04
6,24E-04
6,24E-04
6,26E-04
6,26E-04
6,27E-04
6,27E-04
6,27E-04
6,27E-04
6,27E-04
6,28E-04
6,28E-04
6,31E-04
6,31E-04
6,31E-04
6,32E-04
6,32E-04
6,32E-04
6,32E-04
6,32E-04
6,36E-04
6,36E-04
6,36E-04
6,36E-04
6,36E-04
6,38E-04
6,38E-04
6,47E-04
6,47E-04
6,47E-04
6,47E-04
6,47E-04
6,47E-04

| |
|---|
| 6,47E-04 |
| 6,47E-04 |
| 6,47E-04 |
| 6,55E-04 |
| 6,72E-04 |
| 6,80E-04 |
| 6,84E-04 |
| 6,86E-04 |
| 6,89E-04 |
| 6,91E-04 |
| 6,92E-04 |
| 6,93E-04 |
| 6,93E-04 |
| 6,93E-04 |
| 6,96E-04 |
| 6,96E-04 |
| 6,96E-04 |
| 7,01E-04 |
| 7,05E-04 |
| 7,06E-04 |
| 7,08E-04 |
| 7,08E-04 |
| 7,08E-04 |
| 7,08E-04 |
| 7,08E-04 |
| 7,08E-04 |
| 7,08E-04 |
| 7,22E-04 |
| 7,24E-04 |
| 7,24E-04 |
| 7,27E-04 |
| 7,27E-04 |
| 7,32E-04 |
| 7,34E-04 |
| 7,34E-04 |
| 7,43E-04 |
| 7,58E-04 |
| 7,58E-04 |
| 7,58E-04 |
| 7,59E-04 |
| 7,59E-04 |
| 7,59E-04 |
| 7,59E-04 |
| 7,59E-04 |
| 7,59E-04 |
| 7,59E-04 |

7,59E-04
7,60E-04
7,60E-04
7,65E-04
7,67E-04
7,67E-04
7,67E-04
7,67E-04
7,67E-04
7,91E-04
7,91E-04
7,91E-04
7,91E-04
7,91E-04
7,91E-04
7,91E-04
7,96E-04
7,96E-04
7,96E-04
7,99E-04
7,99E-04
7,99E-04
8,04E-04
8,04E-04
8,04E-04
8,04E-04
8,04E-04
8,04E-04
8,04E-04
8,04E-04
8,04E-04
8,04E-04
8,04E-04
8,04E-04
8,04E-04
8,04E-04
8,04E-04
8,17E-04
8,17E-04
8,18E-04
8,21E-04
8,22E-04
8,42E-04
8,42E-04
8,42E-04
8,47E-04

8,56E-04
8,67E-04
8,75E-04
8,83E-04
8,93E-04
8,94E-04
8,94E-04
8,97E-04
9,09E-04
9,10E-04
9,16E-04
9,19E-04
9,26E-04
9,26E-04
9,27E-04
9,27E-04
9,29E-04
9,29E-04
9,29E-04
9,30E-04
9,32E-04
9,45E-04
9,45E-04
9,50E-04
9,50E-04
9,50E-04
9,50E-04
9,51E-04
9,51E-04
9,63E-04
9,63E-04
9,63E-04
9,63E-04
9,63E-04
9,65E-04
9,68E-04
9,68E-04
9,74E-04
9,79E-04
9,79E-04
9,95E-04
9,96E-04
9,96E-04
9,96E-04
9,96E-04
9,96E-04

9,96E-04
9,96E-04
9,97E-04
9,97E-04
1,00E-03
1,00E-03
1,00E-03
1,01E-03
1,01E-03
1,01E-03
1,01E-03
1,01E-03
1,01E-03
1,01E-03
1,01E-03
1,01E-03
1,01E-03
1,01E-03
1,01E-03
1,02E-03
1,02E-03
1,02E-03
1,02E-03
1,02E-03
1,02E-03
1,03E-03
1,03E-03
1,03E-03
1,03E-03
1,03E-03
1,03E-03
1,04E-03
1,04E-03
1,04E-03
1,04E-03
1,06E-03
1,06E-03
1,06E-03
1,06E-03
1,07E-03
1,07E-03
1,07E-03
1,07E-03
1,07E-03
1,07E-03
1,07E-03

1,07E-03
1,07E-03
1,07E-03
1,07E-03
1,07E-03
1,07E-03
1,07E-03
1,07E-03
1,07E-03
1,07E-03
1,07E-03
1,07E-03
1,07E-03
1,08E-03
1,08E-03
1,08E-03
1,08E-03
1,08E-03
1,10E-03
1,11E-03
1,11E-03
1,11E-03
1,12E-03
1,13E-03
1,13E-03
1,13E-03
1,13E-03
1,13E-03
1,13E-03
1,14E-03
1,14E-03
1,15E-03
1,15E-03
1,15E-03
1,15E-03
1,16E-03
1,17E-03
1,17E-03
1,17E-03
1,18E-03
1,18E-03
1,18E-03
1,18E-03
1,19E-03
1,19E-03
1,19E-03

1,19E-03
1,19E-03
1,19E-03
1,19E-03
1,19E-03
1,19E-03
1,19E-03
1,20E-03
1,20E-03
1,20E-03
1,20E-03
1,20E-03
1,20E-03
1,20E-03
1,20E-03
1,20E-03
1,20E-03
1,20E-03
1,21E-03
1,24E-03
1,24E-03
1,24E-03
1,24E-03
1,24E-03
1,24E-03
1,24E-03
1,25E-03
1,25E-03
1,25E-03
1,25E-03
1,25E-03
1,25E-03
1,25E-03
1,25E-03
1,26E-03
1,27E-03
1,27E-03
1,27E-03
1,29E-03
1,30E-03
1,30E-03
1,31E-03
1,31E-03
1,32E-03
1,32E-03
1,32E-03

1,32E-03
1,32E-03
1,32E-03
1,32E-03
1,32E-03
1,33E-03
1,33E-03
1,33E-03
1,33E-03
1,33E-03
1,34E-03
1,34E-03
1,34E-03
1,35E-03
1,35E-03
1,36E-03
1,37E-03
1,38E-03
1,38E-03
1,38E-03
1,40E-03
1,40E-03
1,40E-03
1,40E-03
1,40E-03
1,40E-03
1,40E-03
1,42E-03
1,42E-03
1,42E-03
1,42E-03
1,42E-03
1,42E-03
1,42E-03
1,43E-03
1,43E-03
1,43E-03
1,43E-03
1,43E-03
1,44E-03
1,44E-03
1,44E-03
1,44E-03
1,45E-03
1,45E-03
1,45E-03

1,45E-03
1,45E-03
1,45E-03
1,45E-03
1,46E-03
1,46E-03
1,47E-03
1,47E-03
1,47E-03
1,48E-03
1,48E-03
1,48E-03
1,48E-03
1,49E-03
1,49E-03
1,49E-03
1,51E-03
1,52E-03
1,52E-03
1,52E-03
1,52E-03
1,52E-03
1,52E-03
1,52E-03
1,52E-03
1,52E-03
1,52E-03
1,53E-03
1,54E-03
1,54E-03
1,54E-03
1,56E-03
1,56E-03
1,57E-03
1,57E-03
1,57E-03
1,58E-03
1,58E-03
1,58E-03
1,59E-03
1,59E-03
1,60E-03
1,60E-03
1,60E-03
1,60E-03
1,60E-03

1,60E-03
1,60E-03
1,60E-03
1,60E-03
1,60E-03
1,60E-03
1,60E-03
1,61E-03
1,61E-03
1,61E-03
1,61E-03
1,62E-03
1,62E-03
1,62E-03
1,62E-03
1,62E-03
1,63E-03
1,63E-03
1,63E-03
1,63E-03
1,63E-03
1,63E-03
1,63E-03
1,63E-03
1,63E-03
1,64E-03
1,65E-03
1,67E-03
1,67E-03
1,69E-03
1,69E-03
1,69E-03
1,69E-03
1,69E-03
1,69E-03
1,69E-03
1,69E-03
1,69E-03
1,69E-03
1,69E-03
1,69E-03
1,70E-03
1,70E-03
1,70E-03
1,71E-03
1,71E-03

1,71E-03
1,71E-03
1,71E-03
1,71E-03
1,71E-03
1,71E-03
1,71E-03
1,71E-03
1,71E-03
1,72E-03
1,72E-03
1,72E-03
1,73E-03
1,73E-03
1,73E-03
1,73E-03
1,73E-03
1,73E-03
1,73E-03
1,73E-03
1,73E-03
1,73E-03
1,74E-03
1,74E-03
1,74E-03
1,75E-03
1,75E-03
1,75E-03
1,76E-03
1,76E-03
1,76E-03
1,76E-03
1,76E-03
1,77E-03
1,77E-03
1,77E-03
1,77E-03
1,77E-03
1,78E-03
1,78E-03
1,78E-03
1,78E-03
1,78E-03
1,79E-03
1,80E-03
1,80E-03

1,80E-03
1,81E-03
1,81E-03
1,82E-03
1,83E-03
1,84E-03
1,84E-03
1,84E-03
1,85E-03
1,85E-03
1,85E-03
1,85E-03
1,85E-03
1,85E-03
1,86E-03
1,86E-03
1,87E-03
1,88E-03
1,88E-03
1,88E-03
1,88E-03
1,88E-03
1,88E-03
1,88E-03
1,88E-03
1,88E-03
1,88E-03
1,90E-03
1,90E-03
1,91E-03
1,91E-03
1,91E-03
1,91E-03
1,91E-03
1,91E-03
1,91E-03
1,91E-03
1,91E-03
1,91E-03
1,92E-03
1,92E-03
1,93E-03
1,93E-03
1,93E-03
1,94E-03
1,94E-03

1,94E-03
1,94E-03
1,94E-03
1,95E-03
1,95E-03
1,96E-03
1,96E-03
1,96E-03
1,96E-03
1,96E-03
1,96E-03
1,97E-03
1,97E-03
1,97E-03
1,97E-03
1,97E-03
1,98E-03
1,98E-03
1,99E-03
2,00E-03
2,02E-03
2,02E-03
2,02E-03
2,02E-03
2,03E-03
2,03E-03
2,03E-03
2,03E-03
2,03E-03
2,03E-03
2,03E-03
2,03E-03
2,03E-03
2,03E-03
2,03E-03
2,03E-03
2,05E-03
2,05E-03
2,05E-03
2,06E-03
2,06E-03
2,06E-03
2,07E-03
2,07E-03
2,07E-03
2,07E-03

2,07E-03
2,09E-03
2,09E-03
2,09E-03
2,09E-03
2,09E-03
2,09E-03
2,09E-03
2,09E-03
2,09E-03
2,09E-03
2,10E-03
2,10E-03
2,10E-03
2,10E-03
2,10E-03
2,10E-03
2,10E-03
2,10E-03
2,10E-03
2,10E-03
2,11E-03
2,11E-03
2,13E-03
2,13E-03
2,13E-03
2,13E-03
2,13E-03
2,13E-03
2,13E-03
2,14E-03
2,14E-03
2,14E-03
2,15E-03
2,15E-03
2,16E-03
2,16E-03
2,16E-03
2,16E-03
2,16E-03
2,16E-03
2,16E-03
2,16E-03
2,17E-03
2,19E-03
2,20E-03

2,21E-03
2,21E-03
2,21E-03
2,21E-03
2,22E-03
2,22E-03
2,22E-03
2,22E-03
2,22E-03
2,22E-03
2,22E-03
2,23E-03
2,23E-03
2,23E-03
2,23E-03
2,23E-03
2,23E-03
2,23E-03
2,23E-03
2,24E-03
2,25E-03
2,26E-03
2,26E-03
2,26E-03
2,26E-03
2,26E-03
2,26E-03
2,26E-03
2,26E-03
2,26E-03
2,26E-03
2,26E-03
2,27E-03
2,27E-03
2,27E-03
2,28E-03
2,28E-03
2,29E-03
2,29E-03
2,29E-03
2,29E-03
2,29E-03
2,29E-03
2,30E-03
2,31E-03
2,31E-03

2,31E-03
2,31E-03
2,31E-03
2,31E-03
2,31E-03
2,31E-03
2,31E-03
2,32E-03
2,32E-03
2,33E-03
2,33E-03
2,33E-03
2,33E-03
2,33E-03
2,33E-03
2,33E-03
2,33E-03
2,34E-03
2,34E-03
2,34E-03
2,35E-03
2,35E-03
2,35E-03
2,35E-03
2,35E-03
2,35E-03
2,37E-03
2,38E-03
2,40E-03
2,40E-03
2,41E-03
2,41E-03
2,41E-03
2,41E-03
2,41E-03
2,41E-03
2,42E-03
2,42E-03
2,44E-03
2,44E-03
2,44E-03
2,44E-03
2,44E-03
2,44E-03
2,44E-03
2,44E-03

2,44E-03
2,44E-03
2,46E-03
2,46E-03
2,46E-03
2,46E-03
2,47E-03
2,49E-03
2,50E-03
2,50E-03
2,51E-03
2,51E-03
2,51E-03
2,51E-03
2,54E-03
2,54E-03
2,55E-03
2,56E-03
2,56E-03
2,56E-03
2,56E-03
2,57E-03
2,57E-03
2,57E-03
2,57E-03
2,58E-03
2,58E-03
2,59E-03
2,59E-03
2,59E-03
2,59E-03
2,59E-03
2,59E-03
2,59E-03
2,59E-03
2,59E-03
2,59E-03
2,59E-03
2,59E-03
2,60E-03
2,60E-03
2,60E-03
2,60E-03
2,60E-03
2,61E-03
2,61E-03

2,61E-03
2,61E-03
2,61E-03
2,62E-03
2,62E-03
2,62E-03
2,63E-03
2,63E-03
2,63E-03
2,63E-03
2,63E-03
2,64E-03
2,64E-03
2,65E-03
2,65E-03
2,66E-03
2,66E-03
2,66E-03
2,66E-03
2,67E-03
2,67E-03
2,69E-03
2,69E-03
2,69E-03
2,70E-03
2,70E-03
2,72E-03
2,72E-03
2,72E-03
2,72E-03
2,72E-03
2,72E-03
2,72E-03
2,72E-03
2,72E-03
2,72E-03
2,72E-03
2,72E-03
2,73E-03
2,73E-03
2,74E-03
2,74E-03
2,74E-03
2,75E-03
2,75E-03
2,75E-03

2,75E-03
2,76E-03
2,76E-03
2,78E-03
2,78E-03
2,78E-03
2,79E-03
2,79E-03
2,80E-03
2,82E-03
2,82E-03
2,83E-03
2,83E-03
2,84E-03
2,84E-03
2,85E-03
2,85E-03
2,85E-03
2,85E-03
2,85E-03
2,85E-03
2,85E-03
2,87E-03
2,87E-03
2,87E-03
2,87E-03
2,87E-03
2,87E-03
2,88E-03
2,88E-03
2,88E-03
2,88E-03
2,90E-03
2,90E-03
2,90E-03
2,90E-03
2,90E-03
2,90E-03
2,90E-03
2,90E-03
2,90E-03
2,90E-03
2,90E-03
2,90E-03
2,90E-03
2,90E-03

2,90E-03
2,90E-03
2,91E-03
2,91E-03
2,92E-03
2,92E-03
2,92E-03
2,92E-03
2,92E-03
2,92E-03
2,92E-03
2,92E-03
2,92E-03
2,92E-03
2,92E-03
2,94E-03
2,94E-03
2,96E-03
2,98E-03
2,98E-03
2,98E-03
3,00E-03
3,02E-03
3,02E-03
3,03E-03
3,04E-03
3,04E-03
3,04E-03
3,04E-03
3,04E-03
3,07E-03
3,07E-03
3,08E-03
3,08E-03
3,08E-03
3,08E-03
3,09E-03
3,09E-03
3,09E-03
3,09E-03
3,12E-03
3,12E-03
3,12E-03
3,13E-03
3,14E-03
3,15E-03

3,15E-03
3,16E-03
3,16E-03
3,16E-03
3,16E-03
3,16E-03
3,16E-03
3,16E-03
3,17E-03
3,17E-03
3,17E-03
3,18E-03
3,18E-03
3,18E-03
3,18E-03
3,18E-03
3,18E-03
3,19E-03
3,19E-03
3,20E-03
3,20E-03
3,20E-03
3,20E-03
3,20E-03
3,20E-03
3,21E-03
3,22E-03
3,23E-03
3,23E-03
3,23E-03
3,23E-03
3,23E-03
3,24E-03
3,26E-03
3,26E-03
3,26E-03
3,26E-03
3,26E-03
3,26E-03
3,26E-03
3,26E-03
3,26E-03
3,27E-03
3,27E-03
3,27E-03
3,28E-03

3,28E-03
3,28E-03
3,30E-03
3,30E-03
3,30E-03
3,32E-03
3,32E-03
3,32E-03
3,33E-03
3,33E-03
3,33E-03
3,35E-03
3,35E-03
3,36E-03
3,36E-03
3,36E-03
3,36E-03
3,37E-03
3,38E-03
3,38E-03
3,38E-03
3,40E-03
3,40E-03
3,40E-03
3,41E-03
3,43E-03
3,43E-03
3,43E-03
3,44E-03
3,45E-03
3,48E-03
3,49E-03
3,49E-03
3,49E-03
3,49E-03
3,51E-03
3,51E-03
3,54E-03
3,54E-03
3,54E-03
3,54E-03
3,56E-03
3,56E-03
3,56E-03
3,56E-03
3,57E-03

3,57E-03
3,57E-03
3,57E-03
3,57E-03
3,58E-03
3,58E-03
3,59E-03
3,59E-03
3,60E-03
3,60E-03
3,60E-03
3,61E-03
3,61E-03
3,61E-03
3,62E-03
3,62E-03
3,62E-03
3,62E-03
3,62E-03
3,66E-03
3,67E-03
3,67E-03
3,67E-03
3,67E-03
3,67E-03
3,68E-03
3,68E-03
3,68E-03
3,68E-03
3,69E-03
3,73E-03
3,73E-03
3,75E-03
3,76E-03
3,77E-03
3,77E-03
3,77E-03
3,77E-03
3,77E-03
3,77E-03
3,77E-03
3,77E-03
3,77E-03
3,79E-03
3,79E-03
3,79E-03

3,81E-03
3,82E-03
3,82E-03
3,82E-03
3,82E-03
3,82E-03
3,83E-03
3,83E-03
3,83E-03
3,84E-03
3,85E-03
3,85E-03
3,85E-03
3,85E-03
3,85E-03
3,86E-03
3,86E-03
3,91E-03
3,93E-03
3,93E-03
3,94E-03
3,94E-03
3,95E-03
3,95E-03
3,95E-03
3,95E-03
3,95E-03
3,96E-03
3,98E-03
3,99E-03
4,01E-03
4,01E-03
4,01E-03
4,01E-03
4,03E-03
4,03E-03
4,04E-03
4,06E-03
4,06E-03
4,06E-03
4,06E-03
4,07E-03
4,07E-03
4,07E-03
4,08E-03
4,09E-03

4,09E-03
4,09E-03
4,11E-03
4,11E-03
4,11E-03
4,11E-03
4,11E-03
4,11E-03
4,11E-03
4,11E-03
4,11E-03
4,11E-03
4,11E-03
4,12E-03
4,13E-03
4,13E-03
4,13E-03
4,13E-03
4,14E-03
4,14E-03
4,15E-03
4,15E-03
4,16E-03
4,16E-03
4,16E-03
4,16E-03
4,16E-03
4,17E-03
4,19E-03
4,19E-03
4,19E-03
4,19E-03
4,19E-03
4,19E-03
4,20E-03
4,20E-03
4,20E-03
4,20E-03
4,20E-03
4,21E-03
4,21E-03
4,23E-03
4,24E-03
4,24E-03
4,24E-03
4,24E-03

4,24E-03
4,24E-03
4,25E-03
4,25E-03
4,27E-03
4,28E-03
4,28E-03
4,28E-03
4,28E-03
4,28E-03
4,29E-03
4,30E-03
4,31E-03
4,31E-03
4,31E-03
4,32E-03
4,32E-03
4,32E-03
4,33E-03
4,34E-03
4,35E-03
4,36E-03
4,36E-03
4,37E-03
4,37E-03
4,38E-03
4,40E-03
4,41E-03
4,42E-03
4,42E-03
4,43E-03
4,43E-03
4,44E-03
4,44E-03
4,45E-03
4,45E-03
4,48E-03
4,50E-03
4,51E-03
4,51E-03
4,52E-03
4,53E-03
4,53E-03
4,54E-03
4,54E-03
4,54E-03

4,54E-03
4,56E-03
4,59E-03
4,59E-03
4,60E-03
4,61E-03
4,61E-03
4,65E-03
4,65E-03
4,65E-03
4,66E-03
4,66E-03
4,66E-03
4,66E-03
4,67E-03
4,67E-03
4,67E-03
4,67E-03
4,72E-03
4,72E-03
4,72E-03
4,72E-03
4,72E-03
4,72E-03
4,72E-03
4,72E-03
4,72E-03
4,72E-03
4,74E-03
4,74E-03
4,74E-03
4,74E-03
4,74E-03
4,75E-03
4,75E-03
4,75E-03
4,77E-03
4,78E-03
4,78E-03
4,78E-03
4,79E-03
4,79E-03
4,82E-03
4,82E-03
4,82E-03
4,82E-03

4,82E-03
4,84E-03
4,85E-03
4,85E-03
4,85E-03
4,86E-03
4,86E-03
4,88E-03
4,88E-03
4,88E-03
4,88E-03
4,88E-03
4,88E-03
4,88E-03
4,90E-03
4,90E-03
4,91E-03
4,91E-03
4,91E-03
4,92E-03
4,92E-03
4,94E-03
4,95E-03
4,97E-03
4,97E-03
4,98E-03
4,98E-03
4,99E-03
5,01E-03
5,01E-03
5,01E-03
5,01E-03
5,01E-03
5,04E-03
5,05E-03
5,05E-03
5,05E-03
5,06E-03
5,06E-03
5,08E-03
5,08E-03
5,09E-03
5,14E-03
5,14E-03
5,14E-03
5,14E-03

5,14E-03
5,14E-03
5,14E-03
5,14E-03
5,14E-03
5,15E-03
5,16E-03
5,17E-03
5,17E-03
5,17E-03
5,17E-03
5,17E-03
5,17E-03
5,17E-03
5,17E-03
5,18E-03
5,18E-03
5,18E-03
5,18E-03
5,18E-03
5,19E-03
5,22E-03
5,22E-03
5,22E-03
5,22E-03
5,23E-03
5,23E-03
5,23E-03
5,24E-03
5,26E-03
5,27E-03
5,29E-03
5,29E-03
5,29E-03
5,29E-03
5,29E-03
5,29E-03
5,29E-03
5,29E-03
5,29E-03
5,30E-03
5,34E-03
5,34E-03
5,34E-03
5,36E-03
5,36E-03

5,36E-03
5,40E-03
5,40E-03
5,40E-03
5,40E-03
5,40E-03
5,41E-03
5,41E-03
5,41E-03
5,42E-03
5,46E-03
5,46E-03
5,46E-03
5,47E-03
5,47E-03
5,47E-03
5,47E-03
5,47E-03
5,47E-03
5,48E-03
5,48E-03
5,51E-03
5,51E-03
5,51E-03
5,51E-03
5,55E-03
5,55E-03
5,56E-03
5,56E-03
5,56E-03
5,56E-03
5,56E-03
5,58E-03
5,59E-03
5,59E-03
5,59E-03
5,59E-03
5,60E-03
5,62E-03
5,63E-03
5,63E-03
5,64E-03
5,64E-03
5,64E-03
5,64E-03
5,64E-03

5,64E-03
5,64E-03
5,64E-03
5,64E-03
5,70E-03
5,70E-03
5,70E-03
5,71E-03
5,73E-03
5,73E-03
5,75E-03
5,77E-03
5,77E-03
5,77E-03
5,79E-03
5,80E-03
5,80E-03
5,82E-03
5,82E-03
5,82E-03
5,82E-03
5,82E-03
5,82E-03
5,82E-03
5,83E-03
5,83E-03
5,85E-03
5,89E-03
5,90E-03
5,90E-03
5,91E-03
5,93E-03
5,93E-03
5,93E-03
5,94E-03
5,94E-03
5,94E-03
5,94E-03
5,94E-03
5,94E-03
6,00E-03
6,00E-03
6,00E-03
6,00E-03
6,01E-03
6,02E-03

6,02E-03
6,06E-03
6,07E-03
6,10E-03
6,10E-03
6,12E-03
6,12E-03
6,16E-03
6,16E-03
6,16E-03
6,16E-03
6,17E-03
6,18E-03
6,18E-03
6,20E-03
6,20E-03
6,21E-03
6,21E-03
6,21E-03
6,23E-03
6,23E-03
6,23E-03
6,24E-03
6,25E-03
6,25E-03
6,25E-03
6,25E-03
6,25E-03
6,26E-03
6,26E-03
6,26E-03
6,28E-03
6,28E-03
6,28E-03
6,28E-03
6,29E-03
6,29E-03
6,30E-03
6,31E-03
6,31E-03
6,31E-03
6,34E-03
6,34E-03
6,34E-03
6,35E-03
6,36E-03

6,36E-03
6,37E-03
6,37E-03
6,37E-03
6,37E-03
6,37E-03
6,37E-03
6,38E-03
6,38E-03
6,38E-03
6,38E-03
6,39E-03
6,39E-03
6,39E-03
6,39E-03
6,39E-03
6,43E-03
6,43E-03
6,43E-03
6,44E-03
6,44E-03
6,44E-03
6,44E-03
6,44E-03
6,44E-03
6,44E-03
6,44E-03
6,46E-03
6,46E-03
6,47E-03
6,47E-03
6,47E-03
6,47E-03
6,48E-03
6,49E-03
6,51E-03
6,51E-03
6,53E-03
6,53E-03
6,53E-03
6,53E-03
6,53E-03
6,54E-03
6,54E-03
6,55E-03
6,55E-03

6,55E-03
6,55E-03
6,56E-03
6,56E-03
6,56E-03
6,57E-03
6,57E-03
6,57E-03
6,58E-03
6,58E-03
6,58E-03
6,58E-03
6,59E-03
6,60E-03
6,60E-03
6,61E-03
6,62E-03
6,62E-03
6,62E-03
6,64E-03
6,64E-03
6,64E-03
6,65E-03
6,65E-03
6,66E-03
6,67E-03
6,67E-03
6,67E-03
6,68E-03
6,69E-03
6,71E-03
6,71E-03
6,72E-03
6,74E-03
6,75E-03
6,76E-03
6,76E-03
6,77E-03
6,77E-03
6,79E-03
6,80E-03
6,80E-03
6,81E-03
6,81E-03
6,82E-03
6,83E-03

6,84E-03
6,84E-03
6,84E-03
6,87E-03
6,94E-03
6,95E-03
6,95E-03
6,98E-03
6,98E-03
7,01E-03
7,03E-03
7,03E-03
7,03E-03
7,05E-03
7,05E-03
7,05E-03
7,06E-03
7,07E-03
7,07E-03
7,07E-03
7,07E-03
7,08E-03
7,11E-03
7,11E-03
7,11E-03
7,11E-03
7,11E-03
7,11E-03
7,11E-03
7,11E-03
7,11E-03
7,11E-03
7,12E-03
7,12E-03
7,13E-03
7,13E-03
7,13E-03
7,13E-03
7,16E-03
7,16E-03
7,16E-03
7,17E-03
7,17E-03
7,17E-03
7,17E-03
7,19E-03

7,21E-03
7,22E-03
7,22E-03
7,23E-03
7,25E-03
7,25E-03
7,25E-03
7,25E-03
7,26E-03
7,26E-03
7,27E-03
7,28E-03
7,33E-03
7,33E-03
7,33E-03
7,35E-03
7,35E-03
7,35E-03
7,36E-03
7,36E-03
7,37E-03
7,39E-03
7,44E-03
7,44E-03
7,45E-03
7,45E-03
7,45E-03
7,45E-03
7,45E-03
7,45E-03
7,45E-03
7,46E-03
7,46E-03
7,46E-03
7,46E-03
7,46E-03
7,49E-03
7,50E-03
7,51E-03
7,51E-03
7,52E-03
7,52E-03
7,52E-03
7,52E-03
7,52E-03
7,55E-03

7,55E-03
7,55E-03
7,55E-03
7,59E-03
7,60E-03
7,60E-03
7,61E-03
7,63E-03
7,63E-03
7,63E-03
7,65E-03
7,67E-03
7,68E-03
7,68E-03
7,69E-03
7,69E-03
7,69E-03
7,69E-03
7,71E-03
7,72E-03
7,75E-03
7,75E-03
7,75E-03
7,75E-03
7,75E-03
7,75E-03
7,77E-03
7,77E-03
7,78E-03
7,78E-03
7,80E-03
7,82E-03
7,84E-03
7,84E-03
7,84E-03
7,85E-03
7,86E-03
7,86E-03
7,86E-03
7,87E-03
7,87E-03
7,88E-03
7,89E-03
7,89E-03
7,89E-03
7,90E-03

7,90E-03
7,92E-03
7,93E-03
7,94E-03
7,98E-03
8,03E-03
8,04E-03
8,04E-03
8,06E-03
8,08E-03
8,08E-03
8,09E-03
8,10E-03
8,10E-03
8,12E-03
8,12E-03
8,12E-03
8,13E-03
8,13E-03
8,13E-03
8,14E-03
8,14E-03
8,15E-03
8,15E-03
8,15E-03
8,15E-03
8,15E-03
8,15E-03
8,16E-03
8,20E-03
8,20E-03
8,20E-03
8,20E-03
8,20E-03
8,22E-03
8,26E-03
8,26E-03
8,27E-03
8,27E-03
8,27E-03
8,27E-03
8,27E-03
8,28E-03
8,28E-03
8,29E-03
8,30E-03

8,30E-03
8,31E-03
8,32E-03
8,34E-03
8,35E-03
8,35E-03
8,35E-03
8,35E-03
8,35E-03
8,35E-03
8,35E-03
8,36E-03
8,36E-03
8,41E-03
8,41E-03
8,42E-03
8,43E-03
8,46E-03
8,46E-03
8,46E-03
8,46E-03
8,46E-03
8,47E-03
8,49E-03
8,49E-03
8,50E-03
8,50E-03
8,51E-03
8,51E-03
8,51E-03
8,51E-03
8,52E-03
8,53E-03
8,53E-03
8,53E-03
8,53E-03
8,54E-03
8,56E-03
8,57E-03
8,59E-03
8,60E-03
8,60E-03
8,60E-03
8,60E-03
8,61E-03
8,63E-03

8,63E-03
8,63E-03
8,63E-03
8,63E-03
8,65E-03
8,68E-03
8,68E-03
8,70E-03
8,70E-03
8,70E-03
8,72E-03
8,72E-03
8,72E-03
8,72E-03
8,73E-03
8,73E-03
8,73E-03
8,73E-03
8,73E-03
8,74E-03
8,74E-03
8,74E-03
8,74E-03
8,75E-03
8,78E-03
8,78E-03
8,78E-03
8,78E-03
8,78E-03
8,80E-03
8,80E-03
8,81E-03
8,81E-03
8,81E-03
8,81E-03
8,82E-03
8,83E-03
8,83E-03
8,86E-03
8,86E-03
8,87E-03
8,87E-03
8,87E-03
8,88E-03
8,92E-03
8,92E-03

8,92E-03
8,92E-03
8,92E-03
8,92E-03
8,94E-03
8,96E-03
8,98E-03
8,98E-03
8,99E-03
8,99E-03
9,01E-03
9,02E-03
9,04E-03
9,06E-03
9,07E-03
9,07E-03
9,07E-03
9,10E-03
9,12E-03
9,12E-03
9,12E-03
9,13E-03
9,13E-03
9,13E-03
9,14E-03
9,15E-03
9,17E-03
9,19E-03
9,21E-03
9,21E-03
9,24E-03
9,24E-03
9,24E-03
9,26E-03
9,27E-03
9,27E-03
9,27E-03
9,27E-03
9,29E-03
9,29E-03
9,29E-03
9,30E-03
9,31E-03
9,31E-03
9,32E-03
9,32E-03

9,33E-03
9,33E-03
9,41E-03
9,41E-03
9,41E-03
9,41E-03
9,41E-03
9,41E-03
9,42E-03
9,48E-03
9,48E-03
9,48E-03
9,53E-03
9,56E-03
9,57E-03
9,60E-03
9,60E-03
9,60E-03
9,60E-03
9,60E-03
9,60E-03
9,60E-03
9,60E-03
9,60E-03
9,60E-03
9,60E-03
9,60E-03
9,62E-03
9,63E-03
9,63E-03
9,65E-03
9,67E-03
9,70E-03
9,71E-03
9,71E-03
9,73E-03
9,74E-03
9,74E-03
9,75E-03
9,81E-03
9,82E-03
9,84E-03
9,85E-03
9,85E-03
9,86E-03
9,87E-03

9,88E-03
9,88E-03
9,88E-03
9,88E-03
9,88E-03
9,90E-03
9,90E-03
9,90E-03
9,93E-03
9,94E-03
9,94E-03
9,95E-03
9,99E-03
9,99E-03
1,00E-02
1,00E-02
1,01E-02
1,01E-02
1,01E-02
1,01E-02
1,01E-02
1,01E-02
1,01E-02
1,01E-02
1,01E-02
1,01E-02
1,02E-02
1,02E-02
1,02E-02
1,02E-02
1,02E-02
1,02E-02
1,02E-02
1,02E-02
1,02E-02
1,03E-02
1,03E-02
1,03E-02
1,03E-02
1,03E-02
1,03E-02
1,03E-02
1,03E-02
1,03E-02
1,03E-02
1,04E-02

1,04E-02
1,04E-02
1,04E-02
1,04E-02
1,04E-02
1,04E-02
1,04E-02
1,04E-02
1,05E-02
1,05E-02
1,05E-02
1,05E-02
1,05E-02
1,05E-02
1,05E-02
1,05E-02
1,05E-02
1,05E-02
1,05E-02
1,05E-02
1,05E-02
1,06E-02
1,06E-02
1,06E-02
1,06E-02
1,06E-02
1,06E-02
1,06E-02
1,06E-02
1,06E-02
1,07E-02
1,07E-02
1,07E-02
1,07E-02
1,07E-02
1,07E-02
1,07E-02
1,07E-02
1,07E-02
1,07E-02
1,08E-02
1,08E-02
1,08E-02
1,08E-02
1,08E-02
1,08E-02

1,08E-02
1,08E-02
1,08E-02
1,09E-02
1,09E-02
1,09E-02
1,09E-02
1,09E-02
1,09E-02
1,09E-02
1,09E-02
1,09E-02
1,09E-02
1,09E-02
1,09E-02
1,09E-02
1,10E-02
1,10E-02
1,10E-02
1,10E-02
1,10E-02
1,10E-02
1,10E-02
1,11E-02
1,11E-02
1,11E-02
1,11E-02
1,11E-02
1,12E-02
1,12E-02
1,12E-02
1,12E-02
1,12E-02
1,12E-02
1,12E-02
1,12E-02
1,12E-02
1,12E-02
1,12E-02
1,13E-02
1,13E-02
1,13E-02
1,13E-02
1,13E-02
1,13E-02
1,13E-02

1,13E-02
1,13E-02
1,13E-02
1,13E-02
1,13E-02
1,13E-02
1,13E-02
1,13E-02
1,14E-02
1,14E-02
1,14E-02
1,14E-02
1,14E-02
1,14E-02
1,14E-02
1,15E-02
1,15E-02
1,15E-02
1,15E-02
1,15E-02
1,15E-02
1,15E-02
1,15E-02
1,16E-02
1,16E-02
1,16E-02
1,16E-02
1,16E-02
1,16E-02
1,16E-02
1,16E-02
1,17E-02
1,17E-02
1,17E-02
1,17E-02
1,17E-02
1,17E-02
1,17E-02
1,17E-02
1,17E-02
1,18E-02
1,18E-02
1,18E-02
1,18E-02
1,18E-02
1,18E-02

| |
|---|
| 1,18E-02 |
| 1,18E-02 |
| 1,18E-02 |
| 1,18E-02 |
| 1,18E-02 |
| 1,18E-02 |
| 1,19E-02 |
| 1,19E-02 |
| 1,19E-02 |
| 1,19E-02 |
| 1,19E-02 |
| 1,19E-02 |
| 1,20E-02 |
| 1,20E-02 |
| 1,20E-02 |
| 1,20E-02 |
| 1,20E-02 |
| 1,20E-02 |
| 1,20E-02 |
| 1,20E-02 |
| 1,20E-02 |
| 1,20E-02 |
| 1,20E-02 |
| 1,20E-02 |
| 1,20E-02 |
| 1,21E-02 |
| 1,21E-02 |
| 1,21E-02 |
| 1,21E-02 |
| 1,21E-02 |
| 1,21E-02 |
| 1,21E-02 |
| 1,21E-02 |
| 1,21E-02 |
| 1,21E-02 |
| 1,21E-02 |
| 1,21E-02 |
| 1,21E-02 |
| 1,21E-02 |
| 1,22E-02 |
| 1,22E-02 |
| 1,22E-02 |
| 1,22E-02 |
| 1,22E-02 |
| 1,22E-02 |
| 1,22E-02 |

| |
|---|
| 1,22E-02 |
| 1,22E-02 |
| 1,22E-02 |
| 1,22E-02 |
| 1,22E-02 |
| 1,23E-02 |
| 1,23E-02 |
| 1,23E-02 |
| 1,23E-02 |
| 1,23E-02 |
| 1,23E-02 |
| 1,23E-02 |
| 1,23E-02 |
| 1,23E-02 |
| 1,23E-02 |
| 1,23E-02 |
| 1,23E-02 |
| 1,23E-02 |
| 1,23E-02 |
| 1,23E-02 |
| 1,23E-02 |
| 1,23E-02 |
| 1,24E-02 |
| 1,24E-02 |
| 1,24E-02 |
| 1,24E-02 |
| 1,24E-02 |
| 1,24E-02 |
| 1,24E-02 |
| 1,24E-02 |
| 1,24E-02 |
| 1,24E-02 |
| 1,24E-02 |
| 1,24E-02 |
| 1,24E-02 |
| 1,25E-02 |
| 1,25E-02 |
| 1,25E-02 |
| 1,25E-02 |
| 1,25E-02 |
| 1,25E-02 |
| 1,25E-02 |
| 1,25E-02 |
| 1,25E-02 |
| 1,25E-02 |
| 1,25E-02 |

| |
|---|
| 1,25E-02 |
| 1,25E-02 |
| 1,26E-02 |
| 1,26E-02 |
| 1,27E-02 |
| 1,27E-02 |
| 1,27E-02 |
| 1,27E-02 |
| 1,27E-02 |
| 1,27E-02 |
| 1,27E-02 |
| 1,27E-02 |
| 1,27E-02 |
| 1,27E-02 |
| 1,28E-02 |
| 1,28E-02 |
| 1,28E-02 |
| 1,28E-02 |
| 1,28E-02 |
| 1,29E-02 |
| 1,29E-02 |
| 1,29E-02 |
| 1,29E-02 |
| 1,29E-02 |
| 1,29E-02 |
| 1,29E-02 |
| 1,29E-02 |
| 1,29E-02 |
| 1,29E-02 |
| 1,29E-02 |
| 1,29E-02 |
| 1,30E-02 |
| 1,30E-02 |
| 1,30E-02 |
| 1,30E-02 |
| 1,30E-02 |
| 1,31E-02 |
| 1,31E-02 |
| 1,31E-02 |
| 1,31E-02 |
| 1,31E-02 |
| 1,31E-02 |
| 1,31E-02 |
| 1,31E-02 |
| 1,31E-02 |
| 1,31E-02 |

1,31E-02
1,31E-02
1,31E-02
1,31E-02
1,32E-02
1,32E-02
1,32E-02
1,32E-02
1,32E-02
1,32E-02
1,33E-02
1,33E-02
1,33E-02
1,33E-02
1,33E-02
1,33E-02
1,33E-02
1,33E-02
1,33E-02
1,33E-02
1,33E-02
1,33E-02
1,34E-02
1,34E-02
1,34E-02
1,34E-02
1,34E-02
1,34E-02
1,34E-02
1,35E-02
1,35E-02
1,35E-02
1,35E-02
1,35E-02
1,35E-02
1,35E-02
1,35E-02
1,36E-02
1,36E-02
1,36E-02
1,36E-02
1,36E-02
1,36E-02
1,36E-02
1,36E-02
1,36E-02

1,36E-02
1,36E-02
1,36E-02
1,37E-02
1,37E-02
1,37E-02
1,37E-02
1,37E-02
1,37E-02
1,37E-02
1,37E-02
1,37E-02
1,37E-02
1,37E-02
1,38E-02
1,38E-02
1,38E-02
1,38E-02
1,38E-02
1,38E-02
1,39E-02
1,39E-02
1,39E-02
1,39E-02
1,39E-02
1,39E-02
1,39E-02
1,39E-02
1,39E-02
1,40E-02
1,40E-02
1,40E-02
1,40E-02
1,40E-02
1,40E-02
1,40E-02
1,40E-02
1,40E-02
1,40E-02
1,40E-02
1,41E-02
1,41E-02
1,41E-02
1,41E-02
1,41E-02
1,41E-02

1,41E-02
1,42E-02
1,42E-02
1,42E-02
1,42E-02
1,42E-02
1,42E-02
1,42E-02
1,42E-02
1,42E-02
1,42E-02
1,42E-02
1,42E-02
1,42E-02
1,42E-02
1,42E-02
1,43E-02
1,43E-02
1,43E-02
1,43E-02
1,43E-02
1,43E-02
1,43E-02
1,43E-02
1,43E-02
1,43E-02
1,44E-02
1,44E-02
1,44E-02
1,44E-02
1,44E-02
1,44E-02
1,44E-02
1,44E-02
1,44E-02
1,44E-02
1,44E-02
1,44E-02
1,44E-02
1,44E-02
1,44E-02
1,44E-02
1,44E-02
1,44E-02
1,44E-02
1,44E-02

1,44E-02
1,44E-02
1,44E-02
1,45E-02
1,45E-02
1,45E-02
1,45E-02
1,45E-02
1,45E-02
1,46E-02
1,46E-02
1,46E-02
1,46E-02
1,46E-02
1,46E-02
1,46E-02
1,46E-02
1,46E-02
1,46E-02
1,46E-02
1,46E-02
1,46E-02
1,46E-02
1,47E-02
1,47E-02
1,47E-02
1,47E-02
1,47E-02
1,47E-02
1,48E-02
1,48E-02
1,48E-02
1,48E-02
1,48E-02
1,48E-02
1,48E-02
1,48E-02
1,48E-02
1,48E-02
1,48E-02
1,48E-02
1,49E-02
1,49E-02
1,49E-02
1,49E-02
1,49E-02

1,49E-02
1,49E-02
1,50E-02
1,50E-02
1,50E-02
1,50E-02
1,50E-02
1,50E-02
1,50E-02
1,50E-02
1,50E-02
1,51E-02
1,51E-02
1,51E-02
1,51E-02
1,51E-02
1,51E-02
1,51E-02
1,52E-02
1,52E-02
1,52E-02
1,52E-02
1,52E-02
1,52E-02
1,52E-02
1,52E-02
1,52E-02
1,53E-02
1,53E-02
1,53E-02
1,53E-02
1,53E-02
1,53E-02
1,53E-02
1,54E-02
1,54E-02
1,54E-02
1,54E-02
1,54E-02
1,54E-02
1,55E-02
1,55E-02
1,55E-02
1,55E-02
1,55E-02
1,55E-02

| |
|---|
| 1,55E-02 |
| 1,55E-02 |
| 1,56E-02 |
| 1,56E-02 |
| 1,56E-02 |
| 1,56E-02 |
| 1,56E-02 |
| 1,56E-02 |
| 1,56E-02 |
| 1,56E-02 |
| 1,56E-02 |
| 1,56E-02 |
| 1,56E-02 |
| 1,56E-02 |
| 1,56E-02 |
| 1,57E-02 |
| 1,58E-02 |
| 1,58E-02 |
| 1,58E-02 |
| 1,58E-02 |
| 1,58E-02 |
| 1,58E-02 |
| 1,58E-02 |
| 1,58E-02 |
| 1,58E-02 |
| 1,58E-02 |
| 1,59E-02 |
| 1,59E-02 |
| 1,59E-02 |
| 1,59E-02 |
| 1,59E-02 |
| 1,59E-02 |
| 1,59E-02 |
| 1,60E-02 |
| 1,60E-02 |
| 1,60E-02 |
| 1,60E-02 |
| 1,60E-02 |
| 1,61E-02 |
| 1,61E-02 |
| 1,62E-02 |
| 1,62E-02 |
| 1,62E-02 |
| 1,62E-02 |
| 1,62E-02 |
| 1,62E-02 |

1,62E-02
1,62E-02
1,62E-02
1,62E-02
1,62E-02
1,62E-02
1,63E-02
1,63E-02
1,63E-02
1,63E-02
1,63E-02
1,63E-02
1,63E-02
1,63E-02
1,63E-02
1,64E-02
1,64E-02
1,64E-02
1,64E-02
1,64E-02
1,64E-02
1,64E-02
1,64E-02
1,65E-02
1,65E-02
1,65E-02
1,65E-02
1,65E-02
1,65E-02
1,65E-02
1,65E-02
1,65E-02
1,65E-02
1,65E-02
1,65E-02
1,65E-02
1,65E-02
1,65E-02
1,66E-02
1,66E-02
1,66E-02
1,66E-02
1,66E-02
1,66E-02
1,66E-02
1,66E-02

1,66E-02
1,66E-02
1,67E-02
1,67E-02
1,67E-02
1,67E-02
1,67E-02
1,67E-02
1,67E-02
1,67E-02
1,67E-02
1,67E-02
1,67E-02
1,67E-02
1,67E-02
1,67E-02
1,68E-02
1,68E-02
1,68E-02
1,68E-02
1,68E-02
1,68E-02
1,68E-02
1,68E-02
1,69E-02
1,69E-02
1,69E-02
1,69E-02
1,69E-02
1,69E-02
1,69E-02
1,69E-02
1,70E-02
1,70E-02
1,70E-02
1,70E-02
1,70E-02
1,70E-02
1,71E-02
1,71E-02
1,71E-02
1,71E-02
1,72E-02
1,72E-02
1,72E-02
1,72E-02

1,73E-02
1,73E-02
1,73E-02
1,73E-02
1,74E-02
1,74E-02
1,74E-02
1,74E-02
1,74E-02
1,74E-02
1,74E-02
1,74E-02
1,75E-02
1,75E-02
1,75E-02
1,75E-02
1,75E-02
1,75E-02
1,75E-02
1,75E-02
1,76E-02
1,76E-02
1,76E-02
1,76E-02
1,76E-02
1,76E-02
1,78E-02
1,78E-02
1,78E-02
1,78E-02
1,78E-02
1,78E-02
1,78E-02
1,78E-02
1,78E-02
1,78E-02
1,79E-02
1,79E-02
1,79E-02
1,79E-02
1,79E-02
1,79E-02
1,79E-02
1,79E-02
1,79E-02
1,79E-02

1,80E-02
1,80E-02
1,80E-02
1,80E-02
1,81E-02
1,81E-02
1,81E-02
1,81E-02
1,81E-02
1,82E-02
1,82E-02
1,82E-02
1,82E-02
1,82E-02
1,83E-02
1,83E-02
1,83E-02
1,83E-02
1,83E-02
1,83E-02
1,84E-02
1,84E-02
1,84E-02
1,84E-02
1,85E-02
1,85E-02
1,85E-02
1,85E-02
1,85E-02
1,85E-02
1,85E-02
1,85E-02
1,85E-02
1,85E-02
1,86E-02
1,86E-02
1,86E-02
1,86E-02
1,86E-02
1,86E-02
1,86E-02
1,86E-02
1,87E-02
1,87E-02
1,87E-02
1,87E-02

1,87E-02
1,87E-02
1,88E-02
1,88E-02
1,88E-02
1,88E-02
1,88E-02
1,88E-02
1,88E-02
1,88E-02
1,88E-02
1,88E-02
1,88E-02
1,88E-02
1,89E-02
1,89E-02
1,89E-02
1,89E-02
1,90E-02
1,90E-02
1,90E-02
1,90E-02
1,90E-02
1,91E-02
1,91E-02
1,91E-02
1,91E-02
1,91E-02
1,91E-02
1,92E-02
1,92E-02
1,92E-02
1,92E-02
1,92E-02
1,92E-02
1,92E-02
1,92E-02
1,93E-02
1,94E-02
1,94E-02
1,94E-02
1,94E-02
1,95E-02
1,95E-02
1,95E-02
1,95E-02

| |
|---|
| 1,96E-02 |
| 1,96E-02 |
| 1,96E-02 |
| 1,96E-02 |
| 1,96E-02 |
| 1,96E-02 |
| 1,96E-02 |
| 1,96E-02 |
| 1,96E-02 |
| 1,97E-02 |
| 1,97E-02 |
| 1,97E-02 |
| 1,97E-02 |
| 1,97E-02 |
| 1,98E-02 |
| 1,99E-02 |
| 1,99E-02 |
| 1,99E-02 |
| 2,00E-02 |
| 2,00E-02 |
| 2,00E-02 |
| 2,01E-02 |
| 2,01E-02 |
| 2,01E-02 |
| 2,01E-02 |
| 2,01E-02 |
| 2,01E-02 |
| 2,01E-02 |
| 2,01E-02 |
| 2,01E-02 |
| 2,02E-02 |
| 2,02E-02 |
| 2,02E-02 |
| 2,02E-02 |
| 2,02E-02 |
| 2,02E-02 |
| 2,02E-02 |
| 2,02E-02 |
| 2,03E-02 |
| 2,03E-02 |
| 2,03E-02 |
| 2,04E-02 |
| 2,04E-02 |
| 2,04E-02 |
| 2,04E-02 |
| 2,04E-02 |

2,05E-02
2,05E-02
2,05E-02
2,05E-02
2,06E-02
2,06E-02
2,06E-02
2,06E-02
2,06E-02
2,06E-02
2,06E-02
2,06E-02
2,06E-02
2,06E-02
2,06E-02
2,06E-02
2,06E-02
2,07E-02
2,07E-02
2,07E-02
2,07E-02
2,07E-02
2,07E-02
2,07E-02
2,07E-02
2,07E-02
2,07E-02
2,07E-02
2,07E-02
2,07E-02
2,07E-02
2,07E-02
2,07E-02
2,08E-02
2,08E-02
2,08E-02
2,08E-02
2,08E-02
2,09E-02
2,09E-02
2,09E-02
2,09E-02
2,10E-02
2,10E-02
2,11E-02
2,11E-02

| |
|---|
| 2,11E-02 |
| 2,11E-02 |
| 2,11E-02 |
| 2,11E-02 |
| 2,11E-02 |
| 2,11E-02 |
| 2,11E-02 |
| 2,11E-02 |
| 2,11E-02 |
| 2,11E-02 |
| 2,11E-02 |
| 2,11E-02 |
| 2,12E-02 |
| 2,12E-02 |
| 2,12E-02 |
| 2,12E-02 |
| 2,12E-02 |
| 2,12E-02 |
| 2,12E-02 |
| 2,12E-02 |
| 2,12E-02 |
| 2,13E-02 |
| 2,13E-02 |
| 2,13E-02 |
| 2,13E-02 |
| 2,13E-02 |
| 2,13E-02 |
| 2,13E-02 |
| 2,13E-02 |
| 2,13E-02 |
| 2,13E-02 |
| 2,13E-02 |
| 2,14E-02 |
| 2,14E-02 |
| 2,14E-02 |
| 2,14E-02 |
| 2,14E-02 |
| 2,14E-02 |
| 2,14E-02 |
| 2,14E-02 |
| 2,14E-02 |
| 2,14E-02 |
| 2,14E-02 |
| 2,14E-02 |
| 2,14E-02 |
| 2,14E-02 |

2,14E-02
2,14E-02
2,14E-02
2,14E-02
2,15E-02
2,15E-02
2,15E-02
2,16E-02
2,16E-02
2,16E-02
2,16E-02
2,17E-02
2,17E-02
2,17E-02
2,17E-02
2,17E-02
2,17E-02
2,17E-02
2,17E-02
2,17E-02
2,18E-02
2,18E-02
2,18E-02
2,19E-02
2,19E-02
2,19E-02
2,19E-02
2,19E-02
2,19E-02
2,19E-02
2,20E-02
2,20E-02
2,20E-02
2,20E-02
2,20E-02
2,20E-02
2,20E-02
2,20E-02
2,20E-02
2,20E-02
2,20E-02
2,20E-02
2,20E-02
2,20E-02
2,20E-02
2,20E-02

2,20E-02
2,20E-02
2,20E-02
2,20E-02
2,21E-02
2,21E-02
2,21E-02
2,21E-02
2,21E-02
2,21E-02
2,21E-02
2,22E-02
2,22E-02
2,22E-02
2,22E-02
2,22E-02
2,22E-02
2,23E-02
2,24E-02
2,24E-02
2,24E-02
2,24E-02
2,24E-02
2,24E-02
2,24E-02
2,24E-02
2,24E-02
2,24E-02
2,24E-02
2,24E-02
2,24E-02
2,25E-02
2,25E-02
2,25E-02
2,25E-02
2,25E-02
2,26E-02
2,26E-02
2,26E-02
2,26E-02
2,26E-02
2,26E-02
2,26E-02
2,26E-02
2,26E-02
2,26E-02

2,26E-02
2,26E-02
2,26E-02
2,26E-02
2,26E-02
2,26E-02
2,27E-02
2,27E-02
2,27E-02
2,27E-02
2,27E-02
2,27E-02
2,27E-02
2,28E-02
2,28E-02
2,28E-02
2,29E-02
2,29E-02
2,29E-02
2,29E-02
2,29E-02
2,29E-02
2,29E-02
2,29E-02
2,29E-02
2,29E-02
2,30E-02
2,30E-02
2,30E-02
2,30E-02
2,30E-02
2,30E-02
2,30E-02
2,30E-02
2,30E-02
2,30E-02
2,30E-02
2,30E-02
2,30E-02
2,30E-02
2,30E-02
2,31E-02
2,31E-02
2,31E-02
2,31E-02
2,31E-02

2,31E-02
2,31E-02
2,32E-02
2,32E-02
2,32E-02
2,32E-02
2,33E-02
2,33E-02
2,33E-02
2,34E-02
2,34E-02
2,34E-02
2,35E-02
2,35E-02
2,35E-02
2,35E-02
2,35E-02
2,35E-02
2,35E-02
2,36E-02
2,36E-02
2,36E-02
2,36E-02
2,36E-02
2,36E-02
2,36E-02
2,36E-02
2,36E-02
2,36E-02
2,36E-02
2,36E-02
2,36E-02
2,37E-02
2,37E-02
2,37E-02
2,37E-02
2,37E-02
2,37E-02
2,37E-02
2,37E-02
2,37E-02
2,38E-02
2,38E-02
2,38E-02
2,38E-02
2,38E-02

2,39E-02
2,39E-02
2,39E-02
2,39E-02
2,39E-02
2,39E-02
2,39E-02
2,39E-02
2,39E-02
2,40E-02
2,40E-02
2,40E-02
2,40E-02
2,40E-02
2,41E-02
2,41E-02
2,41E-02
2,41E-02
2,41E-02
2,41E-02
2,41E-02
2,41E-02
2,41E-02
2,41E-02
2,42E-02
2,42E-02
2,42E-02
2,43E-02
2,43E-02
2,43E-02
2,43E-02
2,43E-02
2,43E-02
2,43E-02
2,43E-02
2,43E-02
2,43E-02
2,43E-02
2,43E-02
2,43E-02
2,43E-02
2,43E-02
2,43E-02
2,43E-02
2,44E-02
2,44E-02

2,44E-02
2,44E-02
2,46E-02
2,46E-02
2,46E-02
2,46E-02
2,46E-02
2,46E-02
2,46E-02
2,46E-02
2,46E-02
2,46E-02
2,46E-02
2,46E-02
2,46E-02
2,46E-02
2,46E-02
2,47E-02
2,47E-02
2,47E-02
2,47E-02
2,48E-02
2,48E-02
2,49E-02
2,50E-02
2,50E-02
2,50E-02
2,50E-02
2,50E-02
2,50E-02
2,50E-02
2,51E-02
2,51E-02
2,51E-02
2,52E-02
2,52E-02
2,52E-02
2,52E-02
2,52E-02
2,52E-02
2,52E-02
2,53E-02
2,53E-02
2,53E-02
2,53E-02
2,54E-02

2,55E-02
2,55E-02
2,55E-02
2,55E-02
2,55E-02
2,55E-02
2,55E-02
2,56E-02
2,56E-02
2,56E-02
2,56E-02
2,56E-02
2,56E-02
2,56E-02
2,56E-02
2,57E-02
2,57E-02
2,57E-02
2,57E-02
2,57E-02
2,57E-02
2,57E-02
2,57E-02
2,57E-02
2,57E-02
2,57E-02
2,57E-02
2,57E-02
2,57E-02
2,57E-02
2,57E-02
2,57E-02
2,57E-02
2,57E-02
2,58E-02
2,58E-02
2,58E-02
2,58E-02
2,58E-02
2,58E-02
2,58E-02
2,58E-02
2,59E-02
2,59E-02
2,60E-02
2,60E-02

2,60E-02
2,60E-02
2,61E-02
2,61E-02
2,61E-02
2,62E-02
2,62E-02
2,62E-02
2,62E-02
2,62E-02
2,62E-02
2,62E-02
2,62E-02
2,63E-02
2,63E-02
2,63E-02
2,63E-02
2,64E-02
2,64E-02
2,64E-02
2,64E-02
2,64E-02
2,64E-02
2,64E-02
2,64E-02
2,65E-02
2,65E-02
2,65E-02
2,65E-02
2,65E-02
2,65E-02
2,65E-02
2,65E-02
2,65E-02
2,66E-02
2,66E-02
2,66E-02
2,66E-02
2,66E-02
2,66E-02
2,67E-02
2,67E-02
2,67E-02
2,67E-02
2,67E-02
2,67E-02

2,67E-02
2,68E-02
2,68E-02
2,68E-02
2,68E-02
2,68E-02
2,69E-02
2,69E-02
2,69E-02
2,69E-02
2,70E-02
2,70E-02
2,70E-02
2,70E-02
2,71E-02
2,71E-02
2,71E-02
2,71E-02
2,71E-02
2,71E-02
2,71E-02
2,71E-02
2,72E-02
2,72E-02
2,72E-02
2,72E-02
2,72E-02
2,72E-02
2,73E-02
2,73E-02
2,73E-02
2,73E-02
2,73E-02
2,73E-02
2,73E-02
2,73E-02
2,73E-02
2,73E-02
2,73E-02
2,73E-02
2,73E-02
2,73E-02
2,73E-02
2,73E-02
2,73E-02
2,73E-02

2,73E-02
2,73E-02
2,73E-02
2,74E-02
2,74E-02
2,75E-02
2,75E-02
2,75E-02
2,75E-02
2,75E-02
2,75E-02
2,76E-02
2,76E-02
2,76E-02
2,76E-02
2,76E-02
2,77E-02
2,77E-02
2,77E-02
2,77E-02
2,77E-02
2,77E-02
2,77E-02
2,77E-02
2,78E-02
2,78E-02
2,78E-02
2,79E-02
2,79E-02
2,79E-02
2,79E-02
2,79E-02
2,79E-02
2,79E-02
2,79E-02
2,79E-02
2,80E-02
2,80E-02
2,80E-02
2,80E-02
2,80E-02
2,81E-02
2,81E-02
2,81E-02
2,81E-02
2,82E-02

2,82E-02
2,82E-02
2,83E-02
2,84E-02
2,84E-02
2,84E-02
2,84E-02
2,84E-02
2,85E-02
2,85E-02
2,85E-02
2,85E-02
2,85E-02
2,85E-02
2,85E-02
2,85E-02
2,85E-02
2,85E-02
2,86E-02
2,86E-02
2,86E-02
2,86E-02
2,86E-02
2,86E-02
2,86E-02
2,86E-02
2,87E-02
2,87E-02
2,87E-02
2,87E-02
2,87E-02
2,87E-02
2,87E-02
2,87E-02
2,87E-02
2,87E-02
2,88E-02
2,88E-02
2,89E-02
2,89E-02
2,89E-02
2,89E-02
2,89E-02
2,89E-02
2,90E-02
2,90E-02

2,90E-02
2,90E-02
2,90E-02
2,91E-02
2,91E-02
2,91E-02
2,91E-02
2,91E-02
2,91E-02
2,91E-02
2,91E-02
2,91E-02
2,92E-02
2,92E-02
2,92E-02
2,92E-02
2,92E-02
2,93E-02
2,93E-02
2,93E-02
2,93E-02
2,94E-02
2,94E-02
2,94E-02
2,95E-02
2,96E-02
2,96E-02
2,97E-02
2,97E-02
2,97E-02
2,97E-02
2,97E-02
2,97E-02
2,97E-02
2,97E-02
2,97E-02
2,97E-02
2,97E-02
2,97E-02
2,97E-02
2,98E-02
2,98E-02
2,98E-02
2,98E-02
2,98E-02
2,99E-02

2,99E-02
2,99E-02
2,99E-02
2,99E-02
2,99E-02
3,00E-02
3,00E-02
3,00E-02
3,00E-02
3,00E-02
3,00E-02
3,00E-02
3,00E-02
3,00E-02
3,01E-02
3,01E-02
3,01E-02
3,01E-02
3,01E-02
3,01E-02
3,01E-02
3,01E-02
3,01E-02
3,01E-02
3,01E-02
3,01E-02
3,01E-02
3,02E-02
3,02E-02
3,02E-02
3,02E-02
3,03E-02
3,03E-02
3,03E-02
3,03E-02
3,03E-02
3,04E-02
3,04E-02
3,04E-02
3,04E-02
3,05E-02
3,05E-02
3,05E-02
3,05E-02
3,05E-02
3,06E-02

3,06E-02
3,06E-02
3,06E-02
3,06E-02
3,06E-02
3,06E-02
3,06E-02
3,06E-02
3,07E-02
3,07E-02
3,07E-02
3,07E-02
3,07E-02
3,07E-02
3,07E-02
3,07E-02
3,07E-02
3,07E-02
3,07E-02
3,07E-02
3,07E-02
3,07E-02
3,07E-02
3,07E-02
3,07E-02
3,08E-02
3,08E-02
3,08E-02
3,08E-02
3,08E-02
3,08E-02
3,08E-02
3,08E-02
3,08E-02
3,09E-02
3,09E-02
3,09E-02
3,09E-02
3,09E-02
3,10E-02
3,10E-02
3,10E-02
3,10E-02
3,10E-02
3,10E-02
3,10E-02

3,10E-02
3,10E-02
3,11E-02
3,11E-02
3,11E-02
3,11E-02
3,11E-02
3,12E-02
3,12E-02
3,12E-02
3,12E-02
3,12E-02
3,12E-02
3,13E-02
3,13E-02
3,13E-02
3,13E-02
3,13E-02
3,14E-02
3,14E-02
3,14E-02
3,14E-02
3,14E-02
3,14E-02
3,14E-02
3,15E-02
3,15E-02
3,15E-02
3,15E-02
3,16E-02
3,16E-02
3,16E-02
3,16E-02
3,16E-02
3,16E-02
3,16E-02
3,17E-02
3,17E-02
3,17E-02
3,17E-02
3,17E-02
3,17E-02
3,17E-02
3,17E-02
3,17E-02
3,19E-02

3,19E-02
3,19E-02
3,19E-02
3,20E-02
3,20E-02
3,21E-02
3,21E-02
3,21E-02
3,21E-02
3,21E-02
3,21E-02
3,21E-02
3,22E-02
3,22E-02
3,22E-02
3,22E-02
3,22E-02
3,22E-02
3,23E-02
3,23E-02
3,23E-02
3,23E-02
3,23E-02
3,23E-02
3,23E-02
3,24E-02
3,24E-02
3,24E-02
3,24E-02
3,24E-02
3,25E-02
3,25E-02
3,25E-02
3,26E-02
3,26E-02
3,26E-02
3,26E-02
3,26E-02
3,26E-02
3,26E-02
3,26E-02
3,27E-02
3,27E-02
3,27E-02
3,27E-02
3,27E-02

3,28E-02
3,28E-02
3,28E-02
3,28E-02
3,28E-02
3,28E-02
3,28E-02
3,28E-02
3,28E-02
3,29E-02
3,29E-02
3,29E-02
3,29E-02
3,29E-02
3,29E-02
3,29E-02
3,29E-02
3,30E-02
3,30E-02
3,31E-02
3,31E-02
3,31E-02
3,31E-02
3,31E-02
3,31E-02
3,31E-02
3,31E-02
3,31E-02
3,31E-02
3,31E-02
3,31E-02
3,31E-02
3,31E-02
3,32E-02
3,32E-02
3,32E-02
3,32E-02
3,32E-02
3,32E-02
3,32E-02
3,32E-02
3,32E-02
3,33E-02
3,33E-02
3,33E-02

3,33E-02
3,33E-02
3,33E-02
3,33E-02
3,33E-02
3,34E-02
3,34E-02
3,34E-02
3,35E-02
3,35E-02
3,35E-02
3,35E-02
3,36E-02
3,36E-02
3,36E-02
3,36E-02
3,36E-02
3,36E-02
3,36E-02
3,36E-02
3,37E-02
3,37E-02
3,38E-02
3,38E-02
3,38E-02
3,38E-02
3,38E-02
3,38E-02
3,39E-02
3,39E-02
3,39E-02
3,39E-02
3,39E-02
3,39E-02
3,40E-02
3,40E-02
3,40E-02
3,40E-02
3,40E-02
3,40E-02
3,40E-02
3,40E-02
3,40E-02
3,40E-02
3,41E-02
3,41E-02

3,41E-02
3,42E-02
3,42E-02
3,42E-02
3,42E-02
3,43E-02
3,44E-02
3,44E-02
3,45E-02
3,45E-02
3,45E-02
3,45E-02
3,46E-02
3,46E-02
3,46E-02
3,46E-02
3,46E-02
3,47E-02
3,47E-02
3,47E-02
3,48E-02
3,48E-02
3,48E-02
3,48E-02
3,48E-02
3,48E-02
3,48E-02
3,48E-02
3,48E-02
3,49E-02
3,49E-02
3,49E-02
3,50E-02
3,50E-02
3,51E-02
3,51E-02
3,51E-02
3,51E-02
3,51E-02
3,51E-02
3,51E-02
3,51E-02
3,52E-02
3,52E-02
3,52E-02
3,52E-02

3,53E-02
3,53E-02
3,53E-02
3,53E-02
3,53E-02
3,53E-02
3,53E-02
3,53E-02
3,54E-02
3,54E-02
3,54E-02
3,55E-02
3,55E-02
3,56E-02
3,56E-02
3,56E-02
3,56E-02
3,57E-02
3,57E-02
3,57E-02
3,57E-02
3,57E-02
3,57E-02
3,57E-02
3,57E-02
3,57E-02
3,58E-02
3,58E-02
3,58E-02
3,58E-02
3,59E-02
3,59E-02
3,59E-02
3,59E-02
3,59E-02
3,60E-02
3,60E-02
3,60E-02
3,60E-02
3,60E-02
3,60E-02
3,60E-02
3,60E-02
3,60E-02
3,60E-02
3,61E-02

3,61E-02
3,61E-02
3,61E-02
3,62E-02
3,62E-02
3,62E-02
3,62E-02
3,62E-02
3,62E-02
3,63E-02
3,63E-02
3,63E-02
3,63E-02
3,63E-02
3,63E-02
3,64E-02
3,64E-02
3,65E-02
3,65E-02
3,66E-02
3,66E-02
3,66E-02
3,66E-02
3,66E-02
3,66E-02
3,67E-02
3,67E-02
3,67E-02
3,67E-02
3,67E-02
3,68E-02
3,68E-02
3,68E-02
3,68E-02
3,68E-02
3,68E-02
3,69E-02
3,69E-02
3,69E-02
3,69E-02
3,69E-02
3,69E-02
3,69E-02
3,69E-02
3,69E-02
3,69E-02

3,69E-02
3,70E-02
3,70E-02
3,70E-02
3,70E-02
3,71E-02
3,71E-02
3,71E-02
3,71E-02
3,71E-02
3,71E-02
3,72E-02
3,72E-02
3,72E-02
3,72E-02
3,72E-02
3,72E-02
3,73E-02
3,73E-02
3,73E-02
3,73E-02
3,73E-02
3,73E-02
3,73E-02
3,73E-02
3,73E-02
3,73E-02
3,74E-02
3,75E-02
3,75E-02
3,75E-02
3,75E-02
3,75E-02
3,75E-02
3,75E-02
3,75E-02
3,76E-02
3,76E-02
3,76E-02
3,76E-02
3,76E-02
3,76E-02
3,76E-02
3,76E-02
3,76E-02
3,77E-02

3,77E-02
3,77E-02
3,77E-02
3,77E-02
3,77E-02
3,78E-02
3,78E-02
3,78E-02
3,79E-02
3,79E-02
3,79E-02
3,79E-02
3,80E-02
3,81E-02
3,81E-02
3,81E-02
3,81E-02
3,81E-02
3,82E-02
3,82E-02
3,82E-02
3,82E-02
3,83E-02
3,83E-02
3,83E-02
3,83E-02
3,84E-02
3,84E-02
3,84E-02
3,84E-02
3,84E-02
3,84E-02
3,85E-02
3,85E-02
3,85E-02
3,85E-02
3,86E-02
3,86E-02
3,86E-02
3,87E-02
3,87E-02
3,87E-02
3,88E-02
3,88E-02
3,88E-02
3,88E-02

3,88E-02
3,89E-02
3,89E-02
3,89E-02
3,89E-02
3,90E-02
3,90E-02
3,90E-02
3,90E-02
3,90E-02
3,90E-02
3,91E-02
3,92E-02
3,92E-02
3,92E-02
3,92E-02
3,92E-02
3,92E-02
3,92E-02
3,92E-02
3,92E-02
3,93E-02
3,93E-02
3,93E-02
3,93E-02
3,93E-02
3,93E-02
3,93E-02
3,93E-02
3,93E-02
3,93E-02
3,94E-02
3,94E-02
3,94E-02
3,94E-02
3,94E-02
3,94E-02
3,95E-02
3,95E-02
3,95E-02
3,96E-02
3,96E-02
3,96E-02
3,96E-02
3,96E-02
3,97E-02

3,98E-02
3,98E-02
3,99E-02
3,99E-02
3,99E-02
3,99E-02
4,01E-02
4,01E-02
4,01E-02
4,02E-02
4,02E-02
4,02E-02
4,02E-02
4,02E-02
4,02E-02
4,03E-02
4,03E-02
4,03E-02
4,03E-02
4,03E-02
4,03E-02
4,04E-02
4,04E-02
4,04E-02
4,04E-02
4,05E-02
4,05E-02
4,05E-02
4,06E-02
4,06E-02
4,06E-02
4,06E-02
4,06E-02
4,06E-02
4,06E-02
4,06E-02
4,07E-02
4,07E-02
4,08E-02
4,09E-02
4,09E-02
4,09E-02
4,10E-02
4,10E-02
4,10E-02
4,11E-02

4,11E-02
4,11E-02
4,12E-02
4,12E-02
4,12E-02
4,12E-02
4,12E-02
4,13E-02
4,13E-02
4,13E-02
4,14E-02
4,14E-02
4,15E-02
4,15E-02
4,16E-02
4,16E-02
4,16E-02
4,16E-02
4,17E-02
4,17E-02
4,17E-02
4,17E-02
4,17E-02
4,17E-02
4,18E-02
4,18E-02
4,18E-02
4,18E-02
4,18E-02
4,18E-02
4,18E-02
4,19E-02
4,19E-02
4,19E-02
4,20E-02
4,20E-02
4,20E-02
4,20E-02
4,20E-02
4,20E-02
4,20E-02
4,20E-02
4,20E-02
4,21E-02
4,21E-02
4,21E-02

4,22E-02
4,22E-02
4,22E-02
4,22E-02
4,22E-02
4,23E-02
4,23E-02
4,23E-02
4,23E-02
4,23E-02
4,23E-02
4,24E-02
4,24E-02
4,25E-02
4,25E-02
4,25E-02
4,25E-02
4,25E-02
4,26E-02
4,26E-02
4,26E-02
4,26E-02
4,27E-02
4,27E-02
4,27E-02
4,27E-02
4,28E-02
4,28E-02
4,28E-02
4,28E-02
4,29E-02
4,30E-02
4,30E-02
4,30E-02
4,30E-02
4,31E-02
4,31E-02
4,32E-02
4,32E-02
4,32E-02
4,32E-02
4,32E-02
4,32E-02
4,33E-02
4,34E-02
4,34E-02

4,35E-02
4,35E-02
4,35E-02
4,36E-02
4,36E-02
4,36E-02
4,36E-02
4,37E-02
4,37E-02
4,37E-02
4,37E-02
4,38E-02
4,39E-02
4,39E-02
4,39E-02
4,40E-02
4,40E-02
4,40E-02
4,41E-02
4,41E-02
4,41E-02
4,41E-02
4,41E-02
4,42E-02
4,42E-02
4,42E-02
4,42E-02
4,42E-02
4,42E-02
4,42E-02
4,42E-02
4,42E-02
4,42E-02
4,42E-02
4,42E-02
4,42E-02
4,42E-02
4,43E-02
4,43E-02
4,43E-02
4,44E-02
4,44E-02
4,44E-02
4,44E-02
4,45E-02
4,45E-02

4,45E-02
4,45E-02
4,45E-02
4,45E-02
4,46E-02
4,46E-02
4,48E-02
4,48E-02
4,48E-02
4,48E-02
4,48E-02
4,48E-02
4,49E-02
4,49E-02
4,49E-02
4,50E-02
4,50E-02
4,51E-02
4,51E-02
4,52E-02
4,52E-02
4,52E-02
4,52E-02
4,53E-02
4,53E-02
4,54E-02
4,54E-02
4,55E-02
4,55E-02
4,55E-02
4,55E-02
4,56E-02
4,56E-02
4,57E-02
4,57E-02
4,57E-02
4,57E-02
4,58E-02
4,58E-02
4,59E-02
4,59E-02
4,59E-02
4,59E-02
4,59E-02
4,59E-02
4,60E-02

4,60E-02
4,60E-02
4,60E-02
4,61E-02
4,61E-02
4,61E-02
4,61E-02
4,62E-02
4,62E-02
4,63E-02
4,64E-02
4,64E-02
4,64E-02
4,65E-02
4,65E-02
4,65E-02
4,66E-02
4,66E-02
4,66E-02
4,66E-02
4,66E-02
4,66E-02
4,67E-02
4,67E-02
4,67E-02
4,67E-02
4,67E-02
4,67E-02
4,67E-02
4,67E-02
4,67E-02
4,67E-02
4,67E-02
4,67E-02
4,68E-02
4,68E-02
4,69E-02
4,69E-02
4,69E-02
4,70E-02
4,70E-02
4,70E-02
4,70E-02
4,70E-02
4,71E-02
4,71E-02

4,71E-02
4,71E-02
4,71E-02
4,71E-02
4,71E-02
4,71E-02
4,71E-02
4,71E-02
4,71E-02
4,71E-02
4,71E-02
4,72E-02
4,73E-02
4,74E-02
4,74E-02
4,74E-02
4,74E-02
4,74E-02
4,74E-02
4,74E-02
4,74E-02
4,74E-02
4,75E-02
4,76E-02
4,76E-02
4,76E-02
4,76E-02
4,76E-02
4,77E-02
4,77E-02
4,77E-02
4,78E-02
4,78E-02
4,78E-02
4,78E-02
4,78E-02
4,78E-02
4,78E-02
4,78E-02
4,78E-02
4,78E-02
4,78E-02
4,79E-02
4,79E-02
4,79E-02
4,79E-02

4,80E-02
4,80E-02
4,80E-02
4,80E-02
4,80E-02
4,80E-02
4,81E-02
4,81E-02
4,82E-02
4,82E-02
4,82E-02
4,83E-02
4,83E-02
4,84E-02
4,84E-02
4,84E-02
4,84E-02
4,85E-02
4,85E-02
4,87E-02
4,88E-02
4,88E-02
4,88E-02
4,88E-02
4,88E-02
4,88E-02
4,88E-02
4,89E-02
4,90E-02
4,90E-02
4,90E-02
4,90E-02
4,90E-02
4,90E-02
4,90E-02
4,92E-02
4,92E-02
4,92E-02
4,92E-02
4,92E-02
4,92E-02
4,92E-02
4,93E-02
4,94E-02
4,94E-02
4,94E-02

4,94E-02
4,95E-02
4,95E-02
4,96E-02
4,96E-02
4,97E-02
4,98E-02
4,98E-02
5,00E-02
5,00E-02
5,00E-02
5,01E-02
5,01E-02
5,01E-02
5,01E-02
5,02E-02
5,02E-02
5,02E-02
5,02E-02
5,02E-02
5,03E-02
5,03E-02
5,03E-02
5,03E-02
5,04E-02
5,04E-02
5,05E-02
5,05E-02
5,06E-02
5,06E-02
5,06E-02
5,07E-02
5,07E-02
5,07E-02
5,07E-02
5,08E-02
5,08E-02
5,08E-02
5,08E-02
5,08E-02
5,09E-02
5,09E-02
5,09E-02
5,10E-02
5,11E-02
5,11E-02

5,11E-02
5,11E-02
5,11E-02
5,11E-02
5,11E-02
5,12E-02
5,12E-02
5,12E-02
5,12E-02
5,13E-02
5,13E-02
5,13E-02
5,15E-02
5,15E-02
5,15E-02
5,15E-02
5,16E-02
5,17E-02
5,17E-02
5,17E-02
5,18E-02
5,18E-02
5,19E-02
5,19E-02
5,20E-02
5,21E-02
5,21E-02
5,21E-02
5,23E-02
5,24E-02
5,24E-02
5,24E-02
5,24E-02
5,24E-02
5,25E-02
5,25E-02
5,25E-02
5,25E-02
5,25E-02
5,26E-02
5,28E-02
5,28E-02
5,28E-02
5,29E-02
5,29E-02
5,29E-02

5,29E-02
5,29E-02
5,29E-02
5,30E-02
5,30E-02
5,31E-02
5,31E-02
5,31E-02
5,31E-02
5,31E-02
5,33E-02
5,33E-02
5,33E-02
5,33E-02
5,33E-02
5,34E-02
5,34E-02
5,34E-02
5,34E-02
5,34E-02
5,34E-02
5,34E-02
5,35E-02
5,35E-02
5,36E-02
5,37E-02
5,37E-02
5,37E-02
5,37E-02
5,37E-02
5,37E-02
5,37E-02
5,37E-02
5,38E-02
5,39E-02
5,39E-02
5,39E-02
5,39E-02
5,39E-02
5,40E-02
5,40E-02
5,40E-02
5,40E-02
5,40E-02
5,40E-02
5,40E-02

5,40E-02
5,40E-02
5,40E-02
5,40E-02
5,40E-02
5,40E-02
5,41E-02
5,41E-02
5,41E-02
5,42E-02
5,43E-02
5,43E-02
5,44E-02
5,44E-02
5,45E-02
5,45E-02
5,45E-02
5,45E-02
5,45E-02
5,45E-02
5,45E-02
5,45E-02
5,45E-02
5,45E-02
5,45E-02
5,45E-02
5,46E-02
5,46E-02
5,46E-02
5,46E-02
5,46E-02
5,46E-02
5,46E-02
5,47E-02
5,47E-02
5,47E-02
5,47E-02
5,47E-02
5,47E-02
5,47E-02
5,47E-02
5,48E-02
5,48E-02
5,48E-02
5,48E-02
5,48E-02

5,49E-02
5,50E-02
5,50E-02
5,50E-02
5,51E-02
5,51E-02
5,51E-02
5,51E-02
5,52E-02
5,52E-02
5,52E-02
5,52E-02
5,53E-02
5,55E-02
5,55E-02
5,55E-02
5,56E-02
5,56E-02
5,56E-02
5,56E-02
5,57E-02
5,57E-02
5,57E-02
5,57E-02
5,57E-02
5,58E-02
5,58E-02
5,58E-02
5,58E-02
5,59E-02
5,59E-02
5,59E-02
5,60E-02
5,61E-02
5,62E-02
5,62E-02
5,62E-02
5,62E-02
5,62E-02
5,62E-02
5,62E-02
5,62E-02
5,63E-02
5,63E-02
5,63E-02
5,64E-02

5,64E-02
5,64E-02
5,65E-02
5,65E-02
5,65E-02
5,66E-02
5,66E-02
5,66E-02
5,66E-02
5,67E-02
5,67E-02
5,67E-02
5,68E-02
5,68E-02
5,68E-02
5,68E-02
5,68E-02
5,68E-02
5,68E-02
5,69E-02
5,70E-02
5,70E-02
5,70E-02
5,70E-02
5,71E-02
5,71E-02
5,71E-02
5,71E-02
5,71E-02
5,71E-02
5,71E-02
5,71E-02
5,71E-02
5,72E-02
5,72E-02
5,72E-02
5,74E-02
5,75E-02
5,75E-02
5,75E-02
5,75E-02
5,76E-02
5,76E-02
5,77E-02
5,77E-02
5,77E-02

5,77E-02
5,77E-02
5,77E-02
5,77E-02
5,78E-02
5,78E-02
5,79E-02
5,79E-02
5,80E-02
5,80E-02
5,81E-02
5,81E-02
5,81E-02
5,81E-02
5,81E-02
5,81E-02
5,82E-02
5,82E-02
5,82E-02
5,82E-02
5,82E-02
5,82E-02
5,83E-02
5,83E-02
5,83E-02
5,84E-02
5,84E-02
5,84E-02
5,85E-02
5,85E-02
5,85E-02
5,87E-02
5,87E-02
5,87E-02
5,88E-02
5,88E-02
5,88E-02
5,88E-02
5,88E-02
5,89E-02
5,89E-02
5,90E-02
5,90E-02
5,90E-02
5,90E-02
5,91E-02

5,91E-02
5,92E-02
5,92E-02
5,93E-02
5,93E-02
5,93E-02
5,93E-02
5,93E-02
5,93E-02
5,93E-02
5,93E-02
5,93E-02
5,94E-02
5,95E-02
5,95E-02
5,95E-02
5,95E-02
5,95E-02
5,95E-02
5,96E-02
5,96E-02
5,97E-02
5,98E-02
5,98E-02
5,98E-02
5,99E-02
6,01E-02
6,01E-02
6,01E-02
6,01E-02
6,01E-02
6,02E-02
6,03E-02
6,03E-02
6,03E-02
6,03E-02
6,03E-02
6,03E-02
6,04E-02
6,04E-02
6,05E-02
6,06E-02
6,06E-02
6,06E-02
6,06E-02
6,07E-02

6,07E-02
6,07E-02
6,07E-02
6,08E-02
6,08E-02
6,08E-02
6,08E-02
6,08E-02
6,09E-02
6,09E-02
6,09E-02
6,10E-02
6,10E-02
6,10E-02
6,10E-02
6,10E-02
6,10E-02
6,10E-02
6,10E-02
6,10E-02
6,10E-02
6,10E-02
6,10E-02
6,11E-02
6,11E-02
6,11E-02
6,11E-02
6,11E-02
6,12E-02
6,12E-02
6,15E-02
6,15E-02
6,15E-02
6,15E-02
6,15E-02
6,16E-02
6,16E-02
6,18E-02
6,18E-02
6,18E-02
6,18E-02
6,19E-02
6,19E-02
6,19E-02
6,19E-02
6,19E-02

6,21E-02
6,21E-02
6,22E-02
6,22E-02
6,23E-02
6,24E-02
6,24E-02
6,25E-02
6,25E-02
6,25E-02
6,25E-02
6,25E-02
6,25E-02
6,25E-02
6,25E-02
6,26E-02
6,27E-02
6,27E-02
6,28E-02
6,28E-02
6,30E-02
6,30E-02
6,30E-02
6,31E-02
6,31E-02
6,32E-02
6,32E-02
6,32E-02
6,32E-02
6,33E-02
6,33E-02
6,33E-02
6,33E-02
6,33E-02
6,33E-02
6,35E-02
6,35E-02
6,35E-02
6,35E-02
6,35E-02
6,35E-02
6,35E-02
6,35E-02
6,35E-02
6,36E-02
6,36E-02

6,37E-02
6,37E-02
6,37E-02
6,37E-02
6,38E-02
6,38E-02
6,39E-02
6,39E-02
6,40E-02
6,40E-02
6,40E-02
6,40E-02
6,40E-02
6,40E-02
6,41E-02
6,41E-02
6,41E-02
6,41E-02
6,41E-02
6,41E-02
6,41E-02
6,41E-02
6,42E-02
6,42E-02
6,43E-02
6,44E-02
6,44E-02
6,44E-02
6,44E-02
6,45E-02
6,45E-02
6,46E-02
6,46E-02
6,46E-02
6,46E-02
6,47E-02
6,48E-02
6,48E-02
6,49E-02
6,49E-02
6,49E-02
6,50E-02
6,50E-02
6,51E-02
6,51E-02
6,51E-02

6,51E-02
6,52E-02
6,52E-02
6,53E-02
6,54E-02
6,54E-02
6,55E-02
6,55E-02
6,56E-02
6,56E-02
6,57E-02
6,57E-02
6,57E-02
6,57E-02
6,57E-02
6,58E-02
6,58E-02
6,58E-02
6,58E-02
6,58E-02
6,58E-02
6,59E-02
6,61E-02
6,61E-02
6,61E-02
6,61E-02
6,61E-02
6,62E-02
6,62E-02
6,63E-02
6,64E-02
6,64E-02
6,64E-02
6,64E-02
6,64E-02
6,66E-02
6,66E-02
6,66E-02
6,66E-02
6,67E-02
6,67E-02
6,67E-02
6,68E-02
6,68E-02
6,69E-02
6,69E-02

6,69E-02
6,70E-02
6,70E-02
6,70E-02
6,70E-02
6,71E-02
6,72E-02
6,72E-02
6,72E-02
6,72E-02
6,72E-02
6,73E-02
6,73E-02
6,73E-02
6,73E-02
6,73E-02
6,73E-02
6,73E-02
6,73E-02
6,73E-02
6,73E-02
6,73E-02
6,73E-02
6,74E-02
6,74E-02
6,74E-02
6,75E-02
6,75E-02
6,75E-02
6,75E-02
6,75E-02
6,75E-02
6,76E-02
6,76E-02
6,77E-02
6,77E-02
6,78E-02
6,78E-02
6,79E-02
6,79E-02
6,79E-02
6,79E-02
6,79E-02
6,80E-02
6,80E-02
6,81E-02

6,81E-02
6,82E-02
6,82E-02
6,82E-02
6,82E-02
6,82E-02
6,82E-02
6,82E-02
6,82E-02
6,82E-02
6,83E-02
6,83E-02
6,83E-02
6,84E-02
6,84E-02
6,84E-02
6,84E-02
6,84E-02
6,84E-02
6,84E-02
6,86E-02
6,86E-02
6,86E-02
6,86E-02
6,87E-02
6,88E-02
6,88E-02
6,88E-02
6,88E-02
6,88E-02
6,89E-02
6,89E-02
6,89E-02
6,89E-02
6,90E-02
6,91E-02
6,91E-02
6,91E-02
6,91E-02
6,92E-02
6,92E-02
6,94E-02
6,95E-02
6,95E-02
6,95E-02
6,95E-02

6,95E-02
6,96E-02
6,96E-02
6,97E-02
6,97E-02
6,97E-02
6,97E-02
6,97E-02
6,97E-02
6,98E-02
6,98E-02
7,00E-02
7,01E-02
7,01E-02
7,01E-02
7,01E-02
7,02E-02
7,02E-02
7,02E-02
7,03E-02
7,05E-02
7,06E-02
7,07E-02
7,08E-02
7,09E-02
7,10E-02
7,10E-02
7,10E-02
7,10E-02
7,10E-02
7,10E-02
7,10E-02
7,10E-02
7,10E-02
7,11E-02
7,11E-02
7,11E-02
7,11E-02
7,12E-02
7,12E-02
7,12E-02
7,12E-02
7,12E-02
7,12E-02
7,12E-02
7,12E-02

| |
|---|
| 7,12E-02 |
| 7,12E-02 |
| 7,12E-02 |
| 7,13E-02 |
| 7,15E-02 |
| 7,17E-02 |
| 7,17E-02 |
| 7,18E-02 |
| 7,19E-02 |
| 7,20E-02 |
| 7,21E-02 |
| 7,21E-02 |
| 7,21E-02 |
| 7,22E-02 |
| 7,22E-02 |
| 7,22E-02 |
| 7,22E-02 |
| 7,22E-02 |
| 7,22E-02 |
| 7,22E-02 |
| 7,22E-02 |
| 7,22E-02 |
| 7,23E-02 |
| 7,23E-02 |
| 7,24E-02 |
| 7,24E-02 |
| 7,24E-02 |
| 7,26E-02 |
| 7,26E-02 |
| 7,26E-02 |
| 7,27E-02 |
| 7,27E-02 |
| 7,27E-02 |
| 7,27E-02 |
| 7,28E-02 |
| 7,29E-02 |
| 7,29E-02 |
| 7,30E-02 |
| 7,30E-02 |
| 7,32E-02 |
| 7,33E-02 |
| 7,33E-02 |
| 7,34E-02 |
| 7,34E-02 |
| 7,35E-02 |
| 7,35E-02 |

7,36E-02
7,36E-02
7,36E-02
7,37E-02
7,38E-02
7,38E-02
7,38E-02
7,38E-02
7,38E-02
7,38E-02
7,39E-02
7,39E-02
7,39E-02
7,40E-02
7,40E-02
7,40E-02
7,41E-02
7,41E-02
7,42E-02
7,42E-02
7,43E-02
7,43E-02
7,45E-02
7,46E-02
7,47E-02
7,47E-02
7,47E-02
7,47E-02
7,48E-02
7,50E-02
7,50E-02
7,50E-02
7,50E-02
7,51E-02
7,51E-02
7,51E-02
7,51E-02
7,51E-02
7,53E-02
7,53E-02
7,54E-02
7,54E-02
7,54E-02
7,54E-02
7,55E-02
7,55E-02

7,55E-02
7,55E-02
7,55E-02
7,55E-02
7,55E-02
7,55E-02
7,55E-02
7,55E-02
7,56E-02
7,56E-02
7,56E-02
7,56E-02
7,57E-02
7,57E-02
7,57E-02
7,58E-02
7,58E-02
7,58E-02
7,58E-02
7,59E-02
7,59E-02
7,59E-02
7,60E-02
7,60E-02
7,60E-02
7,61E-02
7,62E-02
7,62E-02
7,62E-02
7,62E-02
7,63E-02
7,63E-02
7,63E-02
7,63E-02
7,63E-02
7,63E-02
7,63E-02
7,63E-02
7,64E-02
7,64E-02
7,64E-02
7,64E-02
7,64E-02
7,64E-02
7,64E-02
7,65E-02

7,65E-02
7,65E-02
7,65E-02
7,65E-02
7,65E-02
7,66E-02
7,66E-02
7,66E-02
7,67E-02
7,68E-02
7,68E-02
7,68E-02
7,68E-02
7,69E-02
7,70E-02
7,70E-02
7,70E-02
7,71E-02
7,71E-02
7,73E-02
7,73E-02
7,73E-02
7,74E-02
7,74E-02
7,75E-02
7,75E-02
7,75E-02
7,75E-02
7,75E-02
7,75E-02
7,75E-02
7,76E-02
7,76E-02
7,76E-02
7,77E-02
7,78E-02
7,78E-02
7,78E-02
7,79E-02
7,79E-02
7,80E-02
7,80E-02
7,80E-02
7,80E-02
7,81E-02
7,81E-02

| |
|---|
| 7,83E-02 |
| 7,85E-02 |
| 7,85E-02 |
| 7,85E-02 |
| 7,86E-02 |
| 7,86E-02 |
| 7,87E-02 |
| 7,88E-02 |
| 7,88E-02 |
| 7,88E-02 |
| 7,88E-02 |
| 7,88E-02 |
| 7,88E-02 |
| 7,89E-02 |
| 7,89E-02 |
| 7,89E-02 |
| 7,90E-02 |
| 7,92E-02 |
| 7,92E-02 |
| 7,92E-02 |
| 7,93E-02 |
| 7,93E-02 |
| 7,94E-02 |
| 7,94E-02 |
| 7,96E-02 |
| 7,96E-02 |
| 7,96E-02 |
| 7,96E-02 |
| 7,97E-02 |
| 7,97E-02 |
| 7,98E-02 |
| 7,99E-02 |
| 8,00E-02 |
| 8,00E-02 |
| 8,00E-02 |
| 8,01E-02 |
| 8,01E-02 |
| 8,01E-02 |
| 8,01E-02 |
| 8,01E-02 |
| 8,01E-02 |
| 8,01E-02 |
| 8,01E-02 |
| 8,01E-02 |
| 8,02E-02 |
| 8,02E-02 |

| |
|---|
| 8,03E-02 |
| 8,05E-02 |
| 8,05E-02 |
| 8,05E-02 |
| 8,05E-02 |
| 8,05E-02 |
| 8,05E-02 |
| 8,05E-02 |
| 8,05E-02 |
| 8,05E-02 |
| 8,06E-02 |
| 8,07E-02 |
| 8,07E-02 |
| 8,08E-02 |
| 8,08E-02 |
| 8,09E-02 |
| 8,09E-02 |
| 8,09E-02 |
| 8,09E-02 |
| 8,09E-02 |
| 8,09E-02 |
| 8,10E-02 |
| 8,10E-02 |
| 8,10E-02 |
| 8,11E-02 |
| 8,11E-02 |
| 8,11E-02 |
| 8,11E-02 |
| 8,11E-02 |
| 8,11E-02 |
| 8,11E-02 |
| 8,11E-02 |
| 8,11E-02 |
| 8,11E-02 |
| 8,11E-02 |
| 8,11E-02 |
| 8,12E-02 |
| 8,12E-02 |
| 8,12E-02 |
| 8,13E-02 |
| 8,15E-02 |
| 8,16E-02 |
| 8,16E-02 |
| 8,16E-02 |
| 8,16E-02 |
| 8,16E-02 |

8,16E-02
8,17E-02
8,17E-02
8,18E-02
8,18E-02
8,19E-02
8,21E-02
8,22E-02
8,22E-02
8,22E-02
8,22E-02
8,23E-02
8,24E-02
8,24E-02
8,25E-02
8,26E-02
8,26E-02
8,26E-02
8,26E-02
8,27E-02
8,27E-02
8,27E-02
8,27E-02
8,27E-02
8,29E-02
8,29E-02
8,31E-02
8,31E-02
8,32E-02
8,33E-02
8,33E-02
8,35E-02
8,35E-02
8,38E-02
8,39E-02
8,39E-02
8,41E-02
8,41E-02
8,41E-02
8,41E-02
8,41E-02
8,41E-02
8,42E-02
8,42E-02
8,42E-02
8,44E-02

8,44E-02
8,44E-02
8,46E-02
8,47E-02
8,47E-02
8,47E-02
8,47E-02
8,47E-02
8,47E-02
8,47E-02
8,47E-02
8,47E-02
8,48E-02
8,48E-02
8,48E-02
8,49E-02
8,49E-02
8,50E-02
8,52E-02
8,52E-02
8,53E-02
8,53E-02
8,53E-02
8,53E-02
8,54E-02
8,54E-02
8,54E-02
8,54E-02
8,54E-02
8,55E-02
8,56E-02
8,56E-02
8,57E-02
8,59E-02
8,59E-02
8,59E-02
8,59E-02
8,60E-02
8,60E-02
8,60E-02
8,60E-02
8,61E-02
8,63E-02
8,64E-02
8,64E-02
8,64E-02

8,65E-02
8,65E-02
8,65E-02
8,65E-02
8,66E-02
8,66E-02
8,66E-02
8,66E-02
8,66E-02
8,66E-02
8,66E-02
8,67E-02
8,67E-02
8,67E-02
8,68E-02
8,70E-02
8,70E-02
8,70E-02
8,70E-02
8,70E-02
8,70E-02
8,70E-02
8,71E-02
8,71E-02
8,71E-02
8,71E-02
8,72E-02
8,74E-02
8,74E-02
8,74E-02
8,74E-02
8,74E-02
8,74E-02
8,74E-02
8,75E-02
8,75E-02
8,75E-02
8,75E-02
8,75E-02
8,78E-02
8,78E-02
8,79E-02
8,79E-02
8,79E-02
8,79E-02
8,79E-02

8,79E-02
8,80E-02
8,81E-02
8,81E-02
8,82E-02
8,82E-02
8,82E-02
8,84E-02
8,84E-02
8,84E-02
8,84E-02
8,84E-02
8,84E-02
8,84E-02
8,85E-02
8,86E-02
8,86E-02
8,86E-02
8,86E-02
8,87E-02
8,90E-02
8,90E-02
8,91E-02
8,91E-02
8,92E-02
8,95E-02
8,95E-02
8,95E-02
8,95E-02
8,97E-02
8,97E-02
8,97E-02
8,98E-02
8,98E-02
8,98E-02
8,99E-02
8,99E-02
8,99E-02
9,00E-02
9,01E-02
9,01E-02
9,02E-02
9,02E-02
9,02E-02
9,02E-02
9,02E-02

9,03E-02
9,03E-02
9,03E-02
9,04E-02
9,05E-02
9,06E-02
9,06E-02
9,06E-02
9,06E-02
9,06E-02
9,06E-02
9,07E-02
9,07E-02
9,07E-02
9,07E-02
9,07E-02
9,08E-02
9,08E-02
9,09E-02
9,09E-02
9,10E-02
9,10E-02
9,11E-02
9,13E-02
9,13E-02
9,13E-02
9,14E-02
9,14E-02
9,16E-02
9,16E-02
9,17E-02
9,18E-02
9,18E-02
9,18E-02
9,18E-02
9,18E-02
9,18E-02
9,18E-02
9,19E-02
9,20E-02
9,21E-02
9,22E-02
9,22E-02
9,22E-02
9,22E-02
9,23E-02

9,23E-02
9,24E-02
9,24E-02
9,25E-02
9,26E-02
9,26E-02
9,26E-02
9,26E-02
9,26E-02
9,26E-02
9,28E-02
9,29E-02
9,30E-02
9,30E-02
9,32E-02
9,34E-02
9,34E-02
9,35E-02
9,35E-02
9,35E-02
9,36E-02
9,36E-02
9,38E-02
9,38E-02
9,38E-02
9,38E-02
9,39E-02
9,39E-02
9,40E-02
9,41E-02
9,42E-02
9,45E-02
9,45E-02
9,45E-02
9,46E-02
9,48E-02
9,48E-02
9,48E-02
9,49E-02
9,49E-02
9,49E-02
9,49E-02
9,50E-02
9,50E-02
9,50E-02
9,50E-02

9,50E-02
9,50E-02
9,50E-02
9,51E-02
9,51E-02
9,51E-02
9,52E-02
9,52E-02
9,52E-02
9,52E-02
9,53E-02
9,53E-02
9,54E-02
9,54E-02
9,54E-02
9,56E-02
9,56E-02
9,57E-02
9,57E-02
9,57E-02
9,58E-02
9,58E-02
9,58E-02
9,58E-02
9,59E-02
9,59E-02
9,59E-02
9,60E-02
9,60E-02
9,61E-02
9,61E-02
9,61E-02
9,62E-02
9,62E-02
9,66E-02
9,67E-02
9,67E-02
9,67E-02
9,67E-02
9,69E-02
9,69E-02
9,69E-02
9,69E-02
9,69E-02
9,69E-02
9,69E-02

9,69E-02
9,71E-02
9,72E-02
9,72E-02
9,72E-02
9,73E-02
9,75E-02
9,75E-02
9,75E-02
9,76E-02
9,76E-02
9,76E-02
9,76E-02
9,76E-02
9,76E-02
9,76E-02
9,77E-02
9,78E-02
9,78E-02
9,78E-02
9,78E-02
9,79E-02
9,79E-02
9,80E-02
9,80E-02
9,81E-02
9,81E-02
9,81E-02
9,81E-02
9,81E-02
9,82E-02
9,83E-02
9,83E-02
9,83E-02
9,83E-02
9,84E-02
9,85E-02
9,85E-02
9,85E-02
9,85E-02
9,85E-02
9,86E-02
9,89E-02
9,90E-02
9,91E-02
9,93E-02

9,94E-02
9,94E-02
9,95E-02
9,95E-02
9,95E-02
9,96E-02
9,96E-02
9,97E-02
9,98E-02
9,98E-02
9,99E-02
9,99E-02
9,99E-02
1,00E-01
1,00E-01
1,00E-01
1,00E-01
1,00E-01
1,01E-01
1,01E-01
1,01E-01
1,01E-01
1,01E-01
1,01E-01
1,01E-01
1,01E-01
1,01E-01
1,01E-01
1,01E-01
1,01E-01
1,01E-01
1,02E-01
1,02E-01
1,02E-01
1,02E-01
1,02E-01
1,02E-01
1,02E-01
1,02E-01
1,02E-01
1,02E-01
1,02E-01
1,02E-01
1,02E-01
1,02E-01
1,02E-01

1,02E-01
1,02E-01
1,02E-01
1,02E-01
1,02E-01
1,02E-01
1,02E-01
1,03E-01
1,03E-01
1,03E-01
1,03E-01
1,03E-01
1,03E-01
1,03E-01
1,03E-01
1,03E-01
1,03E-01
1,03E-01
1,03E-01
1,03E-01
1,03E-01
1,03E-01
1,03E-01
1,03E-01
1,03E-01
1,03E-01
1,04E-01
1,04E-01
1,04E-01
1,04E-01
1,04E-01
1,04E-01
1,04E-01
1,04E-01
1,04E-01
1,04E-01
1,04E-01
1,04E-01
1,04E-01
1,04E-01
1,04E-01
1,04E-01
1,04E-01
1,04E-01
1,04E-01
1,04E-01

1,04E-01
1,04E-01
1,04E-01
1,04E-01
1,04E-01
1,05E-01
1,05E-01
1,05E-01
1,05E-01
1,05E-01
1,05E-01
1,05E-01
1,05E-01
1,05E-01
1,05E-01
1,05E-01
1,05E-01
1,05E-01
1,05E-01
1,05E-01
1,05E-01
1,05E-01
1,05E-01
1,05E-01
1,05E-01
1,05E-01
1,05E-01
1,06E-01
1,06E-01
1,06E-01
1,06E-01
1,06E-01
1,06E-01
1,06E-01
1,06E-01
1,06E-01
1,06E-01
1,06E-01
1,06E-01
1,06E-01
1,06E-01
1,06E-01
1,06E-01
1,06E-01
1,06E-01
1,06E-01

1,06E-01
1,07E-01
1,07E-01
1,07E-01
1,07E-01
1,07E-01
1,07E-01
1,07E-01
1,07E-01
1,07E-01
1,07E-01
1,07E-01
1,07E-01
1,07E-01
1,07E-01
1,07E-01
1,07E-01
1,07E-01
1,07E-01
1,07E-01
1,07E-01
1,07E-01
1,08E-01
1,08E-01
1,08E-01
1,08E-01
1,08E-01
1,08E-01
1,08E-01
1,08E-01
1,08E-01
1,08E-01
1,08E-01
1,08E-01
1,08E-01
1,08E-01
1,08E-01
1,08E-01
1,08E-01
1,08E-01
1,09E-01
1,09E-01
1,09E-01
1,09E-01
1,09E-01
1,09E-01

1,09E-01
1,09E-01
1,09E-01
1,09E-01
1,09E-01
1,09E-01
1,09E-01
1,09E-01
1,09E-01
1,09E-01
1,09E-01
1,09E-01
1,09E-01
1,09E-01
1,09E-01
1,09E-01
1,09E-01
1,09E-01
1,09E-01
1,09E-01
1,09E-01
1,09E-01
1,09E-01
1,10E-01
1,10E-01
1,10E-01
1,10E-01
1,10E-01
1,10E-01
1,10E-01
1,10E-01
1,10E-01
1,10E-01
1,10E-01
1,10E-01
1,11E-01
1,11E-01
1,11E-01
1,11E-01
1,11E-01
1,11E-01
1,11E-01
1,11E-01
1,11E-01
1,11E-01
1,11E-01

1,11E-01
1,11E-01
1,11E-01
1,11E-01
1,11E-01
1,11E-01
1,12E-01
1,12E-01
1,12E-01
1,12E-01
1,12E-01
1,12E-01
1,12E-01
1,12E-01
1,12E-01
1,12E-01
1,12E-01
1,12E-01
1,12E-01
1,12E-01
1,12E-01
1,12E-01
1,12E-01
1,12E-01
1,12E-01
1,13E-01
1,13E-01
1,13E-01
1,13E-01
1,13E-01
1,13E-01
1,13E-01
1,13E-01
1,13E-01
1,13E-01
1,13E-01
1,13E-01
1,13E-01
1,13E-01
1,13E-01
1,13E-01
1,13E-01
1,13E-01
1,13E-01
1,13E-01

| |
|---|
| 1,13E-01 |
| 1,13E-01 |
| 1,14E-01 |
| 1,14E-01 |
| 1,14E-01 |
| 1,14E-01 |
| 1,14E-01 |
| 1,14E-01 |
| 1,14E-01 |
| 1,14E-01 |
| 1,14E-01 |
| 1,14E-01 |
| 1,14E-01 |
| 1,14E-01 |
| 1,14E-01 |
| 1,14E-01 |
| 1,14E-01 |
| 1,14E-01 |
| 1,14E-01 |
| 1,14E-01 |
| 1,14E-01 |
| 1,14E-01 |
| 1,15E-01 |
| 1,15E-01 |
| 1,15E-01 |
| 1,15E-01 |
| 1,15E-01 |
| 1,15E-01 |
| 1,15E-01 |
| 1,15E-01 |
| 1,15E-01 |
| 1,15E-01 |
| 1,15E-01 |
| 1,15E-01 |
| 1,15E-01 |
| 1,15E-01 |
| 1,15E-01 |
| 1,15E-01 |
| 1,15E-01 |
| 1,15E-01 |
| 1,15E-01 |
| 1,15E-01 |
| 1,15E-01 |
| 1,15E-01 |
| 1,15E-01 |
| 1,15E-01 |

1,16E-01
1,16E-01
1,16E-01
1,16E-01
1,16E-01
1,16E-01
1,16E-01
1,16E-01
1,16E-01
1,16E-01
1,16E-01
1,16E-01
1,16E-01
1,16E-01
1,16E-01
1,17E-01
1,17E-01
1,17E-01
1,17E-01
1,17E-01
1,17E-01
1,17E-01
1,17E-01
1,17E-01
1,17E-01
1,17E-01
1,17E-01
1,17E-01
1,17E-01
1,17E-01
1,17E-01
1,17E-01
1,17E-01
1,17E-01
1,17E-01
1,18E-01
1,18E-01
1,18E-01
1,18E-01
1,18E-01
1,18E-01
1,18E-01
1,18E-01
1,18E-01
1,18E-01
1,18E-01

1,18E-01
1,18E-01
1,19E-01
1,19E-01
1,19E-01
1,19E-01
1,19E-01
1,19E-01
1,19E-01
1,19E-01
1,19E-01
1,19E-01
1,19E-01
1,19E-01
1,19E-01
1,20E-01
1,20E-01
1,20E-01
1,20E-01
1,20E-01
1,20E-01
1,20E-01
1,20E-01
1,20E-01
1,20E-01
1,20E-01
1,20E-01
1,20E-01
1,20E-01
1,20E-01
1,20E-01
1,20E-01
1,20E-01
1,20E-01
1,20E-01
1,20E-01
1,20E-01
1,20E-01
1,20E-01
1,20E-01
1,20E-01
1,21E-01
1,21E-01
1,21E-01
1,21E-01
1,21E-01

1,21E-01
1,21E-01
1,21E-01
1,21E-01
1,21E-01
1,21E-01
1,21E-01
1,21E-01
1,21E-01
1,21E-01
1,21E-01
1,21E-01
1,21E-01
1,21E-01
1,21E-01
1,21E-01
1,21E-01
1,21E-01
1,21E-01
1,21E-01
1,21E-01
1,21E-01
1,21E-01
1,22E-01
1,22E-01
1,22E-01
1,22E-01
1,22E-01
1,22E-01
1,22E-01
1,22E-01
1,22E-01
1,22E-01
1,22E-01
1,22E-01
1,22E-01
1,22E-01
1,22E-01
1,22E-01
1,22E-01
1,23E-01
1,23E-01
1,23E-01
1,23E-01
1,23E-01
1,23E-01

1,23E-01
1,23E-01
1,23E-01
1,23E-01
1,23E-01
1,23E-01
1,23E-01
1,23E-01
1,23E-01
1,23E-01
1,23E-01
1,24E-01
1,24E-01
1,24E-01
1,24E-01
1,24E-01
1,24E-01
1,24E-01
1,24E-01
1,24E-01
1,24E-01
1,24E-01
1,24E-01
1,24E-01
1,24E-01
1,24E-01
1,24E-01
1,24E-01
1,24E-01
1,24E-01
1,24E-01
1,24E-01
1,24E-01
1,25E-01
1,25E-01
1,25E-01
1,25E-01
1,25E-01
1,25E-01
1,25E-01
1,25E-01
1,25E-01
1,26E-01
1,26E-01
1,26E-01
1,26E-01

1,26E-01
1,26E-01
1,26E-01
1,26E-01
1,26E-01
1,26E-01
1,26E-01
1,26E-01
1,26E-01
1,26E-01
1,26E-01
1,26E-01
1,26E-01
1,26E-01
1,26E-01
1,26E-01
1,26E-01
1,26E-01
1,26E-01
1,27E-01
1,27E-01
1,27E-01
1,27E-01
1,27E-01
1,27E-01
1,27E-01
1,27E-01
1,27E-01
1,27E-01
1,27E-01
1,27E-01
1,27E-01
1,27E-01
1,27E-01
1,27E-01
1,27E-01
1,27E-01
1,27E-01
1,28E-01
1,28E-01
1,28E-01
1,28E-01
1,28E-01
1,28E-01
1,28E-01
1,28E-01

| |
|---|
| 1,28E-01 |
| 1,28E-01 |
| 1,28E-01 |
| 1,28E-01 |
| 1,28E-01 |
| 1,28E-01 |
| 1,28E-01 |
| 1,28E-01 |
| 1,28E-01 |
| 1,29E-01 |
| 1,29E-01 |
| 1,29E-01 |
| 1,29E-01 |
| 1,29E-01 |
| 1,29E-01 |
| 1,29E-01 |
| 1,29E-01 |
| 1,29E-01 |
| 1,29E-01 |
| 1,29E-01 |
| 1,29E-01 |
| 1,29E-01 |
| 1,30E-01 |
| 1,30E-01 |
| 1,30E-01 |
| 1,30E-01 |
| 1,30E-01 |
| 1,30E-01 |
| 1,30E-01 |
| 1,30E-01 |
| 1,30E-01 |
| 1,30E-01 |
| 1,30E-01 |
| 1,30E-01 |
| 1,31E-01 |
| 1,31E-01 |
| 1,31E-01 |
| 1,31E-01 |
| 1,31E-01 |
| 1,31E-01 |
| 1,31E-01 |
| 1,31E-01 |
| 1,31E-01 |
| 1,31E-01 |
| 1,31E-01 |
| 1,31E-01 |

1,31E-01
1,31E-01
1,32E-01
1,32E-01
1,32E-01
1,32E-01
1,32E-01
1,32E-01
1,32E-01
1,32E-01
1,32E-01
1,32E-01
1,32E-01
1,32E-01
1,32E-01
1,32E-01
1,32E-01
1,32E-01
1,32E-01
1,33E-01
1,33E-01
1,33E-01
1,33E-01
1,33E-01
1,33E-01
1,33E-01
1,33E-01
1,33E-01
1,33E-01
1,34E-01
1,34E-01
1,34E-01
1,34E-01
1,34E-01
1,34E-01
1,34E-01
1,34E-01
1,34E-01
1,34E-01
1,34E-01
1,34E-01
1,34E-01
1,35E-01
1,35E-01
1,35E-01
1,35E-01

1,35E-01
1,35E-01
1,35E-01
1,35E-01
1,35E-01
1,35E-01
1,35E-01
1,35E-01
1,35E-01
1,35E-01
1,35E-01
1,35E-01
1,35E-01
1,35E-01
1,35E-01
1,35E-01
1,35E-01
1,35E-01
1,35E-01
1,35E-01
1,35E-01
1,36E-01
1,36E-01
1,36E-01
1,36E-01
1,36E-01
1,36E-01
1,36E-01
1,36E-01
1,36E-01
1,36E-01
1,36E-01
1,36E-01
1,36E-01
1,37E-01
1,37E-01
1,37E-01
1,37E-01
1,37E-01
1,37E-01
1,37E-01
1,37E-01
1,37E-01
1,37E-01
1,37E-01
1,37E-01

1,37E-01
1,37E-01
1,37E-01
1,37E-01
1,37E-01
1,37E-01
1,38E-01
1,38E-01
1,38E-01
1,38E-01
1,38E-01
1,38E-01
1,38E-01
1,38E-01
1,38E-01
1,38E-01
1,38E-01
1,39E-01
1,39E-01
1,39E-01
1,39E-01
1,39E-01
1,39E-01
1,39E-01
1,39E-01
1,39E-01
1,39E-01
1,39E-01
1,39E-01
1,39E-01
1,39E-01
1,39E-01
1,39E-01
1,39E-01
1,39E-01
1,39E-01
1,39E-01
1,39E-01
1,39E-01
1,39E-01
1,39E-01
1,39E-01
1,39E-01
1,39E-01
1,39E-01
1,40E-01

| |
|---|
| 1,40E-01 |
| 1,40E-01 |
| 1,40E-01 |
| 1,40E-01 |
| 1,40E-01 |
| 1,40E-01 |
| 1,40E-01 |
| 1,40E-01 |
| 1,40E-01 |
| 1,40E-01 |
| 1,40E-01 |
| 1,40E-01 |
| 1,41E-01 |
| 1,41E-01 |
| 1,41E-01 |
| 1,41E-01 |
| 1,41E-01 |
| 1,41E-01 |
| 1,41E-01 |
| 1,41E-01 |
| 1,41E-01 |
| 1,41E-01 |
| 1,41E-01 |
| 1,41E-01 |
| 1,41E-01 |
| 1,41E-01 |
| 1,41E-01 |
| 1,42E-01 |
| 1,42E-01 |
| 1,42E-01 |
| 1,42E-01 |
| 1,42E-01 |
| 1,42E-01 |
| 1,42E-01 |
| 1,42E-01 |
| 1,42E-01 |
| 1,42E-01 |
| 1,42E-01 |
| 1,42E-01 |
| 1,42E-01 |
| 1,42E-01 |
| 1,42E-01 |
| 1,42E-01 |
| 1,42E-01 |
| 1,42E-01 |
| 1,42E-01 |

1,42E-01
1,42E-01
1,42E-01
1,43E-01
1,43E-01
1,43E-01
1,43E-01
1,43E-01
1,43E-01
1,43E-01
1,43E-01
1,44E-01
1,44E-01
1,44E-01
1,44E-01
1,44E-01
1,44E-01
1,44E-01
1,44E-01
1,44E-01
1,44E-01
1,44E-01
1,44E-01
1,44E-01
1,44E-01
1,44E-01
1,44E-01
1,44E-01
1,44E-01
1,44E-01
1,44E-01
1,44E-01
1,44E-01
1,44E-01
1,45E-01
1,45E-01
1,45E-01
1,45E-01
1,45E-01
1,45E-01
1,45E-01
1,45E-01
1,45E-01
1,45E-01
1,45E-01
1,45E-01

1,45E-01
1,45E-01
1,45E-01
1,45E-01
1,45E-01
1,46E-01
1,46E-01
1,46E-01
1,46E-01
1,46E-01
1,46E-01
1,46E-01
1,46E-01
1,46E-01
1,46E-01
1,46E-01
1,46E-01
1,46E-01
1,46E-01
1,46E-01
1,46E-01
1,46E-01
1,46E-01
1,47E-01
1,47E-01
1,47E-01
1,47E-01
1,47E-01
1,47E-01
1,47E-01
1,47E-01
1,47E-01
1,47E-01
1,47E-01
1,47E-01
1,47E-01
1,47E-01
1,48E-01
1,48E-01
1,48E-01
1,48E-01
1,48E-01
1,48E-01
1,48E-01
1,48E-01
1,48E-01

1,48E-01
1,48E-01
1,48E-01
1,48E-01
1,48E-01
1,48E-01
1,48E-01
1,48E-01
1,49E-01
1,49E-01
1,49E-01
1,49E-01
1,49E-01
1,49E-01
1,49E-01
1,49E-01
1,49E-01
1,50E-01
1,50E-01
1,50E-01
1,50E-01
1,50E-01
1,50E-01
1,50E-01
1,50E-01
1,50E-01
1,50E-01
1,50E-01
1,51E-01
1,51E-01
1,51E-01
1,51E-01
1,51E-01
1,51E-01
1,51E-01
1,52E-01
1,52E-01
1,52E-01
1,52E-01
1,52E-01
1,52E-01
1,52E-01
1,52E-01
1,53E-01
1,53E-01
1,53E-01

| |
|---|
| 1,53E-01 |
| 1,53E-01 |
| 1,53E-01 |
| 1,53E-01 |
| 1,53E-01 |
| 1,53E-01 |
| 1,53E-01 |
| 1,53E-01 |
| 1,53E-01 |
| 1,53E-01 |
| 1,53E-01 |
| 1,53E-01 |
| 1,53E-01 |
| 1,53E-01 |
| 1,53E-01 |
| 1,53E-01 |
| 1,53E-01 |
| 1,53E-01 |
| 1,53E-01 |
| 1,54E-01 |
| 1,54E-01 |
| 1,54E-01 |
| 1,54E-01 |
| 1,54E-01 |
| 1,54E-01 |
| 1,54E-01 |
| 1,54E-01 |
| 1,54E-01 |
| 1,54E-01 |
| 1,54E-01 |
| 1,54E-01 |
| 1,55E-01 |
| 1,55E-01 |
| 1,55E-01 |
| 1,55E-01 |
| 1,55E-01 |
| 1,55E-01 |
| 1,55E-01 |
| 1,55E-01 |
| 1,56E-01 |
| 1,56E-01 |
| 1,56E-01 |
| 1,56E-01 |
| 1,56E-01 |
| 1,56E-01 |
| 1,56E-01 |

1,56E-01
1,56E-01
1,56E-01
1,56E-01
1,56E-01
1,56E-01
1,56E-01
1,56E-01
1,56E-01
1,56E-01
1,57E-01
1,57E-01
1,57E-01
1,57E-01
1,57E-01
1,57E-01
1,57E-01
1,57E-01
1,57E-01
1,57E-01
1,57E-01
1,58E-01
1,58E-01
1,58E-01
1,58E-01
1,58E-01
1,58E-01
1,58E-01
1,58E-01
1,58E-01
1,58E-01
1,58E-01
1,58E-01
1,58E-01
1,59E-01
1,59E-01
1,59E-01
1,59E-01
1,59E-01
1,59E-01
1,59E-01
1,59E-01
1,59E-01
1,59E-01
1,59E-01
1,59E-01

1,59E-01
1,59E-01
1,59E-01
1,59E-01
1,59E-01
1,59E-01
1,59E-01
1,59E-01
1,60E-01
1,60E-01
1,60E-01
1,60E-01
1,60E-01
1,60E-01
1,60E-01
1,60E-01
1,60E-01
1,60E-01
1,60E-01
1,61E-01
1,61E-01
1,61E-01
1,61E-01
1,61E-01
1,61E-01
1,62E-01
1,62E-01
1,62E-01
1,62E-01
1,62E-01
1,62E-01
1,62E-01
1,62E-01
1,62E-01
1,63E-01
1,63E-01
1,63E-01
1,63E-01
1,63E-01
1,63E-01
1,63E-01
1,64E-01
1,64E-01
1,64E-01
1,64E-01
1,64E-01

1,64E-01
1,64E-01
1,64E-01
1,64E-01
1,64E-01
1,64E-01
1,64E-01
1,65E-01
1,65E-01
1,65E-01
1,65E-01
1,65E-01
1,65E-01
1,65E-01
1,65E-01
1,66E-01
1,66E-01
1,66E-01
1,66E-01
1,66E-01
1,66E-01
1,66E-01
1,66E-01
1,66E-01
1,66E-01
1,67E-01
1,67E-01
1,67E-01
1,67E-01
1,67E-01
1,67E-01
1,67E-01
1,68E-01
1,68E-01
1,68E-01
1,68E-01
1,68E-01
1,68E-01
1,68E-01
1,68E-01
1,68E-01
1,68E-01
1,69E-01
1,69E-01
1,69E-01
1,69E-01

1,69E-01
1,69E-01
1,69E-01
1,69E-01
1,69E-01
1,69E-01
1,69E-01
1,69E-01
1,69E-01
1,69E-01
1,69E-01
1,69E-01
1,69E-01
1,70E-01
1,70E-01
1,70E-01
1,70E-01
1,70E-01
1,70E-01
1,70E-01
1,71E-01
1,71E-01
1,71E-01
1,71E-01
1,71E-01
1,71E-01
1,71E-01
1,71E-01
1,71E-01
1,71E-01
1,71E-01
1,72E-01
1,72E-01
1,72E-01
1,72E-01
1,72E-01
1,72E-01
1,72E-01
1,72E-01
1,73E-01
1,73E-01
1,73E-01
1,73E-01
1,73E-01
1,73E-01
1,73E-01

1,73E-01
1,73E-01
1,73E-01
1,73E-01
1,73E-01
1,73E-01
1,73E-01
1,74E-01
1,74E-01
1,74E-01
1,74E-01
1,74E-01
1,74E-01
1,74E-01
1,74E-01
1,74E-01
1,74E-01
1,74E-01
1,74E-01
1,75E-01
1,75E-01
1,75E-01
1,75E-01
1,75E-01
1,75E-01
1,75E-01
1,75E-01
1,75E-01
1,75E-01
1,75E-01
1,75E-01
1,75E-01
1,75E-01
1,75E-01
1,75E-01
1,76E-01
1,76E-01
1,76E-01
1,76E-01
1,76E-01
1,76E-01
1,76E-01
1,76E-01
1,76E-01
1,76E-01
1,76E-01

1,77E-01
1,77E-01
1,77E-01
1,77E-01
1,77E-01
1,77E-01
1,77E-01
1,77E-01
1,78E-01
1,78E-01
1,78E-01
1,78E-01
1,78E-01
1,78E-01
1,78E-01
1,78E-01
1,78E-01
1,79E-01
1,79E-01
1,79E-01
1,79E-01
1,79E-01
1,79E-01
1,79E-01
1,79E-01
1,79E-01
1,80E-01
1,80E-01
1,80E-01
1,80E-01
1,80E-01
1,80E-01
1,80E-01
1,80E-01
1,80E-01
1,80E-01
1,80E-01
1,80E-01
1,81E-01
1,81E-01
1,81E-01
1,81E-01
1,81E-01
1,81E-01
1,81E-01
1,82E-01

1,82E-01
1,82E-01
1,82E-01
1,82E-01
1,82E-01
1,82E-01
1,82E-01
1,83E-01
1,83E-01
1,83E-01
1,83E-01
1,83E-01
1,83E-01
1,83E-01
1,83E-01
1,83E-01
1,83E-01
1,83E-01
1,83E-01
1,83E-01
1,83E-01
1,83E-01
1,83E-01
1,83E-01
1,83E-01
1,83E-01
1,83E-01
1,83E-01
1,84E-01
1,84E-01
1,84E-01
1,84E-01
1,84E-01
1,84E-01
1,84E-01
1,84E-01
1,84E-01
1,84E-01
1,84E-01
1,84E-01
1,84E-01
1,84E-01
1,84E-01
1,84E-01
1,84E-01
1,84E-01

1,85E-01
1,85E-01
1,85E-01
1,85E-01
1,85E-01
1,85E-01
1,85E-01
1,85E-01
1,85E-01
1,85E-01
1,85E-01
1,85E-01
1,85E-01
1,85E-01
1,85E-01
1,85E-01
1,85E-01
1,85E-01
1,86E-01
1,86E-01
1,86E-01
1,86E-01
1,86E-01
1,87E-01
1,87E-01
1,87E-01
1,87E-01
1,87E-01
1,87E-01
1,87E-01
1,87E-01
1,88E-01
1,88E-01
1,88E-01
1,88E-01
1,88E-01
1,88E-01
1,88E-01
1,88E-01
1,88E-01
1,88E-01
1,88E-01
1,88E-01
1,89E-01
1,89E-01
1,89E-01

1,89E-01
1,89E-01
1,89E-01
1,89E-01
1,89E-01
1,89E-01
1,89E-01
1,89E-01
1,89E-01
1,89E-01
1,89E-01
1,89E-01
1,89E-01
1,89E-01
1,89E-01
1,89E-01
1,89E-01
1,90E-01
1,90E-01
1,90E-01
1,90E-01
1,90E-01
1,90E-01
1,90E-01
1,90E-01
1,90E-01
1,90E-01
1,90E-01
1,90E-01
1,90E-01
1,91E-01
1,91E-01
1,91E-01
1,91E-01
1,91E-01
1,91E-01
1,91E-01
1,91E-01
1,91E-01
1,91E-01
1,91E-01
1,91E-01
1,91E-01
1,92E-01
1,92E-01
1,92E-01

| |
|---|
| 1,92E-01 |
| 1,92E-01 |
| 1,92E-01 |
| 1,92E-01 |
| 1,92E-01 |
| 1,92E-01 |
| 1,92E-01 |
| 1,93E-01 |
| 1,93E-01 |
| 1,93E-01 |
| 1,93E-01 |
| 1,93E-01 |
| 1,93E-01 |
| 1,93E-01 |
| 1,93E-01 |
| 1,93E-01 |
| 1,93E-01 |
| 1,93E-01 |
| 1,93E-01 |
| 1,93E-01 |
| 1,93E-01 |
| 1,93E-01 |
| 1,93E-01 |
| 1,94E-01 |
| 1,94E-01 |
| 1,94E-01 |
| 1,94E-01 |
| 1,94E-01 |
| 1,94E-01 |
| 1,94E-01 |
| 1,94E-01 |
| 1,94E-01 |
| 1,95E-01 |
| 1,95E-01 |
| 1,95E-01 |
| 1,95E-01 |
| 1,95E-01 |
| 1,95E-01 |
| 1,95E-01 |
| 1,95E-01 |
| 1,95E-01 |
| 1,95E-01 |
| 1,95E-01 |
| 1,95E-01 |
| 1,95E-01 |
| 1,95E-01 |

1,96E-01
1,96E-01
1,96E-01
1,96E-01
1,96E-01
1,96E-01
1,96E-01
1,96E-01
1,97E-01
1,97E-01
1,97E-01
1,97E-01
1,97E-01
1,97E-01
1,97E-01
1,98E-01
1,98E-01
1,98E-01
1,98E-01
1,98E-01
1,98E-01
1,98E-01
1,98E-01
1,98E-01
1,98E-01
1,98E-01
1,98E-01
1,98E-01
1,98E-01
1,98E-01
1,99E-01
1,99E-01
1,99E-01
1,99E-01
1,99E-01
1,99E-01
2,00E-01
2,00E-01
2,00E-01
2,00E-01
2,00E-01
2,00E-01
2,00E-01
2,00E-01
2,00E-01
2,00E-01

2,00E-01
2,00E-01
2,00E-01
2,00E-01
2,00E-01
2,00E-01
2,01E-01
2,01E-01
2,01E-01
2,01E-01
2,01E-01
2,01E-01
2,01E-01
2,01E-01
2,01E-01
2,02E-01
2,02E-01
2,02E-01
2,03E-01
2,03E-01
2,03E-01
2,03E-01
2,03E-01
2,03E-01
2,03E-01
2,03E-01
2,03E-01
2,03E-01
2,03E-01
2,03E-01
2,04E-01
2,04E-01
2,04E-01
2,04E-01
2,04E-01
2,04E-01
2,04E-01
2,04E-01
2,04E-01
2,04E-01
2,04E-01
2,04E-01
2,05E-01
2,05E-01
2,05E-01
2,05E-01

2,05E-01
2,05E-01
2,05E-01
2,06E-01
2,06E-01
2,06E-01
2,06E-01
2,06E-01
2,06E-01
2,06E-01
2,06E-01
2,07E-01
2,07E-01
2,07E-01
2,07E-01
2,07E-01
2,07E-01
2,07E-01
2,07E-01
2,07E-01
2,07E-01
2,07E-01
2,07E-01
2,07E-01
2,07E-01
2,07E-01
2,07E-01
2,07E-01
2,07E-01
2,07E-01
2,08E-01
2,08E-01
2,08E-01
2,08E-01
2,08E-01
2,08E-01
2,08E-01
2,08E-01
2,08E-01
2,08E-01
2,08E-01
2,08E-01
2,08E-01
2,08E-01
2,08E-01
2,08E-01

2,08E-01
2,08E-01
2,08E-01
2,08E-01
2,08E-01
2,08E-01
2,08E-01
2,09E-01
2,09E-01
2,09E-01
2,09E-01
2,09E-01
2,09E-01
2,09E-01
2,09E-01
2,09E-01
2,09E-01
2,09E-01
2,09E-01
2,09E-01
2,09E-01
2,09E-01
2,09E-01
2,09E-01
2,09E-01
2,09E-01
2,09E-01
2,09E-01
2,10E-01
2,10E-01
2,10E-01
2,10E-01
2,10E-01
2,10E-01
2,10E-01
2,10E-01
2,10E-01
2,10E-01
2,11E-01
2,11E-01
2,11E-01
2,11E-01
2,11E-01
2,11E-01
2,11E-01
2,11E-01

2,11E-01
2,11E-01
2,12E-01
2,12E-01
2,12E-01
2,12E-01
2,12E-01
2,12E-01
2,12E-01
2,12E-01
2,12E-01
2,13E-01
2,13E-01
2,13E-01
2,13E-01
2,13E-01
2,13E-01
2,13E-01
2,14E-01
2,14E-01
2,14E-01
2,14E-01
2,14E-01
2,15E-01
2,15E-01
2,15E-01
2,15E-01
2,15E-01
2,15E-01
2,16E-01
2,16E-01
2,16E-01
2,16E-01
2,16E-01
2,16E-01
2,16E-01
2,16E-01
2,16E-01
2,16E-01
2,16E-01
2,16E-01
2,16E-01
2,16E-01
2,17E-01
2,17E-01
2,17E-01

2,17E-01
2,18E-01
2,18E-01
2,18E-01
2,18E-01
2,18E-01
2,18E-01
2,18E-01
2,18E-01
2,18E-01
2,18E-01
2,18E-01
2,18E-01
2,18E-01
2,19E-01
2,19E-01
2,19E-01
2,19E-01
2,19E-01
2,19E-01
2,19E-01
2,19E-01
2,19E-01
2,20E-01
2,21E-01
2,21E-01
2,21E-01
2,21E-01
2,21E-01
2,21E-01
2,21E-01
2,21E-01
2,21E-01
2,21E-01
2,21E-01
2,21E-01
2,21E-01
2,22E-01
2,22E-01
2,22E-01
2,22E-01
2,22E-01
2,22E-01
2,22E-01
2,22E-01
2,22E-01

2,23E-01
2,23E-01
2,23E-01
2,24E-01
2,24E-01
2,24E-01
2,24E-01
2,24E-01
2,24E-01
2,24E-01
2,24E-01
2,24E-01
2,24E-01
2,24E-01
2,24E-01
2,24E-01
2,25E-01
2,25E-01
2,25E-01
2,25E-01
2,25E-01
2,25E-01
2,25E-01
2,25E-01
2,25E-01
2,26E-01
2,26E-01
2,26E-01
2,26E-01
2,26E-01
2,26E-01
2,26E-01
2,26E-01
2,26E-01
2,26E-01
2,27E-01
2,27E-01
2,27E-01
2,27E-01
2,27E-01
2,27E-01
2,27E-01
2,27E-01
2,27E-01
2,27E-01
2,28E-01

2,28E-01
2,28E-01
2,28E-01
2,28E-01
2,28E-01
2,28E-01
2,28E-01
2,28E-01
2,29E-01
2,29E-01
2,29E-01
2,29E-01
2,29E-01
2,29E-01
2,29E-01
2,29E-01
2,29E-01
2,29E-01
2,29E-01
2,29E-01
2,29E-01
2,29E-01
2,30E-01
2,30E-01
2,30E-01
2,30E-01
2,30E-01
2,30E-01
2,30E-01
2,31E-01
2,31E-01
2,31E-01
2,31E-01
2,31E-01
2,31E-01
2,31E-01
2,31E-01
2,31E-01
2,31E-01
2,31E-01
2,31E-01
2,31E-01
2,31E-01
2,31E-01
2,32E-01
2,32E-01

2,32E-01
2,32E-01
2,32E-01
2,32E-01
2,32E-01
2,32E-01
2,32E-01
2,32E-01
2,32E-01
2,32E-01
2,32E-01
2,32E-01
2,33E-01
2,33E-01
2,33E-01
2,33E-01
2,33E-01
2,33E-01
2,33E-01
2,33E-01
2,33E-01
2,33E-01
2,33E-01
2,33E-01
2,33E-01
2,33E-01
2,33E-01
2,33E-01
2,34E-01
2,34E-01
2,34E-01
2,34E-01
2,34E-01
2,34E-01
2,34E-01
2,34E-01
2,34E-01
2,34E-01
2,34E-01
2,34E-01
2,34E-01
2,34E-01
2,35E-01
2,35E-01
2,35E-01
2,35E-01

2,35E-01
2,35E-01
2,35E-01
2,35E-01
2,35E-01
2,35E-01
2,35E-01
2,35E-01
2,36E-01
2,36E-01
2,36E-01
2,36E-01
2,36E-01
2,37E-01
2,37E-01
2,37E-01
2,37E-01
2,37E-01
2,37E-01
2,37E-01
2,37E-01
2,37E-01
2,37E-01
2,37E-01
2,37E-01
2,37E-01
2,37E-01
2,38E-01
2,38E-01
2,38E-01
2,38E-01
2,38E-01
2,38E-01
2,38E-01
2,38E-01
2,38E-01
2,38E-01
2,38E-01
2,38E-01
2,38E-01
2,38E-01
2,38E-01
2,38E-01
2,38E-01
2,39E-01
2,39E-01

2,39E-01
2,39E-01
2,39E-01
2,39E-01
2,39E-01
2,39E-01
2,39E-01
2,39E-01
2,39E-01
2,39E-01
2,40E-01
2,40E-01
2,40E-01
2,40E-01
2,40E-01
2,40E-01
2,40E-01
2,40E-01
2,40E-01
2,40E-01
2,40E-01
2,41E-01
2,41E-01
2,41E-01
2,41E-01
2,41E-01
2,41E-01
2,41E-01
2,41E-01
2,41E-01
2,41E-01
2,41E-01
2,41E-01
2,41E-01
2,41E-01
2,42E-01
2,42E-01
2,42E-01
2,42E-01
2,42E-01
2,42E-01
2,42E-01
2,42E-01
2,42E-01
2,42E-01
2,42E-01

2,42E-01
2,42E-01
2,42E-01
2,42E-01
2,42E-01
2,42E-01
2,43E-01
2,43E-01
2,43E-01
2,43E-01
2,43E-01
2,43E-01
2,43E-01
2,43E-01
2,43E-01
2,43E-01
2,44E-01
2,44E-01
2,44E-01
2,44E-01
2,44E-01
2,44E-01
2,44E-01
2,44E-01
2,44E-01
2,44E-01
2,44E-01
2,45E-01
2,45E-01
2,45E-01
2,45E-01
2,45E-01
2,45E-01
2,45E-01
2,45E-01
2,45E-01
2,46E-01
2,46E-01
2,46E-01
2,46E-01
2,46E-01
2,46E-01
2,46E-01
2,46E-01
2,47E-01
2,47E-01

| |
|---|
| 2,47E-01 |
| 2,47E-01 |
| 2,47E-01 |
| 2,47E-01 |
| 2,47E-01 |
| 2,48E-01 |
| 2,48E-01 |
| 2,48E-01 |
| 2,48E-01 |
| 2,48E-01 |
| 2,48E-01 |
| 2,48E-01 |
| 2,48E-01 |
| 2,48E-01 |
| 2,48E-01 |
| 2,48E-01 |
| 2,48E-01 |
| 2,48E-01 |
| 2,49E-01 |
| 2,49E-01 |
| 2,49E-01 |
| 2,49E-01 |
| 2,50E-01 |
| 2,50E-01 |
| 2,50E-01 |
| 2,50E-01 |
| 2,50E-01 |
| 2,51E-01 |
| 2,51E-01 |
| 2,51E-01 |
| 2,52E-01 |
| 2,52E-01 |
| 2,52E-01 |
| 2,52E-01 |
| 2,52E-01 |
| 2,52E-01 |
| 2,52E-01 |
| 2,52E-01 |
| 2,52E-01 |
| 2,52E-01 |
| 2,52E-01 |
| 2,52E-01 |
| 2,53E-01 |
| 2,53E-01 |
| 2,53E-01 |
| 2,53E-01 |

2,53E-01
2,53E-01
2,53E-01
2,53E-01
2,53E-01
2,54E-01
2,54E-01
2,54E-01
2,54E-01
2,54E-01
2,54E-01
2,54E-01
2,54E-01
2,54E-01
2,54E-01
2,55E-01
2,55E-01
2,55E-01
2,55E-01
2,55E-01
2,55E-01
2,55E-01
2,55E-01
2,55E-01
2,56E-01
2,56E-01
2,56E-01
2,56E-01
2,56E-01
2,56E-01
2,56E-01
2,56E-01
2,56E-01
2,56E-01
2,56E-01
2,56E-01
2,56E-01
2,56E-01
2,56E-01
2,57E-01
2,57E-01
2,57E-01
2,57E-01
2,57E-01
2,57E-01
2,57E-01

| |
|---|
| 2,57E-01 |
| 2,57E-01 |
| 2,57E-01 |
| 2,57E-01 |
| 2,57E-01 |
| 2,57E-01 |
| 2,57E-01 |
| 2,57E-01 |
| 2,58E-01 |
| 2,58E-01 |
| 2,58E-01 |
| 2,58E-01 |
| 2,58E-01 |
| 2,58E-01 |
| 2,58E-01 |
| 2,58E-01 |
| 2,58E-01 |
| 2,59E-01 |
| 2,59E-01 |
| 2,59E-01 |
| 2,60E-01 |
| 2,60E-01 |
| 2,60E-01 |
| 2,60E-01 |
| 2,60E-01 |
| 2,60E-01 |
| 2,60E-01 |
| 2,60E-01 |
| 2,60E-01 |
| 2,60E-01 |
| 2,60E-01 |
| 2,60E-01 |
| 2,60E-01 |
| 2,60E-01 |
| 2,60E-01 |
| 2,61E-01 |
| 2,61E-01 |
| 2,61E-01 |
| 2,61E-01 |
| 2,61E-01 |
| 2,61E-01 |
| 2,62E-01 |
| 2,62E-01 |
| 2,62E-01 |
| 2,62E-01 |
| 2,62E-01 |

2,62E-01
2,62E-01
2,62E-01
2,63E-01
2,63E-01
2,63E-01
2,63E-01
2,64E-01
2,64E-01
2,64E-01
2,64E-01
2,64E-01
2,64E-01
2,64E-01
2,64E-01
2,64E-01
2,64E-01
2,64E-01
2,64E-01
2,65E-01
2,65E-01
2,65E-01
2,65E-01
2,65E-01
2,65E-01
2,65E-01
2,65E-01
2,65E-01
2,65E-01
2,65E-01
2,65E-01
2,65E-01
2,65E-01
2,65E-01
2,66E-01
2,66E-01
2,66E-01
2,66E-01
2,66E-01
2,66E-01
2,66E-01
2,66E-01
2,66E-01
2,66E-01
2,66E-01
2,67E-01

| |
|---|
| 2,67E-01 |
| 2,67E-01 |
| 2,67E-01 |
| 2,67E-01 |
| 2,67E-01 |
| 2,67E-01 |
| 2,67E-01 |
| 2,67E-01 |
| 2,68E-01 |
| 2,68E-01 |
| 2,68E-01 |
| 2,68E-01 |
| 2,68E-01 |
| 2,68E-01 |
| 2,68E-01 |
| 2,68E-01 |
| 2,68E-01 |
| 2,68E-01 |
| 2,69E-01 |
| 2,69E-01 |
| 2,69E-01 |
| 2,69E-01 |
| 2,69E-01 |
| 2,69E-01 |
| 2,69E-01 |
| 2,69E-01 |
| 2,69E-01 |
| 2,69E-01 |
| 2,69E-01 |
| 2,69E-01 |
| 2,69E-01 |
| 2,70E-01 |
| 2,70E-01 |
| 2,70E-01 |
| 2,70E-01 |
| 2,70E-01 |
| 2,70E-01 |
| 2,71E-01 |
| 2,71E-01 |
| 2,71E-01 |
| 2,71E-01 |
| 2,71E-01 |
| 2,71E-01 |
| 2,71E-01 |
| 2,71E-01 |
| 2,72E-01 |

2,72E-01
2,72E-01
2,72E-01
2,72E-01
2,72E-01
2,72E-01
2,72E-01
2,72E-01
2,72E-01
2,72E-01
2,72E-01
2,72E-01
2,73E-01
2,73E-01
2,73E-01
2,73E-01
2,73E-01
2,73E-01
2,74E-01
2,74E-01
2,74E-01
2,75E-01
2,75E-01
2,75E-01
2,75E-01
2,76E-01
2,76E-01
2,76E-01
2,76E-01
2,76E-01
2,76E-01
2,77E-01
2,77E-01
2,77E-01
2,77E-01
2,77E-01
2,77E-01
2,77E-01
2,77E-01
2,78E-01
2,78E-01
2,78E-01
2,78E-01
2,78E-01
2,79E-01
2,79E-01

2,79E-01
2,79E-01
2,79E-01
2,79E-01
2,79E-01
2,80E-01
2,80E-01
2,80E-01
2,80E-01
2,80E-01
2,80E-01
2,80E-01
2,80E-01
2,80E-01
2,80E-01
2,81E-01
2,81E-01
2,81E-01
2,81E-01
2,81E-01
2,81E-01
2,81E-01
2,81E-01
2,81E-01
2,81E-01
2,81E-01
2,81E-01
2,81E-01
2,81E-01
2,81E-01
2,81E-01
2,81E-01
2,81E-01
2,82E-01
2,82E-01
2,82E-01
2,82E-01
2,82E-01
2,82E-01
2,82E-01
2,82E-01
2,82E-01
2,82E-01
2,83E-01
2,83E-01
2,83E-01

2,83E-01
2,83E-01
2,83E-01
2,83E-01
2,83E-01
2,83E-01
2,83E-01
2,83E-01
2,83E-01
2,83E-01
2,84E-01
2,84E-01
2,84E-01
2,84E-01
2,84E-01
2,84E-01
2,84E-01
2,84E-01
2,84E-01
2,84E-01
2,85E-01
2,85E-01
2,85E-01
2,85E-01
2,85E-01
2,85E-01
2,85E-01
2,85E-01
2,85E-01
2,86E-01
2,86E-01
2,86E-01
2,86E-01
2,86E-01
2,86E-01
2,86E-01
2,87E-01
2,87E-01
2,87E-01
2,87E-01
2,87E-01
2,87E-01
2,87E-01
2,87E-01
2,87E-01
2,88E-01

2,88E-01
2,88E-01
2,88E-01
2,88E-01
2,88E-01
2,88E-01
2,88E-01
2,89E-01
2,89E-01
2,89E-01
2,89E-01
2,89E-01
2,89E-01
2,89E-01
2,89E-01
2,89E-01
2,89E-01
2,89E-01
2,89E-01
2,89E-01
2,89E-01
2,89E-01
2,90E-01
2,90E-01
2,90E-01
2,90E-01
2,90E-01
2,90E-01
2,90E-01
2,90E-01
2,90E-01
2,90E-01
2,90E-01
2,91E-01
2,91E-01
2,91E-01
2,91E-01
2,91E-01
2,91E-01
2,91E-01
2,91E-01
2,91E-01
2,91E-01
2,91E-01
2,92E-01
2,92E-01

2,92E-01
2,92E-01
2,92E-01
2,92E-01
2,92E-01
2,92E-01
2,92E-01
2,92E-01
2,92E-01
2,92E-01
2,92E-01
2,92E-01
2,92E-01
2,92E-01
2,92E-01
2,93E-01
2,93E-01
2,93E-01
2,93E-01
2,93E-01
2,93E-01
2,93E-01
2,93E-01
2,93E-01
2,93E-01
2,93E-01
2,93E-01
2,93E-01
2,94E-01
2,94E-01
2,94E-01
2,94E-01
2,94E-01
2,94E-01
2,94E-01
2,95E-01
2,95E-01
2,95E-01
2,95E-01
2,95E-01
2,95E-01
2,95E-01
2,95E-01
2,96E-01
2,97E-01
2,97E-01

2,97E-01
2,97E-01
2,98E-01
2,98E-01
2,98E-01
2,98E-01
2,98E-01
2,98E-01
2,99E-01
2,99E-01
2,99E-01
2,99E-01
3,00E-01
3,00E-01
3,00E-01
3,00E-01
3,00E-01
3,01E-01
3,01E-01
3,01E-01
3,01E-01
3,01E-01
3,01E-01
3,01E-01
3,02E-01
3,02E-01
3,03E-01
3,03E-01
3,03E-01
3,03E-01
3,03E-01
3,03E-01
3,03E-01
3,04E-01
3,04E-01
3,04E-01
3,04E-01
3,04E-01
3,04E-01
3,04E-01
3,04E-01
3,05E-01
3,05E-01
3,05E-01
3,05E-01
3,05E-01

3,05E-01
3,05E-01
3,06E-01
3,06E-01
3,06E-01
3,06E-01
3,07E-01
3,07E-01
3,07E-01
3,07E-01
3,07E-01
3,07E-01
3,07E-01
3,07E-01
3,07E-01
3,08E-01
3,08E-01
3,08E-01
3,08E-01
3,08E-01
3,08E-01
3,08E-01
3,08E-01
3,08E-01
3,08E-01
3,08E-01
3,08E-01
3,08E-01
3,09E-01
3,09E-01
3,09E-01
3,09E-01
3,09E-01
3,09E-01
3,09E-01
3,09E-01
3,09E-01
3,09E-01
3,10E-01
3,10E-01
3,10E-01
3,11E-01
3,11E-01
3,11E-01
3,11E-01
3,12E-01

3,12E-01
3,12E-01
3,12E-01
3,12E-01
3,12E-01
3,12E-01
3,13E-01
3,13E-01
3,13E-01
3,13E-01
3,13E-01
3,13E-01
3,14E-01
3,14E-01
3,14E-01
3,14E-01
3,14E-01
3,14E-01
3,14E-01
3,14E-01
3,14E-01
3,14E-01
3,15E-01
3,15E-01
3,15E-01
3,15E-01
3,15E-01
3,15E-01
3,15E-01
3,15E-01
3,15E-01
3,15E-01
3,15E-01
3,16E-01
3,16E-01
3,16E-01
3,16E-01
3,17E-01
3,17E-01
3,17E-01
3,17E-01
3,17E-01
3,17E-01
3,17E-01
3,18E-01
3,18E-01

3,18E-01
3,18E-01
3,18E-01
3,18E-01
3,18E-01
3,18E-01
3,19E-01
3,19E-01
3,19E-01
3,19E-01
3,19E-01
3,19E-01
3,19E-01
3,20E-01
3,20E-01
3,20E-01
3,20E-01
3,20E-01
3,20E-01
3,20E-01
3,20E-01
3,21E-01
3,21E-01
3,21E-01
3,21E-01
3,21E-01
3,21E-01
3,21E-01
3,21E-01
3,21E-01
3,21E-01
3,21E-01
3,21E-01
3,21E-01
3,21E-01
3,21E-01
3,21E-01
3,22E-01
3,22E-01
3,23E-01
3,23E-01
3,23E-01
3,23E-01
3,23E-01
3,23E-01
3,23E-01

3,24E-01
3,24E-01
3,24E-01
3,24E-01
3,24E-01
3,24E-01
3,24E-01
3,25E-01
3,25E-01
3,25E-01
3,25E-01
3,25E-01
3,26E-01
3,26E-01
3,26E-01
3,26E-01
3,26E-01
3,26E-01
3,26E-01
3,26E-01
3,26E-01
3,26E-01
3,26E-01
3,26E-01
3,27E-01
3,27E-01
3,27E-01
3,27E-01
3,27E-01
3,27E-01
3,27E-01
3,27E-01
3,27E-01
3,28E-01
3,28E-01
3,28E-01
3,28E-01
3,28E-01
3,28E-01
3,28E-01
3,28E-01
3,28E-01
3,28E-01
3,28E-01
3,28E-01
3,28E-01

| |
|---|
| 3,28E-01 |
| 3,28E-01 |
| 3,29E-01 |
| 3,29E-01 |
| 3,29E-01 |
| 3,29E-01 |
| 3,29E-01 |
| 3,29E-01 |
| 3,29E-01 |
| 3,29E-01 |
| 3,29E-01 |
| 3,29E-01 |
| 3,29E-01 |
| 3,29E-01 |
| 3,30E-01 |
| 3,30E-01 |
| 3,30E-01 |
| 3,31E-01 |
| 3,31E-01 |
| 3,31E-01 |
| 3,31E-01 |
| 3,31E-01 |
| 3,32E-01 |
| 3,32E-01 |
| 3,32E-01 |
| 3,32E-01 |
| 3,32E-01 |
| 3,32E-01 |
| 3,32E-01 |
| 3,32E-01 |
| 3,32E-01 |
| 3,32E-01 |
| 3,32E-01 |
| 3,32E-01 |
| 3,32E-01 |
| 3,33E-01 |
| 3,33E-01 |
| 3,33E-01 |
| 3,33E-01 |
| 3,33E-01 |
| 3,33E-01 |
| 3,34E-01 |
| 3,34E-01 |
| 3,34E-01 |
| 3,34E-01 |
| 3,34E-01 |

3,34E-01
3,34E-01
3,34E-01
3,34E-01
3,34E-01
3,34E-01
3,35E-01
3,35E-01
3,35E-01
3,35E-01
3,35E-01
3,35E-01
3,35E-01
3,35E-01
3,36E-01
3,36E-01
3,36E-01
3,36E-01
3,36E-01
3,36E-01
3,36E-01
3,37E-01
3,37E-01
3,37E-01
3,37E-01
3,37E-01
3,37E-01
3,38E-01
3,38E-01
3,38E-01
3,38E-01
3,38E-01
3,38E-01
3,38E-01
3,39E-01
3,39E-01
3,39E-01
3,39E-01
3,39E-01
3,39E-01
3,40E-01
3,40E-01
3,40E-01
3,40E-01
3,40E-01
3,40E-01

3,40E-01
3,40E-01
3,41E-01
3,41E-01
3,41E-01
3,41E-01
3,41E-01
3,41E-01
3,41E-01
3,41E-01
3,42E-01
3,42E-01
3,42E-01
3,42E-01
3,42E-01
3,42E-01
3,42E-01
3,42E-01
3,42E-01
3,42E-01
3,43E-01
3,43E-01
3,43E-01
3,43E-01
3,43E-01
3,43E-01
3,43E-01
3,43E-01
3,43E-01
3,43E-01
3,44E-01
3,44E-01
3,44E-01
3,44E-01
3,44E-01
3,44E-01
3,44E-01
3,44E-01
3,44E-01
3,44E-01
3,44E-01
3,45E-01
3,45E-01
3,45E-01
3,45E-01
3,45E-01

| |
|---|
| 3,45E-01 |
| 3,45E-01 |
| 3,45E-01 |
| 3,45E-01 |
| 3,45E-01 |
| 3,45E-01 |
| 3,45E-01 |
| 3,45E-01 |
| 3,46E-01 |
| 3,46E-01 |
| 3,46E-01 |
| 3,46E-01 |
| 3,46E-01 |
| 3,46E-01 |
| 3,46E-01 |
| 3,46E-01 |
| 3,46E-01 |
| 3,47E-01 |
| 3,47E-01 |
| 3,47E-01 |
| 3,47E-01 |
| 3,47E-01 |
| 3,48E-01 |
| 3,48E-01 |
| 3,48E-01 |
| 3,49E-01 |
| 3,49E-01 |
| 3,49E-01 |
| 3,49E-01 |
| 3,49E-01 |
| 3,49E-01 |
| 3,49E-01 |
| 3,49E-01 |
| 3,49E-01 |
| 3,49E-01 |
| 3,49E-01 |
| 3,49E-01 |
| 3,50E-01 |
| 3,50E-01 |
| 3,50E-01 |
| 3,50E-01 |
| 3,50E-01 |
| 3,50E-01 |
| 3,50E-01 |
| 3,50E-01 |
| 3,50E-01 |

3,50E-01
3,51E-01
3,51E-01
3,51E-01
3,51E-01
3,51E-01
3,51E-01
3,51E-01
3,51E-01
3,51E-01
3,51E-01
3,51E-01
3,51E-01
3,51E-01
3,52E-01
3,52E-01
3,52E-01
3,52E-01
3,52E-01
3,52E-01
3,52E-01
3,52E-01
3,53E-01
3,53E-01
3,53E-01
3,53E-01
3,53E-01
3,54E-01
3,54E-01
3,54E-01
3,54E-01
3,54E-01
3,54E-01
3,55E-01
3,55E-01
3,55E-01
3,55E-01
3,55E-01
3,55E-01
3,56E-01
3,56E-01
3,56E-01
3,56E-01
3,56E-01
3,56E-01
3,56E-01

3,56E-01
3,56E-01
3,56E-01
3,56E-01
3,56E-01
3,56E-01
3,57E-01
3,57E-01
3,57E-01
3,57E-01
3,58E-01
3,58E-01
3,58E-01
3,58E-01
3,58E-01
3,58E-01
3,58E-01
3,58E-01
3,58E-01
3,58E-01
3,58E-01
3,58E-01
3,59E-01
3,59E-01
3,59E-01
3,59E-01
3,59E-01
3,60E-01
3,60E-01
3,60E-01
3,60E-01
3,60E-01
3,60E-01
3,60E-01
3,61E-01
3,61E-01
3,61E-01
3,61E-01
3,61E-01
3,61E-01
3,61E-01
3,61E-01
3,61E-01
3,61E-01
3,61E-01
3,61E-01

3,61E-01
3,61E-01
3,61E-01
3,62E-01
3,62E-01
3,62E-01
3,62E-01
3,62E-01
3,62E-01
3,63E-01
3,63E-01
3,63E-01
3,63E-01
3,63E-01
3,63E-01
3,63E-01
3,64E-01
3,64E-01
3,64E-01
3,64E-01
3,64E-01
3,65E-01
3,65E-01
3,65E-01
3,65E-01
3,65E-01
3,65E-01
3,65E-01
3,65E-01
3,65E-01
3,66E-01
3,66E-01
3,66E-01
3,66E-01
3,67E-01
3,67E-01
3,67E-01
3,67E-01
3,67E-01
3,67E-01
3,67E-01
3,67E-01
3,67E-01
3,68E-01
3,68E-01
3,69E-01

3,69E-01
3,69E-01
3,69E-01
3,69E-01
3,69E-01
3,69E-01
3,69E-01
3,69E-01
3,70E-01
3,70E-01
3,70E-01
3,70E-01
3,70E-01
3,71E-01
3,71E-01
3,71E-01
3,71E-01
3,71E-01
3,71E-01
3,71E-01
3,71E-01
3,72E-01
3,72E-01
3,72E-01
3,72E-01
3,72E-01
3,72E-01
3,72E-01
3,72E-01
3,72E-01
3,73E-01
3,73E-01
3,73E-01
3,73E-01
3,73E-01
3,73E-01
3,73E-01
3,73E-01
3,73E-01
3,74E-01
3,74E-01
3,74E-01
3,74E-01
3,74E-01
3,74E-01
3,75E-01

3,75E-01
3,75E-01
3,76E-01
3,76E-01
3,76E-01
3,76E-01
3,76E-01
3,76E-01
3,77E-01
3,77E-01
3,77E-01
3,77E-01
3,78E-01
3,78E-01
3,78E-01
3,78E-01
3,78E-01
3,78E-01
3,78E-01
3,78E-01
3,78E-01
3,78E-01
3,79E-01
3,79E-01
3,79E-01
3,80E-01
3,80E-01
3,80E-01
3,80E-01
3,80E-01
3,80E-01
3,80E-01
3,81E-01
3,81E-01
3,81E-01
3,82E-01
3,82E-01
3,82E-01
3,82E-01
3,82E-01
3,82E-01
3,82E-01
3,82E-01
3,82E-01
3,82E-01
3,82E-01

3,82E-01
3,82E-01
3,83E-01
3,83E-01
3,83E-01
3,83E-01
3,83E-01
3,83E-01
3,83E-01
3,83E-01
3,83E-01
3,84E-01
3,84E-01
3,84E-01
3,84E-01
3,85E-01
3,85E-01
3,85E-01
3,85E-01
3,86E-01
3,86E-01
3,86E-01
3,86E-01
3,86E-01
3,86E-01
3,86E-01
3,86E-01
3,86E-01
3,86E-01
3,86E-01
3,86E-01
3,86E-01
3,86E-01
3,86E-01
3,86E-01
3,87E-01
3,87E-01
3,87E-01
3,87E-01
3,87E-01
3,87E-01
3,87E-01
3,88E-01
3,88E-01
3,88E-01
3,88E-01

3,88E-01
3,88E-01
3,88E-01
3,89E-01
3,89E-01
3,89E-01
3,90E-01
3,90E-01
3,90E-01
3,90E-01
3,90E-01
3,90E-01
3,90E-01
3,90E-01
3,90E-01
3,91E-01
3,91E-01
3,91E-01
3,92E-01
3,92E-01
3,92E-01
3,93E-01
3,93E-01
3,93E-01
3,93E-01
3,93E-01
3,94E-01
3,94E-01
3,94E-01
3,94E-01
3,94E-01
3,94E-01
3,94E-01
3,94E-01
3,94E-01
3,94E-01
3,95E-01
3,95E-01
3,95E-01
3,95E-01
3,95E-01
3,95E-01
3,95E-01
3,95E-01
3,96E-01
3,96E-01

3,96E-01
3,96E-01
3,97E-01
3,97E-01
3,97E-01
3,97E-01
3,98E-01
3,98E-01
3,98E-01
3,98E-01
3,98E-01
3,98E-01
3,98E-01
3,99E-01
3,99E-01
3,99E-01
3,99E-01
3,99E-01
3,99E-01
3,99E-01
3,99E-01
4,00E-01
4,00E-01
4,00E-01
4,00E-01
4,00E-01
4,01E-01
4,01E-01
4,01E-01
4,01E-01
4,01E-01
4,01E-01
4,01E-01
4,01E-01
4,01E-01
4,01E-01
4,02E-01
4,02E-01
4,02E-01
4,02E-01
4,02E-01
4,03E-01
4,03E-01
4,03E-01
4,03E-01
4,03E-01

| |
|---|
| 4,03E-01 |
| 4,03E-01 |
| 4,04E-01 |
| 4,04E-01 |
| 4,04E-01 |
| 4,04E-01 |
| 4,04E-01 |
| 4,04E-01 |
| 4,04E-01 |
| 4,04E-01 |
| 4,04E-01 |
| 4,05E-01 |
| 4,05E-01 |
| 4,05E-01 |
| 4,05E-01 |
| 4,05E-01 |
| 4,05E-01 |
| 4,05E-01 |
| 4,05E-01 |
| 4,05E-01 |
| 4,05E-01 |
| 4,05E-01 |
| 4,06E-01 |
| 4,06E-01 |
| 4,06E-01 |
| 4,07E-01 |
| 4,07E-01 |
| 4,07E-01 |
| 4,07E-01 |
| 4,08E-01 |
| 4,08E-01 |
| 4,08E-01 |
| 4,08E-01 |
| 4,08E-01 |
| 4,08E-01 |
| 4,08E-01 |
| 4,09E-01 |
| 4,09E-01 |
| 4,09E-01 |
| 4,09E-01 |
| 4,09E-01 |
| 4,09E-01 |
| 4,09E-01 |
| 4,09E-01 |
| 4,09E-01 |
| 4,09E-01 |

4,10E-01
4,10E-01
4,10E-01
4,10E-01
4,10E-01
4,10E-01
4,10E-01
4,11E-01
4,11E-01
4,11E-01
4,11E-01
4,11E-01
4,11E-01
4,11E-01
4,11E-01
4,11E-01
4,11E-01
4,12E-01
4,12E-01
4,12E-01
4,12E-01
4,12E-01
4,12E-01
4,12E-01
4,12E-01
4,12E-01
4,12E-01
4,13E-01
4,13E-01
4,13E-01
4,13E-01
4,13E-01
4,13E-01
4,14E-01
4,14E-01
4,14E-01
4,14E-01
4,14E-01
4,14E-01
4,15E-01
4,15E-01
4,15E-01
4,15E-01
4,15E-01
4,15E-01
4,15E-01

4,15E-01
4,15E-01
4,15E-01
4,16E-01
4,16E-01
4,16E-01
4,16E-01
4,16E-01
4,16E-01
4,16E-01
4,17E-01
4,17E-01
4,17E-01
4,17E-01
4,18E-01
4,18E-01
4,18E-01
4,18E-01
4,18E-01
4,18E-01
4,19E-01
4,19E-01
4,19E-01
4,19E-01
4,19E-01
4,19E-01
4,19E-01
4,19E-01
4,20E-01
4,20E-01
4,20E-01
4,20E-01
4,20E-01
4,20E-01
4,21E-01
4,21E-01
4,21E-01
4,21E-01
4,21E-01
4,21E-01
4,21E-01
4,21E-01
4,21E-01
4,22E-01
4,22E-01
4,22E-01

4,22E-01
4,22E-01
4,22E-01
4,23E-01
4,23E-01
4,23E-01
4,24E-01
4,24E-01
4,24E-01
4,24E-01
4,24E-01
4,24E-01
4,24E-01
4,24E-01
4,24E-01
4,24E-01
4,24E-01
4,25E-01
4,25E-01
4,25E-01
4,25E-01
4,25E-01
4,25E-01
4,25E-01
4,25E-01
4,25E-01
4,25E-01
4,26E-01
4,26E-01
4,26E-01
4,26E-01
4,26E-01
4,26E-01
4,27E-01
4,27E-01
4,27E-01
4,27E-01
4,27E-01
4,27E-01
4,27E-01
4,27E-01
4,27E-01
4,27E-01
4,27E-01
4,27E-01
4,27E-01

4,28E-01
4,28E-01
4,28E-01
4,29E-01
4,29E-01
4,29E-01
4,29E-01
4,29E-01
4,30E-01
4,30E-01
4,31E-01
4,31E-01
4,31E-01
4,32E-01
4,32E-01
4,32E-01
4,32E-01
4,32E-01
4,32E-01
4,32E-01
4,33E-01
4,33E-01
4,33E-01
4,33E-01
4,33E-01
4,33E-01
4,33E-01
4,33E-01
4,33E-01
4,34E-01
4,34E-01
4,34E-01
4,34E-01
4,34E-01
4,34E-01
4,34E-01
4,34E-01
4,35E-01
4,35E-01
4,35E-01
4,35E-01
4,35E-01
4,36E-01
4,36E-01
4,36E-01
4,36E-01

4,36E-01
4,36E-01
4,36E-01
4,36E-01
4,37E-01
4,37E-01
4,37E-01
4,37E-01
4,37E-01
4,38E-01
4,38E-01
4,38E-01
4,38E-01
4,38E-01
4,38E-01
4,38E-01
4,38E-01
4,38E-01
4,38E-01
4,38E-01
4,38E-01
4,38E-01
4,38E-01
4,39E-01
4,39E-01
4,39E-01
4,39E-01
4,39E-01
4,39E-01
4,40E-01
4,40E-01
4,40E-01
4,40E-01
4,40E-01
4,41E-01
4,41E-01
4,42E-01
4,42E-01
4,42E-01
4,42E-01
4,42E-01
4,42E-01
4,42E-01
4,42E-01
4,43E-01
4,43E-01

4,43E-01
4,43E-01
4,43E-01
4,44E-01
4,45E-01
4,45E-01
4,45E-01
4,45E-01
4,45E-01
4,46E-01
4,46E-01
4,46E-01
4,46E-01
4,46E-01
4,47E-01
4,47E-01
4,47E-01
4,47E-01
4,47E-01
4,47E-01
4,47E-01
4,48E-01
4,48E-01
4,48E-01
4,48E-01
4,48E-01
4,49E-01
4,49E-01
4,49E-01
4,50E-01
4,50E-01
4,50E-01
4,50E-01
4,50E-01
4,50E-01
4,50E-01
4,50E-01
4,50E-01
4,51E-01
4,51E-01
4,51E-01
4,51E-01
4,51E-01
4,51E-01
4,52E-01
4,52E-01

4,52E-01
4,52E-01
4,52E-01
4,52E-01
4,53E-01
4,53E-01
4,53E-01
4,53E-01
4,54E-01
4,54E-01
4,54E-01
4,54E-01
4,54E-01
4,54E-01
4,54E-01
4,54E-01
4,54E-01
4,54E-01
4,55E-01
4,55E-01
4,55E-01
4,55E-01
4,55E-01
4,55E-01
4,55E-01
4,55E-01
4,56E-01
4,56E-01
4,56E-01
4,56E-01
4,56E-01
4,56E-01
4,57E-01
4,57E-01
4,57E-01
4,58E-01
4,58E-01
4,58E-01
4,58E-01
4,58E-01
4,58E-01
4,59E-01
4,59E-01
4,59E-01
4,59E-01
4,59E-01

4,59E-01
4,60E-01
4,60E-01
4,60E-01
4,60E-01
4,61E-01
4,61E-01
4,61E-01
4,61E-01
4,61E-01
4,62E-01
4,62E-01
4,63E-01
4,63E-01
4,63E-01
4,63E-01
4,64E-01
4,64E-01
4,64E-01
4,64E-01
4,64E-01
4,64E-01
4,64E-01
4,64E-01
4,64E-01
4,64E-01
4,64E-01
4,65E-01
4,65E-01
4,65E-01
4,65E-01
4,66E-01
4,66E-01
4,66E-01
4,66E-01
4,66E-01
4,66E-01
4,66E-01
4,66E-01
4,66E-01
4,66E-01
4,67E-01
4,67E-01
4,67E-01
4,67E-01
4,67E-01

4,67E-01
4,68E-01
4,68E-01
4,68E-01
4,68E-01
4,68E-01
4,68E-01
4,68E-01
4,69E-01
4,69E-01
4,69E-01
4,70E-01
4,70E-01
4,70E-01
4,70E-01
4,70E-01
4,71E-01
4,71E-01
4,71E-01
4,71E-01
4,71E-01
4,71E-01
4,72E-01
4,72E-01
4,72E-01
4,72E-01
4,72E-01
4,72E-01
4,72E-01
4,72E-01
4,73E-01
4,73E-01
4,73E-01
4,74E-01
4,75E-01
4,75E-01
4,75E-01
4,76E-01
4,76E-01
4,76E-01
4,76E-01
4,76E-01
4,77E-01
4,78E-01
4,78E-01
4,78E-01

4,78E-01
4,78E-01
4,79E-01
4,79E-01
4,79E-01
4,79E-01
4,79E-01
4,79E-01
4,79E-01
4,79E-01
4,79E-01
4,80E-01
4,80E-01
4,80E-01
4,80E-01
4,80E-01
4,81E-01
4,81E-01
4,81E-01
4,81E-01
4,81E-01
4,82E-01
4,82E-01
4,82E-01
4,82E-01
4,82E-01
4,82E-01
4,83E-01
4,83E-01
4,83E-01
4,83E-01
4,84E-01
4,84E-01
4,84E-01
4,84E-01
4,84E-01
4,84E-01
4,84E-01
4,84E-01
4,85E-01
4,85E-01
4,85E-01
4,85E-01
4,85E-01
4,87E-01
4,87E-01

4,87E-01
4,88E-01
4,88E-01
4,88E-01
4,88E-01
4,88E-01
4,88E-01
4,88E-01
4,88E-01
4,88E-01
4,89E-01
4,89E-01
4,89E-01
4,90E-01
4,90E-01
4,90E-01
4,90E-01
4,90E-01
4,91E-01
4,91E-01
4,91E-01
4,91E-01
4,91E-01
4,92E-01
4,92E-01
4,92E-01
4,93E-01
4,93E-01
4,93E-01
4,93E-01
4,93E-01
4,94E-01
4,94E-01
4,94E-01
4,94E-01
4,94E-01
4,94E-01
4,94E-01
4,94E-01
4,94E-01
4,94E-01
4,95E-01
4,95E-01
4,96E-01
4,96E-01
4,96E-01

4,96E-01
4,96E-01
4,96E-01
4,96E-01
4,96E-01
4,96E-01
4,96E-01
4,96E-01
4,96E-01
4,96E-01
4,96E-01
4,96E-01
4,96E-01
4,96E-01
4,97E-01
4,97E-01
4,97E-01
4,97E-01
4,97E-01
4,98E-01
4,98E-01
4,98E-01
4,99E-01
4,99E-01
4,99E-01
4,99E-01
4,99E-01
4,99E-01
4,99E-01
4,99E-01
4,99E-01
4,99E-01
4,99E-01
5,00E-01
5,00E-01
5,00E-01
5,00E-01
5,00E-01
5,00E-01
5,00E-01
5,01E-01
5,01E-01
5,01E-01
5,01E-01
5,01E-01
5,01E-01

| |
|---|
| 5,01E-01 |
| 5,01E-01 |
| 5,02E-01 |
| 5,02E-01 |
| 5,02E-01 |
| 5,02E-01 |
| 5,02E-01 |
| 5,03E-01 |
| 5,03E-01 |
| 5,03E-01 |
| 5,03E-01 |
| 5,03E-01 |
| 5,03E-01 |
| 5,03E-01 |
| 5,03E-01 |
| 5,04E-01 |
| 5,04E-01 |
| 5,04E-01 |
| 5,04E-01 |
| 5,04E-01 |
| 5,04E-01 |
| 5,05E-01 |
| 5,05E-01 |
| 5,05E-01 |
| 5,05E-01 |
| 5,05E-01 |
| 5,05E-01 |
| 5,05E-01 |
| 5,05E-01 |
| 5,05E-01 |
| 5,06E-01 |
| 5,06E-01 |
| 5,06E-01 |
| 5,06E-01 |
| 5,06E-01 |
| 5,07E-01 |
| 5,07E-01 |
| 5,07E-01 |
| 5,08E-01 |
| 5,08E-01 |
| 5,08E-01 |
| 5,08E-01 |
| 5,08E-01 |
| 5,08E-01 |
| 5,08E-01 |
| 5,08E-01 |

5,08E-01
5,09E-01
5,09E-01
5,09E-01
5,09E-01
5,09E-01
5,09E-01
5,09E-01
5,09E-01
5,09E-01
5,09E-01
5,09E-01
5,09E-01
5,09E-01
5,09E-01
5,09E-01
5,10E-01
5,10E-01
5,10E-01
5,10E-01
5,11E-01
5,11E-01
5,11E-01
5,11E-01
5,12E-01
5,12E-01
5,12E-01
5,12E-01
5,12E-01
5,12E-01
5,12E-01
5,13E-01
5,13E-01
5,13E-01
5,13E-01
5,13E-01
5,13E-01
5,13E-01
5,13E-01
5,13E-01
5,14E-01
5,14E-01
5,14E-01
5,14E-01
5,15E-01
5,15E-01

5,15E-01
5,15E-01
5,15E-01
5,15E-01
5,15E-01
5,15E-01
5,16E-01
5,16E-01
5,16E-01
5,17E-01
5,17E-01
5,18E-01
5,18E-01
5,18E-01
5,18E-01
5,18E-01
5,19E-01
5,19E-01
5,19E-01
5,19E-01
5,19E-01
5,19E-01
5,19E-01
5,19E-01
5,19E-01
5,20E-01
5,20E-01
5,20E-01
5,20E-01
5,20E-01
5,20E-01
5,20E-01
5,21E-01
5,21E-01
5,21E-01
5,21E-01
5,21E-01
5,21E-01
5,21E-01
5,21E-01
5,22E-01
5,22E-01
5,22E-01
5,22E-01
5,22E-01
5,22E-01

5,23E-01
5,23E-01
5,23E-01
5,23E-01
5,23E-01
5,24E-01
5,24E-01
5,24E-01
5,24E-01
5,24E-01
5,24E-01
5,24E-01
5,25E-01
5,25E-01
5,25E-01
5,25E-01
5,25E-01
5,25E-01
5,25E-01
5,26E-01
5,26E-01
5,26E-01
5,26E-01
5,26E-01
5,27E-01
5,27E-01
5,27E-01
5,27E-01
5,27E-01
5,27E-01
5,27E-01
5,27E-01
5,27E-01
5,27E-01
5,28E-01
5,28E-01
5,28E-01
5,28E-01
5,28E-01
5,28E-01
5,28E-01
5,28E-01
5,28E-01
5,29E-01
5,29E-01
5,29E-01

5,29E-01
5,29E-01
5,29E-01
5,30E-01
5,30E-01
5,30E-01
5,30E-01
5,30E-01
5,31E-01
5,31E-01
5,31E-01
5,31E-01
5,31E-01
5,31E-01
5,32E-01
5,32E-01
5,32E-01
5,32E-01
5,32E-01
5,32E-01
5,32E-01
5,33E-01
5,33E-01
5,33E-01
5,34E-01
5,34E-01
5,34E-01
5,34E-01
5,34E-01
5,34E-01
5,34E-01
5,34E-01
5,35E-01
5,35E-01
5,35E-01
5,35E-01
5,35E-01
5,35E-01
5,35E-01
5,36E-01
5,36E-01
5,36E-01
5,36E-01
5,36E-01
5,37E-01
5,37E-01

| |
|---|
| 5,37E-01 |
| 5,37E-01 |
| 5,37E-01 |
| 5,38E-01 |
| 5,38E-01 |
| 5,38E-01 |
| 5,38E-01 |
| 5,38E-01 |
| 5,38E-01 |
| 5,38E-01 |
| 5,38E-01 |
| 5,38E-01 |
| 5,38E-01 |
| 5,38E-01 |
| 5,38E-01 |
| 5,38E-01 |
| 5,38E-01 |
| 5,38E-01 |
| 5,39E-01 |
| 5,39E-01 |
| 5,39E-01 |
| 5,39E-01 |
| 5,39E-01 |
| 5,39E-01 |
| 5,39E-01 |
| 5,39E-01 |
| 5,39E-01 |
| 5,39E-01 |
| 5,39E-01 |
| 5,39E-01 |
| 5,40E-01 |
| 5,40E-01 |
| 5,41E-01 |
| 5,41E-01 |
| 5,41E-01 |
| 5,41E-01 |
| 5,41E-01 |
| 5,41E-01 |
| 5,41E-01 |
| 5,42E-01 |
| 5,42E-01 |
| 5,42E-01 |
| 5,42E-01 |
| 5,42E-01 |
| 5,42E-01 |
| 5,42E-01 |
| 5,42E-01 |

5,43E-01
5,43E-01
5,43E-01
5,43E-01
5,43E-01
5,44E-01
5,44E-01
5,44E-01
5,44E-01
5,45E-01
5,45E-01
5,45E-01
5,46E-01
5,46E-01
5,46E-01
5,46E-01
5,46E-01
5,46E-01
5,47E-01
5,47E-01
5,47E-01
5,47E-01
5,47E-01
5,47E-01
5,48E-01
5,48E-01
5,48E-01
5,48E-01
5,48E-01
5,49E-01
5,49E-01
5,50E-01
5,50E-01
5,50E-01
5,50E-01
5,50E-01
5,50E-01
5,50E-01
5,51E-01
5,51E-01
5,52E-01
5,52E-01
5,52E-01
5,52E-01
5,52E-01
5,52E-01

5,52E-01
5,52E-01
5,52E-01
5,52E-01
5,53E-01
5,53E-01
5,53E-01
5,53E-01
5,53E-01
5,53E-01
5,53E-01
5,53E-01
5,53E-01
5,53E-01
5,53E-01
5,54E-01
5,54E-01
5,55E-01
5,55E-01
5,55E-01
5,55E-01
5,55E-01
5,55E-01
5,56E-01
5,56E-01
5,56E-01
5,56E-01
5,56E-01
5,56E-01
5,56E-01
5,56E-01
5,56E-01
5,57E-01
5,57E-01
5,57E-01
5,57E-01
5,58E-01
5,58E-01
5,58E-01
5,58E-01
5,58E-01
5,59E-01
5,60E-01
5,60E-01
5,60E-01
5,60E-01

5,60E-01
5,60E-01
5,60E-01
5,60E-01
5,60E-01
5,61E-01
5,61E-01
5,61E-01
5,61E-01
5,61E-01
5,62E-01
5,62E-01
5,62E-01
5,62E-01
5,62E-01
5,62E-01
5,62E-01
5,62E-01
5,62E-01
5,62E-01
5,62E-01
5,62E-01
5,62E-01
5,62E-01
5,62E-01
5,62E-01
5,63E-01
5,63E-01
5,64E-01
5,64E-01
5,65E-01
5,66E-01
5,66E-01
5,66E-01
5,66E-01
5,66E-01
5,67E-01
5,67E-01
5,67E-01
5,67E-01
5,67E-01
5,67E-01
5,69E-01
5,69E-01
5,69E-01
5,69E-01

| |
|---|
| 5,69E-01 |
| 5,69E-01 |
| 5,69E-01 |
| 5,69E-01 |
| 5,69E-01 |
| 5,70E-01 |
| 5,70E-01 |
| 5,70E-01 |
| 5,70E-01 |
| 5,71E-01 |
| 5,71E-01 |
| 5,72E-01 |
| 5,72E-01 |
| 5,72E-01 |
| 5,72E-01 |
| 5,73E-01 |
| 5,73E-01 |
| 5,73E-01 |
| 5,73E-01 |
| 5,73E-01 |
| 5,73E-01 |
| 5,73E-01 |
| 5,73E-01 |
| 5,74E-01 |
| 5,74E-01 |
| 5,74E-01 |
| 5,75E-01 |
| 5,75E-01 |
| 5,75E-01 |
| 5,75E-01 |
| 5,75E-01 |
| 5,76E-01 |
| 5,76E-01 |
| 5,76E-01 |
| 5,76E-01 |
| 5,76E-01 |
| 5,76E-01 |
| 5,78E-01 |
| 5,78E-01 |
| 5,78E-01 |
| 5,78E-01 |
| 5,78E-01 |
| 5,78E-01 |
| 5,78E-01 |
| 5,79E-01 |
| 5,79E-01 |

5,79E-01
5,79E-01
5,79E-01
5,79E-01
5,80E-01
5,80E-01
5,80E-01
5,80E-01
5,80E-01
5,80E-01
5,80E-01
5,80E-01
5,80E-01
5,81E-01
5,81E-01
5,81E-01
5,81E-01
5,82E-01
5,82E-01
5,82E-01
5,82E-01
5,82E-01
5,82E-01
5,83E-01
5,83E-01
5,83E-01
5,84E-01
5,84E-01
5,84E-01
5,84E-01
5,84E-01
5,84E-01
5,84E-01
5,84E-01
5,84E-01
5,84E-01
5,85E-01
5,85E-01
5,86E-01
5,86E-01
5,86E-01
5,86E-01
5,86E-01
5,86E-01
5,87E-01
5,87E-01

5,87E-01
5,87E-01
5,87E-01
5,87E-01
5,87E-01
5,87E-01
5,88E-01
5,88E-01
5,88E-01
5,88E-01
5,88E-01
5,88E-01
5,89E-01
5,89E-01
5,89E-01
5,89E-01
5,89E-01
5,89E-01
5,89E-01
5,90E-01
5,90E-01
5,90E-01
5,90E-01
5,91E-01
5,91E-01
5,91E-01
5,92E-01
5,92E-01
5,92E-01
5,92E-01
5,92E-01
5,92E-01
5,92E-01
5,92E-01
5,92E-01
5,92E-01
5,93E-01
5,94E-01
5,95E-01
5,95E-01
5,95E-01
5,95E-01
5,95E-01
5,95E-01
5,95E-01
5,95E-01

5,96E-01
5,96E-01
5,96E-01
5,96E-01
5,96E-01
5,96E-01
5,97E-01
5,97E-01
5,97E-01
5,97E-01
5,97E-01
5,97E-01
5,97E-01
5,97E-01
5,98E-01
5,98E-01
5,99E-01
5,99E-01
5,99E-01
5,99E-01
5,99E-01
5,99E-01
5,99E-01
5,99E-01
6,00E-01
6,00E-01
6,01E-01
6,01E-01
6,01E-01
6,01E-01
6,02E-01
6,02E-01
6,02E-01
6,02E-01
6,02E-01
6,02E-01
6,02E-01
6,03E-01
6,03E-01
6,03E-01
6,03E-01
6,03E-01
6,03E-01
6,03E-01
6,03E-01
6,03E-01

6,03E-01
6,04E-01
6,04E-01
6,04E-01
6,04E-01
6,04E-01
6,04E-01
6,04E-01
6,04E-01
6,05E-01
6,05E-01
6,05E-01
6,05E-01
6,05E-01
6,05E-01
6,05E-01
6,05E-01
6,06E-01
6,06E-01
6,06E-01
6,06E-01
6,06E-01
6,06E-01
6,07E-01
6,07E-01
6,08E-01
6,08E-01
6,08E-01
6,09E-01
6,09E-01
6,09E-01
6,09E-01
6,09E-01
6,09E-01
6,09E-01
6,10E-01
6,11E-01
6,11E-01
6,12E-01
6,12E-01
6,12E-01
6,12E-01
6,13E-01
6,13E-01
6,14E-01
6,14E-01

6,14E-01
6,14E-01
6,15E-01
6,15E-01
6,15E-01
6,16E-01
6,16E-01
6,16E-01
6,16E-01
6,17E-01
6,17E-01
6,17E-01
6,17E-01
6,18E-01
6,19E-01
6,19E-01
6,19E-01
6,19E-01
6,20E-01
6,20E-01
6,20E-01
6,20E-01
6,20E-01
6,21E-01
6,21E-01
6,21E-01
6,21E-01
6,21E-01
6,22E-01
6,23E-01
6,23E-01
6,23E-01
6,23E-01
6,23E-01
6,23E-01
6,24E-01
6,24E-01
6,24E-01
6,25E-01
6,25E-01
6,26E-01
6,26E-01
6,26E-01
6,26E-01
6,26E-01
6,26E-01

6,26E-01
6,26E-01
6,26E-01
6,26E-01
6,26E-01
6,26E-01
6,26E-01
6,26E-01
6,27E-01
6,27E-01
6,27E-01
6,27E-01
6,27E-01
6,27E-01
6,27E-01
6,27E-01
6,27E-01
6,27E-01
6,28E-01
6,28E-01
6,28E-01
6,28E-01
6,28E-01
6,28E-01
6,28E-01
6,29E-01
6,29E-01
6,29E-01
6,29E-01
6,29E-01
6,30E-01
6,30E-01
6,30E-01
6,30E-01
6,30E-01
6,30E-01
6,30E-01
6,30E-01
6,31E-01
6,31E-01
6,31E-01
6,31E-01
6,31E-01
6,31E-01
6,31E-01
6,31E-01

6,31E-01
6,31E-01
6,32E-01
6,32E-01
6,32E-01
6,32E-01
6,32E-01
6,32E-01
6,32E-01
6,32E-01
6,33E-01
6,33E-01
6,33E-01
6,33E-01
6,34E-01
6,34E-01
6,34E-01
6,34E-01
6,34E-01
6,34E-01
6,34E-01
6,34E-01
6,34E-01
6,34E-01
6,34E-01
6,35E-01
6,35E-01
6,35E-01
6,35E-01
6,35E-01
6,36E-01
6,36E-01
6,36E-01
6,36E-01
6,36E-01
6,37E-01
6,37E-01
6,37E-01
6,37E-01
6,38E-01
6,38E-01
6,38E-01
6,38E-01
6,38E-01
6,38E-01
6,38E-01

6,39E-01
6,41E-01
6,42E-01
6,42E-01
6,42E-01
6,43E-01
6,43E-01
6,43E-01
6,43E-01
6,43E-01
6,43E-01
6,44E-01
6,44E-01
6,44E-01
6,45E-01
6,45E-01
6,45E-01
6,45E-01
6,46E-01
6,46E-01
6,46E-01
6,46E-01
6,47E-01
6,47E-01
6,47E-01
6,48E-01
6,48E-01
6,48E-01
6,48E-01
6,48E-01
6,48E-01
6,49E-01
6,49E-01
6,49E-01
6,49E-01
6,49E-01
6,49E-01
6,50E-01
6,50E-01
6,50E-01
6,50E-01
6,50E-01
6,50E-01
6,50E-01
6,50E-01
6,50E-01

6,50E-01
6,50E-01
6,50E-01
6,50E-01
6,50E-01
6,50E-01
6,51E-01
6,51E-01
6,51E-01
6,51E-01
6,51E-01
6,52E-01
6,52E-01
6,52E-01
6,52E-01
6,52E-01
6,52E-01
6,53E-01
6,53E-01
6,53E-01
6,53E-01
6,53E-01
6,53E-01
6,53E-01
6,53E-01
6,53E-01
6,54E-01
6,55E-01
6,55E-01
6,57E-01
6,57E-01
6,57E-01
6,57E-01
6,59E-01
6,59E-01
6,59E-01
6,59E-01
6,59E-01
6,59E-01
6,60E-01
6,60E-01
6,60E-01
6,60E-01
6,60E-01
6,60E-01
6,60E-01

6,60E-01
6,60E-01
6,61E-01
6,61E-01
6,61E-01
6,61E-01
6,61E-01
6,62E-01
6,62E-01
6,62E-01
6,62E-01
6,62E-01
6,62E-01
6,63E-01
6,63E-01
6,63E-01
6,63E-01
6,63E-01
6,63E-01
6,64E-01
6,64E-01
6,64E-01
6,64E-01
6,64E-01
6,65E-01
6,65E-01
6,65E-01
6,65E-01
6,65E-01
6,66E-01
6,66E-01
6,66E-01
6,66E-01
6,67E-01
6,67E-01
6,67E-01
6,67E-01
6,67E-01
6,67E-01
6,67E-01
6,68E-01
6,68E-01
6,69E-01
6,69E-01
6,69E-01
6,70E-01

6,70E-01
6,71E-01
6,72E-01
6,72E-01
6,72E-01
6,73E-01
6,73E-01
6,73E-01
6,73E-01
6,74E-01
6,74E-01
6,75E-01
6,75E-01
6,75E-01
6,75E-01
6,75E-01
6,75E-01
6,75E-01
6,75E-01
6,75E-01
6,75E-01
6,75E-01
6,76E-01
6,76E-01
6,76E-01
6,76E-01
6,76E-01
6,76E-01
6,76E-01
6,76E-01
6,76E-01
6,76E-01
6,77E-01
6,77E-01
6,77E-01
6,77E-01
6,77E-01
6,77E-01
6,77E-01
6,78E-01
6,78E-01
6,78E-01
6,78E-01
6,78E-01
6,78E-01
6,78E-01

6,79E-01
6,79E-01
6,79E-01
6,79E-01
6,80E-01
6,80E-01
6,80E-01
6,80E-01
6,80E-01
6,80E-01
6,80E-01
6,80E-01
6,81E-01
6,81E-01
6,82E-01
6,82E-01
6,82E-01
6,82E-01
6,82E-01
6,82E-01
6,82E-01
6,82E-01
6,82E-01
6,83E-01
6,83E-01
6,83E-01
6,84E-01
6,84E-01
6,84E-01
6,84E-01
6,84E-01
6,85E-01
6,85E-01
6,85E-01
6,85E-01
6,85E-01
6,85E-01
6,85E-01
6,86E-01
6,86E-01
6,86E-01
6,86E-01
6,86E-01
6,87E-01
6,87E-01
6,88E-01

6,88E-01
6,88E-01
6,88E-01
6,88E-01
6,89E-01
6,89E-01
6,89E-01
6,89E-01
6,89E-01
6,89E-01
6,89E-01
6,90E-01
6,90E-01
6,91E-01
6,91E-01
6,91E-01
6,92E-01
6,92E-01
6,92E-01
6,93E-01
6,93E-01
6,93E-01
6,93E-01
6,93E-01
6,93E-01
6,94E-01
6,94E-01
6,94E-01
6,94E-01
6,94E-01
6,94E-01
6,94E-01
6,94E-01
6,94E-01
6,94E-01
6,94E-01
6,95E-01
6,95E-01
6,95E-01
6,95E-01
6,97E-01
6,97E-01
6,97E-01
6,97E-01
6,97E-01
6,97E-01

6,97E-01
6,97E-01
6,97E-01
6,97E-01
6,97E-01
6,98E-01
6,98E-01
6,98E-01
6,98E-01
6,98E-01
6,98E-01
6,98E-01
6,98E-01
6,98E-01
6,98E-01
6,98E-01
6,98E-01
6,99E-01
7,00E-01
7,00E-01
7,00E-01
7,01E-01
7,01E-01
7,02E-01
7,02E-01
7,02E-01
7,02E-01
7,02E-01
7,02E-01
7,02E-01
7,02E-01
7,03E-01
7,03E-01
7,03E-01
7,03E-01
7,03E-01
7,04E-01
7,04E-01
7,04E-01
7,04E-01
7,04E-01
7,04E-01
7,05E-01
7,05E-01
7,05E-01
7,05E-01

7,05E-01
7,05E-01
7,06E-01
7,06E-01
7,06E-01
7,06E-01
7,06E-01
7,06E-01
7,07E-01
7,07E-01
7,07E-01
7,07E-01
7,07E-01
7,07E-01
7,07E-01
7,07E-01
7,07E-01
7,08E-01
7,08E-01
7,08E-01
7,08E-01
7,08E-01
7,08E-01
7,08E-01
7,09E-01
7,09E-01
7,09E-01
7,09E-01
7,10E-01
7,10E-01
7,10E-01
7,10E-01
7,10E-01
7,10E-01
7,11E-01
7,11E-01
7,11E-01
7,11E-01
7,11E-01
7,12E-01
7,12E-01
7,12E-01
7,12E-01
7,13E-01
7,13E-01
7,13E-01

7,13E-01
7,14E-01
7,14E-01
7,14E-01
7,14E-01
7,14E-01
7,15E-01
7,15E-01
7,15E-01
7,16E-01
7,16E-01
7,16E-01
7,16E-01
7,16E-01
7,16E-01
7,17E-01
7,17E-01
7,17E-01
7,17E-01
7,17E-01
7,18E-01
7,18E-01
7,18E-01
7,19E-01
7,19E-01
7,19E-01
7,20E-01
7,20E-01
7,20E-01
7,20E-01
7,20E-01
7,21E-01
7,21E-01
7,21E-01
7,21E-01
7,21E-01
7,21E-01
7,21E-01
7,21E-01
7,22E-01
7,22E-01
7,22E-01
7,22E-01
7,22E-01
7,22E-01
7,22E-01

7,22E-01
7,22E-01
7,23E-01
7,24E-01
7,24E-01
7,24E-01
7,24E-01
7,24E-01
7,24E-01
7,24E-01
7,24E-01
7,24E-01
7,25E-01
7,25E-01
7,25E-01
7,25E-01
7,25E-01
7,25E-01
7,25E-01
7,25E-01
7,26E-01
7,26E-01
7,26E-01
7,26E-01
7,26E-01
7,26E-01
7,26E-01
7,27E-01
7,27E-01
7,27E-01
7,27E-01
7,27E-01
7,27E-01
7,27E-01
7,27E-01
7,27E-01
7,27E-01
7,28E-01
7,28E-01
7,28E-01
7,28E-01
7,28E-01
7,28E-01
7,29E-01
7,29E-01
7,29E-01

7,29E-01
7,30E-01
7,30E-01
7,30E-01
7,31E-01
7,31E-01
7,31E-01
7,31E-01
7,32E-01
7,32E-01
7,32E-01
7,32E-01
7,33E-01
7,34E-01
7,34E-01
7,34E-01
7,34E-01
7,34E-01
7,34E-01
7,35E-01
7,35E-01
7,36E-01
7,36E-01
7,36E-01
7,36E-01
7,36E-01
7,36E-01
7,36E-01
7,37E-01
7,37E-01
7,37E-01
7,38E-01
7,38E-01
7,38E-01
7,38E-01
7,38E-01
7,38E-01
7,38E-01
7,39E-01
7,39E-01
7,39E-01
7,39E-01
7,40E-01
7,40E-01
7,40E-01
7,40E-01

7,40E-01
7,41E-01
7,41E-01
7,41E-01
7,41E-01
7,41E-01
7,42E-01
7,42E-01
7,42E-01
7,42E-01
7,42E-01
7,42E-01
7,43E-01
7,43E-01
7,43E-01
7,43E-01
7,43E-01
7,44E-01
7,44E-01
7,44E-01
7,44E-01
7,44E-01
7,44E-01
7,44E-01
7,44E-01
7,45E-01
7,45E-01
7,45E-01
7,45E-01
7,45E-01
7,45E-01
7,45E-01
7,46E-01
7,46E-01
7,46E-01
7,46E-01
7,46E-01
7,46E-01
7,47E-01
7,47E-01
7,47E-01
7,47E-01
7,48E-01
7,49E-01
7,49E-01
7,49E-01

7,49E-01
7,49E-01
7,49E-01
7,50E-01
7,50E-01
7,50E-01
7,50E-01
7,50E-01
7,50E-01
7,50E-01
7,50E-01
7,51E-01
7,52E-01
7,52E-01
7,52E-01
7,53E-01
7,53E-01
7,53E-01
7,54E-01
7,54E-01
7,54E-01
7,54E-01
7,54E-01
7,55E-01
7,55E-01
7,55E-01
7,55E-01
7,55E-01
7,55E-01
7,55E-01
7,55E-01
7,55E-01
7,56E-01
7,56E-01
7,56E-01
7,56E-01
7,56E-01
7,56E-01
7,57E-01
7,57E-01
7,58E-01
7,58E-01
7,58E-01
7,58E-01
7,58E-01
7,59E-01

| |
|---|
| 7,59E-01 |
| 7,59E-01 |
| 7,59E-01 |
| 7,59E-01 |
| 7,60E-01 |
| 7,60E-01 |
| 7,61E-01 |
| 7,61E-01 |
| 7,62E-01 |
| 7,62E-01 |
| 7,62E-01 |
| 7,62E-01 |
| 7,62E-01 |
| 7,62E-01 |
| 7,62E-01 |
| 7,62E-01 |
| 7,62E-01 |
| 7,63E-01 |
| 7,63E-01 |
| 7,63E-01 |
| 7,63E-01 |
| 7,63E-01 |
| 7,64E-01 |
| 7,64E-01 |
| 7,64E-01 |
| 7,65E-01 |
| 7,65E-01 |
| 7,66E-01 |
| 7,66E-01 |
| 7,66E-01 |
| 7,66E-01 |
| 7,66E-01 |
| 7,66E-01 |
| 7,67E-01 |
| 7,67E-01 |
| 7,67E-01 |
| 7,67E-01 |
| 7,67E-01 |
| 7,67E-01 |
| 7,67E-01 |
| 7,67E-01 |
| 7,67E-01 |
| 7,68E-01 |
| 7,68E-01 |
| 7,68E-01 |
| 7,68E-01 |

7,68E-01
7,68E-01
7,69E-01
7,69E-01
7,69E-01
7,70E-01
7,70E-01
7,71E-01
7,71E-01
7,71E-01
7,72E-01
7,73E-01
7,73E-01
7,73E-01
7,73E-01
7,73E-01
7,73E-01
7,74E-01
7,74E-01
7,74E-01
7,74E-01
7,74E-01
7,74E-01
7,74E-01
7,74E-01
7,74E-01
7,74E-01
7,74E-01
7,74E-01
7,74E-01
7,75E-01
7,76E-01
7,76E-01
7,76E-01
7,76E-01
7,76E-01
7,76E-01
7,76E-01
7,77E-01
7,77E-01
7,77E-01
7,77E-01
7,77E-01
7,77E-01
7,77E-01
7,77E-01

7,77E-01
7,78E-01
7,78E-01
7,79E-01
7,79E-01
7,80E-01
7,80E-01
7,80E-01
7,80E-01
7,81E-01
7,81E-01
7,81E-01
7,81E-01
7,81E-01
7,82E-01
7,82E-01
7,82E-01
7,82E-01
7,83E-01
7,83E-01
7,84E-01
7,84E-01
7,84E-01
7,84E-01
7,84E-01
7,84E-01
7,84E-01
7,85E-01
7,85E-01
7,85E-01
7,85E-01
7,85E-01
7,85E-01
7,86E-01
7,86E-01
7,87E-01
7,87E-01
7,87E-01
7,87E-01
7,87E-01
7,87E-01
7,87E-01
7,88E-01
7,88E-01
7,88E-01
7,88E-01

7,88E-01
7,88E-01
7,88E-01
7,88E-01
7,88E-01
7,88E-01
7,88E-01
7,88E-01
7,88E-01
7,88E-01
7,88E-01
7,88E-01
7,88E-01
7,88E-01
7,88E-01
7,89E-01
7,89E-01
7,89E-01
7,89E-01
7,89E-01
7,90E-01
7,90E-01
7,91E-01
7,91E-01
7,91E-01
7,91E-01
7,91E-01
7,91E-01
7,91E-01
7,92E-01
7,92E-01
7,92E-01
7,92E-01
7,92E-01
7,92E-01
7,92E-01
7,92E-01
7,92E-01
7,92E-01
7,93E-01
7,93E-01
7,93E-01
7,94E-01
7,94E-01
7,94E-01
7,94E-01

7,94E-01
7,94E-01
7,94E-01
7,94E-01
7,94E-01
7,94E-01
7,95E-01
7,95E-01
7,95E-01
7,95E-01
7,95E-01
7,95E-01
7,96E-01
7,96E-01
7,96E-01
7,96E-01
7,96E-01
7,96E-01
7,96E-01
7,96E-01
7,96E-01
7,96E-01
7,96E-01
7,96E-01
7,96E-01
7,97E-01
7,97E-01
7,97E-01
7,97E-01
7,98E-01
7,98E-01
7,98E-01
7,99E-01
7,99E-01
7,99E-01
8,00E-01
8,00E-01
8,00E-01
8,00E-01
8,00E-01
8,00E-01
8,02E-01
8,02E-01
8,02E-01
8,03E-01
8,03E-01

8,05E-01
8,05E-01
8,05E-01
8,05E-01
8,05E-01
8,05E-01
8,06E-01
8,07E-01
8,07E-01
8,07E-01
8,07E-01
8,07E-01
8,07E-01
8,08E-01
8,08E-01
8,08E-01
8,08E-01
8,08E-01
8,08E-01
8,08E-01
8,08E-01
8,08E-01
8,09E-01
8,09E-01
8,09E-01
8,09E-01
8,09E-01
8,09E-01
8,09E-01
8,09E-01
8,10E-01
8,10E-01
8,10E-01
8,10E-01
8,10E-01
8,10E-01
8,11E-01
8,11E-01
8,12E-01
8,12E-01
8,12E-01
8,12E-01
8,13E-01
8,13E-01
8,13E-01
8,13E-01

8,13E-01
8,13E-01
8,13E-01
8,13E-01
8,13E-01
8,14E-01
8,14E-01
8,14E-01
8,14E-01
8,14E-01
8,15E-01
8,16E-01
8,16E-01
8,17E-01
8,17E-01
8,17E-01
8,18E-01
8,18E-01
8,19E-01
8,19E-01
8,19E-01
8,19E-01
8,20E-01
8,20E-01
8,20E-01
8,20E-01
8,21E-01
8,21E-01
8,21E-01
8,21E-01
8,21E-01
8,21E-01
8,21E-01
8,21E-01
8,21E-01
8,21E-01
8,21E-01
8,21E-01
8,23E-01
8,23E-01
8,23E-01
8,23E-01
8,24E-01
8,25E-01
8,25E-01
8,25E-01

| |
|---|
| 8,25E-01 |
| 8,25E-01 |
| 8,25E-01 |
| 8,26E-01 |
| 8,27E-01 |
| 8,27E-01 |
| 8,28E-01 |
| 8,28E-01 |
| 8,28E-01 |
| 8,28E-01 |
| 8,28E-01 |
| 8,29E-01 |
| 8,29E-01 |
| 8,30E-01 |
| 8,30E-01 |
| 8,30E-01 |
| 8,30E-01 |
| 8,30E-01 |
| 8,30E-01 |
| 8,31E-01 |
| 8,31E-01 |
| 8,31E-01 |
| 8,31E-01 |
| 8,31E-01 |
| 8,31E-01 |
| 8,31E-01 |
| 8,31E-01 |
| 8,31E-01 |
| 8,31E-01 |
| 8,31E-01 |
| 8,32E-01 |
| 8,32E-01 |
| 8,32E-01 |
| 8,32E-01 |
| 8,33E-01 |
| 8,33E-01 |
| 8,34E-01 |
| 8,34E-01 |
| 8,34E-01 |
| 8,34E-01 |
| 8,34E-01 |
| 8,34E-01 |
| 8,34E-01 |
| 8,34E-01 |
| 8,34E-01 |
| 8,34E-01 |

8,34E-01
8,34E-01
8,34E-01
8,34E-01
8,35E-01
8,36E-01
8,36E-01
8,37E-01
8,37E-01
8,37E-01
8,37E-01
8,37E-01
8,37E-01
8,37E-01
8,37E-01
8,38E-01
8,38E-01
8,38E-01
8,38E-01
8,38E-01
8,38E-01
8,38E-01
8,38E-01
8,39E-01
8,39E-01
8,39E-01
8,39E-01
8,40E-01
8,40E-01
8,40E-01
8,40E-01
8,41E-01
8,41E-01
8,42E-01
8,42E-01
8,42E-01
8,43E-01
8,43E-01
8,43E-01
8,43E-01
8,43E-01
8,43E-01
8,44E-01
8,44E-01
8,44E-01
8,44E-01

8,45E-01
8,45E-01
8,45E-01
8,45E-01
8,45E-01
8,45E-01
8,45E-01
8,45E-01
8,45E-01
8,45E-01
8,45E-01
8,46E-01
8,46E-01
8,46E-01
8,46E-01
8,46E-01
8,46E-01
8,46E-01
8,47E-01
8,47E-01
8,47E-01
8,47E-01
8,47E-01
8,47E-01
8,47E-01
8,47E-01
8,47E-01
8,47E-01
8,47E-01
8,47E-01
8,48E-01
8,48E-01
8,49E-01
8,49E-01
8,49E-01
8,49E-01
8,50E-01
8,50E-01
8,50E-01
8,51E-01
8,51E-01
8,51E-01
8,51E-01
8,51E-01
8,52E-01
8,53E-01

8,53E-01
8,53E-01
8,53E-01
8,53E-01
8,53E-01
8,53E-01
8,53E-01
8,54E-01
8,54E-01
8,54E-01
8,54E-01
8,54E-01
8,54E-01
8,55E-01
8,55E-01
8,55E-01
8,55E-01
8,55E-01
8,55E-01
8,55E-01
8,55E-01
8,55E-01
8,56E-01
8,56E-01
8,56E-01
8,57E-01
8,57E-01
8,57E-01
8,57E-01
8,57E-01
8,57E-01
8,58E-01
8,58E-01
8,58E-01
8,59E-01
8,59E-01
8,59E-01
8,59E-01
8,59E-01
8,59E-01
8,59E-01
8,59E-01
8,59E-01
8,59E-01
8,60E-01
8,61E-01

8,62E-01
8,62E-01
8,62E-01
8,62E-01
8,62E-01
8,62E-01
8,62E-01
8,63E-01
8,63E-01
8,63E-01
8,63E-01
8,63E-01
8,63E-01
8,63E-01
8,63E-01
8,63E-01
8,64E-01
8,64E-01
8,64E-01
8,64E-01
8,64E-01
8,64E-01
8,65E-01
8,65E-01
8,65E-01
8,66E-01
8,66E-01
8,66E-01
8,67E-01
8,68E-01
8,68E-01
8,68E-01
8,68E-01
8,68E-01
8,68E-01
8,68E-01
8,69E-01
8,69E-01
8,69E-01
8,69E-01
8,69E-01
8,70E-01
8,70E-01
8,70E-01
8,70E-01
8,70E-01

8,71E-01
8,71E-01
8,71E-01
8,71E-01
8,71E-01
8,71E-01
8,72E-01
8,72E-01
8,72E-01
8,72E-01
8,72E-01
8,73E-01
8,74E-01
8,74E-01
8,74E-01
8,75E-01
8,75E-01
8,76E-01
8,76E-01
8,76E-01
8,76E-01
8,76E-01
8,76E-01
8,77E-01
8,77E-01
8,77E-01
8,77E-01
8,77E-01
8,77E-01
8,78E-01
8,78E-01
8,78E-01
8,78E-01
8,78E-01
8,78E-01
8,78E-01
8,79E-01
8,79E-01
8,79E-01
8,80E-01
8,80E-01
8,80E-01
8,80E-01
8,80E-01
8,80E-01
8,81E-01

8,81E-01
8,81E-01
8,81E-01
8,81E-01
8,82E-01
8,82E-01
8,82E-01
8,82E-01
8,82E-01
8,83E-01
8,83E-01
8,83E-01
8,83E-01
8,84E-01
8,85E-01
8,85E-01
8,85E-01
8,86E-01
8,86E-01
8,86E-01
8,86E-01
8,86E-01
8,86E-01
8,86E-01
8,86E-01
8,87E-01
8,87E-01
8,87E-01
8,87E-01
8,87E-01
8,87E-01
8,87E-01
8,87E-01
8,87E-01
8,87E-01
8,88E-01
8,88E-01
8,88E-01
8,88E-01
8,88E-01
8,88E-01
8,89E-01
8,89E-01
8,89E-01
8,89E-01
8,89E-01

8,90E-01
8,90E-01
8,91E-01
8,91E-01
8,91E-01
8,92E-01
8,92E-01
8,92E-01
8,92E-01
8,93E-01
8,93E-01
8,93E-01
8,94E-01
8,94E-01
8,94E-01
8,95E-01
8,95E-01
8,95E-01
8,95E-01
8,95E-01
8,95E-01
8,95E-01
8,95E-01
8,96E-01
8,96E-01
8,97E-01
8,97E-01
8,97E-01
8,97E-01
8,97E-01
8,98E-01
8,98E-01
8,98E-01
8,98E-01
8,98E-01
8,98E-01
8,98E-01
8,99E-01
8,99E-01
8,99E-01
8,99E-01
8,99E-01
8,99E-01
8,99E-01
8,99E-01
8,99E-01

8,99E-01
9,00E-01
9,00E-01
9,00E-01
9,00E-01
9,00E-01
9,00E-01
9,01E-01
9,01E-01
9,01E-01
9,01E-01
9,01E-01
9,01E-01
9,01E-01
9,01E-01
9,01E-01
9,01E-01
9,02E-01
9,02E-01
9,02E-01
9,02E-01
9,02E-01
9,02E-01
9,02E-01
9,03E-01
9,03E-01
9,03E-01
9,03E-01
9,03E-01
9,03E-01
9,03E-01
9,03E-01
9,03E-01
9,04E-01
9,04E-01
9,04E-01
9,05E-01
9,05E-01
9,05E-01
9,05E-01
9,06E-01
9,06E-01
9,06E-01
9,06E-01
9,06E-01
9,06E-01

9,06E-01
9,07E-01
9,07E-01
9,07E-01
9,07E-01
9,07E-01
9,07E-01
9,07E-01
9,08E-01
9,08E-01
9,08E-01
9,08E-01
9,08E-01
9,08E-01
9,08E-01
9,08E-01
9,09E-01
9,09E-01
9,09E-01
9,09E-01
9,10E-01
9,10E-01
9,10E-01
9,10E-01
9,10E-01
9,11E-01
9,11E-01
9,11E-01
9,11E-01
9,11E-01
9,11E-01
9,12E-01
9,12E-01
9,12E-01
9,12E-01
9,12E-01
9,12E-01
9,12E-01
9,12E-01
9,13E-01
9,13E-01
9,13E-01
9,13E-01
9,13E-01
9,13E-01
9,13E-01

9,13E-01
9,13E-01
9,13E-01
9,13E-01
9,13E-01
9,13E-01
9,14E-01
9,14E-01
9,15E-01
9,15E-01
9,15E-01
9,15E-01
9,15E-01
9,15E-01
9,15E-01
9,15E-01
9,16E-01
9,17E-01
9,17E-01
9,17E-01
9,17E-01
9,17E-01
9,18E-01
9,18E-01
9,18E-01
9,18E-01
9,18E-01
9,19E-01
9,19E-01
9,19E-01
9,19E-01
9,19E-01
9,20E-01
9,20E-01
9,21E-01
9,21E-01
9,21E-01
9,21E-01
9,21E-01
9,22E-01
9,22E-01
9,23E-01
9,23E-01
9,24E-01
9,25E-01
9,25E-01

9,25E-01
9,25E-01
9,25E-01
9,25E-01
9,25E-01
9,26E-01
9,26E-01
9,26E-01
9,26E-01
9,26E-01
9,26E-01
9,26E-01
9,26E-01
9,27E-01
9,27E-01
9,27E-01
9,28E-01
9,28E-01
9,28E-01
9,28E-01
9,28E-01
9,29E-01
9,29E-01
9,29E-01
9,29E-01
9,29E-01
9,30E-01
9,30E-01
9,30E-01
9,30E-01
9,30E-01
9,31E-01
9,31E-01
9,31E-01
9,31E-01
9,31E-01
9,31E-01
9,31E-01
9,31E-01
9,32E-01
9,32E-01
9,32E-01
9,32E-01
9,32E-01
9,32E-01
9,33E-01

9,33E-01
9,33E-01
9,33E-01
9,33E-01
9,33E-01
9,33E-01
9,34E-01
9,34E-01
9,34E-01
9,34E-01
9,34E-01
9,34E-01
9,35E-01
9,35E-01
9,35E-01
9,35E-01
9,35E-01
9,35E-01
9,36E-01
9,36E-01
9,36E-01
9,36E-01
9,36E-01
9,36E-01
9,37E-01
9,37E-01
9,38E-01
9,38E-01
9,38E-01
9,38E-01
9,38E-01
9,38E-01
9,38E-01
9,38E-01
9,38E-01
9,38E-01
9,39E-01
9,39E-01
9,39E-01
9,39E-01
9,39E-01
9,39E-01
9,39E-01
9,39E-01
9,39E-01
9,39E-01

9,40E-01
9,40E-01
9,40E-01
9,40E-01
9,40E-01
9,41E-01
9,42E-01
9,42E-01
9,43E-01
9,43E-01
9,43E-01
9,43E-01
9,44E-01
9,44E-01
9,44E-01
9,44E-01
9,44E-01
9,44E-01
9,44E-01
9,44E-01
9,46E-01
9,46E-01
9,46E-01
9,46E-01
9,46E-01
9,47E-01
9,47E-01
9,47E-01
9,47E-01
9,48E-01
9,48E-01
9,48E-01
9,48E-01
9,48E-01
9,48E-01
9,48E-01
9,48E-01
9,48E-01
9,49E-01
9,49E-01
9,49E-01
9,49E-01
9,49E-01
9,50E-01
9,50E-01
9,50E-01

9,51E-01
9,53E-01
9,53E-01
9,53E-01
9,53E-01
9,53E-01
9,53E-01
9,53E-01
9,54E-01
9,54E-01
9,54E-01
9,54E-01
9,54E-01
9,54E-01
9,54E-01
9,55E-01
9,55E-01
9,55E-01
9,55E-01
9,55E-01
9,55E-01
9,55E-01
9,55E-01
9,56E-01
9,56E-01
9,57E-01
9,57E-01
9,57E-01
9,57E-01
9,57E-01
9,57E-01
9,57E-01
9,58E-01
9,58E-01
9,58E-01
9,59E-01
9,59E-01
9,59E-01
9,59E-01
9,59E-01
9,59E-01
9,59E-01
9,59E-01
9,60E-01
9,60E-01
9,60E-01

9,60E-01
9,60E-01
9,61E-01
9,61E-01
9,61E-01
9,61E-01
9,61E-01
9,61E-01
9,61E-01
9,61E-01
9,61E-01
9,62E-01
9,62E-01
9,62E-01
9,63E-01
9,63E-01
9,63E-01
9,63E-01
9,63E-01
9,63E-01
9,63E-01
9,63E-01
9,64E-01
9,64E-01
9,64E-01
9,64E-01
9,64E-01
9,65E-01
9,65E-01
9,65E-01
9,66E-01
9,66E-01
9,66E-01
9,66E-01
9,67E-01
9,67E-01
9,67E-01
9,67E-01
9,67E-01
9,67E-01
9,67E-01
9,67E-01
9,68E-01
9,68E-01
9,68E-01
9,68E-01

9,68E-01
9,68E-01
9,68E-01
9,68E-01
9,68E-01
9,69E-01
9,69E-01
9,69E-01
9,69E-01
9,70E-01
9,70E-01
9,71E-01
9,71E-01
9,71E-01
9,71E-01
9,71E-01
9,71E-01
9,71E-01
9,71E-01
9,71E-01
9,73E-01
9,73E-01
9,74E-01
9,74E-01
9,75E-01
9,75E-01
9,75E-01
9,75E-01
9,75E-01
9,75E-01
9,76E-01
9,76E-01
9,77E-01
9,77E-01
9,77E-01
9,77E-01
9,77E-01
9,78E-01
9,78E-01
9,78E-01
9,78E-01
9,79E-01
9,79E-01
9,80E-01
9,80E-01
9,80E-01

9,80E-01
9,81E-01
9,81E-01
9,81E-01
9,81E-01
9,81E-01
9,81E-01
9,81E-01
9,82E-01
9,82E-01
9,83E-01
9,83E-01
9,83E-01
9,83E-01
9,83E-01
9,83E-01
9,83E-01
9,84E-01
9,84E-01
9,84E-01
9,85E-01
9,85E-01
9,86E-01
9,86E-01
9,86E-01
9,87E-01
9,87E-01
9,87E-01
9,87E-01
9,87E-01
9,87E-01
9,87E-01
9,87E-01
9,88E-01
9,88E-01
9,88E-01
9,89E-01
9,90E-01
9,91E-01
9,91E-01
9,91E-01
9,91E-01
9,91E-01
9,91E-01
9,91E-01
9,92E-01

9,92E-01
9,92E-01
9,92E-01
9,92E-01
9,93E-01
9,93E-01
9,93E-01
9,93E-01
9,93E-01
9,93E-01
9,93E-01
9,93E-01
9,93E-01
9,93E-01
9,93E-01
9,93E-01
9,94E-01
9,94E-01
9,94E-01
9,94E-01
9,94E-01
9,94E-01
9,94E-01
9,94E-01
9,95E-01
9,95E-01
9,96E-01
9,96E-01
9,96E-01
9,96E-01
9,96E-01
9,96E-01
9,97E-01
9,97E-01
9,97E-01
9,97E-01
9,97E-01
9,98E-01
9,98E-01
9,98E-01
9,98E-01
9,98E-01
9,98E-01
9,99E-01
9,99E-01
9,99E-01

| |
|---|
| 9,99E-01 |
| 9,99E-01 |
| 9,99E-01 |
| 9,99E-01 |
| 9,99E-01 |
| 1,00E+00 |
| 1,00E+00 |
| 1,00E+00 |
| 1,00E+00 |
| 1,00E+00 |
| 1,00E+00 |
| 1,00E+00 |
