## Supplementary material for "Longitudinal Assessment of DNA Repair Signature Trajectory in Prodromal versus Established Parkinson’s Disease": Table S3-S8

| | |
|---|---|
| (green) | Top 10 in M12 + M24 + M36 |
| (blue) | Top 10 in M12 + M24 |
| (orange) | Top 10 in M24 + M36 |
| (pink) | Top 10 in M12 + M36 |
| (grey) | Top 10 in one timpoint |

Table S3: mtDNA_rep rankings control vs prodromal PD

| Gene_Name | M12 | M24 | M36 |
|---|---|---|---|
| ALKBH1 | 12 | 6 | 19 |
| APEX1 | 21 | 15 | 25 |
| BRCA1 | 6 | 13 | 7 |
| CRY1 | 3 | 19 | 22 |
| DNA2 | 8 | 0 | 12 |
| DUT | 25 | 26 | 8 |
| ERCC2 | 20 | 10 | 24 |
| ERCC6 | 4 | 7 | 3 |
| LIG3 | 1 | 14 | 11 |
| MGMT | 18 | 23 | 14 |
| MPG | 5 | 11 | 15 |
| MUTYH | 17 | 9 | 5 |
| NEIL1 | 10 | 16 | 6 |
| NEIL2 | 0 | 2 | 17 |
| NTHL1 | 2 | 3 | 16 |
| OGG1 | 11 | 4 | 21 |
| PARK7 | 19 | 20 | 4 |
| PNKP | 22 | 24 | 2 |
| POLB | 24 | 12 | 0 |
| POLG | 23 | 18 | 9 |
| POLQ | 14 | 1 | 20 |
| PRIMPOL | 9 | 8 | 1 |
| RAD23A | 7 | 5 | 13 |
| RECQL4 | 15 | 17 | 26 |
| TP53BP1 | 13 | 22 | 10 |
| UNG | 16 | 21 | 23 |
| YBX1 | 26 | 25 | 18 |

| | |
|---|---|
| (green) | Top 10 in M12 + M24 + M36 |
| (blue) | Top 10 in M12 + M24 |
| (orange) | Top 10 in M24 + M36 |
| (pink) | Top 10 in M12 + M36 |
| (grey) | Top 10 in one timpoint |

Table S4: mtDNA_rep rankings prodromal PD vs established PD

| Gene_Name | M12 | M24 | M36 |
|---|---|---|---|
| ALKBH1 | 5 | 12 | 13 |

| | | | |
|---|---|---|---|
| APEX1 | 20 | 8 | 14 |
| BRCA1 | 16 | 23 | 2 |
| CRY1 | 14 | 4 | 22 |
| DNA2 | 12 | 0 | 4 |
| DUT | 22 | 22 | 8 |
| ERCC2 | 7 | 6 | 19 |
| ERCC6 | 4 | 3 | 5 |
| LIG3 | 2 | 17 | 16 |
| MGMT | 19 | 14 | 6 |
| MPG | 3 | 26 | 17 |
| MUTYH | 17 | 5 | 11 |
| NEIL1 | 15 | 11 | 9 |
| NEIL2 | 0 | 1 | 3 |
| NTHL1 | 1 | 2 | 26 |
| OGG1 | 8 | 13 | 20 |
| PARK7 | 13 | 20 | 7 |
| PNKP | 18 | 25 | 18 |
| POLB | 25 | 19 | 10 |
| POLG | 23 | 18 | 23 |
| POLQ | 21 | 7 | 21 |
| PRIMPOL | 10 | 16 | 0 |
| RAD23A | 6 | 10 | 1 |
| RECQL4 | 11 | 9 | 25 |
| TP53BP1 | 26 | 15 | 12 |
| UNG | 9 | 24 | 15 |
| YBX1 | 24 | 21 | 24 |

| Colour | Legend |
|---|---|
| Green | Top 20 in M12 + M24 + M36 |
| Blue | Top 20 in M12 + M24 |
| Orange | Top 20 in M24 + M36 |
| Pink | Top 20 in M12 + M36 |
| Grey | Top 20 in one timpoint |

Table S5: nDNA_rep rankings healthy vs prodromal PD

| Gene_Name | M12 | M24 | M36 |
|---|---|---|---|
| APEX1 | 14 | 8 | 48 |
| APEX2 | 51 | 90 | 92 |
| APLF | 55 | 74 | 91 |
| APTX | 8 | 10 | 63 |
| BLM | 76 | 53 | 46 |
| CETN2 | 29 | 41 | 72 |
| DCLRE1A | 63 | 85 | 80 |
| DCLRE1C | 30 | 22 | 13 |
| DDB1 | 77 | 47 | 29 |
| DDB2 | 31 | 58 | 60 |
| DTX3L | 27 | 83 | 7 |
| ERCC1 | 67 | 75 | 79 |
| ERCC2 | 61 | 94 | 78 |

| | | | |
|---|---|---|---|
| ERCC3 | 50 | 30 | 23 |
| ERCC4 | 38 | 15 | 56 |
| ERCC5 | 69 | 31 | 19 |
| ERCC6 | 12 | 0 | 27 |
| ERCC8 | 25 | 84 | 25 |
| FAAP20 | 18 | 36 | 77 |
| FEN1 | 73 | 72 | 86 |
| GTF2H1 | 13 | 13 | 49 |
| GTF2H2 | 86 | 45 | 37 |
| GTF2H3 | 40 | 50 | 87 |
| GTF2H4 | 35 | 66 | 40 |
| GTF2H5 | 59 | 65 | 65 |
| H2AX | 2 | 11 | 26 |
| HPF1 | 60 | 91 | 90 |
| LIG1 | 68 | 21 | 93 |
| LIG3 | 78 | 68 | 76 |
| LIG4 | 88 | 87 | 61 |
| MAD2L2 | 4 | 1 | 18 |
| MBD4 | 66 | 12 | 41 |
| MCMDC2 | 90 | 80 | 94 |
| MMS19 | 24 | 18 | 22 |
| MPG | 89 | 71 | 50 |
| MRE11 | 57 | 25 | 83 |
| MUTYH | 84 | 46 | 51 |
| NEIL1 | 39 | 89 | 14 |
| NEIL2 | 11 | 9 | 31 |
| NEIL3 | 28 | 57 | 9 |
| NHEJ1 | 34 | 69 | 57 |
| NTHL1 | 72 | 63 | 38 |
| OGG1 | 41 | 86 | 88 |
| PARP1 | 74 | 77 | 47 |
| PARP2 | 92 | 40 | 21 |
| PARP3 | 53 | 82 | 95 |
| PARP9 | 95 | 93 | 69 |
| PNKP | 79 | 59 | 34 |
| POLA1 | 17 | 34 | 73 |
| POLB | 20 | 17 | 0 |
| POLD1 | 15 | 35 | 66 |
| POLD2 | 81 | 95 | 89 |
| POLD3 | 70 | 81 | 12 |
| POLD4 | 3 | 2 | 2 |
| POLE | 36 | 3 | 36 |
| POLE2 | 19 | 73 | 16 |
| POLE3 | 0 | 5 | 10 |
| POLE4 | 82 | 14 | 17 |
| POLH | 5 | 29 | 28 |
| POLI | 80 | 88 | 3 |
| POLK | 16 | 54 | 20 |

| | | | |
|---|---|---|---|
| POLL | 42 | 24 | 85 |
| POLM | 56 | 43 | 53 |
| POLN | 48 | 64 | 75 |
| POLQ | 43 | 61 | 84 |
| PRIMPOL | 46 | 62 | 35 |
| PRKDC | 23 | 4 | 4 |
| RAD1 | 44 | 56 | 6 |
| RAD23A | 62 | 33 | 67 |
| RAD23B | 85 | 42 | 59 |
| RAD50 | 54 | 7 | 42 |
| RAD9A | 64 | 76 | 44 |
| RBX1 | 75 | 37 | 1 |
| REV1 | 6 | 48 | 24 |
| REV3L | 58 | 6 | 5 |
| RFC4 | 33 | 79 | 52 |
| RIF1 | 22 | 19 | 30 |
| RNF168 | 21 | 20 | 54 |
| RNF8 | 49 | 51 | 82 |
| SHLD1 | 91 | 55 | 15 |
| SHLD2 | 32 | 60 | 64 |
| SHLD3 | 37 | 39 | 45 |
| SLX4 | 52 | 92 | 71 |
| SMUG1 | 9 | 28 | 11 |
| TDG | 65 | 23 | 70 |
| TDP1 | 71 | 26 | 55 |
| TP53BP1 | 7 | 27 | 39 |
| UNG | 83 | 32 | 62 |
| UVSSA | 87 | 52 | 68 |
| XAB2 | 93 | 70 | 58 |
| XPA | 1 | 38 | 32 |
| XPC | 10 | 16 | 8 |
| XRCC1 | 45 | 67 | 81 |
| XRCC4 | 94 | 49 | 74 |
| XRCC5 | 47 | 78 | 33 |
| XRCC6 | 26 | 44 | 43 |

| | |
|---|---|
| (green) | Top 20 in M12 + M24 + M36 |
| (blue) | Top 20 in M12 + M24 |
| (orange) | Top 20 in M24 + M36 |
| (pink) | Top 20 in M12 + M36 |
| (grey) | Top 20 in one timpoint |

Table S6: nDNA_rep rankings prodromal vs established PD

| Gene_Name | M12 | M24 | M36 |
|---|---|---|---|
| APEX1 | 4 | 3 | 60 |
| APEX2 | 83 | 88 | 38 |
| APLF | 40 | 87 | 30 |
| APTX | 27 | 13 | 32 |

| | | | |
|---|---|---|---|
| BLM | 68 | 73 | 27 |
| CETN2 | 17 | 62 | 73 |
| DCLRE1A | 73 | 71 | 90 |
| DCLRE1C | 48 | 41 | 12 |
| DDB1 | 10 | 72 | 43 |
| DDB2 | 29 | 65 | 53 |
| DTX3L | 90 | 6 | 39 |
| ERCC1 | 60 | 44 | 18 |
| ERCC2 | 84 | 70 | 79 |
| ERCC3 | 76 | 37 | 44 |
| ERCC4 | 30 | 29 | 64 |
| ERCC5 | 67 | 27 | 37 |
| ERCC6 | 18 | 8 | 10 |
| ERCC8 | 46 | 30 | 45 |
| FAAP20 | 14 | 81 | 75 |
| FEN1 | 53 | 47 | 91 |
| GTF2H1 | 20 | 90 | 93 |
| GTF2H2 | 87 | 53 | 26 |
| GTF2H3 | 43 | 26 | 61 |
| GTF2H4 | 70 | 64 | 47 |
| GTF2H5 | 72 | 93 | 78 |
| H2AFX | 3 | 18 | 72 |
| HPF1 | 58 | 54 | 87 |
| LIG1 | 51 | 61 | 86 |
| LIG3 | 34 | 91 | 49 |
| LIG4 | 92 | 76 | 54 |
| MAD2L2 | 7 | 1 | 34 |
| MBD4 | 54 | 4 | 9 |
| MCMDC2 | 71 | 85 | 76 |
| MMS19 | 13 | 2 | 80 |
| MPG | 47 | 83 | 67 |
| MRE11 | 65 | 17 | 28 |
| MUTYH | 80 | 24 | 51 |
| NEIL1 | 62 | 67 | 25 |
| NEIL2 | 9 | 45 | 22 |
| NEIL3 | 24 | 23 | 17 |
| NHEJ1 | 22 | 32 | 29 |
| NTHL1 | 8 | 42 | 63 |
| OGG1 | 66 | 84 | 95 |
| PARP1 | 95 | 49 | 62 |
| PARP2 | 93 | 46 | 41 |
| PARP3 | 81 | 79 | 81 |
| PARP9 | 88 | 33 | 55 |
| PNKP | 57 | 59 | 14 |
| POLA1 | 16 | 12 | 66 |
| POLB | 37 | 38 | 3 |
| POLD1 | 12 | 20 | 23 |
| POLD2 | 89 | 63 | 65 |

| | | | |
|---|---|---|---|
| POLD3 | 32 | 94 | 40 |
| POLD4 | 0 | 9 | 1 |
| POLE | 23 | 34 | 21 |
| POLE2 | 38 | 50 | 50 |
| POLE3 | 1 | 0 | 4 |
| POLE4 | 69 | 66 | 11 |
| POLH | 5 | 60 | 84 |
| POLI | 79 | 75 | 94 |
| POLK | 56 | 58 | 33 |
| POLL | 61 | 95 | 2 |
| POLM | 49 | 69 | 5 |
| POLN | 33 | 92 | 69 |
| POLQ | 41 | 68 | 71 |
| PRIMPOL | 78 | 39 | 7 |
| PRKDC | 25 | 19 | 13 |
| RAD1 | 77 | 35 | 19 |
| RAD23A | 64 | 15 | 92 |
| RAD23B | 39 | 7 | 8 |
| RAD50 | 35 | 16 | 24 |
| RAD9A | 28 | 55 | 57 |
| RBX1 | 55 | 56 | 6 |
| REV1 | 6 | 10 | 35 |
| REV3L | 94 | 11 | 15 |
| RFC4 | 19 | 78 | 58 |
| RIF1 | 11 | 14 | 20 |
| RNF168 | 31 | 28 | 83 |
| RNF8 | 82 | 89 | 89 |
| SHLD1 | 52 | 5 | 36 |
| SHLD2 | 36 | 77 | 70 |
| SHLD3 | 26 | 31 | 56 |
| SLX4 | 91 | 82 | 68 |
| SMUG1 | 42 | 48 | 0 |
| TDG | 86 | 22 | 16 |
| TDP1 | 59 | 36 | 77 |
| TP53BP1 | 85 | 74 | 85 |
| UNG | 74 | 43 | 46 |
| UVSSA | 44 | 40 | 59 |
| XAB2 | 75 | 51 | 42 |
| XPA | 2 | 25 | 48 |
| XPC | 63 | 80 | 88 |
| XRCC1 | 50 | 57 | 52 |
| XRCC4 | 15 | 52 | 74 |
| XRCC5 | 45 | 86 | 31 |
| XRCC6 | 21 | 21 | 82 |

| | |
|---|---|
| (green) | Top 20 in M12 + M24 + M36 |
| (blue) | Top 20 in M12 + M24 |
| (orange) | Top 20 in M24 + M36 |

| | |
|---|---|
| (pink) | Top 20 in M12 + M36 |
| (grey) | Top 20 in one timpoint |

Table S7: ISR rankings healthy vs prodromal PD

| Gene_Name | M12 | M24 | M36 |
|---|---|---|---|
| AARS2 | 61 | 52 | 55 |
| ACOT11 | 46 | 46 | 64 |
| APOE | 40 | 48 | 98 |
| ASNS | 60 | 83 | 49 |
| ATF2 | 4 | 17 | 8 |
| ATF3 | 92 | 75 | 75 |
| ATF4 | 9 | 16 | 12 |
| ATF5 | 44 | 33 | 81 |
| ATF6 | 15 | 2 | 19 |
| ATG5 | 39 | 49 | 37 |
| BBC3 | 33 | 27 | 45 |
| BGLAP | 127 | 112 | 114 |
| CA9 | 86 | 100 | 125 |
| CARS1 | 3 | 14 | 24 |
| CARS2 | 2 | 9 | 7 |
| CASP12 | 119 | 109 | 92 |
| CCL2 | 82 | 102 | 110 |
| CCNA2 | 116 | 123 | 71 |
| CCND1 | 53 | 55 | 87 |
| CDC42 | 35 | 36 | 3 |
| CEBPA | 25 | 8 | 25 |
| CEBPB | 0 | 4 | 2 |
| CEBPE | 43 | 77 | 79 |
| CHAC1 | 103 | 106 | 123 |
| CREB1 | 19 | 15 | 27 |
| CREBBP | 47 | 114 | 15 |
| CSF1R | 68 | 29 | 109 |
| CTNNB1 | 30 | 7 | 26 |
| CXCL3 | 94 | 126 | 106 |
| CXCL8 | 5 | 11 | 17 |
| CYP2E1 | 52 | 62 | 78 |
| DARS2 | 85 | 50 | 115 |
| DDIT3 | 57 | 38 | 42 |
| DDIT4 | 83 | 6 | 32 |
| DDR2 | 51 | 60 | 70 |
| DISC1 | 13 | 115 | 23 |
| DKK1 | 98 | 89 | 95 |
| DNAJB9 | 29 | 66 | 83 |
| DRD2 | 63 | 64 | 80 |
| EDN1 | 115 | 111 | 113 |
| EIF2AK2 | 110 | 68 | 22 |
| ELANE | 121 | 54 | 65 |
| ERBB2 | 22 | 22 | 47 |

| | | | |
|---|---|---|---|
| ERVW-1 | 8 | 51 | 11 |
| F7 | 80 | 91 | 119 |
| FGF19 | 99 | 107 | 122 |
| FGF2 | 108 | 121 | 120 |
| FGF21 | 120 | 118 | 108 |
| GNB1L | 50 | 67 | 56 |
| GPT2 | 104 | 92 | 107 |
| GRIN2A | 10 | 35 | 85 |
| HLA-DRB1 | 26 | 58 | 5 |
| HMOX1 | 49 | 31 | 59 |
| HRK | 55 | 117 | 66 |
| HSPA5 | 100 | 19 | 127 |
| IARS1 | 76 | 12 | 18 |
| IGFBP1 | 90 | 97 | 126 |
| IHH | 109 | 105 | 117 |
| IL18 | 71 | 104 | 97 |
| IL1B | 117 | 18 | 43 |
| IL23A | 87 | 59 | 73 |
| IL6 | 78 | 98 | 112 |
| INHBE | 122 | 103 | 99 |
| INS | 88 | 99 | 102 |
| IRF7 | 97 | 127 | 9 |
| ITIH3 | 59 | 74 | 61 |
| KAT2B | 31 | 23 | 0 |
| KDM6B | 11 | 3 | 1 |
| LAMP3 | 69 | 84 | 67 |
| LARS1 | 24 | 80 | 16 |
| LARS2 | 96 | 44 | 57 |
| LHX2 | 77 | 81 | 121 |
| LITAF | 34 | 63 | 6 |
| MAP1LC3B | 21 | 10 | 53 |
| MCL1 | 84 | 65 | 29 |
| MTOR | 89 | 42 | 62 |
| NARS1 | 7 | 39 | 39 |
| NARS2 | 70 | 32 | 84 |
| NDC80 | 42 | 41 | 48 |
| NFE2L2 | 1 | 0 | 13 |
| NRP1 | 45 | 86 | 50 |
| NUPR1 | 38 | 45 | 72 |
| PDGFRA | 48 | 43 | 51 |
| PENK | 56 | 61 | 82 |
| PER2 | 111 | 94 | 68 |
| PLAT | 74 | 71 | 88 |
| PLAU | 113 | 108 | 86 |
| PMAIP1 | 14 | 20 | 41 |
| POLR2C | 20 | 25 | 33 |
| PPARGC1A | 64 | 82 | 94 |
| PPP1R15A | 37 | 56 | 30 |

| | | | |
|---|---|---|---|
| PRKN | 23 | 78 | 58 |
| PSEN1 | 28 | 40 | 10 |
| PTGS2 | 12 | 1 | 21 |
| PTH | 118 | 110 | 111 |
| RPS6KA3 | 16 | 13 | 31 |
| RUNX2 | 58 | 28 | 34 |
| S100A8 | 101 | 125 | 74 |
| S100P | 6 | 21 | 4 |
| SARS2 | 75 | 57 | 89 |
| SCG2 | 125 | 124 | 101 |
| SERPINC1 | 124 | 93 | 116 |
| SIGMAR1 | 79 | 73 | 91 |
| SIRT1 | 114 | 26 | 20 |
| SIRT2 | 17 | 53 | 36 |
| SIRT4 | 102 | 76 | 90 |
| SLC38A2 | 32 | 30 | 14 |
| SLC6A4 | 66 | 69 | 96 |
| SLC7A11 | 105 | 90 | 69 |
| SNCG | 93 | 116 | 124 |
| SP7 | 126 | 119 | 104 |
| SQSTM1 | 18 | 85 | 35 |
| STAT3 | 67 | 47 | 44 |
| TARS2 | 41 | 34 | 46 |
| TH | 72 | 79 | 103 |
| THEG | 65 | 72 | 118 |
| TNC | 91 | 88 | 100 |
| TNF | 123 | 87 | 77 |
| TNFRSF10B | 62 | 5 | 28 |
| TNFRSF11A | 106 | 95 | 105 |
| TNFSF11 | 107 | 122 | 76 |
| TRIB3 | 112 | 120 | 52 |
| TRPV6 | 95 | 96 | 93 |
| USF1 | 36 | 24 | 38 |
| VEGFA | 27 | 70 | 54 |
| VIM | 54 | 101 | 40 |
| WARS2 | 73 | 37 | 63 |
| YARS2 | 81 | 113 | 60 |

| | |
|---|---|
| | Top 20 in M12 + M24 + M36 |
| | Top 20 in M12 + M24 |
| | Top 20 in M24 + M36 |
| | Top 20 in M12 + M36 |
| | Top 20 in one timpoint |

Table S8: ISR rankings prodromal vs established PD

| Gene_Name | M12 | M24 | M36 |
|---|---|---|---|
| AARS2 | 48 | 51 | 52 |

| | | | |
|---|---|---|---|
| ACOT11 | 57 | 64 | 64 |
| APOE | 121 | 38 | 71 |
| ASNS | 11 | 44 | 57 |
| ATF2 | 8 | 36 | 97 |
| ATF3 | 105 | 53 | 88 |
| ATF4 | 26 | 27 | 23 |
| ATF5 | 18 | 23 | 56 |
| ATF6 | 14 | 0 | 10 |
| ATG5 | 120 | 65 | 63 |
| BBC3 | 7 | 3 | 26 |
| BGLAP | 96 | 121 | 126 |
| CA9 | 116 | 114 | 120 |
| CARS1 | 6 | 5 | 25 |
| CARS2 | 39 | 55 | 5 |
| CASP12 | 102 | 120 | 106 |
| CCL2 | 84 | 74 | 65 |
| CCNA2 | 109 | 87 | 105 |
| CCND1 | 20 | 32 | 42 |
| CDC42 | 88 | 34 | 9 |
| CEBPA | 82 | 4 | 73 |
| CEBPB | 16 | 7 | 7 |
| CEBPE | 56 | 102 | 51 |
| CHAC1 | 101 | 113 | 110 |
| CREB1 | 62 | 42 | 28 |
| CREBBP | 72 | 97 | 34 |
| CSF1R | 9 | 16 | 111 |
| CTNNB1 | 60 | 40 | 31 |
| CXCL3 | 124 | 123 | 103 |
| CXCL8 | 17 | 14 | 4 |
| CYP2E1 | 76 | 104 | 75 |
| DARS2 | 87 | 58 | 108 |
| DDIT3 | 27 | 67 | 39 |
| DDIT4 | 50 | 103 | 37 |
| DDR2 | 36 | 52 | 85 |
| DISC1 | 31 | 19 | 15 |
| DKK1 | 107 | 73 | 127 |
| DNAJB9 | 35 | 95 | 59 |
| DRD2 | 63 | 59 | 89 |
| EDN1 | 103 | 117 | 107 |
| EIF2AK2 | 51 | 22 | 17 |
| ELANE | 15 | 45 | 77 |
| ERBB2 | 21 | 50 | 46 |
| ERVW-1 | 37 | 68 | 13 |
| F7 | 83 | 71 | 62 |
| FGF19 | 114 | 93 | 93 |
| FGF2 | 78 | 119 | 109 |
| FGF21 | 106 | 118 | 125 |
| GNB1L | 28 | 89 | 43 |

| | | | |
|---|---|---|---|
| GPT2 | 52 | 109 | 121 |
| GRIN2A | 24 | 10 | 19 |
| HLA-DRB1 | 94 | 35 | 11 |
| HMOX1 | 70 | 43 | 79 |
| HRK | 45 | 127 | 55 |
| HSPA5 | 30 | 25 | 44 |
| IARS1 | 19 | 1 | 16 |
| IGFBP1 | 95 | 90 | 95 |
| IHH | 108 | 101 | 96 |
| IL18 | 61 | 77 | 49 |
| IL1B | 25 | 41 | 68 |
| IL23A | 55 | 69 | 58 |
| IL6 | 79 | 110 | 102 |
| INHBE | 110 | 116 | 101 |
| INS | 73 | 91 | 81 |
| IRF7 | 4 | 57 | 29 |
| ITIH3 | 93 | 60 | 60 |
| KAT2B | 125 | 62 | 1 |
| KDM6B | 59 | 6 | 0 |
| LAMP3 | 75 | 54 | 115 |
| LARS1 | 10 | 84 | 32 |
| LARS2 | 44 | 20 | 40 |
| LHX2 | 92 | 85 | 80 |
| LITAF | 127 | 72 | 18 |
| MAP1LC3B | 47 | 94 | 36 |
| MCL1 | 126 | 81 | 38 |
| MTOR | 33 | 30 | 67 |
| NARS1 | 3 | 26 | 24 |
| NARS2 | 58 | 111 | 53 |
| NDC80 | 49 | 21 | 72 |
| NFE2L2 | 0 | 8 | 2 |
| NRP1 | 64 | 33 | 86 |
| NUPR1 | 32 | 31 | 83 |
| PDGFRA | 23 | 17 | 91 |
| PENK | 97 | 56 | 84 |
| PER2 | 86 | 66 | 119 |
| PLAT | 89 | 80 | 69 |
| PLAU | 85 | 105 | 66 |
| PMAIP1 | 13 | 49 | 90 |
| POLR2C | 2 | 12 | 22 |
| PPARGC1A | 69 | 79 | 98 |
| PPP1R15A | 43 | 18 | 45 |
| PRKN | 112 | 86 | 70 |
| PSEN1 | 5 | 2 | 8 |
| PTGS2 | 1 | 48 | 20 |
| PTH | 100 | 125 | 118 |
| RPS6KA3 | 38 | 9 | 14 |
| RUNX2 | 29 | 82 | 35 |

| | | | |
|---|---|---|---|
| S100A8 | 123 | 122 | 54 |
| S100P | 34 | 39 | 6 |
| SARS2 | 98 | 92 | 50 |
| SCG2 | 111 | 115 | 100 |
| SERPINC1 | 104 | 126 | 124 |
| SIGMAR1 | 67 | 75 | 92 |
| SIRT1 | 40 | 76 | 33 |
| SIRT2 | 42 | 46 | 104 |
| SIRT4 | 71 | 112 | 114 |
| SLC38A2 | 65 | 28 | 21 |
| SLC6A4 | 80 | 88 | 82 |
| SLC7A11 | 113 | 100 | 74 |
| SNCG | 81 | 96 | 99 |
| SP7 | 122 | 124 | 116 |
| SQSTM1 | 54 | 61 | 3 |
| STAT3 | 22 | 11 | 12 |
| TARS2 | 41 | 24 | 87 |
| TH | 91 | 63 | 76 |
| THEG | 99 | 47 | 47 |
| TNC | 118 | 108 | 122 |
| TNF | 77 | 107 | 94 |
| TNFRSF10B | 53 | 15 | 27 |
| TNFRSF11A | 74 | 98 | 113 |
| TNFSF11 | 119 | 83 | 123 |
| TRIB3 | 68 | 78 | 78 |
| TRPV6 | 117 | 99 | 112 |
| USF1 | 46 | 13 | 30 |
| VEGFA | 12 | 29 | 48 |
| VIM | 90 | 106 | 61 |
| WARS2 | 66 | 37 | 41 |
| YARS2 | 115 | 70 | 117 |
